# Small molecule biomarkers predictive of Chagas disease progression

**DOI:** 10.1101/2024.05.13.24307310

**Authors:** Zongyuan Liu, Joseane Godinho, Jarrod A. Laro, Kelly DeToy, Steffany Vucetic, Jordan R. Edens, Jessy Condori, Carolina Duque, Gustavo Durán Saucedo, Manuela Verastegui, Paula Carballo-Jimenez, Brandon N. Mercado-Saavedra, Freddy Tinajeros, Edith S. Málaga-Machaca, Rachel Marcus, Robert H. Gilman, Natalie M. Bowman, Laura-Isobel McCall

**Affiliations:** Department of Chemistry and Biochemistry, University of Oklahoma, Norman, OK, USA; Department of Chemistry & Biochemistry, San Diego State University, San Diego, CA, USA; Department of International Health, Johns Hopkins Bloomberg School of Public Health, Baltimore, MD, USA; Infectious Diseases Research Laboratory, LID, Faculty of Sciences and Engineering, Universidad Peruana Cayetano Heredia, Lima, Peru; Department of Pathology, Johns Hopkins University, Baltimore, MD, USA; IFHAD: Innovation for Health and Development, Prisma NGO, Lima, Peru; FASPA: Faculty of Public Health and Administration, Universidad Peruana Cayetano Heredia, Lima, Peru; Instituto de Investigación de Medicina, Universidad Católica Boliviana San Pablo, Santa Cruz, Bolivia; Medstar Union Memorial Hospital, Baltimore, MD, USA; Division of Infectious Diseases, University of North Carolina at Chapel Hill, Chapel Hill, NC, USA

**Keywords:** Chagas disease, *Trypanosoma cruzi*, disease progression, biomarkers, metabolomics, mass spectrometry

## Abstract

Chagas disease (CD), caused by the protozoan parasite *Trypanosoma cruzi*, affects an estimated 10.5 million people worldwide and remains a leading cause of infectious cardiomyopathy in Latin America. Although infection is lifelong without treatment, only 20–30% of chronically infected individuals progress from an asymptomatic stage to clinically significant cardiac disease, and current antiparasitic therapies are limited by toxicity and variable efficacy. Reliable biomarkers capable of predicting disease progression would improve clinical risk stratification and support therapeutic development. Here, we performed a metabolomics study to identify serum biomarkers predictive of progression from asymptomatic infection to cardiac CD. Untargeted liquid chromatography–mass spectrometry profiling using complementary chromatographic platforms and univariate statistics, machine learning feature selection, and receiver operating characteristic (ROC) analysis were performed to identify candidate biomarkers. These biomarkers were validated by targeted analysis. We identified eight individual metabolite biomarkers and 45 combinatorial panels predictive of progression status, including 10 panels that met sensitivity and specificity criteria defined in the target product profile for CD treatment-response biomarkers. These findings demonstrate that circulating metabolites measured at baseline can predict future cardiac disease progression and provide a foundation for clinically deployable prognostic tools in chronic Chagas disease.

## Introduction

Chagas disease (CD), caused by the protozoan parasite *Trypanosoma cruzi,* is a vector-borne illness and a leading cause of infectious cardiomyopathy worldwide, currently affecting an estimated 10.5 million people with roughly 352,000 new cases annually. Although CD burden remains disproportionately concentrated in Latin America, migration has increased its prevalence in non-endemic regions such as North America and Western Europe, and the age profile of infection has shifted toward older adults, with peak prevalence now observed between 45 and 65 years^1,2^. Clinically, CD follows a complex lifelong course: after an often non-specific or asymptomatic acute phase, individuals enter the chronic phase, which includes a prolonged asymptomatic stage with persistent infection but normal electrocardiographic and cardiac imaging findings (the indeterminate phase), followed in some cases by a symptomatic stage with cardiac, digestive, or mixed organ involvement. Approximately 60-70% of chronically infected individuals remain asymptomatic throughout life, whereas 20-40% progress often decades after infection to end-organ disease, most commonly chronic Chagas cardiomyopathy. This condition, characterized by conduction abnormalities, arrhythmias, apical aneurysms, progressive heart failure, thromboembolism, and sudden death, represents the principal cause of Chagas-related morbidity and mortality. Digestive involvement leads to megaesophagus and megacolon, which cause lower mortality than cardiomyopathy but still cause significant morbidity. Despite the identification of risk factors such as persistent parasitemia, sex, and parasite host genetic interactions, the timing and mechanisms underlying progression remain poorly defined, and no reliable early biomarkers exist to distinguish individuals who will remain clinically stable from those at risk of developing irreversible organ damage, underscoring a major unmet need for prognostic tools^3,4^.

Despite frequent and often severe adverse effects, benznidazole and nifurtimox have remained the only approved antiparasitic therapies for more than five decades and are recommended for acute and indeterminate phases^5,6^. Notably, recent studies such as the BENDITA trial^7^ have shown that substantially shorter benznidazole regimens achieve parasitological clearance comparable to standard 60-day treatments while markedly improving tolerability, reinforcing the feasibility of earlier intervention and optimized treatment strategies^8,9^. During the indeterminate phase of *T. cruzi* infection, treatment recommendations depend strongly on patient age, reproductive status, and the presence of established organ damage. Trypanocidal therapy is strongly recommended for children, immunocompromised patients, and increasingly as a preventive strategy in women of childbearing age to interrupt congenital transmission^10,11^. There is currently no consensus on treatment of asymptomatic adults with regards to the age beyond which treatment is not beneficial. Unlike in Bolivia, Mexican treatment guidelines recommend treatment in asymptomatic patients irrespective of age^12^. Current CDC and Brazilian Society of Cardiology guidelines recommend treating asymptomatic *T. cruzi*-positive individuals 50 years or younger; treatment is optional for patients over 50 years old due to the high risk of side effects^13,14^. In contrast, for adults with established organ damage, particularly advanced Chagas cardiomyopathy, current evidence suggests that trypanocidal therapy does not significantly alter mortality or disease progression, and therefore treatment is generally not recommended in this group. In the BENEFIT clinical trial, no reduction in disease progression or mortality was observed in patients with pre-existing cardiac damage despite PCR negativization^11,15^.

Therapeutic decision-making remains constrained by fundamental limitations as conventional serology cannot serve as a short-term test of cure in the chronic phase, as antibodies persist for decades after successful treatment^5,6,16^. Consequently, surrogate measures of treatment efficacy have relied primarily on anti–*T. cruzi* antibody decline or on qPCR^17^. While antibody decline kinetics show promise as earliest indicators of treatment response, antibody decline does not necessarily predict clinical improvement^18^ and takes at least 6 months of follow-up for the fastest antibody biomarkers^19–25^. The FDA has indicated that monitoring seroreduction may not be sufficient to approve new chemical entities against *T. cruzi* and CD in adults^10^. PCR detection during chronic infection is characterized by low sensitivity (only 30-70% of chronically infected individuals test PCR positive even before treatment) and intermittent positivity, resulting in high false-negative rates^26–29^. However, evidence has shown that parasitological clearance does not reliably predict clinical improvement or prevention of disease progression if treatment is initiated once disease symptoms have emerged, indicating that parasite burden alone does not fully explain chronic organ damage^15,30,31^. These limitations have driven extensive efforts to identify alternative markers of both treatment success and disease progression, including antibodies targeting parasite antigens, host protein fragments, coagulation factors, and immune cell activation profiles^32,20,22,9,23,24,33–35^. However, a critical limitation shared by these approaches is their continued reliance on parasite clearance or surrogate immune activation as proxies for clinical benefit or prognosis, leaving a fundamental gap in the field^36,37^.

Multiple protein and immune biomarkers have been correlated with CD stage, though none yet have proven useful in a clinical context. IFN-γ is essential for the control of *T. cruzi* infection in experimental models. Serum from patients with chronic Chagasic cardiomyopathy had relatively high expression of IFN-γ compared to noncardiac or asymptomatic CD patients^38,39^. A recent study found lower baseline levels of 44 host proteins, predominantly involved in immune activation and chemotaxis, that predicted cardiac functional decline over 10 years in *T. cruzi* seropositive individuals^40^. Other protein biomarkers of CD severity (TNFα, brain natriuretic peptide (BNP), or the MMP-2/MMP-9 ratio, for example), only differentiate between symptomatic and asymptomatic patients or between advanced and early stages of cardiomyopathy. Notably, Clark et al. reported that MMP-2, TIMP-1, and TGF-β were elevated in infected individuals with early ECG changes compared to those with normal ECGs, and explicitly called for longitudinal evaluation of these markers as predictors of progression^41–45^. These biomarkers may not therefore be able to predict disease progression in asymptomatic individuals, where the need is greatest. BNP reflects myocardial wall stress and is elevated in patients with established cardiac structural changes, but its utility for detection of the earliest transitions from asymptomatic infection to initial ECG abnormalities remains unclear^43,46^. These biomarkers are therefore reflections of patients’ current cardiac function rather than predictors of future patient outcomes.

Given the limitations of protein biomarkers, metabolomics offers a complementary approach. Unlike protein markers that primarily reflect immune activation or established cardiac disease, metabolites capture real-time biochemical perturbations in host–pathogen interactions and may detect subclinical changes before structural damage occurs. Serum and urine metabolites have been identified as indicators of *T. cruzi* infection status in human subjects and mouse models^47–50^, and cardiac acylcarnitines and glycerophosphocholines correlate with markers of inflammation and fibrosis in experimental infection^51^. Moreover, the cardiac metabolome predicts infection outcome in acute mouse models^52^, and several metabolite signatures normalize following benznidazole or nifurtimox treatment^50,52,53^. However, despite these advances, studies evaluating metabolites as independent predictors of long-term disease progression in human cohorts remain virtually absent, and no metabolite-based biomarker has yet to be validated for clinical use in this context. To address this need, we analyzed samples from a multiyear cohort of *T. cruzi*–infected individuals from Santa Cruz, Bolivia, comparing asymptomatic individuals who remained stably asymptomatic (non-progressors) with those who subsequently developed cardiac disease (progressors). This analysis identified eight candidate metabolite biomarkers, six of which were elevated in non-progressors and two of which are annotated as PC(30:0) and LPC(14:0). Leveraging the combinatorial nature of liquid chromatography–mass spectrometry, we further identified 45 biomarker panels differentiating progressors from non-progressors, including 10 meeting predefined performance criteria of ≥60% sensitivity and ≥90% specificity. Given the significant adverse effects of antiparasitic drugs and intermittent drug shortages^54^, biomarkers of disease progression could help prioritize treatment and monitoring for individuals most likely to benefit and may also inform the design and analysis of clinical trials for antitrypanosomal therapies.

## Results

### Study participants and clinical classification

The overall goal of our study was to identify metabolite biomarkers that predict disease progression, so we focused on samples from the Johns Hopkins–Universidad Peruana Cayetano Heredia Chagas disease biorepository, collected from a Chagas disease–endemic region in Bolivia at a time when patients were still asymptomatic. A total of 64 *T. cruzi* positive participants were included in this study, across two cohorts (Table 1; Table S1). Baseline banked samples were used for metabolomic profiling.

**Table 1.** Clinical and demographic characteristics of study participants by cohort and clinical group.

| Cohort | Group | N | Female/Male<sup>1</sup> | Age at First Visit<sup>2</sup><br>(mean ± SD) | Follow-up Duration<sup>3</sup><br>(months, mean ± SD) | qPCR Negative<sup>4</sup><br>N (%) | qPCR Positive<br>N (%) | qPCR Not Tested<br>N (%) |
| --- | --- | --- | --- | --- | --- | --- | --- | --- |
| 1 | Stable A (infected) | 42 | 32/10 | 51.9 ± 11.1 | 53.4 ± 25.4 | 4 (9.5%) | 2 (4.8%) | 36 (85.7%) |
| 1 | Progressor from A (infected) | 9 | 5/4 | 57.1 ± 8.3 | 53.4 ± 23.0 | 1 (11.1%) | 0 (0%) | 8 (88.9%) |
| 1 | Stable B (infected) | 1 | 1/0 | 51.0 | 24.0 | - | - | 1 (100%) |
| Cohort | Group | N | Female/Male<sup>5</sup> | Age at First Visit<sup>6</sup><br>(mean ± SD) | Follow-up Duration<sup>7</sup><br>(months, mean ± SD) | qPCR Negative<sup>8</sup><br>N (%) | qPCR Positive<br>N (%) | qPCR Not Tested<br>N (%) |
| 2 | Stable A (infected) | 6 | 3/3 | 64.8 ± 10.7 | 71.0 ± 37.4 | 1 (16.7%) | 0 (0%) | 5 (83.3%) |
| 2 | Progressor from A (infected) | 6 | 2/4 | 66.5 ± 15.3 | 40.7 ± 29.0 | 1 (16.7%) | 1 (16.7%) | 4 (66.7%) |
| 2 | Stable B (infected) | 4 | 1/3 | 54.3 ± 8.5 | 59.3 ± 41.0 | 1 (25%) | 1 (25%) | 2 (50%) |
| 2 | Progressor from B (infected) | 4 | 2/2 | 62.5 ± 13.8 | 50.3 ± 31.6 | 1 (25%) | 1 (25%) | 2 (50%) |

Data are presented as mean ± SD unless otherwise indicated.
SD, standard deviation.
<sup>1</sup>female/male distribution, Fisher's exact test ( $p = 0.4419$ ).
<sup>2</sup> age at first visit, Kruskal–Wallis test ( $p = 0.3615$ ).
<sup>3</sup> duration of follow-up, Kruskal–Wallis test ( $p = 0.2203$ ).
<sup>4</sup> qPCR status, Fisher's exact test ( $p = 1$ ).
<sup>5</sup> female/male distribution, Fisher's exact test ( $p = 0.9357$ ).
<sup>6</sup> age at first visit, Kruskal–Wallis test ( $p = 0.2576$ ).
<sup>7</sup> duration of follow-up, Kruskal–Wallis test ( $p = 0.3764$ ).
<sup>8</sup> qPCR status, Fisher's exact test ( $p = 1$ ).

Cardiac status was assessed using electrocardiography (EKG) and echocardiography, and participants were classified into clinical stages of Chagas cardiomyopathy (Stages A–D) based on established criteria integrating EKG abnormalities and ejection fraction (EF) measurements (Figure 1 and Methods). For the patients with serum samples available for metabolomic analysis and in stage A at baseline, 15 infected individuals progressed to any more advanced cardiac stage and were classified as progressors, while 48 infected individuals remained clinically stable and were classified as non-progressors.

**Figure 1.**
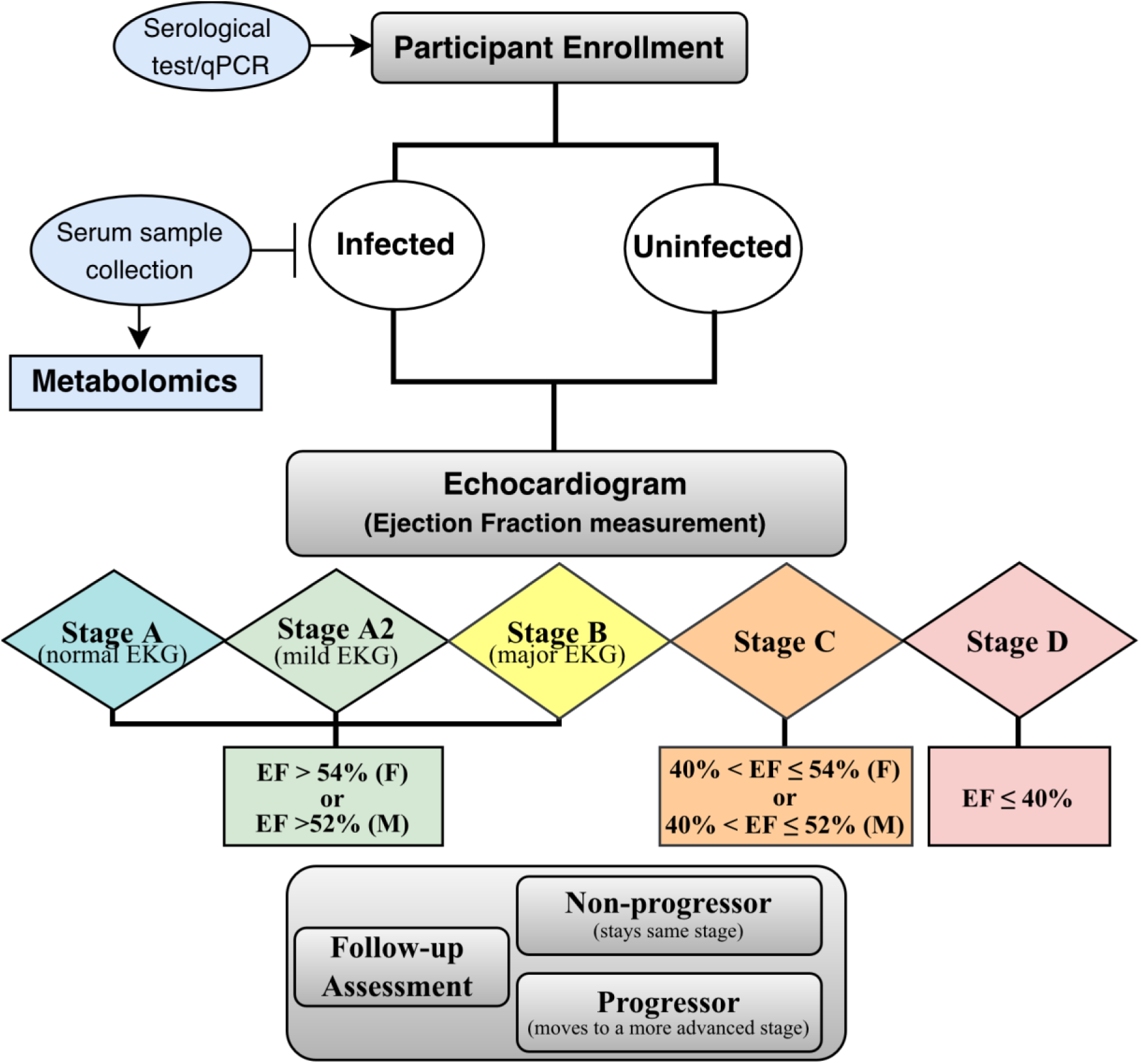
Study workflow for patient classification prior to this metabolomic study. Participants were enrolled and stratified as infected or uninfected based on serological testing and/or quantitative PCR. Serum samples were collected; these are the samples used for metabolomics in this study. Infected individuals underwent electrocardiography (EKG) and echocardiographic evaluation. Based on EF and EKG abnormalities, participants were classified into clinical stages of Chagas cardiomyopathy (Stages A to D). During follow-up, individuals who progressed to a more advanced clinical stage were classified as progressors; those who remained stable were classified as non-progressors. See Methods for details.

Demographic characteristics were comparable across the infected progressors and infected non-progressors from stage A. No significant differences were observed in sex distribution, age at first visit, duration of follow-up, or qPCR status between progressors and non-progressors within either cohort (Table 1). Females represented the majority of participants, consistent with cohort recruitment patterns. qPCR positivity was infrequent and did not differ significantly between groups.

Two independent sample shipments corresponding to Cohorts 1 and 2 were analyzed using identical workflows, including sample extraction protocol, LC gradient, MS acquisition parameters and data processing pipeline. While individual antiparasitic treatment histories and parasite discrete typing units (DTUs) were unavailable, prior studies in this region have reported TcII, TcV, and TcVI^55^. Access to antiparasitic treatment for Chagas disease in Bolivia has improved with the expansion of the National Chagas Network but comprehensive coverage remains limited, especially for adults^56^. As individual treatment histories were unavailable for our cohort, prior benznidazole or nifurtimox therapy is unlikely but cannot be excluded.

### The overall serum metabolome differs between infected progressors and non-progressors

To identify metabolites associated with cardiac disease progression in chronically *Trypanosoma cruzi*–infected individuals, baseline serum samples were analyzed using untargeted HPLC–MS/MS metabolomics and two different chromatography methods (pHILIC and polar C18), and MS data acquisition in positive mode. In the pHILIC dataset, the baseline global metabolome of infected asymptomatic non-progressors from stage A (stable A) differed significantly from infected progressors from stage A (FDR-corrected p = 0.026), infected progressors from stage B (FDR-corrected p = 0.006), and infected non-progressors from stage B (stable B; FDR-corrected p = 0.006). In contrast, infected progressors from stage A, infected progressors from stage B, and infected stable B individuals were not significantly different from each other (Figure 2). These results indicate that the baseline metabolome of stable A individuals is distinct from that of both progressors out of stage A and participants with more advanced cardiac involvement.

**Figure 2.**
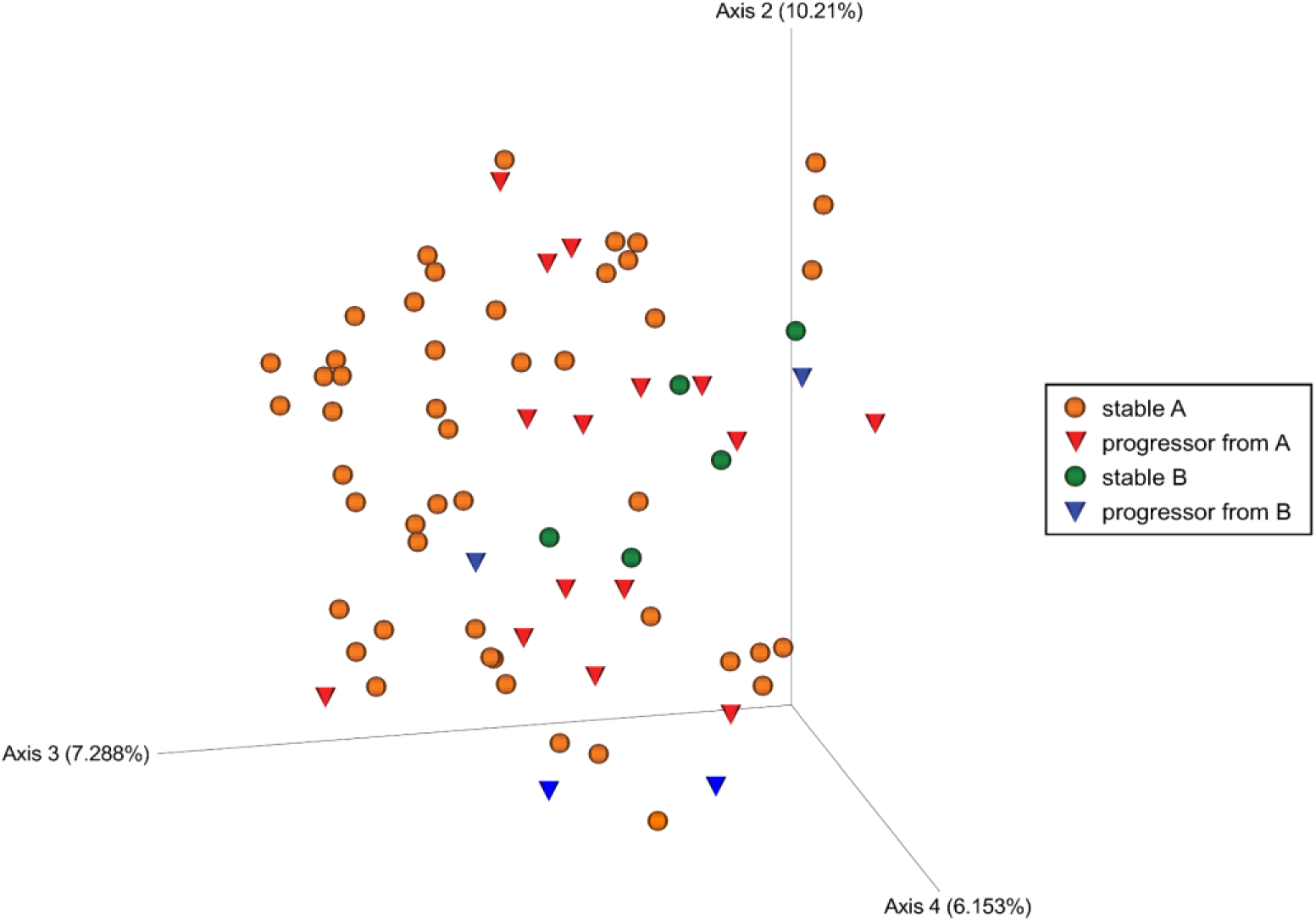
PCoA of Bray-Curtis’s dissimilarities comparing stable (non-progressor) vs. progressor samples analyzed via pHILIC chromatography.

Consistent with this pattern, the polar C18 dataset also showed separation between stable A individuals and groups with more advanced disease. Infected progressors from stage B (FDR-corrected p = 0.015) and infected stable B participants (FDR-corrected p = 0.015) differed significantly from the stable A group, whereas infected progressors from stage A were not significantly different from stable A individuals (Figure S1). Together, these findings suggest that metabolic divergence from the stable A state becomes more pronounced with increasing cardiac involvement and is detected across complementary chromatographic platforms.

We next evaluated potential confounding effects of demographic and study design variables on the overall metabolome. Age and cohort significantly influenced the metabolome in both datasets (age: p = 0.026 and p = 0.002; cohort number: p = 0.001 and p = 0.001 for pHILIC and polar C18 datasets, respectively), while sex influenced only the pHILIC dataset (p = 0.023). Importantly, no interaction was observed between disease progression status and sex, cohort number, duration of follow-up, or age (Table 2), indicating that the progression-associated metabolic differences were independent of these variables.

**Table 2.** Interaction effects of potential confounding variables on disease progression, assessed by PERMANOVA in infected samples. Interactions between patient age and disease progression were evaluated using both a continuous age range (earliest visit: 21-78 years) and discrete age bins. Age bins were defined to ensure approximately equal sample sizes (e.g., bin age range: 55.7-59.6, N = 8). The earliest visit age was categorized into 8 bins (mean N per bin ≈ 9).

| Confounding Variable | HILIC pseudo-F | HILIC R<sup>2</sup> | HILIC p-value | pC18 pseudo-F | pC18 R<sup>2</sup> | pC18 p-value |
| --- | --- | --- | --- | --- | --- | --- |
| Sex | 1.097 | 0.042 | 0.296 | 0.969 | 0.039 | 0.557 |
| Cohort | 1.119 | 0.025 | 0.283 | 1.279 | 0.030 | 0.142 |
| Follow-up duration (months) | 1.092 | 0.042 | 0.302 | 1.055 | 0.043 | 0.386 |
| Age at earliest visit (Continuous) | 1.040 | 0.040 | 0.396 | 1.007 | 0.040 | 0.458 |
| Age at earliest visit (Quantile-binned) | 0.975 | 0.123 | 0.574 | 1.108 | 0.142 | 0.173 |

### Discovery of individual metabolites predictive of progression status

Samples were received in two independent shipments. Cohort 1 (shipment 1) was used for biomarker discovery using untargeted metabolomics, followed by validation in Cohort 2 (shipment 2) using targeted parallel reaction monitoring (PRM). In parallel, discovery was performed independently in Cohort 2 and validated in Cohort 1, enabling bidirectional cross-validation of candidate biomarkers across cohorts (Figure 3A).

**Figure 3.**
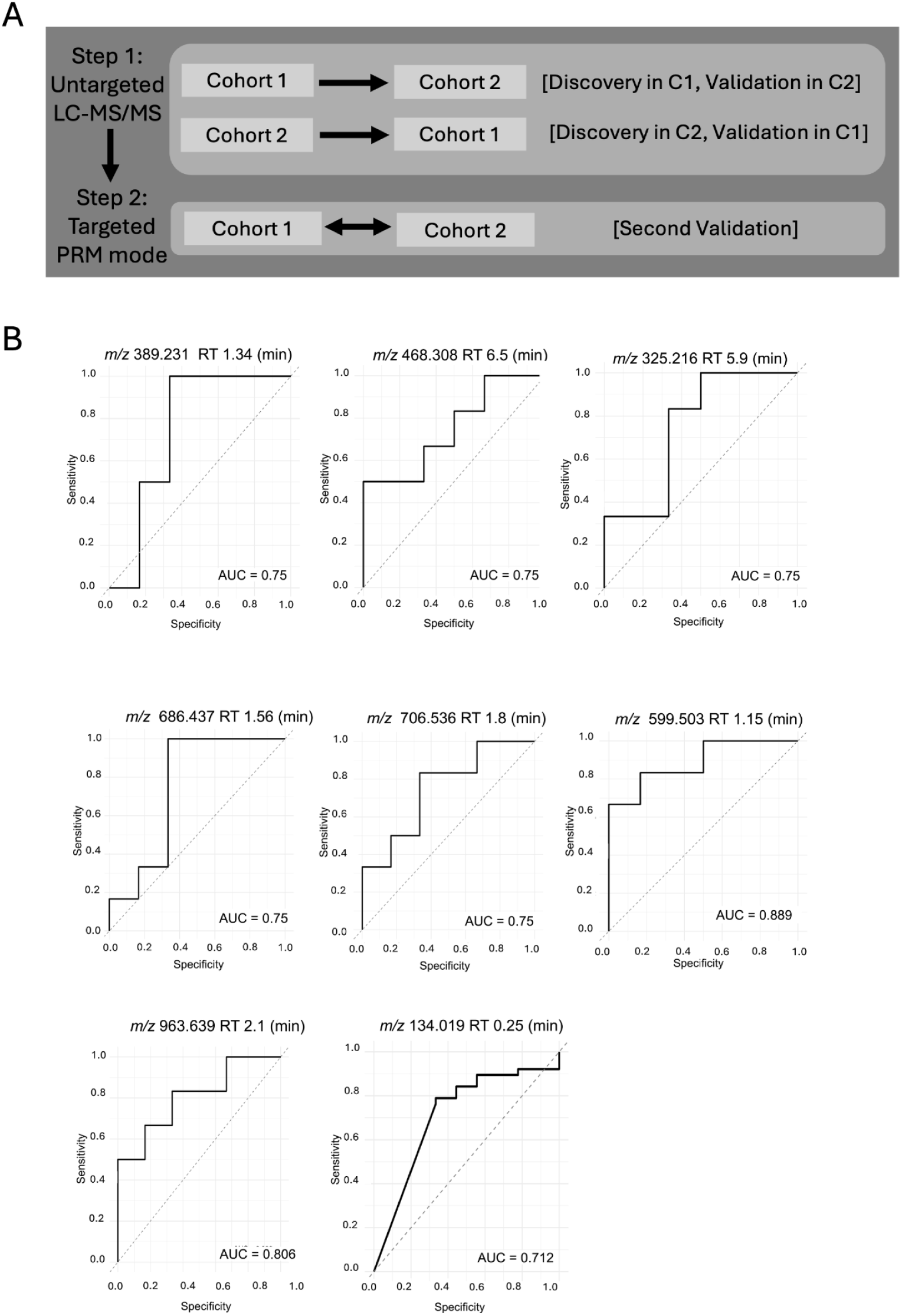
Bidirectional biomarker validation strategy and individual biomarker performance across cohorts. (A) Schematic diagram of the cross-validation design using two independent cohorts (Cohort 1 and Cohort 2). Candidate biomarkers were identified using untargeted LC-MS/MS in one cohort and validated via targeted PRM in the other. The process was repeated in both directions. (B) ROC curves for the single biomarkers validated across cohorts with validation AUC>0.7.

To identify a comprehensive set of candidate metabolites associated with progression status, we combined classical univariate statistics (two-sided Mann–Whitney–Wilcoxon test, uncorrected p < 0.05 for infected progressors from stage A versus infected non-progressors (stable A)) with random forest feature selection (see Methods). This strategy prioritized sensitivity during candidate selection to minimize false negatives at the discovery stage, while independent targeted validation in the reciprocal cohort served to control false positives.

Candidate metabolites meeting these criteria were further evaluated for consistency of effect direction across cohorts, without applying an additional significance threshold. We additionally assessed whether metabolite abundance changes were concordant with levels observed in individuals already presenting with stage B cardiac involvement at enrollment (Cohort 2 only). Receiver operating characteristic (ROC) analysis was then used to refine the target inclusion list, retaining metabolites with area under the curve (AUC) > 0.6 for targeted PRM validation.

Based on prior evidence linking *T. cruzi* infection and Chagas cardiomyopathy severity with perturbations in acylcarnitine and purine metabolism, we additionally included several biologically informed candidates in the targeted inclusion list^57–59^. These included m/z 288.216 (acylcarnitine CAR 8:0), m/z 314.232 (acylcarnitine CAR 10:1), m/z 218.139 (acylcarnitine CAR 3:0), m/z 246.170 (acylcarnitine CAR 5:0), m/z 276.144 (acylcarnitine CAR 5:1;O2), m/z 137.046 (hypoxanthine), m/z 153.041 (xanthine), m/z 169.036 (urate), m/z 268.104 (adenosine, C18), m/z 269.088 (inosine), and m/z 285.083 (xanthosine).

Candidate biomarkers identified in the discovery phase in one cohort were subsequently evaluated in the reciprocal cohort using targeted PRM analysis. Using this bidirectional validation framework, eight metabolites were confirmed as individual predictors distinguishing infected progressors from infected non-progressors, each achieving validation AUC > 0.7 (Figure 3, Table 3). Predictive features were detected in both HILIC and polar C18 datasets, indicating that progression-associated metabolic signatures span multiple chemical classes rather than being restricted to a single metabolite family.

**Table 3.** Performance of individual metabolite biomarkers identified by bidirectional cross-validation across independent cohorts. Metabolites detected by polar C18 (pC18) and HILIC LC–MS are shown with m/z and retention time (RT). Validation performance is reported as sensitivity, specificity, F1 score, and AUC (mean ± SD). Discovery AUC indicates performance in the original feature-selection cohort. Metrics reflect classification of infected progressors from stage A versus non-progressors from stage A at baseline

| LC–MS Method | m/z | RT (min) | Sensitivity | Specificity | F1 Score | Validation AUC | Discovery AUC |
| --- | --- | --- | --- | --- | --- | --- | --- |
| pC18 | 134.019 | 0.25 | 0.83 ± 0.05 | 0.46 ± 0.15 | 0.85 ± 0.01 | 0.712 | 0.64 ± 0.09 |
| pC18 | 325.216 | 5.90 | 0.03 ± 0.11 | 1.00 ± 0.00 | 0.05 ± 0.16 | 0.750 | 0.69 ± 0.09 |
| pC18 | 468.308 | 6.50 | 0.95 ± 0.08 | 0.22 ± 0.08 | 0.69 ± 0.02 | 0.750 | 0.58 ± 0.09 |
| HILIC | 389.231 | 1.34 | 0.52 ± 0.32 | 0.43 ± 0.34 | 0.45 ± 0.22 | 0.750 | 0.58 ± 0.08 |
| HILIC | 599.503 | 1.15 | 0.58 ± 0.44 | 0.68 ± 0.27 | 0.53 ± 0.35 | 0.889 | 0.65 ± 0.09 |
| HILIC | 686.437 | 1.56 | 0.68 ± 0.30 | 0.58 ± 0.14 | 0.62 ± 0.18 | 0.750 | 0.68 ± 0.09 |
| HILIC | 706.536 | 1.80 | 0.48 ± 0.20 | 0.75 ± 0.26 | 0.54 ± 0.06 | 0.750 | 0.68 ± 0.12 |
| HILIC | 963.639 | 2.10 | 0.60 ± 0.49 | 0.40 ± 0.31 | 0.44 ± 0.34 | 0.806 | 0.60 ± 0.11 |

Validation AUC values are shown without SD because values were invariant across validation folds.

None of these biomarkers showed significant correlation with duration of follow-up in the discovery cohort (Spearman *p*>0.05), though the glycerolipid *m/z* 599.503 was positively correlated with the duration of follow-up in the validation cohort (Spearman p=0.006124, rho=0.7381382), suggesting that this biomarker may be more sensitive to confounders. No other candidate biomarkers showed significant correlations in the validation cohort.

### Combinations of metabolite biomarkers show superior performance

The Target Product Profile (TPP) for Chagas disease defines an acceptable test for monitoring treatment response as one that achieves at least 60% sensitivity and 90% specificity, where positive cases correspond to treatment responders (analogous to non-progressors in our study) and negative cases correspond to non-responders (analogous to progressors)^60^. Although the present study focuses on predicting disease progression rather than addressing treatment response, biomarkers capable of identifying patients at increased risk of progression could have important utility for stratifying participants in therapeutic trials and monitoring treatment response. To facilitate comparison with this clinical benchmark, we therefore evaluated performance using the same target thresholds. No individual metabolite biomarker identified in this study met these criteria.

A key advantage of LC–MS–based metabolomics, however, is the ability to combine multiple analytes into multiplexed classifiers. We therefore assessed whether combinations of metabolite biomarkers improved predictive performance. Because the validation cohort 2 included only six progressors and six stable stage A individuals, we defined an acceptable specificity threshold of 83%, corresponding to the misclassification of a single sample on average. When combinatorial analysis was performed using only biomarkers derived from HILIC or only from polar C18 datasets, no metabolite combinations met the desired sensitivity and specificity thresholds. In contrast, integration of features across both chromatographic platforms identified 45 biomarker combinations that satisfied the relaxed threshold criteria (Figure S2, S3, S4), including 10 panels that achieved the stricter 90% specificity threshold (Figure 4, Table 4).

**Figure 4.**
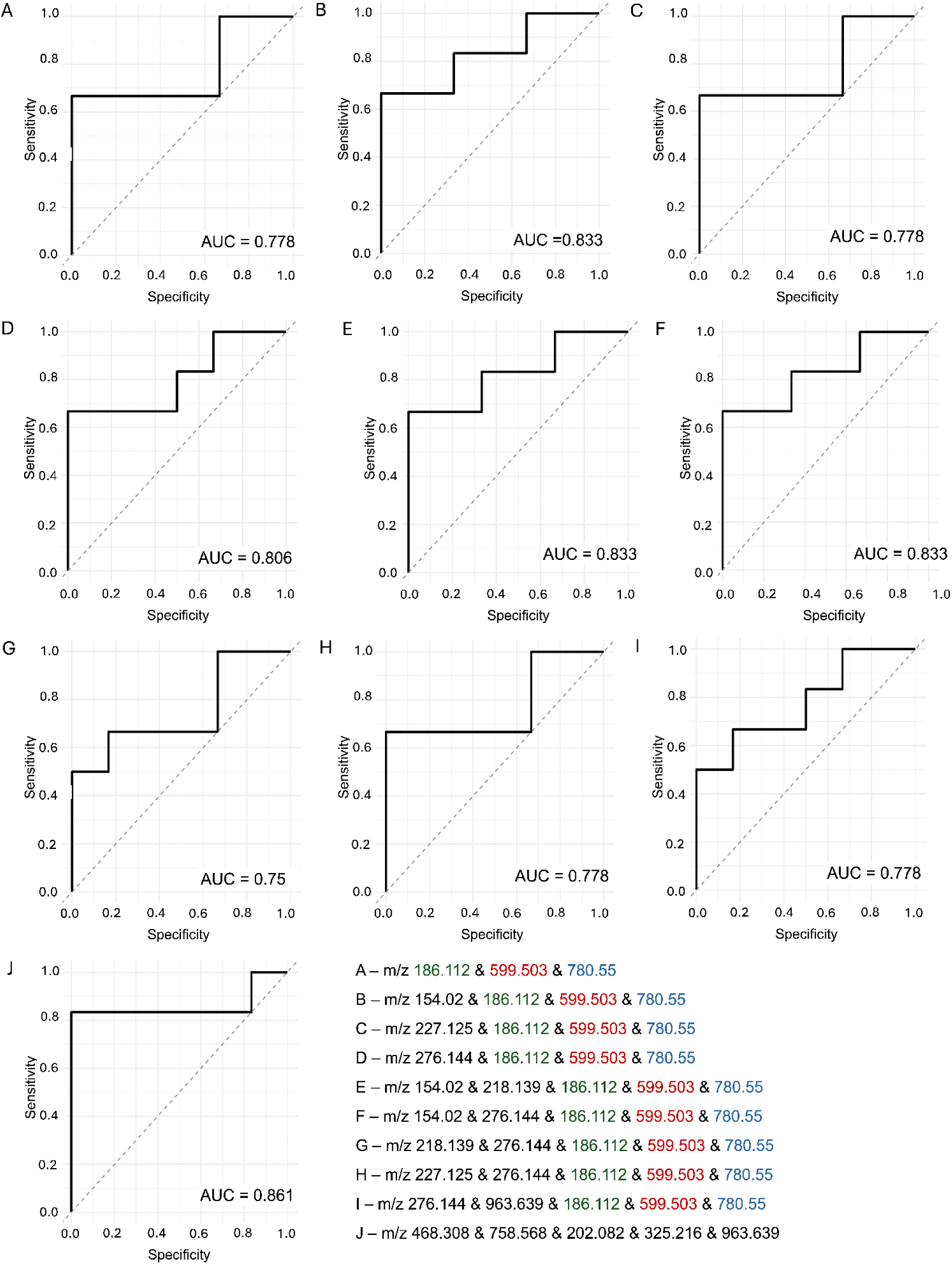
Receiver Operating Characteristic (ROC) curves for the metabolite combinations that met the Chagas disease treatment response test Target Product Profile (sensitivity ≥60% and specificity ≥90%) (A-J). See Table 4 for metabolite retention time and chromatography details.

**Table 4.** Multi-metabolite biomarker combinations with highest diagnostic performance identified by combinatorial ROC analysis. Top-performing metabolite ID panels (combination IDs A–J, corresponding to Fig. 4) identified from integrated pC18 and HILIC LC–MS datasets are shown. Each combination includes metabolites defined by their m/z, retention time (RT), and chromatographic method. Panels were selected based on improved AUC relative to single-metabolite classifiers in bidirectional cross-validated analyses for discrimination of infected progressors versus non-progressors at baseline.

| ID | Feature 1 | Feature 2 | Feature 3 | Feature 4 | Feature 5 |
| --- | --- | --- | --- | --- | --- |
| A | 186.112<br>(5.60, HILIC) | 599.503<br>(1.15, HILIC) | 780.550<br>(8.00, pC18) | – | – |
| B | 154.020<br>(4.55, HILIC) | 186.112<br>(5.60, HILIC) | 599.503<br>(1.15, HILIC) | 780.550<br>(8.00, pC18) | – |
| C | 186.112<br>(5.60, HILIC) | 227.125<br>(4.50, HILIC) | 599.503<br>(1.15, HILIC) | 780.550<br>(8.00, pC18) | – |
| D | 276.144<br>(0.27, pC18) | 186.112<br>(5.60, HILIC) | 599.503<br>(1.15, HILIC) | 780.550<br>(8.00, pC18) | – |
| E | 218.139<br>(0.24, pC18) | 154.020<br>(4.55, HILIC) | 186.112<br>(5.60, HILIC) | 599.503<br>(1.15, HILIC) | 780.550<br>(8.00, pC18) |
| F | 276.144<br>(0.27, pC18) | 154.020<br>(4.55, HILIC) | 186.112<br>(5.60, HILIC) | 599.503<br>(1.15, HILIC) | 780.550<br>(8.00, pC18) |
| G | 218.139<br>(0.24, pC18) | 276.144<br>(0.27, pC18) | 599.503<br>(1.15, HILIC) | 780.550<br>(8.00, pC18) | – |
| H | 276.144<br>(0.27, pC18) | 186.112<br>(5.60, HILIC) | 227.125<br>(4.50, HILIC) | 599.503<br>(1.15, HILIC) | 780.550<br>(8.00, pC18) |
| I | 276.144<br>(0.27, pC18) | 186.112<br>(5.60, HILIC) | 599.503<br>(1.15, HILIC) | 780.550<br>(8.00, pC18) | 963.639<br>(2.10, HILIC) |
| J | 468.308<br>(6.50, pC18) | 758.568<br>(8.70, pC18) | 202.082<br>(6.90, HILIC) | 325.216<br>(5.90, pC18) | 963.639<br>(2.10, HILIC) |

To further quantify the predictive performance of combinatorial metabolite signatures, we evaluated classifier sensitivity, specificity, F1 score, and AUC across discovery and validation cohorts for the top-performing panels (Tables 5 and 6).

**Table 5.** Performance of top multi-metabolite biomarker panels identified by combinatorial analysis across independent cohorts. Performance metrics for metabolite ID combinations (A–J) are shown as sensitivity, specificity, F1 score, and AUC (mean ± SD) in the validation cohort following bidirectional cross-validation. Discovery AUC values indicate classifier performance in the original feature-selection cohort. Panels were selected based on improved discrimination of infected progressors from stage A versus non-progressors from stage A at baseline relative to single-metabolite models and represent candidate multi-analyte signatures with enhanced predictive accuracy.

| ID | Sensitivity | Specificity | F1 Score | Validation AUC | Discovery AUC |
| --- | --- | --- | --- | --- | --- |
| A | 0.67 ± 0.08 | 0.92 ± 0.12 | 0.76 ± 0.06 | 0.776 ± 0.045 | 0.836 ± 0.067 |
| B | 0.62 ± 0.18 | 0.93 ± 0.12 | 0.72 ± 0.16 | 0.757 ± 0.035 | 0.845 ± 0.073 |
| C | 0.62 ± 0.16 | 0.92 ± 0.12 | 0.72 ± 0.13 | 0.779 ± 0.049 | 0.859 ± 0.056 |
| D | 0.68 ± 0.12 | 0.92 ± 0.12 | 0.77 ± 0.07 | 0.788 ± 0.035 | 0.833 ± 0.085 |
| E | 0.62 ± 0.18 | 0.92 ± 0.12 | 0.71 ± 0.16 | 0.761 ± 0.044 | 0.863 ± 0.091 |
| F | 0.60 ± 0.18 | 0.90 ± 0.14 | 0.69 ± 0.16 | 0.769 ± 0.029 | 0.837 ± 0.092 |
| G | 0.65 ± 0.15 | 0.90 ± 0.14 | 0.74 ± 0.08 | 0.786 ± 0.039 | 0.853 ± 0.090 |
| H | 0.62 ± 0.16 | 0.90 ± 0.18 | 0.71 ± 0.13 | 0.776 ± 0.057 | 0.859 ± 0.067 |
| I | 0.63 ± 0.13 | 0.90 ± 0.12 | 0.73 ± 0.11 | 0.757 ± 0.038 | 0.855 ± 0.094 |
| J | 0.63 ± 0.19 | 0.90 ± 0.16 | 0.72 ± 0.17 | 0.833 ± 0.082 | 0.984 ± 0.034 |

**Table 6.** Structural annotation and chemical classification of candidate metabolite biomarkers based on GNPS, SIRIUS, and CANOPUS analyses. Putative metabolite identities are reported with corresponding m/z, retention time (RT), chromatographic method, and detected adduct. Molecular formulas were predicted using SIRIUS, and annotation confidence is indicated by the SIRIUS normalized score. Chemical class assignments were obtained using CANOPUS/ClassyFire, with superclass and class predictions reported together with their associated probabilities (Prob.). Where available, standardized metabolite names from spectral matching are shown; otherwise, features remain annotated as unknown.

| m/z | RT (min) | Method | Adduct | Annotation | Formula | SIRIUS Score | Superclass | Prob. | Class | Prob. |
| --- | --- | --- | --- | --- | --- | --- | --- | --- | --- | --- |
| 154.020 | 4.55 | HILIC | [M+H]<sup>+</sup> | Unknown | C2H8N3OPS | 1.00 | Organoheterocyclic compounds | 0.57 | Azacyclic compounds | 0.58 |
| 186.112 | 5.60 | HILIC | [M+H]<sup>+</sup> | Unknown | C9H15NO3 | 1.00 | Organic acids and derivatives | 1.00 | Carboxylic acids and derivatives | 1.00 |
| 202.082 | 6.90 | HILIC | [M+H]<sup>+</sup> | Unknown | C7H11N3O4 | 1.00 | Organic acids and derivatives | 1.00 | Carboxylic acids and derivatives | 1.00 |
| 227.125 | 4.50 | HILIC | [M+H]<sup>+</sup> | Unknown | C8H14N6O2 | 1.00 | Organoheterocyclic compounds | 0.95 | Triazines | 0.56 |
| 389.231 | 1.34 | HILIC | [M+H]<sup>+</sup> | Unknown | C19H28N6O3 | 0.99 | Organoheterocyclic compounds | 0.99 | Diazines | 0.83 |
| 599.503 | 1.15 | HILIC | [M+H]<sup>+</sup> | Unknown | C39H66O4 | 1.00 | Lipids and lipid-like molecules | 1.00 | Glycerolipids | 1.00 |
| 686.437 | 1.56 | HILIC | [M+H]<sup>+</sup> | Unknown | C36H64NO9P | 0.64 | Lipids and lipid-like molecules | 0.99 | Glycerophospholipids | 1.00 |
| 706.536 | 1.80 | HILIC | [M+H]<sup>+</sup> | PC(30:0) | C38H76NO8P | 1.00 | Lipids and lipid-like molecules | 0.98 | Glycerophospholipids | 1.00 |
| 963.639 | 2.10 | HILIC | [M+H]<sup>+</sup> | Unknown | C46H96N2O14P2 | 0.33 | Lipids and lipid-like molecules | 0.98 | Glycerophospholipids | 1.00 |
| 134.019 | 0.25 | pC18 | [M+H]<sup>+</sup> | Unknown | C4H8NPS | 0.64 | Organic nitrogen compounds | 0.95 | Organonitrogen compounds | 0.94 |
| 218.139 | 0.24 | pC18 | [M+H]<sup>+</sup> | CAR 3:0 | C10H19NO4 | 1.00 | Lipids and lipid-like molecules | 0.98 | Fatty acyls | 0.98 |
| 276.144 | 0.27 | pC18 | [M+H]<sup>+</sup> | CAR DC5:0 | C12H21NO6 | 1.00 | Lipids and lipid-like molecules | 1.00 | Fatty acyls | 1.00 |
| 325.216 | 5.90 | pC18 | [M+H]<sup>+</sup> | Unknown | C22H28O2 | 1.00 | Lipids and lipid-like molecules | 0.90 | Steroids and steroid derivatives | —<sup>a</sup> |
| 468.308 | 6.50 | pC18 | [M+H]<sup>+</sup> | LPC(14:0) | C22H46NO7P | 1.00 | Lipids and lipid-like molecules | 0.98 | Glycerophospholipids | 1.00 |
| **758.568** | 8.70 | pC18 | [M+H]<sup>+</sup> | PC(34:2) | C42H80NO8P | 1.00 | Lipids and lipid-like molecules | 0.99 | Glycerophospholipids | 1.00 |
| **780.550** | 8.00 | pC18 | [M+H]<sup>+</sup> | PC(36:5) | C44H78NO8P | 1.00 | Lipids and lipid-like molecules | 0.98 | Glycerophospholipids | 1.00 |

<sup>a</sup> Manual search, no score

Several panels achieved high specificity while maintaining moderate-to-high sensitivity, highlighting their potential suitability for baseline risk stratification of asymptomatic infected individuals. Notably, panels A-I shared a common metabolite core consisting of m/z 186.112, 599.503, and 780.55, whereas panel J comprised a distinct combination of features. Several metabolites repeatedly detected across high-performing panels including m/z 599.503 and m/z 780.55 were annotated as lipid-related compounds with high ClassyFire classification probabilities (>0.98), supporting their contribution to a shared metabolic progression signature. Consistent with this observation, structural annotation revealed that progression-associated signatures were predominantly enriched in lipids and lipid-like molecules, particularly glycerophospholipids, glycerolipids, and fatty acyl derivatives, with smaller contributions from organic acids and derivatives, organonitrogen compounds, and organoheterocyclic compounds (Table 6). CANOPUS/ClassyFire classification combined with GNPS annotations further identified recurrent phosphatidylcholine species including PC(30:0), PC(34:2), and PC(36:5) as well as LPC(14:0 and short-chain acylcarnitines such as CAR 3:0 and CAR DC5:0, supporting lipid remodeling as a defining feature of early disease progression.

## Discussion

A major need in chronic Chagas disease (CD) management is the ability to predict which asymptomatic infected individuals will progress to cardiomyopathy, enabling clinicians and patients to balance the benefits of antiparasitic therapy against its risk of adverse effects and make an informed decision. Fourteen percent of adults treated with benznidazole, the first line treatment, will suspend it because of adverse effects^61^. Biomarkers predictive of disease progression could also inform clinical trial design, either with regards to initial cohort selection, or with regards to measuring treatment outcomes. Measurements of markers of clinical outcome rather than parasite burden are particularly important given the disconnect observed between parasite levels and outcomes, and in the context of rising interest in host-directed therapeutics for CD^15,62,63^.

Given prior work showing changes in circulating metabolites with *T. cruzi* infection in mouse models^48,58^, their association with disease severity in mouse models^51^, and infection-associated metabolite changes in human CD patients^50,53^, we hypothesized that metabolite levels in the serum of *T. cruzi*-infected people could predict CD progression. Using this approach, we identified multiple individual metabolite biomarkers and combinatorial biomarker panels capable of distinguishing patients who progressed from those who remained asymptomatic during longitudinal follow-up.

Several of the metabolite classes identified in this study, including glycerophosphocholines, and acylcarnitines, have previously been associated with *T. cruzi* infection and disease severity in experimental models^58,64,65^. Alterations in circulating glycerophosphocholines have been linked to inflammatory signaling and membrane remodeling during infection, while changes in acylcarnitines are consistent with impaired mitochondrial fatty acid oxidation and cardiac metabolic stress^66–69^. We observed lower glycerophosphocholines in progressors from stage A compared to non-progressors. This is consistent with our findings in mouse models where mice progressing to lethal disease had lower plasma glycerophosphocholines than uninfected animals or animals in which treatment prevented disease progression^62^.

We envision the greatest utility of these biomarkers in patients who are diagnosed with *T. cruzi* and who must then weigh the benefits of antiparasitic treatment against significant adverse effects in discussion with their clinician, as is the recommendation in many countries for patients above 50 years old^14^. In this context, using existing serological tests to confirm infection status, followed by our biomarkers to assess likelihood of progression, could be a cost-effective approach. Thus, we anticipate that the biomarkers discovered in this study would be implemented once a patient has already been diagnosed with *T. cruzi*. Some biomarkers associated with progression from asymptomatic to symptomatic cardiac disease may reflect shared pathways of cardiac remodeling present in both infected and uninfected patients, as is also observed for indicators of mortality in patients with advanced cardiac disease^45^.

One limitation of this project is the presence of patient comorbidities and behavioral differences that likely increased variability; we could not adjust for these variables in our analyses. Multi-parameter PERMANOVA analysis showed that the different ages of the subjects impact the metabolome but did not confound our interpretation of disease outcomes (Table 2). Although age significantly influenced the overall metabolome, it did not affect classification of progressor status, and we therefore analyzed all samples jointly to preserve statistical power given the limited sample numbers available. Likewise, it is possible that some non-progressors will ultimately progress to symptomatic disease outside this study’s follow-up period, in which case our results may be interpreted as differentiating between faster and slower progressors. However, we did not observe any difference in follow-up duration between progressors and non-progressors (Table 1). The relatively modest number of samples analyzed here reflects the rarity of longitudinally characterized progression cohorts in chronic CD, the challenges of CD classification, and the scarcity of funds available to study this disease, and highlights the importance of future validation in larger prospective cohorts, especially in the context of treatment response where samples may be more accessible.

Given the low parasite burden in chronic CD, the biomarkers identified in this study are almost certainly of host origin. Although multiple DTUs circulate in Bolivia^55^, prior work in animal models has shown concordant changes in glycerophosphocholines and purines across TcI, TcII and TcVI, which appear to be more strongly associated with host responses to the parasite such as inflammation, rather than parasite burden or parasite strain^47,57,58,65^. These observations support the interpretation that the biomarkers identified here capture conserved host metabolic responses associated with cardiac disease progression rather than strain-specific parasite biology.

Assessment of these biomarkers’ performance with a larger sample size and in additional geographic locations would be desirable. However, Bolivia, where our study samples originate, has the highest prevalence of *T. cruzi* infection in the world ^70^ and most *T. cruzi*-infected patients in Spain are of Bolivian origin^71^. Bolivia is also the site of many large-scale clinical trials for new CD therapeutics^7,18,72^. Thus, even if these biomarkers prove to be specific to Bolivian populations, which is unlikely, they will remain highly valuable for clinical trial stratification and therapeutic response assessment in Bolivian settings.

In summary, we have identified eight individual metabolite biomarkers and 45 biomarker combinations that are predictive of progression from asymptomatic to more severe cardiac CD, of which 10 combinations met the criteria of the target product profile for a CD treatment efficacy test^60^. The improved performance of combinatorial metabolite panels compared with individual biomarkers likely reflects the multifactorial pathogenesis of Chagas cardiomyopathy, which involves mitochondrial dysfunction, lipid remodeling, immune activation, and inflammatory signaling^73–75^. These biomarkers show promise for use in human cohorts, either in the context of evaluation of new therapeutic modalities, to prioritize treatment when drug availability is limited, or to increase treatment compliance in the highest-risk group or when patients encounter adverse effects.

## Methods

### Ethics statement

Serum samples were obtained from the Johns Hopkins–Universidad Peruana Cayetano Heredia collaborative Chagas disease biorepository, established under Johns Hopkins Institutional Review Board (IRB) approvals IRB00009799 and IRB00007176. All participants provided written informed consent prior to enrollment.

### Sample collection and diagnostic confirmation

Samples were collected from Chagas disease patients and uninfected controls at San Juan de Dios Hospital in Santa Cruz, Bolivia, as part of multicenter studies on Chagas disease pathogenesis and diagnostics. This is a highly endemic region where community-based surveys have found that approximately one-third of adults are *T. cruzi* seropositive^76^. Chagas disease diagnosis was confirmed by at least two independent positive serological tests. Tests performed included Chagas HAI Polychaco (Laboratorio Lemos S.R.L., Argentina), Chagatest recombinant v.3.0 (Wiener Laboratories SAIC, Argentina), Chagastest lysado (Wiener Laboratories SAIC, Argentina), and Chagas Detect Plus (CDP, InBios International Inc, Seattle, WA, USA), which were performed following manufacturer’s instructions. An IgG TESA blot was also performed on a subset of patients, as previously described^77, 78^.

qPCR was performed for a subset of patients, as follows. Clot samples were processed using a method optimized in^79^, using the High Pure PCR Template Preparation Kit (Roche Diagnostics). The DNA extraction included 300 μL of guanidine 6M and 5 μL of an internal amplification control (IAC). The samples were homogenized using a FastPrep homogenizer and treated with Proteinase K, followed by centrifugation and incubation to complete the DNA purification. Subsequently, a duplex qPCR was performed, targeting the satellite sequence of the nuclear genome of *T. cruzi* and the internal amplification control (IAC), using a previously described protocol with a final volume of 20 μl. The mastermix consisted of 1X FastStart Essential DNA Probes Master (Roche Diagnostics), 0.75 μM of each primer (Cruzi1 and Cruzi2^80^), 0.1 μM of each primer (IACFor and IACRev^80^), and 0.05 μM of each probe (Cruzi3 and IACtq^80^). The cycling conditions included an initial step of 10 minutes at 95 °C, followed by 40 cycles at 95 °C for 15 seconds and 58 °C for 1 minute.

### Participant enrollment and clinical classification

Patients in this cohort were followed longitudinally to evaluate clinical progression. However, because we sought predictive biomarkers of disease progression, only samples from the first visit (prior to progression) were analyzed for metabolomics.

The following electrocardiogram (EKG) and echocardiogram staging criteria was used for determination of cardiac progressors vs cardiac non progressors. EKGs and echocardiograms were read independently by at least one cardiologist familiar with CD and with specific experience treating patients with CD cardiomyopathy. Stages C and D were determined by ejection fraction (EF) on echocardiography. Patients with 40%<EF≤54% for females or 40%<EF≤52% for males were designated stage C, regardless of EKG results. Patients with EF ≤40% were designated stage D, regardless of EKG results.

If EF> 54% for females or EF > 52% for males, then the EKG results were used to guide staging of patients as stage A (fully asymptomatic, normal EKG), stage A2 (mild EKG abnormalities that have not been shown to be significantly more associated with CD than other causes of heart disease or that have not had their association with CD well evaluated^81^), or B (major EKG abnormalities). The following EKG abnormalities were classified as A2: low voltage, left ventricular hypertrophy, ST changes, axis deviation or QT prolongation. To be classified as stage B, patients must present with at least one of the following conditions characteristic of CD^81^: arrhythmias (including atrial fibrillation/flutter, any premature ventricular contractions (PVCs) or ventricular tachycardia), bradycardia (heart rate <50), blocks (including any atrioventricular block, right bundle branch block, left bundle branch block, left anterior fascicular block, left posterior fascicular block, sinus pause/arrest, or bi or tri-fascicular blocks), pathologic Q waves, non-specific intraventricular conduction delay, or A or V pacing. Progressors were defined as patients who progressed to a worse classification over the duration of follow-up (Figure1).

Two separate shipments of samples were received. Full patient clinical metadata is provided in File S1, with information summarized in Table 1.

No *T. cruzi* Discrete Typing Unit (DTU) information was available for these patients^82^. However, a prior cohort study of infected mothers in this region of Bolivia have previously reported circulation of TcII, TcV and TcVI^55,82^. Access to antiparasitic treatment for Chagas disease in Bolivia has improved with the expansion of the National Chagas Network but comprehensive coverage remains limited, especially for adults^56,83^. As individual treatment histories were unavailable for our cohort, prior benznidazole or nifurtimox therapy is unlikely but cannot be excluded.

### Metabolite extraction

Serum metabolites from prior to disease progression were extracted by adding equal volume of 100% methanol with 0.5 μM sulfachloropyridazine (Fisher Optima) as internal standard, vortexed for 15 seconds, followed by centrifugation at 14,800 RPM for 15 minutes. The supernatant was collected into a 96-well plate, then dried down by speedvac, as recommended in Dunn et al^84^. Dried sample plates were stored at -80°C.

### Untargeted LC-MS/MS data acquisition

Before LC-MS/MS instrumental analysis, dried extracts were resuspended with 80% acetonitrile (Fisher Optima; LC-MS grade), spiked with 2 μM sulfadimethoxine (Sigma-Aldrich) as internal standard. Data acquisition was performed under the control of XCalibur and Tune software (ThermoScientific), using a Vanquish LC system and Q-Exactive Plus MS (ThermoFisher). Five pooled quality controls were injected at run start with injection volume 2 μL, 3 μL, 4 μL, and 5 μL to ensure system equilibration. Data was then acquired in randomized order, with a blank followed by a pooled quality control every 12 injections. The injection volume for each sample was 5 μL. We have previously validated this method, demonstrating that it does not lead to retention time shifts or artefactual changes in peak areas^57^ (Figure S5).

A pHILIC LC column (SeQuant® ZIC®-pHILIC, 5 µm polymer, 150 x 2.1 mm) was used, and the column was kept at 40℃ during the run. Liquid chromatography was performed at 0.25 mL/min flow rate for 18 min. LC gradient was as follows, with mobile phase A (water with 20 mM ammonium carbonate, pH adjusted to 9.4) and mobile phase B (acetonitrile): 0-10 min= decrease from 90% B to 30% B; 10-12 min=hold at 30% B; 12-12.666 min=increased to 90% B; 12.666-18 min=hold at 90% B^85^.

After the pHILIC LC-MS run, the same sample plates were dried down and resuspended with 100% water (Fisher Optima; LC-MS grade), spiked with 2 μM sulfadimethoxine (Sigma-Aldrich) as internal standard. A Kinetex polar C18 LC column (Phenomenex; 50 × 2.1 mm, 1.7 μM particle size, 100 Å pore size) was used, and the column was kept at 40℃ during the run. Liquid chromatography was performed at 0.5 mL/min flow rate for 12.5 min. LC gradient was as follows, with mobile phase A (water + 0.1% formic acid) and mobile phase B (acetonitrile +0.1% formic acid): 0-1 min=5% B; 1-9 min=increase to 100% B; 9-11 min=hold at 100% B; 11-11.5 min=decrease to 5% B; and 11.5-12.5 min=5% B. In both cases, mass spectrometry data acquisition was performed in positive polarity and parameters were identical to previously optimized and published parameters^47,57^.

### Untargeted LC-MS/MS data analysis - initial processing

Raw LC–MS/MS data were processed using MZmine (version 2.53)^86^ with parameters as in Table S1. Data were filtered to retain features that were present in at least two samples and had MS2 spectra to enable annotation. Blank removal was performed with a minimum three-fold difference between the blank and serum samples required for a feature to be retained. Feature intensities were normalized using total ion current (TIC) normalization to a constant sum of 1 in Jupyter Notebook using R (version 3.6.1).

Feature tables and MS/MS spectra were analyzed using feature-based molecular networking in GNPS^57,87^ applying parameters consistent with previously described workflows^88^. Annotations were retrieved using a custom script developed in our laboratory^88,89^. Additional annotation of LC–MS/MS features was performed using SIRIUS and CANOPUS to infer molecular formulas and assign putative compound class annotations from fragmentation spectra. Compound superclass and class predictions were assigned using the CANOPUS workflow based on ClassyFire ontology^90–92^. Lipid species, including glycerophosphocholines, glycerophosphoethanolamines, and acylcarnitines, were further supported by database matching against the LIPID MAPS Structure Database (LMSD and COMPDB)^93,94^.

Processed feature tables were integrated with clinical metadata and analyzed in QIIME2^95^ for multivariate analysis. Principal coordinate analysis (PCoA) was performed using Bray–Curtis distance, and group differences were assessed using permutational multivariate analysis of variance (PERMANOVA) with p-values corrected for multiple comparison using Benjamini-Hochberg FDR correction. Ordination plots were visualized using EMPeror^96^.

Statistical analyses and data visualization were performed using Python (version 3.10.14) within Jupyter Notebook environments and RStudio (version 4.1.2).

### Candidate biomarker selection

Candidate biomarkers were selected based on: 1) Mann-Whitney-Wilcoxon test two-sided without correction between infected progressors from stage A and infected non-progressors from stage A, making no assumptions of normality, p<0.05 or 2) random forest variable importance score >1.1 (1,000 trees) for the same comparison. This analysis workflow minimizes false negatives. False positives are reduced by: 1) requiring the same direction of change between non-progressors and progressors from stage A in both cohorts; 2) requiring the same direction of change between patients already in stage B vs stage A, and between progressors from stage A vs patients stably remaining at stage A (cohort 2); 3) initial untargeted ROC analysis and AUC>0.6, implemented using scikit-learn (version 1.0.2). Lastly, validation and false positive minimization was performed using analysis of an independent set of patients, in the targeted PRM analysis workflow (see below).

### Targeted LC-MS/MS data acquisition

Resuspension procedure and LC columns were the same as for untargeted LC-MS/MS data acquisition, but mass spectrometry data acquisition was performed in parallel reaction monitoring (PRM) mode in positive polarity, on an Exploris 240 MS coupled to a Vanquish LC (ThermoFisher). LC conditions were as for untargeted analysis. MS conditions are in Table S2.

### Targeted LC-MS/MS data processing

Collected PRM data was analyzed using the ICIS peak detection algorithm in Xcalibur 4.7 with parameters adjusted on a case-by-case basis to allow for best quantification of the targeted peak. All quantifications were filtered to the targeted parent m/z scan. Exact quantification parameters can be found in Table S3. After quantification, all peaks were normalized to the peak area of the internal standard (m/z 285.0213).

### Combinatorial biomarker analysis

Combinatorial biomarker analysis was performed using the CombiROC package 0.3.4 for R^97^. Preliminary ROC curves were generated for each individual biomarker candidate by training a logistic regression model on cohort 1 data and then applying the model to cohort 2 data. All candidate biomarkers with AUCs > 0.5 from either the HILIC positive mode or pC18 run were included for combinatorial analysis. Combinatorial analysis was performed 10 times, with each “strata” using a random sampling of 8 progressor samples and 8 non-progressor samples from cohort 1 to initially train the model in order to induce variance and balance the classes. The positive class was defined as patients who were stably asymptomatic, and the negative class was defined as patients who progressed to symptomatic disease. All possible combinations of up to 5 biomarkers were considered. After training, all cohort 2 samples were input into the model and predicted as either progressors or non-progressors. Sensitivity, specificity, F1 score, and validated AUC were calculated based on the predicted classes. Afterwards, all 10 strata were averaged, and standard deviations were calculated for each metric. “Good combinations” were defined as those where the averaged validation sensitivity was greater than or equal to 0.6, the averaged validation specificity was greater than or equal to 0.83, and the standard deviation associated with both specificity and sensitivity was less than or equal to 0.2. The sensitivity and specificity cutoff criteria are based on the recommendations of the target product profile for Chagas disease treatment efficacy assessment tests^60^. After good combinations were selected, a logistic regression model for each combination was generated on all cohort 1 data, and ROC curves were created using these trained models on cohort 2 data.

## Supporting information

Supplementary figures and tables referenced in the main text

Participant metadata, including clinical parameters used in the study

MS/MS fragmentation spectra and relative fragment intensities supporting metabolite annotation and biomarker identification

## Data availability

The untargeted MS data generated in this study have been deposited in the MassIVE database for positive mode pHILIC: (MSV000101849) and positive mode polar C18 (MSV000101851). The targeted MS data for PRM validation have been deposited in the MassIVE database for positive mode pHILIC (MSV000101720) and positive mode polar C18 (MSV000101722).

## Code availability

Representative code has been deposited on GitHub, at https://github.com/jarrodRoachChem/ChagasCombinatorialBiomarkerAnalysis.

## Acknowledgments

This project was supported by NIH award number R21AI156669, start-up funds from San Diego State University and the California Metabolic Research Foundation. Samples were from the Johns Hopkins-Universidad Peruana Cayetano Heredia Collaborated Studies Chagas disease biorepository, with prior sample collection, clinical data collection and some salary support to R.H.G and N.M.B. supported by NIH award number R01AI107028 and C.D by F31-HL173973. The content is solely the responsibility of the authors and does not necessarily represent the official views of the National Institutes of Health.

## Author contributions

L-I.M. designed the project. Z.L. performed metabolite extraction. Z.L. and J.G. performed LC-MS data acquisition. Z.L., J.G., J.A.L. and J.R.E. performed LC-MS data analysis. Z.L. drafted the initial manuscript, which was edited by L-I.M., J.G. and J.A.L., with input from all authors. R.H.G. is the PI for the ongoing Bolivian cohort study which provided the samples. R.H.G. and N.M.B. participated in study design, data interpretation, and manuscript editing. K.D., S.V., and C.D. performed data management, cleaning, and staging for the samples. J.C. performed qPCR analyses. G.D.S., M.V., P.C.J., B.M.S., F.T., E.M. and R.M. collected samples and patient data.

