## Supplementary figures and tables referenced in the main text for "Small molecule biomarkers predictive of Chagas disease progression"

### Supplementary information (Files, Figures and Tables)

**File S1. Participant metadata.** Clinical and demographic metadata for all study participants included in the discovery and validation cohorts. Variables include participant identifier, cohort assignment, progression status (progressor vs non-progressor), age, sex, and relevant clinical and diagnostic information used for classification and downstream analyses. All data are de-identified.

**File S2. MS/MS fragmentation patterns for identified biomarkers.**

**Figure S1. PCoA of Bray-Curtis dissimilarities comparing stable (non-progressor) vs. progressor samples analyzed via C18 chromatography (positive ion mode).**

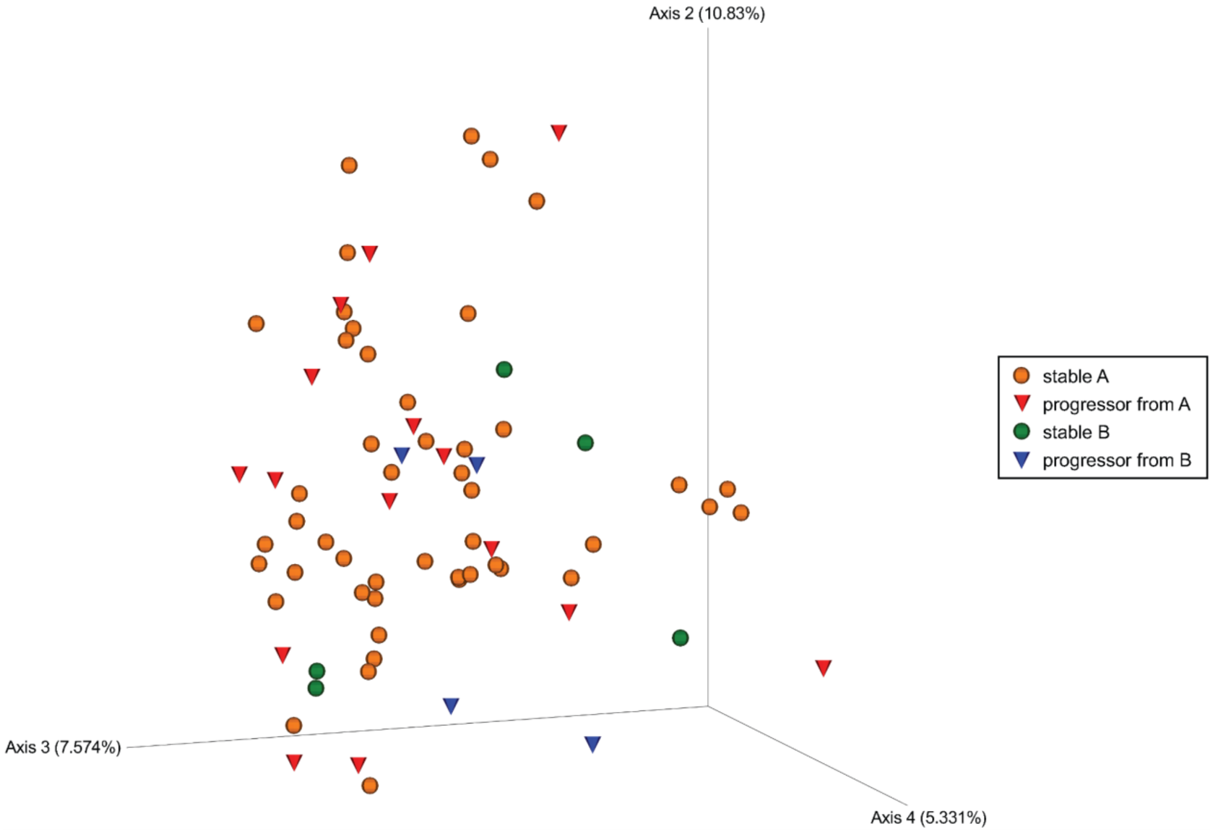

**Figure S2. ROC curves for combinatorial analysis with sensitivity > 0.6 and 0.83 < specificity <0.9**

A) Combination 13632 - Hypoxanthine-154.02-686.437-780.55
B) Combination 13895 - Hypoxanthine-186.112-599.503-780.55
C) Combination 24228 - CAR 3:0-186.112-599.503-780.55
D) Combination 25404 - 186.112-331.226-599.503-780.55
E) Combination 52357 - CAR 5:1;O2-115.063-186.112-599.503-780.55
F) Combination 62242 - Myristoyl-PC-115.063-325.216-780.55-963.639
G) Combination 69916 - 127.05-145.032-584.334-599.503-780.55
H) Combination 76759 - 127.05-186.112-491.373-599.503-780.55
I) Combination 93351 - Hypoxanthine-154.02-186.112-599.503-780.55
J) Combination 96069 - Hypoxanthine-186.112-227.125-599.503-780.55
K) Combination 96583 - Hypoxanthine-186.112-CAR 5:1;O2-599.503-780.55
L) Combination 97369 - Hypoxanthine-Arachidonoylthio PC-186.112-599.503-780.55

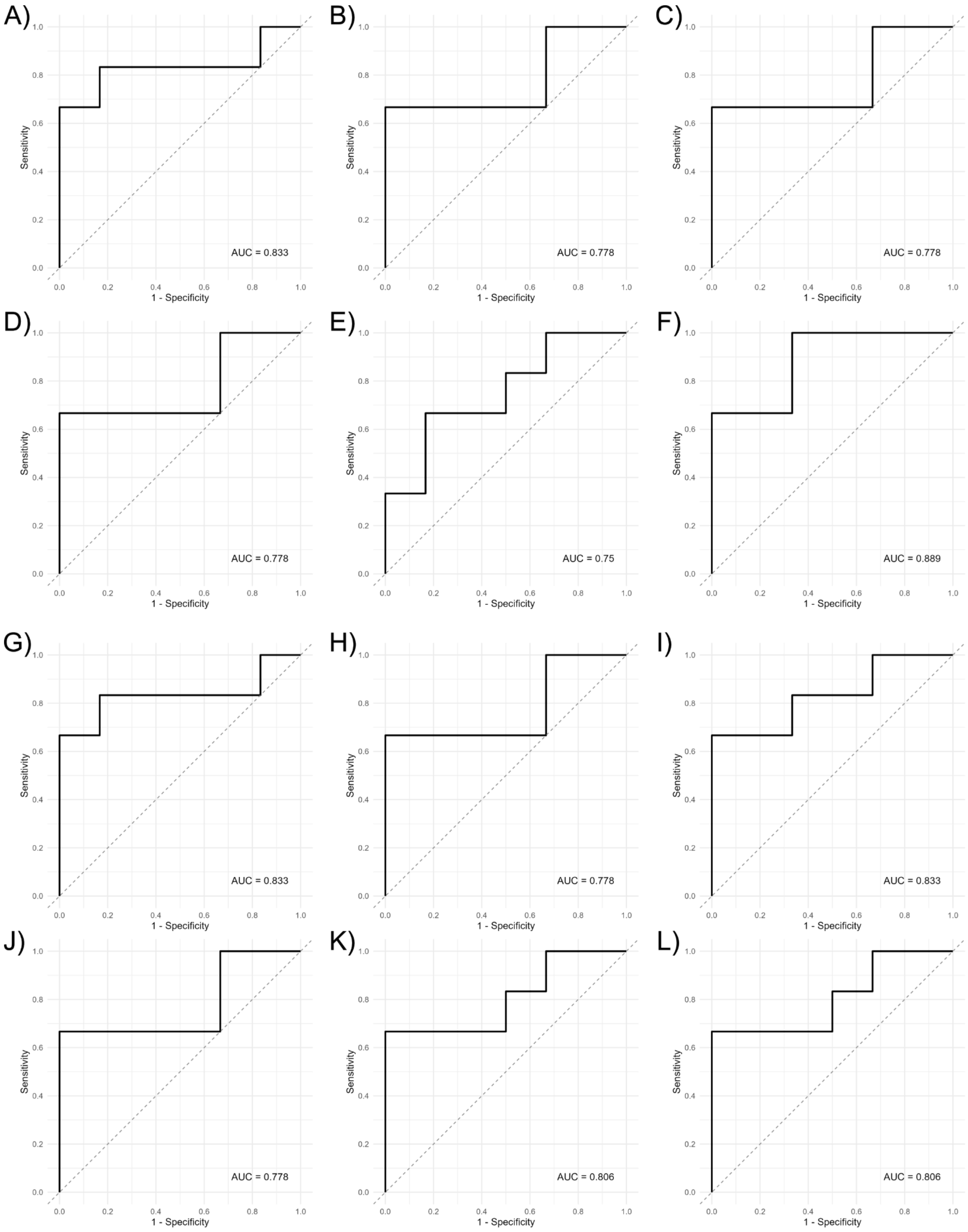
A)
AUC = 0.833
B)
AUC = 0.778
C)
AUC = 0.778
D)
AUC = 0.778
E)
AUC = 0.75
F)
AUC = 0.889
G)
AUC = 0.833
H)
AUC = 0.778
I)
AUC = 0.833
J)
AUC = 0.778
K)
AUC = 0.806
L)
AUC = 0.806
Sensitivity
1 - Specificity
0.0
0.2
0.4
0.6
0.8
1.0

**Figure S3. ROC curves for combinatorial analysis with sensitivity > 0.6 and 0.83 < specificity <0.9**

A) Combination 100716 - Hypoxanthine-CAR 3:0-Arachidonoylthio PC-686.437-780.55
B) Combination 101480 - Hypoxanthine-227.125-319.263-599.503-780.55
C) Combination 101956 - Hypoxanthine-227.125-584.334-599.503-780.55
D) Combination 103589 - Hypoxanthine-265.08-319.263-599.503-780.55
E) Combination 106462 - Hypoxanthine-Myristoyl PC-Palmitoyl PC-325.216-780.55
F) Combination 107907 - Hypoxanthine-584.334-686.437-780.55-963.639
G) Combination 113619 - 145.032-186.112-227.125-599.503-780.55
H) Combination 142263 - 154.02-186.112-389.231-599.503-780.55
I) Combination 142465 - 154.02-186.112-584.334-599.503-780.55
J) Combination 142500 - 154.02-186.112-599.503-638.499-780.55
K) Combination 154130 - CAR 5:1;O2-186.112-202.082-599.503-780.55
L) Combination 156321 - CAR 3:0-186.112-491.373-599.503-780.55

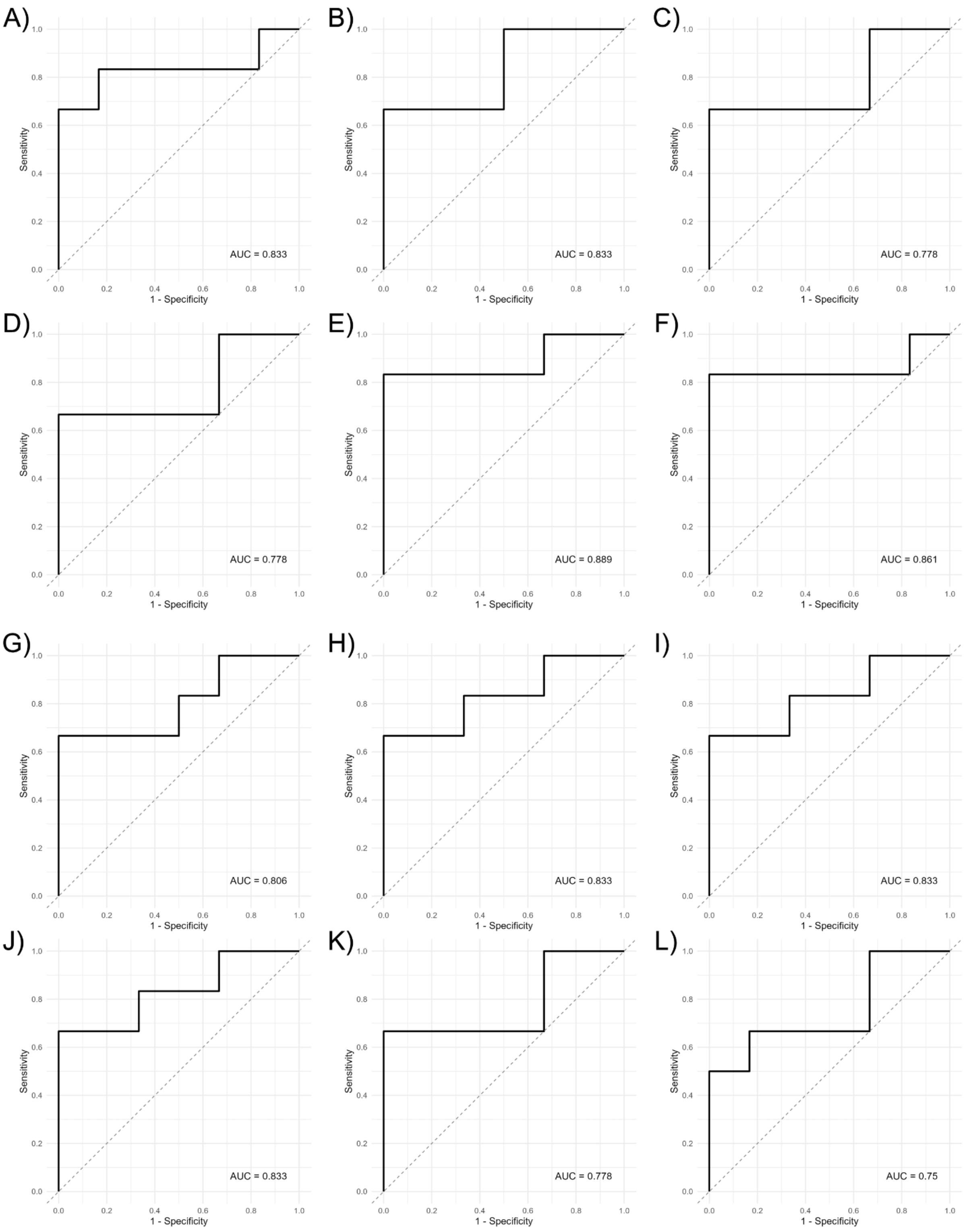
A)
AUC = 0.833
B)
AUC = 0.833
C)
AUC = 0.778
D)
AUC = 0.778
E)
AUC = 0.889
F)
AUC = 0.861
G)
AUC = 0.806
H)
AUC = 0.833
I)
AUC = 0.833
J)
AUC = 0.833
K)
AUC = 0.778
L)
AUC = 0.75
Sensitivity
1 - Specificity
0.0
0.2
0.4
0.6
0.8
1.0

**Figure S4. ROC curves for combinatorial analysis with sensitivity > 0.6 and 0.83 < specificity <0.9**

A) Combination 156402 - CAR 3:0-186.112-584.334-599.503-780.55
B) Combination 156437 - CAR 3:0-186.112-599.503-638.499-780.55
C) Combination 156847 - 186.112-227.125-265.08-599.503-780.55
D) Combination 157732 - 186.112-227.125-584.334-599.503-780.55
E) Combination 159109 - CAR 5:1;O2-186.112-265.08-599.503-780.55
F) Combination 159841 - 186.112-265.08-584.334-599.503-780.55
G) Combination 159876 - 186.112-265.08-599.503-638.499-780.55
H) Combination 160657 - 186.112-CAR 5:1;O2-584.334-599.503-780.55
I) Combination 160711 - CAR 5:1;O2-Arachidonoylthio-PC-186.112-599.503-780.55
J) Combination 163641 - Palmitoyl-PC-186.112-584.334-599.503-780.55
K) Combination 206203 - 1_Myristoyl_2_palmitoyl_sn_glycero_3_phosphocholine-Arachidonoylthio PC-584.334-780.55-963.639

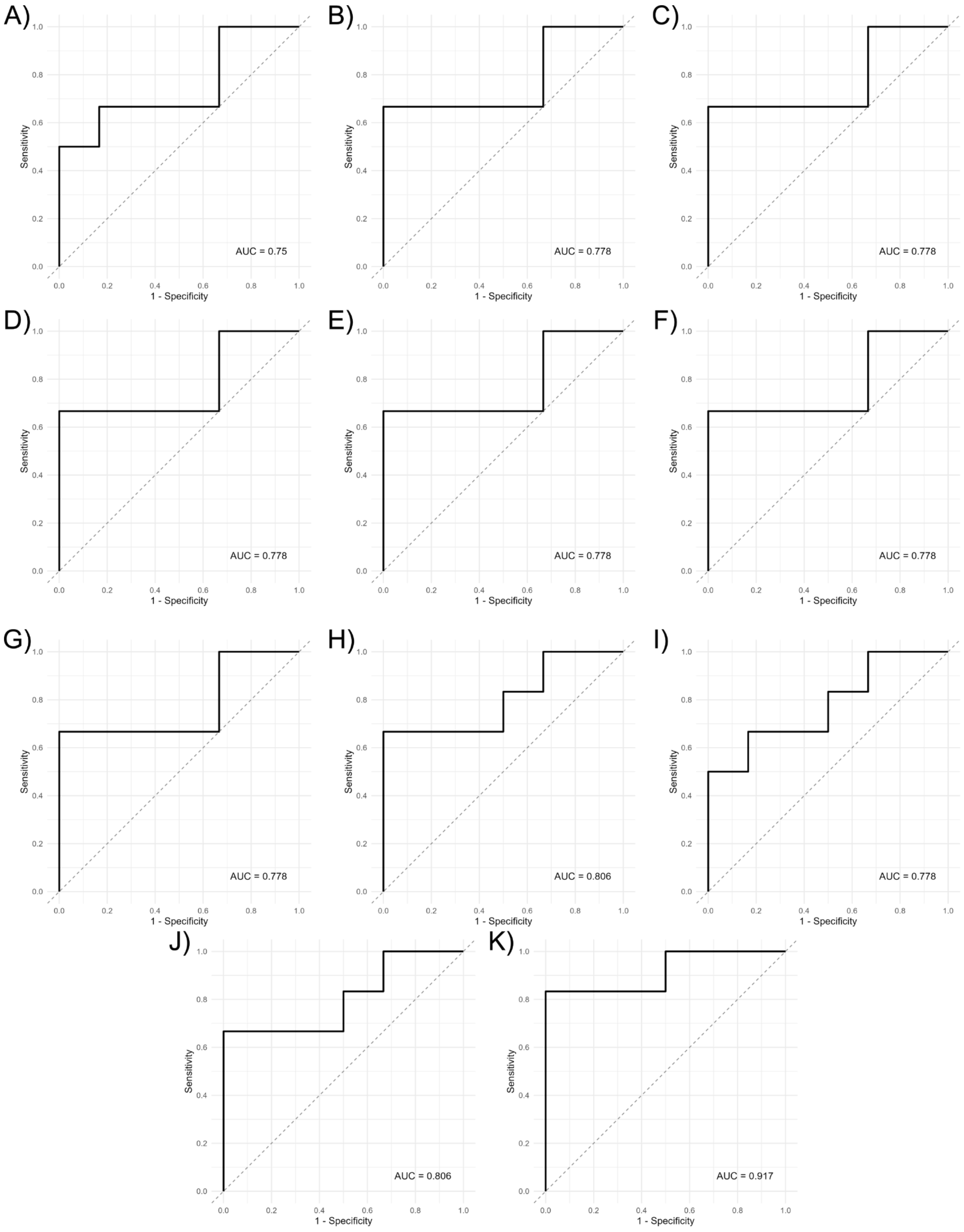
A)
AUC = 0.75
B)
AUC = 0.778
C)
AUC = 0.778
D)
AUC = 0.778
E)
AUC = 0.778
F)
AUC = 0.778
G)
AUC = 0.778
H)
AUC = 0.806
I)
AUC = 0.778
J)
AUC = 0.806
K)
AUC = 0.917
Sensitivity
1 - Specificity
0.0
0.2
0.4
0.6
0.8
1.0

**Figure S5. Comparable retention times and peak areas between samples that are separated by a blank and a pooled quality control (QC) and between consecutive samples.** Note that some variability is expected for the total ion chromatograms, since the samples come from different patients.

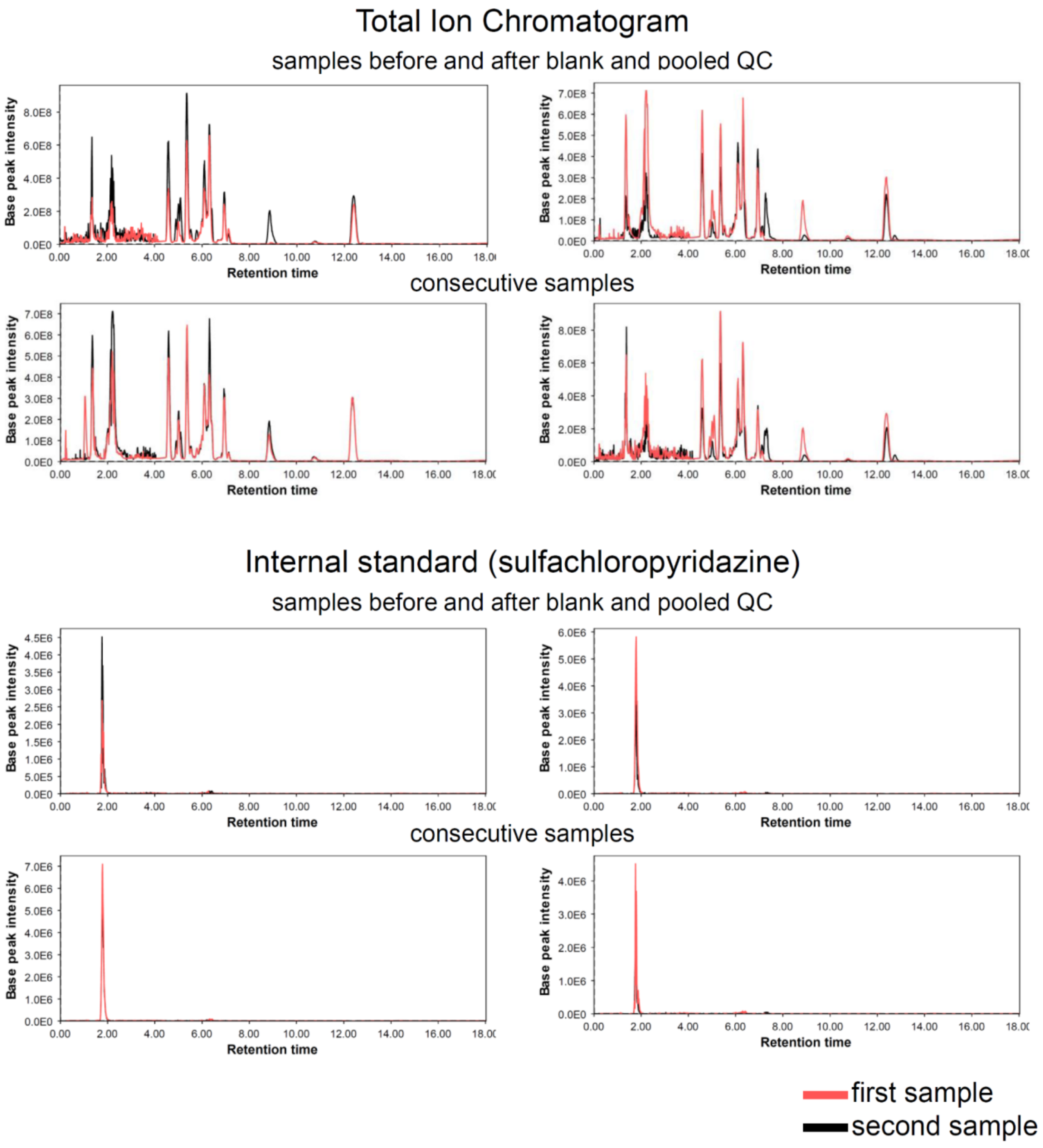

**Figure S6. Differential overall metabolome between shipments.**

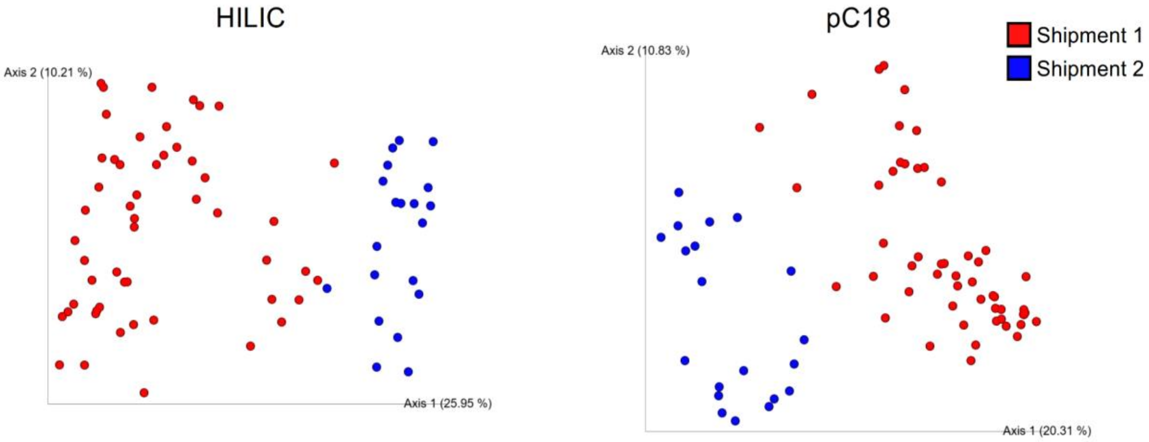

**Table S1. MZmine data processing parameters table for all four datasets.**

| Mzmine parameters | | pHILIC | Polar C18 |
|---|---|---|---|
| $MS^1$ | Retention Time (min) | 0.0 – 18.01 | 0.0 – 12.51 |
| | Noise Level | 1E5 | 1E6 |
| $MS^2$ | Retention Time (min) | 0.0 – 18.01 | 0.0 – 12.51 |
| | Noise Level | 1.0E3 | 1.0E3 |
| Chromatogram Builder ($MS^1$) | Mass List | masses | masses |
| | Minimum Time Span (min) | 0.01 | 0.01 |
| | Minimum Height | 1E5 | 1E6 |
| | *m/z* Tolerance (ppm) | 10.0 | 10.0 |
| Chromatogram Deconvolution | Algorithm | Local minimum search | Local minimum search |
| | *m/z* Range for $MS^2$ Scan Pairing (Da) | 0.01 | 0.01 |
| | RT Range for $MS^2$ Scan Pairing (min) | 0.2 | 0.2 |
| | Chromatographic Threshold | 20% | 20% |
| | Search Minimum in RT Range (min) | 0.07 | 0.07 |
| | Minimum Relative Height | 26% | 26% |
| | Minimum Absolute Height | 1.0E4 | 1.0E4 |
| | Minimum Ratio of Peak Top/Edge | 1 | 1 |
| | Peak Duration Range (min) | 0.01-1.00 | 0.01-1.00 |
| Deisotoping | *m/z* Tolerance (ppm) | 10.0 | 10.0 |
| | Retention Time Tolerance (min) | 0.5 | 0.5 |
| | Monotonic Shape | Checked | Checked |
| | Maximum Charge | 3 | 3 |
| | Representative Isotope | Lowest *m/z* | Lowest *m/z* |

| | | | |
|---|---|---|---|
| Alignment | *m/z* Tolerance (ppm) | 10.0 | 10.0 |
| | Weight for *m/z* | 1 | 1 |
| | Weight for Retention Time | 1 | 1 |
| | Retention Time Tolerance (min) | 0.5 | 0.5 |
| Row Filtering | Minimum Peaks in a Row | 2 | 2 |
| | Retention Time (min) | 0.2 – 17.5 | 0.2 – 12 |
| | Keeps Only Peaks with $MS^2$ Scans | Checked | Checked |
| | Reset Peak No. ID | Checked | Checked |

**Table S2. Targeted analysis MS parameters.**

| Category | Parameter | Value |
|---|---|---|
| Ion Source Parameters | Ion Source Type | H-ESI (Heated Electrospray Ionization) |
| | Spray Voltage (Positive) | 3000 V |
| | Sheath Gas | 50 (arbitrary units) |
| | Aux Gas | 10 (arbitrary units) |
| | Sweep Gas | 1 (arbitrary units) |
| | Ion Transfer Tube Temperature | 325 °C |
| | Vaporizer Temperature | 350 °C |
| MS Global Settings | Method Duration | 12.5 min (polar C18); 18 min (HILIC) |
| | Internal Mass Calibration | EASY-IC™ (Run Start) |
| | Expected LC Peak Width | 6 seconds |
| | Advanced Peak Determination | Enabled |
| | Default Charge State | 1 |
| | Mild Trapping | Disabled |
| Product Ion Scan | Resolution | 15,000 |
| | HCD Collision Energies | 20%, 40%, 60% |
| | Q1 Resolution | 1 m/z |
| | Polarity | Positive |
| | RF Lens | 70% |
| | AGC Target | Standard |
| | Max Injection Time | Auto |
| | Microscans | 1 |
| | Data Type | Centroid |
| | Source Fragmentation | Disabled |

**Table S3. Xcalibur peak area quantification parameters**

| Targeted Run m/z | Annotation | Column | m/z for Quantification | Expected RT | Window | Smoothing Points | Baseline Window | Area Noise Factor | Peak Noise Factor |
|---|---|---|---|---|---|---|---|---|---|
| 134.019 | Unknown | pC18 | 134.019 | 0.25 | 30 | 11 | 30 | 5 | 10 |
| 137.046 | Hypoxanthine | pC18 | 137.046 | 0.38 | 30 | 11 | 30 | 5 | 10 |
| 153.04 | Xanthine | pC18 | 153.04 | 0.39 | 10 | 11 | 30 | 5 | 10 |
| 164.029 | Unknown | pC18 | 164.029 | 0.25 | 30 | 11 | 30 | 5 | 10 |
| 218.139 | Car 3:0 | pC18 | 218.139 | 0.24 | 30 | 11 | 30 | 5 | 10 |
| 246.17 | Car 5:0 | pC18 | 200.164 | 2.4 | 15 | 11 | 30 | 5 | 10 |
| 268.104 | Adenosine | pC18 | 268.104 | 0.5 | 30 | 5 | 30 | 5 | 10 |
| 276.144 | CAR 5:1;O2 | pC18 | 276.144 | 0.27 | 5 | 11 | 30 | 5 | 10 |
| 293.247 | Unknown | pC18 | 293.247 | 7.78 | 30 | 11 | 30 | 5 | 10 |
| 309.242 | Unknown | pC18 | 309.242 | 8.04 | 30 | 11 | 30 | 5 | 10 |
| 319.263 | Unknown | pC18 | 93.0704 | 7.79 | 30 | 11 | 30 | 5 | 10 |
| 325.216 | Unknown | pC18 | 325.215 | 5.9 | 30 | 11 | 30 | 5 | 10 |
| 468.308 | MyristoylPC | pC18 | 468.308 | 6.5 | 30 | 11 | 30 | 5 | 10 |
| 528.308 | Unknown | pC18 | 528.308 | 6.8 | 30 | 11 | 30 | 5 | 10 |
| 658.019 | Unknown | pC18 | 658.019 | 4.04 | 30 | 11 | 30 | 5 | 10 |
| 701.558 | Unknown | pC18 | 701.558 | 7.94 | 30 | 11 | 30 | 5 | 10 |
| 758.568 | PalmitoylPC | pC18 | 758.568 | 8.7 | 30 | 11 | 30 | 5 | 10 |
| 780.55 | Unknown | pC18 | 184.0734 | 8 | 30 | 11 | 30 | 5 | 10 |
| 784.583 | ArachidonoylthioPC | pC18 | 784.583 | 8.96 | 30 | 11 | 30 | 5 | 10 |
| 115.063 | Unknown | HILIC | 70.0659 | 4.58 | 15 | 5 | 30 | 3 | 1 |

| | | | | | | | | | |
|---|---|---|---|---|---|---|---|---|---|
| 127.05 | Unknown | HILIC | 81.0453 | 6.5 | 15 | 9 | 30 | 1 | 1 |
| 137.046 | Hypoxanthine | HILIC | 137.046 | 4.94 | 10 | 5 | 50 | 5 | 10 |
| 140.118 | Unknown | HILIC | 140.118 | 2.1 | 25 | 5 | 30 | 3 | 1 |
| 145.032 | Unknown | HILIC | 73.0113 | 5 | 15 | 5 | 30 | 3 | 1 |
| 153.041 | Xanthine | HILIC | 153.041 | 5.76 | 15 | 15 | 30 | 3 | 1 |
| 154.02 | Unknown | HILIC | 59.0378 | 4.55 | 15 | 5 | 30 | 3 | 1 |
| 169.036 | Urate | HILIC | 169.036 | 6.21 | 15 | 5 | 30 | 3 | 1 |
| 186.112 | Unknown | HILIC | 186.112 | 5.6 | 25 | 5 | 30 | 3 | 1 |
| 202.082 | Unknown | HILIC | 114.0666 | 6.9 | 15 | 11 | 60 | 1 | 1 |
| 209.069 | Unknown | HILIC | 209.069 | 6.71 | 15 | 5 | 30 | 3 | 1 |
| 217.118 | Unknown | HILIC | 217.118 | 4.02 | 25 | 5 | 30 | 3 | 1 |
| 227.125 | Unknown | HILIC | 114.0664 | 4.5 | 25 | 5 | 45 | 3 | 1 |
| 244.154 | Unknown | HILIC | 244.154 | 1.15 | 15 | 5 | 30 | 3 | 1 |
| 252.928 | Unknown | HILIC | 252.928 | 12.37 | 15 | 5 | 30 | 3 | 1 |
| 265.08 | Unknown | HILIC | 152.0476 | 4.5 | 15 | 15 | 30 | 3 | 1 |
| 269.088 | Inosine | HILIC | 269.088 | 5.42 | 15 | 5 | 30 | 3 | 1 |
| 285.083 | Xanthosine | HILIC | 285.083 | 5.9 | 15 | 5 | 30 | 3 | 1 |
| 296.917 | Unknown | HILIC | 296.917 | 12.37 | 15 | 5 | 30 | 3 | 1 |
| 314.232 | CAR 10:1 | HILIC | 85.028 | 2.15 | 15 | 11 | 45 | 5 | 10 |
| 331.226 | Unknown | HILIC | 57.07 | 1.36 | 15 | 9 | 30 | 1 | 1 |
| 389.231 | Unknown | HILIC | 79.0549 | 1.34 | 15 | 5 | 30 | 3 | 10 |
| 397.915 | Unknown | HILIC | 397.915 | 5.58 | 25 | 5 | 30 | 3 | 1 |
| 491.373 | Unknown | HILIC | 81.07 | 1.32 | 10 | 11 | 30 | 1 | 10 |
| 539.43 | Unknown | HILIC | 81.07 | 1.3 | 10 | 9 | 30 | 1 | 10 |
| 584.334 | Unknown | HILIC | 184.073 | 2.1 | 15 | 5 | 30 | 3 | 1 |
| 599.503 | Unknown | HILIC | 599.503 | 1.15 | 15 | 5 | 30 | 3 | 1 |
| 638.499 | Unknown | HILIC | 252.0867 | 1.26 | 15 | 5 | 30 | 1 | 1 |

| | | | | | | | | | |
|---|---|---|---|---|---|---|---|---|---|
| 686.437 | Unknown | HILIC | 184.073 | 1.56 | 15 | 11 | 30 | 1 | 1 |
| 706.536 | 1-Myristoyl-2-palmitoyl-sn-glycero-3-phosphocholine | HILIC | 184.073 | 1.8 | 30 | 11 | 50 | 1 | 1 |
| 834.6 | PC 18:22;6 | HILIC | 184.0732 | 1.44 | 15 | 5 | 60 | 3 | 1 |
| 963.639 | Unknown | HILIC | 184.0735 | 2.1 | 15 | 5 | 30 | 3 | 1 |
| 311.0809 | Sulfadimethoxine | Internal Standard | 156.0766 | 1.66 | 10 | 11 | 50 | 5 | 10 |
| 285.0207 | Sulfachloropyridazine | Internal Standard | 156.0112 | 2.14 | 10 | 11 | 80 | 5 | 10 |
