## Supplementary material for "Small molecule biomarkers predictive of Chagas disease progression": MS/MS fragmentation spectra and relative fragment intensities supporting metabolite annotation and biomarker identification

| LC–MS Metho | Precursor m/z | Fragment m/z | Absolute Inter | Relative Intensity (%) |
|---|---|---|---|---|
| pHILIC | 154.02 | 154.0443 | 43723.2 | 100 |
| pHILIC | 154.02 | 59.0729 | 14339 | 32.8 |
| pHILIC | 154.02 | 58.0651 | 1058.2 | 2.42 |
| pHILIC | 154.02 | 154.1221 | 374.8 | 0.86 |
| pHILIC | 154.02 | 154.0855 | 144.8 | 0.33 |
| pHILIC | 154.02 | 40.9611 | 69.8 | 0.16 |
| pHILIC | 154.02 | 93.4515 | 69.6 | 0.16 |
| pHILIC | 154.02 | 94.4882 | 67.9 | 0.16 |
| pHILIC | 154.02 | 93.385 | 67.6 | 0.15 |
| pHILIC | 154.02 | 91.6338 | 64.7 | 0.15 |
| pHILIC | 154.02 | 150.9799 | 64.7 | 0.15 |
| pHILIC | 154.02 | 75.9762 | 64.4 | 0.15 |
| pHILIC | 154.02 | 100.1332 | 64.2 | 0.15 |
| pHILIC | 154.02 | 154.0596 | 64 | 0.15 |
| pHILIC | 154.02 | 62.803 | 63.7 | 0.15 |
| pHILIC | 154.02 | 64.096 | 62.7 | 0.14 |
| pHILIC | 154.02 | 42.9818 | 62.6 | 0.14 |
| pHILIC | 154.02 | 51.5233 | 62.6 | 0.14 |
| pHILIC | 154.02 | 48.3699 | 62.5 | 0.14 |
| pHILIC | 154.02 | 66.4373 | 61.9 | 0.14 |
| pHILIC | 154.02 | 148.885 | 61.8 | 0.14 |
| pHILIC | 154.02 | 102.7398 | 61.8 | 0.14 |
| pHILIC | 154.02 | 70.6712 | 61.4 | 0.14 |
| pHILIC | 154.02 | 62.0483 | 61.3 | 0.14 |
| pHILIC | 154.02 | 70.0647 | 61.3 | 0.14 |
| pHILIC | 154.02 | 73.976 | 61.3 | 0.14 |
| pHILIC | 154.02 | 44.232 | 61.2 | 0.14 |
| pHILIC | 154.02 | 177.2213 | 61.2 | 0.14 |
| pHILIC | 154.02 | 78.5461 | 61.2 | 0.14 |
| pHILIC | 154.02 | 92.1193 | 60.8 | 0.14 |
| pHILIC | 154.02 | 66.9617 | 60.6 | 0.14 |
| pHILIC | 154.02 | 110.5306 | 60.4 | 0.14 |
| pHILIC | 154.02 | 62.6414 | 60 | 0.14 |
| pHILIC | 154.02 | 80.2827 | 59.7 | 0.14 |
| pHILIC | 154.02 | 89.8683 | 59.6 | 0.14 |
| pHILIC | 154.02 | 50.9117 | 59.6 | 0.14 |
| pHILIC | 154.02 | 119.6329 | 59.4 | 0.14 |
| pHILIC | 154.02 | 58.1161 | 59.1 | 0.14 |
| pHILIC | 154.02 | 61.1122 | 59 | 0.14 |
| pHILIC | 154.02 | 84.0898 | 59 | 0.14 |
| pHILIC | 154.02 | 50.8407 | 58.6 | 0.13 |

| | | | | |
|---|---|---|---|---|
| pHILIC | 154.02 | 41.0151 | 58.4 | 0.13 |
| pHILIC | 154.02 | 87.0351 | 58.2 | 0.13 |
| pHILIC | 154.02 | 108.8162 | 58.2 | 0.13 |
| pHILIC | 154.02 | 142.4893 | 58.2 | 0.13 |
| pHILIC | 154.02 | 149.8057 | 57.8 | 0.13 |
| pHILIC | 154.02 | 94.637 | 57.4 | 0.13 |
| pHILIC | 154.02 | 42.8405 | 56.6 | 0.13 |
| pHILIC | 154.02 | 71.6097 | 56.2 | 0.13 |
| pHILIC | 154.02 | 46.8797 | 55.5 | 0.13 |
| pHILIC | 154.02 | 91.6914 | 46.2 | 0.11 |
| pHILIC | 154.02 | 108.7184 | 44.3 | 0.1 |
| pHILIC | 154.02 | 98.2521 | 42.9 | 0.1 |
| pHILIC | 154.02 | 93.3556 | 42.5 | 0.1 |
| pHILIC | 154.02 | 131.743 | 41.7 | 0.1 |
| pHILIC | 154.02 | 98.8731 | 40.9 | 0.09 |
| pHILIC | 154.02 | 40.9039 | 40.5 | 0.09 |
| pHILIC | 154.02 | 93.2825 | 40.1 | 0.09 |
| pHILIC | 154.02 | 44.0199 | 39.9 | 0.09 |
| pHILIC | 154.02 | 83.3959 | 39.3 | 0.09 |
| pHILIC | 154.02 | 117.3133 | 38.8 | 0.09 |
| pHILIC | 154.02 | 165.499 | 38.6 | 0.09 |
| pHILIC | 154.02 | 123.3878 | 38.5 | 0.09 |
| pHILIC | 154.02 | 143.1989 | 38.3 | 0.09 |
| pHILIC | 154.02 | 110.2434 | 38.3 | 0.09 |
| pHILIC | 154.02 | 54.3604 | 38.3 | 0.09 |
| pHILIC | 154.02 | 46.3281 | 38.3 | 0.09 |
| pHILIC | 154.02 | 110.6053 | 38.2 | 0.09 |
| pHILIC | 154.02 | 56.6597 | 38.1 | 0.09 |
| pHILIC | 154.02 | 51.4805 | 38.1 | 0.09 |
| pHILIC | 154.02 | 171.9109 | 38 | 0.09 |
| pHILIC | 154.02 | 68.0937 | 37.9 | 0.09 |
| pHILIC | 154.02 | 94.1578 | 37.9 | 0.09 |
| pHILIC | 154.02 | 57.3137 | 37.8 | 0.09 |
| pHILIC | 154.02 | 147.6943 | 37.8 | 0.09 |
| pHILIC | 154.02 | 95.9011 | 37.7 | 0.09 |
| pHILIC | 154.02 | 42.9688 | 37.6 | 0.09 |
| pHILIC | 154.02 | 80.1499 | 37.5 | 0.09 |
| pHILIC | 154.02 | 43.2604 | 37.4 | 0.09 |
| pHILIC | 154.02 | 46.8419 | 37.3 | 0.09 |
| pHILIC | 154.02 | 149.4311 | 37.3 | 0.09 |
| pHILIC | 154.02 | 86.9771 | 37.3 | 0.09 |
| pHILIC | 154.02 | 69.8292 | 37.3 | 0.09 |

| | | | | |
|---|---|---|---|---|
| pHILIC | 154.02 | 75.3629 | 37.2 | 0.09 |
| pHILIC | 154.02 | 75.6092 | 37.2 | 0.09 |
| pHILIC | 154.02 | 58.9011 | 37 | 0.08 |
| pHILIC | 154.02 | 96.4812 | 37 | 0.08 |
| pHILIC | 154.02 | 71.719 | 37 | 0.08 |
| pHILIC | 154.02 | 41.1835 | 36.9 | 0.08 |
| pHILIC | 154.02 | 58.1677 | 36.8 | 0.08 |
| pHILIC | 154.02 | 46.4911 | 36.8 | 0.08 |
| pHILIC | 154.02 | 65.4978 | 36.8 | 0.08 |
| pHILIC | 154.02 | 100.032 | 36.7 | 0.08 |
| pHILIC | 154.02 | 59.4869 | 36.7 | 0.08 |
| pHILIC | 154.02 | 50.1628 | 36.6 | 0.08 |
| pHILIC | 154.02 | 47.8761 | 36.6 | 0.08 |
| pHILIC | 154.02 | 70.0788 | 36.6 | 0.08 |
| pHILIC | 154.02 | 65.4638 | 36.5 | 0.08 |
| pHILIC | 154.02 | 85.9156 | 36.4 | 0.08 |
| pHILIC | 154.02 | 151.7611 | 36.3 | 0.08 |
| pHILIC | 154.02 | 125.0043 | 36.3 | 0.08 |
| pHILIC | 154.02 | 125.3792 | 36.3 | 0.08 |
| pHILIC | 154.02 | 93.6585 | 36.3 | 0.08 |
| pHILIC | 154.02 | 60.9005 | 36.2 | 0.08 |
| pHILIC | 154.02 | 85.4066 | 36.2 | 0.08 |
| pHILIC | 154.02 | 122.7789 | 36.2 | 0.08 |
| pHILIC | 154.02 | 45.8042 | 36 | 0.08 |
| pHILIC | 154.02 | 45.798 | 36 | 0.08 |
| pHILIC | 154.02 | 137.1592 | 36 | 0.08 |
| pHILIC | 154.02 | 48.3039 | 36 | 0.08 |
| pHILIC | 154.02 | 48.5097 | 36 | 0.08 |
| pHILIC | 154.02 | 43.5441 | 35.9 | 0.08 |
| pHILIC | 154.02 | 123.2432 | 35.9 | 0.08 |
| pHILIC | 154.02 | 54.447 | 35.8 | 0.08 |
| pHILIC | 154.02 | 79.529 | 35.8 | 0.08 |
| pHILIC | 154.02 | 134.108 | 35.8 | 0.08 |
| pHILIC | 154.02 | 117.8515 | 35.8 | 0.08 |
| pHILIC | 154.02 | 43.5974 | 35.7 | 0.08 |
| pHILIC | 154.02 | 62.6337 | 35.7 | 0.08 |
| pHILIC | 154.02 | 164.5373 | 35.7 | 0.08 |
| pHILIC | 154.02 | 175.1449 | 35.7 | 0.08 |
| pHILIC | 154.02 | 144.1303 | 35.7 | 0.08 |
| pHILIC | 154.02 | 41.9302 | 35.6 | 0.08 |
| pHILIC | 154.02 | 156.8344 | 35.6 | 0.08 |
| pHILIC | 154.02 | 92.6099 | 35.6 | 0.08 |

| | | | | |
|---|---|---|---|---|
| pHILIC | 154.02 | 139.7951 | 35.6 | 0.08 |
| pHILIC | 154.02 | 86.9924 | 35.6 | 0.08 |
| pHILIC | 154.02 | 94.534 | 35.5 | 0.08 |
| pHILIC | 154.02 | 124.4182 | 35.5 | 0.08 |
| pHILIC | 154.02 | 46.4182 | 35.5 | 0.08 |
| pHILIC | 154.02 | 45.5334 | 35.5 | 0.08 |
| pHILIC | 154.02 | 83.1103 | 35.5 | 0.08 |
| pHILIC | 154.02 | 64.0012 | 35.4 | 0.08 |
| pHILIC | 154.02 | 105.4121 | 35.4 | 0.08 |
| pHILIC | 154.02 | 165.6203 | 35.4 | 0.08 |
| pHILIC | 154.02 | 78.9397 | 35.4 | 0.08 |
| pHILIC | 154.02 | 72.2456 | 35.4 | 0.08 |
| pHILIC | 154.02 | 155.5517 | 35.3 | 0.08 |
| pHILIC | 154.02 | 46.8722 | 35.3 | 0.08 |
| pHILIC | 154.02 | 94.6464 | 35.3 | 0.08 |
| pHILIC | 154.02 | 124.4963 | 35.3 | 0.08 |
| pHILIC | 154.02 | 83.3237 | 35.3 | 0.08 |
| pHILIC | 154.02 | 47.9831 | 35.3 | 0.08 |
| pHILIC | 154.02 | 42.122 | 35.3 | 0.08 |
| pHILIC | 154.02 | 44.0807 | 35.3 | 0.08 |
| pHILIC | 154.02 | 43.0325 | 35.3 | 0.08 |
| pHILIC | 154.02 | 40.4659 | 35.3 | 0.08 |
| pHILIC | 154.02 | 47.825 | 35.3 | 0.08 |
| pHILIC | 154.02 | 107.3667 | 35.3 | 0.08 |
| pHILIC | 154.02 | 94.4642 | 35.3 | 0.08 |
| pHILIC | 154.02 | 99.0057 | 35.3 | 0.08 |
| pHILIC | 154.02 | 91.2422 | 35.3 | 0.08 |
| pHILIC | 154.02 | 48.306 | 35.2 | 0.08 |
| pHILIC | 154.02 | 52.6021 | 35.2 | 0.08 |
| pHILIC | 154.02 | 110.8656 | 35.2 | 0.08 |
| pHILIC | 154.02 | 64.2746 | 35.2 | 0.08 |
| pHILIC | 154.02 | 115.138 | 35.2 | 0.08 |
| pHILIC | 154.02 | 81.3732 | 35.2 | 0.08 |
| pHILIC | 154.02 | 133.0856 | 35.2 | 0.08 |
| pHILIC | 154.02 | 67.8521 | 35.2 | 0.08 |
| pHILIC | 154.02 | 112.1526 | 35.2 | 0.08 |
| pHILIC | 154.02 | 61.4449 | 35.1 | 0.08 |
| pHILIC | 154.02 | 45.3781 | 35.1 | 0.08 |
| pHILIC | 154.02 | 87.9994 | 35.1 | 0.08 |
| pHILIC | 154.02 | 52.3307 | 35.1 | 0.08 |
| pHILIC | 154.02 | 141.5225 | 35 | 0.08 |
| pHILIC | 154.02 | 169.0646 | 35 | 0.08 |

| | | | | |
|---|---|---|---|---|
| pHILIC | 154.02 | 45.9927 | 35 | 0.08 |
| pHILIC | 154.02 | 104.2523 | 35 | 0.08 |
| pHILIC | 154.02 | 53.2329 | 35 | 0.08 |
| pHILIC | 154.02 | 141.4522 | 35 | 0.08 |
| pHILIC | 154.02 | 70.1554 | 35 | 0.08 |
| pHILIC | 154.02 | 64.4988 | 35 | 0.08 |
| pHILIC | 154.02 | 44.0656 | 34.9 | 0.08 |
| pHILIC | 154.02 | 116.6384 | 34.9 | 0.08 |
| pHILIC | 154.02 | 66.7373 | 34.9 | 0.08 |
| pHILIC | 154.02 | 84.7429 | 34.9 | 0.08 |
| pHILIC | 154.02 | 123.1467 | 34.8 | 0.08 |
| pHILIC | 154.02 | 43.2314 | 34.8 | 0.08 |
| pHILIC | 154.02 | 125.0481 | 34.8 | 0.08 |
| pHILIC | 154.02 | 90.9978 | 34.8 | 0.08 |
| pHILIC | 154.02 | 47.6485 | 34.8 | 0.08 |
| pHILIC | 154.02 | 64.5243 | 34.8 | 0.08 |
| pHILIC | 154.02 | 47.3998 | 34.7 | 0.08 |
| pHILIC | 154.02 | 93.5011 | 34.7 | 0.08 |
| pHILIC | 154.02 | 60.5629 | 34.7 | 0.08 |
| pHILIC | 154.02 | 82.7483 | 34.7 | 0.08 |
| pHILIC | 154.02 | 97.5395 | 34.7 | 0.08 |
| pHILIC | 154.02 | 46.8395 | 34.6 | 0.08 |
| pHILIC | 154.02 | 115.9223 | 34.6 | 0.08 |
| pHILIC | 154.02 | 67.7894 | 34.6 | 0.08 |
| pHILIC | 154.02 | 92.4463 | 34.6 | 0.08 |
| pHILIC | 154.02 | 63.3708 | 34.6 | 0.08 |
| pHILIC | 154.02 | 162.0535 | 34.5 | 0.08 |
| pHILIC | 154.02 | 105.3261 | 34.5 | 0.08 |
| pHILIC | 154.02 | 42.7272 | 34.5 | 0.08 |
| pHILIC | 154.02 | 100.0104 | 34.5 | 0.08 |
| pHILIC | 154.02 | 110.7889 | 34.5 | 0.08 |
| pHILIC | 154.02 | 65.1993 | 34.5 | 0.08 |
| pHILIC | 154.02 | 93.4444 | 34.5 | 0.08 |
| pHILIC | 154.02 | 44.6655 | 34.5 | 0.08 |
| pHILIC | 154.02 | 93.4251 | 34.5 | 0.08 |
| pHILIC | 154.02 | 74.9165 | 34.5 | 0.08 |
| pHILIC | 154.02 | 40.3095 | 34.5 | 0.08 |
| pHILIC | 154.02 | 82.7626 | 34.5 | 0.08 |
| pHILIC | 154.02 | 81.622 | 34.5 | 0.08 |
| pHILIC | 154.02 | 64.5424 | 34.4 | 0.08 |
| pHILIC | 154.02 | 147.5401 | 34.4 | 0.08 |
| pHILIC | 154.02 | 60.8026 | 34.4 | 0.08 |

| | | | | |
|---|---|---|---|---|
| pHILIC | 154.02 | 77.3007 | 34.4 | 0.08 |
| pHILIC | 154.02 | 50.5917 | 34.4 | 0.08 |
| pHILIC | 154.02 | 43.5896 | 34.4 | 0.08 |
| pHILIC | 154.02 | 45.5889 | 34.4 | 0.08 |
| pHILIC | 154.02 | 136.0631 | 34.4 | 0.08 |
| pHILIC | 154.02 | 62.0044 | 34.4 | 0.08 |
| pHILIC | 154.02 | 86.1704 | 34.4 | 0.08 |
| pHILIC | 154.02 | 41.5812 | 34.4 | 0.08 |
| pHILIC | 154.02 | 47.9286 | 34.4 | 0.08 |
| pHILIC | 154.02 | 71.4405 | 34.4 | 0.08 |
| pHILIC | 154.02 | 83.389 | 34.3 | 0.08 |
| pHILIC | 154.02 | 51.5537 | 34.3 | 0.08 |
| pHILIC | 154.02 | 108.1919 | 34.3 | 0.08 |
| pHILIC | 154.02 | 46.6201 | 34.3 | 0.08 |
| pHILIC | 154.02 | 76.1958 | 34.3 | 0.08 |
| pHILIC | 154.02 | 134.6189 | 34.3 | 0.08 |
| pHILIC | 154.02 | 111.0898 | 34.3 | 0.08 |
| pHILIC | 154.02 | 56.9768 | 34.3 | 0.08 |
| pHILIC | 154.02 | 78.718 | 34.2 | 0.08 |
| pHILIC | 154.02 | 122.3926 | 34.2 | 0.08 |
| pHILIC | 154.02 | 103.404 | 34.2 | 0.08 |
| pHILIC | 154.02 | 50.4272 | 34.2 | 0.08 |
| pHILIC | 154.02 | 71.8927 | 34.2 | 0.08 |
| pHILIC | 154.02 | 108.8639 | 34.2 | 0.08 |
| pHILIC | 154.02 | 83.5803 | 34.1 | 0.08 |
| pHILIC | 154.02 | 50.8771 | 34.1 | 0.08 |
| pHILIC | 154.02 | 79.8407 | 34.1 | 0.08 |
| pHILIC | 154.02 | 49.0091 | 34.1 | 0.08 |
| pHILIC | 154.02 | 58.0028 | 34.1 | 0.08 |
| pHILIC | 154.02 | 40.3676 | 34.1 | 0.08 |
| pHILIC | 154.02 | 141.6466 | 34.1 | 0.08 |
| pHILIC | 154.02 | 43.3594 | 34.1 | 0.08 |
| pHILIC | 154.02 | 42.932 | 34.1 | 0.08 |
| pHILIC | 154.02 | 147.0332 | 34.1 | 0.08 |
| pHILIC | 154.02 | 49.5864 | 34.1 | 0.08 |
| pHILIC | 154.02 | 64.4845 | 34.1 | 0.08 |
| pHILIC | 154.02 | 51.9234 | 34 | 0.08 |
| pHILIC | 154.02 | 144.3722 | 34 | 0.08 |
| pHILIC | 154.02 | 61.2027 | 34 | 0.08 |
| pHILIC | 154.02 | 52.8499 | 34 | 0.08 |
| pHILIC | 154.02 | 62.1653 | 34 | 0.08 |
| pHILIC | 154.02 | 138.133 | 34 | 0.08 |

| | | | | |
|---|---|---|---|---|
| pHILIC | 154.02 | 44.779 | 34 | 0.08 |
| pHILIC | 154.02 | 58.9874 | 34 | 0.08 |
| pHILIC | 154.02 | 60.7751 | 33.9 | 0.08 |
| pHILIC | 154.02 | 54.8405 | 33.9 | 0.08 |
| pHILIC | 154.02 | 40.9251 | 33.9 | 0.08 |
| pHILIC | 154.02 | 99.2191 | 33.9 | 0.08 |
| pHILIC | 154.02 | 68.9335 | 33.9 | 0.08 |
| pHILIC | 154.02 | 134.7662 | 33.9 | 0.08 |
| pHILIC | 154.02 | 75.8369 | 33.9 | 0.08 |
| pHILIC | 154.02 | 94.4737 | 33.9 | 0.08 |
| pHILIC | 154.02 | 61.3533 | 33.8 | 0.08 |
| pHILIC | 154.02 | 83.0651 | 33.8 | 0.08 |
| pHILIC | 154.02 | 65.6062 | 33.8 | 0.08 |
| pHILIC | 154.02 | 94.9459 | 33.8 | 0.08 |
| pHILIC | 154.02 | 119.1939 | 33.8 | 0.08 |
| pHILIC | 154.02 | 44.4257 | 33.8 | 0.08 |
| pHILIC | 154.02 | 53.1171 | 33.8 | 0.08 |
| pHILIC | 154.02 | 48.7291 | 33.7 | 0.08 |
| pHILIC | 154.02 | 140.8983 | 33.7 | 0.08 |
| pHILIC | 154.02 | 43.3705 | 33.7 | 0.08 |
| pHILIC | 154.02 | 41.6172 | 33.7 | 0.08 |
| pHILIC | 154.02 | 103.6641 | 33.7 | 0.08 |
| pHILIC | 154.02 | 137.7085 | 33.7 | 0.08 |
| pHILIC | 154.02 | 66.4265 | 33.7 | 0.08 |
| pHILIC | 154.02 | 45.9688 | 33.7 | 0.08 |
| pHILIC | 154.02 | 42.6735 | 33.7 | 0.08 |
| pHILIC | 154.02 | 158.0257 | 33.7 | 0.08 |
| pHILIC | 154.02 | 133.5726 | 33.7 | 0.08 |
| pHILIC | 154.02 | 44.1658 | 33.7 | 0.08 |
| pHILIC | 154.02 | 90.2374 | 33.6 | 0.08 |
| pHILIC | 154.02 | 43.2027 | 33.6 | 0.08 |
| pHILIC | 154.02 | 74.6861 | 33.6 | 0.08 |
| pHILIC | 154.02 | 105.1945 | 33.6 | 0.08 |
| pHILIC | 154.02 | 176.5759 | 33.6 | 0.08 |
| pHILIC | 154.02 | 71.0685 | 33.6 | 0.08 |
| pHILIC | 154.02 | 42.8098 | 33.6 | 0.08 |
| pHILIC | 154.02 | 166.2335 | 33.6 | 0.08 |
| pHILIC | 154.02 | 49.1939 | 33.6 | 0.08 |
| pHILIC | 154.02 | 45.2287 | 33.6 | 0.08 |
| pHILIC | 154.02 | 157.7712 | 33.6 | 0.08 |
| pHILIC | 154.02 | 66.7255 | 33.6 | 0.08 |
| pHILIC | 154.02 | 122.6406 | 33.6 | 0.08 |

| | | | | |
|---|---|---|---|---|
| pHILIC | 154.02 | 66.988 | 33.6 | 0.08 |
| pHILIC | 154.02 | 54.9453 | 33.6 | 0.08 |
| pHILIC | 154.02 | 109.2555 | 33.5 | 0.08 |
| pHILIC | 154.02 | 101.4406 | 33.5 | 0.08 |
| pHILIC | 154.02 | 87.4112 | 33.5 | 0.08 |
| pHILIC | 154.02 | 56.7996 | 33.5 | 0.08 |
| pHILIC | 154.02 | 92.9792 | 33.5 | 0.08 |
| pHILIC | 154.02 | 132.4671 | 33.5 | 0.08 |
| pHILIC | 154.02 | 140.1033 | 33.5 | 0.08 |
| pHILIC | 154.02 | 65.2387 | 33.5 | 0.08 |
| pHILIC | 154.02 | 93.6288 | 33.5 | 0.08 |
| pHILIC | 154.02 | 62.1913 | 33.5 | 0.08 |
| pHILIC | 154.02 | 98.7666 | 33.4 | 0.08 |
| pHILIC | 154.02 | 54.8355 | 33.4 | 0.08 |
| pHILIC | 154.02 | 118.6379 | 33.4 | 0.08 |
| pHILIC | 154.02 | 144.9333 | 33.4 | 0.08 |
| pHILIC | 154.02 | 86.3585 | 33.4 | 0.08 |
| pHILIC | 154.02 | 79.3981 | 33.4 | 0.08 |
| pHILIC | 154.02 | 75.7655 | 33.4 | 0.08 |
| pHILIC | 154.02 | 172.0266 | 33.4 | 0.08 |
| pHILIC | 154.02 | 63.5477 | 33.3 | 0.08 |
| pHILIC | 154.02 | 161.055 | 33.3 | 0.08 |
| pHILIC | 154.02 | 61.2215 | 33.3 | 0.08 |
| pHILIC | 154.02 | 82.0641 | 33.2 | 0.08 |
| pHILIC | 154.02 | 150.5687 | 33.2 | 0.08 |
| pHILIC | 154.02 | 120.1441 | 33.2 | 0.08 |
| pHILIC | 154.02 | 110.065 | 33.2 | 0.08 |
| pHILIC | 154.02 | 77.3387 | 33.2 | 0.08 |
| pHILIC | 154.02 | 133.5561 | 33.2 | 0.08 |
| pHILIC | 154.02 | 63.2884 | 33.2 | 0.08 |
| pHILIC | 154.02 | 44.8034 | 33.2 | 0.08 |
| pHILIC | 154.02 | 72.2507 | 33.2 | 0.08 |
| pHILIC | 154.02 | 165.7378 | 33.2 | 0.08 |
| pHILIC | 154.02 | 80.7834 | 33.2 | 0.08 |
| pHILIC | 154.02 | 61.9022 | 33.2 | 0.08 |
| pHILIC | 154.02 | 70.3618 | 33.2 | 0.08 |
| pHILIC | 154.02 | 72.9438 | 33.2 | 0.08 |
| pHILIC | 154.02 | 131.2867 | 33.2 | 0.08 |
| pHILIC | 154.02 | 129.7066 | 33.2 | 0.08 |
| pHILIC | 154.02 | 68.5482 | 33.2 | 0.08 |
| pHILIC | 154.02 | 46.9449 | 33.2 | 0.08 |
| pHILIC | 154.02 | 106.8462 | 33.1 | 0.08 |

| | | | | |
|---|---|---|---|---|
| pHILIC | 154.02 | 76.1507 | 33.1 | 0.08 |
| pHILIC | 154.02 | 62.0164 | 33.1 | 0.08 |
| pHILIC | 154.02 | 48.324 | 33.1 | 0.08 |
| pHILIC | 154.02 | 153.0476 | 33.1 | 0.08 |
| pHILIC | 154.02 | 91.255 | 33.1 | 0.08 |
| pHILIC | 154.02 | 46.6735 | 33.1 | 0.08 |
| pHILIC | 154.02 | 153.1234 | 33.1 | 0.08 |
| pHILIC | 154.02 | 71.9167 | 33.1 | 0.08 |
| pHILIC | 154.02 | 72.4887 | 33.1 | 0.08 |
| pHILIC | 154.02 | 154.0698 | 33.1 | 0.08 |
| pHILIC | 154.02 | 103.8169 | 33.1 | 0.08 |
| pHILIC | 154.02 | 57.2922 | 33 | 0.08 |
| pHILIC | 154.02 | 46.8305 | 33 | 0.08 |
| pHILIC | 154.02 | 158.2611 | 33 | 0.08 |
| pHILIC | 154.02 | 73.2974 | 33 | 0.08 |
| pHILIC | 154.02 | 126.9869 | 33 | 0.08 |
| pHILIC | 154.02 | 73.8275 | 33 | 0.08 |
| pHILIC | 154.02 | 131.57 | 33 | 0.08 |
| pHILIC | 154.02 | 126.8893 | 33 | 0.08 |
| pHILIC | 154.02 | 138.417 | 33 | 0.08 |
| pHILIC | 154.02 | 164.096 | 33 | 0.08 |
| pHILIC | 154.02 | 49.8799 | 33 | 0.08 |
| pHILIC | 154.02 | 165.3104 | 33 | 0.08 |
| pHILIC | 154.02 | 79.9646 | 33 | 0.08 |
| pHILIC | 154.02 | 70.2835 | 32.9 | 0.08 |
| pHILIC | 154.02 | 41.0751 | 32.9 | 0.08 |
| pHILIC | 154.02 | 88.7042 | 32.9 | 0.08 |
| pHILIC | 154.02 | 93.8871 | 32.9 | 0.08 |
| pHILIC | 154.02 | 54.1472 | 32.9 | 0.08 |
| pHILIC | 154.02 | 132.4907 | 32.9 | 0.08 |
| pHILIC | 154.02 | 58.4123 | 32.9 | 0.08 |
| pHILIC | 154.02 | 117.2856 | 32.9 | 0.08 |
| pHILIC | 154.02 | 73.6526 | 32.9 | 0.08 |
| pHILIC | 154.02 | 79.6846 | 32.9 | 0.08 |
| pHILIC | 154.02 | 56.9893 | 32.9 | 0.08 |
| pHILIC | 154.02 | 133.0096 | 32.9 | 0.08 |
| pHILIC | 154.02 | 40.1516 | 32.8 | 0.08 |
| pHILIC | 154.02 | 124.615 | 32.8 | 0.08 |
| pHILIC | 154.02 | 173.6833 | 32.8 | 0.08 |
| pHILIC | 154.02 | 63.6181 | 32.8 | 0.08 |
| pHILIC | 154.02 | 159.6243 | 32.8 | 0.08 |
| pHILIC | 154.02 | 50.2016 | 32.8 | 0.08 |

| | | | | |
|---|---|---|---|---|
| pHILIC | 154.02 | 87.1748 | 32.8 | 0.08 |
| pHILIC | 154.02 | 42.1394 | 32.8 | 0.08 |
| pHILIC | 154.02 | 44.9204 | 32.8 | 0.08 |
| pHILIC | 154.02 | 160.3807 | 32.8 | 0.08 |
| pHILIC | 154.02 | 113.3405 | 32.8 | 0.07 |
| pHILIC | 154.02 | 164.5814 | 32.8 | 0.07 |
| pHILIC | 154.02 | 54.8505 | 32.8 | 0.07 |
| pHILIC | 154.02 | 53.7985 | 32.8 | 0.07 |
| pHILIC | 154.02 | 66.5019 | 32.8 | 0.07 |
| pHILIC | 154.02 | 60.6702 | 32.7 | 0.07 |
| pHILIC | 154.02 | 146.4321 | 32.7 | 0.07 |
| pHILIC | 154.02 | 148.0864 | 32.7 | 0.07 |
| pHILIC | 154.02 | 153.0791 | 32.7 | 0.07 |
| pHILIC | 154.02 | 49.5662 | 32.7 | 0.07 |
| pHILIC | 154.02 | 64.377 | 32.7 | 0.07 |
| pHILIC | 154.02 | 133.3309 | 32.6 | 0.07 |
| pHILIC | 154.02 | 96.7609 | 32.6 | 0.07 |
| pHILIC | 154.02 | 114.7002 | 32.6 | 0.07 |
| pHILIC | 154.02 | 63.0768 | 32.6 | 0.07 |
| pHILIC | 154.02 | 103.9542 | 32.6 | 0.07 |
| pHILIC | 154.02 | 63.8287 | 32.6 | 0.07 |
| pHILIC | 154.02 | 164.3373 | 32.6 | 0.07 |
| pHILIC | 154.02 | 75.6137 | 32.6 | 0.07 |
| pHILIC | 154.02 | 67.0361 | 32.6 | 0.07 |
| pHILIC | 154.02 | 69.1991 | 32.6 | 0.07 |
| pHILIC | 154.02 | 52.7667 | 32.6 | 0.07 |
| pHILIC | 154.02 | 79.8104 | 32.6 | 0.07 |
| pHILIC | 154.02 | 42.55 | 32.6 | 0.07 |
| pHILIC | 154.02 | 105.8295 | 32.6 | 0.07 |
| pHILIC | 154.02 | 92.4431 | 32.6 | 0.07 |
| pHILIC | 154.02 | 62.5929 | 32.6 | 0.07 |
| pHILIC | 154.02 | 101.6045 | 32.6 | 0.07 |
| pHILIC | 154.02 | 171.951 | 32.6 | 0.07 |
| pHILIC | 154.02 | 101.2463 | 32.5 | 0.07 |
| pHILIC | 154.02 | 85.6927 | 32.5 | 0.07 |
| pHILIC | 154.02 | 56.2863 | 32.5 | 0.07 |
| pHILIC | 154.02 | 86.6761 | 32.5 | 0.07 |
| pHILIC | 154.02 | 77.2943 | 32.5 | 0.07 |
| pHILIC | 154.02 | 50.804 | 32.5 | 0.07 |
| pHILIC | 154.02 | 93.1622 | 32.5 | 0.07 |
| pHILIC | 154.02 | 155.0272 | 32.5 | 0.07 |
| pHILIC | 154.02 | 177.4539 | 32.5 | 0.07 |

| | | | | |
|---|---|---|---|---|
| pHILIC | 154.02 | 54.6154 | 32.5 | 0.07 |
| pHILIC | 154.02 | 66.0691 | 32.5 | 0.07 |
| pHILIC | 154.02 | 144.6424 | 32.5 | 0.07 |
| pHILIC | 154.02 | 48.4978 | 32.5 | 0.07 |
| pHILIC | 154.02 | 68.117 | 32.5 | 0.07 |
| pHILIC | 154.02 | 50.9741 | 32.5 | 0.07 |
| pHILIC | 154.02 | 40.1472 | 32.5 | 0.07 |
| pHILIC | 154.02 | 77.4533 | 32.5 | 0.07 |
| pHILIC | 154.02 | 46.6136 | 32.4 | 0.07 |
| pHILIC | 154.02 | 86.2953 | 32.4 | 0.07 |
| pHILIC | 154.02 | 114.376 | 32.4 | 0.07 |
| pHILIC | 154.02 | 93.3815 | 32.4 | 0.07 |
| pHILIC | 154.02 | 138.5911 | 32.4 | 0.07 |
| pHILIC | 154.02 | 90.2296 | 32.4 | 0.07 |
| pHILIC | 154.02 | 59.974 | 32.4 | 0.07 |
| pHILIC | 154.02 | 70.4475 | 32.4 | 0.07 |
| pHILIC | 154.02 | 41.143 | 32.4 | 0.07 |
| pHILIC | 154.02 | 45.962 | 32.4 | 0.07 |
| pHILIC | 154.02 | 53.758 | 32.4 | 0.07 |
| pHILIC | 154.02 | 42.7753 | 32.4 | 0.07 |
| pHILIC | 154.02 | 73.3069 | 32.4 | 0.07 |
| pHILIC | 154.02 | 134.7168 | 32.4 | 0.07 |
| pHILIC | 154.02 | 52.103 | 32.3 | 0.07 |
| pHILIC | 154.02 | 51.4899 | 32.3 | 0.07 |
| pHILIC | 154.02 | 75.2192 | 32.3 | 0.07 |
| pHILIC | 154.02 | 91.1274 | 32.3 | 0.07 |
| pHILIC | 154.02 | 79.7581 | 32.3 | 0.07 |
| pHILIC | 154.02 | 59.1031 | 32.3 | 0.07 |
| pHILIC | 154.02 | 66.2582 | 32.3 | 0.07 |
| pHILIC | 154.02 | 65.2078 | 32.3 | 0.07 |
| pHILIC | 154.02 | 56.9094 | 32.3 | 0.07 |
| pHILIC | 154.02 | 85.2132 | 32.3 | 0.07 |
| pHILIC | 154.02 | 97.8075 | 32.3 | 0.07 |
| pHILIC | 154.02 | 125.6843 | 32.3 | 0.07 |
| pHILIC | 154.02 | 103.0349 | 32.3 | 0.07 |
| pHILIC | 154.02 | 56.4122 | 32.3 | 0.07 |
| pHILIC | 154.02 | 114.3644 | 32.3 | 0.07 |
| pHILIC | 154.02 | 66.9247 | 32.2 | 0.07 |
| pHILIC | 154.02 | 145.7075 | 32.2 | 0.07 |
| pHILIC | 154.02 | 62.6794 | 32.2 | 0.07 |
| pHILIC | 154.02 | 55.0803 | 32.2 | 0.07 |
| pHILIC | 154.02 | 86.5698 | 32.2 | 0.07 |

| | | | | |
|---|---|---|---|---|
| pHILIC | 154.02 | 151.3458 | 32.2 | 0.07 |
| pHILIC | 154.02 | 140.6366 | 32.2 | 0.07 |
| pHILIC | 154.02 | 72.0473 | 32.2 | 0.07 |
| pHILIC | 154.02 | 41.5963 | 32.2 | 0.07 |
| pHILIC | 154.02 | 154.7525 | 32.2 | 0.07 |
| pHILIC | 154.02 | 124.9579 | 32.2 | 0.07 |
| pHILIC | 154.02 | 78.2634 | 32.2 | 0.07 |
| pHILIC | 154.02 | 63.9253 | 32.2 | 0.07 |
| pHILIC | 154.02 | 88.3369 | 32.2 | 0.07 |
| pHILIC | 154.02 | 45.9099 | 32.2 | 0.07 |
| pHILIC | 154.02 | 73.0707 | 32.2 | 0.07 |
| pHILIC | 154.02 | 77.2663 | 32.2 | 0.07 |
| pHILIC | 154.02 | 128.7384 | 32.2 | 0.07 |
| pHILIC | 154.02 | 45.9379 | 32.1 | 0.07 |
| pHILIC | 154.02 | 73.3467 | 32.1 | 0.07 |
| pHILIC | 154.02 | 59.8319 | 32.1 | 0.07 |
| pHILIC | 154.02 | 118.5738 | 32.1 | 0.07 |
| pHILIC | 154.02 | 115.0916 | 32.1 | 0.07 |
| pHILIC | 154.02 | 142.7306 | 32.1 | 0.07 |
| pHILIC | 154.02 | 94.0042 | 32.1 | 0.07 |
| pHILIC | 154.02 | 50.745 | 32.1 | 0.07 |
| pHILIC | 154.02 | 110.1768 | 32.1 | 0.07 |
| pHILIC | 154.02 | 112.7605 | 32.1 | 0.07 |
| pHILIC | 154.02 | 66.2251 | 32.1 | 0.07 |
| pHILIC | 154.02 | 85.9392 | 32.1 | 0.07 |
| pHILIC | 154.02 | 102.1673 | 32.1 | 0.07 |
| pHILIC | 154.02 | 43.894 | 32.1 | 0.07 |
| pHILIC | 154.02 | 74.69 | 32 | 0.07 |
| pHILIC | 154.02 | 54.5062 | 32 | 0.07 |
| pHILIC | 154.02 | 123.8769 | 32 | 0.07 |
| pHILIC | 154.02 | 92.7607 | 32 | 0.07 |
| pHILIC | 154.02 | 58.5985 | 32 | 0.07 |
| pHILIC | 154.02 | 45.3228 | 32 | 0.07 |
| pHILIC | 154.02 | 112.4312 | 32 | 0.07 |
| pHILIC | 154.02 | 52.9928 | 32 | 0.07 |
| pHILIC | 154.02 | 89.8217 | 32 | 0.07 |
| pHILIC | 154.02 | 76.0498 | 32 | 0.07 |
| pHILIC | 154.02 | 175.9994 | 32 | 0.07 |
| pHILIC | 154.02 | 90.5576 | 32 | 0.07 |
| pHILIC | 154.02 | 140.4753 | 32 | 0.07 |
| pHILIC | 154.02 | 51.6865 | 32 | 0.07 |
| pHILIC | 154.02 | 84.538 | 32 | 0.07 |

| | | | | |
|---|---|---|---|---|
| pHILIC | 154.02 | 55.122 | 32 | 0.07 |
| pHILIC | 154.02 | 91.3306 | 32 | 0.07 |
| pHILIC | 154.02 | 102.1028 | 32 | 0.07 |
| pHILIC | 154.02 | 157.1694 | 32 | 0.07 |
| pHILIC | 154.02 | 60.8163 | 31.9 | 0.07 |
| pHILIC | 154.02 | 51.1732 | 31.9 | 0.07 |
| pHILIC | 154.02 | 83.7901 | 31.9 | 0.07 |
| pHILIC | 154.02 | 51.24 | 31.9 | 0.07 |
| pHILIC | 154.02 | 121.2225 | 31.9 | 0.07 |
| pHILIC | 154.02 | 54.0821 | 31.9 | 0.07 |
| pHILIC | 154.02 | 47.0222 | 31.9 | 0.07 |
| pHILIC | 154.02 | 62.4157 | 31.9 | 0.07 |
| pHILIC | 154.02 | 73.6904 | 31.9 | 0.07 |
| pHILIC | 154.02 | 65.6882 | 31.9 | 0.07 |
| pHILIC | 154.02 | 44.1392 | 31.9 | 0.07 |
| pHILIC | 154.02 | 42.4781 | 31.9 | 0.07 |
| pHILIC | 154.02 | 45.9534 | 31.9 | 0.07 |
| pHILIC | 154.02 | 77.8499 | 31.8 | 0.07 |
| pHILIC | 154.02 | 168.0533 | 31.8 | 0.07 |
| pHILIC | 154.02 | 77.1619 | 31.8 | 0.07 |
| pHILIC | 154.02 | 122.2175 | 31.8 | 0.07 |
| pHILIC | 154.02 | 99.4049 | 31.8 | 0.07 |
| pHILIC | 154.02 | 52.6463 | 31.8 | 0.07 |
| pHILIC | 154.02 | 58.1089 | 31.8 | 0.07 |
| pHILIC | 154.02 | 62.6387 | 31.8 | 0.07 |
| pHILIC | 154.02 | 57.2579 | 31.8 | 0.07 |
| pHILIC | 154.02 | 51.4414 | 31.8 | 0.07 |
| pHILIC | 154.02 | 119.573 | 31.8 | 0.07 |
| pHILIC | 154.02 | 99.0365 | 31.8 | 0.07 |
| pHILIC | 154.02 | 67.105 | 31.8 | 0.07 |
| pHILIC | 154.02 | 47.2812 | 31.8 | 0.07 |
| pHILIC | 154.02 | 45.7019 | 31.8 | 0.07 |
| pHILIC | 154.02 | 91.7891 | 31.8 | 0.07 |
| pHILIC | 154.02 | 75.7438 | 31.8 | 0.07 |
| pHILIC | 154.02 | 98.8103 | 31.7 | 0.07 |
| pHILIC | 154.02 | 44.2808 | 31.7 | 0.07 |
| pHILIC | 154.02 | 40.5146 | 31.7 | 0.07 |
| pHILIC | 154.02 | 61.526 | 31.7 | 0.07 |
| pHILIC | 154.02 | 46.829 | 31.7 | 0.07 |
| pHILIC | 154.02 | 74.485 | 31.7 | 0.07 |
| pHILIC | 154.02 | 59.9394 | 31.7 | 0.07 |
| pHILIC | 154.02 | 131.7557 | 31.7 | 0.07 |

| | | | | |
|---|---|---|---|---|
| pHILIC | 154.02 | 146.9592 | 31.7 | 0.07 |
| pHILIC | 154.02 | 64.493 | 31.7 | 0.07 |
| pHILIC | 154.02 | 46.4866 | 31.7 | 0.07 |
| pHILIC | 154.02 | 44.5817 | 31.7 | 0.07 |
| pHILIC | 154.02 | 102.6468 | 31.7 | 0.07 |
| pHILIC | 154.02 | 49.3161 | 31.7 | 0.07 |
| pHILIC | 154.02 | 109.0976 | 31.6 | 0.07 |
| pHILIC | 154.02 | 45.9083 | 31.6 | 0.07 |
| pHILIC | 154.02 | 167.4641 | 31.6 | 0.07 |
| pHILIC | 154.02 | 41.513 | 31.6 | 0.07 |
| pHILIC | 154.02 | 102.1121 | 31.6 | 0.07 |
| pHILIC | 154.02 | 63.7528 | 31.6 | 0.07 |
| pHILIC | 154.02 | 131.9444 | 31.6 | 0.07 |
| pHILIC | 154.02 | 60.4739 | 31.6 | 0.07 |
| pHILIC | 154.02 | 139.7766 | 31.6 | 0.07 |
| pHILIC | 154.02 | 76.2754 | 31.6 | 0.07 |
| pHILIC | 154.02 | 55.617 | 31.6 | 0.07 |
| pHILIC | 154.02 | 88.5636 | 31.6 | 0.07 |
| pHILIC | 154.02 | 82.0127 | 31.6 | 0.07 |
| pHILIC | 154.02 | 125.1369 | 31.6 | 0.07 |
| pHILIC | 154.02 | 54.8998 | 31.6 | 0.07 |
| pHILIC | 154.02 | 110.9205 | 31.6 | 0.07 |
| pHILIC | 154.02 | 90.0839 | 31.5 | 0.07 |
| pHILIC | 154.02 | 112.1085 | 31.5 | 0.07 |
| pHILIC | 154.02 | 75.3753 | 31.5 | 0.07 |
| pHILIC | 154.02 | 170.2413 | 31.5 | 0.07 |
| pHILIC | 154.02 | 155.1486 | 31.5 | 0.07 |
| pHILIC | 154.02 | 43.9785 | 31.5 | 0.07 |
| pHILIC | 154.02 | 83.8235 | 31.5 | 0.07 |
| pHILIC | 154.02 | 144.7743 | 31.5 | 0.07 |
| pHILIC | 154.02 | 45.1052 | 31.5 | 0.07 |
| pHILIC | 154.02 | 110.8508 | 31.5 | 0.07 |
| pHILIC | 154.02 | 43.5777 | 31.5 | 0.07 |
| pHILIC | 154.02 | 124.3851 | 31.5 | 0.07 |
| pHILIC | 154.02 | 52.9921 | 31.5 | 0.07 |
| pHILIC | 154.02 | 121.6804 | 31.5 | 0.07 |
| pHILIC | 154.02 | 83.3291 | 31.5 | 0.07 |
| pHILIC | 154.02 | 78.416 | 31.5 | 0.07 |
| pHILIC | 154.02 | 79.6907 | 31.5 | 0.07 |
| pHILIC | 154.02 | 41.7791 | 31.5 | 0.07 |
| pHILIC | 154.02 | 94.2642 | 31.5 | 0.07 |
| pHILIC | 154.02 | 63.4463 | 31.5 | 0.07 |

| | | | | |
|---|---|---|---|---|
| pHILIC | 154.02 | 43.0077 | 31.5 | 0.07 |
| pHILIC | 154.02 | 100.0372 | 31.4 | 0.07 |
| pHILIC | 154.02 | 48.0395 | 31.4 | 0.07 |
| pHILIC | 154.02 | 66.1062 | 31.4 | 0.07 |
| pHILIC | 154.02 | 71.5883 | 31.4 | 0.07 |
| pHILIC | 154.02 | 95.8842 | 31.4 | 0.07 |
| pHILIC | 154.02 | 53.8607 | 31.4 | 0.07 |
| pHILIC | 154.02 | 73.3581 | 31.4 | 0.07 |
| pHILIC | 154.02 | 48.1548 | 31.4 | 0.07 |
| pHILIC | 154.02 | 87.8684 | 31.4 | 0.07 |
| pHILIC | 154.02 | 46.5247 | 31.4 | 0.07 |
| pHILIC | 154.02 | 62.0612 | 31.4 | 0.07 |
| pHILIC | 154.02 | 58.5271 | 31.4 | 0.07 |
| pHILIC | 154.02 | 50.4428 | 31.4 | 0.07 |
| pHILIC | 154.02 | 99.8946 | 31.4 | 0.07 |
| pHILIC | 154.02 | 52.9525 | 31.4 | 0.07 |
| pHILIC | 154.02 | 113.0774 | 31.4 | 0.07 |
| pHILIC | 154.02 | 56.9171 | 31.4 | 0.07 |
| pHILIC | 154.02 | 56.6691 | 31.4 | 0.07 |
| pHILIC | 154.02 | 124.4382 | 31.4 | 0.07 |
| pHILIC | 154.02 | 76.0339 | 31.4 | 0.07 |
| pHILIC | 154.02 | 57.7439 | 31.3 | 0.07 |
| pHILIC | 154.02 | 46.5586 | 31.3 | 0.07 |
| pHILIC | 154.02 | 76.6704 | 31.3 | 0.07 |
| pHILIC | 154.02 | 40.8828 | 31.3 | 0.07 |
| pHILIC | 154.02 | 87.6919 | 31.3 | 0.07 |
| pHILIC | 154.02 | 105.1292 | 31.3 | 0.07 |
| pHILIC | 154.02 | 114.7154 | 31.3 | 0.07 |
| pHILIC | 154.02 | 107.1569 | 31.3 | 0.07 |
| pHILIC | 154.02 | 75.0502 | 31.3 | 0.07 |
| pHILIC | 154.02 | 96.3222 | 31.3 | 0.07 |
| pHILIC | 154.02 | 64.8344 | 31.3 | 0.07 |
| pHILIC | 154.02 | 57.9128 | 31.3 | 0.07 |
| pHILIC | 154.02 | 71.8772 | 31.3 | 0.07 |
| pHILIC | 154.02 | 110.6336 | 31.3 | 0.07 |
| pHILIC | 154.02 | 40.6564 | 31.3 | 0.07 |
| pHILIC | 154.02 | 44.6271 | 31.3 | 0.07 |
| pHILIC | 154.02 | 172.2673 | 31.3 | 0.07 |
| pHILIC | 154.02 | 65.108 | 31.3 | 0.07 |
| pHILIC | 154.02 | 154.1418 | 31.3 | 0.07 |
| pHILIC | 154.02 | 55.1516 | 31.3 | 0.07 |
| pHILIC | 154.02 | 165.9412 | 31.3 | 0.07 |

| | | | | |
|---|---|---|---|---|
| pHILIC | 154.02 | 70.686 | 31.3 | 0.07 |
| pHILIC | 154.02 | 44.2536 | 31.3 | 0.07 |
| pHILIC | 154.02 | 79.1736 | 31.3 | 0.07 |
| pHILIC | 154.02 | 59.2499 | 31.3 | 0.07 |
| pHILIC | 154.02 | 72.6828 | 31.3 | 0.07 |
| pHILIC | 154.02 | 49.8584 | 31.2 | 0.07 |
| pHILIC | 154.02 | 78.6719 | 31.2 | 0.07 |
| pHILIC | 154.02 | 43.2831 | 31.2 | 0.07 |
| pHILIC | 154.02 | 175.7331 | 31.2 | 0.07 |
| pHILIC | 154.02 | 106.3858 | 31.2 | 0.07 |
| pHILIC | 154.02 | 49.2863 | 31.2 | 0.07 |
| pHILIC | 154.02 | 87.2354 | 31.2 | 0.07 |
| pHILIC | 154.02 | 76.4458 | 31.2 | 0.07 |
| pHILIC | 154.02 | 46.1895 | 31.2 | 0.07 |
| pHILIC | 154.02 | 50.3037 | 31.2 | 0.07 |
| pHILIC | 154.02 | 41.9316 | 31.2 | 0.07 |
| pHILIC | 154.02 | 105.0798 | 31.2 | 0.07 |
| pHILIC | 154.02 | 60.3739 | 31.2 | 0.07 |
| pHILIC | 154.02 | 88.8345 | 31.2 | 0.07 |
| pHILIC | 154.02 | 49.8046 | 31.2 | 0.07 |
| pHILIC | 154.02 | 51.0062 | 31.2 | 0.07 |
| pHILIC | 154.02 | 91.5286 | 31.2 | 0.07 |
| pHILIC | 154.02 | 79.5191 | 31.2 | 0.07 |
| pHILIC | 154.02 | 50.2957 | 31.2 | 0.07 |
| pHILIC | 154.02 | 154.9294 | 31.2 | 0.07 |
| pHILIC | 154.02 | 63.853 | 31.2 | 0.07 |
| pHILIC | 154.02 | 71.3979 | 31.2 | 0.07 |
| pHILIC | 154.02 | 59.1144 | 31.1 | 0.07 |
| pHILIC | 154.02 | 91.337 | 31.1 | 0.07 |
| pHILIC | 154.02 | 56.0276 | 31.1 | 0.07 |
| pHILIC | 154.02 | 65.5845 | 31.1 | 0.07 |
| pHILIC | 154.02 | 112.6958 | 31.1 | 0.07 |
| pHILIC | 154.02 | 40.7393 | 31.1 | 0.07 |
| pHILIC | 154.02 | 66.6135 | 31.1 | 0.07 |
| pHILIC | 154.02 | 135.1269 | 31.1 | 0.07 |
| pHILIC | 154.02 | 126.6267 | 31.1 | 0.07 |
| pHILIC | 154.02 | 42.6163 | 31.1 | 0.07 |
| pHILIC | 154.02 | 117.2697 | 31.1 | 0.07 |
| pHILIC | 154.02 | 43.4553 | 31.1 | 0.07 |
| pHILIC | 154.02 | 95.7842 | 31.1 | 0.07 |
| pHILIC | 154.02 | 43.0432 | 31.1 | 0.07 |
| pHILIC | 154.02 | 139.2742 | 31.1 | 0.07 |

| | | | | |
|---|---|---|---|---|
| pHILIC | 154.02 | 106.0864 | 31.1 | 0.07 |
| pHILIC | 154.02 | 116.7234 | 31.1 | 0.07 |
| pHILIC | 154.02 | 106.3505 | 31.1 | 0.07 |
| pHILIC | 154.02 | 86.9726 | 31.1 | 0.07 |
| pHILIC | 154.02 | 54.3813 | 31.1 | 0.07 |
| pHILIC | 154.02 | 119.5266 | 31.1 | 0.07 |
| pHILIC | 154.02 | 109.6553 | 31.1 | 0.07 |
| pHILIC | 154.02 | 72.2051 | 31.1 | 0.07 |
| pHILIC | 154.02 | 57.6055 | 31.1 | 0.07 |
| pHILIC | 154.02 | 73.3988 | 31.1 | 0.07 |
| pHILIC | 154.02 | 77.2246 | 31.1 | 0.07 |
| pHILIC | 154.02 | 64.1793 | 31.1 | 0.07 |
| pHILIC | 154.02 | 53.7799 | 31 | 0.07 |
| pHILIC | 154.02 | 100.4431 | 31 | 0.07 |
| pHILIC | 154.02 | 136.7014 | 31 | 0.07 |
| pHILIC | 154.02 | 93.9944 | 31 | 0.07 |
| pHILIC | 154.02 | 86.1939 | 31 | 0.07 |
| pHILIC | 154.02 | 46.4246 | 31 | 0.07 |
| pHILIC | 154.02 | 43.9065 | 31 | 0.07 |
| pHILIC | 154.02 | 43.9377 | 31 | 0.07 |
| pHILIC | 154.02 | 120.3194 | 31 | 0.07 |
| pHILIC | 154.02 | 49.6971 | 31 | 0.07 |
| pHILIC | 154.02 | 83.031 | 31 | 0.07 |
| pHILIC | 154.02 | 72.2685 | 31 | 0.07 |
| pHILIC | 154.02 | 100.3939 | 31 | 0.07 |
| pHILIC | 154.02 | 79.4001 | 31 | 0.07 |
| pHILIC | 154.02 | 65.7084 | 31 | 0.07 |
| pHILIC | 154.02 | 59.7 | 30.9 | 0.07 |
| pHILIC | 154.02 | 93.7825 | 30.9 | 0.07 |
| pHILIC | 154.02 | 42.8891 | 30.9 | 0.07 |
| pHILIC | 154.02 | 47.1633 | 30.9 | 0.07 |
| pHILIC | 154.02 | 52.4396 | 30.9 | 0.07 |
| pHILIC | 154.02 | 174.413 | 30.9 | 0.07 |
| pHILIC | 154.02 | 78.8004 | 30.9 | 0.07 |
| pHILIC | 154.02 | 40.0632 | 30.9 | 0.07 |
| pHILIC | 154.02 | 121.8208 | 30.9 | 0.07 |
| pHILIC | 154.02 | 46.9207 | 30.9 | 0.07 |
| pHILIC | 154.02 | 47.9419 | 30.9 | 0.07 |
| pHILIC | 154.02 | 79.6082 | 30.9 | 0.07 |
| pHILIC | 154.02 | 90.4956 | 30.9 | 0.07 |
| pHILIC | 154.02 | 58.5072 | 30.9 | 0.07 |
| pHILIC | 154.02 | 61.5615 | 30.9 | 0.07 |

| | | | | |
|---|---|---|---|---|
| pHILIC | 154.02 | 55.069 | 30.9 | 0.07 |
| pHILIC | 154.02 | 158.0475 | 30.9 | 0.07 |
| pHILIC | 154.02 | 154.5396 | 30.9 | 0.07 |
| pHILIC | 154.02 | 48.8447 | 30.9 | 0.07 |
| pHILIC | 154.02 | 114.6662 | 30.8 | 0.07 |
| pHILIC | 154.02 | 66.2021 | 30.8 | 0.07 |
| pHILIC | 154.02 | 53.9926 | 30.8 | 0.07 |
| pHILIC | 154.02 | 47.627 | 30.8 | 0.07 |
| pHILIC | 154.02 | 47.8241 | 30.8 | 0.07 |
| pHILIC | 154.02 | 91.9361 | 30.8 | 0.07 |
| pHILIC | 154.02 | 69.1222 | 30.8 | 0.07 |
| pHILIC | 154.02 | 82.5941 | 30.8 | 0.07 |
| pHILIC | 154.02 | 57.6848 | 30.8 | 0.07 |
| pHILIC | 154.02 | 129.3398 | 30.8 | 0.07 |
| pHILIC | 154.02 | 151.6728 | 30.8 | 0.07 |
| pHILIC | 154.02 | 57.7653 | 30.8 | 0.07 |
| pHILIC | 154.02 | 45.6222 | 30.8 | 0.07 |
| pHILIC | 154.02 | 77.097 | 30.8 | 0.07 |
| pHILIC | 154.02 | 85.5415 | 30.8 | 0.07 |
| pHILIC | 154.02 | 130.1843 | 30.8 | 0.07 |
| pHILIC | 154.02 | 42.083 | 30.8 | 0.07 |
| pHILIC | 154.02 | 57.1489 | 30.8 | 0.07 |
| pHILIC | 154.02 | 156.8203 | 30.8 | 0.07 |
| pHILIC | 154.02 | 166.6679 | 30.8 | 0.07 |
| pHILIC | 154.02 | 126.8369 | 30.7 | 0.07 |
| pHILIC | 154.02 | 82.4716 | 30.7 | 0.07 |
| pHILIC | 154.02 | 79.6733 | 30.7 | 0.07 |
| pHILIC | 154.02 | 110.9588 | 30.7 | 0.07 |
| pHILIC | 154.02 | 53.2926 | 30.7 | 0.07 |
| pHILIC | 154.02 | 60.2889 | 30.7 | 0.07 |
| pHILIC | 154.02 | 43.0936 | 30.7 | 0.07 |
| pHILIC | 154.02 | 61.8858 | 30.7 | 0.07 |
| pHILIC | 154.02 | 46.7141 | 30.7 | 0.07 |
| pHILIC | 154.02 | 55.1654 | 30.7 | 0.07 |
| pHILIC | 154.02 | 66.7313 | 30.7 | 0.07 |
| pHILIC | 154.02 | 84.5514 | 30.7 | 0.07 |
| pHILIC | 154.02 | 136.6342 | 30.7 | 0.07 |
| pHILIC | 154.02 | 60.8311 | 30.7 | 0.07 |
| pHILIC | 154.02 | 88.8024 | 30.7 | 0.07 |
| pHILIC | 154.02 | 56.2452 | 30.7 | 0.07 |
| pHILIC | 154.02 | 42.7133 | 30.7 | 0.07 |
| pHILIC | 154.02 | 120.4463 | 30.7 | 0.07 |

| | | | | |
|---|---|---|---|---|
| pHILIC | 154.02 | 69.4015 | 30.7 | 0.07 |
| pHILIC | 154.02 | 75.3315 | 30.6 | 0.07 |
| pHILIC | 154.02 | 119.6563 | 30.6 | 0.07 |
| pHILIC | 154.02 | 49.4644 | 30.6 | 0.07 |
| pHILIC | 154.02 | 47.5152 | 30.6 | 0.07 |
| pHILIC | 154.02 | 113.7973 | 30.6 | 0.07 |
| pHILIC | 154.02 | 149.2511 | 30.6 | 0.07 |
| pHILIC | 154.02 | 71.2538 | 30.6 | 0.07 |
| pHILIC | 154.02 | 72.3603 | 30.6 | 0.07 |
| pHILIC | 154.02 | 154.1595 | 30.6 | 0.07 |
| pHILIC | 154.02 | 71.2108 | 30.6 | 0.07 |
| pHILIC | 154.02 | 119.5577 | 30.6 | 0.07 |
| pHILIC | 154.02 | 122.8323 | 30.6 | 0.07 |
| pHILIC | 154.02 | 143.0518 | 30.6 | 0.07 |
| pHILIC | 154.02 | 62.1049 | 30.6 | 0.07 |
| pHILIC | 154.02 | 134.0435 | 30.6 | 0.07 |
| pHILIC | 154.02 | 68.1676 | 30.6 | 0.07 |
| pHILIC | 154.02 | 55.2917 | 30.6 | 0.07 |
| pHILIC | 154.02 | 42.158 | 30.6 | 0.07 |
| pHILIC | 154.02 | 83.5182 | 30.5 | 0.07 |
| pHILIC | 154.02 | 86.9696 | 30.5 | 0.07 |
| pHILIC | 154.02 | 129.5535 | 30.5 | 0.07 |
| pHILIC | 154.02 | 110.904 | 30.5 | 0.07 |
| pHILIC | 154.02 | 89.0199 | 30.5 | 0.07 |
| pHILIC | 154.02 | 92.7466 | 30.5 | 0.07 |
| pHILIC | 154.02 | 60.22 | 30.5 | 0.07 |
| pHILIC | 154.02 | 101.3529 | 30.5 | 0.07 |
| pHILIC | 154.02 | 147.1199 | 30.5 | 0.07 |
| pHILIC | 154.02 | 127.1726 | 30.5 | 0.07 |
| pHILIC | 154.02 | 137.3329 | 30.5 | 0.07 |
| pHILIC | 154.02 | 158.1747 | 30.5 | 0.07 |
| pHILIC | 154.02 | 45.4526 | 30.5 | 0.07 |
| pHILIC | 154.02 | 56.0685 | 30.5 | 0.07 |
| pHILIC | 154.02 | 131.7081 | 30.5 | 0.07 |
| pHILIC | 154.02 | 75.9609 | 30.5 | 0.07 |
| pHILIC | 154.02 | 72.5194 | 30.5 | 0.07 |
| pHILIC | 154.02 | 136.3219 | 30.5 | 0.07 |
| pHILIC | 154.02 | 157.7014 | 30.5 | 0.07 |
| pHILIC | 154.02 | 121.2802 | 30.5 | 0.07 |
| pHILIC | 154.02 | 133.8716 | 30.5 | 0.07 |
| pHILIC | 154.02 | 118.9602 | 30.5 | 0.07 |
| pHILIC | 154.02 | 123.5848 | 30.5 | 0.07 |

| | | | | |
|---|---|---|---|---|
| pHILIC | 154.02 | 81.5967 | 30.5 | 0.07 |
| pHILIC | 154.02 | 86.7435 | 30.5 | 0.07 |
| pHILIC | 154.02 | 80.3017 | 30.5 | 0.07 |
| pHILIC | 154.02 | 64.1016 | 30.5 | 0.07 |
| pHILIC | 154.02 | 86.2866 | 30.5 | 0.07 |
| pHILIC | 154.02 | 48.3927 | 30.5 | 0.07 |
| pHILIC | 154.02 | 73.302 | 30.5 | 0.07 |
| pHILIC | 154.02 | 86.6898 | 30.5 | 0.07 |
| pHILIC | 154.02 | 82.217 | 30.4 | 0.07 |
| pHILIC | 154.02 | 49.0473 | 30.4 | 0.07 |
| pHILIC | 154.02 | 42.1071 | 30.4 | 0.07 |
| pHILIC | 154.02 | 49.4935 | 30.4 | 0.07 |
| pHILIC | 154.02 | 118.4015 | 30.4 | 0.07 |
| pHILIC | 154.02 | 155.8723 | 30.4 | 0.07 |
| pHILIC | 154.02 | 40.4854 | 30.4 | 0.07 |
| pHILIC | 154.02 | 59.5723 | 30.4 | 0.07 |
| pHILIC | 154.02 | 145.3434 | 30.4 | 0.07 |
| pHILIC | 154.02 | 50.8327 | 30.4 | 0.07 |
| pHILIC | 154.02 | 63.2592 | 30.4 | 0.07 |
| pHILIC | 154.02 | 64.0065 | 30.4 | 0.07 |
| pHILIC | 154.02 | 75.7348 | 30.4 | 0.07 |
| pHILIC | 154.02 | 106.0536 | 30.4 | 0.07 |
| pHILIC | 154.02 | 138.893 | 30.4 | 0.07 |
| pHILIC | 154.02 | 107.6063 | 30.4 | 0.07 |
| pHILIC | 154.02 | 40.3084 | 30.4 | 0.07 |
| pHILIC | 154.02 | 49.1865 | 30.4 | 0.07 |
| pHILIC | 154.02 | 53.1134 | 30.4 | 0.07 |
| pHILIC | 154.02 | 43.9719 | 30.4 | 0.07 |
| pHILIC | 154.02 | 104.1026 | 30.4 | 0.07 |
| pHILIC | 154.02 | 157.7615 | 30.4 | 0.07 |
| pHILIC | 154.02 | 46.8375 | 30.3 | 0.07 |
| pHILIC | 154.02 | 73.122 | 30.3 | 0.07 |
| pHILIC | 154.02 | 43.429 | 30.3 | 0.07 |
| pHILIC | 154.02 | 73.1158 | 30.3 | 0.07 |
| pHILIC | 154.02 | 144.4547 | 30.3 | 0.07 |
| pHILIC | 154.02 | 119.2638 | 30.3 | 0.07 |
| pHILIC | 154.02 | 44.6729 | 30.3 | 0.07 |
| pHILIC | 154.02 | 105.3743 | 30.3 | 0.07 |
| pHILIC | 154.02 | 49.333 | 30.3 | 0.07 |
| pHILIC | 154.02 | 42.7588 | 30.3 | 0.07 |
| pHILIC | 154.02 | 130.4253 | 30.3 | 0.07 |
| pHILIC | 154.02 | 55.4944 | 30.3 | 0.07 |

| | | | | |
|---|---|---|---|---|
| pHILIC | 154.02 | 174.2692 | 30.3 | 0.07 |
| pHILIC | 154.02 | 64.4135 | 30.3 | 0.07 |
| pHILIC | 154.02 | 84.8989 | 30.3 | 0.07 |
| pHILIC | 154.02 | 170.6564 | 30.3 | 0.07 |
| pHILIC | 154.02 | 101.9623 | 30.3 | 0.07 |
| pHILIC | 154.02 | 68.9569 | 30.3 | 0.07 |
| pHILIC | 154.02 | 107.252 | 30.3 | 0.07 |
| pHILIC | 154.02 | 84.3207 | 30.3 | 0.07 |
| pHILIC | 154.02 | 74.8127 | 30.3 | 0.07 |
| pHILIC | 154.02 | 134.9495 | 30.3 | 0.07 |
| pHILIC | 154.02 | 133.2273 | 30.3 | 0.07 |
| pHILIC | 154.02 | 131.1045 | 30.3 | 0.07 |
| pHILIC | 154.02 | 80.4302 | 30.3 | 0.07 |
| pHILIC | 154.02 | 126.3928 | 30.3 | 0.07 |
| pHILIC | 154.02 | 169.379 | 30.3 | 0.07 |
| pHILIC | 154.02 | 112.5583 | 30.3 | 0.07 |
| pHILIC | 154.02 | 95.483 | 30.2 | 0.07 |
| pHILIC | 154.02 | 123.2213 | 30.2 | 0.07 |
| pHILIC | 154.02 | 60.3268 | 30.2 | 0.07 |
| pHILIC | 154.02 | 73.4887 | 30.2 | 0.07 |
| pHILIC | 154.02 | 74.3965 | 30.2 | 0.07 |
| pHILIC | 154.02 | 62.7962 | 30.2 | 0.07 |
| pHILIC | 154.02 | 97.5432 | 30.2 | 0.07 |
| pHILIC | 154.02 | 76.2291 | 30.2 | 0.07 |
| pHILIC | 154.02 | 86.0788 | 30.2 | 0.07 |
| pHILIC | 154.02 | 90.4796 | 30.2 | 0.07 |
| pHILIC | 154.02 | 46.1686 | 30.2 | 0.07 |
| pHILIC | 154.02 | 93.9219 | 30.2 | 0.07 |
| pHILIC | 154.02 | 123.7072 | 30.2 | 0.07 |
| pHILIC | 154.02 | 92.6284 | 30.2 | 0.07 |
| pHILIC | 154.02 | 81.6444 | 30.2 | 0.07 |
| pHILIC | 154.02 | 150.1031 | 30.2 | 0.07 |
| pHILIC | 154.02 | 64.3425 | 30.2 | 0.07 |
| pHILIC | 154.02 | 40.6574 | 30.2 | 0.07 |
| pHILIC | 154.02 | 117.9594 | 30.2 | 0.07 |
| pHILIC | 154.02 | 70.0877 | 30.2 | 0.07 |
| pHILIC | 154.02 | 134.0613 | 30.2 | 0.07 |
| pHILIC | 154.02 | 84.7644 | 30.2 | 0.07 |
| pHILIC | 154.02 | 52.2925 | 30.2 | 0.07 |
| pHILIC | 154.02 | 162.1402 | 30.2 | 0.07 |
| pHILIC | 154.02 | 165.3272 | 30.1 | 0.07 |
| pHILIC | 154.02 | 137.9008 | 30.1 | 0.07 |

| | | | | |
|---|---|---|---|---|
| pHILIC | 154.02 | 74.7894 | 30.1 | 0.07 |
| pHILIC | 154.02 | 45.7062 | 30.1 | 0.07 |
| pHILIC | 154.02 | 66.0378 | 30.1 | 0.07 |
| pHILIC | 154.02 | 59.8991 | 30.1 | 0.07 |
| pHILIC | 154.02 | 96.4013 | 30.1 | 0.07 |
| pHILIC | 154.02 | 138.2144 | 30.1 | 0.07 |
| pHILIC | 154.02 | 78.5284 | 30.1 | 0.07 |
| pHILIC | 154.02 | 87.3085 | 30.1 | 0.07 |
| pHILIC | 154.02 | 50.5311 | 30.1 | 0.07 |
| pHILIC | 154.02 | 116.8126 | 30.1 | 0.07 |
| pHILIC | 154.02 | 61.3843 | 30.1 | 0.07 |
| pHILIC | 154.02 | 74.9481 | 30.1 | 0.07 |
| pHILIC | 154.02 | 129.3542 | 30.1 | 0.07 |
| pHILIC | 154.02 | 84.3344 | 30.1 | 0.07 |
| pHILIC | 154.02 | 68.3057 | 30.1 | 0.07 |
| pHILIC | 154.02 | 77.7819 | 30.1 | 0.07 |
| pHILIC | 154.02 | 40.582 | 30.1 | 0.07 |
| pHILIC | 154.02 | 111.7317 | 30.1 | 0.07 |
| pHILIC | 154.02 | 88.9535 | 30.1 | 0.07 |
| pHILIC | 154.02 | 83.1875 | 30.1 | 0.07 |
| pHILIC | 154.02 | 152.7413 | 30.1 | 0.07 |
| pHILIC | 154.02 | 113.4044 | 30.1 | 0.07 |
| pHILIC | 154.02 | 107.0173 | 30.1 | 0.07 |
| pHILIC | 154.02 | 69.2718 | 30.1 | 0.07 |
| pHILIC | 154.02 | 45.1319 | 30.1 | 0.07 |
| pHILIC | 154.02 | 97.3609 | 30.1 | 0.07 |
| pHILIC | 154.02 | 49.0545 | 30.1 | 0.07 |
| pHILIC | 154.02 | 40.9162 | 30.1 | 0.07 |
| pHILIC | 154.02 | 45.3039 | 30.1 | 0.07 |
| pHILIC | 154.02 | 159.9574 | 30.1 | 0.07 |
| pHILIC | 154.02 | 72.6002 | 30.1 | 0.07 |
| pHILIC | 154.02 | 79.5907 | 30 | 0.07 |
| pHILIC | 154.02 | 47.8643 | 30 | 0.07 |
| pHILIC | 154.02 | 50.9329 | 30 | 0.07 |
| pHILIC | 154.02 | 162.081 | 30 | 0.07 |
| pHILIC | 154.02 | 124.6318 | 30 | 0.07 |
| pHILIC | 154.02 | 62.9193 | 30 | 0.07 |
| pHILIC | 154.02 | 68.0624 | 30 | 0.07 |
| pHILIC | 154.02 | 131.2347 | 30 | 0.07 |
| pHILIC | 154.02 | 61.7876 | 30 | 0.07 |
| pHILIC | 154.02 | 72.5826 | 30 | 0.07 |
| pHILIC | 154.02 | 48.1249 | 30 | 0.07 |

| | | | | |
|---|---|---|---|---|
| pHILIC | 154.02 | 151.4298 | 30 | 0.07 |
| pHILIC | 154.02 | 53.253 | 30 | 0.07 |
| pHILIC | 154.02 | 91.2175 | 30 | 0.07 |
| pHILIC | 154.02 | 119.2431 | 30 | 0.07 |
| pHILIC | 154.02 | 95.3609 | 30 | 0.07 |
| pHILIC | 154.02 | 146.3456 | 30 | 0.07 |
| pHILIC | 154.02 | 166.19 | 30 | 0.07 |
| pHILIC | 154.02 | 50.2936 | 30 | 0.07 |
| pHILIC | 154.02 | 54.3337 | 30 | 0.07 |
| pHILIC | 154.02 | 57.9381 | 30 | 0.07 |
| pHILIC | 154.02 | 43.124 | 30 | 0.07 |
| pHILIC | 154.02 | 56.9114 | 30 | 0.07 |
| pHILIC | 154.02 | 64.2678 | 30 | 0.07 |
| pHILIC | 154.02 | 43.9514 | 30 | 0.07 |
| pHILIC | 154.02 | 117.2502 | 30 | 0.07 |
| pHILIC | 154.02 | 92.2679 | 30 | 0.07 |
| pHILIC | 154.02 | 159.7873 | 30 | 0.07 |
| pHILIC | 154.02 | 86.8198 | 30 | 0.07 |
| pHILIC | 154.02 | 77.999 | 30 | 0.07 |
| pHILIC | 154.02 | 56.6421 | 30 | 0.07 |
| pHILIC | 154.02 | 61.8891 | 29.9 | 0.07 |
| pHILIC | 154.02 | 87.2243 | 29.9 | 0.07 |
| pHILIC | 154.02 | 73.2084 | 29.9 | 0.07 |
| pHILIC | 154.02 | 81.6239 | 29.9 | 0.07 |
| pHILIC | 154.02 | 81.2544 | 29.9 | 0.07 |
| pHILIC | 154.02 | 150.032 | 29.9 | 0.07 |
| pHILIC | 154.02 | 110.3366 | 29.9 | 0.07 |
| pHILIC | 154.02 | 131.4962 | 29.9 | 0.07 |
| pHILIC | 154.02 | 50.7595 | 29.9 | 0.07 |
| pHILIC | 154.02 | 73.7391 | 29.9 | 0.07 |
| pHILIC | 154.02 | 132.5976 | 29.9 | 0.07 |
| pHILIC | 154.02 | 53.9117 | 29.9 | 0.07 |
| pHILIC | 154.02 | 118.8032 | 29.9 | 0.07 |
| pHILIC | 154.02 | 79.558 | 29.9 | 0.07 |
| pHILIC | 154.02 | 58.5655 | 29.9 | 0.07 |
| pHILIC | 154.02 | 100.6126 | 29.9 | 0.07 |
| pHILIC | 154.02 | 139.9851 | 29.9 | 0.07 |
| pHILIC | 154.02 | 44.9425 | 29.9 | 0.07 |
| pHILIC | 154.02 | 169.7388 | 29.9 | 0.07 |
| pHILIC | 154.02 | 65.5015 | 29.9 | 0.07 |
| pHILIC | 154.02 | 96.8436 | 29.9 | 0.07 |
| pHILIC | 154.02 | 94.2741 | 29.9 | 0.07 |

| | | | | |
|---|---|---|---|---|
| pHILIC | 154.02 | 97.1505 | 29.9 | 0.07 |
| pHILIC | 154.02 | 117.5526 | 29.9 | 0.07 |
| pHILIC | 154.02 | 114.0381 | 29.9 | 0.07 |
| pHILIC | 154.02 | 50.2504 | 29.9 | 0.07 |
| pHILIC | 154.02 | 49.1177 | 29.9 | 0.07 |
| pHILIC | 154.02 | 173.1754 | 29.9 | 0.07 |
| pHILIC | 154.02 | 122.0228 | 29.9 | 0.07 |
| pHILIC | 154.02 | 70.0164 | 29.9 | 0.07 |
| pHILIC | 154.02 | 53.2115 | 29.8 | 0.07 |
| pHILIC | 154.02 | 104.0542 | 29.8 | 0.07 |
| pHILIC | 154.02 | 66.1101 | 29.8 | 0.07 |
| pHILIC | 154.02 | 70.8615 | 29.8 | 0.07 |
| pHILIC | 154.02 | 42.8071 | 29.8 | 0.07 |
| pHILIC | 154.02 | 168.5613 | 29.8 | 0.07 |
| pHILIC | 154.02 | 47.5513 | 29.8 | 0.07 |
| pHILIC | 154.02 | 63.2075 | 29.8 | 0.07 |
| pHILIC | 154.02 | 69.4715 | 29.8 | 0.07 |
| pHILIC | 154.02 | 119.2227 | 29.8 | 0.07 |
| pHILIC | 154.02 | 96.1189 | 29.8 | 0.07 |
| pHILIC | 154.02 | 42.8495 | 29.8 | 0.07 |
| pHILIC | 154.02 | 54.9907 | 29.8 | 0.07 |
| pHILIC | 154.02 | 173.7866 | 29.8 | 0.07 |
| pHILIC | 154.02 | 89.8369 | 29.8 | 0.07 |
| pHILIC | 154.02 | 80.4795 | 29.8 | 0.07 |
| pHILIC | 154.02 | 105.7526 | 29.8 | 0.07 |
| pHILIC | 154.02 | 82.6953 | 29.8 | 0.07 |
| pHILIC | 154.02 | 91.1337 | 29.8 | 0.07 |
| pHILIC | 154.02 | 48.4063 | 29.8 | 0.07 |
| pHILIC | 154.02 | 90.7883 | 29.8 | 0.07 |
| pHILIC | 154.02 | 63.3998 | 29.8 | 0.07 |
| pHILIC | 154.02 | 40.6277 | 29.8 | 0.07 |
| pHILIC | 154.02 | 120.8509 | 29.8 | 0.07 |
| pHILIC | 154.02 | 100.4526 | 29.8 | 0.07 |
| pHILIC | 154.02 | 78.5415 | 29.8 | 0.07 |
| pHILIC | 154.02 | 62.2218 | 29.8 | 0.07 |
| pHILIC | 154.02 | 105.5886 | 29.8 | 0.07 |
| pHILIC | 154.02 | 113.7443 | 29.8 | 0.07 |
| pHILIC | 154.02 | 99.0774 | 29.7 | 0.07 |
| pHILIC | 154.02 | 138.9265 | 29.7 | 0.07 |
| pHILIC | 154.02 | 138.1433 | 29.7 | 0.07 |
| pHILIC | 154.02 | 42.1045 | 29.7 | 0.07 |
| pHILIC | 154.02 | 67.0742 | 29.7 | 0.07 |

| | | | | |
|---|---|---|---|---|
| pHILIC | 154.02 | 41.7655 | 29.7 | 0.07 |
| pHILIC | 154.02 | 50.7484 | 29.7 | 0.07 |
| pHILIC | 154.02 | 85.838 | 29.7 | 0.07 |
| pHILIC | 154.02 | 83.1892 | 29.7 | 0.07 |
| pHILIC | 154.02 | 93.9984 | 29.7 | 0.07 |
| pHILIC | 154.02 | 92.6425 | 29.7 | 0.07 |
| pHILIC | 154.02 | 67.8726 | 29.7 | 0.07 |
| pHILIC | 154.02 | 55.8969 | 29.7 | 0.07 |
| pHILIC | 154.02 | 68.1154 | 29.7 | 0.07 |
| pHILIC | 154.02 | 78.4104 | 29.7 | 0.07 |
| pHILIC | 154.02 | 95.0868 | 29.7 | 0.07 |
| pHILIC | 154.02 | 106.5672 | 29.7 | 0.07 |
| pHILIC | 154.02 | 172.8853 | 29.7 | 0.07 |
| pHILIC | 154.02 | 114.5382 | 29.7 | 0.07 |
| pHILIC | 154.02 | 46.1024 | 29.7 | 0.07 |
| pHILIC | 154.02 | 52.0556 | 29.7 | 0.07 |
| pHILIC | 154.02 | 86.3992 | 29.7 | 0.07 |
| pHILIC | 154.02 | 50.9655 | 29.7 | 0.07 |
| pHILIC | 154.02 | 45.0656 | 29.7 | 0.07 |
| pHILIC | 154.02 | 122.4329 | 29.7 | 0.07 |
| pHILIC | 154.02 | 119.7449 | 29.6 | 0.07 |
| pHILIC | 154.02 | 93.7146 | 29.6 | 0.07 |
| pHILIC | 154.02 | 68.2764 | 29.6 | 0.07 |
| pHILIC | 154.02 | 59.9364 | 29.6 | 0.07 |
| pHILIC | 154.02 | 68.8413 | 29.6 | 0.07 |
| pHILIC | 154.02 | 53.9916 | 29.6 | 0.07 |
| pHILIC | 154.02 | 57.4413 | 29.6 | 0.07 |
| pHILIC | 154.02 | 134.2238 | 29.6 | 0.07 |
| pHILIC | 154.02 | 73.0105 | 29.6 | 0.07 |
| pHILIC | 154.02 | 98.6596 | 29.6 | 0.07 |
| pHILIC | 154.02 | 49.0238 | 29.6 | 0.07 |
| pHILIC | 154.02 | 47.4569 | 29.6 | 0.07 |
| pHILIC | 154.02 | 64.6498 | 29.6 | 0.07 |
| pHILIC | 154.02 | 114.3327 | 29.6 | 0.07 |
| pHILIC | 154.02 | 67.6337 | 29.6 | 0.07 |
| pHILIC | 154.02 | 48.0623 | 29.6 | 0.07 |
| pHILIC | 154.02 | 166.3134 | 29.6 | 0.07 |
| pHILIC | 154.02 | 57.5545 | 29.6 | 0.07 |
| pHILIC | 154.02 | 67.7973 | 29.6 | 0.07 |
| pHILIC | 154.02 | 169.0111 | 29.6 | 0.07 |
| pHILIC | 154.02 | 84.934 | 29.6 | 0.07 |
| pHILIC | 154.02 | 62.0419 | 29.6 | 0.07 |

| | | | | |
|---|---|---|---|---|
| pHILIC | 154.02 | 123.113 | 29.6 | 0.07 |
| pHILIC | 154.02 | 72.9611 | 29.6 | 0.07 |
| pHILIC | 154.02 | 125.4423 | 29.6 | 0.07 |
| pHILIC | 154.02 | 77.7916 | 29.5 | 0.07 |
| pHILIC | 154.02 | 55.7583 | 29.5 | 0.07 |
| pHILIC | 154.02 | 76.0561 | 29.5 | 0.07 |
| pHILIC | 154.02 | 99.6121 | 29.5 | 0.07 |
| pHILIC | 154.02 | 40.7729 | 29.5 | 0.07 |
| pHILIC | 154.02 | 71.4389 | 29.5 | 0.07 |
| pHILIC | 154.02 | 149.3033 | 29.5 | 0.07 |
| pHILIC | 154.02 | 131.0719 | 29.5 | 0.07 |
| pHILIC | 154.02 | 61.7434 | 29.5 | 0.07 |
| pHILIC | 154.02 | 40.1822 | 29.5 | 0.07 |
| pHILIC | 154.02 | 53.9399 | 29.5 | 0.07 |
| pHILIC | 154.02 | 47.744 | 29.5 | 0.07 |
| pHILIC | 154.02 | 127.0901 | 29.5 | 0.07 |
| pHILIC | 154.02 | 50.4346 | 29.5 | 0.07 |
| pHILIC | 154.02 | 87.7326 | 29.5 | 0.07 |
| pHILIC | 154.02 | 91.1243 | 29.5 | 0.07 |
| pHILIC | 154.02 | 56.2002 | 29.5 | 0.07 |
| pHILIC | 154.02 | 108.7828 | 29.5 | 0.07 |
| pHILIC | 154.02 | 58.7924 | 29.5 | 0.07 |
| pHILIC | 154.02 | 74.7655 | 29.5 | 0.07 |
| pHILIC | 154.02 | 151.5775 | 29.5 | 0.07 |
| pHILIC | 154.02 | 49.1047 | 29.5 | 0.07 |
| pHILIC | 154.02 | 93.7858 | 29.5 | 0.07 |
| pHILIC | 154.02 | 62.4874 | 29.5 | 0.07 |
| pHILIC | 154.02 | 58.6599 | 29.5 | 0.07 |
| pHILIC | 154.02 | 42.268 | 29.5 | 0.07 |
| pHILIC | 154.02 | 76.0114 | 29.5 | 0.07 |
| pHILIC | 154.02 | 149.6714 | 29.5 | 0.07 |
| pHILIC | 154.02 | 119.9275 | 29.5 | 0.07 |
| pHILIC | 154.02 | 115.5277 | 29.4 | 0.07 |
| pHILIC | 154.02 | 98.5243 | 29.4 | 0.07 |
| pHILIC | 154.02 | 135.5763 | 29.4 | 0.07 |
| pHILIC | 154.02 | 45.3595 | 29.4 | 0.07 |
| pHILIC | 154.02 | 47.3653 | 29.4 | 0.07 |
| pHILIC | 154.02 | 148.6585 | 29.4 | 0.07 |
| pHILIC | 154.02 | 61.4706 | 29.4 | 0.07 |
| pHILIC | 154.02 | 75.0197 | 29.4 | 0.07 |
| pHILIC | 154.02 | 145.1368 | 29.4 | 0.07 |
| pHILIC | 154.02 | 53.8272 | 29.4 | 0.07 |

| | | | | |
|---|---|---|---|---|
| pHILIC | 154.02 | 61.6496 | 29.4 | 0.07 |
| pHILIC | 154.02 | 97.1176 | 29.4 | 0.07 |
| pHILIC | 154.02 | 138.3661 | 29.4 | 0.07 |
| pHILIC | 154.02 | 60.1169 | 29.4 | 0.07 |
| pHILIC | 154.02 | 97.4087 | 29.4 | 0.07 |
| pHILIC | 154.02 | 44.4001 | 29.4 | 0.07 |
| pHILIC | 154.02 | 47.9091 | 29.4 | 0.07 |
| pHILIC | 154.02 | 54.9407 | 29.4 | 0.07 |
| pHILIC | 154.02 | 61.9694 | 29.4 | 0.07 |
| pHILIC | 154.02 | 70.3121 | 29.4 | 0.07 |
| pHILIC | 154.02 | 54.2696 | 29.4 | 0.07 |
| pHILIC | 154.02 | 111.2493 | 29.4 | 0.07 |
| pHILIC | 154.02 | 66.5281 | 29.4 | 0.07 |
| pHILIC | 154.02 | 94.5191 | 29.4 | 0.07 |
| pHILIC | 154.02 | 85.285 | 29.4 | 0.07 |
| pHILIC | 154.02 | 43.522 | 29.3 | 0.07 |
| pHILIC | 154.02 | 47.5128 | 29.3 | 0.07 |
| pHILIC | 154.02 | 48.3146 | 29.3 | 0.07 |
| pHILIC | 154.02 | 80.3165 | 29.3 | 0.07 |
| pHILIC | 154.02 | 112.7693 | 29.3 | 0.07 |
| pHILIC | 154.02 | 45.7468 | 29.3 | 0.07 |
| pHILIC | 154.02 | 70.9284 | 29.3 | 0.07 |
| pHILIC | 154.02 | 50.1372 | 29.3 | 0.07 |
| pHILIC | 154.02 | 130.2574 | 29.3 | 0.07 |
| pHILIC | 154.02 | 68.849 | 29.3 | 0.07 |
| pHILIC | 154.02 | 74.0527 | 29.3 | 0.07 |
| pHILIC | 154.02 | 58.1637 | 29.3 | 0.07 |
| pHILIC | 154.02 | 158.7519 | 29.3 | 0.07 |
| pHILIC | 154.02 | 66.2562 | 29.3 | 0.07 |
| pHILIC | 154.02 | 71.7791 | 29.3 | 0.07 |
| pHILIC | 154.02 | 52.5197 | 29.3 | 0.07 |
| pHILIC | 154.02 | 173.2323 | 29.3 | 0.07 |
| pHILIC | 154.02 | 65.9236 | 29.3 | 0.07 |
| pHILIC | 154.02 | 47.7091 | 29.3 | 0.07 |
| pHILIC | 154.02 | 40.002 | 29.3 | 0.07 |
| pHILIC | 154.02 | 115.2877 | 29.3 | 0.07 |
| pHILIC | 154.02 | 134.2759 | 29.3 | 0.07 |
| pHILIC | 154.02 | 71.0618 | 29.3 | 0.07 |
| pHILIC | 154.02 | 52.2649 | 29.3 | 0.07 |
| pHILIC | 154.02 | 125.6905 | 29.3 | 0.07 |
| pHILIC | 154.02 | 111.6765 | 29.3 | 0.07 |
| pHILIC | 154.02 | 52.3043 | 29.3 | 0.07 |

| | | | | |
|---|---|---|---|---|
| pHILIC | 154.02 | 46.826 | 29.2 | 0.07 |
| pHILIC | 154.02 | 77.7133 | 29.2 | 0.07 |
| pHILIC | 154.02 | 83.8075 | 29.2 | 0.07 |
| pHILIC | 154.02 | 49.0659 | 29.2 | 0.07 |
| pHILIC | 154.02 | 56.2169 | 29.2 | 0.07 |
| pHILIC | 154.02 | 45.1663 | 29.2 | 0.07 |
| pHILIC | 154.02 | 157.1369 | 29.2 | 0.07 |
| pHILIC | 154.02 | 99.5906 | 29.2 | 0.07 |
| pHILIC | 154.02 | 53.7615 | 29.2 | 0.07 |
| pHILIC | 154.02 | 109.4996 | 29.2 | 0.07 |
| pHILIC | 154.02 | 46.5227 | 29.2 | 0.07 |
| pHILIC | 154.02 | 167.4347 | 29.2 | 0.07 |
| pHILIC | 154.02 | 87.9815 | 29.2 | 0.07 |
| pHILIC | 154.02 | 44.1148 | 29.2 | 0.07 |
| pHILIC | 154.02 | 67.6301 | 29.2 | 0.07 |
| pHILIC | 154.02 | 78.5575 | 29.2 | 0.07 |
| pHILIC | 154.02 | 46.4118 | 29.2 | 0.07 |
| pHILIC | 154.02 | 98.1636 | 29.1 | 0.07 |
| pHILIC | 154.02 | 41.3733 | 29.1 | 0.07 |
| pHILIC | 154.02 | 53.1363 | 29.1 | 0.07 |
| pHILIC | 154.02 | 88.4276 | 29.1 | 0.07 |
| pHILIC | 154.02 | 45.6966 | 29.1 | 0.07 |
| pHILIC | 154.02 | 45.5118 | 29.1 | 0.07 |
| pHILIC | 154.02 | 75.0735 | 29.1 | 0.07 |
| pHILIC | 154.02 | 93.6615 | 29.1 | 0.07 |
| pHILIC | 154.02 | 72.6663 | 29.1 | 0.07 |
| pHILIC | 154.02 | 123.7238 | 29.1 | 0.07 |
| pHILIC | 154.02 | 60.9695 | 29.1 | 0.07 |
| pHILIC | 154.02 | 108.2695 | 29.1 | 0.07 |
| pHILIC | 154.02 | 176.1711 | 29.1 | 0.07 |
| pHILIC | 154.02 | 116.1019 | 29.1 | 0.07 |
| pHILIC | 154.02 | 64.5551 | 29.1 | 0.07 |
| pHILIC | 154.02 | 72.9856 | 29.1 | 0.07 |
| pHILIC | 154.02 | 52.9241 | 29.1 | 0.07 |
| pHILIC | 154.02 | 139.3367 | 29.1 | 0.07 |
| pHILIC | 154.02 | 91.5814 | 29 | 0.07 |
| pHILIC | 154.02 | 45.898 | 29 | 0.07 |
| pHILIC | 154.02 | 47.1881 | 29 | 0.07 |
| pHILIC | 154.02 | 43.7483 | 29 | 0.07 |
| pHILIC | 154.02 | 175.3255 | 29 | 0.07 |
| pHILIC | 154.02 | 119.0932 | 29 | 0.07 |
| pHILIC | 154.02 | 151.1239 | 29 | 0.07 |

| | | | | |
|---|---|---|---|---|
| pHILIC | 154.02 | 133.4306 | 29 | 0.07 |
| pHILIC | 154.02 | 44.968 | 29 | 0.07 |
| pHILIC | 154.02 | 157.3042 | 29 | 0.07 |
| pHILIC | 154.02 | 137.6909 | 29 | 0.07 |
| pHILIC | 154.02 | 69.7165 | 29 | 0.07 |
| pHILIC | 154.02 | 69.7449 | 29 | 0.07 |
| pHILIC | 154.02 | 59.4208 | 29 | 0.07 |
| pHILIC | 154.02 | 60.3211 | 29 | 0.07 |
| pHILIC | 154.02 | 47.8325 | 29 | 0.07 |
| pHILIC | 154.02 | 58.8106 | 29 | 0.07 |
| pHILIC | 154.02 | 126.4882 | 29 | 0.07 |
| pHILIC | 154.02 | 46.9253 | 29 | 0.07 |
| pHILIC | 154.02 | 135.6502 | 29 | 0.07 |
| pHILIC | 154.02 | 50.1953 | 29 | 0.07 |
| pHILIC | 154.02 | 128.3095 | 29 | 0.07 |
| pHILIC | 154.02 | 48.1986 | 29 | 0.07 |
| pHILIC | 154.02 | 76.59 | 29 | 0.07 |
| pHILIC | 154.02 | 123.7415 | 28.9 | 0.07 |
| pHILIC | 154.02 | 109.2951 | 28.9 | 0.07 |
| pHILIC | 154.02 | 175.4123 | 28.9 | 0.07 |
| pHILIC | 154.02 | 67.1309 | 28.9 | 0.07 |
| pHILIC | 154.02 | 77.3878 | 28.9 | 0.07 |
| pHILIC | 154.02 | 171.0982 | 28.9 | 0.07 |
| pHILIC | 154.02 | 66.8184 | 28.9 | 0.07 |
| pHILIC | 154.02 | 51.2242 | 28.9 | 0.07 |
| pHILIC | 154.02 | 64.4955 | 28.9 | 0.07 |
| pHILIC | 154.02 | 118.8747 | 28.9 | 0.07 |
| pHILIC | 154.02 | 70.4437 | 28.9 | 0.07 |
| pHILIC | 154.02 | 120.5204 | 28.9 | 0.07 |
| pHILIC | 154.02 | 110.6283 | 28.9 | 0.07 |
| pHILIC | 154.02 | 46.7837 | 28.9 | 0.07 |
| pHILIC | 154.02 | 46.2154 | 28.9 | 0.07 |
| pHILIC | 154.02 | 72.0405 | 28.9 | 0.07 |
| pHILIC | 154.02 | 48.6709 | 28.9 | 0.07 |
| pHILIC | 154.02 | 53.9746 | 28.9 | 0.07 |
| pHILIC | 154.02 | 42.654 | 28.9 | 0.07 |
| pHILIC | 154.02 | 111.9675 | 28.8 | 0.07 |
| pHILIC | 154.02 | 42.3893 | 28.8 | 0.07 |
| pHILIC | 154.02 | 136.2495 | 28.8 | 0.07 |
| pHILIC | 154.02 | 41.1857 | 28.8 | 0.07 |
| pHILIC | 154.02 | 129.6632 | 28.8 | 0.07 |
| pHILIC | 154.02 | 46.9911 | 28.8 | 0.07 |

| | | | | |
|---|---|---|---|---|
| pHILIC | 154.02 | 52.8785 | 28.8 | 0.07 |
| pHILIC | 154.02 | 95.9395 | 28.8 | 0.07 |
| pHILIC | 154.02 | 45.0016 | 28.8 | 0.07 |
| pHILIC | 154.02 | 110.2857 | 28.8 | 0.07 |
| pHILIC | 154.02 | 51.8761 | 28.8 | 0.07 |
| pHILIC | 154.02 | 61.0914 | 28.8 | 0.07 |
| pHILIC | 154.02 | 41.0049 | 28.8 | 0.07 |
| pHILIC | 154.02 | 40.2566 | 28.8 | 0.07 |
| pHILIC | 154.02 | 66.6685 | 28.8 | 0.07 |
| pHILIC | 154.02 | 80.6678 | 28.8 | 0.07 |
| pHILIC | 154.02 | 162.7397 | 28.8 | 0.07 |
| pHILIC | 154.02 | 56.707 | 28.8 | 0.07 |
| pHILIC | 154.02 | 54.6184 | 28.8 | 0.07 |
| pHILIC | 154.02 | 57.8142 | 28.8 | 0.07 |
| pHILIC | 154.02 | 116.7157 | 28.8 | 0.07 |
| pHILIC | 154.02 | 76.434 | 28.8 | 0.07 |
| pHILIC | 154.02 | 69.6515 | 28.8 | 0.07 |
| pHILIC | 154.02 | 46.9366 | 28.7 | 0.07 |
| pHILIC | 154.02 | 42.3188 | 28.7 | 0.07 |
| pHILIC | 154.02 | 47.6755 | 28.7 | 0.07 |
| pHILIC | 154.02 | 66.8303 | 28.7 | 0.07 |
| pHILIC | 154.02 | 86.4808 | 28.7 | 0.07 |
| pHILIC | 154.02 | 133.6433 | 28.7 | 0.07 |
| pHILIC | 154.02 | 41.6709 | 28.7 | 0.07 |
| pHILIC | 154.02 | 49.5029 | 28.7 | 0.07 |
| pHILIC | 154.02 | 44.8959 | 28.7 | 0.07 |
| pHILIC | 154.02 | 50.6257 | 28.7 | 0.07 |
| pHILIC | 154.02 | 96.1042 | 28.7 | 0.07 |
| pHILIC | 154.02 | 41.7941 | 28.7 | 0.07 |
| pHILIC | 154.02 | 118.36 | 28.7 | 0.07 |
| pHILIC | 154.02 | 79.5403 | 28.7 | 0.07 |
| pHILIC | 154.02 | 135.7642 | 28.7 | 0.07 |
| pHILIC | 154.02 | 106.3941 | 28.7 | 0.07 |
| pHILIC | 154.02 | 177.3589 | 28.7 | 0.07 |
| pHILIC | 154.02 | 66.3593 | 28.7 | 0.07 |
| pHILIC | 154.02 | 45.9735 | 28.7 | 0.07 |
| pHILIC | 154.02 | 116.1514 | 28.7 | 0.07 |
| pHILIC | 154.02 | 42.6363 | 28.7 | 0.07 |
| pHILIC | 154.02 | 55.9296 | 28.7 | 0.07 |
| pHILIC | 154.02 | 125.9911 | 28.7 | 0.07 |
| pHILIC | 154.02 | 114.8487 | 28.7 | 0.07 |
| pHILIC | 154.02 | 171.2353 | 28.7 | 0.07 |

| | | | | |
|---|---|---|---|---|
| pHILIC | 154.02 | 77.3771 | 28.7 | 0.07 |
| pHILIC | 154.02 | 171.2003 | 28.7 | 0.07 |
| pHILIC | 154.02 | 153.2494 | 28.7 | 0.07 |
| pHILIC | 154.02 | 45.1428 | 28.7 | 0.07 |
| pHILIC | 154.02 | 85.3482 | 28.7 | 0.07 |
| pHILIC | 154.02 | 63.6366 | 28.7 | 0.07 |
| pHILIC | 154.02 | 92.542 | 28.6 | 0.07 |
| pHILIC | 154.02 | 51.9655 | 28.6 | 0.07 |
| pHILIC | 154.02 | 43.4798 | 28.6 | 0.07 |
| pHILIC | 154.02 | 74.5778 | 28.6 | 0.07 |
| pHILIC | 154.02 | 83.6284 | 28.6 | 0.07 |
| pHILIC | 154.02 | 63.0846 | 28.6 | 0.07 |
| pHILIC | 154.02 | 161.7523 | 28.6 | 0.07 |
| pHILIC | 154.02 | 70.6679 | 28.6 | 0.07 |
| pHILIC | 154.02 | 49.9863 | 28.6 | 0.07 |
| pHILIC | 154.02 | 51.1327 | 28.6 | 0.07 |
| pHILIC | 154.02 | 75.4578 | 28.6 | 0.07 |
| pHILIC | 154.02 | 62.6504 | 28.6 | 0.07 |
| pHILIC | 154.02 | 48.7624 | 28.6 | 0.07 |
| pHILIC | 154.02 | 48.3974 | 28.6 | 0.07 |
| pHILIC | 154.02 | 116.1768 | 28.6 | 0.07 |
| pHILIC | 154.02 | 63.5843 | 28.6 | 0.07 |
| pHILIC | 154.02 | 115.1215 | 28.6 | 0.07 |
| pHILIC | 154.02 | 89.4823 | 28.6 | 0.07 |
| pHILIC | 154.02 | 117.9424 | 28.6 | 0.07 |
| pHILIC | 154.02 | 148.2615 | 28.5 | 0.07 |
| pHILIC | 154.02 | 62.9027 | 28.5 | 0.07 |
| pHILIC | 154.02 | 45.6812 | 28.5 | 0.07 |
| pHILIC | 154.02 | 144.4968 | 28.5 | 0.07 |
| pHILIC | 154.02 | 69.5616 | 28.5 | 0.07 |
| pHILIC | 154.02 | 63.1145 | 28.5 | 0.07 |
| pHILIC | 154.02 | 87.9202 | 28.5 | 0.07 |
| pHILIC | 154.02 | 54.4542 | 28.5 | 0.07 |
| pHILIC | 154.02 | 61.8957 | 28.5 | 0.07 |
| pHILIC | 154.02 | 52.1734 | 28.5 | 0.07 |
| pHILIC | 154.02 | 48.5493 | 28.5 | 0.07 |
| pHILIC | 154.02 | 119.7335 | 28.5 | 0.07 |
| pHILIC | 154.02 | 46.993 | 28.5 | 0.07 |
| pHILIC | 154.02 | 70.3042 | 28.5 | 0.07 |
| pHILIC | 154.02 | 83.9138 | 28.5 | 0.07 |
| pHILIC | 154.02 | 55.2342 | 28.5 | 0.07 |
| pHILIC | 154.02 | 104.6311 | 28.5 | 0.07 |

| | | | | |
|---|---|---|---|---|
| pHILIC | 154.02 | 82.7197 | 28.5 | 0.07 |
| pHILIC | 154.02 | 46.6445 | 28.5 | 0.07 |
| pHILIC | 154.02 | 43.2541 | 28.5 | 0.07 |
| pHILIC | 154.02 | 65.598 | 28.5 | 0.07 |
| pHILIC | 154.02 | 46.7494 | 28.5 | 0.07 |
| pHILIC | 154.02 | 43.4234 | 28.5 | 0.07 |
| pHILIC | 154.02 | 123.6824 | 28.5 | 0.07 |
| pHILIC | 154.02 | 74.1423 | 28.4 | 0.07 |
| pHILIC | 154.02 | 88.2176 | 28.4 | 0.07 |
| pHILIC | 154.02 | 73.1026 | 28.4 | 0.07 |
| pHILIC | 154.02 | 149.9136 | 28.4 | 0.07 |
| pHILIC | 154.02 | 65.9078 | 28.4 | 0.07 |
| pHILIC | 154.02 | 86.89 | 28.4 | 0.06 |
| pHILIC | 154.02 | 107.7254 | 28.4 | 0.06 |
| pHILIC | 154.02 | 56.5806 | 28.4 | 0.06 |
| pHILIC | 154.02 | 101.9449 | 28.4 | 0.06 |
| pHILIC | 154.02 | 49.1444 | 28.4 | 0.06 |
| pHILIC | 154.02 | 127.4396 | 28.4 | 0.06 |
| pHILIC | 154.02 | 116.3123 | 28.4 | 0.06 |
| pHILIC | 154.02 | 70.8108 | 28.4 | 0.06 |
| pHILIC | 154.02 | 67.9855 | 28.4 | 0.06 |
| pHILIC | 154.02 | 56.5417 | 28.4 | 0.06 |
| pHILIC | 154.02 | 65.4351 | 28.4 | 0.06 |
| pHILIC | 154.02 | 43.217 | 28.4 | 0.06 |
| pHILIC | 154.02 | 40.88 | 28.4 | 0.06 |
| pHILIC | 154.02 | 47.6827 | 28.4 | 0.06 |
| pHILIC | 154.02 | 49.3951 | 28.4 | 0.06 |
| pHILIC | 154.02 | 71.265 | 28.3 | 0.06 |
| pHILIC | 154.02 | 133.2161 | 28.3 | 0.06 |
| pHILIC | 154.02 | 51.264 | 28.3 | 0.06 |
| pHILIC | 154.02 | 126.0898 | 28.3 | 0.06 |
| pHILIC | 154.02 | 98.0927 | 28.3 | 0.06 |
| pHILIC | 154.02 | 176.1591 | 28.3 | 0.06 |
| pHILIC | 154.02 | 40.6846 | 28.3 | 0.06 |
| pHILIC | 154.02 | 59.1304 | 28.3 | 0.06 |
| pHILIC | 154.02 | 90.6927 | 28.3 | 0.06 |
| pHILIC | 154.02 | 91.8165 | 28.3 | 0.06 |
| pHILIC | 154.02 | 41.1265 | 28.3 | 0.06 |
| pHILIC | 154.02 | 150.2932 | 28.3 | 0.06 |
| pHILIC | 154.02 | 100.6247 | 28.3 | 0.06 |
| pHILIC | 154.02 | 117.9376 | 28.2 | 0.06 |
| pHILIC | 154.02 | 85.001 | 28.2 | 0.06 |

| | | | | |
|---|---|---|---|---|
| pHILIC | 154.02 | 134.8685 | 28.2 | 0.06 |
| pHILIC | 154.02 | 61.9214 | 28.2 | 0.06 |
| pHILIC | 154.02 | 40.3416 | 28.2 | 0.06 |
| pHILIC | 154.02 | 108.3464 | 28.2 | 0.06 |
| pHILIC | 154.02 | 43.6499 | 28.2 | 0.06 |
| pHILIC | 154.02 | 42.5358 | 28.2 | 0.06 |
| pHILIC | 154.02 | 64.6911 | 28.2 | 0.06 |
| pHILIC | 154.02 | 88.6418 | 28.2 | 0.06 |
| pHILIC | 154.02 | 58.3821 | 28.2 | 0.06 |
| pHILIC | 154.02 | 77.0703 | 28.2 | 0.06 |
| pHILIC | 154.02 | 55.0795 | 28.2 | 0.06 |
| pHILIC | 154.02 | 139.1948 | 28.2 | 0.06 |
| pHILIC | 154.02 | 71.3178 | 28.2 | 0.06 |
| pHILIC | 154.02 | 59.6505 | 28.2 | 0.06 |
| pHILIC | 154.02 | 47.079 | 28.2 | 0.06 |
| pHILIC | 154.02 | 62.3475 | 28.2 | 0.06 |
| pHILIC | 154.02 | 162.0143 | 28.2 | 0.06 |
| pHILIC | 154.02 | 44.5613 | 28.2 | 0.06 |
| pHILIC | 154.02 | 135.3849 | 28.2 | 0.06 |
| pHILIC | 154.02 | 125.6386 | 28.2 | 0.06 |
| pHILIC | 154.02 | 48.9398 | 28.2 | 0.06 |
| pHILIC | 154.02 | 106.5764 | 28.1 | 0.06 |
| pHILIC | 154.02 | 87.1375 | 28.1 | 0.06 |
| pHILIC | 154.02 | 49.9125 | 28.1 | 0.06 |
| pHILIC | 154.02 | 129.7854 | 28.1 | 0.06 |
| pHILIC | 154.02 | 51.7132 | 28.1 | 0.06 |
| pHILIC | 154.02 | 44.5488 | 28.1 | 0.06 |
| pHILIC | 154.02 | 135.3185 | 28.1 | 0.06 |
| pHILIC | 154.02 | 42.668 | 28.1 | 0.06 |
| pHILIC | 154.02 | 102.9305 | 28.1 | 0.06 |
| pHILIC | 154.02 | 42.8664 | 28.1 | 0.06 |
| pHILIC | 154.02 | 84.4906 | 28.1 | 0.06 |
| pHILIC | 154.02 | 55.9412 | 28.1 | 0.06 |
| pHILIC | 154.02 | 49.6373 | 28.1 | 0.06 |
| pHILIC | 154.02 | 111.3835 | 28.1 | 0.06 |
| pHILIC | 154.02 | 109.2869 | 28.1 | 0.06 |
| pHILIC | 154.02 | 147.1883 | 28.1 | 0.06 |
| pHILIC | 154.02 | 115.5645 | 28 | 0.06 |
| pHILIC | 154.02 | 135.4309 | 28 | 0.06 |
| pHILIC | 154.02 | 42.2743 | 28 | 0.06 |
| pHILIC | 154.02 | 68.2989 | 28 | 0.06 |
| pHILIC | 154.02 | 52.9659 | 28 | 0.06 |

| | | | | |
|---|---|---|---|---|
| pHILIC | 154.02 | 45.3731 | 28 | 0.06 |
| pHILIC | 154.02 | 110.4372 | 28 | 0.06 |
| pHILIC | 154.02 | 105.1731 | 28 | 0.06 |
| pHILIC | 154.02 | 94.7205 | 28 | 0.06 |
| pHILIC | 154.02 | 47.3164 | 28 | 0.06 |
| pHILIC | 154.02 | 48.6795 | 28 | 0.06 |
| pHILIC | 154.02 | 56.7582 | 28 | 0.06 |
| pHILIC | 154.02 | 163.5048 | 28 | 0.06 |
| pHILIC | 154.02 | 66.7451 | 28 | 0.06 |
| pHILIC | 154.02 | 148.633 | 28 | 0.06 |
| pHILIC | 154.02 | 42.5092 | 28 | 0.06 |
| pHILIC | 154.02 | 132.0153 | 28 | 0.06 |
| pHILIC | 154.02 | 41.6336 | 28 | 0.06 |
| pHILIC | 154.02 | 47.1788 | 28 | 0.06 |
| pHILIC | 154.02 | 41.8863 | 28 | 0.06 |
| pHILIC | 154.02 | 68.5086 | 28 | 0.06 |
| pHILIC | 154.02 | 43.4017 | 28 | 0.06 |
| pHILIC | 154.02 | 152.8858 | 28 | 0.06 |
| pHILIC | 154.02 | 121.0194 | 28 | 0.06 |
| pHILIC | 154.02 | 93.402 | 27.9 | 0.06 |
| pHILIC | 154.02 | 55.6197 | 27.9 | 0.06 |
| pHILIC | 154.02 | 43.7216 | 27.9 | 0.06 |
| pHILIC | 154.02 | 56.3732 | 27.9 | 0.06 |
| pHILIC | 154.02 | 152.2047 | 27.9 | 0.06 |
| pHILIC | 154.02 | 53.8785 | 27.9 | 0.06 |
| pHILIC | 154.02 | 82.9758 | 27.9 | 0.06 |
| pHILIC | 154.02 | 73.4564 | 27.9 | 0.06 |
| pHILIC | 154.02 | 149.0625 | 27.9 | 0.06 |
| pHILIC | 154.02 | 63.8713 | 27.9 | 0.06 |
| pHILIC | 154.02 | 69.6766 | 27.9 | 0.06 |
| pHILIC | 154.02 | 55.0651 | 27.9 | 0.06 |
| pHILIC | 154.02 | 43.8019 | 27.9 | 0.06 |
| pHILIC | 154.02 | 74.9238 | 27.9 | 0.06 |
| pHILIC | 154.02 | 59.8591 | 27.9 | 0.06 |
| pHILIC | 154.02 | 40.5733 | 27.9 | 0.06 |
| pHILIC | 154.02 | 50.2383 | 27.9 | 0.06 |
| pHILIC | 154.02 | 59.0764 | 27.9 | 0.06 |
| pHILIC | 154.02 | 148.518 | 27.9 | 0.06 |
| pHILIC | 154.02 | 90.16 | 27.9 | 0.06 |
| pHILIC | 154.02 | 53.9432 | 27.9 | 0.06 |
| pHILIC | 154.02 | 49.2201 | 27.9 | 0.06 |
| pHILIC | 154.02 | 76.4172 | 27.9 | 0.06 |

| | | | | |
|---|---|---|---|---|
| pHILIC | 154.02 | 146.4944 | 27.9 | 0.06 |
| pHILIC | 154.02 | 44.2138 | 27.8 | 0.06 |
| pHILIC | 154.02 | 167.1199 | 27.8 | 0.06 |
| pHILIC | 154.02 | 55.6008 | 27.8 | 0.06 |
| pHILIC | 154.02 | 177.033 | 27.8 | 0.06 |
| pHILIC | 154.02 | 67.4184 | 27.8 | 0.06 |
| pHILIC | 154.02 | 84.668 | 27.8 | 0.06 |
| pHILIC | 154.02 | 60.7866 | 27.8 | 0.06 |
| pHILIC | 154.02 | 41.8297 | 27.8 | 0.06 |
| pHILIC | 154.02 | 62.5567 | 27.8 | 0.06 |
| pHILIC | 154.02 | 43.4563 | 27.8 | 0.06 |
| pHILIC | 154.02 | 65.6409 | 27.8 | 0.06 |
| pHILIC | 154.02 | 45.4083 | 27.8 | 0.06 |
| pHILIC | 154.02 | 46.643 | 27.8 | 0.06 |
| pHILIC | 154.02 | 125.7098 | 27.8 | 0.06 |
| pHILIC | 154.02 | 59.1346 | 27.8 | 0.06 |
| pHILIC | 154.02 | 122.3458 | 27.8 | 0.06 |
| pHILIC | 154.02 | 49.4521 | 27.8 | 0.06 |
| pHILIC | 154.02 | 40.4844 | 27.8 | 0.06 |
| pHILIC | 154.02 | 42.4436 | 27.8 | 0.06 |
| pHILIC | 154.02 | 86.0024 | 27.7 | 0.06 |
| pHILIC | 154.02 | 147.4997 | 27.7 | 0.06 |
| pHILIC | 154.02 | 40.7914 | 27.7 | 0.06 |
| pHILIC | 154.02 | 145.3513 | 27.7 | 0.06 |
| pHILIC | 154.02 | 46.6602 | 27.7 | 0.06 |
| pHILIC | 154.02 | 49.7095 | 27.7 | 0.06 |
| pHILIC | 154.02 | 102.452 | 27.7 | 0.06 |
| pHILIC | 154.02 | 40.7011 | 27.7 | 0.06 |
| pHILIC | 154.02 | 72.4925 | 27.7 | 0.06 |
| pHILIC | 154.02 | 169.6035 | 27.7 | 0.06 |
| pHILIC | 154.02 | 101.8535 | 27.7 | 0.06 |
| pHILIC | 154.02 | 46.8429 | 27.7 | 0.06 |
| pHILIC | 154.02 | 75.8487 | 27.7 | 0.06 |
| pHILIC | 154.02 | 73.7431 | 27.7 | 0.06 |
| pHILIC | 154.02 | 43.8636 | 27.7 | 0.06 |
| pHILIC | 154.02 | 58.4673 | 27.7 | 0.06 |
| pHILIC | 154.02 | 80.5547 | 27.7 | 0.06 |
| pHILIC | 154.02 | 48.7243 | 27.7 | 0.06 |
| pHILIC | 154.02 | 41.1354 | 27.7 | 0.06 |
| pHILIC | 154.02 | 68.725 | 27.7 | 0.06 |
| pHILIC | 154.02 | 87.7775 | 27.7 | 0.06 |
| pHILIC | 154.02 | 70.1933 | 27.6 | 0.06 |

| | | | | |
|---|---|---|---|---|
| pHILIC | 154.02 | 119.1008 | 27.6 | 0.06 |
| pHILIC | 154.02 | 81.1684 | 27.6 | 0.06 |
| pHILIC | 154.02 | 108.1424 | 27.6 | 0.06 |
| pHILIC | 154.02 | 98.1533 | 27.6 | 0.06 |
| pHILIC | 154.02 | 82.6004 | 27.6 | 0.06 |
| pHILIC | 154.02 | 80.7338 | 27.6 | 0.06 |
| pHILIC | 154.02 | 159.7147 | 27.6 | 0.06 |
| pHILIC | 154.02 | 55.5327 | 27.6 | 0.06 |
| pHILIC | 154.02 | 98.677 | 27.6 | 0.06 |
| pHILIC | 154.02 | 51.0399 | 27.6 | 0.06 |
| pHILIC | 154.02 | 40.1855 | 27.6 | 0.06 |
| pHILIC | 154.02 | 43.7377 | 27.6 | 0.06 |
| pHILIC | 154.02 | 43.2282 | 27.6 | 0.06 |
| pHILIC | 154.02 | 61.6508 | 27.6 | 0.06 |
| pHILIC | 154.02 | 80.0176 | 27.6 | 0.06 |
| pHILIC | 154.02 | 54.3751 | 27.5 | 0.06 |
| pHILIC | 154.02 | 47.3304 | 27.5 | 0.06 |
| pHILIC | 154.02 | 66.4743 | 27.5 | 0.06 |
| pHILIC | 154.02 | 52.1201 | 27.5 | 0.06 |
| pHILIC | 154.02 | 56.161 | 27.5 | 0.06 |
| pHILIC | 154.02 | 49.6431 | 27.5 | 0.06 |
| pHILIC | 154.02 | 65.6032 | 27.5 | 0.06 |
| pHILIC | 154.02 | 113.886 | 27.5 | 0.06 |
| pHILIC | 154.02 | 137.449 | 27.5 | 0.06 |
| pHILIC | 154.02 | 89.8187 | 27.5 | 0.06 |
| pHILIC | 154.02 | 103.8815 | 27.5 | 0.06 |
| pHILIC | 154.02 | 147.6029 | 27.5 | 0.06 |
| pHILIC | 154.02 | 81.5457 | 27.5 | 0.06 |
| pHILIC | 154.02 | 71.8721 | 27.5 | 0.06 |
| pHILIC | 154.02 | 117.0874 | 27.5 | 0.06 |
| pHILIC | 154.02 | 42.9832 | 27.5 | 0.06 |
| pHILIC | 154.02 | 48.3585 | 27.5 | 0.06 |
| pHILIC | 154.02 | 111.5916 | 27.5 | 0.06 |
| pHILIC | 154.02 | 53.4509 | 27.5 | 0.06 |
| pHILIC | 154.02 | 45.1166 | 27.5 | 0.06 |
| pHILIC | 154.02 | 49.0344 | 27.5 | 0.06 |
| pHILIC | 154.02 | 44.3638 | 27.5 | 0.06 |
| pHILIC | 154.02 | 67.0926 | 27.5 | 0.06 |
| pHILIC | 154.02 | 40.8625 | 27.5 | 0.06 |
| pHILIC | 154.02 | 73.415 | 27.5 | 0.06 |
| pHILIC | 154.02 | 63.4749 | 27.5 | 0.06 |
| pHILIC | 154.02 | 106.3388 | 27.4 | 0.06 |

| | | | | |
|---|---|---|---|---|
| pHILIC | 154.02 | 167.7681 | 27.4 | 0.06 |
| pHILIC | 154.02 | 123.3772 | 27.4 | 0.06 |
| pHILIC | 154.02 | 47.9148 | 27.4 | 0.06 |
| pHILIC | 154.02 | 61.1323 | 27.4 | 0.06 |
| pHILIC | 154.02 | 43.4282 | 27.4 | 0.06 |
| pHILIC | 154.02 | 158.8189 | 27.4 | 0.06 |
| pHILIC | 154.02 | 64.7785 | 27.4 | 0.06 |
| pHILIC | 154.02 | 66.7765 | 27.4 | 0.06 |
| pHILIC | 154.02 | 63.2038 | 27.4 | 0.06 |
| pHILIC | 154.02 | 54.8602 | 27.4 | 0.06 |
| pHILIC | 154.02 | 67.7144 | 27.4 | 0.06 |
| pHILIC | 154.02 | 40.807 | 27.4 | 0.06 |
| pHILIC | 154.02 | 109.9665 | 27.4 | 0.06 |
| pHILIC | 154.02 | 88.9372 | 27.4 | 0.06 |
| pHILIC | 154.02 | 64.9354 | 27.3 | 0.06 |
| pHILIC | 154.02 | 157.9079 | 27.3 | 0.06 |
| pHILIC | 154.02 | 57.2377 | 27.3 | 0.06 |
| pHILIC | 154.02 | 45.4732 | 27.3 | 0.06 |
| pHILIC | 154.02 | 111.6589 | 27.3 | 0.06 |
| pHILIC | 154.02 | 63.5399 | 27.3 | 0.06 |
| pHILIC | 154.02 | 52.3315 | 27.3 | 0.06 |
| pHILIC | 154.02 | 55.3859 | 27.2 | 0.06 |
| pHILIC | 154.02 | 41.2723 | 27.2 | 0.06 |
| pHILIC | 154.02 | 43.3074 | 27.2 | 0.06 |
| pHILIC | 154.02 | 78.8684 | 27.2 | 0.06 |
| pHILIC | 154.02 | 44.5023 | 27.2 | 0.06 |
| pHILIC | 154.02 | 40.159 | 27.2 | 0.06 |
| pHILIC | 154.02 | 117.9975 | 27.2 | 0.06 |
| pHILIC | 154.02 | 137.0067 | 27.2 | 0.06 |
| pHILIC | 154.02 | 62.6875 | 27.2 | 0.06 |
| pHILIC | 154.02 | 138.9822 | 27.2 | 0.06 |
| pHILIC | 154.02 | 45.3267 | 27.2 | 0.06 |
| pHILIC | 154.02 | 53.1301 | 27.2 | 0.06 |
| pHILIC | 154.02 | 71.0942 | 27.2 | 0.06 |
| pHILIC | 154.02 | 90.7798 | 27.2 | 0.06 |
| pHILIC | 154.02 | 148.9815 | 27.2 | 0.06 |
| pHILIC | 154.02 | 45.2582 | 27.2 | 0.06 |
| pHILIC | 154.02 | 48.6644 | 27.2 | 0.06 |
| pHILIC | 154.02 | 79.4722 | 27.2 | 0.06 |
| pHILIC | 154.02 | 47.6238 | 27.2 | 0.06 |
| pHILIC | 154.02 | 49.5375 | 27.2 | 0.06 |
| pHILIC | 154.02 | 41.2273 | 27.1 | 0.06 |

| | | | | |
|---|---|---|---|---|
| pHILIC | 154.02 | 134.1474 | 27.1 | 0.06 |
| pHILIC | 154.02 | 93.9067 | 27.1 | 0.06 |
| pHILIC | 154.02 | 101.9993 | 27.1 | 0.06 |
| pHILIC | 154.02 | 47.4167 | 27.1 | 0.06 |
| pHILIC | 154.02 | 55.9578 | 27.1 | 0.06 |
| pHILIC | 154.02 | 55.9669 | 27.1 | 0.06 |
| pHILIC | 154.02 | 47.7262 | 27.1 | 0.06 |
| pHILIC | 154.02 | 65.0959 | 27.1 | 0.06 |
| pHILIC | 154.02 | 40.9702 | 27.1 | 0.06 |
| pHILIC | 154.02 | 78.9248 | 27.1 | 0.06 |
| pHILIC | 154.02 | 78.5056 | 27.1 | 0.06 |
| pHILIC | 154.02 | 48.5647 | 27.1 | 0.06 |
| pHILIC | 154.02 | 57.1037 | 27.1 | 0.06 |
| pHILIC | 154.02 | 86.925 | 27.1 | 0.06 |
| pHILIC | 154.02 | 43.5028 | 27.1 | 0.06 |
| pHILIC | 154.02 | 45.6837 | 27.1 | 0.06 |
| pHILIC | 154.02 | 99.3488 | 27.1 | 0.06 |
| pHILIC | 154.02 | 41.1569 | 27.1 | 0.06 |
| pHILIC | 154.02 | 52.8666 | 27.1 | 0.06 |
| pHILIC | 154.02 | 144.3233 | 27 | 0.06 |
| pHILIC | 154.02 | 81.7404 | 27 | 0.06 |
| pHILIC | 154.02 | 42.4664 | 27 | 0.06 |
| pHILIC | 154.02 | 47.0387 | 27 | 0.06 |
| pHILIC | 154.02 | 124.6673 | 27 | 0.06 |
| pHILIC | 154.02 | 98.434 | 27 | 0.06 |
| pHILIC | 154.02 | 66.374 | 27 | 0.06 |
| pHILIC | 154.02 | 149.6889 | 27 | 0.06 |
| pHILIC | 154.02 | 65.5032 | 27 | 0.06 |
| pHILIC | 154.02 | 93.0464 | 27 | 0.06 |
| pHILIC | 154.02 | 49.2229 | 26.9 | 0.06 |
| pHILIC | 154.02 | 54.8959 | 26.9 | 0.06 |
| pHILIC | 154.02 | 40.5039 | 26.9 | 0.06 |
| pHILIC | 154.02 | 43.7474 | 26.9 | 0.06 |
| pHILIC | 154.02 | 88.9774 | 26.9 | 0.06 |
| pHILIC | 154.02 | 114.4716 | 26.9 | 0.06 |
| pHILIC | 154.02 | 98.1438 | 26.9 | 0.06 |
| pHILIC | 154.02 | 41.1332 | 26.9 | 0.06 |
| pHILIC | 154.02 | 45.3131 | 26.9 | 0.06 |
| pHILIC | 154.02 | 62.2897 | 26.9 | 0.06 |
| pHILIC | 154.02 | 113.1354 | 26.9 | 0.06 |
| pHILIC | 154.02 | 84.4701 | 26.8 | 0.06 |
| pHILIC | 154.02 | 42.0199 | 26.8 | 0.06 |

| | | | | |
|---|---|---|---|---|
| pHILIC | 154.02 | 129.7381 | 26.8 | 0.06 |
| pHILIC | 154.02 | 41.8235 | 26.8 | 0.06 |
| pHILIC | 154.02 | 43.288 | 26.8 | 0.06 |
| pHILIC | 154.02 | 124.8966 | 26.8 | 0.06 |
| pHILIC | 154.02 | 44.3772 | 26.8 | 0.06 |
| pHILIC | 154.02 | 44.4572 | 26.7 | 0.06 |
| pHILIC | 154.02 | 69.4321 | 26.7 | 0.06 |
| pHILIC | 154.02 | 46.8029 | 26.7 | 0.06 |
| pHILIC | 154.02 | 58.0436 | 26.7 | 0.06 |
| pHILIC | 154.02 | 40.3023 | 26.7 | 0.06 |
| pHILIC | 154.02 | 41.6927 | 26.6 | 0.06 |
| pHILIC | 154.02 | 89.5768 | 26.6 | 0.06 |
| pHILIC | 154.02 | 79.0867 | 26.6 | 0.06 |
| pHILIC | 154.02 | 123.0718 | 26.6 | 0.06 |
| pHILIC | 154.02 | 51.3646 | 26.6 | 0.06 |
| pHILIC | 154.02 | 50.731 | 26.6 | 0.06 |
| pHILIC | 154.02 | 66.6548 | 26.6 | 0.06 |
| pHILIC | 154.02 | 74.9627 | 26.6 | 0.06 |
| pHILIC | 154.02 | 55.2205 | 26.6 | 0.06 |
| pHILIC | 154.02 | 80.2296 | 26.6 | 0.06 |
| pHILIC | 154.02 | 51.9848 | 26.5 | 0.06 |
| pHILIC | 154.02 | 46.5423 | 26.5 | 0.06 |
| pHILIC | 154.02 | 86.4773 | 26.5 | 0.06 |
| pHILIC | 154.02 | 49.3771 | 26.5 | 0.06 |
| pHILIC | 154.02 | 99.6754 | 26.5 | 0.06 |
| pHILIC | 154.02 | 46.0108 | 26.5 | 0.06 |
| pHILIC | 154.02 | 51.7458 | 26.4 | 0.06 |
| pHILIC | 154.02 | 48.4681 | 26.4 | 0.06 |
| pHILIC | 154.02 | 61.7341 | 26.4 | 0.06 |
| pHILIC | 154.02 | 52.9219 | 26.4 | 0.06 |
| pHILIC | 154.02 | 62.896 | 26.4 | 0.06 |
| pHILIC | 154.02 | 86.8385 | 26.4 | 0.06 |
| pHILIC | 154.02 | 117.4427 | 26.3 | 0.06 |
| pHILIC | 154.02 | 61.5852 | 26.3 | 0.06 |
| pHILIC | 154.02 | 50.8971 | 26.3 | 0.06 |
| pHILIC | 154.02 | 62.4506 | 26.2 | 0.06 |
| pHILIC | 154.02 | 140.5212 | 26.2 | 0.06 |
| pHILIC | 154.02 | 67.1733 | 26.2 | 0.06 |
| pHILIC | 154.02 | 41.7386 | 26.1 | 0.06 |
| pHILIC | 154.02 | 42.0518 | 26.1 | 0.06 |
| pHILIC | 154.02 | 56.8245 | 26 | 0.06 |
| pHILIC | 154.02 | 44.7325 | 26 | 0.06 |

| | | | | |
|---|---|---|---|---|
| pHILIC | 154.02 | 40.9146 | 26 | 0.06 |
| pHILIC | 154.02 | 168.3957 | 26 | 0.06 |
| pHILIC | 154.02 | 40.3737 | 26 | 0.06 |
| pHILIC | 154.02 | 76.7754 | 26 | 0.06 |
| pHILIC | 154.02 | 88.8234 | 25.9 | 0.06 |
| pHILIC | 154.02 | 43.4203 | 25.9 | 0.06 |
| pHILIC | 154.02 | 47.8734 | 25.9 | 0.06 |
| pHILIC | 154.02 | 41.278 | 25.9 | 0.06 |
| pHILIC | 154.02 | 79.8795 | 25.7 | 0.06 |
| pHILIC | 154.02 | 116.7793 | 25.7 | 0.06 |
| pHILIC | 154.02 | 92.754 | 25.6 | 0.06 |
| pHILIC | 154.02 | 44.5245 | 25.6 | 0.06 |
| pHILIC | 154.02 | 40.5882 | 25.6 | 0.06 |
| pHILIC | 154.02 | 48.185 | 25.6 | 0.06 |
| pHILIC | 154.02 | 41.3446 | 25.5 | 0.06 |
| pHILIC | 154.02 | 42.4593 | 25.4 | 0.06 |
| pHILIC | 154.02 | 41.2042 | 25.4 | 0.06 |
| pHILIC | 154.02 | 136.0458 | 25.4 | 0.06 |
| pHILIC | 154.02 | 47.2224 | 25.3 | 0.06 |
| pHILIC | 154.02 | 41.5257 | 25.3 | 0.06 |
| pHILIC | 154.02 | 43.2149 | 25 | 0.06 |
| pHILIC | 154.02 | 40.5612 | 24.9 | 0.06 |
| pHILIC | 154.02 | 64.9813 | 24.6 | 0.06 |
| pHILIC | 154.02 | 40.455 | 22.6 | 0.05 |
| pHILIC | 186.112 | 186.1239 | 110674.8 | 100 |
| pHILIC | 186.112 | 140.1183 | 2801.5 | 2.53 |
| pHILIC | 186.112 | 168.102 | 713.4 | 0.64 |
| pHILIC | 186.112 | 186.2213 | 533.9 | 0.48 |
| pHILIC | 186.112 | 186.1094 | 455.9 | 0.41 |
| pHILIC | 186.112 | 186.1445 | 367.4 | 0.33 |
| pHILIC | 186.112 | 186.0898 | 269.2 | 0.24 |
| pHILIC | 186.112 | 186.1868 | 145.1 | 0.13 |
| pHILIC | 186.112 | 112.896 | 133.5 | 0.12 |
| pHILIC | 186.112 | 57.0698 | 110.9 | 0.1 |
| pHILIC | 186.112 | 109.0134 | 100.5 | 0.09 |
| pHILIC | 186.112 | 59.1056 | 77.6 | 0.07 |
| pHILIC | 186.112 | 93.9606 | 77.4 | 0.07 |
| pHILIC | 186.112 | 70.9384 | 76.2 | 0.07 |
| pHILIC | 186.112 | 99.1871 | 74.7 | 0.07 |
| pHILIC | 186.112 | 54.9981 | 72.9 | 0.07 |
| pHILIC | 186.112 | 131.7527 | 72.5 | 0.07 |
| pHILIC | 186.112 | 55.7303 | 72.1 | 0.07 |

| | | | | |
|---|---|---|---|---|
| pHILIC | 186.112 | 50.9386 | 70.6 | 0.06 |
| pHILIC | 186.112 | 47.7571 | 69.2 | 0.06 |
| pHILIC | 186.112 | 47.2161 | 68.2 | 0.06 |
| pHILIC | 186.112 | 173.517 | 67.7 | 0.06 |
| pHILIC | 186.112 | 45.7434 | 67.6 | 0.06 |
| pHILIC | 186.112 | 45.2942 | 67.4 | 0.06 |
| pHILIC | 186.112 | 132.6136 | 66.9 | 0.06 |
| pHILIC | 186.112 | 84.2036 | 66.5 | 0.06 |
| pHILIC | 186.112 | 155.6782 | 66.2 | 0.06 |
| pHILIC | 186.112 | 64.0793 | 66.1 | 0.06 |
| pHILIC | 186.112 | 46.9495 | 65.9 | 0.06 |
| pHILIC | 186.112 | 146.2507 | 65.7 | 0.06 |
| pHILIC | 186.112 | 46.4716 | 65.2 | 0.06 |
| pHILIC | 186.112 | 120.1674 | 64.9 | 0.06 |
| pHILIC | 186.112 | 62.2049 | 64.8 | 0.06 |
| pHILIC | 186.112 | 104.1551 | 64.4 | 0.06 |
| pHILIC | 186.112 | 128.5514 | 64.1 | 0.06 |
| pHILIC | 186.112 | 115.3928 | 63 | 0.06 |
| pHILIC | 186.112 | 141.8305 | 61.5 | 0.06 |
| pHILIC | 186.112 | 118.3256 | 48.8 | 0.04 |
| pHILIC | 186.112 | 168.2062 | 48.3 | 0.04 |
| pHILIC | 186.112 | 44.0494 | 47.7 | 0.04 |
| pHILIC | 186.112 | 64.7256 | 47.5 | 0.04 |
| pHILIC | 186.112 | 103.5793 | 47.3 | 0.04 |
| pHILIC | 186.112 | 114.3136 | 46.9 | 0.04 |
| pHILIC | 186.112 | 203.3346 | 46.9 | 0.04 |
| pHILIC | 186.112 | 121.6632 | 46.6 | 0.04 |
| pHILIC | 186.112 | 65.588 | 46.5 | 0.04 |
| pHILIC | 186.112 | 88.8057 | 46.2 | 0.04 |
| pHILIC | 186.112 | 64.528 | 46.1 | 0.04 |
| pHILIC | 186.112 | 63.9791 | 45.8 | 0.04 |
| pHILIC | 186.112 | 78.1085 | 45.1 | 0.04 |
| pHILIC | 186.112 | 57.1582 | 45.1 | 0.04 |
| pHILIC | 186.112 | 115.474 | 45 | 0.04 |
| pHILIC | 186.112 | 120.9339 | 44.9 | 0.04 |
| pHILIC | 186.112 | 47.7518 | 44.7 | 0.04 |
| pHILIC | 186.112 | 166.7699 | 44.7 | 0.04 |
| pHILIC | 186.112 | 102.9383 | 44.6 | 0.04 |
| pHILIC | 186.112 | 68.3949 | 44.4 | 0.04 |
| pHILIC | 186.112 | 94.0865 | 44.4 | 0.04 |
| pHILIC | 186.112 | 93.8369 | 44.4 | 0.04 |
| pHILIC | 186.112 | 69.2404 | 44.3 | 0.04 |

| | | | | |
|---|---|---|---|---|
| pHILIC | 186.112 | 42.7755 | 44.3 | 0.04 |
| pHILIC | 186.112 | 176.1949 | 44.1 | 0.04 |
| pHILIC | 186.112 | 45.4345 | 44.1 | 0.04 |
| pHILIC | 186.112 | 75.222 | 44.1 | 0.04 |
| pHILIC | 186.112 | 143.6932 | 43.9 | 0.04 |
| pHILIC | 186.112 | 69.7326 | 43.8 | 0.04 |
| pHILIC | 186.112 | 80.428 | 43.8 | 0.04 |
| pHILIC | 186.112 | 54.6603 | 43.8 | 0.04 |
| pHILIC | 186.112 | 71.7286 | 43.7 | 0.04 |
| pHILIC | 186.112 | 149.7212 | 43.6 | 0.04 |
| pHILIC | 186.112 | 75.4476 | 43.5 | 0.04 |
| pHILIC | 186.112 | 100.6752 | 43.5 | 0.04 |
| pHILIC | 186.112 | 176.0827 | 43.4 | 0.04 |
| pHILIC | 186.112 | 52.8492 | 43.4 | 0.04 |
| pHILIC | 186.112 | 42.7449 | 43.3 | 0.04 |
| pHILIC | 186.112 | 45.5381 | 43.1 | 0.04 |
| pHILIC | 186.112 | 135.0346 | 43.1 | 0.04 |
| pHILIC | 186.112 | 178.6388 | 43 | 0.04 |
| pHILIC | 186.112 | 124.6269 | 43 | 0.04 |
| pHILIC | 186.112 | 206.1543 | 42.9 | 0.04 |
| pHILIC | 186.112 | 64.0818 | 42.8 | 0.04 |
| pHILIC | 186.112 | 87.8423 | 42.7 | 0.04 |
| pHILIC | 186.112 | 180.3901 | 42.6 | 0.04 |
| pHILIC | 186.112 | 175.5728 | 42.6 | 0.04 |
| pHILIC | 186.112 | 72.6071 | 42.4 | 0.04 |
| pHILIC | 186.112 | 93.8792 | 42.4 | 0.04 |
| pHILIC | 186.112 | 111.9607 | 42.4 | 0.04 |
| pHILIC | 186.112 | 51.9316 | 42.4 | 0.04 |
| pHILIC | 186.112 | 106.4247 | 42.2 | 0.04 |
| pHILIC | 186.112 | 169.6992 | 42.2 | 0.04 |
| pHILIC | 186.112 | 91.6884 | 42.1 | 0.04 |
| pHILIC | 186.112 | 94.8409 | 42.1 | 0.04 |
| pHILIC | 186.112 | 162.7038 | 42.1 | 0.04 |
| pHILIC | 186.112 | 63.3515 | 42 | 0.04 |
| pHILIC | 186.112 | 43.0213 | 41.9 | 0.04 |
| pHILIC | 186.112 | 133.6727 | 41.9 | 0.04 |
| pHILIC | 186.112 | 73.0609 | 41.9 | 0.04 |
| pHILIC | 186.112 | 193.813 | 41.8 | 0.04 |
| pHILIC | 186.112 | 77.0528 | 41.8 | 0.04 |
| pHILIC | 186.112 | 45.586 | 41.7 | 0.04 |
| pHILIC | 186.112 | 99.3056 | 41.7 | 0.04 |
| pHILIC | 186.112 | 42.8465 | 41.7 | 0.04 |

| | | | | |
|---|---|---|---|---|
| pHILIC | 186.112 | 50.8279 | 41.7 | 0.04 |
| pHILIC | 186.112 | 59.8222 | 41.7 | 0.04 |
| pHILIC | 186.112 | 139.4174 | 41.6 | 0.04 |
| pHILIC | 186.112 | 93.8621 | 41.6 | 0.04 |
| pHILIC | 186.112 | 43.9909 | 41.5 | 0.04 |
| pHILIC | 186.112 | 44.2111 | 41.5 | 0.04 |
| pHILIC | 186.112 | 89.5645 | 41.4 | 0.04 |
| pHILIC | 186.112 | 167.2982 | 41.4 | 0.04 |
| pHILIC | 186.112 | 46.0927 | 41.4 | 0.04 |
| pHILIC | 186.112 | 111.2482 | 41.4 | 0.04 |
| pHILIC | 186.112 | 59.2154 | 41.3 | 0.04 |
| pHILIC | 186.112 | 132.583 | 41.3 | 0.04 |
| pHILIC | 186.112 | 107.9896 | 41.3 | 0.04 |
| pHILIC | 186.112 | 61.7879 | 41.3 | 0.04 |
| pHILIC | 186.112 | 104.0082 | 41.2 | 0.04 |
| pHILIC | 186.112 | 127.5307 | 41.2 | 0.04 |
| pHILIC | 186.112 | 54.8805 | 41.2 | 0.04 |
| pHILIC | 186.112 | 46.8898 | 41.1 | 0.04 |
| pHILIC | 186.112 | 133.4063 | 41.1 | 0.04 |
| pHILIC | 186.112 | 94.1834 | 41.1 | 0.04 |
| pHILIC | 186.112 | 186.7878 | 41.1 | 0.04 |
| pHILIC | 186.112 | 103.8616 | 41 | 0.04 |
| pHILIC | 186.112 | 72.992 | 41 | 0.04 |
| pHILIC | 186.112 | 50.8776 | 41 | 0.04 |
| pHILIC | 186.112 | 69.8549 | 41 | 0.04 |
| pHILIC | 186.112 | 90.0175 | 40.9 | 0.04 |
| pHILIC | 186.112 | 99.8309 | 40.9 | 0.04 |
| pHILIC | 186.112 | 101.3749 | 40.9 | 0.04 |
| pHILIC | 186.112 | 170.5787 | 40.8 | 0.04 |
| pHILIC | 186.112 | 136.275 | 40.8 | 0.04 |
| pHILIC | 186.112 | 59.0795 | 40.7 | 0.04 |
| pHILIC | 186.112 | 183.4119 | 40.7 | 0.04 |
| pHILIC | 186.112 | 139.8389 | 40.7 | 0.04 |
| pHILIC | 186.112 | 148.5773 | 40.7 | 0.04 |
| pHILIC | 186.112 | 79.476 | 40.7 | 0.04 |
| pHILIC | 186.112 | 141.1637 | 40.7 | 0.04 |
| pHILIC | 186.112 | 150.3473 | 40.7 | 0.04 |
| pHILIC | 186.112 | 51.7648 | 40.6 | 0.04 |
| pHILIC | 186.112 | 93.4375 | 40.6 | 0.04 |
| pHILIC | 186.112 | 96.2991 | 40.6 | 0.04 |
| pHILIC | 186.112 | 188.6149 | 40.6 | 0.04 |
| pHILIC | 186.112 | 63.8177 | 40.5 | 0.04 |

| | | | | |
|---|---|---|---|---|
| pHILIC | 186.112 | 61.0186 | 40.5 | 0.04 |
| pHILIC | 186.112 | 92.0893 | 40.5 | 0.04 |
| pHILIC | 186.112 | 112.7508 | 40.5 | 0.04 |
| pHILIC | 186.112 | 67.1448 | 40.5 | 0.04 |
| pHILIC | 186.112 | 176.2473 | 40.5 | 0.04 |
| pHILIC | 186.112 | 102.151 | 40.5 | 0.04 |
| pHILIC | 186.112 | 195.4251 | 40.5 | 0.04 |
| pHILIC | 186.112 | 118.9346 | 40.4 | 0.04 |
| pHILIC | 186.112 | 106.2371 | 40.4 | 0.04 |
| pHILIC | 186.112 | 183.6564 | 40.4 | 0.04 |
| pHILIC | 186.112 | 206.1674 | 40.4 | 0.04 |
| pHILIC | 186.112 | 92.5778 | 40.4 | 0.04 |
| pHILIC | 186.112 | 78.1849 | 40.4 | 0.04 |
| pHILIC | 186.112 | 45.2513 | 40.4 | 0.04 |
| pHILIC | 186.112 | 122.5339 | 40.3 | 0.04 |
| pHILIC | 186.112 | 89.265 | 40.3 | 0.04 |
| pHILIC | 186.112 | 43.7017 | 40.3 | 0.04 |
| pHILIC | 186.112 | 52.6722 | 40.3 | 0.04 |
| pHILIC | 186.112 | 49.2267 | 40.3 | 0.04 |
| pHILIC | 186.112 | 95.6814 | 40.3 | 0.04 |
| pHILIC | 186.112 | 178.8836 | 40.3 | 0.04 |
| pHILIC | 186.112 | 182.0611 | 40.2 | 0.04 |
| pHILIC | 186.112 | 93.1853 | 40.2 | 0.04 |
| pHILIC | 186.112 | 47.3893 | 40.2 | 0.04 |
| pHILIC | 186.112 | 87.5335 | 40.2 | 0.04 |
| pHILIC | 186.112 | 47.0167 | 40.2 | 0.04 |
| pHILIC | 186.112 | 87.2067 | 40.2 | 0.04 |
| pHILIC | 186.112 | 143.0285 | 40.2 | 0.04 |
| pHILIC | 186.112 | 60.6412 | 40.1 | 0.04 |
| pHILIC | 186.112 | 97.29 | 40.1 | 0.04 |
| pHILIC | 186.112 | 64.9292 | 40.1 | 0.04 |
| pHILIC | 186.112 | 144.0109 | 40 | 0.04 |
| pHILIC | 186.112 | 91.5508 | 40 | 0.04 |
| pHILIC | 186.112 | 116.2152 | 40 | 0.04 |
| pHILIC | 186.112 | 93.972 | 40 | 0.04 |
| pHILIC | 186.112 | 48.2502 | 40 | 0.04 |
| pHILIC | 186.112 | 109.8672 | 39.9 | 0.04 |
| pHILIC | 186.112 | 90.1919 | 39.9 | 0.04 |
| pHILIC | 186.112 | 56.9083 | 39.9 | 0.04 |
| pHILIC | 186.112 | 120.9931 | 39.9 | 0.04 |
| pHILIC | 186.112 | 200.7417 | 39.9 | 0.04 |
| pHILIC | 186.112 | 48.2532 | 39.9 | 0.04 |

| | | | | |
|---|---|---|---|---|
| pHILIC | 186.112 | 52.0462 | 39.8 | 0.04 |
| pHILIC | 186.112 | 48.4302 | 39.8 | 0.04 |
| pHILIC | 186.112 | 47.9124 | 39.8 | 0.04 |
| pHILIC | 186.112 | 45.0235 | 39.8 | 0.04 |
| pHILIC | 186.112 | 122.8074 | 39.7 | 0.04 |
| pHILIC | 186.112 | 51.416 | 39.7 | 0.04 |
| pHILIC | 186.112 | 44.7417 | 39.7 | 0.04 |
| pHILIC | 186.112 | 168.9172 | 39.7 | 0.04 |
| pHILIC | 186.112 | 100.6839 | 39.7 | 0.04 |
| pHILIC | 186.112 | 157.31 | 39.7 | 0.04 |
| pHILIC | 186.112 | 65.0355 | 39.7 | 0.04 |
| pHILIC | 186.112 | 108.0285 | 39.7 | 0.04 |
| pHILIC | 186.112 | 49.0999 | 39.7 | 0.04 |
| pHILIC | 186.112 | 151.2968 | 39.7 | 0.04 |
| pHILIC | 186.112 | 109.4031 | 39.7 | 0.04 |
| pHILIC | 186.112 | 65.1315 | 39.7 | 0.04 |
| pHILIC | 186.112 | 185.9126 | 39.7 | 0.04 |
| pHILIC | 186.112 | 183.1215 | 39.6 | 0.04 |
| pHILIC | 186.112 | 47.1826 | 39.6 | 0.04 |
| pHILIC | 186.112 | 65.3834 | 39.6 | 0.04 |
| pHILIC | 186.112 | 157.3683 | 39.5 | 0.04 |
| pHILIC | 186.112 | 65.9639 | 39.5 | 0.04 |
| pHILIC | 186.112 | 73.0498 | 39.5 | 0.04 |
| pHILIC | 186.112 | 144.7566 | 39.5 | 0.04 |
| pHILIC | 186.112 | 143.7224 | 39.5 | 0.04 |
| pHILIC | 186.112 | 68.2933 | 39.4 | 0.04 |
| pHILIC | 186.112 | 134.3711 | 39.4 | 0.04 |
| pHILIC | 186.112 | 188.6274 | 39.4 | 0.04 |
| pHILIC | 186.112 | 165.8214 | 39.4 | 0.04 |
| pHILIC | 186.112 | 56.9002 | 39.4 | 0.04 |
| pHILIC | 186.112 | 83.2036 | 39.4 | 0.04 |
| pHILIC | 186.112 | 157.5089 | 39.4 | 0.04 |
| pHILIC | 186.112 | 58.0212 | 39.4 | 0.04 |
| pHILIC | 186.112 | 55.6175 | 39.3 | 0.04 |
| pHILIC | 186.112 | 93.3943 | 39.3 | 0.04 |
| pHILIC | 186.112 | 95.9042 | 39.3 | 0.04 |
| pHILIC | 186.112 | 52.6601 | 39.3 | 0.04 |
| pHILIC | 186.112 | 55.2962 | 39.3 | 0.04 |
| pHILIC | 186.112 | 102.5626 | 39.3 | 0.04 |
| pHILIC | 186.112 | 90.4796 | 39.3 | 0.04 |
| pHILIC | 186.112 | 105.2537 | 39.3 | 0.04 |
| pHILIC | 186.112 | 133.1825 | 39.3 | 0.04 |

| | | | | |
|---|---|---|---|---|
| pHILIC | 186.112 | 48.0178 | 39.3 | 0.04 |
| pHILIC | 186.112 | 114.2815 | 39.2 | 0.04 |
| pHILIC | 186.112 | 46.8242 | 39.2 | 0.04 |
| pHILIC | 186.112 | 167.1235 | 39.2 | 0.04 |
| pHILIC | 186.112 | 132.2497 | 39.2 | 0.04 |
| pHILIC | 186.112 | 118.4136 | 39.2 | 0.04 |
| pHILIC | 186.112 | 48.1819 | 39.2 | 0.04 |
| pHILIC | 186.112 | 155.3748 | 39.2 | 0.04 |
| pHILIC | 186.112 | 66.0226 | 39.2 | 0.04 |
| pHILIC | 186.112 | 153.3001 | 39.1 | 0.04 |
| pHILIC | 186.112 | 125.231 | 39.1 | 0.04 |
| pHILIC | 186.112 | 59.0127 | 39.1 | 0.04 |
| pHILIC | 186.112 | 50.6532 | 39.1 | 0.04 |
| pHILIC | 186.112 | 59.9539 | 39.1 | 0.04 |
| pHILIC | 186.112 | 52.3238 | 39.1 | 0.04 |
| pHILIC | 186.112 | 51.7704 | 39.1 | 0.04 |
| pHILIC | 186.112 | 87.5137 | 39.1 | 0.04 |
| pHILIC | 186.112 | 94.1776 | 39.1 | 0.04 |
| pHILIC | 186.112 | 119.2504 | 39 | 0.04 |
| pHILIC | 186.112 | 46.8359 | 39 | 0.04 |
| pHILIC | 186.112 | 42.7642 | 39 | 0.04 |
| pHILIC | 186.112 | 89.8246 | 39 | 0.04 |
| pHILIC | 186.112 | 59.8262 | 39 | 0.04 |
| pHILIC | 186.112 | 65.3057 | 39 | 0.04 |
| pHILIC | 186.112 | 76.639 | 39 | 0.04 |
| pHILIC | 186.112 | 54.0425 | 39 | 0.04 |
| pHILIC | 186.112 | 179.4901 | 39 | 0.04 |
| pHILIC | 186.112 | 158.3932 | 39 | 0.04 |
| pHILIC | 186.112 | 62.7823 | 39 | 0.04 |
| pHILIC | 186.112 | 99.4179 | 38.9 | 0.04 |
| pHILIC | 186.112 | 53.1665 | 38.9 | 0.04 |
| pHILIC | 186.112 | 108.0252 | 38.9 | 0.04 |
| pHILIC | 186.112 | 171.8582 | 38.9 | 0.04 |
| pHILIC | 186.112 | 44.8311 | 38.9 | 0.04 |
| pHILIC | 186.112 | 90.5021 | 38.9 | 0.04 |
| pHILIC | 186.112 | 107.2922 | 38.9 | 0.04 |
| pHILIC | 186.112 | 188.4741 | 38.8 | 0.04 |
| pHILIC | 186.112 | 64.4427 | 38.8 | 0.04 |
| pHILIC | 186.112 | 118.8969 | 38.8 | 0.04 |
| pHILIC | 186.112 | 58.5214 | 38.8 | 0.04 |
| pHILIC | 186.112 | 45.1543 | 38.7 | 0.04 |
| pHILIC | 186.112 | 87.7896 | 38.7 | 0.04 |

| | | | | |
|---|---|---|---|---|
| pHILIC | 186.112 | 102.7713 | 38.7 | 0.03 |
| pHILIC | 186.112 | 210.2648 | 38.7 | 0.03 |
| pHILIC | 186.112 | 172.0826 | 38.7 | 0.03 |
| pHILIC | 186.112 | 115.408 | 38.7 | 0.03 |
| pHILIC | 186.112 | 46.3108 | 38.7 | 0.03 |
| pHILIC | 186.112 | 66.7479 | 38.7 | 0.03 |
| pHILIC | 186.112 | 62.9487 | 38.7 | 0.03 |
| pHILIC | 186.112 | 151.629 | 38.7 | 0.03 |
| pHILIC | 186.112 | 56.944 | 38.7 | 0.03 |
| pHILIC | 186.112 | 172.9646 | 38.7 | 0.03 |
| pHILIC | 186.112 | 97.6014 | 38.6 | 0.03 |
| pHILIC | 186.112 | 60.2461 | 38.6 | 0.03 |
| pHILIC | 186.112 | 63.5515 | 38.6 | 0.03 |
| pHILIC | 186.112 | 86.7179 | 38.6 | 0.03 |
| pHILIC | 186.112 | 71.4787 | 38.6 | 0.03 |
| pHILIC | 186.112 | 51.4799 | 38.6 | 0.03 |
| pHILIC | 186.112 | 138.2685 | 38.6 | 0.03 |
| pHILIC | 186.112 | 104.254 | 38.6 | 0.03 |
| pHILIC | 186.112 | 104.1145 | 38.6 | 0.03 |
| pHILIC | 186.112 | 169.1539 | 38.6 | 0.03 |
| pHILIC | 186.112 | 177.4713 | 38.6 | 0.03 |
| pHILIC | 186.112 | 48.9913 | 38.5 | 0.03 |
| pHILIC | 186.112 | 132.9695 | 38.5 | 0.03 |
| pHILIC | 186.112 | 56.0236 | 38.5 | 0.03 |
| pHILIC | 186.112 | 48.4103 | 38.5 | 0.03 |
| pHILIC | 186.112 | 84.595 | 38.5 | 0.03 |
| pHILIC | 186.112 | 56.0817 | 38.5 | 0.03 |
| pHILIC | 186.112 | 46.842 | 38.5 | 0.03 |
| pHILIC | 186.112 | 126.2609 | 38.4 | 0.03 |
| pHILIC | 186.112 | 60.7987 | 38.4 | 0.03 |
| pHILIC | 186.112 | 94.237 | 38.4 | 0.03 |
| pHILIC | 186.112 | 46.8225 | 38.4 | 0.03 |
| pHILIC | 186.112 | 151.3945 | 38.4 | 0.03 |
| pHILIC | 186.112 | 67.4516 | 38.4 | 0.03 |
| pHILIC | 186.112 | 70.904 | 38.4 | 0.03 |
| pHILIC | 186.112 | 112.7948 | 38.4 | 0.03 |
| pHILIC | 186.112 | 134.3237 | 38.4 | 0.03 |
| pHILIC | 186.112 | 113.1814 | 38.3 | 0.03 |
| pHILIC | 186.112 | 73.0972 | 38.3 | 0.03 |
| pHILIC | 186.112 | 108.5693 | 38.3 | 0.03 |
| pHILIC | 186.112 | 64.6211 | 38.3 | 0.03 |
| pHILIC | 186.112 | 94.7569 | 38.3 | 0.03 |

| | | | | |
|---|---|---|---|---|
| pHILIC | 186.112 | 97.6313 | 38.3 | 0.03 |
| pHILIC | 186.112 | 105.3675 | 38.3 | 0.03 |
| pHILIC | 186.112 | 98.7613 | 38.3 | 0.03 |
| pHILIC | 186.112 | 46.8134 | 38.3 | 0.03 |
| pHILIC | 186.112 | 49.1119 | 38.2 | 0.03 |
| pHILIC | 186.112 | 125.4273 | 38.2 | 0.03 |
| pHILIC | 186.112 | 125.6664 | 38.2 | 0.03 |
| pHILIC | 186.112 | 42.7621 | 38.2 | 0.03 |
| pHILIC | 186.112 | 143.0199 | 38.2 | 0.03 |
| pHILIC | 186.112 | 61.4678 | 38.2 | 0.03 |
| pHILIC | 186.112 | 99.3859 | 38.2 | 0.03 |
| pHILIC | 186.112 | 161.9491 | 38.2 | 0.03 |
| pHILIC | 186.112 | 76.6111 | 38.2 | 0.03 |
| pHILIC | 186.112 | 63.4269 | 38.2 | 0.03 |
| pHILIC | 186.112 | 65.4958 | 38.1 | 0.03 |
| pHILIC | 186.112 | 70.7675 | 38.1 | 0.03 |
| pHILIC | 186.112 | 68.1162 | 38.1 | 0.03 |
| pHILIC | 186.112 | 71.761 | 38.1 | 0.03 |
| pHILIC | 186.112 | 65.3676 | 38.1 | 0.03 |
| pHILIC | 186.112 | 61.0345 | 38.1 | 0.03 |
| pHILIC | 186.112 | 163.1987 | 38 | 0.03 |
| pHILIC | 186.112 | 58.7632 | 38 | 0.03 |
| pHILIC | 186.112 | 172.3652 | 38 | 0.03 |
| pHILIC | 186.112 | 65.2464 | 38 | 0.03 |
| pHILIC | 186.112 | 51.6255 | 38 | 0.03 |
| pHILIC | 186.112 | 102.6134 | 38 | 0.03 |
| pHILIC | 186.112 | 72.6528 | 38 | 0.03 |
| pHILIC | 186.112 | 94.7523 | 38 | 0.03 |
| pHILIC | 186.112 | 42.5724 | 38 | 0.03 |
| pHILIC | 186.112 | 46.6659 | 37.9 | 0.03 |
| pHILIC | 186.112 | 94.4869 | 37.9 | 0.03 |
| pHILIC | 186.112 | 66.2938 | 37.9 | 0.03 |
| pHILIC | 186.112 | 106.6452 | 37.9 | 0.03 |
| pHILIC | 186.112 | 44.7363 | 37.9 | 0.03 |
| pHILIC | 186.112 | 162.6292 | 37.9 | 0.03 |
| pHILIC | 186.112 | 50.3466 | 37.9 | 0.03 |
| pHILIC | 186.112 | 130.4794 | 37.9 | 0.03 |
| pHILIC | 186.112 | 51.2055 | 37.9 | 0.03 |
| pHILIC | 186.112 | 123.1016 | 37.9 | 0.03 |
| pHILIC | 186.112 | 103.6626 | 37.9 | 0.03 |
| pHILIC | 186.112 | 123.424 | 37.8 | 0.03 |
| pHILIC | 186.112 | 43.3386 | 37.8 | 0.03 |

| | | | | |
|---|---|---|---|---|
| pHILIC | 186.112 | 120.0998 | 37.8 | 0.03 |
| pHILIC | 186.112 | 194.1766 | 37.8 | 0.03 |
| pHILIC | 186.112 | 119.2083 | 37.8 | 0.03 |
| pHILIC | 186.112 | 44.2697 | 37.8 | 0.03 |
| pHILIC | 186.112 | 51.4863 | 37.8 | 0.03 |
| pHILIC | 186.112 | 55.0543 | 37.8 | 0.03 |
| pHILIC | 186.112 | 54.3372 | 37.8 | 0.03 |
| pHILIC | 186.112 | 42.7072 | 37.8 | 0.03 |
| pHILIC | 186.112 | 93.3383 | 37.8 | 0.03 |
| pHILIC | 186.112 | 66.3324 | 37.8 | 0.03 |
| pHILIC | 186.112 | 132.4948 | 37.7 | 0.03 |
| pHILIC | 186.112 | 45.6524 | 37.7 | 0.03 |
| pHILIC | 186.112 | 156.8742 | 37.7 | 0.03 |
| pHILIC | 186.112 | 56.9151 | 37.7 | 0.03 |
| pHILIC | 186.112 | 127.9903 | 37.7 | 0.03 |
| pHILIC | 186.112 | 128.9238 | 37.7 | 0.03 |
| pHILIC | 186.112 | 114.6903 | 37.7 | 0.03 |
| pHILIC | 186.112 | 68.4066 | 37.7 | 0.03 |
| pHILIC | 186.112 | 180.3526 | 37.6 | 0.03 |
| pHILIC | 186.112 | 62.0108 | 37.6 | 0.03 |
| pHILIC | 186.112 | 196.4004 | 37.6 | 0.03 |
| pHILIC | 186.112 | 65.4355 | 37.6 | 0.03 |
| pHILIC | 186.112 | 98.7209 | 37.6 | 0.03 |
| pHILIC | 186.112 | 86.4251 | 37.6 | 0.03 |
| pHILIC | 186.112 | 54.1971 | 37.6 | 0.03 |
| pHILIC | 186.112 | 105.2717 | 37.6 | 0.03 |
| pHILIC | 186.112 | 141.5918 | 37.6 | 0.03 |
| pHILIC | 186.112 | 169.735 | 37.5 | 0.03 |
| pHILIC | 186.112 | 70.4928 | 37.5 | 0.03 |
| pHILIC | 186.112 | 79.0554 | 37.5 | 0.03 |
| pHILIC | 186.112 | 43.2544 | 37.5 | 0.03 |
| pHILIC | 186.112 | 63.8524 | 37.5 | 0.03 |
| pHILIC | 186.112 | 134.0045 | 37.5 | 0.03 |
| pHILIC | 186.112 | 53.7584 | 37.5 | 0.03 |
| pHILIC | 186.112 | 140.9725 | 37.5 | 0.03 |
| pHILIC | 186.112 | 171.844 | 37.4 | 0.03 |
| pHILIC | 186.112 | 42.5964 | 37.4 | 0.03 |
| pHILIC | 186.112 | 84.0266 | 37.4 | 0.03 |
| pHILIC | 186.112 | 52.551 | 37.4 | 0.03 |
| pHILIC | 186.112 | 71.6969 | 37.4 | 0.03 |
| pHILIC | 186.112 | 52.5583 | 37.4 | 0.03 |
| pHILIC | 186.112 | 52.3565 | 37.4 | 0.03 |

| | | | | |
|---|---|---|---|---|
| pHILIC | 186.112 | 42.9266 | 37.4 | 0.03 |
| pHILIC | 186.112 | 115.8832 | 37.4 | 0.03 |
| pHILIC | 186.112 | 139.1666 | 37.4 | 0.03 |
| pHILIC | 186.112 | 49.1998 | 37.4 | 0.03 |
| pHILIC | 186.112 | 52.581 | 37.4 | 0.03 |
| pHILIC | 186.112 | 53.0197 | 37.3 | 0.03 |
| pHILIC | 186.112 | 120.7361 | 37.3 | 0.03 |
| pHILIC | 186.112 | 74.3897 | 37.3 | 0.03 |
| pHILIC | 186.112 | 130.463 | 37.3 | 0.03 |
| pHILIC | 186.112 | 42.079 | 37.3 | 0.03 |
| pHILIC | 186.112 | 74.5352 | 37.3 | 0.03 |
| pHILIC | 186.112 | 86.0694 | 37.2 | 0.03 |
| pHILIC | 186.112 | 50.6256 | 37.2 | 0.03 |
| pHILIC | 186.112 | 72.9816 | 37.2 | 0.03 |
| pHILIC | 186.112 | 122.5908 | 37.2 | 0.03 |
| pHILIC | 186.112 | 93.8224 | 37.2 | 0.03 |
| pHILIC | 186.112 | 207.8286 | 37.2 | 0.03 |
| pHILIC | 186.112 | 119.9837 | 37.2 | 0.03 |
| pHILIC | 186.112 | 154.9604 | 37.2 | 0.03 |
| pHILIC | 186.112 | 76.5635 | 37.2 | 0.03 |
| pHILIC | 186.112 | 113.4455 | 37.2 | 0.03 |
| pHILIC | 186.112 | 54.1682 | 37.2 | 0.03 |
| pHILIC | 186.112 | 58.5807 | 37.2 | 0.03 |
| pHILIC | 186.112 | 47.4857 | 37.2 | 0.03 |
| pHILIC | 186.112 | 69.2283 | 37.1 | 0.03 |
| pHILIC | 186.112 | 180.5947 | 37.1 | 0.03 |
| pHILIC | 186.112 | 141.8372 | 37.1 | 0.03 |
| pHILIC | 186.112 | 51.9516 | 37.1 | 0.03 |
| pHILIC | 186.112 | 59.9425 | 37.1 | 0.03 |
| pHILIC | 186.112 | 68.8399 | 37.1 | 0.03 |
| pHILIC | 186.112 | 74.8535 | 37.1 | 0.03 |
| pHILIC | 186.112 | 82.3522 | 37.1 | 0.03 |
| pHILIC | 186.112 | 54.2434 | 37.1 | 0.03 |
| pHILIC | 186.112 | 174.9051 | 37.1 | 0.03 |
| pHILIC | 186.112 | 81.6004 | 37.1 | 0.03 |
| pHILIC | 186.112 | 54.861 | 37.1 | 0.03 |
| pHILIC | 186.112 | 61.9541 | 37.1 | 0.03 |
| pHILIC | 186.112 | 47.8113 | 37.1 | 0.03 |
| pHILIC | 186.112 | 150.1793 | 37.1 | 0.03 |
| pHILIC | 186.112 | 54.3315 | 37 | 0.03 |
| pHILIC | 186.112 | 68.0167 | 37 | 0.03 |
| pHILIC | 186.112 | 89.4965 | 37 | 0.03 |

| | | | | |
|---|---|---|---|---|
| pHILIC | 186.112 | 95.4896 | 37 | 0.03 |
| pHILIC | 186.112 | 73.7692 | 37 | 0.03 |
| pHILIC | 186.112 | 113.6249 | 37 | 0.03 |
| pHILIC | 186.112 | 167.8065 | 37 | 0.03 |
| pHILIC | 186.112 | 57.7883 | 37 | 0.03 |
| pHILIC | 186.112 | 66.1902 | 37 | 0.03 |
| pHILIC | 186.112 | 44.7717 | 37 | 0.03 |
| pHILIC | 186.112 | 98.4021 | 37 | 0.03 |
| pHILIC | 186.112 | 53.9853 | 37 | 0.03 |
| pHILIC | 186.112 | 80.0363 | 37 | 0.03 |
| pHILIC | 186.112 | 59.0007 | 37 | 0.03 |
| pHILIC | 186.112 | 121.8594 | 37 | 0.03 |
| pHILIC | 186.112 | 107.9837 | 37 | 0.03 |
| pHILIC | 186.112 | 55.7014 | 37 | 0.03 |
| pHILIC | 186.112 | 161.6861 | 37 | 0.03 |
| pHILIC | 186.112 | 202.3268 | 37 | 0.03 |
| pHILIC | 186.112 | 50.0915 | 37 | 0.03 |
| pHILIC | 186.112 | 145.4652 | 37 | 0.03 |
| pHILIC | 186.112 | 47.4115 | 37 | 0.03 |
| pHILIC | 186.112 | 96.5866 | 37 | 0.03 |
| pHILIC | 186.112 | 157.2598 | 36.9 | 0.03 |
| pHILIC | 186.112 | 45.9168 | 36.9 | 0.03 |
| pHILIC | 186.112 | 130.1812 | 36.9 | 0.03 |
| pHILIC | 186.112 | 114.6377 | 36.9 | 0.03 |
| pHILIC | 186.112 | 42.4745 | 36.9 | 0.03 |
| pHILIC | 186.112 | 51.6762 | 36.9 | 0.03 |
| pHILIC | 186.112 | 191.2411 | 36.9 | 0.03 |
| pHILIC | 186.112 | 130.8235 | 36.9 | 0.03 |
| pHILIC | 186.112 | 77.1761 | 36.9 | 0.03 |
| pHILIC | 186.112 | 179.2387 | 36.9 | 0.03 |
| pHILIC | 186.112 | 158.955 | 36.9 | 0.03 |
| pHILIC | 186.112 | 204.6005 | 36.9 | 0.03 |
| pHILIC | 186.112 | 83.0095 | 36.9 | 0.03 |
| pHILIC | 186.112 | 105.1256 | 36.9 | 0.03 |
| pHILIC | 186.112 | 150.1927 | 36.9 | 0.03 |
| pHILIC | 186.112 | 172.2811 | 36.8 | 0.03 |
| pHILIC | 186.112 | 56.1781 | 36.8 | 0.03 |
| pHILIC | 186.112 | 64.8597 | 36.8 | 0.03 |
| pHILIC | 186.112 | 55.0038 | 36.8 | 0.03 |
| pHILIC | 186.112 | 205.03 | 36.8 | 0.03 |
| pHILIC | 186.112 | 47.7027 | 36.8 | 0.03 |
| pHILIC | 186.112 | 127.0727 | 36.8 | 0.03 |

| | | | | |
|---|---|---|---|---|
| pHILIC | 186.112 | 161.0145 | 36.8 | 0.03 |
| pHILIC | 186.112 | 131.3021 | 36.8 | 0.03 |
| pHILIC | 186.112 | 85.4241 | 36.7 | 0.03 |
| pHILIC | 186.112 | 69.3958 | 36.7 | 0.03 |
| pHILIC | 186.112 | 85.0001 | 36.7 | 0.03 |
| pHILIC | 186.112 | 57.9332 | 36.7 | 0.03 |
| pHILIC | 186.112 | 46.864 | 36.7 | 0.03 |
| pHILIC | 186.112 | 150.5604 | 36.7 | 0.03 |
| pHILIC | 186.112 | 54.8527 | 36.7 | 0.03 |
| pHILIC | 186.112 | 103.4869 | 36.7 | 0.03 |
| pHILIC | 186.112 | 160.0658 | 36.7 | 0.03 |
| pHILIC | 186.112 | 79.5393 | 36.7 | 0.03 |
| pHILIC | 186.112 | 63.0128 | 36.7 | 0.03 |
| pHILIC | 186.112 | 75.5512 | 36.7 | 0.03 |
| pHILIC | 186.112 | 81.576 | 36.7 | 0.03 |
| pHILIC | 186.112 | 100.0957 | 36.6 | 0.03 |
| pHILIC | 186.112 | 60.9495 | 36.6 | 0.03 |
| pHILIC | 186.112 | 46.7596 | 36.6 | 0.03 |
| pHILIC | 186.112 | 117.5808 | 36.6 | 0.03 |
| pHILIC | 186.112 | 48.4535 | 36.6 | 0.03 |
| pHILIC | 186.112 | 59.1266 | 36.6 | 0.03 |
| pHILIC | 186.112 | 102.4531 | 36.6 | 0.03 |
| pHILIC | 186.112 | 170.1092 | 36.6 | 0.03 |
| pHILIC | 186.112 | 72.8572 | 36.6 | 0.03 |
| pHILIC | 186.112 | 52.6933 | 36.6 | 0.03 |
| pHILIC | 186.112 | 114.0428 | 36.6 | 0.03 |
| pHILIC | 186.112 | 139.4351 | 36.6 | 0.03 |
| pHILIC | 186.112 | 46.0625 | 36.5 | 0.03 |
| pHILIC | 186.112 | 77.4175 | 36.5 | 0.03 |
| pHILIC | 186.112 | 87.8688 | 36.5 | 0.03 |
| pHILIC | 186.112 | 127.1245 | 36.5 | 0.03 |
| pHILIC | 186.112 | 47.9942 | 36.5 | 0.03 |
| pHILIC | 186.112 | 48.955 | 36.5 | 0.03 |
| pHILIC | 186.112 | 77.7227 | 36.5 | 0.03 |
| pHILIC | 186.112 | 62.3187 | 36.5 | 0.03 |
| pHILIC | 186.112 | 55.2564 | 36.5 | 0.03 |
| pHILIC | 186.112 | 76.9519 | 36.5 | 0.03 |
| pHILIC | 186.112 | 72.7283 | 36.5 | 0.03 |
| pHILIC | 186.112 | 42.7299 | 36.5 | 0.03 |
| pHILIC | 186.112 | 124.7825 | 36.5 | 0.03 |
| pHILIC | 186.112 | 96.5949 | 36.5 | 0.03 |
| pHILIC | 186.112 | 79.8437 | 36.5 | 0.03 |

| | | | | |
|---|---|---|---|---|
| pHILIC | 186.112 | 110.349 | 36.5 | 0.03 |
| pHILIC | 186.112 | 117.4963 | 36.5 | 0.03 |
| pHILIC | 186.112 | 181.0083 | 36.5 | 0.03 |
| pHILIC | 186.112 | 132.3023 | 36.5 | 0.03 |
| pHILIC | 186.112 | 126.6185 | 36.5 | 0.03 |
| pHILIC | 186.112 | 43.6843 | 36.5 | 0.03 |
| pHILIC | 186.112 | 141.1378 | 36.4 | 0.03 |
| pHILIC | 186.112 | 55.301 | 36.4 | 0.03 |
| pHILIC | 186.112 | 110.568 | 36.4 | 0.03 |
| pHILIC | 186.112 | 73.7354 | 36.4 | 0.03 |
| pHILIC | 186.112 | 209.6543 | 36.4 | 0.03 |
| pHILIC | 186.112 | 143.8818 | 36.4 | 0.03 |
| pHILIC | 186.112 | 43.674 | 36.4 | 0.03 |
| pHILIC | 186.112 | 42.4787 | 36.4 | 0.03 |
| pHILIC | 186.112 | 51.5293 | 36.4 | 0.03 |
| pHILIC | 186.112 | 55.758 | 36.4 | 0.03 |
| pHILIC | 186.112 | 116.8607 | 36.4 | 0.03 |
| pHILIC | 186.112 | 125.0544 | 36.4 | 0.03 |
| pHILIC | 186.112 | 48.575 | 36.4 | 0.03 |
| pHILIC | 186.112 | 123.7336 | 36.4 | 0.03 |
| pHILIC | 186.112 | 142.5195 | 36.4 | 0.03 |
| pHILIC | 186.112 | 153.3363 | 36.3 | 0.03 |
| pHILIC | 186.112 | 108.5546 | 36.3 | 0.03 |
| pHILIC | 186.112 | 80.2492 | 36.3 | 0.03 |
| pHILIC | 186.112 | 55.7914 | 36.3 | 0.03 |
| pHILIC | 186.112 | 115.48 | 36.3 | 0.03 |
| pHILIC | 186.112 | 99.4262 | 36.3 | 0.03 |
| pHILIC | 186.112 | 44.0933 | 36.3 | 0.03 |
| pHILIC | 186.112 | 133.784 | 36.3 | 0.03 |
| pHILIC | 186.112 | 80.5158 | 36.3 | 0.03 |
| pHILIC | 186.112 | 198.7105 | 36.3 | 0.03 |
| pHILIC | 186.112 | 113.6548 | 36.3 | 0.03 |
| pHILIC | 186.112 | 146.4298 | 36.3 | 0.03 |
| pHILIC | 186.112 | 46.0229 | 36.3 | 0.03 |
| pHILIC | 186.112 | 81.0062 | 36.3 | 0.03 |
| pHILIC | 186.112 | 68.5382 | 36.3 | 0.03 |
| pHILIC | 186.112 | 68.6028 | 36.3 | 0.03 |
| pHILIC | 186.112 | 52.9519 | 36.3 | 0.03 |
| pHILIC | 186.112 | 44.0176 | 36.3 | 0.03 |
| pHILIC | 186.112 | 115.3045 | 36.3 | 0.03 |
| pHILIC | 186.112 | 79.1034 | 36.3 | 0.03 |
| pHILIC | 186.112 | 56.4734 | 36.3 | 0.03 |

| | | | | |
|---|---|---|---|---|
| pHILIC | 186.112 | 90.1814 | 36.3 | 0.03 |
| pHILIC | 186.112 | 88.4371 | 36.3 | 0.03 |
| pHILIC | 186.112 | 85.1435 | 36.2 | 0.03 |
| pHILIC | 186.112 | 87.3581 | 36.2 | 0.03 |
| pHILIC | 186.112 | 100.2607 | 36.2 | 0.03 |
| pHILIC | 186.112 | 101.96 | 36.2 | 0.03 |
| pHILIC | 186.112 | 43.5856 | 36.2 | 0.03 |
| pHILIC | 186.112 | 48.8643 | 36.2 | 0.03 |
| pHILIC | 186.112 | 164.2284 | 36.2 | 0.03 |
| pHILIC | 186.112 | 76.6093 | 36.2 | 0.03 |
| pHILIC | 186.112 | 74.1649 | 36.2 | 0.03 |
| pHILIC | 186.112 | 155.5166 | 36.2 | 0.03 |
| pHILIC | 186.112 | 123.3163 | 36.2 | 0.03 |
| pHILIC | 186.112 | 62.1411 | 36.2 | 0.03 |
| pHILIC | 186.112 | 46.717 | 36.2 | 0.03 |
| pHILIC | 186.112 | 95.893 | 36.2 | 0.03 |
| pHILIC | 186.112 | 54.498 | 36.2 | 0.03 |
| pHILIC | 186.112 | 90.0528 | 36.2 | 0.03 |
| pHILIC | 186.112 | 99.3428 | 36.1 | 0.03 |
| pHILIC | 186.112 | 201.9203 | 36.1 | 0.03 |
| pHILIC | 186.112 | 43.5536 | 36.1 | 0.03 |
| pHILIC | 186.112 | 185.355 | 36.1 | 0.03 |
| pHILIC | 186.112 | 46.1408 | 36.1 | 0.03 |
| pHILIC | 186.112 | 48.0904 | 36.1 | 0.03 |
| pHILIC | 186.112 | 93.7576 | 36.1 | 0.03 |
| pHILIC | 186.112 | 42.5601 | 36.1 | 0.03 |
| pHILIC | 186.112 | 124.1272 | 36.1 | 0.03 |
| pHILIC | 186.112 | 196.6751 | 36.1 | 0.03 |
| pHILIC | 186.112 | 70.2263 | 36.1 | 0.03 |
| pHILIC | 186.112 | 177.7997 | 36.1 | 0.03 |
| pHILIC | 186.112 | 111.6066 | 36.1 | 0.03 |
| pHILIC | 186.112 | 168.7055 | 36.1 | 0.03 |
| pHILIC | 186.112 | 57.4909 | 36.1 | 0.03 |
| pHILIC | 186.112 | 75.426 | 36.1 | 0.03 |
| pHILIC | 186.112 | 58.398 | 36.1 | 0.03 |
| pHILIC | 186.112 | 113.2003 | 36.1 | 0.03 |
| pHILIC | 186.112 | 192.8756 | 36.1 | 0.03 |
| pHILIC | 186.112 | 143.8964 | 36.1 | 0.03 |
| pHILIC | 186.112 | 52.1834 | 36.1 | 0.03 |
| pHILIC | 186.112 | 72.8245 | 36 | 0.03 |
| pHILIC | 186.112 | 185.4544 | 36 | 0.03 |
| pHILIC | 186.112 | 99.3266 | 36 | 0.03 |

| | | | | |
|---|---|---|---|---|
| pHILIC | 186.112 | 203.6989 | 36 | 0.03 |
| pHILIC | 186.112 | 124.841 | 36 | 0.03 |
| pHILIC | 186.112 | 199.1945 | 36 | 0.03 |
| pHILIC | 186.112 | 71.4597 | 36 | 0.03 |
| pHILIC | 186.112 | 181.4072 | 36 | 0.03 |
| pHILIC | 186.112 | 56.1065 | 36 | 0.03 |
| pHILIC | 186.112 | 66.2981 | 36 | 0.03 |
| pHILIC | 186.112 | 116.5663 | 36 | 0.03 |
| pHILIC | 186.112 | 65.9901 | 36 | 0.03 |
| pHILIC | 186.112 | 188.3376 | 36 | 0.03 |
| pHILIC | 186.112 | 118.0157 | 36 | 0.03 |
| pHILIC | 186.112 | 132.6017 | 36 | 0.03 |
| pHILIC | 186.112 | 92.0469 | 36 | 0.03 |
| pHILIC | 186.112 | 82.1743 | 36 | 0.03 |
| pHILIC | 186.112 | 42.2994 | 35.9 | 0.03 |
| pHILIC | 186.112 | 92.9561 | 35.9 | 0.03 |
| pHILIC | 186.112 | 83.3436 | 35.9 | 0.03 |
| pHILIC | 186.112 | 74.5027 | 35.9 | 0.03 |
| pHILIC | 186.112 | 163.4299 | 35.9 | 0.03 |
| pHILIC | 186.112 | 209.7596 | 35.9 | 0.03 |
| pHILIC | 186.112 | 54.6689 | 35.9 | 0.03 |
| pHILIC | 186.112 | 70.2323 | 35.9 | 0.03 |
| pHILIC | 186.112 | 93.991 | 35.9 | 0.03 |
| pHILIC | 186.112 | 42.1714 | 35.9 | 0.03 |
| pHILIC | 186.112 | 67.311 | 35.9 | 0.03 |
| pHILIC | 186.112 | 72.5519 | 35.8 | 0.03 |
| pHILIC | 186.112 | 93.4011 | 35.8 | 0.03 |
| pHILIC | 186.112 | 77.6713 | 35.8 | 0.03 |
| pHILIC | 186.112 | 113.6599 | 35.8 | 0.03 |
| pHILIC | 186.112 | 167.9159 | 35.8 | 0.03 |
| pHILIC | 186.112 | 85.342 | 35.8 | 0.03 |
| pHILIC | 186.112 | 204.0264 | 35.8 | 0.03 |
| pHILIC | 186.112 | 72.4704 | 35.8 | 0.03 |
| pHILIC | 186.112 | 174.3139 | 35.8 | 0.03 |
| pHILIC | 186.112 | 114.2732 | 35.8 | 0.03 |
| pHILIC | 186.112 | 165.2968 | 35.8 | 0.03 |
| pHILIC | 186.112 | 60.257 | 35.8 | 0.03 |
| pHILIC | 186.112 | 205.2361 | 35.8 | 0.03 |
| pHILIC | 186.112 | 62.5674 | 35.8 | 0.03 |
| pHILIC | 186.112 | 148.567 | 35.8 | 0.03 |
| pHILIC | 186.112 | 140.0048 | 35.7 | 0.03 |
| pHILIC | 186.112 | 110.0577 | 35.7 | 0.03 |

| | | | | |
|---|---|---|---|---|
| pHILIC | 186.112 | 63.0248 | 35.7 | 0.03 |
| pHILIC | 186.112 | 65.1651 | 35.7 | 0.03 |
| pHILIC | 186.112 | 61.5514 | 35.7 | 0.03 |
| pHILIC | 186.112 | 72.8787 | 35.7 | 0.03 |
| pHILIC | 186.112 | 93.8258 | 35.7 | 0.03 |
| pHILIC | 186.112 | 166.4658 | 35.7 | 0.03 |
| pHILIC | 186.112 | 190.7174 | 35.7 | 0.03 |
| pHILIC | 186.112 | 42.4057 | 35.7 | 0.03 |
| pHILIC | 186.112 | 48.8539 | 35.7 | 0.03 |
| pHILIC | 186.112 | 74.1229 | 35.7 | 0.03 |
| pHILIC | 186.112 | 43.0535 | 35.7 | 0.03 |
| pHILIC | 186.112 | 55.3398 | 35.7 | 0.03 |
| pHILIC | 186.112 | 202.8174 | 35.6 | 0.03 |
| pHILIC | 186.112 | 130.692 | 35.6 | 0.03 |
| pHILIC | 186.112 | 160.9401 | 35.6 | 0.03 |
| pHILIC | 186.112 | 45.9274 | 35.6 | 0.03 |
| pHILIC | 186.112 | 43.194 | 35.6 | 0.03 |
| pHILIC | 186.112 | 57.2227 | 35.6 | 0.03 |
| pHILIC | 186.112 | 73.7063 | 35.5 | 0.03 |
| pHILIC | 186.112 | 175.0315 | 35.5 | 0.03 |
| pHILIC | 186.112 | 103.8197 | 35.5 | 0.03 |
| pHILIC | 186.112 | 157.7125 | 35.5 | 0.03 |
| pHILIC | 186.112 | 101.2684 | 35.5 | 0.03 |
| pHILIC | 186.112 | 163.0536 | 35.5 | 0.03 |
| pHILIC | 186.112 | 47.5437 | 35.5 | 0.03 |
| pHILIC | 186.112 | 141.2477 | 35.5 | 0.03 |
| pHILIC | 186.112 | 58.216 | 35.5 | 0.03 |
| pHILIC | 186.112 | 80.1043 | 35.5 | 0.03 |
| pHILIC | 186.112 | 74.9192 | 35.5 | 0.03 |
| pHILIC | 186.112 | 45.5132 | 35.5 | 0.03 |
| pHILIC | 186.112 | 112.6514 | 35.5 | 0.03 |
| pHILIC | 186.112 | 145.777 | 35.5 | 0.03 |
| pHILIC | 186.112 | 145.2666 | 35.5 | 0.03 |
| pHILIC | 186.112 | 53.4646 | 35.5 | 0.03 |
| pHILIC | 186.112 | 46.1179 | 35.5 | 0.03 |
| pHILIC | 186.112 | 135.9415 | 35.5 | 0.03 |
| pHILIC | 186.112 | 111.3298 | 35.5 | 0.03 |
| pHILIC | 186.112 | 117.3718 | 35.4 | 0.03 |
| pHILIC | 186.112 | 90.9004 | 35.4 | 0.03 |
| pHILIC | 186.112 | 98.3714 | 35.4 | 0.03 |
| pHILIC | 186.112 | 135.168 | 35.4 | 0.03 |
| pHILIC | 186.112 | 158.7664 | 35.4 | 0.03 |

| | | | | |
|---|---|---|---|---|
| pHILIC | 186.112 | 52.9458 | 35.4 | 0.03 |
| pHILIC | 186.112 | 53.4115 | 35.4 | 0.03 |
| pHILIC | 186.112 | 141.4855 | 35.4 | 0.03 |
| pHILIC | 186.112 | 57.3756 | 35.4 | 0.03 |
| pHILIC | 186.112 | 126.8064 | 35.3 | 0.03 |
| pHILIC | 186.112 | 64.1511 | 35.3 | 0.03 |
| pHILIC | 186.112 | 57.3255 | 35.3 | 0.03 |
| pHILIC | 186.112 | 50.7857 | 35.3 | 0.03 |
| pHILIC | 186.112 | 106.0999 | 35.3 | 0.03 |
| pHILIC | 186.112 | 113.9081 | 35.3 | 0.03 |
| pHILIC | 186.112 | 162.7681 | 35.3 | 0.03 |
| pHILIC | 186.112 | 57.4363 | 35.3 | 0.03 |
| pHILIC | 186.112 | 54.1357 | 35.3 | 0.03 |
| pHILIC | 186.112 | 75.7276 | 35.3 | 0.03 |
| pHILIC | 186.112 | 64.4512 | 35.3 | 0.03 |
| pHILIC | 186.112 | 151.5711 | 35.3 | 0.03 |
| pHILIC | 186.112 | 45.1167 | 35.3 | 0.03 |
| pHILIC | 186.112 | 77.4109 | 35.3 | 0.03 |
| pHILIC | 186.112 | 48.3851 | 35.2 | 0.03 |
| pHILIC | 186.112 | 72.2287 | 35.2 | 0.03 |
| pHILIC | 186.112 | 131.7352 | 35.2 | 0.03 |
| pHILIC | 186.112 | 144.7641 | 35.2 | 0.03 |
| pHILIC | 186.112 | 122.555 | 35.2 | 0.03 |
| pHILIC | 186.112 | 209.5488 | 35.2 | 0.03 |
| pHILIC | 186.112 | 43.9002 | 35.2 | 0.03 |
| pHILIC | 186.112 | 70.1959 | 35.2 | 0.03 |
| pHILIC | 186.112 | 101.4325 | 35.2 | 0.03 |
| pHILIC | 186.112 | 152.5435 | 35.2 | 0.03 |
| pHILIC | 186.112 | 93.5703 | 35.2 | 0.03 |
| pHILIC | 186.112 | 138.0989 | 35.2 | 0.03 |
| pHILIC | 186.112 | 63.4398 | 35.2 | 0.03 |
| pHILIC | 186.112 | 73.1783 | 35.2 | 0.03 |
| pHILIC | 186.112 | 49.0119 | 35.2 | 0.03 |
| pHILIC | 186.112 | 74.7957 | 35.2 | 0.03 |
| pHILIC | 186.112 | 81.9039 | 35.2 | 0.03 |
| pHILIC | 186.112 | 43.0069 | 35.2 | 0.03 |
| pHILIC | 186.112 | 70.7059 | 35.2 | 0.03 |
| pHILIC | 186.112 | 42.0878 | 35.2 | 0.03 |
| pHILIC | 186.112 | 125.1905 | 35.2 | 0.03 |
| pHILIC | 186.112 | 84.7132 | 35.2 | 0.03 |
| pHILIC | 186.112 | 44.7031 | 35.2 | 0.03 |
| pHILIC | 186.112 | 144.6162 | 35.2 | 0.03 |

| | | | | |
|---|---|---|---|---|
| pHILIC | 186.112 | 61.1924 | 35.1 | 0.03 |
| pHILIC | 186.112 | 52.1073 | 35.1 | 0.03 |
| pHILIC | 186.112 | 75.2363 | 35.1 | 0.03 |
| pHILIC | 186.112 | 113.2727 | 35.1 | 0.03 |
| pHILIC | 186.112 | 48.0195 | 35.1 | 0.03 |
| pHILIC | 186.112 | 113.0953 | 35.1 | 0.03 |
| pHILIC | 186.112 | 161.2467 | 35.1 | 0.03 |
| pHILIC | 186.112 | 102.547 | 35.1 | 0.03 |
| pHILIC | 186.112 | 98.1116 | 35.1 | 0.03 |
| pHILIC | 186.112 | 159.3662 | 35.1 | 0.03 |
| pHILIC | 186.112 | 49.5811 | 35.1 | 0.03 |
| pHILIC | 186.112 | 55.4191 | 35.1 | 0.03 |
| pHILIC | 186.112 | 130.6301 | 35.1 | 0.03 |
| pHILIC | 186.112 | 152.8516 | 35.1 | 0.03 |
| pHILIC | 186.112 | 157.774 | 35.1 | 0.03 |
| pHILIC | 186.112 | 117.5665 | 35.1 | 0.03 |
| pHILIC | 186.112 | 183.2945 | 35.1 | 0.03 |
| pHILIC | 186.112 | 96.2594 | 35.1 | 0.03 |
| pHILIC | 186.112 | 130.2312 | 35 | 0.03 |
| pHILIC | 186.112 | 44.8051 | 35 | 0.03 |
| pHILIC | 186.112 | 42.3997 | 35 | 0.03 |
| pHILIC | 186.112 | 178.2541 | 35 | 0.03 |
| pHILIC | 186.112 | 57.4571 | 35 | 0.03 |
| pHILIC | 186.112 | 49.1336 | 35 | 0.03 |
| pHILIC | 186.112 | 42.7923 | 35 | 0.03 |
| pHILIC | 186.112 | 97.9326 | 35 | 0.03 |
| pHILIC | 186.112 | 44.5951 | 35 | 0.03 |
| pHILIC | 186.112 | 55.867 | 35 | 0.03 |
| pHILIC | 186.112 | 64.6936 | 35 | 0.03 |
| pHILIC | 186.112 | 58.9735 | 35 | 0.03 |
| pHILIC | 186.112 | 42.4561 | 35 | 0.03 |
| pHILIC | 186.112 | 59.3565 | 35 | 0.03 |
| pHILIC | 186.112 | 170.5199 | 35 | 0.03 |
| pHILIC | 186.112 | 105.3282 | 35 | 0.03 |
| pHILIC | 186.112 | 107.6299 | 35 | 0.03 |
| pHILIC | 186.112 | 208.2991 | 35 | 0.03 |
| pHILIC | 186.112 | 81.7873 | 35 | 0.03 |
| pHILIC | 186.112 | 57.2029 | 35 | 0.03 |
| pHILIC | 186.112 | 122.717 | 34.9 | 0.03 |
| pHILIC | 186.112 | 162.1186 | 34.9 | 0.03 |
| pHILIC | 186.112 | 94.5529 | 34.9 | 0.03 |
| pHILIC | 186.112 | 151.9716 | 34.9 | 0.03 |

| | | | | |
|---|---|---|---|---|
| pHILIC | 186.112 | 150.6129 | 34.9 | 0.03 |
| pHILIC | 186.112 | 78.8734 | 34.9 | 0.03 |
| pHILIC | 186.112 | 106.5777 | 34.9 | 0.03 |
| pHILIC | 186.112 | 71.3193 | 34.9 | 0.03 |
| pHILIC | 186.112 | 106.0514 | 34.9 | 0.03 |
| pHILIC | 186.112 | 89.5733 | 34.9 | 0.03 |
| pHILIC | 186.112 | 54.1103 | 34.9 | 0.03 |
| pHILIC | 186.112 | 114.4616 | 34.9 | 0.03 |
| pHILIC | 186.112 | 69.3843 | 34.9 | 0.03 |
| pHILIC | 186.112 | 49.4686 | 34.9 | 0.03 |
| pHILIC | 186.112 | 61.8254 | 34.9 | 0.03 |
| pHILIC | 186.112 | 125.8603 | 34.9 | 0.03 |
| pHILIC | 186.112 | 86.6534 | 34.9 | 0.03 |
| pHILIC | 186.112 | 43.8956 | 34.9 | 0.03 |
| pHILIC | 186.112 | 124.5402 | 34.9 | 0.03 |
| pHILIC | 186.112 | 89.6656 | 34.8 | 0.03 |
| pHILIC | 186.112 | 76.1352 | 34.8 | 0.03 |
| pHILIC | 186.112 | 148.6324 | 34.8 | 0.03 |
| pHILIC | 186.112 | 117.183 | 34.8 | 0.03 |
| pHILIC | 186.112 | 123.9619 | 34.8 | 0.03 |
| pHILIC | 186.112 | 142.8514 | 34.8 | 0.03 |
| pHILIC | 186.112 | 52.2255 | 34.8 | 0.03 |
| pHILIC | 186.112 | 174.1084 | 34.8 | 0.03 |
| pHILIC | 186.112 | 77.4576 | 34.8 | 0.03 |
| pHILIC | 186.112 | 47.6523 | 34.8 | 0.03 |
| pHILIC | 186.112 | 153.0096 | 34.8 | 0.03 |
| pHILIC | 186.112 | 107.0742 | 34.8 | 0.03 |
| pHILIC | 186.112 | 85.1605 | 34.8 | 0.03 |
| pHILIC | 186.112 | 66.9276 | 34.8 | 0.03 |
| pHILIC | 186.112 | 153.1792 | 34.8 | 0.03 |
| pHILIC | 186.112 | 120.7785 | 34.8 | 0.03 |
| pHILIC | 186.112 | 129.7115 | 34.8 | 0.03 |
| pHILIC | 186.112 | 46.8435 | 34.8 | 0.03 |
| pHILIC | 186.112 | 53.9574 | 34.8 | 0.03 |
| pHILIC | 186.112 | 73.9637 | 34.8 | 0.03 |
| pHILIC | 186.112 | 44.8955 | 34.8 | 0.03 |
| pHILIC | 186.112 | 60.7844 | 34.8 | 0.03 |
| pHILIC | 186.112 | 93.8643 | 34.8 | 0.03 |
| pHILIC | 186.112 | 159.5349 | 34.8 | 0.03 |
| pHILIC | 186.112 | 47.8102 | 34.7 | 0.03 |
| pHILIC | 186.112 | 150.8186 | 34.7 | 0.03 |
| pHILIC | 186.112 | 43.0645 | 34.7 | 0.03 |

| | | | | |
|---|---|---|---|---|
| pHILIC | 186.112 | 107.2519 | 34.7 | 0.03 |
| pHILIC | 186.112 | 119.7749 | 34.7 | 0.03 |
| pHILIC | 186.112 | 93.7465 | 34.7 | 0.03 |
| pHILIC | 186.112 | 100.3646 | 34.7 | 0.03 |
| pHILIC | 186.112 | 79.513 | 34.7 | 0.03 |
| pHILIC | 186.112 | 57.6352 | 34.7 | 0.03 |
| pHILIC | 186.112 | 57.6222 | 34.7 | 0.03 |
| pHILIC | 186.112 | 195.7635 | 34.7 | 0.03 |
| pHILIC | 186.112 | 125.2459 | 34.7 | 0.03 |
| pHILIC | 186.112 | 49.7587 | 34.7 | 0.03 |
| pHILIC | 186.112 | 134.9213 | 34.7 | 0.03 |
| pHILIC | 186.112 | 118.5876 | 34.7 | 0.03 |
| pHILIC | 186.112 | 170.3677 | 34.7 | 0.03 |
| pHILIC | 186.112 | 92.2425 | 34.7 | 0.03 |
| pHILIC | 186.112 | 107.3505 | 34.7 | 0.03 |
| pHILIC | 186.112 | 94.0984 | 34.7 | 0.03 |
| pHILIC | 186.112 | 64.7598 | 34.7 | 0.03 |
| pHILIC | 186.112 | 89.3869 | 34.6 | 0.03 |
| pHILIC | 186.112 | 72.7943 | 34.6 | 0.03 |
| pHILIC | 186.112 | 73.1964 | 34.6 | 0.03 |
| pHILIC | 186.112 | 192.7444 | 34.6 | 0.03 |
| pHILIC | 186.112 | 44.7426 | 34.6 | 0.03 |
| pHILIC | 186.112 | 93.6017 | 34.6 | 0.03 |
| pHILIC | 186.112 | 128.456 | 34.6 | 0.03 |
| pHILIC | 186.112 | 58.3841 | 34.6 | 0.03 |
| pHILIC | 186.112 | 47.9202 | 34.6 | 0.03 |
| pHILIC | 186.112 | 58.5042 | 34.6 | 0.03 |
| pHILIC | 186.112 | 42.3116 | 34.6 | 0.03 |
| pHILIC | 186.112 | 57.9017 | 34.6 | 0.03 |
| pHILIC | 186.112 | 100.2911 | 34.6 | 0.03 |
| pHILIC | 186.112 | 129.4029 | 34.6 | 0.03 |
| pHILIC | 186.112 | 45.0999 | 34.6 | 0.03 |
| pHILIC | 186.112 | 97.7107 | 34.6 | 0.03 |
| pHILIC | 186.112 | 57.083 | 34.6 | 0.03 |
| pHILIC | 186.112 | 182.9159 | 34.6 | 0.03 |
| pHILIC | 186.112 | 108.712 | 34.6 | 0.03 |
| pHILIC | 186.112 | 171.4288 | 34.6 | 0.03 |
| pHILIC | 186.112 | 128.67 | 34.6 | 0.03 |
| pHILIC | 186.112 | 94.7465 | 34.5 | 0.03 |
| pHILIC | 186.112 | 55.63 | 34.5 | 0.03 |
| pHILIC | 186.112 | 84.338 | 34.5 | 0.03 |
| pHILIC | 186.112 | 80.6024 | 34.5 | 0.03 |

| | | | | |
|---|---|---|---|---|
| pHILIC | 186.112 | 85.323 | 34.5 | 0.03 |
| pHILIC | 186.112 | 81.1007 | 34.5 | 0.03 |
| pHILIC | 186.112 | 61.2415 | 34.5 | 0.03 |
| pHILIC | 186.112 | 50.685 | 34.5 | 0.03 |
| pHILIC | 186.112 | 132.2251 | 34.5 | 0.03 |
| pHILIC | 186.112 | 115.8542 | 34.5 | 0.03 |
| pHILIC | 186.112 | 114.9227 | 34.5 | 0.03 |
| pHILIC | 186.112 | 156.6903 | 34.5 | 0.03 |
| pHILIC | 186.112 | 43.1358 | 34.5 | 0.03 |
| pHILIC | 186.112 | 174.609 | 34.4 | 0.03 |
| pHILIC | 186.112 | 187.1294 | 34.4 | 0.03 |
| pHILIC | 186.112 | 132.3962 | 34.4 | 0.03 |
| pHILIC | 186.112 | 57.4321 | 34.4 | 0.03 |
| pHILIC | 186.112 | 97.9297 | 34.4 | 0.03 |
| pHILIC | 186.112 | 97.041 | 34.4 | 0.03 |
| pHILIC | 186.112 | 200.3373 | 34.4 | 0.03 |
| pHILIC | 186.112 | 161.6512 | 34.4 | 0.03 |
| pHILIC | 186.112 | 71.1567 | 34.4 | 0.03 |
| pHILIC | 186.112 | 50.3374 | 34.4 | 0.03 |
| pHILIC | 186.112 | 151.187 | 34.4 | 0.03 |
| pHILIC | 186.112 | 77.1872 | 34.4 | 0.03 |
| pHILIC | 186.112 | 134.5287 | 34.4 | 0.03 |
| pHILIC | 186.112 | 45.0116 | 34.4 | 0.03 |
| pHILIC | 186.112 | 50.4369 | 34.4 | 0.03 |
| pHILIC | 186.112 | 120.3973 | 34.4 | 0.03 |
| pHILIC | 186.112 | 54.7181 | 34.4 | 0.03 |
| pHILIC | 186.112 | 167.1777 | 34.4 | 0.03 |
| pHILIC | 186.112 | 59.4037 | 34.4 | 0.03 |
| pHILIC | 186.112 | 131.1279 | 34.4 | 0.03 |
| pHILIC | 186.112 | 54.5475 | 34.4 | 0.03 |
| pHILIC | 186.112 | 112.7738 | 34.4 | 0.03 |
| pHILIC | 186.112 | 49.8102 | 34.4 | 0.03 |
| pHILIC | 186.112 | 44.9943 | 34.4 | 0.03 |
| pHILIC | 186.112 | 76.7214 | 34.4 | 0.03 |
| pHILIC | 186.112 | 194.8275 | 34.4 | 0.03 |
| pHILIC | 186.112 | 152.9673 | 34.3 | 0.03 |
| pHILIC | 186.112 | 166.4373 | 34.3 | 0.03 |
| pHILIC | 186.112 | 202.4547 | 34.3 | 0.03 |
| pHILIC | 186.112 | 161.4975 | 34.3 | 0.03 |
| pHILIC | 186.112 | 99.2773 | 34.3 | 0.03 |
| pHILIC | 186.112 | 54.5347 | 34.3 | 0.03 |
| pHILIC | 186.112 | 97.2512 | 34.3 | 0.03 |

| | | | | |
|---|---|---|---|---|
| pHILIC | 186.112 | 112.6005 | 34.3 | 0.03 |
| pHILIC | 186.112 | 102.412 | 34.3 | 0.03 |
| pHILIC | 186.112 | 57.2626 | 34.3 | 0.03 |
| pHILIC | 186.112 | 168.3618 | 34.3 | 0.03 |
| pHILIC | 186.112 | 137.1647 | 34.3 | 0.03 |
| pHILIC | 186.112 | 43.0849 | 34.3 | 0.03 |
| pHILIC | 186.112 | 46.0502 | 34.3 | 0.03 |
| pHILIC | 186.112 | 48.6069 | 34.3 | 0.03 |
| pHILIC | 186.112 | 186.4594 | 34.3 | 0.03 |
| pHILIC | 186.112 | 149.975 | 34.3 | 0.03 |
| pHILIC | 186.112 | 167.9895 | 34.3 | 0.03 |
| pHILIC | 186.112 | 93.7422 | 34.3 | 0.03 |
| pHILIC | 186.112 | 68.8848 | 34.3 | 0.03 |
| pHILIC | 186.112 | 82.1321 | 34.3 | 0.03 |
| pHILIC | 186.112 | 113.7155 | 34.3 | 0.03 |
| pHILIC | 186.112 | 145.9274 | 34.3 | 0.03 |
| pHILIC | 186.112 | 99.4224 | 34.3 | 0.03 |
| pHILIC | 186.112 | 80.8047 | 34.3 | 0.03 |
| pHILIC | 186.112 | 65.5718 | 34.3 | 0.03 |
| pHILIC | 186.112 | 70.6938 | 34.2 | 0.03 |
| pHILIC | 186.112 | 46.8272 | 34.2 | 0.03 |
| pHILIC | 186.112 | 55.7543 | 34.2 | 0.03 |
| pHILIC | 186.112 | 149.5119 | 34.2 | 0.03 |
| pHILIC | 186.112 | 56.7761 | 34.2 | 0.03 |
| pHILIC | 186.112 | 120.7918 | 34.2 | 0.03 |
| pHILIC | 186.112 | 149.1984 | 34.2 | 0.03 |
| pHILIC | 186.112 | 45.9327 | 34.2 | 0.03 |
| pHILIC | 186.112 | 181.5611 | 34.2 | 0.03 |
| pHILIC | 186.112 | 108.8866 | 34.2 | 0.03 |
| pHILIC | 186.112 | 104.0157 | 34.2 | 0.03 |
| pHILIC | 186.112 | 180.9037 | 34.2 | 0.03 |
| pHILIC | 186.112 | 58.1615 | 34.2 | 0.03 |
| pHILIC | 186.112 | 44.9342 | 34.2 | 0.03 |
| pHILIC | 186.112 | 49.2238 | 34.2 | 0.03 |
| pHILIC | 186.112 | 59.1018 | 34.2 | 0.03 |
| pHILIC | 186.112 | 50.7306 | 34.2 | 0.03 |
| pHILIC | 186.112 | 176.776 | 34.2 | 0.03 |
| pHILIC | 186.112 | 148.1448 | 34.2 | 0.03 |
| pHILIC | 186.112 | 119.8318 | 34.2 | 0.03 |
| pHILIC | 186.112 | 42.7813 | 34.2 | 0.03 |
| pHILIC | 186.112 | 81.383 | 34.1 | 0.03 |
| pHILIC | 186.112 | 91.7518 | 34.1 | 0.03 |

| | | | | |
|---|---|---|---|---|
| pHILIC | 186.112 | 79.25 | 34.1 | 0.03 |
| pHILIC | 186.112 | 47.7162 | 34.1 | 0.03 |
| pHILIC | 186.112 | 190.9576 | 34.1 | 0.03 |
| pHILIC | 186.112 | 84.7467 | 34.1 | 0.03 |
| pHILIC | 186.112 | 75.4753 | 34.1 | 0.03 |
| pHILIC | 186.112 | 70.6359 | 34.1 | 0.03 |
| pHILIC | 186.112 | 93.6061 | 34.1 | 0.03 |
| pHILIC | 186.112 | 154.4132 | 34.1 | 0.03 |
| pHILIC | 186.112 | 75.6928 | 34.1 | 0.03 |
| pHILIC | 186.112 | 109.3499 | 34.1 | 0.03 |
| pHILIC | 186.112 | 55.7518 | 34.1 | 0.03 |
| pHILIC | 186.112 | 80.5073 | 34.1 | 0.03 |
| pHILIC | 186.112 | 68.3589 | 34.1 | 0.03 |
| pHILIC | 186.112 | 83.6136 | 34.1 | 0.03 |
| pHILIC | 186.112 | 146.3447 | 34.1 | 0.03 |
| pHILIC | 186.112 | 59.6021 | 34.1 | 0.03 |
| pHILIC | 186.112 | 172.7079 | 34 | 0.03 |
| pHILIC | 186.112 | 84.7337 | 34 | 0.03 |
| pHILIC | 186.112 | 143.1428 | 34 | 0.03 |
| pHILIC | 186.112 | 202.7905 | 34 | 0.03 |
| pHILIC | 186.112 | 146.5123 | 34 | 0.03 |
| pHILIC | 186.112 | 172.2557 | 34 | 0.03 |
| pHILIC | 186.112 | 105.9591 | 34 | 0.03 |
| pHILIC | 186.112 | 106.1313 | 34 | 0.03 |
| pHILIC | 186.112 | 144.7287 | 34 | 0.03 |
| pHILIC | 186.112 | 157.5887 | 34 | 0.03 |
| pHILIC | 186.112 | 92.4266 | 34 | 0.03 |
| pHILIC | 186.112 | 73.2154 | 34 | 0.03 |
| pHILIC | 186.112 | 77.683 | 34 | 0.03 |
| pHILIC | 186.112 | 68.4537 | 34 | 0.03 |
| pHILIC | 186.112 | 137.1492 | 34 | 0.03 |
| pHILIC | 186.112 | 51.6272 | 34 | 0.03 |
| pHILIC | 186.112 | 67.8222 | 34 | 0.03 |
| pHILIC | 186.112 | 74.9396 | 34 | 0.03 |
| pHILIC | 186.112 | 131.8931 | 34 | 0.03 |
| pHILIC | 186.112 | 92.3383 | 34 | 0.03 |
| pHILIC | 186.112 | 193.3472 | 34 | 0.03 |
| pHILIC | 186.112 | 86.5737 | 34 | 0.03 |
| pHILIC | 186.112 | 52.9285 | 34 | 0.03 |
| pHILIC | 186.112 | 68.95 | 34 | 0.03 |
| pHILIC | 186.112 | 53.2329 | 34 | 0.03 |
| pHILIC | 186.112 | 52.6307 | 34 | 0.03 |

| | | | | |
|---|---|---|---|---|
| pHILIC | 186.112 | 86.0424 | 33.9 | 0.03 |
| pHILIC | 186.112 | 165.6753 | 33.9 | 0.03 |
| pHILIC | 186.112 | 97.5836 | 33.9 | 0.03 |
| pHILIC | 186.112 | 138.1517 | 33.9 | 0.03 |
| pHILIC | 186.112 | 56.1385 | 33.9 | 0.03 |
| pHILIC | 186.112 | 95.4672 | 33.9 | 0.03 |
| pHILIC | 186.112 | 144.9272 | 33.9 | 0.03 |
| pHILIC | 186.112 | 86.0811 | 33.9 | 0.03 |
| pHILIC | 186.112 | 146.0201 | 33.9 | 0.03 |
| pHILIC | 186.112 | 68.3932 | 33.9 | 0.03 |
| pHILIC | 186.112 | 205.5424 | 33.9 | 0.03 |
| pHILIC | 186.112 | 51.5254 | 33.9 | 0.03 |
| pHILIC | 186.112 | 171.3886 | 33.9 | 0.03 |
| pHILIC | 186.112 | 68.5599 | 33.9 | 0.03 |
| pHILIC | 186.112 | 120.0536 | 33.9 | 0.03 |
| pHILIC | 186.112 | 143.9899 | 33.9 | 0.03 |
| pHILIC | 186.112 | 59.9588 | 33.9 | 0.03 |
| pHILIC | 186.112 | 139.0061 | 33.9 | 0.03 |
| pHILIC | 186.112 | 105.5858 | 33.9 | 0.03 |
| pHILIC | 186.112 | 44.6765 | 33.9 | 0.03 |
| pHILIC | 186.112 | 51.842 | 33.9 | 0.03 |
| pHILIC | 186.112 | 137.0202 | 33.9 | 0.03 |
| pHILIC | 186.112 | 114.5186 | 33.9 | 0.03 |
| pHILIC | 186.112 | 45.8222 | 33.8 | 0.03 |
| pHILIC | 186.112 | 45.0499 | 33.8 | 0.03 |
| pHILIC | 186.112 | 117.8526 | 33.8 | 0.03 |
| pHILIC | 186.112 | 55.4444 | 33.8 | 0.03 |
| pHILIC | 186.112 | 207.4626 | 33.8 | 0.03 |
| pHILIC | 186.112 | 88.5347 | 33.8 | 0.03 |
| pHILIC | 186.112 | 99.2289 | 33.8 | 0.03 |
| pHILIC | 186.112 | 69.1365 | 33.8 | 0.03 |
| pHILIC | 186.112 | 67.0893 | 33.8 | 0.03 |
| pHILIC | 186.112 | 53.7099 | 33.8 | 0.03 |
| pHILIC | 186.112 | 59.7259 | 33.8 | 0.03 |
| pHILIC | 186.112 | 48.202 | 33.8 | 0.03 |
| pHILIC | 186.112 | 53.9978 | 33.8 | 0.03 |
| pHILIC | 186.112 | 71.0677 | 33.8 | 0.03 |
| pHILIC | 186.112 | 99.6333 | 33.8 | 0.03 |
| pHILIC | 186.112 | 149.8545 | 33.8 | 0.03 |
| pHILIC | 186.112 | 135.569 | 33.8 | 0.03 |
| pHILIC | 186.112 | 76.8636 | 33.8 | 0.03 |
| pHILIC | 186.112 | 69.1705 | 33.8 | 0.03 |

| | | | | |
|---|---|---|---|---|
| pHILIC | 186.112 | 178.046 | 33.8 | 0.03 |
| pHILIC | 186.112 | 49.8931 | 33.8 | 0.03 |
| pHILIC | 186.112 | 149.6736 | 33.8 | 0.03 |
| pHILIC | 186.112 | 67.1831 | 33.8 | 0.03 |
| pHILIC | 186.112 | 61.0622 | 33.8 | 0.03 |
| pHILIC | 186.112 | 61.2891 | 33.8 | 0.03 |
| pHILIC | 186.112 | 62.6256 | 33.8 | 0.03 |
| pHILIC | 186.112 | 162.244 | 33.8 | 0.03 |
| pHILIC | 186.112 | 117.0266 | 33.7 | 0.03 |
| pHILIC | 186.112 | 153.1536 | 33.7 | 0.03 |
| pHILIC | 186.112 | 79.0057 | 33.7 | 0.03 |
| pHILIC | 186.112 | 50.6681 | 33.7 | 0.03 |
| pHILIC | 186.112 | 157.6155 | 33.7 | 0.03 |
| pHILIC | 186.112 | 99.0448 | 33.7 | 0.03 |
| pHILIC | 186.112 | 111.5586 | 33.7 | 0.03 |
| pHILIC | 186.112 | 65.982 | 33.7 | 0.03 |
| pHILIC | 186.112 | 47.0596 | 33.7 | 0.03 |
| pHILIC | 186.112 | 68.1944 | 33.7 | 0.03 |
| pHILIC | 186.112 | 97.732 | 33.7 | 0.03 |
| pHILIC | 186.112 | 73.6791 | 33.7 | 0.03 |
| pHILIC | 186.112 | 94.7805 | 33.7 | 0.03 |
| pHILIC | 186.112 | 59.5353 | 33.7 | 0.03 |
| pHILIC | 186.112 | 138.3974 | 33.7 | 0.03 |
| pHILIC | 186.112 | 80.2184 | 33.6 | 0.03 |
| pHILIC | 186.112 | 44.2018 | 33.6 | 0.03 |
| pHILIC | 186.112 | 47.7919 | 33.6 | 0.03 |
| pHILIC | 186.112 | 54.1572 | 33.6 | 0.03 |
| pHILIC | 186.112 | 122.034 | 33.6 | 0.03 |
| pHILIC | 186.112 | 52.1964 | 33.6 | 0.03 |
| pHILIC | 186.112 | 93.7726 | 33.6 | 0.03 |
| pHILIC | 186.112 | 44.9926 | 33.6 | 0.03 |
| pHILIC | 186.112 | 93.3815 | 33.6 | 0.03 |
| pHILIC | 186.112 | 51.8151 | 33.6 | 0.03 |
| pHILIC | 186.112 | 115.971 | 33.6 | 0.03 |
| pHILIC | 186.112 | 46.996 | 33.6 | 0.03 |
| pHILIC | 186.112 | 51.6748 | 33.6 | 0.03 |
| pHILIC | 186.112 | 166.0053 | 33.6 | 0.03 |
| pHILIC | 186.112 | 69.6277 | 33.6 | 0.03 |
| pHILIC | 186.112 | 44.0122 | 33.6 | 0.03 |
| pHILIC | 186.112 | 84.8463 | 33.6 | 0.03 |
| pHILIC | 186.112 | 47.477 | 33.6 | 0.03 |
| pHILIC | 186.112 | 109.6111 | 33.5 | 0.03 |

| | | | | |
|---|---|---|---|---|
| pHILIC | 186.112 | 117.5617 | 33.5 | 0.03 |
| pHILIC | 186.112 | 111.6124 | 33.5 | 0.03 |
| pHILIC | 186.112 | 114.3532 | 33.5 | 0.03 |
| pHILIC | 186.112 | 86.5159 | 33.5 | 0.03 |
| pHILIC | 186.112 | 132.1989 | 33.5 | 0.03 |
| pHILIC | 186.112 | 129.9391 | 33.5 | 0.03 |
| pHILIC | 186.112 | 42.2235 | 33.5 | 0.03 |
| pHILIC | 186.112 | 201.5755 | 33.5 | 0.03 |
| pHILIC | 186.112 | 85.5258 | 33.5 | 0.03 |
| pHILIC | 186.112 | 49.52 | 33.5 | 0.03 |
| pHILIC | 186.112 | 78.765 | 33.5 | 0.03 |
| pHILIC | 186.112 | 146.9722 | 33.5 | 0.03 |
| pHILIC | 186.112 | 109.9966 | 33.5 | 0.03 |
| pHILIC | 186.112 | 167.8403 | 33.5 | 0.03 |
| pHILIC | 186.112 | 69.3482 | 33.5 | 0.03 |
| pHILIC | 186.112 | 52.0358 | 33.5 | 0.03 |
| pHILIC | 186.112 | 162.7769 | 33.5 | 0.03 |
| pHILIC | 186.112 | 179.7679 | 33.5 | 0.03 |
| pHILIC | 186.112 | 102.5032 | 33.5 | 0.03 |
| pHILIC | 186.112 | 120.1423 | 33.4 | 0.03 |
| pHILIC | 186.112 | 42.7839 | 33.4 | 0.03 |
| pHILIC | 186.112 | 97.7877 | 33.4 | 0.03 |
| pHILIC | 186.112 | 60.4423 | 33.4 | 0.03 |
| pHILIC | 186.112 | 93.4739 | 33.4 | 0.03 |
| pHILIC | 186.112 | 50.354 | 33.4 | 0.03 |
| pHILIC | 186.112 | 196.8472 | 33.4 | 0.03 |
| pHILIC | 186.112 | 67.7633 | 33.4 | 0.03 |
| pHILIC | 186.112 | 123.4829 | 33.4 | 0.03 |
| pHILIC | 186.112 | 46.9357 | 33.4 | 0.03 |
| pHILIC | 186.112 | 128.8295 | 33.4 | 0.03 |
| pHILIC | 186.112 | 187.5401 | 33.4 | 0.03 |
| pHILIC | 186.112 | 58.6342 | 33.4 | 0.03 |
| pHILIC | 186.112 | 131.2428 | 33.4 | 0.03 |
| pHILIC | 186.112 | 44.4137 | 33.4 | 0.03 |
| pHILIC | 186.112 | 63.9086 | 33.4 | 0.03 |
| pHILIC | 186.112 | 51.859 | 33.4 | 0.03 |
| pHILIC | 186.112 | 124.3037 | 33.4 | 0.03 |
| pHILIC | 186.112 | 49.6071 | 33.4 | 0.03 |
| pHILIC | 186.112 | 48.8734 | 33.4 | 0.03 |
| pHILIC | 186.112 | 161.5748 | 33.4 | 0.03 |
| pHILIC | 186.112 | 65.7854 | 33.4 | 0.03 |
| pHILIC | 186.112 | 147.1537 | 33.4 | 0.03 |

| | | | | |
|---|---|---|---|---|
| pHILIC | 186.112 | 119.4228 | 33.4 | 0.03 |
| pHILIC | 186.112 | 70.4845 | 33.4 | 0.03 |
| pHILIC | 186.112 | 193.8995 | 33.4 | 0.03 |
| pHILIC | 186.112 | 102.7231 | 33.4 | 0.03 |
| pHILIC | 186.112 | 111.7355 | 33.3 | 0.03 |
| pHILIC | 186.112 | 79.3017 | 33.3 | 0.03 |
| pHILIC | 186.112 | 44.218 | 33.3 | 0.03 |
| pHILIC | 186.112 | 135.1057 | 33.3 | 0.03 |
| pHILIC | 186.112 | 107.6901 | 33.3 | 0.03 |
| pHILIC | 186.112 | 200.8149 | 33.3 | 0.03 |
| pHILIC | 186.112 | 78.3165 | 33.3 | 0.03 |
| pHILIC | 186.112 | 160.6973 | 33.3 | 0.03 |
| pHILIC | 186.112 | 125.786 | 33.3 | 0.03 |
| pHILIC | 186.112 | 45.4691 | 33.3 | 0.03 |
| pHILIC | 186.112 | 87.0166 | 33.3 | 0.03 |
| pHILIC | 186.112 | 69.6849 | 33.3 | 0.03 |
| pHILIC | 186.112 | 95.872 | 33.3 | 0.03 |
| pHILIC | 186.112 | 47.4762 | 33.3 | 0.03 |
| pHILIC | 186.112 | 51.8433 | 33.3 | 0.03 |
| pHILIC | 186.112 | 42.6467 | 33.3 | 0.03 |
| pHILIC | 186.112 | 43.6533 | 33.2 | 0.03 |
| pHILIC | 186.112 | 51.9359 | 33.2 | 0.03 |
| pHILIC | 186.112 | 144.1924 | 33.2 | 0.03 |
| pHILIC | 186.112 | 93.1402 | 33.2 | 0.03 |
| pHILIC | 186.112 | 82.1876 | 33.2 | 0.03 |
| pHILIC | 186.112 | 123.3806 | 33.2 | 0.03 |
| pHILIC | 186.112 | 88.7059 | 33.2 | 0.03 |
| pHILIC | 186.112 | 79.7812 | 33.2 | 0.03 |
| pHILIC | 186.112 | 47.6396 | 33.2 | 0.03 |
| pHILIC | 186.112 | 48.7451 | 33.2 | 0.03 |
| pHILIC | 186.112 | 127.5137 | 33.2 | 0.03 |
| pHILIC | 186.112 | 94.1025 | 33.2 | 0.03 |
| pHILIC | 186.112 | 68.0346 | 33.2 | 0.03 |
| pHILIC | 186.112 | 60.1219 | 33.2 | 0.03 |
| pHILIC | 186.112 | 158.6372 | 33.2 | 0.03 |
| pHILIC | 186.112 | 65.9993 | 33.2 | 0.03 |
| pHILIC | 186.112 | 50.6746 | 33.2 | 0.03 |
| pHILIC | 186.112 | 93.9171 | 33.2 | 0.03 |
| pHILIC | 186.112 | 51.2953 | 33.2 | 0.03 |
| pHILIC | 186.112 | 54.8178 | 33.2 | 0.03 |
| pHILIC | 186.112 | 64.0217 | 33.2 | 0.03 |
| pHILIC | 186.112 | 154.0042 | 33.2 | 0.03 |

| | | | | |
|---|---|---|---|---|
| pHILIC | 186.112 | 63.0238 | 33.2 | 0.03 |
| pHILIC | 186.112 | 75.1855 | 33.2 | 0.03 |
| pHILIC | 186.112 | 124.5523 | 33.2 | 0.03 |
| pHILIC | 186.112 | 134.237 | 33.1 | 0.03 |
| pHILIC | 186.112 | 93.8705 | 33.1 | 0.03 |
| pHILIC | 186.112 | 96.5595 | 33.1 | 0.03 |
| pHILIC | 186.112 | 118.4436 | 33.1 | 0.03 |
| pHILIC | 186.112 | 69.2989 | 33.1 | 0.03 |
| pHILIC | 186.112 | 145.8895 | 33.1 | 0.03 |
| pHILIC | 186.112 | 102.8494 | 33.1 | 0.03 |
| pHILIC | 186.112 | 60.5992 | 33.1 | 0.03 |
| pHILIC | 186.112 | 52.4674 | 33.1 | 0.03 |
| pHILIC | 186.112 | 109.1293 | 33.1 | 0.03 |
| pHILIC | 186.112 | 48.1739 | 33.1 | 0.03 |
| pHILIC | 186.112 | 126.1775 | 33.1 | 0.03 |
| pHILIC | 186.112 | 45.8923 | 33.1 | 0.03 |
| pHILIC | 186.112 | 99.4584 | 33.1 | 0.03 |
| pHILIC | 186.112 | 131.6657 | 33.1 | 0.03 |
| pHILIC | 186.112 | 193.3376 | 33.1 | 0.03 |
| pHILIC | 186.112 | 43.7756 | 33.1 | 0.03 |
| pHILIC | 186.112 | 146.3219 | 33.1 | 0.03 |
| pHILIC | 186.112 | 123.9781 | 33.1 | 0.03 |
| pHILIC | 186.112 | 54.1803 | 33.1 | 0.03 |
| pHILIC | 186.112 | 48.2247 | 33 | 0.03 |
| pHILIC | 186.112 | 61.5551 | 33 | 0.03 |
| pHILIC | 186.112 | 45.4474 | 33 | 0.03 |
| pHILIC | 186.112 | 79.3738 | 33 | 0.03 |
| pHILIC | 186.112 | 124.1123 | 33 | 0.03 |
| pHILIC | 186.112 | 67.2247 | 33 | 0.03 |
| pHILIC | 186.112 | 184.6028 | 33 | 0.03 |
| pHILIC | 186.112 | 104.2392 | 33 | 0.03 |
| pHILIC | 186.112 | 187.1052 | 33 | 0.03 |
| pHILIC | 186.112 | 59.6954 | 33 | 0.03 |
| pHILIC | 186.112 | 142.3246 | 33 | 0.03 |
| pHILIC | 186.112 | 43.86 | 33 | 0.03 |
| pHILIC | 186.112 | 142.4666 | 33 | 0.03 |
| pHILIC | 186.112 | 96.8642 | 33 | 0.03 |
| pHILIC | 186.112 | 186.8686 | 33 | 0.03 |
| pHILIC | 186.112 | 54.1351 | 33 | 0.03 |
| pHILIC | 186.112 | 44.8552 | 33 | 0.03 |
| pHILIC | 186.112 | 52.1326 | 33 | 0.03 |
| pHILIC | 186.112 | 87.3223 | 33 | 0.03 |

| | | | | |
|---|---|---|---|---|
| pHILIC | 186.112 | 42.5308 | 33 | 0.03 |
| pHILIC | 186.112 | 124.8697 | 33 | 0.03 |
| pHILIC | 186.112 | 44.102 | 32.9 | 0.03 |
| pHILIC | 186.112 | 105.8104 | 32.9 | 0.03 |
| pHILIC | 186.112 | 134.0577 | 32.9 | 0.03 |
| pHILIC | 186.112 | 151.3027 | 32.9 | 0.03 |
| pHILIC | 186.112 | 135.164 | 32.9 | 0.03 |
| pHILIC | 186.112 | 96.457 | 32.9 | 0.03 |
| pHILIC | 186.112 | 102.6845 | 32.9 | 0.03 |
| pHILIC | 186.112 | 172.1972 | 32.9 | 0.03 |
| pHILIC | 186.112 | 45.0531 | 32.9 | 0.03 |
| pHILIC | 186.112 | 47.5391 | 32.9 | 0.03 |
| pHILIC | 186.112 | 48.5842 | 32.9 | 0.03 |
| pHILIC | 186.112 | 101.7981 | 32.9 | 0.03 |
| pHILIC | 186.112 | 80.5929 | 32.9 | 0.03 |
| pHILIC | 186.112 | 55.6104 | 32.9 | 0.03 |
| pHILIC | 186.112 | 111.1143 | 32.9 | 0.03 |
| pHILIC | 186.112 | 49.1257 | 32.8 | 0.03 |
| pHILIC | 186.112 | 61.3659 | 32.8 | 0.03 |
| pHILIC | 186.112 | 96.95 | 32.8 | 0.03 |
| pHILIC | 186.112 | 82.596 | 32.8 | 0.03 |
| pHILIC | 186.112 | 73.7573 | 32.8 | 0.03 |
| pHILIC | 186.112 | 63.2833 | 32.8 | 0.03 |
| pHILIC | 186.112 | 51.6695 | 32.8 | 0.03 |
| pHILIC | 186.112 | 68.9211 | 32.8 | 0.03 |
| pHILIC | 186.112 | 78.729 | 32.8 | 0.03 |
| pHILIC | 186.112 | 68.0913 | 32.8 | 0.03 |
| pHILIC | 186.112 | 187.0349 | 32.8 | 0.03 |
| pHILIC | 186.112 | 153.095 | 32.8 | 0.03 |
| pHILIC | 186.112 | 47.1687 | 32.8 | 0.03 |
| pHILIC | 186.112 | 152.5046 | 32.8 | 0.03 |
| pHILIC | 186.112 | 110.831 | 32.8 | 0.03 |
| pHILIC | 186.112 | 85.1704 | 32.8 | 0.03 |
| pHILIC | 186.112 | 56.6994 | 32.8 | 0.03 |
| pHILIC | 186.112 | 56.8831 | 32.8 | 0.03 |
| pHILIC | 186.112 | 65.0215 | 32.8 | 0.03 |
| pHILIC | 186.112 | 76.5964 | 32.8 | 0.03 |
| pHILIC | 186.112 | 68.3515 | 32.8 | 0.03 |
| pHILIC | 186.112 | 46.9308 | 32.8 | 0.03 |
| pHILIC | 186.112 | 112.1967 | 32.7 | 0.03 |
| pHILIC | 186.112 | 96.5891 | 32.7 | 0.03 |
| pHILIC | 186.112 | 50.016 | 32.7 | 0.03 |

| | | | | |
|---|---|---|---|---|
| pHILIC | 186.112 | 176.7644 | 32.7 | 0.03 |
| pHILIC | 186.112 | 46.8183 | 32.7 | 0.03 |
| pHILIC | 186.112 | 55.562 | 32.7 | 0.03 |
| pHILIC | 186.112 | 68.1857 | 32.7 | 0.03 |
| pHILIC | 186.112 | 42.4214 | 32.7 | 0.03 |
| pHILIC | 186.112 | 49.9836 | 32.7 | 0.03 |
| pHILIC | 186.112 | 147.6755 | 32.7 | 0.03 |
| pHILIC | 186.112 | 70.4781 | 32.7 | 0.03 |
| pHILIC | 186.112 | 147.3602 | 32.7 | 0.03 |
| pHILIC | 186.112 | 53.1482 | 32.7 | 0.03 |
| pHILIC | 186.112 | 46.9686 | 32.7 | 0.03 |
| pHILIC | 186.112 | 157.4874 | 32.7 | 0.03 |
| pHILIC | 186.112 | 89.8401 | 32.7 | 0.03 |
| pHILIC | 186.112 | 90.0407 | 32.7 | 0.03 |
| pHILIC | 186.112 | 84.9258 | 32.7 | 0.03 |
| pHILIC | 186.112 | 107.2265 | 32.7 | 0.03 |
| pHILIC | 186.112 | 85.6864 | 32.7 | 0.03 |
| pHILIC | 186.112 | 64.5309 | 32.7 | 0.03 |
| pHILIC | 186.112 | 75.3146 | 32.7 | 0.03 |
| pHILIC | 186.112 | 116.3713 | 32.7 | 0.03 |
| pHILIC | 186.112 | 55.2318 | 32.7 | 0.03 |
| pHILIC | 186.112 | 101.5003 | 32.7 | 0.03 |
| pHILIC | 186.112 | 101.2872 | 32.7 | 0.03 |
| pHILIC | 186.112 | 94.6021 | 32.7 | 0.03 |
| pHILIC | 186.112 | 58.7681 | 32.6 | 0.03 |
| pHILIC | 186.112 | 52.9909 | 32.6 | 0.03 |
| pHILIC | 186.112 | 49.3784 | 32.6 | 0.03 |
| pHILIC | 186.112 | 47.0901 | 32.6 | 0.03 |
| pHILIC | 186.112 | 79.3159 | 32.6 | 0.03 |
| pHILIC | 186.112 | 101.3241 | 32.6 | 0.03 |
| pHILIC | 186.112 | 50.6573 | 32.6 | 0.03 |
| pHILIC | 186.112 | 144.0934 | 32.6 | 0.03 |
| pHILIC | 186.112 | 46.8967 | 32.6 | 0.03 |
| pHILIC | 186.112 | 78.1643 | 32.6 | 0.03 |
| pHILIC | 186.112 | 111.1019 | 32.6 | 0.03 |
| pHILIC | 186.112 | 66.6771 | 32.6 | 0.03 |
| pHILIC | 186.112 | 79.3039 | 32.6 | 0.03 |
| pHILIC | 186.112 | 93.9844 | 32.6 | 0.03 |
| pHILIC | 186.112 | 65.3493 | 32.6 | 0.03 |
| pHILIC | 186.112 | 124.3851 | 32.6 | 0.03 |
| pHILIC | 186.112 | 73.4782 | 32.6 | 0.03 |
| pHILIC | 186.112 | 51.8743 | 32.6 | 0.03 |

| | | | | |
|---|---|---|---|---|
| pHILIC | 186.112 | 109.3818 | 32.6 | 0.03 |
| pHILIC | 186.112 | 115.7004 | 32.6 | 0.03 |
| pHILIC | 186.112 | 78.093 | 32.6 | 0.03 |
| pHILIC | 186.112 | 82.4412 | 32.6 | 0.03 |
| pHILIC | 186.112 | 183.9476 | 32.6 | 0.03 |
| pHILIC | 186.112 | 126.3191 | 32.6 | 0.03 |
| pHILIC | 186.112 | 152.2971 | 32.6 | 0.03 |
| pHILIC | 186.112 | 52.1413 | 32.6 | 0.03 |
| pHILIC | 186.112 | 102.9806 | 32.5 | 0.03 |
| pHILIC | 186.112 | 101.6442 | 32.5 | 0.03 |
| pHILIC | 186.112 | 156.4032 | 32.5 | 0.03 |
| pHILIC | 186.112 | 154.2584 | 32.5 | 0.03 |
| pHILIC | 186.112 | 59.9884 | 32.5 | 0.03 |
| pHILIC | 186.112 | 164.2019 | 32.5 | 0.03 |
| pHILIC | 186.112 | 93.6363 | 32.5 | 0.03 |
| pHILIC | 186.112 | 173.9952 | 32.5 | 0.03 |
| pHILIC | 186.112 | 208.2867 | 32.5 | 0.03 |
| pHILIC | 186.112 | 101.8978 | 32.5 | 0.03 |
| pHILIC | 186.112 | 167.4033 | 32.5 | 0.03 |
| pHILIC | 186.112 | 43.5434 | 32.5 | 0.03 |
| pHILIC | 186.112 | 124.8965 | 32.5 | 0.03 |
| pHILIC | 186.112 | 42.8284 | 32.5 | 0.03 |
| pHILIC | 186.112 | 44.8065 | 32.5 | 0.03 |
| pHILIC | 186.112 | 93.5223 | 32.5 | 0.03 |
| pHILIC | 186.112 | 68.1559 | 32.5 | 0.03 |
| pHILIC | 186.112 | 52.8858 | 32.4 | 0.03 |
| pHILIC | 186.112 | 69.3316 | 32.4 | 0.03 |
| pHILIC | 186.112 | 196.7243 | 32.4 | 0.03 |
| pHILIC | 186.112 | 77.9674 | 32.4 | 0.03 |
| pHILIC | 186.112 | 119.2882 | 32.4 | 0.03 |
| pHILIC | 186.112 | 69.7689 | 32.4 | 0.03 |
| pHILIC | 186.112 | 46.5197 | 32.4 | 0.03 |
| pHILIC | 186.112 | 135.3133 | 32.4 | 0.03 |
| pHILIC | 186.112 | 78.2333 | 32.4 | 0.03 |
| pHILIC | 186.112 | 123.5734 | 32.4 | 0.03 |
| pHILIC | 186.112 | 49.4297 | 32.4 | 0.03 |
| pHILIC | 186.112 | 192.2032 | 32.4 | 0.03 |
| pHILIC | 186.112 | 46.5111 | 32.4 | 0.03 |
| pHILIC | 186.112 | 130.4571 | 32.4 | 0.03 |
| pHILIC | 186.112 | 109.2609 | 32.4 | 0.03 |
| pHILIC | 186.112 | 49.858 | 32.4 | 0.03 |
| pHILIC | 186.112 | 50.1129 | 32.4 | 0.03 |

| | | | | |
|---|---|---|---|---|
| pHILIC | 186.112 | 145.177 | 32.4 | 0.03 |
| pHILIC | 186.112 | 47.775 | 32.4 | 0.03 |
| pHILIC | 186.112 | 187.7741 | 32.4 | 0.03 |
| pHILIC | 186.112 | 117.4703 | 32.4 | 0.03 |
| pHILIC | 186.112 | 159.5281 | 32.3 | 0.03 |
| pHILIC | 186.112 | 50.2542 | 32.3 | 0.03 |
| pHILIC | 186.112 | 102.7164 | 32.3 | 0.03 |
| pHILIC | 186.112 | 87.5039 | 32.3 | 0.03 |
| pHILIC | 186.112 | 55.289 | 32.3 | 0.03 |
| pHILIC | 186.112 | 93.39 | 32.3 | 0.03 |
| pHILIC | 186.112 | 136.7896 | 32.3 | 0.03 |
| pHILIC | 186.112 | 103.9219 | 32.3 | 0.03 |
| pHILIC | 186.112 | 94.6133 | 32.3 | 0.03 |
| pHILIC | 186.112 | 42.9011 | 32.3 | 0.03 |
| pHILIC | 186.112 | 87.556 | 32.3 | 0.03 |
| pHILIC | 186.112 | 64.2287 | 32.3 | 0.03 |
| pHILIC | 186.112 | 128.43 | 32.3 | 0.03 |
| pHILIC | 186.112 | 119.4635 | 32.3 | 0.03 |
| pHILIC | 186.112 | 82.2007 | 32.3 | 0.03 |
| pHILIC | 186.112 | 131.9153 | 32.3 | 0.03 |
| pHILIC | 186.112 | 43.918 | 32.3 | 0.03 |
| pHILIC | 186.112 | 83.0575 | 32.3 | 0.03 |
| pHILIC | 186.112 | 50.5191 | 32.3 | 0.03 |
| pHILIC | 186.112 | 44.3868 | 32.3 | 0.03 |
| pHILIC | 186.112 | 52.2001 | 32.3 | 0.03 |
| pHILIC | 186.112 | 94.2737 | 32.3 | 0.03 |
| pHILIC | 186.112 | 62.1449 | 32.2 | 0.03 |
| pHILIC | 186.112 | 65.2342 | 32.2 | 0.03 |
| pHILIC | 186.112 | 164.4525 | 32.2 | 0.03 |
| pHILIC | 186.112 | 74.5058 | 32.2 | 0.03 |
| pHILIC | 186.112 | 91.7235 | 32.2 | 0.03 |
| pHILIC | 186.112 | 50.8349 | 32.2 | 0.03 |
| pHILIC | 186.112 | 44.4003 | 32.2 | 0.03 |
| pHILIC | 186.112 | 45.839 | 32.2 | 0.03 |
| pHILIC | 186.112 | 87.7461 | 32.2 | 0.03 |
| pHILIC | 186.112 | 77.5123 | 32.2 | 0.03 |
| pHILIC | 186.112 | 151.5957 | 32.2 | 0.03 |
| pHILIC | 186.112 | 173.3207 | 32.2 | 0.03 |
| pHILIC | 186.112 | 49.9138 | 32.2 | 0.03 |
| pHILIC | 186.112 | 49.7153 | 32.2 | 0.03 |
| pHILIC | 186.112 | 146.6709 | 32.2 | 0.03 |
| pHILIC | 186.112 | 152.3198 | 32.2 | 0.03 |

| | | | | |
|---|---|---|---|---|
| pHILIC | 186.112 | 43.4364 | 32.1 | 0.03 |
| pHILIC | 186.112 | 52.984 | 32.1 | 0.03 |
| pHILIC | 186.112 | 43.3096 | 32.1 | 0.03 |
| pHILIC | 186.112 | 77.715 | 32.1 | 0.03 |
| pHILIC | 186.112 | 107.1945 | 32.1 | 0.03 |
| pHILIC | 186.112 | 116.5844 | 32.1 | 0.03 |
| pHILIC | 186.112 | 128.5331 | 32.1 | 0.03 |
| pHILIC | 186.112 | 54.392 | 32.1 | 0.03 |
| pHILIC | 186.112 | 114.2446 | 32.1 | 0.03 |
| pHILIC | 186.112 | 188.6639 | 32.1 | 0.03 |
| pHILIC | 186.112 | 65.5451 | 32.1 | 0.03 |
| pHILIC | 186.112 | 42.8061 | 32.1 | 0.03 |
| pHILIC | 186.112 | 82.2735 | 32.1 | 0.03 |
| pHILIC | 186.112 | 44.4646 | 32.1 | 0.03 |
| pHILIC | 186.112 | 74.5941 | 32.1 | 0.03 |
| pHILIC | 186.112 | 84.7028 | 32.1 | 0.03 |
| pHILIC | 186.112 | 97.2121 | 32.1 | 0.03 |
| pHILIC | 186.112 | 120.419 | 32.1 | 0.03 |
| pHILIC | 186.112 | 123.1981 | 32.1 | 0.03 |
| pHILIC | 186.112 | 142.436 | 32.1 | 0.03 |
| pHILIC | 186.112 | 76.6963 | 32.1 | 0.03 |
| pHILIC | 186.112 | 56.3915 | 32.1 | 0.03 |
| pHILIC | 186.112 | 87.7302 | 32.1 | 0.03 |
| pHILIC | 186.112 | 83.9167 | 32 | 0.03 |
| pHILIC | 186.112 | 51.2367 | 32 | 0.03 |
| pHILIC | 186.112 | 119.8044 | 32 | 0.03 |
| pHILIC | 186.112 | 45.579 | 32 | 0.03 |
| pHILIC | 186.112 | 68.7961 | 32 | 0.03 |
| pHILIC | 186.112 | 113.9535 | 32 | 0.03 |
| pHILIC | 186.112 | 108.3876 | 32 | 0.03 |
| pHILIC | 186.112 | 53.3885 | 32 | 0.03 |
| pHILIC | 186.112 | 190.6575 | 32 | 0.03 |
| pHILIC | 186.112 | 198.2437 | 32 | 0.03 |
| pHILIC | 186.112 | 53.656 | 32 | 0.03 |
| pHILIC | 186.112 | 157.755 | 32 | 0.03 |
| pHILIC | 186.112 | 101.6565 | 32 | 0.03 |
| pHILIC | 186.112 | 44.888 | 32 | 0.03 |
| pHILIC | 186.112 | 173.3058 | 32 | 0.03 |
| pHILIC | 186.112 | 67.8485 | 32 | 0.03 |
| pHILIC | 186.112 | 71.4845 | 31.9 | 0.03 |
| pHILIC | 186.112 | 146.7796 | 31.9 | 0.03 |
| pHILIC | 186.112 | 163.9148 | 31.9 | 0.03 |

| | | | | |
|---|---|---|---|---|
| pHILIC | 186.112 | 101.358 | 31.9 | 0.03 |
| pHILIC | 186.112 | 42.6944 | 31.9 | 0.03 |
| pHILIC | 186.112 | 45.2772 | 31.9 | 0.03 |
| pHILIC | 186.112 | 44.6489 | 31.9 | 0.03 |
| pHILIC | 186.112 | 56.5944 | 31.9 | 0.03 |
| pHILIC | 186.112 | 127.1348 | 31.9 | 0.03 |
| pHILIC | 186.112 | 49.4852 | 31.9 | 0.03 |
| pHILIC | 186.112 | 68.7196 | 31.9 | 0.03 |
| pHILIC | 186.112 | 150.1605 | 31.9 | 0.03 |
| pHILIC | 186.112 | 93.3336 | 31.9 | 0.03 |
| pHILIC | 186.112 | 54.1156 | 31.8 | 0.03 |
| pHILIC | 186.112 | 120.8022 | 31.8 | 0.03 |
| pHILIC | 186.112 | 67.3727 | 31.8 | 0.03 |
| pHILIC | 186.112 | 157.449 | 31.8 | 0.03 |
| pHILIC | 186.112 | 199.1107 | 31.8 | 0.03 |
| pHILIC | 186.112 | 123.4067 | 31.8 | 0.03 |
| pHILIC | 186.112 | 50.0428 | 31.8 | 0.03 |
| pHILIC | 186.112 | 163.6656 | 31.8 | 0.03 |
| pHILIC | 186.112 | 72.5258 | 31.8 | 0.03 |
| pHILIC | 186.112 | 50.8927 | 31.8 | 0.03 |
| pHILIC | 186.112 | 71.6828 | 31.8 | 0.03 |
| pHILIC | 186.112 | 53.6976 | 31.8 | 0.03 |
| pHILIC | 186.112 | 107.7801 | 31.8 | 0.03 |
| pHILIC | 186.112 | 92.0651 | 31.8 | 0.03 |
| pHILIC | 186.112 | 75.2291 | 31.8 | 0.03 |
| pHILIC | 186.112 | 47.5966 | 31.7 | 0.03 |
| pHILIC | 186.112 | 93.9233 | 31.7 | 0.03 |
| pHILIC | 186.112 | 56.6945 | 31.7 | 0.03 |
| pHILIC | 186.112 | 102.6755 | 31.7 | 0.03 |
| pHILIC | 186.112 | 185.7276 | 31.7 | 0.03 |
| pHILIC | 186.112 | 90.1464 | 31.7 | 0.03 |
| pHILIC | 186.112 | 71.242 | 31.7 | 0.03 |
| pHILIC | 186.112 | 85.099 | 31.7 | 0.03 |
| pHILIC | 186.112 | 124.8162 | 31.7 | 0.03 |
| pHILIC | 186.112 | 44.0631 | 31.7 | 0.03 |
| pHILIC | 186.112 | 49.7454 | 31.7 | 0.03 |
| pHILIC | 186.112 | 61.6873 | 31.7 | 0.03 |
| pHILIC | 186.112 | 102.8518 | 31.7 | 0.03 |
| pHILIC | 186.112 | 99.4796 | 31.7 | 0.03 |
| pHILIC | 186.112 | 100.2392 | 31.6 | 0.03 |
| pHILIC | 186.112 | 58.7744 | 31.6 | 0.03 |
| pHILIC | 186.112 | 69.4587 | 31.6 | 0.03 |

| | | | | |
|---|---|---|---|---|
| pHILIC | 186.112 | 189.0745 | 31.6 | 0.03 |
| pHILIC | 186.112 | 119.8407 | 31.6 | 0.03 |
| pHILIC | 186.112 | 128.6102 | 31.6 | 0.03 |
| pHILIC | 186.112 | 110.3129 | 31.6 | 0.03 |
| pHILIC | 186.112 | 81.3557 | 31.6 | 0.03 |
| pHILIC | 186.112 | 47.9904 | 31.6 | 0.03 |
| pHILIC | 186.112 | 161.5228 | 31.6 | 0.03 |
| pHILIC | 186.112 | 137.7417 | 31.6 | 0.03 |
| pHILIC | 186.112 | 147.814 | 31.6 | 0.03 |
| pHILIC | 186.112 | 140.8414 | 31.6 | 0.03 |
| pHILIC | 186.112 | 125.2139 | 31.6 | 0.03 |
| pHILIC | 186.112 | 76.2251 | 31.6 | 0.03 |
| pHILIC | 186.112 | 42.8392 | 31.5 | 0.03 |
| pHILIC | 186.112 | 43.0019 | 31.5 | 0.03 |
| pHILIC | 186.112 | 77.4846 | 31.5 | 0.03 |
| pHILIC | 186.112 | 100.8822 | 31.5 | 0.03 |
| pHILIC | 186.112 | 48.5034 | 31.5 | 0.03 |
| pHILIC | 186.112 | 117.1389 | 31.5 | 0.03 |
| pHILIC | 186.112 | 207.0869 | 31.5 | 0.03 |
| pHILIC | 186.112 | 85.3633 | 31.5 | 0.03 |
| pHILIC | 186.112 | 43.3642 | 31.5 | 0.03 |
| pHILIC | 186.112 | 61.7218 | 31.5 | 0.03 |
| pHILIC | 186.112 | 64.271 | 31.5 | 0.03 |
| pHILIC | 186.112 | 72.9766 | 31.5 | 0.03 |
| pHILIC | 186.112 | 70.0016 | 31.5 | 0.03 |
| pHILIC | 186.112 | 61.2373 | 31.4 | 0.03 |
| pHILIC | 186.112 | 45.3743 | 31.4 | 0.03 |
| pHILIC | 186.112 | 160.1955 | 31.4 | 0.03 |
| pHILIC | 186.112 | 112.0912 | 31.4 | 0.03 |
| pHILIC | 186.112 | 83.0886 | 31.4 | 0.03 |
| pHILIC | 186.112 | 93.4312 | 31.4 | 0.03 |
| pHILIC | 186.112 | 157.5748 | 31.4 | 0.03 |
| pHILIC | 186.112 | 91.6087 | 31.4 | 0.03 |
| pHILIC | 186.112 | 110.3034 | 31.3 | 0.03 |
| pHILIC | 186.112 | 120.3811 | 31.3 | 0.03 |
| pHILIC | 186.112 | 141.1831 | 31.3 | 0.03 |
| pHILIC | 186.112 | 55.1074 | 31.3 | 0.03 |
| pHILIC | 186.112 | 88.5718 | 31.3 | 0.03 |
| pHILIC | 186.112 | 80.7253 | 31.2 | 0.03 |
| pHILIC | 186.112 | 104.0301 | 31.2 | 0.03 |
| pHILIC | 186.112 | 78.7676 | 31.2 | 0.03 |
| pHILIC | 186.112 | 130.1575 | 31.2 | 0.03 |

| | | | | |
|---|---|---|---|---|
| pHILIC | 186.112 | 47.6455 | 31.2 | 0.03 |
| pHILIC | 186.112 | 45.3948 | 31.2 | 0.03 |
| pHILIC | 186.112 | 62.0836 | 31.2 | 0.03 |
| pHILIC | 186.112 | 146.1542 | 31.1 | 0.03 |
| pHILIC | 186.112 | 111.1412 | 31.1 | 0.03 |
| pHILIC | 186.112 | 88.8798 | 31.1 | 0.03 |
| pHILIC | 186.112 | 46.0405 | 31.1 | 0.03 |
| pHILIC | 186.112 | 43.2371 | 31.1 | 0.03 |
| pHILIC | 186.112 | 94.7755 | 31.1 | 0.03 |
| pHILIC | 186.112 | 46.7085 | 31.1 | 0.03 |
| pHILIC | 186.112 | 43.4914 | 31.1 | 0.03 |
| pHILIC | 186.112 | 80.5176 | 31.1 | 0.03 |
| pHILIC | 186.112 | 45.5147 | 31 | 0.03 |
| pHILIC | 186.112 | 47.0309 | 31 | 0.03 |
| pHILIC | 186.112 | 54.3083 | 31 | 0.03 |
| pHILIC | 186.112 | 59.2266 | 31 | 0.03 |
| pHILIC | 186.112 | 83.6642 | 31 | 0.03 |
| pHILIC | 186.112 | 94.5625 | 31 | 0.03 |
| pHILIC | 186.112 | 47.4126 | 31 | 0.03 |
| pHILIC | 186.112 | 68.8639 | 31 | 0.03 |
| pHILIC | 186.112 | 61.1426 | 31 | 0.03 |
| pHILIC | 186.112 | 91.5478 | 31 | 0.03 |
| pHILIC | 186.112 | 59.7942 | 31 | 0.03 |
| pHILIC | 186.112 | 42.9558 | 30.9 | 0.03 |
| pHILIC | 186.112 | 117.3082 | 30.9 | 0.03 |
| pHILIC | 186.112 | 110.7176 | 30.9 | 0.03 |
| pHILIC | 186.112 | 56.8986 | 30.9 | 0.03 |
| pHILIC | 186.112 | 67.5789 | 30.9 | 0.03 |
| pHILIC | 186.112 | 55.2224 | 30.8 | 0.03 |
| pHILIC | 186.112 | 43.4258 | 30.8 | 0.03 |
| pHILIC | 186.112 | 108.3981 | 30.8 | 0.03 |
| pHILIC | 186.112 | 53.3717 | 30.8 | 0.03 |
| pHILIC | 186.112 | 44.8004 | 30.8 | 0.03 |
| pHILIC | 186.112 | 53.2195 | 30.8 | 0.03 |
| pHILIC | 186.112 | 54.1927 | 30.8 | 0.03 |
| pHILIC | 186.112 | 110.0024 | 30.8 | 0.03 |
| pHILIC | 186.112 | 49.5772 | 30.8 | 0.03 |
| pHILIC | 186.112 | 79.2701 | 30.7 | 0.03 |
| pHILIC | 186.112 | 57.478 | 30.7 | 0.03 |
| pHILIC | 186.112 | 87.6417 | 30.7 | 0.03 |
| pHILIC | 186.112 | 49.8956 | 30.7 | 0.03 |
| pHILIC | 186.112 | 53.5049 | 30.7 | 0.03 |

| | | | | |
|---|---|---|---|---|
| pHILIC | 186.112 | 181.9423 | 30.7 | 0.03 |
| pHILIC | 186.112 | 117.0599 | 30.7 | 0.03 |
| pHILIC | 186.112 | 122.7266 | 30.6 | 0.03 |
| pHILIC | 186.112 | 59.0862 | 30.6 | 0.03 |
| pHILIC | 186.112 | 53.7799 | 30.6 | 0.03 |
| pHILIC | 186.112 | 69.9855 | 30.6 | 0.03 |
| pHILIC | 186.112 | 171.7206 | 30.6 | 0.03 |
| pHILIC | 186.112 | 100.2706 | 30.6 | 0.03 |
| pHILIC | 186.112 | 89.4336 | 30.6 | 0.03 |
| pHILIC | 186.112 | 91.6288 | 30.6 | 0.03 |
| pHILIC | 186.112 | 46.3499 | 30.6 | 0.03 |
| pHILIC | 186.112 | 52.8015 | 30.6 | 0.03 |
| pHILIC | 186.112 | 90.0779 | 30.6 | 0.03 |
| pHILIC | 186.112 | 44.5373 | 30.5 | 0.03 |
| pHILIC | 186.112 | 49.264 | 30.5 | 0.03 |
| pHILIC | 186.112 | 64.116 | 30.5 | 0.03 |
| pHILIC | 186.112 | 51.6646 | 30.5 | 0.03 |
| pHILIC | 186.112 | 95.3108 | 30.5 | 0.03 |
| pHILIC | 186.112 | 91.6656 | 30.5 | 0.03 |
| pHILIC | 186.112 | 84.9211 | 30.5 | 0.03 |
| pHILIC | 186.112 | 55.16 | 30.4 | 0.03 |
| pHILIC | 186.112 | 79.1003 | 30.4 | 0.03 |
| pHILIC | 186.112 | 65.0581 | 30.4 | 0.03 |
| pHILIC | 186.112 | 129.6653 | 30.4 | 0.03 |
| pHILIC | 186.112 | 75.1626 | 30.3 | 0.03 |
| pHILIC | 186.112 | 149.0339 | 30.3 | 0.03 |
| pHILIC | 186.112 | 46.8037 | 30.3 | 0.03 |
| pHILIC | 186.112 | 44.2466 | 30.3 | 0.03 |
| pHILIC | 186.112 | 84.6037 | 30.2 | 0.03 |
| pHILIC | 186.112 | 61.6292 | 30.2 | 0.03 |
| pHILIC | 186.112 | 118.8267 | 30.2 | 0.03 |
| pHILIC | 186.112 | 73.1686 | 30.2 | 0.03 |
| pHILIC | 186.112 | 107.4094 | 30.2 | 0.03 |
| pHILIC | 186.112 | 100.7798 | 30.1 | 0.03 |
| pHILIC | 186.112 | 68.3245 | 30.1 | 0.03 |
| pHILIC | 186.112 | 140.4808 | 30.1 | 0.03 |
| pHILIC | 186.112 | 70.8823 | 30 | 0.03 |
| pHILIC | 186.112 | 56.3224 | 29.9 | 0.03 |
| pHILIC | 186.112 | 66.0005 | 29.8 | 0.03 |
| pHILIC | 186.112 | 48.3444 | 29.5 | 0.03 |
| pHILIC | 186.112 | 48.3465 | 29.4 | 0.03 |
| pHILIC | 186.112 | 64.3006 | 29.4 | 0.03 |

| | | | | |
|---|---|---|---|---|
| pHILIC | 186.112 | 81.2036 | 29.3 | 0.03 |
| pHILIC | 186.112 | 65.7309 | 29.2 | 0.03 |
| pHILIC | 186.112 | 197.2561 | 28.1 | 0.03 |
| pHILIC | 202.082 | 114.0662 | 5062.1 | 100 |
| pHILIC | 202.082 | 174.0549 | 1240.3 | 24.5 |
| pHILIC | 202.082 | 202.0502 | 928.5 | 18.34 |
| pHILIC | 202.082 | 202.0826 | 682.3 | 13.48 |
| pHILIC | 202.082 | 202.1438 | 534 | 10.55 |
| pHILIC | 202.082 | 146.0603 | 302 | 5.97 |
| pHILIC | 202.082 | 156.0448 | 216.9 | 4.28 |
| pHILIC | 202.082 | 132.0455 | 168.2 | 3.32 |
| pHILIC | 202.082 | 145.0058 | 164 | 3.24 |
| pHILIC | 202.082 | 202.1087 | 138.5 | 2.74 |
| pHILIC | 202.082 | 126.9957 | 121.7 | 2.4 |
| pHILIC | 202.082 | 67.801 | 119.5 | 2.36 |
| pHILIC | 202.082 | 175.6056 | 97.8 | 1.93 |
| pHILIC | 202.082 | 144.0658 | 84.4 | 1.67 |
| pHILIC | 202.082 | 138.1763 | 81 | 1.6 |
| pHILIC | 202.082 | 202.1784 | 79 | 1.56 |
| pHILIC | 202.082 | 186.489 | 76.8 | 1.52 |
| pHILIC | 202.082 | 133.4699 | 73.7 | 1.46 |
| pHILIC | 202.082 | 68.4387 | 73.6 | 1.45 |
| pHILIC | 202.082 | 219.4735 | 73.3 | 1.45 |
| pHILIC | 202.082 | 128.9142 | 72.7 | 1.44 |
| pHILIC | 202.082 | 200.7453 | 72.1 | 1.42 |
| pHILIC | 202.082 | 195.374 | 71.4 | 1.41 |
| pHILIC | 202.082 | 90.1432 | 71 | 1.4 |
| pHILIC | 202.082 | 66.0407 | 70.3 | 1.39 |
| pHILIC | 202.082 | 112.7232 | 70.3 | 1.39 |
| pHILIC | 202.082 | 203.2078 | 70.1 | 1.38 |
| pHILIC | 202.082 | 78.2802 | 69.3 | 1.37 |
| pHILIC | 202.082 | 138.8114 | 68.8 | 1.36 |
| pHILIC | 202.082 | 189.5611 | 68.1 | 1.35 |
| pHILIC | 202.082 | 179.9938 | 68.1 | 1.35 |
| pHILIC | 202.082 | 110.5263 | 67.8 | 1.34 |
| pHILIC | 202.082 | 213.0888 | 67.5 | 1.33 |
| pHILIC | 202.082 | 138.6344 | 66.4 | 1.31 |
| pHILIC | 202.082 | 51.6465 | 66.3 | 1.31 |
| pHILIC | 202.082 | 126.5312 | 65.8 | 1.3 |
| pHILIC | 202.082 | 60.0974 | 65.8 | 1.3 |
| pHILIC | 202.082 | 171.0697 | 65.7 | 1.3 |
| pHILIC | 202.082 | 155.4114 | 65.4 | 1.29 |

| | | | | |
|---|---|---|---|---|
| pHILIC | 202.082 | 139.4778 | 65.4 | 1.29 |
| pHILIC | 202.082 | 145.0662 | 64.6 | 1.28 |
| pHILIC | 202.082 | 172.9429 | 62.9 | 1.24 |
| pHILIC | 202.082 | 83.4335 | 62.7 | 1.24 |
| pHILIC | 202.082 | 109.3858 | 61.5 | 1.22 |
| pHILIC | 202.082 | 83.7531 | 60.9 | 1.2 |
| pHILIC | 202.082 | 131.1741 | 60.3 | 1.19 |
| pHILIC | 202.082 | 65.9773 | 59.9 | 1.18 |
| pHILIC | 202.082 | 179.8457 | 59.6 | 1.18 |
| pHILIC | 202.082 | 75.2273 | 57.6 | 1.14 |
| pHILIC | 202.082 | 130.8633 | 56.8 | 1.12 |
| pHILIC | 202.082 | 195.5381 | 56.2 | 1.11 |
| pHILIC | 202.082 | 130.065 | 56.1 | 1.11 |
| pHILIC | 202.082 | 60.6558 | 55.7 | 1.1 |
| pHILIC | 202.082 | 77.2285 | 54 | 1.07 |
| pHILIC | 202.082 | 178.6863 | 54 | 1.07 |
| pHILIC | 202.082 | 213.8329 | 54 | 1.07 |
| pHILIC | 202.082 | 95.9334 | 53.7 | 1.06 |
| pHILIC | 202.082 | 118.9231 | 53.6 | 1.06 |
| pHILIC | 202.082 | 141.9452 | 53.5 | 1.06 |
| pHILIC | 202.082 | 135.7855 | 53.2 | 1.05 |
| pHILIC | 202.082 | 98.6411 | 53 | 1.05 |
| pHILIC | 202.082 | 87.6225 | 52.8 | 1.04 |
| pHILIC | 202.082 | 136.1874 | 52.6 | 1.04 |
| pHILIC | 202.082 | 223.6765 | 52.3 | 1.03 |
| pHILIC | 202.082 | 62.1971 | 52.3 | 1.03 |
| pHILIC | 202.082 | 192.698 | 51.9 | 1.02 |
| pHILIC | 202.082 | 200.426 | 51.7 | 1.02 |
| pHILIC | 202.082 | 84.1979 | 51.7 | 1.02 |
| pHILIC | 202.082 | 177.4886 | 51.6 | 1.02 |
| pHILIC | 202.082 | 174.9518 | 51.6 | 1.02 |
| pHILIC | 202.082 | 77.2364 | 51.6 | 1.02 |
| pHILIC | 202.082 | 45.7012 | 51.5 | 1.02 |
| pHILIC | 202.082 | 174.292 | 51.4 | 1.02 |
| pHILIC | 202.082 | 99.3337 | 51.3 | 1.01 |
| pHILIC | 202.082 | 115.0557 | 51.2 | 1.01 |
| pHILIC | 202.082 | 48.0584 | 50.9 | 1.01 |
| pHILIC | 202.082 | 112.6349 | 50.8 | 1 |
| pHILIC | 202.082 | 215.0899 | 50.7 | 1 |
| pHILIC | 202.082 | 141.4068 | 50.6 | 1 |
| pHILIC | 202.082 | 177.0065 | 50.5 | 1 |
| pHILIC | 202.082 | 48.6069 | 50.4 | 1 |

| | | | | |
|---|---|---|---|---|
| pHILIC | 202.082 | 179.6705 | 50.4 | 0.99 |
| pHILIC | 202.082 | 194.5558 | 50.4 | 0.99 |
| pHILIC | 202.082 | 102.7258 | 50.4 | 0.99 |
| pHILIC | 202.082 | 176.29 | 50.3 | 0.99 |
| pHILIC | 202.082 | 94.5677 | 50.3 | 0.99 |
| pHILIC | 202.082 | 183.9796 | 50.1 | 0.99 |
| pHILIC | 202.082 | 129.9419 | 50.1 | 0.99 |
| pHILIC | 202.082 | 58.5755 | 50 | 0.99 |
| pHILIC | 202.082 | 175.4506 | 49.8 | 0.98 |
| pHILIC | 202.082 | 115.5895 | 49.7 | 0.98 |
| pHILIC | 202.082 | 151.1204 | 49.7 | 0.98 |
| pHILIC | 202.082 | 225.8498 | 49.7 | 0.98 |
| pHILIC | 202.082 | 94.4733 | 49.4 | 0.98 |
| pHILIC | 202.082 | 60.8285 | 49.4 | 0.98 |
| pHILIC | 202.082 | 138.7309 | 49.3 | 0.97 |
| pHILIC | 202.082 | 57.6101 | 49.3 | 0.97 |
| pHILIC | 202.082 | 75.0559 | 49.3 | 0.97 |
| pHILIC | 202.082 | 70.0633 | 49.1 | 0.97 |
| pHILIC | 202.082 | 170.3268 | 49.1 | 0.97 |
| pHILIC | 202.082 | 107.1391 | 48.9 | 0.97 |
| pHILIC | 202.082 | 72.1317 | 48.9 | 0.97 |
| pHILIC | 202.082 | 214.6325 | 48.8 | 0.96 |
| pHILIC | 202.082 | 169.605 | 48.8 | 0.96 |
| pHILIC | 202.082 | 129.6608 | 48.8 | 0.96 |
| pHILIC | 202.082 | 79.306 | 48.8 | 0.96 |
| pHILIC | 202.082 | 205.2653 | 48.8 | 0.96 |
| pHILIC | 202.082 | 118.2679 | 48.8 | 0.96 |
| pHILIC | 202.082 | 45.5862 | 48.8 | 0.96 |
| pHILIC | 202.082 | 189.1344 | 48.7 | 0.96 |
| pHILIC | 202.082 | 80.5055 | 48.7 | 0.96 |
| pHILIC | 202.082 | 49.2694 | 48.7 | 0.96 |
| pHILIC | 202.082 | 53.0317 | 48.7 | 0.96 |
| pHILIC | 202.082 | 87.468 | 48.7 | 0.96 |
| pHILIC | 202.082 | 158.7227 | 48.6 | 0.96 |
| pHILIC | 202.082 | 158.6028 | 48.5 | 0.96 |
| pHILIC | 202.082 | 70.0947 | 48.5 | 0.96 |
| pHILIC | 202.082 | 213.5907 | 48.5 | 0.96 |
| pHILIC | 202.082 | 99.4613 | 48.4 | 0.96 |
| pHILIC | 202.082 | 197.9275 | 48.4 | 0.96 |
| pHILIC | 202.082 | 200.0857 | 48.4 | 0.96 |
| pHILIC | 202.082 | 88.0774 | 48.3 | 0.96 |
| pHILIC | 202.082 | 160.4433 | 48.3 | 0.95 |

| | | | | |
|---|---|---|---|---|
| pHILIC | 202.082 | 131.0096 | 48.3 | 0.95 |
| pHILIC | 202.082 | 136.9821 | 48.2 | 0.95 |
| pHILIC | 202.082 | 60.7373 | 48 | 0.95 |
| pHILIC | 202.082 | 76.7146 | 48 | 0.95 |
| pHILIC | 202.082 | 135.031 | 48 | 0.95 |
| pHILIC | 202.082 | 160.5009 | 47.9 | 0.95 |
| pHILIC | 202.082 | 49.3395 | 47.9 | 0.95 |
| pHILIC | 202.082 | 71.5419 | 47.9 | 0.95 |
| pHILIC | 202.082 | 49.5718 | 47.8 | 0.94 |
| pHILIC | 202.082 | 113.895 | 47.7 | 0.94 |
| pHILIC | 202.082 | 63.4148 | 47.6 | 0.94 |
| pHILIC | 202.082 | 87.8481 | 47.6 | 0.94 |
| pHILIC | 202.082 | 166.9614 | 47.5 | 0.94 |
| pHILIC | 202.082 | 101.1759 | 47.5 | 0.94 |
| pHILIC | 202.082 | 196.7943 | 47.4 | 0.94 |
| pHILIC | 202.082 | 67.4198 | 47.4 | 0.94 |
| pHILIC | 202.082 | 60.319 | 47.4 | 0.94 |
| pHILIC | 202.082 | 221.507 | 47.3 | 0.93 |
| pHILIC | 202.082 | 53.1306 | 47.3 | 0.93 |
| pHILIC | 202.082 | 67.2806 | 47.3 | 0.93 |
| pHILIC | 202.082 | 70.0314 | 47.3 | 0.93 |
| pHILIC | 202.082 | 190.2791 | 47.2 | 0.93 |
| pHILIC | 202.082 | 107.2402 | 47.2 | 0.93 |
| pHILIC | 202.082 | 83.1071 | 47.2 | 0.93 |
| pHILIC | 202.082 | 108.4707 | 47.1 | 0.93 |
| pHILIC | 202.082 | 84.0829 | 47.1 | 0.93 |
| pHILIC | 202.082 | 59.4787 | 47.1 | 0.93 |
| pHILIC | 202.082 | 55.6355 | 47 | 0.93 |
| pHILIC | 202.082 | 121.1488 | 47 | 0.93 |
| pHILIC | 202.082 | 58.9691 | 46.9 | 0.93 |
| pHILIC | 202.082 | 176.3087 | 46.9 | 0.93 |
| pHILIC | 202.082 | 106.3457 | 46.9 | 0.93 |
| pHILIC | 202.082 | 223.994 | 46.8 | 0.93 |
| pHILIC | 202.082 | 140.9556 | 46.8 | 0.92 |
| pHILIC | 202.082 | 65.9368 | 46.8 | 0.92 |
| pHILIC | 202.082 | 71.0105 | 46.8 | 0.92 |
| pHILIC | 202.082 | 56.5077 | 46.8 | 0.92 |
| pHILIC | 202.082 | 166.8166 | 46.8 | 0.92 |
| pHILIC | 202.082 | 61.0793 | 46.7 | 0.92 |
| pHILIC | 202.082 | 219.619 | 46.7 | 0.92 |
| pHILIC | 202.082 | 126.9797 | 46.7 | 0.92 |
| pHILIC | 202.082 | 87.7516 | 46.6 | 0.92 |

| | | | | |
|---|---|---|---|---|
| pHILIC | 202.082 | 88.8896 | 46.5 | 0.92 |
| pHILIC | 202.082 | 148.8344 | 46.5 | 0.92 |
| pHILIC | 202.082 | 58.3883 | 46.5 | 0.92 |
| pHILIC | 202.082 | 174.7693 | 46.4 | 0.92 |
| pHILIC | 202.082 | 72.7373 | 46.4 | 0.92 |
| pHILIC | 202.082 | 65.5008 | 46.3 | 0.92 |
| pHILIC | 202.082 | 80.2343 | 46.3 | 0.91 |
| pHILIC | 202.082 | 62.2441 | 46.3 | 0.91 |
| pHILIC | 202.082 | 185.5993 | 46.2 | 0.91 |
| pHILIC | 202.082 | 186.5432 | 46.2 | 0.91 |
| pHILIC | 202.082 | 72.7283 | 46.2 | 0.91 |
| pHILIC | 202.082 | 78.2861 | 46.2 | 0.91 |
| pHILIC | 202.082 | 53.26 | 46.1 | 0.91 |
| pHILIC | 202.082 | 177.8966 | 46.1 | 0.91 |
| pHILIC | 202.082 | 66.3608 | 46.1 | 0.91 |
| pHILIC | 202.082 | 91.1051 | 46 | 0.91 |
| pHILIC | 202.082 | 65.8839 | 46 | 0.91 |
| pHILIC | 202.082 | 162.6969 | 46 | 0.91 |
| pHILIC | 202.082 | 215.6276 | 46 | 0.91 |
| pHILIC | 202.082 | 66.7774 | 45.9 | 0.91 |
| pHILIC | 202.082 | 85.5824 | 45.9 | 0.91 |
| pHILIC | 202.082 | 134.1164 | 45.9 | 0.91 |
| pHILIC | 202.082 | 54.8848 | 45.8 | 0.9 |
| pHILIC | 202.082 | 59.0033 | 45.8 | 0.9 |
| pHILIC | 202.082 | 83.1105 | 45.8 | 0.9 |
| pHILIC | 202.082 | 80.6519 | 45.8 | 0.9 |
| pHILIC | 202.082 | 96.6212 | 45.7 | 0.9 |
| pHILIC | 202.082 | 68.5491 | 45.7 | 0.9 |
| pHILIC | 202.082 | 105.77 | 45.7 | 0.9 |
| pHILIC | 202.082 | 127.3188 | 45.7 | 0.9 |
| pHILIC | 202.082 | 57.7781 | 45.6 | 0.9 |
| pHILIC | 202.082 | 73.0845 | 45.6 | 0.9 |
| pHILIC | 202.082 | 68.013 | 45.6 | 0.9 |
| pHILIC | 202.082 | 54.2703 | 45.5 | 0.9 |
| pHILIC | 202.082 | 94.236 | 45.5 | 0.9 |
| pHILIC | 202.082 | 69.3982 | 45.5 | 0.9 |
| pHILIC | 202.082 | 77.6851 | 45.4 | 0.9 |
| pHILIC | 202.082 | 45.7791 | 45.3 | 0.9 |
| pHILIC | 202.082 | 106.2115 | 45.3 | 0.89 |
| pHILIC | 202.082 | 126.1463 | 45.3 | 0.89 |
| pHILIC | 202.082 | 125.6957 | 45.3 | 0.89 |
| pHILIC | 202.082 | 105.4468 | 45.2 | 0.89 |

| | | | | |
|---|---|---|---|---|
| pHILIC | 202.082 | 215.9117 | 45.2 | 0.89 |
| pHILIC | 202.082 | 131.6419 | 45.2 | 0.89 |
| pHILIC | 202.082 | 87.5671 | 45.2 | 0.89 |
| pHILIC | 202.082 | 91.751 | 45.1 | 0.89 |
| pHILIC | 202.082 | 213.6318 | 45 | 0.89 |
| pHILIC | 202.082 | 55.7732 | 44.9 | 0.89 |
| pHILIC | 202.082 | 86.1714 | 44.9 | 0.89 |
| pHILIC | 202.082 | 121.2861 | 44.9 | 0.89 |
| pHILIC | 202.082 | 77.8228 | 44.9 | 0.89 |
| pHILIC | 202.082 | 69.576 | 44.8 | 0.89 |
| pHILIC | 202.082 | 123.2996 | 44.8 | 0.88 |
| pHILIC | 202.082 | 136.8852 | 44.8 | 0.88 |
| pHILIC | 202.082 | 55.8447 | 44.8 | 0.88 |
| pHILIC | 202.082 | 160.3431 | 44.7 | 0.88 |
| pHILIC | 202.082 | 223.0703 | 44.7 | 0.88 |
| pHILIC | 202.082 | 108.8987 | 44.7 | 0.88 |
| pHILIC | 202.082 | 105.0697 | 44.6 | 0.88 |
| pHILIC | 202.082 | 208.4446 | 44.6 | 0.88 |
| pHILIC | 202.082 | 130.7059 | 44.6 | 0.88 |
| pHILIC | 202.082 | 89.6621 | 44.5 | 0.88 |
| pHILIC | 202.082 | 47.5808 | 44.5 | 0.88 |
| pHILIC | 202.082 | 147.8712 | 44.5 | 0.88 |
| pHILIC | 202.082 | 90.2887 | 44.5 | 0.88 |
| pHILIC | 202.082 | 191.3647 | 44.5 | 0.88 |
| pHILIC | 202.082 | 45.6136 | 44.5 | 0.88 |
| pHILIC | 202.082 | 225.2661 | 44.4 | 0.88 |
| pHILIC | 202.082 | 62.4477 | 44.4 | 0.88 |
| pHILIC | 202.082 | 53.1288 | 44.3 | 0.88 |
| pHILIC | 202.082 | 206.2134 | 44.3 | 0.88 |
| pHILIC | 202.082 | 78.0564 | 44.2 | 0.87 |
| pHILIC | 202.082 | 123.0479 | 44.2 | 0.87 |
| pHILIC | 202.082 | 121.1302 | 44 | 0.87 |
| pHILIC | 202.082 | 93.6413 | 44 | 0.87 |
| pHILIC | 202.082 | 50.8913 | 43.9 | 0.87 |
| pHILIC | 202.082 | 50.7847 | 43.9 | 0.87 |
| pHILIC | 202.082 | 87.3619 | 43.8 | 0.87 |
| pHILIC | 202.082 | 78.3975 | 43.8 | 0.87 |
| pHILIC | 202.082 | 138.6097 | 43.8 | 0.87 |
| pHILIC | 202.082 | 147.5505 | 43.8 | 0.86 |
| pHILIC | 202.082 | 54.1929 | 43.8 | 0.86 |
| pHILIC | 202.082 | 71.3896 | 43.7 | 0.86 |
| pHILIC | 202.082 | 168.8627 | 43.6 | 0.86 |

| | | | | |
|---|---|---|---|---|
| pHILIC | 202.082 | 71.6344 | 43.5 | 0.86 |
| pHILIC | 202.082 | 158.1513 | 43.5 | 0.86 |
| pHILIC | 202.082 | 219.4223 | 43.5 | 0.86 |
| pHILIC | 202.082 | 204.7902 | 43.5 | 0.86 |
| pHILIC | 202.082 | 97.4916 | 43.3 | 0.86 |
| pHILIC | 202.082 | 164.0588 | 43.3 | 0.86 |
| pHILIC | 202.082 | 77.2791 | 43.3 | 0.85 |
| pHILIC | 202.082 | 164.0811 | 43.2 | 0.85 |
| pHILIC | 202.082 | 84.2502 | 43.2 | 0.85 |
| pHILIC | 202.082 | 68.0053 | 43.2 | 0.85 |
| pHILIC | 202.082 | 170.5294 | 43.1 | 0.85 |
| pHILIC | 202.082 | 107.8588 | 43.1 | 0.85 |
| pHILIC | 202.082 | 59.991 | 43.1 | 0.85 |
| pHILIC | 202.082 | 89.707 | 43.1 | 0.85 |
| pHILIC | 202.082 | 146.1613 | 43 | 0.85 |
| pHILIC | 202.082 | 71.3051 | 42.9 | 0.85 |
| pHILIC | 202.082 | 53.5027 | 42.9 | 0.85 |
| pHILIC | 202.082 | 98.7218 | 42.9 | 0.85 |
| pHILIC | 202.082 | 139.7675 | 42.8 | 0.85 |
| pHILIC | 202.082 | 192.7683 | 42.8 | 0.84 |
| pHILIC | 202.082 | 59.7554 | 42.8 | 0.84 |
| pHILIC | 202.082 | 70.3501 | 42.6 | 0.84 |
| pHILIC | 202.082 | 69.3351 | 42.6 | 0.84 |
| pHILIC | 202.082 | 173.9551 | 42.6 | 0.84 |
| pHILIC | 202.082 | 83.8276 | 42.6 | 0.84 |
| pHILIC | 202.082 | 151.1387 | 42.5 | 0.84 |
| pHILIC | 202.082 | 140.5197 | 42.5 | 0.84 |
| pHILIC | 202.082 | 51.368 | 42.4 | 0.84 |
| pHILIC | 202.082 | 104.8479 | 42.4 | 0.84 |
| pHILIC | 202.082 | 130.1375 | 42.4 | 0.84 |
| pHILIC | 202.082 | 210.155 | 42.4 | 0.84 |
| pHILIC | 202.082 | 59.7058 | 42.3 | 0.84 |
| pHILIC | 202.082 | 95.6263 | 42.3 | 0.83 |
| pHILIC | 202.082 | 46.7355 | 42.2 | 0.83 |
| pHILIC | 202.082 | 184.1917 | 42.2 | 0.83 |
| pHILIC | 202.082 | 115.8584 | 42.2 | 0.83 |
| pHILIC | 202.082 | 58.601 | 42.2 | 0.83 |
| pHILIC | 202.082 | 124.7384 | 42.1 | 0.83 |
| pHILIC | 202.082 | 55.1832 | 42.1 | 0.83 |
| pHILIC | 202.082 | 138.7107 | 42.1 | 0.83 |
| pHILIC | 202.082 | 96.4534 | 42 | 0.83 |
| pHILIC | 202.082 | 199.6706 | 42 | 0.83 |

| | | | | |
|---|---|---|---|---|
| pHILIC | 202.082 | 90.5407 | 42 | 0.83 |
| pHILIC | 202.082 | 106.5502 | 41.9 | 0.83 |
| pHILIC | 202.082 | 146.1343 | 41.9 | 0.83 |
| pHILIC | 202.082 | 78.4363 | 41.9 | 0.83 |
| pHILIC | 202.082 | 58.9163 | 41.9 | 0.83 |
| pHILIC | 202.082 | 120.1416 | 41.9 | 0.83 |
| pHILIC | 202.082 | 91.2574 | 41.9 | 0.83 |
| pHILIC | 202.082 | 166.3141 | 41.8 | 0.83 |
| pHILIC | 202.082 | 55.2916 | 41.8 | 0.83 |
| pHILIC | 202.082 | 75.4597 | 41.7 | 0.82 |
| pHILIC | 202.082 | 51.0883 | 41.7 | 0.82 |
| pHILIC | 202.082 | 73.3604 | 41.7 | 0.82 |
| pHILIC | 202.082 | 109.5389 | 41.7 | 0.82 |
| pHILIC | 202.082 | 80.1934 | 41.5 | 0.82 |
| pHILIC | 202.082 | 127.981 | 41.5 | 0.82 |
| pHILIC | 202.082 | 60.5431 | 41.5 | 0.82 |
| pHILIC | 202.082 | 156.6974 | 41.5 | 0.82 |
| pHILIC | 202.082 | 149.9901 | 41.4 | 0.82 |
| pHILIC | 202.082 | 188.0561 | 41.3 | 0.82 |
| pHILIC | 202.082 | 124.8716 | 41.3 | 0.82 |
| pHILIC | 202.082 | 82.1025 | 41.3 | 0.82 |
| pHILIC | 202.082 | 142.7102 | 41.2 | 0.81 |
| pHILIC | 202.082 | 46.8034 | 41.2 | 0.81 |
| pHILIC | 202.082 | 76.5174 | 41.1 | 0.81 |
| pHILIC | 202.082 | 187.845 | 41.1 | 0.81 |
| pHILIC | 202.082 | 119.5869 | 41.1 | 0.81 |
| pHILIC | 202.082 | 52.3146 | 41 | 0.81 |
| pHILIC | 202.082 | 143.7096 | 40.9 | 0.81 |
| pHILIC | 202.082 | 105.2249 | 40.9 | 0.81 |
| pHILIC | 202.082 | 57.6712 | 40.9 | 0.81 |
| pHILIC | 202.082 | 61.4063 | 40.8 | 0.81 |
| pHILIC | 202.082 | 209.9021 | 40.8 | 0.81 |
| pHILIC | 202.082 | 130.9455 | 40.7 | 0.8 |
| pHILIC | 202.082 | 122.3854 | 40.7 | 0.8 |
| pHILIC | 202.082 | 106.2297 | 40.5 | 0.8 |
| pHILIC | 202.082 | 186.2505 | 40.4 | 0.8 |
| pHILIC | 202.082 | 145.7277 | 40.4 | 0.8 |
| pHILIC | 202.082 | 46.5237 | 40.4 | 0.8 |
| pHILIC | 202.082 | 52.5743 | 40.4 | 0.8 |
| pHILIC | 202.082 | 151.217 | 40.3 | 0.8 |
| pHILIC | 202.082 | 58.9943 | 40.3 | 0.8 |
| pHILIC | 202.082 | 210.6226 | 40.1 | 0.79 |

| | | | | |
|---|---|---|---|---|
| pHILIC | 202.082 | 64.726 | 40 | 0.79 |
| pHILIC | 202.082 | 210.8517 | 40 | 0.79 |
| pHILIC | 202.082 | 66.7895 | 39.9 | 0.79 |
| pHILIC | 202.082 | 71.6734 | 39.9 | 0.79 |
| pHILIC | 202.082 | 101.5099 | 39.8 | 0.79 |
| pHILIC | 202.082 | 91.5278 | 39.7 | 0.79 |
| pHILIC | 202.082 | 157.5922 | 39.7 | 0.78 |
| pHILIC | 202.082 | 121.2597 | 39.7 | 0.78 |
| pHILIC | 202.082 | 57.7928 | 39.6 | 0.78 |
| pHILIC | 202.082 | 80.5381 | 39.6 | 0.78 |
| pHILIC | 202.082 | 55.8883 | 39.6 | 0.78 |
| pHILIC | 202.082 | 88.5035 | 39.4 | 0.78 |
| pHILIC | 202.082 | 154.8996 | 39.3 | 0.78 |
| pHILIC | 202.082 | 81.6816 | 39.2 | 0.77 |
| pHILIC | 202.082 | 92.781 | 39.1 | 0.77 |
| pHILIC | 202.082 | 211.1908 | 39 | 0.77 |
| pHILIC | 202.082 | 136.386 | 38.9 | 0.77 |
| pHILIC | 202.082 | 89.2416 | 38.7 | 0.76 |
| pHILIC | 202.082 | 82.6848 | 38.6 | 0.76 |
| pHILIC | 202.082 | 53.4691 | 38.6 | 0.76 |
| pHILIC | 202.082 | 160.7561 | 38.5 | 0.76 |
| pHILIC | 202.082 | 222.6491 | 38.3 | 0.76 |
| pHILIC | 202.082 | 90.6871 | 38.3 | 0.76 |
| pHILIC | 202.082 | 58.0799 | 38.1 | 0.75 |
| pHILIC | 202.082 | 74.9887 | 38 | 0.75 |
| pHILIC | 202.082 | 72.1624 | 38 | 0.75 |
| pHILIC | 202.082 | 92.6063 | 38 | 0.75 |
| pHILIC | 202.082 | 223.4666 | 38 | 0.75 |
| pHILIC | 202.082 | 137.8576 | 38 | 0.75 |
| pHILIC | 202.082 | 47.9827 | 37.9 | 0.75 |
| pHILIC | 202.082 | 112.4972 | 37.8 | 0.75 |
| pHILIC | 202.082 | 153.6404 | 37.2 | 0.73 |
| pHILIC | 202.082 | 53.5217 | 37.1 | 0.73 |
| pHILIC | 202.082 | 190.9734 | 37 | 0.73 |
| pHILIC | 202.082 | 97.6085 | 37 | 0.73 |
| pHILIC | 202.082 | 108.5516 | 36.5 | 0.72 |
| pHILIC | 202.082 | 67.7148 | 36.4 | 0.72 |
| pHILIC | 202.082 | 182.6688 | 36.3 | 0.72 |
| pHILIC | 202.082 | 67.8281 | 35.8 | 0.71 |
| pHILIC | 202.082 | 78.6811 | 35.4 | 0.7 |
| pHILIC | 202.082 | 50.8203 | 35.4 | 0.7 |
| pHILIC | 202.082 | 73.3833 | 35.4 | 0.7 |

| | | | | |
|---|---|---|---|---|
| pHILIC | 202.082 | 51.4512 | 34.1 | 0.67 |
| pHILIC | 202.082 | 173.5961 | 33.6 | 0.66 |
| pHILIC | 202.082 | 162.5272 | 33.3 | 0.66 |
| pHILIC | 202.082 | 61.0534 | 33.1 | 0.65 |
| pHILIC | 202.082 | 221.0322 | 33 | 0.65 |
| pHILIC | 202.082 | 85.949 | 32.4 | 0.64 |
| pHILIC | 202.082 | 62.4623 | 32.3 | 0.64 |
| pHILIC | 202.082 | 179.2268 | 32 | 0.63 |
| pHILIC | 202.082 | 199.7118 | 31.7 | 0.63 |
| pHILIC | 202.082 | 148.1613 | 31.3 | 0.62 |
| pHILIC | 202.082 | 224.5006 | 31.2 | 0.62 |
| pHILIC | 202.082 | 224.6946 | 31.1 | 0.62 |
| pHILIC | 202.082 | 144.8996 | 31 | 0.61 |
| pHILIC | 202.082 | 66.7981 | 31 | 0.61 |
| pHILIC | 202.082 | 60.4238 | 30.6 | 0.6 |
| pHILIC | 202.082 | 67.9727 | 30.6 | 0.6 |
| pHILIC | 202.082 | 99.7614 | 30.4 | 0.6 |
| pHILIC | 202.082 | 67.4854 | 30.4 | 0.6 |
| pHILIC | 202.082 | 50.9075 | 30.2 | 0.6 |
| pHILIC | 202.082 | 146.8147 | 30.2 | 0.6 |
| pHILIC | 202.082 | 52.1081 | 30.1 | 0.59 |
| pHILIC | 202.082 | 85.5269 | 30 | 0.59 |
| pHILIC | 202.082 | 78.9816 | 30 | 0.59 |
| pHILIC | 202.082 | 80.3909 | 30 | 0.59 |
| pHILIC | 202.082 | 104.9824 | 30 | 0.59 |
| pHILIC | 202.082 | 127.6592 | 29.9 | 0.59 |
| pHILIC | 202.082 | 60.5987 | 29.8 | 0.59 |
| pHILIC | 202.082 | 139.103 | 29.8 | 0.59 |
| pHILIC | 202.082 | 182.3717 | 29.8 | 0.59 |
| pHILIC | 202.082 | 111.3383 | 29.8 | 0.59 |
| pHILIC | 202.082 | 93.3748 | 29.7 | 0.59 |
| pHILIC | 202.082 | 210.8081 | 29.6 | 0.58 |
| pHILIC | 202.082 | 99.0553 | 29.5 | 0.58 |
| pHILIC | 202.082 | 89.1766 | 29.5 | 0.58 |
| pHILIC | 202.082 | 60.0538 | 29.5 | 0.58 |
| pHILIC | 202.082 | 57.1917 | 29.5 | 0.58 |
| pHILIC | 202.082 | 62.1389 | 29.5 | 0.58 |
| pHILIC | 202.082 | 81.1375 | 29.4 | 0.58 |
| pHILIC | 202.082 | 123.6359 | 29.3 | 0.58 |
| pHILIC | 202.082 | 191.0027 | 29.3 | 0.58 |
| pHILIC | 202.082 | 185.56 | 29.2 | 0.58 |
| pHILIC | 202.082 | 58.5654 | 29.1 | 0.58 |

| | | | | |
|---|---|---|---|---|
| pHILIC | 202.082 | 204.6168 | 29.1 | 0.57 |
| pHILIC | 202.082 | 135.9507 | 29 | 0.57 |
| pHILIC | 202.082 | 195.7671 | 29 | 0.57 |
| pHILIC | 202.082 | 60.1621 | 29 | 0.57 |
| pHILIC | 202.082 | 50.5198 | 29 | 0.57 |
| pHILIC | 202.082 | 50.9689 | 29 | 0.57 |
| pHILIC | 202.082 | 82.8227 | 28.9 | 0.57 |
| pHILIC | 202.082 | 161.6576 | 28.9 | 0.57 |
| pHILIC | 202.082 | 71.0537 | 28.9 | 0.57 |
| pHILIC | 202.082 | 60.1717 | 28.8 | 0.57 |
| pHILIC | 202.082 | 77.8415 | 28.8 | 0.57 |
| pHILIC | 202.082 | 91.8019 | 28.8 | 0.57 |
| pHILIC | 202.082 | 113.0054 | 28.8 | 0.57 |
| pHILIC | 202.082 | 195.6449 | 28.7 | 0.57 |
| pHILIC | 202.082 | 199.8039 | 28.7 | 0.57 |
| pHILIC | 202.082 | 222.4975 | 28.6 | 0.57 |
| pHILIC | 202.082 | 128.9708 | 28.6 | 0.56 |
| pHILIC | 202.082 | 53.6066 | 28.6 | 0.56 |
| pHILIC | 202.082 | 61.7752 | 28.6 | 0.56 |
| pHILIC | 202.082 | 80.8054 | 28.6 | 0.56 |
| pHILIC | 202.082 | 75.043 | 28.5 | 0.56 |
| pHILIC | 202.082 | 182.8636 | 28.5 | 0.56 |
| pHILIC | 202.082 | 92.8024 | 28.5 | 0.56 |
| pHILIC | 202.082 | 113.9614 | 28.5 | 0.56 |
| pHILIC | 202.082 | 141.8341 | 28.5 | 0.56 |
| pHILIC | 202.082 | 186.0723 | 28.4 | 0.56 |
| pHILIC | 202.082 | 97.3189 | 28.4 | 0.56 |
| pHILIC | 202.082 | 85.0985 | 28.3 | 0.56 |
| pHILIC | 202.082 | 51.6712 | 28.3 | 0.56 |
| pHILIC | 202.082 | 191.7274 | 28.3 | 0.56 |
| pHILIC | 202.082 | 166.0574 | 28.3 | 0.56 |
| pHILIC | 202.082 | 206.2417 | 28.3 | 0.56 |
| pHILIC | 202.082 | 82.5284 | 28.3 | 0.56 |
| pHILIC | 202.082 | 219.1924 | 28.3 | 0.56 |
| pHILIC | 202.082 | 85.2036 | 28.3 | 0.56 |
| pHILIC | 202.082 | 55.0058 | 28.2 | 0.56 |
| pHILIC | 202.082 | 104.1575 | 28.2 | 0.56 |
| pHILIC | 202.082 | 99.6742 | 28.1 | 0.56 |
| pHILIC | 202.082 | 115.6192 | 28.1 | 0.56 |
| pHILIC | 202.082 | 74.1246 | 28.1 | 0.55 |
| pHILIC | 202.082 | 211.2553 | 28 | 0.55 |
| pHILIC | 202.082 | 199.9559 | 28 | 0.55 |

| | | | | |
|---|---|---|---|---|
| pHILIC | 202.082 | 142.0629 | 28 | 0.55 |
| pHILIC | 202.082 | 56.0689 | 28 | 0.55 |
| pHILIC | 202.082 | 108.4884 | 28 | 0.55 |
| pHILIC | 202.082 | 99.8123 | 28 | 0.55 |
| pHILIC | 202.082 | 50.8933 | 27.9 | 0.55 |
| pHILIC | 202.082 | 167.2275 | 27.9 | 0.55 |
| pHILIC | 202.082 | 69.9983 | 27.9 | 0.55 |
| pHILIC | 202.082 | 203.6387 | 27.9 | 0.55 |
| pHILIC | 202.082 | 95.5328 | 27.9 | 0.55 |
| pHILIC | 202.082 | 190.4076 | 27.9 | 0.55 |
| pHILIC | 202.082 | 136.8407 | 27.9 | 0.55 |
| pHILIC | 202.082 | 64.8102 | 27.9 | 0.55 |
| pHILIC | 202.082 | 90.8065 | 27.9 | 0.55 |
| pHILIC | 202.082 | 62.4718 | 27.8 | 0.55 |
| pHILIC | 202.082 | 46.3541 | 27.8 | 0.55 |
| pHILIC | 202.082 | 56.6724 | 27.8 | 0.55 |
| pHILIC | 202.082 | 117.3095 | 27.8 | 0.55 |
| pHILIC | 202.082 | 199.0073 | 27.8 | 0.55 |
| pHILIC | 202.082 | 45.9909 | 27.8 | 0.55 |
| pHILIC | 202.082 | 88.4599 | 27.8 | 0.55 |
| pHILIC | 202.082 | 58.3766 | 27.8 | 0.55 |
| pHILIC | 202.082 | 102.8173 | 27.7 | 0.55 |
| pHILIC | 202.082 | 95.3514 | 27.7 | 0.55 |
| pHILIC | 202.082 | 196.1642 | 27.7 | 0.55 |
| pHILIC | 202.082 | 166.8425 | 27.7 | 0.55 |
| pHILIC | 202.082 | 95.8643 | 27.7 | 0.55 |
| pHILIC | 202.082 | 61.5499 | 27.7 | 0.55 |
| pHILIC | 202.082 | 124.4567 | 27.7 | 0.55 |
| pHILIC | 202.082 | 118.6328 | 27.7 | 0.55 |
| pHILIC | 202.082 | 46.2533 | 27.7 | 0.55 |
| pHILIC | 202.082 | 106.2808 | 27.7 | 0.55 |
| pHILIC | 202.082 | 100.452 | 27.7 | 0.55 |
| pHILIC | 202.082 | 50.1554 | 27.7 | 0.55 |
| pHILIC | 202.082 | 115.8335 | 27.7 | 0.55 |
| pHILIC | 202.082 | 128.0872 | 27.7 | 0.55 |
| pHILIC | 202.082 | 55.4808 | 27.6 | 0.55 |
| pHILIC | 202.082 | 140.5517 | 27.6 | 0.55 |
| pHILIC | 202.082 | 47.2451 | 27.6 | 0.55 |
| pHILIC | 202.082 | 86.3788 | 27.6 | 0.55 |
| pHILIC | 202.082 | 92.0036 | 27.6 | 0.55 |
| pHILIC | 202.082 | 49.6458 | 27.6 | 0.55 |
| pHILIC | 202.082 | 92.5443 | 27.6 | 0.55 |

| | | | | |
|---|---|---|---|---|
| pHILIC | 202.082 | 92.7972 | 27.6 | 0.54 |
| pHILIC | 202.082 | 54.142 | 27.5 | 0.54 |
| pHILIC | 202.082 | 114.7504 | 27.5 | 0.54 |
| pHILIC | 202.082 | 48.2582 | 27.5 | 0.54 |
| pHILIC | 202.082 | 56.0603 | 27.5 | 0.54 |
| pHILIC | 202.082 | 215.6718 | 27.5 | 0.54 |
| pHILIC | 202.082 | 102.1334 | 27.5 | 0.54 |
| pHILIC | 202.082 | 47.8609 | 27.5 | 0.54 |
| pHILIC | 202.082 | 84.318 | 27.5 | 0.54 |
| pHILIC | 202.082 | 83.4198 | 27.5 | 0.54 |
| pHILIC | 202.082 | 65.0777 | 27.5 | 0.54 |
| pHILIC | 202.082 | 180.09 | 27.5 | 0.54 |
| pHILIC | 202.082 | 136.5471 | 27.4 | 0.54 |
| pHILIC | 202.082 | 87.4987 | 27.4 | 0.54 |
| pHILIC | 202.082 | 50.6562 | 27.4 | 0.54 |
| pHILIC | 202.082 | 131.097 | 27.4 | 0.54 |
| pHILIC | 202.082 | 159.7113 | 27.4 | 0.54 |
| pHILIC | 202.082 | 49.3716 | 27.4 | 0.54 |
| pHILIC | 202.082 | 89.7907 | 27.3 | 0.54 |
| pHILIC | 202.082 | 58.2285 | 27.3 | 0.54 |
| pHILIC | 202.082 | 90.2582 | 27.3 | 0.54 |
| pHILIC | 202.082 | 123.8321 | 27.3 | 0.54 |
| pHILIC | 202.082 | 68.4017 | 27.3 | 0.54 |
| pHILIC | 202.082 | 45.9472 | 27.3 | 0.54 |
| pHILIC | 202.082 | 116.0639 | 27.3 | 0.54 |
| pHILIC | 202.082 | 95.1481 | 27.3 | 0.54 |
| pHILIC | 202.082 | 60.0334 | 27.3 | 0.54 |
| pHILIC | 202.082 | 105.1304 | 27.3 | 0.54 |
| pHILIC | 202.082 | 67.9269 | 27.2 | 0.54 |
| pHILIC | 202.082 | 72.8008 | 27.2 | 0.54 |
| pHILIC | 202.082 | 56.4514 | 27.2 | 0.54 |
| pHILIC | 202.082 | 101.1946 | 27.2 | 0.54 |
| pHILIC | 202.082 | 62.8934 | 27.2 | 0.54 |
| pHILIC | 202.082 | 126.0345 | 27.2 | 0.54 |
| pHILIC | 202.082 | 51.7865 | 27.2 | 0.54 |
| pHILIC | 202.082 | 97.3985 | 27.1 | 0.54 |
| pHILIC | 202.082 | 214.2142 | 27.1 | 0.54 |
| pHILIC | 202.082 | 126.7022 | 27.1 | 0.53 |
| pHILIC | 202.082 | 47.9626 | 27.1 | 0.53 |
| pHILIC | 202.082 | 96.2898 | 27.1 | 0.53 |
| pHILIC | 202.082 | 81.8058 | 27.1 | 0.53 |
| pHILIC | 202.082 | 79.7254 | 27.1 | 0.53 |

| | | | | |
|---|---|---|---|---|
| pHILIC | 202.082 | 49.4413 | 27 | 0.53 |
| pHILIC | 202.082 | 67.0337 | 27 | 0.53 |
| pHILIC | 202.082 | 65.3276 | 27 | 0.53 |
| pHILIC | 202.082 | 130.3091 | 27 | 0.53 |
| pHILIC | 202.082 | 71.45 | 27 | 0.53 |
| pHILIC | 202.082 | 139.1401 | 27 | 0.53 |
| pHILIC | 202.082 | 50.0454 | 27 | 0.53 |
| pHILIC | 202.082 | 118.1389 | 27 | 0.53 |
| pHILIC | 202.082 | 75.622 | 27 | 0.53 |
| pHILIC | 202.082 | 70.977 | 27 | 0.53 |
| pHILIC | 202.082 | 66.7358 | 26.9 | 0.53 |
| pHILIC | 202.082 | 57.4169 | 26.9 | 0.53 |
| pHILIC | 202.082 | 188.7178 | 26.9 | 0.53 |
| pHILIC | 202.082 | 134.9358 | 26.9 | 0.53 |
| pHILIC | 202.082 | 138.7677 | 26.9 | 0.53 |
| pHILIC | 202.082 | 57.756 | 26.9 | 0.53 |
| pHILIC | 202.082 | 138.9675 | 26.9 | 0.53 |
| pHILIC | 202.082 | 55.6092 | 26.9 | 0.53 |
| pHILIC | 202.082 | 51.1267 | 26.9 | 0.53 |
| pHILIC | 202.082 | 183.7627 | 26.8 | 0.53 |
| pHILIC | 202.082 | 91.3993 | 26.8 | 0.53 |
| pHILIC | 202.082 | 220.9422 | 26.8 | 0.53 |
| pHILIC | 202.082 | 56.3199 | 26.8 | 0.53 |
| pHILIC | 202.082 | 114.7901 | 26.8 | 0.53 |
| pHILIC | 202.082 | 70.3042 | 26.7 | 0.53 |
| pHILIC | 202.082 | 51.9487 | 26.7 | 0.53 |
| pHILIC | 202.082 | 53.4065 | 26.7 | 0.53 |
| pHILIC | 202.082 | 65.6258 | 26.7 | 0.53 |
| pHILIC | 202.082 | 56.3255 | 26.7 | 0.53 |
| pHILIC | 202.082 | 213.7533 | 26.7 | 0.53 |
| pHILIC | 202.082 | 66.8332 | 26.7 | 0.53 |
| pHILIC | 202.082 | 76.076 | 26.7 | 0.53 |
| pHILIC | 202.082 | 213.9512 | 26.6 | 0.53 |
| pHILIC | 202.082 | 213.6866 | 26.6 | 0.53 |
| pHILIC | 202.082 | 50.3035 | 26.6 | 0.53 |
| pHILIC | 202.082 | 45.3782 | 26.6 | 0.53 |
| pHILIC | 202.082 | 201.251 | 26.6 | 0.53 |
| pHILIC | 202.082 | 67.8112 | 26.6 | 0.53 |
| pHILIC | 202.082 | 63.5958 | 26.6 | 0.53 |
| pHILIC | 202.082 | 184.1346 | 26.6 | 0.53 |
| pHILIC | 202.082 | 70.7252 | 26.6 | 0.53 |
| pHILIC | 202.082 | 51.6423 | 26.6 | 0.53 |

| | | | | |
|---|---|---|---|---|
| pHILIC | 202.082 | 65.1121 | 26.6 | 0.53 |
| pHILIC | 202.082 | 152.1668 | 26.6 | 0.52 |
| pHILIC | 202.082 | 51.58 | 26.6 | 0.52 |
| pHILIC | 202.082 | 72.2755 | 26.6 | 0.52 |
| pHILIC | 202.082 | 79.5815 | 26.6 | 0.52 |
| pHILIC | 202.082 | 191.4731 | 26.5 | 0.52 |
| pHILIC | 202.082 | 47.1836 | 26.5 | 0.52 |
| pHILIC | 202.082 | 69.3721 | 26.5 | 0.52 |
| pHILIC | 202.082 | 81.6918 | 26.5 | 0.52 |
| pHILIC | 202.082 | 62.6362 | 26.5 | 0.52 |
| pHILIC | 202.082 | 47.8501 | 26.5 | 0.52 |
| pHILIC | 202.082 | 129.4872 | 26.5 | 0.52 |
| pHILIC | 202.082 | 159.764 | 26.5 | 0.52 |
| pHILIC | 202.082 | 63.8213 | 26.5 | 0.52 |
| pHILIC | 202.082 | 62.0904 | 26.5 | 0.52 |
| pHILIC | 202.082 | 126.0002 | 26.5 | 0.52 |
| pHILIC | 202.082 | 51.9599 | 26.5 | 0.52 |
| pHILIC | 202.082 | 81.1975 | 26.4 | 0.52 |
| pHILIC | 202.082 | 68.0398 | 26.4 | 0.52 |
| pHILIC | 202.082 | 96.2961 | 26.4 | 0.52 |
| pHILIC | 202.082 | 63.6976 | 26.4 | 0.52 |
| pHILIC | 202.082 | 88.6849 | 26.4 | 0.52 |
| pHILIC | 202.082 | 52.9571 | 26.4 | 0.52 |
| pHILIC | 202.082 | 54.6199 | 26.4 | 0.52 |
| pHILIC | 202.082 | 132.0794 | 26.4 | 0.52 |
| pHILIC | 202.082 | 78.3032 | 26.4 | 0.52 |
| pHILIC | 202.082 | 122.9888 | 26.4 | 0.52 |
| pHILIC | 202.082 | 102.978 | 26.4 | 0.52 |
| pHILIC | 202.082 | 47.9295 | 26.4 | 0.52 |
| pHILIC | 202.082 | 112.743 | 26.4 | 0.52 |
| pHILIC | 202.082 | 91.4868 | 26.4 | 0.52 |
| pHILIC | 202.082 | 50.5045 | 26.3 | 0.52 |
| pHILIC | 202.082 | 67.0477 | 26.3 | 0.52 |
| pHILIC | 202.082 | 46.9601 | 26.3 | 0.52 |
| pHILIC | 202.082 | 163.9949 | 26.3 | 0.52 |
| pHILIC | 202.082 | 60.9263 | 26.3 | 0.52 |
| pHILIC | 202.082 | 66.584 | 26.3 | 0.52 |
| pHILIC | 202.082 | 49.5649 | 26.3 | 0.52 |
| pHILIC | 202.082 | 145.4487 | 26.3 | 0.52 |
| pHILIC | 202.082 | 153.8287 | 26.3 | 0.52 |
| pHILIC | 202.082 | 59.4914 | 26.3 | 0.52 |
| pHILIC | 202.082 | 48.3801 | 26.3 | 0.52 |

| | | | | |
|---|---|---|---|---|
| pHILIC | 202.082 | 49.8679 | 26.3 | 0.52 |
| pHILIC | 202.082 | 188.3698 | 26.3 | 0.52 |
| pHILIC | 202.082 | 58.4762 | 26.3 | 0.52 |
| pHILIC | 202.082 | 149.5085 | 26.3 | 0.52 |
| pHILIC | 202.082 | 145.5373 | 26.3 | 0.52 |
| pHILIC | 202.082 | 149.5714 | 26.3 | 0.52 |
| pHILIC | 202.082 | 67.415 | 26.3 | 0.52 |
| pHILIC | 202.082 | 46.5422 | 26.2 | 0.52 |
| pHILIC | 202.082 | 50.1257 | 26.2 | 0.52 |
| pHILIC | 202.082 | 48.9357 | 26.2 | 0.52 |
| pHILIC | 202.082 | 53.1851 | 26.2 | 0.52 |
| pHILIC | 202.082 | 187.6542 | 26.2 | 0.52 |
| pHILIC | 202.082 | 71.367 | 26.2 | 0.52 |
| pHILIC | 202.082 | 67.3172 | 26.2 | 0.52 |
| pHILIC | 202.082 | 72.5437 | 26.2 | 0.52 |
| pHILIC | 202.082 | 87.8342 | 26.2 | 0.52 |
| pHILIC | 202.082 | 64.034 | 26.2 | 0.52 |
| pHILIC | 202.082 | 97.4796 | 26.2 | 0.52 |
| pHILIC | 202.082 | 62.8282 | 26.2 | 0.52 |
| pHILIC | 202.082 | 51.8301 | 26.2 | 0.52 |
| pHILIC | 202.082 | 75.9524 | 26.2 | 0.52 |
| pHILIC | 202.082 | 198.7499 | 26.2 | 0.52 |
| pHILIC | 202.082 | 205.4041 | 26.2 | 0.52 |
| pHILIC | 202.082 | 174.4241 | 26.1 | 0.52 |
| pHILIC | 202.082 | 114.4877 | 26.1 | 0.52 |
| pHILIC | 202.082 | 48.4753 | 26.1 | 0.52 |
| pHILIC | 202.082 | 123.9548 | 26.1 | 0.52 |
| pHILIC | 202.082 | 73.3382 | 26.1 | 0.52 |
| pHILIC | 202.082 | 116.2031 | 26.1 | 0.52 |
| pHILIC | 202.082 | 173.6736 | 26.1 | 0.52 |
| pHILIC | 202.082 | 58.5373 | 26.1 | 0.52 |
| pHILIC | 202.082 | 96.7837 | 26.1 | 0.52 |
| pHILIC | 202.082 | 65.1674 | 26.1 | 0.52 |
| pHILIC | 202.082 | 86.5921 | 26.1 | 0.52 |
| pHILIC | 202.082 | 146.0907 | 26.1 | 0.51 |
| pHILIC | 202.082 | 77.0629 | 26.1 | 0.51 |
| pHILIC | 202.082 | 102.0084 | 26.1 | 0.51 |
| pHILIC | 202.082 | 49.4714 | 26.1 | 0.51 |
| pHILIC | 202.082 | 95.3604 | 26.1 | 0.51 |
| pHILIC | 202.082 | 142.0511 | 26 | 0.51 |
| pHILIC | 202.082 | 51.6674 | 26 | 0.51 |
| pHILIC | 202.082 | 53.226 | 26 | 0.51 |

| | | | | |
|---|---|---|---|---|
| pHILIC | 202.082 | 96.4456 | 26 | 0.51 |
| pHILIC | 202.082 | 55.2765 | 26 | 0.51 |
| pHILIC | 202.082 | 75.1795 | 26 | 0.51 |
| pHILIC | 202.082 | 153.7503 | 26 | 0.51 |
| pHILIC | 202.082 | 74.028 | 26 | 0.51 |
| pHILIC | 202.082 | 98.8195 | 26 | 0.51 |
| pHILIC | 202.082 | 149.2965 | 26 | 0.51 |
| pHILIC | 202.082 | 51.9517 | 26 | 0.51 |
| pHILIC | 202.082 | 87.206 | 26 | 0.51 |
| pHILIC | 202.082 | 64.148 | 26 | 0.51 |
| pHILIC | 202.082 | 85.4351 | 26 | 0.51 |
| pHILIC | 202.082 | 66.0925 | 26 | 0.51 |
| pHILIC | 202.082 | 171.4355 | 26 | 0.51 |
| pHILIC | 202.082 | 72.319 | 26 | 0.51 |
| pHILIC | 202.082 | 77.3737 | 26 | 0.51 |
| pHILIC | 202.082 | 49.5602 | 25.9 | 0.51 |
| pHILIC | 202.082 | 57.0147 | 25.9 | 0.51 |
| pHILIC | 202.082 | 66.9284 | 25.9 | 0.51 |
| pHILIC | 202.082 | 61.6646 | 25.9 | 0.51 |
| pHILIC | 202.082 | 51.7152 | 25.9 | 0.51 |
| pHILIC | 202.082 | 109.6946 | 25.9 | 0.51 |
| pHILIC | 202.082 | 116.0514 | 25.9 | 0.51 |
| pHILIC | 202.082 | 104.3754 | 25.9 | 0.51 |
| pHILIC | 202.082 | 158.7663 | 25.9 | 0.51 |
| pHILIC | 202.082 | 49.6081 | 25.9 | 0.51 |
| pHILIC | 202.082 | 131.6202 | 25.9 | 0.51 |
| pHILIC | 202.082 | 54.4055 | 25.9 | 0.51 |
| pHILIC | 202.082 | 148.2166 | 25.9 | 0.51 |
| pHILIC | 202.082 | 140.6181 | 25.9 | 0.51 |
| pHILIC | 202.082 | 48.5663 | 25.9 | 0.51 |
| pHILIC | 202.082 | 51.5415 | 25.9 | 0.51 |
| pHILIC | 202.082 | 105.8418 | 25.9 | 0.51 |
| pHILIC | 202.082 | 68.7484 | 25.9 | 0.51 |
| pHILIC | 202.082 | 126.7282 | 25.9 | 0.51 |
| pHILIC | 202.082 | 139.2943 | 25.9 | 0.51 |
| pHILIC | 202.082 | 52.3825 | 25.8 | 0.51 |
| pHILIC | 202.082 | 87.0569 | 25.8 | 0.51 |
| pHILIC | 202.082 | 73.8073 | 25.8 | 0.51 |
| pHILIC | 202.082 | 46.1763 | 25.8 | 0.51 |
| pHILIC | 202.082 | 70.3211 | 25.8 | 0.51 |
| pHILIC | 202.082 | 52.5363 | 25.8 | 0.51 |
| pHILIC | 202.082 | 51.757 | 25.8 | 0.51 |

| | | | | |
|---|---|---|---|---|
| pHILIC | 202.082 | 84.3336 | 25.8 | 0.51 |
| pHILIC | 202.082 | 84.581 | 25.8 | 0.51 |
| pHILIC | 202.082 | 70.2169 | 25.8 | 0.51 |
| pHILIC | 202.082 | 51.7252 | 25.8 | 0.51 |
| pHILIC | 202.082 | 57.9918 | 25.8 | 0.51 |
| pHILIC | 202.082 | 55.9951 | 25.8 | 0.51 |
| pHILIC | 202.082 | 61.2467 | 25.8 | 0.51 |
| pHILIC | 202.082 | 157.386 | 25.8 | 0.51 |
| pHILIC | 202.082 | 94.1306 | 25.8 | 0.51 |
| pHILIC | 202.082 | 68.4495 | 25.8 | 0.51 |
| pHILIC | 202.082 | 55.103 | 25.8 | 0.51 |
| pHILIC | 202.082 | 68.4778 | 25.8 | 0.51 |
| pHILIC | 202.082 | 98.3365 | 25.8 | 0.51 |
| pHILIC | 202.082 | 70.3834 | 25.8 | 0.51 |
| pHILIC | 202.082 | 215.5941 | 25.8 | 0.51 |
| pHILIC | 202.082 | 116.5222 | 25.7 | 0.51 |
| pHILIC | 202.082 | 70.0468 | 25.7 | 0.51 |
| pHILIC | 202.082 | 177.6656 | 25.7 | 0.51 |
| pHILIC | 202.082 | 94.9228 | 25.7 | 0.51 |
| pHILIC | 202.082 | 46.5598 | 25.7 | 0.51 |
| pHILIC | 202.082 | 212.3328 | 25.7 | 0.51 |
| pHILIC | 202.082 | 143.6853 | 25.7 | 0.51 |
| pHILIC | 202.082 | 139.5927 | 25.7 | 0.51 |
| pHILIC | 202.082 | 80.8268 | 25.7 | 0.51 |
| pHILIC | 202.082 | 50.1358 | 25.7 | 0.51 |
| pHILIC | 202.082 | 174.5817 | 25.7 | 0.51 |
| pHILIC | 202.082 | 76.5136 | 25.7 | 0.51 |
| pHILIC | 202.082 | 191.2944 | 25.7 | 0.51 |
| pHILIC | 202.082 | 69.8981 | 25.7 | 0.51 |
| pHILIC | 202.082 | 56.8503 | 25.7 | 0.51 |
| pHILIC | 202.082 | 148.7137 | 25.7 | 0.51 |
| pHILIC | 202.082 | 204.9989 | 25.6 | 0.51 |
| pHILIC | 202.082 | 185.9223 | 25.6 | 0.51 |
| pHILIC | 202.082 | 111.9266 | 25.6 | 0.51 |
| pHILIC | 202.082 | 177.1722 | 25.6 | 0.51 |
| pHILIC | 202.082 | 127.5109 | 25.6 | 0.51 |
| pHILIC | 202.082 | 51.2456 | 25.6 | 0.51 |
| pHILIC | 202.082 | 135.7372 | 25.6 | 0.51 |
| pHILIC | 202.082 | 120.4374 | 25.6 | 0.51 |
| pHILIC | 202.082 | 224.2603 | 25.6 | 0.51 |
| pHILIC | 202.082 | 119.8952 | 25.6 | 0.51 |
| pHILIC | 202.082 | 45.9286 | 25.6 | 0.51 |

| | | | | |
|---|---|---|---|---|
| pHILIC | 202.082 | 52.1739 | 25.6 | 0.51 |
| pHILIC | 202.082 | 97.0157 | 25.6 | 0.51 |
| pHILIC | 202.082 | 200.6981 | 25.6 | 0.51 |
| pHILIC | 202.082 | 161.139 | 25.6 | 0.51 |
| pHILIC | 202.082 | 148.297 | 25.6 | 0.51 |
| pHILIC | 202.082 | 103.9128 | 25.6 | 0.5 |
| pHILIC | 202.082 | 110.6998 | 25.6 | 0.5 |
| pHILIC | 202.082 | 59.4236 | 25.5 | 0.5 |
| pHILIC | 202.082 | 66.9886 | 25.5 | 0.5 |
| pHILIC | 202.082 | 61.5778 | 25.5 | 0.5 |
| pHILIC | 202.082 | 100.6347 | 25.5 | 0.5 |
| pHILIC | 202.082 | 53.8562 | 25.5 | 0.5 |
| pHILIC | 202.082 | 130.8237 | 25.5 | 0.5 |
| pHILIC | 202.082 | 46.8263 | 25.5 | 0.5 |
| pHILIC | 202.082 | 73.971 | 25.5 | 0.5 |
| pHILIC | 202.082 | 48.2286 | 25.5 | 0.5 |
| pHILIC | 202.082 | 159.2882 | 25.5 | 0.5 |
| pHILIC | 202.082 | 175.7833 | 25.5 | 0.5 |
| pHILIC | 202.082 | 120.7017 | 25.5 | 0.5 |
| pHILIC | 202.082 | 205.9377 | 25.5 | 0.5 |
| pHILIC | 202.082 | 183.9985 | 25.5 | 0.5 |
| pHILIC | 202.082 | 62.2732 | 25.5 | 0.5 |
| pHILIC | 202.082 | 141.1122 | 25.5 | 0.5 |
| pHILIC | 202.082 | 76.8103 | 25.5 | 0.5 |
| pHILIC | 202.082 | 65.6125 | 25.5 | 0.5 |
| pHILIC | 202.082 | 120.3105 | 25.5 | 0.5 |
| pHILIC | 202.082 | 79.64 | 25.4 | 0.5 |
| pHILIC | 202.082 | 51.8526 | 25.4 | 0.5 |
| pHILIC | 202.082 | 51.3474 | 25.4 | 0.5 |
| pHILIC | 202.082 | 105.3729 | 25.4 | 0.5 |
| pHILIC | 202.082 | 71.64 | 25.4 | 0.5 |
| pHILIC | 202.082 | 185.1453 | 25.4 | 0.5 |
| pHILIC | 202.082 | 171.5005 | 25.4 | 0.5 |
| pHILIC | 202.082 | 65.6593 | 25.4 | 0.5 |
| pHILIC | 202.082 | 137.3278 | 25.4 | 0.5 |
| pHILIC | 202.082 | 81.8592 | 25.4 | 0.5 |
| pHILIC | 202.082 | 111.7132 | 25.4 | 0.5 |
| pHILIC | 202.082 | 222.215 | 25.4 | 0.5 |
| pHILIC | 202.082 | 149.0117 | 25.4 | 0.5 |
| pHILIC | 202.082 | 57.5655 | 25.4 | 0.5 |
| pHILIC | 202.082 | 107.2487 | 25.3 | 0.5 |
| pHILIC | 202.082 | 69.1258 | 25.3 | 0.5 |

| | | | | |
|---|---|---|---|---|
| pHILIC | 202.082 | 76.1961 | 25.3 | 0.5 |
| pHILIC | 202.082 | 77.0543 | 25.3 | 0.5 |
| pHILIC | 202.082 | 79.5579 | 25.3 | 0.5 |
| pHILIC | 202.082 | 131.4245 | 25.3 | 0.5 |
| pHILIC | 202.082 | 133.9074 | 25.3 | 0.5 |
| pHILIC | 202.082 | 52.4513 | 25.3 | 0.5 |
| pHILIC | 202.082 | 79.5079 | 25.3 | 0.5 |
| pHILIC | 202.082 | 74.501 | 25.3 | 0.5 |
| pHILIC | 202.082 | 55.9113 | 25.3 | 0.5 |
| pHILIC | 202.082 | 49.701 | 25.3 | 0.5 |
| pHILIC | 202.082 | 119.1602 | 25.3 | 0.5 |
| pHILIC | 202.082 | 50.183 | 25.3 | 0.5 |
| pHILIC | 202.082 | 46.8757 | 25.3 | 0.5 |
| pHILIC | 202.082 | 117.4411 | 25.3 | 0.5 |
| pHILIC | 202.082 | 59.8867 | 25.3 | 0.5 |
| pHILIC | 202.082 | 132.9587 | 25.3 | 0.5 |
| pHILIC | 202.082 | 66.2884 | 25.3 | 0.5 |
| pHILIC | 202.082 | 107.3264 | 25.3 | 0.5 |
| pHILIC | 202.082 | 144.7034 | 25.3 | 0.5 |
| pHILIC | 202.082 | 109.9387 | 25.3 | 0.5 |
| pHILIC | 202.082 | 64.6408 | 25.3 | 0.5 |
| pHILIC | 202.082 | 134.3152 | 25.3 | 0.5 |
| pHILIC | 202.082 | 216.0782 | 25.3 | 0.5 |
| pHILIC | 202.082 | 46.6633 | 25.2 | 0.5 |
| pHILIC | 202.082 | 129.3558 | 25.2 | 0.5 |
| pHILIC | 202.082 | 73.9154 | 25.2 | 0.5 |
| pHILIC | 202.082 | 163.2361 | 25.2 | 0.5 |
| pHILIC | 202.082 | 82.9812 | 25.2 | 0.5 |
| pHILIC | 202.082 | 51.6498 | 25.2 | 0.5 |
| pHILIC | 202.082 | 60.4789 | 25.2 | 0.5 |
| pHILIC | 202.082 | 84.9991 | 25.2 | 0.5 |
| pHILIC | 202.082 | 72.3771 | 25.2 | 0.5 |
| pHILIC | 202.082 | 47.3643 | 25.2 | 0.5 |
| pHILIC | 202.082 | 213.1953 | 25.2 | 0.5 |
| pHILIC | 202.082 | 45.6068 | 25.2 | 0.5 |
| pHILIC | 202.082 | 207.6942 | 25.2 | 0.5 |
| pHILIC | 202.082 | 176.3987 | 25.2 | 0.5 |
| pHILIC | 202.082 | 102.4003 | 25.2 | 0.5 |
| pHILIC | 202.082 | 102.121 | 25.2 | 0.5 |
| pHILIC | 202.082 | 52.5533 | 25.2 | 0.5 |
| pHILIC | 202.082 | 206.1518 | 25.2 | 0.5 |
| pHILIC | 202.082 | 103.4968 | 25.2 | 0.5 |

| | | | | |
|---|---|---|---|---|
| pHILIC | 202.082 | 98.4818 | 25.2 | 0.5 |
| pHILIC | 202.082 | 91.8554 | 25.2 | 0.5 |
| pHILIC | 202.082 | 96.0974 | 25.2 | 0.5 |
| pHILIC | 202.082 | 74.4634 | 25.2 | 0.5 |
| pHILIC | 202.082 | 54.7252 | 25.2 | 0.5 |
| pHILIC | 202.082 | 97.0845 | 25.1 | 0.5 |
| pHILIC | 202.082 | 51.5081 | 25.1 | 0.5 |
| pHILIC | 202.082 | 49.5883 | 25.1 | 0.5 |
| pHILIC | 202.082 | 120.5872 | 25.1 | 0.5 |
| pHILIC | 202.082 | 71.1976 | 25.1 | 0.5 |
| pHILIC | 202.082 | 63.5408 | 25.1 | 0.5 |
| pHILIC | 202.082 | 157.8437 | 25.1 | 0.5 |
| pHILIC | 202.082 | 158.9433 | 25.1 | 0.5 |
| pHILIC | 202.082 | 52.134 | 25.1 | 0.5 |
| pHILIC | 202.082 | 60.0814 | 25.1 | 0.5 |
| pHILIC | 202.082 | 75.8958 | 25.1 | 0.5 |
| pHILIC | 202.082 | 122.6998 | 25.1 | 0.5 |
| pHILIC | 202.082 | 79.1831 | 25.1 | 0.5 |
| pHILIC | 202.082 | 77.0032 | 25.1 | 0.5 |
| pHILIC | 202.082 | 108.6263 | 25.1 | 0.5 |
| pHILIC | 202.082 | 46.4402 | 25.1 | 0.5 |
| pHILIC | 202.082 | 169.524 | 25.1 | 0.5 |
| pHILIC | 202.082 | 57.2033 | 25.1 | 0.5 |
| pHILIC | 202.082 | 49.0299 | 25.1 | 0.5 |
| pHILIC | 202.082 | 107.4697 | 25.1 | 0.5 |
| pHILIC | 202.082 | 47.7189 | 25.1 | 0.5 |
| pHILIC | 202.082 | 102.1005 | 25.1 | 0.5 |
| pHILIC | 202.082 | 46.4544 | 25.1 | 0.5 |
| pHILIC | 202.082 | 59.2049 | 25.1 | 0.5 |
| pHILIC | 202.082 | 80.8926 | 25.1 | 0.5 |
| pHILIC | 202.082 | 178.1721 | 25.1 | 0.5 |
| pHILIC | 202.082 | 59.5261 | 25.1 | 0.5 |
| pHILIC | 202.082 | 56.0748 | 25.1 | 0.5 |
| pHILIC | 202.082 | 55.6686 | 25.1 | 0.5 |
| pHILIC | 202.082 | 136.5739 | 25.1 | 0.5 |
| pHILIC | 202.082 | 130.0985 | 25.1 | 0.5 |
| pHILIC | 202.082 | 74.4458 | 25.1 | 0.5 |
| pHILIC | 202.082 | 61.5366 | 25.1 | 0.49 |
| pHILIC | 202.082 | 207.9372 | 25.1 | 0.49 |
| pHILIC | 202.082 | 53.4277 | 25.1 | 0.49 |
| pHILIC | 202.082 | 56.6357 | 25.1 | 0.49 |
| pHILIC | 202.082 | 112.0191 | 25 | 0.49 |

| | | | | |
|---|---|---|---|---|
| pHILIC | 202.082 | 59.7603 | 25 | 0.49 |
| pHILIC | 202.082 | 52.2715 | 25 | 0.49 |
| pHILIC | 202.082 | 64.1832 | 25 | 0.49 |
| pHILIC | 202.082 | 176.0812 | 25 | 0.49 |
| pHILIC | 202.082 | 48.4465 | 25 | 0.49 |
| pHILIC | 202.082 | 109.9162 | 25 | 0.49 |
| pHILIC | 202.082 | 177.3835 | 25 | 0.49 |
| pHILIC | 202.082 | 113.1325 | 25 | 0.49 |
| pHILIC | 202.082 | 221.5624 | 25 | 0.49 |
| pHILIC | 202.082 | 224.8448 | 25 | 0.49 |
| pHILIC | 202.082 | 157.7101 | 25 | 0.49 |
| pHILIC | 202.082 | 65.6858 | 25 | 0.49 |
| pHILIC | 202.082 | 87.4626 | 25 | 0.49 |
| pHILIC | 202.082 | 165.5654 | 25 | 0.49 |
| pHILIC | 202.082 | 130.488 | 25 | 0.49 |
| pHILIC | 202.082 | 148.4729 | 25 | 0.49 |
| pHILIC | 202.082 | 129.7583 | 25 | 0.49 |
| pHILIC | 202.082 | 47.1558 | 25 | 0.49 |
| pHILIC | 202.082 | 73.6756 | 25 | 0.49 |
| pHILIC | 202.082 | 161.407 | 25 | 0.49 |
| pHILIC | 202.082 | 138.0314 | 25 | 0.49 |
| pHILIC | 202.082 | 77.1884 | 25 | 0.49 |
| pHILIC | 202.082 | 65.7058 | 24.9 | 0.49 |
| pHILIC | 202.082 | 46.6499 | 24.9 | 0.49 |
| pHILIC | 202.082 | 78.3101 | 24.9 | 0.49 |
| pHILIC | 202.082 | 102.2834 | 24.9 | 0.49 |
| pHILIC | 202.082 | 66.3705 | 24.9 | 0.49 |
| pHILIC | 202.082 | 89.6549 | 24.9 | 0.49 |
| pHILIC | 202.082 | 103.2963 | 24.9 | 0.49 |
| pHILIC | 202.082 | 91.6387 | 24.9 | 0.49 |
| pHILIC | 202.082 | 83.1593 | 24.9 | 0.49 |
| pHILIC | 202.082 | 51.0718 | 24.9 | 0.49 |
| pHILIC | 202.082 | 147.3007 | 24.9 | 0.49 |
| pHILIC | 202.082 | 168.3468 | 24.9 | 0.49 |
| pHILIC | 202.082 | 210.9828 | 24.9 | 0.49 |
| pHILIC | 202.082 | 57.4461 | 24.9 | 0.49 |
| pHILIC | 202.082 | 209.1519 | 24.9 | 0.49 |
| pHILIC | 202.082 | 51.8348 | 24.9 | 0.49 |
| pHILIC | 202.082 | 56.2608 | 24.9 | 0.49 |
| pHILIC | 202.082 | 172.4848 | 24.9 | 0.49 |
| pHILIC | 202.082 | 123.7671 | 24.9 | 0.49 |
| pHILIC | 202.082 | 216.5117 | 24.9 | 0.49 |

| | | | | |
|---|---|---|---|---|
| pHILIC | 202.082 | 58.3282 | 24.9 | 0.49 |
| pHILIC | 202.082 | 184.7716 | 24.9 | 0.49 |
| pHILIC | 202.082 | 45.7576 | 24.9 | 0.49 |
| pHILIC | 202.082 | 58.2719 | 24.9 | 0.49 |
| pHILIC | 202.082 | 117.4635 | 24.9 | 0.49 |
| pHILIC | 202.082 | 71.3282 | 24.9 | 0.49 |
| pHILIC | 202.082 | 83.5732 | 24.9 | 0.49 |
| pHILIC | 202.082 | 109.084 | 24.9 | 0.49 |
| pHILIC | 202.082 | 125.4208 | 24.8 | 0.49 |
| pHILIC | 202.082 | 136.7802 | 24.8 | 0.49 |
| pHILIC | 202.082 | 223.7738 | 24.8 | 0.49 |
| pHILIC | 202.082 | 90.3663 | 24.8 | 0.49 |
| pHILIC | 202.082 | 221.0543 | 24.8 | 0.49 |
| pHILIC | 202.082 | 90.4587 | 24.8 | 0.49 |
| pHILIC | 202.082 | 178.0216 | 24.8 | 0.49 |
| pHILIC | 202.082 | 131.7665 | 24.8 | 0.49 |
| pHILIC | 202.082 | 108.7431 | 24.8 | 0.49 |
| pHILIC | 202.082 | 46.5834 | 24.8 | 0.49 |
| pHILIC | 202.082 | 53.4817 | 24.8 | 0.49 |
| pHILIC | 202.082 | 131.9422 | 24.8 | 0.49 |
| pHILIC | 202.082 | 57.0547 | 24.8 | 0.49 |
| pHILIC | 202.082 | 85.6858 | 24.8 | 0.49 |
| pHILIC | 202.082 | 179.6425 | 24.8 | 0.49 |
| pHILIC | 202.082 | 159.9231 | 24.8 | 0.49 |
| pHILIC | 202.082 | 67.8943 | 24.8 | 0.49 |
| pHILIC | 202.082 | 146.2199 | 24.8 | 0.49 |
| pHILIC | 202.082 | 103.7242 | 24.8 | 0.49 |
| pHILIC | 202.082 | 97.4679 | 24.8 | 0.49 |
| pHILIC | 202.082 | 88.5168 | 24.8 | 0.49 |
| pHILIC | 202.082 | 149.4849 | 24.8 | 0.49 |
| pHILIC | 202.082 | 86.4217 | 24.8 | 0.49 |
| pHILIC | 202.082 | 53.939 | 24.8 | 0.49 |
| pHILIC | 202.082 | 95.0099 | 24.8 | 0.49 |
| pHILIC | 202.082 | 179.366 | 24.8 | 0.49 |
| pHILIC | 202.082 | 73.3882 | 24.8 | 0.49 |
| pHILIC | 202.082 | 131.8873 | 24.8 | 0.49 |
| pHILIC | 202.082 | 90.4651 | 24.8 | 0.49 |
| pHILIC | 202.082 | 76.9335 | 24.7 | 0.49 |
| pHILIC | 202.082 | 217.0961 | 24.7 | 0.49 |
| pHILIC | 202.082 | 67.9791 | 24.7 | 0.49 |
| pHILIC | 202.082 | 165.2768 | 24.7 | 0.49 |
| pHILIC | 202.082 | 48.5446 | 24.7 | 0.49 |

| | | | | |
|---|---|---|---|---|
| pHILIC | 202.082 | 64.376 | 24.7 | 0.49 |
| pHILIC | 202.082 | 150.0857 | 24.7 | 0.49 |
| pHILIC | 202.082 | 133.8795 | 24.7 | 0.49 |
| pHILIC | 202.082 | 89.1065 | 24.7 | 0.49 |
| pHILIC | 202.082 | 189.5308 | 24.7 | 0.49 |
| pHILIC | 202.082 | 85.6472 | 24.7 | 0.49 |
| pHILIC | 202.082 | 48.8071 | 24.7 | 0.49 |
| pHILIC | 202.082 | 99.6394 | 24.7 | 0.49 |
| pHILIC | 202.082 | 146.9827 | 24.7 | 0.49 |
| pHILIC | 202.082 | 55.2587 | 24.7 | 0.49 |
| pHILIC | 202.082 | 79.7635 | 24.7 | 0.49 |
| pHILIC | 202.082 | 82.7138 | 24.7 | 0.49 |
| pHILIC | 202.082 | 198.6541 | 24.7 | 0.49 |
| pHILIC | 202.082 | 49.7219 | 24.7 | 0.49 |
| pHILIC | 202.082 | 90.7395 | 24.7 | 0.49 |
| pHILIC | 202.082 | 46.7455 | 24.7 | 0.49 |
| pHILIC | 202.082 | 114.287 | 24.7 | 0.49 |
| pHILIC | 202.082 | 159.2175 | 24.7 | 0.49 |
| pHILIC | 202.082 | 46.4605 | 24.7 | 0.49 |
| pHILIC | 202.082 | 88.9978 | 24.7 | 0.49 |
| pHILIC | 202.082 | 61.6162 | 24.7 | 0.49 |
| pHILIC | 202.082 | 159.0933 | 24.7 | 0.49 |
| pHILIC | 202.082 | 196.4043 | 24.7 | 0.49 |
| pHILIC | 202.082 | 63.2345 | 24.7 | 0.49 |
| pHILIC | 202.082 | 122.4481 | 24.7 | 0.49 |
| pHILIC | 202.082 | 161.2107 | 24.7 | 0.49 |
| pHILIC | 202.082 | 180.1244 | 24.6 | 0.49 |
| pHILIC | 202.082 | 152.9773 | 24.6 | 0.49 |
| pHILIC | 202.082 | 95.2924 | 24.6 | 0.49 |
| pHILIC | 202.082 | 56.0848 | 24.6 | 0.49 |
| pHILIC | 202.082 | 148.9779 | 24.6 | 0.49 |
| pHILIC | 202.082 | 52.7602 | 24.6 | 0.49 |
| pHILIC | 202.082 | 55.989 | 24.6 | 0.49 |
| pHILIC | 202.082 | 112.2236 | 24.6 | 0.49 |
| pHILIC | 202.082 | 98.9486 | 24.6 | 0.49 |
| pHILIC | 202.082 | 69.0871 | 24.6 | 0.49 |
| pHILIC | 202.082 | 96.3075 | 24.6 | 0.49 |
| pHILIC | 202.082 | 121.2499 | 24.6 | 0.49 |
| pHILIC | 202.082 | 80.734 | 24.6 | 0.49 |
| pHILIC | 202.082 | 128.5355 | 24.6 | 0.49 |
| pHILIC | 202.082 | 93.7053 | 24.6 | 0.49 |
| pHILIC | 202.082 | 200.5421 | 24.6 | 0.49 |

| | | | | |
|---|---|---|---|---|
| pHILIC | 202.082 | 208.9074 | 24.6 | 0.49 |
| pHILIC | 202.082 | 195.7977 | 24.6 | 0.49 |
| pHILIC | 202.082 | 117.9984 | 24.6 | 0.49 |
| pHILIC | 202.082 | 71.0183 | 24.6 | 0.49 |
| pHILIC | 202.082 | 78.9344 | 24.6 | 0.49 |
| pHILIC | 202.082 | 62.6968 | 24.6 | 0.49 |
| pHILIC | 202.082 | 55.9236 | 24.6 | 0.49 |
| pHILIC | 202.082 | 49.6635 | 24.6 | 0.49 |
| pHILIC | 202.082 | 129.8887 | 24.6 | 0.49 |
| pHILIC | 202.082 | 64.3726 | 24.6 | 0.49 |
| pHILIC | 202.082 | 110.0848 | 24.6 | 0.49 |
| pHILIC | 202.082 | 56.2003 | 24.6 | 0.49 |
| pHILIC | 202.082 | 66.2941 | 24.6 | 0.49 |
| pHILIC | 202.082 | 50.4774 | 24.6 | 0.49 |
| pHILIC | 202.082 | 125.2403 | 24.6 | 0.49 |
| pHILIC | 202.082 | 77.7399 | 24.6 | 0.49 |
| pHILIC | 202.082 | 59.3415 | 24.6 | 0.49 |
| pHILIC | 202.082 | 131.5516 | 24.5 | 0.48 |
| pHILIC | 202.082 | 53.2891 | 24.5 | 0.48 |
| pHILIC | 202.082 | 62.7088 | 24.5 | 0.48 |
| pHILIC | 202.082 | 116.8193 | 24.5 | 0.48 |
| pHILIC | 202.082 | 176.4842 | 24.5 | 0.48 |
| pHILIC | 202.082 | 81.5427 | 24.5 | 0.48 |
| pHILIC | 202.082 | 48.3088 | 24.5 | 0.48 |
| pHILIC | 202.082 | 145.9311 | 24.5 | 0.48 |
| pHILIC | 202.082 | 59.3522 | 24.5 | 0.48 |
| pHILIC | 202.082 | 199.1557 | 24.5 | 0.48 |
| pHILIC | 202.082 | 144.651 | 24.5 | 0.48 |
| pHILIC | 202.082 | 120.6201 | 24.5 | 0.48 |
| pHILIC | 202.082 | 79.718 | 24.5 | 0.48 |
| pHILIC | 202.082 | 128.4431 | 24.5 | 0.48 |
| pHILIC | 202.082 | 103.9987 | 24.5 | 0.48 |
| pHILIC | 202.082 | 176.8827 | 24.5 | 0.48 |
| pHILIC | 202.082 | 173.4737 | 24.5 | 0.48 |
| pHILIC | 202.082 | 75.1582 | 24.5 | 0.48 |
| pHILIC | 202.082 | 46.7941 | 24.5 | 0.48 |
| pHILIC | 202.082 | 87.8549 | 24.5 | 0.48 |
| pHILIC | 202.082 | 86.3008 | 24.5 | 0.48 |
| pHILIC | 202.082 | 132.5489 | 24.5 | 0.48 |
| pHILIC | 202.082 | 55.4617 | 24.5 | 0.48 |
| pHILIC | 202.082 | 50.3939 | 24.5 | 0.48 |
| pHILIC | 202.082 | 79.8677 | 24.5 | 0.48 |

| | | | | |
|---|---|---|---|---|
| pHILIC | 202.082 | 63.9569 | 24.4 | 0.48 |
| pHILIC | 202.082 | 147.1113 | 24.4 | 0.48 |
| pHILIC | 202.082 | 72.486 | 24.4 | 0.48 |
| pHILIC | 202.082 | 57.9481 | 24.4 | 0.48 |
| pHILIC | 202.082 | 50.9875 | 24.4 | 0.48 |
| pHILIC | 202.082 | 224.1566 | 24.4 | 0.48 |
| pHILIC | 202.082 | 90.0444 | 24.4 | 0.48 |
| pHILIC | 202.082 | 50.9099 | 24.4 | 0.48 |
| pHILIC | 202.082 | 49.0721 | 24.4 | 0.48 |
| pHILIC | 202.082 | 139.8939 | 24.4 | 0.48 |
| pHILIC | 202.082 | 49.2418 | 24.4 | 0.48 |
| pHILIC | 202.082 | 96.978 | 24.4 | 0.48 |
| pHILIC | 202.082 | 63.8243 | 24.4 | 0.48 |
| pHILIC | 202.082 | 193.2307 | 24.4 | 0.48 |
| pHILIC | 202.082 | 55.3904 | 24.4 | 0.48 |
| pHILIC | 202.082 | 81.5358 | 24.4 | 0.48 |
| pHILIC | 202.082 | 60.8908 | 24.4 | 0.48 |
| pHILIC | 202.082 | 183.4553 | 24.4 | 0.48 |
| pHILIC | 202.082 | 207.2622 | 24.4 | 0.48 |
| pHILIC | 202.082 | 150.12 | 24.4 | 0.48 |
| pHILIC | 202.082 | 75.8353 | 24.4 | 0.48 |
| pHILIC | 202.082 | 68.621 | 24.4 | 0.48 |
| pHILIC | 202.082 | 59.6282 | 24.4 | 0.48 |
| pHILIC | 202.082 | 192.4891 | 24.4 | 0.48 |
| pHILIC | 202.082 | 60.6934 | 24.4 | 0.48 |
| pHILIC | 202.082 | 120.6583 | 24.4 | 0.48 |
| pHILIC | 202.082 | 74.9614 | 24.4 | 0.48 |
| pHILIC | 202.082 | 54.694 | 24.4 | 0.48 |
| pHILIC | 202.082 | 224.4591 | 24.4 | 0.48 |
| pHILIC | 202.082 | 71.2294 | 24.4 | 0.48 |
| pHILIC | 202.082 | 78.2251 | 24.4 | 0.48 |
| pHILIC | 202.082 | 58.1769 | 24.4 | 0.48 |
| pHILIC | 202.082 | 117.015 | 24.3 | 0.48 |
| pHILIC | 202.082 | 76.2717 | 24.3 | 0.48 |
| pHILIC | 202.082 | 71.7027 | 24.3 | 0.48 |
| pHILIC | 202.082 | 173.0444 | 24.3 | 0.48 |
| pHILIC | 202.082 | 177.6149 | 24.3 | 0.48 |
| pHILIC | 202.082 | 67.6692 | 24.3 | 0.48 |
| pHILIC | 202.082 | 203.5021 | 24.3 | 0.48 |
| pHILIC | 202.082 | 102.03 | 24.3 | 0.48 |
| pHILIC | 202.082 | 206.9259 | 24.3 | 0.48 |
| pHILIC | 202.082 | 225.5436 | 24.3 | 0.48 |

| | | | | |
|---|---|---|---|---|
| pHILIC | 202.082 | 88.4918 | 24.3 | 0.48 |
| pHILIC | 202.082 | 150.0989 | 24.3 | 0.48 |
| pHILIC | 202.082 | 74.5499 | 24.3 | 0.48 |
| pHILIC | 202.082 | 149.0883 | 24.3 | 0.48 |
| pHILIC | 202.082 | 135.1562 | 24.3 | 0.48 |
| pHILIC | 202.082 | 75.5011 | 24.3 | 0.48 |
| pHILIC | 202.082 | 108.9298 | 24.3 | 0.48 |
| pHILIC | 202.082 | 47.5211 | 24.3 | 0.48 |
| pHILIC | 202.082 | 52.1137 | 24.3 | 0.48 |
| pHILIC | 202.082 | 50.6351 | 24.3 | 0.48 |
| pHILIC | 202.082 | 221.9092 | 24.2 | 0.48 |
| pHILIC | 202.082 | 176.0457 | 24.2 | 0.48 |
| pHILIC | 202.082 | 93.4173 | 24.2 | 0.48 |
| pHILIC | 202.082 | 76.8158 | 24.2 | 0.48 |
| pHILIC | 202.082 | 88.0474 | 24.2 | 0.48 |
| pHILIC | 202.082 | 140.5405 | 24.2 | 0.48 |
| pHILIC | 202.082 | 55.8276 | 24.2 | 0.48 |
| pHILIC | 202.082 | 191.1198 | 24.2 | 0.48 |
| pHILIC | 202.082 | 172.3146 | 24.2 | 0.48 |
| pHILIC | 202.082 | 100.1789 | 24.2 | 0.48 |
| pHILIC | 202.082 | 157.8093 | 24.2 | 0.48 |
| pHILIC | 202.082 | 62.9315 | 24.2 | 0.48 |
| pHILIC | 202.082 | 86.5703 | 24.2 | 0.48 |
| pHILIC | 202.082 | 206.2907 | 24.2 | 0.48 |
| pHILIC | 202.082 | 76.8565 | 24.2 | 0.48 |
| pHILIC | 202.082 | 115.5786 | 24.2 | 0.48 |
| pHILIC | 202.082 | 74.4243 | 24.2 | 0.48 |
| pHILIC | 202.082 | 125.059 | 24.2 | 0.48 |
| pHILIC | 202.082 | 53.2218 | 24.2 | 0.48 |
| pHILIC | 202.082 | 161.597 | 24.2 | 0.48 |
| pHILIC | 202.082 | 76.5424 | 24.2 | 0.48 |
| pHILIC | 202.082 | 56.1394 | 24.2 | 0.48 |
| pHILIC | 202.082 | 110.7482 | 24.2 | 0.48 |
| pHILIC | 202.082 | 120.1708 | 24.2 | 0.48 |
| pHILIC | 202.082 | 65.8459 | 24.2 | 0.48 |
| pHILIC | 202.082 | 120.5231 | 24.2 | 0.48 |
| pHILIC | 202.082 | 103.9193 | 24.2 | 0.48 |
| pHILIC | 202.082 | 94.8017 | 24.2 | 0.48 |
| pHILIC | 202.082 | 87.9081 | 24.2 | 0.48 |
| pHILIC | 202.082 | 79.9287 | 24.2 | 0.48 |
| pHILIC | 202.082 | 107.8117 | 24.2 | 0.48 |
| pHILIC | 202.082 | 198.8085 | 24.2 | 0.48 |

| | | | | |
|---|---|---|---|---|
| pHILIC | 202.082 | 194.5021 | 24.2 | 0.48 |
| pHILIC | 202.082 | 70.1559 | 24.2 | 0.48 |
| pHILIC | 202.082 | 46.8926 | 24.2 | 0.48 |
| pHILIC | 202.082 | 191.6592 | 24.2 | 0.48 |
| pHILIC | 202.082 | 103.3707 | 24.1 | 0.48 |
| pHILIC | 202.082 | 165.6576 | 24.1 | 0.48 |
| pHILIC | 202.082 | 107.0494 | 24.1 | 0.48 |
| pHILIC | 202.082 | 81.4935 | 24.1 | 0.48 |
| pHILIC | 202.082 | 60.6789 | 24.1 | 0.48 |
| pHILIC | 202.082 | 192.107 | 24.1 | 0.48 |
| pHILIC | 202.082 | 66.2057 | 24.1 | 0.48 |
| pHILIC | 202.082 | 60.4616 | 24.1 | 0.48 |
| pHILIC | 202.082 | 99.3859 | 24.1 | 0.48 |
| pHILIC | 202.082 | 138.3387 | 24.1 | 0.48 |
| pHILIC | 202.082 | 83.7925 | 24.1 | 0.48 |
| pHILIC | 202.082 | 101.3012 | 24.1 | 0.48 |
| pHILIC | 202.082 | 48.5478 | 24.1 | 0.48 |
| pHILIC | 202.082 | 176.5836 | 24.1 | 0.48 |
| pHILIC | 202.082 | 176.5077 | 24.1 | 0.48 |
| pHILIC | 202.082 | 78.995 | 24.1 | 0.48 |
| pHILIC | 202.082 | 57.6363 | 24.1 | 0.48 |
| pHILIC | 202.082 | 119.9769 | 24.1 | 0.48 |
| pHILIC | 202.082 | 83.451 | 24.1 | 0.48 |
| pHILIC | 202.082 | 106.1853 | 24.1 | 0.48 |
| pHILIC | 202.082 | 105.5962 | 24.1 | 0.48 |
| pHILIC | 202.082 | 118.6056 | 24.1 | 0.48 |
| pHILIC | 202.082 | 100.2426 | 24.1 | 0.48 |
| pHILIC | 202.082 | 50.9756 | 24.1 | 0.48 |
| pHILIC | 202.082 | 55.8735 | 24.1 | 0.48 |
| pHILIC | 202.082 | 67.6575 | 24.1 | 0.48 |
| pHILIC | 202.082 | 50.5726 | 24.1 | 0.48 |
| pHILIC | 202.082 | 120.9864 | 24 | 0.47 |
| pHILIC | 202.082 | 45.7878 | 24 | 0.47 |
| pHILIC | 202.082 | 92.9109 | 24 | 0.47 |
| pHILIC | 202.082 | 174.3175 | 24 | 0.47 |
| pHILIC | 202.082 | 57.8694 | 24 | 0.47 |
| pHILIC | 202.082 | 91.8417 | 24 | 0.47 |
| pHILIC | 202.082 | 73.1906 | 24 | 0.47 |
| pHILIC | 202.082 | 106.2695 | 24 | 0.47 |
| pHILIC | 202.082 | 154.3486 | 24 | 0.47 |
| pHILIC | 202.082 | 123.3958 | 24 | 0.47 |
| pHILIC | 202.082 | 102.5181 | 24 | 0.47 |

| | | | | |
|---|---|---|---|---|
| pHILIC | 202.082 | 46.4255 | 24 | 0.47 |
| pHILIC | 202.082 | 223.1131 | 24 | 0.47 |
| pHILIC | 202.082 | 54.2575 | 24 | 0.47 |
| pHILIC | 202.082 | 83.2605 | 24 | 0.47 |
| pHILIC | 202.082 | 109.7859 | 24 | 0.47 |
| pHILIC | 202.082 | 48.4995 | 24 | 0.47 |
| pHILIC | 202.082 | 88.7191 | 24 | 0.47 |
| pHILIC | 202.082 | 81.5995 | 24 | 0.47 |
| pHILIC | 202.082 | 87.5571 | 24 | 0.47 |
| pHILIC | 202.082 | 93.4501 | 24 | 0.47 |
| pHILIC | 202.082 | 190.1576 | 24 | 0.47 |
| pHILIC | 202.082 | 103.3064 | 24 | 0.47 |
| pHILIC | 202.082 | 209.1188 | 24 | 0.47 |
| pHILIC | 202.082 | 178.3847 | 24 | 0.47 |
| pHILIC | 202.082 | 127.1619 | 24 | 0.47 |
| pHILIC | 202.082 | 196.43 | 24 | 0.47 |
| pHILIC | 202.082 | 192.1429 | 24 | 0.47 |
| pHILIC | 202.082 | 115.9421 | 24 | 0.47 |
| pHILIC | 202.082 | 99.5255 | 24 | 0.47 |
| pHILIC | 202.082 | 110.6226 | 24 | 0.47 |
| pHILIC | 202.082 | 53.8958 | 24 | 0.47 |
| pHILIC | 202.082 | 70.2763 | 24 | 0.47 |
| pHILIC | 202.082 | 69.8675 | 24 | 0.47 |
| pHILIC | 202.082 | 173.4295 | 24 | 0.47 |
| pHILIC | 202.082 | 45.4813 | 24 | 0.47 |
| pHILIC | 202.082 | 141.864 | 24 | 0.47 |
| pHILIC | 202.082 | 178.5079 | 24 | 0.47 |
| pHILIC | 202.082 | 128.5981 | 23.9 | 0.47 |
| pHILIC | 202.082 | 55.5918 | 23.9 | 0.47 |
| pHILIC | 202.082 | 45.9309 | 23.9 | 0.47 |
| pHILIC | 202.082 | 122.6342 | 23.9 | 0.47 |
| pHILIC | 202.082 | 61.6909 | 23.9 | 0.47 |
| pHILIC | 202.082 | 101.5948 | 23.9 | 0.47 |
| pHILIC | 202.082 | 108.1129 | 23.9 | 0.47 |
| pHILIC | 202.082 | 88.5618 | 23.9 | 0.47 |
| pHILIC | 202.082 | 74.6316 | 23.9 | 0.47 |
| pHILIC | 202.082 | 106.5642 | 23.9 | 0.47 |
| pHILIC | 202.082 | 57.9584 | 23.9 | 0.47 |
| pHILIC | 202.082 | 47.1753 | 23.9 | 0.47 |
| pHILIC | 202.082 | 172.2515 | 23.9 | 0.47 |
| pHILIC | 202.082 | 121.0343 | 23.9 | 0.47 |
| pHILIC | 202.082 | 90.2086 | 23.9 | 0.47 |

| | | | | |
|---|---|---|---|---|
| pHILIC | 202.082 | 218.3149 | 23.9 | 0.47 |
| pHILIC | 202.082 | 52.4174 | 23.9 | 0.47 |
| pHILIC | 202.082 | 129.1867 | 23.9 | 0.47 |
| pHILIC | 202.082 | 64.9959 | 23.9 | 0.47 |
| pHILIC | 202.082 | 50.4834 | 23.9 | 0.47 |
| pHILIC | 202.082 | 106.0647 | 23.9 | 0.47 |
| pHILIC | 202.082 | 180.0344 | 23.9 | 0.47 |
| pHILIC | 202.082 | 51.5672 | 23.9 | 0.47 |
| pHILIC | 202.082 | 84.0026 | 23.9 | 0.47 |
| pHILIC | 202.082 | 153.1786 | 23.9 | 0.47 |
| pHILIC | 202.082 | 46.6775 | 23.9 | 0.47 |
| pHILIC | 202.082 | 89.6127 | 23.9 | 0.47 |
| pHILIC | 202.082 | 60.3093 | 23.9 | 0.47 |
| pHILIC | 202.082 | 72.4212 | 23.9 | 0.47 |
| pHILIC | 202.082 | 61.6865 | 23.9 | 0.47 |
| pHILIC | 202.082 | 121.0165 | 23.9 | 0.47 |
| pHILIC | 202.082 | 64.8237 | 23.9 | 0.47 |
| pHILIC | 202.082 | 152.4172 | 23.9 | 0.47 |
| pHILIC | 202.082 | 48.6798 | 23.8 | 0.47 |
| pHILIC | 202.082 | 215.43 | 23.8 | 0.47 |
| pHILIC | 202.082 | 213.2599 | 23.8 | 0.47 |
| pHILIC | 202.082 | 142.8432 | 23.8 | 0.47 |
| pHILIC | 202.082 | 64.9229 | 23.8 | 0.47 |
| pHILIC | 202.082 | 64.9718 | 23.8 | 0.47 |
| pHILIC | 202.082 | 159.9402 | 23.8 | 0.47 |
| pHILIC | 202.082 | 104.4936 | 23.8 | 0.47 |
| pHILIC | 202.082 | 72.2554 | 23.8 | 0.47 |
| pHILIC | 202.082 | 123.5997 | 23.8 | 0.47 |
| pHILIC | 202.082 | 69.4109 | 23.8 | 0.47 |
| pHILIC | 202.082 | 46.9649 | 23.8 | 0.47 |
| pHILIC | 202.082 | 66.8677 | 23.8 | 0.47 |
| pHILIC | 202.082 | 93.5548 | 23.8 | 0.47 |
| pHILIC | 202.082 | 78.2357 | 23.8 | 0.47 |
| pHILIC | 202.082 | 198.4622 | 23.8 | 0.47 |
| pHILIC | 202.082 | 75.2475 | 23.8 | 0.47 |
| pHILIC | 202.082 | 79.3535 | 23.8 | 0.47 |
| pHILIC | 202.082 | 106.467 | 23.8 | 0.47 |
| pHILIC | 202.082 | 64.0306 | 23.8 | 0.47 |
| pHILIC | 202.082 | 175.3465 | 23.8 | 0.47 |
| pHILIC | 202.082 | 68.4741 | 23.8 | 0.47 |
| pHILIC | 202.082 | 102.598 | 23.8 | 0.47 |
| pHILIC | 202.082 | 54.4653 | 23.8 | 0.47 |

| | | | | |
|---|---|---|---|---|
| pHILIC | 202.082 | 117.2459 | 23.8 | 0.47 |
| pHILIC | 202.082 | 76.336 | 23.8 | 0.47 |
| pHILIC | 202.082 | 51.5997 | 23.8 | 0.47 |
| pHILIC | 202.082 | 50.642 | 23.8 | 0.47 |
| pHILIC | 202.082 | 66.9459 | 23.8 | 0.47 |
| pHILIC | 202.082 | 124.8403 | 23.8 | 0.47 |
| pHILIC | 202.082 | 67.0837 | 23.8 | 0.47 |
| pHILIC | 202.082 | 50.4087 | 23.8 | 0.47 |
| pHILIC | 202.082 | 74.8741 | 23.8 | 0.47 |
| pHILIC | 202.082 | 117.1137 | 23.8 | 0.47 |
| pHILIC | 202.082 | 130.648 | 23.8 | 0.47 |
| pHILIC | 202.082 | 66.8368 | 23.7 | 0.47 |
| pHILIC | 202.082 | 51.6514 | 23.7 | 0.47 |
| pHILIC | 202.082 | 112.9941 | 23.7 | 0.47 |
| pHILIC | 202.082 | 76.947 | 23.7 | 0.47 |
| pHILIC | 202.082 | 87.9782 | 23.7 | 0.47 |
| pHILIC | 202.082 | 67.5649 | 23.7 | 0.47 |
| pHILIC | 202.082 | 47.3314 | 23.7 | 0.47 |
| pHILIC | 202.082 | 60.4136 | 23.7 | 0.47 |
| pHILIC | 202.082 | 99.1566 | 23.7 | 0.47 |
| pHILIC | 202.082 | 58.4212 | 23.7 | 0.47 |
| pHILIC | 202.082 | 50.0225 | 23.7 | 0.47 |
| pHILIC | 202.082 | 50.1134 | 23.7 | 0.47 |
| pHILIC | 202.082 | 48.0488 | 23.7 | 0.47 |
| pHILIC | 202.082 | 63.3393 | 23.7 | 0.47 |
| pHILIC | 202.082 | 49.8164 | 23.7 | 0.47 |
| pHILIC | 202.082 | 96.428 | 23.7 | 0.47 |
| pHILIC | 202.082 | 57.738 | 23.7 | 0.47 |
| pHILIC | 202.082 | 66.5452 | 23.7 | 0.47 |
| pHILIC | 202.082 | 63.3713 | 23.7 | 0.47 |
| pHILIC | 202.082 | 85.907 | 23.7 | 0.47 |
| pHILIC | 202.082 | 50.3356 | 23.7 | 0.47 |
| pHILIC | 202.082 | 89.596 | 23.6 | 0.47 |
| pHILIC | 202.082 | 153.5025 | 23.6 | 0.47 |
| pHILIC | 202.082 | 64.0752 | 23.6 | 0.47 |
| pHILIC | 202.082 | 118.1766 | 23.6 | 0.47 |
| pHILIC | 202.082 | 73.6458 | 23.6 | 0.47 |
| pHILIC | 202.082 | 64.5422 | 23.6 | 0.47 |
| pHILIC | 202.082 | 117.976 | 23.6 | 0.47 |
| pHILIC | 202.082 | 90.8265 | 23.6 | 0.47 |
| pHILIC | 202.082 | 94.0965 | 23.6 | 0.47 |
| pHILIC | 202.082 | 76.7224 | 23.6 | 0.47 |

| | | | | |
|---|---|---|---|---|
| pHILIC | 202.082 | 64.9614 | 23.6 | 0.47 |
| pHILIC | 202.082 | 154.0404 | 23.6 | 0.47 |
| pHILIC | 202.082 | 81.6562 | 23.6 | 0.47 |
| pHILIC | 202.082 | 112.8938 | 23.6 | 0.47 |
| pHILIC | 202.082 | 145.23 | 23.6 | 0.47 |
| pHILIC | 202.082 | 64.2082 | 23.6 | 0.47 |
| pHILIC | 202.082 | 117.3895 | 23.6 | 0.47 |
| pHILIC | 202.082 | 124.4753 | 23.6 | 0.47 |
| pHILIC | 202.082 | 63.1816 | 23.6 | 0.47 |
| pHILIC | 202.082 | 196.0799 | 23.6 | 0.47 |
| pHILIC | 202.082 | 98.158 | 23.6 | 0.47 |
| pHILIC | 202.082 | 71.7163 | 23.6 | 0.47 |
| pHILIC | 202.082 | 171.1199 | 23.6 | 0.47 |
| pHILIC | 202.082 | 52.4156 | 23.6 | 0.47 |
| pHILIC | 202.082 | 70.3369 | 23.6 | 0.47 |
| pHILIC | 202.082 | 223.3597 | 23.6 | 0.47 |
| pHILIC | 202.082 | 113.1219 | 23.6 | 0.47 |
| pHILIC | 202.082 | 164.7446 | 23.6 | 0.47 |
| pHILIC | 202.082 | 80.0677 | 23.6 | 0.47 |
| pHILIC | 202.082 | 66.2007 | 23.6 | 0.47 |
| pHILIC | 202.082 | 190.6557 | 23.6 | 0.47 |
| pHILIC | 202.082 | 61.7376 | 23.6 | 0.47 |
| pHILIC | 202.082 | 71.3624 | 23.6 | 0.47 |
| pHILIC | 202.082 | 192.925 | 23.6 | 0.47 |
| pHILIC | 202.082 | 213.2225 | 23.6 | 0.47 |
| pHILIC | 202.082 | 78.6255 | 23.6 | 0.47 |
| pHILIC | 202.082 | 53.3161 | 23.5 | 0.47 |
| pHILIC | 202.082 | 131.0496 | 23.5 | 0.47 |
| pHILIC | 202.082 | 76.1696 | 23.5 | 0.47 |
| pHILIC | 202.082 | 53.1168 | 23.5 | 0.47 |
| pHILIC | 202.082 | 78.074 | 23.5 | 0.47 |
| pHILIC | 202.082 | 90.1557 | 23.5 | 0.47 |
| pHILIC | 202.082 | 58.9578 | 23.5 | 0.47 |
| pHILIC | 202.082 | 181.9988 | 23.5 | 0.46 |
| pHILIC | 202.082 | 111.9464 | 23.5 | 0.46 |
| pHILIC | 202.082 | 62.3383 | 23.5 | 0.46 |
| pHILIC | 202.082 | 50.6004 | 23.5 | 0.46 |
| pHILIC | 202.082 | 106.3247 | 23.5 | 0.46 |
| pHILIC | 202.082 | 112.0019 | 23.5 | 0.46 |
| pHILIC | 202.082 | 103.2821 | 23.5 | 0.46 |
| pHILIC | 202.082 | 69.8839 | 23.5 | 0.46 |
| pHILIC | 202.082 | 49.6041 | 23.5 | 0.46 |

| | | | | |
|---|---|---|---|---|
| pHILIC | 202.082 | 90.0029 | 23.5 | 0.46 |
| pHILIC | 202.082 | 131.3896 | 23.5 | 0.46 |
| pHILIC | 202.082 | 177.7784 | 23.5 | 0.46 |
| pHILIC | 202.082 | 65.8893 | 23.5 | 0.46 |
| pHILIC | 202.082 | 98.3601 | 23.5 | 0.46 |
| pHILIC | 202.082 | 78.652 | 23.5 | 0.46 |
| pHILIC | 202.082 | 104.8194 | 23.5 | 0.46 |
| pHILIC | 202.082 | 61.2414 | 23.5 | 0.46 |
| pHILIC | 202.082 | 48.5253 | 23.5 | 0.46 |
| pHILIC | 202.082 | 224.9886 | 23.5 | 0.46 |
| pHILIC | 202.082 | 103.0164 | 23.5 | 0.46 |
| pHILIC | 202.082 | 103.6352 | 23.5 | 0.46 |
| pHILIC | 202.082 | 62.1915 | 23.5 | 0.46 |
| pHILIC | 202.082 | 73.1613 | 23.5 | 0.46 |
| pHILIC | 202.082 | 108.3428 | 23.5 | 0.46 |
| pHILIC | 202.082 | 102.833 | 23.5 | 0.46 |
| pHILIC | 202.082 | 56.1347 | 23.5 | 0.46 |
| pHILIC | 202.082 | 72.4365 | 23.5 | 0.46 |
| pHILIC | 202.082 | 93.4107 | 23.5 | 0.46 |
| pHILIC | 202.082 | 74.6772 | 23.5 | 0.46 |
| pHILIC | 202.082 | 181.3049 | 23.5 | 0.46 |
| pHILIC | 202.082 | 56.3662 | 23.4 | 0.46 |
| pHILIC | 202.082 | 64.383 | 23.4 | 0.46 |
| pHILIC | 202.082 | 214.7129 | 23.4 | 0.46 |
| pHILIC | 202.082 | 182.6161 | 23.4 | 0.46 |
| pHILIC | 202.082 | 75.8684 | 23.4 | 0.46 |
| pHILIC | 202.082 | 45.9245 | 23.4 | 0.46 |
| pHILIC | 202.082 | 78.8905 | 23.4 | 0.46 |
| pHILIC | 202.082 | 82.7335 | 23.4 | 0.46 |
| pHILIC | 202.082 | 97.8772 | 23.4 | 0.46 |
| pHILIC | 202.082 | 48.742 | 23.4 | 0.46 |
| pHILIC | 202.082 | 107.4361 | 23.4 | 0.46 |
| pHILIC | 202.082 | 125.1483 | 23.4 | 0.46 |
| pHILIC | 202.082 | 106.4883 | 23.4 | 0.46 |
| pHILIC | 202.082 | 101.7639 | 23.4 | 0.46 |
| pHILIC | 202.082 | 101.4504 | 23.4 | 0.46 |
| pHILIC | 202.082 | 214.1533 | 23.4 | 0.46 |
| pHILIC | 202.082 | 88.7702 | 23.4 | 0.46 |
| pHILIC | 202.082 | 52.4476 | 23.4 | 0.46 |
| pHILIC | 202.082 | 214.416 | 23.4 | 0.46 |
| pHILIC | 202.082 | 56.0515 | 23.4 | 0.46 |
| pHILIC | 202.082 | 94.9592 | 23.4 | 0.46 |

| | | | | |
|---|---|---|---|---|
| pHILIC | 202.082 | 111.4076 | 23.4 | 0.46 |
| pHILIC | 202.082 | 131.4103 | 23.4 | 0.46 |
| pHILIC | 202.082 | 154.7779 | 23.4 | 0.46 |
| pHILIC | 202.082 | 113.8734 | 23.4 | 0.46 |
| pHILIC | 202.082 | 46.8449 | 23.4 | 0.46 |
| pHILIC | 202.082 | 62.5361 | 23.4 | 0.46 |
| pHILIC | 202.082 | 106.6528 | 23.4 | 0.46 |
| pHILIC | 202.082 | 222.3972 | 23.4 | 0.46 |
| pHILIC | 202.082 | 89.2978 | 23.4 | 0.46 |
| pHILIC | 202.082 | 52.4387 | 23.4 | 0.46 |
| pHILIC | 202.082 | 64.6225 | 23.4 | 0.46 |
| pHILIC | 202.082 | 65.2173 | 23.4 | 0.46 |
| pHILIC | 202.082 | 203.8766 | 23.4 | 0.46 |
| pHILIC | 202.082 | 45.8659 | 23.4 | 0.46 |
| pHILIC | 202.082 | 150.4552 | 23.4 | 0.46 |
| pHILIC | 202.082 | 76.9941 | 23.4 | 0.46 |
| pHILIC | 202.082 | 47.4296 | 23.4 | 0.46 |
| pHILIC | 202.082 | 46.0636 | 23.4 | 0.46 |
| pHILIC | 202.082 | 46.1187 | 23.4 | 0.46 |
| pHILIC | 202.082 | 46.9456 | 23.4 | 0.46 |
| pHILIC | 202.082 | 119.1482 | 23.4 | 0.46 |
| pHILIC | 202.082 | 78.7189 | 23.4 | 0.46 |
| pHILIC | 202.082 | 54.2067 | 23.3 | 0.46 |
| pHILIC | 202.082 | 110.3816 | 23.3 | 0.46 |
| pHILIC | 202.082 | 165.8264 | 23.3 | 0.46 |
| pHILIC | 202.082 | 57.2694 | 23.3 | 0.46 |
| pHILIC | 202.082 | 100.8554 | 23.3 | 0.46 |
| pHILIC | 202.082 | 135.1779 | 23.3 | 0.46 |
| pHILIC | 202.082 | 143.3689 | 23.3 | 0.46 |
| pHILIC | 202.082 | 77.043 | 23.3 | 0.46 |
| pHILIC | 202.082 | 82.1419 | 23.3 | 0.46 |
| pHILIC | 202.082 | 97.2835 | 23.3 | 0.46 |
| pHILIC | 202.082 | 58.6368 | 23.3 | 0.46 |
| pHILIC | 202.082 | 61.1283 | 23.3 | 0.46 |
| pHILIC | 202.082 | 59.5391 | 23.3 | 0.46 |
| pHILIC | 202.082 | 84.6091 | 23.3 | 0.46 |
| pHILIC | 202.082 | 74.5209 | 23.3 | 0.46 |
| pHILIC | 202.082 | 114.244 | 23.3 | 0.46 |
| pHILIC | 202.082 | 92.3492 | 23.3 | 0.46 |
| pHILIC | 202.082 | 61.9683 | 23.3 | 0.46 |
| pHILIC | 202.082 | 132.9843 | 23.3 | 0.46 |
| pHILIC | 202.082 | 55.6643 | 23.3 | 0.46 |

| | | | | |
|---|---|---|---|---|
| pHILIC | 202.082 | 137.2728 | 23.3 | 0.46 |
| pHILIC | 202.082 | 70.3315 | 23.3 | 0.46 |
| pHILIC | 202.082 | 192.7927 | 23.3 | 0.46 |
| pHILIC | 202.082 | 217.3422 | 23.3 | 0.46 |
| pHILIC | 202.082 | 110.1276 | 23.3 | 0.46 |
| pHILIC | 202.082 | 56.4923 | 23.3 | 0.46 |
| pHILIC | 202.082 | 55.2552 | 23.3 | 0.46 |
| pHILIC | 202.082 | 192.6063 | 23.3 | 0.46 |
| pHILIC | 202.082 | 146.331 | 23.3 | 0.46 |
| pHILIC | 202.082 | 53.1531 | 23.3 | 0.46 |
| pHILIC | 202.082 | 192.0094 | 23.3 | 0.46 |
| pHILIC | 202.082 | 103.7938 | 23.3 | 0.46 |
| pHILIC | 202.082 | 131.2198 | 23.3 | 0.46 |
| pHILIC | 202.082 | 65.0511 | 23.3 | 0.46 |
| pHILIC | 202.082 | 128.898 | 23.2 | 0.46 |
| pHILIC | 202.082 | 56.9555 | 23.2 | 0.46 |
| pHILIC | 202.082 | 68.2508 | 23.2 | 0.46 |
| pHILIC | 202.082 | 49.1737 | 23.2 | 0.46 |
| pHILIC | 202.082 | 45.3631 | 23.2 | 0.46 |
| pHILIC | 202.082 | 53.9518 | 23.2 | 0.46 |
| pHILIC | 202.082 | 185.3515 | 23.2 | 0.46 |
| pHILIC | 202.082 | 130.9206 | 23.2 | 0.46 |
| pHILIC | 202.082 | 54.2048 | 23.2 | 0.46 |
| pHILIC | 202.082 | 79.6636 | 23.2 | 0.46 |
| pHILIC | 202.082 | 169.8013 | 23.2 | 0.46 |
| pHILIC | 202.082 | 217.3957 | 23.2 | 0.46 |
| pHILIC | 202.082 | 62.9599 | 23.2 | 0.46 |
| pHILIC | 202.082 | 189.3159 | 23.2 | 0.46 |
| pHILIC | 202.082 | 54.892 | 23.2 | 0.46 |
| pHILIC | 202.082 | 223.8581 | 23.2 | 0.46 |
| pHILIC | 202.082 | 92.737 | 23.2 | 0.46 |
| pHILIC | 202.082 | 108.5245 | 23.2 | 0.46 |
| pHILIC | 202.082 | 78.899 | 23.2 | 0.46 |
| pHILIC | 202.082 | 48.9009 | 23.2 | 0.46 |
| pHILIC | 202.082 | 89.6842 | 23.2 | 0.46 |
| pHILIC | 202.082 | 221.6213 | 23.2 | 0.46 |
| pHILIC | 202.082 | 80.8492 | 23.2 | 0.46 |
| pHILIC | 202.082 | 50.4124 | 23.2 | 0.46 |
| pHILIC | 202.082 | 147.9267 | 23.2 | 0.46 |
| pHILIC | 202.082 | 74.8702 | 23.2 | 0.46 |
| pHILIC | 202.082 | 88.8467 | 23.2 | 0.46 |
| pHILIC | 202.082 | 169.6749 | 23.2 | 0.46 |

| | | | | |
|---|---|---|---|---|
| pHILIC | 202.082 | 107.772 | 23.2 | 0.46 |
| pHILIC | 202.082 | 52.2949 | 23.2 | 0.46 |
| pHILIC | 202.082 | 56.2723 | 23.2 | 0.46 |
| pHILIC | 202.082 | 122.2014 | 23.2 | 0.46 |
| pHILIC | 202.082 | 78.7966 | 23.2 | 0.46 |
| pHILIC | 202.082 | 59.8738 | 23.2 | 0.46 |
| pHILIC | 202.082 | 132.3714 | 23.2 | 0.46 |
| pHILIC | 202.082 | 73.8726 | 23.2 | 0.46 |
| pHILIC | 202.082 | 97.4623 | 23.2 | 0.46 |
| pHILIC | 202.082 | 57.1162 | 23.2 | 0.46 |
| pHILIC | 202.082 | 64.7103 | 23.2 | 0.46 |
| pHILIC | 202.082 | 112.2442 | 23.2 | 0.46 |
| pHILIC | 202.082 | 63.2565 | 23.2 | 0.46 |
| pHILIC | 202.082 | 122.2759 | 23.2 | 0.46 |
| pHILIC | 202.082 | 169.6313 | 23.2 | 0.46 |
| pHILIC | 202.082 | 171.4 | 23.2 | 0.46 |
| pHILIC | 202.082 | 101.9933 | 23.2 | 0.46 |
| pHILIC | 202.082 | 136.1237 | 23.2 | 0.46 |
| pHILIC | 202.082 | 109.3453 | 23.2 | 0.46 |
| pHILIC | 202.082 | 137.5283 | 23.2 | 0.46 |
| pHILIC | 202.082 | 154.3251 | 23.2 | 0.46 |
| pHILIC | 202.082 | 120.6415 | 23.2 | 0.46 |
| pHILIC | 202.082 | 61.7203 | 23.2 | 0.46 |
| pHILIC | 202.082 | 100.1516 | 23.2 | 0.46 |
| pHILIC | 202.082 | 213.8937 | 23.2 | 0.46 |
| pHILIC | 202.082 | 159.3683 | 23.1 | 0.46 |
| pHILIC | 202.082 | 104.9546 | 23.1 | 0.46 |
| pHILIC | 202.082 | 179.7536 | 23.1 | 0.46 |
| pHILIC | 202.082 | 90.519 | 23.1 | 0.46 |
| pHILIC | 202.082 | 60.2123 | 23.1 | 0.46 |
| pHILIC | 202.082 | 65.3197 | 23.1 | 0.46 |
| pHILIC | 202.082 | 170.8262 | 23.1 | 0.46 |
| pHILIC | 202.082 | 49.1242 | 23.1 | 0.46 |
| pHILIC | 202.082 | 212.4004 | 23.1 | 0.46 |
| pHILIC | 202.082 | 58.1564 | 23.1 | 0.46 |
| pHILIC | 202.082 | 88.6009 | 23.1 | 0.46 |
| pHILIC | 202.082 | 49.8617 | 23.1 | 0.46 |
| pHILIC | 202.082 | 81.3481 | 23.1 | 0.46 |
| pHILIC | 202.082 | 119.359 | 23.1 | 0.46 |
| pHILIC | 202.082 | 129.0126 | 23.1 | 0.46 |
| pHILIC | 202.082 | 83.0642 | 23.1 | 0.46 |
| pHILIC | 202.082 | 65.7611 | 23.1 | 0.46 |

| | | | | |
|---|---|---|---|---|
| pHILIC | 202.082 | 60.1644 | 23.1 | 0.46 |
| pHILIC | 202.082 | 116.4095 | 23.1 | 0.46 |
| pHILIC | 202.082 | 108.7909 | 23.1 | 0.46 |
| pHILIC | 202.082 | 138.2308 | 23.1 | 0.46 |
| pHILIC | 202.082 | 58.5061 | 23.1 | 0.46 |
| pHILIC | 202.082 | 120.9523 | 23.1 | 0.46 |
| pHILIC | 202.082 | 126.1134 | 23.1 | 0.46 |
| pHILIC | 202.082 | 77.6611 | 23.1 | 0.46 |
| pHILIC | 202.082 | 61.3775 | 23.1 | 0.46 |
| pHILIC | 202.082 | 106.4234 | 23.1 | 0.46 |
| pHILIC | 202.082 | 109.312 | 23.1 | 0.46 |
| pHILIC | 202.082 | 53.7776 | 23.1 | 0.46 |
| pHILIC | 202.082 | 75.3634 | 23.1 | 0.46 |
| pHILIC | 202.082 | 108.1419 | 23.1 | 0.46 |
| pHILIC | 202.082 | 51.0918 | 23.1 | 0.46 |
| pHILIC | 202.082 | 53.0642 | 23.1 | 0.46 |
| pHILIC | 202.082 | 186.3613 | 23.1 | 0.46 |
| pHILIC | 202.082 | 130.0177 | 23.1 | 0.46 |
| pHILIC | 202.082 | 103.5797 | 23.1 | 0.46 |
| pHILIC | 202.082 | 117.3589 | 23.1 | 0.46 |
| pHILIC | 202.082 | 80.9061 | 23.1 | 0.46 |
| pHILIC | 202.082 | 64.25 | 23.1 | 0.46 |
| pHILIC | 202.082 | 62.2219 | 23 | 0.46 |
| pHILIC | 202.082 | 91.0381 | 23 | 0.46 |
| pHILIC | 202.082 | 186.2029 | 23 | 0.46 |
| pHILIC | 202.082 | 211.4436 | 23 | 0.46 |
| pHILIC | 202.082 | 200.9312 | 23 | 0.46 |
| pHILIC | 202.082 | 54.9184 | 23 | 0.45 |
| pHILIC | 202.082 | 46.0542 | 23 | 0.45 |
| pHILIC | 202.082 | 66.986 | 23 | 0.45 |
| pHILIC | 202.082 | 128.5737 | 23 | 0.45 |
| pHILIC | 202.082 | 52.2642 | 23 | 0.45 |
| pHILIC | 202.082 | 80.0171 | 23 | 0.45 |
| pHILIC | 202.082 | 118.8 | 23 | 0.45 |
| pHILIC | 202.082 | 66.2827 | 23 | 0.45 |
| pHILIC | 202.082 | 72.7817 | 23 | 0.45 |
| pHILIC | 202.082 | 68.8006 | 23 | 0.45 |
| pHILIC | 202.082 | 198.1176 | 23 | 0.45 |
| pHILIC | 202.082 | 188.3226 | 23 | 0.45 |
| pHILIC | 202.082 | 133.7237 | 23 | 0.45 |
| pHILIC | 202.082 | 71.1505 | 23 | 0.45 |
| pHILIC | 202.082 | 186.3027 | 23 | 0.45 |

| | | | | |
|---|---|---|---|---|
| pHILIC | 202.082 | 46.0128 | 23 | 0.45 |
| pHILIC | 202.082 | 123.8919 | 23 | 0.45 |
| pHILIC | 202.082 | 201.3743 | 23 | 0.45 |
| pHILIC | 202.082 | 75.9101 | 23 | 0.45 |
| pHILIC | 202.082 | 56.4151 | 23 | 0.45 |
| pHILIC | 202.082 | 56.8882 | 23 | 0.45 |
| pHILIC | 202.082 | 49.2746 | 23 | 0.45 |
| pHILIC | 202.082 | 134.424 | 23 | 0.45 |
| pHILIC | 202.082 | 57.4542 | 23 | 0.45 |
| pHILIC | 202.082 | 73.8933 | 23 | 0.45 |
| pHILIC | 202.082 | 46.1342 | 23 | 0.45 |
| pHILIC | 202.082 | 170.1914 | 23 | 0.45 |
| pHILIC | 202.082 | 212.1611 | 23 | 0.45 |
| pHILIC | 202.082 | 48.1009 | 23 | 0.45 |
| pHILIC | 202.082 | 175.0566 | 23 | 0.45 |
| pHILIC | 202.082 | 80.2305 | 23 | 0.45 |
| pHILIC | 202.082 | 172.5233 | 23 | 0.45 |
| pHILIC | 202.082 | 79.1334 | 23 | 0.45 |
| pHILIC | 202.082 | 173.6223 | 23 | 0.45 |
| pHILIC | 202.082 | 49.0272 | 23 | 0.45 |
| pHILIC | 202.082 | 63.0738 | 23 | 0.45 |
| pHILIC | 202.082 | 66.2102 | 23 | 0.45 |
| pHILIC | 202.082 | 155.0217 | 23 | 0.45 |
| pHILIC | 202.082 | 173.2141 | 23 | 0.45 |
| pHILIC | 202.082 | 163.2172 | 23 | 0.45 |
| pHILIC | 202.082 | 130.34 | 22.9 | 0.45 |
| pHILIC | 202.082 | 148.0541 | 22.9 | 0.45 |
| pHILIC | 202.082 | 64.859 | 22.9 | 0.45 |
| pHILIC | 202.082 | 68.1258 | 22.9 | 0.45 |
| pHILIC | 202.082 | 142.9355 | 22.9 | 0.45 |
| pHILIC | 202.082 | 113.2453 | 22.9 | 0.45 |
| pHILIC | 202.082 | 118.0754 | 22.9 | 0.45 |
| pHILIC | 202.082 | 70.9823 | 22.9 | 0.45 |
| pHILIC | 202.082 | 54.6545 | 22.9 | 0.45 |
| pHILIC | 202.082 | 82.8155 | 22.9 | 0.45 |
| pHILIC | 202.082 | 72.0516 | 22.9 | 0.45 |
| pHILIC | 202.082 | 149.7599 | 22.9 | 0.45 |
| pHILIC | 202.082 | 108.4282 | 22.9 | 0.45 |
| pHILIC | 202.082 | 72.9342 | 22.9 | 0.45 |
| pHILIC | 202.082 | 169.2312 | 22.9 | 0.45 |
| pHILIC | 202.082 | 52.7699 | 22.9 | 0.45 |
| pHILIC | 202.082 | 67.571 | 22.9 | 0.45 |

| | | | | |
|---|---|---|---|---|
| pHILIC | 202.082 | 177.7138 | 22.9 | 0.45 |
| pHILIC | 202.082 | 48.7903 | 22.9 | 0.45 |
| pHILIC | 202.082 | 112.46 | 22.9 | 0.45 |
| pHILIC | 202.082 | 145.1674 | 22.9 | 0.45 |
| pHILIC | 202.082 | 97.0502 | 22.9 | 0.45 |
| pHILIC | 202.082 | 48.3578 | 22.9 | 0.45 |
| pHILIC | 202.082 | 57.0465 | 22.9 | 0.45 |
| pHILIC | 202.082 | 54.0881 | 22.9 | 0.45 |
| pHILIC | 202.082 | 46.4821 | 22.9 | 0.45 |
| pHILIC | 202.082 | 209.3162 | 22.9 | 0.45 |
| pHILIC | 202.082 | 83.3028 | 22.9 | 0.45 |
| pHILIC | 202.082 | 155.23 | 22.8 | 0.45 |
| pHILIC | 202.082 | 63.446 | 22.8 | 0.45 |
| pHILIC | 202.082 | 170.1515 | 22.8 | 0.45 |
| pHILIC | 202.082 | 107.9204 | 22.8 | 0.45 |
| pHILIC | 202.082 | 60.2623 | 22.8 | 0.45 |
| pHILIC | 202.082 | 148.3874 | 22.8 | 0.45 |
| pHILIC | 202.082 | 54.6458 | 22.8 | 0.45 |
| pHILIC | 202.082 | 127.1244 | 22.8 | 0.45 |
| pHILIC | 202.082 | 80.5317 | 22.8 | 0.45 |
| pHILIC | 202.082 | 51.3032 | 22.8 | 0.45 |
| pHILIC | 202.082 | 177.2319 | 22.8 | 0.45 |
| pHILIC | 202.082 | 47.5541 | 22.8 | 0.45 |
| pHILIC | 202.082 | 73.0098 | 22.8 | 0.45 |
| pHILIC | 202.082 | 95.5457 | 22.8 | 0.45 |
| pHILIC | 202.082 | 60.4995 | 22.8 | 0.45 |
| pHILIC | 202.082 | 193.843 | 22.8 | 0.45 |
| pHILIC | 202.082 | 72.3476 | 22.8 | 0.45 |
| pHILIC | 202.082 | 50.0387 | 22.8 | 0.45 |
| pHILIC | 202.082 | 81.7385 | 22.8 | 0.45 |
| pHILIC | 202.082 | 100.4167 | 22.8 | 0.45 |
| pHILIC | 202.082 | 162.5012 | 22.8 | 0.45 |
| pHILIC | 202.082 | 134.7035 | 22.8 | 0.45 |
| pHILIC | 202.082 | 51.0835 | 22.8 | 0.45 |
| pHILIC | 202.082 | 91.6549 | 22.8 | 0.45 |
| pHILIC | 202.082 | 199.297 | 22.8 | 0.45 |
| pHILIC | 202.082 | 83.6938 | 22.8 | 0.45 |
| pHILIC | 202.082 | 65.9478 | 22.8 | 0.45 |
| pHILIC | 202.082 | 163.0621 | 22.8 | 0.45 |
| pHILIC | 202.082 | 83.5652 | 22.8 | 0.45 |
| pHILIC | 202.082 | 57.9369 | 22.8 | 0.45 |
| pHILIC | 202.082 | 55.4991 | 22.8 | 0.45 |

| | | | | |
|---|---|---|---|---|
| pHILIC | 202.082 | 150.3888 | 22.8 | 0.45 |
| pHILIC | 202.082 | 66.2678 | 22.8 | 0.45 |
| pHILIC | 202.082 | 62.3186 | 22.8 | 0.45 |
| pHILIC | 202.082 | 143.7436 | 22.8 | 0.45 |
| pHILIC | 202.082 | 47.47 | 22.8 | 0.45 |
| pHILIC | 202.082 | 51.0024 | 22.8 | 0.45 |
| pHILIC | 202.082 | 55.6023 | 22.8 | 0.45 |
| pHILIC | 202.082 | 77.0223 | 22.8 | 0.45 |
| pHILIC | 202.082 | 76.4625 | 22.8 | 0.45 |
| pHILIC | 202.082 | 103.668 | 22.8 | 0.45 |
| pHILIC | 202.082 | 178.8288 | 22.8 | 0.45 |
| pHILIC | 202.082 | 163.1919 | 22.8 | 0.45 |
| pHILIC | 202.082 | 50.2707 | 22.8 | 0.45 |
| pHILIC | 202.082 | 112.5359 | 22.8 | 0.45 |
| pHILIC | 202.082 | 69.6718 | 22.8 | 0.45 |
| pHILIC | 202.082 | 170.2465 | 22.8 | 0.45 |
| pHILIC | 202.082 | 201.0031 | 22.8 | 0.45 |
| pHILIC | 202.082 | 69.479 | 22.8 | 0.45 |
| pHILIC | 202.082 | 94.5744 | 22.8 | 0.45 |
| pHILIC | 202.082 | 154.1009 | 22.8 | 0.45 |
| pHILIC | 202.082 | 204.925 | 22.8 | 0.45 |
| pHILIC | 202.082 | 109.0555 | 22.7 | 0.45 |
| pHILIC | 202.082 | 46.0214 | 22.7 | 0.45 |
| pHILIC | 202.082 | 216.043 | 22.7 | 0.45 |
| pHILIC | 202.082 | 114.9469 | 22.7 | 0.45 |
| pHILIC | 202.082 | 49.9471 | 22.7 | 0.45 |
| pHILIC | 202.082 | 65.7747 | 22.7 | 0.45 |
| pHILIC | 202.082 | 223.9359 | 22.7 | 0.45 |
| pHILIC | 202.082 | 74.4807 | 22.7 | 0.45 |
| pHILIC | 202.082 | 51.7136 | 22.7 | 0.45 |
| pHILIC | 202.082 | 89.7698 | 22.7 | 0.45 |
| pHILIC | 202.082 | 72.8938 | 22.7 | 0.45 |
| pHILIC | 202.082 | 143.5453 | 22.7 | 0.45 |
| pHILIC | 202.082 | 88.7576 | 22.7 | 0.45 |
| pHILIC | 202.082 | 103.2903 | 22.7 | 0.45 |
| pHILIC | 202.082 | 61.0738 | 22.7 | 0.45 |
| pHILIC | 202.082 | 77.1117 | 22.7 | 0.45 |
| pHILIC | 202.082 | 183.2252 | 22.7 | 0.45 |
| pHILIC | 202.082 | 119.5771 | 22.7 | 0.45 |
| pHILIC | 202.082 | 110.3651 | 22.7 | 0.45 |
| pHILIC | 202.082 | 73.5467 | 22.7 | 0.45 |
| pHILIC | 202.082 | 178.7789 | 22.7 | 0.45 |

| | | | | |
|---|---|---|---|---|
| pHILIC | 202.082 | 78.642 | 22.7 | 0.45 |
| pHILIC | 202.082 | 172.8206 | 22.7 | 0.45 |
| pHILIC | 202.082 | 156.5874 | 22.7 | 0.45 |
| pHILIC | 202.082 | 143.1413 | 22.7 | 0.45 |
| pHILIC | 202.082 | 105.1591 | 22.7 | 0.45 |
| pHILIC | 202.082 | 75.2364 | 22.7 | 0.45 |
| pHILIC | 202.082 | 119.2133 | 22.7 | 0.45 |
| pHILIC | 202.082 | 74.7472 | 22.7 | 0.45 |
| pHILIC | 202.082 | 205.344 | 22.7 | 0.45 |
| pHILIC | 202.082 | 49.6723 | 22.7 | 0.45 |
| pHILIC | 202.082 | 55.1882 | 22.7 | 0.45 |
| pHILIC | 202.082 | 51.2081 | 22.7 | 0.45 |
| pHILIC | 202.082 | 53.956 | 22.7 | 0.45 |
| pHILIC | 202.082 | 210.0339 | 22.7 | 0.45 |
| pHILIC | 202.082 | 98.9218 | 22.7 | 0.45 |
| pHILIC | 202.082 | 66.2233 | 22.7 | 0.45 |
| pHILIC | 202.082 | 91.5783 | 22.7 | 0.45 |
| pHILIC | 202.082 | 96.6431 | 22.7 | 0.45 |
| pHILIC | 202.082 | 53.5063 | 22.7 | 0.45 |
| pHILIC | 202.082 | 116.1702 | 22.7 | 0.45 |
| pHILIC | 202.082 | 132.2594 | 22.7 | 0.45 |
| pHILIC | 202.082 | 48.501 | 22.7 | 0.45 |
| pHILIC | 202.082 | 69.6078 | 22.7 | 0.45 |
| pHILIC | 202.082 | 55.6557 | 22.7 | 0.45 |
| pHILIC | 202.082 | 59.6111 | 22.7 | 0.45 |
| pHILIC | 202.082 | 106.2442 | 22.7 | 0.45 |
| pHILIC | 202.082 | 79.6103 | 22.6 | 0.45 |
| pHILIC | 202.082 | 129.5986 | 22.6 | 0.45 |
| pHILIC | 202.082 | 51.0593 | 22.6 | 0.45 |
| pHILIC | 202.082 | 58.6352 | 22.6 | 0.45 |
| pHILIC | 202.082 | 85.2341 | 22.6 | 0.45 |
| pHILIC | 202.082 | 128.2067 | 22.6 | 0.45 |
| pHILIC | 202.082 | 72.5721 | 22.6 | 0.45 |
| pHILIC | 202.082 | 51.8972 | 22.6 | 0.45 |
| pHILIC | 202.082 | 115.9837 | 22.6 | 0.45 |
| pHILIC | 202.082 | 58.5492 | 22.6 | 0.45 |
| pHILIC | 202.082 | 149.1545 | 22.6 | 0.45 |
| pHILIC | 202.082 | 55.3009 | 22.6 | 0.45 |
| pHILIC | 202.082 | 123.0021 | 22.6 | 0.45 |
| pHILIC | 202.082 | 154.792 | 22.6 | 0.45 |
| pHILIC | 202.082 | 170.67 | 22.6 | 0.45 |
| pHILIC | 202.082 | 121.2997 | 22.6 | 0.45 |

| | | | | |
|---|---|---|---|---|
| pHILIC | 202.082 | 93.7182 | 22.6 | 0.45 |
| pHILIC | 202.082 | 88.7412 | 22.6 | 0.45 |
| pHILIC | 202.082 | 59.6695 | 22.6 | 0.45 |
| pHILIC | 202.082 | 139.5621 | 22.6 | 0.45 |
| pHILIC | 202.082 | 63.155 | 22.6 | 0.45 |
| pHILIC | 202.082 | 53.1558 | 22.6 | 0.45 |
| pHILIC | 202.082 | 60.709 | 22.6 | 0.45 |
| pHILIC | 202.082 | 53.7326 | 22.6 | 0.45 |
| pHILIC | 202.082 | 85.3354 | 22.6 | 0.45 |
| pHILIC | 202.082 | 104.8968 | 22.6 | 0.45 |
| pHILIC | 202.082 | 145.4065 | 22.6 | 0.45 |
| pHILIC | 202.082 | 54.8506 | 22.6 | 0.45 |
| pHILIC | 202.082 | 58.8223 | 22.6 | 0.45 |
| pHILIC | 202.082 | 55.7027 | 22.6 | 0.45 |
| pHILIC | 202.082 | 118.0944 | 22.6 | 0.45 |
| pHILIC | 202.082 | 168.05 | 22.6 | 0.45 |
| pHILIC | 202.082 | 47.6019 | 22.6 | 0.45 |
| pHILIC | 202.082 | 82.058 | 22.6 | 0.45 |
| pHILIC | 202.082 | 51.8121 | 22.6 | 0.45 |
| pHILIC | 202.082 | 146.3754 | 22.6 | 0.45 |
| pHILIC | 202.082 | 171.6755 | 22.6 | 0.45 |
| pHILIC | 202.082 | 68.3546 | 22.6 | 0.45 |
| pHILIC | 202.082 | 153.6257 | 22.6 | 0.45 |
| pHILIC | 202.082 | 63.3787 | 22.6 | 0.45 |
| pHILIC | 202.082 | 82.8633 | 22.6 | 0.45 |
| pHILIC | 202.082 | 175.8742 | 22.6 | 0.45 |
| pHILIC | 202.082 | 84.2851 | 22.5 | 0.45 |
| pHILIC | 202.082 | 186.5907 | 22.5 | 0.45 |
| pHILIC | 202.082 | 71.0806 | 22.5 | 0.45 |
| pHILIC | 202.082 | 59.5342 | 22.5 | 0.45 |
| pHILIC | 202.082 | 123.9801 | 22.5 | 0.45 |
| pHILIC | 202.082 | 217.303 | 22.5 | 0.45 |
| pHILIC | 202.082 | 68.1176 | 22.5 | 0.45 |
| pHILIC | 202.082 | 145.6089 | 22.5 | 0.45 |
| pHILIC | 202.082 | 118.35 | 22.5 | 0.45 |
| pHILIC | 202.082 | 70.9412 | 22.5 | 0.45 |
| pHILIC | 202.082 | 105.752 | 22.5 | 0.45 |
| pHILIC | 202.082 | 63.7655 | 22.5 | 0.45 |
| pHILIC | 202.082 | 56.5562 | 22.5 | 0.44 |
| pHILIC | 202.082 | 90.4434 | 22.5 | 0.44 |
| pHILIC | 202.082 | 137.705 | 22.5 | 0.44 |
| pHILIC | 202.082 | 213.5332 | 22.5 | 0.44 |

| | | | | |
|---|---|---|---|---|
| pHILIC | 202.082 | 177.4041 | 22.5 | 0.44 |
| pHILIC | 202.082 | 57.4498 | 22.5 | 0.44 |
| pHILIC | 202.082 | 166.4872 | 22.5 | 0.44 |
| pHILIC | 202.082 | 110.1355 | 22.5 | 0.44 |
| pHILIC | 202.082 | 97.904 | 22.5 | 0.44 |
| pHILIC | 202.082 | 54.2215 | 22.5 | 0.44 |
| pHILIC | 202.082 | 207.8223 | 22.5 | 0.44 |
| pHILIC | 202.082 | 52.4972 | 22.5 | 0.44 |
| pHILIC | 202.082 | 67.5443 | 22.5 | 0.44 |
| pHILIC | 202.082 | 133.438 | 22.5 | 0.44 |
| pHILIC | 202.082 | 64.7877 | 22.5 | 0.44 |
| pHILIC | 202.082 | 75.0229 | 22.5 | 0.44 |
| pHILIC | 202.082 | 88.4301 | 22.5 | 0.44 |
| pHILIC | 202.082 | 72.111 | 22.5 | 0.44 |
| pHILIC | 202.082 | 67.4485 | 22.5 | 0.44 |
| pHILIC | 202.082 | 58.8673 | 22.5 | 0.44 |
| pHILIC | 202.082 | 121.3367 | 22.5 | 0.44 |
| pHILIC | 202.082 | 211.1595 | 22.5 | 0.44 |
| pHILIC | 202.082 | 179.621 | 22.5 | 0.44 |
| pHILIC | 202.082 | 84.117 | 22.5 | 0.44 |
| pHILIC | 202.082 | 160.9042 | 22.5 | 0.44 |
| pHILIC | 202.082 | 147.8382 | 22.5 | 0.44 |
| pHILIC | 202.082 | 46.8133 | 22.5 | 0.44 |
| pHILIC | 202.082 | 72.0319 | 22.4 | 0.44 |
| pHILIC | 202.082 | 88.8416 | 22.4 | 0.44 |
| pHILIC | 202.082 | 113.0364 | 22.4 | 0.44 |
| pHILIC | 202.082 | 215.2297 | 22.4 | 0.44 |
| pHILIC | 202.082 | 66.2985 | 22.4 | 0.44 |
| pHILIC | 202.082 | 98.6018 | 22.4 | 0.44 |
| pHILIC | 202.082 | 203.0932 | 22.4 | 0.44 |
| pHILIC | 202.082 | 82.5485 | 22.4 | 0.44 |
| pHILIC | 202.082 | 48.2261 | 22.4 | 0.44 |
| pHILIC | 202.082 | 71.4883 | 22.4 | 0.44 |
| pHILIC | 202.082 | 124.1088 | 22.4 | 0.44 |
| pHILIC | 202.082 | 139.842 | 22.4 | 0.44 |
| pHILIC | 202.082 | 93.6686 | 22.4 | 0.44 |
| pHILIC | 202.082 | 118.3962 | 22.4 | 0.44 |
| pHILIC | 202.082 | 62.2144 | 22.4 | 0.44 |
| pHILIC | 202.082 | 58.573 | 22.4 | 0.44 |
| pHILIC | 202.082 | 85.6225 | 22.4 | 0.44 |
| pHILIC | 202.082 | 73.3697 | 22.4 | 0.44 |
| pHILIC | 202.082 | 48.7875 | 22.4 | 0.44 |

| | | | | |
|---|---|---|---|---|
| pHILIC | 202.082 | 89.2634 | 22.4 | 0.44 |
| pHILIC | 202.082 | 54.2992 | 22.4 | 0.44 |
| pHILIC | 202.082 | 58.5001 | 22.4 | 0.44 |
| pHILIC | 202.082 | 57.4029 | 22.4 | 0.44 |
| pHILIC | 202.082 | 120.0636 | 22.4 | 0.44 |
| pHILIC | 202.082 | 64.3212 | 22.4 | 0.44 |
| pHILIC | 202.082 | 82.2908 | 22.4 | 0.44 |
| pHILIC | 202.082 | 59.8087 | 22.4 | 0.44 |
| pHILIC | 202.082 | 88.9757 | 22.4 | 0.44 |
| pHILIC | 202.082 | 45.8628 | 22.4 | 0.44 |
| pHILIC | 202.082 | 54.5384 | 22.4 | 0.44 |
| pHILIC | 202.082 | 52.0471 | 22.4 | 0.44 |
| pHILIC | 202.082 | 91.159 | 22.4 | 0.44 |
| pHILIC | 202.082 | 119.2712 | 22.4 | 0.44 |
| pHILIC | 202.082 | 118.2402 | 22.4 | 0.44 |
| pHILIC | 202.082 | 87.0637 | 22.4 | 0.44 |
| pHILIC | 202.082 | 50.3386 | 22.4 | 0.44 |
| pHILIC | 202.082 | 99.3492 | 22.4 | 0.44 |
| pHILIC | 202.082 | 78.2195 | 22.4 | 0.44 |
| pHILIC | 202.082 | 129.0296 | 22.4 | 0.44 |
| pHILIC | 202.082 | 71.7756 | 22.4 | 0.44 |
| pHILIC | 202.082 | 65.1035 | 22.4 | 0.44 |
| pHILIC | 202.082 | 164.6014 | 22.4 | 0.44 |
| pHILIC | 202.082 | 95.5898 | 22.4 | 0.44 |
| pHILIC | 202.082 | 57.3762 | 22.4 | 0.44 |
| pHILIC | 202.082 | 81.0432 | 22.4 | 0.44 |
| pHILIC | 202.082 | 83.1749 | 22.4 | 0.44 |
| pHILIC | 202.082 | 79.0851 | 22.4 | 0.44 |
| pHILIC | 202.082 | 61.3887 | 22.4 | 0.44 |
| pHILIC | 202.082 | 53.049 | 22.4 | 0.44 |
| pHILIC | 202.082 | 46.8801 | 22.4 | 0.44 |
| pHILIC | 202.082 | 193.4935 | 22.4 | 0.44 |
| pHILIC | 202.082 | 63.9144 | 22.4 | 0.44 |
| pHILIC | 202.082 | 65.989 | 22.4 | 0.44 |
| pHILIC | 202.082 | 122.9359 | 22.4 | 0.44 |
| pHILIC | 202.082 | 198.846 | 22.4 | 0.44 |
| pHILIC | 202.082 | 218.9105 | 22.4 | 0.44 |
| pHILIC | 202.082 | 95.7743 | 22.4 | 0.44 |
| pHILIC | 202.082 | 97.3006 | 22.3 | 0.44 |
| pHILIC | 202.082 | 145.6929 | 22.3 | 0.44 |
| pHILIC | 202.082 | 56.7353 | 22.3 | 0.44 |
| pHILIC | 202.082 | 116.0424 | 22.3 | 0.44 |

| | | | | |
|---|---|---|---|---|
| pHILIC | 202.082 | 138.1653 | 22.3 | 0.44 |
| pHILIC | 202.082 | 48.3974 | 22.3 | 0.44 |
| pHILIC | 202.082 | 46.6458 | 22.3 | 0.44 |
| pHILIC | 202.082 | 50.5628 | 22.3 | 0.44 |
| pHILIC | 202.082 | 195.5678 | 22.3 | 0.44 |
| pHILIC | 202.082 | 79.4786 | 22.3 | 0.44 |
| pHILIC | 202.082 | 99.1865 | 22.3 | 0.44 |
| pHILIC | 202.082 | 69.5888 | 22.3 | 0.44 |
| pHILIC | 202.082 | 106.4791 | 22.3 | 0.44 |
| pHILIC | 202.082 | 133.3642 | 22.3 | 0.44 |
| pHILIC | 202.082 | 59.9867 | 22.3 | 0.44 |
| pHILIC | 202.082 | 72.4891 | 22.3 | 0.44 |
| pHILIC | 202.082 | 75.4359 | 22.3 | 0.44 |
| pHILIC | 202.082 | 123.6547 | 22.3 | 0.44 |
| pHILIC | 202.082 | 116.8705 | 22.3 | 0.44 |
| pHILIC | 202.082 | 45.4143 | 22.3 | 0.44 |
| pHILIC | 202.082 | 64.8407 | 22.3 | 0.44 |
| pHILIC | 202.082 | 205.4416 | 22.3 | 0.44 |
| pHILIC | 202.082 | 72.1376 | 22.3 | 0.44 |
| pHILIC | 202.082 | 56.0404 | 22.3 | 0.44 |
| pHILIC | 202.082 | 185.6734 | 22.3 | 0.44 |
| pHILIC | 202.082 | 129.9796 | 22.3 | 0.44 |
| pHILIC | 202.082 | 98.856 | 22.3 | 0.44 |
| pHILIC | 202.082 | 182.4363 | 22.3 | 0.44 |
| pHILIC | 202.082 | 120.8043 | 22.3 | 0.44 |
| pHILIC | 202.082 | 54.9287 | 22.3 | 0.44 |
| pHILIC | 202.082 | 79.5307 | 22.3 | 0.44 |
| pHILIC | 202.082 | 56.8586 | 22.3 | 0.44 |
| pHILIC | 202.082 | 60.7599 | 22.3 | 0.44 |
| pHILIC | 202.082 | 119.6195 | 22.3 | 0.44 |
| pHILIC | 202.082 | 106.6918 | 22.3 | 0.44 |
| pHILIC | 202.082 | 133.696 | 22.3 | 0.44 |
| pHILIC | 202.082 | 57.2773 | 22.3 | 0.44 |
| pHILIC | 202.082 | 73.0436 | 22.3 | 0.44 |
| pHILIC | 202.082 | 115.425 | 22.2 | 0.44 |
| pHILIC | 202.082 | 150.9462 | 22.2 | 0.44 |
| pHILIC | 202.082 | 136.4391 | 22.2 | 0.44 |
| pHILIC | 202.082 | 115.6678 | 22.2 | 0.44 |
| pHILIC | 202.082 | 95.4609 | 22.2 | 0.44 |
| pHILIC | 202.082 | 58.6724 | 22.2 | 0.44 |
| pHILIC | 202.082 | 77.8552 | 22.2 | 0.44 |
| pHILIC | 202.082 | 132.6492 | 22.2 | 0.44 |

| | | | | |
|---|---|---|---|---|
| pHILIC | 202.082 | 78.3199 | 22.2 | 0.44 |
| pHILIC | 202.082 | 130.7723 | 22.2 | 0.44 |
| pHILIC | 202.082 | 146.5005 | 22.2 | 0.44 |
| pHILIC | 202.082 | 116.5091 | 22.2 | 0.44 |
| pHILIC | 202.082 | 148.1771 | 22.2 | 0.44 |
| pHILIC | 202.082 | 62.6741 | 22.2 | 0.44 |
| pHILIC | 202.082 | 52.9198 | 22.2 | 0.44 |
| pHILIC | 202.082 | 77.5789 | 22.2 | 0.44 |
| pHILIC | 202.082 | 151.3583 | 22.2 | 0.44 |
| pHILIC | 202.082 | 164.3976 | 22.2 | 0.44 |
| pHILIC | 202.082 | 123.3727 | 22.2 | 0.44 |
| pHILIC | 202.082 | 51.235 | 22.2 | 0.44 |
| pHILIC | 202.082 | 117.7303 | 22.2 | 0.44 |
| pHILIC | 202.082 | 53.7255 | 22.2 | 0.44 |
| pHILIC | 202.082 | 183.3257 | 22.2 | 0.44 |
| pHILIC | 202.082 | 76.1596 | 22.2 | 0.44 |
| pHILIC | 202.082 | 121.0649 | 22.2 | 0.44 |
| pHILIC | 202.082 | 165.2611 | 22.2 | 0.44 |
| pHILIC | 202.082 | 90.2637 | 22.2 | 0.44 |
| pHILIC | 202.082 | 65.3729 | 22.2 | 0.44 |
| pHILIC | 202.082 | 137.447 | 22.2 | 0.44 |
| pHILIC | 202.082 | 47.1629 | 22.2 | 0.44 |
| pHILIC | 202.082 | 103.4529 | 22.2 | 0.44 |
| pHILIC | 202.082 | 141.2419 | 22.2 | 0.44 |
| pHILIC | 202.082 | 187.4936 | 22.2 | 0.44 |
| pHILIC | 202.082 | 184.8408 | 22.2 | 0.44 |
| pHILIC | 202.082 | 76.2996 | 22.2 | 0.44 |
| pHILIC | 202.082 | 48.8886 | 22.2 | 0.44 |
| pHILIC | 202.082 | 105.7321 | 22.2 | 0.44 |
| pHILIC | 202.082 | 51.5355 | 22.2 | 0.44 |
| pHILIC | 202.082 | 47.7677 | 22.2 | 0.44 |
| pHILIC | 202.082 | 57.4725 | 22.2 | 0.44 |
| pHILIC | 202.082 | 73.7477 | 22.2 | 0.44 |
| pHILIC | 202.082 | 52.7717 | 22.2 | 0.44 |
| pHILIC | 202.082 | 59.2443 | 22.2 | 0.44 |
| pHILIC | 202.082 | 56.8617 | 22.1 | 0.44 |
| pHILIC | 202.082 | 72.6862 | 22.1 | 0.44 |
| pHILIC | 202.082 | 110.4639 | 22.1 | 0.44 |
| pHILIC | 202.082 | 70.1019 | 22.1 | 0.44 |
| pHILIC | 202.082 | 86.8537 | 22.1 | 0.44 |
| pHILIC | 202.082 | 66.8507 | 22.1 | 0.44 |
| pHILIC | 202.082 | 110.4316 | 22.1 | 0.44 |

| | | | | |
|---|---|---|---|---|
| pHILIC | 202.082 | 54.3432 | 22.1 | 0.44 |
| pHILIC | 202.082 | 45.987 | 22.1 | 0.44 |
| pHILIC | 202.082 | 92.5005 | 22.1 | 0.44 |
| pHILIC | 202.082 | 46.3592 | 22.1 | 0.44 |
| pHILIC | 202.082 | 105.3391 | 22.1 | 0.44 |
| pHILIC | 202.082 | 141.5243 | 22.1 | 0.44 |
| pHILIC | 202.082 | 132.4972 | 22.1 | 0.44 |
| pHILIC | 202.082 | 55.6124 | 22.1 | 0.44 |
| pHILIC | 202.082 | 62.0784 | 22.1 | 0.44 |
| pHILIC | 202.082 | 137.6582 | 22.1 | 0.44 |
| pHILIC | 202.082 | 124.9215 | 22.1 | 0.44 |
| pHILIC | 202.082 | 52.42 | 22.1 | 0.44 |
| pHILIC | 202.082 | 112.886 | 22.1 | 0.44 |
| pHILIC | 202.082 | 195.9329 | 22.1 | 0.44 |
| pHILIC | 202.082 | 114.692 | 22.1 | 0.44 |
| pHILIC | 202.082 | 47.4614 | 22.1 | 0.44 |
| pHILIC | 202.082 | 133.5044 | 22.1 | 0.44 |
| pHILIC | 202.082 | 58.7914 | 22.1 | 0.44 |
| pHILIC | 202.082 | 86.2299 | 22.1 | 0.44 |
| pHILIC | 202.082 | 114.966 | 22.1 | 0.44 |
| pHILIC | 202.082 | 77.8701 | 22.1 | 0.44 |
| pHILIC | 202.082 | 103.0476 | 22.1 | 0.44 |
| pHILIC | 202.082 | 52.0986 | 22.1 | 0.44 |
| pHILIC | 202.082 | 68.4604 | 22.1 | 0.44 |
| pHILIC | 202.082 | 59.0143 | 22.1 | 0.44 |
| pHILIC | 202.082 | 53.6472 | 22.1 | 0.44 |
| pHILIC | 202.082 | 90.1078 | 22.1 | 0.44 |
| pHILIC | 202.082 | 46.2473 | 22.1 | 0.44 |
| pHILIC | 202.082 | 147.6272 | 22.1 | 0.44 |
| pHILIC | 202.082 | 130.5456 | 22.1 | 0.44 |
| pHILIC | 202.082 | 84.0571 | 22.1 | 0.44 |
| pHILIC | 202.082 | 75.3023 | 22.1 | 0.44 |
| pHILIC | 202.082 | 117.1655 | 22.1 | 0.44 |
| pHILIC | 202.082 | 140.4312 | 22.1 | 0.44 |
| pHILIC | 202.082 | 50.1595 | 22 | 0.44 |
| pHILIC | 202.082 | 105.1772 | 22 | 0.44 |
| pHILIC | 202.082 | 56.2152 | 22 | 0.44 |
| pHILIC | 202.082 | 225.3412 | 22 | 0.44 |
| pHILIC | 202.082 | 145.0286 | 22 | 0.44 |
| pHILIC | 202.082 | 97.4051 | 22 | 0.44 |
| pHILIC | 202.082 | 56.1673 | 22 | 0.44 |
| pHILIC | 202.082 | 54.7225 | 22 | 0.43 |

| | | | | |
|---|---|---|---|---|
| pHILIC | 202.082 | 60.9742 | 22 | 0.43 |
| pHILIC | 202.082 | 61.4718 | 22 | 0.43 |
| pHILIC | 202.082 | 147.53 | 22 | 0.43 |
| pHILIC | 202.082 | 141.0147 | 22 | 0.43 |
| pHILIC | 202.082 | 54.2777 | 22 | 0.43 |
| pHILIC | 202.082 | 58.1845 | 22 | 0.43 |
| pHILIC | 202.082 | 112.4182 | 22 | 0.43 |
| pHILIC | 202.082 | 82.3641 | 22 | 0.43 |
| pHILIC | 202.082 | 55.0967 | 22 | 0.43 |
| pHILIC | 202.082 | 77.861 | 22 | 0.43 |
| pHILIC | 202.082 | 112.167 | 22 | 0.43 |
| pHILIC | 202.082 | 129.0515 | 22 | 0.43 |
| pHILIC | 202.082 | 224.7368 | 22 | 0.43 |
| pHILIC | 202.082 | 103.5144 | 22 | 0.43 |
| pHILIC | 202.082 | 221.4408 | 22 | 0.43 |
| pHILIC | 202.082 | 104.0987 | 22 | 0.43 |
| pHILIC | 202.082 | 93.6074 | 22 | 0.43 |
| pHILIC | 202.082 | 84.0718 | 22 | 0.43 |
| pHILIC | 202.082 | 54.2469 | 22 | 0.43 |
| pHILIC | 202.082 | 97.7218 | 22 | 0.43 |
| pHILIC | 202.082 | 62.162 | 22 | 0.43 |
| pHILIC | 202.082 | 74.5927 | 22 | 0.43 |
| pHILIC | 202.082 | 95.4186 | 22 | 0.43 |
| pHILIC | 202.082 | 169.0832 | 22 | 0.43 |
| pHILIC | 202.082 | 57.5874 | 21.9 | 0.43 |
| pHILIC | 202.082 | 145.7498 | 21.9 | 0.43 |
| pHILIC | 202.082 | 151.7002 | 21.9 | 0.43 |
| pHILIC | 202.082 | 51.0624 | 21.9 | 0.43 |
| pHILIC | 202.082 | 46.6381 | 21.9 | 0.43 |
| pHILIC | 202.082 | 100.0744 | 21.9 | 0.43 |
| pHILIC | 202.082 | 74.9084 | 21.9 | 0.43 |
| pHILIC | 202.082 | 115.1258 | 21.9 | 0.43 |
| pHILIC | 202.082 | 48.4382 | 21.9 | 0.43 |
| pHILIC | 202.082 | 69.9507 | 21.9 | 0.43 |
| pHILIC | 202.082 | 210.484 | 21.9 | 0.43 |
| pHILIC | 202.082 | 54.5833 | 21.9 | 0.43 |
| pHILIC | 202.082 | 161.5063 | 21.9 | 0.43 |
| pHILIC | 202.082 | 46.6396 | 21.9 | 0.43 |
| pHILIC | 202.082 | 135.2713 | 21.9 | 0.43 |
| pHILIC | 202.082 | 48.6412 | 21.9 | 0.43 |
| pHILIC | 202.082 | 69.3864 | 21.9 | 0.43 |
| pHILIC | 202.082 | 121.9373 | 21.9 | 0.43 |

| | | | | |
|---|---|---|---|---|
| pHILIC | 202.082 | 169.157 | 21.9 | 0.43 |
| pHILIC | 202.082 | 212.6236 | 21.9 | 0.43 |
| pHILIC | 202.082 | 132.3911 | 21.9 | 0.43 |
| pHILIC | 202.082 | 101.104 | 21.9 | 0.43 |
| pHILIC | 202.082 | 50.7112 | 21.9 | 0.43 |
| pHILIC | 202.082 | 105.9269 | 21.9 | 0.43 |
| pHILIC | 202.082 | 62.5681 | 21.9 | 0.43 |
| pHILIC | 202.082 | 112.5886 | 21.9 | 0.43 |
| pHILIC | 202.082 | 47.114 | 21.9 | 0.43 |
| pHILIC | 202.082 | 119.121 | 21.9 | 0.43 |
| pHILIC | 202.082 | 100.3671 | 21.9 | 0.43 |
| pHILIC | 202.082 | 84.3382 | 21.9 | 0.43 |
| pHILIC | 202.082 | 63.192 | 21.9 | 0.43 |
| pHILIC | 202.082 | 65.4943 | 21.9 | 0.43 |
| pHILIC | 202.082 | 159.1662 | 21.9 | 0.43 |
| pHILIC | 202.082 | 141.7508 | 21.9 | 0.43 |
| pHILIC | 202.082 | 46.8196 | 21.9 | 0.43 |
| pHILIC | 202.082 | 67.4432 | 21.9 | 0.43 |
| pHILIC | 202.082 | 137.0381 | 21.8 | 0.43 |
| pHILIC | 202.082 | 54.3661 | 21.8 | 0.43 |
| pHILIC | 202.082 | 50.8336 | 21.8 | 0.43 |
| pHILIC | 202.082 | 63.395 | 21.8 | 0.43 |
| pHILIC | 202.082 | 57.3114 | 21.8 | 0.43 |
| pHILIC | 202.082 | 59.8709 | 21.8 | 0.43 |
| pHILIC | 202.082 | 57.89 | 21.8 | 0.43 |
| pHILIC | 202.082 | 96.7582 | 21.8 | 0.43 |
| pHILIC | 202.082 | 53.9094 | 21.8 | 0.43 |
| pHILIC | 202.082 | 68.8141 | 21.8 | 0.43 |
| pHILIC | 202.082 | 212.7383 | 21.8 | 0.43 |
| pHILIC | 202.082 | 118.5737 | 21.8 | 0.43 |
| pHILIC | 202.082 | 56.5797 | 21.8 | 0.43 |
| pHILIC | 202.082 | 86.3203 | 21.8 | 0.43 |
| pHILIC | 202.082 | 100.401 | 21.8 | 0.43 |
| pHILIC | 202.082 | 64.2365 | 21.8 | 0.43 |
| pHILIC | 202.082 | 67.2237 | 21.8 | 0.43 |
| pHILIC | 202.082 | 102.9182 | 21.8 | 0.43 |
| pHILIC | 202.082 | 64.1019 | 21.8 | 0.43 |
| pHILIC | 202.082 | 67.2343 | 21.8 | 0.43 |
| pHILIC | 202.082 | 132.0117 | 21.8 | 0.43 |
| pHILIC | 202.082 | 52.9767 | 21.8 | 0.43 |
| pHILIC | 202.082 | 67.0448 | 21.8 | 0.43 |
| pHILIC | 202.082 | 120.1936 | 21.8 | 0.43 |

| | | | | |
|---|---|---|---|---|
| pHILIC | 202.082 | 123.3526 | 21.8 | 0.43 |
| pHILIC | 202.082 | 108.2726 | 21.8 | 0.43 |
| pHILIC | 202.082 | 69.4363 | 21.8 | 0.43 |
| pHILIC | 202.082 | 62.7822 | 21.8 | 0.43 |
| pHILIC | 202.082 | 49.4273 | 21.8 | 0.43 |
| pHILIC | 202.082 | 84.1748 | 21.8 | 0.43 |
| pHILIC | 202.082 | 143.8032 | 21.8 | 0.43 |
| pHILIC | 202.082 | 146.076 | 21.8 | 0.43 |
| pHILIC | 202.082 | 81.2495 | 21.8 | 0.43 |
| pHILIC | 202.082 | 66.154 | 21.8 | 0.43 |
| pHILIC | 202.082 | 91.3831 | 21.8 | 0.43 |
| pHILIC | 202.082 | 50.7011 | 21.8 | 0.43 |
| pHILIC | 202.082 | 60.3825 | 21.8 | 0.43 |
| pHILIC | 202.082 | 79.4208 | 21.7 | 0.43 |
| pHILIC | 202.082 | 205.5503 | 21.7 | 0.43 |
| pHILIC | 202.082 | 166.5435 | 21.7 | 0.43 |
| pHILIC | 202.082 | 104.6763 | 21.7 | 0.43 |
| pHILIC | 202.082 | 47.858 | 21.7 | 0.43 |
| pHILIC | 202.082 | 160.4116 | 21.7 | 0.43 |
| pHILIC | 202.082 | 177.0869 | 21.7 | 0.43 |
| pHILIC | 202.082 | 61.8497 | 21.7 | 0.43 |
| pHILIC | 202.082 | 70.1454 | 21.7 | 0.43 |
| pHILIC | 202.082 | 47.1346 | 21.7 | 0.43 |
| pHILIC | 202.082 | 107.9012 | 21.7 | 0.43 |
| pHILIC | 202.082 | 51.3876 | 21.7 | 0.43 |
| pHILIC | 202.082 | 55.3483 | 21.7 | 0.43 |
| pHILIC | 202.082 | 136.3971 | 21.7 | 0.43 |
| pHILIC | 202.082 | 164.1092 | 21.7 | 0.43 |
| pHILIC | 202.082 | 83.2833 | 21.7 | 0.43 |
| pHILIC | 202.082 | 146.0229 | 21.7 | 0.43 |
| pHILIC | 202.082 | 106.4987 | 21.7 | 0.43 |
| pHILIC | 202.082 | 49.399 | 21.7 | 0.43 |
| pHILIC | 202.082 | 102.8859 | 21.7 | 0.43 |
| pHILIC | 202.082 | 110.2806 | 21.7 | 0.43 |
| pHILIC | 202.082 | 117.8211 | 21.7 | 0.43 |
| pHILIC | 202.082 | 185.0557 | 21.7 | 0.43 |
| pHILIC | 202.082 | 66.3223 | 21.7 | 0.43 |
| pHILIC | 202.082 | 63.277 | 21.7 | 0.43 |
| pHILIC | 202.082 | 58.8557 | 21.7 | 0.43 |
| pHILIC | 202.082 | 89.1421 | 21.7 | 0.43 |
| pHILIC | 202.082 | 73.2584 | 21.7 | 0.43 |
| pHILIC | 202.082 | 149.865 | 21.7 | 0.43 |

| | | | | |
|---|---|---|---|---|
| pHILIC | 202.082 | 50.5803 | 21.7 | 0.43 |
| pHILIC | 202.082 | 61.9267 | 21.7 | 0.43 |
| pHILIC | 202.082 | 133.5771 | 21.7 | 0.43 |
| pHILIC | 202.082 | 46.3478 | 21.7 | 0.43 |
| pHILIC | 202.082 | 99.5694 | 21.6 | 0.43 |
| pHILIC | 202.082 | 99.9869 | 21.6 | 0.43 |
| pHILIC | 202.082 | 96.8781 | 21.6 | 0.43 |
| pHILIC | 202.082 | 214.4857 | 21.6 | 0.43 |
| pHILIC | 202.082 | 219.7105 | 21.6 | 0.43 |
| pHILIC | 202.082 | 60.3465 | 21.6 | 0.43 |
| pHILIC | 202.082 | 70.5203 | 21.6 | 0.43 |
| pHILIC | 202.082 | 47.6581 | 21.6 | 0.43 |
| pHILIC | 202.082 | 61.7352 | 21.6 | 0.43 |
| pHILIC | 202.082 | 80.5511 | 21.6 | 0.43 |
| pHILIC | 202.082 | 83.6111 | 21.6 | 0.43 |
| pHILIC | 202.082 | 225.45 | 21.6 | 0.43 |
| pHILIC | 202.082 | 57.8323 | 21.6 | 0.43 |
| pHILIC | 202.082 | 90.7872 | 21.6 | 0.43 |
| pHILIC | 202.082 | 175.561 | 21.6 | 0.43 |
| pHILIC | 202.082 | 48.8005 | 21.6 | 0.43 |
| pHILIC | 202.082 | 184.3367 | 21.6 | 0.43 |
| pHILIC | 202.082 | 204.0608 | 21.6 | 0.43 |
| pHILIC | 202.082 | 50.1113 | 21.6 | 0.43 |
| pHILIC | 202.082 | 109.7481 | 21.6 | 0.43 |
| pHILIC | 202.082 | 104.4155 | 21.6 | 0.43 |
| pHILIC | 202.082 | 53.2774 | 21.6 | 0.43 |
| pHILIC | 202.082 | 61.145 | 21.6 | 0.43 |
| pHILIC | 202.082 | 80.9304 | 21.6 | 0.43 |
| pHILIC | 202.082 | 61.7138 | 21.6 | 0.43 |
| pHILIC | 202.082 | 56.245 | 21.5 | 0.43 |
| pHILIC | 202.082 | 75.6415 | 21.5 | 0.43 |
| pHILIC | 202.082 | 48.7444 | 21.5 | 0.43 |
| pHILIC | 202.082 | 71.861 | 21.5 | 0.43 |
| pHILIC | 202.082 | 75.2442 | 21.5 | 0.43 |
| pHILIC | 202.082 | 49.1349 | 21.5 | 0.43 |
| pHILIC | 202.082 | 226.2818 | 21.5 | 0.43 |
| pHILIC | 202.082 | 60.7668 | 21.5 | 0.43 |
| pHILIC | 202.082 | 47.2399 | 21.5 | 0.43 |
| pHILIC | 202.082 | 205.1726 | 21.5 | 0.43 |
| pHILIC | 202.082 | 63.6731 | 21.5 | 0.43 |
| pHILIC | 202.082 | 97.76 | 21.5 | 0.42 |
| pHILIC | 202.082 | 49.2558 | 21.5 | 0.42 |

| | | | | |
|---|---|---|---|---|
| pHILIC | 202.082 | 58.4353 | 21.5 | 0.42 |
| pHILIC | 202.082 | 103.8468 | 21.5 | 0.42 |
| pHILIC | 202.082 | 74.9692 | 21.5 | 0.42 |
| pHILIC | 202.082 | 182.7348 | 21.5 | 0.42 |
| pHILIC | 202.082 | 97.8396 | 21.5 | 0.42 |
| pHILIC | 202.082 | 97.5248 | 21.5 | 0.42 |
| pHILIC | 202.082 | 152.0721 | 21.5 | 0.42 |
| pHILIC | 202.082 | 74.4707 | 21.5 | 0.42 |
| pHILIC | 202.082 | 75.1183 | 21.5 | 0.42 |
| pHILIC | 202.082 | 72.4691 | 21.5 | 0.42 |
| pHILIC | 202.082 | 66.6118 | 21.5 | 0.42 |
| pHILIC | 202.082 | 106.0842 | 21.5 | 0.42 |
| pHILIC | 202.082 | 73.3924 | 21.5 | 0.42 |
| pHILIC | 202.082 | 73.1048 | 21.5 | 0.42 |
| pHILIC | 202.082 | 73.5754 | 21.5 | 0.42 |
| pHILIC | 202.082 | 62.3819 | 21.5 | 0.42 |
| pHILIC | 202.082 | 108.8741 | 21.5 | 0.42 |
| pHILIC | 202.082 | 98.6902 | 21.5 | 0.42 |
| pHILIC | 202.082 | 50.4417 | 21.5 | 0.42 |
| pHILIC | 202.082 | 57.9409 | 21.5 | 0.42 |
| pHILIC | 202.082 | 94.4259 | 21.5 | 0.42 |
| pHILIC | 202.082 | 47.32 | 21.5 | 0.42 |
| pHILIC | 202.082 | 195.7023 | 21.5 | 0.42 |
| pHILIC | 202.082 | 101.4036 | 21.4 | 0.42 |
| pHILIC | 202.082 | 69.9669 | 21.4 | 0.42 |
| pHILIC | 202.082 | 65.0211 | 21.4 | 0.42 |
| pHILIC | 202.082 | 211.7037 | 21.4 | 0.42 |
| pHILIC | 202.082 | 153.8434 | 21.4 | 0.42 |
| pHILIC | 202.082 | 99.838 | 21.4 | 0.42 |
| pHILIC | 202.082 | 71.5645 | 21.4 | 0.42 |
| pHILIC | 202.082 | 142.9769 | 21.4 | 0.42 |
| pHILIC | 202.082 | 58.033 | 21.4 | 0.42 |
| pHILIC | 202.082 | 138.5408 | 21.4 | 0.42 |
| pHILIC | 202.082 | 109.5679 | 21.4 | 0.42 |
| pHILIC | 202.082 | 47.0151 | 21.4 | 0.42 |
| pHILIC | 202.082 | 111.6086 | 21.4 | 0.42 |
| pHILIC | 202.082 | 70.2944 | 21.4 | 0.42 |
| pHILIC | 202.082 | 55.9775 | 21.4 | 0.42 |
| pHILIC | 202.082 | 95.3278 | 21.4 | 0.42 |
| pHILIC | 202.082 | 90.1604 | 21.4 | 0.42 |
| pHILIC | 202.082 | 93.8674 | 21.4 | 0.42 |
| pHILIC | 202.082 | 51.1823 | 21.4 | 0.42 |

| | | | | |
|---|---|---|---|---|
| pHILIC | 202.082 | 51.1136 | 21.4 | 0.42 |
| pHILIC | 202.082 | 62.5158 | 21.4 | 0.42 |
| pHILIC | 202.082 | 91.0798 | 21.4 | 0.42 |
| pHILIC | 202.082 | 73.539 | 21.4 | 0.42 |
| pHILIC | 202.082 | 222.5967 | 21.4 | 0.42 |
| pHILIC | 202.082 | 60.4349 | 21.4 | 0.42 |
| pHILIC | 202.082 | 45.9792 | 21.4 | 0.42 |
| pHILIC | 202.082 | 57.6575 | 21.4 | 0.42 |
| pHILIC | 202.082 | 54.5751 | 21.4 | 0.42 |
| pHILIC | 202.082 | 95.9769 | 21.4 | 0.42 |
| pHILIC | 202.082 | 61.7731 | 21.3 | 0.42 |
| pHILIC | 202.082 | 97.2341 | 21.3 | 0.42 |
| pHILIC | 202.082 | 56.6785 | 21.3 | 0.42 |
| pHILIC | 202.082 | 88.8717 | 21.3 | 0.42 |
| pHILIC | 202.082 | 71.3513 | 21.3 | 0.42 |
| pHILIC | 202.082 | 85.9421 | 21.3 | 0.42 |
| pHILIC | 202.082 | 125.8641 | 21.3 | 0.42 |
| pHILIC | 202.082 | 50.474 | 21.3 | 0.42 |
| pHILIC | 202.082 | 93.5753 | 21.3 | 0.42 |
| pHILIC | 202.082 | 57.2361 | 21.3 | 0.42 |
| pHILIC | 202.082 | 76.4558 | 21.3 | 0.42 |
| pHILIC | 202.082 | 125.0886 | 21.3 | 0.42 |
| pHILIC | 202.082 | 52.9535 | 21.3 | 0.42 |
| pHILIC | 202.082 | 81.2863 | 21.3 | 0.42 |
| pHILIC | 202.082 | 79.3117 | 21.3 | 0.42 |
| pHILIC | 202.082 | 63.2741 | 21.3 | 0.42 |
| pHILIC | 202.082 | 78.8569 | 21.3 | 0.42 |
| pHILIC | 202.082 | 97.359 | 21.3 | 0.42 |
| pHILIC | 202.082 | 50.6627 | 21.3 | 0.42 |
| pHILIC | 202.082 | 48.0122 | 21.3 | 0.42 |
| pHILIC | 202.082 | 85.5913 | 21.3 | 0.42 |
| pHILIC | 202.082 | 120.4783 | 21.3 | 0.42 |
| pHILIC | 202.082 | 162.4481 | 21.3 | 0.42 |
| pHILIC | 202.082 | 46.283 | 21.3 | 0.42 |
| pHILIC | 202.082 | 142.9134 | 21.3 | 0.42 |
| pHILIC | 202.082 | 116.3279 | 21.3 | 0.42 |
| pHILIC | 202.082 | 62.4254 | 21.2 | 0.42 |
| pHILIC | 202.082 | 152.4513 | 21.2 | 0.42 |
| pHILIC | 202.082 | 88.5259 | 21.2 | 0.42 |
| pHILIC | 202.082 | 118.1648 | 21.2 | 0.42 |
| pHILIC | 202.082 | 114.1405 | 21.2 | 0.42 |
| pHILIC | 202.082 | 73.1447 | 21.2 | 0.42 |

| | | | | |
|---|---|---|---|---|
| pHILIC | 202.082 | 223.2211 | 21.2 | 0.42 |
| pHILIC | 202.082 | 52.5273 | 21.2 | 0.42 |
| pHILIC | 202.082 | 76.7571 | 21.2 | 0.42 |
| pHILIC | 202.082 | 71.2147 | 21.2 | 0.42 |
| pHILIC | 202.082 | 70.2464 | 21.2 | 0.42 |
| pHILIC | 202.082 | 53.284 | 21.2 | 0.42 |
| pHILIC | 202.082 | 100.9065 | 21.2 | 0.42 |
| pHILIC | 202.082 | 72.2989 | 21.2 | 0.42 |
| pHILIC | 202.082 | 118.3031 | 21.2 | 0.42 |
| pHILIC | 202.082 | 115.0249 | 21.2 | 0.42 |
| pHILIC | 202.082 | 127.7736 | 21.2 | 0.42 |
| pHILIC | 202.082 | 137.8951 | 21.2 | 0.42 |
| pHILIC | 202.082 | 165.1337 | 21.2 | 0.42 |
| pHILIC | 202.082 | 47.3751 | 21.2 | 0.42 |
| pHILIC | 202.082 | 80.3706 | 21.2 | 0.42 |
| pHILIC | 202.082 | 77.4806 | 21.2 | 0.42 |
| pHILIC | 202.082 | 108.9737 | 21.2 | 0.42 |
| pHILIC | 202.082 | 135.3701 | 21.2 | 0.42 |
| pHILIC | 202.082 | 75.307 | 21.2 | 0.42 |
| pHILIC | 202.082 | 54.1082 | 21.2 | 0.42 |
| pHILIC | 202.082 | 133.041 | 21.2 | 0.42 |
| pHILIC | 202.082 | 56.1863 | 21.2 | 0.42 |
| pHILIC | 202.082 | 67.4711 | 21.2 | 0.42 |
| pHILIC | 202.082 | 46.43 | 21.2 | 0.42 |
| pHILIC | 202.082 | 67.1964 | 21.1 | 0.42 |
| pHILIC | 202.082 | 212.8282 | 21.1 | 0.42 |
| pHILIC | 202.082 | 166.6616 | 21.1 | 0.42 |
| pHILIC | 202.082 | 73.0619 | 21.1 | 0.42 |
| pHILIC | 202.082 | 57.2056 | 21.1 | 0.42 |
| pHILIC | 202.082 | 197.6917 | 21.1 | 0.42 |
| pHILIC | 202.082 | 119.5189 | 21.1 | 0.42 |
| pHILIC | 202.082 | 58.9655 | 21.1 | 0.42 |
| pHILIC | 202.082 | 157.8771 | 21.1 | 0.42 |
| pHILIC | 202.082 | 66.3762 | 21.1 | 0.42 |
| pHILIC | 202.082 | 83.6582 | 21.1 | 0.42 |
| pHILIC | 202.082 | 118.9637 | 21.1 | 0.42 |
| pHILIC | 202.082 | 74.554 | 21.1 | 0.42 |
| pHILIC | 202.082 | 157.938 | 21.1 | 0.42 |
| pHILIC | 202.082 | 53.3878 | 21.1 | 0.42 |
| pHILIC | 202.082 | 47.0286 | 21.1 | 0.42 |
| pHILIC | 202.082 | 101.8673 | 21.1 | 0.42 |
| pHILIC | 202.082 | 177.8135 | 21.1 | 0.42 |

| | | | | |
|---|---|---|---|---|
| pHILIC | 202.082 | 65.2001 | 21.1 | 0.42 |
| pHILIC | 202.082 | 193.0663 | 21.1 | 0.42 |
| pHILIC | 202.082 | 47.1688 | 21.1 | 0.42 |
| pHILIC | 202.082 | 46.6695 | 21.1 | 0.42 |
| pHILIC | 202.082 | 206.9904 | 21.1 | 0.42 |
| pHILIC | 202.082 | 133.6752 | 21.1 | 0.42 |
| pHILIC | 202.082 | 181.6286 | 21.1 | 0.42 |
| pHILIC | 202.082 | 59.2901 | 21.1 | 0.42 |
| pHILIC | 202.082 | 58.6203 | 21.1 | 0.42 |
| pHILIC | 202.082 | 66.7596 | 21.1 | 0.42 |
| pHILIC | 202.082 | 55.7631 | 21 | 0.42 |
| pHILIC | 202.082 | 173.0711 | 21 | 0.42 |
| pHILIC | 202.082 | 137.7199 | 21 | 0.42 |
| pHILIC | 202.082 | 74.0088 | 21 | 0.42 |
| pHILIC | 202.082 | 74.2428 | 21 | 0.42 |
| pHILIC | 202.082 | 59.44 | 21 | 0.42 |
| pHILIC | 202.082 | 61.8729 | 21 | 0.42 |
| pHILIC | 202.082 | 82.7915 | 21 | 0.42 |
| pHILIC | 202.082 | 54.007 | 21 | 0.42 |
| pHILIC | 202.082 | 45.5942 | 21 | 0.42 |
| pHILIC | 202.082 | 125.7689 | 21 | 0.42 |
| pHILIC | 202.082 | 163.7545 | 21 | 0.41 |
| pHILIC | 202.082 | 185.3245 | 21 | 0.41 |
| pHILIC | 202.082 | 74.3638 | 21 | 0.41 |
| pHILIC | 202.082 | 47.4751 | 21 | 0.41 |
| pHILIC | 202.082 | 70.5313 | 21 | 0.41 |
| pHILIC | 202.082 | 53.3456 | 21 | 0.41 |
| pHILIC | 202.082 | 56.6681 | 21 | 0.41 |
| pHILIC | 202.082 | 91.044 | 21 | 0.41 |
| pHILIC | 202.082 | 141.1965 | 21 | 0.41 |
| pHILIC | 202.082 | 74.6061 | 21 | 0.41 |
| pHILIC | 202.082 | 52.2029 | 21 | 0.41 |
| pHILIC | 202.082 | 52.6944 | 21 | 0.41 |
| pHILIC | 202.082 | 164.6477 | 21 | 0.41 |
| pHILIC | 202.082 | 49.3731 | 21 | 0.41 |
| pHILIC | 202.082 | 118.6211 | 21 | 0.41 |
| pHILIC | 202.082 | 134.658 | 21 | 0.41 |
| pHILIC | 202.082 | 71.2978 | 21 | 0.41 |
| pHILIC | 202.082 | 57.3028 | 21 | 0.41 |
| pHILIC | 202.082 | 51.427 | 21 | 0.41 |
| pHILIC | 202.082 | 49.4506 | 20.9 | 0.41 |
| pHILIC | 202.082 | 83.5414 | 20.9 | 0.41 |

| | | | | |
|---|---|---|---|---|
| pHILIC | 202.082 | 77.9031 | 20.9 | 0.41 |
| pHILIC | 202.082 | 125.6322 | 20.9 | 0.41 |
| pHILIC | 202.082 | 182.165 | 20.9 | 0.41 |
| pHILIC | 202.082 | 220.3192 | 20.9 | 0.41 |
| pHILIC | 202.082 | 49.7653 | 20.9 | 0.41 |
| pHILIC | 202.082 | 111.7707 | 20.9 | 0.41 |
| pHILIC | 202.082 | 75.7958 | 20.9 | 0.41 |
| pHILIC | 202.082 | 59.1823 | 20.9 | 0.41 |
| pHILIC | 202.082 | 49.0481 | 20.9 | 0.41 |
| pHILIC | 202.082 | 151.7692 | 20.9 | 0.41 |
| pHILIC | 202.082 | 221.4025 | 20.9 | 0.41 |
| pHILIC | 202.082 | 75.205 | 20.9 | 0.41 |
| pHILIC | 202.082 | 46.1366 | 20.9 | 0.41 |
| pHILIC | 202.082 | 113.9037 | 20.9 | 0.41 |
| pHILIC | 202.082 | 63.1412 | 20.9 | 0.41 |
| pHILIC | 202.082 | 61.4001 | 20.9 | 0.41 |
| pHILIC | 202.082 | 55.0786 | 20.9 | 0.41 |
| pHILIC | 202.082 | 46.3844 | 20.9 | 0.41 |
| pHILIC | 202.082 | 225.0891 | 20.9 | 0.41 |
| pHILIC | 202.082 | 73.2513 | 20.9 | 0.41 |
| pHILIC | 202.082 | 174.8334 | 20.9 | 0.41 |
| pHILIC | 202.082 | 86.9007 | 20.9 | 0.41 |
| pHILIC | 202.082 | 77.4946 | 20.9 | 0.41 |
| pHILIC | 202.082 | 88.1254 | 20.9 | 0.41 |
| pHILIC | 202.082 | 78.5678 | 20.9 | 0.41 |
| pHILIC | 202.082 | 64.7449 | 20.9 | 0.41 |
| pHILIC | 202.082 | 168.4606 | 20.9 | 0.41 |
| pHILIC | 202.082 | 103.3136 | 20.8 | 0.41 |
| pHILIC | 202.082 | 172.0298 | 20.8 | 0.41 |
| pHILIC | 202.082 | 61.5553 | 20.8 | 0.41 |
| pHILIC | 202.082 | 220.5186 | 20.8 | 0.41 |
| pHILIC | 202.082 | 65.1153 | 20.8 | 0.41 |
| pHILIC | 202.082 | 183.1729 | 20.8 | 0.41 |
| pHILIC | 202.082 | 78.9081 | 20.8 | 0.41 |
| pHILIC | 202.082 | 56.9332 | 20.8 | 0.41 |
| pHILIC | 202.082 | 71.0913 | 20.8 | 0.41 |
| pHILIC | 202.082 | 134.2411 | 20.8 | 0.41 |
| pHILIC | 202.082 | 47.9188 | 20.8 | 0.41 |
| pHILIC | 202.082 | 187.7295 | 20.8 | 0.41 |
| pHILIC | 202.082 | 93.2571 | 20.8 | 0.41 |
| pHILIC | 202.082 | 61.2041 | 20.8 | 0.41 |
| pHILIC | 202.082 | 63.5497 | 20.8 | 0.41 |

| | | | | |
|---|---|---|---|---|
| pHILIC | 202.082 | 73.1537 | 20.8 | 0.41 |
| pHILIC | 202.082 | 152.5922 | 20.8 | 0.41 |
| pHILIC | 202.082 | 152.716 | 20.8 | 0.41 |
| pHILIC | 202.082 | 58.5826 | 20.8 | 0.41 |
| pHILIC | 202.082 | 111.5256 | 20.8 | 0.41 |
| pHILIC | 202.082 | 57.2138 | 20.8 | 0.41 |
| pHILIC | 202.082 | 49.5988 | 20.8 | 0.41 |
| pHILIC | 202.082 | 48.2535 | 20.7 | 0.41 |
| pHILIC | 202.082 | 117.2116 | 20.7 | 0.41 |
| pHILIC | 202.082 | 204.4574 | 20.7 | 0.41 |
| pHILIC | 202.082 | 78.1475 | 20.7 | 0.41 |
| pHILIC | 202.082 | 162.1139 | 20.7 | 0.41 |
| pHILIC | 202.082 | 56.6129 | 20.7 | 0.41 |
| pHILIC | 202.082 | 51.2578 | 20.7 | 0.41 |
| pHILIC | 202.082 | 165.2078 | 20.7 | 0.41 |
| pHILIC | 202.082 | 48.9712 | 20.7 | 0.41 |
| pHILIC | 202.082 | 93.3881 | 20.7 | 0.41 |
| pHILIC | 202.082 | 117.5913 | 20.7 | 0.41 |
| pHILIC | 202.082 | 162.0717 | 20.7 | 0.41 |
| pHILIC | 202.082 | 50.0837 | 20.7 | 0.41 |
| pHILIC | 202.082 | 66.6947 | 20.7 | 0.41 |
| pHILIC | 202.082 | 217.4759 | 20.7 | 0.41 |
| pHILIC | 202.082 | 117.5164 | 20.7 | 0.41 |
| pHILIC | 202.082 | 99.0762 | 20.7 | 0.41 |
| pHILIC | 202.082 | 72.0846 | 20.7 | 0.41 |
| pHILIC | 202.082 | 69.3142 | 20.7 | 0.41 |
| pHILIC | 202.082 | 164.4327 | 20.7 | 0.41 |
| pHILIC | 202.082 | 66.2149 | 20.6 | 0.41 |
| pHILIC | 202.082 | 145.984 | 20.6 | 0.41 |
| pHILIC | 202.082 | 158.7019 | 20.6 | 0.41 |
| pHILIC | 202.082 | 47.8812 | 20.6 | 0.41 |
| pHILIC | 202.082 | 50.3778 | 20.6 | 0.41 |
| pHILIC | 202.082 | 63.2044 | 20.6 | 0.41 |
| pHILIC | 202.082 | 154.5921 | 20.6 | 0.41 |
| pHILIC | 202.082 | 49.7893 | 20.6 | 0.41 |
| pHILIC | 202.082 | 48.7489 | 20.6 | 0.41 |
| pHILIC | 202.082 | 91.2047 | 20.6 | 0.41 |
| pHILIC | 202.082 | 92.9946 | 20.6 | 0.41 |
| pHILIC | 202.082 | 220.0738 | 20.6 | 0.41 |
| pHILIC | 202.082 | 99.0934 | 20.6 | 0.41 |
| pHILIC | 202.082 | 153.7736 | 20.5 | 0.41 |
| pHILIC | 202.082 | 82.6914 | 20.5 | 0.41 |

| | | | | |
|---|---|---|---|---|
| pHILIC | 202.082 | 89.9107 | 20.5 | 0.41 |
| pHILIC | 202.082 | 82.073 | 20.5 | 0.41 |
| pHILIC | 202.082 | 64.7547 | 20.5 | 0.41 |
| pHILIC | 202.082 | 114.0207 | 20.5 | 0.41 |
| pHILIC | 202.082 | 121.0454 | 20.5 | 0.41 |
| pHILIC | 202.082 | 74.3317 | 20.5 | 0.41 |
| pHILIC | 202.082 | 76.3229 | 20.5 | 0.4 |
| pHILIC | 202.082 | 85.6698 | 20.5 | 0.4 |
| pHILIC | 202.082 | 125.4462 | 20.5 | 0.4 |
| pHILIC | 202.082 | 81.1099 | 20.5 | 0.4 |
| pHILIC | 202.082 | 93.1794 | 20.5 | 0.4 |
| pHILIC | 202.082 | 138.5236 | 20.5 | 0.4 |
| pHILIC | 202.082 | 157.7931 | 20.5 | 0.4 |
| pHILIC | 202.082 | 70.6609 | 20.5 | 0.4 |
| pHILIC | 202.082 | 55.7575 | 20.5 | 0.4 |
| pHILIC | 202.082 | 60.3683 | 20.4 | 0.4 |
| pHILIC | 202.082 | 62.1665 | 20.4 | 0.4 |
| pHILIC | 202.082 | 123.1964 | 20.4 | 0.4 |
| pHILIC | 202.082 | 139.1185 | 20.4 | 0.4 |
| pHILIC | 202.082 | 89.2683 | 20.4 | 0.4 |
| pHILIC | 202.082 | 184.5669 | 20.4 | 0.4 |
| pHILIC | 202.082 | 195.7355 | 20.4 | 0.4 |
| pHILIC | 202.082 | 65.5345 | 20.4 | 0.4 |
| pHILIC | 202.082 | 58.9722 | 20.4 | 0.4 |
| pHILIC | 202.082 | 102.4338 | 20.4 | 0.4 |
| pHILIC | 202.082 | 107.2135 | 20.3 | 0.4 |
| pHILIC | 202.082 | 109.1166 | 20.3 | 0.4 |
| pHILIC | 202.082 | 72.0275 | 20.3 | 0.4 |
| pHILIC | 202.082 | 61.758 | 20.3 | 0.4 |
| pHILIC | 202.082 | 49.2813 | 20.3 | 0.4 |
| pHILIC | 202.082 | 134.7864 | 20.3 | 0.4 |
| pHILIC | 202.082 | 45.9588 | 20.3 | 0.4 |
| pHILIC | 202.082 | 161.7888 | 20.3 | 0.4 |
| pHILIC | 202.082 | 107.3514 | 20.3 | 0.4 |
| pHILIC | 202.082 | 55.722 | 20.3 | 0.4 |
| pHILIC | 202.082 | 64.5063 | 20.3 | 0.4 |
| pHILIC | 202.082 | 112.0842 | 20.3 | 0.4 |
| pHILIC | 202.082 | 50.325 | 20.3 | 0.4 |
| pHILIC | 202.082 | 103.8614 | 20.3 | 0.4 |
| pHILIC | 202.082 | 82.5747 | 20.2 | 0.4 |
| pHILIC | 202.082 | 63.6198 | 20.2 | 0.4 |
| pHILIC | 202.082 | 45.7204 | 20.2 | 0.4 |

| | | | | |
|---|---|---|---|---|
| pHILIC | 202.082 | 50.7903 | 20.2 | 0.4 |
| pHILIC | 202.082 | 78.2515 | 20.2 | 0.4 |
| pHILIC | 202.082 | 120.0209 | 20.2 | 0.4 |
| pHILIC | 202.082 | 90.5472 | 20.2 | 0.4 |
| pHILIC | 202.082 | 115.1349 | 20.2 | 0.4 |
| pHILIC | 202.082 | 62.796 | 20.2 | 0.4 |
| pHILIC | 202.082 | 100.7328 | 20.2 | 0.4 |
| pHILIC | 202.082 | 98.7033 | 20.2 | 0.4 |
| pHILIC | 202.082 | 55.4302 | 20.1 | 0.4 |
| pHILIC | 202.082 | 68.3051 | 20.1 | 0.4 |
| pHILIC | 202.082 | 59.3851 | 20.1 | 0.4 |
| pHILIC | 202.082 | 121.7074 | 20.1 | 0.4 |
| pHILIC | 202.082 | 111.6268 | 20.1 | 0.4 |
| pHILIC | 202.082 | 96.7983 | 20.1 | 0.4 |
| pHILIC | 202.082 | 63.0841 | 20.1 | 0.4 |
| pHILIC | 202.082 | 112.276 | 20.1 | 0.4 |
| pHILIC | 202.082 | 49.8834 | 20.1 | 0.4 |
| pHILIC | 202.082 | 150.5115 | 20.1 | 0.4 |
| pHILIC | 202.082 | 105.2433 | 20.1 | 0.4 |
| pHILIC | 202.082 | 71.4821 | 20.1 | 0.4 |
| pHILIC | 202.082 | 104.2242 | 20.1 | 0.4 |
| pHILIC | 202.082 | 62.4962 | 20.1 | 0.4 |
| pHILIC | 202.082 | 84.9608 | 20.1 | 0.4 |
| pHILIC | 202.082 | 73.1767 | 20.1 | 0.4 |
| pHILIC | 202.082 | 148.1454 | 20.1 | 0.4 |
| pHILIC | 202.082 | 194.1313 | 20.1 | 0.4 |
| pHILIC | 202.082 | 161.0441 | 20.1 | 0.4 |
| pHILIC | 202.082 | 52.3576 | 20.1 | 0.4 |
| pHILIC | 202.082 | 140.6935 | 20 | 0.4 |
| pHILIC | 202.082 | 141.9675 | 20 | 0.4 |
| pHILIC | 202.082 | 86.6241 | 20 | 0.4 |
| pHILIC | 202.082 | 98.5737 | 20 | 0.4 |
| pHILIC | 202.082 | 71.6132 | 20 | 0.4 |
| pHILIC | 202.082 | 46.4114 | 20 | 0.4 |
| pHILIC | 202.082 | 56.5296 | 20 | 0.39 |
| pHILIC | 202.082 | 46.3826 | 20 | 0.39 |
| pHILIC | 202.082 | 98.8024 | 20 | 0.39 |
| pHILIC | 202.082 | 104.174 | 20 | 0.39 |
| pHILIC | 202.082 | 48.1911 | 20 | 0.39 |
| pHILIC | 202.082 | 128.3306 | 20 | 0.39 |
| pHILIC | 202.082 | 80.0302 | 20 | 0.39 |
| pHILIC | 202.082 | 188.5674 | 20 | 0.39 |

| | | | | |
|---|---|---|---|---|
| pHILIC | 202.082 | 99.0168 | 19.9 | 0.39 |
| pHILIC | 202.082 | 99.102 | 19.9 | 0.39 |
| pHILIC | 202.082 | 221.3569 | 19.9 | 0.39 |
| pHILIC | 202.082 | 45.9358 | 19.9 | 0.39 |
| pHILIC | 202.082 | 106.3361 | 19.9 | 0.39 |
| pHILIC | 202.082 | 167.3673 | 19.9 | 0.39 |
| pHILIC | 202.082 | 173.2365 | 19.9 | 0.39 |
| pHILIC | 202.082 | 54.5572 | 19.9 | 0.39 |
| pHILIC | 202.082 | 165.6415 | 19.9 | 0.39 |
| pHILIC | 202.082 | 92.9642 | 19.9 | 0.39 |
| pHILIC | 202.082 | 97.5715 | 19.9 | 0.39 |
| pHILIC | 202.082 | 45.6894 | 19.9 | 0.39 |
| pHILIC | 202.082 | 130.9671 | 19.9 | 0.39 |
| pHILIC | 202.082 | 86.077 | 19.9 | 0.39 |
| pHILIC | 202.082 | 84.4262 | 19.9 | 0.39 |
| pHILIC | 202.082 | 78.8734 | 19.9 | 0.39 |
| pHILIC | 202.082 | 122.1058 | 19.9 | 0.39 |
| pHILIC | 202.082 | 90.5966 | 19.9 | 0.39 |
| pHILIC | 202.082 | 200.3441 | 19.9 | 0.39 |
| pHILIC | 202.082 | 61.6194 | 19.9 | 0.39 |
| pHILIC | 202.082 | 192.2767 | 19.9 | 0.39 |
| pHILIC | 202.082 | 51.1439 | 19.8 | 0.39 |
| pHILIC | 202.082 | 91.4485 | 19.8 | 0.39 |
| pHILIC | 202.082 | 51.3343 | 19.8 | 0.39 |
| pHILIC | 202.082 | 193.8966 | 19.8 | 0.39 |
| pHILIC | 202.082 | 57.6056 | 19.8 | 0.39 |
| pHILIC | 202.082 | 216.6168 | 19.8 | 0.39 |
| pHILIC | 202.082 | 74.6198 | 19.8 | 0.39 |
| pHILIC | 202.082 | 160.1615 | 19.8 | 0.39 |
| pHILIC | 202.082 | 104.5626 | 19.8 | 0.39 |
| pHILIC | 202.082 | 49.0151 | 19.8 | 0.39 |
| pHILIC | 202.082 | 219.5112 | 19.8 | 0.39 |
| pHILIC | 202.082 | 189.942 | 19.8 | 0.39 |
| pHILIC | 202.082 | 59.5083 | 19.8 | 0.39 |
| pHILIC | 202.082 | 46.7488 | 19.8 | 0.39 |
| pHILIC | 202.082 | 96.1761 | 19.8 | 0.39 |
| pHILIC | 202.082 | 62.4028 | 19.8 | 0.39 |
| pHILIC | 202.082 | 69.8361 | 19.7 | 0.39 |
| pHILIC | 202.082 | 71.7663 | 19.7 | 0.39 |
| pHILIC | 202.082 | 203.5807 | 19.7 | 0.39 |
| pHILIC | 202.082 | 69.8278 | 19.7 | 0.39 |
| pHILIC | 202.082 | 48.1523 | 19.7 | 0.39 |

| | | | | |
|---|---|---|---|---|
| pHILIC | 202.082 | 98.9842 | 19.7 | 0.39 |
| pHILIC | 202.082 | 211.2882 | 19.7 | 0.39 |
| pHILIC | 202.082 | 70.5904 | 19.7 | 0.39 |
| pHILIC | 202.082 | 124.9053 | 19.7 | 0.39 |
| pHILIC | 202.082 | 46.9626 | 19.7 | 0.39 |
| pHILIC | 202.082 | 155.7488 | 19.7 | 0.39 |
| pHILIC | 202.082 | 113.2822 | 19.7 | 0.39 |
| pHILIC | 202.082 | 116.703 | 19.7 | 0.39 |
| pHILIC | 202.082 | 59.9026 | 19.7 | 0.39 |
| pHILIC | 202.082 | 109.213 | 19.7 | 0.39 |
| pHILIC | 202.082 | 177.3555 | 19.7 | 0.39 |
| pHILIC | 202.082 | 68.1537 | 19.7 | 0.39 |
| pHILIC | 202.082 | 56.0268 | 19.7 | 0.39 |
| pHILIC | 202.082 | 85.2125 | 19.7 | 0.39 |
| pHILIC | 202.082 | 89.8557 | 19.7 | 0.39 |
| pHILIC | 202.082 | 67.1206 | 19.7 | 0.39 |
| pHILIC | 202.082 | 76.9019 | 19.7 | 0.39 |
| pHILIC | 202.082 | 106.8281 | 19.6 | 0.39 |
| pHILIC | 202.082 | 101.671 | 19.6 | 0.39 |
| pHILIC | 202.082 | 58.9838 | 19.6 | 0.39 |
| pHILIC | 202.082 | 73.7747 | 19.6 | 0.39 |
| pHILIC | 202.082 | 106.7143 | 19.6 | 0.39 |
| pHILIC | 202.082 | 221.1518 | 19.6 | 0.39 |
| pHILIC | 202.082 | 84.6826 | 19.6 | 0.39 |
| pHILIC | 202.082 | 62.4393 | 19.6 | 0.39 |
| pHILIC | 202.082 | 50.2177 | 19.6 | 0.39 |
| pHILIC | 202.082 | 135.4238 | 19.6 | 0.39 |
| pHILIC | 202.082 | 55.3938 | 19.6 | 0.39 |
| pHILIC | 202.082 | 49.5858 | 19.6 | 0.39 |
| pHILIC | 202.082 | 55.9039 | 19.6 | 0.39 |
| pHILIC | 202.082 | 89.4927 | 19.6 | 0.39 |
| pHILIC | 202.082 | 215.1981 | 19.6 | 0.39 |
| pHILIC | 202.082 | 84.1288 | 19.6 | 0.39 |
| pHILIC | 202.082 | 50.7776 | 19.6 | 0.39 |
| pHILIC | 202.082 | 82.7858 | 19.6 | 0.39 |
| pHILIC | 202.082 | 73.535 | 19.6 | 0.39 |
| pHILIC | 202.082 | 147.1954 | 19.5 | 0.39 |
| pHILIC | 202.082 | 129.1408 | 19.5 | 0.39 |
| pHILIC | 202.082 | 122.4242 | 19.5 | 0.39 |
| pHILIC | 202.082 | 65.4728 | 19.5 | 0.39 |
| pHILIC | 202.082 | 156.0616 | 19.5 | 0.39 |
| pHILIC | 202.082 | 45.8536 | 19.5 | 0.39 |

| | | | | |
|---|---|---|---|---|
| pHILIC | 202.082 | 63.5072 | 19.5 | 0.39 |
| pHILIC | 202.082 | 69.2148 | 19.5 | 0.39 |
| pHILIC | 202.082 | 165.4198 | 19.5 | 0.39 |
| pHILIC | 202.082 | 83.0498 | 19.5 | 0.39 |
| pHILIC | 202.082 | 198.3282 | 19.5 | 0.39 |
| pHILIC | 202.082 | 105.3988 | 19.5 | 0.39 |
| pHILIC | 202.082 | 77.848 | 19.5 | 0.39 |
| pHILIC | 202.082 | 217.6254 | 19.5 | 0.39 |
| pHILIC | 202.082 | 192.4128 | 19.5 | 0.38 |
| pHILIC | 202.082 | 48.349 | 19.5 | 0.38 |
| pHILIC | 202.082 | 114.3312 | 19.5 | 0.38 |
| pHILIC | 202.082 | 90.7449 | 19.5 | 0.38 |
| pHILIC | 202.082 | 108.1608 | 19.4 | 0.38 |
| pHILIC | 202.082 | 117.237 | 19.4 | 0.38 |
| pHILIC | 202.082 | 64.176 | 19.4 | 0.38 |
| pHILIC | 202.082 | 57.1987 | 19.4 | 0.38 |
| pHILIC | 202.082 | 142.2707 | 19.4 | 0.38 |
| pHILIC | 202.082 | 138.3691 | 19.4 | 0.38 |
| pHILIC | 202.082 | 55.5125 | 19.4 | 0.38 |
| pHILIC | 202.082 | 76.111 | 19.4 | 0.38 |
| pHILIC | 202.082 | 54.3064 | 19.4 | 0.38 |
| pHILIC | 202.082 | 48.1286 | 19.4 | 0.38 |
| pHILIC | 202.082 | 128.4848 | 19.4 | 0.38 |
| pHILIC | 202.082 | 220.1027 | 19.4 | 0.38 |
| pHILIC | 202.082 | 133.2189 | 19.4 | 0.38 |
| pHILIC | 202.082 | 46.7071 | 19.4 | 0.38 |
| pHILIC | 202.082 | 186.7752 | 19.4 | 0.38 |
| pHILIC | 202.082 | 94.3377 | 19.4 | 0.38 |
| pHILIC | 202.082 | 63.6763 | 19.3 | 0.38 |
| pHILIC | 202.082 | 89.2124 | 19.3 | 0.38 |
| pHILIC | 202.082 | 55.4679 | 19.3 | 0.38 |
| pHILIC | 202.082 | 163.274 | 19.3 | 0.38 |
| pHILIC | 202.082 | 188.8573 | 19.3 | 0.38 |
| pHILIC | 202.082 | 107.2742 | 19.3 | 0.38 |
| pHILIC | 202.082 | 180.1828 | 19.3 | 0.38 |
| pHILIC | 202.082 | 56.2834 | 19.3 | 0.38 |
| pHILIC | 202.082 | 46.158 | 19.3 | 0.38 |
| pHILIC | 202.082 | 83.3208 | 19.3 | 0.38 |
| pHILIC | 202.082 | 79.0522 | 19.3 | 0.38 |
| pHILIC | 202.082 | 45.916 | 19.3 | 0.38 |
| pHILIC | 202.082 | 55.2417 | 19.3 | 0.38 |
| pHILIC | 202.082 | 190.078 | 19.3 | 0.38 |

| | | | | |
|---|---|---|---|---|
| pHILIC | 202.082 | 202.795 | 19.3 | 0.38 |
| pHILIC | 202.082 | 47.5742 | 19.2 | 0.38 |
| pHILIC | 202.082 | 67.4798 | 19.2 | 0.38 |
| pHILIC | 202.082 | 219.8784 | 19.2 | 0.38 |
| pHILIC | 202.082 | 48.313 | 19.2 | 0.38 |
| pHILIC | 202.082 | 87.9912 | 19.2 | 0.38 |
| pHILIC | 202.082 | 69.0914 | 19.2 | 0.38 |
| pHILIC | 202.082 | 90.8514 | 19.2 | 0.38 |
| pHILIC | 202.082 | 84.5989 | 19.2 | 0.38 |
| pHILIC | 202.082 | 63.6485 | 19.2 | 0.38 |
| pHILIC | 202.082 | 204.5415 | 19.2 | 0.38 |
| pHILIC | 202.082 | 47.0503 | 19.2 | 0.38 |
| pHILIC | 202.082 | 46.6413 | 19.2 | 0.38 |
| pHILIC | 202.082 | 60.235 | 19.2 | 0.38 |
| pHILIC | 202.082 | 185.4159 | 19.2 | 0.38 |
| pHILIC | 202.082 | 199.7773 | 19.2 | 0.38 |
| pHILIC | 202.082 | 77.2557 | 19.2 | 0.38 |
| pHILIC | 202.082 | 49.9143 | 19.2 | 0.38 |
| pHILIC | 202.082 | 76.627 | 19.2 | 0.38 |
| pHILIC | 202.082 | 78.757 | 19.2 | 0.38 |
| pHILIC | 202.082 | 50.3636 | 19.2 | 0.38 |
| pHILIC | 202.082 | 70.7576 | 19.1 | 0.38 |
| pHILIC | 202.082 | 183.7835 | 19.1 | 0.38 |
| pHILIC | 202.082 | 79.4001 | 19.1 | 0.38 |
| pHILIC | 202.082 | 146.8953 | 19.1 | 0.38 |
| pHILIC | 202.082 | 69.1683 | 19.1 | 0.38 |
| pHILIC | 202.082 | 79.7579 | 19.1 | 0.38 |
| pHILIC | 202.082 | 97.8887 | 19.1 | 0.38 |
| pHILIC | 202.082 | 90.5269 | 19.1 | 0.38 |
| pHILIC | 202.082 | 144.1473 | 19.1 | 0.38 |
| pHILIC | 202.082 | 108.6155 | 19.1 | 0.38 |
| pHILIC | 202.082 | 125.3875 | 19.1 | 0.38 |
| pHILIC | 202.082 | 106.7916 | 19.1 | 0.38 |
| pHILIC | 202.082 | 53.6443 | 19 | 0.38 |
| pHILIC | 202.082 | 53.7406 | 19 | 0.38 |
| pHILIC | 202.082 | 53.3918 | 19 | 0.38 |
| pHILIC | 202.082 | 48.6598 | 19 | 0.38 |
| pHILIC | 202.082 | 60.8618 | 19 | 0.38 |
| pHILIC | 202.082 | 186.2742 | 19 | 0.38 |
| pHILIC | 202.082 | 184.4329 | 19 | 0.38 |
| pHILIC | 202.082 | 90.2004 | 19 | 0.38 |
| pHILIC | 202.082 | 46.9527 | 19 | 0.38 |

| | | | | |
|---|---|---|---|---|
| pHILIC | 202.082 | 77.2133 | 19 | 0.38 |
| pHILIC | 202.082 | 69.2922 | 19 | 0.37 |
| pHILIC | 202.082 | 45.5441 | 19 | 0.37 |
| pHILIC | 202.082 | 63.0271 | 19 | 0.37 |
| pHILIC | 202.082 | 107.0054 | 19 | 0.37 |
| pHILIC | 202.082 | 66.4573 | 18.9 | 0.37 |
| pHILIC | 202.082 | 174.3721 | 18.9 | 0.37 |
| pHILIC | 202.082 | 67.6197 | 18.9 | 0.37 |
| pHILIC | 202.082 | 99.1437 | 18.9 | 0.37 |
| pHILIC | 202.082 | 52.9395 | 18.9 | 0.37 |
| pHILIC | 202.082 | 112.1442 | 18.9 | 0.37 |
| pHILIC | 202.082 | 48.706 | 18.9 | 0.37 |
| pHILIC | 202.082 | 177.1472 | 18.9 | 0.37 |
| pHILIC | 202.082 | 115.892 | 18.9 | 0.37 |
| pHILIC | 202.082 | 68.5213 | 18.9 | 0.37 |
| pHILIC | 202.082 | 94.876 | 18.9 | 0.37 |
| pHILIC | 202.082 | 90.186 | 18.9 | 0.37 |
| pHILIC | 202.082 | 66.0081 | 18.9 | 0.37 |
| pHILIC | 202.082 | 80.4205 | 18.9 | 0.37 |
| pHILIC | 202.082 | 95.4017 | 18.9 | 0.37 |
| pHILIC | 202.082 | 84.2732 | 18.8 | 0.37 |
| pHILIC | 202.082 | 151.0593 | 18.8 | 0.37 |
| pHILIC | 202.082 | 145.3837 | 18.8 | 0.37 |
| pHILIC | 202.082 | 178.7069 | 18.8 | 0.37 |
| pHILIC | 202.082 | 197.1442 | 18.8 | 0.37 |
| pHILIC | 202.082 | 108.5971 | 18.8 | 0.37 |
| pHILIC | 202.082 | 124.8213 | 18.8 | 0.37 |
| pHILIC | 202.082 | 49.9881 | 18.8 | 0.37 |
| pHILIC | 202.082 | 125.4939 | 18.8 | 0.37 |
| pHILIC | 202.082 | 72.7623 | 18.8 | 0.37 |
| pHILIC | 202.082 | 52.5007 | 18.8 | 0.37 |
| pHILIC | 202.082 | 57.4107 | 18.8 | 0.37 |
| pHILIC | 202.082 | 71.9453 | 18.8 | 0.37 |
| pHILIC | 202.082 | 84.5615 | 18.8 | 0.37 |
| pHILIC | 202.082 | 61.2514 | 18.8 | 0.37 |
| pHILIC | 202.082 | 101.0413 | 18.8 | 0.37 |
| pHILIC | 202.082 | 156.8608 | 18.8 | 0.37 |
| pHILIC | 202.082 | 51.6976 | 18.8 | 0.37 |
| pHILIC | 202.082 | 66.1379 | 18.8 | 0.37 |
| pHILIC | 202.082 | 102.9469 | 18.8 | 0.37 |
| pHILIC | 202.082 | 78.6762 | 18.8 | 0.37 |
| pHILIC | 202.082 | 62.6018 | 18.8 | 0.37 |

| | | | | |
|---|---|---|---|---|
| pHILIC | 202.082 | 87.4284 | 18.7 | 0.37 |
| pHILIC | 202.082 | 74.0582 | 18.7 | 0.37 |
| pHILIC | 202.082 | 119.2226 | 18.7 | 0.37 |
| pHILIC | 202.082 | 83.7675 | 18.7 | 0.37 |
| pHILIC | 202.082 | 49.2027 | 18.7 | 0.37 |
| pHILIC | 202.082 | 59.8117 | 18.7 | 0.37 |
| pHILIC | 202.082 | 106.7748 | 18.7 | 0.37 |
| pHILIC | 202.082 | 48.0629 | 18.7 | 0.37 |
| pHILIC | 202.082 | 49.5026 | 18.7 | 0.37 |
| pHILIC | 202.082 | 120.7611 | 18.7 | 0.37 |
| pHILIC | 202.082 | 200.13 | 18.7 | 0.37 |
| pHILIC | 202.082 | 67.3714 | 18.7 | 0.37 |
| pHILIC | 202.082 | 132.9017 | 18.7 | 0.37 |
| pHILIC | 202.082 | 87.2971 | 18.7 | 0.37 |
| pHILIC | 202.082 | 132.5341 | 18.6 | 0.37 |
| pHILIC | 202.082 | 202.2304 | 18.6 | 0.37 |
| pHILIC | 202.082 | 142.1228 | 18.6 | 0.37 |
| pHILIC | 202.082 | 105.2057 | 18.6 | 0.37 |
| pHILIC | 202.082 | 109.9908 | 18.6 | 0.37 |
| pHILIC | 202.082 | 60.7139 | 18.6 | 0.37 |
| pHILIC | 202.082 | 148.7781 | 18.6 | 0.37 |
| pHILIC | 202.082 | 57.1009 | 18.6 | 0.37 |
| pHILIC | 202.082 | 224.5618 | 18.6 | 0.37 |
| pHILIC | 202.082 | 83.5028 | 18.6 | 0.37 |
| pHILIC | 202.082 | 78.4839 | 18.6 | 0.37 |
| pHILIC | 202.082 | 202.951 | 18.6 | 0.37 |
| pHILIC | 202.082 | 53.2685 | 18.6 | 0.37 |
| pHILIC | 202.082 | 66.3527 | 18.6 | 0.37 |
| pHILIC | 202.082 | 70.8949 | 18.6 | 0.37 |
| pHILIC | 202.082 | 139.995 | 18.6 | 0.37 |
| pHILIC | 202.082 | 196.9313 | 18.6 | 0.37 |
| pHILIC | 202.082 | 77.4872 | 18.6 | 0.37 |
| pHILIC | 202.082 | 48.6522 | 18.6 | 0.37 |
| pHILIC | 202.082 | 102.2263 | 18.6 | 0.37 |
| pHILIC | 202.082 | 45.7814 | 18.5 | 0.37 |
| pHILIC | 202.082 | 54.8116 | 18.5 | 0.37 |
| pHILIC | 202.082 | 53.6736 | 18.5 | 0.37 |
| pHILIC | 202.082 | 71.0434 | 18.5 | 0.37 |
| pHILIC | 202.082 | 173.3451 | 18.5 | 0.37 |
| pHILIC | 202.082 | 136.008 | 18.5 | 0.37 |
| pHILIC | 202.082 | 63.1162 | 18.5 | 0.37 |
| pHILIC | 202.082 | 53.0582 | 18.5 | 0.37 |

| | | | | |
|---|---|---|---|---|
| pHILIC | 202.082 | 46.4912 | 18.5 | 0.37 |
| pHILIC | 202.082 | 75.2111 | 18.5 | 0.37 |
| pHILIC | 202.082 | 189.7269 | 18.5 | 0.37 |
| pHILIC | 202.082 | 91.9148 | 18.5 | 0.37 |
| pHILIC | 202.082 | 194.2735 | 18.5 | 0.37 |
| pHILIC | 202.082 | 122.7512 | 18.5 | 0.37 |
| pHILIC | 202.082 | 52.0511 | 18.5 | 0.36 |
| pHILIC | 202.082 | 74.9015 | 18.5 | 0.36 |
| pHILIC | 202.082 | 59.3752 | 18.5 | 0.36 |
| pHILIC | 202.082 | 168.4009 | 18.5 | 0.36 |
| pHILIC | 202.082 | 52.3389 | 18.5 | 0.36 |
| pHILIC | 202.082 | 90.9577 | 18.5 | 0.36 |
| pHILIC | 202.082 | 77.5667 | 18.5 | 0.36 |
| pHILIC | 202.082 | 109.198 | 18.4 | 0.36 |
| pHILIC | 202.082 | 57.245 | 18.4 | 0.36 |
| pHILIC | 202.082 | 71.781 | 18.4 | 0.36 |
| pHILIC | 202.082 | 74.715 | 18.4 | 0.36 |
| pHILIC | 202.082 | 81.1571 | 18.4 | 0.36 |
| pHILIC | 202.082 | 48.6029 | 18.4 | 0.36 |
| pHILIC | 202.082 | 120.6312 | 18.4 | 0.36 |
| pHILIC | 202.082 | 98.1706 | 18.4 | 0.36 |
| pHILIC | 202.082 | 62.5037 | 18.4 | 0.36 |
| pHILIC | 202.082 | 175.8017 | 18.4 | 0.36 |
| pHILIC | 202.082 | 100.5578 | 18.4 | 0.36 |
| pHILIC | 202.082 | 152.3469 | 18.4 | 0.36 |
| pHILIC | 202.082 | 60.2856 | 18.4 | 0.36 |
| pHILIC | 202.082 | 98.8399 | 18.4 | 0.36 |
| pHILIC | 202.082 | 172.6153 | 18.3 | 0.36 |
| pHILIC | 202.082 | 79.8538 | 18.3 | 0.36 |
| pHILIC | 202.082 | 81.4098 | 18.3 | 0.36 |
| pHILIC | 202.082 | 108.3767 | 18.3 | 0.36 |
| pHILIC | 202.082 | 59.7382 | 18.3 | 0.36 |
| pHILIC | 202.082 | 59.8632 | 18.3 | 0.36 |
| pHILIC | 202.082 | 61.8226 | 18.3 | 0.36 |
| pHILIC | 202.082 | 72.5262 | 18.3 | 0.36 |
| pHILIC | 202.082 | 46.4573 | 18.3 | 0.36 |
| pHILIC | 202.082 | 124.2278 | 18.3 | 0.36 |
| pHILIC | 202.082 | 224.6278 | 18.3 | 0.36 |
| pHILIC | 202.082 | 180.3295 | 18.3 | 0.36 |
| pHILIC | 202.082 | 48.5771 | 18.3 | 0.36 |
| pHILIC | 202.082 | 57.8387 | 18.3 | 0.36 |
| pHILIC | 202.082 | 82.8294 | 18.3 | 0.36 |

| | | | | |
|---|---|---|---|---|
| pHILIC | 202.082 | 125.1252 | 18.3 | 0.36 |
| pHILIC | 202.082 | 132.4776 | 18.3 | 0.36 |
| pHILIC | 202.082 | 84.8232 | 18.3 | 0.36 |
| pHILIC | 202.082 | 86.7385 | 18.3 | 0.36 |
| pHILIC | 202.082 | 47.9977 | 18.3 | 0.36 |
| pHILIC | 202.082 | 53.4214 | 18.3 | 0.36 |
| pHILIC | 202.082 | 130.4092 | 18.3 | 0.36 |
| pHILIC | 202.082 | 163.6053 | 18.2 | 0.36 |
| pHILIC | 202.082 | 152.8999 | 18.2 | 0.36 |
| pHILIC | 202.082 | 50.8953 | 18.2 | 0.36 |
| pHILIC | 202.082 | 159.8049 | 18.2 | 0.36 |
| pHILIC | 202.082 | 56.1509 | 18.2 | 0.36 |
| pHILIC | 202.082 | 46.1035 | 18.2 | 0.36 |
| pHILIC | 202.082 | 163.1678 | 18.2 | 0.36 |
| pHILIC | 202.082 | 49.4212 | 18.2 | 0.36 |
| pHILIC | 202.082 | 204.1259 | 18.2 | 0.36 |
| pHILIC | 202.082 | 46.0068 | 18.2 | 0.36 |
| pHILIC | 202.082 | 87.5483 | 18.2 | 0.36 |
| pHILIC | 202.082 | 47.6106 | 18.2 | 0.36 |
| pHILIC | 202.082 | 69.0367 | 18.2 | 0.36 |
| pHILIC | 202.082 | 203.9683 | 18.2 | 0.36 |
| pHILIC | 202.082 | 69.2833 | 18.2 | 0.36 |
| pHILIC | 202.082 | 156.1406 | 18.2 | 0.36 |
| pHILIC | 202.082 | 61.6961 | 18.2 | 0.36 |
| pHILIC | 202.082 | 77.0924 | 18.1 | 0.36 |
| pHILIC | 202.082 | 91.2177 | 18.1 | 0.36 |
| pHILIC | 202.082 | 49.2451 | 18.1 | 0.36 |
| pHILIC | 202.082 | 133.9635 | 18.1 | 0.36 |
| pHILIC | 202.082 | 82.6403 | 18.1 | 0.36 |
| pHILIC | 202.082 | 96.8864 | 18.1 | 0.36 |
| pHILIC | 202.082 | 77.3055 | 18.1 | 0.36 |
| pHILIC | 202.082 | 178.6329 | 18.1 | 0.36 |
| pHILIC | 202.082 | 68.8731 | 18.1 | 0.36 |
| pHILIC | 202.082 | 49.1721 | 18.1 | 0.36 |
| pHILIC | 202.082 | 127.2953 | 18.1 | 0.36 |
| pHILIC | 202.082 | 69.9048 | 18.1 | 0.36 |
| pHILIC | 202.082 | 100.7185 | 18.1 | 0.36 |
| pHILIC | 202.082 | 48.5213 | 18.1 | 0.36 |
| pHILIC | 202.082 | 217.0683 | 18.1 | 0.36 |
| pHILIC | 202.082 | 91.2722 | 18.1 | 0.36 |
| pHILIC | 202.082 | 106.2195 | 18.1 | 0.36 |
| pHILIC | 202.082 | 221.1919 | 18.1 | 0.36 |

| | | | | |
|---|---|---|---|---|
| pHILIC | 202.082 | 92.3174 | 18.1 | 0.36 |
| pHILIC | 202.082 | 59.048 | 18.1 | 0.36 |
| pHILIC | 202.082 | 45.5954 | 18 | 0.36 |
| pHILIC | 202.082 | 107.3811 | 18 | 0.36 |
| pHILIC | 202.082 | 63.9305 | 18 | 0.36 |
| pHILIC | 202.082 | 220.5506 | 18 | 0.36 |
| pHILIC | 202.082 | 66.5531 | 18 | 0.36 |
| pHILIC | 202.082 | 90.1283 | 18 | 0.36 |
| pHILIC | 202.082 | 62.7337 | 18 | 0.36 |
| pHILIC | 202.082 | 145.3289 | 18 | 0.36 |
| pHILIC | 202.082 | 129.8522 | 18 | 0.36 |
| pHILIC | 202.082 | 58.5341 | 18 | 0.36 |
| pHILIC | 202.082 | 108.1954 | 18 | 0.36 |
| pHILIC | 202.082 | 111.2967 | 18 | 0.36 |
| pHILIC | 202.082 | 102.4837 | 18 | 0.36 |
| pHILIC | 202.082 | 49.2127 | 18 | 0.36 |
| pHILIC | 202.082 | 134.1852 | 18 | 0.35 |
| pHILIC | 202.082 | 47.4814 | 18 | 0.35 |
| pHILIC | 202.082 | 116.3682 | 18 | 0.35 |
| pHILIC | 202.082 | 94.8914 | 18 | 0.35 |
| pHILIC | 202.082 | 69.8431 | 18 | 0.35 |
| pHILIC | 202.082 | 98.4707 | 17.9 | 0.35 |
| pHILIC | 202.082 | 72.9692 | 17.9 | 0.35 |
| pHILIC | 202.082 | 46.8884 | 17.9 | 0.35 |
| pHILIC | 202.082 | 64.1513 | 17.9 | 0.35 |
| pHILIC | 202.082 | 48.0097 | 17.9 | 0.35 |
| pHILIC | 202.082 | 50.9584 | 17.9 | 0.35 |
| pHILIC | 202.082 | 55.132 | 17.9 | 0.35 |
| pHILIC | 202.082 | 65.8607 | 17.9 | 0.35 |
| pHILIC | 202.082 | 110.7274 | 17.9 | 0.35 |
| pHILIC | 202.082 | 70.8625 | 17.9 | 0.35 |
| pHILIC | 202.082 | 70.5355 | 17.9 | 0.35 |
| pHILIC | 202.082 | 71.8077 | 17.9 | 0.35 |
| pHILIC | 202.082 | 174.7127 | 17.9 | 0.35 |
| pHILIC | 202.082 | 50.7042 | 17.9 | 0.35 |
| pHILIC | 202.082 | 92.4582 | 17.9 | 0.35 |
| pHILIC | 202.082 | 47.0071 | 17.9 | 0.35 |
| pHILIC | 202.082 | 66.8718 | 17.9 | 0.35 |
| pHILIC | 202.082 | 49.1288 | 17.9 | 0.35 |
| pHILIC | 202.082 | 51.9229 | 17.9 | 0.35 |
| pHILIC | 202.082 | 59.8331 | 17.8 | 0.35 |
| pHILIC | 202.082 | 58.9457 | 17.8 | 0.35 |

| | | | | |
|---|---|---|---|---|
| pHILIC | 202.082 | 45.8376 | 17.8 | 0.35 |
| pHILIC | 202.082 | 50.6047 | 17.8 | 0.35 |
| pHILIC | 202.082 | 120.7132 | 17.8 | 0.35 |
| pHILIC | 202.082 | 77.4479 | 17.8 | 0.35 |
| pHILIC | 202.082 | 183.8158 | 17.8 | 0.35 |
| pHILIC | 202.082 | 157.5181 | 17.8 | 0.35 |
| pHILIC | 202.082 | 50.1933 | 17.8 | 0.35 |
| pHILIC | 202.082 | 104.549 | 17.8 | 0.35 |
| pHILIC | 202.082 | 69.0469 | 17.8 | 0.35 |
| pHILIC | 202.082 | 74.5655 | 17.8 | 0.35 |
| pHILIC | 202.082 | 51.3721 | 17.8 | 0.35 |
| pHILIC | 202.082 | 62.5942 | 17.7 | 0.35 |
| pHILIC | 202.082 | 111.8211 | 17.7 | 0.35 |
| pHILIC | 202.082 | 121.5162 | 17.7 | 0.35 |
| pHILIC | 202.082 | 51.5713 | 17.7 | 0.35 |
| pHILIC | 202.082 | 196.5493 | 17.7 | 0.35 |
| pHILIC | 202.082 | 45.3293 | 17.7 | 0.35 |
| pHILIC | 202.082 | 139.6456 | 17.7 | 0.35 |
| pHILIC | 202.082 | 50.1394 | 17.7 | 0.35 |
| pHILIC | 202.082 | 47.6223 | 17.7 | 0.35 |
| pHILIC | 202.082 | 47.2127 | 17.7 | 0.35 |
| pHILIC | 202.082 | 102.2038 | 17.7 | 0.35 |
| pHILIC | 202.082 | 55.3308 | 17.7 | 0.35 |
| pHILIC | 202.082 | 56.5674 | 17.7 | 0.35 |
| pHILIC | 202.082 | 58.3244 | 17.7 | 0.35 |
| pHILIC | 202.082 | 200.5688 | 17.7 | 0.35 |
| pHILIC | 202.082 | 66.4301 | 17.7 | 0.35 |
| pHILIC | 202.082 | 70.4964 | 17.7 | 0.35 |
| pHILIC | 202.082 | 54.942 | 17.7 | 0.35 |
| pHILIC | 202.082 | 147.7996 | 17.6 | 0.35 |
| pHILIC | 202.082 | 62.0405 | 17.6 | 0.35 |
| pHILIC | 202.082 | 113.9506 | 17.6 | 0.35 |
| pHILIC | 202.082 | 71.4126 | 17.6 | 0.35 |
| pHILIC | 202.082 | 56.1549 | 17.6 | 0.35 |
| pHILIC | 202.082 | 61.7905 | 17.6 | 0.35 |
| pHILIC | 202.082 | 58.3956 | 17.6 | 0.35 |
| pHILIC | 202.082 | 66.9782 | 17.6 | 0.35 |
| pHILIC | 202.082 | 83.9443 | 17.6 | 0.35 |
| pHILIC | 202.082 | 88.7888 | 17.6 | 0.35 |
| pHILIC | 202.082 | 130.5025 | 17.6 | 0.35 |
| pHILIC | 202.082 | 220.8685 | 17.6 | 0.35 |
| pHILIC | 202.082 | 183.4814 | 17.5 | 0.35 |

| | | | | |
|---|---|---|---|---|
| pHILIC | 202.082 | 97.1315 | 17.5 | 0.35 |
| pHILIC | 202.082 | 185.2062 | 17.5 | 0.35 |
| pHILIC | 202.082 | 92.0507 | 17.5 | 0.35 |
| pHILIC | 202.082 | 64.0673 | 17.5 | 0.35 |
| pHILIC | 202.082 | 47.3485 | 17.5 | 0.35 |
| pHILIC | 202.082 | 163.8194 | 17.5 | 0.35 |
| pHILIC | 202.082 | 80.3858 | 17.5 | 0.35 |
| pHILIC | 202.082 | 138.8604 | 17.5 | 0.35 |
| pHILIC | 202.082 | 104.1835 | 17.5 | 0.35 |
| pHILIC | 202.082 | 121.5553 | 17.5 | 0.35 |
| pHILIC | 202.082 | 122.0169 | 17.5 | 0.35 |
| pHILIC | 202.082 | 84.516 | 17.5 | 0.35 |
| pHILIC | 202.082 | 201.8164 | 17.5 | 0.34 |
| pHILIC | 202.082 | 126.1017 | 17.5 | 0.34 |
| pHILIC | 202.082 | 63.7466 | 17.4 | 0.34 |
| pHILIC | 202.082 | 179.1328 | 17.4 | 0.34 |
| pHILIC | 202.082 | 86.9991 | 17.4 | 0.34 |
| pHILIC | 202.082 | 66.7013 | 17.4 | 0.34 |
| pHILIC | 202.082 | 83.3384 | 17.4 | 0.34 |
| pHILIC | 202.082 | 179.894 | 17.4 | 0.34 |
| pHILIC | 202.082 | 166.289 | 17.4 | 0.34 |
| pHILIC | 202.082 | 102.0502 | 17.4 | 0.34 |
| pHILIC | 202.082 | 201.0304 | 17.4 | 0.34 |
| pHILIC | 202.082 | 122.5626 | 17.4 | 0.34 |
| pHILIC | 202.082 | 59.5817 | 17.4 | 0.34 |
| pHILIC | 202.082 | 77.7522 | 17.4 | 0.34 |
| pHILIC | 202.082 | 68.0659 | 17.4 | 0.34 |
| pHILIC | 202.082 | 56.2187 | 17.4 | 0.34 |
| pHILIC | 202.082 | 122.1466 | 17.4 | 0.34 |
| pHILIC | 202.082 | 55.4429 | 17.4 | 0.34 |
| pHILIC | 202.082 | 54.8663 | 17.4 | 0.34 |
| pHILIC | 202.082 | 116.1069 | 17.4 | 0.34 |
| pHILIC | 202.082 | 118.1936 | 17.3 | 0.34 |
| pHILIC | 202.082 | 45.4077 | 17.3 | 0.34 |
| pHILIC | 202.082 | 47.2809 | 17.3 | 0.34 |
| pHILIC | 202.082 | 145.5707 | 17.3 | 0.34 |
| pHILIC | 202.082 | 97.6183 | 17.3 | 0.34 |
| pHILIC | 202.082 | 70.9185 | 17.3 | 0.34 |
| pHILIC | 202.082 | 211.0797 | 17.3 | 0.34 |
| pHILIC | 202.082 | 62.8843 | 17.3 | 0.34 |
| pHILIC | 202.082 | 48.8989 | 17.3 | 0.34 |
| pHILIC | 202.082 | 72.0415 | 17.3 | 0.34 |

| | | | | |
|---|---|---|---|---|
| pHILIC | 202.082 | 95.6485 | 17.3 | 0.34 |
| pHILIC | 202.082 | 50.8241 | 17.2 | 0.34 |
| pHILIC | 202.082 | 109.1654 | 17.2 | 0.34 |
| pHILIC | 202.082 | 145.5584 | 17.2 | 0.34 |
| pHILIC | 202.082 | 87.3352 | 17.2 | 0.34 |
| pHILIC | 202.082 | 48.3739 | 17.2 | 0.34 |
| pHILIC | 202.082 | 54.6834 | 17.2 | 0.34 |
| pHILIC | 202.082 | 46.3554 | 17.2 | 0.34 |
| pHILIC | 202.082 | 143.4083 | 17.2 | 0.34 |
| pHILIC | 202.082 | 205.5156 | 17.2 | 0.34 |
| pHILIC | 202.082 | 81.4502 | 17.2 | 0.34 |
| pHILIC | 202.082 | 103.4797 | 17.2 | 0.34 |
| pHILIC | 202.082 | 142.5996 | 17.2 | 0.34 |
| pHILIC | 202.082 | 102.6568 | 17.2 | 0.34 |
| pHILIC | 202.082 | 101.8061 | 17.2 | 0.34 |
| pHILIC | 202.082 | 58.6243 | 17.2 | 0.34 |
| pHILIC | 202.082 | 210.2025 | 17.1 | 0.34 |
| pHILIC | 202.082 | 168.6662 | 17.1 | 0.34 |
| pHILIC | 202.082 | 74.6926 | 17.1 | 0.34 |
| pHILIC | 202.082 | 86.0112 | 17.1 | 0.34 |
| pHILIC | 202.082 | 63.8845 | 17.1 | 0.34 |
| pHILIC | 202.082 | 127.6821 | 17.1 | 0.34 |
| pHILIC | 202.082 | 186.8663 | 17.1 | 0.34 |
| pHILIC | 202.082 | 50.2377 | 17.1 | 0.34 |
| pHILIC | 202.082 | 109.8795 | 17.1 | 0.34 |
| pHILIC | 202.082 | 105.4885 | 17.1 | 0.34 |
| pHILIC | 202.082 | 59.6808 | 17.1 | 0.34 |
| pHILIC | 202.082 | 75.7088 | 17 | 0.34 |
| pHILIC | 202.082 | 156.0148 | 17 | 0.34 |
| pHILIC | 202.082 | 95.7989 | 17 | 0.34 |
| pHILIC | 202.082 | 219.1369 | 17 | 0.34 |
| pHILIC | 202.082 | 53.6868 | 17 | 0.34 |
| pHILIC | 202.082 | 70.1976 | 17 | 0.34 |
| pHILIC | 202.082 | 83.1878 | 17 | 0.34 |
| pHILIC | 202.082 | 93.6561 | 17 | 0.34 |
| pHILIC | 202.082 | 70.0158 | 17 | 0.34 |
| pHILIC | 202.082 | 179.4241 | 17 | 0.33 |
| pHILIC | 202.082 | 75.2693 | 17 | 0.33 |
| pHILIC | 202.082 | 101.0867 | 16.9 | 0.33 |
| pHILIC | 202.082 | 53.7483 | 16.9 | 0.33 |
| pHILIC | 202.082 | 136.9954 | 16.9 | 0.33 |
| pHILIC | 202.082 | 138.4115 | 16.9 | 0.33 |

| | | | | |
|---|---|---|---|---|
| pHILIC | 202.082 | 81.2214 | 16.9 | 0.33 |
| pHILIC | 202.082 | 88.3213 | 16.9 | 0.33 |
| pHILIC | 202.082 | 66.7092 | 16.9 | 0.33 |
| pHILIC | 202.082 | 73.8336 | 16.9 | 0.33 |
| pHILIC | 202.082 | 51.6179 | 16.9 | 0.33 |
| pHILIC | 202.082 | 65.6339 | 16.9 | 0.33 |
| pHILIC | 202.082 | 66.9118 | 16.9 | 0.33 |
| pHILIC | 202.082 | 208.0064 | 16.8 | 0.33 |
| pHILIC | 202.082 | 221.6508 | 16.8 | 0.33 |
| pHILIC | 202.082 | 175.4186 | 16.8 | 0.33 |
| pHILIC | 202.082 | 69.4058 | 16.8 | 0.33 |
| pHILIC | 202.082 | 68.7307 | 16.8 | 0.33 |
| pHILIC | 202.082 | 51.4389 | 16.7 | 0.33 |
| pHILIC | 202.082 | 54.4259 | 16.7 | 0.33 |
| pHILIC | 202.082 | 114.3563 | 16.7 | 0.33 |
| pHILIC | 202.082 | 46.7653 | 16.7 | 0.33 |
| pHILIC | 202.082 | 65.2949 | 16.7 | 0.33 |
| pHILIC | 202.082 | 67.5897 | 16.7 | 0.33 |
| pHILIC | 202.082 | 95.1804 | 16.7 | 0.33 |
| pHILIC | 202.082 | 57.0036 | 16.6 | 0.33 |
| pHILIC | 202.082 | 177.1068 | 16.6 | 0.33 |
| pHILIC | 202.082 | 166.9963 | 16.6 | 0.33 |
| pHILIC | 202.082 | 156.802 | 16.6 | 0.33 |
| pHILIC | 202.082 | 80.4701 | 16.6 | 0.33 |
| pHILIC | 202.082 | 95.7673 | 16.6 | 0.33 |
| pHILIC | 202.082 | 72.2827 | 16.6 | 0.33 |
| pHILIC | 202.082 | 60.3494 | 16.6 | 0.33 |
| pHILIC | 202.082 | 70.3745 | 16.6 | 0.33 |
| pHILIC | 202.082 | 114.044 | 16.6 | 0.33 |
| pHILIC | 202.082 | 78.052 | 16.6 | 0.33 |
| pHILIC | 202.082 | 66.718 | 16.6 | 0.33 |
| pHILIC | 202.082 | 55.6666 | 16.5 | 0.33 |
| pHILIC | 202.082 | 93.1912 | 16.5 | 0.33 |
| pHILIC | 202.082 | 91.3681 | 16.5 | 0.33 |
| pHILIC | 202.082 | 73.5208 | 16.5 | 0.33 |
| pHILIC | 202.082 | 114.9885 | 16.4 | 0.32 |
| pHILIC | 202.082 | 203.7857 | 16.4 | 0.32 |
| pHILIC | 202.082 | 96.3914 | 16.4 | 0.32 |
| pHILIC | 202.082 | 113.5283 | 16.4 | 0.32 |
| pHILIC | 202.082 | 123.2692 | 16.4 | 0.32 |
| pHILIC | 202.082 | 218.2021 | 16.4 | 0.32 |
| pHILIC | 202.082 | 207.0948 | 16.4 | 0.32 |

| | | | | |
|---|---|---|---|---|
| pHILIC | 202.082 | 55.8696 | 16.4 | 0.32 |
| pHILIC | 202.082 | 46.2265 | 16.4 | 0.32 |
| pHILIC | 202.082 | 160.6346 | 16.3 | 0.32 |
| pHILIC | 202.082 | 83.8761 | 16.3 | 0.32 |
| pHILIC | 202.082 | 57.6649 | 16.3 | 0.32 |
| pHILIC | 202.082 | 61.4567 | 16.3 | 0.32 |
| pHILIC | 202.082 | 56.7218 | 16.3 | 0.32 |
| pHILIC | 202.082 | 190.5919 | 16.2 | 0.32 |
| pHILIC | 202.082 | 58.4277 | 16.2 | 0.32 |
| pHILIC | 202.082 | 67.6445 | 16.2 | 0.32 |
| pHILIC | 202.082 | 112.7669 | 16.1 | 0.32 |
| pHILIC | 202.082 | 115.8057 | 16.1 | 0.32 |
| pHILIC | 202.082 | 57.4787 | 16.1 | 0.32 |
| pHILIC | 202.082 | 73.997 | 16.1 | 0.32 |
| pHILIC | 202.082 | 61.7103 | 16.1 | 0.32 |
| pHILIC | 202.082 | 194.9516 | 16 | 0.32 |
| pHILIC | 202.082 | 100.5111 | 16 | 0.32 |
| pHILIC | 202.082 | 46.7791 | 16 | 0.32 |
| pHILIC | 202.082 | 59.1883 | 16 | 0.32 |
| pHILIC | 202.082 | 55.6851 | 16 | 0.32 |
| pHILIC | 202.082 | 118.7478 | 16 | 0.32 |
| pHILIC | 202.082 | 140.0981 | 16 | 0.32 |
| pHILIC | 202.082 | 145.7942 | 15.9 | 0.31 |
| pHILIC | 202.082 | 47.3877 | 15.9 | 0.31 |
| pHILIC | 202.082 | 80.5732 | 15.9 | 0.31 |
| pHILIC | 202.082 | 77.1434 | 15.8 | 0.31 |
| pHILIC | 202.082 | 59.2563 | 15.8 | 0.31 |
| pHILIC | 202.082 | 50.3841 | 15.8 | 0.31 |
| pHILIC | 202.082 | 124.7477 | 15.7 | 0.31 |
| pHILIC | 202.082 | 127.8295 | 15.6 | 0.31 |
| pHILIC | 202.082 | 52.5914 | 15.5 | 0.31 |
| pHILIC | 202.082 | 52.405 | 15.4 | 0.3 |
| pHILIC | 202.082 | 61.3574 | 15.4 | 0.3 |
| pHILIC | 202.082 | 65.0293 | 15.3 | 0.3 |
| pHILIC | 202.082 | 48.452 | 15.2 | 0.3 |
| pHILIC | 202.082 | 67.5064 | 15.1 | 0.3 |
| pHILIC | 202.082 | 102.7492 | 15 | 0.3 |
| pHILIC | 202.082 | 212.3595 | 15 | 0.3 |
| pHILIC | 202.082 | 57.8234 | 14.9 | 0.29 |
| pHILIC | 202.082 | 46.1646 | 14.8 | 0.29 |
| pHILIC | 202.082 | 46.2581 | 14.7 | 0.29 |
| pHILIC | 202.082 | 51.4873 | 14.3 | 0.28 |

| | | | | |
|---|---|---|---|---|
| pHILIC | 227.125 | 114.0661 | 29226.1 | 100 |
| pHILIC | 227.125 | 110.0713 | 1982.9 | 6.78 |
| pHILIC | 227.125 | 227.1266 | 719.4 | 2.46 |
| pHILIC | 227.125 | 114.0758 | 249 | 0.85 |
| pHILIC | 227.125 | 93.9334 | 137.5 | 0.47 |
| pHILIC | 227.125 | 96.2494 | 119.9 | 0.41 |
| pHILIC | 227.125 | 176.3078 | 119.2 | 0.41 |
| pHILIC | 227.125 | 94.1183 | 111.7 | 0.38 |
| pHILIC | 227.125 | 227.1752 | 110 | 0.38 |
| pHILIC | 227.125 | 114.0559 | 104.3 | 0.36 |
| pHILIC | 227.125 | 58.2552 | 98.7 | 0.34 |
| pHILIC | 227.125 | 52.5457 | 96.2 | 0.33 |
| pHILIC | 227.125 | 239.849 | 91.8 | 0.31 |
| pHILIC | 227.125 | 170.5533 | 89.9 | 0.31 |
| pHILIC | 227.125 | 109.9413 | 87.2 | 0.3 |
| pHILIC | 227.125 | 53.1136 | 84.9 | 0.29 |
| pHILIC | 227.125 | 70.6219 | 80.6 | 0.28 |
| pHILIC | 227.125 | 241.6226 | 78.9 | 0.27 |
| pHILIC | 227.125 | 91.1148 | 74.3 | 0.25 |
| pHILIC | 227.125 | 94.0205 | 71.8 | 0.25 |
| pHILIC | 227.125 | 241.6633 | 71.6 | 0.25 |
| pHILIC | 227.125 | 155.1405 | 70.6 | 0.24 |
| pHILIC | 227.125 | 60.2039 | 70.2 | 0.24 |
| pHILIC | 227.125 | 185.1039 | 70 | 0.24 |
| pHILIC | 227.125 | 65.9103 | 69 | 0.24 |
| pHILIC | 227.125 | 167.726 | 68.6 | 0.23 |
| pHILIC | 227.125 | 93.4001 | 68.5 | 0.23 |
| pHILIC | 227.125 | 130.1629 | 68.5 | 0.23 |
| pHILIC | 227.125 | 186.6044 | 68.2 | 0.23 |
| pHILIC | 227.125 | 84.3374 | 67.9 | 0.23 |
| pHILIC | 227.125 | 194.4253 | 67.7 | 0.23 |
| pHILIC | 227.125 | 154.4361 | 67.5 | 0.23 |
| pHILIC | 227.125 | 65.5333 | 67.5 | 0.23 |
| pHILIC | 227.125 | 213.6134 | 67.3 | 0.23 |
| pHILIC | 227.125 | 84.2448 | 67 | 0.23 |
| pHILIC | 227.125 | 89.8162 | 66.7 | 0.23 |
| pHILIC | 227.125 | 173.176 | 66.6 | 0.23 |
| pHILIC | 227.125 | 70.6931 | 66.2 | 0.23 |
| pHILIC | 227.125 | 93.7736 | 66.1 | 0.23 |
| pHILIC | 227.125 | 57.2543 | 66 | 0.23 |
| pHILIC | 227.125 | 211.681 | 65.9 | 0.23 |
| pHILIC | 227.125 | 150.846 | 65.7 | 0.22 |

| | | | | |
|---|---|---|---|---|
| pHILIC | 227.125 | 93.6613 | 65.7 | 0.22 |
| pHILIC | 227.125 | 221.522 | 65.5 | 0.22 |
| pHILIC | 227.125 | 93.9234 | 65.4 | 0.22 |
| pHILIC | 227.125 | 74.7793 | 65.2 | 0.22 |
| pHILIC | 227.125 | 167.4179 | 65.2 | 0.22 |
| pHILIC | 227.125 | 57.9798 | 65.1 | 0.22 |
| pHILIC | 227.125 | 157.2406 | 64.9 | 0.22 |
| pHILIC | 227.125 | 167.0536 | 64.7 | 0.22 |
| pHILIC | 227.125 | 113.9258 | 64.6 | 0.22 |
| pHILIC | 227.125 | 246.4995 | 64.6 | 0.22 |
| pHILIC | 227.125 | 93.3727 | 64.4 | 0.22 |
| pHILIC | 227.125 | 108.8063 | 64.4 | 0.22 |
| pHILIC | 227.125 | 153.8681 | 64.3 | 0.22 |
| pHILIC | 227.125 | 101.7983 | 64.2 | 0.22 |
| pHILIC | 227.125 | 87.5977 | 64.2 | 0.22 |
| pHILIC | 227.125 | 135.0828 | 64.1 | 0.22 |
| pHILIC | 227.125 | 89.7473 | 63.9 | 0.22 |
| pHILIC | 227.125 | 220.6829 | 63.9 | 0.22 |
| pHILIC | 227.125 | 185.633 | 63.9 | 0.22 |
| pHILIC | 227.125 | 245.5436 | 63.5 | 0.22 |
| pHILIC | 227.125 | 92.1478 | 63.4 | 0.22 |
| pHILIC | 227.125 | 116.9488 | 63.4 | 0.22 |
| pHILIC | 227.125 | 57.8739 | 63.3 | 0.22 |
| pHILIC | 227.125 | 65.9045 | 63.3 | 0.22 |
| pHILIC | 227.125 | 61.9788 | 63.3 | 0.22 |
| pHILIC | 227.125 | 121.0671 | 63.2 | 0.22 |
| pHILIC | 227.125 | 166.2944 | 63.2 | 0.22 |
| pHILIC | 227.125 | 106.2009 | 63 | 0.22 |
| pHILIC | 227.125 | 78.1495 | 62.6 | 0.21 |
| pHILIC | 227.125 | 78.3038 | 62.6 | 0.21 |
| pHILIC | 227.125 | 56.9444 | 62.6 | 0.21 |
| pHILIC | 227.125 | 154.7262 | 62.5 | 0.21 |
| pHILIC | 227.125 | 94.1066 | 62.4 | 0.21 |
| pHILIC | 227.125 | 181.2241 | 62.3 | 0.21 |
| pHILIC | 227.125 | 183.5324 | 62.3 | 0.21 |
| pHILIC | 227.125 | 190.5394 | 62.1 | 0.21 |
| pHILIC | 227.125 | 128.5205 | 62 | 0.21 |
| pHILIC | 227.125 | 133.3315 | 61.8 | 0.21 |
| pHILIC | 227.125 | 87.0442 | 61.8 | 0.21 |
| pHILIC | 227.125 | 93.8596 | 61.8 | 0.21 |
| pHILIC | 227.125 | 235.0752 | 61.7 | 0.21 |
| pHILIC | 227.125 | 90.8348 | 61.6 | 0.21 |

| | | | | |
|---|---|---|---|---|
| pHILIC | 227.125 | 58.108 | 61.5 | 0.21 |
| pHILIC | 227.125 | 56.1076 | 61.4 | 0.21 |
| pHILIC | 227.125 | 172.0698 | 61.4 | 0.21 |
| pHILIC | 227.125 | 79.6387 | 61.4 | 0.21 |
| pHILIC | 227.125 | 90.4398 | 61.2 | 0.21 |
| pHILIC | 227.125 | 58.3838 | 61.1 | 0.21 |
| pHILIC | 227.125 | 50.4313 | 61 | 0.21 |
| pHILIC | 227.125 | 97.4961 | 60.8 | 0.21 |
| pHILIC | 227.125 | 60.3861 | 60.6 | 0.21 |
| pHILIC | 227.125 | 204.7371 | 60.5 | 0.21 |
| pHILIC | 227.125 | 79.1792 | 60.5 | 0.21 |
| pHILIC | 227.125 | 164.7913 | 60.4 | 0.21 |
| pHILIC | 227.125 | 111.3327 | 60.4 | 0.21 |
| pHILIC | 227.125 | 59.4244 | 60.3 | 0.21 |
| pHILIC | 227.125 | 158.703 | 60.1 | 0.21 |
| pHILIC | 227.125 | 164.4924 | 59.9 | 0.21 |
| pHILIC | 227.125 | 237.9619 | 59.9 | 0.2 |
| pHILIC | 227.125 | 53.0603 | 59.6 | 0.2 |
| pHILIC | 227.125 | 97.2443 | 59.1 | 0.2 |
| pHILIC | 227.125 | 128.8845 | 59 | 0.2 |
| pHILIC | 227.125 | 59.818 | 58.9 | 0.2 |
| pHILIC | 227.125 | 56.9237 | 58.9 | 0.2 |
| pHILIC | 227.125 | 227.2472 | 58.8 | 0.2 |
| pHILIC | 227.125 | 108.8554 | 58.8 | 0.2 |
| pHILIC | 227.125 | 146.4935 | 58.6 | 0.2 |
| pHILIC | 227.125 | 60.8358 | 58.6 | 0.2 |
| pHILIC | 227.125 | 246.3165 | 58.5 | 0.2 |
| pHILIC | 227.125 | 103.4503 | 58.2 | 0.2 |
| pHILIC | 227.125 | 94.1647 | 58.2 | 0.2 |
| pHILIC | 227.125 | 56.8782 | 58.1 | 0.2 |
| pHILIC | 227.125 | 228.2213 | 58 | 0.2 |
| pHILIC | 227.125 | 107.2287 | 58 | 0.2 |
| pHILIC | 227.125 | 69.5371 | 57.9 | 0.2 |
| pHILIC | 227.125 | 91.3221 | 57.9 | 0.2 |
| pHILIC | 227.125 | 96.3961 | 57.9 | 0.2 |
| pHILIC | 227.125 | 195.2203 | 57.9 | 0.2 |
| pHILIC | 227.125 | 222.3257 | 57.8 | 0.2 |
| pHILIC | 227.125 | 51.3796 | 57.8 | 0.2 |
| pHILIC | 227.125 | 115.7596 | 57.7 | 0.2 |
| pHILIC | 227.125 | 65.2937 | 57.5 | 0.2 |
| pHILIC | 227.125 | 136.4734 | 57.4 | 0.2 |
| pHILIC | 227.125 | 93.8966 | 57.3 | 0.2 |

| | | | | |
|---|---|---|---|---|
| pHILIC | 227.125 | 78.1007 | 57.3 | 0.2 |
| pHILIC | 227.125 | 115.0672 | 57.3 | 0.2 |
| pHILIC | 227.125 | 110.6435 | 57.3 | 0.2 |
| pHILIC | 227.125 | 72.9698 | 57.1 | 0.2 |
| pHILIC | 227.125 | 121.3314 | 57 | 0.2 |
| pHILIC | 227.125 | 76.8723 | 56.7 | 0.19 |
| pHILIC | 227.125 | 88.3967 | 56.7 | 0.19 |
| pHILIC | 227.125 | 209.8963 | 56.5 | 0.19 |
| pHILIC | 227.125 | 233.293 | 56.4 | 0.19 |
| pHILIC | 227.125 | 154.4533 | 56.4 | 0.19 |
| pHILIC | 227.125 | 72.5195 | 56.4 | 0.19 |
| pHILIC | 227.125 | 65.8597 | 56.4 | 0.19 |
| pHILIC | 227.125 | 50.8774 | 56.1 | 0.19 |
| pHILIC | 227.125 | 96.9555 | 55.9 | 0.19 |
| pHILIC | 227.125 | 52.6949 | 55.8 | 0.19 |
| pHILIC | 227.125 | 98.8411 | 55.3 | 0.19 |
| pHILIC | 227.125 | 250.2481 | 55.2 | 0.19 |
| pHILIC | 227.125 | 225.0066 | 55.1 | 0.19 |
| pHILIC | 227.125 | 109.4648 | 55.1 | 0.19 |
| pHILIC | 227.125 | 238.501 | 55.1 | 0.19 |
| pHILIC | 227.125 | 74.8815 | 55.1 | 0.19 |
| pHILIC | 227.125 | 159.4997 | 54.7 | 0.19 |
| pHILIC | 227.125 | 75.6007 | 54.6 | 0.19 |
| pHILIC | 227.125 | 200.6643 | 54.6 | 0.19 |
| pHILIC | 227.125 | 120.3129 | 54.5 | 0.19 |
| pHILIC | 227.125 | 57.5398 | 54.5 | 0.19 |
| pHILIC | 227.125 | 98.6186 | 54.5 | 0.19 |
| pHILIC | 227.125 | 95.2233 | 54.5 | 0.19 |
| pHILIC | 227.125 | 60.1233 | 54.5 | 0.19 |
| pHILIC | 227.125 | 73.6931 | 54.3 | 0.19 |
| pHILIC | 227.125 | 201.0679 | 54.3 | 0.19 |
| pHILIC | 227.125 | 224.9171 | 54.2 | 0.19 |
| pHILIC | 227.125 | 109.7568 | 54.1 | 0.19 |
| pHILIC | 227.125 | 63.7725 | 54 | 0.18 |
| pHILIC | 227.125 | 67.1888 | 54 | 0.18 |
| pHILIC | 227.125 | 141.995 | 54 | 0.18 |
| pHILIC | 227.125 | 77.279 | 53.9 | 0.18 |
| pHILIC | 227.125 | 166.7609 | 53.8 | 0.18 |
| pHILIC | 227.125 | 113.8271 | 53.7 | 0.18 |
| pHILIC | 227.125 | 77.3985 | 53.7 | 0.18 |
| pHILIC | 227.125 | 109.8901 | 53.5 | 0.18 |
| pHILIC | 227.125 | 78.4475 | 53.5 | 0.18 |

| | | | | |
|---|---|---|---|---|
| pHILIC | 227.125 | 56.1017 | 53.4 | 0.18 |
| pHILIC | 227.125 | 88.4595 | 53.2 | 0.18 |
| pHILIC | 227.125 | 57.9921 | 53.2 | 0.18 |
| pHILIC | 227.125 | 54.4716 | 53.1 | 0.18 |
| pHILIC | 227.125 | 102.0254 | 53 | 0.18 |
| pHILIC | 227.125 | 111.1974 | 53 | 0.18 |
| pHILIC | 227.125 | 51.6066 | 52.8 | 0.18 |
| pHILIC | 227.125 | 90.0536 | 52.6 | 0.18 |
| pHILIC | 227.125 | 133.7072 | 52.6 | 0.18 |
| pHILIC | 227.125 | 110.2881 | 52.5 | 0.18 |
| pHILIC | 227.125 | 63.4055 | 52.4 | 0.18 |
| pHILIC | 227.125 | 199.0955 | 51.7 | 0.18 |
| pHILIC | 227.125 | 175.9832 | 51.7 | 0.18 |
| pHILIC | 227.125 | 199.9051 | 51.5 | 0.18 |
| pHILIC | 227.125 | 75.0826 | 51.4 | 0.18 |
| pHILIC | 227.125 | 72.6177 | 51.4 | 0.18 |
| pHILIC | 227.125 | 65.0343 | 50.8 | 0.17 |
| pHILIC | 227.125 | 138.5924 | 50.7 | 0.17 |
| pHILIC | 227.125 | 159.9864 | 50.5 | 0.17 |
| pHILIC | 227.125 | 103.0913 | 50.4 | 0.17 |
| pHILIC | 227.125 | 105.7242 | 50.3 | 0.17 |
| pHILIC | 227.125 | 159.2799 | 49.8 | 0.17 |
| pHILIC | 227.125 | 61.8162 | 49.8 | 0.17 |
| pHILIC | 227.125 | 243.9685 | 49.4 | 0.17 |
| pHILIC | 227.125 | 91.9326 | 48.9 | 0.17 |
| pHILIC | 227.125 | 244.0788 | 47.8 | 0.16 |
| pHILIC | 227.125 | 244.417 | 47.5 | 0.16 |
| pHILIC | 227.125 | 170.1868 | 47.1 | 0.16 |
| pHILIC | 227.125 | 224.6609 | 46.7 | 0.16 |
| pHILIC | 227.125 | 247.2778 | 46 | 0.16 |
| pHILIC | 227.125 | 186.7322 | 45.5 | 0.16 |
| pHILIC | 227.125 | 93.5258 | 44 | 0.15 |
| pHILIC | 227.125 | 51.5669 | 43.3 | 0.15 |
| pHILIC | 227.125 | 139.6424 | 42 | 0.14 |
| pHILIC | 227.125 | 132.2297 | 41.7 | 0.14 |
| pHILIC | 227.125 | 60.4639 | 41.6 | 0.14 |
| pHILIC | 227.125 | 234.1939 | 41.5 | 0.14 |
| pHILIC | 227.125 | 147.6647 | 41.5 | 0.14 |
| pHILIC | 227.125 | 94.5201 | 40.8 | 0.14 |
| pHILIC | 227.125 | 67.354 | 40.4 | 0.14 |
| pHILIC | 227.125 | 193.0989 | 40 | 0.14 |
| pHILIC | 227.125 | 94.3358 | 39.9 | 0.14 |

| | | | | |
|---|---|---|---|---|
| pHILIC | 227.125 | 66.6122 | 39.9 | 0.14 |
| pHILIC | 227.125 | 93.9642 | 39.8 | 0.14 |
| pHILIC | 227.125 | 173.4692 | 39.5 | 0.14 |
| pHILIC | 227.125 | 134.8867 | 39.4 | 0.13 |
| pHILIC | 227.125 | 95.3394 | 39.1 | 0.13 |
| pHILIC | 227.125 | 210.8511 | 39.1 | 0.13 |
| pHILIC | 227.125 | 65.2859 | 39 | 0.13 |
| pHILIC | 227.125 | 183.9262 | 39 | 0.13 |
| pHILIC | 227.125 | 50.1325 | 38.8 | 0.13 |
| pHILIC | 227.125 | 51.9459 | 38.8 | 0.13 |
| pHILIC | 227.125 | 218.2626 | 38.8 | 0.13 |
| pHILIC | 227.125 | 201.7909 | 38.7 | 0.13 |
| pHILIC | 227.125 | 164.8515 | 38.6 | 0.13 |
| pHILIC | 227.125 | 132.3839 | 38.5 | 0.13 |
| pHILIC | 227.125 | 53.9837 | 38.4 | 0.13 |
| pHILIC | 227.125 | 248.4953 | 38.3 | 0.13 |
| pHILIC | 227.125 | 69.1353 | 38.2 | 0.13 |
| pHILIC | 227.125 | 194.9621 | 38.2 | 0.13 |
| pHILIC | 227.125 | 130.7032 | 37.9 | 0.13 |
| pHILIC | 227.125 | 93.3605 | 37.9 | 0.13 |
| pHILIC | 227.125 | 105.9193 | 37.8 | 0.13 |
| pHILIC | 227.125 | 240.2971 | 37.6 | 0.13 |
| pHILIC | 227.125 | 79.5389 | 37.6 | 0.13 |
| pHILIC | 227.125 | 167.3373 | 37.5 | 0.13 |
| pHILIC | 227.125 | 68.3283 | 37.5 | 0.13 |
| pHILIC | 227.125 | 69.2608 | 37.4 | 0.13 |
| pHILIC | 227.125 | 65.6448 | 37.3 | 0.13 |
| pHILIC | 227.125 | 51.0901 | 37.3 | 0.13 |
| pHILIC | 227.125 | 53.5009 | 37.3 | 0.13 |
| pHILIC | 227.125 | 104.8177 | 37.2 | 0.13 |
| pHILIC | 227.125 | 227.3704 | 37.2 | 0.13 |
| pHILIC | 227.125 | 113.0347 | 37.1 | 0.13 |
| pHILIC | 227.125 | 59.1054 | 37.1 | 0.13 |
| pHILIC | 227.125 | 96.5939 | 37 | 0.13 |
| pHILIC | 227.125 | 118.6682 | 37 | 0.13 |
| pHILIC | 227.125 | 64.3814 | 37 | 0.13 |
| pHILIC | 227.125 | 157.7867 | 37 | 0.13 |
| pHILIC | 227.125 | 54.9148 | 36.9 | 0.13 |
| pHILIC | 227.125 | 50.4205 | 36.9 | 0.13 |
| pHILIC | 227.125 | 95.1939 | 36.9 | 0.13 |
| pHILIC | 227.125 | 94.6332 | 36.8 | 0.13 |
| pHILIC | 227.125 | 146.3248 | 36.8 | 0.13 |

| | | | | |
|---|---|---|---|---|
| pHILIC | 227.125 | 115.9672 | 36.8 | 0.13 |
| pHILIC | 227.125 | 191.2266 | 36.7 | 0.13 |
| pHILIC | 227.125 | 90.9663 | 36.7 | 0.13 |
| pHILIC | 227.125 | 103.753 | 36.7 | 0.13 |
| pHILIC | 227.125 | 99.156 | 36.7 | 0.13 |
| pHILIC | 227.125 | 132.8315 | 36.7 | 0.13 |
| pHILIC | 227.125 | 117.1142 | 36.6 | 0.13 |
| pHILIC | 227.125 | 107.6437 | 36.6 | 0.13 |
| pHILIC | 227.125 | 71.8888 | 36.6 | 0.13 |
| pHILIC | 227.125 | 222.65 | 36.6 | 0.13 |
| pHILIC | 227.125 | 143.3105 | 36.6 | 0.13 |
| pHILIC | 227.125 | 214.942 | 36.6 | 0.13 |
| pHILIC | 227.125 | 153.9165 | 36.6 | 0.13 |
| pHILIC | 227.125 | 151.0887 | 36.5 | 0.13 |
| pHILIC | 227.125 | 212.3958 | 36.5 | 0.12 |
| pHILIC | 227.125 | 76.7173 | 36.5 | 0.12 |
| pHILIC | 227.125 | 230.6778 | 36.5 | 0.12 |
| pHILIC | 227.125 | 127.1111 | 36.5 | 0.12 |
| pHILIC | 227.125 | 105.9715 | 36.4 | 0.12 |
| pHILIC | 227.125 | 78.9201 | 36.4 | 0.12 |
| pHILIC | 227.125 | 151.2659 | 36.4 | 0.12 |
| pHILIC | 227.125 | 92.0155 | 36.4 | 0.12 |
| pHILIC | 227.125 | 208.1845 | 36.4 | 0.12 |
| pHILIC | 227.125 | 54.4124 | 36.3 | 0.12 |
| pHILIC | 227.125 | 94.6911 | 36.3 | 0.12 |
| pHILIC | 227.125 | 137.6946 | 36.3 | 0.12 |
| pHILIC | 227.125 | 197.5811 | 36.2 | 0.12 |
| pHILIC | 227.125 | 138.7053 | 36.2 | 0.12 |
| pHILIC | 227.125 | 51.5482 | 36.2 | 0.12 |
| pHILIC | 227.125 | 109.6494 | 36.1 | 0.12 |
| pHILIC | 227.125 | 82.0352 | 36.1 | 0.12 |
| pHILIC | 227.125 | 51.4742 | 36 | 0.12 |
| pHILIC | 227.125 | 90.5738 | 36 | 0.12 |
| pHILIC | 227.125 | 94.4364 | 36 | 0.12 |
| pHILIC | 227.125 | 225.3278 | 35.9 | 0.12 |
| pHILIC | 227.125 | 151.9017 | 35.9 | 0.12 |
| pHILIC | 227.125 | 85.7967 | 35.9 | 0.12 |
| pHILIC | 227.125 | 128.9392 | 35.9 | 0.12 |
| pHILIC | 227.125 | 139.481 | 35.9 | 0.12 |
| pHILIC | 227.125 | 62.4964 | 35.9 | 0.12 |
| pHILIC | 227.125 | 51.5082 | 35.9 | 0.12 |
| pHILIC | 227.125 | 55.1399 | 35.8 | 0.12 |

| | | | | |
|---|---|---|---|---|
| pHILIC | 227.125 | 135.5362 | 35.8 | 0.12 |
| pHILIC | 227.125 | 237.3251 | 35.8 | 0.12 |
| pHILIC | 227.125 | 104.4996 | 35.8 | 0.12 |
| pHILIC | 227.125 | 126.2167 | 35.8 | 0.12 |
| pHILIC | 227.125 | 206.8452 | 35.8 | 0.12 |
| pHILIC | 227.125 | 166.2588 | 35.7 | 0.12 |
| pHILIC | 227.125 | 67.4967 | 35.7 | 0.12 |
| pHILIC | 227.125 | 67.7621 | 35.7 | 0.12 |
| pHILIC | 227.125 | 239.1475 | 35.7 | 0.12 |
| pHILIC | 227.125 | 76.0719 | 35.7 | 0.12 |
| pHILIC | 227.125 | 85.1518 | 35.7 | 0.12 |
| pHILIC | 227.125 | 50.499 | 35.7 | 0.12 |
| pHILIC | 227.125 | 184.2511 | 35.7 | 0.12 |
| pHILIC | 227.125 | 53.2025 | 35.7 | 0.12 |
| pHILIC | 227.125 | 73.0281 | 35.7 | 0.12 |
| pHILIC | 227.125 | 131.2749 | 35.6 | 0.12 |
| pHILIC | 227.125 | 99.1675 | 35.6 | 0.12 |
| pHILIC | 227.125 | 93.0539 | 35.6 | 0.12 |
| pHILIC | 227.125 | 82.1952 | 35.6 | 0.12 |
| pHILIC | 227.125 | 95.4609 | 35.6 | 0.12 |
| pHILIC | 227.125 | 92.837 | 35.5 | 0.12 |
| pHILIC | 227.125 | 55.0232 | 35.5 | 0.12 |
| pHILIC | 227.125 | 142.4466 | 35.5 | 0.12 |
| pHILIC | 227.125 | 67.2575 | 35.5 | 0.12 |
| pHILIC | 227.125 | 78.0489 | 35.5 | 0.12 |
| pHILIC | 227.125 | 191.9633 | 35.4 | 0.12 |
| pHILIC | 227.125 | 84.2771 | 35.4 | 0.12 |
| pHILIC | 227.125 | 216.0315 | 35.4 | 0.12 |
| pHILIC | 227.125 | 59.8102 | 35.3 | 0.12 |
| pHILIC | 227.125 | 117.2739 | 35.3 | 0.12 |
| pHILIC | 227.125 | 52.6621 | 35.3 | 0.12 |
| pHILIC | 227.125 | 102.9402 | 35.3 | 0.12 |
| pHILIC | 227.125 | 237.7213 | 35.3 | 0.12 |
| pHILIC | 227.125 | 156.7704 | 35.3 | 0.12 |
| pHILIC | 227.125 | 84.3455 | 35.3 | 0.12 |
| pHILIC | 227.125 | 178.3751 | 35.3 | 0.12 |
| pHILIC | 227.125 | 109.2748 | 35.2 | 0.12 |
| pHILIC | 227.125 | 143.0072 | 35.2 | 0.12 |
| pHILIC | 227.125 | 221.2433 | 35.2 | 0.12 |
| pHILIC | 227.125 | 148.4234 | 35.2 | 0.12 |
| pHILIC | 227.125 | 171.6301 | 35.2 | 0.12 |
| pHILIC | 227.125 | 89.6687 | 35.1 | 0.12 |

| | | | | |
|---|---|---|---|---|
| pHILIC | 227.125 | 92.4466 | 35.1 | 0.12 |
| pHILIC | 227.125 | 71.0362 | 35.1 | 0.12 |
| pHILIC | 227.125 | 86.0417 | 35.1 | 0.12 |
| pHILIC | 227.125 | 143.3988 | 35.1 | 0.12 |
| pHILIC | 227.125 | 52.8926 | 35.1 | 0.12 |
| pHILIC | 227.125 | 51.0252 | 35 | 0.12 |
| pHILIC | 227.125 | 131.1699 | 35 | 0.12 |
| pHILIC | 227.125 | 207.6665 | 35 | 0.12 |
| pHILIC | 227.125 | 60.867 | 35 | 0.12 |
| pHILIC | 227.125 | 51.0089 | 35 | 0.12 |
| pHILIC | 227.125 | 80.7127 | 35 | 0.12 |
| pHILIC | 227.125 | 201.4257 | 35 | 0.12 |
| pHILIC | 227.125 | 88.5296 | 34.9 | 0.12 |
| pHILIC | 227.125 | 61.6642 | 34.9 | 0.12 |
| pHILIC | 227.125 | 69.4989 | 34.9 | 0.12 |
| pHILIC | 227.125 | 85.9477 | 34.9 | 0.12 |
| pHILIC | 227.125 | 78.4851 | 34.9 | 0.12 |
| pHILIC | 227.125 | 94.9937 | 34.8 | 0.12 |
| pHILIC | 227.125 | 101.2227 | 34.8 | 0.12 |
| pHILIC | 227.125 | 103.4743 | 34.8 | 0.12 |
| pHILIC | 227.125 | 248.8096 | 34.8 | 0.12 |
| pHILIC | 227.125 | 61.1556 | 34.8 | 0.12 |
| pHILIC | 227.125 | 56.9342 | 34.8 | 0.12 |
| pHILIC | 227.125 | 69.7964 | 34.8 | 0.12 |
| pHILIC | 227.125 | 227.2082 | 34.8 | 0.12 |
| pHILIC | 227.125 | 96.4977 | 34.8 | 0.12 |
| pHILIC | 227.125 | 90.2627 | 34.8 | 0.12 |
| pHILIC | 227.125 | 59.9249 | 34.7 | 0.12 |
| pHILIC | 227.125 | 125.1004 | 34.7 | 0.12 |
| pHILIC | 227.125 | 96.8371 | 34.7 | 0.12 |
| pHILIC | 227.125 | 90.7007 | 34.7 | 0.12 |
| pHILIC | 227.125 | 102.8764 | 34.7 | 0.12 |
| pHILIC | 227.125 | 127.489 | 34.7 | 0.12 |
| pHILIC | 227.125 | 68.6582 | 34.7 | 0.12 |
| pHILIC | 227.125 | 106.4837 | 34.7 | 0.12 |
| pHILIC | 227.125 | 72.8844 | 34.7 | 0.12 |
| pHILIC | 227.125 | 53.5097 | 34.7 | 0.12 |
| pHILIC | 227.125 | 87.5818 | 34.7 | 0.12 |
| pHILIC | 227.125 | 181.3877 | 34.6 | 0.12 |
| pHILIC | 227.125 | 140.4665 | 34.6 | 0.12 |
| pHILIC | 227.125 | 102.4064 | 34.6 | 0.12 |
| pHILIC | 227.125 | 194.2503 | 34.6 | 0.12 |

| | | | | |
|---|---|---|---|---|
| pHILIC | 227.125 | 91.1788 | 34.6 | 0.12 |
| pHILIC | 227.125 | 64.8356 | 34.6 | 0.12 |
| pHILIC | 227.125 | 54.0183 | 34.6 | 0.12 |
| pHILIC | 227.125 | 56.8016 | 34.6 | 0.12 |
| pHILIC | 227.125 | 143.0818 | 34.6 | 0.12 |
| pHILIC | 227.125 | 118.3301 | 34.6 | 0.12 |
| pHILIC | 227.125 | 189.5322 | 34.6 | 0.12 |
| pHILIC | 227.125 | 168.8987 | 34.6 | 0.12 |
| pHILIC | 227.125 | 75.1982 | 34.5 | 0.12 |
| pHILIC | 227.125 | 163.1313 | 34.5 | 0.12 |
| pHILIC | 227.125 | 96.4336 | 34.5 | 0.12 |
| pHILIC | 227.125 | 120.5214 | 34.5 | 0.12 |
| pHILIC | 227.125 | 63.7797 | 34.5 | 0.12 |
| pHILIC | 227.125 | 53.1977 | 34.5 | 0.12 |
| pHILIC | 227.125 | 71.6901 | 34.4 | 0.12 |
| pHILIC | 227.125 | 180.4299 | 34.4 | 0.12 |
| pHILIC | 227.125 | 221.9938 | 34.4 | 0.12 |
| pHILIC | 227.125 | 182.2735 | 34.4 | 0.12 |
| pHILIC | 227.125 | 52.5167 | 34.4 | 0.12 |
| pHILIC | 227.125 | 139.8289 | 34.4 | 0.12 |
| pHILIC | 227.125 | 127.0786 | 34.4 | 0.12 |
| pHILIC | 227.125 | 96.2755 | 34.3 | 0.12 |
| pHILIC | 227.125 | 115.3891 | 34.3 | 0.12 |
| pHILIC | 227.125 | 251.506 | 34.3 | 0.12 |
| pHILIC | 227.125 | 55.8183 | 34.3 | 0.12 |
| pHILIC | 227.125 | 51.6478 | 34.3 | 0.12 |
| pHILIC | 227.125 | 137.2295 | 34.3 | 0.12 |
| pHILIC | 227.125 | 84.8076 | 34.3 | 0.12 |
| pHILIC | 227.125 | 63.6119 | 34.3 | 0.12 |
| pHILIC | 227.125 | 90.33 | 34.3 | 0.12 |
| pHILIC | 227.125 | 78.7746 | 34.3 | 0.12 |
| pHILIC | 227.125 | 51.9163 | 34.3 | 0.12 |
| pHILIC | 227.125 | 100.0274 | 34.3 | 0.12 |
| pHILIC | 227.125 | 128.8716 | 34.3 | 0.12 |
| pHILIC | 227.125 | 93.5145 | 34.3 | 0.12 |
| pHILIC | 227.125 | 65.4163 | 34.3 | 0.12 |
| pHILIC | 227.125 | 178.4946 | 34.2 | 0.12 |
| pHILIC | 227.125 | 213.883 | 34.2 | 0.12 |
| pHILIC | 227.125 | 86.1517 | 34.2 | 0.12 |
| pHILIC | 227.125 | 87.5861 | 34.2 | 0.12 |
| pHILIC | 227.125 | 130.0425 | 34.2 | 0.12 |
| pHILIC | 227.125 | 71.5944 | 34.2 | 0.12 |

| | | | | |
|---|---|---|---|---|
| pHILIC | 227.125 | 64.7459 | 34.2 | 0.12 |
| pHILIC | 227.125 | 52.4332 | 34.2 | 0.12 |
| pHILIC | 227.125 | 79.3619 | 34.2 | 0.12 |
| pHILIC | 227.125 | 151.3495 | 34.2 | 0.12 |
| pHILIC | 227.125 | 69.6262 | 34.2 | 0.12 |
| pHILIC | 227.125 | 96.518 | 34.2 | 0.12 |
| pHILIC | 227.125 | 221.7423 | 34.2 | 0.12 |
| pHILIC | 227.125 | 150.8621 | 34.2 | 0.12 |
| pHILIC | 227.125 | 78.0692 | 34.1 | 0.12 |
| pHILIC | 227.125 | 237.2701 | 34.1 | 0.12 |
| pHILIC | 227.125 | 72.1433 | 34.1 | 0.12 |
| pHILIC | 227.125 | 65.7832 | 34.1 | 0.12 |
| pHILIC | 227.125 | 63.0204 | 34.1 | 0.12 |
| pHILIC | 227.125 | 150.1428 | 34.1 | 0.12 |
| pHILIC | 227.125 | 167.3124 | 34.1 | 0.12 |
| pHILIC | 227.125 | 203.7132 | 34.1 | 0.12 |
| pHILIC | 227.125 | 82.1407 | 34.1 | 0.12 |
| pHILIC | 227.125 | 71.1127 | 34 | 0.12 |
| pHILIC | 227.125 | 61.2605 | 34 | 0.12 |
| pHILIC | 227.125 | 81.7907 | 34 | 0.12 |
| pHILIC | 227.125 | 53.4347 | 34 | 0.12 |
| pHILIC | 227.125 | 54.9806 | 34 | 0.12 |
| pHILIC | 227.125 | 95.0086 | 34 | 0.12 |
| pHILIC | 227.125 | 75.0132 | 34 | 0.12 |
| pHILIC | 227.125 | 135.4425 | 34 | 0.12 |
| pHILIC | 227.125 | 73.2992 | 34 | 0.12 |
| pHILIC | 227.125 | 61.8352 | 34 | 0.12 |
| pHILIC | 227.125 | 94.4785 | 33.9 | 0.12 |
| pHILIC | 227.125 | 108.6061 | 33.9 | 0.12 |
| pHILIC | 227.125 | 53.7457 | 33.9 | 0.12 |
| pHILIC | 227.125 | 61.6659 | 33.9 | 0.12 |
| pHILIC | 227.125 | 138.6059 | 33.9 | 0.12 |
| pHILIC | 227.125 | 73.1948 | 33.9 | 0.12 |
| pHILIC | 227.125 | 131.2938 | 33.9 | 0.12 |
| pHILIC | 227.125 | 194.3031 | 33.9 | 0.12 |
| pHILIC | 227.125 | 83.2713 | 33.9 | 0.12 |
| pHILIC | 227.125 | 58.521 | 33.9 | 0.12 |
| pHILIC | 227.125 | 53.0992 | 33.9 | 0.12 |
| pHILIC | 227.125 | 124.743 | 33.9 | 0.12 |
| pHILIC | 227.125 | 199.7017 | 33.9 | 0.12 |
| pHILIC | 227.125 | 53.5986 | 33.8 | 0.12 |
| pHILIC | 227.125 | 116.324 | 33.8 | 0.12 |

| | | | | |
|---|---|---|---|---|
| pHILIC | 227.125 | 93.69 | 33.8 | 0.12 |
| pHILIC | 227.125 | 73.7659 | 33.8 | 0.12 |
| pHILIC | 227.125 | 157.686 | 33.8 | 0.12 |
| pHILIC | 227.125 | 81.235 | 33.8 | 0.12 |
| pHILIC | 227.125 | 110.7375 | 33.8 | 0.12 |
| pHILIC | 227.125 | 187.4807 | 33.8 | 0.12 |
| pHILIC | 227.125 | 251.3223 | 33.7 | 0.12 |
| pHILIC | 227.125 | 185.485 | 33.7 | 0.12 |
| pHILIC | 227.125 | 148.0038 | 33.7 | 0.12 |
| pHILIC | 227.125 | 206.766 | 33.7 | 0.12 |
| pHILIC | 227.125 | 142.3862 | 33.7 | 0.12 |
| pHILIC | 227.125 | 157.9254 | 33.7 | 0.12 |
| pHILIC | 227.125 | 111.6284 | 33.7 | 0.12 |
| pHILIC | 227.125 | 68.1718 | 33.7 | 0.12 |
| pHILIC | 227.125 | 113.06 | 33.7 | 0.12 |
| pHILIC | 227.125 | 89.6373 | 33.7 | 0.12 |
| pHILIC | 227.125 | 99.9135 | 33.7 | 0.12 |
| pHILIC | 227.125 | 129.7306 | 33.7 | 0.12 |
| pHILIC | 227.125 | 74.1541 | 33.7 | 0.12 |
| pHILIC | 227.125 | 251.9682 | 33.7 | 0.12 |
| pHILIC | 227.125 | 85.0266 | 33.7 | 0.12 |
| pHILIC | 227.125 | 58.9291 | 33.7 | 0.12 |
| pHILIC | 227.125 | 86.0716 | 33.7 | 0.12 |
| pHILIC | 227.125 | 60.4983 | 33.7 | 0.12 |
| pHILIC | 227.125 | 63.9725 | 33.7 | 0.12 |
| pHILIC | 227.125 | 57.141 | 33.6 | 0.12 |
| pHILIC | 227.125 | 88.6904 | 33.6 | 0.12 |
| pHILIC | 227.125 | 210.583 | 33.6 | 0.12 |
| pHILIC | 227.125 | 95.5884 | 33.6 | 0.12 |
| pHILIC | 227.125 | 98.1785 | 33.6 | 0.12 |
| pHILIC | 227.125 | 129.4645 | 33.6 | 0.12 |
| pHILIC | 227.125 | 91.4069 | 33.6 | 0.11 |
| pHILIC | 227.125 | 155.2834 | 33.6 | 0.11 |
| pHILIC | 227.125 | 88.0181 | 33.6 | 0.11 |
| pHILIC | 227.125 | 92.0594 | 33.6 | 0.11 |
| pHILIC | 227.125 | 77.8822 | 33.6 | 0.11 |
| pHILIC | 227.125 | 94.8448 | 33.6 | 0.11 |
| pHILIC | 227.125 | 157.1009 | 33.6 | 0.11 |
| pHILIC | 227.125 | 114.759 | 33.6 | 0.11 |
| pHILIC | 227.125 | 221.0968 | 33.6 | 0.11 |
| pHILIC | 227.125 | 57.9894 | 33.5 | 0.11 |
| pHILIC | 227.125 | 196.6162 | 33.5 | 0.11 |

| | | | | |
|---|---|---|---|---|
| pHILIC | 227.125 | 62.6021 | 33.5 | 0.11 |
| pHILIC | 227.125 | 65.1 | 33.5 | 0.11 |
| pHILIC | 227.125 | 87.6805 | 33.5 | 0.11 |
| pHILIC | 227.125 | 248.259 | 33.5 | 0.11 |
| pHILIC | 227.125 | 119.6522 | 33.5 | 0.11 |
| pHILIC | 227.125 | 51.0918 | 33.5 | 0.11 |
| pHILIC | 227.125 | 92.0429 | 33.5 | 0.11 |
| pHILIC | 227.125 | 250.1734 | 33.5 | 0.11 |
| pHILIC | 227.125 | 132.1899 | 33.5 | 0.11 |
| pHILIC | 227.125 | 98.4219 | 33.5 | 0.11 |
| pHILIC | 227.125 | 125.3551 | 33.5 | 0.11 |
| pHILIC | 227.125 | 78.6275 | 33.5 | 0.11 |
| pHILIC | 227.125 | 79.173 | 33.5 | 0.11 |
| pHILIC | 227.125 | 50.7801 | 33.4 | 0.11 |
| pHILIC | 227.125 | 54.9918 | 33.4 | 0.11 |
| pHILIC | 227.125 | 97.0382 | 33.4 | 0.11 |
| pHILIC | 227.125 | 62.3662 | 33.4 | 0.11 |
| pHILIC | 227.125 | 92.6534 | 33.4 | 0.11 |
| pHILIC | 227.125 | 90.6852 | 33.4 | 0.11 |
| pHILIC | 227.125 | 112.1334 | 33.4 | 0.11 |
| pHILIC | 227.125 | 58.8136 | 33.4 | 0.11 |
| pHILIC | 227.125 | 141.39 | 33.4 | 0.11 |
| pHILIC | 227.125 | 242.3076 | 33.4 | 0.11 |
| pHILIC | 227.125 | 226.3714 | 33.4 | 0.11 |
| pHILIC | 227.125 | 62.303 | 33.4 | 0.11 |
| pHILIC | 227.125 | 173.9402 | 33.4 | 0.11 |
| pHILIC | 227.125 | 72.2776 | 33.4 | 0.11 |
| pHILIC | 227.125 | 114.8157 | 33.3 | 0.11 |
| pHILIC | 227.125 | 52.2172 | 33.3 | 0.11 |
| pHILIC | 227.125 | 50.0256 | 33.3 | 0.11 |
| pHILIC | 227.125 | 114.4779 | 33.3 | 0.11 |
| pHILIC | 227.125 | 127.7846 | 33.3 | 0.11 |
| pHILIC | 227.125 | 148.7141 | 33.3 | 0.11 |
| pHILIC | 227.125 | 51.0447 | 33.3 | 0.11 |
| pHILIC | 227.125 | 52.8697 | 33.2 | 0.11 |
| pHILIC | 227.125 | 67.6485 | 33.2 | 0.11 |
| pHILIC | 227.125 | 56.0122 | 33.2 | 0.11 |
| pHILIC | 227.125 | 62.6721 | 33.2 | 0.11 |
| pHILIC | 227.125 | 103.9871 | 33.2 | 0.11 |
| pHILIC | 227.125 | 55.9322 | 33.2 | 0.11 |
| pHILIC | 227.125 | 162.5005 | 33.2 | 0.11 |
| pHILIC | 227.125 | 98.4588 | 33.2 | 0.11 |

| | | | | |
|---|---|---|---|---|
| pHILIC | 227.125 | 71.8141 | 33.2 | 0.11 |
| pHILIC | 227.125 | 73.3823 | 33.1 | 0.11 |
| pHILIC | 227.125 | 70.7864 | 33.1 | 0.11 |
| pHILIC | 227.125 | 147.2978 | 33.1 | 0.11 |
| pHILIC | 227.125 | 144.4653 | 33.1 | 0.11 |
| pHILIC | 227.125 | 184.0464 | 33.1 | 0.11 |
| pHILIC | 227.125 | 63.8427 | 33.1 | 0.11 |
| pHILIC | 227.125 | 187.4324 | 33.1 | 0.11 |
| pHILIC | 227.125 | 193.3125 | 33.1 | 0.11 |
| pHILIC | 227.125 | 236.8787 | 33.1 | 0.11 |
| pHILIC | 227.125 | 108.5663 | 33.1 | 0.11 |
| pHILIC | 227.125 | 162.9114 | 33.1 | 0.11 |
| pHILIC | 227.125 | 193.4966 | 33.1 | 0.11 |
| pHILIC | 227.125 | 169.9392 | 33.1 | 0.11 |
| pHILIC | 227.125 | 178.5506 | 33.1 | 0.11 |
| pHILIC | 227.125 | 83.5474 | 33.1 | 0.11 |
| pHILIC | 227.125 | 58.6228 | 33.1 | 0.11 |
| pHILIC | 227.125 | 83.9432 | 33.1 | 0.11 |
| pHILIC | 227.125 | 147.6483 | 33.1 | 0.11 |
| pHILIC | 227.125 | 243.1894 | 33 | 0.11 |
| pHILIC | 227.125 | 64.0086 | 33 | 0.11 |
| pHILIC | 227.125 | 88.2259 | 33 | 0.11 |
| pHILIC | 227.125 | 181.3627 | 33 | 0.11 |
| pHILIC | 227.125 | 56.2103 | 33 | 0.11 |
| pHILIC | 227.125 | 135.8416 | 33 | 0.11 |
| pHILIC | 227.125 | 61.2512 | 33 | 0.11 |
| pHILIC | 227.125 | 132.8437 | 33 | 0.11 |
| pHILIC | 227.125 | 69.4784 | 33 | 0.11 |
| pHILIC | 227.125 | 82.1811 | 33 | 0.11 |
| pHILIC | 227.125 | 109.562 | 33 | 0.11 |
| pHILIC | 227.125 | 95.1866 | 33 | 0.11 |
| pHILIC | 227.125 | 129.1794 | 32.9 | 0.11 |
| pHILIC | 227.125 | 68.9929 | 32.9 | 0.11 |
| pHILIC | 227.125 | 60.1633 | 32.9 | 0.11 |
| pHILIC | 227.125 | 99.1036 | 32.9 | 0.11 |
| pHILIC | 227.125 | 77.8411 | 32.9 | 0.11 |
| pHILIC | 227.125 | 51.5242 | 32.9 | 0.11 |
| pHILIC | 227.125 | 168.2287 | 32.9 | 0.11 |
| pHILIC | 227.125 | 184.4285 | 32.9 | 0.11 |
| pHILIC | 227.125 | 51.7332 | 32.9 | 0.11 |
| pHILIC | 227.125 | 55.0805 | 32.9 | 0.11 |
| pHILIC | 227.125 | 212.0203 | 32.9 | 0.11 |

| | | | | |
|---|---|---|---|---|
| pHILIC | 227.125 | 177.1295 | 32.9 | 0.11 |
| pHILIC | 227.125 | 52.0218 | 32.9 | 0.11 |
| pHILIC | 227.125 | 196.4771 | 32.8 | 0.11 |
| pHILIC | 227.125 | 51.5184 | 32.8 | 0.11 |
| pHILIC | 227.125 | 175.348 | 32.8 | 0.11 |
| pHILIC | 227.125 | 202.2969 | 32.8 | 0.11 |
| pHILIC | 227.125 | 60.541 | 32.8 | 0.11 |
| pHILIC | 227.125 | 70.9246 | 32.8 | 0.11 |
| pHILIC | 227.125 | 132.8096 | 32.8 | 0.11 |
| pHILIC | 227.125 | 97.8286 | 32.8 | 0.11 |
| pHILIC | 227.125 | 203.1344 | 32.8 | 0.11 |
| pHILIC | 227.125 | 115.3229 | 32.8 | 0.11 |
| pHILIC | 227.125 | 221.4142 | 32.8 | 0.11 |
| pHILIC | 227.125 | 242.0397 | 32.8 | 0.11 |
| pHILIC | 227.125 | 208.8463 | 32.8 | 0.11 |
| pHILIC | 227.125 | 119.7082 | 32.8 | 0.11 |
| pHILIC | 227.125 | 112.3105 | 32.8 | 0.11 |
| pHILIC | 227.125 | 53.6313 | 32.7 | 0.11 |
| pHILIC | 227.125 | 93.4203 | 32.7 | 0.11 |
| pHILIC | 227.125 | 60.3758 | 32.7 | 0.11 |
| pHILIC | 227.125 | 149.2505 | 32.7 | 0.11 |
| pHILIC | 227.125 | 117.2968 | 32.7 | 0.11 |
| pHILIC | 227.125 | 59.9896 | 32.7 | 0.11 |
| pHILIC | 227.125 | 64.6499 | 32.7 | 0.11 |
| pHILIC | 227.125 | 139.3339 | 32.7 | 0.11 |
| pHILIC | 227.125 | 112.9204 | 32.7 | 0.11 |
| pHILIC | 227.125 | 89.6515 | 32.7 | 0.11 |
| pHILIC | 227.125 | 120.731 | 32.7 | 0.11 |
| pHILIC | 227.125 | 55.9704 | 32.7 | 0.11 |
| pHILIC | 227.125 | 144.4841 | 32.7 | 0.11 |
| pHILIC | 227.125 | 66.2702 | 32.7 | 0.11 |
| pHILIC | 227.125 | 58.5413 | 32.7 | 0.11 |
| pHILIC | 227.125 | 73.2606 | 32.7 | 0.11 |
| pHILIC | 227.125 | 59.5342 | 32.6 | 0.11 |
| pHILIC | 227.125 | 51.6784 | 32.6 | 0.11 |
| pHILIC | 227.125 | 173.5319 | 32.6 | 0.11 |
| pHILIC | 227.125 | 51.5148 | 32.6 | 0.11 |
| pHILIC | 227.125 | 76.2287 | 32.6 | 0.11 |
| pHILIC | 227.125 | 112.2035 | 32.6 | 0.11 |
| pHILIC | 227.125 | 68.4812 | 32.6 | 0.11 |
| pHILIC | 227.125 | 125.8032 | 32.6 | 0.11 |
| pHILIC | 227.125 | 137.4277 | 32.6 | 0.11 |

| | | | | |
|---|---|---|---|---|
| pHILIC | 227.125 | 60.4321 | 32.6 | 0.11 |
| pHILIC | 227.125 | 142.088 | 32.6 | 0.11 |
| pHILIC | 227.125 | 181.1668 | 32.6 | 0.11 |
| pHILIC | 227.125 | 62.3126 | 32.6 | 0.11 |
| pHILIC | 227.125 | 210.0712 | 32.6 | 0.11 |
| pHILIC | 227.125 | 51.1499 | 32.6 | 0.11 |
| pHILIC | 227.125 | 114.674 | 32.6 | 0.11 |
| pHILIC | 227.125 | 57.2232 | 32.6 | 0.11 |
| pHILIC | 227.125 | 53.277 | 32.6 | 0.11 |
| pHILIC | 227.125 | 57.258 | 32.6 | 0.11 |
| pHILIC | 227.125 | 65.8888 | 32.6 | 0.11 |
| pHILIC | 227.125 | 133.5212 | 32.6 | 0.11 |
| pHILIC | 227.125 | 91.9412 | 32.5 | 0.11 |
| pHILIC | 227.125 | 71.162 | 32.5 | 0.11 |
| pHILIC | 227.125 | 51.1339 | 32.5 | 0.11 |
| pHILIC | 227.125 | 51.5503 | 32.5 | 0.11 |
| pHILIC | 227.125 | 170.2611 | 32.5 | 0.11 |
| pHILIC | 227.125 | 65.3942 | 32.5 | 0.11 |
| pHILIC | 227.125 | 92.5564 | 32.5 | 0.11 |
| pHILIC | 227.125 | 184.1988 | 32.5 | 0.11 |
| pHILIC | 227.125 | 88.494 | 32.5 | 0.11 |
| pHILIC | 227.125 | 166.6162 | 32.5 | 0.11 |
| pHILIC | 227.125 | 143.8693 | 32.5 | 0.11 |
| pHILIC | 227.125 | 72.094 | 32.5 | 0.11 |
| pHILIC | 227.125 | 134.8974 | 32.5 | 0.11 |
| pHILIC | 227.125 | 58.7945 | 32.5 | 0.11 |
| pHILIC | 227.125 | 53.8089 | 32.5 | 0.11 |
| pHILIC | 227.125 | 61.8887 | 32.5 | 0.11 |
| pHILIC | 227.125 | 62.117 | 32.5 | 0.11 |
| pHILIC | 227.125 | 53.5661 | 32.5 | 0.11 |
| pHILIC | 227.125 | 136.3016 | 32.5 | 0.11 |
| pHILIC | 227.125 | 77.4672 | 32.5 | 0.11 |
| pHILIC | 227.125 | 56.9101 | 32.5 | 0.11 |
| pHILIC | 227.125 | 78.8306 | 32.5 | 0.11 |
| pHILIC | 227.125 | 139.6314 | 32.5 | 0.11 |
| pHILIC | 227.125 | 209.1913 | 32.4 | 0.11 |
| pHILIC | 227.125 | 105.4529 | 32.4 | 0.11 |
| pHILIC | 227.125 | 193.6743 | 32.4 | 0.11 |
| pHILIC | 227.125 | 209.3625 | 32.4 | 0.11 |
| pHILIC | 227.125 | 65.4218 | 32.4 | 0.11 |
| pHILIC | 227.125 | 94.6122 | 32.4 | 0.11 |
| pHILIC | 227.125 | 51.9912 | 32.4 | 0.11 |

| | | | | |
|---|---|---|---|---|
| pHILIC | 227.125 | 173.577 | 32.4 | 0.11 |
| pHILIC | 227.125 | 152.3928 | 32.4 | 0.11 |
| pHILIC | 227.125 | 61.4093 | 32.4 | 0.11 |
| pHILIC | 227.125 | 54.1584 | 32.4 | 0.11 |
| pHILIC | 227.125 | 232.6046 | 32.4 | 0.11 |
| pHILIC | 227.125 | 71.5647 | 32.4 | 0.11 |
| pHILIC | 227.125 | 120.5444 | 32.4 | 0.11 |
| pHILIC | 227.125 | 214.4475 | 32.4 | 0.11 |
| pHILIC | 227.125 | 118.7461 | 32.4 | 0.11 |
| pHILIC | 227.125 | 87.4124 | 32.4 | 0.11 |
| pHILIC | 227.125 | 107.1449 | 32.4 | 0.11 |
| pHILIC | 227.125 | 84.0101 | 32.4 | 0.11 |
| pHILIC | 227.125 | 230.39 | 32.4 | 0.11 |
| pHILIC | 227.125 | 223.4586 | 32.4 | 0.11 |
| pHILIC | 227.125 | 74.5966 | 32.4 | 0.11 |
| pHILIC | 227.125 | 64.666 | 32.4 | 0.11 |
| pHILIC | 227.125 | 145.8528 | 32.4 | 0.11 |
| pHILIC | 227.125 | 57.4267 | 32.4 | 0.11 |
| pHILIC | 227.125 | 206.6592 | 32.4 | 0.11 |
| pHILIC | 227.125 | 163.2964 | 32.4 | 0.11 |
| pHILIC | 227.125 | 100.2439 | 32.4 | 0.11 |
| pHILIC | 227.125 | 50.2578 | 32.3 | 0.11 |
| pHILIC | 227.125 | 186.5123 | 32.3 | 0.11 |
| pHILIC | 227.125 | 60.6536 | 32.3 | 0.11 |
| pHILIC | 227.125 | 91.7255 | 32.3 | 0.11 |
| pHILIC | 227.125 | 58.885 | 32.3 | 0.11 |
| pHILIC | 227.125 | 163.6022 | 32.3 | 0.11 |
| pHILIC | 227.125 | 109.4135 | 32.3 | 0.11 |
| pHILIC | 227.125 | 50.6498 | 32.3 | 0.11 |
| pHILIC | 227.125 | 238.714 | 32.2 | 0.11 |
| pHILIC | 227.125 | 95.2974 | 32.2 | 0.11 |
| pHILIC | 227.125 | 100.2369 | 32.2 | 0.11 |
| pHILIC | 227.125 | 218.6195 | 32.2 | 0.11 |
| pHILIC | 227.125 | 64.4611 | 32.2 | 0.11 |
| pHILIC | 227.125 | 58.6055 | 32.2 | 0.11 |
| pHILIC | 227.125 | 248.9309 | 32.2 | 0.11 |
| pHILIC | 227.125 | 70.0256 | 32.2 | 0.11 |
| pHILIC | 227.125 | 79.7791 | 32.2 | 0.11 |
| pHILIC | 227.125 | 58.4128 | 32.2 | 0.11 |
| pHILIC | 227.125 | 50.8874 | 32.2 | 0.11 |
| pHILIC | 227.125 | 217.078 | 32.2 | 0.11 |
| pHILIC | 227.125 | 106.2889 | 32.1 | 0.11 |

| | | | | |
|---|---|---|---|---|
| pHILIC | 227.125 | 126.6635 | 32.1 | 0.11 |
| pHILIC | 227.125 | 204.5135 | 32.1 | 0.11 |
| pHILIC | 227.125 | 79.0235 | 32.1 | 0.11 |
| pHILIC | 227.125 | 66.6183 | 32.1 | 0.11 |
| pHILIC | 227.125 | 54.788 | 32.1 | 0.11 |
| pHILIC | 227.125 | 199.6009 | 32.1 | 0.11 |
| pHILIC | 227.125 | 78.3128 | 32.1 | 0.11 |
| pHILIC | 227.125 | 54.2654 | 32.1 | 0.11 |
| pHILIC | 227.125 | 106.9737 | 32.1 | 0.11 |
| pHILIC | 227.125 | 146.5372 | 32.1 | 0.11 |
| pHILIC | 227.125 | 99.6775 | 32.1 | 0.11 |
| pHILIC | 227.125 | 130.3338 | 32.1 | 0.11 |
| pHILIC | 227.125 | 64.5235 | 32.1 | 0.11 |
| pHILIC | 227.125 | 66.1435 | 32.1 | 0.11 |
| pHILIC | 227.125 | 81.7001 | 32 | 0.11 |
| pHILIC | 227.125 | 85.6808 | 32 | 0.11 |
| pHILIC | 227.125 | 156.4212 | 32 | 0.11 |
| pHILIC | 227.125 | 144.44 | 32 | 0.11 |
| pHILIC | 227.125 | 65.1774 | 32 | 0.11 |
| pHILIC | 227.125 | 65.2897 | 32 | 0.11 |
| pHILIC | 227.125 | 74.7422 | 32 | 0.11 |
| pHILIC | 227.125 | 144.4991 | 32 | 0.11 |
| pHILIC | 227.125 | 104.7164 | 32 | 0.11 |
| pHILIC | 227.125 | 73.5011 | 32 | 0.11 |
| pHILIC | 227.125 | 83.2384 | 32 | 0.11 |
| pHILIC | 227.125 | 66.9788 | 32 | 0.11 |
| pHILIC | 227.125 | 64.9052 | 32 | 0.11 |
| pHILIC | 227.125 | 57.3866 | 32 | 0.11 |
| pHILIC | 227.125 | 51.5025 | 32 | 0.11 |
| pHILIC | 227.125 | 210.4075 | 32 | 0.11 |
| pHILIC | 227.125 | 250.3891 | 32 | 0.11 |
| pHILIC | 227.125 | 209.8031 | 31.9 | 0.11 |
| pHILIC | 227.125 | 120.4879 | 31.9 | 0.11 |
| pHILIC | 227.125 | 86.4232 | 31.9 | 0.11 |
| pHILIC | 227.125 | 152.3147 | 31.9 | 0.11 |
| pHILIC | 227.125 | 240.128 | 31.9 | 0.11 |
| pHILIC | 227.125 | 140.0011 | 31.9 | 0.11 |
| pHILIC | 227.125 | 63.5774 | 31.9 | 0.11 |
| pHILIC | 227.125 | 78.9671 | 31.9 | 0.11 |
| pHILIC | 227.125 | 116.8116 | 31.9 | 0.11 |
| pHILIC | 227.125 | 66.6491 | 31.9 | 0.11 |
| pHILIC | 227.125 | 243.3641 | 31.9 | 0.11 |

| | | | | |
|---|---|---|---|---|
| pHILIC | 227.125 | 58.8671 | 31.9 | 0.11 |
| pHILIC | 227.125 | 73.9491 | 31.9 | 0.11 |
| pHILIC | 227.125 | 89.3431 | 31.9 | 0.11 |
| pHILIC | 227.125 | 138.3289 | 31.9 | 0.11 |
| pHILIC | 227.125 | 145.5569 | 31.8 | 0.11 |
| pHILIC | 227.125 | 88.049 | 31.8 | 0.11 |
| pHILIC | 227.125 | 222.1476 | 31.8 | 0.11 |
| pHILIC | 227.125 | 74.4812 | 31.8 | 0.11 |
| pHILIC | 227.125 | 160.4409 | 31.8 | 0.11 |
| pHILIC | 227.125 | 70.8624 | 31.8 | 0.11 |
| pHILIC | 227.125 | 61.5283 | 31.8 | 0.11 |
| pHILIC | 227.125 | 172.7906 | 31.8 | 0.11 |
| pHILIC | 227.125 | 84.999 | 31.8 | 0.11 |
| pHILIC | 227.125 | 181.9311 | 31.8 | 0.11 |
| pHILIC | 227.125 | 110.0896 | 31.8 | 0.11 |
| pHILIC | 227.125 | 107.1608 | 31.8 | 0.11 |
| pHILIC | 227.125 | 223.4863 | 31.8 | 0.11 |
| pHILIC | 227.125 | 53.3955 | 31.8 | 0.11 |
| pHILIC | 227.125 | 62.3927 | 31.8 | 0.11 |
| pHILIC | 227.125 | 50.7101 | 31.8 | 0.11 |
| pHILIC | 227.125 | 83.4471 | 31.7 | 0.11 |
| pHILIC | 227.125 | 251.0145 | 31.7 | 0.11 |
| pHILIC | 227.125 | 216.1259 | 31.7 | 0.11 |
| pHILIC | 227.125 | 51.5168 | 31.7 | 0.11 |
| pHILIC | 227.125 | 200.4159 | 31.7 | 0.11 |
| pHILIC | 227.125 | 104.2474 | 31.7 | 0.11 |
| pHILIC | 227.125 | 78.5782 | 31.7 | 0.11 |
| pHILIC | 227.125 | 205.4331 | 31.7 | 0.11 |
| pHILIC | 227.125 | 57.7915 | 31.7 | 0.11 |
| pHILIC | 227.125 | 50.3683 | 31.7 | 0.11 |
| pHILIC | 227.125 | 51.5554 | 31.7 | 0.11 |
| pHILIC | 227.125 | 99.9779 | 31.7 | 0.11 |
| pHILIC | 227.125 | 78.0572 | 31.7 | 0.11 |
| pHILIC | 227.125 | 83.9475 | 31.7 | 0.11 |
| pHILIC | 227.125 | 70.3213 | 31.7 | 0.11 |
| pHILIC | 227.125 | 85.7081 | 31.7 | 0.11 |
| pHILIC | 227.125 | 59.313 | 31.7 | 0.11 |
| pHILIC | 227.125 | 156.8125 | 31.7 | 0.11 |
| pHILIC | 227.125 | 66.305 | 31.7 | 0.11 |
| pHILIC | 227.125 | 63.7239 | 31.7 | 0.11 |
| pHILIC | 227.125 | 209.4832 | 31.7 | 0.11 |
| pHILIC | 227.125 | 81.0447 | 31.7 | 0.11 |

| | | | | |
|---|---|---|---|---|
| pHILIC | 227.125 | 51.0973 | 31.7 | 0.11 |
| pHILIC | 227.125 | 189.4714 | 31.6 | 0.11 |
| pHILIC | 227.125 | 93.2981 | 31.6 | 0.11 |
| pHILIC | 227.125 | 87.2991 | 31.6 | 0.11 |
| pHILIC | 227.125 | 55.7252 | 31.6 | 0.11 |
| pHILIC | 227.125 | 99.952 | 31.6 | 0.11 |
| pHILIC | 227.125 | 100.2488 | 31.6 | 0.11 |
| pHILIC | 227.125 | 70.5458 | 31.6 | 0.11 |
| pHILIC | 227.125 | 80.8235 | 31.6 | 0.11 |
| pHILIC | 227.125 | 82.0128 | 31.6 | 0.11 |
| pHILIC | 227.125 | 70.3453 | 31.6 | 0.11 |
| pHILIC | 227.125 | 174.5488 | 31.6 | 0.11 |
| pHILIC | 227.125 | 73.3188 | 31.6 | 0.11 |
| pHILIC | 227.125 | 103.8377 | 31.6 | 0.11 |
| pHILIC | 227.125 | 69.0002 | 31.6 | 0.11 |
| pHILIC | 227.125 | 65.111 | 31.6 | 0.11 |
| pHILIC | 227.125 | 124.9417 | 31.6 | 0.11 |
| pHILIC | 227.125 | 126.3463 | 31.6 | 0.11 |
| pHILIC | 227.125 | 246.7313 | 31.6 | 0.11 |
| pHILIC | 227.125 | 67.7514 | 31.6 | 0.11 |
| pHILIC | 227.125 | 83.0695 | 31.6 | 0.11 |
| pHILIC | 227.125 | 82.6248 | 31.5 | 0.11 |
| pHILIC | 227.125 | 116.7168 | 31.5 | 0.11 |
| pHILIC | 227.125 | 107.6218 | 31.5 | 0.11 |
| pHILIC | 227.125 | 73.377 | 31.5 | 0.11 |
| pHILIC | 227.125 | 143.5222 | 31.5 | 0.11 |
| pHILIC | 227.125 | 183.8435 | 31.5 | 0.11 |
| pHILIC | 227.125 | 68.8474 | 31.5 | 0.11 |
| pHILIC | 227.125 | 138.5528 | 31.5 | 0.11 |
| pHILIC | 227.125 | 105.2972 | 31.5 | 0.11 |
| pHILIC | 227.125 | 128.7567 | 31.5 | 0.11 |
| pHILIC | 227.125 | 136.2838 | 31.5 | 0.11 |
| pHILIC | 227.125 | 152.2591 | 31.5 | 0.11 |
| pHILIC | 227.125 | 126.7245 | 31.5 | 0.11 |
| pHILIC | 227.125 | 95.1009 | 31.5 | 0.11 |
| pHILIC | 227.125 | 53.4591 | 31.5 | 0.11 |
| pHILIC | 227.125 | 99.0745 | 31.5 | 0.11 |
| pHILIC | 227.125 | 112.853 | 31.5 | 0.11 |
| pHILIC | 227.125 | 134.3786 | 31.5 | 0.11 |
| pHILIC | 227.125 | 64.2414 | 31.5 | 0.11 |
| pHILIC | 227.125 | 107.1866 | 31.5 | 0.11 |
| pHILIC | 227.125 | 68.2834 | 31.5 | 0.11 |

| | | | | |
|---|---|---|---|---|
| pHILIC | 227.125 | 195.594 | 31.4 | 0.11 |
| pHILIC | 227.125 | 71.9191 | 31.4 | 0.11 |
| pHILIC | 227.125 | 69.4466 | 31.4 | 0.11 |
| pHILIC | 227.125 | 148.1385 | 31.4 | 0.11 |
| pHILIC | 227.125 | 86.2549 | 31.4 | 0.11 |
| pHILIC | 227.125 | 221.671 | 31.4 | 0.11 |
| pHILIC | 227.125 | 84.2882 | 31.4 | 0.11 |
| pHILIC | 227.125 | 83.7936 | 31.4 | 0.11 |
| pHILIC | 227.125 | 168.5202 | 31.4 | 0.11 |
| pHILIC | 227.125 | 52.9871 | 31.4 | 0.11 |
| pHILIC | 227.125 | 57.0909 | 31.3 | 0.11 |
| pHILIC | 227.125 | 127.1849 | 31.3 | 0.11 |
| pHILIC | 227.125 | 206.2339 | 31.3 | 0.11 |
| pHILIC | 227.125 | 80.7001 | 31.3 | 0.11 |
| pHILIC | 227.125 | 82.7077 | 31.3 | 0.11 |
| pHILIC | 227.125 | 195.8792 | 31.3 | 0.11 |
| pHILIC | 227.125 | 50.4782 | 31.3 | 0.11 |
| pHILIC | 227.125 | 52.0059 | 31.3 | 0.11 |
| pHILIC | 227.125 | 63.0437 | 31.3 | 0.11 |
| pHILIC | 227.125 | 74.485 | 31.3 | 0.11 |
| pHILIC | 227.125 | 177.8291 | 31.3 | 0.11 |
| pHILIC | 227.125 | 74.6756 | 31.3 | 0.11 |
| pHILIC | 227.125 | 207.2045 | 31.3 | 0.11 |
| pHILIC | 227.125 | 65.472 | 31.3 | 0.11 |
| pHILIC | 227.125 | 72.2249 | 31.3 | 0.11 |
| pHILIC | 227.125 | 210.6963 | 31.3 | 0.11 |
| pHILIC | 227.125 | 94.2354 | 31.2 | 0.11 |
| pHILIC | 227.125 | 56.3518 | 31.2 | 0.11 |
| pHILIC | 227.125 | 75.8884 | 31.2 | 0.11 |
| pHILIC | 227.125 | 190.7017 | 31.2 | 0.11 |
| pHILIC | 227.125 | 56.2585 | 31.2 | 0.11 |
| pHILIC | 227.125 | 52.3371 | 31.2 | 0.11 |
| pHILIC | 227.125 | 97.7135 | 31.2 | 0.11 |
| pHILIC | 227.125 | 165.0027 | 31.2 | 0.11 |
| pHILIC | 227.125 | 75.6633 | 31.2 | 0.11 |
| pHILIC | 227.125 | 51.597 | 31.2 | 0.11 |
| pHILIC | 227.125 | 146.1029 | 31.2 | 0.11 |
| pHILIC | 227.125 | 107.3721 | 31.2 | 0.11 |
| pHILIC | 227.125 | 99.1753 | 31.2 | 0.11 |
| pHILIC | 227.125 | 145.5219 | 31.2 | 0.11 |
| pHILIC | 227.125 | 198.4017 | 31.2 | 0.11 |
| pHILIC | 227.125 | 179.8106 | 31.2 | 0.11 |

| | | | | |
|---|---|---|---|---|
| pHILIC | 227.125 | 214.8613 | 31.2 | 0.11 |
| pHILIC | 227.125 | 107.6886 | 31.2 | 0.11 |
| pHILIC | 227.125 | 67.1639 | 31.2 | 0.11 |
| pHILIC | 227.125 | 72.1677 | 31.2 | 0.11 |
| pHILIC | 227.125 | 210.2122 | 31.2 | 0.11 |
| pHILIC | 227.125 | 61.8855 | 31.2 | 0.11 |
| pHILIC | 227.125 | 128.282 | 31.2 | 0.11 |
| pHILIC | 227.125 | 251.6425 | 31.2 | 0.11 |
| pHILIC | 227.125 | 207.6313 | 31.2 | 0.11 |
| pHILIC | 227.125 | 57.9074 | 31.2 | 0.11 |
| pHILIC | 227.125 | 105.608 | 31.2 | 0.11 |
| pHILIC | 227.125 | 129.0002 | 31.2 | 0.11 |
| pHILIC | 227.125 | 106.9998 | 31.2 | 0.11 |
| pHILIC | 227.125 | 57.9736 | 31.2 | 0.11 |
| pHILIC | 227.125 | 61.1466 | 31.2 | 0.11 |
| pHILIC | 227.125 | 50.6 | 31.1 | 0.11 |
| pHILIC | 227.125 | 133.9149 | 31.1 | 0.11 |
| pHILIC | 227.125 | 149.5017 | 31.1 | 0.11 |
| pHILIC | 227.125 | 64.2073 | 31.1 | 0.11 |
| pHILIC | 227.125 | 82.983 | 31.1 | 0.11 |
| pHILIC | 227.125 | 53.0464 | 31.1 | 0.11 |
| pHILIC | 227.125 | 54.5721 | 31.1 | 0.11 |
| pHILIC | 227.125 | 76.9148 | 31.1 | 0.11 |
| pHILIC | 227.125 | 57.4105 | 31.1 | 0.11 |
| pHILIC | 227.125 | 143.8365 | 31.1 | 0.11 |
| pHILIC | 227.125 | 119.9526 | 31.1 | 0.11 |
| pHILIC | 227.125 | 84.157 | 31.1 | 0.11 |
| pHILIC | 227.125 | 134.3221 | 31.1 | 0.11 |
| pHILIC | 227.125 | 171.079 | 31.1 | 0.11 |
| pHILIC | 227.125 | 57.7126 | 31.1 | 0.11 |
| pHILIC | 227.125 | 56.2465 | 31.1 | 0.11 |
| pHILIC | 227.125 | 71.4713 | 31.1 | 0.11 |
| pHILIC | 227.125 | 134.2462 | 31.1 | 0.11 |
| pHILIC | 227.125 | 156.8957 | 31.1 | 0.11 |
| pHILIC | 227.125 | 146.2699 | 31.1 | 0.11 |
| pHILIC | 227.125 | 75.9963 | 31 | 0.11 |
| pHILIC | 227.125 | 61.2908 | 31 | 0.11 |
| pHILIC | 227.125 | 145.1393 | 31 | 0.11 |
| pHILIC | 227.125 | 80.8715 | 31 | 0.11 |
| pHILIC | 227.125 | 200.7371 | 31 | 0.11 |
| pHILIC | 227.125 | 92.9169 | 31 | 0.11 |
| pHILIC | 227.125 | 53.615 | 31 | 0.11 |

| | | | | |
|---|---|---|---|---|
| pHILIC | 227.125 | 50.0175 | 31 | 0.11 |
| pHILIC | 227.125 | 125.5055 | 31 | 0.11 |
| pHILIC | 227.125 | 50.7118 | 31 | 0.11 |
| pHILIC | 227.125 | 75.2655 | 31 | 0.11 |
| pHILIC | 227.125 | 94.809 | 31 | 0.11 |
| pHILIC | 227.125 | 88.9858 | 31 | 0.11 |
| pHILIC | 227.125 | 68.277 | 31 | 0.11 |
| pHILIC | 227.125 | 51.977 | 31 | 0.11 |
| pHILIC | 227.125 | 75.5121 | 31 | 0.11 |
| pHILIC | 227.125 | 198.2341 | 31 | 0.11 |
| pHILIC | 227.125 | 87.8012 | 31 | 0.11 |
| pHILIC | 227.125 | 91.6676 | 31 | 0.11 |
| pHILIC | 227.125 | 82.0848 | 31 | 0.11 |
| pHILIC | 227.125 | 113.4155 | 31 | 0.11 |
| pHILIC | 227.125 | 79.694 | 31 | 0.11 |
| pHILIC | 227.125 | 145.4897 | 31 | 0.11 |
| pHILIC | 227.125 | 51.6307 | 31 | 0.11 |
| pHILIC | 227.125 | 143.245 | 30.9 | 0.11 |
| pHILIC | 227.125 | 88.7264 | 30.9 | 0.11 |
| pHILIC | 227.125 | 95.7905 | 30.9 | 0.11 |
| pHILIC | 227.125 | 93.7376 | 30.9 | 0.11 |
| pHILIC | 227.125 | 59.5795 | 30.9 | 0.11 |
| pHILIC | 227.125 | 50.7189 | 30.9 | 0.11 |
| pHILIC | 227.125 | 83.2606 | 30.9 | 0.11 |
| pHILIC | 227.125 | 186.9 | 30.9 | 0.11 |
| pHILIC | 227.125 | 59.0284 | 30.9 | 0.11 |
| pHILIC | 227.125 | 63.649 | 30.9 | 0.11 |
| pHILIC | 227.125 | 75.9121 | 30.9 | 0.11 |
| pHILIC | 227.125 | 178.7351 | 30.9 | 0.11 |
| pHILIC | 227.125 | 196.8034 | 30.9 | 0.11 |
| pHILIC | 227.125 | 140.4531 | 30.9 | 0.11 |
| pHILIC | 227.125 | 71.5591 | 30.9 | 0.11 |
| pHILIC | 227.125 | 62.2603 | 30.9 | 0.11 |
| pHILIC | 227.125 | 52.1301 | 30.9 | 0.11 |
| pHILIC | 227.125 | 125.998 | 30.8 | 0.11 |
| pHILIC | 227.125 | 93.1458 | 30.8 | 0.11 |
| pHILIC | 227.125 | 111.7645 | 30.8 | 0.11 |
| pHILIC | 227.125 | 74.3466 | 30.8 | 0.11 |
| pHILIC | 227.125 | 71.2112 | 30.8 | 0.11 |
| pHILIC | 227.125 | 56.1289 | 30.8 | 0.11 |
| pHILIC | 227.125 | 183.6638 | 30.8 | 0.11 |
| pHILIC | 227.125 | 140.7288 | 30.8 | 0.11 |

| | | | | |
|---|---|---|---|---|
| pHILIC | 227.125 | 71.7993 | 30.8 | 0.11 |
| pHILIC | 227.125 | 104.6034 | 30.8 | 0.11 |
| pHILIC | 227.125 | 95.6554 | 30.8 | 0.11 |
| pHILIC | 227.125 | 51.3659 | 30.8 | 0.11 |
| pHILIC | 227.125 | 76.282 | 30.8 | 0.11 |
| pHILIC | 227.125 | 87.006 | 30.8 | 0.11 |
| pHILIC | 227.125 | 98.8976 | 30.8 | 0.11 |
| pHILIC | 227.125 | 62.6482 | 30.8 | 0.11 |
| pHILIC | 227.125 | 195.1707 | 30.8 | 0.11 |
| pHILIC | 227.125 | 96.2694 | 30.8 | 0.11 |
| pHILIC | 227.125 | 127.1638 | 30.8 | 0.11 |
| pHILIC | 227.125 | 94.6629 | 30.8 | 0.11 |
| pHILIC | 227.125 | 50.4382 | 30.8 | 0.11 |
| pHILIC | 227.125 | 56.9068 | 30.8 | 0.11 |
| pHILIC | 227.125 | 51.0635 | 30.8 | 0.11 |
| pHILIC | 227.125 | 86.2761 | 30.8 | 0.11 |
| pHILIC | 227.125 | 128.7264 | 30.8 | 0.11 |
| pHILIC | 227.125 | 105.552 | 30.8 | 0.11 |
| pHILIC | 227.125 | 177.8609 | 30.8 | 0.11 |
| pHILIC | 227.125 | 60.7128 | 30.8 | 0.11 |
| pHILIC | 227.125 | 65.7976 | 30.8 | 0.11 |
| pHILIC | 227.125 | 53.1306 | 30.7 | 0.11 |
| pHILIC | 227.125 | 57.8706 | 30.7 | 0.11 |
| pHILIC | 227.125 | 155.3985 | 30.7 | 0.11 |
| pHILIC | 227.125 | 52.7587 | 30.7 | 0.11 |
| pHILIC | 227.125 | 61.645 | 30.7 | 0.11 |
| pHILIC | 227.125 | 78.3706 | 30.7 | 0.11 |
| pHILIC | 227.125 | 65.6218 | 30.7 | 0.11 |
| pHILIC | 227.125 | 51.2503 | 30.7 | 0.11 |
| pHILIC | 227.125 | 149.4462 | 30.7 | 0.11 |
| pHILIC | 227.125 | 175.6528 | 30.7 | 0.11 |
| pHILIC | 227.125 | 131.0496 | 30.7 | 0.11 |
| pHILIC | 227.125 | 111.106 | 30.7 | 0.11 |
| pHILIC | 227.125 | 69.31 | 30.7 | 0.11 |
| pHILIC | 227.125 | 88.2139 | 30.7 | 0.11 |
| pHILIC | 227.125 | 127.4419 | 30.7 | 0.1 |
| pHILIC | 227.125 | 77.0581 | 30.7 | 0.1 |
| pHILIC | 227.125 | 61.572 | 30.7 | 0.1 |
| pHILIC | 227.125 | 53.6706 | 30.7 | 0.1 |
| pHILIC | 227.125 | 56.8713 | 30.7 | 0.1 |
| pHILIC | 227.125 | 98.7679 | 30.7 | 0.1 |
| pHILIC | 227.125 | 99.4881 | 30.7 | 0.1 |

| | | | | |
|---|---|---|---|---|
| pHILIC | 227.125 | 139.4618 | 30.7 | 0.1 |
| pHILIC | 227.125 | 108.7292 | 30.7 | 0.1 |
| pHILIC | 227.125 | 68.054 | 30.6 | 0.1 |
| pHILIC | 227.125 | 189.7004 | 30.6 | 0.1 |
| pHILIC | 227.125 | 56.6944 | 30.6 | 0.1 |
| pHILIC | 227.125 | 70.7578 | 30.6 | 0.1 |
| pHILIC | 227.125 | 137.028 | 30.6 | 0.1 |
| pHILIC | 227.125 | 215.096 | 30.6 | 0.1 |
| pHILIC | 227.125 | 79.4695 | 30.6 | 0.1 |
| pHILIC | 227.125 | 94.1011 | 30.6 | 0.1 |
| pHILIC | 227.125 | 152.9603 | 30.6 | 0.1 |
| pHILIC | 227.125 | 62.2088 | 30.6 | 0.1 |
| pHILIC | 227.125 | 51.4334 | 30.6 | 0.1 |
| pHILIC | 227.125 | 56.5922 | 30.6 | 0.1 |
| pHILIC | 227.125 | 199.1511 | 30.6 | 0.1 |
| pHILIC | 227.125 | 66.9732 | 30.6 | 0.1 |
| pHILIC | 227.125 | 104.4723 | 30.6 | 0.1 |
| pHILIC | 227.125 | 73.8883 | 30.6 | 0.1 |
| pHILIC | 227.125 | 72.8436 | 30.6 | 0.1 |
| pHILIC | 227.125 | 118.1037 | 30.6 | 0.1 |
| pHILIC | 227.125 | 78.1934 | 30.6 | 0.1 |
| pHILIC | 227.125 | 140.4055 | 30.6 | 0.1 |
| pHILIC | 227.125 | 70.2073 | 30.6 | 0.1 |
| pHILIC | 227.125 | 72.2295 | 30.6 | 0.1 |
| pHILIC | 227.125 | 56.427 | 30.5 | 0.1 |
| pHILIC | 227.125 | 173.4215 | 30.5 | 0.1 |
| pHILIC | 227.125 | 75.0027 | 30.5 | 0.1 |
| pHILIC | 227.125 | 173.2576 | 30.5 | 0.1 |
| pHILIC | 227.125 | 154.8748 | 30.5 | 0.1 |
| pHILIC | 227.125 | 60.4573 | 30.5 | 0.1 |
| pHILIC | 227.125 | 60.0581 | 30.5 | 0.1 |
| pHILIC | 227.125 | 116.6778 | 30.5 | 0.1 |
| pHILIC | 227.125 | 138.9492 | 30.5 | 0.1 |
| pHILIC | 227.125 | 94.2484 | 30.5 | 0.1 |
| pHILIC | 227.125 | 56.7719 | 30.5 | 0.1 |
| pHILIC | 227.125 | 73.4175 | 30.5 | 0.1 |
| pHILIC | 227.125 | 93.1177 | 30.5 | 0.1 |
| pHILIC | 227.125 | 107.5607 | 30.5 | 0.1 |
| pHILIC | 227.125 | 104.5698 | 30.5 | 0.1 |
| pHILIC | 227.125 | 103.3733 | 30.5 | 0.1 |
| pHILIC | 227.125 | 225.2546 | 30.5 | 0.1 |
| pHILIC | 227.125 | 50.595 | 30.5 | 0.1 |

| | | | | |
|---|---|---|---|---|
| pHILIC | 227.125 | 54.42 | 30.5 | 0.1 |
| pHILIC | 227.125 | 151.2809 | 30.5 | 0.1 |
| pHILIC | 227.125 | 136.665 | 30.5 | 0.1 |
| pHILIC | 227.125 | 59.5542 | 30.5 | 0.1 |
| pHILIC | 227.125 | 60.4693 | 30.5 | 0.1 |
| pHILIC | 227.125 | 93.9091 | 30.5 | 0.1 |
| pHILIC | 227.125 | 67.685 | 30.5 | 0.1 |
| pHILIC | 227.125 | 66.5446 | 30.4 | 0.1 |
| pHILIC | 227.125 | 51.8107 | 30.4 | 0.1 |
| pHILIC | 227.125 | 57.6038 | 30.4 | 0.1 |
| pHILIC | 227.125 | 62.435 | 30.4 | 0.1 |
| pHILIC | 227.125 | 197.7861 | 30.4 | 0.1 |
| pHILIC | 227.125 | 127.3328 | 30.4 | 0.1 |
| pHILIC | 227.125 | 106.0394 | 30.4 | 0.1 |
| pHILIC | 227.125 | 102.2518 | 30.4 | 0.1 |
| pHILIC | 227.125 | 202.9109 | 30.4 | 0.1 |
| pHILIC | 227.125 | 60.6257 | 30.4 | 0.1 |
| pHILIC | 227.125 | 226.4705 | 30.4 | 0.1 |
| pHILIC | 227.125 | 240.9496 | 30.4 | 0.1 |
| pHILIC | 227.125 | 107.6983 | 30.4 | 0.1 |
| pHILIC | 227.125 | 69.3213 | 30.4 | 0.1 |
| pHILIC | 227.125 | 82.5744 | 30.4 | 0.1 |
| pHILIC | 227.125 | 53.0359 | 30.4 | 0.1 |
| pHILIC | 227.125 | 56.8502 | 30.4 | 0.1 |
| pHILIC | 227.125 | 203.7728 | 30.4 | 0.1 |
| pHILIC | 227.125 | 213.5191 | 30.4 | 0.1 |
| pHILIC | 227.125 | 53.7533 | 30.4 | 0.1 |
| pHILIC | 227.125 | 63.1229 | 30.4 | 0.1 |
| pHILIC | 227.125 | 123.4972 | 30.4 | 0.1 |
| pHILIC | 227.125 | 75.5496 | 30.4 | 0.1 |
| pHILIC | 227.125 | 66.4392 | 30.4 | 0.1 |
| pHILIC | 227.125 | 83.5285 | 30.4 | 0.1 |
| pHILIC | 227.125 | 128.6044 | 30.4 | 0.1 |
| pHILIC | 227.125 | 157.2876 | 30.4 | 0.1 |
| pHILIC | 227.125 | 55.0186 | 30.4 | 0.1 |
| pHILIC | 227.125 | 116.3577 | 30.4 | 0.1 |
| pHILIC | 227.125 | 82.613 | 30.4 | 0.1 |
| pHILIC | 227.125 | 213.8006 | 30.4 | 0.1 |
| pHILIC | 227.125 | 240.8371 | 30.4 | 0.1 |
| pHILIC | 227.125 | 245.1201 | 30.4 | 0.1 |
| pHILIC | 227.125 | 130.9923 | 30.4 | 0.1 |
| pHILIC | 227.125 | 86.4428 | 30.4 | 0.1 |

| | | | | |
|---|---|---|---|---|
| pHILIC | 227.125 | 140.6209 | 30.3 | 0.1 |
| pHILIC | 227.125 | 51.9187 | 30.3 | 0.1 |
| pHILIC | 227.125 | 54.4373 | 30.3 | 0.1 |
| pHILIC | 227.125 | 58.8253 | 30.3 | 0.1 |
| pHILIC | 227.125 | 51.8039 | 30.3 | 0.1 |
| pHILIC | 227.125 | 55.6333 | 30.3 | 0.1 |
| pHILIC | 227.125 | 70.9275 | 30.3 | 0.1 |
| pHILIC | 227.125 | 216.5392 | 30.3 | 0.1 |
| pHILIC | 227.125 | 203.6383 | 30.3 | 0.1 |
| pHILIC | 227.125 | 74.7257 | 30.3 | 0.1 |
| pHILIC | 227.125 | 54.6675 | 30.3 | 0.1 |
| pHILIC | 227.125 | 70.2997 | 30.3 | 0.1 |
| pHILIC | 227.125 | 93.6525 | 30.3 | 0.1 |
| pHILIC | 227.125 | 87.9618 | 30.3 | 0.1 |
| pHILIC | 227.125 | 129.7077 | 30.3 | 0.1 |
| pHILIC | 227.125 | 64.2702 | 30.3 | 0.1 |
| pHILIC | 227.125 | 193.0339 | 30.3 | 0.1 |
| pHILIC | 227.125 | 222.9196 | 30.3 | 0.1 |
| pHILIC | 227.125 | 125.6929 | 30.3 | 0.1 |
| pHILIC | 227.125 | 78.4781 | 30.3 | 0.1 |
| pHILIC | 227.125 | 50.0789 | 30.3 | 0.1 |
| pHILIC | 227.125 | 88.1976 | 30.3 | 0.1 |
| pHILIC | 227.125 | 58.1298 | 30.3 | 0.1 |
| pHILIC | 227.125 | 85.2518 | 30.3 | 0.1 |
| pHILIC | 227.125 | 191.5769 | 30.3 | 0.1 |
| pHILIC | 227.125 | 99.0383 | 30.3 | 0.1 |
| pHILIC | 227.125 | 243.8806 | 30.3 | 0.1 |
| pHILIC | 227.125 | 123.9433 | 30.3 | 0.1 |
| pHILIC | 227.125 | 55.9937 | 30.3 | 0.1 |
| pHILIC | 227.125 | 59.7526 | 30.2 | 0.1 |
| pHILIC | 227.125 | 65.0045 | 30.2 | 0.1 |
| pHILIC | 227.125 | 127.2712 | 30.2 | 0.1 |
| pHILIC | 227.125 | 165.3222 | 30.2 | 0.1 |
| pHILIC | 227.125 | 69.7522 | 30.2 | 0.1 |
| pHILIC | 227.125 | 54.8551 | 30.2 | 0.1 |
| pHILIC | 227.125 | 89.3837 | 30.2 | 0.1 |
| pHILIC | 227.125 | 64.7773 | 30.2 | 0.1 |
| pHILIC | 227.125 | 58.4538 | 30.2 | 0.1 |
| pHILIC | 227.125 | 166.528 | 30.2 | 0.1 |
| pHILIC | 227.125 | 83.8488 | 30.2 | 0.1 |
| pHILIC | 227.125 | 53.7841 | 30.2 | 0.1 |
| pHILIC | 227.125 | 91.4824 | 30.2 | 0.1 |

| | | | | |
|---|---|---|---|---|
| pHILIC | 227.125 | 83.1091 | 30.2 | 0.1 |
| pHILIC | 227.125 | 188.213 | 30.2 | 0.1 |
| pHILIC | 227.125 | 137.7955 | 30.2 | 0.1 |
| pHILIC | 227.125 | 164.5876 | 30.2 | 0.1 |
| pHILIC | 227.125 | 174.875 | 30.1 | 0.1 |
| pHILIC | 227.125 | 51.8017 | 30.1 | 0.1 |
| pHILIC | 227.125 | 101.0781 | 30.1 | 0.1 |
| pHILIC | 227.125 | 165.5853 | 30.1 | 0.1 |
| pHILIC | 227.125 | 139.8177 | 30.1 | 0.1 |
| pHILIC | 227.125 | 63.8255 | 30.1 | 0.1 |
| pHILIC | 227.125 | 51.4823 | 30.1 | 0.1 |
| pHILIC | 227.125 | 105.0308 | 30.1 | 0.1 |
| pHILIC | 227.125 | 109.5874 | 30.1 | 0.1 |
| pHILIC | 227.125 | 241.4659 | 30.1 | 0.1 |
| pHILIC | 227.125 | 116.4119 | 30.1 | 0.1 |
| pHILIC | 227.125 | 61.0467 | 30.1 | 0.1 |
| pHILIC | 227.125 | 50.4154 | 30.1 | 0.1 |
| pHILIC | 227.125 | 69.9313 | 30.1 | 0.1 |
| pHILIC | 227.125 | 160.0294 | 30.1 | 0.1 |
| pHILIC | 227.125 | 153.933 | 30.1 | 0.1 |
| pHILIC | 227.125 | 60.5659 | 30.1 | 0.1 |
| pHILIC | 227.125 | 104.3312 | 30.1 | 0.1 |
| pHILIC | 227.125 | 50.7601 | 30.1 | 0.1 |
| pHILIC | 227.125 | 79.4139 | 30.1 | 0.1 |
| pHILIC | 227.125 | 93.5659 | 30.1 | 0.1 |
| pHILIC | 227.125 | 50.1526 | 30 | 0.1 |
| pHILIC | 227.125 | 66.1038 | 30 | 0.1 |
| pHILIC | 227.125 | 108.6374 | 30 | 0.1 |
| pHILIC | 227.125 | 52.4659 | 30 | 0.1 |
| pHILIC | 227.125 | 55.4052 | 30 | 0.1 |
| pHILIC | 227.125 | 70.6593 | 30 | 0.1 |
| pHILIC | 227.125 | 83.7571 | 30 | 0.1 |
| pHILIC | 227.125 | 55.8141 | 30 | 0.1 |
| pHILIC | 227.125 | 73.8842 | 30 | 0.1 |
| pHILIC | 227.125 | 65.0851 | 30 | 0.1 |
| pHILIC | 227.125 | 93.1567 | 30 | 0.1 |
| pHILIC | 227.125 | 78.4088 | 30 | 0.1 |
| pHILIC | 227.125 | 66.8143 | 30 | 0.1 |
| pHILIC | 227.125 | 184.3072 | 30 | 0.1 |
| pHILIC | 227.125 | 55.9879 | 30 | 0.1 |
| pHILIC | 227.125 | 87.4409 | 30 | 0.1 |
| pHILIC | 227.125 | 52.3561 | 30 | 0.1 |

| | | | | |
|---|---|---|---|---|
| pHILIC | 227.125 | 130.2687 | 30 | 0.1 |
| pHILIC | 227.125 | 59.5098 | 30 | 0.1 |
| pHILIC | 227.125 | 103.0263 | 30 | 0.1 |
| pHILIC | 227.125 | 111.1576 | 30 | 0.1 |
| pHILIC | 227.125 | 187.2254 | 30 | 0.1 |
| pHILIC | 227.125 | 115.0485 | 30 | 0.1 |
| pHILIC | 227.125 | 54.1652 | 29.9 | 0.1 |
| pHILIC | 227.125 | 52.6525 | 29.9 | 0.1 |
| pHILIC | 227.125 | 50.561 | 29.9 | 0.1 |
| pHILIC | 227.125 | 54.6831 | 29.9 | 0.1 |
| pHILIC | 227.125 | 138.964 | 29.9 | 0.1 |
| pHILIC | 227.125 | 238.8884 | 29.9 | 0.1 |
| pHILIC | 227.125 | 57.0469 | 29.9 | 0.1 |
| pHILIC | 227.125 | 192.7321 | 29.9 | 0.1 |
| pHILIC | 227.125 | 55.2324 | 29.9 | 0.1 |
| pHILIC | 227.125 | 53.8026 | 29.9 | 0.1 |
| pHILIC | 227.125 | 55.6064 | 29.9 | 0.1 |
| pHILIC | 227.125 | 239.1802 | 29.9 | 0.1 |
| pHILIC | 227.125 | 76.1062 | 29.9 | 0.1 |
| pHILIC | 227.125 | 61.4451 | 29.9 | 0.1 |
| pHILIC | 227.125 | 120.2748 | 29.9 | 0.1 |
| pHILIC | 227.125 | 103.2673 | 29.9 | 0.1 |
| pHILIC | 227.125 | 62.4692 | 29.9 | 0.1 |
| pHILIC | 227.125 | 112.2724 | 29.9 | 0.1 |
| pHILIC | 227.125 | 180.9357 | 29.9 | 0.1 |
| pHILIC | 227.125 | 134.514 | 29.9 | 0.1 |
| pHILIC | 227.125 | 149.3406 | 29.9 | 0.1 |
| pHILIC | 227.125 | 65.727 | 29.8 | 0.1 |
| pHILIC | 227.125 | 120.8898 | 29.8 | 0.1 |
| pHILIC | 227.125 | 88.9196 | 29.8 | 0.1 |
| pHILIC | 227.125 | 62.6661 | 29.8 | 0.1 |
| pHILIC | 227.125 | 147.1221 | 29.8 | 0.1 |
| pHILIC | 227.125 | 241.8135 | 29.8 | 0.1 |
| pHILIC | 227.125 | 239.8086 | 29.8 | 0.1 |
| pHILIC | 227.125 | 68.3423 | 29.8 | 0.1 |
| pHILIC | 227.125 | 132.7762 | 29.8 | 0.1 |
| pHILIC | 227.125 | 58.0715 | 29.8 | 0.1 |
| pHILIC | 227.125 | 64.5278 | 29.8 | 0.1 |
| pHILIC | 227.125 | 109.1005 | 29.8 | 0.1 |
| pHILIC | 227.125 | 192.6194 | 29.8 | 0.1 |
| pHILIC | 227.125 | 175.2453 | 29.8 | 0.1 |
| pHILIC | 227.125 | 200.4358 | 29.8 | 0.1 |

| | | | | |
|---|---|---|---|---|
| pHILIC | 227.125 | 55.8378 | 29.8 | 0.1 |
| pHILIC | 227.125 | 62.4426 | 29.8 | 0.1 |
| pHILIC | 227.125 | 75.3971 | 29.8 | 0.1 |
| pHILIC | 227.125 | 50.5251 | 29.8 | 0.1 |
| pHILIC | 227.125 | 136.6863 | 29.8 | 0.1 |
| pHILIC | 227.125 | 52.0418 | 29.8 | 0.1 |
| pHILIC | 227.125 | 102.8039 | 29.8 | 0.1 |
| pHILIC | 227.125 | 106.8381 | 29.8 | 0.1 |
| pHILIC | 227.125 | 122.3881 | 29.7 | 0.1 |
| pHILIC | 227.125 | 81.1941 | 29.7 | 0.1 |
| pHILIC | 227.125 | 59.0993 | 29.7 | 0.1 |
| pHILIC | 227.125 | 61.3135 | 29.7 | 0.1 |
| pHILIC | 227.125 | 188.3281 | 29.7 | 0.1 |
| pHILIC | 227.125 | 82.4764 | 29.7 | 0.1 |
| pHILIC | 227.125 | 117.8546 | 29.7 | 0.1 |
| pHILIC | 227.125 | 71.993 | 29.7 | 0.1 |
| pHILIC | 227.125 | 215.3275 | 29.7 | 0.1 |
| pHILIC | 227.125 | 95.1693 | 29.7 | 0.1 |
| pHILIC | 227.125 | 57.5634 | 29.7 | 0.1 |
| pHILIC | 227.125 | 79.2191 | 29.7 | 0.1 |
| pHILIC | 227.125 | 151.766 | 29.7 | 0.1 |
| pHILIC | 227.125 | 158.1729 | 29.7 | 0.1 |
| pHILIC | 227.125 | 150.6285 | 29.7 | 0.1 |
| pHILIC | 227.125 | 52.4966 | 29.7 | 0.1 |
| pHILIC | 227.125 | 108.2293 | 29.7 | 0.1 |
| pHILIC | 227.125 | 85.9163 | 29.7 | 0.1 |
| pHILIC | 227.125 | 74.9045 | 29.7 | 0.1 |
| pHILIC | 227.125 | 147.5615 | 29.7 | 0.1 |
| pHILIC | 227.125 | 99.11 | 29.7 | 0.1 |
| pHILIC | 227.125 | 159.6468 | 29.7 | 0.1 |
| pHILIC | 227.125 | 80.5592 | 29.7 | 0.1 |
| pHILIC | 227.125 | 123.6342 | 29.6 | 0.1 |
| pHILIC | 227.125 | 68.5222 | 29.6 | 0.1 |
| pHILIC | 227.125 | 80.2286 | 29.6 | 0.1 |
| pHILIC | 227.125 | 81.4262 | 29.6 | 0.1 |
| pHILIC | 227.125 | 120.302 | 29.6 | 0.1 |
| pHILIC | 227.125 | 88.3454 | 29.6 | 0.1 |
| pHILIC | 227.125 | 202.7731 | 29.6 | 0.1 |
| pHILIC | 227.125 | 98.1646 | 29.6 | 0.1 |
| pHILIC | 227.125 | 58.7662 | 29.6 | 0.1 |
| pHILIC | 227.125 | 63.9819 | 29.6 | 0.1 |
| pHILIC | 227.125 | 51.637 | 29.6 | 0.1 |

| | | | | |
|---|---|---|---|---|
| pHILIC | 227.125 | 52.3689 | 29.6 | 0.1 |
| pHILIC | 227.125 | 112.1925 | 29.6 | 0.1 |
| pHILIC | 227.125 | 52.4309 | 29.6 | 0.1 |
| pHILIC | 227.125 | 111.5006 | 29.6 | 0.1 |
| pHILIC | 227.125 | 201.8338 | 29.6 | 0.1 |
| pHILIC | 227.125 | 59.4651 | 29.6 | 0.1 |
| pHILIC | 227.125 | 105.2635 | 29.6 | 0.1 |
| pHILIC | 227.125 | 106.6612 | 29.6 | 0.1 |
| pHILIC | 227.125 | 87.5206 | 29.6 | 0.1 |
| pHILIC | 227.125 | 111.2466 | 29.6 | 0.1 |
| pHILIC | 227.125 | 52.5317 | 29.6 | 0.1 |
| pHILIC | 227.125 | 56.394 | 29.6 | 0.1 |
| pHILIC | 227.125 | 163.9302 | 29.6 | 0.1 |
| pHILIC | 227.125 | 180.764 | 29.5 | 0.1 |
| pHILIC | 227.125 | 108.002 | 29.5 | 0.1 |
| pHILIC | 227.125 | 127.3063 | 29.5 | 0.1 |
| pHILIC | 227.125 | 76.4191 | 29.5 | 0.1 |
| pHILIC | 227.125 | 146.3621 | 29.5 | 0.1 |
| pHILIC | 227.125 | 94.5079 | 29.5 | 0.1 |
| pHILIC | 227.125 | 106.3427 | 29.5 | 0.1 |
| pHILIC | 227.125 | 167.4031 | 29.5 | 0.1 |
| pHILIC | 227.125 | 67.5935 | 29.5 | 0.1 |
| pHILIC | 227.125 | 91.915 | 29.5 | 0.1 |
| pHILIC | 227.125 | 58.2202 | 29.5 | 0.1 |
| pHILIC | 227.125 | 174.3494 | 29.5 | 0.1 |
| pHILIC | 227.125 | 50.5476 | 29.5 | 0.1 |
| pHILIC | 227.125 | 54.0996 | 29.5 | 0.1 |
| pHILIC | 227.125 | 85.1992 | 29.5 | 0.1 |
| pHILIC | 227.125 | 73.8007 | 29.5 | 0.1 |
| pHILIC | 227.125 | 92.7144 | 29.5 | 0.1 |
| pHILIC | 227.125 | 110.998 | 29.5 | 0.1 |
| pHILIC | 227.125 | 182.4891 | 29.5 | 0.1 |
| pHILIC | 227.125 | 70.1181 | 29.5 | 0.1 |
| pHILIC | 227.125 | 112.9307 | 29.5 | 0.1 |
| pHILIC | 227.125 | 51.9282 | 29.5 | 0.1 |
| pHILIC | 227.125 | 100.0936 | 29.5 | 0.1 |
| pHILIC | 227.125 | 211.2255 | 29.5 | 0.1 |
| pHILIC | 227.125 | 79.2862 | 29.5 | 0.1 |
| pHILIC | 227.125 | 92.853 | 29.4 | 0.1 |
| pHILIC | 227.125 | 220.2162 | 29.4 | 0.1 |
| pHILIC | 227.125 | 175.6031 | 29.4 | 0.1 |
| pHILIC | 227.125 | 58.4752 | 29.4 | 0.1 |

| | | | | |
|---|---|---|---|---|
| pHILIC | 227.125 | 115.2569 | 29.4 | 0.1 |
| pHILIC | 227.125 | 71.244 | 29.4 | 0.1 |
| pHILIC | 227.125 | 101.4966 | 29.4 | 0.1 |
| pHILIC | 227.125 | 54.8344 | 29.4 | 0.1 |
| pHILIC | 227.125 | 90.5219 | 29.4 | 0.1 |
| pHILIC | 227.125 | 66.9319 | 29.4 | 0.1 |
| pHILIC | 227.125 | 77.9014 | 29.4 | 0.1 |
| pHILIC | 227.125 | 56.1952 | 29.4 | 0.1 |
| pHILIC | 227.125 | 78.354 | 29.4 | 0.1 |
| pHILIC | 227.125 | 172.2431 | 29.4 | 0.1 |
| pHILIC | 227.125 | 125.4265 | 29.4 | 0.1 |
| pHILIC | 227.125 | 92.255 | 29.4 | 0.1 |
| pHILIC | 227.125 | 141.6855 | 29.4 | 0.1 |
| pHILIC | 227.125 | 76.7043 | 29.3 | 0.1 |
| pHILIC | 227.125 | 121.3541 | 29.3 | 0.1 |
| pHILIC | 227.125 | 85.3551 | 29.3 | 0.1 |
| pHILIC | 227.125 | 153.1294 | 29.3 | 0.1 |
| pHILIC | 227.125 | 95.9476 | 29.3 | 0.1 |
| pHILIC | 227.125 | 130.481 | 29.3 | 0.1 |
| pHILIC | 227.125 | 86.7684 | 29.3 | 0.1 |
| pHILIC | 227.125 | 92.5062 | 29.3 | 0.1 |
| pHILIC | 227.125 | 121.6228 | 29.3 | 0.1 |
| pHILIC | 227.125 | 63.6915 | 29.3 | 0.1 |
| pHILIC | 227.125 | 59.2038 | 29.3 | 0.1 |
| pHILIC | 227.125 | 246.1661 | 29.3 | 0.1 |
| pHILIC | 227.125 | 54.4963 | 29.3 | 0.1 |
| pHILIC | 227.125 | 56.7899 | 29.3 | 0.1 |
| pHILIC | 227.125 | 90.7048 | 29.3 | 0.1 |
| pHILIC | 227.125 | 88.8416 | 29.3 | 0.1 |
| pHILIC | 227.125 | 60.8242 | 29.3 | 0.1 |
| pHILIC | 227.125 | 199.4017 | 29.3 | 0.1 |
| pHILIC | 227.125 | 139.2264 | 29.3 | 0.1 |
| pHILIC | 227.125 | 97.6622 | 29.3 | 0.1 |
| pHILIC | 227.125 | 90.0665 | 29.3 | 0.1 |
| pHILIC | 227.125 | 141.6518 | 29.3 | 0.1 |
| pHILIC | 227.125 | 162.5702 | 29.3 | 0.1 |
| pHILIC | 227.125 | 51.9422 | 29.3 | 0.1 |
| pHILIC | 227.125 | 213.0334 | 29.3 | 0.1 |
| pHILIC | 227.125 | 99.8141 | 29.3 | 0.1 |
| pHILIC | 227.125 | 84.0928 | 29.3 | 0.1 |
| pHILIC | 227.125 | 84.5604 | 29.3 | 0.1 |
| pHILIC | 227.125 | 60.7696 | 29.3 | 0.1 |

| | | | | |
|---|---|---|---|---|
| pHILIC | 227.125 | 107.8864 | 29.3 | 0.1 |
| pHILIC | 227.125 | 106.904 | 29.3 | 0.1 |
| pHILIC | 227.125 | 113.2197 | 29.3 | 0.1 |
| pHILIC | 227.125 | 152.6021 | 29.3 | 0.1 |
| pHILIC | 227.125 | 70.1807 | 29.3 | 0.1 |
| pHILIC | 227.125 | 92.9449 | 29.3 | 0.1 |
| pHILIC | 227.125 | 70.5938 | 29.3 | 0.1 |
| pHILIC | 227.125 | 58.2292 | 29.2 | 0.1 |
| pHILIC | 227.125 | 149.8035 | 29.2 | 0.1 |
| pHILIC | 227.125 | 52.7827 | 29.2 | 0.1 |
| pHILIC | 227.125 | 138.3408 | 29.2 | 0.1 |
| pHILIC | 227.125 | 56.0463 | 29.2 | 0.1 |
| pHILIC | 227.125 | 88.1242 | 29.2 | 0.1 |
| pHILIC | 227.125 | 101.7831 | 29.2 | 0.1 |
| pHILIC | 227.125 | 215.469 | 29.2 | 0.1 |
| pHILIC | 227.125 | 143.7819 | 29.2 | 0.1 |
| pHILIC | 227.125 | 105.7581 | 29.2 | 0.1 |
| pHILIC | 227.125 | 66.0101 | 29.2 | 0.1 |
| pHILIC | 227.125 | 51.5933 | 29.2 | 0.1 |
| pHILIC | 227.125 | 187.2831 | 29.2 | 0.1 |
| pHILIC | 227.125 | 129.4489 | 29.2 | 0.1 |
| pHILIC | 227.125 | 79.3142 | 29.2 | 0.1 |
| pHILIC | 227.125 | 199.2126 | 29.2 | 0.1 |
| pHILIC | 227.125 | 130.1824 | 29.2 | 0.1 |
| pHILIC | 227.125 | 57.2443 | 29.2 | 0.1 |
| pHILIC | 227.125 | 66.8762 | 29.2 | 0.1 |
| pHILIC | 227.125 | 151.4743 | 29.2 | 0.1 |
| pHILIC | 227.125 | 59.4742 | 29.2 | 0.1 |
| pHILIC | 227.125 | 65.7116 | 29.2 | 0.1 |
| pHILIC | 227.125 | 84.0567 | 29.2 | 0.1 |
| pHILIC | 227.125 | 226.8081 | 29.2 | 0.1 |
| pHILIC | 227.125 | 176.6637 | 29.2 | 0.1 |
| pHILIC | 227.125 | 63.8554 | 29.2 | 0.1 |
| pHILIC | 227.125 | 54.6884 | 29.2 | 0.1 |
| pHILIC | 227.125 | 66.3741 | 29.2 | 0.1 |
| pHILIC | 227.125 | 132.9483 | 29.2 | 0.1 |
| pHILIC | 227.125 | 83.489 | 29.2 | 0.1 |
| pHILIC | 227.125 | 80.126 | 29.2 | 0.1 |
| pHILIC | 227.125 | 101.8743 | 29.1 | 0.1 |
| pHILIC | 227.125 | 78.1337 | 29.1 | 0.1 |
| pHILIC | 227.125 | 150.103 | 29.1 | 0.1 |
| pHILIC | 227.125 | 61.5116 | 29.1 | 0.1 |

| | | | | |
|---|---|---|---|---|
| pHILIC | 227.125 | 94.9176 | 29.1 | 0.1 |
| pHILIC | 227.125 | 61.87 | 29.1 | 0.1 |
| pHILIC | 227.125 | 141.7245 | 29.1 | 0.1 |
| pHILIC | 227.125 | 235.0214 | 29.1 | 0.1 |
| pHILIC | 227.125 | 154.0862 | 29.1 | 0.1 |
| pHILIC | 227.125 | 71.883 | 29.1 | 0.1 |
| pHILIC | 227.125 | 212.172 | 29.1 | 0.1 |
| pHILIC | 227.125 | 106.5739 | 29.1 | 0.1 |
| pHILIC | 227.125 | 56.5021 | 29.1 | 0.1 |
| pHILIC | 227.125 | 89.4135 | 29.1 | 0.1 |
| pHILIC | 227.125 | 61.1488 | 29.1 | 0.1 |
| pHILIC | 227.125 | 52.698 | 29.1 | 0.1 |
| pHILIC | 227.125 | 249.508 | 29.1 | 0.1 |
| pHILIC | 227.125 | 59.2397 | 29.1 | 0.1 |
| pHILIC | 227.125 | 65.3278 | 29.1 | 0.1 |
| pHILIC | 227.125 | 216.6846 | 29.1 | 0.1 |
| pHILIC | 227.125 | 63.8702 | 29.1 | 0.1 |
| pHILIC | 227.125 | 93.1963 | 29.1 | 0.1 |
| pHILIC | 227.125 | 70.0686 | 29.1 | 0.1 |
| pHILIC | 227.125 | 71.7476 | 29.1 | 0.1 |
| pHILIC | 227.125 | 138.1802 | 29.1 | 0.1 |
| pHILIC | 227.125 | 151.953 | 29.1 | 0.1 |
| pHILIC | 227.125 | 81.9002 | 29 | 0.1 |
| pHILIC | 227.125 | 74.8539 | 29 | 0.1 |
| pHILIC | 227.125 | 82.1858 | 29 | 0.1 |
| pHILIC | 227.125 | 143.8092 | 29 | 0.1 |
| pHILIC | 227.125 | 55.3834 | 29 | 0.1 |
| pHILIC | 227.125 | 130.7943 | 29 | 0.1 |
| pHILIC | 227.125 | 68.3129 | 29 | 0.1 |
| pHILIC | 227.125 | 57.0749 | 29 | 0.1 |
| pHILIC | 227.125 | 79.8948 | 29 | 0.1 |
| pHILIC | 227.125 | 81.883 | 29 | 0.1 |
| pHILIC | 227.125 | 133.1148 | 29 | 0.1 |
| pHILIC | 227.125 | 69.0842 | 29 | 0.1 |
| pHILIC | 227.125 | 88.3256 | 29 | 0.1 |
| pHILIC | 227.125 | 94.1901 | 29 | 0.1 |
| pHILIC | 227.125 | 54.173 | 29 | 0.1 |
| pHILIC | 227.125 | 159.9617 | 29 | 0.1 |
| pHILIC | 227.125 | 109.9537 | 29 | 0.1 |
| pHILIC | 227.125 | 83.8713 | 29 | 0.1 |
| pHILIC | 227.125 | 54.0941 | 29 | 0.1 |
| pHILIC | 227.125 | 180.4493 | 29 | 0.1 |

| | | | | |
|---|---|---|---|---|
| pHILIC | 227.125 | 128.3916 | 29 | 0.1 |
| pHILIC | 227.125 | 126.1625 | 29 | 0.1 |
| pHILIC | 227.125 | 189.2964 | 29 | 0.1 |
| pHILIC | 227.125 | 62.1618 | 29 | 0.1 |
| pHILIC | 227.125 | 228.0899 | 29 | 0.1 |
| pHILIC | 227.125 | 55.0542 | 29 | 0.1 |
| pHILIC | 227.125 | 169.5504 | 29 | 0.1 |
| pHILIC | 227.125 | 206.0461 | 29 | 0.1 |
| pHILIC | 227.125 | 186.8782 | 29 | 0.1 |
| pHILIC | 227.125 | 50.8274 | 28.9 | 0.1 |
| pHILIC | 227.125 | 221.0618 | 28.9 | 0.1 |
| pHILIC | 227.125 | 128.0987 | 28.9 | 0.1 |
| pHILIC | 227.125 | 87.928 | 28.9 | 0.1 |
| pHILIC | 227.125 | 125.2917 | 28.9 | 0.1 |
| pHILIC | 227.125 | 135.1544 | 28.9 | 0.1 |
| pHILIC | 227.125 | 111.0416 | 28.9 | 0.1 |
| pHILIC | 227.125 | 197.9559 | 28.9 | 0.1 |
| pHILIC | 227.125 | 55.0388 | 28.9 | 0.1 |
| pHILIC | 227.125 | 215.5901 | 28.9 | 0.1 |
| pHILIC | 227.125 | 135.7734 | 28.9 | 0.1 |
| pHILIC | 227.125 | 120.7722 | 28.9 | 0.1 |
| pHILIC | 227.125 | 232.2235 | 28.9 | 0.1 |
| pHILIC | 227.125 | 126.9093 | 28.9 | 0.1 |
| pHILIC | 227.125 | 136.2434 | 28.9 | 0.1 |
| pHILIC | 227.125 | 71.6384 | 28.9 | 0.1 |
| pHILIC | 227.125 | 85.651 | 28.9 | 0.1 |
| pHILIC | 227.125 | 65.4409 | 28.9 | 0.1 |
| pHILIC | 227.125 | 73.8273 | 28.9 | 0.1 |
| pHILIC | 227.125 | 96.3809 | 28.9 | 0.1 |
| pHILIC | 227.125 | 69.5177 | 28.9 | 0.1 |
| pHILIC | 227.125 | 55.5157 | 28.9 | 0.1 |
| pHILIC | 227.125 | 56.609 | 28.9 | 0.1 |
| pHILIC | 227.125 | 233.7559 | 28.8 | 0.1 |
| pHILIC | 227.125 | 88.1491 | 28.8 | 0.1 |
| pHILIC | 227.125 | 95.3744 | 28.8 | 0.1 |
| pHILIC | 227.125 | 111.0822 | 28.8 | 0.1 |
| pHILIC | 227.125 | 210.9133 | 28.8 | 0.1 |
| pHILIC | 227.125 | 109.853 | 28.8 | 0.1 |
| pHILIC | 227.125 | 51.0935 | 28.8 | 0.1 |
| pHILIC | 227.125 | 95.8953 | 28.8 | 0.1 |
| pHILIC | 227.125 | 152.4892 | 28.8 | 0.1 |
| pHILIC | 227.125 | 103.0472 | 28.8 | 0.1 |

| | | | | |
|---|---|---|---|---|
| pHILIC | 227.125 | 75.916 | 28.8 | 0.1 |
| pHILIC | 227.125 | 78.5747 | 28.8 | 0.1 |
| pHILIC | 227.125 | 116.631 | 28.8 | 0.1 |
| pHILIC | 227.125 | 177.6366 | 28.8 | 0.1 |
| pHILIC | 227.125 | 78.6483 | 28.8 | 0.1 |
| pHILIC | 227.125 | 238.5926 | 28.8 | 0.1 |
| pHILIC | 227.125 | 66.0327 | 28.8 | 0.1 |
| pHILIC | 227.125 | 175.9247 | 28.8 | 0.1 |
| pHILIC | 227.125 | 95.9281 | 28.8 | 0.1 |
| pHILIC | 227.125 | 134.97 | 28.7 | 0.1 |
| pHILIC | 227.125 | 98.7042 | 28.7 | 0.1 |
| pHILIC | 227.125 | 73.6512 | 28.7 | 0.1 |
| pHILIC | 227.125 | 193.3816 | 28.7 | 0.1 |
| pHILIC | 227.125 | 69.2788 | 28.7 | 0.1 |
| pHILIC | 227.125 | 102.5979 | 28.7 | 0.1 |
| pHILIC | 227.125 | 73.9876 | 28.7 | 0.1 |
| pHILIC | 227.125 | 186.8202 | 28.7 | 0.1 |
| pHILIC | 227.125 | 59.6845 | 28.7 | 0.1 |
| pHILIC | 227.125 | 181.2868 | 28.7 | 0.1 |
| pHILIC | 227.125 | 118.0177 | 28.7 | 0.1 |
| pHILIC | 227.125 | 73.8647 | 28.7 | 0.1 |
| pHILIC | 227.125 | 148.9795 | 28.7 | 0.1 |
| pHILIC | 227.125 | 56.5603 | 28.7 | 0.1 |
| pHILIC | 227.125 | 68.4592 | 28.7 | 0.1 |
| pHILIC | 227.125 | 141.9557 | 28.7 | 0.1 |
| pHILIC | 227.125 | 56.5528 | 28.7 | 0.1 |
| pHILIC | 227.125 | 163.7011 | 28.7 | 0.1 |
| pHILIC | 227.125 | 82.8236 | 28.6 | 0.1 |
| pHILIC | 227.125 | 89.2746 | 28.6 | 0.1 |
| pHILIC | 227.125 | 67.9574 | 28.6 | 0.1 |
| pHILIC | 227.125 | 154.5606 | 28.6 | 0.1 |
| pHILIC | 227.125 | 71.7378 | 28.6 | 0.1 |
| pHILIC | 227.125 | 86.2596 | 28.6 | 0.1 |
| pHILIC | 227.125 | 79.7372 | 28.6 | 0.1 |
| pHILIC | 227.125 | 73.5117 | 28.6 | 0.1 |
| pHILIC | 227.125 | 52.6237 | 28.6 | 0.1 |
| pHILIC | 227.125 | 53.8367 | 28.6 | 0.1 |
| pHILIC | 227.125 | 84.2228 | 28.6 | 0.1 |
| pHILIC | 227.125 | 90.8733 | 28.6 | 0.1 |
| pHILIC | 227.125 | 158.997 | 28.6 | 0.1 |
| pHILIC | 227.125 | 80.0984 | 28.6 | 0.1 |
| pHILIC | 227.125 | 139.4215 | 28.6 | 0.1 |

| | | | | |
|---|---|---|---|---|
| pHILIC | 227.125 | 58.9311 | 28.6 | 0.1 |
| pHILIC | 227.125 | 101.4572 | 28.6 | 0.1 |
| pHILIC | 227.125 | 168.1713 | 28.6 | 0.1 |
| pHILIC | 227.125 | 110.9096 | 28.6 | 0.1 |
| pHILIC | 227.125 | 204.9392 | 28.6 | 0.1 |
| pHILIC | 227.125 | 123.6211 | 28.6 | 0.1 |
| pHILIC | 227.125 | 59.6058 | 28.6 | 0.1 |
| pHILIC | 227.125 | 223.7694 | 28.6 | 0.1 |
| pHILIC | 227.125 | 208.8764 | 28.6 | 0.1 |
| pHILIC | 227.125 | 60.6502 | 28.6 | 0.1 |
| pHILIC | 227.125 | 61.0854 | 28.6 | 0.1 |
| pHILIC | 227.125 | 119.8284 | 28.6 | 0.1 |
| pHILIC | 227.125 | 119.4432 | 28.5 | 0.1 |
| pHILIC | 227.125 | 70.1707 | 28.5 | 0.1 |
| pHILIC | 227.125 | 85.7248 | 28.5 | 0.1 |
| pHILIC | 227.125 | 74.8406 | 28.5 | 0.1 |
| pHILIC | 227.125 | 240.3279 | 28.5 | 0.1 |
| pHILIC | 227.125 | 141.4855 | 28.5 | 0.1 |
| pHILIC | 227.125 | 182.5835 | 28.5 | 0.1 |
| pHILIC | 227.125 | 161.3695 | 28.5 | 0.1 |
| pHILIC | 227.125 | 67.3294 | 28.5 | 0.1 |
| pHILIC | 227.125 | 56.9286 | 28.5 | 0.1 |
| pHILIC | 227.125 | 53.4715 | 28.5 | 0.1 |
| pHILIC | 227.125 | 84.4329 | 28.5 | 0.1 |
| pHILIC | 227.125 | 96.5051 | 28.5 | 0.1 |
| pHILIC | 227.125 | 153.2107 | 28.5 | 0.1 |
| pHILIC | 227.125 | 95.0877 | 28.5 | 0.1 |
| pHILIC | 227.125 | 197.8187 | 28.5 | 0.1 |
| pHILIC | 227.125 | 101.4627 | 28.5 | 0.1 |
| pHILIC | 227.125 | 151.3094 | 28.5 | 0.1 |
| pHILIC | 227.125 | 90.915 | 28.5 | 0.1 |
| pHILIC | 227.125 | 94.8699 | 28.5 | 0.1 |
| pHILIC | 227.125 | 136.5626 | 28.5 | 0.1 |
| pHILIC | 227.125 | 98.2571 | 28.5 | 0.1 |
| pHILIC | 227.125 | 60.4434 | 28.5 | 0.1 |
| pHILIC | 227.125 | 82.8656 | 28.5 | 0.1 |
| pHILIC | 227.125 | 52.8573 | 28.4 | 0.1 |
| pHILIC | 227.125 | 121.0244 | 28.4 | 0.1 |
| pHILIC | 227.125 | 72.3853 | 28.4 | 0.1 |
| pHILIC | 227.125 | 67.7654 | 28.4 | 0.1 |
| pHILIC | 227.125 | 173.7017 | 28.4 | 0.1 |
| pHILIC | 227.125 | 223.3258 | 28.4 | 0.1 |

| | | | | |
|---|---|---|---|---|
| pHILIC | 227.125 | 120.3921 | 28.4 | 0.1 |
| pHILIC | 227.125 | 61.9283 | 28.4 | 0.1 |
| pHILIC | 227.125 | 50.9185 | 28.4 | 0.1 |
| pHILIC | 227.125 | 82.9285 | 28.4 | 0.1 |
| pHILIC | 227.125 | 50.7831 | 28.4 | 0.1 |
| pHILIC | 227.125 | 56.957 | 28.4 | 0.1 |
| pHILIC | 227.125 | 151.6787 | 28.4 | 0.1 |
| pHILIC | 227.125 | 68.7503 | 28.4 | 0.1 |
| pHILIC | 227.125 | 80.1657 | 28.4 | 0.1 |
| pHILIC | 227.125 | 221.5747 | 28.4 | 0.1 |
| pHILIC | 227.125 | 194.4718 | 28.4 | 0.1 |
| pHILIC | 227.125 | 83.2552 | 28.4 | 0.1 |
| pHILIC | 227.125 | 56.7762 | 28.4 | 0.1 |
| pHILIC | 227.125 | 125.1517 | 28.4 | 0.1 |
| pHILIC | 227.125 | 124.4222 | 28.4 | 0.1 |
| pHILIC | 227.125 | 77.618 | 28.4 | 0.1 |
| pHILIC | 227.125 | 157.9055 | 28.4 | 0.1 |
| pHILIC | 227.125 | 137.5378 | 28.4 | 0.1 |
| pHILIC | 227.125 | 57.006 | 28.4 | 0.1 |
| pHILIC | 227.125 | 90.9814 | 28.4 | 0.1 |
| pHILIC | 227.125 | 159.4674 | 28.4 | 0.1 |
| pHILIC | 227.125 | 103.3848 | 28.3 | 0.1 |
| pHILIC | 227.125 | 152.5458 | 28.3 | 0.1 |
| pHILIC | 227.125 | 69.6041 | 28.3 | 0.1 |
| pHILIC | 227.125 | 68.0462 | 28.3 | 0.1 |
| pHILIC | 227.125 | 52.4049 | 28.3 | 0.1 |
| pHILIC | 227.125 | 251.4723 | 28.3 | 0.1 |
| pHILIC | 227.125 | 52.4466 | 28.3 | 0.1 |
| pHILIC | 227.125 | 58.4075 | 28.3 | 0.1 |
| pHILIC | 227.125 | 142.6469 | 28.3 | 0.1 |
| pHILIC | 227.125 | 61.3345 | 28.3 | 0.1 |
| pHILIC | 227.125 | 64.135 | 28.3 | 0.1 |
| pHILIC | 227.125 | 119.6981 | 28.3 | 0.1 |
| pHILIC | 227.125 | 108.6848 | 28.3 | 0.1 |
| pHILIC | 227.125 | 104.7778 | 28.3 | 0.1 |
| pHILIC | 227.125 | 56.8081 | 28.3 | 0.1 |
| pHILIC | 227.125 | 200.2897 | 28.3 | 0.1 |
| pHILIC | 227.125 | 106.8699 | 28.3 | 0.1 |
| pHILIC | 227.125 | 250.9298 | 28.3 | 0.1 |
| pHILIC | 227.125 | 66.2555 | 28.3 | 0.1 |
| pHILIC | 227.125 | 83.3626 | 28.3 | 0.1 |
| pHILIC | 227.125 | 177.0042 | 28.3 | 0.1 |

| | | | | |
|---|---|---|---|---|
| pHILIC | 227.125 | 83.3911 | 28.2 | 0.1 |
| pHILIC | 227.125 | 111.266 | 28.2 | 0.1 |
| pHILIC | 227.125 | 54.5213 | 28.2 | 0.1 |
| pHILIC | 227.125 | 64.1535 | 28.2 | 0.1 |
| pHILIC | 227.125 | 82.5912 | 28.2 | 0.1 |
| pHILIC | 227.125 | 53.4942 | 28.2 | 0.1 |
| pHILIC | 227.125 | 142.4319 | 28.2 | 0.1 |
| pHILIC | 227.125 | 138.7209 | 28.2 | 0.1 |
| pHILIC | 227.125 | 143.2105 | 28.2 | 0.1 |
| pHILIC | 227.125 | 63.1008 | 28.2 | 0.1 |
| pHILIC | 227.125 | 233.147 | 28.2 | 0.1 |
| pHILIC | 227.125 | 129.1684 | 28.2 | 0.1 |
| pHILIC | 227.125 | 91.7058 | 28.2 | 0.1 |
| pHILIC | 227.125 | 112.6164 | 28.2 | 0.1 |
| pHILIC | 227.125 | 115.2224 | 28.2 | 0.1 |
| pHILIC | 227.125 | 50.5631 | 28.2 | 0.1 |
| pHILIC | 227.125 | 224.8397 | 28.2 | 0.1 |
| pHILIC | 227.125 | 225.5233 | 28.2 | 0.1 |
| pHILIC | 227.125 | 51.5418 | 28.2 | 0.1 |
| pHILIC | 227.125 | 51.297 | 28.2 | 0.1 |
| pHILIC | 227.125 | 94.0938 | 28.2 | 0.1 |
| pHILIC | 227.125 | 79.7212 | 28.2 | 0.1 |
| pHILIC | 227.125 | 148.6418 | 28.2 | 0.1 |
| pHILIC | 227.125 | 56.5724 | 28.2 | 0.1 |
| pHILIC | 227.125 | 115.1371 | 28.2 | 0.1 |
| pHILIC | 227.125 | 175.218 | 28.2 | 0.1 |
| pHILIC | 227.125 | 51.0798 | 28.2 | 0.1 |
| pHILIC | 227.125 | 214.6887 | 28.2 | 0.1 |
| pHILIC | 227.125 | 224.6985 | 28.2 | 0.1 |
| pHILIC | 227.125 | 69.9964 | 28.2 | 0.1 |
| pHILIC | 227.125 | 72.5534 | 28.1 | 0.1 |
| pHILIC | 227.125 | 111.378 | 28.1 | 0.1 |
| pHILIC | 227.125 | 159.5765 | 28.1 | 0.1 |
| pHILIC | 227.125 | 150.0166 | 28.1 | 0.1 |
| pHILIC | 227.125 | 183.4378 | 28.1 | 0.1 |
| pHILIC | 227.125 | 66.7921 | 28.1 | 0.1 |
| pHILIC | 227.125 | 191.4405 | 28.1 | 0.1 |
| pHILIC | 227.125 | 240.5237 | 28.1 | 0.1 |
| pHILIC | 227.125 | 56.2284 | 28.1 | 0.1 |
| pHILIC | 227.125 | 240.1564 | 28.1 | 0.1 |
| pHILIC | 227.125 | 52.8819 | 28.1 | 0.1 |
| pHILIC | 227.125 | 62.1394 | 28.1 | 0.1 |

| | | | | |
|---|---|---|---|---|
| pHILIC | 227.125 | 60.6032 | 28.1 | 0.1 |
| pHILIC | 227.125 | 197.3768 | 28.1 | 0.1 |
| pHILIC | 227.125 | 82.0094 | 28.1 | 0.1 |
| pHILIC | 227.125 | 181.8924 | 28.1 | 0.1 |
| pHILIC | 227.125 | 69.8352 | 28.1 | 0.1 |
| pHILIC | 227.125 | 79.6811 | 28.1 | 0.1 |
| pHILIC | 227.125 | 54.9266 | 28.1 | 0.1 |
| pHILIC | 227.125 | 179.976 | 28.1 | 0.1 |
| pHILIC | 227.125 | 65.6313 | 28.1 | 0.1 |
| pHILIC | 227.125 | 169.7151 | 28.1 | 0.1 |
| pHILIC | 227.125 | 56.84 | 28 | 0.1 |
| pHILIC | 227.125 | 98.0972 | 28 | 0.1 |
| pHILIC | 227.125 | 76.8328 | 28 | 0.1 |
| pHILIC | 227.125 | 76.8514 | 28 | 0.1 |
| pHILIC | 227.125 | 99.6588 | 28 | 0.1 |
| pHILIC | 227.125 | 117.1634 | 28 | 0.1 |
| pHILIC | 227.125 | 64.1665 | 28 | 0.1 |
| pHILIC | 227.125 | 54.2867 | 28 | 0.1 |
| pHILIC | 227.125 | 116.0286 | 28 | 0.1 |
| pHILIC | 227.125 | 119.2395 | 28 | 0.1 |
| pHILIC | 227.125 | 77.5909 | 28 | 0.1 |
| pHILIC | 227.125 | 63.7936 | 28 | 0.1 |
| pHILIC | 227.125 | 56.7342 | 28 | 0.1 |
| pHILIC | 227.125 | 114.5469 | 28 | 0.1 |
| pHILIC | 227.125 | 51.2182 | 28 | 0.1 |
| pHILIC | 227.125 | 58.5975 | 28 | 0.1 |
| pHILIC | 227.125 | 239.0167 | 28 | 0.1 |
| pHILIC | 227.125 | 106.8236 | 28 | 0.1 |
| pHILIC | 227.125 | 72.7303 | 28 | 0.1 |
| pHILIC | 227.125 | 239.2439 | 28 | 0.1 |
| pHILIC | 227.125 | 209.4537 | 28 | 0.1 |
| pHILIC | 227.125 | 174.208 | 28 | 0.1 |
| pHILIC | 227.125 | 52.9326 | 28 | 0.1 |
| pHILIC | 227.125 | 130.379 | 28 | 0.1 |
| pHILIC | 227.125 | 62.8681 | 28 | 0.1 |
| pHILIC | 227.125 | 93.5398 | 27.9 | 0.1 |
| pHILIC | 227.125 | 130.694 | 27.9 | 0.1 |
| pHILIC | 227.125 | 85.6768 | 27.9 | 0.1 |
| pHILIC | 227.125 | 63.8763 | 27.9 | 0.1 |
| pHILIC | 227.125 | 58.1322 | 27.9 | 0.1 |
| pHILIC | 227.125 | 202.8043 | 27.9 | 0.1 |
| pHILIC | 227.125 | 93.2059 | 27.9 | 0.1 |

| | | | | |
|---|---|---|---|---|
| pHILIC | 227.125 | 198.8143 | 27.9 | 0.1 |
| pHILIC | 227.125 | 170.9453 | 27.9 | 0.1 |
| pHILIC | 227.125 | 86.3566 | 27.9 | 0.1 |
| pHILIC | 227.125 | 163.8191 | 27.9 | 0.1 |
| pHILIC | 227.125 | 95.6099 | 27.9 | 0.1 |
| pHILIC | 227.125 | 132.4436 | 27.9 | 0.1 |
| pHILIC | 227.125 | 86.532 | 27.9 | 0.1 |
| pHILIC | 227.125 | 61.5393 | 27.9 | 0.1 |
| pHILIC | 227.125 | 246.2185 | 27.9 | 0.1 |
| pHILIC | 227.125 | 108.3834 | 27.9 | 0.1 |
| pHILIC | 227.125 | 57.4509 | 27.9 | 0.1 |
| pHILIC | 227.125 | 51.4115 | 27.9 | 0.1 |
| pHILIC | 227.125 | 76.023 | 27.9 | 0.1 |
| pHILIC | 227.125 | 91.0489 | 27.9 | 0.1 |
| pHILIC | 227.125 | 53.6658 | 27.9 | 0.1 |
| pHILIC | 227.125 | 92.5248 | 27.9 | 0.1 |
| pHILIC | 227.125 | 70.6404 | 27.8 | 0.1 |
| pHILIC | 227.125 | 146.8865 | 27.8 | 0.1 |
| pHILIC | 227.125 | 111.8035 | 27.8 | 0.1 |
| pHILIC | 227.125 | 222.6874 | 27.8 | 0.1 |
| pHILIC | 227.125 | 55.1505 | 27.8 | 0.1 |
| pHILIC | 227.125 | 180.8167 | 27.8 | 0.1 |
| pHILIC | 227.125 | 56.077 | 27.8 | 0.1 |
| pHILIC | 227.125 | 222.6043 | 27.8 | 0.1 |
| pHILIC | 227.125 | 171.867 | 27.8 | 0.1 |
| pHILIC | 227.125 | 57.0859 | 27.8 | 0.1 |
| pHILIC | 227.125 | 56.1675 | 27.8 | 0.1 |
| pHILIC | 227.125 | 90.4524 | 27.8 | 0.1 |
| pHILIC | 227.125 | 59.156 | 27.8 | 0.1 |
| pHILIC | 227.125 | 78.8222 | 27.8 | 0.1 |
| pHILIC | 227.125 | 56.6353 | 27.8 | 0.1 |
| pHILIC | 227.125 | 202.015 | 27.8 | 0.1 |
| pHILIC | 227.125 | 107.92 | 27.8 | 0.1 |
| pHILIC | 227.125 | 204.9123 | 27.7 | 0.09 |
| pHILIC | 227.125 | 109.9299 | 27.7 | 0.09 |
| pHILIC | 227.125 | 63.5004 | 27.7 | 0.09 |
| pHILIC | 227.125 | 137.7305 | 27.7 | 0.09 |
| pHILIC | 227.125 | 160.2402 | 27.7 | 0.09 |
| pHILIC | 227.125 | 62.6174 | 27.7 | 0.09 |
| pHILIC | 227.125 | 53.8813 | 27.7 | 0.09 |
| pHILIC | 227.125 | 178.0152 | 27.7 | 0.09 |
| pHILIC | 227.125 | 164.4558 | 27.7 | 0.09 |

| | | | | |
|---|---|---|---|---|
| pHILIC | 227.125 | 139.9189 | 27.7 | 0.09 |
| pHILIC | 227.125 | 133.6786 | 27.7 | 0.09 |
| pHILIC | 227.125 | 190.568 | 27.7 | 0.09 |
| pHILIC | 227.125 | 138.2032 | 27.7 | 0.09 |
| pHILIC | 227.125 | 233.1768 | 27.7 | 0.09 |
| pHILIC | 227.125 | 147.4376 | 27.7 | 0.09 |
| pHILIC | 227.125 | 139.9826 | 27.7 | 0.09 |
| pHILIC | 227.125 | 97.8803 | 27.7 | 0.09 |
| pHILIC | 227.125 | 87.1967 | 27.7 | 0.09 |
| pHILIC | 227.125 | 90.313 | 27.7 | 0.09 |
| pHILIC | 227.125 | 88.5027 | 27.7 | 0.09 |
| pHILIC | 227.125 | 78.2693 | 27.6 | 0.09 |
| pHILIC | 227.125 | 91.7301 | 27.6 | 0.09 |
| pHILIC | 227.125 | 70.9311 | 27.6 | 0.09 |
| pHILIC | 227.125 | 178.2548 | 27.6 | 0.09 |
| pHILIC | 227.125 | 65.6378 | 27.6 | 0.09 |
| pHILIC | 227.125 | 130.1486 | 27.6 | 0.09 |
| pHILIC | 227.125 | 61.7887 | 27.6 | 0.09 |
| pHILIC | 227.125 | 62.3074 | 27.6 | 0.09 |
| pHILIC | 227.125 | 99.6123 | 27.6 | 0.09 |
| pHILIC | 227.125 | 236.8131 | 27.6 | 0.09 |
| pHILIC | 227.125 | 71.6445 | 27.6 | 0.09 |
| pHILIC | 227.125 | 218.3433 | 27.6 | 0.09 |
| pHILIC | 227.125 | 204.0439 | 27.5 | 0.09 |
| pHILIC | 227.125 | 154.8075 | 27.5 | 0.09 |
| pHILIC | 227.125 | 92.6439 | 27.5 | 0.09 |
| pHILIC | 227.125 | 117.5311 | 27.5 | 0.09 |
| pHILIC | 227.125 | 73.0687 | 27.5 | 0.09 |
| pHILIC | 227.125 | 178.1379 | 27.5 | 0.09 |
| pHILIC | 227.125 | 70.4949 | 27.5 | 0.09 |
| pHILIC | 227.125 | 76.3958 | 27.5 | 0.09 |
| pHILIC | 227.125 | 105.9386 | 27.5 | 0.09 |
| pHILIC | 227.125 | 127.7377 | 27.5 | 0.09 |
| pHILIC | 227.125 | 161.1692 | 27.5 | 0.09 |
| pHILIC | 227.125 | 55.0505 | 27.5 | 0.09 |
| pHILIC | 227.125 | 89.4171 | 27.5 | 0.09 |
| pHILIC | 227.125 | 107.2208 | 27.5 | 0.09 |
| pHILIC | 227.125 | 64.4058 | 27.5 | 0.09 |
| pHILIC | 227.125 | 128.0046 | 27.5 | 0.09 |
| pHILIC | 227.125 | 63.2738 | 27.5 | 0.09 |
| pHILIC | 227.125 | 53.5769 | 27.5 | 0.09 |
| pHILIC | 227.125 | 68.6161 | 27.5 | 0.09 |

| | | | | |
|---|---|---|---|---|
| pHILIC | 227.125 | 69.0242 | 27.4 | 0.09 |
| pHILIC | 227.125 | 79.7408 | 27.4 | 0.09 |
| pHILIC | 227.125 | 57.3607 | 27.4 | 0.09 |
| pHILIC | 227.125 | 53.6422 | 27.4 | 0.09 |
| pHILIC | 227.125 | 54.2276 | 27.4 | 0.09 |
| pHILIC | 227.125 | 62.1767 | 27.4 | 0.09 |
| pHILIC | 227.125 | 57.9686 | 27.4 | 0.09 |
| pHILIC | 227.125 | 75.7379 | 27.4 | 0.09 |
| pHILIC | 227.125 | 72.9885 | 27.4 | 0.09 |
| pHILIC | 227.125 | 53.2218 | 27.4 | 0.09 |
| pHILIC | 227.125 | 52.6901 | 27.4 | 0.09 |
| pHILIC | 227.125 | 130.5936 | 27.4 | 0.09 |
| pHILIC | 227.125 | 93.6331 | 27.4 | 0.09 |
| pHILIC | 227.125 | 213.2543 | 27.4 | 0.09 |
| pHILIC | 227.125 | 64.442 | 27.4 | 0.09 |
| pHILIC | 227.125 | 116.3387 | 27.3 | 0.09 |
| pHILIC | 227.125 | 56.8426 | 27.3 | 0.09 |
| pHILIC | 227.125 | 231.9106 | 27.3 | 0.09 |
| pHILIC | 227.125 | 87.4814 | 27.3 | 0.09 |
| pHILIC | 227.125 | 150.611 | 27.3 | 0.09 |
| pHILIC | 227.125 | 61.9378 | 27.3 | 0.09 |
| pHILIC | 227.125 | 141.4454 | 27.3 | 0.09 |
| pHILIC | 227.125 | 62.4215 | 27.3 | 0.09 |
| pHILIC | 227.125 | 104.9118 | 27.3 | 0.09 |
| pHILIC | 227.125 | 95.3523 | 27.3 | 0.09 |
| pHILIC | 227.125 | 68.2613 | 27.3 | 0.09 |
| pHILIC | 227.125 | 57.3924 | 27.3 | 0.09 |
| pHILIC | 227.125 | 213.4883 | 27.3 | 0.09 |
| pHILIC | 227.125 | 242.7861 | 27.2 | 0.09 |
| pHILIC | 227.125 | 234.8192 | 27.2 | 0.09 |
| pHILIC | 227.125 | 64.8163 | 27.2 | 0.09 |
| pHILIC | 227.125 | 114.1006 | 27.2 | 0.09 |
| pHILIC | 227.125 | 105.7807 | 27.2 | 0.09 |
| pHILIC | 227.125 | 71.068 | 27.2 | 0.09 |
| pHILIC | 227.125 | 117.4387 | 27.2 | 0.09 |
| pHILIC | 227.125 | 76.8141 | 27.2 | 0.09 |
| pHILIC | 227.125 | 66.2279 | 27.2 | 0.09 |
| pHILIC | 227.125 | 64.6139 | 27.2 | 0.09 |
| pHILIC | 227.125 | 175.9016 | 27.2 | 0.09 |
| pHILIC | 227.125 | 115.9746 | 27.2 | 0.09 |
| pHILIC | 227.125 | 75.8519 | 27.2 | 0.09 |
| pHILIC | 227.125 | 71.3815 | 27.2 | 0.09 |

| | | | | |
|---|---|---|---|---|
| pHILIC | 227.125 | 72.86 | 27.1 | 0.09 |
| pHILIC | 227.125 | 74.6523 | 27.1 | 0.09 |
| pHILIC | 227.125 | 89.9423 | 27.1 | 0.09 |
| pHILIC | 227.125 | 80.76 | 27.1 | 0.09 |
| pHILIC | 227.125 | 144.5397 | 27.1 | 0.09 |
| pHILIC | 227.125 | 56.4785 | 27.1 | 0.09 |
| pHILIC | 227.125 | 86.0892 | 27.1 | 0.09 |
| pHILIC | 227.125 | 106.4657 | 27.1 | 0.09 |
| pHILIC | 227.125 | 99.856 | 27.1 | 0.09 |
| pHILIC | 227.125 | 60.0816 | 27.1 | 0.09 |
| pHILIC | 227.125 | 97.5022 | 27.1 | 0.09 |
| pHILIC | 227.125 | 63.9842 | 27.1 | 0.09 |
| pHILIC | 227.125 | 88.1929 | 27.1 | 0.09 |
| pHILIC | 227.125 | 58.436 | 27.1 | 0.09 |
| pHILIC | 227.125 | 52.4546 | 27.1 | 0.09 |
| pHILIC | 227.125 | 107.3514 | 27.1 | 0.09 |
| pHILIC | 227.125 | 55.7006 | 27.1 | 0.09 |
| pHILIC | 227.125 | 55.5227 | 27.1 | 0.09 |
| pHILIC | 227.125 | 163.8824 | 27.1 | 0.09 |
| pHILIC | 227.125 | 62.4025 | 27.1 | 0.09 |
| pHILIC | 227.125 | 132.6577 | 27 | 0.09 |
| pHILIC | 227.125 | 124.9261 | 27 | 0.09 |
| pHILIC | 227.125 | 67.1233 | 27 | 0.09 |
| pHILIC | 227.125 | 85.5828 | 27 | 0.09 |
| pHILIC | 227.125 | 101.8068 | 27 | 0.09 |
| pHILIC | 227.125 | 50.2153 | 27 | 0.09 |
| pHILIC | 227.125 | 203.8059 | 27 | 0.09 |
| pHILIC | 227.125 | 246.9365 | 27 | 0.09 |
| pHILIC | 227.125 | 99.9927 | 27 | 0.09 |
| pHILIC | 227.125 | 72.0176 | 27 | 0.09 |
| pHILIC | 227.125 | 94.0129 | 27 | 0.09 |
| pHILIC | 227.125 | 97.1442 | 27 | 0.09 |
| pHILIC | 227.125 | 193.5908 | 27 | 0.09 |
| pHILIC | 227.125 | 67.5167 | 27 | 0.09 |
| pHILIC | 227.125 | 224.5136 | 26.9 | 0.09 |
| pHILIC | 227.125 | 67.8156 | 26.9 | 0.09 |
| pHILIC | 227.125 | 154.1141 | 26.9 | 0.09 |
| pHILIC | 227.125 | 103.1532 | 26.9 | 0.09 |
| pHILIC | 227.125 | 62.5215 | 26.9 | 0.09 |
| pHILIC | 227.125 | 51.3937 | 26.9 | 0.09 |
| pHILIC | 227.125 | 51.5039 | 26.9 | 0.09 |
| pHILIC | 227.125 | 68.5187 | 26.9 | 0.09 |

| | | | | |
|---|---|---|---|---|
| pHILIC | 227.125 | 78.7383 | 26.9 | 0.09 |
| pHILIC | 227.125 | 82.46 | 26.9 | 0.09 |
| pHILIC | 227.125 | 117.6044 | 26.9 | 0.09 |
| pHILIC | 227.125 | 64.5298 | 26.9 | 0.09 |
| pHILIC | 227.125 | 90.4041 | 26.9 | 0.09 |
| pHILIC | 227.125 | 64.4488 | 26.8 | 0.09 |
| pHILIC | 227.125 | 54.9372 | 26.8 | 0.09 |
| pHILIC | 227.125 | 167.6066 | 26.8 | 0.09 |
| pHILIC | 227.125 | 61.0513 | 26.8 | 0.09 |
| pHILIC | 227.125 | 185.0479 | 26.8 | 0.09 |
| pHILIC | 227.125 | 62.962 | 26.8 | 0.09 |
| pHILIC | 227.125 | 94.6212 | 26.8 | 0.09 |
| pHILIC | 227.125 | 89.6592 | 26.8 | 0.09 |
| pHILIC | 227.125 | 236.3743 | 26.8 | 0.09 |
| pHILIC | 227.125 | 59.2202 | 26.8 | 0.09 |
| pHILIC | 227.125 | 141.0446 | 26.8 | 0.09 |
| pHILIC | 227.125 | 124.2842 | 26.8 | 0.09 |
| pHILIC | 227.125 | 114.2345 | 26.8 | 0.09 |
| pHILIC | 227.125 | 72.8809 | 26.8 | 0.09 |
| pHILIC | 227.125 | 59.2337 | 26.8 | 0.09 |
| pHILIC | 227.125 | 173.0298 | 26.8 | 0.09 |
| pHILIC | 227.125 | 136.7359 | 26.8 | 0.09 |
| pHILIC | 227.125 | 100.0423 | 26.7 | 0.09 |
| pHILIC | 227.125 | 103.2446 | 26.7 | 0.09 |
| pHILIC | 227.125 | 109.188 | 26.7 | 0.09 |
| pHILIC | 227.125 | 65.617 | 26.7 | 0.09 |
| pHILIC | 227.125 | 81.2416 | 26.7 | 0.09 |
| pHILIC | 227.125 | 163.3272 | 26.7 | 0.09 |
| pHILIC | 227.125 | 184.8189 | 26.7 | 0.09 |
| pHILIC | 227.125 | 135.2534 | 26.7 | 0.09 |
| pHILIC | 227.125 | 124.9969 | 26.7 | 0.09 |
| pHILIC | 227.125 | 69.0202 | 26.7 | 0.09 |
| pHILIC | 227.125 | 165.6688 | 26.7 | 0.09 |
| pHILIC | 227.125 | 142.9558 | 26.7 | 0.09 |
| pHILIC | 227.125 | 203.4228 | 26.7 | 0.09 |
| pHILIC | 227.125 | 236.9958 | 26.7 | 0.09 |
| pHILIC | 227.125 | 69.1194 | 26.7 | 0.09 |
| pHILIC | 227.125 | 149.5182 | 26.7 | 0.09 |
| pHILIC | 227.125 | 84.468 | 26.7 | 0.09 |
| pHILIC | 227.125 | 58.3366 | 26.7 | 0.09 |
| pHILIC | 227.125 | 122.1348 | 26.7 | 0.09 |
| pHILIC | 227.125 | 87.9754 | 26.7 | 0.09 |

| | | | | |
|---|---|---|---|---|
| pHILIC | 227.125 | 63.22 | 26.6 | 0.09 |
| pHILIC | 227.125 | 100.1344 | 26.6 | 0.09 |
| pHILIC | 227.125 | 142.1791 | 26.6 | 0.09 |
| pHILIC | 227.125 | 236.7479 | 26.6 | 0.09 |
| pHILIC | 227.125 | 108.3427 | 26.6 | 0.09 |
| pHILIC | 227.125 | 152.6363 | 26.6 | 0.09 |
| pHILIC | 227.125 | 58.6594 | 26.6 | 0.09 |
| pHILIC | 227.125 | 76.6052 | 26.6 | 0.09 |
| pHILIC | 227.125 | 125.4397 | 26.6 | 0.09 |
| pHILIC | 227.125 | 205.1775 | 26.6 | 0.09 |
| pHILIC | 227.125 | 61.6622 | 26.5 | 0.09 |
| pHILIC | 227.125 | 72.1161 | 26.5 | 0.09 |
| pHILIC | 227.125 | 55.6638 | 26.5 | 0.09 |
| pHILIC | 227.125 | 235.1474 | 26.5 | 0.09 |
| pHILIC | 227.125 | 54.1683 | 26.5 | 0.09 |
| pHILIC | 227.125 | 123.2502 | 26.5 | 0.09 |
| pHILIC | 227.125 | 111.9696 | 26.5 | 0.09 |
| pHILIC | 227.125 | 161.4365 | 26.5 | 0.09 |
| pHILIC | 227.125 | 88.4044 | 26.5 | 0.09 |
| pHILIC | 227.125 | 122.9576 | 26.5 | 0.09 |
| pHILIC | 227.125 | 52.6181 | 26.5 | 0.09 |
| pHILIC | 227.125 | 122.4712 | 26.5 | 0.09 |
| pHILIC | 227.125 | 74.0691 | 26.5 | 0.09 |
| pHILIC | 227.125 | 95.4909 | 26.4 | 0.09 |
| pHILIC | 227.125 | 216.7432 | 26.4 | 0.09 |
| pHILIC | 227.125 | 245.1643 | 26.4 | 0.09 |
| pHILIC | 227.125 | 82.5324 | 26.4 | 0.09 |
| pHILIC | 227.125 | 87.8478 | 26.4 | 0.09 |
| pHILIC | 227.125 | 54.3837 | 26.4 | 0.09 |
| pHILIC | 227.125 | 53.2687 | 26.4 | 0.09 |
| pHILIC | 227.125 | 55.7068 | 26.4 | 0.09 |
| pHILIC | 227.125 | 93.1615 | 26.4 | 0.09 |
| pHILIC | 227.125 | 238.0287 | 26.4 | 0.09 |
| pHILIC | 227.125 | 83.1243 | 26.4 | 0.09 |
| pHILIC | 227.125 | 52.9637 | 26.4 | 0.09 |
| pHILIC | 227.125 | 67.9264 | 26.4 | 0.09 |
| pHILIC | 227.125 | 104.1424 | 26.4 | 0.09 |
| pHILIC | 227.125 | 60.4902 | 26.4 | 0.09 |
| pHILIC | 227.125 | 99.4991 | 26.3 | 0.09 |
| pHILIC | 227.125 | 54.2741 | 26.3 | 0.09 |
| pHILIC | 227.125 | 173.9777 | 26.3 | 0.09 |
| pHILIC | 227.125 | 244.3625 | 26.3 | 0.09 |

| | | | | |
|---|---|---|---|---|
| pHILIC | 227.125 | 209.3331 | 26.3 | 0.09 |
| pHILIC | 227.125 | 86.3223 | 26.3 | 0.09 |
| pHILIC | 227.125 | 56.3245 | 26.3 | 0.09 |
| pHILIC | 227.125 | 113.4341 | 26.3 | 0.09 |
| pHILIC | 227.125 | 101.3973 | 26.3 | 0.09 |
| pHILIC | 227.125 | 56.9643 | 26.3 | 0.09 |
| pHILIC | 227.125 | 53.9141 | 26.3 | 0.09 |
| pHILIC | 227.125 | 157.6004 | 26.2 | 0.09 |
| pHILIC | 227.125 | 105.4363 | 26.2 | 0.09 |
| pHILIC | 227.125 | 134.0955 | 26.2 | 0.09 |
| pHILIC | 227.125 | 148.4383 | 26.2 | 0.09 |
| pHILIC | 227.125 | 197.6689 | 26.2 | 0.09 |
| pHILIC | 227.125 | 66.3455 | 26.2 | 0.09 |
| pHILIC | 227.125 | 152.5742 | 26.2 | 0.09 |
| pHILIC | 227.125 | 64.1128 | 26.2 | 0.09 |
| pHILIC | 227.125 | 89.4832 | 26.2 | 0.09 |
| pHILIC | 227.125 | 73.9058 | 26.2 | 0.09 |
| pHILIC | 227.125 | 71.1865 | 26.2 | 0.09 |
| pHILIC | 227.125 | 121.7885 | 26.2 | 0.09 |
| pHILIC | 227.125 | 62.6845 | 26.2 | 0.09 |
| pHILIC | 227.125 | 113.8839 | 26.2 | 0.09 |
| pHILIC | 227.125 | 225.0572 | 26.2 | 0.09 |
| pHILIC | 227.125 | 53.212 | 26.2 | 0.09 |
| pHILIC | 227.125 | 51.1887 | 26.1 | 0.09 |
| pHILIC | 227.125 | 209.2987 | 26.1 | 0.09 |
| pHILIC | 227.125 | 101.5161 | 26.1 | 0.09 |
| pHILIC | 227.125 | 58.6619 | 26.1 | 0.09 |
| pHILIC | 227.125 | 101.4046 | 26.1 | 0.09 |
| pHILIC | 227.125 | 226.9006 | 26.1 | 0.09 |
| pHILIC | 227.125 | 85.4447 | 26.1 | 0.09 |
| pHILIC | 227.125 | 110.5711 | 26.1 | 0.09 |
| pHILIC | 227.125 | 50.3646 | 26.1 | 0.09 |
| pHILIC | 227.125 | 166.8315 | 26.1 | 0.09 |
| pHILIC | 227.125 | 76.8399 | 26.1 | 0.09 |
| pHILIC | 227.125 | 178.9478 | 26.1 | 0.09 |
| pHILIC | 227.125 | 65.6841 | 26.1 | 0.09 |
| pHILIC | 227.125 | 72.8168 | 26.1 | 0.09 |
| pHILIC | 227.125 | 184.5728 | 26.1 | 0.09 |
| pHILIC | 227.125 | 125.616 | 26 | 0.09 |
| pHILIC | 227.125 | 69.854 | 26 | 0.09 |
| pHILIC | 227.125 | 50.166 | 26 | 0.09 |
| pHILIC | 227.125 | 104.2868 | 26 | 0.09 |

| | | | | |
|---|---|---|---|---|
| pHILIC | 227.125 | 201.5084 | 26 | 0.09 |
| pHILIC | 227.125 | 87.8225 | 26 | 0.09 |
| pHILIC | 227.125 | 95.72 | 26 | 0.09 |
| pHILIC | 227.125 | 56.822 | 25.9 | 0.09 |
| pHILIC | 227.125 | 51.0423 | 25.9 | 0.09 |
| pHILIC | 227.125 | 132.0572 | 25.9 | 0.09 |
| pHILIC | 227.125 | 97.1052 | 25.9 | 0.09 |
| pHILIC | 227.125 | 170.4868 | 25.9 | 0.09 |
| pHILIC | 227.125 | 115.119 | 25.9 | 0.09 |
| pHILIC | 227.125 | 140.2702 | 25.9 | 0.09 |
| pHILIC | 227.125 | 81.4078 | 25.9 | 0.09 |
| pHILIC | 227.125 | 52.4747 | 25.9 | 0.09 |
| pHILIC | 227.125 | 87.6661 | 25.8 | 0.09 |
| pHILIC | 227.125 | 50.8708 | 25.8 | 0.09 |
| pHILIC | 227.125 | 168.8239 | 25.8 | 0.09 |
| pHILIC | 227.125 | 231.8193 | 25.8 | 0.09 |
| pHILIC | 227.125 | 169.2135 | 25.8 | 0.09 |
| pHILIC | 227.125 | 74.7596 | 25.8 | 0.09 |
| pHILIC | 227.125 | 246.7853 | 25.8 | 0.09 |
| pHILIC | 227.125 | 77.427 | 25.8 | 0.09 |
| pHILIC | 227.125 | 192.9013 | 25.8 | 0.09 |
| pHILIC | 227.125 | 50.0299 | 25.8 | 0.09 |
| pHILIC | 227.125 | 228.651 | 25.7 | 0.09 |
| pHILIC | 227.125 | 64.4281 | 25.7 | 0.09 |
| pHILIC | 227.125 | 88.2967 | 25.7 | 0.09 |
| pHILIC | 227.125 | 186.066 | 25.7 | 0.09 |
| pHILIC | 227.125 | 54.1027 | 25.7 | 0.09 |
| pHILIC | 227.125 | 202.5586 | 25.7 | 0.09 |
| pHILIC | 227.125 | 53.9763 | 25.6 | 0.09 |
| pHILIC | 227.125 | 63.3957 | 25.6 | 0.09 |
| pHILIC | 227.125 | 156.3557 | 25.6 | 0.09 |
| pHILIC | 227.125 | 57.6746 | 25.6 | 0.09 |
| pHILIC | 227.125 | 66.4191 | 25.6 | 0.09 |
| pHILIC | 227.125 | 84.8727 | 25.6 | 0.09 |
| pHILIC | 227.125 | 148.1897 | 25.6 | 0.09 |
| pHILIC | 227.125 | 51.0071 | 25.6 | 0.09 |
| pHILIC | 227.125 | 64.1314 | 25.6 | 0.09 |
| pHILIC | 227.125 | 233.6944 | 25.6 | 0.09 |
| pHILIC | 227.125 | 51.887 | 25.6 | 0.09 |
| pHILIC | 227.125 | 244.3293 | 25.6 | 0.09 |
| pHILIC | 227.125 | 75.2419 | 25.6 | 0.09 |
| pHILIC | 227.125 | 74.2187 | 25.5 | 0.09 |

| | | | | |
|---|---|---|---|---|
| pHILIC | 227.125 | 77.6322 | 25.5 | 0.09 |
| pHILIC | 227.125 | 131.1532 | 25.5 | 0.09 |
| pHILIC | 227.125 | 82.7372 | 25.5 | 0.09 |
| pHILIC | 227.125 | 178.2918 | 25.5 | 0.09 |
| pHILIC | 227.125 | 151.6154 | 25.5 | 0.09 |
| pHILIC | 227.125 | 55.5371 | 25.5 | 0.09 |
| pHILIC | 227.125 | 96.6928 | 25.5 | 0.09 |
| pHILIC | 227.125 | 54.0001 | 25.5 | 0.09 |
| pHILIC | 227.125 | 50.3316 | 25.5 | 0.09 |
| pHILIC | 227.125 | 133.1732 | 25.4 | 0.09 |
| pHILIC | 227.125 | 54.4532 | 25.4 | 0.09 |
| pHILIC | 227.125 | 176.2046 | 25.4 | 0.09 |
| pHILIC | 227.125 | 227.0219 | 25.4 | 0.09 |
| pHILIC | 227.125 | 78.2491 | 25.4 | 0.09 |
| pHILIC | 227.125 | 82.9108 | 25.4 | 0.09 |
| pHILIC | 227.125 | 79.1179 | 25.4 | 0.09 |
| pHILIC | 227.125 | 101.2877 | 25.4 | 0.09 |
| pHILIC | 227.125 | 123.5359 | 25.4 | 0.09 |
| pHILIC | 227.125 | 132.9912 | 25.4 | 0.09 |
| pHILIC | 227.125 | 241.2876 | 25.4 | 0.09 |
| pHILIC | 227.125 | 130.568 | 25.4 | 0.09 |
| pHILIC | 227.125 | 214.1538 | 25.4 | 0.09 |
| pHILIC | 227.125 | 84.5383 | 25.3 | 0.09 |
| pHILIC | 227.125 | 81.5749 | 25.3 | 0.09 |
| pHILIC | 227.125 | 77.1773 | 25.3 | 0.09 |
| pHILIC | 227.125 | 77.7457 | 25.3 | 0.09 |
| pHILIC | 227.125 | 116.8869 | 25.3 | 0.09 |
| pHILIC | 227.125 | 99.5884 | 25.3 | 0.09 |
| pHILIC | 227.125 | 222.8321 | 25.3 | 0.09 |
| pHILIC | 227.125 | 99.1336 | 25.3 | 0.09 |
| pHILIC | 227.125 | 66.3797 | 25.3 | 0.09 |
| pHILIC | 227.125 | 103.9535 | 25.3 | 0.09 |
| pHILIC | 227.125 | 196.0632 | 25.3 | 0.09 |
| pHILIC | 227.125 | 59.115 | 25.2 | 0.09 |
| pHILIC | 227.125 | 52.9916 | 25.2 | 0.09 |
| pHILIC | 227.125 | 91.5471 | 25.2 | 0.09 |
| pHILIC | 227.125 | 230.0027 | 25.2 | 0.09 |
| pHILIC | 227.125 | 82.2923 | 25.2 | 0.09 |
| pHILIC | 227.125 | 218.7067 | 25.2 | 0.09 |
| pHILIC | 227.125 | 145.5351 | 25.2 | 0.09 |
| pHILIC | 227.125 | 174.2912 | 25.2 | 0.09 |
| pHILIC | 227.125 | 51.2417 | 25.2 | 0.09 |

| | | | | |
|---|---|---|---|---|
| pHILIC | 227.125 | 195.1056 | 25.2 | 0.09 |
| pHILIC | 227.125 | 69.4323 | 25.2 | 0.09 |
| pHILIC | 227.125 | 67.0654 | 25.2 | 0.09 |
| pHILIC | 227.125 | 63.8881 | 25.2 | 0.09 |
| pHILIC | 227.125 | 69.2857 | 25.2 | 0.09 |
| pHILIC | 227.125 | 78.3391 | 25.1 | 0.09 |
| pHILIC | 227.125 | 84.2929 | 25.1 | 0.09 |
| pHILIC | 227.125 | 68.4783 | 25.1 | 0.09 |
| pHILIC | 227.125 | 220.8827 | 25.1 | 0.09 |
| pHILIC | 227.125 | 126.0095 | 25.1 | 0.09 |
| pHILIC | 227.125 | 180.2866 | 25.1 | 0.09 |
| pHILIC | 227.125 | 237.1964 | 25.1 | 0.09 |
| pHILIC | 227.125 | 221.0207 | 25.1 | 0.09 |
| pHILIC | 227.125 | 91.0428 | 25.1 | 0.09 |
| pHILIC | 227.125 | 63.9173 | 25.1 | 0.09 |
| pHILIC | 227.125 | 75.9417 | 25.1 | 0.09 |
| pHILIC | 227.125 | 81.6074 | 25.1 | 0.09 |
| pHILIC | 227.125 | 219.0283 | 25.1 | 0.09 |
| pHILIC | 227.125 | 152.8648 | 25.1 | 0.09 |
| pHILIC | 227.125 | 108.8225 | 25 | 0.09 |
| pHILIC | 227.125 | 180.9854 | 25 | 0.09 |
| pHILIC | 227.125 | 147.2401 | 25 | 0.09 |
| pHILIC | 227.125 | 134.034 | 25 | 0.09 |
| pHILIC | 227.125 | 116.3661 | 25 | 0.09 |
| pHILIC | 227.125 | 135.5983 | 25 | 0.09 |
| pHILIC | 227.125 | 114.2486 | 25 | 0.09 |
| pHILIC | 227.125 | 141.6697 | 25 | 0.09 |
| pHILIC | 227.125 | 214.0306 | 25 | 0.09 |
| pHILIC | 227.125 | 235.4382 | 25 | 0.09 |
| pHILIC | 227.125 | 248.6202 | 25 | 0.09 |
| pHILIC | 227.125 | 181.6474 | 25 | 0.09 |
| pHILIC | 227.125 | 51.3688 | 24.9 | 0.09 |
| pHILIC | 227.125 | 133.3053 | 24.9 | 0.09 |
| pHILIC | 227.125 | 60.7577 | 24.9 | 0.09 |
| pHILIC | 227.125 | 109.771 | 24.9 | 0.09 |
| pHILIC | 227.125 | 81.9742 | 24.9 | 0.09 |
| pHILIC | 227.125 | 96.0985 | 24.9 | 0.09 |
| pHILIC | 227.125 | 109.3484 | 24.9 | 0.09 |
| pHILIC | 227.125 | 202.1504 | 24.9 | 0.09 |
| pHILIC | 227.125 | 68.4406 | 24.8 | 0.09 |
| pHILIC | 227.125 | 102.5066 | 24.8 | 0.08 |
| pHILIC | 227.125 | 186.386 | 24.8 | 0.08 |

| | | | | |
|---|---|---|---|---|
| pHILIC | 227.125 | 89.5157 | 24.8 | 0.08 |
| pHILIC | 227.125 | 62.5761 | 24.8 | 0.08 |
| pHILIC | 227.125 | 57.8236 | 24.8 | 0.08 |
| pHILIC | 227.125 | 241.9837 | 24.8 | 0.08 |
| pHILIC | 227.125 | 230.6039 | 24.8 | 0.08 |
| pHILIC | 227.125 | 80.3496 | 24.8 | 0.08 |
| pHILIC | 227.125 | 141.8625 | 24.7 | 0.08 |
| pHILIC | 227.125 | 93.9982 | 24.7 | 0.08 |
| pHILIC | 227.125 | 202.3645 | 24.7 | 0.08 |
| pHILIC | 227.125 | 237.081 | 24.7 | 0.08 |
| pHILIC | 227.125 | 217.8988 | 24.7 | 0.08 |
| pHILIC | 227.125 | 65.7346 | 24.7 | 0.08 |
| pHILIC | 227.125 | 61.3008 | 24.7 | 0.08 |
| pHILIC | 227.125 | 51.808 | 24.7 | 0.08 |
| pHILIC | 227.125 | 83.1541 | 24.7 | 0.08 |
| pHILIC | 227.125 | 66.2038 | 24.7 | 0.08 |
| pHILIC | 227.125 | 82.4021 | 24.7 | 0.08 |
| pHILIC | 227.125 | 53.6856 | 24.7 | 0.08 |
| pHILIC | 227.125 | 134.342 | 24.7 | 0.08 |
| pHILIC | 227.125 | 68.0915 | 24.7 | 0.08 |
| pHILIC | 227.125 | 61.1729 | 24.7 | 0.08 |
| pHILIC | 227.125 | 107.073 | 24.7 | 0.08 |
| pHILIC | 227.125 | 157.2245 | 24.6 | 0.08 |
| pHILIC | 227.125 | 89.728 | 24.6 | 0.08 |
| pHILIC | 227.125 | 75.8795 | 24.6 | 0.08 |
| pHILIC | 227.125 | 125.0304 | 24.6 | 0.08 |
| pHILIC | 227.125 | 88.5109 | 24.6 | 0.08 |
| pHILIC | 227.125 | 192.8246 | 24.6 | 0.08 |
| pHILIC | 227.125 | 87.3202 | 24.6 | 0.08 |
| pHILIC | 227.125 | 183.1981 | 24.6 | 0.08 |
| pHILIC | 227.125 | 190.3018 | 24.6 | 0.08 |
| pHILIC | 227.125 | 52.334 | 24.6 | 0.08 |
| pHILIC | 227.125 | 60.4166 | 24.6 | 0.08 |
| pHILIC | 227.125 | 219.7375 | 24.6 | 0.08 |
| pHILIC | 227.125 | 52.7338 | 24.6 | 0.08 |
| pHILIC | 227.125 | 251.196 | 24.6 | 0.08 |
| pHILIC | 227.125 | 162.3087 | 24.5 | 0.08 |
| pHILIC | 227.125 | 131.6278 | 24.5 | 0.08 |
| pHILIC | 227.125 | 120.6501 | 24.5 | 0.08 |
| pHILIC | 227.125 | 53.6439 | 24.5 | 0.08 |
| pHILIC | 227.125 | 139.2504 | 24.5 | 0.08 |
| pHILIC | 227.125 | 152.5866 | 24.5 | 0.08 |

| | | | | |
|---|---|---|---|---|
| pHILIC | 227.125 | 54.4604 | 24.5 | 0.08 |
| pHILIC | 227.125 | 66.2453 | 24.5 | 0.08 |
| pHILIC | 227.125 | 192.1897 | 24.5 | 0.08 |
| pHILIC | 227.125 | 88.5669 | 24.5 | 0.08 |
| pHILIC | 227.125 | 72.7995 | 24.5 | 0.08 |
| pHILIC | 227.125 | 113.4775 | 24.5 | 0.08 |
| pHILIC | 227.125 | 57.5514 | 24.5 | 0.08 |
| pHILIC | 227.125 | 70.5007 | 24.5 | 0.08 |
| pHILIC | 227.125 | 87.4713 | 24.4 | 0.08 |
| pHILIC | 227.125 | 52.9772 | 24.4 | 0.08 |
| pHILIC | 227.125 | 96.9916 | 24.4 | 0.08 |
| pHILIC | 227.125 | 165.4034 | 24.4 | 0.08 |
| pHILIC | 227.125 | 55.9238 | 24.4 | 0.08 |
| pHILIC | 227.125 | 197.09 | 24.4 | 0.08 |
| pHILIC | 227.125 | 139.7663 | 24.3 | 0.08 |
| pHILIC | 227.125 | 58.3159 | 24.3 | 0.08 |
| pHILIC | 227.125 | 170.5374 | 24.3 | 0.08 |
| pHILIC | 227.125 | 73.4003 | 24.3 | 0.08 |
| pHILIC | 227.125 | 86.5789 | 24.3 | 0.08 |
| pHILIC | 227.125 | 151.433 | 24.3 | 0.08 |
| pHILIC | 227.125 | 107.764 | 24.3 | 0.08 |
| pHILIC | 227.125 | 95.8142 | 24.3 | 0.08 |
| pHILIC | 227.125 | 154.332 | 24.2 | 0.08 |
| pHILIC | 227.125 | 60.4371 | 24.2 | 0.08 |
| pHILIC | 227.125 | 50.8127 | 24.2 | 0.08 |
| pHILIC | 227.125 | 142.5134 | 24.2 | 0.08 |
| pHILIC | 227.125 | 141.5237 | 24.2 | 0.08 |
| pHILIC | 227.125 | 76.4102 | 24.2 | 0.08 |
| pHILIC | 227.125 | 159.561 | 24.2 | 0.08 |
| pHILIC | 227.125 | 58.0016 | 24.2 | 0.08 |
| pHILIC | 227.125 | 211.6179 | 24.2 | 0.08 |
| pHILIC | 227.125 | 83.5079 | 24.2 | 0.08 |
| pHILIC | 227.125 | 124.1382 | 24.2 | 0.08 |
| pHILIC | 227.125 | 136.9302 | 24.1 | 0.08 |
| pHILIC | 227.125 | 130.9291 | 24.1 | 0.08 |
| pHILIC | 227.125 | 54.929 | 24.1 | 0.08 |
| pHILIC | 227.125 | 64.1386 | 24.1 | 0.08 |
| pHILIC | 227.125 | 137.6135 | 24.1 | 0.08 |
| pHILIC | 227.125 | 60.3953 | 24.1 | 0.08 |
| pHILIC | 227.125 | 236.717 | 24.1 | 0.08 |
| pHILIC | 227.125 | 56.8765 | 24 | 0.08 |
| pHILIC | 227.125 | 75.4616 | 24 | 0.08 |

| | | | | |
|---|---|---|---|---|
| pHILIC | 227.125 | 55.0945 | 24 | 0.08 |
| pHILIC | 227.125 | 233.5182 | 24 | 0.08 |
| pHILIC | 227.125 | 224.4291 | 24 | 0.08 |
| pHILIC | 227.125 | 126.381 | 24 | 0.08 |
| pHILIC | 227.125 | 63.385 | 24 | 0.08 |
| pHILIC | 227.125 | 229.6246 | 24 | 0.08 |
| pHILIC | 227.125 | 58.69 | 24 | 0.08 |
| pHILIC | 227.125 | 69.2547 | 24 | 0.08 |
| pHILIC | 227.125 | 84.5221 | 23.9 | 0.08 |
| pHILIC | 227.125 | 199.7539 | 23.9 | 0.08 |
| pHILIC | 227.125 | 208.8158 | 23.9 | 0.08 |
| pHILIC | 227.125 | 54.9181 | 23.9 | 0.08 |
| pHILIC | 227.125 | 72.8524 | 23.9 | 0.08 |
| pHILIC | 227.125 | 124.654 | 23.9 | 0.08 |
| pHILIC | 227.125 | 54.312 | 23.9 | 0.08 |
| pHILIC | 227.125 | 71.5267 | 23.9 | 0.08 |
| pHILIC | 227.125 | 114.9661 | 23.8 | 0.08 |
| pHILIC | 227.125 | 65.8309 | 23.8 | 0.08 |
| pHILIC | 227.125 | 127.5721 | 23.8 | 0.08 |
| pHILIC | 227.125 | 199.0476 | 23.8 | 0.08 |
| pHILIC | 227.125 | 83.0798 | 23.8 | 0.08 |
| pHILIC | 227.125 | 57.8201 | 23.8 | 0.08 |
| pHILIC | 227.125 | 220.8173 | 23.8 | 0.08 |
| pHILIC | 227.125 | 136.7048 | 23.8 | 0.08 |
| pHILIC | 227.125 | 60.6229 | 23.8 | 0.08 |
| pHILIC | 227.125 | 146.3789 | 23.8 | 0.08 |
| pHILIC | 227.125 | 67.2726 | 23.7 | 0.08 |
| pHILIC | 227.125 | 57.0987 | 23.7 | 0.08 |
| pHILIC | 227.125 | 118.1366 | 23.7 | 0.08 |
| pHILIC | 227.125 | 228.8456 | 23.7 | 0.08 |
| pHILIC | 227.125 | 76.71 | 23.7 | 0.08 |
| pHILIC | 227.125 | 173.5065 | 23.7 | 0.08 |
| pHILIC | 227.125 | 69.9686 | 23.7 | 0.08 |
| pHILIC | 227.125 | 78.5517 | 23.7 | 0.08 |
| pHILIC | 227.125 | 65.7298 | 23.7 | 0.08 |
| pHILIC | 227.125 | 155.2508 | 23.6 | 0.08 |
| pHILIC | 227.125 | 99.672 | 23.6 | 0.08 |
| pHILIC | 227.125 | 116.4706 | 23.6 | 0.08 |
| pHILIC | 227.125 | 99.3663 | 23.6 | 0.08 |
| pHILIC | 227.125 | 103.9979 | 23.6 | 0.08 |
| pHILIC | 227.125 | 236.0278 | 23.6 | 0.08 |
| pHILIC | 227.125 | 99.8319 | 23.6 | 0.08 |

| | | | | |
|---|---|---|---|---|
| pHILIC | 227.125 | 165.9083 | 23.6 | 0.08 |
| pHILIC | 227.125 | 129.3328 | 23.6 | 0.08 |
| pHILIC | 227.125 | 66.1319 | 23.6 | 0.08 |
| pHILIC | 227.125 | 56.5489 | 23.6 | 0.08 |
| pHILIC | 227.125 | 141.6335 | 23.6 | 0.08 |
| pHILIC | 227.125 | 52.9362 | 23.6 | 0.08 |
| pHILIC | 227.125 | 207.2844 | 23.6 | 0.08 |
| pHILIC | 227.125 | 118.4825 | 23.6 | 0.08 |
| pHILIC | 227.125 | 153.1835 | 23.6 | 0.08 |
| pHILIC | 227.125 | 86.2134 | 23.6 | 0.08 |
| pHILIC | 227.125 | 68.6301 | 23.5 | 0.08 |
| pHILIC | 227.125 | 53.8057 | 23.5 | 0.08 |
| pHILIC | 227.125 | 54.0299 | 23.5 | 0.08 |
| pHILIC | 227.125 | 244.2283 | 23.5 | 0.08 |
| pHILIC | 227.125 | 115.8546 | 23.5 | 0.08 |
| pHILIC | 227.125 | 100.0107 | 23.5 | 0.08 |
| pHILIC | 227.125 | 180.04 | 23.5 | 0.08 |
| pHILIC | 227.125 | 117.9436 | 23.5 | 0.08 |
| pHILIC | 227.125 | 81.7591 | 23.5 | 0.08 |
| pHILIC | 227.125 | 87.2266 | 23.5 | 0.08 |
| pHILIC | 227.125 | 93.2621 | 23.5 | 0.08 |
| pHILIC | 227.125 | 60.6626 | 23.4 | 0.08 |
| pHILIC | 227.125 | 50.0976 | 23.4 | 0.08 |
| pHILIC | 227.125 | 239.1111 | 23.4 | 0.08 |
| pHILIC | 227.125 | 58.1037 | 23.4 | 0.08 |
| pHILIC | 227.125 | 110.5598 | 23.4 | 0.08 |
| pHILIC | 227.125 | 57.0203 | 23.4 | 0.08 |
| pHILIC | 227.125 | 88.0822 | 23.4 | 0.08 |
| pHILIC | 227.125 | 53.34 | 23.4 | 0.08 |
| pHILIC | 227.125 | 85.6596 | 23.4 | 0.08 |
| pHILIC | 227.125 | 220.628 | 23.4 | 0.08 |
| pHILIC | 227.125 | 132.4698 | 23.4 | 0.08 |
| pHILIC | 227.125 | 177.921 | 23.4 | 0.08 |
| pHILIC | 227.125 | 65.3214 | 23.4 | 0.08 |
| pHILIC | 227.125 | 50.6252 | 23.4 | 0.08 |
| pHILIC | 227.125 | 85.1573 | 23.3 | 0.08 |
| pHILIC | 227.125 | 149.4611 | 23.3 | 0.08 |
| pHILIC | 227.125 | 87.5265 | 23.3 | 0.08 |
| pHILIC | 227.125 | 179.2906 | 23.3 | 0.08 |
| pHILIC | 227.125 | 94.4269 | 23.3 | 0.08 |
| pHILIC | 227.125 | 73.2055 | 23.3 | 0.08 |
| pHILIC | 227.125 | 84.7138 | 23.3 | 0.08 |

| | | | | |
|---|---|---|---|---|
| pHILIC | 227.125 | 68.9788 | 23.3 | 0.08 |
| pHILIC | 227.125 | 73.0246 | 23.3 | 0.08 |
| pHILIC | 227.125 | 110.6493 | 23.3 | 0.08 |
| pHILIC | 227.125 | 228.3379 | 23.2 | 0.08 |
| pHILIC | 227.125 | 75.0081 | 23.2 | 0.08 |
| pHILIC | 227.125 | 71.2168 | 23.2 | 0.08 |
| pHILIC | 227.125 | 55.6586 | 23.2 | 0.08 |
| pHILIC | 227.125 | 54.417 | 23.2 | 0.08 |
| pHILIC | 227.125 | 90.3575 | 23.2 | 0.08 |
| pHILIC | 227.125 | 50.5773 | 23.2 | 0.08 |
| pHILIC | 227.125 | 53.5162 | 23.2 | 0.08 |
| pHILIC | 227.125 | 214.2014 | 23.2 | 0.08 |
| pHILIC | 227.125 | 54.8616 | 23.2 | 0.08 |
| pHILIC | 227.125 | 226.7146 | 23.2 | 0.08 |
| pHILIC | 227.125 | 93.4161 | 23.1 | 0.08 |
| pHILIC | 227.125 | 189.8139 | 23.1 | 0.08 |
| pHILIC | 227.125 | 52.55 | 23.1 | 0.08 |
| pHILIC | 227.125 | 110.3685 | 23.1 | 0.08 |
| pHILIC | 227.125 | 105.5731 | 23.1 | 0.08 |
| pHILIC | 227.125 | 50.9906 | 23.1 | 0.08 |
| pHILIC | 227.125 | 79.5328 | 23.1 | 0.08 |
| pHILIC | 227.125 | 66.7049 | 23.1 | 0.08 |
| pHILIC | 227.125 | 118.9873 | 23 | 0.08 |
| pHILIC | 227.125 | 108.8982 | 23 | 0.08 |
| pHILIC | 227.125 | 191.8874 | 23 | 0.08 |
| pHILIC | 227.125 | 55.414 | 23 | 0.08 |
| pHILIC | 227.125 | 111.1205 | 23 | 0.08 |
| pHILIC | 227.125 | 223.6971 | 23 | 0.08 |
| pHILIC | 227.125 | 140.8577 | 23 | 0.08 |
| pHILIC | 227.125 | 84.6883 | 23 | 0.08 |
| pHILIC | 227.125 | 55.7332 | 23 | 0.08 |
| pHILIC | 227.125 | 230.6329 | 23 | 0.08 |
| pHILIC | 227.125 | 57.3989 | 23 | 0.08 |
| pHILIC | 227.125 | 122.7788 | 23 | 0.08 |
| pHILIC | 227.125 | 143.1697 | 23 | 0.08 |
| pHILIC | 227.125 | 85.5199 | 23 | 0.08 |
| pHILIC | 227.125 | 53.3101 | 23 | 0.08 |
| pHILIC | 227.125 | 98.5854 | 22.9 | 0.08 |
| pHILIC | 227.125 | 56.8658 | 22.9 | 0.08 |
| pHILIC | 227.125 | 224.7498 | 22.9 | 0.08 |
| pHILIC | 227.125 | 62.7938 | 22.9 | 0.08 |
| pHILIC | 227.125 | 74.0158 | 22.9 | 0.08 |

| | | | | |
|---|---|---|---|---|
| pHILIC | 227.125 | 84.2096 | 22.9 | 0.08 |
| pHILIC | 227.125 | 54.9832 | 22.9 | 0.08 |
| pHILIC | 227.125 | 83.8351 | 22.9 | 0.08 |
| pHILIC | 227.125 | 86.4348 | 22.9 | 0.08 |
| pHILIC | 227.125 | 178.0447 | 22.9 | 0.08 |
| pHILIC | 227.125 | 56.6634 | 22.8 | 0.08 |
| pHILIC | 227.125 | 99.3761 | 22.8 | 0.08 |
| pHILIC | 227.125 | 90.6212 | 22.8 | 0.08 |
| pHILIC | 227.125 | 129.4794 | 22.8 | 0.08 |
| pHILIC | 227.125 | 101.9683 | 22.8 | 0.08 |
| pHILIC | 227.125 | 124.3738 | 22.8 | 0.08 |
| pHILIC | 227.125 | 60.0397 | 22.8 | 0.08 |
| pHILIC | 227.125 | 148.7248 | 22.8 | 0.08 |
| pHILIC | 227.125 | 61.1982 | 22.8 | 0.08 |
| pHILIC | 227.125 | 242.6148 | 22.7 | 0.08 |
| pHILIC | 227.125 | 182.1845 | 22.7 | 0.08 |
| pHILIC | 227.125 | 50.3605 | 22.7 | 0.08 |
| pHILIC | 227.125 | 55.5709 | 22.7 | 0.08 |
| pHILIC | 227.125 | 67.2136 | 22.7 | 0.08 |
| pHILIC | 227.125 | 68.7714 | 22.7 | 0.08 |
| pHILIC | 227.125 | 53.359 | 22.7 | 0.08 |
| pHILIC | 227.125 | 58.018 | 22.7 | 0.08 |
| pHILIC | 227.125 | 75.9448 | 22.7 | 0.08 |
| pHILIC | 227.125 | 65.4129 | 22.7 | 0.08 |
| pHILIC | 227.125 | 53.8404 | 22.7 | 0.08 |
| pHILIC | 227.125 | 159.1178 | 22.7 | 0.08 |
| pHILIC | 227.125 | 64.8638 | 22.6 | 0.08 |
| pHILIC | 227.125 | 145.2901 | 22.6 | 0.08 |
| pHILIC | 227.125 | 67.2266 | 22.6 | 0.08 |
| pHILIC | 227.125 | 66.3707 | 22.6 | 0.08 |
| pHILIC | 227.125 | 79.319 | 22.6 | 0.08 |
| pHILIC | 227.125 | 220.0732 | 22.6 | 0.08 |
| pHILIC | 227.125 | 51.4846 | 22.6 | 0.08 |
| pHILIC | 227.125 | 78.6036 | 22.6 | 0.08 |
| pHILIC | 227.125 | 176.5806 | 22.6 | 0.08 |
| pHILIC | 227.125 | 72.0435 | 22.6 | 0.08 |
| pHILIC | 227.125 | 54.3649 | 22.6 | 0.08 |
| pHILIC | 227.125 | 127.2143 | 22.6 | 0.08 |
| pHILIC | 227.125 | 71.6049 | 22.6 | 0.08 |
| pHILIC | 227.125 | 161.6783 | 22.6 | 0.08 |
| pHILIC | 227.125 | 97.0521 | 22.6 | 0.08 |
| pHILIC | 227.125 | 121.4473 | 22.5 | 0.08 |

| | | | | |
|---|---|---|---|---|
| pHILIC | 227.125 | 65.307 | 22.5 | 0.08 |
| pHILIC | 227.125 | 86.9813 | 22.5 | 0.08 |
| pHILIC | 227.125 | 121.8081 | 22.5 | 0.08 |
| pHILIC | 227.125 | 53.4879 | 22.5 | 0.08 |
| pHILIC | 227.125 | 215.7682 | 22.5 | 0.08 |
| pHILIC | 227.125 | 128.9539 | 22.5 | 0.08 |
| pHILIC | 227.125 | 162.7567 | 22.5 | 0.08 |
| pHILIC | 227.125 | 56.1356 | 22.5 | 0.08 |
| pHILIC | 227.125 | 61.1678 | 22.5 | 0.08 |
| pHILIC | 227.125 | 61.4526 | 22.5 | 0.08 |
| pHILIC | 227.125 | 88.2734 | 22.5 | 0.08 |
| pHILIC | 227.125 | 72.9466 | 22.4 | 0.08 |
| pHILIC | 227.125 | 60.1739 | 22.4 | 0.08 |
| pHILIC | 227.125 | 165.7823 | 22.4 | 0.08 |
| pHILIC | 227.125 | 61.3089 | 22.4 | 0.08 |
| pHILIC | 227.125 | 63.5825 | 22.4 | 0.08 |
| pHILIC | 227.125 | 85.8677 | 22.4 | 0.08 |
| pHILIC | 227.125 | 94.8298 | 22.3 | 0.08 |
| pHILIC | 227.125 | 71.7048 | 22.3 | 0.08 |
| pHILIC | 227.125 | 58.8081 | 22.3 | 0.08 |
| pHILIC | 227.125 | 60.6458 | 22.3 | 0.08 |
| pHILIC | 227.125 | 76.501 | 22.3 | 0.08 |
| pHILIC | 227.125 | 119.4341 | 22.3 | 0.08 |
| pHILIC | 227.125 | 77.9498 | 22.2 | 0.08 |
| pHILIC | 227.125 | 150.331 | 22.2 | 0.08 |
| pHILIC | 227.125 | 84.7759 | 22.2 | 0.08 |
| pHILIC | 227.125 | 58.6125 | 22.2 | 0.08 |
| pHILIC | 227.125 | 166.3114 | 22.2 | 0.08 |
| pHILIC | 227.125 | 63.7697 | 22.2 | 0.08 |
| pHILIC | 227.125 | 60.7264 | 22.2 | 0.08 |
| pHILIC | 227.125 | 73.409 | 22.2 | 0.08 |
| pHILIC | 227.125 | 189.5561 | 22.2 | 0.08 |
| pHILIC | 227.125 | 55.6556 | 22.2 | 0.08 |
| pHILIC | 227.125 | 58.3232 | 22.2 | 0.08 |
| pHILIC | 227.125 | 51.5724 | 22.1 | 0.08 |
| pHILIC | 227.125 | 192.853 | 22.1 | 0.08 |
| pHILIC | 227.125 | 62.2467 | 22.1 | 0.08 |
| pHILIC | 227.125 | 74.6971 | 22 | 0.08 |
| pHILIC | 227.125 | 112.7105 | 22 | 0.08 |
| pHILIC | 227.125 | 51.1135 | 22 | 0.08 |
| pHILIC | 227.125 | 72.206 | 22 | 0.08 |
| pHILIC | 227.125 | 153.6673 | 22 | 0.08 |

| | | | | |
|---|---|---|---|---|
| pHILIC | 227.125 | 58.7891 | 22 | 0.08 |
| pHILIC | 227.125 | 73.137 | 22 | 0.08 |
| pHILIC | 227.125 | 74.639 | 22 | 0.08 |
| pHILIC | 227.125 | 116.266 | 22 | 0.08 |
| pHILIC | 227.125 | 73.9676 | 22 | 0.08 |
| pHILIC | 227.125 | 77.0068 | 22 | 0.08 |
| pHILIC | 227.125 | 59.7137 | 22 | 0.08 |
| pHILIC | 227.125 | 79.0115 | 22 | 0.08 |
| pHILIC | 227.125 | 60.4049 | 22 | 0.08 |
| pHILIC | 227.125 | 108.5 | 21.9 | 0.08 |
| pHILIC | 227.125 | 77.5329 | 21.9 | 0.08 |
| pHILIC | 227.125 | 59.7264 | 21.9 | 0.08 |
| pHILIC | 227.125 | 131.5864 | 21.9 | 0.07 |
| pHILIC | 227.125 | 88.5219 | 21.9 | 0.07 |
| pHILIC | 227.125 | 167.7649 | 21.9 | 0.07 |
| pHILIC | 227.125 | 121.4908 | 21.8 | 0.07 |
| pHILIC | 227.125 | 51.7047 | 21.8 | 0.07 |
| pHILIC | 227.125 | 87.7206 | 21.8 | 0.07 |
| pHILIC | 227.125 | 195.9801 | 21.7 | 0.07 |
| pHILIC | 227.125 | 78.0931 | 21.7 | 0.07 |
| pHILIC | 227.125 | 211.8499 | 21.7 | 0.07 |
| pHILIC | 227.125 | 190.6075 | 21.7 | 0.07 |
| pHILIC | 227.125 | 167.9214 | 21.7 | 0.07 |
| pHILIC | 227.125 | 52.8083 | 21.7 | 0.07 |
| pHILIC | 227.125 | 60.1661 | 21.7 | 0.07 |
| pHILIC | 227.125 | 51.8576 | 21.6 | 0.07 |
| pHILIC | 227.125 | 80.6711 | 21.6 | 0.07 |
| pHILIC | 227.125 | 57.3147 | 21.6 | 0.07 |
| pHILIC | 227.125 | 87.0683 | 21.6 | 0.07 |
| pHILIC | 227.125 | 83.7738 | 21.5 | 0.07 |
| pHILIC | 227.125 | 73.2144 | 21.5 | 0.07 |
| pHILIC | 227.125 | 147.3237 | 21.5 | 0.07 |
| pHILIC | 227.125 | 85.0331 | 21.5 | 0.07 |
| pHILIC | 227.125 | 186.94 | 21.5 | 0.07 |
| pHILIC | 227.125 | 64.7436 | 21.4 | 0.07 |
| pHILIC | 227.125 | 76.66 | 21.4 | 0.07 |
| pHILIC | 227.125 | 139.9614 | 21.4 | 0.07 |
| pHILIC | 227.125 | 153.5001 | 21.4 | 0.07 |
| pHILIC | 227.125 | 163.4244 | 21.4 | 0.07 |
| pHILIC | 227.125 | 121.8855 | 21.4 | 0.07 |
| pHILIC | 227.125 | 179.242 | 21.4 | 0.07 |
| pHILIC | 227.125 | 51.8554 | 21.4 | 0.07 |

| | | | | |
|---|---|---|---|---|
| pHILIC | 227.125 | 81.0063 | 21.4 | 0.07 |
| pHILIC | 227.125 | 61.3712 | 21.4 | 0.07 |
| pHILIC | 227.125 | 232.8948 | 21.3 | 0.07 |
| pHILIC | 227.125 | 156.7282 | 21.3 | 0.07 |
| pHILIC | 227.125 | 71.683 | 21.3 | 0.07 |
| pHILIC | 227.125 | 51.8542 | 21.3 | 0.07 |
| pHILIC | 227.125 | 54.0341 | 21.2 | 0.07 |
| pHILIC | 227.125 | 89.3372 | 21.2 | 0.07 |
| pHILIC | 227.125 | 71.2471 | 21.1 | 0.07 |
| pHILIC | 227.125 | 63.0634 | 21 | 0.07 |
| pHILIC | 227.125 | 53.5457 | 21 | 0.07 |
| pHILIC | 227.125 | 89.4268 | 21 | 0.07 |
| pHILIC | 227.125 | 65.7029 | 21 | 0.07 |
| pHILIC | 227.125 | 166.4812 | 21 | 0.07 |
| pHILIC | 227.125 | 92.7197 | 20.9 | 0.07 |
| pHILIC | 227.125 | 57.1935 | 20.9 | 0.07 |
| pHILIC | 227.125 | 219.6871 | 20.9 | 0.07 |
| pHILIC | 227.125 | 119.9652 | 20.9 | 0.07 |
| pHILIC | 227.125 | 82.2636 | 20.8 | 0.07 |
| pHILIC | 227.125 | 71.9767 | 20.7 | 0.07 |
| pHILIC | 227.125 | 63.5411 | 20.7 | 0.07 |
| pHILIC | 227.125 | 99.8033 | 20.6 | 0.07 |
| pHILIC | 227.125 | 166.1107 | 20.5 | 0.07 |
| pHILIC | 227.125 | 77.7834 | 20.4 | 0.07 |
| pHILIC | 227.125 | 116.5056 | 20.4 | 0.07 |
| pHILIC | 227.125 | 131.1381 | 20.4 | 0.07 |
| pHILIC | 227.125 | 89.6013 | 20.3 | 0.07 |
| pHILIC | 227.125 | 70.6153 | 20.2 | 0.07 |
| pHILIC | 227.125 | 89.1045 | 20.2 | 0.07 |
| pHILIC | 227.125 | 140.6443 | 20.2 | 0.07 |
| pHILIC | 227.125 | 97.1809 | 20.1 | 0.07 |
| pHILIC | 227.125 | 50.3776 | 20 | 0.07 |
| pHILIC | 227.125 | 81.7474 | 19.7 | 0.07 |
| pHILIC | 227.125 | 55.8284 | 19.6 | 0.07 |
| pHILIC | 227.125 | 55.4379 | 19.6 | 0.07 |
| pHILIC | 227.125 | 87.8971 | 19.1 | 0.07 |
| pHILIC | 389.231 | 105.0698 | 272323.8 | 100 |
| pHILIC | 389.231 | 106.0731 | 155734.7 | 57.19 |
| pHILIC | 389.231 | 107.0765 | 36628.7 | 13.45 |
| pHILIC | 389.231 | 89.0596 | 24318.2 | 8.93 |
| pHILIC | 389.231 | 389.1772 | 12404.6 | 4.56 |
| pHILIC | 389.231 | 372.2304 | 10376.8 | 3.81 |

| | | | | |
|---|---|---|---|---|
| pHILIC | 389.231 | 228.2196 | 6825.8 | 2.51 |
| pHILIC | 389.231 | 133.0859 | 5554.8 | 2.04 |
| pHILIC | 389.231 | 119.0492 | 3486.7 | 1.28 |
| pHILIC | 389.231 | 79.0541 | 2988.1 | 1.1 |
| pHILIC | 389.231 | 90.063 | 2327 | 0.85 |
| pHILIC | 389.231 | 176.8753 | 2117.1 | 0.78 |
| pHILIC | 389.231 | 389.0949 | 1987.5 | 0.73 |
| pHILIC | 389.231 | 120.0524 | 1984.8 | 0.73 |
| pHILIC | 389.231 | 121.0648 | 1744.8 | 0.64 |
| pHILIC | 389.231 | 134.0894 | 1741.6 | 0.64 |
| pHILIC | 389.231 | 211.0789 | 1415.5 | 0.52 |
| pHILIC | 389.231 | 103.0541 | 1404.2 | 0.52 |
| pHILIC | 389.231 | 57.0698 | 1090.7 | 0.4 |
| pHILIC | 389.231 | 105.0336 | 1084.1 | 0.4 |
| pHILIC | 389.231 | 122.0679 | 900.8 | 0.33 |
| pHILIC | 389.231 | 81.0334 | 858.4 | 0.32 |
| pHILIC | 389.231 | 80.0575 | 849.3 | 0.31 |
| pHILIC | 389.231 | 93.0698 | 846.6 | 0.31 |
| pHILIC | 389.231 | 178.1155 | 808.7 | 0.3 |
| pHILIC | 389.231 | 177.112 | 795.7 | 0.29 |
| pHILIC | 389.231 | 69.0335 | 747.5 | 0.27 |
| pHILIC | 389.231 | 104.0574 | 610.4 | 0.22 |
| pHILIC | 389.231 | 177.0888 | 346 | 0.13 |
| pHILIC | 389.231 | 121.0549 | 344.9 | 0.13 |
| pHILIC | 389.231 | 105.0783 | 160.5 | 0.06 |
| pHILIC | 389.231 | 232.8096 | 153 | 0.06 |
| pHILIC | 389.231 | 107.0687 | 143.9 | 0.05 |
| pHILIC | 389.231 | 148.5711 | 138.2 | 0.05 |
| pHILIC | 389.231 | 88.6934 | 135.2 | 0.05 |
| pHILIC | 389.231 | 94.8619 | 125.1 | 0.05 |
| pHILIC | 389.231 | 107.0845 | 122.2 | 0.04 |
| pHILIC | 389.231 | 53.0385 | 114.7 | 0.04 |
| pHILIC | 389.231 | 91.0539 | 112.1 | 0.04 |
| pHILIC | 389.231 | 105.0613 | 110 | 0.04 |
| pHILIC | 389.231 | 248.2546 | 106.9 | 0.04 |
| pHILIC | 389.231 | 85.0281 | 105 | 0.04 |
| pHILIC | 389.231 | 362.0918 | 103.1 | 0.04 |
| pHILIC | 389.231 | 259.6483 | 102.5 | 0.04 |
| pHILIC | 389.231 | 381.0045 | 100 | 0.04 |
| pHILIC | 389.231 | 65.5846 | 99.5 | 0.04 |
| pHILIC | 389.231 | 359.4657 | 98.3 | 0.04 |
| pHILIC | 389.231 | 318.1005 | 97.6 | 0.04 |

| | | | | |
|---|---|---|---|---|
| pHILIC | 389.231 | 200.2634 | 97.6 | 0.04 |
| pHILIC | 389.231 | 253.3633 | 97.5 | 0.04 |
| pHILIC | 389.231 | 275.735 | 97.3 | 0.04 |
| pHILIC | 389.231 | 78.8153 | 95.6 | 0.04 |
| pHILIC | 389.231 | 306.5265 | 95.6 | 0.04 |
| pHILIC | 389.231 | 263.3948 | 95.4 | 0.04 |
| pHILIC | 389.231 | 181.4679 | 94.5 | 0.03 |
| pHILIC | 389.231 | 136.6762 | 94.3 | 0.03 |
| pHILIC | 389.231 | 251.6072 | 94 | 0.03 |
| pHILIC | 389.231 | 232.044 | 93.4 | 0.03 |
| pHILIC | 389.231 | 309.6367 | 93.4 | 0.03 |
| pHILIC | 389.231 | 298.6619 | 92.3 | 0.03 |
| pHILIC | 389.231 | 133.2652 | 91.2 | 0.03 |
| pHILIC | 389.231 | 251.7645 | 91.1 | 0.03 |
| pHILIC | 389.231 | 204.947 | 91 | 0.03 |
| pHILIC | 389.231 | 65.9965 | 90.8 | 0.03 |
| pHILIC | 389.231 | 61.6362 | 90.7 | 0.03 |
| pHILIC | 389.231 | 61.3873 | 90.7 | 0.03 |
| pHILIC | 389.231 | 69.6158 | 90.5 | 0.03 |
| pHILIC | 389.231 | 310.2867 | 90.2 | 0.03 |
| pHILIC | 389.231 | 55.952 | 90.1 | 0.03 |
| pHILIC | 389.231 | 114.5268 | 89.6 | 0.03 |
| pHILIC | 389.231 | 121.8331 | 89.4 | 0.03 |
| pHILIC | 389.231 | 140.3863 | 89.1 | 0.03 |
| pHILIC | 389.231 | 178.5811 | 89 | 0.03 |
| pHILIC | 389.231 | 295.4026 | 88.6 | 0.03 |
| pHILIC | 389.231 | 76.3699 | 88.6 | 0.03 |
| pHILIC | 389.231 | 120.8424 | 88.4 | 0.03 |
| pHILIC | 389.231 | 338.3366 | 88.4 | 0.03 |
| pHILIC | 389.231 | 83.7986 | 87.7 | 0.03 |
| pHILIC | 389.231 | 56.7461 | 87.7 | 0.03 |
| pHILIC | 389.231 | 93.5528 | 87.6 | 0.03 |
| pHILIC | 389.231 | 201.1855 | 87.4 | 0.03 |
| pHILIC | 389.231 | 98.1892 | 86.8 | 0.03 |
| pHILIC | 389.231 | 184.5917 | 86.1 | 0.03 |
| pHILIC | 389.231 | 63.7007 | 85.9 | 0.03 |
| pHILIC | 389.231 | 56.451 | 84.7 | 0.03 |
| pHILIC | 389.231 | 139.3448 | 83.7 | 0.03 |
| pHILIC | 389.231 | 115.29 | 83.4 | 0.03 |
| pHILIC | 389.231 | 172.2554 | 83.2 | 0.03 |
| pHILIC | 389.231 | 53.7834 | 82.9 | 0.03 |
| pHILIC | 389.231 | 94.9478 | 81.5 | 0.03 |

| | | | | |
|---|---|---|---|---|
| pHILIC | 389.231 | 82.5934 | 79.9 | 0.03 |
| pHILIC | 389.231 | 85.794 | 79.7 | 0.03 |
| pHILIC | 389.231 | 83.0486 | 73.5 | 0.03 |
| pHILIC | 389.231 | 106.0919 | 69.1 | 0.03 |
| pHILIC | 389.231 | 371.2279 | 62.5 | 0.02 |
| pHILIC | 389.231 | 60.21 | 61.5 | 0.02 |
| pHILIC | 389.231 | 403.5479 | 61.3 | 0.02 |
| pHILIC | 389.231 | 70.8659 | 60.5 | 0.02 |
| pHILIC | 389.231 | 289.1151 | 60.5 | 0.02 |
| pHILIC | 389.231 | 373.966 | 60.4 | 0.02 |
| pHILIC | 389.231 | 294.3953 | 60.3 | 0.02 |
| pHILIC | 389.231 | 156.1932 | 60 | 0.02 |
| pHILIC | 389.231 | 55.4837 | 59.6 | 0.02 |
| pHILIC | 389.231 | 347.2767 | 59.6 | 0.02 |
| pHILIC | 389.231 | 92.886 | 59.4 | 0.02 |
| pHILIC | 389.231 | 62.4788 | 59.2 | 0.02 |
| pHILIC | 389.231 | 412.0104 | 58.3 | 0.02 |
| pHILIC | 389.231 | 410.0841 | 58.3 | 0.02 |
| pHILIC | 389.231 | 70.0366 | 58.2 | 0.02 |
| pHILIC | 389.231 | 211.2533 | 58 | 0.02 |
| pHILIC | 389.231 | 316.3996 | 57.3 | 0.02 |
| pHILIC | 389.231 | 222.1511 | 57.2 | 0.02 |
| pHILIC | 389.231 | 148.7817 | 57.2 | 0.02 |
| pHILIC | 389.231 | 211.0987 | 57.1 | 0.02 |
| pHILIC | 389.231 | 220.7899 | 57 | 0.02 |
| pHILIC | 389.231 | 57.5914 | 57 | 0.02 |
| pHILIC | 389.231 | 336.3451 | 56.9 | 0.02 |
| pHILIC | 389.231 | 177.9386 | 56.9 | 0.02 |
| pHILIC | 389.231 | 216.5697 | 56.8 | 0.02 |
| pHILIC | 389.231 | 138.1838 | 56.7 | 0.02 |
| pHILIC | 389.231 | 136.6537 | 56.6 | 0.02 |
| pHILIC | 389.231 | 73.6697 | 55.9 | 0.02 |
| pHILIC | 389.231 | 390.1392 | 55.9 | 0.02 |
| pHILIC | 389.231 | 85.9251 | 55.8 | 0.02 |
| pHILIC | 389.231 | 121.418 | 55.7 | 0.02 |
| pHILIC | 389.231 | 393.7257 | 55.7 | 0.02 |
| pHILIC | 389.231 | 389.2555 | 55.4 | 0.02 |
| pHILIC | 389.231 | 84.7735 | 55.3 | 0.02 |
| pHILIC | 389.231 | 93.5263 | 55.3 | 0.02 |
| pHILIC | 389.231 | 56.4194 | 55.3 | 0.02 |
| pHILIC | 389.231 | 81.5846 | 55.2 | 0.02 |
| pHILIC | 389.231 | 198.1682 | 55.1 | 0.02 |

| | | | | |
|---|---|---|---|---|
| pHILIC | 389.231 | 68.1685 | 55.1 | 0.02 |
| pHILIC | 389.231 | 340.0468 | 55 | 0.02 |
| pHILIC | 389.231 | 91.7587 | 55 | 0.02 |
| pHILIC | 389.231 | 328.411 | 54.9 | 0.02 |
| pHILIC | 389.231 | 243.4544 | 54.8 | 0.02 |
| pHILIC | 389.231 | 395.0221 | 54.8 | 0.02 |
| pHILIC | 389.231 | 192.6141 | 54.7 | 0.02 |
| pHILIC | 389.231 | 108.6395 | 54.7 | 0.02 |
| pHILIC | 389.231 | 94.1811 | 54.6 | 0.02 |
| pHILIC | 389.231 | 69.1905 | 54.5 | 0.02 |
| pHILIC | 389.231 | 162.1933 | 54.5 | 0.02 |
| pHILIC | 389.231 | 94.2077 | 54.4 | 0.02 |
| pHILIC | 389.231 | 69.0701 | 54.4 | 0.02 |
| pHILIC | 389.231 | 187.2251 | 54.4 | 0.02 |
| pHILIC | 389.231 | 217.4698 | 54.4 | 0.02 |
| pHILIC | 389.231 | 51.1985 | 54.3 | 0.02 |
| pHILIC | 389.231 | 62.5331 | 54.3 | 0.02 |
| pHILIC | 389.231 | 125.2687 | 54.2 | 0.02 |
| pHILIC | 389.231 | 72.0079 | 54.1 | 0.02 |
| pHILIC | 389.231 | 280.7056 | 53.9 | 0.02 |
| pHILIC | 389.231 | 92.0539 | 53.9 | 0.02 |
| pHILIC | 389.231 | 401.1622 | 53.8 | 0.02 |
| pHILIC | 389.231 | 378.7753 | 53.7 | 0.02 |
| pHILIC | 389.231 | 136.6943 | 53.7 | 0.02 |
| pHILIC | 389.231 | 197.5655 | 53.7 | 0.02 |
| pHILIC | 389.231 | 73.81 | 53.6 | 0.02 |
| pHILIC | 389.231 | 79.9997 | 53.5 | 0.02 |
| pHILIC | 389.231 | 89.866 | 53.3 | 0.02 |
| pHILIC | 389.231 | 287.0043 | 53.3 | 0.02 |
| pHILIC | 389.231 | 212.1745 | 53.1 | 0.02 |
| pHILIC | 389.231 | 158.8689 | 53.1 | 0.02 |
| pHILIC | 389.231 | 392.5118 | 53.1 | 0.02 |
| pHILIC | 389.231 | 82.0918 | 53 | 0.02 |
| pHILIC | 389.231 | 179.9093 | 53 | 0.02 |
| pHILIC | 389.231 | 111.7666 | 53 | 0.02 |
| pHILIC | 389.231 | 354.6545 | 53 | 0.02 |
| pHILIC | 389.231 | 154.4057 | 52.9 | 0.02 |
| pHILIC | 389.231 | 135.5923 | 52.9 | 0.02 |
| pHILIC | 389.231 | 105.3476 | 52.9 | 0.02 |
| pHILIC | 389.231 | 51.5626 | 52.7 | 0.02 |
| pHILIC | 389.231 | 103.3227 | 52.7 | 0.02 |
| pHILIC | 389.231 | 87.3235 | 52.7 | 0.02 |

| | | | | |
|---|---|---|---|---|
| pHILIC | 389.231 | 258.5884 | 52.6 | 0.02 |
| pHILIC | 389.231 | 56.9187 | 52.5 | 0.02 |
| pHILIC | 389.231 | 50.5829 | 52.5 | 0.02 |
| pHILIC | 389.231 | 411.5993 | 52.5 | 0.02 |
| pHILIC | 389.231 | 94.4795 | 52.4 | 0.02 |
| pHILIC | 389.231 | 286.7886 | 52.3 | 0.02 |
| pHILIC | 389.231 | 223.3683 | 52.3 | 0.02 |
| pHILIC | 389.231 | 332.449 | 52.3 | 0.02 |
| pHILIC | 389.231 | 115.9914 | 52.2 | 0.02 |
| pHILIC | 389.231 | 219.3994 | 52.2 | 0.02 |
| pHILIC | 389.231 | 269.6048 | 52.1 | 0.02 |
| pHILIC | 389.231 | 384.694 | 52.1 | 0.02 |
| pHILIC | 389.231 | 137.06 | 52.1 | 0.02 |
| pHILIC | 389.231 | 107.6839 | 52.1 | 0.02 |
| pHILIC | 389.231 | 66.777 | 52 | 0.02 |
| pHILIC | 389.231 | 65.105 | 52 | 0.02 |
| pHILIC | 389.231 | 75.2367 | 51.9 | 0.02 |
| pHILIC | 389.231 | 248.7569 | 51.9 | 0.02 |
| pHILIC | 389.231 | 274.6011 | 51.9 | 0.02 |
| pHILIC | 389.231 | 246.7411 | 51.9 | 0.02 |
| pHILIC | 389.231 | 339.4695 | 51.9 | 0.02 |
| pHILIC | 389.231 | 219.1777 | 51.9 | 0.02 |
| pHILIC | 389.231 | 249.606 | 51.8 | 0.02 |
| pHILIC | 389.231 | 220.8472 | 51.8 | 0.02 |
| pHILIC | 389.231 | 200.5474 | 51.8 | 0.02 |
| pHILIC | 389.231 | 171.1753 | 51.8 | 0.02 |
| pHILIC | 389.231 | 72.5221 | 51.8 | 0.02 |
| pHILIC | 389.231 | 65.2444 | 51.7 | 0.02 |
| pHILIC | 389.231 | 51.5017 | 51.7 | 0.02 |
| pHILIC | 389.231 | 319.4427 | 51.7 | 0.02 |
| pHILIC | 389.231 | 102.4675 | 51.7 | 0.02 |
| pHILIC | 389.231 | 124.5517 | 51.6 | 0.02 |
| pHILIC | 389.231 | 79.3371 | 51.6 | 0.02 |
| pHILIC | 389.231 | 361.1868 | 51.6 | 0.02 |
| pHILIC | 389.231 | 77.1939 | 51.6 | 0.02 |
| pHILIC | 389.231 | 54.752 | 51.5 | 0.02 |
| pHILIC | 389.231 | 148.5224 | 51.5 | 0.02 |
| pHILIC | 389.231 | 166.0762 | 51.5 | 0.02 |
| pHILIC | 389.231 | 122.2414 | 51.5 | 0.02 |
| pHILIC | 389.231 | 69.3548 | 51.5 | 0.02 |
| pHILIC | 389.231 | 94.1023 | 51.5 | 0.02 |
| pHILIC | 389.231 | 93.2477 | 51.5 | 0.02 |

| | | | | |
|---|---|---|---|---|
| pHILIC | 389.231 | 54.6372 | 51.5 | 0.02 |
| pHILIC | 389.231 | 50.2977 | 51.4 | 0.02 |
| pHILIC | 389.231 | 106.2985 | 51.4 | 0.02 |
| pHILIC | 389.231 | 269.0465 | 51.4 | 0.02 |
| pHILIC | 389.231 | 108.8224 | 51.4 | 0.02 |
| pHILIC | 389.231 | 153.2172 | 51.4 | 0.02 |
| pHILIC | 389.231 | 88.7676 | 51.4 | 0.02 |
| pHILIC | 389.231 | 146.4054 | 51.4 | 0.02 |
| pHILIC | 389.231 | 60.9044 | 51.4 | 0.02 |
| pHILIC | 389.231 | 149.9268 | 51.3 | 0.02 |
| pHILIC | 389.231 | 151.8728 | 51.3 | 0.02 |
| pHILIC | 389.231 | 170.0282 | 51.3 | 0.02 |
| pHILIC | 389.231 | 120.7222 | 51.2 | 0.02 |
| pHILIC | 389.231 | 323.5779 | 51.2 | 0.02 |
| pHILIC | 389.231 | 76.217 | 51.2 | 0.02 |
| pHILIC | 389.231 | 51.7544 | 51.2 | 0.02 |
| pHILIC | 389.231 | 78.3686 | 51.1 | 0.02 |
| pHILIC | 389.231 | 411.0672 | 51.1 | 0.02 |
| pHILIC | 389.231 | 279.693 | 51 | 0.02 |
| pHILIC | 389.231 | 53.4076 | 51 | 0.02 |
| pHILIC | 389.231 | 85.7056 | 51 | 0.02 |
| pHILIC | 389.231 | 335.8193 | 51 | 0.02 |
| pHILIC | 389.231 | 242.7723 | 51 | 0.02 |
| pHILIC | 389.231 | 95.0022 | 51 | 0.02 |
| pHILIC | 389.231 | 120.0419 | 50.9 | 0.02 |
| pHILIC | 389.231 | 76.6763 | 50.9 | 0.02 |
| pHILIC | 389.231 | 152.6493 | 50.9 | 0.02 |
| pHILIC | 389.231 | 69.1461 | 50.8 | 0.02 |
| pHILIC | 389.231 | 258.9818 | 50.7 | 0.02 |
| pHILIC | 389.231 | 69.9002 | 50.7 | 0.02 |
| pHILIC | 389.231 | 150.3693 | 50.7 | 0.02 |
| pHILIC | 389.231 | 61.0378 | 50.6 | 0.02 |
| pHILIC | 389.231 | 99.4404 | 50.6 | 0.02 |
| pHILIC | 389.231 | 268.5277 | 50.6 | 0.02 |
| pHILIC | 389.231 | 99.8106 | 50.6 | 0.02 |
| pHILIC | 389.231 | 51.3307 | 50.5 | 0.02 |
| pHILIC | 389.231 | 60.1011 | 50.5 | 0.02 |
| pHILIC | 389.231 | 202.5076 | 50.5 | 0.02 |
| pHILIC | 389.231 | 57.9092 | 50.5 | 0.02 |
| pHILIC | 389.231 | 190.0675 | 50.5 | 0.02 |
| pHILIC | 389.231 | 295.8694 | 50.5 | 0.02 |
| pHILIC | 389.231 | 395.776 | 50.5 | 0.02 |

| | | | | |
|---|---|---|---|---|
| pHILIC | 389.231 | 57.1909 | 50.4 | 0.02 |
| pHILIC | 389.231 | 80.819 | 50.4 | 0.02 |
| pHILIC | 389.231 | 50.5841 | 50.4 | 0.02 |
| pHILIC | 389.231 | 246.1237 | 50.4 | 0.02 |
| pHILIC | 389.231 | 82.9109 | 50.4 | 0.02 |
| pHILIC | 389.231 | 157.4687 | 50.3 | 0.02 |
| pHILIC | 389.231 | 89.1345 | 50.3 | 0.02 |
| pHILIC | 389.231 | 121.9312 | 50.3 | 0.02 |
| pHILIC | 389.231 | 143.6066 | 50.3 | 0.02 |
| pHILIC | 389.231 | 97.029 | 50.3 | 0.02 |
| pHILIC | 389.231 | 258.2167 | 50.3 | 0.02 |
| pHILIC | 389.231 | 311.495 | 50.3 | 0.02 |
| pHILIC | 389.231 | 227.7894 | 50.3 | 0.02 |
| pHILIC | 389.231 | 288.6942 | 50.3 | 0.02 |
| pHILIC | 389.231 | 409.5761 | 50.3 | 0.02 |
| pHILIC | 389.231 | 123.1028 | 50.3 | 0.02 |
| pHILIC | 389.231 | 376.6011 | 50.3 | 0.02 |
| pHILIC | 389.231 | 202.7503 | 50.2 | 0.02 |
| pHILIC | 389.231 | 93.3463 | 50.2 | 0.02 |
| pHILIC | 389.231 | 91.0358 | 50.2 | 0.02 |
| pHILIC | 389.231 | 66.3818 | 50.2 | 0.02 |
| pHILIC | 389.231 | 122.4943 | 50.1 | 0.02 |
| pHILIC | 389.231 | 253.3262 | 50.1 | 0.02 |
| pHILIC | 389.231 | 216.2074 | 50.1 | 0.02 |
| pHILIC | 389.231 | 69.3814 | 50.1 | 0.02 |
| pHILIC | 389.231 | 306.7661 | 50 | 0.02 |
| pHILIC | 389.231 | 56.2976 | 50 | 0.02 |
| pHILIC | 389.231 | 191.95 | 49.9 | 0.02 |
| pHILIC | 389.231 | 140.3623 | 49.9 | 0.02 |
| pHILIC | 389.231 | 98.1637 | 49.9 | 0.02 |
| pHILIC | 389.231 | 127.8441 | 49.9 | 0.02 |
| pHILIC | 389.231 | 381.9552 | 49.9 | 0.02 |
| pHILIC | 389.231 | 57.6441 | 49.9 | 0.02 |
| pHILIC | 389.231 | 196.8197 | 49.9 | 0.02 |
| pHILIC | 389.231 | 203.0912 | 49.9 | 0.02 |
| pHILIC | 389.231 | 132.8444 | 49.9 | 0.02 |
| pHILIC | 389.231 | 94.4701 | 49.8 | 0.02 |
| pHILIC | 389.231 | 65.9522 | 49.8 | 0.02 |
| pHILIC | 389.231 | 125.5288 | 49.8 | 0.02 |
| pHILIC | 389.231 | 65.6448 | 49.8 | 0.02 |
| pHILIC | 389.231 | 180.4682 | 49.8 | 0.02 |
| pHILIC | 389.231 | 62.571 | 49.8 | 0.02 |

| | | | | |
|---|---|---|---|---|
| pHILIC | 389.231 | 139.0407 | 49.8 | 0.02 |
| pHILIC | 389.231 | 173.9799 | 49.8 | 0.02 |
| pHILIC | 389.231 | 74.2158 | 49.8 | 0.02 |
| pHILIC | 389.231 | 193.5799 | 49.7 | 0.02 |
| pHILIC | 389.231 | 102.3113 | 49.7 | 0.02 |
| pHILIC | 389.231 | 223.126 | 49.7 | 0.02 |
| pHILIC | 389.231 | 51.2614 | 49.7 | 0.02 |
| pHILIC | 389.231 | 417.4707 | 49.7 | 0.02 |
| pHILIC | 389.231 | 65.7531 | 49.7 | 0.02 |
| pHILIC | 389.231 | 384.0508 | 49.7 | 0.02 |
| pHILIC | 389.231 | 68.0188 | 49.7 | 0.02 |
| pHILIC | 389.231 | 50.6334 | 49.7 | 0.02 |
| pHILIC | 389.231 | 52.0224 | 49.7 | 0.02 |
| pHILIC | 389.231 | 53.9406 | 49.7 | 0.02 |
| pHILIC | 389.231 | 52.7797 | 49.6 | 0.02 |
| pHILIC | 389.231 | 82.0364 | 49.6 | 0.02 |
| pHILIC | 389.231 | 131.0593 | 49.6 | 0.02 |
| pHILIC | 389.231 | 211.2312 | 49.6 | 0.02 |
| pHILIC | 389.231 | 356.072 | 49.6 | 0.02 |
| pHILIC | 389.231 | 96.0329 | 49.6 | 0.02 |
| pHILIC | 389.231 | 123.9726 | 49.5 | 0.02 |
| pHILIC | 389.231 | 60.6355 | 49.5 | 0.02 |
| pHILIC | 389.231 | 71.5757 | 49.5 | 0.02 |
| pHILIC | 389.231 | 51.5059 | 49.5 | 0.02 |
| pHILIC | 389.231 | 76.8528 | 49.5 | 0.02 |
| pHILIC | 389.231 | 102.5312 | 49.4 | 0.02 |
| pHILIC | 389.231 | 172.1348 | 49.4 | 0.02 |
| pHILIC | 389.231 | 375.7841 | 49.4 | 0.02 |
| pHILIC | 389.231 | 134.9193 | 49.4 | 0.02 |
| pHILIC | 389.231 | 179.0383 | 49.4 | 0.02 |
| pHILIC | 389.231 | 241.3722 | 49.4 | 0.02 |
| pHILIC | 389.231 | 57.6603 | 49.3 | 0.02 |
| pHILIC | 389.231 | 56.8086 | 49.3 | 0.02 |
| pHILIC | 389.231 | 106.0166 | 49.3 | 0.02 |
| pHILIC | 389.231 | 66.7946 | 49.3 | 0.02 |
| pHILIC | 389.231 | 91.0598 | 49.3 | 0.02 |
| pHILIC | 389.231 | 139.2761 | 49.3 | 0.02 |
| pHILIC | 389.231 | 84.7174 | 49.3 | 0.02 |
| pHILIC | 389.231 | 189.491 | 49.3 | 0.02 |
| pHILIC | 389.231 | 98.4782 | 49.3 | 0.02 |
| pHILIC | 389.231 | 179.0251 | 49.3 | 0.02 |
| pHILIC | 389.231 | 350.5074 | 49.2 | 0.02 |

| | | | | |
|---|---|---|---|---|
| pHILIC | 389.231 | 376.4689 | 49.2 | 0.02 |
| pHILIC | 389.231 | 112.6458 | 49.2 | 0.02 |
| pHILIC | 389.231 | 204.1298 | 49.2 | 0.02 |
| pHILIC | 389.231 | 156.4546 | 49.2 | 0.02 |
| pHILIC | 389.231 | 58.9986 | 49.2 | 0.02 |
| pHILIC | 389.231 | 87.1826 | 49.1 | 0.02 |
| pHILIC | 389.231 | 51.158 | 49.1 | 0.02 |
| pHILIC | 389.231 | 95.2872 | 49.1 | 0.02 |
| pHILIC | 389.231 | 346.2472 | 49.1 | 0.02 |
| pHILIC | 389.231 | 140.2758 | 49.1 | 0.02 |
| pHILIC | 389.231 | 101.533 | 49.1 | 0.02 |
| pHILIC | 389.231 | 144.522 | 49.1 | 0.02 |
| pHILIC | 389.231 | 62.4756 | 49.1 | 0.02 |
| pHILIC | 389.231 | 124.0704 | 49.1 | 0.02 |
| pHILIC | 389.231 | 94.9342 | 49 | 0.02 |
| pHILIC | 389.231 | 203.9865 | 49 | 0.02 |
| pHILIC | 389.231 | 275.4862 | 49 | 0.02 |
| pHILIC | 389.231 | 279.6617 | 49 | 0.02 |
| pHILIC | 389.231 | 61.7723 | 49 | 0.02 |
| pHILIC | 389.231 | 75.9055 | 48.9 | 0.02 |
| pHILIC | 389.231 | 155.0995 | 48.9 | 0.02 |
| pHILIC | 389.231 | 51.1487 | 48.9 | 0.02 |
| pHILIC | 389.231 | 146.2385 | 48.9 | 0.02 |
| pHILIC | 389.231 | 51.5176 | 48.9 | 0.02 |
| pHILIC | 389.231 | 114.2541 | 48.9 | 0.02 |
| pHILIC | 389.231 | 228.427 | 48.9 | 0.02 |
| pHILIC | 389.231 | 270.6328 | 48.9 | 0.02 |
| pHILIC | 389.231 | 138.134 | 48.9 | 0.02 |
| pHILIC | 389.231 | 81.2918 | 48.8 | 0.02 |
| pHILIC | 389.231 | 64.5864 | 48.8 | 0.02 |
| pHILIC | 389.231 | 51.3965 | 48.8 | 0.02 |
| pHILIC | 389.231 | 101.1232 | 48.8 | 0.02 |
| pHILIC | 389.231 | 78.6577 | 48.8 | 0.02 |
| pHILIC | 389.231 | 248.0609 | 48.8 | 0.02 |
| pHILIC | 389.231 | 107.3074 | 48.8 | 0.02 |
| pHILIC | 389.231 | 193.1391 | 48.8 | 0.02 |
| pHILIC | 389.231 | 232.5465 | 48.8 | 0.02 |
| pHILIC | 389.231 | 127.5015 | 48.8 | 0.02 |
| pHILIC | 389.231 | 55.6227 | 48.8 | 0.02 |
| pHILIC | 389.231 | 59.231 | 48.7 | 0.02 |
| pHILIC | 389.231 | 52.1287 | 48.7 | 0.02 |
| pHILIC | 389.231 | 154.334 | 48.7 | 0.02 |

| | | | | |
|---|---|---|---|---|
| pHILIC | 389.231 | 213.2497 | 48.7 | 0.02 |
| pHILIC | 389.231 | 55.7231 | 48.7 | 0.02 |
| pHILIC | 389.231 | 52.6127 | 48.7 | 0.02 |
| pHILIC | 389.231 | 116.543 | 48.7 | 0.02 |
| pHILIC | 389.231 | 194.0432 | 48.7 | 0.02 |
| pHILIC | 389.231 | 121.2808 | 48.6 | 0.02 |
| pHILIC | 389.231 | 220.0598 | 48.6 | 0.02 |
| pHILIC | 389.231 | 55.0461 | 48.6 | 0.02 |
| pHILIC | 389.231 | 274.8615 | 48.6 | 0.02 |
| pHILIC | 389.231 | 171.9988 | 48.6 | 0.02 |
| pHILIC | 389.231 | 82.7614 | 48.6 | 0.02 |
| pHILIC | 389.231 | 74.6105 | 48.6 | 0.02 |
| pHILIC | 389.231 | 137.6362 | 48.6 | 0.02 |
| pHILIC | 389.231 | 150.4823 | 48.6 | 0.02 |
| pHILIC | 389.231 | 84.9909 | 48.6 | 0.02 |
| pHILIC | 389.231 | 122.2484 | 48.6 | 0.02 |
| pHILIC | 389.231 | 56.062 | 48.6 | 0.02 |
| pHILIC | 389.231 | 377.6883 | 48.6 | 0.02 |
| pHILIC | 389.231 | 310.6305 | 48.6 | 0.02 |
| pHILIC | 389.231 | 127.788 | 48.5 | 0.02 |
| pHILIC | 389.231 | 50.5682 | 48.5 | 0.02 |
| pHILIC | 389.231 | 363.4186 | 48.5 | 0.02 |
| pHILIC | 389.231 | 96.2867 | 48.5 | 0.02 |
| pHILIC | 389.231 | 407.3572 | 48.5 | 0.02 |
| pHILIC | 389.231 | 186.5303 | 48.5 | 0.02 |
| pHILIC | 389.231 | 94.8512 | 48.5 | 0.02 |
| pHILIC | 389.231 | 95.7608 | 48.5 | 0.02 |
| pHILIC | 389.231 | 177.7318 | 48.5 | 0.02 |
| pHILIC | 389.231 | 67.3829 | 48.5 | 0.02 |
| pHILIC | 389.231 | 118.8857 | 48.5 | 0.02 |
| pHILIC | 389.231 | 94.3058 | 48.4 | 0.02 |
| pHILIC | 389.231 | 68.8186 | 48.4 | 0.02 |
| pHILIC | 389.231 | 354.1653 | 48.4 | 0.02 |
| pHILIC | 389.231 | 53.8821 | 48.4 | 0.02 |
| pHILIC | 389.231 | 102.2582 | 48.4 | 0.02 |
| pHILIC | 389.231 | 410.018 | 48.4 | 0.02 |
| pHILIC | 389.231 | 54.8363 | 48.4 | 0.02 |
| pHILIC | 389.231 | 84.6108 | 48.3 | 0.02 |
| pHILIC | 389.231 | 51.5329 | 48.3 | 0.02 |
| pHILIC | 389.231 | 94.0521 | 48.3 | 0.02 |
| pHILIC | 389.231 | 398.839 | 48.3 | 0.02 |
| pHILIC | 389.231 | 131.7817 | 48.3 | 0.02 |

| | | | | |
|---|---|---|---|---|
| pHILIC | 389.231 | 93.6756 | 48.3 | 0.02 |
| pHILIC | 389.231 | 145.0131 | 48.3 | 0.02 |
| pHILIC | 389.231 | 103.2917 | 48.3 | 0.02 |
| pHILIC | 389.231 | 92.5266 | 48.2 | 0.02 |
| pHILIC | 389.231 | 83.7113 | 48.2 | 0.02 |
| pHILIC | 389.231 | 56.0116 | 48.2 | 0.02 |
| pHILIC | 389.231 | 152.548 | 48.2 | 0.02 |
| pHILIC | 389.231 | 62.8187 | 48.2 | 0.02 |
| pHILIC | 389.231 | 173.4398 | 48.2 | 0.02 |
| pHILIC | 389.231 | 192.7769 | 48.2 | 0.02 |
| pHILIC | 389.231 | 128.9578 | 48.2 | 0.02 |
| pHILIC | 389.231 | 73.1965 | 48.2 | 0.02 |
| pHILIC | 389.231 | 85.4959 | 48.1 | 0.02 |
| pHILIC | 389.231 | 66.5355 | 48.1 | 0.02 |
| pHILIC | 389.231 | 68.5816 | 48.1 | 0.02 |
| pHILIC | 389.231 | 89.5691 | 48.1 | 0.02 |
| pHILIC | 389.231 | 288.903 | 48.1 | 0.02 |
| pHILIC | 389.231 | 164.2928 | 48.1 | 0.02 |
| pHILIC | 389.231 | 288.2989 | 48.1 | 0.02 |
| pHILIC | 389.231 | 195.6758 | 48.1 | 0.02 |
| pHILIC | 389.231 | 272.7029 | 48.1 | 0.02 |
| pHILIC | 389.231 | 167.5306 | 48 | 0.02 |
| pHILIC | 389.231 | 50.5662 | 48 | 0.02 |
| pHILIC | 389.231 | 185.7173 | 48 | 0.02 |
| pHILIC | 389.231 | 175.3014 | 48 | 0.02 |
| pHILIC | 389.231 | 111.9358 | 48 | 0.02 |
| pHILIC | 389.231 | 99.5032 | 48 | 0.02 |
| pHILIC | 389.231 | 123.2102 | 48 | 0.02 |
| pHILIC | 389.231 | 277.6436 | 48 | 0.02 |
| pHILIC | 389.231 | 162.1184 | 48 | 0.02 |
| pHILIC | 389.231 | 125.5494 | 48 | 0.02 |
| pHILIC | 389.231 | 333.5498 | 48 | 0.02 |
| pHILIC | 389.231 | 117.3813 | 48 | 0.02 |
| pHILIC | 389.231 | 275.9321 | 48 | 0.02 |
| pHILIC | 389.231 | 61.1387 | 47.9 | 0.02 |
| pHILIC | 389.231 | 299.2705 | 47.9 | 0.02 |
| pHILIC | 389.231 | 52.6229 | 47.9 | 0.02 |
| pHILIC | 389.231 | 75.6185 | 47.9 | 0.02 |
| pHILIC | 389.231 | 322.9268 | 47.9 | 0.02 |
| pHILIC | 389.231 | 62.4201 | 47.9 | 0.02 |
| pHILIC | 389.231 | 129.7284 | 47.8 | 0.02 |
| pHILIC | 389.231 | 363.9432 | 47.8 | 0.02 |

| | | | | |
|---|---|---|---|---|
| pHILIC | 389.231 | 83.1333 | 47.8 | 0.02 |
| pHILIC | 389.231 | 75.8466 | 47.8 | 0.02 |
| pHILIC | 389.231 | 69.4532 | 47.8 | 0.02 |
| pHILIC | 389.231 | 120.5817 | 47.8 | 0.02 |
| pHILIC | 389.231 | 123.5308 | 47.8 | 0.02 |
| pHILIC | 389.231 | 97.2892 | 47.8 | 0.02 |
| pHILIC | 389.231 | 206.5892 | 47.8 | 0.02 |
| pHILIC | 389.231 | 80.9828 | 47.8 | 0.02 |
| pHILIC | 389.231 | 90.9878 | 47.8 | 0.02 |
| pHILIC | 389.231 | 143.4492 | 47.8 | 0.02 |
| pHILIC | 389.231 | 290.4736 | 47.8 | 0.02 |
| pHILIC | 389.231 | 89.0264 | 47.7 | 0.02 |
| pHILIC | 389.231 | 93.8372 | 47.7 | 0.02 |
| pHILIC | 389.231 | 309.1847 | 47.7 | 0.02 |
| pHILIC | 389.231 | 120.23 | 47.7 | 0.02 |
| pHILIC | 389.231 | 249.3603 | 47.7 | 0.02 |
| pHILIC | 389.231 | 193.6539 | 47.7 | 0.02 |
| pHILIC | 389.231 | 261.399 | 47.7 | 0.02 |
| pHILIC | 389.231 | 54.9559 | 47.6 | 0.02 |
| pHILIC | 389.231 | 85.2182 | 47.6 | 0.02 |
| pHILIC | 389.231 | 402.2191 | 47.6 | 0.02 |
| pHILIC | 389.231 | 69.7743 | 47.6 | 0.02 |
| pHILIC | 389.231 | 121.9112 | 47.6 | 0.02 |
| pHILIC | 389.231 | 244.7948 | 47.6 | 0.02 |
| pHILIC | 389.231 | 129.0543 | 47.6 | 0.02 |
| pHILIC | 389.231 | 245.5777 | 47.6 | 0.02 |
| pHILIC | 389.231 | 52.145 | 47.6 | 0.02 |
| pHILIC | 389.231 | 62.5127 | 47.6 | 0.02 |
| pHILIC | 389.231 | 58.7951 | 47.5 | 0.02 |
| pHILIC | 389.231 | 71.0491 | 47.5 | 0.02 |
| pHILIC | 389.231 | 196.2803 | 47.5 | 0.02 |
| pHILIC | 389.231 | 148.5948 | 47.5 | 0.02 |
| pHILIC | 389.231 | 135.6503 | 47.5 | 0.02 |
| pHILIC | 389.231 | 62.8947 | 47.5 | 0.02 |
| pHILIC | 389.231 | 105.6065 | 47.5 | 0.02 |
| pHILIC | 389.231 | 164.1525 | 47.4 | 0.02 |
| pHILIC | 389.231 | 179.3888 | 47.4 | 0.02 |
| pHILIC | 389.231 | 88.4088 | 47.4 | 0.02 |
| pHILIC | 389.231 | 83.982 | 47.4 | 0.02 |
| pHILIC | 389.231 | 64.1901 | 47.4 | 0.02 |
| pHILIC | 389.231 | 282.0664 | 47.4 | 0.02 |
| pHILIC | 389.231 | 54.7045 | 47.4 | 0.02 |

| | | | | |
|---|---|---|---|---|
| pHILIC | 389.231 | 207.6673 | 47.4 | 0.02 |
| pHILIC | 389.231 | 50.8671 | 47.4 | 0.02 |
| pHILIC | 389.231 | 129.0366 | 47.4 | 0.02 |
| pHILIC | 389.231 | 61.9788 | 47.4 | 0.02 |
| pHILIC | 389.231 | 167.2085 | 47.4 | 0.02 |
| pHILIC | 389.231 | 113.1335 | 47.4 | 0.02 |
| pHILIC | 389.231 | 250.8297 | 47.3 | 0.02 |
| pHILIC | 389.231 | 132.0734 | 47.3 | 0.02 |
| pHILIC | 389.231 | 111.5832 | 47.3 | 0.02 |
| pHILIC | 389.231 | 300.4272 | 47.3 | 0.02 |
| pHILIC | 389.231 | 78.9623 | 47.3 | 0.02 |
| pHILIC | 389.231 | 397.9044 | 47.3 | 0.02 |
| pHILIC | 389.231 | 67.3476 | 47.3 | 0.02 |
| pHILIC | 389.231 | 115.9247 | 47.3 | 0.02 |
| pHILIC | 389.231 | 223.1028 | 47.3 | 0.02 |
| pHILIC | 389.231 | 81.6126 | 47.3 | 0.02 |
| pHILIC | 389.231 | 80.0948 | 47.3 | 0.02 |
| pHILIC | 389.231 | 337.7788 | 47.3 | 0.02 |
| pHILIC | 389.231 | 171.3023 | 47.3 | 0.02 |
| pHILIC | 389.231 | 131.4626 | 47.2 | 0.02 |
| pHILIC | 389.231 | 62.3178 | 47.2 | 0.02 |
| pHILIC | 389.231 | 55.1447 | 47.2 | 0.02 |
| pHILIC | 389.231 | 178.5335 | 47.2 | 0.02 |
| pHILIC | 389.231 | 116.7783 | 47.2 | 0.02 |
| pHILIC | 389.231 | 212.5657 | 47.2 | 0.02 |
| pHILIC | 389.231 | 88.7218 | 47.2 | 0.02 |
| pHILIC | 389.231 | 76.5254 | 47.2 | 0.02 |
| pHILIC | 389.231 | 57.329 | 47.2 | 0.02 |
| pHILIC | 389.231 | 98.5621 | 47.2 | 0.02 |
| pHILIC | 389.231 | 65.6547 | 47.2 | 0.02 |
| pHILIC | 389.231 | 58.4529 | 47.2 | 0.02 |
| pHILIC | 389.231 | 74.979 | 47.2 | 0.02 |
| pHILIC | 389.231 | 141.8233 | 47.2 | 0.02 |
| pHILIC | 389.231 | 51.3022 | 47.1 | 0.02 |
| pHILIC | 389.231 | 53.5035 | 47.1 | 0.02 |
| pHILIC | 389.231 | 143.8993 | 47.1 | 0.02 |
| pHILIC | 389.231 | 66.5634 | 47.1 | 0.02 |
| pHILIC | 389.231 | 176.9024 | 47.1 | 0.02 |
| pHILIC | 389.231 | 270.829 | 47.1 | 0.02 |
| pHILIC | 389.231 | 224.6279 | 47.1 | 0.02 |
| pHILIC | 389.231 | 56.9934 | 47.1 | 0.02 |
| pHILIC | 389.231 | 277.4727 | 47.1 | 0.02 |

| | | | | |
|---|---|---|---|---|
| pHILIC | 389.231 | 112.5337 | 47.1 | 0.02 |
| pHILIC | 389.231 | 50.225 | 47.1 | 0.02 |
| pHILIC | 389.231 | 51.4717 | 47 | 0.02 |
| pHILIC | 389.231 | 136.8487 | 47 | 0.02 |
| pHILIC | 389.231 | 64.11 | 47 | 0.02 |
| pHILIC | 389.231 | 181.7419 | 47 | 0.02 |
| pHILIC | 389.231 | 68.0554 | 47 | 0.02 |
| pHILIC | 389.231 | 335.9765 | 47 | 0.02 |
| pHILIC | 389.231 | 94.9769 | 47 | 0.02 |
| pHILIC | 389.231 | 221.136 | 47 | 0.02 |
| pHILIC | 389.231 | 95.0575 | 47 | 0.02 |
| pHILIC | 389.231 | 62.734 | 47 | 0.02 |
| pHILIC | 389.231 | 77.5829 | 47 | 0.02 |
| pHILIC | 389.231 | 197.1935 | 47 | 0.02 |
| pHILIC | 389.231 | 72.3879 | 46.9 | 0.02 |
| pHILIC | 389.231 | 135.0627 | 46.9 | 0.02 |
| pHILIC | 389.231 | 91.149 | 46.9 | 0.02 |
| pHILIC | 389.231 | 75.7567 | 46.9 | 0.02 |
| pHILIC | 389.231 | 167.6276 | 46.9 | 0.02 |
| pHILIC | 389.231 | 55.8033 | 46.9 | 0.02 |
| pHILIC | 389.231 | 80.4638 | 46.9 | 0.02 |
| pHILIC | 389.231 | 52.8058 | 46.9 | 0.02 |
| pHILIC | 389.231 | 84.2048 | 46.9 | 0.02 |
| pHILIC | 389.231 | 157.0825 | 46.9 | 0.02 |
| pHILIC | 389.231 | 89.0095 | 46.9 | 0.02 |
| pHILIC | 389.231 | 61.2652 | 46.9 | 0.02 |
| pHILIC | 389.231 | 186.5025 | 46.9 | 0.02 |
| pHILIC | 389.231 | 66.9045 | 46.9 | 0.02 |
| pHILIC | 389.231 | 377.213 | 46.9 | 0.02 |
| pHILIC | 389.231 | 90.1653 | 46.9 | 0.02 |
| pHILIC | 389.231 | 108.298 | 46.9 | 0.02 |
| pHILIC | 389.231 | 185.2513 | 46.9 | 0.02 |
| pHILIC | 389.231 | 50.2939 | 46.9 | 0.02 |
| pHILIC | 389.231 | 66.0067 | 46.9 | 0.02 |
| pHILIC | 389.231 | 75.2698 | 46.8 | 0.02 |
| pHILIC | 389.231 | 116.4681 | 46.8 | 0.02 |
| pHILIC | 389.231 | 94.1544 | 46.8 | 0.02 |
| pHILIC | 389.231 | 95.0384 | 46.8 | 0.02 |
| pHILIC | 389.231 | 330.5989 | 46.8 | 0.02 |
| pHILIC | 389.231 | 95.5275 | 46.8 | 0.02 |
| pHILIC | 389.231 | 341.3076 | 46.7 | 0.02 |
| pHILIC | 389.231 | 87.1042 | 46.7 | 0.02 |

| | | | | |
|---|---|---|---|---|
| pHILIC | 389.231 | 292.687 | 46.7 | 0.02 |
| pHILIC | 389.231 | 99.0888 | 46.7 | 0.02 |
| pHILIC | 389.231 | 75.6211 | 46.7 | 0.02 |
| pHILIC | 389.231 | 298.2398 | 46.7 | 0.02 |
| pHILIC | 389.231 | 368.1151 | 46.7 | 0.02 |
| pHILIC | 389.231 | 91.2953 | 46.7 | 0.02 |
| pHILIC | 389.231 | 63.0316 | 46.7 | 0.02 |
| pHILIC | 389.231 | 88.882 | 46.7 | 0.02 |
| pHILIC | 389.231 | 247.2374 | 46.7 | 0.02 |
| pHILIC | 389.231 | 60.7281 | 46.7 | 0.02 |
| pHILIC | 389.231 | 73.8226 | 46.7 | 0.02 |
| pHILIC | 389.231 | 70.1348 | 46.7 | 0.02 |
| pHILIC | 389.231 | 142.506 | 46.6 | 0.02 |
| pHILIC | 389.231 | 363.2856 | 46.6 | 0.02 |
| pHILIC | 389.231 | 197.6651 | 46.6 | 0.02 |
| pHILIC | 389.231 | 51.6493 | 46.6 | 0.02 |
| pHILIC | 389.231 | 182.7455 | 46.6 | 0.02 |
| pHILIC | 389.231 | 56.8864 | 46.6 | 0.02 |
| pHILIC | 389.231 | 65.5058 | 46.6 | 0.02 |
| pHILIC | 389.231 | 96.3994 | 46.6 | 0.02 |
| pHILIC | 389.231 | 120.7971 | 46.6 | 0.02 |
| pHILIC | 389.231 | 66.8986 | 46.5 | 0.02 |
| pHILIC | 389.231 | 276.1769 | 46.5 | 0.02 |
| pHILIC | 389.231 | 114.0659 | 46.5 | 0.02 |
| pHILIC | 389.231 | 202.915 | 46.5 | 0.02 |
| pHILIC | 389.231 | 87.0434 | 46.5 | 0.02 |
| pHILIC | 389.231 | 61.758 | 46.5 | 0.02 |
| pHILIC | 389.231 | 232.7303 | 46.5 | 0.02 |
| pHILIC | 389.231 | 348.563 | 46.4 | 0.02 |
| pHILIC | 389.231 | 83.273 | 46.4 | 0.02 |
| pHILIC | 389.231 | 173.4273 | 46.4 | 0.02 |
| pHILIC | 389.231 | 136.8736 | 46.4 | 0.02 |
| pHILIC | 389.231 | 413.0448 | 46.4 | 0.02 |
| pHILIC | 389.231 | 51.6448 | 46.4 | 0.02 |
| pHILIC | 389.231 | 77.6938 | 46.4 | 0.02 |
| pHILIC | 389.231 | 88.9113 | 46.4 | 0.02 |
| pHILIC | 389.231 | 281.2605 | 46.4 | 0.02 |
| pHILIC | 389.231 | 180.6012 | 46.4 | 0.02 |
| pHILIC | 389.231 | 116.8276 | 46.3 | 0.02 |
| pHILIC | 389.231 | 184.576 | 46.3 | 0.02 |
| pHILIC | 389.231 | 275.3445 | 46.3 | 0.02 |
| pHILIC | 389.231 | 206.3265 | 46.3 | 0.02 |

| | | | | |
|---|---|---|---|---|
| pHILIC | 389.231 | 65.2361 | 46.3 | 0.02 |
| pHILIC | 389.231 | 60.3722 | 46.3 | 0.02 |
| pHILIC | 389.231 | 295.317 | 46.3 | 0.02 |
| pHILIC | 389.231 | 212.6218 | 46.3 | 0.02 |
| pHILIC | 389.231 | 105.1367 | 46.3 | 0.02 |
| pHILIC | 389.231 | 63.0233 | 46.3 | 0.02 |
| pHILIC | 389.231 | 352.1094 | 46.3 | 0.02 |
| pHILIC | 389.231 | 383.3361 | 46.3 | 0.02 |
| pHILIC | 389.231 | 100.9907 | 46.3 | 0.02 |
| pHILIC | 389.231 | 62.5402 | 46.3 | 0.02 |
| pHILIC | 389.231 | 111.4686 | 46.3 | 0.02 |
| pHILIC | 389.231 | 143.8748 | 46.3 | 0.02 |
| pHILIC | 389.231 | 157.7144 | 46.3 | 0.02 |
| pHILIC | 389.231 | 124.7161 | 46.3 | 0.02 |
| pHILIC | 389.231 | 74.6299 | 46.2 | 0.02 |
| pHILIC | 389.231 | 363.067 | 46.2 | 0.02 |
| pHILIC | 389.231 | 82.3279 | 46.2 | 0.02 |
| pHILIC | 389.231 | 61.0184 | 46.2 | 0.02 |
| pHILIC | 389.231 | 77.2666 | 46.2 | 0.02 |
| pHILIC | 389.231 | 78.3608 | 46.2 | 0.02 |
| pHILIC | 389.231 | 94.1956 | 46.2 | 0.02 |
| pHILIC | 389.231 | 116.5178 | 46.1 | 0.02 |
| pHILIC | 389.231 | 93.6223 | 46.1 | 0.02 |
| pHILIC | 389.231 | 54.5836 | 46.1 | 0.02 |
| pHILIC | 389.231 | 335.238 | 46.1 | 0.02 |
| pHILIC | 389.231 | 81.9607 | 46.1 | 0.02 |
| pHILIC | 389.231 | 234.7365 | 46.1 | 0.02 |
| pHILIC | 389.231 | 60.9961 | 46.1 | 0.02 |
| pHILIC | 389.231 | 56.5069 | 46.1 | 0.02 |
| pHILIC | 389.231 | 166.0367 | 46.1 | 0.02 |
| pHILIC | 389.231 | 101.0681 | 46 | 0.02 |
| pHILIC | 389.231 | 348.3041 | 46 | 0.02 |
| pHILIC | 389.231 | 204.5847 | 46 | 0.02 |
| pHILIC | 389.231 | 241.7633 | 46 | 0.02 |
| pHILIC | 389.231 | 129.781 | 46 | 0.02 |
| pHILIC | 389.231 | 50.8519 | 46 | 0.02 |
| pHILIC | 389.231 | 398.0258 | 46 | 0.02 |
| pHILIC | 389.231 | 202.5719 | 46 | 0.02 |
| pHILIC | 389.231 | 79.0809 | 46 | 0.02 |
| pHILIC | 389.231 | 106.1525 | 46 | 0.02 |
| pHILIC | 389.231 | 52.8688 | 46 | 0.02 |
| pHILIC | 389.231 | 148.6743 | 46 | 0.02 |

| | | | | |
|---|---|---|---|---|
| pHILIC | 389.231 | 389.6931 | 46 | 0.02 |
| pHILIC | 389.231 | 76.341 | 46 | 0.02 |
| pHILIC | 389.231 | 210.5513 | 45.9 | 0.02 |
| pHILIC | 389.231 | 310.4305 | 45.9 | 0.02 |
| pHILIC | 389.231 | 92.2231 | 45.9 | 0.02 |
| pHILIC | 389.231 | 88.4285 | 45.9 | 0.02 |
| pHILIC | 389.231 | 59.3163 | 45.9 | 0.02 |
| pHILIC | 389.231 | 236.7138 | 45.9 | 0.02 |
| pHILIC | 389.231 | 209.6582 | 45.9 | 0.02 |
| pHILIC | 389.231 | 110.1156 | 45.9 | 0.02 |
| pHILIC | 389.231 | 132.081 | 45.9 | 0.02 |
| pHILIC | 389.231 | 79.0299 | 45.9 | 0.02 |
| pHILIC | 389.231 | 52.1476 | 45.9 | 0.02 |
| pHILIC | 389.231 | 72.7659 | 45.9 | 0.02 |
| pHILIC | 389.231 | 76.1036 | 45.9 | 0.02 |
| pHILIC | 389.231 | 175.0746 | 45.9 | 0.02 |
| pHILIC | 389.231 | 228.1342 | 45.9 | 0.02 |
| pHILIC | 389.231 | 109.1007 | 45.9 | 0.02 |
| pHILIC | 389.231 | 287.8819 | 45.8 | 0.02 |
| pHILIC | 389.231 | 76.6469 | 45.8 | 0.02 |
| pHILIC | 389.231 | 83.3562 | 45.8 | 0.02 |
| pHILIC | 389.231 | 108.4458 | 45.8 | 0.02 |
| pHILIC | 389.231 | 410.6479 | 45.8 | 0.02 |
| pHILIC | 389.231 | 413.4276 | 45.8 | 0.02 |
| pHILIC | 389.231 | 166.4868 | 45.8 | 0.02 |
| pHILIC | 389.231 | 199.259 | 45.8 | 0.02 |
| pHILIC | 389.231 | 155.5807 | 45.8 | 0.02 |
| pHILIC | 389.231 | 260.2224 | 45.8 | 0.02 |
| pHILIC | 389.231 | 102.9756 | 45.8 | 0.02 |
| pHILIC | 389.231 | 162.6183 | 45.8 | 0.02 |
| pHILIC | 389.231 | 111.9707 | 45.8 | 0.02 |
| pHILIC | 389.231 | 70.4827 | 45.7 | 0.02 |
| pHILIC | 389.231 | 273.6511 | 45.7 | 0.02 |
| pHILIC | 389.231 | 124.9129 | 45.7 | 0.02 |
| pHILIC | 389.231 | 183.0576 | 45.7 | 0.02 |
| pHILIC | 389.231 | 250.2418 | 45.7 | 0.02 |
| pHILIC | 389.231 | 243.2137 | 45.7 | 0.02 |
| pHILIC | 389.231 | 336.7922 | 45.7 | 0.02 |
| pHILIC | 389.231 | 58.0882 | 45.7 | 0.02 |
| pHILIC | 389.231 | 389.3467 | 45.7 | 0.02 |
| pHILIC | 389.231 | 72.2229 | 45.7 | 0.02 |
| pHILIC | 389.231 | 80.9431 | 45.6 | 0.02 |

| | | | | |
|---|---|---|---|---|
| pHILIC | 389.231 | 54.4818 | 45.6 | 0.02 |
| pHILIC | 389.231 | 113.9618 | 45.6 | 0.02 |
| pHILIC | 389.231 | 95.1111 | 45.6 | 0.02 |
| pHILIC | 389.231 | 160.7867 | 45.6 | 0.02 |
| pHILIC | 389.231 | 77.3563 | 45.6 | 0.02 |
| pHILIC | 389.231 | 109.5862 | 45.6 | 0.02 |
| pHILIC | 389.231 | 274.9894 | 45.6 | 0.02 |
| pHILIC | 389.231 | 52.2364 | 45.6 | 0.02 |
| pHILIC | 389.231 | 305.3565 | 45.6 | 0.02 |
| pHILIC | 389.231 | 161.5888 | 45.6 | 0.02 |
| pHILIC | 389.231 | 87.1099 | 45.5 | 0.02 |
| pHILIC | 389.231 | 60.5787 | 45.5 | 0.02 |
| pHILIC | 389.231 | 129.5662 | 45.5 | 0.02 |
| pHILIC | 389.231 | 65.7718 | 45.5 | 0.02 |
| pHILIC | 389.231 | 77.2409 | 45.5 | 0.02 |
| pHILIC | 389.231 | 78.957 | 45.5 | 0.02 |
| pHILIC | 389.231 | 208.3677 | 45.5 | 0.02 |
| pHILIC | 389.231 | 120.8102 | 45.5 | 0.02 |
| pHILIC | 389.231 | 59.1444 | 45.5 | 0.02 |
| pHILIC | 389.231 | 70.3164 | 45.5 | 0.02 |
| pHILIC | 389.231 | 88.4487 | 45.5 | 0.02 |
| pHILIC | 389.231 | 82.3212 | 45.5 | 0.02 |
| pHILIC | 389.231 | 117.5648 | 45.5 | 0.02 |
| pHILIC | 389.231 | 134.0728 | 45.5 | 0.02 |
| pHILIC | 389.231 | 279.7772 | 45.5 | 0.02 |
| pHILIC | 389.231 | 173.7036 | 45.4 | 0.02 |
| pHILIC | 389.231 | 101.0613 | 45.4 | 0.02 |
| pHILIC | 389.231 | 88.6176 | 45.4 | 0.02 |
| pHILIC | 389.231 | 86.8596 | 45.4 | 0.02 |
| pHILIC | 389.231 | 347.2 | 45.4 | 0.02 |
| pHILIC | 389.231 | 212.0626 | 45.4 | 0.02 |
| pHILIC | 389.231 | 66.4808 | 45.4 | 0.02 |
| pHILIC | 389.231 | 211.4033 | 45.4 | 0.02 |
| pHILIC | 389.231 | 60.529 | 45.4 | 0.02 |
| pHILIC | 389.231 | 391.6247 | 45.4 | 0.02 |
| pHILIC | 389.231 | 163.1511 | 45.4 | 0.02 |
| pHILIC | 389.231 | 104.7255 | 45.4 | 0.02 |
| pHILIC | 389.231 | 138.4805 | 45.4 | 0.02 |
| pHILIC | 389.231 | 148.3774 | 45.3 | 0.02 |
| pHILIC | 389.231 | 67.5667 | 45.3 | 0.02 |
| pHILIC | 389.231 | 378.9264 | 45.3 | 0.02 |
| pHILIC | 389.231 | 68.6056 | 45.3 | 0.02 |

| | | | | |
|---|---|---|---|---|
| pHILIC | 389.231 | 179.0013 | 45.3 | 0.02 |
| pHILIC | 389.231 | 134.153 | 45.3 | 0.02 |
| pHILIC | 389.231 | 81.057 | 45.3 | 0.02 |
| pHILIC | 389.231 | 142.1973 | 45.3 | 0.02 |
| pHILIC | 389.231 | 350.1469 | 45.3 | 0.02 |
| pHILIC | 389.231 | 95.3019 | 45.3 | 0.02 |
| pHILIC | 389.231 | 198.5288 | 45.3 | 0.02 |
| pHILIC | 389.231 | 50.9097 | 45.3 | 0.02 |
| pHILIC | 389.231 | 71.1274 | 45.3 | 0.02 |
| pHILIC | 389.231 | 223.8885 | 45.3 | 0.02 |
| pHILIC | 389.231 | 66.9298 | 45.3 | 0.02 |
| pHILIC | 389.231 | 59.4977 | 45.3 | 0.02 |
| pHILIC | 389.231 | 91.8469 | 45.3 | 0.02 |
| pHILIC | 389.231 | 205.1468 | 45.3 | 0.02 |
| pHILIC | 389.231 | 50.0631 | 45.3 | 0.02 |
| pHILIC | 389.231 | 123.6883 | 45.2 | 0.02 |
| pHILIC | 389.231 | 397.037 | 45.2 | 0.02 |
| pHILIC | 389.231 | 64.7794 | 45.2 | 0.02 |
| pHILIC | 389.231 | 65.6101 | 45.2 | 0.02 |
| pHILIC | 389.231 | 78.768 | 45.2 | 0.02 |
| pHILIC | 389.231 | 62.6925 | 45.2 | 0.02 |
| pHILIC | 389.231 | 230.3007 | 45.2 | 0.02 |
| pHILIC | 389.231 | 189.4224 | 45.2 | 0.02 |
| pHILIC | 389.231 | 331.627 | 45.2 | 0.02 |
| pHILIC | 389.231 | 93.295 | 45.2 | 0.02 |
| pHILIC | 389.231 | 55.4937 | 45.1 | 0.02 |
| pHILIC | 389.231 | 67.277 | 45.1 | 0.02 |
| pHILIC | 389.231 | 52.8301 | 45.1 | 0.02 |
| pHILIC | 389.231 | 297.0799 | 45.1 | 0.02 |
| pHILIC | 389.231 | 59.7521 | 45.1 | 0.02 |
| pHILIC | 389.231 | 124.4707 | 45.1 | 0.02 |
| pHILIC | 389.231 | 52.2513 | 45.1 | 0.02 |
| pHILIC | 389.231 | 416.4712 | 45.1 | 0.02 |
| pHILIC | 389.231 | 218.6595 | 45.1 | 0.02 |
| pHILIC | 389.231 | 314.6266 | 45 | 0.02 |
| pHILIC | 389.231 | 338.2271 | 45 | 0.02 |
| pHILIC | 389.231 | 55.0598 | 45 | 0.02 |
| pHILIC | 389.231 | 81.822 | 45 | 0.02 |
| pHILIC | 389.231 | 63.1642 | 45 | 0.02 |
| pHILIC | 389.231 | 100.5728 | 45 | 0.02 |
| pHILIC | 389.231 | 58.3515 | 45 | 0.02 |
| pHILIC | 389.231 | 50.1887 | 45 | 0.02 |

| | | | | |
|---|---|---|---|---|
| pHILIC | 389.231 | 138.9331 | 45 | 0.02 |
| pHILIC | 389.231 | 50.7522 | 45 | 0.02 |
| pHILIC | 389.231 | 133.1008 | 45 | 0.02 |
| pHILIC | 389.231 | 50.3249 | 45 | 0.02 |
| pHILIC | 389.231 | 156.762 | 45 | 0.02 |
| pHILIC | 389.231 | 325.73 | 45 | 0.02 |
| pHILIC | 389.231 | 108.5396 | 45 | 0.02 |
| pHILIC | 389.231 | 112.0501 | 44.9 | 0.02 |
| pHILIC | 389.231 | 194.5151 | 44.9 | 0.02 |
| pHILIC | 389.231 | 80.3298 | 44.9 | 0.02 |
| pHILIC | 389.231 | 169.8746 | 44.9 | 0.02 |
| pHILIC | 389.231 | 329.6866 | 44.9 | 0.02 |
| pHILIC | 389.231 | 85.2937 | 44.9 | 0.02 |
| pHILIC | 389.231 | 159.0832 | 44.9 | 0.02 |
| pHILIC | 389.231 | 110.1234 | 44.9 | 0.02 |
| pHILIC | 389.231 | 51.0186 | 44.9 | 0.02 |
| pHILIC | 389.231 | 57.8858 | 44.9 | 0.02 |
| pHILIC | 389.231 | 269.5297 | 44.9 | 0.02 |
| pHILIC | 389.231 | 125.3496 | 44.9 | 0.02 |
| pHILIC | 389.231 | 61.7368 | 44.9 | 0.02 |
| pHILIC | 389.231 | 208.485 | 44.9 | 0.02 |
| pHILIC | 389.231 | 71.7586 | 44.9 | 0.02 |
| pHILIC | 389.231 | 85.6043 | 44.9 | 0.02 |
| pHILIC | 389.231 | 160.9698 | 44.9 | 0.02 |
| pHILIC | 389.231 | 161.7765 | 44.9 | 0.02 |
| pHILIC | 389.231 | 177.8642 | 44.8 | 0.02 |
| pHILIC | 389.231 | 139.9673 | 44.8 | 0.02 |
| pHILIC | 389.231 | 165.0166 | 44.8 | 0.02 |
| pHILIC | 389.231 | 79.8176 | 44.8 | 0.02 |
| pHILIC | 389.231 | 213.3927 | 44.8 | 0.02 |
| pHILIC | 389.231 | 214.5707 | 44.8 | 0.02 |
| pHILIC | 389.231 | 51.1589 | 44.8 | 0.02 |
| pHILIC | 389.231 | 339.2508 | 44.8 | 0.02 |
| pHILIC | 389.231 | 77.371 | 44.8 | 0.02 |
| pHILIC | 389.231 | 379.0571 | 44.8 | 0.02 |
| pHILIC | 389.231 | 403.6909 | 44.8 | 0.02 |
| pHILIC | 389.231 | 126.1002 | 44.8 | 0.02 |
| pHILIC | 389.231 | 123.7568 | 44.8 | 0.02 |
| pHILIC | 389.231 | 93.592 | 44.7 | 0.02 |
| pHILIC | 389.231 | 51.9658 | 44.7 | 0.02 |
| pHILIC | 389.231 | 291.1471 | 44.7 | 0.02 |
| pHILIC | 389.231 | 104.8262 | 44.7 | 0.02 |

| | | | | |
|---|---|---|---|---|
| pHILIC | 389.231 | 199.6224 | 44.7 | 0.02 |
| pHILIC | 389.231 | 185.4142 | 44.7 | 0.02 |
| pHILIC | 389.231 | 134.3582 | 44.7 | 0.02 |
| pHILIC | 389.231 | 51.5 | 44.7 | 0.02 |
| pHILIC | 389.231 | 236.2726 | 44.7 | 0.02 |
| pHILIC | 389.231 | 110.2378 | 44.7 | 0.02 |
| pHILIC | 389.231 | 61.9712 | 44.7 | 0.02 |
| pHILIC | 389.231 | 138.9783 | 44.7 | 0.02 |
| pHILIC | 389.231 | 90.9228 | 44.7 | 0.02 |
| pHILIC | 389.231 | 233.3575 | 44.7 | 0.02 |
| pHILIC | 389.231 | 249.8075 | 44.6 | 0.02 |
| pHILIC | 389.231 | 54.3627 | 44.6 | 0.02 |
| pHILIC | 389.231 | 151.6736 | 44.6 | 0.02 |
| pHILIC | 389.231 | 68.8682 | 44.6 | 0.02 |
| pHILIC | 389.231 | 330.9521 | 44.6 | 0.02 |
| pHILIC | 389.231 | 73.9784 | 44.6 | 0.02 |
| pHILIC | 389.231 | 114.3574 | 44.6 | 0.02 |
| pHILIC | 389.231 | 207.022 | 44.6 | 0.02 |
| pHILIC | 389.231 | 394.0446 | 44.6 | 0.02 |
| pHILIC | 389.231 | 65.5018 | 44.6 | 0.02 |
| pHILIC | 389.231 | 63.0432 | 44.6 | 0.02 |
| pHILIC | 389.231 | 66.9077 | 44.6 | 0.02 |
| pHILIC | 389.231 | 313.5662 | 44.6 | 0.02 |
| pHILIC | 389.231 | 204.844 | 44.6 | 0.02 |
| pHILIC | 389.231 | 216.1111 | 44.5 | 0.02 |
| pHILIC | 389.231 | 152.0466 | 44.5 | 0.02 |
| pHILIC | 389.231 | 147.8036 | 44.5 | 0.02 |
| pHILIC | 389.231 | 93.7956 | 44.5 | 0.02 |
| pHILIC | 389.231 | 323.2851 | 44.5 | 0.02 |
| pHILIC | 389.231 | 92.369 | 44.5 | 0.02 |
| pHILIC | 389.231 | 101.8402 | 44.5 | 0.02 |
| pHILIC | 389.231 | 103.1293 | 44.5 | 0.02 |
| pHILIC | 389.231 | 51.4707 | 44.5 | 0.02 |
| pHILIC | 389.231 | 86.486 | 44.5 | 0.02 |
| pHILIC | 389.231 | 73.0501 | 44.5 | 0.02 |
| pHILIC | 389.231 | 159.2788 | 44.5 | 0.02 |
| pHILIC | 389.231 | 239.1488 | 44.5 | 0.02 |
| pHILIC | 389.231 | 132.7697 | 44.5 | 0.02 |
| pHILIC | 389.231 | 70.5995 | 44.5 | 0.02 |
| pHILIC | 389.231 | 213.9687 | 44.5 | 0.02 |
| pHILIC | 389.231 | 233.2466 | 44.5 | 0.02 |
| pHILIC | 389.231 | 78.5858 | 44.4 | 0.02 |

| | | | | |
|---|---|---|---|---|
| pHILIC | 389.231 | 52.4997 | 44.4 | 0.02 |
| pHILIC | 389.231 | 114.7382 | 44.4 | 0.02 |
| pHILIC | 389.231 | 367.3 | 44.4 | 0.02 |
| pHILIC | 389.231 | 84.2235 | 44.4 | 0.02 |
| pHILIC | 389.231 | 68.921 | 44.4 | 0.02 |
| pHILIC | 389.231 | 62.2013 | 44.4 | 0.02 |
| pHILIC | 389.231 | 61.6724 | 44.4 | 0.02 |
| pHILIC | 389.231 | 250.9574 | 44.3 | 0.02 |
| pHILIC | 389.231 | 358.5829 | 44.3 | 0.02 |
| pHILIC | 389.231 | 309.5697 | 44.3 | 0.02 |
| pHILIC | 389.231 | 94.3947 | 44.3 | 0.02 |
| pHILIC | 389.231 | 122.8802 | 44.3 | 0.02 |
| pHILIC | 389.231 | 83.9386 | 44.3 | 0.02 |
| pHILIC | 389.231 | 53.1068 | 44.3 | 0.02 |
| pHILIC | 389.231 | 67.0313 | 44.3 | 0.02 |
| pHILIC | 389.231 | 115.0694 | 44.3 | 0.02 |
| pHILIC | 389.231 | 60.2886 | 44.3 | 0.02 |
| pHILIC | 389.231 | 167.2785 | 44.3 | 0.02 |
| pHILIC | 389.231 | 142.171 | 44.3 | 0.02 |
| pHILIC | 389.231 | 123.0667 | 44.3 | 0.02 |
| pHILIC | 389.231 | 57.8808 | 44.3 | 0.02 |
| pHILIC | 389.231 | 66.0649 | 44.3 | 0.02 |
| pHILIC | 389.231 | 77.1693 | 44.3 | 0.02 |
| pHILIC | 389.231 | 181.9279 | 44.3 | 0.02 |
| pHILIC | 389.231 | 217.6333 | 44.2 | 0.02 |
| pHILIC | 389.231 | 88.5416 | 44.2 | 0.02 |
| pHILIC | 389.231 | 168.3714 | 44.2 | 0.02 |
| pHILIC | 389.231 | 86.8258 | 44.2 | 0.02 |
| pHILIC | 389.231 | 70.5588 | 44.2 | 0.02 |
| pHILIC | 389.231 | 111.0537 | 44.2 | 0.02 |
| pHILIC | 389.231 | 378.847 | 44.2 | 0.02 |
| pHILIC | 389.231 | 343.7646 | 44.2 | 0.02 |
| pHILIC | 389.231 | 68.2198 | 44.1 | 0.02 |
| pHILIC | 389.231 | 163.8241 | 44.1 | 0.02 |
| pHILIC | 389.231 | 74.9097 | 44.1 | 0.02 |
| pHILIC | 389.231 | 91.4416 | 44.1 | 0.02 |
| pHILIC | 389.231 | 62.0032 | 44.1 | 0.02 |
| pHILIC | 389.231 | 86.1277 | 44.1 | 0.02 |
| pHILIC | 389.231 | 191.3459 | 44.1 | 0.02 |
| pHILIC | 389.231 | 255.7747 | 44.1 | 0.02 |
| pHILIC | 389.231 | 182.789 | 44.1 | 0.02 |
| pHILIC | 389.231 | 93.715 | 44.1 | 0.02 |

| | | | | |
|---|---|---|---|---|
| pHILIC | 389.231 | 76.4831 | 44.1 | 0.02 |
| pHILIC | 389.231 | 215.8098 | 44.1 | 0.02 |
| pHILIC | 389.231 | 98.9603 | 44 | 0.02 |
| pHILIC | 389.231 | 215.2036 | 44 | 0.02 |
| pHILIC | 389.231 | 89.2247 | 44 | 0.02 |
| pHILIC | 389.231 | 90.4794 | 44 | 0.02 |
| pHILIC | 389.231 | 126.6608 | 44 | 0.02 |
| pHILIC | 389.231 | 222.7952 | 44 | 0.02 |
| pHILIC | 389.231 | 56.1881 | 44 | 0.02 |
| pHILIC | 389.231 | 121.6576 | 44 | 0.02 |
| pHILIC | 389.231 | 165.0649 | 44 | 0.02 |
| pHILIC | 389.231 | 54.5397 | 44 | 0.02 |
| pHILIC | 389.231 | 289.2528 | 44 | 0.02 |
| pHILIC | 389.231 | 92.337 | 44 | 0.02 |
| pHILIC | 389.231 | 90.8379 | 43.9 | 0.02 |
| pHILIC | 389.231 | 328.0687 | 43.9 | 0.02 |
| pHILIC | 389.231 | 357.4994 | 43.9 | 0.02 |
| pHILIC | 389.231 | 257.6897 | 43.9 | 0.02 |
| pHILIC | 389.231 | 59.9154 | 43.9 | 0.02 |
| pHILIC | 389.231 | 50.7388 | 43.9 | 0.02 |
| pHILIC | 389.231 | 99.1211 | 43.9 | 0.02 |
| pHILIC | 389.231 | 64.0348 | 43.9 | 0.02 |
| pHILIC | 389.231 | 187.3521 | 43.9 | 0.02 |
| pHILIC | 389.231 | 82.5629 | 43.9 | 0.02 |
| pHILIC | 389.231 | 167.557 | 43.8 | 0.02 |
| pHILIC | 389.231 | 72.2037 | 43.8 | 0.02 |
| pHILIC | 389.231 | 72.6716 | 43.8 | 0.02 |
| pHILIC | 389.231 | 283.7809 | 43.8 | 0.02 |
| pHILIC | 389.231 | 111.3059 | 43.8 | 0.02 |
| pHILIC | 389.231 | 238.9729 | 43.8 | 0.02 |
| pHILIC | 389.231 | 249.1624 | 43.8 | 0.02 |
| pHILIC | 389.231 | 118.9452 | 43.8 | 0.02 |
| pHILIC | 389.231 | 64.0456 | 43.8 | 0.02 |
| pHILIC | 389.231 | 179.4587 | 43.8 | 0.02 |
| pHILIC | 389.231 | 194.1956 | 43.8 | 0.02 |
| pHILIC | 389.231 | 79.5289 | 43.8 | 0.02 |
| pHILIC | 389.231 | 94.6771 | 43.7 | 0.02 |
| pHILIC | 389.231 | 62.25 | 43.7 | 0.02 |
| pHILIC | 389.231 | 234.9102 | 43.7 | 0.02 |
| pHILIC | 389.231 | 95.4268 | 43.7 | 0.02 |
| pHILIC | 389.231 | 98.0856 | 43.7 | 0.02 |
| pHILIC | 389.231 | 125.1031 | 43.7 | 0.02 |

| | | | | |
|---|---|---|---|---|
| pHILIC | 389.231 | 144.6912 | 43.7 | 0.02 |
| pHILIC | 389.231 | 58.4919 | 43.7 | 0.02 |
| pHILIC | 389.231 | 218.4791 | 43.7 | 0.02 |
| pHILIC | 389.231 | 58.9893 | 43.7 | 0.02 |
| pHILIC | 389.231 | 229.9784 | 43.7 | 0.02 |
| pHILIC | 389.231 | 188.5986 | 43.7 | 0.02 |
| pHILIC | 389.231 | 195.6987 | 43.7 | 0.02 |
| pHILIC | 389.231 | 51.0751 | 43.6 | 0.02 |
| pHILIC | 389.231 | 65.3292 | 43.6 | 0.02 |
| pHILIC | 389.231 | 225.8363 | 43.6 | 0.02 |
| pHILIC | 389.231 | 119.4344 | 43.6 | 0.02 |
| pHILIC | 389.231 | 96.1007 | 43.6 | 0.02 |
| pHILIC | 389.231 | 356.2961 | 43.6 | 0.02 |
| pHILIC | 389.231 | 338.1662 | 43.6 | 0.02 |
| pHILIC | 389.231 | 315.3122 | 43.6 | 0.02 |
| pHILIC | 389.231 | 262.0658 | 43.6 | 0.02 |
| pHILIC | 389.231 | 410.3085 | 43.6 | 0.02 |
| pHILIC | 389.231 | 126.4403 | 43.6 | 0.02 |
| pHILIC | 389.231 | 173.2648 | 43.6 | 0.02 |
| pHILIC | 389.231 | 51.808 | 43.5 | 0.02 |
| pHILIC | 389.231 | 74.4743 | 43.5 | 0.02 |
| pHILIC | 389.231 | 266.4405 | 43.5 | 0.02 |
| pHILIC | 389.231 | 65.0324 | 43.5 | 0.02 |
| pHILIC | 389.231 | 132.5251 | 43.5 | 0.02 |
| pHILIC | 389.231 | 121.8401 | 43.5 | 0.02 |
| pHILIC | 389.231 | 142.5455 | 43.5 | 0.02 |
| pHILIC | 389.231 | 253.0848 | 43.5 | 0.02 |
| pHILIC | 389.231 | 103.6753 | 43.5 | 0.02 |
| pHILIC | 389.231 | 161.3824 | 43.5 | 0.02 |
| pHILIC | 389.231 | 237.7439 | 43.5 | 0.02 |
| pHILIC | 389.231 | 93.8817 | 43.4 | 0.02 |
| pHILIC | 389.231 | 217.3865 | 43.4 | 0.02 |
| pHILIC | 389.231 | 89.6257 | 43.4 | 0.02 |
| pHILIC | 389.231 | 240.644 | 43.4 | 0.02 |
| pHILIC | 389.231 | 91.9847 | 43.4 | 0.02 |
| pHILIC | 389.231 | 78.8801 | 43.4 | 0.02 |
| pHILIC | 389.231 | 307.0501 | 43.4 | 0.02 |
| pHILIC | 389.231 | 349.744 | 43.4 | 0.02 |
| pHILIC | 389.231 | 90.5061 | 43.4 | 0.02 |
| pHILIC | 389.231 | 108.4524 | 43.4 | 0.02 |
| pHILIC | 389.231 | 92.5899 | 43.4 | 0.02 |
| pHILIC | 389.231 | 56.3087 | 43.4 | 0.02 |

| | | | | |
|---|---|---|---|---|
| pHILIC | 389.231 | 93.6083 | 43.3 | 0.02 |
| pHILIC | 389.231 | 81.71 | 43.3 | 0.02 |
| pHILIC | 389.231 | 56.0342 | 43.3 | 0.02 |
| pHILIC | 389.231 | 126.5502 | 43.3 | 0.02 |
| pHILIC | 389.231 | 148.7503 | 43.3 | 0.02 |
| pHILIC | 389.231 | 62.2596 | 43.3 | 0.02 |
| pHILIC | 389.231 | 292.7689 | 43.3 | 0.02 |
| pHILIC | 389.231 | 396.9339 | 43.3 | 0.02 |
| pHILIC | 389.231 | 116.6383 | 43.3 | 0.02 |
| pHILIC | 389.231 | 121.6144 | 43.3 | 0.02 |
| pHILIC | 389.231 | 87.2166 | 43.3 | 0.02 |
| pHILIC | 389.231 | 262.7391 | 43.3 | 0.02 |
| pHILIC | 389.231 | 113.8026 | 43.3 | 0.02 |
| pHILIC | 389.231 | 142.6266 | 43.3 | 0.02 |
| pHILIC | 389.231 | 56.6062 | 43.3 | 0.02 |
| pHILIC | 389.231 | 50.8565 | 43.2 | 0.02 |
| pHILIC | 389.231 | 186.734 | 43.2 | 0.02 |
| pHILIC | 389.231 | 68.1324 | 43.2 | 0.02 |
| pHILIC | 389.231 | 141.65 | 43.2 | 0.02 |
| pHILIC | 389.231 | 159.2552 | 43.2 | 0.02 |
| pHILIC | 389.231 | 85.2102 | 43.2 | 0.02 |
| pHILIC | 389.231 | 121.1559 | 43.2 | 0.02 |
| pHILIC | 389.231 | 71.5222 | 43.2 | 0.02 |
| pHILIC | 389.231 | 102.5055 | 43.2 | 0.02 |
| pHILIC | 389.231 | 140.4829 | 43.2 | 0.02 |
| pHILIC | 389.231 | 59.8649 | 43.2 | 0.02 |
| pHILIC | 389.231 | 98.6286 | 43.2 | 0.02 |
| pHILIC | 389.231 | 53.8709 | 43.2 | 0.02 |
| pHILIC | 389.231 | 327.774 | 43.2 | 0.02 |
| pHILIC | 389.231 | 53.5962 | 43.2 | 0.02 |
| pHILIC | 389.231 | 80.5625 | 43.2 | 0.02 |
| pHILIC | 389.231 | 128.8857 | 43.2 | 0.02 |
| pHILIC | 389.231 | 74.8148 | 43.2 | 0.02 |
| pHILIC | 389.231 | 71.2925 | 43.2 | 0.02 |
| pHILIC | 389.231 | 51.4001 | 43.1 | 0.02 |
| pHILIC | 389.231 | 160.2099 | 43.1 | 0.02 |
| pHILIC | 389.231 | 304.2461 | 43.1 | 0.02 |
| pHILIC | 389.231 | 374.6044 | 43.1 | 0.02 |
| pHILIC | 389.231 | 379.6075 | 43.1 | 0.02 |
| pHILIC | 389.231 | 188.3899 | 43.1 | 0.02 |
| pHILIC | 389.231 | 156.2043 | 43.1 | 0.02 |
| pHILIC | 389.231 | 61.5398 | 43.1 | 0.02 |

| | | | | |
|---|---|---|---|---|
| pHILIC | 389.231 | 68.9342 | 43.1 | 0.02 |
| pHILIC | 389.231 | 72.7437 | 43.1 | 0.02 |
| pHILIC | 389.231 | 156.9261 | 43.1 | 0.02 |
| pHILIC | 389.231 | 184.0433 | 43.1 | 0.02 |
| pHILIC | 389.231 | 158.7664 | 43.1 | 0.02 |
| pHILIC | 389.231 | 66.4912 | 43 | 0.02 |
| pHILIC | 389.231 | 93.6578 | 43 | 0.02 |
| pHILIC | 389.231 | 148.8315 | 43 | 0.02 |
| pHILIC | 389.231 | 84.2377 | 43 | 0.02 |
| pHILIC | 389.231 | 126.9768 | 43 | 0.02 |
| pHILIC | 389.231 | 190.1815 | 43 | 0.02 |
| pHILIC | 389.231 | 100.8209 | 43 | 0.02 |
| pHILIC | 389.231 | 381.6923 | 43 | 0.02 |
| pHILIC | 389.231 | 80.7191 | 43 | 0.02 |
| pHILIC | 389.231 | 76.2132 | 43 | 0.02 |
| pHILIC | 389.231 | 58.3692 | 43 | 0.02 |
| pHILIC | 389.231 | 54.1314 | 43 | 0.02 |
| pHILIC | 389.231 | 267.1915 | 43 | 0.02 |
| pHILIC | 389.231 | 66.6314 | 43 | 0.02 |
| pHILIC | 389.231 | 67.6163 | 43 | 0.02 |
| pHILIC | 389.231 | 195.1515 | 43 | 0.02 |
| pHILIC | 389.231 | 76.3276 | 43 | 0.02 |
| pHILIC | 389.231 | 72.1354 | 43 | 0.02 |
| pHILIC | 389.231 | 110.6464 | 43 | 0.02 |
| pHILIC | 389.231 | 164.7725 | 43 | 0.02 |
| pHILIC | 389.231 | 55.038 | 42.9 | 0.02 |
| pHILIC | 389.231 | 159.7583 | 42.9 | 0.02 |
| pHILIC | 389.231 | 104.8861 | 42.9 | 0.02 |
| pHILIC | 389.231 | 73.3635 | 42.9 | 0.02 |
| pHILIC | 389.231 | 108.1882 | 42.9 | 0.02 |
| pHILIC | 389.231 | 93.1383 | 42.9 | 0.02 |
| pHILIC | 389.231 | 410.2357 | 42.9 | 0.02 |
| pHILIC | 389.231 | 73.0996 | 42.9 | 0.02 |
| pHILIC | 389.231 | 110.1505 | 42.9 | 0.02 |
| pHILIC | 389.231 | 112.3721 | 42.9 | 0.02 |
| pHILIC | 389.231 | 137.0898 | 42.8 | 0.02 |
| pHILIC | 389.231 | 72.5696 | 42.8 | 0.02 |
| pHILIC | 389.231 | 75.8789 | 42.8 | 0.02 |
| pHILIC | 389.231 | 114.4982 | 42.8 | 0.02 |
| pHILIC | 389.231 | 95.3857 | 42.8 | 0.02 |
| pHILIC | 389.231 | 67.9809 | 42.8 | 0.02 |
| pHILIC | 389.231 | 55.5321 | 42.8 | 0.02 |

| | | | | |
|---|---|---|---|---|
| pHILIC | 389.231 | 92.378 | 42.7 | 0.02 |
| pHILIC | 389.231 | 367.1301 | 42.7 | 0.02 |
| pHILIC | 389.231 | 327.3268 | 42.7 | 0.02 |
| pHILIC | 389.231 | 123.8079 | 42.7 | 0.02 |
| pHILIC | 389.231 | 69.5134 | 42.7 | 0.02 |
| pHILIC | 389.231 | 85.427 | 42.7 | 0.02 |
| pHILIC | 389.231 | 66.8683 | 42.7 | 0.02 |
| pHILIC | 389.231 | 151.8203 | 42.7 | 0.02 |
| pHILIC | 389.231 | 62.6267 | 42.7 | 0.02 |
| pHILIC | 389.231 | 58.6039 | 42.7 | 0.02 |
| pHILIC | 389.231 | 89.9412 | 42.7 | 0.02 |
| pHILIC | 389.231 | 156.0591 | 42.7 | 0.02 |
| pHILIC | 389.231 | 50.1458 | 42.7 | 0.02 |
| pHILIC | 389.231 | 263.982 | 42.7 | 0.02 |
| pHILIC | 389.231 | 59.1314 | 42.7 | 0.02 |
| pHILIC | 389.231 | 69.1287 | 42.7 | 0.02 |
| pHILIC | 389.231 | 77.3954 | 42.7 | 0.02 |
| pHILIC | 389.231 | 290.7134 | 42.6 | 0.02 |
| pHILIC | 389.231 | 393.9148 | 42.6 | 0.02 |
| pHILIC | 389.231 | 145.3874 | 42.6 | 0.02 |
| pHILIC | 389.231 | 244.4655 | 42.6 | 0.02 |
| pHILIC | 389.231 | 146.5456 | 42.6 | 0.02 |
| pHILIC | 389.231 | 189.616 | 42.6 | 0.02 |
| pHILIC | 389.231 | 58.053 | 42.6 | 0.02 |
| pHILIC | 389.231 | 126.9938 | 42.6 | 0.02 |
| pHILIC | 389.231 | 64.7256 | 42.6 | 0.02 |
| pHILIC | 389.231 | 111.3483 | 42.6 | 0.02 |
| pHILIC | 389.231 | 54.899 | 42.6 | 0.02 |
| pHILIC | 389.231 | 206.9617 | 42.6 | 0.02 |
| pHILIC | 389.231 | 116.2944 | 42.6 | 0.02 |
| pHILIC | 389.231 | 151.3967 | 42.6 | 0.02 |
| pHILIC | 389.231 | 54.906 | 42.6 | 0.02 |
| pHILIC | 389.231 | 75.9812 | 42.5 | 0.02 |
| pHILIC | 389.231 | 89.5843 | 42.5 | 0.02 |
| pHILIC | 389.231 | 194.1729 | 42.5 | 0.02 |
| pHILIC | 389.231 | 99.3903 | 42.5 | 0.02 |
| pHILIC | 389.231 | 78.6387 | 42.5 | 0.02 |
| pHILIC | 389.231 | 78.3386 | 42.5 | 0.02 |
| pHILIC | 389.231 | 274.6312 | 42.5 | 0.02 |
| pHILIC | 389.231 | 94.0356 | 42.5 | 0.02 |
| pHILIC | 389.231 | 56.8193 | 42.5 | 0.02 |
| pHILIC | 389.231 | 329.964 | 42.5 | 0.02 |

| | | | | |
|---|---|---|---|---|
| pHILIC | 389.231 | 52.7499 | 42.5 | 0.02 |
| pHILIC | 389.231 | 174.0068 | 42.5 | 0.02 |
| pHILIC | 389.231 | 78.929 | 42.5 | 0.02 |
| pHILIC | 389.231 | 237.0715 | 42.5 | 0.02 |
| pHILIC | 389.231 | 353.176 | 42.5 | 0.02 |
| pHILIC | 389.231 | 94.7921 | 42.5 | 0.02 |
| pHILIC | 389.231 | 176.371 | 42.5 | 0.02 |
| pHILIC | 389.231 | 51.0589 | 42.4 | 0.02 |
| pHILIC | 389.231 | 210.5093 | 42.4 | 0.02 |
| pHILIC | 389.231 | 95.8211 | 42.4 | 0.02 |
| pHILIC | 389.231 | 126.2414 | 42.4 | 0.02 |
| pHILIC | 389.231 | 112.6286 | 42.4 | 0.02 |
| pHILIC | 389.231 | 112.7478 | 42.4 | 0.02 |
| pHILIC | 389.231 | 90.1165 | 42.4 | 0.02 |
| pHILIC | 389.231 | 63.658 | 42.4 | 0.02 |
| pHILIC | 389.231 | 59.9838 | 42.4 | 0.02 |
| pHILIC | 389.231 | 86.4031 | 42.4 | 0.02 |
| pHILIC | 389.231 | 101.9375 | 42.4 | 0.02 |
| pHILIC | 389.231 | 50.2434 | 42.4 | 0.02 |
| pHILIC | 389.231 | 78.1625 | 42.4 | 0.02 |
| pHILIC | 389.231 | 56.9236 | 42.4 | 0.02 |
| pHILIC | 389.231 | 81.8397 | 42.4 | 0.02 |
| pHILIC | 389.231 | 181.9871 | 42.4 | 0.02 |
| pHILIC | 389.231 | 101.6503 | 42.4 | 0.02 |
| pHILIC | 389.231 | 181.5717 | 42.4 | 0.02 |
| pHILIC | 389.231 | 217.336 | 42.4 | 0.02 |
| pHILIC | 389.231 | 254.7407 | 42.4 | 0.02 |
| pHILIC | 389.231 | 51.661 | 42.3 | 0.02 |
| pHILIC | 389.231 | 214.2627 | 42.3 | 0.02 |
| pHILIC | 389.231 | 55.9192 | 42.3 | 0.02 |
| pHILIC | 389.231 | 78.6094 | 42.3 | 0.02 |
| pHILIC | 389.231 | 109.032 | 42.3 | 0.02 |
| pHILIC | 389.231 | 94.6916 | 42.3 | 0.02 |
| pHILIC | 389.231 | 76.3052 | 42.3 | 0.02 |
| pHILIC | 389.231 | 75.5413 | 42.3 | 0.02 |
| pHILIC | 389.231 | 63.2635 | 42.3 | 0.02 |
| pHILIC | 389.231 | 113.6283 | 42.3 | 0.02 |
| pHILIC | 389.231 | 158.7889 | 42.3 | 0.02 |
| pHILIC | 389.231 | 382.7706 | 42.3 | 0.02 |
| pHILIC | 389.231 | 338.8486 | 42.3 | 0.02 |
| pHILIC | 389.231 | 76.6296 | 42.3 | 0.02 |
| pHILIC | 389.231 | 122.5409 | 42.3 | 0.02 |

| | | | | |
|---|---|---|---|---|
| pHILIC | 389.231 | 50.7744 | 42.3 | 0.02 |
| pHILIC | 389.231 | 68.9479 | 42.2 | 0.02 |
| pHILIC | 389.231 | 197.1023 | 42.2 | 0.02 |
| pHILIC | 389.231 | 91.3218 | 42.2 | 0.02 |
| pHILIC | 389.231 | 51.5853 | 42.2 | 0.02 |
| pHILIC | 389.231 | 60.8596 | 42.2 | 0.02 |
| pHILIC | 389.231 | 110.1936 | 42.2 | 0.02 |
| pHILIC | 389.231 | 191.5581 | 42.2 | 0.02 |
| pHILIC | 389.231 | 87.9138 | 42.2 | 0.02 |
| pHILIC | 389.231 | 61.173 | 42.2 | 0.02 |
| pHILIC | 389.231 | 128.2953 | 42.2 | 0.02 |
| pHILIC | 389.231 | 287.2733 | 42.1 | 0.02 |
| pHILIC | 389.231 | 221.5865 | 42.1 | 0.02 |
| pHILIC | 389.231 | 55.9719 | 42.1 | 0.02 |
| pHILIC | 389.231 | 52.1313 | 42.1 | 0.02 |
| pHILIC | 389.231 | 79.184 | 42.1 | 0.02 |
| pHILIC | 389.231 | 156.111 | 42.1 | 0.02 |
| pHILIC | 389.231 | 83.5218 | 42.1 | 0.02 |
| pHILIC | 389.231 | 66.5255 | 42.1 | 0.02 |
| pHILIC | 389.231 | 110.9824 | 42.1 | 0.02 |
| pHILIC | 389.231 | 97.0387 | 42.1 | 0.02 |
| pHILIC | 389.231 | 279.2123 | 42.1 | 0.02 |
| pHILIC | 389.231 | 71.3808 | 42.1 | 0.02 |
| pHILIC | 389.231 | 53.2881 | 42.1 | 0.02 |
| pHILIC | 389.231 | 71.8421 | 42 | 0.02 |
| pHILIC | 389.231 | 126.8783 | 42 | 0.02 |
| pHILIC | 389.231 | 167.9584 | 42 | 0.02 |
| pHILIC | 389.231 | 64.7336 | 42 | 0.02 |
| pHILIC | 389.231 | 78.1589 | 42 | 0.02 |
| pHILIC | 389.231 | 116.6281 | 42 | 0.02 |
| pHILIC | 389.231 | 196.5994 | 42 | 0.02 |
| pHILIC | 389.231 | 290.5399 | 42 | 0.02 |
| pHILIC | 389.231 | 188.0334 | 42 | 0.02 |
| pHILIC | 389.231 | 279.8115 | 42 | 0.02 |
| pHILIC | 389.231 | 102.8996 | 42 | 0.02 |
| pHILIC | 389.231 | 58.6 | 42 | 0.02 |
| pHILIC | 389.231 | 51.7257 | 42 | 0.02 |
| pHILIC | 389.231 | 155.6417 | 41.9 | 0.02 |
| pHILIC | 389.231 | 345.3188 | 41.9 | 0.02 |
| pHILIC | 389.231 | 358.1837 | 41.9 | 0.02 |
| pHILIC | 389.231 | 173.3012 | 41.9 | 0.02 |
| pHILIC | 389.231 | 54.2781 | 41.9 | 0.02 |

| | | | | |
|---|---|---|---|---|
| pHILIC | 389.231 | 59.1266 | 41.9 | 0.02 |
| pHILIC | 389.231 | 86.4957 | 41.9 | 0.02 |
| pHILIC | 389.231 | 93.5638 | 41.9 | 0.02 |
| pHILIC | 389.231 | 162.5204 | 41.9 | 0.02 |
| pHILIC | 389.231 | 106.0526 | 41.9 | 0.02 |
| pHILIC | 389.231 | 171.2351 | 41.9 | 0.02 |
| pHILIC | 389.231 | 131.5648 | 41.9 | 0.02 |
| pHILIC | 389.231 | 164.2303 | 41.9 | 0.02 |
| pHILIC | 389.231 | 73.3137 | 41.9 | 0.02 |
| pHILIC | 389.231 | 234.1468 | 41.8 | 0.02 |
| pHILIC | 389.231 | 151.055 | 41.8 | 0.02 |
| pHILIC | 389.231 | 181.1817 | 41.8 | 0.02 |
| pHILIC | 389.231 | 225.7666 | 41.8 | 0.02 |
| pHILIC | 389.231 | 148.5628 | 41.8 | 0.02 |
| pHILIC | 389.231 | 340.1622 | 41.8 | 0.02 |
| pHILIC | 389.231 | 51.7285 | 41.8 | 0.02 |
| pHILIC | 389.231 | 400.3823 | 41.8 | 0.02 |
| pHILIC | 389.231 | 83.296 | 41.8 | 0.02 |
| pHILIC | 389.231 | 71.061 | 41.8 | 0.02 |
| pHILIC | 389.231 | 114.7872 | 41.7 | 0.02 |
| pHILIC | 389.231 | 139.396 | 41.7 | 0.02 |
| pHILIC | 389.231 | 74.4313 | 41.7 | 0.02 |
| pHILIC | 389.231 | 130.8753 | 41.7 | 0.02 |
| pHILIC | 389.231 | 195.5701 | 41.7 | 0.02 |
| pHILIC | 389.231 | 138.6246 | 41.7 | 0.02 |
| pHILIC | 389.231 | 54.7909 | 41.7 | 0.02 |
| pHILIC | 389.231 | 66.0727 | 41.7 | 0.02 |
| pHILIC | 389.231 | 247.5791 | 41.7 | 0.02 |
| pHILIC | 389.231 | 139.1119 | 41.7 | 0.02 |
| pHILIC | 389.231 | 86.0281 | 41.7 | 0.02 |
| pHILIC | 389.231 | 52.3707 | 41.7 | 0.02 |
| pHILIC | 389.231 | 84.7626 | 41.7 | 0.02 |
| pHILIC | 389.231 | 131.0316 | 41.7 | 0.02 |
| pHILIC | 389.231 | 150.754 | 41.7 | 0.02 |
| pHILIC | 389.231 | 85.6233 | 41.6 | 0.02 |
| pHILIC | 389.231 | 97.9108 | 41.6 | 0.02 |
| pHILIC | 389.231 | 182.4055 | 41.6 | 0.02 |
| pHILIC | 389.231 | 196.1227 | 41.6 | 0.02 |
| pHILIC | 389.231 | 159.9179 | 41.6 | 0.02 |
| pHILIC | 389.231 | 88.1261 | 41.6 | 0.02 |
| pHILIC | 389.231 | 160.642 | 41.6 | 0.02 |
| pHILIC | 389.231 | 263.0053 | 41.6 | 0.02 |

| | | | | |
|---|---|---|---|---|
| pHILIC | 389.231 | 181.9694 | 41.6 | 0.02 |
| pHILIC | 389.231 | 120.2909 | 41.6 | 0.02 |
| pHILIC | 389.231 | 106.4195 | 41.6 | 0.02 |
| pHILIC | 389.231 | 58.273 | 41.5 | 0.02 |
| pHILIC | 389.231 | 58.7484 | 41.5 | 0.02 |
| pHILIC | 389.231 | 161.6163 | 41.5 | 0.02 |
| pHILIC | 389.231 | 117.2413 | 41.5 | 0.02 |
| pHILIC | 389.231 | 60.0279 | 41.5 | 0.02 |
| pHILIC | 389.231 | 66.7193 | 41.5 | 0.02 |
| pHILIC | 389.231 | 76.2732 | 41.5 | 0.02 |
| pHILIC | 389.231 | 54.3525 | 41.5 | 0.02 |
| pHILIC | 389.231 | 181.2402 | 41.5 | 0.02 |
| pHILIC | 389.231 | 85.9968 | 41.5 | 0.02 |
| pHILIC | 389.231 | 255.4073 | 41.5 | 0.02 |
| pHILIC | 389.231 | 54.302 | 41.5 | 0.02 |
| pHILIC | 389.231 | 103.0008 | 41.4 | 0.02 |
| pHILIC | 389.231 | 126.337 | 41.4 | 0.02 |
| pHILIC | 389.231 | 80.0462 | 41.4 | 0.02 |
| pHILIC | 389.231 | 147.3494 | 41.4 | 0.02 |
| pHILIC | 389.231 | 71.8084 | 41.4 | 0.02 |
| pHILIC | 389.231 | 102.9404 | 41.4 | 0.02 |
| pHILIC | 389.231 | 66.384 | 41.3 | 0.02 |
| pHILIC | 389.231 | 160.4535 | 41.3 | 0.02 |
| pHILIC | 389.231 | 104.9662 | 41.3 | 0.02 |
| pHILIC | 389.231 | 106.327 | 41.3 | 0.02 |
| pHILIC | 389.231 | 377.4761 | 41.3 | 0.02 |
| pHILIC | 389.231 | 66.5295 | 41.3 | 0.02 |
| pHILIC | 389.231 | 50.0113 | 41.3 | 0.02 |
| pHILIC | 389.231 | 70.1896 | 41.3 | 0.02 |
| pHILIC | 389.231 | 133.3398 | 41.3 | 0.02 |
| pHILIC | 389.231 | 86.3758 | 41.3 | 0.02 |
| pHILIC | 389.231 | 103.0874 | 41.3 | 0.02 |
| pHILIC | 389.231 | 104.2957 | 41.3 | 0.02 |
| pHILIC | 389.231 | 190.0874 | 41.3 | 0.02 |
| pHILIC | 389.231 | 101.8919 | 41.3 | 0.02 |
| pHILIC | 389.231 | 78.1114 | 41.3 | 0.02 |
| pHILIC | 389.231 | 191.6382 | 41.3 | 0.02 |
| pHILIC | 389.231 | 119.146 | 41.3 | 0.02 |
| pHILIC | 389.231 | 82.2915 | 41.3 | 0.02 |
| pHILIC | 389.231 | 290.4282 | 41.2 | 0.02 |
| pHILIC | 389.231 | 66.0197 | 41.2 | 0.02 |
| pHILIC | 389.231 | 320.1452 | 41.2 | 0.02 |

| | | | | |
|---|---|---|---|---|
| pHILIC | 389.231 | 180.3213 | 41.2 | 0.02 |
| pHILIC | 389.231 | 58.5485 | 41.2 | 0.02 |
| pHILIC | 389.231 | 233.0106 | 41.2 | 0.02 |
| pHILIC | 389.231 | 123.8242 | 41.1 | 0.02 |
| pHILIC | 389.231 | 152.7925 | 41.1 | 0.02 |
| pHILIC | 389.231 | 177.3249 | 41.1 | 0.02 |
| pHILIC | 389.231 | 121.1027 | 41.1 | 0.02 |
| pHILIC | 389.231 | 154.8767 | 41.1 | 0.02 |
| pHILIC | 389.231 | 199.0742 | 41.1 | 0.02 |
| pHILIC | 389.231 | 129.3114 | 41.1 | 0.02 |
| pHILIC | 389.231 | 301.5872 | 41.1 | 0.02 |
| pHILIC | 389.231 | 276.36 | 41.1 | 0.02 |
| pHILIC | 389.231 | 150.4723 | 41.1 | 0.02 |
| pHILIC | 389.231 | 146.1329 | 41 | 0.02 |
| pHILIC | 389.231 | 102.2719 | 41 | 0.02 |
| pHILIC | 389.231 | 61.8921 | 41 | 0.02 |
| pHILIC | 389.231 | 53.2212 | 41 | 0.02 |
| pHILIC | 389.231 | 51.5156 | 41 | 0.02 |
| pHILIC | 389.231 | 126.7586 | 41 | 0.02 |
| pHILIC | 389.231 | 102.0251 | 41 | 0.02 |
| pHILIC | 389.231 | 113.7273 | 41 | 0.02 |
| pHILIC | 389.231 | 74.3417 | 41 | 0.02 |
| pHILIC | 389.231 | 137.3675 | 41 | 0.02 |
| pHILIC | 389.231 | 178.3554 | 41 | 0.02 |
| pHILIC | 389.231 | 194.3509 | 41 | 0.02 |
| pHILIC | 389.231 | 56.6451 | 41 | 0.02 |
| pHILIC | 389.231 | 91.9921 | 41 | 0.02 |
| pHILIC | 389.231 | 93.0216 | 41 | 0.02 |
| pHILIC | 389.231 | 76.1333 | 40.9 | 0.02 |
| pHILIC | 389.231 | 238.5875 | 40.9 | 0.02 |
| pHILIC | 389.231 | 73.4491 | 40.9 | 0.02 |
| pHILIC | 389.231 | 117.781 | 40.9 | 0.02 |
| pHILIC | 389.231 | 77.0329 | 40.9 | 0.02 |
| pHILIC | 389.231 | 274.3373 | 40.9 | 0.02 |
| pHILIC | 389.231 | 132.6201 | 40.9 | 0.02 |
| pHILIC | 389.231 | 105.7579 | 40.9 | 0.02 |
| pHILIC | 389.231 | 102.5106 | 40.9 | 0.02 |
| pHILIC | 389.231 | 61.5631 | 40.8 | 0.01 |
| pHILIC | 389.231 | 103.6173 | 40.8 | 0.01 |
| pHILIC | 389.231 | 300.1901 | 40.8 | 0.01 |
| pHILIC | 389.231 | 149.4697 | 40.8 | 0.01 |
| pHILIC | 389.231 | 178.8395 | 40.8 | 0.01 |

| | | | | |
|---|---|---|---|---|
| pHILIC | 389.231 | 222.0303 | 40.8 | 0.01 |
| pHILIC | 389.231 | 129.8639 | 40.8 | 0.01 |
| pHILIC | 389.231 | 62.1624 | 40.8 | 0.01 |
| pHILIC | 389.231 | 372.9832 | 40.8 | 0.01 |
| pHILIC | 389.231 | 186.1503 | 40.8 | 0.01 |
| pHILIC | 389.231 | 284.8756 | 40.7 | 0.01 |
| pHILIC | 389.231 | 106.931 | 40.7 | 0.01 |
| pHILIC | 389.231 | 92.0015 | 40.7 | 0.01 |
| pHILIC | 389.231 | 58.411 | 40.7 | 0.01 |
| pHILIC | 389.231 | 116.6731 | 40.7 | 0.01 |
| pHILIC | 389.231 | 67.8559 | 40.7 | 0.01 |
| pHILIC | 389.231 | 97.2686 | 40.7 | 0.01 |
| pHILIC | 389.231 | 100.4335 | 40.7 | 0.01 |
| pHILIC | 389.231 | 314.7554 | 40.7 | 0.01 |
| pHILIC | 389.231 | 103.5248 | 40.7 | 0.01 |
| pHILIC | 389.231 | 184.2181 | 40.7 | 0.01 |
| pHILIC | 389.231 | 285.1587 | 40.7 | 0.01 |
| pHILIC | 389.231 | 199.1581 | 40.7 | 0.01 |
| pHILIC | 389.231 | 92.3256 | 40.6 | 0.01 |
| pHILIC | 389.231 | 72.5623 | 40.6 | 0.01 |
| pHILIC | 389.231 | 97.9806 | 40.6 | 0.01 |
| pHILIC | 389.231 | 199.9839 | 40.6 | 0.01 |
| pHILIC | 389.231 | 57.0674 | 40.6 | 0.01 |
| pHILIC | 389.231 | 119.1319 | 40.6 | 0.01 |
| pHILIC | 389.231 | 84.9879 | 40.6 | 0.01 |
| pHILIC | 389.231 | 65.4224 | 40.6 | 0.01 |
| pHILIC | 389.231 | 63.694 | 40.5 | 0.01 |
| pHILIC | 389.231 | 177.6103 | 40.5 | 0.01 |
| pHILIC | 389.231 | 87.4979 | 40.5 | 0.01 |
| pHILIC | 389.231 | 102.0437 | 40.4 | 0.01 |
| pHILIC | 389.231 | 62.425 | 40.4 | 0.01 |
| pHILIC | 389.231 | 94.1431 | 40.4 | 0.01 |
| pHILIC | 389.231 | 187.9691 | 40.4 | 0.01 |
| pHILIC | 389.231 | 100.8819 | 40.4 | 0.01 |
| pHILIC | 389.231 | 148.864 | 40.4 | 0.01 |
| pHILIC | 389.231 | 174.2103 | 40.4 | 0.01 |
| pHILIC | 389.231 | 120.1739 | 40.4 | 0.01 |
| pHILIC | 389.231 | 149.1019 | 40.3 | 0.01 |
| pHILIC | 389.231 | 88.1848 | 40.3 | 0.01 |
| pHILIC | 389.231 | 59.0121 | 40.3 | 0.01 |
| pHILIC | 389.231 | 88.0636 | 40.3 | 0.01 |
| pHILIC | 389.231 | 101.724 | 40.3 | 0.01 |

| | | | | |
|---|---|---|---|---|
| pHILIC | 389.231 | 56.1563 | 40.3 | 0.01 |
| pHILIC | 389.231 | 170.5866 | 40.3 | 0.01 |
| pHILIC | 389.231 | 106.906 | 40.3 | 0.01 |
| pHILIC | 389.231 | 65.6045 | 40.3 | 0.01 |
| pHILIC | 389.231 | 84.22 | 40.2 | 0.01 |
| pHILIC | 389.231 | 112.8386 | 40.2 | 0.01 |
| pHILIC | 389.231 | 70.0165 | 40.2 | 0.01 |
| pHILIC | 389.231 | 287.449 | 40.2 | 0.01 |
| pHILIC | 389.231 | 112.3592 | 40.2 | 0.01 |
| pHILIC | 389.231 | 54.2919 | 40.2 | 0.01 |
| pHILIC | 389.231 | 50.9009 | 40.1 | 0.01 |
| pHILIC | 389.231 | 63.5588 | 40.1 | 0.01 |
| pHILIC | 389.231 | 153.6439 | 40.1 | 0.01 |
| pHILIC | 389.231 | 91.0887 | 40.1 | 0.01 |
| pHILIC | 389.231 | 50.8814 | 40 | 0.01 |
| pHILIC | 389.231 | 284.7074 | 40 | 0.01 |
| pHILIC | 389.231 | 100.9855 | 40 | 0.01 |
| pHILIC | 389.231 | 193.7669 | 40 | 0.01 |
| pHILIC | 389.231 | 191.8582 | 40 | 0.01 |
| pHILIC | 389.231 | 183.1084 | 40 | 0.01 |
| pHILIC | 389.231 | 70.6895 | 40 | 0.01 |
| pHILIC | 389.231 | 233.053 | 39.9 | 0.01 |
| pHILIC | 389.231 | 58.7246 | 39.9 | 0.01 |
| pHILIC | 389.231 | 89.928 | 39.9 | 0.01 |
| pHILIC | 389.231 | 54.4451 | 39.9 | 0.01 |
| pHILIC | 389.231 | 118.1872 | 39.9 | 0.01 |
| pHILIC | 389.231 | 167.977 | 39.9 | 0.01 |
| pHILIC | 389.231 | 74.6971 | 39.9 | 0.01 |
| pHILIC | 389.231 | 59.1669 | 39.8 | 0.01 |
| pHILIC | 389.231 | 60.6169 | 39.8 | 0.01 |
| pHILIC | 389.231 | 91.299 | 39.7 | 0.01 |
| pHILIC | 389.231 | 117.121 | 39.7 | 0.01 |
| pHILIC | 389.231 | 102.9068 | 39.7 | 0.01 |
| pHILIC | 389.231 | 58.3402 | 39.7 | 0.01 |
| pHILIC | 389.231 | 117.6398 | 39.7 | 0.01 |
| pHILIC | 389.231 | 56.4739 | 39.6 | 0.01 |
| pHILIC | 389.231 | 69.0213 | 39.6 | 0.01 |
| pHILIC | 389.231 | 74.9729 | 39.6 | 0.01 |
| pHILIC | 389.231 | 82.3613 | 39.6 | 0.01 |
| pHILIC | 389.231 | 64.3476 | 39.6 | 0.01 |
| pHILIC | 389.231 | 64.4047 | 39.6 | 0.01 |
| pHILIC | 389.231 | 92.4152 | 39.6 | 0.01 |

| | | | | |
|---|---|---|---|---|
| pHILIC | 389.231 | 51.6512 | 39.6 | 0.01 |
| pHILIC | 389.231 | 56.9214 | 39.5 | 0.01 |
| pHILIC | 389.231 | 50.0695 | 39.5 | 0.01 |
| pHILIC | 389.231 | 57.0918 | 39.5 | 0.01 |
| pHILIC | 389.231 | 53.4228 | 39.4 | 0.01 |
| pHILIC | 389.231 | 299.616 | 39.4 | 0.01 |
| pHILIC | 389.231 | 75.711 | 39.4 | 0.01 |
| pHILIC | 389.231 | 258.1283 | 39.4 | 0.01 |
| pHILIC | 389.231 | 120.4631 | 39.4 | 0.01 |
| pHILIC | 389.231 | 183.5227 | 39.4 | 0.01 |
| pHILIC | 389.231 | 203.401 | 39.4 | 0.01 |
| pHILIC | 389.231 | 220.0901 | 39.3 | 0.01 |
| pHILIC | 389.231 | 305.4151 | 39.3 | 0.01 |
| pHILIC | 389.231 | 64.5577 | 39.3 | 0.01 |
| pHILIC | 389.231 | 162.4212 | 39.2 | 0.01 |
| pHILIC | 389.231 | 77.0296 | 39.2 | 0.01 |
| pHILIC | 389.231 | 82.1581 | 39.2 | 0.01 |
| pHILIC | 389.231 | 81.7021 | 39.2 | 0.01 |
| pHILIC | 389.231 | 110.9602 | 39.2 | 0.01 |
| pHILIC | 389.231 | 173.6879 | 39.1 | 0.01 |
| pHILIC | 389.231 | 117.232 | 39.1 | 0.01 |
| pHILIC | 389.231 | 50.5566 | 39.1 | 0.01 |
| pHILIC | 389.231 | 59.9966 | 39.1 | 0.01 |
| pHILIC | 389.231 | 55.6451 | 39.1 | 0.01 |
| pHILIC | 389.231 | 61.0692 | 39.1 | 0.01 |
| pHILIC | 389.231 | 330.7231 | 39 | 0.01 |
| pHILIC | 389.231 | 126.4763 | 39 | 0.01 |
| pHILIC | 389.231 | 74.7358 | 39 | 0.01 |
| pHILIC | 389.231 | 90.5682 | 38.9 | 0.01 |
| pHILIC | 389.231 | 117.4717 | 38.8 | 0.01 |
| pHILIC | 389.231 | 70.9118 | 38.8 | 0.01 |
| pHILIC | 389.231 | 118.1561 | 38.8 | 0.01 |
| pHILIC | 389.231 | 116.5978 | 38.7 | 0.01 |
| pHILIC | 389.231 | 61.9195 | 38.7 | 0.01 |
| pHILIC | 389.231 | 63.8156 | 38.7 | 0.01 |
| pHILIC | 389.231 | 117.1728 | 38.7 | 0.01 |
| pHILIC | 389.231 | 71.9759 | 38.7 | 0.01 |
| pHILIC | 389.231 | 213.197 | 38.7 | 0.01 |
| pHILIC | 389.231 | 53.487 | 38.7 | 0.01 |
| pHILIC | 389.231 | 55.6371 | 38.6 | 0.01 |
| pHILIC | 389.231 | 84.9089 | 38.6 | 0.01 |
| pHILIC | 389.231 | 75.2214 | 38.5 | 0.01 |

| | | | | |
|---|---|---|---|---|
| pHILIC | 389.231 | 115.5534 | 38.5 | 0.01 |
| pHILIC | 389.231 | 164.5718 | 38.5 | 0.01 |
| pHILIC | 389.231 | 208.8978 | 38.5 | 0.01 |
| pHILIC | 389.231 | 50.5329 | 38.5 | 0.01 |
| pHILIC | 389.231 | 65.5529 | 38.5 | 0.01 |
| pHILIC | 389.231 | 67.0701 | 38.4 | 0.01 |
| pHILIC | 389.231 | 95.5133 | 38.3 | 0.01 |
| pHILIC | 389.231 | 118.8702 | 38.3 | 0.01 |
| pHILIC | 389.231 | 61.5795 | 38.3 | 0.01 |
| pHILIC | 389.231 | 115.2225 | 38.2 | 0.01 |
| pHILIC | 389.231 | 185.6421 | 38.2 | 0.01 |
| pHILIC | 389.231 | 65.8713 | 38.2 | 0.01 |
| pHILIC | 389.231 | 89.6411 | 38.2 | 0.01 |
| pHILIC | 389.231 | 71.2326 | 38.2 | 0.01 |
| pHILIC | 389.231 | 59.9817 | 38.1 | 0.01 |
| pHILIC | 389.231 | 76.6881 | 38 | 0.01 |
| pHILIC | 389.231 | 59.6517 | 37.9 | 0.01 |
| pHILIC | 389.231 | 105.0527 | 37.8 | 0.01 |
| pHILIC | 389.231 | 59.8347 | 37.7 | 0.01 |
| pHILIC | 389.231 | 85.2622 | 37.7 | 0.01 |
| pHILIC | 389.231 | 240.4781 | 37.4 | 0.01 |
| pHILIC | 389.231 | 76.0225 | 37.2 | 0.01 |
| pHILIC | 389.231 | 168.9821 | 36.9 | 0.01 |
| pHILIC | 389.231 | 63.5119 | 36.7 | 0.01 |
| pHILIC | 389.231 | 66.6449 | 36.6 | 0.01 |
| pHILIC | 389.231 | 109.0954 | 36.3 | 0.01 |
| pHILIC | 389.231 | 52.6824 | 36.3 | 0.01 |
| pHILIC | 599.503 | 105.0334 | 56986.6 | 100 |
| pHILIC | 599.503 | 89.0597 | 16923.3 | 29.7 |
| pHILIC | 599.503 | 133.0859 | 4792.3 | 8.41 |
| pHILIC | 599.503 | 599.2477 | 4025 | 7.06 |
| pHILIC | 599.503 | 228.219 | 2768.4 | 4.86 |
| pHILIC | 599.503 | 225.0756 | 2005.3 | 3.52 |
| pHILIC | 599.503 | 77.0385 | 861.4 | 1.51 |
| pHILIC | 599.503 | 95.0491 | 764.6 | 1.34 |
| pHILIC | 599.503 | 73.0283 | 583.5 | 1.02 |
| pHILIC | 599.503 | 71.0856 | 566.5 | 0.99 |
| pHILIC | 599.503 | 421.1468 | 555.2 | 0.97 |
| pHILIC | 599.503 | 161.0598 | 549.9 | 0.96 |
| pHILIC | 599.503 | 106.0373 | 451.4 | 0.79 |
| pHILIC | 599.503 | 269.1531 | 425.9 | 0.75 |
| pHILIC | 599.503 | 177.1123 | 408.9 | 0.72 |

| | | | | |
|---|---|---|---|---|
| pHILIC | 599.503 | 309.2787 | 381.5 | 0.67 |
| pHILIC | 599.503 | 179.0699 | 375.4 | 0.66 |
| pHILIC | 599.503 | 207.0655 | 357.1 | 0.63 |
| pHILIC | 599.503 | 133.0649 | 355.4 | 0.62 |
| pHILIC | 599.503 | 469.1099 | 275.5 | 0.48 |
| pHILIC | 599.503 | 389.175 | 264.6 | 0.46 |
| pHILIC | 599.503 | 243.0468 | 251.4 | 0.44 |
| pHILIC | 599.503 | 481.7955 | 194.9 | 0.34 |
| pHILIC | 599.503 | 564.3028 | 179.7 | 0.32 |
| pHILIC | 599.503 | 461.2348 | 175.3 | 0.31 |
| pHILIC | 599.503 | 471.1239 | 166.6 | 0.29 |
| pHILIC | 599.503 | 157.1259 | 160.4 | 0.28 |
| pHILIC | 599.503 | 299.1367 | 155.6 | 0.27 |
| pHILIC | 599.503 | 477.2133 | 154.6 | 0.27 |
| pHILIC | 599.503 | 87.0439 | 138.7 | 0.24 |
| pHILIC | 599.503 | 112.896 | 135.8 | 0.24 |
| pHILIC | 599.503 | 80.0543 | 135.4 | 0.24 |
| pHILIC | 599.503 | 583.2482 | 132.9 | 0.23 |
| pHILIC | 599.503 | 411.2762 | 129.7 | 0.23 |
| pHILIC | 599.503 | 453.9038 | 128.7 | 0.23 |
| pHILIC | 599.503 | 612.4784 | 127.5 | 0.22 |
| pHILIC | 599.503 | 412.0469 | 123.5 | 0.22 |
| pHILIC | 599.503 | 229.0912 | 114.6 | 0.2 |
| pHILIC | 599.503 | 478.2143 | 107.2 | 0.19 |
| pHILIC | 599.503 | 199.1235 | 106.9 | 0.19 |
| pHILIC | 599.503 | 572.9505 | 101.2 | 0.18 |
| pHILIC | 599.503 | 97.4217 | 99.9 | 0.18 |
| pHILIC | 599.503 | 130.9182 | 98.8 | 0.17 |
| pHILIC | 599.503 | 563.542 | 98.6 | 0.17 |
| pHILIC | 599.503 | 543.191 | 98.4 | 0.17 |
| pHILIC | 599.503 | 162.8746 | 98.1 | 0.17 |
| pHILIC | 599.503 | 338.8593 | 97.6 | 0.17 |
| pHILIC | 599.503 | 277.641 | 96.6 | 0.17 |
| pHILIC | 599.503 | 192.9851 | 96.6 | 0.17 |
| pHILIC | 599.503 | 553.6306 | 95.8 | 0.17 |
| pHILIC | 599.503 | 385.9775 | 95.3 | 0.17 |
| pHILIC | 599.503 | 477.6283 | 95 | 0.17 |
| pHILIC | 599.503 | 340.8581 | 94.8 | 0.17 |
| pHILIC | 599.503 | 560.6662 | 94.5 | 0.17 |
| pHILIC | 599.503 | 116.3417 | 94.4 | 0.17 |
| pHILIC | 599.503 | 288.2388 | 94.3 | 0.17 |
| pHILIC | 599.503 | 534.2556 | 94.2 | 0.17 |

| | | | | |
|---|---|---|---|---|
| pHILIC | 599.503 | 482.7986 | 93.8 | 0.16 |
| pHILIC | 599.503 | 91.1293 | 93.7 | 0.16 |
| pHILIC | 599.503 | 127.9278 | 93.3 | 0.16 |
| pHILIC | 599.503 | 309.4194 | 93.1 | 0.16 |
| pHILIC | 599.503 | 82.0466 | 92.6 | 0.16 |
| pHILIC | 599.503 | 409.318 | 92.6 | 0.16 |
| pHILIC | 599.503 | 372.5722 | 92.3 | 0.16 |
| pHILIC | 599.503 | 505.7446 | 92.1 | 0.16 |
| pHILIC | 599.503 | 133.4715 | 91.8 | 0.16 |
| pHILIC | 599.503 | 122.0539 | 91.6 | 0.16 |
| pHILIC | 599.503 | 520.095 | 91.6 | 0.16 |
| pHILIC | 599.503 | 152.1025 | 91.5 | 0.16 |
| pHILIC | 599.503 | 327.1973 | 91.2 | 0.16 |
| pHILIC | 599.503 | 616.5701 | 91 | 0.16 |
| pHILIC | 599.503 | 143.5682 | 90.9 | 0.16 |
| pHILIC | 599.503 | 94.7349 | 90.6 | 0.16 |
| pHILIC | 599.503 | 119.6012 | 90.5 | 0.16 |
| pHILIC | 599.503 | 356.0532 | 90.3 | 0.16 |
| pHILIC | 599.503 | 282.2602 | 90.2 | 0.16 |
| pHILIC | 599.503 | 189.98 | 89.8 | 0.16 |
| pHILIC | 599.503 | 249.0531 | 89.7 | 0.16 |
| pHILIC | 599.503 | 513.0142 | 89.5 | 0.16 |
| pHILIC | 599.503 | 146.6658 | 89.4 | 0.16 |
| pHILIC | 599.503 | 134.4321 | 89.4 | 0.16 |
| pHILIC | 599.503 | 94.4754 | 89.3 | 0.16 |
| pHILIC | 599.503 | 139.6057 | 89.1 | 0.16 |
| pHILIC | 599.503 | 255.9112 | 89.1 | 0.16 |
| pHILIC | 599.503 | 124.4925 | 88.9 | 0.16 |
| pHILIC | 599.503 | 292.8209 | 88.7 | 0.16 |
| pHILIC | 599.503 | 394.8818 | 88.5 | 0.16 |
| pHILIC | 599.503 | 222.3923 | 88.2 | 0.15 |
| pHILIC | 599.503 | 289.9928 | 88 | 0.15 |
| pHILIC | 599.503 | 175.8396 | 87.7 | 0.15 |
| pHILIC | 599.503 | 124.2663 | 87.6 | 0.15 |
| pHILIC | 599.503 | 584.3178 | 87.6 | 0.15 |
| pHILIC | 599.503 | 160.1377 | 87.5 | 0.15 |
| pHILIC | 599.503 | 154.231 | 87.4 | 0.15 |
| pHILIC | 599.503 | 75.9674 | 87.3 | 0.15 |
| pHILIC | 599.503 | 73.809 | 87 | 0.15 |
| pHILIC | 599.503 | 64.9665 | 86.9 | 0.15 |
| pHILIC | 599.503 | 238.9202 | 86.4 | 0.15 |
| pHILIC | 599.503 | 108.4524 | 86.1 | 0.15 |

| | | | | |
|---|---|---|---|---|
| pHILIC | 599.503 | 419.6443 | 85.9 | 0.15 |
| pHILIC | 599.503 | 85.7912 | 85.9 | 0.15 |
| pHILIC | 599.503 | 409.9856 | 85.7 | 0.15 |
| pHILIC | 599.503 | 200.0067 | 85.5 | 0.15 |
| pHILIC | 599.503 | 359.4967 | 85.4 | 0.15 |
| pHILIC | 599.503 | 236.4428 | 85.4 | 0.15 |
| pHILIC | 599.503 | 451.7623 | 85.1 | 0.15 |
| pHILIC | 599.503 | 240.1569 | 84.7 | 0.15 |
| pHILIC | 599.503 | 303.2545 | 84.7 | 0.15 |
| pHILIC | 599.503 | 93.5571 | 84.5 | 0.15 |
| pHILIC | 599.503 | 119.132 | 84 | 0.15 |
| pHILIC | 599.503 | 375.5962 | 83.9 | 0.15 |
| pHILIC | 599.503 | 248.2387 | 83.2 | 0.15 |
| pHILIC | 599.503 | 101.4233 | 83 | 0.15 |
| pHILIC | 599.503 | 321.2775 | 83 | 0.15 |
| pHILIC | 599.503 | 80.4338 | 82.8 | 0.15 |
| pHILIC | 599.503 | 124.4444 | 82.7 | 0.15 |
| pHILIC | 599.503 | 193.6133 | 82.6 | 0.14 |
| pHILIC | 599.503 | 107.2411 | 82.4 | 0.14 |
| pHILIC | 599.503 | 305.4627 | 82.3 | 0.14 |
| pHILIC | 599.503 | 113.6225 | 82.3 | 0.14 |
| pHILIC | 599.503 | 132.3282 | 82.1 | 0.14 |
| pHILIC | 599.503 | 78.0186 | 82.1 | 0.14 |
| pHILIC | 599.503 | 163.4219 | 81.6 | 0.14 |
| pHILIC | 599.503 | 77.1696 | 81.6 | 0.14 |
| pHILIC | 599.503 | 146.1825 | 81.4 | 0.14 |
| pHILIC | 599.503 | 125.0321 | 80.2 | 0.14 |
| pHILIC | 599.503 | 77.3014 | 80.2 | 0.14 |
| pHILIC | 599.503 | 552.2818 | 79.9 | 0.14 |
| pHILIC | 599.503 | 100.2127 | 79.5 | 0.14 |
| pHILIC | 599.503 | 304.9272 | 79.5 | 0.14 |
| pHILIC | 599.503 | 79.1002 | 79.4 | 0.14 |
| pHILIC | 599.503 | 149.548 | 79 | 0.14 |
| pHILIC | 599.503 | 245.2808 | 78.8 | 0.14 |
| pHILIC | 599.503 | 127.5044 | 78.4 | 0.14 |
| pHILIC | 599.503 | 67.0542 | 77.2 | 0.14 |
| pHILIC | 599.503 | 110.4117 | 76.9 | 0.14 |
| pHILIC | 599.503 | 94.1775 | 75 | 0.13 |
| pHILIC | 599.503 | 105.0696 | 74.3 | 0.13 |
| pHILIC | 599.503 | 63.3059 | 74.2 | 0.13 |
| pHILIC | 599.503 | 164.5668 | 73.9 | 0.13 |
| pHILIC | 599.503 | 226.5596 | 72.6 | 0.13 |

| | | | | |
|---|---|---|---|---|
| pHILIC | 599.503 | 81.6224 | 72.6 | 0.13 |
| pHILIC | 599.503 | 375.7846 | 63.9 | 0.11 |
| pHILIC | 599.503 | 64.5709 | 61.6 | 0.11 |
| pHILIC | 599.503 | 290.704 | 61.2 | 0.11 |
| pHILIC | 599.503 | 94.5759 | 60.8 | 0.11 |
| pHILIC | 599.503 | 120.9101 | 60.2 | 0.11 |
| pHILIC | 599.503 | 116.9129 | 59.3 | 0.1 |
| pHILIC | 599.503 | 509.1974 | 58.9 | 0.1 |
| pHILIC | 599.503 | 461.4127 | 58.7 | 0.1 |
| pHILIC | 599.503 | 306.9919 | 57.8 | 0.1 |
| pHILIC | 599.503 | 366.3914 | 56.6 | 0.1 |
| pHILIC | 599.503 | 78.5951 | 56.5 | 0.1 |
| pHILIC | 599.503 | 75.8402 | 56.3 | 0.1 |
| pHILIC | 599.503 | 66.0318 | 56.2 | 0.1 |
| pHILIC | 599.503 | 75.0628 | 56.2 | 0.1 |
| pHILIC | 599.503 | 119.5471 | 56 | 0.1 |
| pHILIC | 599.503 | 205.6109 | 55.9 | 0.1 |
| pHILIC | 599.503 | 565.7297 | 55.7 | 0.1 |
| pHILIC | 599.503 | 365.0833 | 55.7 | 0.1 |
| pHILIC | 599.503 | 236.9557 | 55.7 | 0.1 |
| pHILIC | 599.503 | 86.5678 | 55.7 | 0.1 |
| pHILIC | 599.503 | 496.178 | 55.6 | 0.1 |
| pHILIC | 599.503 | 281.1387 | 55.4 | 0.1 |
| pHILIC | 599.503 | 581.0292 | 55.4 | 0.1 |
| pHILIC | 599.503 | 442.9479 | 55.3 | 0.1 |
| pHILIC | 599.503 | 151.8648 | 55.2 | 0.1 |
| pHILIC | 599.503 | 145.0769 | 55.1 | 0.1 |
| pHILIC | 599.503 | 63.3231 | 55.1 | 0.1 |
| pHILIC | 599.503 | 192.2695 | 55.1 | 0.1 |
| pHILIC | 599.503 | 66.8218 | 54.8 | 0.1 |
| pHILIC | 599.503 | 83.8771 | 54.7 | 0.1 |
| pHILIC | 599.503 | 462.5287 | 54.6 | 0.1 |
| pHILIC | 599.503 | 289.5576 | 54.6 | 0.1 |
| pHILIC | 599.503 | 154.303 | 54.5 | 0.1 |
| pHILIC | 599.503 | 449.8581 | 54.4 | 0.1 |
| pHILIC | 599.503 | 466.5116 | 54.3 | 0.1 |
| pHILIC | 599.503 | 223.0408 | 54.3 | 0.1 |
| pHILIC | 599.503 | 249.2931 | 54.3 | 0.1 |
| pHILIC | 599.503 | 121.0111 | 54.2 | 0.1 |
| pHILIC | 599.503 | 199.5046 | 53.8 | 0.09 |
| pHILIC | 599.503 | 192.3022 | 53.8 | 0.09 |
| pHILIC | 599.503 | 117.4128 | 53.5 | 0.09 |

| | | | | |
|---|---|---|---|---|
| pHILIC | 599.503 | 543.553 | 53.2 | 0.09 |
| pHILIC | 599.503 | 208.1446 | 53.2 | 0.09 |
| pHILIC | 599.503 | 300.2692 | 53.1 | 0.09 |
| pHILIC | 599.503 | 126.2342 | 53 | 0.09 |
| pHILIC | 599.503 | 492.5283 | 52.9 | 0.09 |
| pHILIC | 599.503 | 159.3784 | 52.8 | 0.09 |
| pHILIC | 599.503 | 156.2269 | 52.8 | 0.09 |
| pHILIC | 599.503 | 550.9049 | 52.6 | 0.09 |
| pHILIC | 599.503 | 72.4314 | 52.4 | 0.09 |
| pHILIC | 599.503 | 488.4653 | 52.4 | 0.09 |
| pHILIC | 599.503 | 95.4451 | 52.3 | 0.09 |
| pHILIC | 599.503 | 117.6285 | 52.3 | 0.09 |
| pHILIC | 599.503 | 319.1499 | 52.3 | 0.09 |
| pHILIC | 599.503 | 426.555 | 52.2 | 0.09 |
| pHILIC | 599.503 | 440.3318 | 52.2 | 0.09 |
| pHILIC | 599.503 | 94.5948 | 52.2 | 0.09 |
| pHILIC | 599.503 | 107.4646 | 52 | 0.09 |
| pHILIC | 599.503 | 76.7972 | 52 | 0.09 |
| pHILIC | 599.503 | 404.9878 | 52 | 0.09 |
| pHILIC | 599.503 | 120.0564 | 51.9 | 0.09 |
| pHILIC | 599.503 | 100.6728 | 51.9 | 0.09 |
| pHILIC | 599.503 | 91.0276 | 51.8 | 0.09 |
| pHILIC | 599.503 | 86.6972 | 51.8 | 0.09 |
| pHILIC | 599.503 | 290.2737 | 51.8 | 0.09 |
| pHILIC | 599.503 | 261.5679 | 51.7 | 0.09 |
| pHILIC | 599.503 | 109.0999 | 51.7 | 0.09 |
| pHILIC | 599.503 | 470.6907 | 51.6 | 0.09 |
| pHILIC | 599.503 | 227.867 | 51.5 | 0.09 |
| pHILIC | 599.503 | 148.3104 | 51.4 | 0.09 |
| pHILIC | 599.503 | 471.0196 | 51.4 | 0.09 |
| pHILIC | 599.503 | 110.6263 | 51.4 | 0.09 |
| pHILIC | 599.503 | 95.4617 | 51.4 | 0.09 |
| pHILIC | 599.503 | 269.4898 | 51.4 | 0.09 |
| pHILIC | 599.503 | 487.1201 | 51.4 | 0.09 |
| pHILIC | 599.503 | 64.7251 | 51.3 | 0.09 |
| pHILIC | 599.503 | 151.756 | 51.3 | 0.09 |
| pHILIC | 599.503 | 313.0288 | 51.2 | 0.09 |
| pHILIC | 599.503 | 611.1641 | 51.2 | 0.09 |
| pHILIC | 599.503 | 283.9464 | 51.2 | 0.09 |
| pHILIC | 599.503 | 102.0664 | 51.2 | 0.09 |
| pHILIC | 599.503 | 304.7969 | 51.1 | 0.09 |
| pHILIC | 599.503 | 92.0569 | 51.1 | 0.09 |

| | | | | |
|---|---|---|---|---|
| pHILIC | 599.503 | 243.6701 | 51 | 0.09 |
| pHILIC | 599.503 | 77.4839 | 51 | 0.09 |
| pHILIC | 599.503 | 90.1891 | 51 | 0.09 |
| pHILIC | 599.503 | 339.3123 | 51 | 0.09 |
| pHILIC | 599.503 | 91.4857 | 50.9 | 0.09 |
| pHILIC | 599.503 | 284.4333 | 50.8 | 0.09 |
| pHILIC | 599.503 | 263.0805 | 50.6 | 0.09 |
| pHILIC | 599.503 | 94.1128 | 50.6 | 0.09 |
| pHILIC | 599.503 | 69.3083 | 50.6 | 0.09 |
| pHILIC | 599.503 | 213.4281 | 50.5 | 0.09 |
| pHILIC | 599.503 | 277.2855 | 50.4 | 0.09 |
| pHILIC | 599.503 | 74.9554 | 50.4 | 0.09 |
| pHILIC | 599.503 | 248.6402 | 50.4 | 0.09 |
| pHILIC | 599.503 | 68.2303 | 50.4 | 0.09 |
| pHILIC | 599.503 | 72.3051 | 50.4 | 0.09 |
| pHILIC | 599.503 | 71.5935 | 50.3 | 0.09 |
| pHILIC | 599.503 | 358.2537 | 50.3 | 0.09 |
| pHILIC | 599.503 | 74.5661 | 50.3 | 0.09 |
| pHILIC | 599.503 | 270.3714 | 50.2 | 0.09 |
| pHILIC | 599.503 | 371.4981 | 50.2 | 0.09 |
| pHILIC | 599.503 | 67.3165 | 50.1 | 0.09 |
| pHILIC | 599.503 | 204.5407 | 50 | 0.09 |
| pHILIC | 599.503 | 416.9839 | 50 | 0.09 |
| pHILIC | 599.503 | 109.3136 | 49.9 | 0.09 |
| pHILIC | 599.503 | 180.0736 | 49.9 | 0.09 |
| pHILIC | 599.503 | 351.6864 | 49.8 | 0.09 |
| pHILIC | 599.503 | 170.506 | 49.8 | 0.09 |
| pHILIC | 599.503 | 469.825 | 49.8 | 0.09 |
| pHILIC | 599.503 | 119.3415 | 49.7 | 0.09 |
| pHILIC | 599.503 | 124.5293 | 49.7 | 0.09 |
| pHILIC | 599.503 | 175.2813 | 49.7 | 0.09 |
| pHILIC | 599.503 | 222.4783 | 49.7 | 0.09 |
| pHILIC | 599.503 | 76.2773 | 49.7 | 0.09 |
| pHILIC | 599.503 | 109.1249 | 49.7 | 0.09 |
| pHILIC | 599.503 | 579.1895 | 49.6 | 0.09 |
| pHILIC | 599.503 | 117.7506 | 49.6 | 0.09 |
| pHILIC | 599.503 | 160.806 | 49.6 | 0.09 |
| pHILIC | 599.503 | 212.1649 | 49.5 | 0.09 |
| pHILIC | 599.503 | 97.6387 | 49.5 | 0.09 |
| pHILIC | 599.503 | 128.5617 | 49.5 | 0.09 |
| pHILIC | 599.503 | 87.1076 | 49.4 | 0.09 |
| pHILIC | 599.503 | 95.0168 | 49.4 | 0.09 |

| | | | | |
|---|---|---|---|---|
| pHILIC | 599.503 | 173.6618 | 49.4 | 0.09 |
| pHILIC | 599.503 | 254.7576 | 49.4 | 0.09 |
| pHILIC | 599.503 | 202.1947 | 49.4 | 0.09 |
| pHILIC | 599.503 | 67.7868 | 49.3 | 0.09 |
| pHILIC | 599.503 | 443.3999 | 49.3 | 0.09 |
| pHILIC | 599.503 | 382.4733 | 49.2 | 0.09 |
| pHILIC | 599.503 | 161.5192 | 49.2 | 0.09 |
| pHILIC | 599.503 | 105.7406 | 49.2 | 0.09 |
| pHILIC | 599.503 | 396.413 | 49.1 | 0.09 |
| pHILIC | 599.503 | 281.6068 | 49.1 | 0.09 |
| pHILIC | 599.503 | 65.5779 | 49.1 | 0.09 |
| pHILIC | 599.503 | 97.5796 | 49.1 | 0.09 |
| pHILIC | 599.503 | 613.5195 | 49.1 | 0.09 |
| pHILIC | 599.503 | 222.5458 | 49 | 0.09 |
| pHILIC | 599.503 | 112.7775 | 49 | 0.09 |
| pHILIC | 599.503 | 129.5572 | 49 | 0.09 |
| pHILIC | 599.503 | 75.6307 | 49 | 0.09 |
| pHILIC | 599.503 | 169.6728 | 49 | 0.09 |
| pHILIC | 599.503 | 74.1265 | 49 | 0.09 |
| pHILIC | 599.503 | 229.287 | 49 | 0.09 |
| pHILIC | 599.503 | 96.7823 | 49 | 0.09 |
| pHILIC | 599.503 | 534.5764 | 48.9 | 0.09 |
| pHILIC | 599.503 | 537.1242 | 48.9 | 0.09 |
| pHILIC | 599.503 | 481.1697 | 48.9 | 0.09 |
| pHILIC | 599.503 | 153.9087 | 48.8 | 0.09 |
| pHILIC | 599.503 | 71.8067 | 48.8 | 0.09 |
| pHILIC | 599.503 | 472.5497 | 48.8 | 0.09 |
| pHILIC | 599.503 | 80.2404 | 48.8 | 0.09 |
| pHILIC | 599.503 | 66.397 | 48.8 | 0.09 |
| pHILIC | 599.503 | 66.2032 | 48.8 | 0.09 |
| pHILIC | 599.503 | 63.8497 | 48.8 | 0.09 |
| pHILIC | 599.503 | 313.8063 | 48.8 | 0.09 |
| pHILIC | 599.503 | 140.4119 | 48.8 | 0.09 |
| pHILIC | 599.503 | 285.4876 | 48.7 | 0.09 |
| pHILIC | 599.503 | 255.0977 | 48.7 | 0.09 |
| pHILIC | 599.503 | 268.2356 | 48.7 | 0.09 |
| pHILIC | 599.503 | 86.2805 | 48.7 | 0.09 |
| pHILIC | 599.503 | 74.6412 | 48.7 | 0.09 |
| pHILIC | 599.503 | 91.9887 | 48.7 | 0.09 |
| pHILIC | 599.503 | 392.657 | 48.7 | 0.09 |
| pHILIC | 599.503 | 488.1958 | 48.6 | 0.09 |
| pHILIC | 599.503 | 176.8652 | 48.6 | 0.09 |

| | | | | |
|---|---|---|---|---|
| pHILIC | 599.503 | 272.7309 | 48.6 | 0.09 |
| pHILIC | 599.503 | 90.5046 | 48.6 | 0.09 |
| pHILIC | 599.503 | 95.0272 | 48.6 | 0.09 |
| pHILIC | 599.503 | 132.6186 | 48.6 | 0.09 |
| pHILIC | 599.503 | 187.4183 | 48.6 | 0.09 |
| pHILIC | 599.503 | 100.5533 | 48.6 | 0.09 |
| pHILIC | 599.503 | 299.0494 | 48.5 | 0.09 |
| pHILIC | 599.503 | 321.3343 | 48.5 | 0.09 |
| pHILIC | 599.503 | 124.3417 | 48.5 | 0.09 |
| pHILIC | 599.503 | 94.1626 | 48.5 | 0.09 |
| pHILIC | 599.503 | 65.3169 | 48.4 | 0.08 |
| pHILIC | 599.503 | 380.9537 | 48.4 | 0.08 |
| pHILIC | 599.503 | 109.7666 | 48.4 | 0.08 |
| pHILIC | 599.503 | 419.4544 | 48.4 | 0.08 |
| pHILIC | 599.503 | 193.4949 | 48.4 | 0.08 |
| pHILIC | 599.503 | 93.614 | 48.3 | 0.08 |
| pHILIC | 599.503 | 265.3398 | 48.3 | 0.08 |
| pHILIC | 599.503 | 162.1234 | 48.3 | 0.08 |
| pHILIC | 599.503 | 101.8752 | 48.3 | 0.08 |
| pHILIC | 599.503 | 594.4434 | 48.3 | 0.08 |
| pHILIC | 599.503 | 280.4912 | 48.2 | 0.08 |
| pHILIC | 599.503 | 70.0101 | 48.2 | 0.08 |
| pHILIC | 599.503 | 135.9041 | 48.2 | 0.08 |
| pHILIC | 599.503 | 90.2436 | 48.2 | 0.08 |
| pHILIC | 599.503 | 76.2391 | 48.2 | 0.08 |
| pHILIC | 599.503 | 95.9616 | 48.2 | 0.08 |
| pHILIC | 599.503 | 72.7807 | 48.1 | 0.08 |
| pHILIC | 599.503 | 67.9457 | 48.1 | 0.08 |
| pHILIC | 599.503 | 96.01 | 48.1 | 0.08 |
| pHILIC | 599.503 | 167.7648 | 48.1 | 0.08 |
| pHILIC | 599.503 | 142.1233 | 48.1 | 0.08 |
| pHILIC | 599.503 | 122.3779 | 48.1 | 0.08 |
| pHILIC | 599.503 | 629.0496 | 48.1 | 0.08 |
| pHILIC | 599.503 | 198.7703 | 48.1 | 0.08 |
| pHILIC | 599.503 | 94.7927 | 48.1 | 0.08 |
| pHILIC | 599.503 | 550.7405 | 48.1 | 0.08 |
| pHILIC | 599.503 | 184.8274 | 48 | 0.08 |
| pHILIC | 599.503 | 109.5283 | 48 | 0.08 |
| pHILIC | 599.503 | 213.7284 | 48 | 0.08 |
| pHILIC | 599.503 | 66.9073 | 48 | 0.08 |
| pHILIC | 599.503 | 92.948 | 48 | 0.08 |
| pHILIC | 599.503 | 64.6548 | 48 | 0.08 |

| | | | | |
|---|---|---|---|---|
| pHILIC | 599.503 | 203.4587 | 48 | 0.08 |
| pHILIC | 599.503 | 92.2138 | 47.9 | 0.08 |
| pHILIC | 599.503 | 87.2134 | 47.9 | 0.08 |
| pHILIC | 599.503 | 94.4037 | 47.9 | 0.08 |
| pHILIC | 599.503 | 253.0121 | 47.9 | 0.08 |
| pHILIC | 599.503 | 216.702 | 47.8 | 0.08 |
| pHILIC | 599.503 | 90.5722 | 47.8 | 0.08 |
| pHILIC | 599.503 | 126.4287 | 47.8 | 0.08 |
| pHILIC | 599.503 | 75.1291 | 47.8 | 0.08 |
| pHILIC | 599.503 | 140.8064 | 47.8 | 0.08 |
| pHILIC | 599.503 | 596.1166 | 47.8 | 0.08 |
| pHILIC | 599.503 | 97.6277 | 47.8 | 0.08 |
| pHILIC | 599.503 | 134.2905 | 47.8 | 0.08 |
| pHILIC | 599.503 | 467.1062 | 47.7 | 0.08 |
| pHILIC | 599.503 | 268.095 | 47.7 | 0.08 |
| pHILIC | 599.503 | 131.2301 | 47.7 | 0.08 |
| pHILIC | 599.503 | 151.9867 | 47.7 | 0.08 |
| pHILIC | 599.503 | 77.4082 | 47.7 | 0.08 |
| pHILIC | 599.503 | 114.304 | 47.7 | 0.08 |
| pHILIC | 599.503 | 65.6889 | 47.7 | 0.08 |
| pHILIC | 599.503 | 109.4247 | 47.7 | 0.08 |
| pHILIC | 599.503 | 214.209 | 47.6 | 0.08 |
| pHILIC | 599.503 | 169.0185 | 47.6 | 0.08 |
| pHILIC | 599.503 | 519.6183 | 47.6 | 0.08 |
| pHILIC | 599.503 | 108.4759 | 47.6 | 0.08 |
| pHILIC | 599.503 | 66.3343 | 47.6 | 0.08 |
| pHILIC | 599.503 | 488.3545 | 47.5 | 0.08 |
| pHILIC | 599.503 | 228.3576 | 47.5 | 0.08 |
| pHILIC | 599.503 | 342.2873 | 47.5 | 0.08 |
| pHILIC | 599.503 | 99.9598 | 47.5 | 0.08 |
| pHILIC | 599.503 | 112.1143 | 47.5 | 0.08 |
| pHILIC | 599.503 | 66.7344 | 47.5 | 0.08 |
| pHILIC | 599.503 | 188.3777 | 47.5 | 0.08 |
| pHILIC | 599.503 | 525.726 | 47.4 | 0.08 |
| pHILIC | 599.503 | 86.8483 | 47.4 | 0.08 |
| pHILIC | 599.503 | 369.9775 | 47.4 | 0.08 |
| pHILIC | 599.503 | 108.555 | 47.4 | 0.08 |
| pHILIC | 599.503 | 213.2958 | 47.4 | 0.08 |
| pHILIC | 599.503 | 385.7353 | 47.4 | 0.08 |
| pHILIC | 599.503 | 465.3676 | 47.3 | 0.08 |
| pHILIC | 599.503 | 114.2518 | 47.3 | 0.08 |
| pHILIC | 599.503 | 582.2206 | 47.3 | 0.08 |

| | | | | |
|---|---|---|---|---|
| pHILIC | 599.503 | 438.2435 | 47.3 | 0.08 |
| pHILIC | 599.503 | 128.663 | 47.3 | 0.08 |
| pHILIC | 599.503 | 77.8269 | 47.3 | 0.08 |
| pHILIC | 599.503 | 454.6989 | 47.3 | 0.08 |
| pHILIC | 599.503 | 308.6001 | 47.3 | 0.08 |
| pHILIC | 599.503 | 90.6626 | 47.3 | 0.08 |
| pHILIC | 599.503 | 229.2071 | 47.2 | 0.08 |
| pHILIC | 599.503 | 216.5482 | 47.2 | 0.08 |
| pHILIC | 599.503 | 203.0804 | 47.2 | 0.08 |
| pHILIC | 599.503 | 309.8513 | 47.2 | 0.08 |
| pHILIC | 599.503 | 161.133 | 47.2 | 0.08 |
| pHILIC | 599.503 | 94.1419 | 47.2 | 0.08 |
| pHILIC | 599.503 | 217.6445 | 47.2 | 0.08 |
| pHILIC | 599.503 | 98.0942 | 47.1 | 0.08 |
| pHILIC | 599.503 | 70.0487 | 47.1 | 0.08 |
| pHILIC | 599.503 | 196.7678 | 47.1 | 0.08 |
| pHILIC | 599.503 | 308.2944 | 47.1 | 0.08 |
| pHILIC | 599.503 | 81.8344 | 47.1 | 0.08 |
| pHILIC | 599.503 | 126.1391 | 47.1 | 0.08 |
| pHILIC | 599.503 | 117.2134 | 47 | 0.08 |
| pHILIC | 599.503 | 67.3016 | 47 | 0.08 |
| pHILIC | 599.503 | 149.2516 | 47 | 0.08 |
| pHILIC | 599.503 | 67.9342 | 47 | 0.08 |
| pHILIC | 599.503 | 361.7247 | 47 | 0.08 |
| pHILIC | 599.503 | 147.8287 | 47 | 0.08 |
| pHILIC | 599.503 | 618.579 | 47 | 0.08 |
| pHILIC | 599.503 | 266.053 | 47 | 0.08 |
| pHILIC | 599.503 | 271.0107 | 47 | 0.08 |
| pHILIC | 599.503 | 138.1968 | 46.9 | 0.08 |
| pHILIC | 599.503 | 329.5083 | 46.9 | 0.08 |
| pHILIC | 599.503 | 379.03 | 46.9 | 0.08 |
| pHILIC | 599.503 | 78.8629 | 46.9 | 0.08 |
| pHILIC | 599.503 | 148.8628 | 46.8 | 0.08 |
| pHILIC | 599.503 | 164.5346 | 46.8 | 0.08 |
| pHILIC | 599.503 | 176.5931 | 46.8 | 0.08 |
| pHILIC | 599.503 | 158.2247 | 46.8 | 0.08 |
| pHILIC | 599.503 | 64.8518 | 46.7 | 0.08 |
| pHILIC | 599.503 | 552.9318 | 46.7 | 0.08 |
| pHILIC | 599.503 | 238.1144 | 46.7 | 0.08 |
| pHILIC | 599.503 | 136.4156 | 46.7 | 0.08 |
| pHILIC | 599.503 | 214.3879 | 46.7 | 0.08 |
| pHILIC | 599.503 | 128.1888 | 46.7 | 0.08 |

| | | | | |
|---|---|---|---|---|
| pHILIC | 599.503 | 91.5311 | 46.7 | 0.08 |
| pHILIC | 599.503 | 273.6783 | 46.7 | 0.08 |
| pHILIC | 599.503 | 288.8875 | 46.7 | 0.08 |
| pHILIC | 599.503 | 420.6741 | 46.7 | 0.08 |
| pHILIC | 599.503 | 102.599 | 46.6 | 0.08 |
| pHILIC | 599.503 | 406.7298 | 46.6 | 0.08 |
| pHILIC | 599.503 | 601.9999 | 46.6 | 0.08 |
| pHILIC | 599.503 | 102.3918 | 46.6 | 0.08 |
| pHILIC | 599.503 | 141.9795 | 46.6 | 0.08 |
| pHILIC | 599.503 | 134.021 | 46.6 | 0.08 |
| pHILIC | 599.503 | 158.3451 | 46.6 | 0.08 |
| pHILIC | 599.503 | 115.8948 | 46.6 | 0.08 |
| pHILIC | 599.503 | 242.0604 | 46.6 | 0.08 |
| pHILIC | 599.503 | 552.4525 | 46.6 | 0.08 |
| pHILIC | 599.503 | 99.5849 | 46.6 | 0.08 |
| pHILIC | 599.503 | 140.1026 | 46.6 | 0.08 |
| pHILIC | 599.503 | 195.0562 | 46.6 | 0.08 |
| pHILIC | 599.503 | 174.9233 | 46.6 | 0.08 |
| pHILIC | 599.503 | 131.3187 | 46.6 | 0.08 |
| pHILIC | 599.503 | 285.7885 | 46.6 | 0.08 |
| pHILIC | 599.503 | 97.1853 | 46.5 | 0.08 |
| pHILIC | 599.503 | 266.3228 | 46.5 | 0.08 |
| pHILIC | 599.503 | 285.405 | 46.5 | 0.08 |
| pHILIC | 599.503 | 111.1618 | 46.5 | 0.08 |
| pHILIC | 599.503 | 130.2561 | 46.5 | 0.08 |
| pHILIC | 599.503 | 79.9912 | 46.5 | 0.08 |
| pHILIC | 599.503 | 78.1336 | 46.4 | 0.08 |
| pHILIC | 599.503 | 161.7015 | 46.4 | 0.08 |
| pHILIC | 599.503 | 92.7596 | 46.4 | 0.08 |
| pHILIC | 599.503 | 88.6173 | 46.4 | 0.08 |
| pHILIC | 599.503 | 84.3712 | 46.4 | 0.08 |
| pHILIC | 599.503 | 485.8201 | 46.4 | 0.08 |
| pHILIC | 599.503 | 241.8126 | 46.4 | 0.08 |
| pHILIC | 599.503 | 193.8295 | 46.4 | 0.08 |
| pHILIC | 599.503 | 271.9043 | 46.3 | 0.08 |
| pHILIC | 599.503 | 66.6516 | 46.3 | 0.08 |
| pHILIC | 599.503 | 85.8102 | 46.3 | 0.08 |
| pHILIC | 599.503 | 137.611 | 46.3 | 0.08 |
| pHILIC | 599.503 | 541.2097 | 46.3 | 0.08 |
| pHILIC | 599.503 | 596.7564 | 46.3 | 0.08 |
| pHILIC | 599.503 | 423.9999 | 46.3 | 0.08 |
| pHILIC | 599.503 | 545.2195 | 46.3 | 0.08 |

| | | | | |
|---|---|---|---|---|
| pHILIC | 599.503 | 365.3272 | 46.2 | 0.08 |
| pHILIC | 599.503 | 320.7294 | 46.2 | 0.08 |
| pHILIC | 599.503 | 266.8362 | 46.2 | 0.08 |
| pHILIC | 599.503 | 259.0974 | 46.2 | 0.08 |
| pHILIC | 599.503 | 287.8354 | 46.2 | 0.08 |
| pHILIC | 599.503 | 484.7459 | 46.2 | 0.08 |
| pHILIC | 599.503 | 85.0581 | 46.2 | 0.08 |
| pHILIC | 599.503 | 179.3916 | 46.2 | 0.08 |
| pHILIC | 599.503 | 228.4512 | 46.2 | 0.08 |
| pHILIC | 599.503 | 163.8991 | 46.2 | 0.08 |
| pHILIC | 599.503 | 338.4666 | 46.2 | 0.08 |
| pHILIC | 599.503 | 95.2476 | 46.2 | 0.08 |
| pHILIC | 599.503 | 77.2396 | 46.1 | 0.08 |
| pHILIC | 599.503 | 109.8435 | 46.1 | 0.08 |
| pHILIC | 599.503 | 462.1424 | 46.1 | 0.08 |
| pHILIC | 599.503 | 326.8409 | 46.1 | 0.08 |
| pHILIC | 599.503 | 213.3304 | 46.1 | 0.08 |
| pHILIC | 599.503 | 75.7152 | 46.1 | 0.08 |
| pHILIC | 599.503 | 373.7881 | 46.1 | 0.08 |
| pHILIC | 599.503 | 168.9377 | 46.1 | 0.08 |
| pHILIC | 599.503 | 171.3948 | 46.1 | 0.08 |
| pHILIC | 599.503 | 67.9181 | 46.1 | 0.08 |
| pHILIC | 599.503 | 142.1041 | 46.1 | 0.08 |
| pHILIC | 599.503 | 91.973 | 46.1 | 0.08 |
| pHILIC | 599.503 | 339.5306 | 46 | 0.08 |
| pHILIC | 599.503 | 73.3903 | 46 | 0.08 |
| pHILIC | 599.503 | 602.4759 | 46 | 0.08 |
| pHILIC | 599.503 | 258.9166 | 46 | 0.08 |
| pHILIC | 599.503 | 147.9631 | 46 | 0.08 |
| pHILIC | 599.503 | 468.0227 | 46 | 0.08 |
| pHILIC | 599.503 | 147.157 | 46 | 0.08 |
| pHILIC | 599.503 | 77.755 | 46 | 0.08 |
| pHILIC | 599.503 | 93.9071 | 46 | 0.08 |
| pHILIC | 599.503 | 80.5998 | 46 | 0.08 |
| pHILIC | 599.503 | 93.9204 | 45.9 | 0.08 |
| pHILIC | 599.503 | 181.3762 | 45.9 | 0.08 |
| pHILIC | 599.503 | 463.854 | 45.9 | 0.08 |
| pHILIC | 599.503 | 244.4799 | 45.9 | 0.08 |
| pHILIC | 599.503 | 370.6141 | 45.9 | 0.08 |
| pHILIC | 599.503 | 113.4842 | 45.9 | 0.08 |
| pHILIC | 599.503 | 72.0558 | 45.9 | 0.08 |
| pHILIC | 599.503 | 84.016 | 45.9 | 0.08 |

| | | | | |
|---|---|---|---|---|
| pHILIC | 599.503 | 237.9676 | 45.8 | 0.08 |
| pHILIC | 599.503 | 491.5833 | 45.8 | 0.08 |
| pHILIC | 599.503 | 103.9125 | 45.8 | 0.08 |
| pHILIC | 599.503 | 173.107 | 45.8 | 0.08 |
| pHILIC | 599.503 | 143.8822 | 45.8 | 0.08 |
| pHILIC | 599.503 | 180.4397 | 45.8 | 0.08 |
| pHILIC | 599.503 | 67.9433 | 45.8 | 0.08 |
| pHILIC | 599.503 | 245.874 | 45.8 | 0.08 |
| pHILIC | 599.503 | 242.915 | 45.8 | 0.08 |
| pHILIC | 599.503 | 263.9608 | 45.8 | 0.08 |
| pHILIC | 599.503 | 346.1392 | 45.8 | 0.08 |
| pHILIC | 599.503 | 340.0104 | 45.8 | 0.08 |
| pHILIC | 599.503 | 145.2713 | 45.8 | 0.08 |
| pHILIC | 599.503 | 269.9251 | 45.8 | 0.08 |
| pHILIC | 599.503 | 89.0513 | 45.8 | 0.08 |
| pHILIC | 599.503 | 85.4213 | 45.7 | 0.08 |
| pHILIC | 599.503 | 114.5834 | 45.7 | 0.08 |
| pHILIC | 599.503 | 392.0893 | 45.6 | 0.08 |
| pHILIC | 599.503 | 281.8339 | 45.6 | 0.08 |
| pHILIC | 599.503 | 590.3677 | 45.6 | 0.08 |
| pHILIC | 599.503 | 221.8655 | 45.6 | 0.08 |
| pHILIC | 599.503 | 115.4778 | 45.6 | 0.08 |
| pHILIC | 599.503 | 71.1171 | 45.6 | 0.08 |
| pHILIC | 599.503 | 228.6105 | 45.6 | 0.08 |
| pHILIC | 599.503 | 365.7137 | 45.6 | 0.08 |
| pHILIC | 599.503 | 235.2414 | 45.5 | 0.08 |
| pHILIC | 599.503 | 76.9706 | 45.5 | 0.08 |
| pHILIC | 599.503 | 74.1413 | 45.5 | 0.08 |
| pHILIC | 599.503 | 219.4754 | 45.5 | 0.08 |
| pHILIC | 599.503 | 87.0222 | 45.5 | 0.08 |
| pHILIC | 599.503 | 464.5611 | 45.5 | 0.08 |
| pHILIC | 599.503 | 121.9914 | 45.5 | 0.08 |
| pHILIC | 599.503 | 278.3941 | 45.5 | 0.08 |
| pHILIC | 599.503 | 117.4022 | 45.5 | 0.08 |
| pHILIC | 599.503 | 174.9458 | 45.4 | 0.08 |
| pHILIC | 599.503 | 190.5081 | 45.4 | 0.08 |
| pHILIC | 599.503 | 199.7632 | 45.4 | 0.08 |
| pHILIC | 599.503 | 159.8062 | 45.4 | 0.08 |
| pHILIC | 599.503 | 185.2007 | 45.4 | 0.08 |
| pHILIC | 599.503 | 117.2229 | 45.4 | 0.08 |
| pHILIC | 599.503 | 88.2103 | 45.4 | 0.08 |
| pHILIC | 599.503 | 122.6659 | 45.4 | 0.08 |

| | | | | |
|---|---|---|---|---|
| pHILIC | 599.503 | 85.0196 | 45.4 | 0.08 |
| pHILIC | 599.503 | 282.325 | 45.3 | 0.08 |
| pHILIC | 599.503 | 415.9981 | 45.3 | 0.08 |
| pHILIC | 599.503 | 94.8812 | 45.3 | 0.08 |
| pHILIC | 599.503 | 64.0314 | 45.3 | 0.08 |
| pHILIC | 599.503 | 81.3565 | 45.3 | 0.08 |
| pHILIC | 599.503 | 259.7654 | 45.3 | 0.08 |
| pHILIC | 599.503 | 178.0985 | 45.3 | 0.08 |
| pHILIC | 599.503 | 112.3675 | 45.3 | 0.08 |
| pHILIC | 599.503 | 417.2291 | 45.3 | 0.08 |
| pHILIC | 599.503 | 168.7447 | 45.3 | 0.08 |
| pHILIC | 599.503 | 107.1715 | 45.3 | 0.08 |
| pHILIC | 599.503 | 348.5868 | 45.3 | 0.08 |
| pHILIC | 599.503 | 435.2088 | 45.3 | 0.08 |
| pHILIC | 599.503 | 355.5636 | 45.2 | 0.08 |
| pHILIC | 599.503 | 120.1487 | 45.2 | 0.08 |
| pHILIC | 599.503 | 209.4343 | 45.2 | 0.08 |
| pHILIC | 599.503 | 126.6097 | 45.2 | 0.08 |
| pHILIC | 599.503 | 234.3403 | 45.2 | 0.08 |
| pHILIC | 599.503 | 94.2181 | 45.2 | 0.08 |
| pHILIC | 599.503 | 139.1341 | 45.2 | 0.08 |
| pHILIC | 599.503 | 390.8633 | 45.2 | 0.08 |
| pHILIC | 599.503 | 87.761 | 45.2 | 0.08 |
| pHILIC | 599.503 | 298.3492 | 45.2 | 0.08 |
| pHILIC | 599.503 | 434.1971 | 45.2 | 0.08 |
| pHILIC | 599.503 | 130.0575 | 45.2 | 0.08 |
| pHILIC | 599.503 | 346.6961 | 45.2 | 0.08 |
| pHILIC | 599.503 | 572.0923 | 45.2 | 0.08 |
| pHILIC | 599.503 | 145.6185 | 45.2 | 0.08 |
| pHILIC | 599.503 | 468.3239 | 45.2 | 0.08 |
| pHILIC | 599.503 | 288.741 | 45.1 | 0.08 |
| pHILIC | 599.503 | 81.4693 | 45.1 | 0.08 |
| pHILIC | 599.503 | 151.9428 | 45.1 | 0.08 |
| pHILIC | 599.503 | 354.1612 | 45.1 | 0.08 |
| pHILIC | 599.503 | 80.8386 | 45.1 | 0.08 |
| pHILIC | 599.503 | 455.2891 | 45.1 | 0.08 |
| pHILIC | 599.503 | 91.5021 | 45.1 | 0.08 |
| pHILIC | 599.503 | 79.5609 | 45.1 | 0.08 |
| pHILIC | 599.503 | 192.5161 | 45.1 | 0.08 |
| pHILIC | 599.503 | 250.8531 | 45.1 | 0.08 |
| pHILIC | 599.503 | 73.7361 | 45.1 | 0.08 |
| pHILIC | 599.503 | 608.6509 | 45 | 0.08 |

| | | | | |
|---|---|---|---|---|
| pHILIC | 599.503 | 408.9674 | 45 | 0.08 |
| pHILIC | 599.503 | 165.2707 | 45 | 0.08 |
| pHILIC | 599.503 | 141.7708 | 45 | 0.08 |
| pHILIC | 599.503 | 89.6457 | 45 | 0.08 |
| pHILIC | 599.503 | 267.8893 | 45 | 0.08 |
| pHILIC | 599.503 | 64.2487 | 45 | 0.08 |
| pHILIC | 599.503 | 546.7181 | 45 | 0.08 |
| pHILIC | 599.503 | 119.514 | 45 | 0.08 |
| pHILIC | 599.503 | 450.836 | 45 | 0.08 |
| pHILIC | 599.503 | 286.0811 | 45 | 0.08 |
| pHILIC | 599.503 | 80.2034 | 44.9 | 0.08 |
| pHILIC | 599.503 | 382.9341 | 44.9 | 0.08 |
| pHILIC | 599.503 | 224.6185 | 44.9 | 0.08 |
| pHILIC | 599.503 | 383.1591 | 44.9 | 0.08 |
| pHILIC | 599.503 | 92.721 | 44.9 | 0.08 |
| pHILIC | 599.503 | 96.8404 | 44.9 | 0.08 |
| pHILIC | 599.503 | 250.5579 | 44.9 | 0.08 |
| pHILIC | 599.503 | 121.3845 | 44.9 | 0.08 |
| pHILIC | 599.503 | 169.5212 | 44.9 | 0.08 |
| pHILIC | 599.503 | 273.2275 | 44.9 | 0.08 |
| pHILIC | 599.503 | 187.3394 | 44.9 | 0.08 |
| pHILIC | 599.503 | 100.5983 | 44.9 | 0.08 |
| pHILIC | 599.503 | 251.3107 | 44.9 | 0.08 |
| pHILIC | 599.503 | 151.4432 | 44.9 | 0.08 |
| pHILIC | 599.503 | 74.4333 | 44.8 | 0.08 |
| pHILIC | 599.503 | 110.7447 | 44.8 | 0.08 |
| pHILIC | 599.503 | 478.461 | 44.8 | 0.08 |
| pHILIC | 599.503 | 105.2495 | 44.8 | 0.08 |
| pHILIC | 599.503 | 571.077 | 44.8 | 0.08 |
| pHILIC | 599.503 | 133.6578 | 44.8 | 0.08 |
| pHILIC | 599.503 | 399.2413 | 44.8 | 0.08 |
| pHILIC | 599.503 | 96.2229 | 44.8 | 0.08 |
| pHILIC | 599.503 | 107.6299 | 44.8 | 0.08 |
| pHILIC | 599.503 | 75.2335 | 44.8 | 0.08 |
| pHILIC | 599.503 | 149.0648 | 44.8 | 0.08 |
| pHILIC | 599.503 | 359.3742 | 44.8 | 0.08 |
| pHILIC | 599.503 | 232.2475 | 44.7 | 0.08 |
| pHILIC | 599.503 | 406.2753 | 44.7 | 0.08 |
| pHILIC | 599.503 | 424.8687 | 44.7 | 0.08 |
| pHILIC | 599.503 | 386.5208 | 44.7 | 0.08 |
| pHILIC | 599.503 | 519.8404 | 44.7 | 0.08 |
| pHILIC | 599.503 | 139.0685 | 44.7 | 0.08 |

| | | | | |
|---|---|---|---|---|
| pHILIC | 599.503 | 446.1963 | 44.7 | 0.08 |
| pHILIC | 599.503 | 111.6847 | 44.7 | 0.08 |
| pHILIC | 599.503 | 130.3463 | 44.7 | 0.08 |
| pHILIC | 599.503 | 270.577 | 44.7 | 0.08 |
| pHILIC | 599.503 | 86.52 | 44.7 | 0.08 |
| pHILIC | 599.503 | 224.4489 | 44.7 | 0.08 |
| pHILIC | 599.503 | 195.6414 | 44.7 | 0.08 |
| pHILIC | 599.503 | 107.8053 | 44.7 | 0.08 |
| pHILIC | 599.503 | 75.5737 | 44.6 | 0.08 |
| pHILIC | 599.503 | 540.4547 | 44.6 | 0.08 |
| pHILIC | 599.503 | 69.3177 | 44.6 | 0.08 |
| pHILIC | 599.503 | 219.3983 | 44.6 | 0.08 |
| pHILIC | 599.503 | 440.045 | 44.6 | 0.08 |
| pHILIC | 599.503 | 71.4105 | 44.6 | 0.08 |
| pHILIC | 599.503 | 99.1834 | 44.6 | 0.08 |
| pHILIC | 599.503 | 79.2181 | 44.6 | 0.08 |
| pHILIC | 599.503 | 547.6706 | 44.6 | 0.08 |
| pHILIC | 599.503 | 97.4141 | 44.6 | 0.08 |
| pHILIC | 599.503 | 413.123 | 44.6 | 0.08 |
| pHILIC | 599.503 | 227.1503 | 44.6 | 0.08 |
| pHILIC | 599.503 | 93.8548 | 44.6 | 0.08 |
| pHILIC | 599.503 | 130.1913 | 44.6 | 0.08 |
| pHILIC | 599.503 | 614.3846 | 44.6 | 0.08 |
| pHILIC | 599.503 | 151.0746 | 44.6 | 0.08 |
| pHILIC | 599.503 | 498.0549 | 44.6 | 0.08 |
| pHILIC | 599.503 | 325.6423 | 44.6 | 0.08 |
| pHILIC | 599.503 | 161.2817 | 44.6 | 0.08 |
| pHILIC | 599.503 | 135.2318 | 44.6 | 0.08 |
| pHILIC | 599.503 | 221.2509 | 44.6 | 0.08 |
| pHILIC | 599.503 | 139.6883 | 44.5 | 0.08 |
| pHILIC | 599.503 | 527.8536 | 44.5 | 0.08 |
| pHILIC | 599.503 | 257.2206 | 44.5 | 0.08 |
| pHILIC | 599.503 | 67.3452 | 44.5 | 0.08 |
| pHILIC | 599.503 | 95.8066 | 44.5 | 0.08 |
| pHILIC | 599.503 | 129.4823 | 44.5 | 0.08 |
| pHILIC | 599.503 | 251.3726 | 44.5 | 0.08 |
| pHILIC | 599.503 | 89.5117 | 44.5 | 0.08 |
| pHILIC | 599.503 | 357.7709 | 44.5 | 0.08 |
| pHILIC | 599.503 | 325.5515 | 44.5 | 0.08 |
| pHILIC | 599.503 | 88.0012 | 44.5 | 0.08 |
| pHILIC | 599.503 | 63.523 | 44.5 | 0.08 |
| pHILIC | 599.503 | 209.0762 | 44.5 | 0.08 |

| | | | | |
|---|---|---|---|---|
| pHILIC | 599.503 | 66.5615 | 44.5 | 0.08 |
| pHILIC | 599.503 | 94.1322 | 44.5 | 0.08 |
| pHILIC | 599.503 | 106.3122 | 44.5 | 0.08 |
| pHILIC | 599.503 | 125.4668 | 44.5 | 0.08 |
| pHILIC | 599.503 | 85.7787 | 44.5 | 0.08 |
| pHILIC | 599.503 | 99.6494 | 44.5 | 0.08 |
| pHILIC | 599.503 | 207.3716 | 44.5 | 0.08 |
| pHILIC | 599.503 | 124.9034 | 44.5 | 0.08 |
| pHILIC | 599.503 | 84.9942 | 44.5 | 0.08 |
| pHILIC | 599.503 | 243.8431 | 44.5 | 0.08 |
| pHILIC | 599.503 | 136.9707 | 44.5 | 0.08 |
| pHILIC | 599.503 | 78.5008 | 44.4 | 0.08 |
| pHILIC | 599.503 | 492.4071 | 44.4 | 0.08 |
| pHILIC | 599.503 | 78.0673 | 44.4 | 0.08 |
| pHILIC | 599.503 | 157.663 | 44.4 | 0.08 |
| pHILIC | 599.503 | 254.5574 | 44.4 | 0.08 |
| pHILIC | 599.503 | 336.9246 | 44.4 | 0.08 |
| pHILIC | 599.503 | 119.0197 | 44.4 | 0.08 |
| pHILIC | 599.503 | 82.0616 | 44.4 | 0.08 |
| pHILIC | 599.503 | 579.8902 | 44.4 | 0.08 |
| pHILIC | 599.503 | 77.4164 | 44.4 | 0.08 |
| pHILIC | 599.503 | 346.6029 | 44.3 | 0.08 |
| pHILIC | 599.503 | 72.0875 | 44.3 | 0.08 |
| pHILIC | 599.503 | 109.9137 | 44.3 | 0.08 |
| pHILIC | 599.503 | 164.8608 | 44.3 | 0.08 |
| pHILIC | 599.503 | 89.4158 | 44.3 | 0.08 |
| pHILIC | 599.503 | 135.8708 | 44.3 | 0.08 |
| pHILIC | 599.503 | 95.3623 | 44.3 | 0.08 |
| pHILIC | 599.503 | 93.9946 | 44.3 | 0.08 |
| pHILIC | 599.503 | 470.1027 | 44.3 | 0.08 |
| pHILIC | 599.503 | 151.4799 | 44.3 | 0.08 |
| pHILIC | 599.503 | 204.1944 | 44.2 | 0.08 |
| pHILIC | 599.503 | 235.999 | 44.2 | 0.08 |
| pHILIC | 599.503 | 247.3332 | 44.2 | 0.08 |
| pHILIC | 599.503 | 537.9136 | 44.2 | 0.08 |
| pHILIC | 599.503 | 117.1326 | 44.2 | 0.08 |
| pHILIC | 599.503 | 96.6779 | 44.2 | 0.08 |
| pHILIC | 599.503 | 250.9241 | 44.2 | 0.08 |
| pHILIC | 599.503 | 230.4058 | 44.2 | 0.08 |
| pHILIC | 599.503 | 244.292 | 44.2 | 0.08 |
| pHILIC | 599.503 | 124.2073 | 44.2 | 0.08 |
| pHILIC | 599.503 | 167.4703 | 44.2 | 0.08 |

| | | | | |
|---|---|---|---|---|
| pHILIC | 599.503 | 90.5777 | 44.1 | 0.08 |
| pHILIC | 599.503 | 98.4354 | 44.1 | 0.08 |
| pHILIC | 599.503 | 118.968 | 44.1 | 0.08 |
| pHILIC | 599.503 | 124.9849 | 44.1 | 0.08 |
| pHILIC | 599.503 | 85.4371 | 44.1 | 0.08 |
| pHILIC | 599.503 | 98.6293 | 44.1 | 0.08 |
| pHILIC | 599.503 | 91.0347 | 44.1 | 0.08 |
| pHILIC | 599.503 | 558.9694 | 44.1 | 0.08 |
| pHILIC | 599.503 | 390.1681 | 44.1 | 0.08 |
| pHILIC | 599.503 | 119.3211 | 44.1 | 0.08 |
| pHILIC | 599.503 | 198.5045 | 44.1 | 0.08 |
| pHILIC | 599.503 | 126.4845 | 44.1 | 0.08 |
| pHILIC | 599.503 | 118.553 | 44.1 | 0.08 |
| pHILIC | 599.503 | 407.5444 | 44.1 | 0.08 |
| pHILIC | 599.503 | 304.4636 | 44 | 0.08 |
| pHILIC | 599.503 | 133.8852 | 44 | 0.08 |
| pHILIC | 599.503 | 202.3546 | 44 | 0.08 |
| pHILIC | 599.503 | 70.6989 | 44 | 0.08 |
| pHILIC | 599.503 | 350.5707 | 44 | 0.08 |
| pHILIC | 599.503 | 253.3746 | 44 | 0.08 |
| pHILIC | 599.503 | 82.6909 | 44 | 0.08 |
| pHILIC | 599.503 | 88.0402 | 44 | 0.08 |
| pHILIC | 599.503 | 426.7169 | 43.9 | 0.08 |
| pHILIC | 599.503 | 627.2101 | 43.9 | 0.08 |
| pHILIC | 599.503 | 186.2544 | 43.9 | 0.08 |
| pHILIC | 599.503 | 108.8735 | 43.9 | 0.08 |
| pHILIC | 599.503 | 364.2147 | 43.9 | 0.08 |
| pHILIC | 599.503 | 85.2206 | 43.9 | 0.08 |
| pHILIC | 599.503 | 78.34 | 43.9 | 0.08 |
| pHILIC | 599.503 | 509.8985 | 43.9 | 0.08 |
| pHILIC | 599.503 | 139.0173 | 43.9 | 0.08 |
| pHILIC | 599.503 | 280.946 | 43.9 | 0.08 |
| pHILIC | 599.503 | 284.7997 | 43.9 | 0.08 |
| pHILIC | 599.503 | 243.0054 | 43.9 | 0.08 |
| pHILIC | 599.503 | 65.3582 | 43.9 | 0.08 |
| pHILIC | 599.503 | 173.9522 | 43.9 | 0.08 |
| pHILIC | 599.503 | 167.8062 | 43.8 | 0.08 |
| pHILIC | 599.503 | 135.0661 | 43.8 | 0.08 |
| pHILIC | 599.503 | 101.7256 | 43.8 | 0.08 |
| pHILIC | 599.503 | 104.7184 | 43.8 | 0.08 |
| pHILIC | 599.503 | 72.8204 | 43.8 | 0.08 |
| pHILIC | 599.503 | 68.6766 | 43.8 | 0.08 |

| | | | | |
|---|---|---|---|---|
| pHILIC | 599.503 | 101.1855 | 43.8 | 0.08 |
| pHILIC | 599.503 | 176.0893 | 43.8 | 0.08 |
| pHILIC | 599.503 | 69.128 | 43.8 | 0.08 |
| pHILIC | 599.503 | 82.0258 | 43.8 | 0.08 |
| pHILIC | 599.503 | 384.0161 | 43.8 | 0.08 |
| pHILIC | 599.503 | 104.1081 | 43.8 | 0.08 |
| pHILIC | 599.503 | 201.2941 | 43.8 | 0.08 |
| pHILIC | 599.503 | 270.1326 | 43.8 | 0.08 |
| pHILIC | 599.503 | 197.3353 | 43.8 | 0.08 |
| pHILIC | 599.503 | 215.6835 | 43.8 | 0.08 |
| pHILIC | 599.503 | 74.1148 | 43.7 | 0.08 |
| pHILIC | 599.503 | 453.5938 | 43.7 | 0.08 |
| pHILIC | 599.503 | 173.5594 | 43.7 | 0.08 |
| pHILIC | 599.503 | 120.0187 | 43.7 | 0.08 |
| pHILIC | 599.503 | 422.2938 | 43.7 | 0.08 |
| pHILIC | 599.503 | 83.2839 | 43.7 | 0.08 |
| pHILIC | 599.503 | 196.8581 | 43.7 | 0.08 |
| pHILIC | 599.503 | 495.6892 | 43.7 | 0.08 |
| pHILIC | 599.503 | 430.3172 | 43.7 | 0.08 |
| pHILIC | 599.503 | 168.5595 | 43.7 | 0.08 |
| pHILIC | 599.503 | 409.7268 | 43.6 | 0.08 |
| pHILIC | 599.503 | 99.113 | 43.6 | 0.08 |
| pHILIC | 599.503 | 141.6617 | 43.6 | 0.08 |
| pHILIC | 599.503 | 146.7185 | 43.6 | 0.08 |
| pHILIC | 599.503 | 123.5485 | 43.6 | 0.08 |
| pHILIC | 599.503 | 431.8427 | 43.6 | 0.08 |
| pHILIC | 599.503 | 70.4724 | 43.6 | 0.08 |
| pHILIC | 599.503 | 100.6672 | 43.6 | 0.08 |
| pHILIC | 599.503 | 175.3647 | 43.6 | 0.08 |
| pHILIC | 599.503 | 153.4062 | 43.6 | 0.08 |
| pHILIC | 599.503 | 107.3956 | 43.6 | 0.08 |
| pHILIC | 599.503 | 198.3598 | 43.6 | 0.08 |
| pHILIC | 599.503 | 378.8026 | 43.6 | 0.08 |
| pHILIC | 599.503 | 96.211 | 43.6 | 0.08 |
| pHILIC | 599.503 | 94.2025 | 43.6 | 0.08 |
| pHILIC | 599.503 | 205.5323 | 43.6 | 0.08 |
| pHILIC | 599.503 | 81.0796 | 43.6 | 0.08 |
| pHILIC | 599.503 | 118.6015 | 43.6 | 0.08 |
| pHILIC | 599.503 | 135.1055 | 43.6 | 0.08 |
| pHILIC | 599.503 | 99.3406 | 43.6 | 0.08 |
| pHILIC | 599.503 | 583.7177 | 43.6 | 0.08 |
| pHILIC | 599.503 | 323.7528 | 43.6 | 0.08 |

| | | | | |
|---|---|---|---|---|
| pHILIC | 599.503 | 149.5113 | 43.5 | 0.08 |
| pHILIC | 599.503 | 494.3505 | 43.5 | 0.08 |
| pHILIC | 599.503 | 299.5976 | 43.5 | 0.08 |
| pHILIC | 599.503 | 72.2606 | 43.5 | 0.08 |
| pHILIC | 599.503 | 76.3854 | 43.5 | 0.08 |
| pHILIC | 599.503 | 105.3379 | 43.5 | 0.08 |
| pHILIC | 599.503 | 133.7067 | 43.5 | 0.08 |
| pHILIC | 599.503 | 132.864 | 43.5 | 0.08 |
| pHILIC | 599.503 | 166.1257 | 43.4 | 0.08 |
| pHILIC | 599.503 | 351.6111 | 43.4 | 0.08 |
| pHILIC | 599.503 | 216.8183 | 43.4 | 0.08 |
| pHILIC | 599.503 | 87.9967 | 43.4 | 0.08 |
| pHILIC | 599.503 | 166.882 | 43.4 | 0.08 |
| pHILIC | 599.503 | 119.6852 | 43.4 | 0.08 |
| pHILIC | 599.503 | 377.6351 | 43.4 | 0.08 |
| pHILIC | 599.503 | 373.9628 | 43.4 | 0.08 |
| pHILIC | 599.503 | 72.05 | 43.4 | 0.08 |
| pHILIC | 599.503 | 216.9945 | 43.4 | 0.08 |
| pHILIC | 599.503 | 77.6181 | 43.4 | 0.08 |
| pHILIC | 599.503 | 202.7218 | 43.4 | 0.08 |
| pHILIC | 599.503 | 437.8051 | 43.4 | 0.08 |
| pHILIC | 599.503 | 129.3586 | 43.3 | 0.08 |
| pHILIC | 599.503 | 355.0704 | 43.3 | 0.08 |
| pHILIC | 599.503 | 353.1821 | 43.3 | 0.08 |
| pHILIC | 599.503 | 333.1158 | 43.3 | 0.08 |
| pHILIC | 599.503 | 169.4052 | 43.3 | 0.08 |
| pHILIC | 599.503 | 355.2966 | 43.3 | 0.08 |
| pHILIC | 599.503 | 73.6178 | 43.3 | 0.08 |
| pHILIC | 599.503 | 78.1977 | 43.3 | 0.08 |
| pHILIC | 599.503 | 189.7343 | 43.3 | 0.08 |
| pHILIC | 599.503 | 527.1998 | 43.2 | 0.08 |
| pHILIC | 599.503 | 90.2336 | 43.2 | 0.08 |
| pHILIC | 599.503 | 266.182 | 43.2 | 0.08 |
| pHILIC | 599.503 | 311.5264 | 43.2 | 0.08 |
| pHILIC | 599.503 | 93.9874 | 43.2 | 0.08 |
| pHILIC | 599.503 | 110.169 | 43.2 | 0.08 |
| pHILIC | 599.503 | 94.9375 | 43.2 | 0.08 |
| pHILIC | 599.503 | 211.674 | 43.2 | 0.08 |
| pHILIC | 599.503 | 176.2806 | 43.2 | 0.08 |
| pHILIC | 599.503 | 72.9648 | 43.2 | 0.08 |
| pHILIC | 599.503 | 334.8754 | 43.2 | 0.08 |
| pHILIC | 599.503 | 221.4564 | 43.2 | 0.08 |

| | | | | |
|---|---|---|---|---|
| pHILIC | 599.503 | 185.0945 | 43.1 | 0.08 |
| pHILIC | 599.503 | 66.532 | 43.1 | 0.08 |
| pHILIC | 599.503 | 529.7252 | 43.1 | 0.08 |
| pHILIC | 599.503 | 78.4576 | 43.1 | 0.08 |
| pHILIC | 599.503 | 551.7747 | 43.1 | 0.08 |
| pHILIC | 599.503 | 83.6392 | 43.1 | 0.08 |
| pHILIC | 599.503 | 190.8247 | 43.1 | 0.08 |
| pHILIC | 599.503 | 78.1038 | 43.1 | 0.08 |
| pHILIC | 599.503 | 86.1718 | 43.1 | 0.08 |
| pHILIC | 599.503 | 130.1083 | 43.1 | 0.08 |
| pHILIC | 599.503 | 179.375 | 43.1 | 0.08 |
| pHILIC | 599.503 | 105.9322 | 43.1 | 0.08 |
| pHILIC | 599.503 | 103.3427 | 43.1 | 0.08 |
| pHILIC | 599.503 | 462.4305 | 43.1 | 0.08 |
| pHILIC | 599.503 | 134.3531 | 43.1 | 0.08 |
| pHILIC | 599.503 | 168.0728 | 43.1 | 0.08 |
| pHILIC | 599.503 | 148.7568 | 43.1 | 0.08 |
| pHILIC | 599.503 | 215.3161 | 43.1 | 0.08 |
| pHILIC | 599.503 | 94.7482 | 43.1 | 0.08 |
| pHILIC | 599.503 | 218.6746 | 43.1 | 0.08 |
| pHILIC | 599.503 | 104.322 | 43.1 | 0.08 |
| pHILIC | 599.503 | 245.8444 | 43 | 0.08 |
| pHILIC | 599.503 | 101.0137 | 43 | 0.08 |
| pHILIC | 599.503 | 301.4417 | 43 | 0.08 |
| pHILIC | 599.503 | 173.8767 | 43 | 0.08 |
| pHILIC | 599.503 | 86.8405 | 43 | 0.08 |
| pHILIC | 599.503 | 94.6319 | 43 | 0.08 |
| pHILIC | 599.503 | 119.3961 | 43 | 0.08 |
| pHILIC | 599.503 | 70.5384 | 43 | 0.08 |
| pHILIC | 599.503 | 105.4757 | 43 | 0.08 |
| pHILIC | 599.503 | 385.0807 | 43 | 0.08 |
| pHILIC | 599.503 | 190.4118 | 43 | 0.08 |
| pHILIC | 599.503 | 175.8954 | 42.9 | 0.08 |
| pHILIC | 599.503 | 473.673 | 42.9 | 0.08 |
| pHILIC | 599.503 | 101.9167 | 42.9 | 0.08 |
| pHILIC | 599.503 | 140.1455 | 42.9 | 0.08 |
| pHILIC | 599.503 | 339.2081 | 42.9 | 0.08 |
| pHILIC | 599.503 | 506.5147 | 42.9 | 0.08 |
| pHILIC | 599.503 | 173.3388 | 42.9 | 0.08 |
| pHILIC | 599.503 | 114.2256 | 42.9 | 0.08 |
| pHILIC | 599.503 | 138.8363 | 42.9 | 0.08 |
| pHILIC | 599.503 | 86.7821 | 42.9 | 0.08 |

| | | | | |
|---|---|---|---|---|
| pHILIC | 599.503 | 407.9238 | 42.9 | 0.08 |
| pHILIC | 599.503 | 106.3485 | 42.9 | 0.08 |
| pHILIC | 599.503 | 132.4077 | 42.9 | 0.08 |
| pHILIC | 599.503 | 199.1855 | 42.9 | 0.08 |
| pHILIC | 599.503 | 132.7925 | 42.8 | 0.08 |
| pHILIC | 599.503 | 140.5137 | 42.8 | 0.08 |
| pHILIC | 599.503 | 122.0829 | 42.8 | 0.08 |
| pHILIC | 599.503 | 205.4161 | 42.8 | 0.08 |
| pHILIC | 599.503 | 163.0763 | 42.8 | 0.08 |
| pHILIC | 599.503 | 335.0041 | 42.8 | 0.08 |
| pHILIC | 599.503 | 100.5747 | 42.8 | 0.08 |
| pHILIC | 599.503 | 104.7763 | 42.8 | 0.08 |
| pHILIC | 599.503 | 68.1215 | 42.8 | 0.08 |
| pHILIC | 599.503 | 78.5766 | 42.8 | 0.08 |
| pHILIC | 599.503 | 64.4333 | 42.8 | 0.08 |
| pHILIC | 599.503 | 165.8777 | 42.8 | 0.08 |
| pHILIC | 599.503 | 154.3663 | 42.7 | 0.08 |
| pHILIC | 599.503 | 102.6318 | 42.7 | 0.08 |
| pHILIC | 599.503 | 178.8731 | 42.7 | 0.08 |
| pHILIC | 599.503 | 131.2415 | 42.7 | 0.08 |
| pHILIC | 599.503 | 82.3021 | 42.7 | 0.07 |
| pHILIC | 599.503 | 87.8298 | 42.7 | 0.07 |
| pHILIC | 599.503 | 76.6174 | 42.7 | 0.07 |
| pHILIC | 599.503 | 94.7019 | 42.7 | 0.07 |
| pHILIC | 599.503 | 71.9528 | 42.7 | 0.07 |
| pHILIC | 599.503 | 74.1079 | 42.7 | 0.07 |
| pHILIC | 599.503 | 298.0422 | 42.7 | 0.07 |
| pHILIC | 599.503 | 118.4036 | 42.7 | 0.07 |
| pHILIC | 599.503 | 577.8515 | 42.7 | 0.07 |
| pHILIC | 599.503 | 157.8437 | 42.7 | 0.07 |
| pHILIC | 599.503 | 192.8922 | 42.7 | 0.07 |
| pHILIC | 599.503 | 155.6782 | 42.7 | 0.07 |
| pHILIC | 599.503 | 179.6284 | 42.7 | 0.07 |
| pHILIC | 599.503 | 311.5946 | 42.7 | 0.07 |
| pHILIC | 599.503 | 177.8961 | 42.7 | 0.07 |
| pHILIC | 599.503 | 145.1281 | 42.6 | 0.07 |
| pHILIC | 599.503 | 188.8571 | 42.6 | 0.07 |
| pHILIC | 599.503 | 94.6531 | 42.6 | 0.07 |
| pHILIC | 599.503 | 537.4473 | 42.6 | 0.07 |
| pHILIC | 599.503 | 222.352 | 42.6 | 0.07 |
| pHILIC | 599.503 | 63.997 | 42.6 | 0.07 |
| pHILIC | 599.503 | 484.8961 | 42.6 | 0.07 |

| | | | | |
|---|---|---|---|---|
| pHILIC | 599.503 | 75.5947 | 42.6 | 0.07 |
| pHILIC | 599.503 | 576.1217 | 42.6 | 0.07 |
| pHILIC | 599.503 | 96.3912 | 42.6 | 0.07 |
| pHILIC | 599.503 | 264.2068 | 42.6 | 0.07 |
| pHILIC | 599.503 | 491.1973 | 42.6 | 0.07 |
| pHILIC | 599.503 | 112.5853 | 42.6 | 0.07 |
| pHILIC | 599.503 | 174.4961 | 42.6 | 0.07 |
| pHILIC | 599.503 | 64.2448 | 42.6 | 0.07 |
| pHILIC | 599.503 | 212.6031 | 42.6 | 0.07 |
| pHILIC | 599.503 | 280.2451 | 42.6 | 0.07 |
| pHILIC | 599.503 | 496.7211 | 42.6 | 0.07 |
| pHILIC | 599.503 | 74.0387 | 42.6 | 0.07 |
| pHILIC | 599.503 | 216.5656 | 42.6 | 0.07 |
| pHILIC | 599.503 | 81.4344 | 42.5 | 0.07 |
| pHILIC | 599.503 | 103.5451 | 42.5 | 0.07 |
| pHILIC | 599.503 | 196.8368 | 42.5 | 0.07 |
| pHILIC | 599.503 | 125.1211 | 42.5 | 0.07 |
| pHILIC | 599.503 | 339.8017 | 42.5 | 0.07 |
| pHILIC | 599.503 | 77.8721 | 42.5 | 0.07 |
| pHILIC | 599.503 | 292.6671 | 42.5 | 0.07 |
| pHILIC | 599.503 | 79.3688 | 42.5 | 0.07 |
| pHILIC | 599.503 | 63.6806 | 42.5 | 0.07 |
| pHILIC | 599.503 | 537.6884 | 42.5 | 0.07 |
| pHILIC | 599.503 | 174.2052 | 42.5 | 0.07 |
| pHILIC | 599.503 | 75.0112 | 42.5 | 0.07 |
| pHILIC | 599.503 | 401.5682 | 42.5 | 0.07 |
| pHILIC | 599.503 | 87.3499 | 42.5 | 0.07 |
| pHILIC | 599.503 | 85.7163 | 42.5 | 0.07 |
| pHILIC | 599.503 | 178.7672 | 42.4 | 0.07 |
| pHILIC | 599.503 | 79.8463 | 42.4 | 0.07 |
| pHILIC | 599.503 | 473.8325 | 42.4 | 0.07 |
| pHILIC | 599.503 | 198.304 | 42.4 | 0.07 |
| pHILIC | 599.503 | 95.9167 | 42.4 | 0.07 |
| pHILIC | 599.503 | 247.224 | 42.4 | 0.07 |
| pHILIC | 599.503 | 183.8011 | 42.4 | 0.07 |
| pHILIC | 599.503 | 337.0573 | 42.4 | 0.07 |
| pHILIC | 599.503 | 210.3786 | 42.4 | 0.07 |
| pHILIC | 599.503 | 71.0813 | 42.4 | 0.07 |
| pHILIC | 599.503 | 102.0822 | 42.4 | 0.07 |
| pHILIC | 599.503 | 74.211 | 42.4 | 0.07 |
| pHILIC | 599.503 | 527.3851 | 42.4 | 0.07 |
| pHILIC | 599.503 | 383.7264 | 42.4 | 0.07 |

| | | | | |
|---|---|---|---|---|
| pHILIC | 599.503 | 121.8614 | 42.4 | 0.07 |
| pHILIC | 599.503 | 533.4 | 42.4 | 0.07 |
| pHILIC | 599.503 | 93.9395 | 42.4 | 0.07 |
| pHILIC | 599.503 | 436.021 | 42.4 | 0.07 |
| pHILIC | 599.503 | 105.3583 | 42.3 | 0.07 |
| pHILIC | 599.503 | 626.3228 | 42.3 | 0.07 |
| pHILIC | 599.503 | 70.8209 | 42.3 | 0.07 |
| pHILIC | 599.503 | 199.7166 | 42.3 | 0.07 |
| pHILIC | 599.503 | 583.551 | 42.3 | 0.07 |
| pHILIC | 599.503 | 120.8315 | 42.3 | 0.07 |
| pHILIC | 599.503 | 111.5697 | 42.3 | 0.07 |
| pHILIC | 599.503 | 373.4223 | 42.3 | 0.07 |
| pHILIC | 599.503 | 66.8927 | 42.3 | 0.07 |
| pHILIC | 599.503 | 220.5016 | 42.3 | 0.07 |
| pHILIC | 599.503 | 125.5436 | 42.3 | 0.07 |
| pHILIC | 599.503 | 93.07 | 42.3 | 0.07 |
| pHILIC | 599.503 | 363.2158 | 42.3 | 0.07 |
| pHILIC | 599.503 | 66.4721 | 42.3 | 0.07 |
| pHILIC | 599.503 | 118.3568 | 42.3 | 0.07 |
| pHILIC | 599.503 | 70.454 | 42.3 | 0.07 |
| pHILIC | 599.503 | 324.5101 | 42.3 | 0.07 |
| pHILIC | 599.503 | 123.1327 | 42.3 | 0.07 |
| pHILIC | 599.503 | 76.5231 | 42.2 | 0.07 |
| pHILIC | 599.503 | 136.6855 | 42.2 | 0.07 |
| pHILIC | 599.503 | 580.1266 | 42.2 | 0.07 |
| pHILIC | 599.503 | 79.1139 | 42.2 | 0.07 |
| pHILIC | 599.503 | 76.1703 | 42.2 | 0.07 |
| pHILIC | 599.503 | 198.6668 | 42.2 | 0.07 |
| pHILIC | 599.503 | 151.1572 | 42.2 | 0.07 |
| pHILIC | 599.503 | 291.6524 | 42.2 | 0.07 |
| pHILIC | 599.503 | 293.4636 | 42.2 | 0.07 |
| pHILIC | 599.503 | 305.9472 | 42.2 | 0.07 |
| pHILIC | 599.503 | 85.8732 | 42.2 | 0.07 |
| pHILIC | 599.503 | 169.3868 | 42.2 | 0.07 |
| pHILIC | 599.503 | 182.7704 | 42.2 | 0.07 |
| pHILIC | 599.503 | 94.2112 | 42.2 | 0.07 |
| pHILIC | 599.503 | 92.3181 | 42.2 | 0.07 |
| pHILIC | 599.503 | 244.674 | 42.2 | 0.07 |
| pHILIC | 599.503 | 200.9102 | 42.2 | 0.07 |
| pHILIC | 599.503 | 73.5783 | 42.2 | 0.07 |
| pHILIC | 599.503 | 586.3135 | 42.2 | 0.07 |
| pHILIC | 599.503 | 112.5697 | 42.1 | 0.07 |

| | | | | |
|---|---|---|---|---|
| pHILIC | 599.503 | 85.8483 | 42.1 | 0.07 |
| pHILIC | 599.503 | 260.3488 | 42.1 | 0.07 |
| pHILIC | 599.503 | 230.0078 | 42.1 | 0.07 |
| pHILIC | 599.503 | 191.1071 | 42.1 | 0.07 |
| pHILIC | 599.503 | 150.8214 | 42.1 | 0.07 |
| pHILIC | 599.503 | 240.4175 | 42.1 | 0.07 |
| pHILIC | 599.503 | 542.9636 | 42.1 | 0.07 |
| pHILIC | 599.503 | 179.2789 | 42.1 | 0.07 |
| pHILIC | 599.503 | 369.5154 | 42.1 | 0.07 |
| pHILIC | 599.503 | 127.2053 | 42.1 | 0.07 |
| pHILIC | 599.503 | 434.6979 | 42.1 | 0.07 |
| pHILIC | 599.503 | 91.3139 | 42.1 | 0.07 |
| pHILIC | 599.503 | 101.4551 | 42.1 | 0.07 |
| pHILIC | 599.503 | 321.0148 | 42 | 0.07 |
| pHILIC | 599.503 | 69.801 | 42 | 0.07 |
| pHILIC | 599.503 | 185.8068 | 42 | 0.07 |
| pHILIC | 599.503 | 166.6394 | 42 | 0.07 |
| pHILIC | 599.503 | 96.8702 | 42 | 0.07 |
| pHILIC | 599.503 | 303.4204 | 42 | 0.07 |
| pHILIC | 599.503 | 151.3252 | 42 | 0.07 |
| pHILIC | 599.503 | 120.1404 | 42 | 0.07 |
| pHILIC | 599.503 | 228.2623 | 42 | 0.07 |
| pHILIC | 599.503 | 350.7099 | 42 | 0.07 |
| pHILIC | 599.503 | 88.6759 | 42 | 0.07 |
| pHILIC | 599.503 | 498.8129 | 42 | 0.07 |
| pHILIC | 599.503 | 99.3938 | 42 | 0.07 |
| pHILIC | 599.503 | 65.2969 | 42 | 0.07 |
| pHILIC | 599.503 | 64.8345 | 41.9 | 0.07 |
| pHILIC | 599.503 | 322.088 | 41.9 | 0.07 |
| pHILIC | 599.503 | 310.0407 | 41.9 | 0.07 |
| pHILIC | 599.503 | 70.3398 | 41.9 | 0.07 |
| pHILIC | 599.503 | 70.0846 | 41.9 | 0.07 |
| pHILIC | 599.503 | 181.3428 | 41.9 | 0.07 |
| pHILIC | 599.503 | 105.011 | 41.9 | 0.07 |
| pHILIC | 599.503 | 276.1558 | 41.9 | 0.07 |
| pHILIC | 599.503 | 348.6861 | 41.9 | 0.07 |
| pHILIC | 599.503 | 93.0778 | 41.9 | 0.07 |
| pHILIC | 599.503 | 208.688 | 41.9 | 0.07 |
| pHILIC | 599.503 | 123.6243 | 41.9 | 0.07 |
| pHILIC | 599.503 | 455.027 | 41.9 | 0.07 |
| pHILIC | 599.503 | 213.3719 | 41.9 | 0.07 |
| pHILIC | 599.503 | 70.6609 | 41.9 | 0.07 |

| | | | | |
|---|---|---|---|---|
| pHILIC | 599.503 | 67.048 | 41.9 | 0.07 |
| pHILIC | 599.503 | 104.7597 | 41.9 | 0.07 |
| pHILIC | 599.503 | 216.454 | 41.9 | 0.07 |
| pHILIC | 599.503 | 512.6143 | 41.9 | 0.07 |
| pHILIC | 599.503 | 137.7007 | 41.9 | 0.07 |
| pHILIC | 599.503 | 268.1531 | 41.8 | 0.07 |
| pHILIC | 599.503 | 164.6808 | 41.8 | 0.07 |
| pHILIC | 599.503 | 70.6395 | 41.8 | 0.07 |
| pHILIC | 599.503 | 182.0835 | 41.8 | 0.07 |
| pHILIC | 599.503 | 64.4784 | 41.8 | 0.07 |
| pHILIC | 599.503 | 140.659 | 41.8 | 0.07 |
| pHILIC | 599.503 | 75.4523 | 41.8 | 0.07 |
| pHILIC | 599.503 | 284.8626 | 41.8 | 0.07 |
| pHILIC | 599.503 | 128.0027 | 41.8 | 0.07 |
| pHILIC | 599.503 | 106.8968 | 41.8 | 0.07 |
| pHILIC | 599.503 | 77.333 | 41.8 | 0.07 |
| pHILIC | 599.503 | 177.7113 | 41.8 | 0.07 |
| pHILIC | 599.503 | 103.303 | 41.8 | 0.07 |
| pHILIC | 599.503 | 497.361 | 41.8 | 0.07 |
| pHILIC | 599.503 | 177.876 | 41.8 | 0.07 |
| pHILIC | 599.503 | 93.9335 | 41.8 | 0.07 |
| pHILIC | 599.503 | 246.573 | 41.8 | 0.07 |
| pHILIC | 599.503 | 94.6485 | 41.8 | 0.07 |
| pHILIC | 599.503 | 429.4482 | 41.8 | 0.07 |
| pHILIC | 599.503 | 363.3264 | 41.8 | 0.07 |
| pHILIC | 599.503 | 167.8237 | 41.7 | 0.07 |
| pHILIC | 599.503 | 94.361 | 41.7 | 0.07 |
| pHILIC | 599.503 | 73.9089 | 41.7 | 0.07 |
| pHILIC | 599.503 | 64.0118 | 41.7 | 0.07 |
| pHILIC | 599.503 | 77.7072 | 41.7 | 0.07 |
| pHILIC | 599.503 | 588.1414 | 41.7 | 0.07 |
| pHILIC | 599.503 | 72.6468 | 41.7 | 0.07 |
| pHILIC | 599.503 | 181.6097 | 41.7 | 0.07 |
| pHILIC | 599.503 | 83.0891 | 41.7 | 0.07 |
| pHILIC | 599.503 | 76.0077 | 41.7 | 0.07 |
| pHILIC | 599.503 | 97.8064 | 41.7 | 0.07 |
| pHILIC | 599.503 | 305.0513 | 41.7 | 0.07 |
| pHILIC | 599.503 | 214.1111 | 41.7 | 0.07 |
| pHILIC | 599.503 | 459.1635 | 41.7 | 0.07 |
| pHILIC | 599.503 | 183.7383 | 41.7 | 0.07 |
| pHILIC | 599.503 | 413.7315 | 41.7 | 0.07 |
| pHILIC | 599.503 | 295.5618 | 41.7 | 0.07 |

| | | | | |
|---|---|---|---|---|
| pHILIC | 599.503 | 138.2066 | 41.7 | 0.07 |
| pHILIC | 599.503 | 263.5578 | 41.6 | 0.07 |
| pHILIC | 599.503 | 160.1028 | 41.6 | 0.07 |
| pHILIC | 599.503 | 274.7573 | 41.6 | 0.07 |
| pHILIC | 599.503 | 142.2279 | 41.6 | 0.07 |
| pHILIC | 599.503 | 219.2727 | 41.6 | 0.07 |
| pHILIC | 599.503 | 103.4203 | 41.6 | 0.07 |
| pHILIC | 599.503 | 184.3758 | 41.6 | 0.07 |
| pHILIC | 599.503 | 156.9878 | 41.6 | 0.07 |
| pHILIC | 599.503 | 452.2854 | 41.6 | 0.07 |
| pHILIC | 599.503 | 452.4126 | 41.6 | 0.07 |
| pHILIC | 599.503 | 111.8576 | 41.6 | 0.07 |
| pHILIC | 599.503 | 74.2387 | 41.6 | 0.07 |
| pHILIC | 599.503 | 80.7287 | 41.6 | 0.07 |
| pHILIC | 599.503 | 84.9231 | 41.6 | 0.07 |
| pHILIC | 599.503 | 260.6302 | 41.6 | 0.07 |
| pHILIC | 599.503 | 217.487 | 41.6 | 0.07 |
| pHILIC | 599.503 | 114.4441 | 41.6 | 0.07 |
| pHILIC | 599.503 | 108.8069 | 41.6 | 0.07 |
| pHILIC | 599.503 | 279.3794 | 41.6 | 0.07 |
| pHILIC | 599.503 | 324.6848 | 41.6 | 0.07 |
| pHILIC | 599.503 | 90.3367 | 41.5 | 0.07 |
| pHILIC | 599.503 | 63.2472 | 41.5 | 0.07 |
| pHILIC | 599.503 | 407.4068 | 41.5 | 0.07 |
| pHILIC | 599.503 | 68.6647 | 41.5 | 0.07 |
| pHILIC | 599.503 | 127.0981 | 41.5 | 0.07 |
| pHILIC | 599.503 | 95.7334 | 41.5 | 0.07 |
| pHILIC | 599.503 | 539.3489 | 41.5 | 0.07 |
| pHILIC | 599.503 | 150.4108 | 41.5 | 0.07 |
| pHILIC | 599.503 | 64.0456 | 41.5 | 0.07 |
| pHILIC | 599.503 | 63.5669 | 41.5 | 0.07 |
| pHILIC | 599.503 | 102.6731 | 41.5 | 0.07 |
| pHILIC | 599.503 | 153.0946 | 41.4 | 0.07 |
| pHILIC | 599.503 | 98.7206 | 41.4 | 0.07 |
| pHILIC | 599.503 | 113.9619 | 41.4 | 0.07 |
| pHILIC | 599.503 | 527.0557 | 41.4 | 0.07 |
| pHILIC | 599.503 | 108.7625 | 41.4 | 0.07 |
| pHILIC | 599.503 | 71.0359 | 41.4 | 0.07 |
| pHILIC | 599.503 | 178.4898 | 41.4 | 0.07 |
| pHILIC | 599.503 | 294.0866 | 41.4 | 0.07 |
| pHILIC | 599.503 | 156.9066 | 41.4 | 0.07 |
| pHILIC | 599.503 | 81.604 | 41.4 | 0.07 |

| | | | | |
|---|---|---|---|---|
| pHILIC | 599.503 | 141.272 | 41.4 | 0.07 |
| pHILIC | 599.503 | 140.6908 | 41.4 | 0.07 |
| pHILIC | 599.503 | 79.7727 | 41.4 | 0.07 |
| pHILIC | 599.503 | 68.858 | 41.4 | 0.07 |
| pHILIC | 599.503 | 82.3336 | 41.3 | 0.07 |
| pHILIC | 599.503 | 313.1794 | 41.3 | 0.07 |
| pHILIC | 599.503 | 209.224 | 41.3 | 0.07 |
| pHILIC | 599.503 | 215.8792 | 41.3 | 0.07 |
| pHILIC | 599.503 | 114.7176 | 41.3 | 0.07 |
| pHILIC | 599.503 | 101.7007 | 41.3 | 0.07 |
| pHILIC | 599.503 | 237.2035 | 41.3 | 0.07 |
| pHILIC | 599.503 | 182.3648 | 41.3 | 0.07 |
| pHILIC | 599.503 | 263.2879 | 41.3 | 0.07 |
| pHILIC | 599.503 | 87.4284 | 41.3 | 0.07 |
| pHILIC | 599.503 | 177.2399 | 41.3 | 0.07 |
| pHILIC | 599.503 | 90.5478 | 41.3 | 0.07 |
| pHILIC | 599.503 | 63.4165 | 41.3 | 0.07 |
| pHILIC | 599.503 | 194.231 | 41.3 | 0.07 |
| pHILIC | 599.503 | 116.2977 | 41.3 | 0.07 |
| pHILIC | 599.503 | 395.2197 | 41.3 | 0.07 |
| pHILIC | 599.503 | 64.6761 | 41.2 | 0.07 |
| pHILIC | 599.503 | 147.5701 | 41.2 | 0.07 |
| pHILIC | 599.503 | 623.8702 | 41.2 | 0.07 |
| pHILIC | 599.503 | 220.8087 | 41.2 | 0.07 |
| pHILIC | 599.503 | 84.1568 | 41.2 | 0.07 |
| pHILIC | 599.503 | 364.7571 | 41.2 | 0.07 |
| pHILIC | 599.503 | 138.9305 | 41.2 | 0.07 |
| pHILIC | 599.503 | 144.2786 | 41.2 | 0.07 |
| pHILIC | 599.503 | 517.8385 | 41.2 | 0.07 |
| pHILIC | 599.503 | 239.7065 | 41.2 | 0.07 |
| pHILIC | 599.503 | 441.6124 | 41.2 | 0.07 |
| pHILIC | 599.503 | 127.4603 | 41.2 | 0.07 |
| pHILIC | 599.503 | 67.6865 | 41.2 | 0.07 |
| pHILIC | 599.503 | 335.3328 | 41.2 | 0.07 |
| pHILIC | 599.503 | 66.1192 | 41.2 | 0.07 |
| pHILIC | 599.503 | 233.6808 | 41.1 | 0.07 |
| pHILIC | 599.503 | 131.9189 | 41.1 | 0.07 |
| pHILIC | 599.503 | 395.661 | 41.1 | 0.07 |
| pHILIC | 599.503 | 140.6015 | 41.1 | 0.07 |
| pHILIC | 599.503 | 86.1324 | 41.1 | 0.07 |
| pHILIC | 599.503 | 168.3192 | 41.1 | 0.07 |
| pHILIC | 599.503 | 115.2704 | 41.1 | 0.07 |

| | | | | |
|---|---|---|---|---|
| pHILIC | 599.503 | 125.7165 | 41.1 | 0.07 |
| pHILIC | 599.503 | 252.3717 | 41.1 | 0.07 |
| pHILIC | 599.503 | 101.8969 | 41.1 | 0.07 |
| pHILIC | 599.503 | 96.9334 | 41.1 | 0.07 |
| pHILIC | 599.503 | 386.6591 | 41.1 | 0.07 |
| pHILIC | 599.503 | 71.0323 | 41.1 | 0.07 |
| pHILIC | 599.503 | 75.0563 | 41.1 | 0.07 |
| pHILIC | 599.503 | 89.8595 | 41.1 | 0.07 |
| pHILIC | 599.503 | 78.4046 | 41.1 | 0.07 |
| pHILIC | 599.503 | 125.7334 | 41.1 | 0.07 |
| pHILIC | 599.503 | 625.8015 | 41.1 | 0.07 |
| pHILIC | 599.503 | 319.6034 | 41.1 | 0.07 |
| pHILIC | 599.503 | 176.3313 | 41.1 | 0.07 |
| pHILIC | 599.503 | 159.2307 | 41.1 | 0.07 |
| pHILIC | 599.503 | 76.5545 | 41.1 | 0.07 |
| pHILIC | 599.503 | 69.4535 | 41 | 0.07 |
| pHILIC | 599.503 | 442.7068 | 41 | 0.07 |
| pHILIC | 599.503 | 123.4874 | 41 | 0.07 |
| pHILIC | 599.503 | 124.1674 | 41 | 0.07 |
| pHILIC | 599.503 | 286.2182 | 41 | 0.07 |
| pHILIC | 599.503 | 133.6803 | 41 | 0.07 |
| pHILIC | 599.503 | 150.0721 | 41 | 0.07 |
| pHILIC | 599.503 | 259.2915 | 41 | 0.07 |
| pHILIC | 599.503 | 91.3786 | 41 | 0.07 |
| pHILIC | 599.503 | 100.5239 | 40.9 | 0.07 |
| pHILIC | 599.503 | 139.1436 | 40.9 | 0.07 |
| pHILIC | 599.503 | 374.9196 | 40.9 | 0.07 |
| pHILIC | 599.503 | 240.014 | 40.9 | 0.07 |
| pHILIC | 599.503 | 95.0857 | 40.9 | 0.07 |
| pHILIC | 599.503 | 145.6989 | 40.9 | 0.07 |
| pHILIC | 599.503 | 276.531 | 40.9 | 0.07 |
| pHILIC | 599.503 | 70.0725 | 40.9 | 0.07 |
| pHILIC | 599.503 | 483.3359 | 40.9 | 0.07 |
| pHILIC | 599.503 | 86.4782 | 40.9 | 0.07 |
| pHILIC | 599.503 | 574.963 | 40.9 | 0.07 |
| pHILIC | 599.503 | 90.018 | 40.9 | 0.07 |
| pHILIC | 599.503 | 123.3004 | 40.9 | 0.07 |
| pHILIC | 599.503 | 75.0752 | 40.9 | 0.07 |
| pHILIC | 599.503 | 270.4618 | 40.9 | 0.07 |
| pHILIC | 599.503 | 99.8368 | 40.9 | 0.07 |
| pHILIC | 599.503 | 151.9251 | 40.9 | 0.07 |
| pHILIC | 599.503 | 129.1248 | 40.9 | 0.07 |

| | | | | |
|---|---|---|---|---|
| pHILIC | 599.503 | 198.5689 | 40.8 | 0.07 |
| pHILIC | 599.503 | 137.4104 | 40.8 | 0.07 |
| pHILIC | 599.503 | 97.0949 | 40.8 | 0.07 |
| pHILIC | 599.503 | 432.2784 | 40.8 | 0.07 |
| pHILIC | 599.503 | 200.3824 | 40.8 | 0.07 |
| pHILIC | 599.503 | 307.288 | 40.8 | 0.07 |
| pHILIC | 599.503 | 511.5285 | 40.8 | 0.07 |
| pHILIC | 599.503 | 150.3109 | 40.8 | 0.07 |
| pHILIC | 599.503 | 397.6947 | 40.8 | 0.07 |
| pHILIC | 599.503 | 138.5756 | 40.8 | 0.07 |
| pHILIC | 599.503 | 174.8832 | 40.8 | 0.07 |
| pHILIC | 599.503 | 200.4975 | 40.8 | 0.07 |
| pHILIC | 599.503 | 69.1848 | 40.8 | 0.07 |
| pHILIC | 599.503 | 67.244 | 40.8 | 0.07 |
| pHILIC | 599.503 | 231.1847 | 40.7 | 0.07 |
| pHILIC | 599.503 | 64.5314 | 40.7 | 0.07 |
| pHILIC | 599.503 | 163.47 | 40.7 | 0.07 |
| pHILIC | 599.503 | 237.6747 | 40.7 | 0.07 |
| pHILIC | 599.503 | 101.7982 | 40.7 | 0.07 |
| pHILIC | 599.503 | 301.8503 | 40.7 | 0.07 |
| pHILIC | 599.503 | 75.4484 | 40.7 | 0.07 |
| pHILIC | 599.503 | 172.9813 | 40.7 | 0.07 |
| pHILIC | 599.503 | 132.0988 | 40.6 | 0.07 |
| pHILIC | 599.503 | 66.4781 | 40.6 | 0.07 |
| pHILIC | 599.503 | 82.3605 | 40.6 | 0.07 |
| pHILIC | 599.503 | 210.88 | 40.6 | 0.07 |
| pHILIC | 599.503 | 90.6358 | 40.6 | 0.07 |
| pHILIC | 599.503 | 261.9698 | 40.6 | 0.07 |
| pHILIC | 599.503 | 72.2744 | 40.6 | 0.07 |
| pHILIC | 599.503 | 79.9611 | 40.6 | 0.07 |
| pHILIC | 599.503 | 100.2623 | 40.6 | 0.07 |
| pHILIC | 599.503 | 87.9755 | 40.6 | 0.07 |
| pHILIC | 599.503 | 176.7173 | 40.6 | 0.07 |
| pHILIC | 599.503 | 195.0296 | 40.6 | 0.07 |
| pHILIC | 599.503 | 135.597 | 40.6 | 0.07 |
| pHILIC | 599.503 | 202.2161 | 40.6 | 0.07 |
| pHILIC | 599.503 | 335.9755 | 40.6 | 0.07 |
| pHILIC | 599.503 | 119.9999 | 40.6 | 0.07 |
| pHILIC | 599.503 | 319.3917 | 40.5 | 0.07 |
| pHILIC | 599.503 | 68.3177 | 40.5 | 0.07 |
| pHILIC | 599.503 | 158.9855 | 40.5 | 0.07 |
| pHILIC | 599.503 | 79.5328 | 40.5 | 0.07 |

| | | | | |
|---|---|---|---|---|
| pHILIC | 599.503 | 153.0059 | 40.5 | 0.07 |
| pHILIC | 599.503 | 194.3868 | 40.5 | 0.07 |
| pHILIC | 599.503 | 417.5248 | 40.5 | 0.07 |
| pHILIC | 599.503 | 190.8785 | 40.5 | 0.07 |
| pHILIC | 599.503 | 69.85 | 40.5 | 0.07 |
| pHILIC | 599.503 | 65.6969 | 40.5 | 0.07 |
| pHILIC | 599.503 | 124.684 | 40.5 | 0.07 |
| pHILIC | 599.503 | 106.8434 | 40.5 | 0.07 |
| pHILIC | 599.503 | 80.5696 | 40.5 | 0.07 |
| pHILIC | 599.503 | 76.4622 | 40.4 | 0.07 |
| pHILIC | 599.503 | 211.0457 | 40.4 | 0.07 |
| pHILIC | 599.503 | 161.0232 | 40.4 | 0.07 |
| pHILIC | 599.503 | 231.8658 | 40.4 | 0.07 |
| pHILIC | 599.503 | 64.2876 | 40.4 | 0.07 |
| pHILIC | 599.503 | 121.1683 | 40.4 | 0.07 |
| pHILIC | 599.503 | 126.3346 | 40.4 | 0.07 |
| pHILIC | 599.503 | 71.2694 | 40.4 | 0.07 |
| pHILIC | 599.503 | 106.1206 | 40.4 | 0.07 |
| pHILIC | 599.503 | 85.5892 | 40.4 | 0.07 |
| pHILIC | 599.503 | 81.8302 | 40.4 | 0.07 |
| pHILIC | 599.503 | 146.9467 | 40.4 | 0.07 |
| pHILIC | 599.503 | 230.8103 | 40.4 | 0.07 |
| pHILIC | 599.503 | 221.0459 | 40.4 | 0.07 |
| pHILIC | 599.503 | 366.205 | 40.4 | 0.07 |
| pHILIC | 599.503 | 70.0783 | 40.3 | 0.07 |
| pHILIC | 599.503 | 206.1208 | 40.3 | 0.07 |
| pHILIC | 599.503 | 280.0708 | 40.3 | 0.07 |
| pHILIC | 599.503 | 218.0773 | 40.3 | 0.07 |
| pHILIC | 599.503 | 224.0588 | 40.3 | 0.07 |
| pHILIC | 599.503 | 170.1596 | 40.3 | 0.07 |
| pHILIC | 599.503 | 169.9731 | 40.3 | 0.07 |
| pHILIC | 599.503 | 148.0767 | 40.3 | 0.07 |
| pHILIC | 599.503 | 601.6938 | 40.3 | 0.07 |
| pHILIC | 599.503 | 265.1481 | 40.3 | 0.07 |
| pHILIC | 599.503 | 69.5886 | 40.3 | 0.07 |
| pHILIC | 599.503 | 209.3166 | 40.3 | 0.07 |
| pHILIC | 599.503 | 104.7937 | 40.3 | 0.07 |
| pHILIC | 599.503 | 141.1509 | 40.3 | 0.07 |
| pHILIC | 599.503 | 181.9818 | 40.3 | 0.07 |
| pHILIC | 599.503 | 90.7924 | 40.3 | 0.07 |
| pHILIC | 599.503 | 393.6971 | 40.2 | 0.07 |
| pHILIC | 599.503 | 358.8133 | 40.2 | 0.07 |

| | | | | |
|---|---|---|---|---|
| pHILIC | 599.503 | 139.7622 | 40.2 | 0.07 |
| pHILIC | 599.503 | 281.1996 | 40.2 | 0.07 |
| pHILIC | 599.503 | 205.2818 | 40.2 | 0.07 |
| pHILIC | 599.503 | 77.2548 | 40.2 | 0.07 |
| pHILIC | 599.503 | 416.4709 | 40.2 | 0.07 |
| pHILIC | 599.503 | 152.8309 | 40.2 | 0.07 |
| pHILIC | 599.503 | 195.5447 | 40.2 | 0.07 |
| pHILIC | 599.503 | 124.7192 | 40.2 | 0.07 |
| pHILIC | 599.503 | 73.9578 | 40.2 | 0.07 |
| pHILIC | 599.503 | 109.7989 | 40.2 | 0.07 |
| pHILIC | 599.503 | 436.4768 | 40.2 | 0.07 |
| pHILIC | 599.503 | 174.0533 | 40.2 | 0.07 |
| pHILIC | 599.503 | 85.7672 | 40.1 | 0.07 |
| pHILIC | 599.503 | 125.1294 | 40.1 | 0.07 |
| pHILIC | 599.503 | 80.2889 | 40.1 | 0.07 |
| pHILIC | 599.503 | 84.3441 | 40.1 | 0.07 |
| pHILIC | 599.503 | 71.7997 | 40.1 | 0.07 |
| pHILIC | 599.503 | 138.7211 | 40.1 | 0.07 |
| pHILIC | 599.503 | 112.1949 | 40.1 | 0.07 |
| pHILIC | 599.503 | 295.3665 | 40.1 | 0.07 |
| pHILIC | 599.503 | 374.1865 | 40.1 | 0.07 |
| pHILIC | 599.503 | 487.7974 | 40.1 | 0.07 |
| pHILIC | 599.503 | 73.8717 | 40.1 | 0.07 |
| pHILIC | 599.503 | 294.6606 | 40 | 0.07 |
| pHILIC | 599.503 | 107.093 | 40 | 0.07 |
| pHILIC | 599.503 | 66.8165 | 40 | 0.07 |
| pHILIC | 599.503 | 162.842 | 40 | 0.07 |
| pHILIC | 599.503 | 178.2488 | 40 | 0.07 |
| pHILIC | 599.503 | 217.4561 | 40 | 0.07 |
| pHILIC | 599.503 | 194.511 | 40 | 0.07 |
| pHILIC | 599.503 | 83.2697 | 40 | 0.07 |
| pHILIC | 599.503 | 71.7413 | 40 | 0.07 |
| pHILIC | 599.503 | 164.3562 | 40 | 0.07 |
| pHILIC | 599.503 | 125.2291 | 39.9 | 0.07 |
| pHILIC | 599.503 | 137.7918 | 39.9 | 0.07 |
| pHILIC | 599.503 | 65.3614 | 39.9 | 0.07 |
| pHILIC | 599.503 | 112.7936 | 39.9 | 0.07 |
| pHILIC | 599.503 | 77.3429 | 39.9 | 0.07 |
| pHILIC | 599.503 | 65.2233 | 39.9 | 0.07 |
| pHILIC | 599.503 | 78.9787 | 39.9 | 0.07 |
| pHILIC | 599.503 | 238.2954 | 39.9 | 0.07 |
| pHILIC | 599.503 | 74.8165 | 39.9 | 0.07 |

| | | | | |
|---|---|---|---|---|
| pHILIC | 599.503 | 169.3547 | 39.9 | 0.07 |
| pHILIC | 599.503 | 311.741 | 39.9 | 0.07 |
| pHILIC | 599.503 | 178.0007 | 39.9 | 0.07 |
| pHILIC | 599.503 | 93.4994 | 39.9 | 0.07 |
| pHILIC | 599.503 | 149.9405 | 39.8 | 0.07 |
| pHILIC | 599.503 | 192.5522 | 39.8 | 0.07 |
| pHILIC | 599.503 | 95.4036 | 39.8 | 0.07 |
| pHILIC | 599.503 | 119.9638 | 39.8 | 0.07 |
| pHILIC | 599.503 | 98.5076 | 39.8 | 0.07 |
| pHILIC | 599.503 | 119.7194 | 39.8 | 0.07 |
| pHILIC | 599.503 | 149.6869 | 39.8 | 0.07 |
| pHILIC | 599.503 | 145.007 | 39.8 | 0.07 |
| pHILIC | 599.503 | 237.3792 | 39.7 | 0.07 |
| pHILIC | 599.503 | 197.6989 | 39.7 | 0.07 |
| pHILIC | 599.503 | 297.8819 | 39.7 | 0.07 |
| pHILIC | 599.503 | 70.1621 | 39.7 | 0.07 |
| pHILIC | 599.503 | 105.8132 | 39.7 | 0.07 |
| pHILIC | 599.503 | 99.0618 | 39.7 | 0.07 |
| pHILIC | 599.503 | 95.9688 | 39.7 | 0.07 |
| pHILIC | 599.503 | 196.9141 | 39.7 | 0.07 |
| pHILIC | 599.503 | 159.7002 | 39.7 | 0.07 |
| pHILIC | 599.503 | 193.984 | 39.7 | 0.07 |
| pHILIC | 599.503 | 204.7773 | 39.7 | 0.07 |
| pHILIC | 599.503 | 66.2743 | 39.7 | 0.07 |
| pHILIC | 599.503 | 190.4424 | 39.7 | 0.07 |
| pHILIC | 599.503 | 142.1832 | 39.6 | 0.07 |
| pHILIC | 599.503 | 82.9267 | 39.6 | 0.07 |
| pHILIC | 599.503 | 176.9393 | 39.6 | 0.07 |
| pHILIC | 599.503 | 83.2916 | 39.6 | 0.07 |
| pHILIC | 599.503 | 206.4721 | 39.6 | 0.07 |
| pHILIC | 599.503 | 476.9586 | 39.6 | 0.07 |
| pHILIC | 599.503 | 175.4039 | 39.6 | 0.07 |
| pHILIC | 599.503 | 80.8525 | 39.6 | 0.07 |
| pHILIC | 599.503 | 536.9528 | 39.6 | 0.07 |
| pHILIC | 599.503 | 132.358 | 39.6 | 0.07 |
| pHILIC | 599.503 | 145.6882 | 39.6 | 0.07 |
| pHILIC | 599.503 | 66.5933 | 39.6 | 0.07 |
| pHILIC | 599.503 | 90.8212 | 39.6 | 0.07 |
| pHILIC | 599.503 | 204.0021 | 39.5 | 0.07 |
| pHILIC | 599.503 | 68.5236 | 39.5 | 0.07 |
| pHILIC | 599.503 | 131.3638 | 39.5 | 0.07 |
| pHILIC | 599.503 | 67.1573 | 39.5 | 0.07 |

| | | | | |
|---|---|---|---|---|
| pHILIC | 599.503 | 121.9566 | 39.5 | 0.07 |
| pHILIC | 599.503 | 85.4658 | 39.5 | 0.07 |
| pHILIC | 599.503 | 103.563 | 39.5 | 0.07 |
| pHILIC | 599.503 | 94.3452 | 39.5 | 0.07 |
| pHILIC | 599.503 | 97.7399 | 39.5 | 0.07 |
| pHILIC | 599.503 | 405.935 | 39.5 | 0.07 |
| pHILIC | 599.503 | 67.9877 | 39.5 | 0.07 |
| pHILIC | 599.503 | 220.6869 | 39.5 | 0.07 |
| pHILIC | 599.503 | 101.0756 | 39.5 | 0.07 |
| pHILIC | 599.503 | 483.095 | 39.5 | 0.07 |
| pHILIC | 599.503 | 135.92 | 39.5 | 0.07 |
| pHILIC | 599.503 | 68.8462 | 39.4 | 0.07 |
| pHILIC | 599.503 | 203.1445 | 39.4 | 0.07 |
| pHILIC | 599.503 | 106.6436 | 39.4 | 0.07 |
| pHILIC | 599.503 | 66.6625 | 39.4 | 0.07 |
| pHILIC | 599.503 | 72.4977 | 39.4 | 0.07 |
| pHILIC | 599.503 | 142.3527 | 39.4 | 0.07 |
| pHILIC | 599.503 | 109.4149 | 39.4 | 0.07 |
| pHILIC | 599.503 | 215.6514 | 39.4 | 0.07 |
| pHILIC | 599.503 | 143.3032 | 39.4 | 0.07 |
| pHILIC | 599.503 | 72.6255 | 39.4 | 0.07 |
| pHILIC | 599.503 | 134.6465 | 39.3 | 0.07 |
| pHILIC | 599.503 | 105.7932 | 39.3 | 0.07 |
| pHILIC | 599.503 | 68.7812 | 39.3 | 0.07 |
| pHILIC | 599.503 | 127.585 | 39.3 | 0.07 |
| pHILIC | 599.503 | 191.4183 | 39.3 | 0.07 |
| pHILIC | 599.503 | 242.7762 | 39.3 | 0.07 |
| pHILIC | 599.503 | 73.5631 | 39.3 | 0.07 |
| pHILIC | 599.503 | 95.0976 | 39.3 | 0.07 |
| pHILIC | 599.503 | 212.065 | 39.3 | 0.07 |
| pHILIC | 599.503 | 97.158 | 39.2 | 0.07 |
| pHILIC | 599.503 | 164.18 | 39.2 | 0.07 |
| pHILIC | 599.503 | 86.3412 | 39.2 | 0.07 |
| pHILIC | 599.503 | 203.2142 | 39.2 | 0.07 |
| pHILIC | 599.503 | 110.589 | 39.2 | 0.07 |
| pHILIC | 599.503 | 78.259 | 39.2 | 0.07 |
| pHILIC | 599.503 | 191.1451 | 39.2 | 0.07 |
| pHILIC | 599.503 | 622.115 | 39.2 | 0.07 |
| pHILIC | 599.503 | 71.9664 | 39.2 | 0.07 |
| pHILIC | 599.503 | 157.6508 | 39.2 | 0.07 |
| pHILIC | 599.503 | 327.9791 | 39.2 | 0.07 |
| pHILIC | 599.503 | 586.0413 | 39.2 | 0.07 |

| | | | | |
|---|---|---|---|---|
| pHILIC | 599.503 | 78.8769 | 39.1 | 0.07 |
| pHILIC | 599.503 | 509.3279 | 39.1 | 0.07 |
| pHILIC | 599.503 | 264.4865 | 39.1 | 0.07 |
| pHILIC | 599.503 | 65.6014 | 39.1 | 0.07 |
| pHILIC | 599.503 | 328.689 | 39.1 | 0.07 |
| pHILIC | 599.503 | 71.1778 | 39.1 | 0.07 |
| pHILIC | 599.503 | 111.0558 | 39.1 | 0.07 |
| pHILIC | 599.503 | 251.6371 | 39 | 0.07 |
| pHILIC | 599.503 | 103.8735 | 39 | 0.07 |
| pHILIC | 599.503 | 300.6282 | 39 | 0.07 |
| pHILIC | 599.503 | 67.5176 | 39 | 0.07 |
| pHILIC | 599.503 | 369.3186 | 39 | 0.07 |
| pHILIC | 599.503 | 148.4011 | 39 | 0.07 |
| pHILIC | 599.503 | 91.7267 | 38.9 | 0.07 |
| pHILIC | 599.503 | 84.9334 | 38.9 | 0.07 |
| pHILIC | 599.503 | 70.2299 | 38.9 | 0.07 |
| pHILIC | 599.503 | 69.7129 | 38.9 | 0.07 |
| pHILIC | 599.503 | 92.7456 | 38.9 | 0.07 |
| pHILIC | 599.503 | 126.5103 | 38.9 | 0.07 |
| pHILIC | 599.503 | 161.8013 | 38.9 | 0.07 |
| pHILIC | 599.503 | 131.7205 | 38.9 | 0.07 |
| pHILIC | 599.503 | 87.1524 | 38.9 | 0.07 |
| pHILIC | 599.503 | 102.1394 | 38.8 | 0.07 |
| pHILIC | 599.503 | 390.7925 | 38.8 | 0.07 |
| pHILIC | 599.503 | 128.0721 | 38.8 | 0.07 |
| pHILIC | 599.503 | 188.3095 | 38.8 | 0.07 |
| pHILIC | 599.503 | 552.604 | 38.8 | 0.07 |
| pHILIC | 599.503 | 273.7695 | 38.8 | 0.07 |
| pHILIC | 599.503 | 67.968 | 38.8 | 0.07 |
| pHILIC | 599.503 | 74.032 | 38.8 | 0.07 |
| pHILIC | 599.503 | 64.5882 | 38.8 | 0.07 |
| pHILIC | 599.503 | 170.2255 | 38.8 | 0.07 |
| pHILIC | 599.503 | 86.8714 | 38.8 | 0.07 |
| pHILIC | 599.503 | 114.8937 | 38.8 | 0.07 |
| pHILIC | 599.503 | 182.9007 | 38.8 | 0.07 |
| pHILIC | 599.503 | 125.9231 | 38.7 | 0.07 |
| pHILIC | 599.503 | 94.6894 | 38.7 | 0.07 |
| pHILIC | 599.503 | 344.6043 | 38.7 | 0.07 |
| pHILIC | 599.503 | 138.1208 | 38.7 | 0.07 |
| pHILIC | 599.503 | 112.5313 | 38.7 | 0.07 |
| pHILIC | 599.503 | 67.5828 | 38.7 | 0.07 |
| pHILIC | 599.503 | 74.4874 | 38.7 | 0.07 |

| | | | | |
|---|---|---|---|---|
| pHILIC | 599.503 | 310.5349 | 38.7 | 0.07 |
| pHILIC | 599.503 | 67.7405 | 38.7 | 0.07 |
| pHILIC | 599.503 | 101.4915 | 38.6 | 0.07 |
| pHILIC | 599.503 | 115.9024 | 38.6 | 0.07 |
| pHILIC | 599.503 | 84.6344 | 38.6 | 0.07 |
| pHILIC | 599.503 | 67.2221 | 38.6 | 0.07 |
| pHILIC | 599.503 | 90.1518 | 38.6 | 0.07 |
| pHILIC | 599.503 | 66.5836 | 38.6 | 0.07 |
| pHILIC | 599.503 | 70.3428 | 38.5 | 0.07 |
| pHILIC | 599.503 | 84.6998 | 38.5 | 0.07 |
| pHILIC | 599.503 | 104.1627 | 38.5 | 0.07 |
| pHILIC | 599.503 | 123.9702 | 38.5 | 0.07 |
| pHILIC | 599.503 | 114.1545 | 38.5 | 0.07 |
| pHILIC | 599.503 | 196.6951 | 38.5 | 0.07 |
| pHILIC | 599.503 | 422.1819 | 38.5 | 0.07 |
| pHILIC | 599.503 | 191.5233 | 38.4 | 0.07 |
| pHILIC | 599.503 | 272.5774 | 38.4 | 0.07 |
| pHILIC | 599.503 | 138.4169 | 38.4 | 0.07 |
| pHILIC | 599.503 | 68.6033 | 38.3 | 0.07 |
| pHILIC | 599.503 | 494.7469 | 38.3 | 0.07 |
| pHILIC | 599.503 | 73.0477 | 38.3 | 0.07 |
| pHILIC | 599.503 | 84.1253 | 38.3 | 0.07 |
| pHILIC | 599.503 | 67.8502 | 38.2 | 0.07 |
| pHILIC | 599.503 | 151.3717 | 38.2 | 0.07 |
| pHILIC | 599.503 | 127.9607 | 38.2 | 0.07 |
| pHILIC | 599.503 | 118.6215 | 38.2 | 0.07 |
| pHILIC | 599.503 | 466.0071 | 38.2 | 0.07 |
| pHILIC | 599.503 | 135.8158 | 38.2 | 0.07 |
| pHILIC | 599.503 | 88.4449 | 38.2 | 0.07 |
| pHILIC | 599.503 | 308.6832 | 38.2 | 0.07 |
| pHILIC | 599.503 | 82.733 | 38.1 | 0.07 |
| pHILIC | 599.503 | 91.8274 | 38.1 | 0.07 |
| pHILIC | 599.503 | 90.706 | 38.1 | 0.07 |
| pHILIC | 599.503 | 366.4561 | 38.1 | 0.07 |
| pHILIC | 599.503 | 291.5787 | 38.1 | 0.07 |
| pHILIC | 599.503 | 71.1735 | 38 | 0.07 |
| pHILIC | 599.503 | 155.793 | 38 | 0.07 |
| pHILIC | 599.503 | 77.8156 | 38 | 0.07 |
| pHILIC | 599.503 | 164.0561 | 38 | 0.07 |
| pHILIC | 599.503 | 74.6482 | 38 | 0.07 |
| pHILIC | 599.503 | 276.8792 | 37.9 | 0.07 |
| pHILIC | 599.503 | 414.49 | 37.9 | 0.07 |

| | | | | |
|---|---|---|---|---|
| pHILIC | 599.503 | 393.7793 | 37.9 | 0.07 |
| pHILIC | 599.503 | 106.4487 | 37.9 | 0.07 |
| pHILIC | 599.503 | 238.8653 | 37.9 | 0.07 |
| pHILIC | 599.503 | 170.1911 | 37.9 | 0.07 |
| pHILIC | 599.503 | 104.2291 | 37.8 | 0.07 |
| pHILIC | 599.503 | 66.5172 | 37.8 | 0.07 |
| pHILIC | 599.503 | 140.7215 | 37.8 | 0.07 |
| pHILIC | 599.503 | 128.959 | 37.8 | 0.07 |
| pHILIC | 599.503 | 178.9085 | 37.7 | 0.07 |
| pHILIC | 599.503 | 218.9028 | 37.7 | 0.07 |
| pHILIC | 599.503 | 87.5658 | 37.7 | 0.07 |
| pHILIC | 599.503 | 77.8221 | 37.7 | 0.07 |
| pHILIC | 599.503 | 84.1607 | 37.6 | 0.07 |
| pHILIC | 599.503 | 82.3183 | 37.6 | 0.07 |
| pHILIC | 599.503 | 164.3839 | 37.6 | 0.07 |
| pHILIC | 599.503 | 203.6093 | 37.6 | 0.07 |
| pHILIC | 599.503 | 75.2558 | 37.6 | 0.07 |
| pHILIC | 599.503 | 69.8908 | 37.5 | 0.07 |
| pHILIC | 599.503 | 85.6188 | 37.5 | 0.07 |
| pHILIC | 599.503 | 65.5394 | 37.5 | 0.07 |
| pHILIC | 599.503 | 279.1889 | 37.5 | 0.07 |
| pHILIC | 599.503 | 116.8443 | 37.5 | 0.07 |
| pHILIC | 599.503 | 73.1766 | 37.5 | 0.07 |
| pHILIC | 599.503 | 584.7268 | 37.5 | 0.07 |
| pHILIC | 599.503 | 266.4648 | 37.4 | 0.07 |
| pHILIC | 599.503 | 565.8334 | 37.4 | 0.07 |
| pHILIC | 599.503 | 70.9211 | 37.4 | 0.07 |
| pHILIC | 599.503 | 344.088 | 37.3 | 0.07 |
| pHILIC | 599.503 | 418.5894 | 37.3 | 0.07 |
| pHILIC | 599.503 | 120.3066 | 37.3 | 0.07 |
| pHILIC | 599.503 | 156.7185 | 37.3 | 0.07 |
| pHILIC | 599.503 | 475.6693 | 37.2 | 0.07 |
| pHILIC | 599.503 | 73.9987 | 37.2 | 0.07 |
| pHILIC | 599.503 | 91.0397 | 37.2 | 0.07 |
| pHILIC | 599.503 | 136.8479 | 37.2 | 0.07 |
| pHILIC | 599.503 | 169.0372 | 37.1 | 0.07 |
| pHILIC | 599.503 | 83.0555 | 37.1 | 0.07 |
| pHILIC | 599.503 | 197.2729 | 37 | 0.06 |
| pHILIC | 599.503 | 519.0257 | 37 | 0.06 |
| pHILIC | 599.503 | 99.9918 | 36.9 | 0.06 |
| pHILIC | 599.503 | 297.767 | 36.9 | 0.06 |
| pHILIC | 599.503 | 68.3272 | 36.9 | 0.06 |

| | | | | |
|---|---|---|---|---|
| pHILIC | 599.503 | 69.5769 | 36.8 | 0.06 |
| pHILIC | 599.503 | 70.9433 | 36.8 | 0.06 |
| pHILIC | 599.503 | 63.2756 | 36.8 | 0.06 |
| pHILIC | 599.503 | 65.5547 | 36.8 | 0.06 |
| pHILIC | 599.503 | 97.1385 | 36.8 | 0.06 |
| pHILIC | 599.503 | 73.1295 | 36.8 | 0.06 |
| pHILIC | 599.503 | 73.9853 | 36.8 | 0.06 |
| pHILIC | 599.503 | 177.1429 | 36.7 | 0.06 |
| pHILIC | 599.503 | 130.5077 | 36.7 | 0.06 |
| pHILIC | 599.503 | 149.6717 | 36.7 | 0.06 |
| pHILIC | 599.503 | 93.2189 | 36.6 | 0.06 |
| pHILIC | 599.503 | 96.7105 | 36.6 | 0.06 |
| pHILIC | 599.503 | 75.1034 | 36.5 | 0.06 |
| pHILIC | 599.503 | 422.4036 | 36.5 | 0.06 |
| pHILIC | 599.503 | 92.4842 | 36.5 | 0.06 |
| pHILIC | 599.503 | 328.3015 | 36.4 | 0.06 |
| pHILIC | 599.503 | 172.1276 | 36.4 | 0.06 |
| pHILIC | 599.503 | 198.7272 | 36.4 | 0.06 |
| pHILIC | 599.503 | 133.6374 | 36.4 | 0.06 |
| pHILIC | 599.503 | 103.7743 | 36.3 | 0.06 |
| pHILIC | 599.503 | 294.2636 | 36.3 | 0.06 |
| pHILIC | 599.503 | 83.6944 | 36.3 | 0.06 |
| pHILIC | 599.503 | 579.6819 | 36.3 | 0.06 |
| pHILIC | 599.503 | 139.037 | 36.2 | 0.06 |
| pHILIC | 599.503 | 90.9375 | 36.2 | 0.06 |
| pHILIC | 599.503 | 195.7677 | 36.1 | 0.06 |
| pHILIC | 599.503 | 123.8973 | 36.1 | 0.06 |
| pHILIC | 599.503 | 77.0199 | 36.1 | 0.06 |
| pHILIC | 599.503 | 619.8489 | 36 | 0.06 |
| pHILIC | 599.503 | 107.3715 | 35.9 | 0.06 |
| pHILIC | 599.503 | 195.31 | 35.9 | 0.06 |
| pHILIC | 599.503 | 68.8923 | 35.9 | 0.06 |
| pHILIC | 599.503 | 108.8631 | 35.8 | 0.06 |
| pHILIC | 599.503 | 441.9557 | 35.6 | 0.06 |
| pHILIC | 599.503 | 296.9457 | 35.6 | 0.06 |
| pHILIC | 599.503 | 95.5595 | 35.6 | 0.06 |
| pHILIC | 599.503 | 85.1158 | 35.6 | 0.06 |
| pHILIC | 599.503 | 155.1417 | 35.6 | 0.06 |
| pHILIC | 599.503 | 84.8985 | 35.4 | 0.06 |
| pHILIC | 599.503 | 93.8384 | 35.2 | 0.06 |
| pHILIC | 599.503 | 144.6317 | 35.2 | 0.06 |
| pHILIC | 599.503 | 88.1037 | 35 | 0.06 |

| | | | | |
|---|---|---|---|---|
| pHILIC | 599.503 | 356.6296 | 35 | 0.06 |
| pHILIC | 599.503 | 288.6735 | 35 | 0.06 |
| pHILIC | 599.503 | 85.0886 | 35 | 0.06 |
| pHILIC | 599.503 | 128.1768 | 34.9 | 0.06 |
| pHILIC | 599.503 | 158.5141 | 34.9 | 0.06 |
| pHILIC | 599.503 | 199.8315 | 34.9 | 0.06 |
| pHILIC | 599.503 | 74.8605 | 34.7 | 0.06 |
| pHILIC | 599.503 | 68.0018 | 34.7 | 0.06 |
| pHILIC | 599.503 | 67.1013 | 34.7 | 0.06 |
| pHILIC | 599.503 | 63.4979 | 34.6 | 0.06 |
| pHILIC | 599.503 | 72.8722 | 34.6 | 0.06 |
| pHILIC | 599.503 | 76.2708 | 34.6 | 0.06 |
| pHILIC | 599.503 | 220.8532 | 34.5 | 0.06 |
| pHILIC | 599.503 | 81.3596 | 34.5 | 0.06 |
| pHILIC | 599.503 | 199.9376 | 34.5 | 0.06 |
| pHILIC | 599.503 | 113.8155 | 34.4 | 0.06 |
| pHILIC | 599.503 | 84.2053 | 34.4 | 0.06 |
| pHILIC | 599.503 | 89.746 | 34.4 | 0.06 |
| pHILIC | 599.503 | 63.5156 | 34.3 | 0.06 |
| pHILIC | 599.503 | 403.6449 | 34.3 | 0.06 |
| pHILIC | 599.503 | 78.0397 | 34.2 | 0.06 |
| pHILIC | 599.503 | 116.0582 | 34.2 | 0.06 |
| pHILIC | 599.503 | 66.1354 | 34.2 | 0.06 |
| pHILIC | 599.503 | 164.605 | 34.1 | 0.06 |
| pHILIC | 599.503 | 171.6535 | 34.1 | 0.06 |
| pHILIC | 599.503 | 161.3452 | 34 | 0.06 |
| pHILIC | 599.503 | 317.3807 | 34 | 0.06 |
| pHILIC | 599.503 | 292.6143 | 33.9 | 0.06 |
| pHILIC | 599.503 | 71.2329 | 33.9 | 0.06 |
| pHILIC | 599.503 | 114.1402 | 33.8 | 0.06 |
| pHILIC | 599.503 | 630.886 | 33.6 | 0.06 |
| pHILIC | 599.503 | 73.5992 | 33.6 | 0.06 |
| pHILIC | 599.503 | 101.9625 | 33.5 | 0.06 |
| pHILIC | 599.503 | 442.3518 | 33.5 | 0.06 |
| pHILIC | 599.503 | 538.5088 | 33.4 | 0.06 |
| pHILIC | 599.503 | 375.1887 | 33.4 | 0.06 |
| pHILIC | 599.503 | 532.1351 | 33.4 | 0.06 |
| pHILIC | 599.503 | 75.1267 | 33.3 | 0.06 |
| pHILIC | 599.503 | 186.4857 | 33.3 | 0.06 |
| pHILIC | 599.503 | 77.719 | 33.2 | 0.06 |
| pHILIC | 599.503 | 296.321 | 33.2 | 0.06 |
| pHILIC | 599.503 | 65.2328 | 33.1 | 0.06 |

| | | | | |
|---|---|---|---|---|
| pHILIC | 599.503 | 86.721 | 33.1 | 0.06 |
| pHILIC | 599.503 | 176.2916 | 33.1 | 0.06 |
| pHILIC | 599.503 | 70.8362 | 33.1 | 0.06 |
| pHILIC | 599.503 | 282.391 | 33 | 0.06 |
| pHILIC | 599.503 | 385.1921 | 32.9 | 0.06 |
| pHILIC | 599.503 | 129.1832 | 32.9 | 0.06 |
| pHILIC | 599.503 | 72.1904 | 32.8 | 0.06 |
| pHILIC | 599.503 | 116.1814 | 32.7 | 0.06 |
| pHILIC | 599.503 | 67.8946 | 32.5 | 0.06 |
| pHILIC | 599.503 | 71.6132 | 32.3 | 0.06 |
| pHILIC | 599.503 | 117.4338 | 32.3 | 0.06 |
| pHILIC | 599.503 | 133.9668 | 32.1 | 0.06 |
| pHILIC | 599.503 | 158.9205 | 32.1 | 0.06 |
| pHILIC | 599.503 | 158.7972 | 32 | 0.06 |
| pHILIC | 599.503 | 507.2543 | 31.9 | 0.06 |
| pHILIC | 599.503 | 78.8254 | 31.8 | 0.06 |
| pHILIC | 599.503 | 78.4455 | 31.8 | 0.06 |
| pHILIC | 599.503 | 71.2815 | 31.8 | 0.06 |
| pHILIC | 599.503 | 123.0605 | 31.8 | 0.06 |
| pHILIC | 599.503 | 422.8604 | 31.7 | 0.06 |
| pHILIC | 599.503 | 201.9083 | 31.6 | 0.06 |
| pHILIC | 599.503 | 123.5668 | 31.6 | 0.06 |
| pHILIC | 599.503 | 78.3281 | 31.5 | 0.06 |
| pHILIC | 599.503 | 65.8281 | 31.5 | 0.06 |
| pHILIC | 599.503 | 68.1103 | 31.4 | 0.06 |
| pHILIC | 599.503 | 161.8339 | 31.4 | 0.06 |
| pHILIC | 599.503 | 156.1669 | 31.3 | 0.05 |
| pHILIC | 599.503 | 134.041 | 31.3 | 0.05 |
| pHILIC | 599.503 | 213.1552 | 31.2 | 0.05 |
| pHILIC | 599.503 | 77.5007 | 31.2 | 0.05 |
| pHILIC | 599.503 | 208.0419 | 31.2 | 0.05 |
| pHILIC | 599.503 | 74.6978 | 31.1 | 0.05 |
| pHILIC | 599.503 | 192.6296 | 30.7 | 0.05 |
| pHILIC | 599.503 | 67.1486 | 30.7 | 0.05 |
| pHILIC | 599.503 | 97.2255 | 30.7 | 0.05 |
| pHILIC | 599.503 | 74.3849 | 30.5 | 0.05 |
| pHILIC | 599.503 | 70.9641 | 30.3 | 0.05 |
| pHILIC | 599.503 | 366.1344 | 30.1 | 0.05 |
| pHILIC | 599.503 | 65.0781 | 30 | 0.05 |
| pHILIC | 599.503 | 109.1992 | 29.7 | 0.05 |
| pHILIC | 599.503 | 73.5565 | 29.4 | 0.05 |
| pHILIC | 599.503 | 81.9642 | 29.2 | 0.05 |

| | | | | |
|---|---|---|---|---|
| pHILIC | 599.503 | 246.8331 | 29.1 | 0.05 |
| pHILIC | 599.503 | 82.2897 | 28.5 | 0.05 |
| pHILIC | 686.437 | 413.3068 | 32279.1 | 100 |
| pHILIC | 686.437 | 251.0911 | 22684.3 | 70.28 |
| pHILIC | 686.437 | 101.0403 | 20723 | 64.2 |
| pHILIC | 686.437 | 651.5355 | 20612.2 | 63.86 |
| pHILIC | 686.437 | 73.0467 | 19448.3 | 60.25 |
| pHILIC | 686.437 | 325.1643 | 18070.4 | 55.98 |
| pHILIC | 686.437 | 269.1018 | 17263.9 | 53.48 |
| pHILIC | 686.437 | 445.185 | 14954.1 | 46.33 |
| pHILIC | 686.437 | 205.0858 | 13099.9 | 40.58 |
| pHILIC | 686.437 | 157.0666 | 12537.8 | 38.84 |
| pHILIC | 686.437 | 431.0864 | 11287.1 | 34.97 |
| pHILIC | 686.437 | 279.1226 | 9951.7 | 30.83 |
| pHILIC | 686.437 | 233.0809 | 7122.7 | 22.07 |
| pHILIC | 686.437 | 187.0751 | 6333.8 | 19.62 |
| pHILIC | 686.437 | 107.0491 | 6206 | 19.23 |
| pHILIC | 686.437 | 313.2736 | 6027.7 | 18.67 |
| pHILIC | 686.437 | 219.1743 | 5924.1 | 18.35 |
| pHILIC | 686.437 | 149.0448 | 5580.4 | 17.29 |
| pHILIC | 686.437 | 159.0803 | 5564.4 | 17.24 |
| pHILIC | 686.437 | 551.5036 | 4895.4 | 15.17 |
| pHILIC | 686.437 | 429.0873 | 4222.3 | 13.08 |
| pHILIC | 686.437 | 95.0855 | 4027.9 | 12.48 |
| pHILIC | 686.437 | 163.1117 | 3973.2 | 12.31 |
| pHILIC | 686.437 | 686.2016 | 3963.2 | 12.28 |
| pHILIC | 686.437 | 686.4675 | 3917.3 | 12.14 |
| pHILIC | 686.437 | 225.0427 | 3795.9 | 11.76 |
| pHILIC | 686.437 | 430.0916 | 3663.5 | 11.35 |
| pHILIC | 686.437 | 357.0675 | 3137.2 | 9.72 |
| pHILIC | 686.437 | 81.0699 | 3102.6 | 9.61 |
| pHILIC | 686.437 | 149.0598 | 3037.2 | 9.41 |
| pHILIC | 686.437 | 131.0857 | 2918.3 | 9.04 |
| pHILIC | 686.437 | 355.0703 | 2909.3 | 9.01 |
| pHILIC | 686.437 | 501.2469 | 2841.3 | 8.8 |
| pHILIC | 686.437 | 221.0841 | 2729 | 8.45 |
| pHILIC | 686.437 | 359.028 | 2657.4 | 8.23 |
| pHILIC | 686.437 | 361.0262 | 2487.1 | 7.71 |
| pHILIC | 686.437 | 91.0572 | 2368.4 | 7.34 |
| pHILIC | 686.437 | 177.091 | 2323.5 | 7.2 |
| pHILIC | 686.437 | 83.0491 | 2310.4 | 7.16 |
| pHILIC | 686.437 | 105.0699 | 2228.5 | 6.9 |

| | | | | |
|---|---|---|---|---|
| pHILIC | 686.437 | 133.1013 | 2116.3 | 6.56 |
| pHILIC | 686.437 | 223.0967 | 2015.7 | 6.24 |
| pHILIC | 686.437 | 437.2898 | 1994.8 | 6.18 |
| pHILIC | 686.437 | 161.096 | 1869.2 | 5.79 |
| pHILIC | 686.437 | 381.227 | 1704.1 | 5.28 |
| pHILIC | 686.437 | 335.1856 | 1670.5 | 5.18 |
| pHILIC | 686.437 | 285.0101 | 1626.1 | 5.04 |
| pHILIC | 686.437 | 100.0441 | 1620.9 | 5.02 |
| pHILIC | 686.437 | 343.0156 | 1459.6 | 4.52 |
| pHILIC | 686.437 | 109.1012 | 1455.4 | 4.51 |
| pHILIC | 686.437 | 341.0181 | 1448.9 | 4.49 |
| pHILIC | 686.437 | 356.0703 | 1398.3 | 4.33 |
| pHILIC | 686.437 | 189.0911 | 1336.6 | 4.14 |
| pHILIC | 686.437 | 241.0699 | 1270.5 | 3.94 |
| pHILIC | 686.437 | 85.1011 | 1269.6 | 3.93 |
| pHILIC | 686.437 | 215.0699 | 1229.4 | 3.81 |
| pHILIC | 686.437 | 83.0855 | 1219 | 3.78 |
| pHILIC | 686.437 | 239.0951 | 1146.2 | 3.55 |
| pHILIC | 686.437 | 75.0436 | 1132.7 | 3.51 |
| pHILIC | 686.437 | 360.0286 | 1106.4 | 3.43 |
| pHILIC | 686.437 | 147.0803 | 996.5 | 3.09 |
| pHILIC | 686.437 | 307.1539 | 940 | 2.91 |
| pHILIC | 686.437 | 114.0734 | 895.2 | 2.77 |
| pHILIC | 686.437 | 299.0616 | 885.5 | 2.74 |
| pHILIC | 686.437 | 287.0071 | 858.1 | 2.66 |
| pHILIC | 686.437 | 281.0513 | 792 | 2.45 |
| pHILIC | 686.437 | 205.1226 | 774.4 | 2.4 |
| pHILIC | 686.437 | 342.0183 | 756 | 2.34 |
| pHILIC | 686.437 | 167.0549 | 754.8 | 2.34 |
| pHILIC | 686.437 | 227.0409 | 746.1 | 2.31 |
| pHILIC | 686.437 | 121.0648 | 741.8 | 2.3 |
| pHILIC | 686.437 | 412.3107 | 734 | 2.27 |
| pHILIC | 686.437 | 326.9848 | 693.2 | 2.15 |
| pHILIC | 686.437 | 93.0697 | 691.9 | 2.14 |
| pHILIC | 686.437 | 97.1012 | 673.3 | 2.09 |
| pHILIC | 686.437 | 104.1069 | 663.4 | 2.06 |
| pHILIC | 686.437 | 367.1748 | 657.6 | 2.04 |
| pHILIC | 686.437 | 226.0435 | 649.7 | 2.01 |
| pHILIC | 686.437 | 311.1119 | 606.2 | 1.88 |
| pHILIC | 686.437 | 151.0415 | 596 | 1.85 |
| pHILIC | 686.437 | 381.1596 | 575.9 | 1.78 |
| pHILIC | 686.437 | 229.1227 | 536.5 | 1.66 |

| | | | | |
|---|---|---|---|---|
| pHILIC | 686.437 | 74.0461 | 534.3 | 1.66 |
| pHILIC | 686.437 | 89.0595 | 525.5 | 1.63 |
| pHILIC | 686.437 | 222.0847 | 515.9 | 1.6 |
| pHILIC | 686.437 | 176.8754 | 514.3 | 1.59 |
| pHILIC | 686.437 | 534.2917 | 511.2 | 1.58 |
| pHILIC | 686.437 | 195.0648 | 469.8 | 1.46 |
| pHILIC | 686.437 | 97.0285 | 461.6 | 1.43 |
| pHILIC | 686.437 | 151.0739 | 452 | 1.4 |
| pHILIC | 686.437 | 156.07 | 446.7 | 1.38 |
| pHILIC | 686.437 | 301.0607 | 403.8 | 1.25 |
| pHILIC | 686.437 | 150.0453 | 400 | 1.24 |
| pHILIC | 686.437 | 286.0099 | 385.9 | 1.2 |
| pHILIC | 686.437 | 169.0495 | 383.2 | 1.19 |
| pHILIC | 686.437 | 143.086 | 380.4 | 1.18 |
| pHILIC | 686.437 | 315.143 | 327.2 | 1.01 |
| pHILIC | 686.437 | 402.9831 | 325.6 | 1.01 |
| pHILIC | 686.437 | 101.0485 | 310.5 | 0.96 |
| pHILIC | 686.437 | 223.0605 | 310.1 | 0.96 |
| pHILIC | 686.437 | 324.9859 | 289 | 0.9 |
| pHILIC | 686.437 | 557.3139 | 284.5 | 0.88 |
| pHILIC | 686.437 | 325.2101 | 273.9 | 0.85 |
| pHILIC | 686.437 | 123.1167 | 270 | 0.84 |
| pHILIC | 686.437 | 669.5476 | 261.7 | 0.81 |
| pHILIC | 686.437 | 357.2797 | 254.9 | 0.79 |
| pHILIC | 686.437 | 95.0488 | 239.3 | 0.74 |
| pHILIC | 686.437 | 117.0712 | 218.2 | 0.68 |
| pHILIC | 686.437 | 127.0558 | 217.7 | 0.67 |
| pHILIC | 686.437 | 552.5381 | 213.6 | 0.66 |
| pHILIC | 686.437 | 141.0695 | 211.6 | 0.66 |
| pHILIC | 686.437 | 268.9832 | 208.4 | 0.65 |
| pHILIC | 686.437 | 112.8959 | 207.7 | 0.64 |
| pHILIC | 686.437 | 147.0661 | 200.4 | 0.62 |
| pHILIC | 686.437 | 123.0804 | 199.7 | 0.62 |
| pHILIC | 686.437 | 105.0336 | 191 | 0.59 |
| pHILIC | 686.437 | 415.1746 | 189.4 | 0.59 |
| pHILIC | 686.437 | 497.3059 | 189.3 | 0.59 |
| pHILIC | 686.437 | 395.1662 | 187.3 | 0.58 |
| pHILIC | 686.437 | 289.1452 | 187.1 | 0.58 |
| pHILIC | 686.437 | 609.4348 | 184.9 | 0.57 |
| pHILIC | 686.437 | 207.1022 | 183.6 | 0.57 |
| pHILIC | 686.437 | 481.7648 | 173.9 | 0.54 |
| pHILIC | 686.437 | 133.0649 | 171.6 | 0.53 |

| | | | | |
|---|---|---|---|---|
| pHILIC | 686.437 | 119.0857 | 169 | 0.52 |
| pHILIC | 686.437 | 167.0706 | 164.9 | 0.51 |
| pHILIC | 686.437 | 239.2358 | 163.3 | 0.51 |
| pHILIC | 686.437 | 145.0651 | 161.9 | 0.5 |
| pHILIC | 686.437 | 257.1156 | 159.1 | 0.49 |
| pHILIC | 686.437 | 503.9213 | 154.3 | 0.48 |
| pHILIC | 686.437 | 139.0583 | 152.3 | 0.47 |
| pHILIC | 686.437 | 300.0611 | 151.2 | 0.47 |
| pHILIC | 686.437 | 115.0757 | 150.1 | 0.47 |
| pHILIC | 686.437 | 184.0727 | 148.6 | 0.46 |
| pHILIC | 686.437 | 177.0545 | 144.1 | 0.45 |
| pHILIC | 686.437 | 346.6185 | 143 | 0.44 |
| pHILIC | 686.437 | 107.0857 | 141.8 | 0.44 |
| pHILIC | 686.437 | 173.0811 | 141.7 | 0.44 |
| pHILIC | 686.437 | 93.8217 | 141.5 | 0.44 |
| pHILIC | 686.437 | 82.5584 | 136.8 | 0.42 |
| pHILIC | 686.437 | 79.6038 | 135 | 0.42 |
| pHILIC | 686.437 | 287.4526 | 134 | 0.42 |
| pHILIC | 686.437 | 295.5064 | 133.9 | 0.41 |
| pHILIC | 686.437 | 149.0939 | 133 | 0.41 |
| pHILIC | 686.437 | 124.9749 | 132 | 0.41 |
| pHILIC | 686.437 | 626.6223 | 130.2 | 0.4 |
| pHILIC | 686.437 | 100.8689 | 129.6 | 0.4 |
| pHILIC | 686.437 | 697.6124 | 128.7 | 0.4 |
| pHILIC | 686.437 | 358.1779 | 128.2 | 0.4 |
| pHILIC | 686.437 | 245.1261 | 128.1 | 0.4 |
| pHILIC | 686.437 | 402.4922 | 128 | 0.4 |
| pHILIC | 686.437 | 267.453 | 127.8 | 0.4 |
| pHILIC | 686.437 | 116.5684 | 126.2 | 0.39 |
| pHILIC | 686.437 | 436.3858 | 125.9 | 0.39 |
| pHILIC | 686.437 | 302.6664 | 125.7 | 0.39 |
| pHILIC | 686.437 | 74.1788 | 125.6 | 0.39 |
| pHILIC | 686.437 | 129.9662 | 125.5 | 0.39 |
| pHILIC | 686.437 | 487.7503 | 124.9 | 0.39 |
| pHILIC | 686.437 | 425.8938 | 124.9 | 0.39 |
| pHILIC | 686.437 | 136.7708 | 123.9 | 0.38 |
| pHILIC | 686.437 | 154.6173 | 123.7 | 0.38 |
| pHILIC | 686.437 | 114.9021 | 123.4 | 0.38 |
| pHILIC | 686.437 | 72.543 | 122.8 | 0.38 |
| pHILIC | 686.437 | 79.6933 | 121.8 | 0.38 |
| pHILIC | 686.437 | 98.9387 | 121.5 | 0.38 |
| pHILIC | 686.437 | 702.3206 | 121 | 0.37 |

| | | | | |
|---|---|---|---|---|
| pHILIC | 686.437 | 509.0594 | 120.8 | 0.37 |
| pHILIC | 686.437 | 209.9755 | 120.6 | 0.37 |
| pHILIC | 686.437 | 477.2028 | 119.7 | 0.37 |
| pHILIC | 686.437 | 259.55 | 119.3 | 0.37 |
| pHILIC | 686.437 | 210.5622 | 118.6 | 0.37 |
| pHILIC | 686.437 | 404.9633 | 118.3 | 0.37 |
| pHILIC | 686.437 | 300.5492 | 117.9 | 0.37 |
| pHILIC | 686.437 | 169.2432 | 117.4 | 0.36 |
| pHILIC | 686.437 | 170.5539 | 117.2 | 0.36 |
| pHILIC | 686.437 | 134.0815 | 114.9 | 0.36 |
| pHILIC | 686.437 | 102.2519 | 114.3 | 0.35 |
| pHILIC | 686.437 | 155.4595 | 114.2 | 0.35 |
| pHILIC | 686.437 | 243.0345 | 113 | 0.35 |
| pHILIC | 686.437 | 193.4546 | 112.8 | 0.35 |
| pHILIC | 686.437 | 80.557 | 112.5 | 0.35 |
| pHILIC | 686.437 | 602.5937 | 111.8 | 0.35 |
| pHILIC | 686.437 | 307.834 | 110.8 | 0.34 |
| pHILIC | 686.437 | 394.1723 | 110.6 | 0.34 |
| pHILIC | 686.437 | 94.1553 | 110.1 | 0.34 |
| pHILIC | 686.437 | 621.1747 | 110.1 | 0.34 |
| pHILIC | 686.437 | 324.4536 | 110 | 0.34 |
| pHILIC | 686.437 | 163.1538 | 109.8 | 0.34 |
| pHILIC | 686.437 | 461.5158 | 108.7 | 0.34 |
| pHILIC | 686.437 | 266.9993 | 108.2 | 0.34 |
| pHILIC | 686.437 | 604.1941 | 107.8 | 0.33 |
| pHILIC | 686.437 | 457.493 | 107.8 | 0.33 |
| pHILIC | 686.437 | 665.8966 | 107.4 | 0.33 |
| pHILIC | 686.437 | 167.9606 | 107.2 | 0.33 |
| pHILIC | 686.437 | 93.4769 | 103.8 | 0.32 |
| pHILIC | 686.437 | 80.5227 | 101.8 | 0.32 |
| pHILIC | 686.437 | 269.6625 | 100.3 | 0.31 |
| pHILIC | 686.437 | 270.0294 | 100.2 | 0.31 |
| pHILIC | 686.437 | 303.1214 | 99.5 | 0.31 |
| pHILIC | 686.437 | 197.0802 | 99 | 0.31 |
| pHILIC | 686.437 | 297.1348 | 98.3 | 0.3 |
| pHILIC | 686.437 | 157.0816 | 94.1 | 0.29 |
| pHILIC | 686.437 | 536.9205 | 88.4 | 0.27 |
| pHILIC | 686.437 | 283.035 | 87 | 0.27 |
| pHILIC | 686.437 | 109.0647 | 86.1 | 0.27 |
| pHILIC | 686.437 | 367.4804 | 84.9 | 0.26 |
| pHILIC | 686.437 | 275.1255 | 83.2 | 0.26 |
| pHILIC | 686.437 | 373.3326 | 83 | 0.26 |

| | | | | |
|---|---|---|---|---|
| pHILIC | 686.437 | 74.0505 | 81.8 | 0.25 |
| pHILIC | 686.437 | 241.096 | 81.1 | 0.25 |
| pHILIC | 686.437 | 438.1194 | 80.7 | 0.25 |
| pHILIC | 686.437 | 346.8885 | 80.6 | 0.25 |
| pHILIC | 686.437 | 304.936 | 79.3 | 0.25 |
| pHILIC | 686.437 | 447.1149 | 79.3 | 0.25 |
| pHILIC | 686.437 | 163.0746 | 78.3 | 0.24 |
| pHILIC | 686.437 | 480.668 | 78.1 | 0.24 |
| pHILIC | 686.437 | 106.3907 | 77.9 | 0.24 |
| pHILIC | 686.437 | 389.4245 | 77.8 | 0.24 |
| pHILIC | 686.437 | 111.324 | 77.6 | 0.24 |
| pHILIC | 686.437 | 168.6205 | 77.3 | 0.24 |
| pHILIC | 686.437 | 103.8066 | 77.1 | 0.24 |
| pHILIC | 686.437 | 257.2623 | 76.5 | 0.24 |
| pHILIC | 686.437 | 608.8656 | 76.4 | 0.24 |
| pHILIC | 686.437 | 93.9954 | 76.2 | 0.24 |
| pHILIC | 686.437 | 385.1818 | 76 | 0.24 |
| pHILIC | 686.437 | 80.8678 | 75.9 | 0.24 |
| pHILIC | 686.437 | 514.9222 | 75.9 | 0.24 |
| pHILIC | 686.437 | 306.7921 | 75.8 | 0.23 |
| pHILIC | 686.437 | 104.4082 | 75.8 | 0.23 |
| pHILIC | 686.437 | 511.6217 | 75.6 | 0.23 |
| pHILIC | 686.437 | 90.7229 | 75.4 | 0.23 |
| pHILIC | 686.437 | 159.8413 | 75.4 | 0.23 |
| pHILIC | 686.437 | 478.3863 | 75.2 | 0.23 |
| pHILIC | 686.437 | 431.4128 | 75.1 | 0.23 |
| pHILIC | 686.437 | 199.2926 | 75.1 | 0.23 |
| pHILIC | 686.437 | 216.394 | 75 | 0.23 |
| pHILIC | 686.437 | 262.1337 | 74.9 | 0.23 |
| pHILIC | 686.437 | 451.6324 | 74.9 | 0.23 |
| pHILIC | 686.437 | 256.6695 | 74.8 | 0.23 |
| pHILIC | 686.437 | 81.9 | 74.8 | 0.23 |
| pHILIC | 686.437 | 323.2282 | 74.6 | 0.23 |
| pHILIC | 686.437 | 137.0847 | 74.5 | 0.23 |
| pHILIC | 686.437 | 344.1927 | 74.4 | 0.23 |
| pHILIC | 686.437 | 431.243 | 74.3 | 0.23 |
| pHILIC | 686.437 | 249.968 | 74.2 | 0.23 |
| pHILIC | 686.437 | 541.4356 | 74.2 | 0.23 |
| pHILIC | 686.437 | 601.1548 | 74.1 | 0.23 |
| pHILIC | 686.437 | 212.0669 | 74 | 0.23 |
| pHILIC | 686.437 | 104.3076 | 74 | 0.23 |
| pHILIC | 686.437 | 115.372 | 74 | 0.23 |

| | | | | |
|---|---|---|---|---|
| pHILIC | 686.437 | 90.2754 | 73.9 | 0.23 |
| pHILIC | 686.437 | 181.4156 | 73.9 | 0.23 |
| pHILIC | 686.437 | 252.0784 | 73.9 | 0.23 |
| pHILIC | 686.437 | 132.9472 | 73.8 | 0.23 |
| pHILIC | 686.437 | 601.4517 | 73.8 | 0.23 |
| pHILIC | 686.437 | 302.7886 | 73.7 | 0.23 |
| pHILIC | 686.437 | 75.404 | 73.6 | 0.23 |
| pHILIC | 686.437 | 496.2797 | 73.3 | 0.23 |
| pHILIC | 686.437 | 354.237 | 73.2 | 0.23 |
| pHILIC | 686.437 | 183.1646 | 73.2 | 0.23 |
| pHILIC | 686.437 | 208.9714 | 73.1 | 0.23 |
| pHILIC | 686.437 | 93.7558 | 73 | 0.23 |
| pHILIC | 686.437 | 458.7545 | 72.9 | 0.23 |
| pHILIC | 686.437 | 158.4341 | 72.9 | 0.23 |
| pHILIC | 686.437 | 302.8321 | 72.8 | 0.23 |
| pHILIC | 686.437 | 208.5145 | 72.8 | 0.23 |
| pHILIC | 686.437 | 76.9342 | 72.8 | 0.23 |
| pHILIC | 686.437 | 268.1262 | 72.7 | 0.23 |
| pHILIC | 686.437 | 420.9935 | 72.7 | 0.23 |
| pHILIC | 686.437 | 520.3829 | 72.6 | 0.23 |
| pHILIC | 686.437 | 77.5137 | 72.6 | 0.22 |
| pHILIC | 686.437 | 656.9528 | 72.6 | 0.22 |
| pHILIC | 686.437 | 315.0474 | 72.5 | 0.22 |
| pHILIC | 686.437 | 391.226 | 72.5 | 0.22 |
| pHILIC | 686.437 | 403.6317 | 72 | 0.22 |
| pHILIC | 686.437 | 365.1493 | 71.6 | 0.22 |
| pHILIC | 686.437 | 611.1277 | 71.5 | 0.22 |
| pHILIC | 686.437 | 216.6378 | 71.5 | 0.22 |
| pHILIC | 686.437 | 131.8825 | 71.3 | 0.22 |
| pHILIC | 686.437 | 286.6882 | 71.3 | 0.22 |
| pHILIC | 686.437 | 682.196 | 71.3 | 0.22 |
| pHILIC | 686.437 | 201.1292 | 71.2 | 0.22 |
| pHILIC | 686.437 | 127.7552 | 71.2 | 0.22 |
| pHILIC | 686.437 | 92.4907 | 71.1 | 0.22 |
| pHILIC | 686.437 | 434.4731 | 71.1 | 0.22 |
| pHILIC | 686.437 | 320.5672 | 71.1 | 0.22 |
| pHILIC | 686.437 | 380.2188 | 71.1 | 0.22 |
| pHILIC | 686.437 | 696.2107 | 71.1 | 0.22 |
| pHILIC | 686.437 | 626.8702 | 71 | 0.22 |
| pHILIC | 686.437 | 509.1969 | 71 | 0.22 |
| pHILIC | 686.437 | 122.2282 | 70.9 | 0.22 |
| pHILIC | 686.437 | 163.2828 | 70.9 | 0.22 |

| | | | | |
|---|---|---|---|---|
| pHILIC | 686.437 | 208.0117 | 70.9 | 0.22 |
| pHILIC | 686.437 | 350.8087 | 70.9 | 0.22 |
| pHILIC | 686.437 | 201.3856 | 70.8 | 0.22 |
| pHILIC | 686.437 | 119.529 | 70.8 | 0.22 |
| pHILIC | 686.437 | 283.3708 | 70.7 | 0.22 |
| pHILIC | 686.437 | 287.2156 | 70.7 | 0.22 |
| pHILIC | 686.437 | 122.161 | 70.7 | 0.22 |
| pHILIC | 686.437 | 104.5677 | 70.7 | 0.22 |
| pHILIC | 686.437 | 130.6889 | 70.7 | 0.22 |
| pHILIC | 686.437 | 506.0118 | 70.7 | 0.22 |
| pHILIC | 686.437 | 325.3588 | 70.7 | 0.22 |
| pHILIC | 686.437 | 491.0549 | 70.6 | 0.22 |
| pHILIC | 686.437 | 410.0093 | 70.6 | 0.22 |
| pHILIC | 686.437 | 386.6061 | 70.6 | 0.22 |
| pHILIC | 686.437 | 714.79 | 70.5 | 0.22 |
| pHILIC | 686.437 | 601.8246 | 70.4 | 0.22 |
| pHILIC | 686.437 | 92.2378 | 70.4 | 0.22 |
| pHILIC | 686.437 | 561.6932 | 70.3 | 0.22 |
| pHILIC | 686.437 | 135.1537 | 70.3 | 0.22 |
| pHILIC | 686.437 | 477.4171 | 70.2 | 0.22 |
| pHILIC | 686.437 | 127.2048 | 70.1 | 0.22 |
| pHILIC | 686.437 | 452.4168 | 70.1 | 0.22 |
| pHILIC | 686.437 | 175.1093 | 70.1 | 0.22 |
| pHILIC | 686.437 | 80.5078 | 69.9 | 0.22 |
| pHILIC | 686.437 | 93.9593 | 69.9 | 0.22 |
| pHILIC | 686.437 | 91.9671 | 69.6 | 0.22 |
| pHILIC | 686.437 | 220.1053 | 69.6 | 0.22 |
| pHILIC | 686.437 | 162.0869 | 69.6 | 0.22 |
| pHILIC | 686.437 | 135.8659 | 69.5 | 0.22 |
| pHILIC | 686.437 | 91.9109 | 69.5 | 0.22 |
| pHILIC | 686.437 | 391.375 | 69.5 | 0.22 |
| pHILIC | 686.437 | 657.7878 | 69.5 | 0.22 |
| pHILIC | 686.437 | 121.641 | 69.4 | 0.22 |
| pHILIC | 686.437 | 156.0059 | 69.4 | 0.21 |
| pHILIC | 686.437 | 85.4399 | 69.4 | 0.21 |
| pHILIC | 686.437 | 85.029 | 69.3 | 0.21 |
| pHILIC | 686.437 | 408.1583 | 69.2 | 0.21 |
| pHILIC | 686.437 | 128.7135 | 69.2 | 0.21 |
| pHILIC | 686.437 | 77.7205 | 69.2 | 0.21 |
| pHILIC | 686.437 | 151.8642 | 69.2 | 0.21 |
| pHILIC | 686.437 | 130.8002 | 69.1 | 0.21 |
| pHILIC | 686.437 | 224.719 | 69.1 | 0.21 |

| | | | | |
|---|---|---|---|---|
| pHILIC | 686.437 | 76.9538 | 69.1 | 0.21 |
| pHILIC | 686.437 | 100.2241 | 69.1 | 0.21 |
| pHILIC | 686.437 | 239.8709 | 69.1 | 0.21 |
| pHILIC | 686.437 | 76.8767 | 69 | 0.21 |
| pHILIC | 686.437 | 505.7784 | 69 | 0.21 |
| pHILIC | 686.437 | 75.8624 | 69 | 0.21 |
| pHILIC | 686.437 | 578.8902 | 68.9 | 0.21 |
| pHILIC | 686.437 | 79.2157 | 68.9 | 0.21 |
| pHILIC | 686.437 | 413.5649 | 68.9 | 0.21 |
| pHILIC | 686.437 | 715.7431 | 68.9 | 0.21 |
| pHILIC | 686.437 | 89.765 | 68.9 | 0.21 |
| pHILIC | 686.437 | 109.8568 | 68.8 | 0.21 |
| pHILIC | 686.437 | 534.6747 | 68.8 | 0.21 |
| pHILIC | 686.437 | 131.376 | 68.8 | 0.21 |
| pHILIC | 686.437 | 98.1095 | 68.8 | 0.21 |
| pHILIC | 686.437 | 175.076 | 68.7 | 0.21 |
| pHILIC | 686.437 | 665.5336 | 68.7 | 0.21 |
| pHILIC | 686.437 | 117.9272 | 68.6 | 0.21 |
| pHILIC | 686.437 | 122.4684 | 68.5 | 0.21 |
| pHILIC | 686.437 | 386.3556 | 68.5 | 0.21 |
| pHILIC | 686.437 | 82.364 | 68.3 | 0.21 |
| pHILIC | 686.437 | 375.5139 | 68.3 | 0.21 |
| pHILIC | 686.437 | 78.8818 | 68.3 | 0.21 |
| pHILIC | 686.437 | 543.0521 | 68.3 | 0.21 |
| pHILIC | 686.437 | 108.4677 | 68.3 | 0.21 |
| pHILIC | 686.437 | 123.3935 | 68.2 | 0.21 |
| pHILIC | 686.437 | 279.5516 | 68.2 | 0.21 |
| pHILIC | 686.437 | 117.8117 | 68.2 | 0.21 |
| pHILIC | 686.437 | 129.901 | 68.1 | 0.21 |
| pHILIC | 686.437 | 263.3396 | 68.1 | 0.21 |
| pHILIC | 686.437 | 211.7161 | 68.1 | 0.21 |
| pHILIC | 686.437 | 206.7042 | 68.1 | 0.21 |
| pHILIC | 686.437 | 121.7027 | 68.1 | 0.21 |
| pHILIC | 686.437 | 359.3195 | 68 | 0.21 |
| pHILIC | 686.437 | 372.5216 | 68 | 0.21 |
| pHILIC | 686.437 | 115.2286 | 68 | 0.21 |
| pHILIC | 686.437 | 137.1328 | 68 | 0.21 |
| pHILIC | 686.437 | 160.127 | 67.9 | 0.21 |
| pHILIC | 686.437 | 105.3775 | 67.9 | 0.21 |
| pHILIC | 686.437 | 314.3538 | 67.8 | 0.21 |
| pHILIC | 686.437 | 172.817 | 67.8 | 0.21 |
| pHILIC | 686.437 | 93.7652 | 67.8 | 0.21 |

| | | | | |
|---|---|---|---|---|
| pHILIC | 686.437 | 115.5626 | 67.8 | 0.21 |
| pHILIC | 686.437 | 469.0534 | 67.7 | 0.21 |
| pHILIC | 686.437 | 370.5568 | 67.7 | 0.21 |
| pHILIC | 686.437 | 72.809 | 67.7 | 0.21 |
| pHILIC | 686.437 | 646.2683 | 67.7 | 0.21 |
| pHILIC | 686.437 | 274.6289 | 67.7 | 0.21 |
| pHILIC | 686.437 | 568.4293 | 67.6 | 0.21 |
| pHILIC | 686.437 | 97.3516 | 67.6 | 0.21 |
| pHILIC | 686.437 | 661.9702 | 67.5 | 0.21 |
| pHILIC | 686.437 | 72.6219 | 67.4 | 0.21 |
| pHILIC | 686.437 | 185.6715 | 67.4 | 0.21 |
| pHILIC | 686.437 | 454.0039 | 67.4 | 0.21 |
| pHILIC | 686.437 | 148.6148 | 67.4 | 0.21 |
| pHILIC | 686.437 | 82.0409 | 67.4 | 0.21 |
| pHILIC | 686.437 | 678.8738 | 67.4 | 0.21 |
| pHILIC | 686.437 | 654.8773 | 67.4 | 0.21 |
| pHILIC | 686.437 | 259.3358 | 67.3 | 0.21 |
| pHILIC | 686.437 | 225.6759 | 67.3 | 0.21 |
| pHILIC | 686.437 | 259.2974 | 67.2 | 0.21 |
| pHILIC | 686.437 | 137.6428 | 67.2 | 0.21 |
| pHILIC | 686.437 | 119.6357 | 67.2 | 0.21 |
| pHILIC | 686.437 | 194.8467 | 67.2 | 0.21 |
| pHILIC | 686.437 | 483.3768 | 67.2 | 0.21 |
| pHILIC | 686.437 | 490.029 | 67.2 | 0.21 |
| pHILIC | 686.437 | 104.7032 | 67.1 | 0.21 |
| pHILIC | 686.437 | 345.1312 | 67.1 | 0.21 |
| pHILIC | 686.437 | 691.7004 | 67.1 | 0.21 |
| pHILIC | 686.437 | 604.5184 | 67 | 0.21 |
| pHILIC | 686.437 | 673.7002 | 67 | 0.21 |
| pHILIC | 686.437 | 643.6683 | 67 | 0.21 |
| pHILIC | 686.437 | 134.2328 | 67 | 0.21 |
| pHILIC | 686.437 | 228.0298 | 67 | 0.21 |
| pHILIC | 686.437 | 94.1904 | 67 | 0.21 |
| pHILIC | 686.437 | 642.5095 | 67 | 0.21 |
| pHILIC | 686.437 | 184.8601 | 66.9 | 0.21 |
| pHILIC | 686.437 | 419.0987 | 66.9 | 0.21 |
| pHILIC | 686.437 | 243.4498 | 66.9 | 0.21 |
| pHILIC | 686.437 | 160.2002 | 66.8 | 0.21 |
| pHILIC | 686.437 | 192.524 | 66.8 | 0.21 |
| pHILIC | 686.437 | 658.9476 | 66.8 | 0.21 |
| pHILIC | 686.437 | 295.0705 | 66.8 | 0.21 |
| pHILIC | 686.437 | 193.0573 | 66.8 | 0.21 |

| | | | | |
|---|---|---|---|---|
| pHILIC | 686.437 | 148.0529 | 66.8 | 0.21 |
| pHILIC | 686.437 | 131.2912 | 66.7 | 0.21 |
| pHILIC | 686.437 | 143.6427 | 66.7 | 0.21 |
| pHILIC | 686.437 | 540.3043 | 66.7 | 0.21 |
| pHILIC | 686.437 | 395.5273 | 66.7 | 0.21 |
| pHILIC | 686.437 | 73.8947 | 66.7 | 0.21 |
| pHILIC | 686.437 | 111.7081 | 66.7 | 0.21 |
| pHILIC | 686.437 | 621.399 | 66.7 | 0.21 |
| pHILIC | 686.437 | 406.9973 | 66.7 | 0.21 |
| pHILIC | 686.437 | 647.3976 | 66.7 | 0.21 |
| pHILIC | 686.437 | 169.087 | 66.6 | 0.21 |
| pHILIC | 686.437 | 652.7307 | 66.6 | 0.21 |
| pHILIC | 686.437 | 616.1646 | 66.6 | 0.21 |
| pHILIC | 686.437 | 157.6371 | 66.5 | 0.21 |
| pHILIC | 686.437 | 333.7989 | 66.5 | 0.21 |
| pHILIC | 686.437 | 357.8771 | 66.5 | 0.21 |
| pHILIC | 686.437 | 389.5562 | 66.5 | 0.21 |
| pHILIC | 686.437 | 354.9222 | 66.5 | 0.21 |
| pHILIC | 686.437 | 183.0405 | 66.5 | 0.21 |
| pHILIC | 686.437 | 284.5142 | 66.5 | 0.21 |
| pHILIC | 686.437 | 249.1391 | 66.5 | 0.21 |
| pHILIC | 686.437 | 591.6193 | 66.5 | 0.21 |
| pHILIC | 686.437 | 602.8801 | 66.4 | 0.21 |
| pHILIC | 686.437 | 305.8247 | 66.3 | 0.21 |
| pHILIC | 686.437 | 82.3042 | 66.3 | 0.21 |
| pHILIC | 686.437 | 110.175 | 66.3 | 0.21 |
| pHILIC | 686.437 | 453.8855 | 66.2 | 0.21 |
| pHILIC | 686.437 | 303.3498 | 66.2 | 0.21 |
| pHILIC | 686.437 | 150.462 | 66.2 | 0.21 |
| pHILIC | 686.437 | 85.7673 | 66.2 | 0.2 |
| pHILIC | 686.437 | 91.3507 | 66.2 | 0.2 |
| pHILIC | 686.437 | 98.4794 | 66.1 | 0.2 |
| pHILIC | 686.437 | 86.3832 | 66.1 | 0.2 |
| pHILIC | 686.437 | 335.6905 | 66.1 | 0.2 |
| pHILIC | 686.437 | 477.6213 | 66.1 | 0.2 |
| pHILIC | 686.437 | 175.3124 | 66.1 | 0.2 |
| pHILIC | 686.437 | 73.4751 | 66.1 | 0.2 |
| pHILIC | 686.437 | 113.3948 | 66 | 0.2 |
| pHILIC | 686.437 | 304.3644 | 66 | 0.2 |
| pHILIC | 686.437 | 76.5472 | 66 | 0.2 |
| pHILIC | 686.437 | 112.4636 | 66 | 0.2 |
| pHILIC | 686.437 | 160.554 | 66 | 0.2 |

| | | | | |
|---|---|---|---|---|
| pHILIC | 686.437 | 158.0054 | 65.9 | 0.2 |
| pHILIC | 686.437 | 219.9662 | 65.9 | 0.2 |
| pHILIC | 686.437 | 211.9531 | 65.9 | 0.2 |
| pHILIC | 686.437 | 141.055 | 65.9 | 0.2 |
| pHILIC | 686.437 | 206.4282 | 65.8 | 0.2 |
| pHILIC | 686.437 | 95.7534 | 65.8 | 0.2 |
| pHILIC | 686.437 | 255.0557 | 65.8 | 0.2 |
| pHILIC | 686.437 | 263.0511 | 65.8 | 0.2 |
| pHILIC | 686.437 | 113.6966 | 65.8 | 0.2 |
| pHILIC | 686.437 | 620.4589 | 65.7 | 0.2 |
| pHILIC | 686.437 | 92.6316 | 65.6 | 0.2 |
| pHILIC | 686.437 | 74.8086 | 65.6 | 0.2 |
| pHILIC | 686.437 | 212.7629 | 65.6 | 0.2 |
| pHILIC | 686.437 | 366.2137 | 65.6 | 0.2 |
| pHILIC | 686.437 | 339.0648 | 65.6 | 0.2 |
| pHILIC | 686.437 | 143.1178 | 65.6 | 0.2 |
| pHILIC | 686.437 | 84.5834 | 65.5 | 0.2 |
| pHILIC | 686.437 | 78.0768 | 65.5 | 0.2 |
| pHILIC | 686.437 | 102.7163 | 65.5 | 0.2 |
| pHILIC | 686.437 | 168.2695 | 65.4 | 0.2 |
| pHILIC | 686.437 | 226.6383 | 65.4 | 0.2 |
| pHILIC | 686.437 | 189.2815 | 65.4 | 0.2 |
| pHILIC | 686.437 | 364.2227 | 65.3 | 0.2 |
| pHILIC | 686.437 | 412.1497 | 65.3 | 0.2 |
| pHILIC | 686.437 | 374.054 | 65.3 | 0.2 |
| pHILIC | 686.437 | 95.0659 | 65.3 | 0.2 |
| pHILIC | 686.437 | 102.2455 | 65.3 | 0.2 |
| pHILIC | 686.437 | 562.1555 | 65.3 | 0.2 |
| pHILIC | 686.437 | 145.1012 | 65.3 | 0.2 |
| pHILIC | 686.437 | 155.5138 | 65.2 | 0.2 |
| pHILIC | 686.437 | 212.9841 | 65.2 | 0.2 |
| pHILIC | 686.437 | 128.4083 | 65.2 | 0.2 |
| pHILIC | 686.437 | 176.4092 | 65.2 | 0.2 |
| pHILIC | 686.437 | 82.8234 | 65.2 | 0.2 |
| pHILIC | 686.437 | 233.7544 | 65.2 | 0.2 |
| pHILIC | 686.437 | 123.8549 | 65.1 | 0.2 |
| pHILIC | 686.437 | 492.6631 | 65.1 | 0.2 |
| pHILIC | 686.437 | 102.0513 | 65.1 | 0.2 |
| pHILIC | 686.437 | 122.3817 | 65.1 | 0.2 |
| pHILIC | 686.437 | 348.2427 | 65.1 | 0.2 |
| pHILIC | 686.437 | 219.1265 | 65 | 0.2 |
| pHILIC | 686.437 | 216.7233 | 65 | 0.2 |

| | | | | |
|---|---|---|---|---|
| pHILIC | 686.437 | 595.366 | 65 | 0.2 |
| pHILIC | 686.437 | 79.7987 | 65 | 0.2 |
| pHILIC | 686.437 | 73.544 | 65 | 0.2 |
| pHILIC | 686.437 | 155.1999 | 65 | 0.2 |
| pHILIC | 686.437 | 659.5497 | 64.9 | 0.2 |
| pHILIC | 686.437 | 406.533 | 64.9 | 0.2 |
| pHILIC | 686.437 | 80.5826 | 64.9 | 0.2 |
| pHILIC | 686.437 | 705.439 | 64.8 | 0.2 |
| pHILIC | 686.437 | 588.9479 | 64.8 | 0.2 |
| pHILIC | 686.437 | 341.7876 | 64.8 | 0.2 |
| pHILIC | 686.437 | 167.484 | 64.8 | 0.2 |
| pHILIC | 686.437 | 485.1877 | 64.8 | 0.2 |
| pHILIC | 686.437 | 135.3453 | 64.7 | 0.2 |
| pHILIC | 686.437 | 168.7552 | 64.7 | 0.2 |
| pHILIC | 686.437 | 121.1009 | 64.7 | 0.2 |
| pHILIC | 686.437 | 182.4184 | 64.7 | 0.2 |
| pHILIC | 686.437 | 110.3661 | 64.7 | 0.2 |
| pHILIC | 686.437 | 226.3515 | 64.7 | 0.2 |
| pHILIC | 686.437 | 87.1397 | 64.7 | 0.2 |
| pHILIC | 686.437 | 147.6141 | 64.7 | 0.2 |
| pHILIC | 686.437 | 590.4633 | 64.7 | 0.2 |
| pHILIC | 686.437 | 226.7191 | 64.6 | 0.2 |
| pHILIC | 686.437 | 363.4339 | 64.6 | 0.2 |
| pHILIC | 686.437 | 92.0131 | 64.6 | 0.2 |
| pHILIC | 686.437 | 192.903 | 64.6 | 0.2 |
| pHILIC | 686.437 | 472.2367 | 64.6 | 0.2 |
| pHILIC | 686.437 | 464.5854 | 64.6 | 0.2 |
| pHILIC | 686.437 | 105.4991 | 64.6 | 0.2 |
| pHILIC | 686.437 | 99.3308 | 64.6 | 0.2 |
| pHILIC | 686.437 | 118.5613 | 64.6 | 0.2 |
| pHILIC | 686.437 | 86.2527 | 64.6 | 0.2 |
| pHILIC | 686.437 | 106.526 | 64.5 | 0.2 |
| pHILIC | 686.437 | 708.04 | 64.5 | 0.2 |
| pHILIC | 686.437 | 198.1252 | 64.5 | 0.2 |
| pHILIC | 686.437 | 245.0673 | 64.5 | 0.2 |
| pHILIC | 686.437 | 98.4068 | 64.4 | 0.2 |
| pHILIC | 686.437 | 75.8826 | 64.4 | 0.2 |
| pHILIC | 686.437 | 131.5815 | 64.4 | 0.2 |
| pHILIC | 686.437 | 379.5103 | 64.4 | 0.2 |
| pHILIC | 686.437 | 95.6667 | 64.4 | 0.2 |
| pHILIC | 686.437 | 589.1396 | 64.4 | 0.2 |
| pHILIC | 686.437 | 111.8172 | 64.4 | 0.2 |

| | | | | |
|---|---|---|---|---|
| pHILIC | 686.437 | 126.7089 | 64.4 | 0.2 |
| pHILIC | 686.437 | 142.6358 | 64.4 | 0.2 |
| pHILIC | 686.437 | 565.2376 | 64.3 | 0.2 |
| pHILIC | 686.437 | 134.2002 | 64.3 | 0.2 |
| pHILIC | 686.437 | 75.177 | 64.3 | 0.2 |
| pHILIC | 686.437 | 84.6992 | 64.3 | 0.2 |
| pHILIC | 686.437 | 106.9882 | 64.2 | 0.2 |
| pHILIC | 686.437 | 242.3564 | 64.2 | 0.2 |
| pHILIC | 686.437 | 656.2434 | 64.2 | 0.2 |
| pHILIC | 686.437 | 434.7294 | 64.2 | 0.2 |
| pHILIC | 686.437 | 163.3714 | 64.1 | 0.2 |
| pHILIC | 686.437 | 76.6864 | 64.1 | 0.2 |
| pHILIC | 686.437 | 327.6721 | 64.1 | 0.2 |
| pHILIC | 686.437 | 214.7215 | 64.1 | 0.2 |
| pHILIC | 686.437 | 87.6387 | 64.1 | 0.2 |
| pHILIC | 686.437 | 88.6926 | 64.1 | 0.2 |
| pHILIC | 686.437 | 98.3846 | 64 | 0.2 |
| pHILIC | 686.437 | 459.1378 | 64 | 0.2 |
| pHILIC | 686.437 | 719.3908 | 64 | 0.2 |
| pHILIC | 686.437 | 176.8015 | 64 | 0.2 |
| pHILIC | 686.437 | 103.7235 | 64 | 0.2 |
| pHILIC | 686.437 | 177.217 | 64 | 0.2 |
| pHILIC | 686.437 | 362.8186 | 64 | 0.2 |
| pHILIC | 686.437 | 151.8018 | 64 | 0.2 |
| pHILIC | 686.437 | 91.1732 | 64 | 0.2 |
| pHILIC | 686.437 | 135.5157 | 64 | 0.2 |
| pHILIC | 686.437 | 487.3781 | 64 | 0.2 |
| pHILIC | 686.437 | 224.3529 | 63.9 | 0.2 |
| pHILIC | 686.437 | 86.7655 | 63.9 | 0.2 |
| pHILIC | 686.437 | 246.9639 | 63.9 | 0.2 |
| pHILIC | 686.437 | 130.7572 | 63.8 | 0.2 |
| pHILIC | 686.437 | 335.9322 | 63.8 | 0.2 |
| pHILIC | 686.437 | 708.9825 | 63.8 | 0.2 |
| pHILIC | 686.437 | 425.647 | 63.8 | 0.2 |
| pHILIC | 686.437 | 257.9285 | 63.8 | 0.2 |
| pHILIC | 686.437 | 96.1464 | 63.8 | 0.2 |
| pHILIC | 686.437 | 72.2058 | 63.8 | 0.2 |
| pHILIC | 686.437 | 129.6823 | 63.8 | 0.2 |
| pHILIC | 686.437 | 93.9686 | 63.7 | 0.2 |
| pHILIC | 686.437 | 341.6301 | 63.7 | 0.2 |
| pHILIC | 686.437 | 89.474 | 63.7 | 0.2 |
| pHILIC | 686.437 | 686.7479 | 63.6 | 0.2 |

| | | | | |
|---|---|---|---|---|
| pHILIC | 686.437 | 115.9308 | 63.6 | 0.2 |
| pHILIC | 686.437 | 99.9849 | 63.6 | 0.2 |
| pHILIC | 686.437 | 179.2729 | 63.6 | 0.2 |
| pHILIC | 686.437 | 582.6172 | 63.6 | 0.2 |
| pHILIC | 686.437 | 348.6763 | 63.6 | 0.2 |
| pHILIC | 686.437 | 255.0206 | 63.6 | 0.2 |
| pHILIC | 686.437 | 143.6876 | 63.5 | 0.2 |
| pHILIC | 686.437 | 88.3326 | 63.5 | 0.2 |
| pHILIC | 686.437 | 136.1433 | 63.4 | 0.2 |
| pHILIC | 686.437 | 121.3308 | 63.4 | 0.2 |
| pHILIC | 686.437 | 599.2173 | 63.4 | 0.2 |
| pHILIC | 686.437 | 215.8131 | 63.4 | 0.2 |
| pHILIC | 686.437 | 281.6985 | 63.4 | 0.2 |
| pHILIC | 686.437 | 82.5775 | 63.4 | 0.2 |
| pHILIC | 686.437 | 208.5695 | 63.4 | 0.2 |
| pHILIC | 686.437 | 132.0949 | 63.4 | 0.2 |
| pHILIC | 686.437 | 324.889 | 63.4 | 0.2 |
| pHILIC | 686.437 | 131.9754 | 63.4 | 0.2 |
| pHILIC | 686.437 | 535.5554 | 63.3 | 0.2 |
| pHILIC | 686.437 | 73.3562 | 63.3 | 0.2 |
| pHILIC | 686.437 | 146.1838 | 63.3 | 0.2 |
| pHILIC | 686.437 | 152.2122 | 63.3 | 0.2 |
| pHILIC | 686.437 | 588.3972 | 63.3 | 0.2 |
| pHILIC | 686.437 | 411.7947 | 63.3 | 0.2 |
| pHILIC | 686.437 | 347.6166 | 63.3 | 0.2 |
| pHILIC | 686.437 | 369.9043 | 63.3 | 0.2 |
| pHILIC | 686.437 | 75.6285 | 63.3 | 0.2 |
| pHILIC | 686.437 | 85.0478 | 63.3 | 0.2 |
| pHILIC | 686.437 | 96.6543 | 63.2 | 0.2 |
| pHILIC | 686.437 | 99.7384 | 63.2 | 0.2 |
| pHILIC | 686.437 | 208.1028 | 63.2 | 0.2 |
| pHILIC | 686.437 | 225.133 | 63.2 | 0.2 |
| pHILIC | 686.437 | 121.3619 | 63.2 | 0.2 |
| pHILIC | 686.437 | 145.3307 | 63.2 | 0.2 |
| pHILIC | 686.437 | 269.5885 | 63.2 | 0.2 |
| pHILIC | 686.437 | 159.4504 | 63.2 | 0.2 |
| pHILIC | 686.437 | 122.7606 | 63.2 | 0.2 |
| pHILIC | 686.437 | 617.0109 | 63.1 | 0.2 |
| pHILIC | 686.437 | 208.1505 | 63.1 | 0.2 |
| pHILIC | 686.437 | 559.0344 | 63.1 | 0.2 |
| pHILIC | 686.437 | 87.0077 | 63.1 | 0.2 |
| pHILIC | 686.437 | 84.8391 | 63.1 | 0.2 |

| | | | | |
|---|---|---|---|---|
| pHILIC | 686.437 | 524.6849 | 63.1 | 0.2 |
| pHILIC | 686.437 | 198.9289 | 63.1 | 0.2 |
| pHILIC | 686.437 | 635.2462 | 63.1 | 0.2 |
| pHILIC | 686.437 | 155.4322 | 63.1 | 0.2 |
| pHILIC | 686.437 | 644.7475 | 63.1 | 0.2 |
| pHILIC | 686.437 | 93.5864 | 63.1 | 0.2 |
| pHILIC | 686.437 | 82.0616 | 63.1 | 0.2 |
| pHILIC | 686.437 | 79.9855 | 63 | 0.2 |
| pHILIC | 686.437 | 571.5378 | 63 | 0.2 |
| pHILIC | 686.437 | 140.6554 | 63 | 0.2 |
| pHILIC | 686.437 | 510.0464 | 63 | 0.2 |
| pHILIC | 686.437 | 468.3631 | 63 | 0.2 |
| pHILIC | 686.437 | 97.6527 | 63 | 0.2 |
| pHILIC | 686.437 | 287.7262 | 63 | 0.2 |
| pHILIC | 686.437 | 102.096 | 63 | 0.2 |
| pHILIC | 686.437 | 486.4864 | 62.9 | 0.19 |
| pHILIC | 686.437 | 338.849 | 62.9 | 0.19 |
| pHILIC | 686.437 | 286.0491 | 62.9 | 0.19 |
| pHILIC | 686.437 | 445.4586 | 62.9 | 0.19 |
| pHILIC | 686.437 | 92.1773 | 62.9 | 0.19 |
| pHILIC | 686.437 | 147.1241 | 62.9 | 0.19 |
| pHILIC | 686.437 | 93.8414 | 62.9 | 0.19 |
| pHILIC | 686.437 | 142.8351 | 62.9 | 0.19 |
| pHILIC | 686.437 | 678.1667 | 62.8 | 0.19 |
| pHILIC | 686.437 | 127.2218 | 62.8 | 0.19 |
| pHILIC | 686.437 | 78.6224 | 62.8 | 0.19 |
| pHILIC | 686.437 | 131.1946 | 62.8 | 0.19 |
| pHILIC | 686.437 | 164.0763 | 62.8 | 0.19 |
| pHILIC | 686.437 | 102.5938 | 62.8 | 0.19 |
| pHILIC | 686.437 | 204.7451 | 62.8 | 0.19 |
| pHILIC | 686.437 | 72.8213 | 62.7 | 0.19 |
| pHILIC | 686.437 | 154.5506 | 62.7 | 0.19 |
| pHILIC | 686.437 | 128.7479 | 62.6 | 0.19 |
| pHILIC | 686.437 | 158.9848 | 62.6 | 0.19 |
| pHILIC | 686.437 | 704.8205 | 62.6 | 0.19 |
| pHILIC | 686.437 | 157.9128 | 62.6 | 0.19 |
| pHILIC | 686.437 | 119.3584 | 62.6 | 0.19 |
| pHILIC | 686.437 | 95.5481 | 62.5 | 0.19 |
| pHILIC | 686.437 | 116.4266 | 62.5 | 0.19 |
| pHILIC | 686.437 | 224.4872 | 62.5 | 0.19 |
| pHILIC | 686.437 | 74.4506 | 62.5 | 0.19 |
| pHILIC | 686.437 | 238.4242 | 62.5 | 0.19 |

| | | | | |
|---|---|---|---|---|
| pHILIC | 686.437 | 103.3656 | 62.5 | 0.19 |
| pHILIC | 686.437 | 178.903 | 62.5 | 0.19 |
| pHILIC | 686.437 | 162.9184 | 62.5 | 0.19 |
| pHILIC | 686.437 | 75.3494 | 62.4 | 0.19 |
| pHILIC | 686.437 | 165.6743 | 62.4 | 0.19 |
| pHILIC | 686.437 | 394.4109 | 62.4 | 0.19 |
| pHILIC | 686.437 | 558.2882 | 62.4 | 0.19 |
| pHILIC | 686.437 | 369.4481 | 62.4 | 0.19 |
| pHILIC | 686.437 | 118.374 | 62.3 | 0.19 |
| pHILIC | 686.437 | 196.1462 | 62.3 | 0.19 |
| pHILIC | 686.437 | 95.8743 | 62.3 | 0.19 |
| pHILIC | 686.437 | 669.8486 | 62.3 | 0.19 |
| pHILIC | 686.437 | 126.7445 | 62.3 | 0.19 |
| pHILIC | 686.437 | 280.6477 | 62.3 | 0.19 |
| pHILIC | 686.437 | 105.8721 | 62.3 | 0.19 |
| pHILIC | 686.437 | 334.2385 | 62.2 | 0.19 |
| pHILIC | 686.437 | 709.8939 | 62.2 | 0.19 |
| pHILIC | 686.437 | 97.7903 | 62.2 | 0.19 |
| pHILIC | 686.437 | 75.8708 | 62.2 | 0.19 |
| pHILIC | 686.437 | 88.6807 | 62.2 | 0.19 |
| pHILIC | 686.437 | 117.6387 | 62.2 | 0.19 |
| pHILIC | 686.437 | 115.4993 | 62.2 | 0.19 |
| pHILIC | 686.437 | 557.4818 | 62.2 | 0.19 |
| pHILIC | 686.437 | 661.4019 | 62.2 | 0.19 |
| pHILIC | 686.437 | 96.3848 | 62.1 | 0.19 |
| pHILIC | 686.437 | 295.9172 | 62.1 | 0.19 |
| pHILIC | 686.437 | 88.146 | 62.1 | 0.19 |
| pHILIC | 686.437 | 89.4794 | 62.1 | 0.19 |
| pHILIC | 686.437 | 629.9983 | 62.1 | 0.19 |
| pHILIC | 686.437 | 198.6334 | 62 | 0.19 |
| pHILIC | 686.437 | 683.604 | 62 | 0.19 |
| pHILIC | 686.437 | 269.1321 | 62 | 0.19 |
| pHILIC | 686.437 | 229.915 | 62 | 0.19 |
| pHILIC | 686.437 | 88.9239 | 62 | 0.19 |
| pHILIC | 686.437 | 109.7811 | 62 | 0.19 |
| pHILIC | 686.437 | 98.081 | 61.9 | 0.19 |
| pHILIC | 686.437 | 292.5789 | 61.9 | 0.19 |
| pHILIC | 686.437 | 79.999 | 61.9 | 0.19 |
| pHILIC | 686.437 | 111.8461 | 61.9 | 0.19 |
| pHILIC | 686.437 | 88.1413 | 61.9 | 0.19 |
| pHILIC | 686.437 | 245.7375 | 61.9 | 0.19 |
| pHILIC | 686.437 | 158.548 | 61.9 | 0.19 |

| | | | | |
|---|---|---|---|---|
| pHILIC | 686.437 | 282.5629 | 61.9 | 0.19 |
| pHILIC | 686.437 | 96.6651 | 61.9 | 0.19 |
| pHILIC | 686.437 | 83.2138 | 61.8 | 0.19 |
| pHILIC | 686.437 | 100.432 | 61.8 | 0.19 |
| pHILIC | 686.437 | 403.1585 | 61.8 | 0.19 |
| pHILIC | 686.437 | 79.8058 | 61.7 | 0.19 |
| pHILIC | 686.437 | 156.3101 | 61.7 | 0.19 |
| pHILIC | 686.437 | 304.4686 | 61.7 | 0.19 |
| pHILIC | 686.437 | 113.0071 | 61.7 | 0.19 |
| pHILIC | 686.437 | 92.1573 | 61.7 | 0.19 |
| pHILIC | 686.437 | 177.0248 | 61.6 | 0.19 |
| pHILIC | 686.437 | 125.2726 | 61.6 | 0.19 |
| pHILIC | 686.437 | 97.6151 | 61.6 | 0.19 |
| pHILIC | 686.437 | 86.6793 | 61.6 | 0.19 |
| pHILIC | 686.437 | 120.8085 | 61.6 | 0.19 |
| pHILIC | 686.437 | 150.6677 | 61.6 | 0.19 |
| pHILIC | 686.437 | 213.9644 | 61.6 | 0.19 |
| pHILIC | 686.437 | 136.9573 | 61.6 | 0.19 |
| pHILIC | 686.437 | 182.291 | 61.6 | 0.19 |
| pHILIC | 686.437 | 564.2917 | 61.6 | 0.19 |
| pHILIC | 686.437 | 266.1762 | 61.6 | 0.19 |
| pHILIC | 686.437 | 514.3325 | 61.6 | 0.19 |
| pHILIC | 686.437 | 76.0872 | 61.6 | 0.19 |
| pHILIC | 686.437 | 385.2631 | 61.6 | 0.19 |
| pHILIC | 686.437 | 102.6866 | 61.6 | 0.19 |
| pHILIC | 686.437 | 95.1995 | 61.6 | 0.19 |
| pHILIC | 686.437 | 241.3598 | 61.6 | 0.19 |
| pHILIC | 686.437 | 84.0812 | 61.6 | 0.19 |
| pHILIC | 686.437 | 437.9283 | 61.5 | 0.19 |
| pHILIC | 686.437 | 291.4111 | 61.5 | 0.19 |
| pHILIC | 686.437 | 73.207 | 61.5 | 0.19 |
| pHILIC | 686.437 | 90.8154 | 61.5 | 0.19 |
| pHILIC | 686.437 | 239.9729 | 61.5 | 0.19 |
| pHILIC | 686.437 | 233.7972 | 61.5 | 0.19 |
| pHILIC | 686.437 | 463.1711 | 61.5 | 0.19 |
| pHILIC | 686.437 | 399.2868 | 61.5 | 0.19 |
| pHILIC | 686.437 | 202.6224 | 61.5 | 0.19 |
| pHILIC | 686.437 | 221.4248 | 61.5 | 0.19 |
| pHILIC | 686.437 | 264.0654 | 61.5 | 0.19 |
| pHILIC | 686.437 | 79.5252 | 61.5 | 0.19 |
| pHILIC | 686.437 | 223.0066 | 61.5 | 0.19 |
| pHILIC | 686.437 | 470.1269 | 61.5 | 0.19 |

| | | | | |
|---|---|---|---|---|
| pHILIC | 686.437 | 415.6033 | 61.5 | 0.19 |
| pHILIC | 686.437 | 201.5038 | 61.4 | 0.19 |
| pHILIC | 686.437 | 192.2314 | 61.4 | 0.19 |
| pHILIC | 686.437 | 121.2231 | 61.4 | 0.19 |
| pHILIC | 686.437 | 171.9856 | 61.4 | 0.19 |
| pHILIC | 686.437 | 199.1996 | 61.4 | 0.19 |
| pHILIC | 686.437 | 238.3209 | 61.3 | 0.19 |
| pHILIC | 686.437 | 370.2375 | 61.3 | 0.19 |
| pHILIC | 686.437 | 156.5089 | 61.3 | 0.19 |
| pHILIC | 686.437 | 279.4084 | 61.3 | 0.19 |
| pHILIC | 686.437 | 254.4431 | 61.3 | 0.19 |
| pHILIC | 686.437 | 227.5416 | 61.3 | 0.19 |
| pHILIC | 686.437 | 664.038 | 61.3 | 0.19 |
| pHILIC | 686.437 | 145.461 | 61.2 | 0.19 |
| pHILIC | 686.437 | 87.3369 | 61.2 | 0.19 |
| pHILIC | 686.437 | 92.0049 | 61.2 | 0.19 |
| pHILIC | 686.437 | 196.2426 | 61.2 | 0.19 |
| pHILIC | 686.437 | 90.78 | 61.2 | 0.19 |
| pHILIC | 686.437 | 701.1057 | 61.2 | 0.19 |
| pHILIC | 686.437 | 140.9456 | 61.2 | 0.19 |
| pHILIC | 686.437 | 624.8426 | 61.2 | 0.19 |
| pHILIC | 686.437 | 550.4374 | 61.1 | 0.19 |
| pHILIC | 686.437 | 87.1304 | 61.1 | 0.19 |
| pHILIC | 686.437 | 79.4076 | 61.1 | 0.19 |
| pHILIC | 686.437 | 236.0712 | 61.1 | 0.19 |
| pHILIC | 686.437 | 211.1424 | 61.1 | 0.19 |
| pHILIC | 686.437 | 217.5253 | 61.1 | 0.19 |
| pHILIC | 686.437 | 135.0793 | 61.1 | 0.19 |
| pHILIC | 686.437 | 525.9499 | 61.1 | 0.19 |
| pHILIC | 686.437 | 385.5472 | 61.1 | 0.19 |
| pHILIC | 686.437 | 116.1398 | 61.1 | 0.19 |
| pHILIC | 686.437 | 113.7375 | 61 | 0.19 |
| pHILIC | 686.437 | 264.7901 | 61 | 0.19 |
| pHILIC | 686.437 | 163.6281 | 61 | 0.19 |
| pHILIC | 686.437 | 129.8679 | 61 | 0.19 |
| pHILIC | 686.437 | 116.5249 | 61 | 0.19 |
| pHILIC | 686.437 | 95.8234 | 60.9 | 0.19 |
| pHILIC | 686.437 | 219.6432 | 60.9 | 0.19 |
| pHILIC | 686.437 | 127.6851 | 60.9 | 0.19 |
| pHILIC | 686.437 | 116.7334 | 60.9 | 0.19 |
| pHILIC | 686.437 | 75.9137 | 60.8 | 0.19 |
| pHILIC | 686.437 | 193.2855 | 60.8 | 0.19 |

| | | | | |
|---|---|---|---|---|
| pHILIC | 686.437 | 134.9613 | 60.8 | 0.19 |
| pHILIC | 686.437 | 82.0089 | 60.8 | 0.19 |
| pHILIC | 686.437 | 102.9156 | 60.8 | 0.19 |
| pHILIC | 686.437 | 160.1029 | 60.8 | 0.19 |
| pHILIC | 686.437 | 84.2398 | 60.8 | 0.19 |
| pHILIC | 686.437 | 705.9604 | 60.8 | 0.19 |
| pHILIC | 686.437 | 259.9898 | 60.8 | 0.19 |
| pHILIC | 686.437 | 150.6461 | 60.8 | 0.19 |
| pHILIC | 686.437 | 87.2322 | 60.8 | 0.19 |
| pHILIC | 686.437 | 95.1133 | 60.8 | 0.19 |
| pHILIC | 686.437 | 455.3648 | 60.8 | 0.19 |
| pHILIC | 686.437 | 254.6931 | 60.7 | 0.19 |
| pHILIC | 686.437 | 209.9539 | 60.7 | 0.19 |
| pHILIC | 686.437 | 578.4559 | 60.7 | 0.19 |
| pHILIC | 686.437 | 316.2939 | 60.7 | 0.19 |
| pHILIC | 686.437 | 173.0604 | 60.7 | 0.19 |
| pHILIC | 686.437 | 354.5347 | 60.7 | 0.19 |
| pHILIC | 686.437 | 120.0636 | 60.7 | 0.19 |
| pHILIC | 686.437 | 91.2453 | 60.7 | 0.19 |
| pHILIC | 686.437 | 195.8342 | 60.7 | 0.19 |
| pHILIC | 686.437 | 212.1477 | 60.6 | 0.19 |
| pHILIC | 686.437 | 236.6119 | 60.6 | 0.19 |
| pHILIC | 686.437 | 286.0841 | 60.6 | 0.19 |
| pHILIC | 686.437 | 83.6971 | 60.6 | 0.19 |
| pHILIC | 686.437 | 95.5303 | 60.6 | 0.19 |
| pHILIC | 686.437 | 99.3217 | 60.6 | 0.19 |
| pHILIC | 686.437 | 258.6069 | 60.6 | 0.19 |
| pHILIC | 686.437 | 634.6915 | 60.6 | 0.19 |
| pHILIC | 686.437 | 542.4749 | 60.6 | 0.19 |
| pHILIC | 686.437 | 124.0653 | 60.5 | 0.19 |
| pHILIC | 686.437 | 85.4821 | 60.5 | 0.19 |
| pHILIC | 686.437 | 190.2864 | 60.5 | 0.19 |
| pHILIC | 686.437 | 333.119 | 60.5 | 0.19 |
| pHILIC | 686.437 | 164.717 | 60.5 | 0.19 |
| pHILIC | 686.437 | 80.309 | 60.5 | 0.19 |
| pHILIC | 686.437 | 74.7907 | 60.4 | 0.19 |
| pHILIC | 686.437 | 137.4581 | 60.4 | 0.19 |
| pHILIC | 686.437 | 263.6001 | 60.4 | 0.19 |
| pHILIC | 686.437 | 158.9662 | 60.4 | 0.19 |
| pHILIC | 686.437 | 301.4493 | 60.3 | 0.19 |
| pHILIC | 686.437 | 143.1991 | 60.3 | 0.19 |
| pHILIC | 686.437 | 142.7013 | 60.3 | 0.19 |

| | | | | |
|---|---|---|---|---|
| pHILIC | 686.437 | 77.9425 | 60.3 | 0.19 |
| pHILIC | 686.437 | 125.0104 | 60.3 | 0.19 |
| pHILIC | 686.437 | 204.0373 | 60.2 | 0.19 |
| pHILIC | 686.437 | 74.5887 | 60.2 | 0.19 |
| pHILIC | 686.437 | 97.2866 | 60.2 | 0.19 |
| pHILIC | 686.437 | 186.1595 | 60.2 | 0.19 |
| pHILIC | 686.437 | 267.1969 | 60.2 | 0.19 |
| pHILIC | 686.437 | 289.4912 | 60.2 | 0.19 |
| pHILIC | 686.437 | 305.8951 | 60.2 | 0.19 |
| pHILIC | 686.437 | 140.02 | 60.2 | 0.19 |
| pHILIC | 686.437 | 157.0105 | 60.1 | 0.19 |
| pHILIC | 686.437 | 308.9053 | 60.1 | 0.19 |
| pHILIC | 686.437 | 176.0616 | 60.1 | 0.19 |
| pHILIC | 686.437 | 191.4347 | 60.1 | 0.19 |
| pHILIC | 686.437 | 84.0959 | 60.1 | 0.19 |
| pHILIC | 686.437 | 95.2478 | 60.1 | 0.19 |
| pHILIC | 686.437 | 298.6291 | 60.1 | 0.19 |
| pHILIC | 686.437 | 104.3659 | 60.1 | 0.19 |
| pHILIC | 686.437 | 148.4786 | 60.1 | 0.19 |
| pHILIC | 686.437 | 283.3309 | 60 | 0.19 |
| pHILIC | 686.437 | 648.7978 | 60 | 0.19 |
| pHILIC | 686.437 | 337.5925 | 60 | 0.19 |
| pHILIC | 686.437 | 485.3658 | 60 | 0.19 |
| pHILIC | 686.437 | 221.1324 | 60 | 0.19 |
| pHILIC | 686.437 | 132.3985 | 59.9 | 0.19 |
| pHILIC | 686.437 | 77.8013 | 59.9 | 0.19 |
| pHILIC | 686.437 | 500.4504 | 59.9 | 0.19 |
| pHILIC | 686.437 | 129.1944 | 59.9 | 0.19 |
| pHILIC | 686.437 | 79.8026 | 59.9 | 0.19 |
| pHILIC | 686.437 | 134.4298 | 59.9 | 0.19 |
| pHILIC | 686.437 | 211.8387 | 59.9 | 0.19 |
| pHILIC | 686.437 | 82.5988 | 59.9 | 0.19 |
| pHILIC | 686.437 | 184.7596 | 59.8 | 0.19 |
| pHILIC | 686.437 | 403.2918 | 59.8 | 0.19 |
| pHILIC | 686.437 | 103.4086 | 59.8 | 0.19 |
| pHILIC | 686.437 | 263.4714 | 59.8 | 0.19 |
| pHILIC | 686.437 | 81.7565 | 59.8 | 0.19 |
| pHILIC | 686.437 | 152.7015 | 59.8 | 0.19 |
| pHILIC | 686.437 | 106.1892 | 59.8 | 0.19 |
| pHILIC | 686.437 | 99.6237 | 59.8 | 0.19 |
| pHILIC | 686.437 | 108.3871 | 59.7 | 0.19 |
| pHILIC | 686.437 | 143.9207 | 59.7 | 0.18 |

| | | | | |
|---|---|---|---|---|
| pHILIC | 686.437 | 161.9939 | 59.7 | 0.18 |
| pHILIC | 686.437 | 312.5809 | 59.7 | 0.18 |
| pHILIC | 686.437 | 102.6426 | 59.7 | 0.18 |
| pHILIC | 686.437 | 107.5551 | 59.7 | 0.18 |
| pHILIC | 686.437 | 159.7128 | 59.7 | 0.18 |
| pHILIC | 686.437 | 248.7481 | 59.6 | 0.18 |
| pHILIC | 686.437 | 273.6396 | 59.6 | 0.18 |
| pHILIC | 686.437 | 105.1843 | 59.6 | 0.18 |
| pHILIC | 686.437 | 317.0681 | 59.6 | 0.18 |
| pHILIC | 686.437 | 79.324 | 59.6 | 0.18 |
| pHILIC | 686.437 | 135.6465 | 59.6 | 0.18 |
| pHILIC | 686.437 | 142.4679 | 59.6 | 0.18 |
| pHILIC | 686.437 | 86.1385 | 59.6 | 0.18 |
| pHILIC | 686.437 | 372.9049 | 59.6 | 0.18 |
| pHILIC | 686.437 | 224.8756 | 59.6 | 0.18 |
| pHILIC | 686.437 | 184.5821 | 59.6 | 0.18 |
| pHILIC | 686.437 | 130.6777 | 59.5 | 0.18 |
| pHILIC | 686.437 | 169.8485 | 59.5 | 0.18 |
| pHILIC | 686.437 | 573.3798 | 59.5 | 0.18 |
| pHILIC | 686.437 | 130.0223 | 59.5 | 0.18 |
| pHILIC | 686.437 | 148.7914 | 59.5 | 0.18 |
| pHILIC | 686.437 | 85.6176 | 59.5 | 0.18 |
| pHILIC | 686.437 | 211.6619 | 59.4 | 0.18 |
| pHILIC | 686.437 | 352.1022 | 59.4 | 0.18 |
| pHILIC | 686.437 | 172.0417 | 59.4 | 0.18 |
| pHILIC | 686.437 | 73.5035 | 59.4 | 0.18 |
| pHILIC | 686.437 | 285.6894 | 59.4 | 0.18 |
| pHILIC | 686.437 | 417.4516 | 59.4 | 0.18 |
| pHILIC | 686.437 | 390.6119 | 59.4 | 0.18 |
| pHILIC | 686.437 | 104.1699 | 59.4 | 0.18 |
| pHILIC | 686.437 | 105.9581 | 59.4 | 0.18 |
| pHILIC | 686.437 | 244.9896 | 59.3 | 0.18 |
| pHILIC | 686.437 | 74.7328 | 59.3 | 0.18 |
| pHILIC | 686.437 | 163.6981 | 59.3 | 0.18 |
| pHILIC | 686.437 | 170.1784 | 59.3 | 0.18 |
| pHILIC | 686.437 | 360.6322 | 59.3 | 0.18 |
| pHILIC | 686.437 | 206.8796 | 59.3 | 0.18 |
| pHILIC | 686.437 | 90.1547 | 59.3 | 0.18 |
| pHILIC | 686.437 | 454.8358 | 59.3 | 0.18 |
| pHILIC | 686.437 | 79.4973 | 59.2 | 0.18 |
| pHILIC | 686.437 | 221.1689 | 59.2 | 0.18 |
| pHILIC | 686.437 | 169.6466 | 59.2 | 0.18 |

| | | | | |
|---|---|---|---|---|
| pHILIC | 686.437 | 148.5365 | 59.2 | 0.18 |
| pHILIC | 686.437 | 84.07 | 59.2 | 0.18 |
| pHILIC | 686.437 | 523.9623 | 59.2 | 0.18 |
| pHILIC | 686.437 | 150.9259 | 59.2 | 0.18 |
| pHILIC | 686.437 | 83.5313 | 59.2 | 0.18 |
| pHILIC | 686.437 | 621.556 | 59.1 | 0.18 |
| pHILIC | 686.437 | 326.7806 | 59.1 | 0.18 |
| pHILIC | 686.437 | 76.4581 | 59.1 | 0.18 |
| pHILIC | 686.437 | 84.2007 | 59.1 | 0.18 |
| pHILIC | 686.437 | 158.781 | 59.1 | 0.18 |
| pHILIC | 686.437 | 245.6771 | 59.1 | 0.18 |
| pHILIC | 686.437 | 291.1469 | 59.1 | 0.18 |
| pHILIC | 686.437 | 82.285 | 59.1 | 0.18 |
| pHILIC | 686.437 | 307.5595 | 59.1 | 0.18 |
| pHILIC | 686.437 | 77.9995 | 59.1 | 0.18 |
| pHILIC | 686.437 | 104.4885 | 59.1 | 0.18 |
| pHILIC | 686.437 | 118.0174 | 59.1 | 0.18 |
| pHILIC | 686.437 | 482.1286 | 59.1 | 0.18 |
| pHILIC | 686.437 | 377.3605 | 59 | 0.18 |
| pHILIC | 686.437 | 296.8097 | 59 | 0.18 |
| pHILIC | 686.437 | 524.8119 | 59 | 0.18 |
| pHILIC | 686.437 | 503.2832 | 59 | 0.18 |
| pHILIC | 686.437 | 167.3072 | 59 | 0.18 |
| pHILIC | 686.437 | 224.3939 | 58.9 | 0.18 |
| pHILIC | 686.437 | 169.8907 | 58.9 | 0.18 |
| pHILIC | 686.437 | 549.8813 | 58.9 | 0.18 |
| pHILIC | 686.437 | 269.244 | 58.9 | 0.18 |
| pHILIC | 686.437 | 550.5695 | 58.8 | 0.18 |
| pHILIC | 686.437 | 77.6955 | 58.8 | 0.18 |
| pHILIC | 686.437 | 76.709 | 58.8 | 0.18 |
| pHILIC | 686.437 | 280.4151 | 58.8 | 0.18 |
| pHILIC | 686.437 | 195.8038 | 58.8 | 0.18 |
| pHILIC | 686.437 | 119.6421 | 58.8 | 0.18 |
| pHILIC | 686.437 | 346.6763 | 58.8 | 0.18 |
| pHILIC | 686.437 | 508.6389 | 58.8 | 0.18 |
| pHILIC | 686.437 | 340.4771 | 58.8 | 0.18 |
| pHILIC | 686.437 | 160.7563 | 58.8 | 0.18 |
| pHILIC | 686.437 | 262.4586 | 58.7 | 0.18 |
| pHILIC | 686.437 | 126.9265 | 58.7 | 0.18 |
| pHILIC | 686.437 | 158.4041 | 58.7 | 0.18 |
| pHILIC | 686.437 | 460.526 | 58.7 | 0.18 |
| pHILIC | 686.437 | 117.2256 | 58.7 | 0.18 |

| | | | | |
|---|---|---|---|---|
| pHILIC | 686.437 | 161.4167 | 58.7 | 0.18 |
| pHILIC | 686.437 | 98.3515 | 58.7 | 0.18 |
| pHILIC | 686.437 | 79.8599 | 58.7 | 0.18 |
| pHILIC | 686.437 | 279.6148 | 58.7 | 0.18 |
| pHILIC | 686.437 | 119.9795 | 58.7 | 0.18 |
| pHILIC | 686.437 | 95.1268 | 58.7 | 0.18 |
| pHILIC | 686.437 | 98.0399 | 58.7 | 0.18 |
| pHILIC | 686.437 | 288.2459 | 58.7 | 0.18 |
| pHILIC | 686.437 | 137.4953 | 58.6 | 0.18 |
| pHILIC | 686.437 | 180.208 | 58.6 | 0.18 |
| pHILIC | 686.437 | 91.5677 | 58.6 | 0.18 |
| pHILIC | 686.437 | 139.4535 | 58.6 | 0.18 |
| pHILIC | 686.437 | 592.5688 | 58.6 | 0.18 |
| pHILIC | 686.437 | 397.9315 | 58.6 | 0.18 |
| pHILIC | 686.437 | 286.9168 | 58.5 | 0.18 |
| pHILIC | 686.437 | 416.3484 | 58.5 | 0.18 |
| pHILIC | 686.437 | 106.3021 | 58.5 | 0.18 |
| pHILIC | 686.437 | 311.6596 | 58.5 | 0.18 |
| pHILIC | 686.437 | 169.4497 | 58.5 | 0.18 |
| pHILIC | 686.437 | 396.0033 | 58.5 | 0.18 |
| pHILIC | 686.437 | 440.4998 | 58.5 | 0.18 |
| pHILIC | 686.437 | 109.3036 | 58.4 | 0.18 |
| pHILIC | 686.437 | 154.3504 | 58.4 | 0.18 |
| pHILIC | 686.437 | 86.0089 | 58.4 | 0.18 |
| pHILIC | 686.437 | 86.4241 | 58.4 | 0.18 |
| pHILIC | 686.437 | 456.167 | 58.3 | 0.18 |
| pHILIC | 686.437 | 185.3213 | 58.3 | 0.18 |
| pHILIC | 686.437 | 133.7435 | 58.3 | 0.18 |
| pHILIC | 686.437 | 200.5806 | 58.2 | 0.18 |
| pHILIC | 686.437 | 94.951 | 58.2 | 0.18 |
| pHILIC | 686.437 | 103.8647 | 58.2 | 0.18 |
| pHILIC | 686.437 | 78.5441 | 58.2 | 0.18 |
| pHILIC | 686.437 | 304.8798 | 58.2 | 0.18 |
| pHILIC | 686.437 | 91.4919 | 58.2 | 0.18 |
| pHILIC | 686.437 | 168.5242 | 58.2 | 0.18 |
| pHILIC | 686.437 | 215.9706 | 58.2 | 0.18 |
| pHILIC | 686.437 | 215.1464 | 58.2 | 0.18 |
| pHILIC | 686.437 | 121.3157 | 58.2 | 0.18 |
| pHILIC | 686.437 | 150.0789 | 58.2 | 0.18 |
| pHILIC | 686.437 | 77.3249 | 58.2 | 0.18 |
| pHILIC | 686.437 | 154.4548 | 58.2 | 0.18 |
| pHILIC | 686.437 | 84.4722 | 58.1 | 0.18 |

| | | | | |
|---|---|---|---|---|
| pHILIC | 686.437 | 91.2351 | 58.1 | 0.18 |
| pHILIC | 686.437 | 195.2752 | 58.1 | 0.18 |
| pHILIC | 686.437 | 292.387 | 58.1 | 0.18 |
| pHILIC | 686.437 | 108.2155 | 58.1 | 0.18 |
| pHILIC | 686.437 | 81.0938 | 58.1 | 0.18 |
| pHILIC | 686.437 | 112.285 | 58.1 | 0.18 |
| pHILIC | 686.437 | 80.4447 | 58.1 | 0.18 |
| pHILIC | 686.437 | 286.2129 | 58.1 | 0.18 |
| pHILIC | 686.437 | 243.2187 | 58.1 | 0.18 |
| pHILIC | 686.437 | 389.9624 | 58 | 0.18 |
| pHILIC | 686.437 | 177.1407 | 58 | 0.18 |
| pHILIC | 686.437 | 139.1437 | 58 | 0.18 |
| pHILIC | 686.437 | 268.8568 | 58 | 0.18 |
| pHILIC | 686.437 | 119.1334 | 58 | 0.18 |
| pHILIC | 686.437 | 300.3662 | 58 | 0.18 |
| pHILIC | 686.437 | 418.0064 | 58 | 0.18 |
| pHILIC | 686.437 | 101.4549 | 58 | 0.18 |
| pHILIC | 686.437 | 320.8426 | 58 | 0.18 |
| pHILIC | 686.437 | 282.9851 | 57.9 | 0.18 |
| pHILIC | 686.437 | 140.5828 | 57.9 | 0.18 |
| pHILIC | 686.437 | 205.8726 | 57.9 | 0.18 |
| pHILIC | 686.437 | 261.8116 | 57.9 | 0.18 |
| pHILIC | 686.437 | 346.2613 | 57.9 | 0.18 |
| pHILIC | 686.437 | 95.5524 | 57.9 | 0.18 |
| pHILIC | 686.437 | 368.3748 | 57.9 | 0.18 |
| pHILIC | 686.437 | 465.1394 | 57.9 | 0.18 |
| pHILIC | 686.437 | 91.2103 | 57.9 | 0.18 |
| pHILIC | 686.437 | 112.7392 | 57.9 | 0.18 |
| pHILIC | 686.437 | 259.7636 | 57.9 | 0.18 |
| pHILIC | 686.437 | 347.1745 | 57.9 | 0.18 |
| pHILIC | 686.437 | 491.8499 | 57.9 | 0.18 |
| pHILIC | 686.437 | 127.6677 | 57.9 | 0.18 |
| pHILIC | 686.437 | 94.8567 | 57.9 | 0.18 |
| pHILIC | 686.437 | 96.083 | 57.8 | 0.18 |
| pHILIC | 686.437 | 158.7211 | 57.8 | 0.18 |
| pHILIC | 686.437 | 321.8578 | 57.8 | 0.18 |
| pHILIC | 686.437 | 598.7302 | 57.8 | 0.18 |
| pHILIC | 686.437 | 125.4277 | 57.8 | 0.18 |
| pHILIC | 686.437 | 288.6175 | 57.8 | 0.18 |
| pHILIC | 686.437 | 187.1606 | 57.7 | 0.18 |
| pHILIC | 686.437 | 196.8707 | 57.7 | 0.18 |
| pHILIC | 686.437 | 288.7464 | 57.7 | 0.18 |

| | | | | |
|---|---|---|---|---|
| pHILIC | 686.437 | 201.3647 | 57.7 | 0.18 |
| pHILIC | 686.437 | 77.1343 | 57.6 | 0.18 |
| pHILIC | 686.437 | 529.2992 | 57.6 | 0.18 |
| pHILIC | 686.437 | 110.4483 | 57.6 | 0.18 |
| pHILIC | 686.437 | 76.4068 | 57.6 | 0.18 |
| pHILIC | 686.437 | 82.9285 | 57.6 | 0.18 |
| pHILIC | 686.437 | 93.948 | 57.5 | 0.18 |
| pHILIC | 686.437 | 78.8734 | 57.5 | 0.18 |
| pHILIC | 686.437 | 145.9972 | 57.5 | 0.18 |
| pHILIC | 686.437 | 73.746 | 57.5 | 0.18 |
| pHILIC | 686.437 | 214.3831 | 57.5 | 0.18 |
| pHILIC | 686.437 | 199.6523 | 57.5 | 0.18 |
| pHILIC | 686.437 | 272.3362 | 57.5 | 0.18 |
| pHILIC | 686.437 | 251.1205 | 57.5 | 0.18 |
| pHILIC | 686.437 | 141.1816 | 57.5 | 0.18 |
| pHILIC | 686.437 | 242.2291 | 57.5 | 0.18 |
| pHILIC | 686.437 | 129.7328 | 57.4 | 0.18 |
| pHILIC | 686.437 | 73.4927 | 57.4 | 0.18 |
| pHILIC | 686.437 | 108.7609 | 57.4 | 0.18 |
| pHILIC | 686.437 | 300.8354 | 57.4 | 0.18 |
| pHILIC | 686.437 | 90.17 | 57.4 | 0.18 |
| pHILIC | 686.437 | 106.5202 | 57.3 | 0.18 |
| pHILIC | 686.437 | 406.2462 | 57.3 | 0.18 |
| pHILIC | 686.437 | 96.5819 | 57.2 | 0.18 |
| pHILIC | 686.437 | 94.1311 | 57.2 | 0.18 |
| pHILIC | 686.437 | 543.4503 | 57.2 | 0.18 |
| pHILIC | 686.437 | 90.1465 | 57.2 | 0.18 |
| pHILIC | 686.437 | 176.5982 | 57.2 | 0.18 |
| pHILIC | 686.437 | 158.7378 | 57.2 | 0.18 |
| pHILIC | 686.437 | 80.3419 | 57.1 | 0.18 |
| pHILIC | 686.437 | 339.8085 | 57.1 | 0.18 |
| pHILIC | 686.437 | 74.4335 | 57.1 | 0.18 |
| pHILIC | 686.437 | 424.0409 | 57.1 | 0.18 |
| pHILIC | 686.437 | 270.9699 | 57.1 | 0.18 |
| pHILIC | 686.437 | 246.8102 | 57.1 | 0.18 |
| pHILIC | 686.437 | 227.6427 | 57 | 0.18 |
| pHILIC | 686.437 | 112.3679 | 57 | 0.18 |
| pHILIC | 686.437 | 150.5145 | 57 | 0.18 |
| pHILIC | 686.437 | 247.4496 | 57 | 0.18 |
| pHILIC | 686.437 | 258.3818 | 57 | 0.18 |
| pHILIC | 686.437 | 295.7321 | 57 | 0.18 |
| pHILIC | 686.437 | 445.8337 | 57 | 0.18 |

| | | | | |
|---|---|---|---|---|
| pHILIC | 686.437 | 110.0286 | 56.9 | 0.18 |
| pHILIC | 686.437 | 140.3137 | 56.9 | 0.18 |
| pHILIC | 686.437 | 104.5799 | 56.9 | 0.18 |
| pHILIC | 686.437 | 137.7724 | 56.9 | 0.18 |
| pHILIC | 686.437 | 577.4136 | 56.9 | 0.18 |
| pHILIC | 686.437 | 119.1744 | 56.8 | 0.18 |
| pHILIC | 686.437 | 242.9038 | 56.8 | 0.18 |
| pHILIC | 686.437 | 98.0663 | 56.8 | 0.18 |
| pHILIC | 686.437 | 103.6871 | 56.8 | 0.18 |
| pHILIC | 686.437 | 93.0043 | 56.8 | 0.18 |
| pHILIC | 686.437 | 318.2283 | 56.8 | 0.18 |
| pHILIC | 686.437 | 203.2175 | 56.8 | 0.18 |
| pHILIC | 686.437 | 110.5415 | 56.7 | 0.18 |
| pHILIC | 686.437 | 209.3791 | 56.7 | 0.18 |
| pHILIC | 686.437 | 281.2114 | 56.7 | 0.18 |
| pHILIC | 686.437 | 130.1664 | 56.6 | 0.18 |
| pHILIC | 686.437 | 221.4836 | 56.6 | 0.18 |
| pHILIC | 686.437 | 132.6144 | 56.6 | 0.18 |
| pHILIC | 686.437 | 356.6115 | 56.6 | 0.18 |
| pHILIC | 686.437 | 401.3941 | 56.5 | 0.18 |
| pHILIC | 686.437 | 199.7671 | 56.5 | 0.18 |
| pHILIC | 686.437 | 115.2636 | 56.5 | 0.18 |
| pHILIC | 686.437 | 233.8802 | 56.5 | 0.17 |
| pHILIC | 686.437 | 93.8946 | 56.4 | 0.17 |
| pHILIC | 686.437 | 166.6004 | 56.4 | 0.17 |
| pHILIC | 686.437 | 132.7319 | 56.4 | 0.17 |
| pHILIC | 686.437 | 242.1303 | 56.4 | 0.17 |
| pHILIC | 686.437 | 298.7603 | 56.3 | 0.17 |
| pHILIC | 686.437 | 572.8766 | 56.3 | 0.17 |
| pHILIC | 686.437 | 150.2958 | 56.3 | 0.17 |
| pHILIC | 686.437 | 129.352 | 56.3 | 0.17 |
| pHILIC | 686.437 | 109.3447 | 56.2 | 0.17 |
| pHILIC | 686.437 | 590.2905 | 56.2 | 0.17 |
| pHILIC | 686.437 | 106.3113 | 56.2 | 0.17 |
| pHILIC | 686.437 | 98.656 | 56.2 | 0.17 |
| pHILIC | 686.437 | 73.7893 | 56.2 | 0.17 |
| pHILIC | 686.437 | 366.6151 | 56.1 | 0.17 |
| pHILIC | 686.437 | 126.5172 | 56.1 | 0.17 |
| pHILIC | 686.437 | 223.556 | 56.1 | 0.17 |
| pHILIC | 686.437 | 129.2585 | 56 | 0.17 |
| pHILIC | 686.437 | 562.4405 | 56 | 0.17 |
| pHILIC | 686.437 | 252.4196 | 56 | 0.17 |

| | | | | |
|---|---|---|---|---|
| pHILIC | 686.437 | 93.5962 | 56 | 0.17 |
| pHILIC | 686.437 | 113.8294 | 55.9 | 0.17 |
| pHILIC | 686.437 | 81.2432 | 55.9 | 0.17 |
| pHILIC | 686.437 | 118.7567 | 55.9 | 0.17 |
| pHILIC | 686.437 | 166.6595 | 55.9 | 0.17 |
| pHILIC | 686.437 | 76.8708 | 55.9 | 0.17 |
| pHILIC | 686.437 | 140.7459 | 55.9 | 0.17 |
| pHILIC | 686.437 | 276.3337 | 55.9 | 0.17 |
| pHILIC | 686.437 | 74.4667 | 55.9 | 0.17 |
| pHILIC | 686.437 | 80.6646 | 55.8 | 0.17 |
| pHILIC | 686.437 | 296.2553 | 55.8 | 0.17 |
| pHILIC | 686.437 | 82.8199 | 55.8 | 0.17 |
| pHILIC | 686.437 | 178.6009 | 55.8 | 0.17 |
| pHILIC | 686.437 | 152.3513 | 55.8 | 0.17 |
| pHILIC | 686.437 | 105.1243 | 55.7 | 0.17 |
| pHILIC | 686.437 | 80.1295 | 55.7 | 0.17 |
| pHILIC | 686.437 | 81.3347 | 55.7 | 0.17 |
| pHILIC | 686.437 | 165.9668 | 55.7 | 0.17 |
| pHILIC | 686.437 | 112.1539 | 55.7 | 0.17 |
| pHILIC | 686.437 | 629.5026 | 55.7 | 0.17 |
| pHILIC | 686.437 | 325.5619 | 55.6 | 0.17 |
| pHILIC | 686.437 | 117.241 | 55.6 | 0.17 |
| pHILIC | 686.437 | 170.7256 | 55.6 | 0.17 |
| pHILIC | 686.437 | 78.6478 | 55.6 | 0.17 |
| pHILIC | 686.437 | 206.7582 | 55.6 | 0.17 |
| pHILIC | 686.437 | 301.2095 | 55.6 | 0.17 |
| pHILIC | 686.437 | 108.2076 | 55.6 | 0.17 |
| pHILIC | 686.437 | 328.3733 | 55.6 | 0.17 |
| pHILIC | 686.437 | 80.8097 | 55.5 | 0.17 |
| pHILIC | 686.437 | 109.4066 | 55.5 | 0.17 |
| pHILIC | 686.437 | 96.2924 | 55.5 | 0.17 |
| pHILIC | 686.437 | 83.3245 | 55.5 | 0.17 |
| pHILIC | 686.437 | 131.1862 | 55.5 | 0.17 |
| pHILIC | 686.437 | 103.6568 | 55.5 | 0.17 |
| pHILIC | 686.437 | 134.7504 | 55.4 | 0.17 |
| pHILIC | 686.437 | 202.8289 | 55.4 | 0.17 |
| pHILIC | 686.437 | 102.4757 | 55.4 | 0.17 |
| pHILIC | 686.437 | 75.0316 | 55.3 | 0.17 |
| pHILIC | 686.437 | 244.618 | 55.3 | 0.17 |
| pHILIC | 686.437 | 88.8256 | 55.3 | 0.17 |
| pHILIC | 686.437 | 93.5734 | 55.3 | 0.17 |
| pHILIC | 686.437 | 75.3178 | 55.3 | 0.17 |

| | | | | |
|---|---|---|---|---|
| pHILIC | 686.437 | 96.2505 | 55.3 | 0.17 |
| pHILIC | 686.437 | 79.6503 | 55.3 | 0.17 |
| pHILIC | 686.437 | 213.9008 | 55.2 | 0.17 |
| pHILIC | 686.437 | 96.2651 | 55.2 | 0.17 |
| pHILIC | 686.437 | 393.1365 | 55.2 | 0.17 |
| pHILIC | 686.437 | 112.9333 | 55.1 | 0.17 |
| pHILIC | 686.437 | 351.2244 | 55.1 | 0.17 |
| pHILIC | 686.437 | 107.9081 | 55.1 | 0.17 |
| pHILIC | 686.437 | 492.036 | 55.1 | 0.17 |
| pHILIC | 686.437 | 154.7074 | 55.1 | 0.17 |
| pHILIC | 686.437 | 85.9129 | 55.1 | 0.17 |
| pHILIC | 686.437 | 74.91 | 55 | 0.17 |
| pHILIC | 686.437 | 72.882 | 55 | 0.17 |
| pHILIC | 686.437 | 80.7582 | 55 | 0.17 |
| pHILIC | 686.437 | 113.1107 | 55 | 0.17 |
| pHILIC | 686.437 | 203.6218 | 54.9 | 0.17 |
| pHILIC | 686.437 | 463.4251 | 54.9 | 0.17 |
| pHILIC | 686.437 | 232.3679 | 54.9 | 0.17 |
| pHILIC | 686.437 | 124.1033 | 54.7 | 0.17 |
| pHILIC | 686.437 | 124.9127 | 54.7 | 0.17 |
| pHILIC | 686.437 | 93.6168 | 54.7 | 0.17 |
| pHILIC | 686.437 | 298.8782 | 54.7 | 0.17 |
| pHILIC | 686.437 | 74.9386 | 54.7 | 0.17 |
| pHILIC | 686.437 | 97.294 | 54.7 | 0.17 |
| pHILIC | 686.437 | 73.4254 | 54.7 | 0.17 |
| pHILIC | 686.437 | 156.3286 | 54.6 | 0.17 |
| pHILIC | 686.437 | 114.4736 | 54.6 | 0.17 |
| pHILIC | 686.437 | 86.8531 | 54.5 | 0.17 |
| pHILIC | 686.437 | 130.9328 | 54.5 | 0.17 |
| pHILIC | 686.437 | 104.4272 | 54.5 | 0.17 |
| pHILIC | 686.437 | 178.0125 | 54.5 | 0.17 |
| pHILIC | 686.437 | 142.1349 | 54.5 | 0.17 |
| pHILIC | 686.437 | 94.8301 | 54.5 | 0.17 |
| pHILIC | 686.437 | 128.2223 | 54.4 | 0.17 |
| pHILIC | 686.437 | 277.7896 | 54.4 | 0.17 |
| pHILIC | 686.437 | 108.4903 | 54.4 | 0.17 |
| pHILIC | 686.437 | 235.067 | 54.4 | 0.17 |
| pHILIC | 686.437 | 106.0844 | 54.4 | 0.17 |
| pHILIC | 686.437 | 154.2999 | 54.3 | 0.17 |
| pHILIC | 686.437 | 97.6046 | 54.2 | 0.17 |
| pHILIC | 686.437 | 137.7609 | 54.1 | 0.17 |
| pHILIC | 686.437 | 74.3772 | 54.1 | 0.17 |

| | | | | |
|---|---|---|---|---|
| pHILIC | 686.437 | 77.3104 | 54.1 | 0.17 |
| pHILIC | 686.437 | 121.3015 | 54.1 | 0.17 |
| pHILIC | 686.437 | 96.3429 | 54 | 0.17 |
| pHILIC | 686.437 | 91.4007 | 54 | 0.17 |
| pHILIC | 686.437 | 81.9405 | 53.9 | 0.17 |
| pHILIC | 686.437 | 164.255 | 53.9 | 0.17 |
| pHILIC | 686.437 | 167.7727 | 53.8 | 0.17 |
| pHILIC | 686.437 | 211.2261 | 53.7 | 0.17 |
| pHILIC | 686.437 | 85.9426 | 53.7 | 0.17 |
| pHILIC | 686.437 | 270.933 | 53.6 | 0.17 |
| pHILIC | 686.437 | 316.4231 | 53.6 | 0.17 |
| pHILIC | 686.437 | 72.7897 | 53.6 | 0.17 |
| pHILIC | 686.437 | 111.669 | 53.6 | 0.17 |
| pHILIC | 686.437 | 164.2727 | 53.6 | 0.17 |
| pHILIC | 686.437 | 84.5406 | 53.5 | 0.17 |
| pHILIC | 686.437 | 162.374 | 53.5 | 0.17 |
| pHILIC | 686.437 | 152.552 | 53.5 | 0.17 |
| pHILIC | 686.437 | 104.9263 | 53.4 | 0.17 |
| pHILIC | 686.437 | 79.4242 | 53.4 | 0.17 |
| pHILIC | 686.437 | 640.8844 | 53.3 | 0.17 |
| pHILIC | 686.437 | 121.9293 | 53.3 | 0.17 |
| pHILIC | 686.437 | 173.7704 | 53.3 | 0.17 |
| pHILIC | 686.437 | 376.4382 | 53.3 | 0.17 |
| pHILIC | 686.437 | 398.4579 | 53.3 | 0.17 |
| pHILIC | 686.437 | 514.1318 | 53.2 | 0.16 |
| pHILIC | 686.437 | 87.5628 | 53.2 | 0.16 |
| pHILIC | 686.437 | 117.0358 | 53.1 | 0.16 |
| pHILIC | 686.437 | 145.0435 | 53.1 | 0.16 |
| pHILIC | 686.437 | 276.1627 | 53 | 0.16 |
| pHILIC | 686.437 | 648.5198 | 53 | 0.16 |
| pHILIC | 686.437 | 552.9294 | 52.9 | 0.16 |
| pHILIC | 686.437 | 84.8884 | 52.8 | 0.16 |
| pHILIC | 686.437 | 92.0658 | 52.8 | 0.16 |
| pHILIC | 686.437 | 82.2233 | 52.8 | 0.16 |
| pHILIC | 686.437 | 191.2373 | 52.8 | 0.16 |
| pHILIC | 686.437 | 77.3666 | 52.7 | 0.16 |
| pHILIC | 686.437 | 501.9767 | 52.7 | 0.16 |
| pHILIC | 686.437 | 261.6243 | 52.7 | 0.16 |
| pHILIC | 686.437 | 404.391 | 52.7 | 0.16 |
| pHILIC | 686.437 | 214.4174 | 52.7 | 0.16 |
| pHILIC | 686.437 | 522.0549 | 52.6 | 0.16 |
| pHILIC | 686.437 | 107.9945 | 52.6 | 0.16 |

| | | | | |
|---|---|---|---|---|
| pHILIC | 686.437 | 362.0836 | 52.5 | 0.16 |
| pHILIC | 686.437 | 97.5834 | 52.5 | 0.16 |
| pHILIC | 686.437 | 99.3023 | 52.5 | 0.16 |
| pHILIC | 686.437 | 149.0806 | 52.4 | 0.16 |
| pHILIC | 686.437 | 642.3615 | 52.4 | 0.16 |
| pHILIC | 686.437 | 93.6454 | 52.3 | 0.16 |
| pHILIC | 686.437 | 218.432 | 52.2 | 0.16 |
| pHILIC | 686.437 | 352.6762 | 52.1 | 0.16 |
| pHILIC | 686.437 | 198.1756 | 52.1 | 0.16 |
| pHILIC | 686.437 | 646.9417 | 52.1 | 0.16 |
| pHILIC | 686.437 | 267.1359 | 52 | 0.16 |
| pHILIC | 686.437 | 160.6864 | 51.9 | 0.16 |
| pHILIC | 686.437 | 243.7397 | 51.9 | 0.16 |
| pHILIC | 686.437 | 361.5228 | 51.9 | 0.16 |
| pHILIC | 686.437 | 161.1123 | 51.9 | 0.16 |
| pHILIC | 686.437 | 137.5206 | 51.9 | 0.16 |
| pHILIC | 686.437 | 89.0938 | 51.8 | 0.16 |
| pHILIC | 686.437 | 166.4801 | 51.8 | 0.16 |
| pHILIC | 686.437 | 298.3392 | 51.8 | 0.16 |
| pHILIC | 686.437 | 101.9076 | 51.8 | 0.16 |
| pHILIC | 686.437 | 180.8654 | 51.6 | 0.16 |
| pHILIC | 686.437 | 105.2867 | 51.6 | 0.16 |
| pHILIC | 686.437 | 384.9622 | 51.5 | 0.16 |
| pHILIC | 686.437 | 711.2745 | 51.4 | 0.16 |
| pHILIC | 686.437 | 525.7455 | 51.4 | 0.16 |
| pHILIC | 686.437 | 236.3693 | 51.4 | 0.16 |
| pHILIC | 686.437 | 138.9979 | 51.4 | 0.16 |
| pHILIC | 686.437 | 141.1415 | 51.3 | 0.16 |
| pHILIC | 686.437 | 197.976 | 51.3 | 0.16 |
| pHILIC | 686.437 | 278.1944 | 51.2 | 0.16 |
| pHILIC | 686.437 | 444.5236 | 51.2 | 0.16 |
| pHILIC | 686.437 | 98.6855 | 51.2 | 0.16 |
| pHILIC | 686.437 | 153.4754 | 51.1 | 0.16 |
| pHILIC | 686.437 | 76.8175 | 51.1 | 0.16 |
| pHILIC | 686.437 | 164.9179 | 51.1 | 0.16 |
| pHILIC | 686.437 | 690.0493 | 51.1 | 0.16 |
| pHILIC | 686.437 | 77.9787 | 51 | 0.16 |
| pHILIC | 686.437 | 126.0042 | 50.9 | 0.16 |
| pHILIC | 686.437 | 73.7241 | 50.9 | 0.16 |
| pHILIC | 686.437 | 125.5461 | 50.8 | 0.16 |
| pHILIC | 686.437 | 93.0258 | 50.8 | 0.16 |
| pHILIC | 686.437 | 128.9596 | 50.7 | 0.16 |

| | | | | |
|---|---|---|---|---|
| pHILIC | 686.437 | 114.3033 | 50.6 | 0.16 |
| pHILIC | 686.437 | 200.1894 | 50.6 | 0.16 |
| pHILIC | 686.437 | 562.7339 | 50.6 | 0.16 |
| pHILIC | 686.437 | 122.0734 | 50.6 | 0.16 |
| pHILIC | 686.437 | 107.4278 | 50.5 | 0.16 |
| pHILIC | 686.437 | 515.6095 | 50.5 | 0.16 |
| pHILIC | 686.437 | 440.8535 | 50.4 | 0.16 |
| pHILIC | 686.437 | 80.5529 | 50.4 | 0.16 |
| pHILIC | 686.437 | 300.2157 | 50.4 | 0.16 |
| pHILIC | 686.437 | 164.3826 | 50.3 | 0.16 |
| pHILIC | 686.437 | 678.4663 | 50.3 | 0.16 |
| pHILIC | 686.437 | 89.84 | 50.3 | 0.16 |
| pHILIC | 686.437 | 116.5467 | 50.3 | 0.16 |
| pHILIC | 686.437 | 187.7642 | 50.3 | 0.16 |
| pHILIC | 686.437 | 705.6928 | 50.3 | 0.16 |
| pHILIC | 686.437 | 209.843 | 50.3 | 0.16 |
| pHILIC | 686.437 | 152.6048 | 50.2 | 0.16 |
| pHILIC | 686.437 | 85.9633 | 50.2 | 0.16 |
| pHILIC | 686.437 | 274.6884 | 50.2 | 0.16 |
| pHILIC | 686.437 | 449.2075 | 50.2 | 0.16 |
| pHILIC | 686.437 | 78.5256 | 50.1 | 0.16 |
| pHILIC | 686.437 | 202.1049 | 50.1 | 0.16 |
| pHILIC | 686.437 | 86.8639 | 50.1 | 0.16 |
| pHILIC | 686.437 | 210.6236 | 50 | 0.16 |
| pHILIC | 686.437 | 439.717 | 50 | 0.15 |
| pHILIC | 686.437 | 85.3783 | 50 | 0.15 |
| pHILIC | 686.437 | 554.9642 | 49.9 | 0.15 |
| pHILIC | 686.437 | 74.2433 | 49.8 | 0.15 |
| pHILIC | 686.437 | 522.5583 | 49.6 | 0.15 |
| pHILIC | 686.437 | 294.5211 | 49.5 | 0.15 |
| pHILIC | 686.437 | 253.2333 | 49.5 | 0.15 |
| pHILIC | 686.437 | 545.4808 | 49.5 | 0.15 |
| pHILIC | 686.437 | 284.5858 | 49.5 | 0.15 |
| pHILIC | 686.437 | 228.5691 | 49.5 | 0.15 |
| pHILIC | 686.437 | 82.0019 | 49.5 | 0.15 |
| pHILIC | 686.437 | 531.204 | 49.5 | 0.15 |
| pHILIC | 686.437 | 319.2657 | 49.3 | 0.15 |
| pHILIC | 686.437 | 156.4484 | 49.3 | 0.15 |
| pHILIC | 686.437 | 207.6055 | 49.3 | 0.15 |
| pHILIC | 686.437 | 593.8714 | 49.3 | 0.15 |
| pHILIC | 686.437 | 119.8925 | 49.2 | 0.15 |
| pHILIC | 686.437 | 150.8194 | 49.2 | 0.15 |

| | | | | |
|---|---|---|---|---|
| pHILIC | 686.437 | 304.2891 | 49.2 | 0.15 |
| pHILIC | 686.437 | 132.6806 | 49.1 | 0.15 |
| pHILIC | 686.437 | 304.3276 | 49 | 0.15 |
| pHILIC | 686.437 | 211.0859 | 49 | 0.15 |
| pHILIC | 686.437 | 106.4693 | 49 | 0.15 |
| pHILIC | 686.437 | 86.2135 | 48.8 | 0.15 |
| pHILIC | 686.437 | 239.6517 | 48.7 | 0.15 |
| pHILIC | 686.437 | 97.7212 | 48.7 | 0.15 |
| pHILIC | 686.437 | 116.2993 | 48.7 | 0.15 |
| pHILIC | 686.437 | 115.8244 | 48.7 | 0.15 |
| pHILIC | 686.437 | 261.5216 | 48.7 | 0.15 |
| pHILIC | 686.437 | 688.4203 | 48.7 | 0.15 |
| pHILIC | 686.437 | 538.3755 | 48.6 | 0.15 |
| pHILIC | 686.437 | 107.7802 | 48.5 | 0.15 |
| pHILIC | 686.437 | 461.2594 | 48.5 | 0.15 |
| pHILIC | 686.437 | 94.5559 | 48.3 | 0.15 |
| pHILIC | 686.437 | 116.2436 | 48.3 | 0.15 |
| pHILIC | 686.437 | 119.4013 | 48.2 | 0.15 |
| pHILIC | 686.437 | 74.8937 | 48.2 | 0.15 |
| pHILIC | 686.437 | 648.0717 | 48 | 0.15 |
| pHILIC | 686.437 | 115.2996 | 48 | 0.15 |
| pHILIC | 686.437 | 494.2991 | 48 | 0.15 |
| pHILIC | 686.437 | 155.7992 | 47.9 | 0.15 |
| pHILIC | 686.437 | 328.6808 | 47.8 | 0.15 |
| pHILIC | 686.437 | 297.0261 | 47.8 | 0.15 |
| pHILIC | 686.437 | 303.2747 | 47.8 | 0.15 |
| pHILIC | 686.437 | 254.8248 | 47.8 | 0.15 |
| pHILIC | 686.437 | 381.4244 | 47.8 | 0.15 |
| pHILIC | 686.437 | 576.3055 | 47.7 | 0.15 |
| pHILIC | 686.437 | 343.171 | 47.7 | 0.15 |
| pHILIC | 686.437 | 86.0972 | 47.6 | 0.15 |
| pHILIC | 686.437 | 114.4369 | 47.6 | 0.15 |
| pHILIC | 686.437 | 282.4586 | 47.6 | 0.15 |
| pHILIC | 686.437 | 462.1324 | 47.6 | 0.15 |
| pHILIC | 686.437 | 273.1196 | 47.6 | 0.15 |
| pHILIC | 686.437 | 335.4668 | 47.5 | 0.15 |
| pHILIC | 686.437 | 93.5688 | 47.5 | 0.15 |
| pHILIC | 686.437 | 77.2925 | 47.5 | 0.15 |
| pHILIC | 686.437 | 264.0184 | 47.4 | 0.15 |
| pHILIC | 686.437 | 518.2319 | 47.4 | 0.15 |
| pHILIC | 686.437 | 75.7182 | 47.4 | 0.15 |
| pHILIC | 686.437 | 123.3001 | 47.3 | 0.15 |

| | | | | |
|---|---|---|---|---|
| pHILIC | 686.437 | 73.1992 | 47.3 | 0.15 |
| pHILIC | 686.437 | 86.4417 | 47.3 | 0.15 |
| pHILIC | 686.437 | 120.8571 | 47.3 | 0.15 |
| pHILIC | 686.437 | 92.2021 | 47.3 | 0.15 |
| pHILIC | 686.437 | 147.9154 | 47.2 | 0.15 |
| pHILIC | 686.437 | 168.9129 | 47.1 | 0.15 |
| pHILIC | 686.437 | 280.2401 | 47.1 | 0.15 |
| pHILIC | 686.437 | 384.7192 | 47.1 | 0.15 |
| pHILIC | 686.437 | 404.2792 | 47.1 | 0.15 |
| pHILIC | 686.437 | 73.5805 | 47.1 | 0.15 |
| pHILIC | 686.437 | 108.683 | 47.1 | 0.15 |
| pHILIC | 686.437 | 280.1383 | 47 | 0.15 |
| pHILIC | 686.437 | 365.8144 | 46.9 | 0.15 |
| pHILIC | 686.437 | 101.309 | 46.8 | 0.14 |
| pHILIC | 686.437 | 150.0348 | 46.8 | 0.14 |
| pHILIC | 686.437 | 93.1921 | 46.7 | 0.14 |
| pHILIC | 686.437 | 334.1788 | 46.7 | 0.14 |
| pHILIC | 686.437 | 300.4029 | 46.6 | 0.14 |
| pHILIC | 686.437 | 417.2389 | 46.4 | 0.14 |
| pHILIC | 686.437 | 259.1221 | 46.4 | 0.14 |
| pHILIC | 686.437 | 84.5017 | 46.4 | 0.14 |
| pHILIC | 686.437 | 74.9137 | 46.3 | 0.14 |
| pHILIC | 686.437 | 149.106 | 46.3 | 0.14 |
| pHILIC | 686.437 | 92.1117 | 46.2 | 0.14 |
| pHILIC | 686.437 | 78.5084 | 46.2 | 0.14 |
| pHILIC | 686.437 | 351.7874 | 46.2 | 0.14 |
| pHILIC | 686.437 | 618.075 | 46.1 | 0.14 |
| pHILIC | 686.437 | 102.739 | 46.1 | 0.14 |
| pHILIC | 686.437 | 209.3961 | 46.1 | 0.14 |
| pHILIC | 686.437 | 101.5346 | 46 | 0.14 |
| pHILIC | 686.437 | 72.8539 | 46 | 0.14 |
| pHILIC | 686.437 | 89.0025 | 46 | 0.14 |
| pHILIC | 686.437 | 164.1925 | 45.9 | 0.14 |
| pHILIC | 686.437 | 73.9432 | 45.9 | 0.14 |
| pHILIC | 686.437 | 314.8094 | 45.9 | 0.14 |
| pHILIC | 686.437 | 248.5416 | 45.8 | 0.14 |
| pHILIC | 686.437 | 217.3721 | 45.8 | 0.14 |
| pHILIC | 686.437 | 155.8721 | 45.7 | 0.14 |
| pHILIC | 686.437 | 513.2709 | 45.7 | 0.14 |
| pHILIC | 686.437 | 72.8705 | 45.7 | 0.14 |
| pHILIC | 686.437 | 196.4963 | 45.6 | 0.14 |
| pHILIC | 686.437 | 313.6725 | 45.6 | 0.14 |

| | | | | |
|---|---|---|---|---|
| pHILIC | 686.437 | 351.1223 | 45.6 | 0.14 |
| pHILIC | 686.437 | 537.8389 | 45.6 | 0.14 |
| pHILIC | 686.437 | 95.3664 | 45.6 | 0.14 |
| pHILIC | 686.437 | 117.2045 | 45.6 | 0.14 |
| pHILIC | 686.437 | 77.59 | 45.5 | 0.14 |
| pHILIC | 686.437 | 93.5362 | 45.5 | 0.14 |
| pHILIC | 686.437 | 378.9981 | 45.5 | 0.14 |
| pHILIC | 686.437 | 548.7811 | 45.3 | 0.14 |
| pHILIC | 686.437 | 210.5056 | 45.3 | 0.14 |
| pHILIC | 686.437 | 240.5186 | 45.3 | 0.14 |
| pHILIC | 686.437 | 85.5438 | 45.3 | 0.14 |
| pHILIC | 686.437 | 183.4094 | 45.3 | 0.14 |
| pHILIC | 686.437 | 77.5984 | 45.2 | 0.14 |
| pHILIC | 686.437 | 221.7688 | 45.2 | 0.14 |
| pHILIC | 686.437 | 126.5633 | 45.2 | 0.14 |
| pHILIC | 686.437 | 72.9542 | 45.2 | 0.14 |
| pHILIC | 686.437 | 132.7096 | 45.1 | 0.14 |
| pHILIC | 686.437 | 75.661 | 45.1 | 0.14 |
| pHILIC | 686.437 | 134.9737 | 45 | 0.14 |
| pHILIC | 686.437 | 658.6539 | 44.9 | 0.14 |
| pHILIC | 686.437 | 148.4658 | 44.9 | 0.14 |
| pHILIC | 686.437 | 104.1372 | 44.8 | 0.14 |
| pHILIC | 686.437 | 106.269 | 44.7 | 0.14 |
| pHILIC | 686.437 | 162.9566 | 44.5 | 0.14 |
| pHILIC | 686.437 | 112.603 | 44.5 | 0.14 |
| pHILIC | 686.437 | 79.7484 | 44.5 | 0.14 |
| pHILIC | 686.437 | 215.0248 | 44.4 | 0.14 |
| pHILIC | 686.437 | 99.7159 | 44.3 | 0.14 |
| pHILIC | 686.437 | 641.3058 | 44.2 | 0.14 |
| pHILIC | 686.437 | 160.9473 | 44.2 | 0.14 |
| pHILIC | 686.437 | 117.9114 | 44.1 | 0.14 |
| pHILIC | 686.437 | 112.4568 | 44.1 | 0.14 |
| pHILIC | 686.437 | 302.5277 | 44 | 0.14 |
| pHILIC | 686.437 | 120.2087 | 44 | 0.14 |
| pHILIC | 686.437 | 148.7658 | 43.7 | 0.14 |
| pHILIC | 686.437 | 239.5272 | 43.7 | 0.14 |
| pHILIC | 686.437 | 88.0228 | 43.5 | 0.13 |
| pHILIC | 686.437 | 104.0935 | 43.4 | 0.13 |
| pHILIC | 686.437 | 95.7302 | 43.3 | 0.13 |
| pHILIC | 686.437 | 134.6388 | 43.3 | 0.13 |
| pHILIC | 686.437 | 163.1364 | 43 | 0.13 |
| pHILIC | 686.437 | 335.6129 | 42.8 | 0.13 |

| | | | | |
|---|---|---|---|---|
| pHILIC | 686.437 | 83.49 | 42.7 | 0.13 |
| pHILIC | 686.437 | 106.0395 | 42.7 | 0.13 |
| pHILIC | 686.437 | 380.3997 | 42.7 | 0.13 |
| pHILIC | 686.437 | 84.8104 | 42.6 | 0.13 |
| pHILIC | 686.437 | 85.8639 | 42.4 | 0.13 |
| pHILIC | 686.437 | 195.5672 | 42.4 | 0.13 |
| pHILIC | 686.437 | 210.1327 | 42.2 | 0.13 |
| pHILIC | 686.437 | 85.9526 | 42 | 0.13 |
| pHILIC | 686.437 | 75.7837 | 40.8 | 0.13 |
| pHILIC | 686.437 | 91.4655 | 40.6 | 0.13 |
| pHILIC | 686.437 | 178.3668 | 40.5 | 0.13 |
| pHILIC | 686.437 | 124.1393 | 40.5 | 0.13 |
| pHILIC | 686.437 | 98.0585 | 39.1 | 0.12 |
| pHILIC | 706.536 | 184.0733 | 47357.1 | 100 |
| pHILIC | 706.536 | 706.5008 | 35936.7 | 75.88 |
| pHILIC | 706.536 | 86.0963 | 8465.9 | 17.88 |
| pHILIC | 706.536 | 112.8958 | 5566.9 | 11.76 |
| pHILIC | 706.536 | 124.9999 | 4932.8 | 10.42 |
| pHILIC | 706.536 | 228.2198 | 4120.2 | 8.7 |
| pHILIC | 706.536 | 707.4637 | 2597.7 | 5.49 |
| pHILIC | 706.536 | 323.295 | 2208.5 | 4.66 |
| pHILIC | 706.536 | 327.0782 | 2084.6 | 4.4 |
| pHILIC | 706.536 | 309.2778 | 1633.6 | 3.45 |
| pHILIC | 706.536 | 580.3613 | 1286.1 | 2.72 |
| pHILIC | 706.536 | 328.0809 | 1153.5 | 2.44 |
| pHILIC | 706.536 | 89.0597 | 417.7 | 0.88 |
| pHILIC | 706.536 | 454.2338 | 285.4 | 0.6 |
| pHILIC | 706.536 | 453.2105 | 190.9 | 0.4 |
| pHILIC | 706.536 | 93.6578 | 189 | 0.4 |
| pHILIC | 706.536 | 95.0857 | 186.9 | 0.39 |
| pHILIC | 706.536 | 114.8926 | 179.5 | 0.38 |
| pHILIC | 706.536 | 81.0697 | 157.6 | 0.33 |
| pHILIC | 706.536 | 176.8756 | 144.7 | 0.31 |
| pHILIC | 706.536 | 310.2809 | 144.6 | 0.31 |
| pHILIC | 706.536 | 93.6002 | 144 | 0.3 |
| pHILIC | 706.536 | 93.6717 | 136 | 0.29 |
| pHILIC | 706.536 | 185.077 | 132.8 | 0.28 |
| pHILIC | 706.536 | 254.7986 | 128.8 | 0.27 |
| pHILIC | 706.536 | 623.0293 | 126.2 | 0.27 |
| pHILIC | 706.536 | 531.8515 | 125.7 | 0.27 |
| pHILIC | 706.536 | 247.0641 | 125.6 | 0.27 |
| pHILIC | 706.536 | 238.9302 | 125 | 0.26 |

| | | | | |
|---|---|---|---|---|
| pHILIC | 706.536 | 245.5808 | 124.8 | 0.26 |
| pHILIC | 706.536 | 300.3943 | 123.9 | 0.26 |
| pHILIC | 706.536 | 425.7558 | 122.7 | 0.26 |
| pHILIC | 706.536 | 104.1309 | 122.4 | 0.26 |
| pHILIC | 706.536 | 376.984 | 121.9 | 0.26 |
| pHILIC | 706.536 | 226.0976 | 121.5 | 0.26 |
| pHILIC | 706.536 | 153.1598 | 119.5 | 0.25 |
| pHILIC | 706.536 | 607.083 | 119.4 | 0.25 |
| pHILIC | 706.536 | 81.2946 | 117.9 | 0.25 |
| pHILIC | 706.536 | 350.5749 | 117.8 | 0.25 |
| pHILIC | 706.536 | 233.2815 | 117.6 | 0.25 |
| pHILIC | 706.536 | 149.822 | 117.4 | 0.25 |
| pHILIC | 706.536 | 93.9579 | 116.1 | 0.25 |
| pHILIC | 706.536 | 88.7719 | 115.8 | 0.24 |
| pHILIC | 706.536 | 151.3 | 115.7 | 0.24 |
| pHILIC | 706.536 | 232.3358 | 114.7 | 0.24 |
| pHILIC | 706.536 | 357.0722 | 114.5 | 0.24 |
| pHILIC | 706.536 | 172.4001 | 112.3 | 0.24 |
| pHILIC | 706.536 | 94.101 | 112.2 | 0.24 |
| pHILIC | 706.536 | 400.6926 | 112.1 | 0.24 |
| pHILIC | 706.536 | 203.6456 | 110.2 | 0.23 |
| pHILIC | 706.536 | 137.1281 | 109.7 | 0.23 |
| pHILIC | 706.536 | 101.4058 | 109 | 0.23 |
| pHILIC | 706.536 | 112.6431 | 109 | 0.23 |
| pHILIC | 706.536 | 119.1509 | 107.1 | 0.23 |
| pHILIC | 706.536 | 104.3907 | 107 | 0.23 |
| pHILIC | 706.536 | 91.3503 | 104.4 | 0.22 |
| pHILIC | 706.536 | 384.4496 | 80.4 | 0.17 |
| pHILIC | 706.536 | 229.9219 | 80.2 | 0.17 |
| pHILIC | 706.536 | 516.8276 | 78.3 | 0.17 |
| pHILIC | 706.536 | 558.3018 | 78.2 | 0.17 |
| pHILIC | 706.536 | 316.0818 | 77.9 | 0.16 |
| pHILIC | 706.536 | 74.3471 | 77.8 | 0.16 |
| pHILIC | 706.536 | 122.7179 | 76.7 | 0.16 |
| pHILIC | 706.536 | 85.0579 | 76.3 | 0.16 |
| pHILIC | 706.536 | 593.7695 | 75.3 | 0.16 |
| pHILIC | 706.536 | 100.2592 | 75.1 | 0.16 |
| pHILIC | 706.536 | 246.9966 | 74.9 | 0.16 |
| pHILIC | 706.536 | 451.6272 | 74.9 | 0.16 |
| pHILIC | 706.536 | 93.731 | 74.8 | 0.16 |
| pHILIC | 706.536 | 104.3793 | 74.8 | 0.16 |
| pHILIC | 706.536 | 160.7306 | 74.5 | 0.16 |

| | | | | |
|---|---|---|---|---|
| pHILIC | 706.536 | 349.8427 | 74 | 0.16 |
| pHILIC | 706.536 | 92.299 | 73.7 | 0.16 |
| pHILIC | 706.536 | 93.941 | 73.5 | 0.16 |
| pHILIC | 706.536 | 490.3346 | 73.1 | 0.15 |
| pHILIC | 706.536 | 269.0132 | 73 | 0.15 |
| pHILIC | 706.536 | 614.72 | 73 | 0.15 |
| pHILIC | 706.536 | 168.3938 | 72.9 | 0.15 |
| pHILIC | 706.536 | 511.4719 | 72.9 | 0.15 |
| pHILIC | 706.536 | 627.7296 | 72.9 | 0.15 |
| pHILIC | 706.536 | 99.7346 | 72.8 | 0.15 |
| pHILIC | 706.536 | 419.4638 | 72.7 | 0.15 |
| pHILIC | 706.536 | 549.726 | 72.6 | 0.15 |
| pHILIC | 706.536 | 83.085 | 72.5 | 0.15 |
| pHILIC | 706.536 | 603.0279 | 72.5 | 0.15 |
| pHILIC | 706.536 | 409.0898 | 72.5 | 0.15 |
| pHILIC | 706.536 | 522.4031 | 72.2 | 0.15 |
| pHILIC | 706.536 | 137.8705 | 72.1 | 0.15 |
| pHILIC | 706.536 | 184.1248 | 72.1 | 0.15 |
| pHILIC | 706.536 | 93.7778 | 72.1 | 0.15 |
| pHILIC | 706.536 | 105.6327 | 71.9 | 0.15 |
| pHILIC | 706.536 | 79.0688 | 71.9 | 0.15 |
| pHILIC | 706.536 | 173.7396 | 71.9 | 0.15 |
| pHILIC | 706.536 | 90.9398 | 71.8 | 0.15 |
| pHILIC | 706.536 | 152.57 | 71.6 | 0.15 |
| pHILIC | 706.536 | 148.8521 | 71.3 | 0.15 |
| pHILIC | 706.536 | 82.9338 | 71.3 | 0.15 |
| pHILIC | 706.536 | 87.4568 | 71.2 | 0.15 |
| pHILIC | 706.536 | 577.2224 | 71 | 0.15 |
| pHILIC | 706.536 | 76.4586 | 70.9 | 0.15 |
| pHILIC | 706.536 | 168.1557 | 70.8 | 0.15 |
| pHILIC | 706.536 | 388.725 | 70.7 | 0.15 |
| pHILIC | 706.536 | 83.9575 | 70.7 | 0.15 |
| pHILIC | 706.536 | 400.0394 | 70.5 | 0.15 |
| pHILIC | 706.536 | 377.548 | 70.5 | 0.15 |
| pHILIC | 706.536 | 424.8167 | 70.4 | 0.15 |
| pHILIC | 706.536 | 221.2404 | 70.4 | 0.15 |
| pHILIC | 706.536 | 123.4177 | 70.4 | 0.15 |
| pHILIC | 706.536 | 92.9454 | 70.3 | 0.15 |
| pHILIC | 706.536 | 315.5227 | 70.2 | 0.15 |
| pHILIC | 706.536 | 93.3687 | 70.2 | 0.15 |
| pHILIC | 706.536 | 81.2456 | 70.1 | 0.15 |
| pHILIC | 706.536 | 451.8916 | 70.1 | 0.15 |

| | | | | |
|---|---|---|---|---|
| pHILIC | 706.536 | 139.076 | 70 | 0.15 |
| pHILIC | 706.536 | 426.6825 | 70 | 0.15 |
| pHILIC | 706.536 | 324.0952 | 69.7 | 0.15 |
| pHILIC | 706.536 | 104.8801 | 69.7 | 0.15 |
| pHILIC | 706.536 | 93.8686 | 69.6 | 0.15 |
| pHILIC | 706.536 | 175.4415 | 69.6 | 0.15 |
| pHILIC | 706.536 | 606.8403 | 69.6 | 0.15 |
| pHILIC | 706.536 | 186.0783 | 69.6 | 0.15 |
| pHILIC | 706.536 | 98.9544 | 69.4 | 0.15 |
| pHILIC | 706.536 | 427.2772 | 69.3 | 0.15 |
| pHILIC | 706.536 | 201.8546 | 69.3 | 0.15 |
| pHILIC | 706.536 | 173.1598 | 69.3 | 0.15 |
| pHILIC | 706.536 | 105.1331 | 69.3 | 0.15 |
| pHILIC | 706.536 | 493.7554 | 69.2 | 0.15 |
| pHILIC | 706.536 | 630.846 | 69.2 | 0.15 |
| pHILIC | 706.536 | 74.2717 | 69.1 | 0.15 |
| pHILIC | 706.536 | 93.701 | 69 | 0.15 |
| pHILIC | 706.536 | 180.0386 | 68.9 | 0.15 |
| pHILIC | 706.536 | 117.2543 | 68.9 | 0.15 |
| pHILIC | 706.536 | 103.8319 | 68.9 | 0.15 |
| pHILIC | 706.536 | 118.8057 | 68.8 | 0.15 |
| pHILIC | 706.536 | 198.2078 | 68.8 | 0.15 |
| pHILIC | 706.536 | 394.6955 | 68.8 | 0.15 |
| pHILIC | 706.536 | 199.9192 | 68.8 | 0.15 |
| pHILIC | 706.536 | 287.3814 | 68.6 | 0.14 |
| pHILIC | 706.536 | 131.4387 | 68.5 | 0.14 |
| pHILIC | 706.536 | 81.9028 | 68.4 | 0.14 |
| pHILIC | 706.536 | 140.5502 | 68.3 | 0.14 |
| pHILIC | 706.536 | 334.979 | 68.1 | 0.14 |
| pHILIC | 706.536 | 390.2082 | 68.1 | 0.14 |
| pHILIC | 706.536 | 330.8877 | 68.1 | 0.14 |
| pHILIC | 706.536 | 77.1521 | 68 | 0.14 |
| pHILIC | 706.536 | 427.9059 | 68 | 0.14 |
| pHILIC | 706.536 | 424.4224 | 68 | 0.14 |
| pHILIC | 706.536 | 76.2003 | 67.9 | 0.14 |
| pHILIC | 706.536 | 134.6136 | 67.9 | 0.14 |
| pHILIC | 706.536 | 407.8086 | 67.9 | 0.14 |
| pHILIC | 706.536 | 174.2699 | 67.8 | 0.14 |
| pHILIC | 706.536 | 139.5905 | 67.8 | 0.14 |
| pHILIC | 706.536 | 710.9937 | 67.8 | 0.14 |
| pHILIC | 706.536 | 83.423 | 67.8 | 0.14 |
| pHILIC | 706.536 | 368.8913 | 67.7 | 0.14 |

| | | | | |
|---|---|---|---|---|
| pHILIC | 706.536 | 84.193 | 67.7 | 0.14 |
| pHILIC | 706.536 | 112.8792 | 67.5 | 0.14 |
| pHILIC | 706.536 | 163.7445 | 67.3 | 0.14 |
| pHILIC | 706.536 | 199.5829 | 67.2 | 0.14 |
| pHILIC | 706.536 | 84.3475 | 67.2 | 0.14 |
| pHILIC | 706.536 | 675.7796 | 67.2 | 0.14 |
| pHILIC | 706.536 | 369.3881 | 67.2 | 0.14 |
| pHILIC | 706.536 | 478.0041 | 67.2 | 0.14 |
| pHILIC | 706.536 | 374.9988 | 67.1 | 0.14 |
| pHILIC | 706.536 | 162.1326 | 67 | 0.14 |
| pHILIC | 706.536 | 683.0455 | 66.9 | 0.14 |
| pHILIC | 706.536 | 299.3341 | 66.9 | 0.14 |
| pHILIC | 706.536 | 91.6472 | 66.9 | 0.14 |
| pHILIC | 706.536 | 618.5893 | 66.8 | 0.14 |
| pHILIC | 706.536 | 476.6546 | 66.7 | 0.14 |
| pHILIC | 706.536 | 301.6658 | 66.7 | 0.14 |
| pHILIC | 706.536 | 75.9534 | 66.7 | 0.14 |
| pHILIC | 706.536 | 80.6082 | 66.7 | 0.14 |
| pHILIC | 706.536 | 166.3383 | 66.6 | 0.14 |
| pHILIC | 706.536 | 366.2706 | 66.5 | 0.14 |
| pHILIC | 706.536 | 116.4486 | 66.5 | 0.14 |
| pHILIC | 706.536 | 551.5615 | 66.5 | 0.14 |
| pHILIC | 706.536 | 122.0804 | 66.4 | 0.14 |
| pHILIC | 706.536 | 507.3201 | 66.4 | 0.14 |
| pHILIC | 706.536 | 214.1326 | 66.4 | 0.14 |
| pHILIC | 706.536 | 681.3629 | 66.3 | 0.14 |
| pHILIC | 706.536 | 590.8962 | 66.2 | 0.14 |
| pHILIC | 706.536 | 256.2773 | 66.1 | 0.14 |
| pHILIC | 706.536 | 123.0273 | 66 | 0.14 |
| pHILIC | 706.536 | 109.2755 | 66 | 0.14 |
| pHILIC | 706.536 | 123.4741 | 65.9 | 0.14 |
| pHILIC | 706.536 | 183.7275 | 65.9 | 0.14 |
| pHILIC | 706.536 | 690.0847 | 65.9 | 0.14 |
| pHILIC | 706.536 | 310.1534 | 65.9 | 0.14 |
| pHILIC | 706.536 | 306.1326 | 65.8 | 0.14 |
| pHILIC | 706.536 | 151.5349 | 65.8 | 0.14 |
| pHILIC | 706.536 | 93.9704 | 65.8 | 0.14 |
| pHILIC | 706.536 | 329.417 | 65.7 | 0.14 |
| pHILIC | 706.536 | 159.1926 | 65.6 | 0.14 |
| pHILIC | 706.536 | 399.8214 | 65.6 | 0.14 |
| pHILIC | 706.536 | 667.6289 | 65.5 | 0.14 |
| pHILIC | 706.536 | 288.7285 | 65.5 | 0.14 |

| | | | | |
|---|---|---|---|---|
| pHILIC | 706.536 | 105.5578 | 65.5 | 0.14 |
| pHILIC | 706.536 | 123.9419 | 65.5 | 0.14 |
| pHILIC | 706.536 | 354.3354 | 65.4 | 0.14 |
| pHILIC | 706.536 | 154.0949 | 65.4 | 0.14 |
| pHILIC | 706.536 | 93.6461 | 65.2 | 0.14 |
| pHILIC | 706.536 | 83.0495 | 65.2 | 0.14 |
| pHILIC | 706.536 | 294.571 | 65.2 | 0.14 |
| pHILIC | 706.536 | 104.1068 | 65.1 | 0.14 |
| pHILIC | 706.536 | 473.6021 | 65.1 | 0.14 |
| pHILIC | 706.536 | 620.6771 | 65.1 | 0.14 |
| pHILIC | 706.536 | 78.7581 | 65.1 | 0.14 |
| pHILIC | 706.536 | 283.6993 | 65.1 | 0.14 |
| pHILIC | 706.536 | 148.7241 | 65.1 | 0.14 |
| pHILIC | 706.536 | 691.7502 | 65.1 | 0.14 |
| pHILIC | 706.536 | 398.7751 | 65.1 | 0.14 |
| pHILIC | 706.536 | 114.7582 | 65.1 | 0.14 |
| pHILIC | 706.536 | 171.3608 | 65.1 | 0.14 |
| pHILIC | 706.536 | 97.9602 | 65 | 0.14 |
| pHILIC | 706.536 | 175.638 | 65 | 0.14 |
| pHILIC | 706.536 | 172.5903 | 65 | 0.14 |
| pHILIC | 706.536 | 702.89 | 65 | 0.14 |
| pHILIC | 706.536 | 459.1883 | 64.9 | 0.14 |
| pHILIC | 706.536 | 733.0342 | 64.9 | 0.14 |
| pHILIC | 706.536 | 302.6739 | 64.9 | 0.14 |
| pHILIC | 706.536 | 183.5612 | 64.9 | 0.14 |
| pHILIC | 706.536 | 620.0224 | 64.8 | 0.14 |
| pHILIC | 706.536 | 195.7228 | 64.8 | 0.14 |
| pHILIC | 706.536 | 622.3957 | 64.8 | 0.14 |
| pHILIC | 706.536 | 159.4137 | 64.8 | 0.14 |
| pHILIC | 706.536 | 728.9811 | 64.8 | 0.14 |
| pHILIC | 706.536 | 89.3719 | 64.8 | 0.14 |
| pHILIC | 706.536 | 449.0663 | 64.7 | 0.14 |
| pHILIC | 706.536 | 83.7741 | 64.7 | 0.14 |
| pHILIC | 706.536 | 289.1988 | 64.7 | 0.14 |
| pHILIC | 706.536 | 144.4311 | 64.7 | 0.14 |
| pHILIC | 706.536 | 736.3216 | 64.7 | 0.14 |
| pHILIC | 706.536 | 93.8609 | 64.7 | 0.14 |
| pHILIC | 706.536 | 139.9503 | 64.7 | 0.14 |
| pHILIC | 706.536 | 79.0307 | 64.7 | 0.14 |
| pHILIC | 706.536 | 452.3375 | 64.6 | 0.14 |
| pHILIC | 706.536 | 93.6319 | 64.6 | 0.14 |
| pHILIC | 706.536 | 319.8557 | 64.6 | 0.14 |

| | | | | |
|---|---|---|---|---|
| pHILIC | 706.536 | 425.251 | 64.5 | 0.14 |
| pHILIC | 706.536 | 445.6219 | 64.5 | 0.14 |
| pHILIC | 706.536 | 524.0186 | 64.4 | 0.14 |
| pHILIC | 706.536 | 396.1388 | 64.4 | 0.14 |
| pHILIC | 706.536 | 201.188 | 64.4 | 0.14 |
| pHILIC | 706.536 | 636.871 | 64.4 | 0.14 |
| pHILIC | 706.536 | 99.4893 | 64.3 | 0.14 |
| pHILIC | 706.536 | 98.0099 | 64.3 | 0.14 |
| pHILIC | 706.536 | 127.6425 | 64.2 | 0.14 |
| pHILIC | 706.536 | 528.4481 | 64.2 | 0.14 |
| pHILIC | 706.536 | 374.0394 | 64.2 | 0.14 |
| pHILIC | 706.536 | 547.9648 | 64.2 | 0.14 |
| pHILIC | 706.536 | 76.6236 | 64.2 | 0.14 |
| pHILIC | 706.536 | 556.0881 | 64.1 | 0.14 |
| pHILIC | 706.536 | 440.3732 | 64.1 | 0.14 |
| pHILIC | 706.536 | 542.1492 | 64.1 | 0.14 |
| pHILIC | 706.536 | 572.732 | 64.1 | 0.14 |
| pHILIC | 706.536 | 227.2535 | 64 | 0.14 |
| pHILIC | 706.536 | 191.951 | 64 | 0.14 |
| pHILIC | 706.536 | 613.0356 | 64 | 0.14 |
| pHILIC | 706.536 | 93.8904 | 64 | 0.14 |
| pHILIC | 706.536 | 75.7787 | 63.9 | 0.13 |
| pHILIC | 706.536 | 136.6624 | 63.9 | 0.13 |
| pHILIC | 706.536 | 187.4386 | 63.9 | 0.13 |
| pHILIC | 706.536 | 206.8241 | 63.9 | 0.13 |
| pHILIC | 706.536 | 310.3871 | 63.9 | 0.13 |
| pHILIC | 706.536 | 386.4625 | 63.9 | 0.13 |
| pHILIC | 706.536 | 150.5587 | 63.9 | 0.13 |
| pHILIC | 706.536 | 102.4163 | 63.9 | 0.13 |
| pHILIC | 706.536 | 123.5503 | 63.8 | 0.13 |
| pHILIC | 706.536 | 103.5792 | 63.8 | 0.13 |
| pHILIC | 706.536 | 139.3249 | 63.8 | 0.13 |
| pHILIC | 706.536 | 422.8585 | 63.8 | 0.13 |
| pHILIC | 706.536 | 383.5708 | 63.8 | 0.13 |
| pHILIC | 706.536 | 137.6495 | 63.7 | 0.13 |
| pHILIC | 706.536 | 501.7319 | 63.7 | 0.13 |
| pHILIC | 706.536 | 617.72 | 63.7 | 0.13 |
| pHILIC | 706.536 | 384.812 | 63.7 | 0.13 |
| pHILIC | 706.536 | 614.0221 | 63.7 | 0.13 |
| pHILIC | 706.536 | 311.2298 | 63.6 | 0.13 |
| pHILIC | 706.536 | 508.8597 | 63.6 | 0.13 |
| pHILIC | 706.536 | 76.5583 | 63.6 | 0.13 |

| | | | | |
|---|---|---|---|---|
| pHILIC | 706.536 | 222.9767 | 63.6 | 0.13 |
| pHILIC | 706.536 | 438.033 | 63.6 | 0.13 |
| pHILIC | 706.536 | 172.8501 | 63.6 | 0.13 |
| pHILIC | 706.536 | 123.5619 | 63.6 | 0.13 |
| pHILIC | 706.536 | 120.4216 | 63.6 | 0.13 |
| pHILIC | 706.536 | 87.8902 | 63.6 | 0.13 |
| pHILIC | 706.536 | 319.6759 | 63.5 | 0.13 |
| pHILIC | 706.536 | 124.7923 | 63.4 | 0.13 |
| pHILIC | 706.536 | 195.2539 | 63.4 | 0.13 |
| pHILIC | 706.536 | 75.4897 | 63.4 | 0.13 |
| pHILIC | 706.536 | 727.9717 | 63.4 | 0.13 |
| pHILIC | 706.536 | 353.8924 | 63.3 | 0.13 |
| pHILIC | 706.536 | 129.9765 | 63.3 | 0.13 |
| pHILIC | 706.536 | 394.909 | 63.3 | 0.13 |
| pHILIC | 706.536 | 163.6902 | 63.3 | 0.13 |
| pHILIC | 706.536 | 175.5264 | 63.3 | 0.13 |
| pHILIC | 706.536 | 687.0939 | 63.3 | 0.13 |
| pHILIC | 706.536 | 191.1 | 63.2 | 0.13 |
| pHILIC | 706.536 | 391.7569 | 63.2 | 0.13 |
| pHILIC | 706.536 | 335.9822 | 63.2 | 0.13 |
| pHILIC | 706.536 | 391.9808 | 63.2 | 0.13 |
| pHILIC | 706.536 | 230.9449 | 63.1 | 0.13 |
| pHILIC | 706.536 | 685.8699 | 63.1 | 0.13 |
| pHILIC | 706.536 | 198.9524 | 63.1 | 0.13 |
| pHILIC | 706.536 | 243.9208 | 63.1 | 0.13 |
| pHILIC | 706.536 | 147.4634 | 63.1 | 0.13 |
| pHILIC | 706.536 | 201.7696 | 63.1 | 0.13 |
| pHILIC | 706.536 | 224.4546 | 63.1 | 0.13 |
| pHILIC | 706.536 | 79.6224 | 63 | 0.13 |
| pHILIC | 706.536 | 382.1018 | 63 | 0.13 |
| pHILIC | 706.536 | 124.6023 | 63 | 0.13 |
| pHILIC | 706.536 | 376.6953 | 63 | 0.13 |
| pHILIC | 706.536 | 449.959 | 63 | 0.13 |
| pHILIC | 706.536 | 110.6549 | 63 | 0.13 |
| pHILIC | 706.536 | 94.0765 | 62.9 | 0.13 |
| pHILIC | 706.536 | 651.7056 | 62.9 | 0.13 |
| pHILIC | 706.536 | 85.1015 | 62.9 | 0.13 |
| pHILIC | 706.536 | 499.2198 | 62.9 | 0.13 |
| pHILIC | 706.536 | 519.9506 | 62.9 | 0.13 |
| pHILIC | 706.536 | 277.6965 | 62.9 | 0.13 |
| pHILIC | 706.536 | 76.4648 | 62.8 | 0.13 |
| pHILIC | 706.536 | 383.1737 | 62.8 | 0.13 |

| | | | | |
|---|---|---|---|---|
| pHILIC | 706.536 | 102.9268 | 62.8 | 0.13 |
| pHILIC | 706.536 | 467.5088 | 62.8 | 0.13 |
| pHILIC | 706.536 | 90.9501 | 62.8 | 0.13 |
| pHILIC | 706.536 | 81.3432 | 62.8 | 0.13 |
| pHILIC | 706.536 | 535.4591 | 62.8 | 0.13 |
| pHILIC | 706.536 | 207.7752 | 62.7 | 0.13 |
| pHILIC | 706.536 | 313.5357 | 62.6 | 0.13 |
| pHILIC | 706.536 | 84.9734 | 62.6 | 0.13 |
| pHILIC | 706.536 | 91.1293 | 62.6 | 0.13 |
| pHILIC | 706.536 | 81.5596 | 62.6 | 0.13 |
| pHILIC | 706.536 | 232.112 | 62.6 | 0.13 |
| pHILIC | 706.536 | 636.3199 | 62.5 | 0.13 |
| pHILIC | 706.536 | 135.9895 | 62.5 | 0.13 |
| pHILIC | 706.536 | 281.8652 | 62.5 | 0.13 |
| pHILIC | 706.536 | 107.635 | 62.5 | 0.13 |
| pHILIC | 706.536 | 348.0431 | 62.5 | 0.13 |
| pHILIC | 706.536 | 135.1872 | 62.4 | 0.13 |
| pHILIC | 706.536 | 321.1188 | 62.4 | 0.13 |
| pHILIC | 706.536 | 81.5207 | 62.4 | 0.13 |
| pHILIC | 706.536 | 208.7759 | 62.4 | 0.13 |
| pHILIC | 706.536 | 377.509 | 62.3 | 0.13 |
| pHILIC | 706.536 | 114.5289 | 62.3 | 0.13 |
| pHILIC | 706.536 | 94.3763 | 62.3 | 0.13 |
| pHILIC | 706.536 | 82.4895 | 62.3 | 0.13 |
| pHILIC | 706.536 | 142.2917 | 62.3 | 0.13 |
| pHILIC | 706.536 | 106.7002 | 62.3 | 0.13 |
| pHILIC | 706.536 | 662.872 | 62.3 | 0.13 |
| pHILIC | 706.536 | 112.4348 | 62.3 | 0.13 |
| pHILIC | 706.536 | 85.5126 | 62.3 | 0.13 |
| pHILIC | 706.536 | 159.6303 | 62.3 | 0.13 |
| pHILIC | 706.536 | 486.5285 | 62.2 | 0.13 |
| pHILIC | 706.536 | 381.6842 | 62.2 | 0.13 |
| pHILIC | 706.536 | 124.3453 | 62.2 | 0.13 |
| pHILIC | 706.536 | 248.8693 | 62.1 | 0.13 |
| pHILIC | 706.536 | 567.1298 | 62.1 | 0.13 |
| pHILIC | 706.536 | 141.7039 | 62.1 | 0.13 |
| pHILIC | 706.536 | 254.9656 | 62.1 | 0.13 |
| pHILIC | 706.536 | 652.0467 | 62.1 | 0.13 |
| pHILIC | 706.536 | 309.0922 | 62.1 | 0.13 |
| pHILIC | 706.536 | 103.9625 | 62 | 0.13 |
| pHILIC | 706.536 | 432.9662 | 62 | 0.13 |
| pHILIC | 706.536 | 86.4451 | 62 | 0.13 |

| | | | | |
|---|---|---|---|---|
| pHILIC | 706.536 | 241.7169 | 62 | 0.13 |
| pHILIC | 706.536 | 116.8068 | 62 | 0.13 |
| pHILIC | 706.536 | 126.9765 | 62 | 0.13 |
| pHILIC | 706.536 | 164.2121 | 61.9 | 0.13 |
| pHILIC | 706.536 | 188.293 | 61.9 | 0.13 |
| pHILIC | 706.536 | 365.327 | 61.9 | 0.13 |
| pHILIC | 706.536 | 93.7369 | 61.9 | 0.13 |
| pHILIC | 706.536 | 162.3561 | 61.9 | 0.13 |
| pHILIC | 706.536 | 200.5188 | 61.9 | 0.13 |
| pHILIC | 706.536 | 690.3809 | 61.9 | 0.13 |
| pHILIC | 706.536 | 492.4733 | 61.9 | 0.13 |
| pHILIC | 706.536 | 610.0505 | 61.9 | 0.13 |
| pHILIC | 706.536 | 108.6924 | 61.9 | 0.13 |
| pHILIC | 706.536 | 520.9772 | 61.8 | 0.13 |
| pHILIC | 706.536 | 182.3345 | 61.8 | 0.13 |
| pHILIC | 706.536 | 145.4309 | 61.8 | 0.13 |
| pHILIC | 706.536 | 88.1602 | 61.8 | 0.13 |
| pHILIC | 706.536 | 156.3523 | 61.8 | 0.13 |
| pHILIC | 706.536 | 238.1755 | 61.8 | 0.13 |
| pHILIC | 706.536 | 107.9927 | 61.8 | 0.13 |
| pHILIC | 706.536 | 76.0184 | 61.8 | 0.13 |
| pHILIC | 706.536 | 463.5889 | 61.8 | 0.13 |
| pHILIC | 706.536 | 586.686 | 61.7 | 0.13 |
| pHILIC | 706.536 | 489.8202 | 61.7 | 0.13 |
| pHILIC | 706.536 | 251.0139 | 61.7 | 0.13 |
| pHILIC | 706.536 | 562.056 | 61.7 | 0.13 |
| pHILIC | 706.536 | 710.3489 | 61.7 | 0.13 |
| pHILIC | 706.536 | 205.6058 | 61.7 | 0.13 |
| pHILIC | 706.536 | 634.8052 | 61.6 | 0.13 |
| pHILIC | 706.536 | 225.5561 | 61.6 | 0.13 |
| pHILIC | 706.536 | 78.0551 | 61.6 | 0.13 |
| pHILIC | 706.536 | 324.2392 | 61.6 | 0.13 |
| pHILIC | 706.536 | 222.4562 | 61.6 | 0.13 |
| pHILIC | 706.536 | 76.6189 | 61.6 | 0.13 |
| pHILIC | 706.536 | 112.5799 | 61.6 | 0.13 |
| pHILIC | 706.536 | 189.8294 | 61.6 | 0.13 |
| pHILIC | 706.536 | 128.0144 | 61.6 | 0.13 |
| pHILIC | 706.536 | 294.2399 | 61.5 | 0.13 |
| pHILIC | 706.536 | 93.546 | 61.5 | 0.13 |
| pHILIC | 706.536 | 245.453 | 61.5 | 0.13 |
| pHILIC | 706.536 | 187.3661 | 61.4 | 0.13 |
| pHILIC | 706.536 | 187.3796 | 61.3 | 0.13 |

| | | | | |
|---|---|---|---|---|
| pHILIC | 706.536 | 370.9201 | 61.3 | 0.13 |
| pHILIC | 706.536 | 363.7617 | 61.3 | 0.13 |
| pHILIC | 706.536 | 199.8255 | 61.3 | 0.13 |
| pHILIC | 706.536 | 161.713 | 61.2 | 0.13 |
| pHILIC | 706.536 | 393.428 | 61.2 | 0.13 |
| pHILIC | 706.536 | 396.2301 | 61.2 | 0.13 |
| pHILIC | 706.536 | 171.1985 | 61.2 | 0.13 |
| pHILIC | 706.536 | 326.5977 | 61.2 | 0.13 |
| pHILIC | 706.536 | 174.8061 | 61.2 | 0.13 |
| pHILIC | 706.536 | 260.779 | 61.2 | 0.13 |
| pHILIC | 706.536 | 93.801 | 61.1 | 0.13 |
| pHILIC | 706.536 | 510.5996 | 61.1 | 0.13 |
| pHILIC | 706.536 | 303.7257 | 61.1 | 0.13 |
| pHILIC | 706.536 | 118.5927 | 61.1 | 0.13 |
| pHILIC | 706.536 | 293.7759 | 61.1 | 0.13 |
| pHILIC | 706.536 | 163.7229 | 61 | 0.13 |
| pHILIC | 706.536 | 332.6501 | 61 | 0.13 |
| pHILIC | 706.536 | 188.415 | 61 | 0.13 |
| pHILIC | 706.536 | 273.1928 | 61 | 0.13 |
| pHILIC | 706.536 | 75.8139 | 61 | 0.13 |
| pHILIC | 706.536 | 106.0357 | 61 | 0.13 |
| pHILIC | 706.536 | 107.024 | 60.9 | 0.13 |
| pHILIC | 706.536 | 730.7607 | 60.9 | 0.13 |
| pHILIC | 706.536 | 506.7415 | 60.9 | 0.13 |
| pHILIC | 706.536 | 91.6444 | 60.9 | 0.13 |
| pHILIC | 706.536 | 106.4414 | 60.9 | 0.13 |
| pHILIC | 706.536 | 75.8042 | 60.8 | 0.13 |
| pHILIC | 706.536 | 397.8279 | 60.8 | 0.13 |
| pHILIC | 706.536 | 100.4954 | 60.8 | 0.13 |
| pHILIC | 706.536 | 118.0226 | 60.8 | 0.13 |
| pHILIC | 706.536 | 93.6257 | 60.8 | 0.13 |
| pHILIC | 706.536 | 411.8685 | 60.8 | 0.13 |
| pHILIC | 706.536 | 382.9309 | 60.8 | 0.13 |
| pHILIC | 706.536 | 682.5757 | 60.8 | 0.13 |
| pHILIC | 706.536 | 82.9247 | 60.8 | 0.13 |
| pHILIC | 706.536 | 269.54 | 60.8 | 0.13 |
| pHILIC | 706.536 | 140.5301 | 60.7 | 0.13 |
| pHILIC | 706.536 | 210.4648 | 60.7 | 0.13 |
| pHILIC | 706.536 | 75.6227 | 60.7 | 0.13 |
| pHILIC | 706.536 | 200.9595 | 60.7 | 0.13 |
| pHILIC | 706.536 | 675.1697 | 60.7 | 0.13 |
| pHILIC | 706.536 | 119.0326 | 60.7 | 0.13 |

| | | | | |
|---|---|---|---|---|
| pHILIC | 706.536 | 80.06 | 60.7 | 0.13 |
| pHILIC | 706.536 | 606.0089 | 60.7 | 0.13 |
| pHILIC | 706.536 | 207.0975 | 60.6 | 0.13 |
| pHILIC | 706.536 | 90.1958 | 60.6 | 0.13 |
| pHILIC | 706.536 | 83.1475 | 60.6 | 0.13 |
| pHILIC | 706.536 | 225.0211 | 60.6 | 0.13 |
| pHILIC | 706.536 | 264.9384 | 60.6 | 0.13 |
| pHILIC | 706.536 | 83.2838 | 60.6 | 0.13 |
| pHILIC | 706.536 | 487.8716 | 60.6 | 0.13 |
| pHILIC | 706.536 | 81.5905 | 60.6 | 0.13 |
| pHILIC | 706.536 | 76.1408 | 60.6 | 0.13 |
| pHILIC | 706.536 | 87.7659 | 60.5 | 0.13 |
| pHILIC | 706.536 | 603.9126 | 60.5 | 0.13 |
| pHILIC | 706.536 | 304.3749 | 60.5 | 0.13 |
| pHILIC | 706.536 | 307.1539 | 60.4 | 0.13 |
| pHILIC | 706.536 | 561.6239 | 60.4 | 0.13 |
| pHILIC | 706.536 | 718.6946 | 60.4 | 0.13 |
| pHILIC | 706.536 | 724.4934 | 60.4 | 0.13 |
| pHILIC | 706.536 | 356.3242 | 60.4 | 0.13 |
| pHILIC | 706.536 | 210.561 | 60.4 | 0.13 |
| pHILIC | 706.536 | 110.3903 | 60.3 | 0.13 |
| pHILIC | 706.536 | 126.7275 | 60.3 | 0.13 |
| pHILIC | 706.536 | 182.7029 | 60.3 | 0.13 |
| pHILIC | 706.536 | 321.3046 | 60.3 | 0.13 |
| pHILIC | 706.536 | 78.3664 | 60.2 | 0.13 |
| pHILIC | 706.536 | 136.8649 | 60.2 | 0.13 |
| pHILIC | 706.536 | 89.2633 | 60.2 | 0.13 |
| pHILIC | 706.536 | 169.0937 | 60.2 | 0.13 |
| pHILIC | 706.536 | 380.2848 | 60.2 | 0.13 |
| pHILIC | 706.536 | 92.9688 | 60.2 | 0.13 |
| pHILIC | 706.536 | 186.4341 | 60.2 | 0.13 |
| pHILIC | 706.536 | 124.7328 | 60.2 | 0.13 |
| pHILIC | 706.536 | 378.3625 | 60.2 | 0.13 |
| pHILIC | 706.536 | 734.2948 | 60.1 | 0.13 |
| pHILIC | 706.536 | 86.5662 | 60.1 | 0.13 |
| pHILIC | 706.536 | 661.5655 | 60.1 | 0.13 |
| pHILIC | 706.536 | 268.9663 | 60.1 | 0.13 |
| pHILIC | 706.536 | 148.7003 | 60 | 0.13 |
| pHILIC | 706.536 | 243.2066 | 60 | 0.13 |
| pHILIC | 706.536 | 480.0007 | 60 | 0.13 |
| pHILIC | 706.536 | 231.1471 | 60 | 0.13 |
| pHILIC | 706.536 | 387.9094 | 60 | 0.13 |

| | | | | |
|---|---|---|---|---|
| pHILIC | 706.536 | 150.9733 | 60 | 0.13 |
| pHILIC | 706.536 | 88.4146 | 60 | 0.13 |
| pHILIC | 706.536 | 269.2228 | 60 | 0.13 |
| pHILIC | 706.536 | 117.0917 | 60 | 0.13 |
| pHILIC | 706.536 | 116.8912 | 60 | 0.13 |
| pHILIC | 706.536 | 90.7537 | 60 | 0.13 |
| pHILIC | 706.536 | 668.8707 | 60 | 0.13 |
| pHILIC | 706.536 | 80.5525 | 59.9 | 0.13 |
| pHILIC | 706.536 | 383.3925 | 59.9 | 0.13 |
| pHILIC | 706.536 | 118.3777 | 59.9 | 0.13 |
| pHILIC | 706.536 | 323.7174 | 59.9 | 0.13 |
| pHILIC | 706.536 | 645.5347 | 59.9 | 0.13 |
| pHILIC | 706.536 | 393.84 | 59.9 | 0.13 |
| pHILIC | 706.536 | 93.9161 | 59.9 | 0.13 |
| pHILIC | 706.536 | 391.6229 | 59.9 | 0.13 |
| pHILIC | 706.536 | 109.2956 | 59.9 | 0.13 |
| pHILIC | 706.536 | 91.436 | 59.9 | 0.13 |
| pHILIC | 706.536 | 188.7886 | 59.9 | 0.13 |
| pHILIC | 706.536 | 144.7586 | 59.9 | 0.13 |
| pHILIC | 706.536 | 555.444 | 59.8 | 0.13 |
| pHILIC | 706.536 | 657.5192 | 59.8 | 0.13 |
| pHILIC | 706.536 | 116.721 | 59.8 | 0.13 |
| pHILIC | 706.536 | 139.5463 | 59.8 | 0.13 |
| pHILIC | 706.536 | 119.3848 | 59.8 | 0.13 |
| pHILIC | 706.536 | 112.4865 | 59.8 | 0.13 |
| pHILIC | 706.536 | 89.0309 | 59.8 | 0.13 |
| pHILIC | 706.536 | 107.8815 | 59.7 | 0.13 |
| pHILIC | 706.536 | 97.6088 | 59.7 | 0.13 |
| pHILIC | 706.536 | 185.715 | 59.7 | 0.13 |
| pHILIC | 706.536 | 227.2104 | 59.7 | 0.13 |
| pHILIC | 706.536 | 125.5227 | 59.6 | 0.13 |
| pHILIC | 706.536 | 354.8166 | 59.6 | 0.13 |
| pHILIC | 706.536 | 84.9299 | 59.6 | 0.13 |
| pHILIC | 706.536 | 166.2206 | 59.6 | 0.13 |
| pHILIC | 706.536 | 156.5592 | 59.6 | 0.13 |
| pHILIC | 706.536 | 82.3184 | 59.6 | 0.13 |
| pHILIC | 706.536 | 150.608 | 59.5 | 0.13 |
| pHILIC | 706.536 | 569.9338 | 59.5 | 0.13 |
| pHILIC | 706.536 | 572.4066 | 59.5 | 0.13 |
| pHILIC | 706.536 | 77.8752 | 59.5 | 0.13 |
| pHILIC | 706.536 | 156.3011 | 59.5 | 0.13 |
| pHILIC | 706.536 | 401.3106 | 59.4 | 0.13 |

| | | | | |
|---|---|---|---|---|
| pHILIC | 706.536 | 79.4529 | 59.4 | 0.13 |
| pHILIC | 706.536 | 390.1096 | 59.4 | 0.13 |
| pHILIC | 706.536 | 80.8465 | 59.4 | 0.13 |
| pHILIC | 706.536 | 190.9024 | 59.4 | 0.13 |
| pHILIC | 706.536 | 200.6643 | 59.3 | 0.13 |
| pHILIC | 706.536 | 98.6696 | 59.3 | 0.13 |
| pHILIC | 706.536 | 109.4013 | 59.3 | 0.13 |
| pHILIC | 706.536 | 249.7933 | 59.3 | 0.13 |
| pHILIC | 706.536 | 233.4653 | 59.3 | 0.13 |
| pHILIC | 706.536 | 650.6055 | 59.3 | 0.13 |
| pHILIC | 706.536 | 250.3475 | 59.3 | 0.13 |
| pHILIC | 706.536 | 94.8458 | 59.3 | 0.13 |
| pHILIC | 706.536 | 124.4023 | 59.3 | 0.13 |
| pHILIC | 706.536 | 74.4628 | 59.2 | 0.13 |
| pHILIC | 706.536 | 88.9595 | 59.2 | 0.13 |
| pHILIC | 706.536 | 130.8516 | 59.2 | 0.13 |
| pHILIC | 706.536 | 232.0874 | 59.2 | 0.13 |
| pHILIC | 706.536 | 133.0687 | 59.2 | 0.13 |
| pHILIC | 706.536 | 519.2845 | 59.2 | 0.13 |
| pHILIC | 706.536 | 305.595 | 59.2 | 0.13 |
| pHILIC | 706.536 | 107.4522 | 59.2 | 0.13 |
| pHILIC | 706.536 | 267.14 | 59.2 | 0.13 |
| pHILIC | 706.536 | 156.5074 | 59.2 | 0.12 |
| pHILIC | 706.536 | 387.8324 | 59.2 | 0.12 |
| pHILIC | 706.536 | 296.7372 | 59.2 | 0.12 |
| pHILIC | 706.536 | 204.8394 | 59.2 | 0.12 |
| pHILIC | 706.536 | 631.1484 | 59.2 | 0.12 |
| pHILIC | 706.536 | 387.17 | 59.1 | 0.12 |
| pHILIC | 706.536 | 351.7766 | 59.1 | 0.12 |
| pHILIC | 706.536 | 167.51 | 59.1 | 0.12 |
| pHILIC | 706.536 | 109.7974 | 59.1 | 0.12 |
| pHILIC | 706.536 | 654.6069 | 59.1 | 0.12 |
| pHILIC | 706.536 | 113.219 | 59 | 0.12 |
| pHILIC | 706.536 | 321.753 | 59 | 0.12 |
| pHILIC | 706.536 | 168.1798 | 59 | 0.12 |
| pHILIC | 706.536 | 96.9341 | 59 | 0.12 |
| pHILIC | 706.536 | 435.3111 | 59 | 0.12 |
| pHILIC | 706.536 | 89.6073 | 59 | 0.12 |
| pHILIC | 706.536 | 90.4113 | 59 | 0.12 |
| pHILIC | 706.536 | 94.3892 | 59 | 0.12 |
| pHILIC | 706.536 | 170.6963 | 59 | 0.12 |
| pHILIC | 706.536 | 262.7741 | 59 | 0.12 |

| | | | | |
|---|---|---|---|---|
| pHILIC | 706.536 | 99.656 | 59 | 0.12 |
| pHILIC | 706.536 | 154.2498 | 58.9 | 0.12 |
| pHILIC | 706.536 | 301.4506 | 58.9 | 0.12 |
| pHILIC | 706.536 | 374.4425 | 58.9 | 0.12 |
| pHILIC | 706.536 | 246.0005 | 58.9 | 0.12 |
| pHILIC | 706.536 | 158.271 | 58.9 | 0.12 |
| pHILIC | 706.536 | 91.1862 | 58.9 | 0.12 |
| pHILIC | 706.536 | 82.9961 | 58.8 | 0.12 |
| pHILIC | 706.536 | 277.8299 | 58.8 | 0.12 |
| pHILIC | 706.536 | 508.2542 | 58.8 | 0.12 |
| pHILIC | 706.536 | 352.5972 | 58.8 | 0.12 |
| pHILIC | 706.536 | 635.6718 | 58.7 | 0.12 |
| pHILIC | 706.536 | 153.414 | 58.7 | 0.12 |
| pHILIC | 706.536 | 130.1114 | 58.7 | 0.12 |
| pHILIC | 706.536 | 328.4231 | 58.7 | 0.12 |
| pHILIC | 706.536 | 101.3984 | 58.7 | 0.12 |
| pHILIC | 706.536 | 127.7666 | 58.7 | 0.12 |
| pHILIC | 706.536 | 240.3399 | 58.6 | 0.12 |
| pHILIC | 706.536 | 93.6138 | 58.6 | 0.12 |
| pHILIC | 706.536 | 717.0825 | 58.6 | 0.12 |
| pHILIC | 706.536 | 184.5385 | 58.6 | 0.12 |
| pHILIC | 706.536 | 456.3331 | 58.6 | 0.12 |
| pHILIC | 706.536 | 121.7614 | 58.6 | 0.12 |
| pHILIC | 706.536 | 556.9098 | 58.5 | 0.12 |
| pHILIC | 706.536 | 324.9079 | 58.5 | 0.12 |
| pHILIC | 706.536 | 269.3008 | 58.5 | 0.12 |
| pHILIC | 706.536 | 86.3529 | 58.5 | 0.12 |
| pHILIC | 706.536 | 630.5582 | 58.5 | 0.12 |
| pHILIC | 706.536 | 112.9637 | 58.4 | 0.12 |
| pHILIC | 706.536 | 385.5434 | 58.4 | 0.12 |
| pHILIC | 706.536 | 505.1057 | 58.4 | 0.12 |
| pHILIC | 706.536 | 222.9024 | 58.4 | 0.12 |
| pHILIC | 706.536 | 75.2082 | 58.4 | 0.12 |
| pHILIC | 706.536 | 188.4708 | 58.4 | 0.12 |
| pHILIC | 706.536 | 647.3118 | 58.4 | 0.12 |
| pHILIC | 706.536 | 315.2602 | 58.4 | 0.12 |
| pHILIC | 706.536 | 163.2072 | 58.4 | 0.12 |
| pHILIC | 706.536 | 624.6589 | 58.3 | 0.12 |
| pHILIC | 706.536 | 235.38 | 58.3 | 0.12 |
| pHILIC | 706.536 | 446.3897 | 58.3 | 0.12 |
| pHILIC | 706.536 | 87.7838 | 58.3 | 0.12 |
| pHILIC | 706.536 | 255.5885 | 58.3 | 0.12 |

| | | | | |
|---|---|---|---|---|
| pHILIC | 706.536 | 106.6745 | 58.3 | 0.12 |
| pHILIC | 706.536 | 117.9024 | 58.3 | 0.12 |
| pHILIC | 706.536 | 134.8162 | 58.2 | 0.12 |
| pHILIC | 706.536 | 145.5787 | 58.2 | 0.12 |
| pHILIC | 706.536 | 90.6149 | 58.2 | 0.12 |
| pHILIC | 706.536 | 537.906 | 58.2 | 0.12 |
| pHILIC | 706.536 | 146.6577 | 58.2 | 0.12 |
| pHILIC | 706.536 | 105.4655 | 58.1 | 0.12 |
| pHILIC | 706.536 | 644.4643 | 58.1 | 0.12 |
| pHILIC | 706.536 | 110.0992 | 58.1 | 0.12 |
| pHILIC | 706.536 | 333.5737 | 58.1 | 0.12 |
| pHILIC | 706.536 | 89.9778 | 58.1 | 0.12 |
| pHILIC | 706.536 | 283.4198 | 58 | 0.12 |
| pHILIC | 706.536 | 170.1776 | 58 | 0.12 |
| pHILIC | 706.536 | 200.4404 | 58 | 0.12 |
| pHILIC | 706.536 | 75.9775 | 58 | 0.12 |
| pHILIC | 706.536 | 164.4557 | 58 | 0.12 |
| pHILIC | 706.536 | 102.5169 | 58 | 0.12 |
| pHILIC | 706.536 | 392.7815 | 58 | 0.12 |
| pHILIC | 706.536 | 117.6803 | 58 | 0.12 |
| pHILIC | 706.536 | 80.3692 | 58 | 0.12 |
| pHILIC | 706.536 | 82.2415 | 58 | 0.12 |
| pHILIC | 706.536 | 118.4456 | 57.9 | 0.12 |
| pHILIC | 706.536 | 112.9867 | 57.9 | 0.12 |
| pHILIC | 706.536 | 81.0326 | 57.9 | 0.12 |
| pHILIC | 706.536 | 87.2978 | 57.9 | 0.12 |
| pHILIC | 706.536 | 429.6422 | 57.9 | 0.12 |
| pHILIC | 706.536 | 182.6806 | 57.9 | 0.12 |
| pHILIC | 706.536 | 351.643 | 57.9 | 0.12 |
| pHILIC | 706.536 | 276.1351 | 57.9 | 0.12 |
| pHILIC | 706.536 | 738.4819 | 57.9 | 0.12 |
| pHILIC | 706.536 | 448.2933 | 57.9 | 0.12 |
| pHILIC | 706.536 | 116.3535 | 57.9 | 0.12 |
| pHILIC | 706.536 | 151.2333 | 57.9 | 0.12 |
| pHILIC | 706.536 | 274.5443 | 57.9 | 0.12 |
| pHILIC | 706.536 | 199.6207 | 57.9 | 0.12 |
| pHILIC | 706.536 | 153.0904 | 57.9 | 0.12 |
| pHILIC | 706.536 | 278.8487 | 57.8 | 0.12 |
| pHILIC | 706.536 | 433.1904 | 57.8 | 0.12 |
| pHILIC | 706.536 | 111.7218 | 57.8 | 0.12 |
| pHILIC | 706.536 | 83.6892 | 57.8 | 0.12 |
| pHILIC | 706.536 | 198.7402 | 57.8 | 0.12 |

| | | | | |
|---|---|---|---|---|
| pHILIC | 706.536 | 96.0092 | 57.8 | 0.12 |
| pHILIC | 706.536 | 283.1695 | 57.7 | 0.12 |
| pHILIC | 706.536 | 114.5972 | 57.7 | 0.12 |
| pHILIC | 706.536 | 104.3834 | 57.7 | 0.12 |
| pHILIC | 706.536 | 112.7871 | 57.7 | 0.12 |
| pHILIC | 706.536 | 97.9528 | 57.7 | 0.12 |
| pHILIC | 706.536 | 79.812 | 57.7 | 0.12 |
| pHILIC | 706.536 | 310.5747 | 57.7 | 0.12 |
| pHILIC | 706.536 | 190.2472 | 57.7 | 0.12 |
| pHILIC | 706.536 | 90.2024 | 57.7 | 0.12 |
| pHILIC | 706.536 | 274.2497 | 57.7 | 0.12 |
| pHILIC | 706.536 | 559.6862 | 57.6 | 0.12 |
| pHILIC | 706.536 | 137.4141 | 57.6 | 0.12 |
| pHILIC | 706.536 | 98.5927 | 57.6 | 0.12 |
| pHILIC | 706.536 | 98.1719 | 57.6 | 0.12 |
| pHILIC | 706.536 | 110.7879 | 57.6 | 0.12 |
| pHILIC | 706.536 | 94.8258 | 57.6 | 0.12 |
| pHILIC | 706.536 | 79.3056 | 57.6 | 0.12 |
| pHILIC | 706.536 | 224.1283 | 57.6 | 0.12 |
| pHILIC | 706.536 | 84.3252 | 57.5 | 0.12 |
| pHILIC | 706.536 | 395.4296 | 57.5 | 0.12 |
| pHILIC | 706.536 | 75.9947 | 57.5 | 0.12 |
| pHILIC | 706.536 | 340.5895 | 57.5 | 0.12 |
| pHILIC | 706.536 | 91.8935 | 57.5 | 0.12 |
| pHILIC | 706.536 | 98.8737 | 57.5 | 0.12 |
| pHILIC | 706.536 | 200.8968 | 57.5 | 0.12 |
| pHILIC | 706.536 | 180.2308 | 57.5 | 0.12 |
| pHILIC | 706.536 | 83.8848 | 57.4 | 0.12 |
| pHILIC | 706.536 | 243.5786 | 57.4 | 0.12 |
| pHILIC | 706.536 | 133.6233 | 57.4 | 0.12 |
| pHILIC | 706.536 | 93.7138 | 57.4 | 0.12 |
| pHILIC | 706.536 | 236.6043 | 57.4 | 0.12 |
| pHILIC | 706.536 | 171.0048 | 57.4 | 0.12 |
| pHILIC | 706.536 | 336.922 | 57.4 | 0.12 |
| pHILIC | 706.536 | 247.0239 | 57.4 | 0.12 |
| pHILIC | 706.536 | 539.6533 | 57.3 | 0.12 |
| pHILIC | 706.536 | 380.6497 | 57.3 | 0.12 |
| pHILIC | 706.536 | 89.6002 | 57.3 | 0.12 |
| pHILIC | 706.536 | 252.9447 | 57.3 | 0.12 |
| pHILIC | 706.536 | 311.1671 | 57.3 | 0.12 |
| pHILIC | 706.536 | 136.385 | 57.3 | 0.12 |
| pHILIC | 706.536 | 158.8841 | 57.3 | 0.12 |

| | | | | |
|---|---|---|---|---|
| pHILIC | 706.536 | 507.239 | 57.3 | 0.12 |
| pHILIC | 706.536 | 233.057 | 57.3 | 0.12 |
| pHILIC | 706.536 | 138.7209 | 57.3 | 0.12 |
| pHILIC | 706.536 | 286.6921 | 57.3 | 0.12 |
| pHILIC | 706.536 | 493.218 | 57.3 | 0.12 |
| pHILIC | 706.536 | 284.9918 | 57.2 | 0.12 |
| pHILIC | 706.536 | 207.6383 | 57.2 | 0.12 |
| pHILIC | 706.536 | 191.7906 | 57.2 | 0.12 |
| pHILIC | 706.536 | 93.0908 | 57.2 | 0.12 |
| pHILIC | 706.536 | 126.7725 | 57.2 | 0.12 |
| pHILIC | 706.536 | 171.3907 | 57.2 | 0.12 |
| pHILIC | 706.536 | 130.4453 | 57.1 | 0.12 |
| pHILIC | 706.536 | 93.7582 | 57.1 | 0.12 |
| pHILIC | 706.536 | 112.7795 | 57.1 | 0.12 |
| pHILIC | 706.536 | 628.9144 | 57.1 | 0.12 |
| pHILIC | 706.536 | 193.771 | 57.1 | 0.12 |
| pHILIC | 706.536 | 93.1588 | 57.1 | 0.12 |
| pHILIC | 706.536 | 649.5127 | 57.1 | 0.12 |
| pHILIC | 706.536 | 92.481 | 57.1 | 0.12 |
| pHILIC | 706.536 | 446.7307 | 57 | 0.12 |
| pHILIC | 706.536 | 77.7363 | 57 | 0.12 |
| pHILIC | 706.536 | 198.8078 | 57 | 0.12 |
| pHILIC | 706.536 | 123.2327 | 57 | 0.12 |
| pHILIC | 706.536 | 234.7229 | 57 | 0.12 |
| pHILIC | 706.536 | 469.2511 | 57 | 0.12 |
| pHILIC | 706.536 | 99.3179 | 57 | 0.12 |
| pHILIC | 706.536 | 300.1168 | 57 | 0.12 |
| pHILIC | 706.536 | 110.4176 | 57 | 0.12 |
| pHILIC | 706.536 | 116.7292 | 56.9 | 0.12 |
| pHILIC | 706.536 | 217.8147 | 56.9 | 0.12 |
| pHILIC | 706.536 | 246.6158 | 56.9 | 0.12 |
| pHILIC | 706.536 | 175.8663 | 56.9 | 0.12 |
| pHILIC | 706.536 | 76.4911 | 56.9 | 0.12 |
| pHILIC | 706.536 | 711.9882 | 56.9 | 0.12 |
| pHILIC | 706.536 | 210.3305 | 56.9 | 0.12 |
| pHILIC | 706.536 | 83.3828 | 56.9 | 0.12 |
| pHILIC | 706.536 | 155.536 | 56.9 | 0.12 |
| pHILIC | 706.536 | 152.8424 | 56.9 | 0.12 |
| pHILIC | 706.536 | 168.653 | 56.9 | 0.12 |
| pHILIC | 706.536 | 347.6633 | 56.8 | 0.12 |
| pHILIC | 706.536 | 241.0966 | 56.8 | 0.12 |
| pHILIC | 706.536 | 431.9182 | 56.8 | 0.12 |

| | | | | |
|---|---|---|---|---|
| pHILIC | 706.536 | 318.0936 | 56.8 | 0.12 |
| pHILIC | 706.536 | 372.4797 | 56.8 | 0.12 |
| pHILIC | 706.536 | 146.0244 | 56.8 | 0.12 |
| pHILIC | 706.536 | 247.9345 | 56.7 | 0.12 |
| pHILIC | 706.536 | 402.7118 | 56.7 | 0.12 |
| pHILIC | 706.536 | 85.6217 | 56.7 | 0.12 |
| pHILIC | 706.536 | 147.1158 | 56.7 | 0.12 |
| pHILIC | 706.536 | 623.905 | 56.7 | 0.12 |
| pHILIC | 706.536 | 256.3945 | 56.6 | 0.12 |
| pHILIC | 706.536 | 474.8923 | 56.6 | 0.12 |
| pHILIC | 706.536 | 140.4653 | 56.6 | 0.12 |
| pHILIC | 706.536 | 246.9504 | 56.6 | 0.12 |
| pHILIC | 706.536 | 346.8734 | 56.6 | 0.12 |
| pHILIC | 706.536 | 391.8386 | 56.6 | 0.12 |
| pHILIC | 706.536 | 167.1537 | 56.5 | 0.12 |
| pHILIC | 706.536 | 85.4023 | 56.5 | 0.12 |
| pHILIC | 706.536 | 100.2336 | 56.5 | 0.12 |
| pHILIC | 706.536 | 250.7379 | 56.5 | 0.12 |
| pHILIC | 706.536 | 86.0531 | 56.5 | 0.12 |
| pHILIC | 706.536 | 116.2434 | 56.5 | 0.12 |
| pHILIC | 706.536 | 186.6458 | 56.5 | 0.12 |
| pHILIC | 706.536 | 262.036 | 56.5 | 0.12 |
| pHILIC | 706.536 | 172.6999 | 56.4 | 0.12 |
| pHILIC | 706.536 | 490.6912 | 56.4 | 0.12 |
| pHILIC | 706.536 | 283.1447 | 56.4 | 0.12 |
| pHILIC | 706.536 | 238.7866 | 56.4 | 0.12 |
| pHILIC | 706.536 | 567.4379 | 56.4 | 0.12 |
| pHILIC | 706.536 | 82.0295 | 56.4 | 0.12 |
| pHILIC | 706.536 | 119.1911 | 56.4 | 0.12 |
| pHILIC | 706.536 | 119.8391 | 56.4 | 0.12 |
| pHILIC | 706.536 | 274.6217 | 56.4 | 0.12 |
| pHILIC | 706.536 | 304.0111 | 56.3 | 0.12 |
| pHILIC | 706.536 | 259.9808 | 56.3 | 0.12 |
| pHILIC | 706.536 | 568.7726 | 56.3 | 0.12 |
| pHILIC | 706.536 | 133.5188 | 56.3 | 0.12 |
| pHILIC | 706.536 | 84.7262 | 56.3 | 0.12 |
| pHILIC | 706.536 | 113.463 | 56.3 | 0.12 |
| pHILIC | 706.536 | 107.9848 | 56.2 | 0.12 |
| pHILIC | 706.536 | 532.0129 | 56.2 | 0.12 |
| pHILIC | 706.536 | 251.5412 | 56.2 | 0.12 |
| pHILIC | 706.536 | 232.5547 | 56.2 | 0.12 |
| pHILIC | 706.536 | 96.8928 | 56.2 | 0.12 |

| | | | | |
|---|---|---|---|---|
| pHILIC | 706.536 | 506.1292 | 56.2 | 0.12 |
| pHILIC | 706.536 | 500.3335 | 56.2 | 0.12 |
| pHILIC | 706.536 | 151.1092 | 56.1 | 0.12 |
| pHILIC | 706.536 | 452.5414 | 56.1 | 0.12 |
| pHILIC | 706.536 | 134.5471 | 56.1 | 0.12 |
| pHILIC | 706.536 | 250.8058 | 56.1 | 0.12 |
| pHILIC | 706.536 | 188.7191 | 56.1 | 0.12 |
| pHILIC | 706.536 | 104.7458 | 56.1 | 0.12 |
| pHILIC | 706.536 | 114.225 | 56.1 | 0.12 |
| pHILIC | 706.536 | 167.0146 | 56 | 0.12 |
| pHILIC | 706.536 | 154.5947 | 56 | 0.12 |
| pHILIC | 706.536 | 582.736 | 56 | 0.12 |
| pHILIC | 706.536 | 166.5737 | 56 | 0.12 |
| pHILIC | 706.536 | 713.2256 | 56 | 0.12 |
| pHILIC | 706.536 | 211.3999 | 56 | 0.12 |
| pHILIC | 706.536 | 437.0158 | 56 | 0.12 |
| pHILIC | 706.536 | 79.428 | 55.9 | 0.12 |
| pHILIC | 706.536 | 160.3705 | 55.9 | 0.12 |
| pHILIC | 706.536 | 177.7552 | 55.9 | 0.12 |
| pHILIC | 706.536 | 664.1204 | 55.9 | 0.12 |
| pHILIC | 706.536 | 142.8146 | 55.9 | 0.12 |
| pHILIC | 706.536 | 120.7182 | 55.9 | 0.12 |
| pHILIC | 706.536 | 513.3389 | 55.9 | 0.12 |
| pHILIC | 706.536 | 88.7229 | 55.9 | 0.12 |
| pHILIC | 706.536 | 85.2709 | 55.9 | 0.12 |
| pHILIC | 706.536 | 167.3432 | 55.9 | 0.12 |
| pHILIC | 706.536 | 220.7497 | 55.9 | 0.12 |
| pHILIC | 706.536 | 118.2764 | 55.9 | 0.12 |
| pHILIC | 706.536 | 112.709 | 55.9 | 0.12 |
| pHILIC | 706.536 | 608.7737 | 55.9 | 0.12 |
| pHILIC | 706.536 | 76.3226 | 55.8 | 0.12 |
| pHILIC | 706.536 | 84.9046 | 55.8 | 0.12 |
| pHILIC | 706.536 | 565.628 | 55.8 | 0.12 |
| pHILIC | 706.536 | 230.2263 | 55.8 | 0.12 |
| pHILIC | 706.536 | 402.8759 | 55.8 | 0.12 |
| pHILIC | 706.536 | 207.7174 | 55.8 | 0.12 |
| pHILIC | 706.536 | 362.3896 | 55.8 | 0.12 |
| pHILIC | 706.536 | 204.8519 | 55.8 | 0.12 |
| pHILIC | 706.536 | 441.1441 | 55.8 | 0.12 |
| pHILIC | 706.536 | 93.7444 | 55.7 | 0.12 |
| pHILIC | 706.536 | 80.8045 | 55.7 | 0.12 |
| pHILIC | 706.536 | 250.1913 | 55.7 | 0.12 |

| | | | | |
|---|---|---|---|---|
| pHILIC | 706.536 | 262.5863 | 55.7 | 0.12 |
| pHILIC | 706.536 | 198.623 | 55.6 | 0.12 |
| pHILIC | 706.536 | 91.2216 | 55.6 | 0.12 |
| pHILIC | 706.536 | 81.3631 | 55.6 | 0.12 |
| pHILIC | 706.536 | 128.7992 | 55.6 | 0.12 |
| pHILIC | 706.536 | 291.2369 | 55.6 | 0.12 |
| pHILIC | 706.536 | 91.2365 | 55.6 | 0.12 |
| pHILIC | 706.536 | 135.038 | 55.5 | 0.12 |
| pHILIC | 706.536 | 551.867 | 55.5 | 0.12 |
| pHILIC | 706.536 | 167.946 | 55.5 | 0.12 |
| pHILIC | 706.536 | 485.731 | 55.5 | 0.12 |
| pHILIC | 706.536 | 291.0883 | 55.5 | 0.12 |
| pHILIC | 706.536 | 86.41 | 55.5 | 0.12 |
| pHILIC | 706.536 | 93.3274 | 55.5 | 0.12 |
| pHILIC | 706.536 | 491.9049 | 55.5 | 0.12 |
| pHILIC | 706.536 | 83.3808 | 55.5 | 0.12 |
| pHILIC | 706.536 | 309.9444 | 55.4 | 0.12 |
| pHILIC | 706.536 | 88.2682 | 55.4 | 0.12 |
| pHILIC | 706.536 | 528.3055 | 55.4 | 0.12 |
| pHILIC | 706.536 | 178.3383 | 55.4 | 0.12 |
| pHILIC | 706.536 | 108.5796 | 55.4 | 0.12 |
| pHILIC | 706.536 | 92.7259 | 55.4 | 0.12 |
| pHILIC | 706.536 | 294.8514 | 55.3 | 0.12 |
| pHILIC | 706.536 | 487.0585 | 55.3 | 0.12 |
| pHILIC | 706.536 | 191.3072 | 55.3 | 0.12 |
| pHILIC | 706.536 | 79.9475 | 55.3 | 0.12 |
| pHILIC | 706.536 | 145.2317 | 55.3 | 0.12 |
| pHILIC | 706.536 | 584.412 | 55.3 | 0.12 |
| pHILIC | 706.536 | 194.7641 | 55.3 | 0.12 |
| pHILIC | 706.536 | 87.4225 | 55.3 | 0.12 |
| pHILIC | 706.536 | 336.0345 | 55.3 | 0.12 |
| pHILIC | 706.536 | 100.3791 | 55.3 | 0.12 |
| pHILIC | 706.536 | 110.5973 | 55.2 | 0.12 |
| pHILIC | 706.536 | 262.2397 | 55.2 | 0.12 |
| pHILIC | 706.536 | 364.0409 | 55.2 | 0.12 |
| pHILIC | 706.536 | 92.8033 | 55.2 | 0.12 |
| pHILIC | 706.536 | 125.7082 | 55.2 | 0.12 |
| pHILIC | 706.536 | 120.961 | 55.2 | 0.12 |
| pHILIC | 706.536 | 80.5056 | 55.2 | 0.12 |
| pHILIC | 706.536 | 182.7404 | 55.2 | 0.12 |
| pHILIC | 706.536 | 137.8856 | 55.2 | 0.12 |
| pHILIC | 706.536 | 93.6831 | 55.2 | 0.12 |

| | | | | |
|---|---|---|---|---|
| pHILIC | 706.536 | 624.1139 | 55.2 | 0.12 |
| pHILIC | 706.536 | 356.596 | 55.2 | 0.12 |
| pHILIC | 706.536 | 81.9189 | 55.2 | 0.12 |
| pHILIC | 706.536 | 98.4382 | 55.1 | 0.12 |
| pHILIC | 706.536 | 87.1365 | 55.1 | 0.12 |
| pHILIC | 706.536 | 103.5298 | 55.1 | 0.12 |
| pHILIC | 706.536 | 362.8878 | 55.1 | 0.12 |
| pHILIC | 706.536 | 367.1884 | 55 | 0.12 |
| pHILIC | 706.536 | 286.2249 | 55 | 0.12 |
| pHILIC | 706.536 | 80.6616 | 55 | 0.12 |
| pHILIC | 706.536 | 155.949 | 55 | 0.12 |
| pHILIC | 706.536 | 374.3095 | 55 | 0.12 |
| pHILIC | 706.536 | 226.4245 | 55 | 0.12 |
| pHILIC | 706.536 | 206.2664 | 55 | 0.12 |
| pHILIC | 706.536 | 157.2576 | 55 | 0.12 |
| pHILIC | 706.536 | 201.9785 | 54.9 | 0.12 |
| pHILIC | 706.536 | 414.3666 | 54.9 | 0.12 |
| pHILIC | 706.536 | 162.6896 | 54.9 | 0.12 |
| pHILIC | 706.536 | 88.9158 | 54.9 | 0.12 |
| pHILIC | 706.536 | 86.2719 | 54.9 | 0.12 |
| pHILIC | 706.536 | 104.2734 | 54.8 | 0.12 |
| pHILIC | 706.536 | 293.7343 | 54.8 | 0.12 |
| pHILIC | 706.536 | 89.9702 | 54.8 | 0.12 |
| pHILIC | 706.536 | 302.0667 | 54.8 | 0.12 |
| pHILIC | 706.536 | 547.7325 | 54.8 | 0.12 |
| pHILIC | 706.536 | 634.0381 | 54.8 | 0.12 |
| pHILIC | 706.536 | 116.9964 | 54.8 | 0.12 |
| pHILIC | 706.536 | 145.2713 | 54.8 | 0.12 |
| pHILIC | 706.536 | 336.2994 | 54.7 | 0.12 |
| pHILIC | 706.536 | 159.1728 | 54.7 | 0.12 |
| pHILIC | 706.536 | 232.3707 | 54.7 | 0.12 |
| pHILIC | 706.536 | 211.185 | 54.7 | 0.12 |
| pHILIC | 706.536 | 186.692 | 54.7 | 0.12 |
| pHILIC | 706.536 | 160.0079 | 54.6 | 0.12 |
| pHILIC | 706.536 | 241.6874 | 54.6 | 0.12 |
| pHILIC | 706.536 | 124.749 | 54.5 | 0.12 |
| pHILIC | 706.536 | 719.3011 | 54.5 | 0.12 |
| pHILIC | 706.536 | 87.5582 | 54.5 | 0.12 |
| pHILIC | 706.536 | 676.4988 | 54.5 | 0.11 |
| pHILIC | 706.536 | 237.6914 | 54.4 | 0.11 |
| pHILIC | 706.536 | 147.255 | 54.4 | 0.11 |
| pHILIC | 706.536 | 441.8239 | 54.4 | 0.11 |

| | | | | |
|---|---|---|---|---|
| pHILIC | 706.536 | 271.5222 | 54.4 | 0.11 |
| pHILIC | 706.536 | 204.77 | 54.4 | 0.11 |
| pHILIC | 706.536 | 374.6474 | 54.4 | 0.11 |
| pHILIC | 706.536 | 98.9386 | 54.4 | 0.11 |
| pHILIC | 706.536 | 93.6081 | 54.4 | 0.11 |
| pHILIC | 706.536 | 104.298 | 54.3 | 0.11 |
| pHILIC | 706.536 | 332.7531 | 54.3 | 0.11 |
| pHILIC | 706.536 | 93.5279 | 54.3 | 0.11 |
| pHILIC | 706.536 | 117.8174 | 54.3 | 0.11 |
| pHILIC | 706.536 | 101.0244 | 54.3 | 0.11 |
| pHILIC | 706.536 | 312.4561 | 54.3 | 0.11 |
| pHILIC | 706.536 | 419.3306 | 54.3 | 0.11 |
| pHILIC | 706.536 | 336.746 | 54.3 | 0.11 |
| pHILIC | 706.536 | 264.8019 | 54.3 | 0.11 |
| pHILIC | 706.536 | 116.5991 | 54.3 | 0.11 |
| pHILIC | 706.536 | 253.0905 | 54.2 | 0.11 |
| pHILIC | 706.536 | 195.9546 | 54.2 | 0.11 |
| pHILIC | 706.536 | 114.5382 | 54.2 | 0.11 |
| pHILIC | 706.536 | 152.6925 | 54.2 | 0.11 |
| pHILIC | 706.536 | 213.5479 | 54.2 | 0.11 |
| pHILIC | 706.536 | 124.1704 | 54.2 | 0.11 |
| pHILIC | 706.536 | 254.4927 | 54.2 | 0.11 |
| pHILIC | 706.536 | 88.6458 | 54.2 | 0.11 |
| pHILIC | 706.536 | 359.0083 | 54.1 | 0.11 |
| pHILIC | 706.536 | 93.6918 | 54.1 | 0.11 |
| pHILIC | 706.536 | 329.1897 | 54.1 | 0.11 |
| pHILIC | 706.536 | 142.0504 | 54.1 | 0.11 |
| pHILIC | 706.536 | 159.8897 | 54.1 | 0.11 |
| pHILIC | 706.536 | 77.9749 | 54.1 | 0.11 |
| pHILIC | 706.536 | 84.6504 | 54.1 | 0.11 |
| pHILIC | 706.536 | 117.8749 | 54.1 | 0.11 |
| pHILIC | 706.536 | 230.347 | 54 | 0.11 |
| pHILIC | 706.536 | 155.5163 | 54 | 0.11 |
| pHILIC | 706.536 | 100.4612 | 54 | 0.11 |
| pHILIC | 706.536 | 128.034 | 54 | 0.11 |
| pHILIC | 706.536 | 122.4465 | 54 | 0.11 |
| pHILIC | 706.536 | 191.0659 | 54 | 0.11 |
| pHILIC | 706.536 | 130.6315 | 53.9 | 0.11 |
| pHILIC | 706.536 | 90.001 | 53.9 | 0.11 |
| pHILIC | 706.536 | 83.0456 | 53.9 | 0.11 |
| pHILIC | 706.536 | 330.0019 | 53.9 | 0.11 |
| pHILIC | 706.536 | 95.8322 | 53.9 | 0.11 |

| | | | | |
|---|---|---|---|---|
| pHILIC | 706.536 | 246.8424 | 53.8 | 0.11 |
| pHILIC | 706.536 | 88.8317 | 53.8 | 0.11 |
| pHILIC | 706.536 | 126.9918 | 53.8 | 0.11 |
| pHILIC | 706.536 | 79.8555 | 53.8 | 0.11 |
| pHILIC | 706.536 | 520.5649 | 53.8 | 0.11 |
| pHILIC | 706.536 | 163.9705 | 53.8 | 0.11 |
| pHILIC | 706.536 | 99.2808 | 53.8 | 0.11 |
| pHILIC | 706.536 | 95.9769 | 53.7 | 0.11 |
| pHILIC | 706.536 | 92.2723 | 53.7 | 0.11 |
| pHILIC | 706.536 | 116.0867 | 53.7 | 0.11 |
| pHILIC | 706.536 | 97.5766 | 53.6 | 0.11 |
| pHILIC | 706.536 | 111.5263 | 53.6 | 0.11 |
| pHILIC | 706.536 | 83.092 | 53.6 | 0.11 |
| pHILIC | 706.536 | 355.5302 | 53.6 | 0.11 |
| pHILIC | 706.536 | 101.0805 | 53.6 | 0.11 |
| pHILIC | 706.536 | 641.0313 | 53.6 | 0.11 |
| pHILIC | 706.536 | 205.3025 | 53.6 | 0.11 |
| pHILIC | 706.536 | 119.0632 | 53.6 | 0.11 |
| pHILIC | 706.536 | 84.0152 | 53.6 | 0.11 |
| pHILIC | 706.536 | 162.8797 | 53.6 | 0.11 |
| pHILIC | 706.536 | 123.2033 | 53.5 | 0.11 |
| pHILIC | 706.536 | 148.6157 | 53.5 | 0.11 |
| pHILIC | 706.536 | 121.3064 | 53.5 | 0.11 |
| pHILIC | 706.536 | 117.7472 | 53.4 | 0.11 |
| pHILIC | 706.536 | 99.9376 | 53.4 | 0.11 |
| pHILIC | 706.536 | 283.593 | 53.4 | 0.11 |
| pHILIC | 706.536 | 133.3655 | 53.4 | 0.11 |
| pHILIC | 706.536 | 159.7171 | 53.4 | 0.11 |
| pHILIC | 706.536 | 84.5839 | 53.4 | 0.11 |
| pHILIC | 706.536 | 108.573 | 53.4 | 0.11 |
| pHILIC | 706.536 | 226.4987 | 53.4 | 0.11 |
| pHILIC | 706.536 | 219.9329 | 53.3 | 0.11 |
| pHILIC | 706.536 | 142.1974 | 53.3 | 0.11 |
| pHILIC | 706.536 | 289.1061 | 53.3 | 0.11 |
| pHILIC | 706.536 | 75.8422 | 53.3 | 0.11 |
| pHILIC | 706.536 | 107.8599 | 53.3 | 0.11 |
| pHILIC | 706.536 | 152.27 | 53.3 | 0.11 |
| pHILIC | 706.536 | 82.3154 | 53.3 | 0.11 |
| pHILIC | 706.536 | 109.4394 | 53.2 | 0.11 |
| pHILIC | 706.536 | 177.1634 | 53.2 | 0.11 |
| pHILIC | 706.536 | 262.0912 | 53.2 | 0.11 |
| pHILIC | 706.536 | 82.6168 | 53.2 | 0.11 |

| | | | | |
|---|---|---|---|---|
| pHILIC | 706.536 | 177.3615 | 53.2 | 0.11 |
| pHILIC | 706.536 | 162.7198 | 53.2 | 0.11 |
| pHILIC | 706.536 | 105.8169 | 53.1 | 0.11 |
| pHILIC | 706.536 | 83.6845 | 53 | 0.11 |
| pHILIC | 706.536 | 100.3514 | 53 | 0.11 |
| pHILIC | 706.536 | 209.2114 | 53 | 0.11 |
| pHILIC | 706.536 | 89.1975 | 53 | 0.11 |
| pHILIC | 706.536 | 104.1832 | 53 | 0.11 |
| pHILIC | 706.536 | 80.9223 | 52.9 | 0.11 |
| pHILIC | 706.536 | 174.6067 | 52.9 | 0.11 |
| pHILIC | 706.536 | 174.7692 | 52.9 | 0.11 |
| pHILIC | 706.536 | 354.7591 | 52.9 | 0.11 |
| pHILIC | 706.536 | 126.073 | 52.9 | 0.11 |
| pHILIC | 706.536 | 77.1481 | 52.8 | 0.11 |
| pHILIC | 706.536 | 126.3365 | 52.8 | 0.11 |
| pHILIC | 706.536 | 257.0162 | 52.8 | 0.11 |
| pHILIC | 706.536 | 670.2288 | 52.8 | 0.11 |
| pHILIC | 706.536 | 78.9695 | 52.7 | 0.11 |
| pHILIC | 706.536 | 101.7574 | 52.7 | 0.11 |
| pHILIC | 706.536 | 457.3517 | 52.7 | 0.11 |
| pHILIC | 706.536 | 187.8014 | 52.7 | 0.11 |
| pHILIC | 706.536 | 324.6626 | 52.7 | 0.11 |
| pHILIC | 706.536 | 138.3355 | 52.7 | 0.11 |
| pHILIC | 706.536 | 153.9962 | 52.6 | 0.11 |
| pHILIC | 706.536 | 305.6783 | 52.6 | 0.11 |
| pHILIC | 706.536 | 89.0733 | 52.6 | 0.11 |
| pHILIC | 706.536 | 170.6667 | 52.6 | 0.11 |
| pHILIC | 706.536 | 165.4317 | 52.5 | 0.11 |
| pHILIC | 706.536 | 80.4994 | 52.5 | 0.11 |
| pHILIC | 706.536 | 80.5287 | 52.5 | 0.11 |
| pHILIC | 706.536 | 115.8875 | 52.5 | 0.11 |
| pHILIC | 706.536 | 111.1007 | 52.5 | 0.11 |
| pHILIC | 706.536 | 336.4347 | 52.4 | 0.11 |
| pHILIC | 706.536 | 168.5797 | 52.4 | 0.11 |
| pHILIC | 706.536 | 187.7334 | 52.4 | 0.11 |
| pHILIC | 706.536 | 512.866 | 52.4 | 0.11 |
| pHILIC | 706.536 | 429.9198 | 52.3 | 0.11 |
| pHILIC | 706.536 | 76.3484 | 52.3 | 0.11 |
| pHILIC | 706.536 | 237.6124 | 52.3 | 0.11 |
| pHILIC | 706.536 | 234.9965 | 52.3 | 0.11 |
| pHILIC | 706.536 | 145.1306 | 52.3 | 0.11 |
| pHILIC | 706.536 | 155.3876 | 52.3 | 0.11 |

| | | | | |
|---|---|---|---|---|
| pHILIC | 706.536 | 125.8403 | 52.3 | 0.11 |
| pHILIC | 706.536 | 94.353 | 52.2 | 0.11 |
| pHILIC | 706.536 | 112.1578 | 52.2 | 0.11 |
| pHILIC | 706.536 | 82.4574 | 52.2 | 0.11 |
| pHILIC | 706.536 | 113.9519 | 52.2 | 0.11 |
| pHILIC | 706.536 | 74.3588 | 52.2 | 0.11 |
| pHILIC | 706.536 | 209.8474 | 52.2 | 0.11 |
| pHILIC | 706.536 | 189.798 | 52.1 | 0.11 |
| pHILIC | 706.536 | 79.1831 | 52.1 | 0.11 |
| pHILIC | 706.536 | 151.4366 | 52.1 | 0.11 |
| pHILIC | 706.536 | 242.2695 | 52.1 | 0.11 |
| pHILIC | 706.536 | 109.4079 | 52.1 | 0.11 |
| pHILIC | 706.536 | 77.6231 | 52 | 0.11 |
| pHILIC | 706.536 | 149.8522 | 52 | 0.11 |
| pHILIC | 706.536 | 459.6372 | 51.9 | 0.11 |
| pHILIC | 706.536 | 77.5615 | 51.9 | 0.11 |
| pHILIC | 706.536 | 121.8613 | 51.9 | 0.11 |
| pHILIC | 706.536 | 77.9896 | 51.9 | 0.11 |
| pHILIC | 706.536 | 76.0961 | 51.8 | 0.11 |
| pHILIC | 706.536 | 113.4509 | 51.8 | 0.11 |
| pHILIC | 706.536 | 136.307 | 51.8 | 0.11 |
| pHILIC | 706.536 | 154.694 | 51.8 | 0.11 |
| pHILIC | 706.536 | 135.935 | 51.8 | 0.11 |
| pHILIC | 706.536 | 379.7046 | 51.7 | 0.11 |
| pHILIC | 706.536 | 191.2398 | 51.6 | 0.11 |
| pHILIC | 706.536 | 145.2569 | 51.6 | 0.11 |
| pHILIC | 706.536 | 93.9078 | 51.6 | 0.11 |
| pHILIC | 706.536 | 113.7871 | 51.5 | 0.11 |
| pHILIC | 706.536 | 152.2162 | 51.5 | 0.11 |
| pHILIC | 706.536 | 188.0157 | 51.4 | 0.11 |
| pHILIC | 706.536 | 290.6356 | 51.3 | 0.11 |
| pHILIC | 706.536 | 161.1906 | 51.3 | 0.11 |
| pHILIC | 706.536 | 76.5964 | 51.2 | 0.11 |
| pHILIC | 706.536 | 183.8693 | 51.2 | 0.11 |
| pHILIC | 706.536 | 223.3798 | 51.1 | 0.11 |
| pHILIC | 706.536 | 97.7933 | 51.1 | 0.11 |
| pHILIC | 706.536 | 327.8889 | 51 | 0.11 |
| pHILIC | 706.536 | 91.3286 | 51 | 0.11 |
| pHILIC | 706.536 | 83.2752 | 50.9 | 0.11 |
| pHILIC | 706.536 | 155.5966 | 50.9 | 0.11 |
| pHILIC | 706.536 | 89.7881 | 50.9 | 0.11 |
| pHILIC | 706.536 | 89.7288 | 50.9 | 0.11 |

| | | | | |
|---|---|---|---|---|
| pHILIC | 706.536 | 102.5332 | 50.8 | 0.11 |
| pHILIC | 706.536 | 146.9271 | 50.7 | 0.11 |
| pHILIC | 706.536 | 175.1733 | 50.6 | 0.11 |
| pHILIC | 706.536 | 80.0539 | 50.6 | 0.11 |
| pHILIC | 706.536 | 104.2163 | 50.6 | 0.11 |
| pHILIC | 706.536 | 104.0771 | 50.6 | 0.11 |
| pHILIC | 706.536 | 270.6 | 50.5 | 0.11 |
| pHILIC | 706.536 | 175.1006 | 50.5 | 0.11 |
| pHILIC | 706.536 | 140.0862 | 50.5 | 0.11 |
| pHILIC | 706.536 | 106.5022 | 50.3 | 0.11 |
| pHILIC | 706.536 | 132.6257 | 50.3 | 0.11 |
| pHILIC | 706.536 | 79.7913 | 50.2 | 0.11 |
| pHILIC | 706.536 | 143.4949 | 50.2 | 0.11 |
| pHILIC | 706.536 | 96.3856 | 50.1 | 0.11 |
| pHILIC | 706.536 | 167.3879 | 49.8 | 0.11 |
| pHILIC | 706.536 | 86.1029 | 49.6 | 0.1 |
| pHILIC | 706.536 | 168.545 | 49.4 | 0.1 |
| pHILIC | 706.536 | 151.5086 | 49.1 | 0.1 |
| pHILIC | 706.536 | 74.673 | 49 | 0.1 |
| pHILIC | 706.536 | 116.0911 | 48.5 | 0.1 |
| pHILIC | 706.536 | 75.1799 | 48.2 | 0.1 |
| pHILIC | 706.536 | 165.3044 | 48.1 | 0.1 |
| pHILIC | 706.536 | 138.8306 | 46.8 | 0.1 |
| pHILIC | 963.639 | 112.8958 | 6258.1 | 100 |
| pHILIC | 963.639 | 228.2204 | 2974.7 | 47.53 |
| pHILIC | 963.639 | 114.8929 | 330.3 | 5.28 |
| pHILIC | 963.639 | 867.1497 | 174.6 | 2.79 |
| pHILIC | 963.639 | 977.5437 | 154.9 | 2.48 |
| pHILIC | 963.639 | 832.5957 | 128.1 | 2.05 |
| pHILIC | 963.639 | 743.6396 | 124.1 | 1.98 |
| pHILIC | 963.639 | 547.29 | 121 | 1.93 |
| pHILIC | 963.639 | 829.9103 | 120 | 1.92 |
| pHILIC | 963.639 | 765.3357 | 119.1 | 1.9 |
| pHILIC | 963.639 | 528.1236 | 117.9 | 1.88 |
| pHILIC | 963.639 | 345.6246 | 117.4 | 1.88 |
| pHILIC | 963.639 | 295.7993 | 116.9 | 1.87 |
| pHILIC | 963.639 | 144.0186 | 115.4 | 1.84 |
| pHILIC | 963.639 | 989.3438 | 114.9 | 1.84 |
| pHILIC | 963.639 | 994.0153 | 113.2 | 1.81 |
| pHILIC | 963.639 | 445.7715 | 112.7 | 1.8 |
| pHILIC | 963.639 | 995.2298 | 112 | 1.79 |
| pHILIC | 963.639 | 708.4456 | 112 | 1.79 |

| | | | | |
|---|---|---|---|---|
| pHILIC | 963.639 | 483.7216 | 111.7 | 1.78 |
| pHILIC | 963.639 | 796.2287 | 111 | 1.77 |
| pHILIC | 963.639 | 223.3698 | 109.8 | 1.75 |
| pHILIC | 963.639 | 226.3601 | 109.5 | 1.75 |
| pHILIC | 963.639 | 322.8168 | 105.7 | 1.69 |
| pHILIC | 963.639 | 158.2016 | 104.3 | 1.67 |
| pHILIC | 963.639 | 476.9483 | 104.1 | 1.66 |
| pHILIC | 963.639 | 179.9219 | 102.7 | 1.64 |
| pHILIC | 963.639 | 271.9709 | 98.7 | 1.58 |
| pHILIC | 963.639 | 771.5771 | 98.6 | 1.58 |
| pHILIC | 963.639 | 833.4716 | 97.4 | 1.56 |
| pHILIC | 963.639 | 366.8642 | 95 | 1.52 |
| pHILIC | 963.639 | 557.7557 | 92.5 | 1.48 |
| pHILIC | 963.639 | 810.8619 | 91 | 1.45 |
| pHILIC | 963.639 | 856.967 | 90.5 | 1.45 |
| pHILIC | 963.639 | 180.3664 | 87.9 | 1.41 |
| pHILIC | 963.639 | 857.5575 | 87.5 | 1.4 |
| pHILIC | 963.639 | 947.7546 | 87.4 | 1.4 |
| pHILIC | 963.639 | 237.5867 | 86.8 | 1.39 |
| pHILIC | 963.639 | 929.1961 | 86.4 | 1.38 |
| pHILIC | 963.639 | 121.2656 | 86.3 | 1.38 |
| pHILIC | 963.639 | 103.3548 | 85.6 | 1.37 |
| pHILIC | 963.639 | 981.9328 | 84.6 | 1.35 |
| pHILIC | 963.639 | 219.9201 | 84.5 | 1.35 |
| pHILIC | 963.639 | 212.6519 | 83.8 | 1.34 |
| pHILIC | 963.639 | 988.4671 | 83.5 | 1.33 |
| pHILIC | 963.639 | 736.8593 | 83.1 | 1.33 |
| pHILIC | 963.639 | 195.2071 | 83.1 | 1.33 |
| pHILIC | 963.639 | 472.4188 | 83 | 1.33 |
| pHILIC | 963.639 | 995.6187 | 82.9 | 1.32 |
| pHILIC | 963.639 | 578.1272 | 82.8 | 1.32 |
| pHILIC | 963.639 | 199.8871 | 82.6 | 1.32 |
| pHILIC | 963.639 | 822.6075 | 82.6 | 1.32 |
| pHILIC | 963.639 | 102.7195 | 82.5 | 1.32 |
| pHILIC | 963.639 | 447.7499 | 82.3 | 1.31 |
| pHILIC | 963.639 | 151.9851 | 82.2 | 1.31 |
| pHILIC | 963.639 | 287.9129 | 82.2 | 1.31 |
| pHILIC | 963.639 | 144.7427 | 82.2 | 1.31 |
| pHILIC | 963.639 | 144.9342 | 82.1 | 1.31 |
| pHILIC | 963.639 | 124.9477 | 81.7 | 1.31 |
| pHILIC | 963.639 | 843.4045 | 81.6 | 1.3 |
| pHILIC | 963.639 | 635.0567 | 81.5 | 1.3 |

| | | | | |
|---|---|---|---|---|
| pHILIC | 963.639 | 627.317 | 81.4 | 1.3 |
| pHILIC | 963.639 | 537.9396 | 81.4 | 1.3 |
| pHILIC | 963.639 | 240.7869 | 81.3 | 1.3 |
| pHILIC | 963.639 | 685.3003 | 81.3 | 1.3 |
| pHILIC | 963.639 | 248.0117 | 81.1 | 1.3 |
| pHILIC | 963.639 | 803.7643 | 80.6 | 1.29 |
| pHILIC | 963.639 | 327.7379 | 80.6 | 1.29 |
| pHILIC | 963.639 | 209.2038 | 80.5 | 1.29 |
| pHILIC | 963.639 | 113.7447 | 80.4 | 1.29 |
| pHILIC | 963.639 | 262.7608 | 80.2 | 1.28 |
| pHILIC | 963.639 | 213.1992 | 80.2 | 1.28 |
| pHILIC | 963.639 | 486.4943 | 80.1 | 1.28 |
| pHILIC | 963.639 | 828.5447 | 79.8 | 1.28 |
| pHILIC | 963.639 | 704.8822 | 79.8 | 1.27 |
| pHILIC | 963.639 | 783.0788 | 79.7 | 1.27 |
| pHILIC | 963.639 | 146.7888 | 79.6 | 1.27 |
| pHILIC | 963.639 | 771.2511 | 79.6 | 1.27 |
| pHILIC | 963.639 | 178.0091 | 79.4 | 1.27 |
| pHILIC | 963.639 | 807.0078 | 79.4 | 1.27 |
| pHILIC | 963.639 | 311.1648 | 79.3 | 1.27 |
| pHILIC | 963.639 | 237.3774 | 79.3 | 1.27 |
| pHILIC | 963.639 | 465.1402 | 79.2 | 1.27 |
| pHILIC | 963.639 | 738.9754 | 79.2 | 1.27 |
| pHILIC | 963.639 | 350.8772 | 79.1 | 1.26 |
| pHILIC | 963.639 | 536.5004 | 79 | 1.26 |
| pHILIC | 963.639 | 548.962 | 78.8 | 1.26 |
| pHILIC | 963.639 | 403.759 | 78.6 | 1.26 |
| pHILIC | 963.639 | 188.588 | 78.6 | 1.26 |
| pHILIC | 963.639 | 465.0046 | 78.6 | 1.26 |
| pHILIC | 963.639 | 805.0936 | 78.5 | 1.26 |
| pHILIC | 963.639 | 160.495 | 78.5 | 1.25 |
| pHILIC | 963.639 | 775.7611 | 78.4 | 1.25 |
| pHILIC | 963.639 | 535.5674 | 78.3 | 1.25 |
| pHILIC | 963.639 | 669.3755 | 78.3 | 1.25 |
| pHILIC | 963.639 | 115.0065 | 78.2 | 1.25 |
| pHILIC | 963.639 | 276.7213 | 77.8 | 1.24 |
| pHILIC | 963.639 | 451.1173 | 77.8 | 1.24 |
| pHILIC | 963.639 | 703.5044 | 77.7 | 1.24 |
| pHILIC | 963.639 | 307.4901 | 77.6 | 1.24 |
| pHILIC | 963.639 | 208.8267 | 77.5 | 1.24 |
| pHILIC | 963.639 | 824.4607 | 77.3 | 1.24 |
| pHILIC | 963.639 | 309.8101 | 77.2 | 1.23 |

| | | | | |
|---|---|---|---|---|
| pHILIC | 963.639 | 758.4076 | 77.2 | 1.23 |
| pHILIC | 963.639 | 132.5057 | 77.1 | 1.23 |
| pHILIC | 963.639 | 458.1982 | 77.1 | 1.23 |
| pHILIC | 963.639 | 477.1588 | 77 | 1.23 |
| pHILIC | 963.639 | 260.6682 | 76.9 | 1.23 |
| pHILIC | 963.639 | 208.5339 | 76.9 | 1.23 |
| pHILIC | 963.639 | 355.4529 | 76.7 | 1.22 |
| pHILIC | 963.639 | 968.8081 | 76.4 | 1.22 |
| pHILIC | 963.639 | 182.426 | 76.3 | 1.22 |
| pHILIC | 963.639 | 278.881 | 76.1 | 1.22 |
| pHILIC | 963.639 | 256.2876 | 76 | 1.22 |
| pHILIC | 963.639 | 240.0951 | 75.9 | 1.21 |
| pHILIC | 963.639 | 278.6363 | 75.8 | 1.21 |
| pHILIC | 963.639 | 142.5686 | 75.7 | 1.21 |
| pHILIC | 963.639 | 283.7116 | 75.7 | 1.21 |
| pHILIC | 963.639 | 124.0313 | 75.6 | 1.21 |
| pHILIC | 963.639 | 135.2648 | 75.6 | 1.21 |
| pHILIC | 963.639 | 174.1738 | 75.4 | 1.21 |
| pHILIC | 963.639 | 587.3994 | 75.4 | 1.21 |
| pHILIC | 963.639 | 398.7181 | 75.4 | 1.2 |
| pHILIC | 963.639 | 502.6179 | 75.2 | 1.2 |
| pHILIC | 963.639 | 212.049 | 75.1 | 1.2 |
| pHILIC | 963.639 | 131.9649 | 75 | 1.2 |
| pHILIC | 963.639 | 142.0062 | 74.9 | 1.2 |
| pHILIC | 963.639 | 234.9342 | 74.9 | 1.2 |
| pHILIC | 963.639 | 331.1518 | 74.8 | 1.2 |
| pHILIC | 963.639 | 322.6149 | 74.8 | 1.19 |
| pHILIC | 963.639 | 107.8024 | 74.7 | 1.19 |
| pHILIC | 963.639 | 584.5681 | 74.6 | 1.19 |
| pHILIC | 963.639 | 233.9117 | 74.6 | 1.19 |
| pHILIC | 963.639 | 484.6612 | 74.6 | 1.19 |
| pHILIC | 963.639 | 603.0973 | 74.5 | 1.19 |
| pHILIC | 963.639 | 116.418 | 74.3 | 1.19 |
| pHILIC | 963.639 | 531.9594 | 74.3 | 1.19 |
| pHILIC | 963.639 | 705.5792 | 74.3 | 1.19 |
| pHILIC | 963.639 | 549.8118 | 74.1 | 1.18 |
| pHILIC | 963.639 | 207.5115 | 73.9 | 1.18 |
| pHILIC | 963.639 | 190.2236 | 73.9 | 1.18 |
| pHILIC | 963.639 | 204.9744 | 73.9 | 1.18 |
| pHILIC | 963.639 | 284.155 | 73.8 | 1.18 |
| pHILIC | 963.639 | 293.9977 | 73.6 | 1.18 |
| pHILIC | 963.639 | 266.7549 | 73.5 | 1.17 |

| | | | | |
|---|---|---|---|---|
| pHILIC | 963.639 | 121.8731 | 73.4 | 1.17 |
| pHILIC | 963.639 | 394.2637 | 73.3 | 1.17 |
| pHILIC | 963.639 | 478.9254 | 73.3 | 1.17 |
| pHILIC | 963.639 | 344.0327 | 73.1 | 1.17 |
| pHILIC | 963.639 | 431.6548 | 73 | 1.17 |
| pHILIC | 963.639 | 227.2187 | 72.9 | 1.16 |
| pHILIC | 963.639 | 704.579 | 72.8 | 1.16 |
| pHILIC | 963.639 | 631.7102 | 72.8 | 1.16 |
| pHILIC | 963.639 | 798.9279 | 72.8 | 1.16 |
| pHILIC | 963.639 | 907.6115 | 72.6 | 1.16 |
| pHILIC | 963.639 | 694.744 | 72.5 | 1.16 |
| pHILIC | 963.639 | 702.0373 | 72.3 | 1.16 |
| pHILIC | 963.639 | 136.9192 | 72.3 | 1.15 |
| pHILIC | 963.639 | 248.4137 | 72.3 | 1.15 |
| pHILIC | 963.639 | 191.1596 | 72.2 | 1.15 |
| pHILIC | 963.639 | 132.1998 | 72.2 | 1.15 |
| pHILIC | 963.639 | 113.1008 | 71.8 | 1.15 |
| pHILIC | 963.639 | 252.1527 | 71.8 | 1.15 |
| pHILIC | 963.639 | 341.7367 | 71.6 | 1.14 |
| pHILIC | 963.639 | 369.9637 | 71.6 | 1.14 |
| pHILIC | 963.639 | 607.1629 | 71.5 | 1.14 |
| pHILIC | 963.639 | 464.4237 | 71.4 | 1.14 |
| pHILIC | 963.639 | 576.2579 | 71.3 | 1.14 |
| pHILIC | 963.639 | 111.9741 | 71.3 | 1.14 |
| pHILIC | 963.639 | 957.3658 | 71.2 | 1.14 |
| pHILIC | 963.639 | 253.8678 | 71.2 | 1.14 |
| pHILIC | 963.639 | 719.7317 | 71 | 1.13 |
| pHILIC | 963.639 | 133.085 | 70.8 | 1.13 |
| pHILIC | 963.639 | 136.9731 | 70.6 | 1.13 |
| pHILIC | 963.639 | 250.0538 | 70.6 | 1.13 |
| pHILIC | 963.639 | 214.6597 | 70.5 | 1.13 |
| pHILIC | 963.639 | 354.8246 | 70.5 | 1.13 |
| pHILIC | 963.639 | 217.5824 | 70.4 | 1.13 |
| pHILIC | 963.639 | 102.4652 | 70.4 | 1.13 |
| pHILIC | 963.639 | 212.3766 | 70.4 | 1.13 |
| pHILIC | 963.639 | 387.4591 | 70.4 | 1.12 |
| pHILIC | 963.639 | 715.4428 | 70.4 | 1.12 |
| pHILIC | 963.639 | 435.8289 | 70.4 | 1.12 |
| pHILIC | 963.639 | 473.5789 | 70.3 | 1.12 |
| pHILIC | 963.639 | 196.3898 | 70.2 | 1.12 |
| pHILIC | 963.639 | 125.7708 | 70 | 1.12 |
| pHILIC | 963.639 | 351.7171 | 69.9 | 1.12 |

| | | | | |
|---|---|---|---|---|
| pHILIC | 963.639 | 184.821 | 69.9 | 1.12 |
| pHILIC | 963.639 | 342.3466 | 69.9 | 1.12 |
| pHILIC | 963.639 | 120.2981 | 69.8 | 1.12 |
| pHILIC | 963.639 | 159.4077 | 69.5 | 1.11 |
| pHILIC | 963.639 | 339.0364 | 69.5 | 1.11 |
| pHILIC | 963.639 | 485.0692 | 69.5 | 1.11 |
| pHILIC | 963.639 | 713.2088 | 69.4 | 1.11 |
| pHILIC | 963.639 | 770.9106 | 69.4 | 1.11 |
| pHILIC | 963.639 | 393.8164 | 69.2 | 1.11 |
| pHILIC | 963.639 | 489.1351 | 69.1 | 1.1 |
| pHILIC | 963.639 | 472.1029 | 69.1 | 1.1 |
| pHILIC | 963.639 | 145.5447 | 69 | 1.1 |
| pHILIC | 963.639 | 103.6238 | 69 | 1.1 |
| pHILIC | 963.639 | 773.4942 | 68.9 | 1.1 |
| pHILIC | 963.639 | 652.8505 | 68.8 | 1.1 |
| pHILIC | 963.639 | 383.0381 | 68.6 | 1.1 |
| pHILIC | 963.639 | 527.8783 | 68.4 | 1.09 |
| pHILIC | 963.639 | 521.8789 | 68.3 | 1.09 |
| pHILIC | 963.639 | 461.2165 | 68.2 | 1.09 |
| pHILIC | 963.639 | 406.0019 | 68.1 | 1.09 |
| pHILIC | 963.639 | 107.9172 | 67.9 | 1.09 |
| pHILIC | 963.639 | 539.2285 | 67.6 | 1.08 |
| pHILIC | 963.639 | 284.8636 | 67.5 | 1.08 |
| pHILIC | 963.639 | 427.6464 | 67.3 | 1.08 |
| pHILIC | 963.639 | 708.0713 | 67.2 | 1.07 |
| pHILIC | 963.639 | 276.0795 | 67.2 | 1.07 |
| pHILIC | 963.639 | 171.4963 | 66.9 | 1.07 |
| pHILIC | 963.639 | 401.7823 | 66.5 | 1.06 |
| pHILIC | 963.639 | 141.7619 | 66.2 | 1.06 |
| pHILIC | 963.639 | 385.335 | 65.9 | 1.05 |
| pHILIC | 963.639 | 381.3242 | 65.9 | 1.05 |
| pHILIC | 963.639 | 540.449 | 65.7 | 1.05 |
| pHILIC | 963.639 | 398.9128 | 65.6 | 1.05 |
| pHILIC | 963.639 | 146.3979 | 65.3 | 1.04 |
| pHILIC | 963.639 | 176.9608 | 64.9 | 1.04 |
| pHILIC | 963.639 | 143.6245 | 64.6 | 1.03 |
| pHILIC | 963.639 | 277.8386 | 64.4 | 1.03 |
| pHILIC | 963.639 | 113.4146 | 64.4 | 1.03 |
| pHILIC | 963.639 | 304.2203 | 63.6 | 1.02 |
| pHILIC | 963.639 | 159.5844 | 62.9 | 1.01 |
| pHILIC | 963.639 | 236.4395 | 62.8 | 1 |
| pHILIC | 963.639 | 123.4401 | 62.7 | 1 |

| | | | | |
|---|---|---|---|---|
| pHILIC | 963.639 | 590.5679 | 61.8 | 0.99 |
| pHILIC | 963.639 | 139.8396 | 61.5 | 0.98 |
| pHILIC | 963.639 | 116.6845 | 61.4 | 0.98 |
| pHILIC | 963.639 | 448.4882 | 61.4 | 0.98 |
| pHILIC | 963.639 | 225.7941 | 61.4 | 0.98 |
| pHILIC | 963.639 | 150.8971 | 61.1 | 0.98 |
| pHILIC | 963.639 | 100.373 | 60.5 | 0.97 |
| pHILIC | 963.639 | 126.0191 | 60.4 | 0.97 |
| pHILIC | 963.639 | 147.4945 | 60 | 0.96 |
| pHILIC | 963.639 | 431.0994 | 59.4 | 0.95 |
| pHILIC | 963.639 | 731.6984 | 59 | 0.94 |
| pHILIC | 963.639 | 305.6019 | 59 | 0.94 |
| pHILIC | 963.639 | 212.5462 | 58.8 | 0.94 |
| pHILIC | 963.639 | 546.7202 | 57.9 | 0.93 |
| pHILIC | 963.639 | 283.1874 | 57.7 | 0.92 |
| pHILIC | 963.639 | 116.8146 | 57.6 | 0.92 |
| pHILIC | 963.639 | 345.2548 | 57.4 | 0.92 |
| pHILIC | 963.639 | 116.5873 | 56.9 | 0.91 |
| pHILIC | 963.639 | 360.6004 | 55.1 | 0.88 |
| pHILIC | 963.639 | 353.2487 | 54.4 | 0.87 |
| pHILIC | 963.639 | 195.0325 | 53.2 | 0.85 |
| pHILIC | 963.639 | 349.6901 | 53.1 | 0.85 |
| pHILIC | 963.639 | 436.6694 | 52.8 | 0.84 |
| pHILIC | 963.639 | 864.8091 | 52.7 | 0.84 |
| pHILIC | 963.639 | 123.6816 | 52.5 | 0.84 |
| pHILIC | 963.639 | 367.6469 | 52 | 0.83 |
| pHILIC | 963.639 | 542.4541 | 51 | 0.82 |
| pHILIC | 963.639 | 260.7942 | 50.9 | 0.81 |
| pHILIC | 963.639 | 128.7421 | 50.3 | 0.8 |
| pHILIC | 963.639 | 122.492 | 50.3 | 0.8 |
| pHILIC | 963.639 | 364.5322 | 50.3 | 0.8 |
| pHILIC | 963.639 | 268.6285 | 49.7 | 0.79 |
| pHILIC | 963.639 | 200.4869 | 49.3 | 0.79 |
| pHILIC | 963.639 | 493.3737 | 49.3 | 0.79 |
| pHILIC | 963.639 | 307.765 | 49.1 | 0.78 |
| pHILIC | 963.639 | 298.6449 | 48.9 | 0.78 |
| pHILIC | 963.639 | 487.0218 | 48.9 | 0.78 |
| pHILIC | 963.639 | 340.5695 | 48.8 | 0.78 |
| pHILIC | 963.639 | 678.809 | 48.5 | 0.78 |
| pHILIC | 963.639 | 857.9708 | 48.5 | 0.77 |
| pHILIC | 963.639 | 219.7848 | 48.5 | 0.77 |
| pHILIC | 963.639 | 257.3686 | 48.4 | 0.77 |

| | | | | |
|---|---|---|---|---|
| pHILIC | 963.639 | 429.4459 | 47.9 | 0.76 |
| pHILIC | 963.639 | 650.3227 | 47.8 | 0.76 |
| pHILIC | 963.639 | 448.241 | 47.8 | 0.76 |
| pHILIC | 963.639 | 173.9867 | 47.8 | 0.76 |
| pHILIC | 963.639 | 849.1714 | 47.7 | 0.76 |
| pHILIC | 963.639 | 141.4682 | 47.7 | 0.76 |
| pHILIC | 963.639 | 134.3724 | 47.6 | 0.76 |
| pHILIC | 963.639 | 721.2546 | 47.6 | 0.76 |
| pHILIC | 963.639 | 815.8365 | 47.6 | 0.76 |
| pHILIC | 963.639 | 101.7451 | 47.5 | 0.76 |
| pHILIC | 963.639 | 281.538 | 47.5 | 0.76 |
| pHILIC | 963.639 | 935.5787 | 47.5 | 0.76 |
| pHILIC | 963.639 | 346.1988 | 47.4 | 0.76 |
| pHILIC | 963.639 | 210.3213 | 47.3 | 0.76 |
| pHILIC | 963.639 | 251.6526 | 47.2 | 0.75 |
| pHILIC | 963.639 | 213.0088 | 47.2 | 0.75 |
| pHILIC | 963.639 | 940.4279 | 47.2 | 0.75 |
| pHILIC | 963.639 | 712.6039 | 47.1 | 0.75 |
| pHILIC | 963.639 | 878.7195 | 47 | 0.75 |
| pHILIC | 963.639 | 277.2207 | 47 | 0.75 |
| pHILIC | 963.639 | 183.4295 | 47 | 0.75 |
| pHILIC | 963.639 | 692.0385 | 46.9 | 0.75 |
| pHILIC | 963.639 | 249.9122 | 46.9 | 0.75 |
| pHILIC | 963.639 | 746.4411 | 46.9 | 0.75 |
| pHILIC | 963.639 | 126.9844 | 46.9 | 0.75 |
| pHILIC | 963.639 | 178.7457 | 46.9 | 0.75 |
| pHILIC | 963.639 | 920.0501 | 46.9 | 0.75 |
| pHILIC | 963.639 | 674.3962 | 46.8 | 0.75 |
| pHILIC | 963.639 | 796.9885 | 46.8 | 0.75 |
| pHILIC | 963.639 | 852.8124 | 46.8 | 0.75 |
| pHILIC | 963.639 | 405.028 | 46.7 | 0.75 |
| pHILIC | 963.639 | 166.2565 | 46.7 | 0.75 |
| pHILIC | 963.639 | 130.0251 | 46.7 | 0.75 |
| pHILIC | 963.639 | 533.9664 | 46.7 | 0.75 |
| pHILIC | 963.639 | 100.6001 | 46.5 | 0.74 |
| pHILIC | 963.639 | 622.2011 | 46.5 | 0.74 |
| pHILIC | 963.639 | 776.8383 | 46.5 | 0.74 |
| pHILIC | 963.639 | 115.2215 | 46.5 | 0.74 |
| pHILIC | 963.639 | 586.5153 | 46.4 | 0.74 |
| pHILIC | 963.639 | 428.6774 | 46.4 | 0.74 |
| pHILIC | 963.639 | 167.8165 | 46.4 | 0.74 |
| pHILIC | 963.639 | 124.1225 | 46.4 | 0.74 |

| | | | | |
|---|---|---|---|---|
| pHILIC | 963.639 | 776.4958 | 46.3 | 0.74 |
| pHILIC | 963.639 | 388.7282 | 46.3 | 0.74 |
| pHILIC | 963.639 | 195.6058 | 46.3 | 0.74 |
| pHILIC | 963.639 | 195.1515 | 46.2 | 0.74 |
| pHILIC | 963.639 | 789.9998 | 46.2 | 0.74 |
| pHILIC | 963.639 | 792.5208 | 46.2 | 0.74 |
| pHILIC | 963.639 | 801.7491 | 46.2 | 0.74 |
| pHILIC | 963.639 | 165.0145 | 46.1 | 0.74 |
| pHILIC | 963.639 | 665.3998 | 46.1 | 0.74 |
| pHILIC | 963.639 | 465.5277 | 46.1 | 0.74 |
| pHILIC | 963.639 | 255.3607 | 46.1 | 0.74 |
| pHILIC | 963.639 | 277.4735 | 46 | 0.74 |
| pHILIC | 963.639 | 479.1413 | 46 | 0.74 |
| pHILIC | 963.639 | 258.292 | 46 | 0.74 |
| pHILIC | 963.639 | 882.7352 | 46 | 0.73 |
| pHILIC | 963.639 | 226.4908 | 45.9 | 0.73 |
| pHILIC | 963.639 | 505.0707 | 45.9 | 0.73 |
| pHILIC | 963.639 | 583.0542 | 45.9 | 0.73 |
| pHILIC | 963.639 | 177.7857 | 45.9 | 0.73 |
| pHILIC | 963.639 | 294.1757 | 45.9 | 0.73 |
| pHILIC | 963.639 | 566.3763 | 45.8 | 0.73 |
| pHILIC | 963.639 | 574.709 | 45.8 | 0.73 |
| pHILIC | 963.639 | 708.7666 | 45.8 | 0.73 |
| pHILIC | 963.639 | 265.6013 | 45.7 | 0.73 |
| pHILIC | 963.639 | 238.5279 | 45.7 | 0.73 |
| pHILIC | 963.639 | 303.7287 | 45.5 | 0.73 |
| pHILIC | 963.639 | 403.2074 | 45.4 | 0.73 |
| pHILIC | 963.639 | 130.8792 | 45.3 | 0.72 |
| pHILIC | 963.639 | 150.2329 | 45.3 | 0.72 |
| pHILIC | 963.639 | 219.5984 | 45.3 | 0.72 |
| pHILIC | 963.639 | 755.3823 | 45.2 | 0.72 |
| pHILIC | 963.639 | 804.6759 | 45.2 | 0.72 |
| pHILIC | 963.639 | 209.0355 | 45.2 | 0.72 |
| pHILIC | 963.639 | 638.8521 | 45.2 | 0.72 |
| pHILIC | 963.639 | 454.2956 | 45.2 | 0.72 |
| pHILIC | 963.639 | 202.9523 | 45.1 | 0.72 |
| pHILIC | 963.639 | 328.6041 | 45.1 | 0.72 |
| pHILIC | 963.639 | 142.9324 | 45.1 | 0.72 |
| pHILIC | 963.639 | 143.1356 | 45.1 | 0.72 |
| pHILIC | 963.639 | 141.0111 | 45.1 | 0.72 |
| pHILIC | 963.639 | 109.8587 | 45 | 0.72 |
| pHILIC | 963.639 | 137.098 | 45 | 0.72 |

| | | | | |
|---|---|---|---|---|
| pHILIC | 963.639 | 790.2508 | 45 | 0.72 |
| pHILIC | 963.639 | 828.8931 | 45 | 0.72 |
| pHILIC | 963.639 | 155.2467 | 45 | 0.72 |
| pHILIC | 963.639 | 183.817 | 45 | 0.72 |
| pHILIC | 963.639 | 784.7834 | 44.9 | 0.72 |
| pHILIC | 963.639 | 244.008 | 44.9 | 0.72 |
| pHILIC | 963.639 | 324.2573 | 44.9 | 0.72 |
| pHILIC | 963.639 | 498.1815 | 44.9 | 0.72 |
| pHILIC | 963.639 | 402.4962 | 44.9 | 0.72 |
| pHILIC | 963.639 | 140.1431 | 44.9 | 0.72 |
| pHILIC | 963.639 | 159.9205 | 44.8 | 0.72 |
| pHILIC | 963.639 | 389.1762 | 44.8 | 0.72 |
| pHILIC | 963.639 | 950.4005 | 44.7 | 0.72 |
| pHILIC | 963.639 | 991.287 | 44.7 | 0.71 |
| pHILIC | 963.639 | 480.8116 | 44.7 | 0.71 |
| pHILIC | 963.639 | 187.3179 | 44.7 | 0.71 |
| pHILIC | 963.639 | 620.5345 | 44.6 | 0.71 |
| pHILIC | 963.639 | 464.6312 | 44.6 | 0.71 |
| pHILIC | 963.639 | 285.7832 | 44.6 | 0.71 |
| pHILIC | 963.639 | 629.6815 | 44.6 | 0.71 |
| pHILIC | 963.639 | 157.4128 | 44.5 | 0.71 |
| pHILIC | 963.639 | 119.4274 | 44.5 | 0.71 |
| pHILIC | 963.639 | 523.1998 | 44.5 | 0.71 |
| pHILIC | 963.639 | 414.3689 | 44.5 | 0.71 |
| pHILIC | 963.639 | 106.9495 | 44.5 | 0.71 |
| pHILIC | 963.639 | 607.8135 | 44.5 | 0.71 |
| pHILIC | 963.639 | 506.1934 | 44.5 | 0.71 |
| pHILIC | 963.639 | 393.5316 | 44.5 | 0.71 |
| pHILIC | 963.639 | 200.6951 | 44.5 | 0.71 |
| pHILIC | 963.639 | 383.8785 | 44.5 | 0.71 |
| pHILIC | 963.639 | 266.0356 | 44.5 | 0.71 |
| pHILIC | 963.639 | 718.556 | 44.4 | 0.71 |
| pHILIC | 963.639 | 192.1684 | 44.4 | 0.71 |
| pHILIC | 963.639 | 559.4985 | 44.4 | 0.71 |
| pHILIC | 963.639 | 204.9104 | 44.3 | 0.71 |
| pHILIC | 963.639 | 111.3555 | 44.3 | 0.71 |
| pHILIC | 963.639 | 497.7924 | 44.2 | 0.71 |
| pHILIC | 963.639 | 641.6273 | 44.2 | 0.71 |
| pHILIC | 963.639 | 142.6789 | 44.2 | 0.71 |
| pHILIC | 963.639 | 560.8556 | 44.2 | 0.71 |
| pHILIC | 963.639 | 284.2328 | 44.1 | 0.71 |
| pHILIC | 963.639 | 155.9842 | 44.1 | 0.71 |

| | | | | |
|---|---|---|---|---|
| pHILIC | 963.639 | 820.0302 | 44.1 | 0.71 |
| pHILIC | 963.639 | 455.7968 | 44.1 | 0.7 |
| pHILIC | 963.639 | 122.3401 | 44.1 | 0.7 |
| pHILIC | 963.639 | 115.0167 | 44.1 | 0.7 |
| pHILIC | 963.639 | 506.9323 | 44.1 | 0.7 |
| pHILIC | 963.639 | 107.2435 | 44.1 | 0.7 |
| pHILIC | 963.639 | 316.4883 | 44.1 | 0.7 |
| pHILIC | 963.639 | 438.0414 | 44.1 | 0.7 |
| pHILIC | 963.639 | 134.4148 | 44.1 | 0.7 |
| pHILIC | 963.639 | 510.0162 | 44.1 | 0.7 |
| pHILIC | 963.639 | 944.8395 | 44 | 0.7 |
| pHILIC | 963.639 | 140.7573 | 44 | 0.7 |
| pHILIC | 963.639 | 148.0235 | 44 | 0.7 |
| pHILIC | 963.639 | 913.6663 | 44 | 0.7 |
| pHILIC | 963.639 | 398.1816 | 44 | 0.7 |
| pHILIC | 963.639 | 319.7958 | 43.9 | 0.7 |
| pHILIC | 963.639 | 709.4185 | 43.9 | 0.7 |
| pHILIC | 963.639 | 134.7685 | 43.9 | 0.7 |
| pHILIC | 963.639 | 235.0181 | 43.9 | 0.7 |
| pHILIC | 963.639 | 937.7107 | 43.9 | 0.7 |
| pHILIC | 963.639 | 466.7543 | 43.9 | 0.7 |
| pHILIC | 963.639 | 762.7402 | 43.8 | 0.7 |
| pHILIC | 963.639 | 506.3457 | 43.8 | 0.7 |
| pHILIC | 963.639 | 219.1734 | 43.8 | 0.7 |
| pHILIC | 963.639 | 538.5063 | 43.8 | 0.7 |
| pHILIC | 963.639 | 117.5109 | 43.8 | 0.7 |
| pHILIC | 963.639 | 561.6269 | 43.8 | 0.7 |
| pHILIC | 963.639 | 153.0955 | 43.8 | 0.7 |
| pHILIC | 963.639 | 101.444 | 43.8 | 0.7 |
| pHILIC | 963.639 | 142.9655 | 43.7 | 0.7 |
| pHILIC | 963.639 | 284.4038 | 43.7 | 0.7 |
| pHILIC | 963.639 | 519.6553 | 43.7 | 0.7 |
| pHILIC | 963.639 | 667.3738 | 43.7 | 0.7 |
| pHILIC | 963.639 | 139.2369 | 43.7 | 0.7 |
| pHILIC | 963.639 | 142.7719 | 43.6 | 0.7 |
| pHILIC | 963.639 | 442.136 | 43.6 | 0.7 |
| pHILIC | 963.639 | 135.9667 | 43.6 | 0.7 |
| pHILIC | 963.639 | 855.3255 | 43.6 | 0.7 |
| pHILIC | 963.639 | 200.1398 | 43.6 | 0.7 |
| pHILIC | 963.639 | 143.6543 | 43.6 | 0.7 |
| pHILIC | 963.639 | 542.0724 | 43.6 | 0.7 |
| pHILIC | 963.639 | 146.9842 | 43.6 | 0.7 |

| | | | | |
|---|---|---|---|---|
| pHILIC | 963.639 | 630.5032 | 43.6 | 0.7 |
| pHILIC | 963.639 | 987.5246 | 43.6 | 0.7 |
| pHILIC | 963.639 | 241.7509 | 43.5 | 0.7 |
| pHILIC | 963.639 | 471.8159 | 43.5 | 0.7 |
| pHILIC | 963.639 | 111.6645 | 43.5 | 0.7 |
| pHILIC | 963.639 | 860.9476 | 43.5 | 0.7 |
| pHILIC | 963.639 | 195.3871 | 43.5 | 0.69 |
| pHILIC | 963.639 | 393.9669 | 43.5 | 0.69 |
| pHILIC | 963.639 | 902.4321 | 43.5 | 0.69 |
| pHILIC | 963.639 | 722.2487 | 43.4 | 0.69 |
| pHILIC | 963.639 | 567.6467 | 43.4 | 0.69 |
| pHILIC | 963.639 | 545.5886 | 43.4 | 0.69 |
| pHILIC | 963.639 | 154.5731 | 43.4 | 0.69 |
| pHILIC | 963.639 | 310.7416 | 43.4 | 0.69 |
| pHILIC | 963.639 | 952.9402 | 43.4 | 0.69 |
| pHILIC | 963.639 | 111.4657 | 43.3 | 0.69 |
| pHILIC | 963.639 | 432.3167 | 43.3 | 0.69 |
| pHILIC | 963.639 | 118.2986 | 43.3 | 0.69 |
| pHILIC | 963.639 | 303.2462 | 43.3 | 0.69 |
| pHILIC | 963.639 | 100.4706 | 43.3 | 0.69 |
| pHILIC | 963.639 | 140.6778 | 43.3 | 0.69 |
| pHILIC | 963.639 | 627.6451 | 43.3 | 0.69 |
| pHILIC | 963.639 | 578.4824 | 43.3 | 0.69 |
| pHILIC | 963.639 | 780.0331 | 43.3 | 0.69 |
| pHILIC | 963.639 | 110.1648 | 43.2 | 0.69 |
| pHILIC | 963.639 | 358.7982 | 43.2 | 0.69 |
| pHILIC | 963.639 | 132.4356 | 43.2 | 0.69 |
| pHILIC | 963.639 | 277.4452 | 43.2 | 0.69 |
| pHILIC | 963.639 | 126.9519 | 43.1 | 0.69 |
| pHILIC | 963.639 | 105.8301 | 43.1 | 0.69 |
| pHILIC | 963.639 | 308.8437 | 43.1 | 0.69 |
| pHILIC | 963.639 | 682.6356 | 43.1 | 0.69 |
| pHILIC | 963.639 | 357.3331 | 43.1 | 0.69 |
| pHILIC | 963.639 | 321.7395 | 43 | 0.69 |
| pHILIC | 963.639 | 273.7763 | 43 | 0.69 |
| pHILIC | 963.639 | 409.2981 | 43 | 0.69 |
| pHILIC | 963.639 | 439.0035 | 43 | 0.69 |
| pHILIC | 963.639 | 689.3347 | 42.9 | 0.69 |
| pHILIC | 963.639 | 160.0216 | 42.9 | 0.69 |
| pHILIC | 963.639 | 478.0746 | 42.9 | 0.69 |
| pHILIC | 963.639 | 380.6419 | 42.9 | 0.69 |
| pHILIC | 963.639 | 137.1683 | 42.9 | 0.69 |

| | | | | |
|---|---|---|---|---|
| pHILIC | 963.639 | 187.0081 | 42.9 | 0.69 |
| pHILIC | 963.639 | 101.9193 | 42.9 | 0.68 |
| pHILIC | 963.639 | 170.7731 | 42.8 | 0.68 |
| pHILIC | 963.639 | 486.877 | 42.8 | 0.68 |
| pHILIC | 963.639 | 151.3951 | 42.8 | 0.68 |
| pHILIC | 963.639 | 400.8306 | 42.8 | 0.68 |
| pHILIC | 963.639 | 809.2179 | 42.8 | 0.68 |
| pHILIC | 963.639 | 947.2272 | 42.7 | 0.68 |
| pHILIC | 963.639 | 191.4381 | 42.7 | 0.68 |
| pHILIC | 963.639 | 145.4311 | 42.7 | 0.68 |
| pHILIC | 963.639 | 807.6688 | 42.7 | 0.68 |
| pHILIC | 963.639 | 531.6194 | 42.7 | 0.68 |
| pHILIC | 963.639 | 116.0597 | 42.7 | 0.68 |
| pHILIC | 963.639 | 280.7123 | 42.7 | 0.68 |
| pHILIC | 963.639 | 146.9141 | 42.7 | 0.68 |
| pHILIC | 963.639 | 118.9727 | 42.7 | 0.68 |
| pHILIC | 963.639 | 729.2988 | 42.7 | 0.68 |
| pHILIC | 963.639 | 889.4804 | 42.6 | 0.68 |
| pHILIC | 963.639 | 687.741 | 42.6 | 0.68 |
| pHILIC | 963.639 | 163.462 | 42.6 | 0.68 |
| pHILIC | 963.639 | 652.251 | 42.6 | 0.68 |
| pHILIC | 963.639 | 347.6456 | 42.6 | 0.68 |
| pHILIC | 963.639 | 501.5402 | 42.6 | 0.68 |
| pHILIC | 963.639 | 521.2097 | 42.6 | 0.68 |
| pHILIC | 963.639 | 323.1737 | 42.6 | 0.68 |
| pHILIC | 963.639 | 836.9291 | 42.5 | 0.68 |
| pHILIC | 963.639 | 416.7764 | 42.5 | 0.68 |
| pHILIC | 963.639 | 330.0375 | 42.5 | 0.68 |
| pHILIC | 963.639 | 105.5706 | 42.4 | 0.68 |
| pHILIC | 963.639 | 186.5105 | 42.4 | 0.68 |
| pHILIC | 963.639 | 235.3238 | 42.4 | 0.68 |
| pHILIC | 963.639 | 490.5888 | 42.4 | 0.68 |
| pHILIC | 963.639 | 616.0089 | 42.4 | 0.68 |
| pHILIC | 963.639 | 903.7137 | 42.4 | 0.68 |
| pHILIC | 963.639 | 292.4521 | 42.4 | 0.68 |
| pHILIC | 963.639 | 193.2189 | 42.4 | 0.68 |
| pHILIC | 963.639 | 292.6714 | 42.4 | 0.68 |
| pHILIC | 963.639 | 339.9408 | 42.4 | 0.68 |
| pHILIC | 963.639 | 180.4908 | 42.3 | 0.68 |
| pHILIC | 963.639 | 581.8267 | 42.3 | 0.68 |
| pHILIC | 963.639 | 105.1688 | 42.3 | 0.68 |
| pHILIC | 963.639 | 488.0042 | 42.3 | 0.68 |

| | | | | |
|---|---|---|---|---|
| pHILIC | 963.639 | 105.7249 | 42.3 | 0.68 |
| pHILIC | 963.639 | 218.3284 | 42.3 | 0.68 |
| pHILIC | 963.639 | 180.5382 | 42.3 | 0.68 |
| pHILIC | 963.639 | 719.9394 | 42.3 | 0.68 |
| pHILIC | 963.639 | 104.1048 | 42.3 | 0.68 |
| pHILIC | 963.639 | 769.6486 | 42.3 | 0.68 |
| pHILIC | 963.639 | 140.9729 | 42.2 | 0.68 |
| pHILIC | 963.639 | 851.8797 | 42.2 | 0.67 |
| pHILIC | 963.639 | 314.8729 | 42.2 | 0.67 |
| pHILIC | 963.639 | 751.3256 | 42.2 | 0.67 |
| pHILIC | 963.639 | 280.571 | 42.2 | 0.67 |
| pHILIC | 963.639 | 636.7523 | 42.2 | 0.67 |
| pHILIC | 963.639 | 225.4609 | 42.2 | 0.67 |
| pHILIC | 963.639 | 673.2637 | 42.2 | 0.67 |
| pHILIC | 963.639 | 143.8253 | 42.1 | 0.67 |
| pHILIC | 963.639 | 168.9629 | 42.1 | 0.67 |
| pHILIC | 963.639 | 271.6037 | 42.1 | 0.67 |
| pHILIC | 963.639 | 740.3683 | 42.1 | 0.67 |
| pHILIC | 963.639 | 267.6108 | 42.1 | 0.67 |
| pHILIC | 963.639 | 262.2327 | 42.1 | 0.67 |
| pHILIC | 963.639 | 467.3819 | 42.1 | 0.67 |
| pHILIC | 963.639 | 491.0523 | 42 | 0.67 |
| pHILIC | 963.639 | 252.2723 | 42 | 0.67 |
| pHILIC | 963.639 | 163.8339 | 42 | 0.67 |
| pHILIC | 963.639 | 593.849 | 42 | 0.67 |
| pHILIC | 963.639 | 385.0894 | 42 | 0.67 |
| pHILIC | 963.639 | 219.1494 | 42 | 0.67 |
| pHILIC | 963.639 | 666.0985 | 42 | 0.67 |
| pHILIC | 963.639 | 464.0602 | 42 | 0.67 |
| pHILIC | 963.639 | 695.4924 | 41.9 | 0.67 |
| pHILIC | 963.639 | 114.7963 | 41.9 | 0.67 |
| pHILIC | 963.639 | 347.7328 | 41.9 | 0.67 |
| pHILIC | 963.639 | 629.8245 | 41.9 | 0.67 |
| pHILIC | 963.639 | 935.2234 | 41.9 | 0.67 |
| pHILIC | 963.639 | 732.8713 | 41.9 | 0.67 |
| pHILIC | 963.639 | 325.0502 | 41.9 | 0.67 |
| pHILIC | 963.639 | 364.0247 | 41.9 | 0.67 |
| pHILIC | 963.639 | 103.8035 | 41.9 | 0.67 |
| pHILIC | 963.639 | 175.0807 | 41.8 | 0.67 |
| pHILIC | 963.639 | 304.6754 | 41.8 | 0.67 |
| pHILIC | 963.639 | 605.7652 | 41.8 | 0.67 |
| pHILIC | 963.639 | 820.4369 | 41.8 | 0.67 |

| | | | | |
|---|---|---|---|---|
| pHILIC | 963.639 | 115.9804 | 41.8 | 0.67 |
| pHILIC | 963.639 | 152.9123 | 41.8 | 0.67 |
| pHILIC | 963.639 | 157.3575 | 41.8 | 0.67 |
| pHILIC | 963.639 | 985.2476 | 41.8 | 0.67 |
| pHILIC | 963.639 | 104.3283 | 41.8 | 0.67 |
| pHILIC | 963.639 | 352.8636 | 41.8 | 0.67 |
| pHILIC | 963.639 | 275.9902 | 41.8 | 0.67 |
| pHILIC | 963.639 | 747.4224 | 41.8 | 0.67 |
| pHILIC | 963.639 | 116.9973 | 41.7 | 0.67 |
| pHILIC | 963.639 | 180.577 | 41.7 | 0.67 |
| pHILIC | 963.639 | 284.9911 | 41.7 | 0.67 |
| pHILIC | 963.639 | 257.26 | 41.7 | 0.67 |
| pHILIC | 963.639 | 171.531 | 41.7 | 0.67 |
| pHILIC | 963.639 | 185.142 | 41.7 | 0.67 |
| pHILIC | 963.639 | 189.4552 | 41.7 | 0.67 |
| pHILIC | 963.639 | 123.753 | 41.6 | 0.66 |
| pHILIC | 963.639 | 974.6785 | 41.6 | 0.66 |
| pHILIC | 963.639 | 553.827 | 41.6 | 0.66 |
| pHILIC | 963.639 | 665.046 | 41.6 | 0.66 |
| pHILIC | 963.639 | 156.6017 | 41.5 | 0.66 |
| pHILIC | 963.639 | 582.4342 | 41.5 | 0.66 |
| pHILIC | 963.639 | 577.7651 | 41.5 | 0.66 |
| pHILIC | 963.639 | 146.9282 | 41.5 | 0.66 |
| pHILIC | 963.639 | 152.841 | 41.5 | 0.66 |
| pHILIC | 963.639 | 641.4273 | 41.5 | 0.66 |
| pHILIC | 963.639 | 167.2674 | 41.5 | 0.66 |
| pHILIC | 963.639 | 780.5708 | 41.5 | 0.66 |
| pHILIC | 963.639 | 168.7003 | 41.5 | 0.66 |
| pHILIC | 963.639 | 881.7432 | 41.5 | 0.66 |
| pHILIC | 963.639 | 102.7511 | 41.5 | 0.66 |
| pHILIC | 963.639 | 107.3168 | 41.5 | 0.66 |
| pHILIC | 963.639 | 466.2551 | 41.4 | 0.66 |
| pHILIC | 963.639 | 206.9905 | 41.4 | 0.66 |
| pHILIC | 963.639 | 114.3102 | 41.4 | 0.66 |
| pHILIC | 963.639 | 107.6035 | 41.4 | 0.66 |
| pHILIC | 963.639 | 608.8301 | 41.4 | 0.66 |
| pHILIC | 963.639 | 243.1386 | 41.4 | 0.66 |
| pHILIC | 963.639 | 120.0514 | 41.4 | 0.66 |
| pHILIC | 963.639 | 393.7332 | 41.4 | 0.66 |
| pHILIC | 963.639 | 179.076 | 41.4 | 0.66 |
| pHILIC | 963.639 | 299.8404 | 41.3 | 0.66 |
| pHILIC | 963.639 | 368.5847 | 41.3 | 0.66 |

| | | | | |
|---|---|---|---|---|
| pHILIC | 963.639 | 435.472 | 41.3 | 0.66 |
| pHILIC | 963.639 | 649.0432 | 41.3 | 0.66 |
| pHILIC | 963.639 | 163.0374 | 41.3 | 0.66 |
| pHILIC | 963.639 | 129.5702 | 41.3 | 0.66 |
| pHILIC | 963.639 | 538.8272 | 41.2 | 0.66 |
| pHILIC | 963.639 | 155.609 | 41.2 | 0.66 |
| pHILIC | 963.639 | 144.424 | 41.2 | 0.66 |
| pHILIC | 963.639 | 121.3689 | 41.2 | 0.66 |
| pHILIC | 963.639 | 255.3886 | 41.2 | 0.66 |
| pHILIC | 963.639 | 230.0491 | 41.2 | 0.66 |
| pHILIC | 963.639 | 140.4966 | 41.2 | 0.66 |
| pHILIC | 963.639 | 152.941 | 41.2 | 0.66 |
| pHILIC | 963.639 | 168.2496 | 41.2 | 0.66 |
| pHILIC | 963.639 | 133.2047 | 41.2 | 0.66 |
| pHILIC | 963.639 | 141.1067 | 41.2 | 0.66 |
| pHILIC | 963.639 | 852.4414 | 41.2 | 0.66 |
| pHILIC | 963.639 | 175.5045 | 41.2 | 0.66 |
| pHILIC | 963.639 | 196.3741 | 41.2 | 0.66 |
| pHILIC | 963.639 | 163.5245 | 41.2 | 0.66 |
| pHILIC | 963.639 | 223.8105 | 41.2 | 0.66 |
| pHILIC | 963.639 | 463.5036 | 41.1 | 0.66 |
| pHILIC | 963.639 | 203.7425 | 41.1 | 0.66 |
| pHILIC | 963.639 | 913.9542 | 41.1 | 0.66 |
| pHILIC | 963.639 | 188.2141 | 41.1 | 0.66 |
| pHILIC | 963.639 | 326.9348 | 41.1 | 0.66 |
| pHILIC | 963.639 | 557.902 | 41.1 | 0.66 |
| pHILIC | 963.639 | 192.9927 | 41.1 | 0.66 |
| pHILIC | 963.639 | 285.999 | 41.1 | 0.66 |
| pHILIC | 963.639 | 294.6916 | 41.1 | 0.66 |
| pHILIC | 963.639 | 178.4385 | 41.1 | 0.66 |
| pHILIC | 963.639 | 337.1762 | 41.1 | 0.66 |
| pHILIC | 963.639 | 205.3229 | 41.1 | 0.66 |
| pHILIC | 963.639 | 107.4061 | 41.1 | 0.66 |
| pHILIC | 963.639 | 283.437 | 41.1 | 0.66 |
| pHILIC | 963.639 | 482.9567 | 41 | 0.66 |
| pHILIC | 963.639 | 643.0606 | 41 | 0.66 |
| pHILIC | 963.639 | 394.5351 | 41 | 0.66 |
| pHILIC | 963.639 | 483.3002 | 41 | 0.66 |
| pHILIC | 963.639 | 107.663 | 41 | 0.65 |
| pHILIC | 963.639 | 132.139 | 41 | 0.65 |
| pHILIC | 963.639 | 327.4604 | 41 | 0.65 |
| pHILIC | 963.639 | 127.4099 | 41 | 0.65 |

| | | | | |
|---|---|---|---|---|
| pHILIC | 963.639 | 325.48 | 41 | 0.65 |
| pHILIC | 963.639 | 951.748 | 41 | 0.65 |
| pHILIC | 963.639 | 209.8832 | 40.9 | 0.65 |
| pHILIC | 963.639 | 396.4881 | 40.9 | 0.65 |
| pHILIC | 963.639 | 868.6274 | 40.9 | 0.65 |
| pHILIC | 963.639 | 232.8468 | 40.9 | 0.65 |
| pHILIC | 963.639 | 165.2046 | 40.9 | 0.65 |
| pHILIC | 963.639 | 120.7056 | 40.9 | 0.65 |
| pHILIC | 963.639 | 133.8403 | 40.9 | 0.65 |
| pHILIC | 963.639 | 542.2824 | 40.9 | 0.65 |
| pHILIC | 963.639 | 958.2456 | 40.9 | 0.65 |
| pHILIC | 963.639 | 126.8189 | 40.9 | 0.65 |
| pHILIC | 963.639 | 105.4364 | 40.9 | 0.65 |
| pHILIC | 963.639 | 166.9049 | 40.9 | 0.65 |
| pHILIC | 963.639 | 373.8438 | 40.9 | 0.65 |
| pHILIC | 963.639 | 675.8566 | 40.9 | 0.65 |
| pHILIC | 963.639 | 759.5082 | 40.9 | 0.65 |
| pHILIC | 963.639 | 119.0039 | 40.9 | 0.65 |
| pHILIC | 963.639 | 192.7468 | 40.9 | 0.65 |
| pHILIC | 963.639 | 277.9525 | 40.9 | 0.65 |
| pHILIC | 963.639 | 171.3103 | 40.9 | 0.65 |
| pHILIC | 963.639 | 492.6279 | 40.8 | 0.65 |
| pHILIC | 963.639 | 723.4126 | 40.8 | 0.65 |
| pHILIC | 963.639 | 246.8839 | 40.8 | 0.65 |
| pHILIC | 963.639 | 895.2214 | 40.8 | 0.65 |
| pHILIC | 963.639 | 158.5622 | 40.8 | 0.65 |
| pHILIC | 963.639 | 522.9706 | 40.8 | 0.65 |
| pHILIC | 963.639 | 130.972 | 40.8 | 0.65 |
| pHILIC | 963.639 | 327.5627 | 40.8 | 0.65 |
| pHILIC | 963.639 | 230.1122 | 40.8 | 0.65 |
| pHILIC | 963.639 | 491.5434 | 40.8 | 0.65 |
| pHILIC | 963.639 | 900.2606 | 40.7 | 0.65 |
| pHILIC | 963.639 | 317.0587 | 40.7 | 0.65 |
| pHILIC | 963.639 | 865.7761 | 40.7 | 0.65 |
| pHILIC | 963.639 | 157.1698 | 40.7 | 0.65 |
| pHILIC | 963.639 | 546.5243 | 40.7 | 0.65 |
| pHILIC | 963.639 | 342.3896 | 40.7 | 0.65 |
| pHILIC | 963.639 | 833.0365 | 40.7 | 0.65 |
| pHILIC | 963.639 | 565.5454 | 40.7 | 0.65 |
| pHILIC | 963.639 | 360.7685 | 40.7 | 0.65 |
| pHILIC | 963.639 | 256.3737 | 40.7 | 0.65 |
| pHILIC | 963.639 | 270.079 | 40.7 | 0.65 |

| | | | | |
|---|---|---|---|---|
| pHILIC | 963.639 | 922.3517 | 40.7 | 0.65 |
| pHILIC | 963.639 | 800.0987 | 40.7 | 0.65 |
| pHILIC | 963.639 | 155.0509 | 40.7 | 0.65 |
| pHILIC | 963.639 | 173.6897 | 40.7 | 0.65 |
| pHILIC | 963.639 | 390.0744 | 40.7 | 0.65 |
| pHILIC | 963.639 | 352.9453 | 40.7 | 0.65 |
| pHILIC | 963.639 | 637.382 | 40.7 | 0.65 |
| pHILIC | 963.639 | 156.0453 | 40.6 | 0.65 |
| pHILIC | 963.639 | 846.9391 | 40.6 | 0.65 |
| pHILIC | 963.639 | 197.4057 | 40.6 | 0.65 |
| pHILIC | 963.639 | 142.3572 | 40.6 | 0.65 |
| pHILIC | 963.639 | 475.2261 | 40.6 | 0.65 |
| pHILIC | 963.639 | 441.9141 | 40.6 | 0.65 |
| pHILIC | 963.639 | 158.3492 | 40.6 | 0.65 |
| pHILIC | 963.639 | 644.3065 | 40.6 | 0.65 |
| pHILIC | 963.639 | 172.9458 | 40.6 | 0.65 |
| pHILIC | 963.639 | 106.5435 | 40.6 | 0.65 |
| pHILIC | 963.639 | 132.2242 | 40.6 | 0.65 |
| pHILIC | 963.639 | 292.5665 | 40.6 | 0.65 |
| pHILIC | 963.639 | 351.5988 | 40.5 | 0.65 |
| pHILIC | 963.639 | 371.9846 | 40.5 | 0.65 |
| pHILIC | 963.639 | 323.4625 | 40.5 | 0.65 |
| pHILIC | 963.639 | 986.6681 | 40.5 | 0.65 |
| pHILIC | 963.639 | 363.8799 | 40.5 | 0.65 |
| pHILIC | 963.639 | 420.7251 | 40.5 | 0.65 |
| pHILIC | 963.639 | 129.4433 | 40.5 | 0.65 |
| pHILIC | 963.639 | 909.6699 | 40.5 | 0.65 |
| pHILIC | 963.639 | 632.4172 | 40.4 | 0.65 |
| pHILIC | 963.639 | 716.5042 | 40.4 | 0.65 |
| pHILIC | 963.639 | 164.8633 | 40.4 | 0.65 |
| pHILIC | 963.639 | 453.8723 | 40.4 | 0.65 |
| pHILIC | 963.639 | 182.4408 | 40.4 | 0.65 |
| pHILIC | 963.639 | 298.8133 | 40.4 | 0.64 |
| pHILIC | 963.639 | 275.5622 | 40.4 | 0.64 |
| pHILIC | 963.639 | 325.1319 | 40.3 | 0.64 |
| pHILIC | 963.639 | 287.9897 | 40.3 | 0.64 |
| pHILIC | 963.639 | 818.0587 | 40.3 | 0.64 |
| pHILIC | 963.639 | 600.8885 | 40.3 | 0.64 |
| pHILIC | 963.639 | 699.1294 | 40.3 | 0.64 |
| pHILIC | 963.639 | 120.4218 | 40.3 | 0.64 |
| pHILIC | 963.639 | 108.8545 | 40.3 | 0.64 |
| pHILIC | 963.639 | 265.4659 | 40.3 | 0.64 |

| | | | | |
|---|---|---|---|---|
| pHILIC | 963.639 | 300.6992 | 40.3 | 0.64 |
| pHILIC | 963.639 | 502.2136 | 40.3 | 0.64 |
| pHILIC | 963.639 | 143.3422 | 40.3 | 0.64 |
| pHILIC | 963.639 | 479.0511 | 40.3 | 0.64 |
| pHILIC | 963.639 | 136.3111 | 40.3 | 0.64 |
| pHILIC | 963.639 | 872.7513 | 40.3 | 0.64 |
| pHILIC | 963.639 | 175.6725 | 40.3 | 0.64 |
| pHILIC | 963.639 | 120.1783 | 40.3 | 0.64 |
| pHILIC | 963.639 | 173.3506 | 40.3 | 0.64 |
| pHILIC | 963.639 | 555.7628 | 40.2 | 0.64 |
| pHILIC | 963.639 | 372.8311 | 40.2 | 0.64 |
| pHILIC | 963.639 | 227.3317 | 40.2 | 0.64 |
| pHILIC | 963.639 | 187.1844 | 40.2 | 0.64 |
| pHILIC | 963.639 | 469.2752 | 40.2 | 0.64 |
| pHILIC | 963.639 | 159.9812 | 40.2 | 0.64 |
| pHILIC | 963.639 | 127.0929 | 40.2 | 0.64 |
| pHILIC | 963.639 | 154.8719 | 40.2 | 0.64 |
| pHILIC | 963.639 | 850.3735 | 40.2 | 0.64 |
| pHILIC | 963.639 | 138.0717 | 40.2 | 0.64 |
| pHILIC | 963.639 | 128.3272 | 40.1 | 0.64 |
| pHILIC | 963.639 | 384.214 | 40.1 | 0.64 |
| pHILIC | 963.639 | 148.209 | 40.1 | 0.64 |
| pHILIC | 963.639 | 123.7008 | 40.1 | 0.64 |
| pHILIC | 963.639 | 107.0611 | 40.1 | 0.64 |
| pHILIC | 963.639 | 351.4297 | 40.1 | 0.64 |
| pHILIC | 963.639 | 142.2857 | 40.1 | 0.64 |
| pHILIC | 963.639 | 274.0204 | 40.1 | 0.64 |
| pHILIC | 963.639 | 422.4203 | 40.1 | 0.64 |
| pHILIC | 963.639 | 419.66 | 40.1 | 0.64 |
| pHILIC | 963.639 | 103.3203 | 40.1 | 0.64 |
| pHILIC | 963.639 | 175.5882 | 40.1 | 0.64 |
| pHILIC | 963.639 | 571.2682 | 40.1 | 0.64 |
| pHILIC | 963.639 | 230.4198 | 40.1 | 0.64 |
| pHILIC | 963.639 | 304.7866 | 40.1 | 0.64 |
| pHILIC | 963.639 | 319.3382 | 40 | 0.64 |
| pHILIC | 963.639 | 104.6649 | 40 | 0.64 |
| pHILIC | 963.639 | 180.6528 | 40 | 0.64 |
| pHILIC | 963.639 | 277.075 | 40 | 0.64 |
| pHILIC | 963.639 | 289.9932 | 40 | 0.64 |
| pHILIC | 963.639 | 223.9891 | 40 | 0.64 |
| pHILIC | 963.639 | 197.6757 | 40 | 0.64 |
| pHILIC | 963.639 | 165.7854 | 40 | 0.64 |

| | | | | |
|---|---|---|---|---|
| pHILIC | 963.639 | 117.7488 | 40 | 0.64 |
| pHILIC | 963.639 | 225.9092 | 40 | 0.64 |
| pHILIC | 963.639 | 147.3271 | 40 | 0.64 |
| pHILIC | 963.639 | 148.9734 | 40 | 0.64 |
| pHILIC | 963.639 | 196.2647 | 40 | 0.64 |
| pHILIC | 963.639 | 227.9074 | 40 | 0.64 |
| pHILIC | 963.639 | 208.3745 | 40 | 0.64 |
| pHILIC | 963.639 | 591.7004 | 40 | 0.64 |
| pHILIC | 963.639 | 682.8769 | 40 | 0.64 |
| pHILIC | 963.639 | 721.9633 | 40 | 0.64 |
| pHILIC | 963.639 | 458.9662 | 39.9 | 0.64 |
| pHILIC | 963.639 | 181.278 | 39.9 | 0.64 |
| pHILIC | 963.639 | 275.6347 | 39.9 | 0.64 |
| pHILIC | 963.639 | 119.3434 | 39.9 | 0.64 |
| pHILIC | 963.639 | 188.7698 | 39.9 | 0.64 |
| pHILIC | 963.639 | 232.1669 | 39.9 | 0.64 |
| pHILIC | 963.639 | 744.8874 | 39.9 | 0.64 |
| pHILIC | 963.639 | 420.5917 | 39.9 | 0.64 |
| pHILIC | 963.639 | 103.7598 | 39.9 | 0.64 |
| pHILIC | 963.639 | 110.0023 | 39.9 | 0.64 |
| pHILIC | 963.639 | 125.4157 | 39.9 | 0.64 |
| pHILIC | 963.639 | 312.5383 | 39.9 | 0.64 |
| pHILIC | 963.639 | 132.8421 | 39.8 | 0.64 |
| pHILIC | 963.639 | 124.5991 | 39.8 | 0.64 |
| pHILIC | 963.639 | 494.8131 | 39.8 | 0.64 |
| pHILIC | 963.639 | 227.5378 | 39.8 | 0.64 |
| pHILIC | 963.639 | 509.2751 | 39.8 | 0.64 |
| pHILIC | 963.639 | 121.0376 | 39.8 | 0.64 |
| pHILIC | 963.639 | 871.6126 | 39.8 | 0.64 |
| pHILIC | 963.639 | 311.0493 | 39.8 | 0.64 |
| pHILIC | 963.639 | 655.5551 | 39.8 | 0.64 |
| pHILIC | 963.639 | 589.2581 | 39.8 | 0.64 |
| pHILIC | 963.639 | 205.2982 | 39.8 | 0.64 |
| pHILIC | 963.639 | 346.4372 | 39.8 | 0.64 |
| pHILIC | 963.639 | 519.3726 | 39.8 | 0.64 |
| pHILIC | 963.639 | 204.792 | 39.8 | 0.64 |
| pHILIC | 963.639 | 434.122 | 39.8 | 0.64 |
| pHILIC | 963.639 | 350.4597 | 39.8 | 0.64 |
| pHILIC | 963.639 | 566.2544 | 39.7 | 0.64 |
| pHILIC | 963.639 | 194.4095 | 39.7 | 0.64 |
| pHILIC | 963.639 | 297.9876 | 39.7 | 0.63 |
| pHILIC | 963.639 | 164.142 | 39.7 | 0.63 |

| | | | | |
|---|---|---|---|---|
| pHILIC | 963.639 | 686.1923 | 39.7 | 0.63 |
| pHILIC | 963.639 | 323.9983 | 39.7 | 0.63 |
| pHILIC | 963.639 | 336.976 | 39.7 | 0.63 |
| pHILIC | 963.639 | 273.3763 | 39.7 | 0.63 |
| pHILIC | 963.639 | 249.7311 | 39.7 | 0.63 |
| pHILIC | 963.639 | 217.9537 | 39.7 | 0.63 |
| pHILIC | 963.639 | 133.0167 | 39.7 | 0.63 |
| pHILIC | 963.639 | 134.9534 | 39.7 | 0.63 |
| pHILIC | 963.639 | 398.0635 | 39.7 | 0.63 |
| pHILIC | 963.639 | 346.3674 | 39.7 | 0.63 |
| pHILIC | 963.639 | 106.4731 | 39.7 | 0.63 |
| pHILIC | 963.639 | 140.6853 | 39.6 | 0.63 |
| pHILIC | 963.639 | 546.9824 | 39.6 | 0.63 |
| pHILIC | 963.639 | 549.1462 | 39.6 | 0.63 |
| pHILIC | 963.639 | 127.7802 | 39.6 | 0.63 |
| pHILIC | 963.639 | 217.694 | 39.6 | 0.63 |
| pHILIC | 963.639 | 424.0709 | 39.6 | 0.63 |
| pHILIC | 963.639 | 307.049 | 39.6 | 0.63 |
| pHILIC | 963.639 | 153.2306 | 39.6 | 0.63 |
| pHILIC | 963.639 | 494.7069 | 39.6 | 0.63 |
| pHILIC | 963.639 | 189.3401 | 39.6 | 0.63 |
| pHILIC | 963.639 | 188.192 | 39.6 | 0.63 |
| pHILIC | 963.639 | 140.8302 | 39.6 | 0.63 |
| pHILIC | 963.639 | 716.6896 | 39.5 | 0.63 |
| pHILIC | 963.639 | 141.2737 | 39.5 | 0.63 |
| pHILIC | 963.639 | 946.7278 | 39.5 | 0.63 |
| pHILIC | 963.639 | 324.7458 | 39.5 | 0.63 |
| pHILIC | 963.639 | 137.6966 | 39.5 | 0.63 |
| pHILIC | 963.639 | 121.8515 | 39.5 | 0.63 |
| pHILIC | 963.639 | 859.4754 | 39.5 | 0.63 |
| pHILIC | 963.639 | 383.2212 | 39.5 | 0.63 |
| pHILIC | 963.639 | 414.9264 | 39.5 | 0.63 |
| pHILIC | 963.639 | 129.6421 | 39.5 | 0.63 |
| pHILIC | 963.639 | 500.4951 | 39.5 | 0.63 |
| pHILIC | 963.639 | 836.3904 | 39.5 | 0.63 |
| pHILIC | 963.639 | 236.6261 | 39.5 | 0.63 |
| pHILIC | 963.639 | 157.8455 | 39.5 | 0.63 |
| pHILIC | 963.639 | 579.4714 | 39.4 | 0.63 |
| pHILIC | 963.639 | 640.7012 | 39.4 | 0.63 |
| pHILIC | 963.639 | 101.01 | 39.4 | 0.63 |
| pHILIC | 963.639 | 306.8293 | 39.4 | 0.63 |
| pHILIC | 963.639 | 235.8102 | 39.4 | 0.63 |

| | | | | |
|---|---|---|---|---|
| pHILIC | 963.639 | 237.7648 | 39.4 | 0.63 |
| pHILIC | 963.639 | 164.7283 | 39.4 | 0.63 |
| pHILIC | 963.639 | 100.6814 | 39.4 | 0.63 |
| pHILIC | 963.639 | 375.7419 | 39.4 | 0.63 |
| pHILIC | 963.639 | 205.3522 | 39.4 | 0.63 |
| pHILIC | 963.639 | 277.5713 | 39.4 | 0.63 |
| pHILIC | 963.639 | 157.9866 | 39.4 | 0.63 |
| pHILIC | 963.639 | 807.9537 | 39.4 | 0.63 |
| pHILIC | 963.639 | 224.1474 | 39.4 | 0.63 |
| pHILIC | 963.639 | 941.3588 | 39.4 | 0.63 |
| pHILIC | 963.639 | 675.587 | 39.4 | 0.63 |
| pHILIC | 963.639 | 751.1167 | 39.4 | 0.63 |
| pHILIC | 963.639 | 729.108 | 39.4 | 0.63 |
| pHILIC | 963.639 | 759.7875 | 39.3 | 0.63 |
| pHILIC | 963.639 | 402.9913 | 39.3 | 0.63 |
| pHILIC | 963.639 | 252.2181 | 39.3 | 0.63 |
| pHILIC | 963.639 | 120.9399 | 39.3 | 0.63 |
| pHILIC | 963.639 | 101.115 | 39.3 | 0.63 |
| pHILIC | 963.639 | 147.8663 | 39.3 | 0.63 |
| pHILIC | 963.639 | 364.2441 | 39.3 | 0.63 |
| pHILIC | 963.639 | 258.57 | 39.3 | 0.63 |
| pHILIC | 963.639 | 293.8264 | 39.3 | 0.63 |
| pHILIC | 963.639 | 261.9428 | 39.3 | 0.63 |
| pHILIC | 963.639 | 361.2878 | 39.3 | 0.63 |
| pHILIC | 963.639 | 222.7612 | 39.3 | 0.63 |
| pHILIC | 963.639 | 652.4203 | 39.3 | 0.63 |
| pHILIC | 963.639 | 745.4884 | 39.3 | 0.63 |
| pHILIC | 963.639 | 131.3381 | 39.3 | 0.63 |
| pHILIC | 963.639 | 375.0417 | 39.3 | 0.63 |
| pHILIC | 963.639 | 885.6391 | 39.3 | 0.63 |
| pHILIC | 963.639 | 686.4747 | 39.3 | 0.63 |
| pHILIC | 963.639 | 217.6428 | 39.3 | 0.63 |
| pHILIC | 963.639 | 415.2375 | 39.2 | 0.63 |
| pHILIC | 963.639 | 218.896 | 39.2 | 0.63 |
| pHILIC | 963.639 | 260.6403 | 39.2 | 0.63 |
| pHILIC | 963.639 | 419.2688 | 39.2 | 0.63 |
| pHILIC | 963.639 | 406.4129 | 39.2 | 0.63 |
| pHILIC | 963.639 | 896.3489 | 39.2 | 0.63 |
| pHILIC | 963.639 | 234.649 | 39.2 | 0.63 |
| pHILIC | 963.639 | 106.8498 | 39.2 | 0.63 |
| pHILIC | 963.639 | 242.0671 | 39.2 | 0.63 |
| pHILIC | 963.639 | 161.9402 | 39.2 | 0.63 |

| | | | | |
|---|---|---|---|---|
| pHILIC | 963.639 | 210.2759 | 39.2 | 0.63 |
| pHILIC | 963.639 | 446.6743 | 39.2 | 0.63 |
| pHILIC | 963.639 | 969.8101 | 39.2 | 0.63 |
| pHILIC | 963.639 | 196.9429 | 39.2 | 0.63 |
| pHILIC | 963.639 | 174.1501 | 39.2 | 0.63 |
| pHILIC | 963.639 | 130.7284 | 39.2 | 0.63 |
| pHILIC | 963.639 | 149.3856 | 39.2 | 0.63 |
| pHILIC | 963.639 | 201.1218 | 39.2 | 0.63 |
| pHILIC | 963.639 | 644.7291 | 39.2 | 0.63 |
| pHILIC | 963.639 | 227.3557 | 39.2 | 0.63 |
| pHILIC | 963.639 | 101.6876 | 39.1 | 0.63 |
| pHILIC | 963.639 | 142.7942 | 39.1 | 0.63 |
| pHILIC | 963.639 | 163.4288 | 39.1 | 0.63 |
| pHILIC | 963.639 | 104.7062 | 39.1 | 0.63 |
| pHILIC | 963.639 | 140.1062 | 39.1 | 0.63 |
| pHILIC | 963.639 | 155.0306 | 39.1 | 0.63 |
| pHILIC | 963.639 | 153.953 | 39.1 | 0.63 |
| pHILIC | 963.639 | 662.2777 | 39.1 | 0.63 |
| pHILIC | 963.639 | 292.2601 | 39.1 | 0.63 |
| pHILIC | 963.639 | 128.7681 | 39.1 | 0.63 |
| pHILIC | 963.639 | 892.576 | 39.1 | 0.63 |
| pHILIC | 963.639 | 538.1652 | 39.1 | 0.63 |
| pHILIC | 963.639 | 314.6833 | 39.1 | 0.63 |
| pHILIC | 963.639 | 641.1284 | 39.1 | 0.62 |
| pHILIC | 963.639 | 552.6155 | 39.1 | 0.62 |
| pHILIC | 963.639 | 146.9684 | 39.1 | 0.62 |
| pHILIC | 963.639 | 111.0588 | 39.1 | 0.62 |
| pHILIC | 963.639 | 100.6514 | 39.1 | 0.62 |
| pHILIC | 963.639 | 314.2857 | 39.1 | 0.62 |
| pHILIC | 963.639 | 217.7967 | 39.1 | 0.62 |
| pHILIC | 963.639 | 273.07 | 39.1 | 0.62 |
| pHILIC | 963.639 | 224.1696 | 39 | 0.62 |
| pHILIC | 963.639 | 391.4989 | 39 | 0.62 |
| pHILIC | 963.639 | 294.6082 | 39 | 0.62 |
| pHILIC | 963.639 | 173.7037 | 39 | 0.62 |
| pHILIC | 963.639 | 411.1577 | 39 | 0.62 |
| pHILIC | 963.639 | 924.5038 | 39 | 0.62 |
| pHILIC | 963.639 | 150.0614 | 39 | 0.62 |
| pHILIC | 963.639 | 389.8234 | 39 | 0.62 |
| pHILIC | 963.639 | 136.3986 | 39 | 0.62 |
| pHILIC | 963.639 | 309.1111 | 39 | 0.62 |
| pHILIC | 963.639 | 592.6709 | 39 | 0.62 |

| | | | | |
|---|---|---|---|---|
| pHILIC | 963.639 | 168.1539 | 39 | 0.62 |
| pHILIC | 963.639 | 433.7858 | 39 | 0.62 |
| pHILIC | 963.639 | 118.5488 | 39 | 0.62 |
| pHILIC | 963.639 | 639.0518 | 39 | 0.62 |
| pHILIC | 963.639 | 396.6468 | 38.9 | 0.62 |
| pHILIC | 963.639 | 104.5196 | 38.9 | 0.62 |
| pHILIC | 963.639 | 653.4246 | 38.9 | 0.62 |
| pHILIC | 963.639 | 914.5612 | 38.9 | 0.62 |
| pHILIC | 963.639 | 749.9573 | 38.9 | 0.62 |
| pHILIC | 963.639 | 485.9534 | 38.9 | 0.62 |
| pHILIC | 963.639 | 175.401 | 38.9 | 0.62 |
| pHILIC | 963.639 | 905.4568 | 38.9 | 0.62 |
| pHILIC | 963.639 | 160.5515 | 38.9 | 0.62 |
| pHILIC | 963.639 | 574.3307 | 38.9 | 0.62 |
| pHILIC | 963.639 | 429.2164 | 38.9 | 0.62 |
| pHILIC | 963.639 | 149.0689 | 38.9 | 0.62 |
| pHILIC | 963.639 | 148.7894 | 38.9 | 0.62 |
| pHILIC | 963.639 | 269.982 | 38.9 | 0.62 |
| pHILIC | 963.639 | 213.3022 | 38.9 | 0.62 |
| pHILIC | 963.639 | 280.2401 | 38.9 | 0.62 |
| pHILIC | 963.639 | 381.4964 | 38.9 | 0.62 |
| pHILIC | 963.639 | 231.354 | 38.9 | 0.62 |
| pHILIC | 963.639 | 439.1958 | 38.8 | 0.62 |
| pHILIC | 963.639 | 399.6826 | 38.8 | 0.62 |
| pHILIC | 963.639 | 103.7424 | 38.8 | 0.62 |
| pHILIC | 963.639 | 286.9585 | 38.8 | 0.62 |
| pHILIC | 963.639 | 291.4407 | 38.8 | 0.62 |
| pHILIC | 963.639 | 369.8305 | 38.8 | 0.62 |
| pHILIC | 963.639 | 101.1987 | 38.8 | 0.62 |
| pHILIC | 963.639 | 949.0026 | 38.8 | 0.62 |
| pHILIC | 963.639 | 585.36 | 38.8 | 0.62 |
| pHILIC | 963.639 | 136.9487 | 38.8 | 0.62 |
| pHILIC | 963.639 | 975.1567 | 38.8 | 0.62 |
| pHILIC | 963.639 | 114.0755 | 38.7 | 0.62 |
| pHILIC | 963.639 | 118.369 | 38.7 | 0.62 |
| pHILIC | 963.639 | 565.1436 | 38.7 | 0.62 |
| pHILIC | 963.639 | 696.2505 | 38.7 | 0.62 |
| pHILIC | 963.639 | 178.1006 | 38.7 | 0.62 |
| pHILIC | 963.639 | 157.3036 | 38.7 | 0.62 |
| pHILIC | 963.639 | 153.9873 | 38.7 | 0.62 |
| pHILIC | 963.639 | 329.1707 | 38.7 | 0.62 |
| pHILIC | 963.639 | 106.4954 | 38.7 | 0.62 |

| | | | | |
|---|---|---|---|---|
| pHILIC | 963.639 | 177.7643 | 38.7 | 0.62 |
| pHILIC | 963.639 | 121.5067 | 38.7 | 0.62 |
| pHILIC | 963.639 | 185.697 | 38.7 | 0.62 |
| pHILIC | 963.639 | 434.7434 | 38.7 | 0.62 |
| pHILIC | 963.639 | 151.3316 | 38.7 | 0.62 |
| pHILIC | 963.639 | 288.6999 | 38.7 | 0.62 |
| pHILIC | 963.639 | 234.2453 | 38.7 | 0.62 |
| pHILIC | 963.639 | 240.2993 | 38.7 | 0.62 |
| pHILIC | 963.639 | 343.731 | 38.7 | 0.62 |
| pHILIC | 963.639 | 135.7588 | 38.7 | 0.62 |
| pHILIC | 963.639 | 301.8752 | 38.7 | 0.62 |
| pHILIC | 963.639 | 102.5852 | 38.7 | 0.62 |
| pHILIC | 963.639 | 118.5853 | 38.7 | 0.62 |
| pHILIC | 963.639 | 737.7146 | 38.6 | 0.62 |
| pHILIC | 963.639 | 141.0747 | 38.6 | 0.62 |
| pHILIC | 963.639 | 293.8689 | 38.6 | 0.62 |
| pHILIC | 963.639 | 104.1204 | 38.6 | 0.62 |
| pHILIC | 963.639 | 171.6217 | 38.6 | 0.62 |
| pHILIC | 963.639 | 216.7345 | 38.6 | 0.62 |
| pHILIC | 963.639 | 123.9722 | 38.6 | 0.62 |
| pHILIC | 963.639 | 194.3116 | 38.6 | 0.62 |
| pHILIC | 963.639 | 209.6544 | 38.6 | 0.62 |
| pHILIC | 963.639 | 825.5745 | 38.6 | 0.62 |
| pHILIC | 963.639 | 176.5505 | 38.6 | 0.62 |
| pHILIC | 963.639 | 148.9273 | 38.6 | 0.62 |
| pHILIC | 963.639 | 372.2226 | 38.6 | 0.62 |
| pHILIC | 963.639 | 274.4889 | 38.6 | 0.62 |
| pHILIC | 963.639 | 121.8808 | 38.6 | 0.62 |
| pHILIC | 963.639 | 260.031 | 38.6 | 0.62 |
| pHILIC | 963.639 | 404.4128 | 38.6 | 0.62 |
| pHILIC | 963.639 | 398.6394 | 38.6 | 0.62 |
| pHILIC | 963.639 | 437.0686 | 38.6 | 0.62 |
| pHILIC | 963.639 | 432.4955 | 38.6 | 0.62 |
| pHILIC | 963.639 | 166.1743 | 38.6 | 0.62 |
| pHILIC | 963.639 | 106.5568 | 38.6 | 0.62 |
| pHILIC | 963.639 | 138.5017 | 38.6 | 0.62 |
| pHILIC | 963.639 | 774.4625 | 38.6 | 0.62 |
| pHILIC | 963.639 | 294.3211 | 38.6 | 0.62 |
| pHILIC | 963.639 | 333.6743 | 38.6 | 0.62 |
| pHILIC | 963.639 | 450.119 | 38.5 | 0.62 |
| pHILIC | 963.639 | 442.0245 | 38.5 | 0.62 |
| pHILIC | 963.639 | 303.3442 | 38.5 | 0.62 |

| | | | | |
|---|---|---|---|---|
| pHILIC | 963.639 | 149.1764 | 38.5 | 0.62 |
| pHILIC | 963.639 | 376.6335 | 38.5 | 0.62 |
| pHILIC | 963.639 | 453.0258 | 38.5 | 0.62 |
| pHILIC | 963.639 | 129.0837 | 38.5 | 0.62 |
| pHILIC | 963.639 | 490.2498 | 38.5 | 0.62 |
| pHILIC | 963.639 | 105.2066 | 38.5 | 0.61 |
| pHILIC | 963.639 | 177.1641 | 38.5 | 0.61 |
| pHILIC | 963.639 | 125.1265 | 38.5 | 0.61 |
| pHILIC | 963.639 | 102.5109 | 38.5 | 0.61 |
| pHILIC | 963.639 | 525.9838 | 38.5 | 0.61 |
| pHILIC | 963.639 | 679.9974 | 38.4 | 0.61 |
| pHILIC | 963.639 | 139.3996 | 38.4 | 0.61 |
| pHILIC | 963.639 | 403.9254 | 38.4 | 0.61 |
| pHILIC | 963.639 | 309.4329 | 38.4 | 0.61 |
| pHILIC | 963.639 | 410.2972 | 38.4 | 0.61 |
| pHILIC | 963.639 | 106.7737 | 38.4 | 0.61 |
| pHILIC | 963.639 | 214.8587 | 38.4 | 0.61 |
| pHILIC | 963.639 | 268.6814 | 38.4 | 0.61 |
| pHILIC | 963.639 | 327.2427 | 38.4 | 0.61 |
| pHILIC | 963.639 | 997.9973 | 38.4 | 0.61 |
| pHILIC | 963.639 | 906.5637 | 38.4 | 0.61 |
| pHILIC | 963.639 | 422.8685 | 38.4 | 0.61 |
| pHILIC | 963.639 | 100.9401 | 38.4 | 0.61 |
| pHILIC | 963.639 | 242.3134 | 38.4 | 0.61 |
| pHILIC | 963.639 | 188.0205 | 38.4 | 0.61 |
| pHILIC | 963.639 | 228.8615 | 38.4 | 0.61 |
| pHILIC | 963.639 | 330.3918 | 38.4 | 0.61 |
| pHILIC | 963.639 | 303.8446 | 38.4 | 0.61 |
| pHILIC | 963.639 | 102.5509 | 38.3 | 0.61 |
| pHILIC | 963.639 | 120.8754 | 38.3 | 0.61 |
| pHILIC | 963.639 | 159.6489 | 38.3 | 0.61 |
| pHILIC | 963.639 | 114.5198 | 38.3 | 0.61 |
| pHILIC | 963.639 | 345.3887 | 38.3 | 0.61 |
| pHILIC | 963.639 | 111.6835 | 38.3 | 0.61 |
| pHILIC | 963.639 | 999.0531 | 38.3 | 0.61 |
| pHILIC | 963.639 | 168.483 | 38.3 | 0.61 |
| pHILIC | 963.639 | 231.0298 | 38.3 | 0.61 |
| pHILIC | 963.639 | 954.2635 | 38.3 | 0.61 |
| pHILIC | 963.639 | 704.1863 | 38.3 | 0.61 |
| pHILIC | 963.639 | 113.2135 | 38.3 | 0.61 |
| pHILIC | 963.639 | 271.5227 | 38.3 | 0.61 |
| pHILIC | 963.639 | 550.1744 | 38.3 | 0.61 |

| | | | | |
|---|---|---|---|---|
| pHILIC | 963.639 | 121.1974 | 38.3 | 0.61 |
| pHILIC | 963.639 | 720.457 | 38.3 | 0.61 |
| pHILIC | 963.639 | 230.9073 | 38.3 | 0.61 |
| pHILIC | 963.639 | 580.1607 | 38.2 | 0.61 |
| pHILIC | 963.639 | 748.806 | 38.2 | 0.61 |
| pHILIC | 963.639 | 151.8093 | 38.2 | 0.61 |
| pHILIC | 963.639 | 204.3141 | 38.2 | 0.61 |
| pHILIC | 963.639 | 163.8209 | 38.2 | 0.61 |
| pHILIC | 963.639 | 288.2413 | 38.2 | 0.61 |
| pHILIC | 963.639 | 795.0116 | 38.2 | 0.61 |
| pHILIC | 963.639 | 271.3676 | 38.2 | 0.61 |
| pHILIC | 963.639 | 235.8851 | 38.2 | 0.61 |
| pHILIC | 963.639 | 181.9528 | 38.2 | 0.61 |
| pHILIC | 963.639 | 157.804 | 38.2 | 0.61 |
| pHILIC | 963.639 | 944.1479 | 38.2 | 0.61 |
| pHILIC | 963.639 | 394.0544 | 38.2 | 0.61 |
| pHILIC | 963.639 | 102.3625 | 38.2 | 0.61 |
| pHILIC | 963.639 | 152.8904 | 38.2 | 0.61 |
| pHILIC | 963.639 | 151.2709 | 38.2 | 0.61 |
| pHILIC | 963.639 | 167.3018 | 38.2 | 0.61 |
| pHILIC | 963.639 | 871.2061 | 38.2 | 0.61 |
| pHILIC | 963.639 | 102.2808 | 38.2 | 0.61 |
| pHILIC | 963.639 | 358.6553 | 38.2 | 0.61 |
| pHILIC | 963.639 | 158.8358 | 38.2 | 0.61 |
| pHILIC | 963.639 | 310.546 | 38.2 | 0.61 |
| pHILIC | 963.639 | 135.7864 | 38.2 | 0.61 |
| pHILIC | 963.639 | 101.8796 | 38.2 | 0.61 |
| pHILIC | 963.639 | 222.1496 | 38.2 | 0.61 |
| pHILIC | 963.639 | 207.0433 | 38.1 | 0.61 |
| pHILIC | 963.639 | 262.585 | 38.1 | 0.61 |
| pHILIC | 963.639 | 626.0853 | 38.1 | 0.61 |
| pHILIC | 963.639 | 112.0607 | 38.1 | 0.61 |
| pHILIC | 963.639 | 880.6671 | 38.1 | 0.61 |
| pHILIC | 963.639 | 343.9343 | 38.1 | 0.61 |
| pHILIC | 963.639 | 259.5744 | 38.1 | 0.61 |
| pHILIC | 963.639 | 296.6749 | 38.1 | 0.61 |
| pHILIC | 963.639 | 908.2094 | 38.1 | 0.61 |
| pHILIC | 963.639 | 121.1124 | 38.1 | 0.61 |
| pHILIC | 963.639 | 310.3658 | 38.1 | 0.61 |
| pHILIC | 963.639 | 370.8742 | 38.1 | 0.61 |
| pHILIC | 963.639 | 318.3921 | 38.1 | 0.61 |
| pHILIC | 963.639 | 494.9313 | 38.1 | 0.61 |

| | | | | |
|---|---|---|---|---|
| pHILIC | 963.639 | 283.0795 | 38 | 0.61 |
| pHILIC | 963.639 | 224.8223 | 38 | 0.61 |
| pHILIC | 963.639 | 146.6133 | 38 | 0.61 |
| pHILIC | 963.639 | 115.1648 | 38 | 0.61 |
| pHILIC | 963.639 | 363.6293 | 38 | 0.61 |
| pHILIC | 963.639 | 104.8831 | 38 | 0.61 |
| pHILIC | 963.639 | 125.1482 | 38 | 0.61 |
| pHILIC | 963.639 | 321.1168 | 38 | 0.61 |
| pHILIC | 963.639 | 317.2556 | 38 | 0.61 |
| pHILIC | 963.639 | 865.17 | 38 | 0.61 |
| pHILIC | 963.639 | 606.3829 | 38 | 0.61 |
| pHILIC | 963.639 | 443.6724 | 38 | 0.61 |
| pHILIC | 963.639 | 499.5531 | 38 | 0.61 |
| pHILIC | 963.639 | 115.4335 | 38 | 0.61 |
| pHILIC | 963.639 | 624.1255 | 38 | 0.61 |
| pHILIC | 963.639 | 559.7761 | 38 | 0.61 |
| pHILIC | 963.639 | 256.321 | 38 | 0.61 |
| pHILIC | 963.639 | 170.0387 | 38 | 0.61 |
| pHILIC | 963.639 | 178.5465 | 38 | 0.61 |
| pHILIC | 963.639 | 210.1325 | 37.9 | 0.61 |
| pHILIC | 963.639 | 323.0189 | 37.9 | 0.61 |
| pHILIC | 963.639 | 354.0287 | 37.9 | 0.61 |
| pHILIC | 963.639 | 355.5788 | 37.9 | 0.61 |
| pHILIC | 963.639 | 107.3329 | 37.9 | 0.61 |
| pHILIC | 963.639 | 324.9032 | 37.9 | 0.61 |
| pHILIC | 963.639 | 609.5095 | 37.9 | 0.61 |
| pHILIC | 963.639 | 162.9749 | 37.9 | 0.61 |
| pHILIC | 963.639 | 162.6469 | 37.9 | 0.61 |
| pHILIC | 963.639 | 385.6559 | 37.9 | 0.61 |
| pHILIC | 963.639 | 151.0884 | 37.9 | 0.61 |
| pHILIC | 963.639 | 440.1997 | 37.9 | 0.61 |
| pHILIC | 963.639 | 245.8493 | 37.9 | 0.61 |
| pHILIC | 963.639 | 295.464 | 37.9 | 0.61 |
| pHILIC | 963.639 | 253.6551 | 37.9 | 0.61 |
| pHILIC | 963.639 | 456.4001 | 37.9 | 0.61 |
| pHILIC | 963.639 | 228.2392 | 37.9 | 0.61 |
| pHILIC | 963.639 | 133.0426 | 37.9 | 0.61 |
| pHILIC | 963.639 | 140.9962 | 37.9 | 0.61 |
| pHILIC | 963.639 | 557.2123 | 37.9 | 0.61 |
| pHILIC | 963.639 | 356.7549 | 37.9 | 0.6 |
| pHILIC | 963.639 | 120.1441 | 37.9 | 0.6 |
| pHILIC | 963.639 | 144.4037 | 37.9 | 0.6 |

| | | | | |
|---|---|---|---|---|
| pHILIC | 963.639 | 308.0654 | 37.8 | 0.6 |
| pHILIC | 963.639 | 116.2885 | 37.8 | 0.6 |
| pHILIC | 963.639 | 385.5722 | 37.8 | 0.6 |
| pHILIC | 963.639 | 157.7889 | 37.8 | 0.6 |
| pHILIC | 963.639 | 141.1728 | 37.8 | 0.6 |
| pHILIC | 963.639 | 980.299 | 37.8 | 0.6 |
| pHILIC | 963.639 | 139.0119 | 37.8 | 0.6 |
| pHILIC | 963.639 | 486.0573 | 37.8 | 0.6 |
| pHILIC | 963.639 | 423.8026 | 37.8 | 0.6 |
| pHILIC | 963.639 | 736.3178 | 37.8 | 0.6 |
| pHILIC | 963.639 | 166.4199 | 37.8 | 0.6 |
| pHILIC | 963.639 | 372.1554 | 37.8 | 0.6 |
| pHILIC | 963.639 | 435.1756 | 37.8 | 0.6 |
| pHILIC | 963.639 | 320.9792 | 37.8 | 0.6 |
| pHILIC | 963.639 | 151.9072 | 37.8 | 0.6 |
| pHILIC | 963.639 | 367.0706 | 37.8 | 0.6 |
| pHILIC | 963.639 | 197.0998 | 37.7 | 0.6 |
| pHILIC | 963.639 | 158.0001 | 37.7 | 0.6 |
| pHILIC | 963.639 | 208.0561 | 37.7 | 0.6 |
| pHILIC | 963.639 | 393.5868 | 37.7 | 0.6 |
| pHILIC | 963.639 | 833.8926 | 37.7 | 0.6 |
| pHILIC | 963.639 | 404.7572 | 37.7 | 0.6 |
| pHILIC | 963.639 | 116.1818 | 37.7 | 0.6 |
| pHILIC | 963.639 | 725.2762 | 37.7 | 0.6 |
| pHILIC | 963.639 | 101.3863 | 37.7 | 0.6 |
| pHILIC | 963.639 | 859.2312 | 37.7 | 0.6 |
| pHILIC | 963.639 | 391.6416 | 37.7 | 0.6 |
| pHILIC | 963.639 | 228.7746 | 37.7 | 0.6 |
| pHILIC | 963.639 | 735.0008 | 37.7 | 0.6 |
| pHILIC | 963.639 | 117.063 | 37.7 | 0.6 |
| pHILIC | 963.639 | 174.463 | 37.7 | 0.6 |
| pHILIC | 963.639 | 109.1318 | 37.7 | 0.6 |
| pHILIC | 963.639 | 182.0316 | 37.7 | 0.6 |
| pHILIC | 963.639 | 676.2632 | 37.7 | 0.6 |
| pHILIC | 963.639 | 160.9236 | 37.6 | 0.6 |
| pHILIC | 963.639 | 355.0884 | 37.6 | 0.6 |
| pHILIC | 963.639 | 257.3083 | 37.6 | 0.6 |
| pHILIC | 963.639 | 122.0441 | 37.6 | 0.6 |
| pHILIC | 963.639 | 108.7954 | 37.6 | 0.6 |
| pHILIC | 963.639 | 258.9532 | 37.6 | 0.6 |
| pHILIC | 963.639 | 363.0938 | 37.6 | 0.6 |
| pHILIC | 963.639 | 169.7972 | 37.6 | 0.6 |

| | | | | |
|---|---|---|---|---|
| pHILIC | 963.639 | 305.3257 | 37.6 | 0.6 |
| pHILIC | 963.639 | 589.6354 | 37.6 | 0.6 |
| pHILIC | 963.639 | 315.5328 | 37.6 | 0.6 |
| pHILIC | 963.639 | 250.1 | 37.6 | 0.6 |
| pHILIC | 963.639 | 125.2859 | 37.6 | 0.6 |
| pHILIC | 963.639 | 657.9598 | 37.6 | 0.6 |
| pHILIC | 963.639 | 571.6476 | 37.6 | 0.6 |
| pHILIC | 963.639 | 243.8611 | 37.6 | 0.6 |
| pHILIC | 963.639 | 105.7791 | 37.6 | 0.6 |
| pHILIC | 963.639 | 120.6971 | 37.6 | 0.6 |
| pHILIC | 963.639 | 918.459 | 37.6 | 0.6 |
| pHILIC | 963.639 | 452.2072 | 37.6 | 0.6 |
| pHILIC | 963.639 | 477.2762 | 37.5 | 0.6 |
| pHILIC | 963.639 | 265.2326 | 37.5 | 0.6 |
| pHILIC | 963.639 | 352.0895 | 37.5 | 0.6 |
| pHILIC | 963.639 | 286.6211 | 37.5 | 0.6 |
| pHILIC | 963.639 | 115.2148 | 37.5 | 0.6 |
| pHILIC | 963.639 | 738.2794 | 37.5 | 0.6 |
| pHILIC | 963.639 | 114.9521 | 37.5 | 0.6 |
| pHILIC | 963.639 | 235.8555 | 37.5 | 0.6 |
| pHILIC | 963.639 | 233.2895 | 37.5 | 0.6 |
| pHILIC | 963.639 | 180.0074 | 37.5 | 0.6 |
| pHILIC | 963.639 | 420.8186 | 37.5 | 0.6 |
| pHILIC | 963.639 | 175.9718 | 37.5 | 0.6 |
| pHILIC | 963.639 | 581.3387 | 37.5 | 0.6 |
| pHILIC | 963.639 | 185.5638 | 37.5 | 0.6 |
| pHILIC | 963.639 | 317.3516 | 37.5 | 0.6 |
| pHILIC | 963.639 | 185.639 | 37.5 | 0.6 |
| pHILIC | 963.639 | 310.0156 | 37.5 | 0.6 |
| pHILIC | 963.639 | 111.7684 | 37.5 | 0.6 |
| pHILIC | 963.639 | 165.2427 | 37.5 | 0.6 |
| pHILIC | 963.639 | 274.527 | 37.5 | 0.6 |
| pHILIC | 963.639 | 123.3787 | 37.5 | 0.6 |
| pHILIC | 963.639 | 135.5559 | 37.5 | 0.6 |
| pHILIC | 963.639 | 217.7685 | 37.5 | 0.6 |
| pHILIC | 963.639 | 198.5214 | 37.5 | 0.6 |
| pHILIC | 963.639 | 213.8413 | 37.5 | 0.6 |
| pHILIC | 963.639 | 219.0774 | 37.5 | 0.6 |
| pHILIC | 963.639 | 281.8215 | 37.4 | 0.6 |
| pHILIC | 963.639 | 352.8096 | 37.4 | 0.6 |
| pHILIC | 963.639 | 106.934 | 37.4 | 0.6 |
| pHILIC | 963.639 | 128.1734 | 37.4 | 0.6 |

| | | | | |
|---|---|---|---|---|
| pHILIC | 963.639 | 102.0292 | 37.4 | 0.6 |
| pHILIC | 963.639 | 151.3429 | 37.4 | 0.6 |
| pHILIC | 963.639 | 119.4466 | 37.4 | 0.6 |
| pHILIC | 963.639 | 147.6086 | 37.4 | 0.6 |
| pHILIC | 963.639 | 689.1534 | 37.4 | 0.6 |
| pHILIC | 963.639 | 190.5473 | 37.4 | 0.6 |
| pHILIC | 963.639 | 124.9631 | 37.4 | 0.6 |
| pHILIC | 963.639 | 476.37 | 37.4 | 0.6 |
| pHILIC | 963.639 | 116.7169 | 37.4 | 0.6 |
| pHILIC | 963.639 | 618.6494 | 37.4 | 0.6 |
| pHILIC | 963.639 | 118.7627 | 37.4 | 0.6 |
| pHILIC | 963.639 | 568.754 | 37.4 | 0.6 |
| pHILIC | 963.639 | 190.2001 | 37.4 | 0.6 |
| pHILIC | 963.639 | 220.1364 | 37.4 | 0.6 |
| pHILIC | 963.639 | 164.5715 | 37.4 | 0.6 |
| pHILIC | 963.639 | 516.2291 | 37.4 | 0.6 |
| pHILIC | 963.639 | 598.7325 | 37.4 | 0.6 |
| pHILIC | 963.639 | 106.3918 | 37.3 | 0.6 |
| pHILIC | 963.639 | 478.6364 | 37.3 | 0.6 |
| pHILIC | 963.639 | 174.7399 | 37.3 | 0.6 |
| pHILIC | 963.639 | 303.7866 | 37.3 | 0.6 |
| pHILIC | 963.639 | 540.2577 | 37.3 | 0.6 |
| pHILIC | 963.639 | 169.6462 | 37.3 | 0.6 |
| pHILIC | 963.639 | 112.3207 | 37.3 | 0.6 |
| pHILIC | 963.639 | 118.7821 | 37.3 | 0.6 |
| pHILIC | 963.639 | 117.3445 | 37.3 | 0.6 |
| pHILIC | 963.639 | 956.5232 | 37.3 | 0.6 |
| pHILIC | 963.639 | 761.8848 | 37.3 | 0.6 |
| pHILIC | 963.639 | 276.8632 | 37.3 | 0.6 |
| pHILIC | 963.639 | 165.9555 | 37.3 | 0.6 |
| pHILIC | 963.639 | 101.7364 | 37.3 | 0.6 |
| pHILIC | 963.639 | 133.5245 | 37.3 | 0.6 |
| pHILIC | 963.639 | 247.432 | 37.3 | 0.6 |
| pHILIC | 963.639 | 888.3739 | 37.3 | 0.6 |
| pHILIC | 963.639 | 432.7793 | 37.3 | 0.6 |
| pHILIC | 963.639 | 126.7192 | 37.3 | 0.6 |
| pHILIC | 963.639 | 379.3568 | 37.3 | 0.6 |
| pHILIC | 963.639 | 263.6777 | 37.2 | 0.6 |
| pHILIC | 963.639 | 409.1302 | 37.2 | 0.6 |
| pHILIC | 963.639 | 136.4256 | 37.2 | 0.6 |
| pHILIC | 963.639 | 654.8741 | 37.2 | 0.59 |
| pHILIC | 963.639 | 139.7841 | 37.2 | 0.59 |

| | | | | |
|---|---|---|---|---|
| pHILIC | 963.639 | 624.6225 | 37.2 | 0.59 |
| pHILIC | 963.639 | 240.7021 | 37.2 | 0.59 |
| pHILIC | 963.639 | 100.6731 | 37.2 | 0.59 |
| pHILIC | 963.639 | 404.112 | 37.2 | 0.59 |
| pHILIC | 963.639 | 128.0764 | 37.2 | 0.59 |
| pHILIC | 963.639 | 284.202 | 37.2 | 0.59 |
| pHILIC | 963.639 | 102.1133 | 37.2 | 0.59 |
| pHILIC | 963.639 | 355.8661 | 37.2 | 0.59 |
| pHILIC | 963.639 | 504.7114 | 37.2 | 0.59 |
| pHILIC | 963.639 | 138.5798 | 37.1 | 0.59 |
| pHILIC | 963.639 | 373.6856 | 37.1 | 0.59 |
| pHILIC | 963.639 | 204.2441 | 37.1 | 0.59 |
| pHILIC | 963.639 | 537.1496 | 37.1 | 0.59 |
| pHILIC | 963.639 | 825.3064 | 37.1 | 0.59 |
| pHILIC | 963.639 | 174.0734 | 37.1 | 0.59 |
| pHILIC | 963.639 | 142.6566 | 37.1 | 0.59 |
| pHILIC | 963.639 | 207.4693 | 37.1 | 0.59 |
| pHILIC | 963.639 | 132.5887 | 37.1 | 0.59 |
| pHILIC | 963.639 | 307.5968 | 37.1 | 0.59 |
| pHILIC | 963.639 | 311.9528 | 37.1 | 0.59 |
| pHILIC | 963.639 | 118.6637 | 37.1 | 0.59 |
| pHILIC | 963.639 | 684.639 | 37.1 | 0.59 |
| pHILIC | 963.639 | 158.4158 | 37.1 | 0.59 |
| pHILIC | 963.639 | 225.1125 | 37.1 | 0.59 |
| pHILIC | 963.639 | 116.3698 | 37.1 | 0.59 |
| pHILIC | 963.639 | 145.8885 | 37.1 | 0.59 |
| pHILIC | 963.639 | 519.5013 | 37.1 | 0.59 |
| pHILIC | 963.639 | 280.4985 | 37.1 | 0.59 |
| pHILIC | 963.639 | 105.4292 | 37.1 | 0.59 |
| pHILIC | 963.639 | 260.1557 | 37.1 | 0.59 |
| pHILIC | 963.639 | 115.8713 | 37.1 | 0.59 |
| pHILIC | 963.639 | 116.952 | 37 | 0.59 |
| pHILIC | 963.639 | 193.1888 | 37 | 0.59 |
| pHILIC | 963.639 | 117.525 | 37 | 0.59 |
| pHILIC | 963.639 | 161.4172 | 37 | 0.59 |
| pHILIC | 963.639 | 155.4035 | 37 | 0.59 |
| pHILIC | 963.639 | 219.7381 | 37 | 0.59 |
| pHILIC | 963.639 | 111.7413 | 37 | 0.59 |
| pHILIC | 963.639 | 162.5698 | 37 | 0.59 |
| pHILIC | 963.639 | 201.1924 | 37 | 0.59 |
| pHILIC | 963.639 | 485.6292 | 37 | 0.59 |
| pHILIC | 963.639 | 101.0352 | 37 | 0.59 |

| | | | | |
|---|---|---|---|---|
| pHILIC | 963.639 | 932.0682 | 37 | 0.59 |
| pHILIC | 963.639 | 143.4537 | 37 | 0.59 |
| pHILIC | 963.639 | 121.5563 | 37 | 0.59 |
| pHILIC | 963.639 | 528.744 | 37 | 0.59 |
| pHILIC | 963.639 | 745.8832 | 37 | 0.59 |
| pHILIC | 963.639 | 677.3543 | 37 | 0.59 |
| pHILIC | 963.639 | 386.5765 | 37 | 0.59 |
| pHILIC | 963.639 | 133.562 | 36.9 | 0.59 |
| pHILIC | 963.639 | 150.0178 | 36.9 | 0.59 |
| pHILIC | 963.639 | 332.8855 | 36.9 | 0.59 |
| pHILIC | 963.639 | 363.7413 | 36.9 | 0.59 |
| pHILIC | 963.639 | 436.3382 | 36.9 | 0.59 |
| pHILIC | 963.639 | 223.6768 | 36.9 | 0.59 |
| pHILIC | 963.639 | 699.4302 | 36.9 | 0.59 |
| pHILIC | 963.639 | 355.9598 | 36.9 | 0.59 |
| pHILIC | 963.639 | 112.6771 | 36.9 | 0.59 |
| pHILIC | 963.639 | 331.5841 | 36.9 | 0.59 |
| pHILIC | 963.639 | 338.8705 | 36.9 | 0.59 |
| pHILIC | 963.639 | 405.4098 | 36.9 | 0.59 |
| pHILIC | 963.639 | 146.0373 | 36.9 | 0.59 |
| pHILIC | 963.639 | 130.2758 | 36.9 | 0.59 |
| pHILIC | 963.639 | 144.2525 | 36.8 | 0.59 |
| pHILIC | 963.639 | 347.2299 | 36.8 | 0.59 |
| pHILIC | 963.639 | 371.3896 | 36.8 | 0.59 |
| pHILIC | 963.639 | 230.5917 | 36.8 | 0.59 |
| pHILIC | 963.639 | 206.0247 | 36.8 | 0.59 |
| pHILIC | 963.639 | 154.2498 | 36.8 | 0.59 |
| pHILIC | 963.639 | 103.8445 | 36.8 | 0.59 |
| pHILIC | 963.639 | 432.4254 | 36.8 | 0.59 |
| pHILIC | 963.639 | 107.1853 | 36.8 | 0.59 |
| pHILIC | 963.639 | 604.5764 | 36.8 | 0.59 |
| pHILIC | 963.639 | 132.767 | 36.8 | 0.59 |
| pHILIC | 963.639 | 189.8316 | 36.8 | 0.59 |
| pHILIC | 963.639 | 171.6797 | 36.8 | 0.59 |
| pHILIC | 963.639 | 616.5826 | 36.8 | 0.59 |
| pHILIC | 963.639 | 501.823 | 36.8 | 0.59 |
| pHILIC | 963.639 | 465.8417 | 36.8 | 0.59 |
| pHILIC | 963.639 | 105.6106 | 36.8 | 0.59 |
| pHILIC | 963.639 | 166.9875 | 36.8 | 0.59 |
| pHILIC | 963.639 | 206.3981 | 36.8 | 0.59 |
| pHILIC | 963.639 | 192.443 | 36.8 | 0.59 |
| pHILIC | 963.639 | 943.3516 | 36.8 | 0.59 |

| | | | | |
|---|---|---|---|---|
| pHILIC | 963.639 | 901.4585 | 36.8 | 0.59 |
| pHILIC | 963.639 | 197.7514 | 36.8 | 0.59 |
| pHILIC | 963.639 | 291.325 | 36.7 | 0.59 |
| pHILIC | 963.639 | 178.9543 | 36.7 | 0.59 |
| pHILIC | 963.639 | 221.4131 | 36.7 | 0.59 |
| pHILIC | 963.639 | 345.7729 | 36.7 | 0.59 |
| pHILIC | 963.639 | 103.1164 | 36.7 | 0.59 |
| pHILIC | 963.639 | 180.8483 | 36.7 | 0.59 |
| pHILIC | 963.639 | 131.855 | 36.7 | 0.59 |
| pHILIC | 963.639 | 213.0606 | 36.7 | 0.59 |
| pHILIC | 963.639 | 141.2513 | 36.7 | 0.59 |
| pHILIC | 963.639 | 138.7866 | 36.7 | 0.59 |
| pHILIC | 963.639 | 200.362 | 36.7 | 0.59 |
| pHILIC | 963.639 | 224.1186 | 36.7 | 0.59 |
| pHILIC | 963.639 | 104.184 | 36.7 | 0.59 |
| pHILIC | 963.639 | 623.0299 | 36.7 | 0.59 |
| pHILIC | 963.639 | 114.5627 | 36.7 | 0.59 |
| pHILIC | 963.639 | 110.9738 | 36.7 | 0.59 |
| pHILIC | 963.639 | 416.8438 | 36.7 | 0.59 |
| pHILIC | 963.639 | 109.2775 | 36.7 | 0.59 |
| pHILIC | 963.639 | 268.0068 | 36.7 | 0.59 |
| pHILIC | 963.639 | 309.7213 | 36.6 | 0.59 |
| pHILIC | 963.639 | 122.1528 | 36.6 | 0.59 |
| pHILIC | 963.639 | 295.2764 | 36.6 | 0.59 |
| pHILIC | 963.639 | 453.5578 | 36.6 | 0.58 |
| pHILIC | 963.639 | 717.2434 | 36.6 | 0.58 |
| pHILIC | 963.639 | 198.6563 | 36.6 | 0.58 |
| pHILIC | 963.639 | 237.9301 | 36.6 | 0.58 |
| pHILIC | 963.639 | 215.7368 | 36.6 | 0.58 |
| pHILIC | 963.639 | 396.1932 | 36.6 | 0.58 |
| pHILIC | 963.639 | 690.6115 | 36.6 | 0.58 |
| pHILIC | 963.639 | 133.4877 | 36.6 | 0.58 |
| pHILIC | 963.639 | 623.7935 | 36.6 | 0.58 |
| pHILIC | 963.639 | 919.5782 | 36.6 | 0.58 |
| pHILIC | 963.639 | 320.84 | 36.6 | 0.58 |
| pHILIC | 963.639 | 847.8875 | 36.6 | 0.58 |
| pHILIC | 963.639 | 145.022 | 36.6 | 0.58 |
| pHILIC | 963.639 | 783.877 | 36.5 | 0.58 |
| pHILIC | 963.639 | 135.5099 | 36.5 | 0.58 |
| pHILIC | 963.639 | 320.1322 | 36.5 | 0.58 |
| pHILIC | 963.639 | 104.0821 | 36.5 | 0.58 |
| pHILIC | 963.639 | 193.4659 | 36.5 | 0.58 |

| | | | | |
|---|---|---|---|---|
| pHILIC | 963.639 | 490.6917 | 36.5 | 0.58 |
| pHILIC | 963.639 | 111.4939 | 36.5 | 0.58 |
| pHILIC | 963.639 | 128.6659 | 36.5 | 0.58 |
| pHILIC | 963.639 | 223.7528 | 36.5 | 0.58 |
| pHILIC | 963.639 | 213.867 | 36.5 | 0.58 |
| pHILIC | 963.639 | 837.3454 | 36.5 | 0.58 |
| pHILIC | 963.639 | 191.9643 | 36.5 | 0.58 |
| pHILIC | 963.639 | 456.2531 | 36.5 | 0.58 |
| pHILIC | 963.639 | 145.1396 | 36.5 | 0.58 |
| pHILIC | 963.639 | 353.4711 | 36.5 | 0.58 |
| pHILIC | 963.639 | 114.2354 | 36.5 | 0.58 |
| pHILIC | 963.639 | 478.4026 | 36.5 | 0.58 |
| pHILIC | 963.639 | 161.873 | 36.4 | 0.58 |
| pHILIC | 963.639 | 108.3358 | 36.4 | 0.58 |
| pHILIC | 963.639 | 111.0316 | 36.4 | 0.58 |
| pHILIC | 963.639 | 357.8047 | 36.4 | 0.58 |
| pHILIC | 963.639 | 155.3405 | 36.4 | 0.58 |
| pHILIC | 963.639 | 389.7217 | 36.4 | 0.58 |
| pHILIC | 963.639 | 292.373 | 36.4 | 0.58 |
| pHILIC | 963.639 | 195.55 | 36.4 | 0.58 |
| pHILIC | 963.639 | 247.5048 | 36.4 | 0.58 |
| pHILIC | 963.639 | 186.9864 | 36.4 | 0.58 |
| pHILIC | 963.639 | 633.2125 | 36.4 | 0.58 |
| pHILIC | 963.639 | 145.2102 | 36.4 | 0.58 |
| pHILIC | 963.639 | 300.4344 | 36.4 | 0.58 |
| pHILIC | 963.639 | 225.219 | 36.4 | 0.58 |
| pHILIC | 963.639 | 198.3444 | 36.4 | 0.58 |
| pHILIC | 963.639 | 114.604 | 36.4 | 0.58 |
| pHILIC | 963.639 | 177.0368 | 36.4 | 0.58 |
| pHILIC | 963.639 | 100.9515 | 36.4 | 0.58 |
| pHILIC | 963.639 | 396.279 | 36.4 | 0.58 |
| pHILIC | 963.639 | 512.7825 | 36.4 | 0.58 |
| pHILIC | 963.639 | 223.096 | 36.3 | 0.58 |
| pHILIC | 963.639 | 307.1275 | 36.3 | 0.58 |
| pHILIC | 963.639 | 182.8906 | 36.3 | 0.58 |
| pHILIC | 963.639 | 313.2248 | 36.3 | 0.58 |
| pHILIC | 963.639 | 109.4452 | 36.3 | 0.58 |
| pHILIC | 963.639 | 122.4188 | 36.3 | 0.58 |
| pHILIC | 963.639 | 273.5809 | 36.3 | 0.58 |
| pHILIC | 963.639 | 651.3015 | 36.3 | 0.58 |
| pHILIC | 963.639 | 104.5826 | 36.3 | 0.58 |
| pHILIC | 963.639 | 216.8642 | 36.3 | 0.58 |

| | | | | |
|---|---|---|---|---|
| pHILIC | 963.639 | 204.8412 | 36.3 | 0.58 |
| pHILIC | 963.639 | 102.775 | 36.3 | 0.58 |
| pHILIC | 963.639 | 260.8854 | 36.2 | 0.58 |
| pHILIC | 963.639 | 745.6851 | 36.2 | 0.58 |
| pHILIC | 963.639 | 144.3735 | 36.2 | 0.58 |
| pHILIC | 963.639 | 266.1256 | 36.2 | 0.58 |
| pHILIC | 963.639 | 103.8879 | 36.2 | 0.58 |
| pHILIC | 963.639 | 206.8619 | 36.2 | 0.58 |
| pHILIC | 963.639 | 107.8192 | 36.2 | 0.58 |
| pHILIC | 963.639 | 785.4211 | 36.2 | 0.58 |
| pHILIC | 963.639 | 121.1474 | 36.2 | 0.58 |
| pHILIC | 963.639 | 124.5149 | 36.2 | 0.58 |
| pHILIC | 963.639 | 196.8806 | 36.2 | 0.58 |
| pHILIC | 963.639 | 228.4461 | 36.2 | 0.58 |
| pHILIC | 963.639 | 160.6566 | 36.2 | 0.58 |
| pHILIC | 963.639 | 105.8193 | 36.2 | 0.58 |
| pHILIC | 963.639 | 198.2542 | 36.2 | 0.58 |
| pHILIC | 963.639 | 715.1646 | 36.1 | 0.58 |
| pHILIC | 963.639 | 119.2349 | 36.1 | 0.58 |
| pHILIC | 963.639 | 396.8295 | 36.1 | 0.58 |
| pHILIC | 963.639 | 552.2837 | 36.1 | 0.58 |
| pHILIC | 963.639 | 100.6186 | 36.1 | 0.58 |
| pHILIC | 963.639 | 672.8243 | 36.1 | 0.58 |
| pHILIC | 963.639 | 132.1591 | 36.1 | 0.58 |
| pHILIC | 963.639 | 112.9407 | 36.1 | 0.58 |
| pHILIC | 963.639 | 158.1388 | 36.1 | 0.58 |
| pHILIC | 963.639 | 204.8756 | 36.1 | 0.58 |
| pHILIC | 963.639 | 470.5553 | 36.1 | 0.58 |
| pHILIC | 963.639 | 105.6808 | 36.1 | 0.58 |
| pHILIC | 963.639 | 108.893 | 36.1 | 0.58 |
| pHILIC | 963.639 | 517.1893 | 36.1 | 0.58 |
| pHILIC | 963.639 | 119.7182 | 36.1 | 0.58 |
| pHILIC | 963.639 | 613.1075 | 36.1 | 0.58 |
| pHILIC | 963.639 | 512.3229 | 36 | 0.58 |
| pHILIC | 963.639 | 121.8373 | 36 | 0.58 |
| pHILIC | 963.639 | 106.5208 | 36 | 0.58 |
| pHILIC | 963.639 | 349.0695 | 36 | 0.58 |
| pHILIC | 963.639 | 207.7834 | 36 | 0.58 |
| pHILIC | 963.639 | 154.0863 | 36 | 0.58 |
| pHILIC | 963.639 | 182.1167 | 36 | 0.58 |
| pHILIC | 963.639 | 411.7873 | 36 | 0.58 |
| pHILIC | 963.639 | 152.1593 | 36 | 0.58 |

| | | | | |
|---|---|---|---|---|
| pHILIC | 963.639 | 137.615 | 36 | 0.58 |
| pHILIC | 963.639 | 248.1003 | 36 | 0.57 |
| pHILIC | 963.639 | 302.5323 | 36 | 0.57 |
| pHILIC | 963.639 | 111.7071 | 36 | 0.57 |
| pHILIC | 963.639 | 109.2951 | 36 | 0.57 |
| pHILIC | 963.639 | 214.0514 | 35.9 | 0.57 |
| pHILIC | 963.639 | 594.84 | 35.9 | 0.57 |
| pHILIC | 963.639 | 232.6652 | 35.9 | 0.57 |
| pHILIC | 963.639 | 349.6095 | 35.9 | 0.57 |
| pHILIC | 963.639 | 169.1832 | 35.9 | 0.57 |
| pHILIC | 963.639 | 108.7022 | 35.9 | 0.57 |
| pHILIC | 963.639 | 110.6041 | 35.9 | 0.57 |
| pHILIC | 963.639 | 212.4252 | 35.9 | 0.57 |
| pHILIC | 963.639 | 409.9678 | 35.9 | 0.57 |
| pHILIC | 963.639 | 145.9557 | 35.9 | 0.57 |
| pHILIC | 963.639 | 302.7877 | 35.9 | 0.57 |
| pHILIC | 963.639 | 103.2865 | 35.9 | 0.57 |
| pHILIC | 963.639 | 343.1641 | 35.9 | 0.57 |
| pHILIC | 963.639 | 186.7619 | 35.9 | 0.57 |
| pHILIC | 963.639 | 112.6576 | 35.9 | 0.57 |
| pHILIC | 963.639 | 228.5578 | 35.9 | 0.57 |
| pHILIC | 963.639 | 876.8564 | 35.9 | 0.57 |
| pHILIC | 963.639 | 118.8907 | 35.9 | 0.57 |
| pHILIC | 963.639 | 122.9586 | 35.9 | 0.57 |
| pHILIC | 963.639 | 110.8578 | 35.8 | 0.57 |
| pHILIC | 963.639 | 129.2212 | 35.8 | 0.57 |
| pHILIC | 963.639 | 129.9807 | 35.8 | 0.57 |
| pHILIC | 963.639 | 137.5517 | 35.8 | 0.57 |
| pHILIC | 963.639 | 105.8791 | 35.8 | 0.57 |
| pHILIC | 963.639 | 423.435 | 35.8 | 0.57 |
| pHILIC | 963.639 | 173.4922 | 35.8 | 0.57 |
| pHILIC | 963.639 | 913.1938 | 35.8 | 0.57 |
| pHILIC | 963.639 | 110.2964 | 35.8 | 0.57 |
| pHILIC | 963.639 | 323.6114 | 35.8 | 0.57 |
| pHILIC | 963.639 | 198.7557 | 35.8 | 0.57 |
| pHILIC | 963.639 | 133.4309 | 35.8 | 0.57 |
| pHILIC | 963.639 | 170.7253 | 35.8 | 0.57 |
| pHILIC | 963.639 | 481.7603 | 35.8 | 0.57 |
| pHILIC | 963.639 | 287.7197 | 35.8 | 0.57 |
| pHILIC | 963.639 | 140.872 | 35.8 | 0.57 |
| pHILIC | 963.639 | 144.6264 | 35.8 | 0.57 |
| pHILIC | 963.639 | 136.6804 | 35.7 | 0.57 |

| | | | | |
|---|---|---|---|---|
| pHILIC | 963.639 | 137.1112 | 35.7 | 0.57 |
| pHILIC | 963.639 | 264.9773 | 35.7 | 0.57 |
| pHILIC | 963.639 | 397.3725 | 35.7 | 0.57 |
| pHILIC | 963.639 | 119.9399 | 35.7 | 0.57 |
| pHILIC | 963.639 | 224.018 | 35.7 | 0.57 |
| pHILIC | 963.639 | 132.404 | 35.7 | 0.57 |
| pHILIC | 963.639 | 164.0225 | 35.7 | 0.57 |
| pHILIC | 963.639 | 431.9985 | 35.7 | 0.57 |
| pHILIC | 963.639 | 266.0678 | 35.7 | 0.57 |
| pHILIC | 963.639 | 262.3645 | 35.7 | 0.57 |
| pHILIC | 963.639 | 189.9083 | 35.7 | 0.57 |
| pHILIC | 963.639 | 151.771 | 35.7 | 0.57 |
| pHILIC | 963.639 | 756.8671 | 35.7 | 0.57 |
| pHILIC | 963.639 | 245.8253 | 35.7 | 0.57 |
| pHILIC | 963.639 | 196.724 | 35.7 | 0.57 |
| pHILIC | 963.639 | 158.0558 | 35.7 | 0.57 |
| pHILIC | 963.639 | 279.0214 | 35.6 | 0.57 |
| pHILIC | 963.639 | 205.1829 | 35.6 | 0.57 |
| pHILIC | 963.639 | 189.5555 | 35.6 | 0.57 |
| pHILIC | 963.639 | 963.3145 | 35.6 | 0.57 |
| pHILIC | 963.639 | 206.7407 | 35.6 | 0.57 |
| pHILIC | 963.639 | 113.7522 | 35.6 | 0.57 |
| pHILIC | 963.639 | 205.493 | 35.6 | 0.57 |
| pHILIC | 963.639 | 214.9979 | 35.6 | 0.57 |
| pHILIC | 963.639 | 161.4365 | 35.6 | 0.57 |
| pHILIC | 963.639 | 184.6388 | 35.6 | 0.57 |
| pHILIC | 963.639 | 397.6578 | 35.6 | 0.57 |
| pHILIC | 963.639 | 236.2697 | 35.6 | 0.57 |
| pHILIC | 963.639 | 225.592 | 35.5 | 0.57 |
| pHILIC | 963.639 | 177.8904 | 35.5 | 0.57 |
| pHILIC | 963.639 | 136.0188 | 35.5 | 0.57 |
| pHILIC | 963.639 | 246.0108 | 35.5 | 0.57 |
| pHILIC | 963.639 | 135.1183 | 35.5 | 0.57 |
| pHILIC | 963.639 | 255.6521 | 35.5 | 0.57 |
| pHILIC | 963.639 | 117.9192 | 35.5 | 0.57 |
| pHILIC | 963.639 | 169.1168 | 35.5 | 0.57 |
| pHILIC | 963.639 | 104.2454 | 35.5 | 0.57 |
| pHILIC | 963.639 | 136.7146 | 35.5 | 0.57 |
| pHILIC | 963.639 | 125.1047 | 35.5 | 0.57 |
| pHILIC | 963.639 | 234.9869 | 35.5 | 0.57 |
| pHILIC | 963.639 | 119.8645 | 35.4 | 0.57 |
| pHILIC | 963.639 | 633.544 | 35.4 | 0.57 |

| | | | | |
|---|---|---|---|---|
| pHILIC | 963.639 | 901.8433 | 35.4 | 0.57 |
| pHILIC | 963.639 | 211.3502 | 35.4 | 0.57 |
| pHILIC | 963.639 | 183.2808 | 35.4 | 0.57 |
| pHILIC | 963.639 | 304.8657 | 35.4 | 0.57 |
| pHILIC | 963.639 | 123.7643 | 35.4 | 0.57 |
| pHILIC | 963.639 | 116.3638 | 35.4 | 0.57 |
| pHILIC | 963.639 | 180.7566 | 35.4 | 0.57 |
| pHILIC | 963.639 | 166.366 | 35.4 | 0.57 |
| pHILIC | 963.639 | 693.2849 | 35.4 | 0.57 |
| pHILIC | 963.639 | 155.1037 | 35.4 | 0.57 |
| pHILIC | 963.639 | 103.1845 | 35.4 | 0.57 |
| pHILIC | 963.639 | 116.2313 | 35.4 | 0.57 |
| pHILIC | 963.639 | 124.5639 | 35.4 | 0.57 |
| pHILIC | 963.639 | 126.9386 | 35.4 | 0.57 |
| pHILIC | 963.639 | 128.1141 | 35.4 | 0.57 |
| pHILIC | 963.639 | 392.6168 | 35.4 | 0.57 |
| pHILIC | 963.639 | 114.2705 | 35.4 | 0.57 |
| pHILIC | 963.639 | 475.399 | 35.4 | 0.57 |
| pHILIC | 963.639 | 100.7652 | 35.4 | 0.56 |
| pHILIC | 963.639 | 271.669 | 35.3 | 0.56 |
| pHILIC | 963.639 | 104.4195 | 35.3 | 0.56 |
| pHILIC | 963.639 | 533.6219 | 35.3 | 0.56 |
| pHILIC | 963.639 | 366.0172 | 35.3 | 0.56 |
| pHILIC | 963.639 | 189.3529 | 35.3 | 0.56 |
| pHILIC | 963.639 | 266.5737 | 35.3 | 0.56 |
| pHILIC | 963.639 | 108.3766 | 35.3 | 0.56 |
| pHILIC | 963.639 | 138.4449 | 35.3 | 0.56 |
| pHILIC | 963.639 | 308.6385 | 35.3 | 0.56 |
| pHILIC | 963.639 | 295.1489 | 35.3 | 0.56 |
| pHILIC | 963.639 | 113.1325 | 35.3 | 0.56 |
| pHILIC | 963.639 | 216.1533 | 35.3 | 0.56 |
| pHILIC | 963.639 | 366.5846 | 35.3 | 0.56 |
| pHILIC | 963.639 | 290.6251 | 35.3 | 0.56 |
| pHILIC | 963.639 | 142.802 | 35.2 | 0.56 |
| pHILIC | 963.639 | 890.7568 | 35.2 | 0.56 |
| pHILIC | 963.639 | 121.5825 | 35.2 | 0.56 |
| pHILIC | 963.639 | 142.4341 | 35.2 | 0.56 |
| pHILIC | 963.639 | 114.0152 | 35.2 | 0.56 |
| pHILIC | 963.639 | 131.6782 | 35.2 | 0.56 |
| pHILIC | 963.639 | 108.3305 | 35.2 | 0.56 |
| pHILIC | 963.639 | 135.4247 | 35.2 | 0.56 |
| pHILIC | 963.639 | 275.4632 | 35.2 | 0.56 |

| | | | | |
|---|---|---|---|---|
| pHILIC | 963.639 | 234.7437 | 35.2 | 0.56 |
| pHILIC | 963.639 | 555.5195 | 35.2 | 0.56 |
| pHILIC | 963.639 | 198.6761 | 35.2 | 0.56 |
| pHILIC | 963.639 | 376.257 | 35.2 | 0.56 |
| pHILIC | 963.639 | 248.0438 | 35.2 | 0.56 |
| pHILIC | 963.639 | 238.1399 | 35.2 | 0.56 |
| pHILIC | 963.639 | 146.8071 | 35.2 | 0.56 |
| pHILIC | 963.639 | 495.3976 | 35.2 | 0.56 |
| pHILIC | 963.639 | 414.7074 | 35.2 | 0.56 |
| pHILIC | 963.639 | 279.3954 | 35.1 | 0.56 |
| pHILIC | 963.639 | 219.2871 | 35.1 | 0.56 |
| pHILIC | 963.639 | 216.4434 | 35.1 | 0.56 |
| pHILIC | 963.639 | 288.5524 | 35.1 | 0.56 |
| pHILIC | 963.639 | 123.9301 | 35.1 | 0.56 |
| pHILIC | 963.639 | 180.9964 | 35.1 | 0.56 |
| pHILIC | 963.639 | 329.0161 | 35.1 | 0.56 |
| pHILIC | 963.639 | 146.6969 | 35.1 | 0.56 |
| pHILIC | 963.639 | 265.6834 | 35.1 | 0.56 |
| pHILIC | 963.639 | 388.5912 | 35.1 | 0.56 |
| pHILIC | 963.639 | 320.3664 | 35.1 | 0.56 |
| pHILIC | 963.639 | 136.5673 | 35.1 | 0.56 |
| pHILIC | 963.639 | 425.5225 | 35.1 | 0.56 |
| pHILIC | 963.639 | 120.597 | 35 | 0.56 |
| pHILIC | 963.639 | 171.9289 | 35 | 0.56 |
| pHILIC | 963.639 | 306.6241 | 35 | 0.56 |
| pHILIC | 963.639 | 149.8525 | 35 | 0.56 |
| pHILIC | 963.639 | 189.2906 | 35 | 0.56 |
| pHILIC | 963.639 | 139.7203 | 35 | 0.56 |
| pHILIC | 963.639 | 102.5784 | 35 | 0.56 |
| pHILIC | 963.639 | 256.7745 | 35 | 0.56 |
| pHILIC | 963.639 | 121.2499 | 35 | 0.56 |
| pHILIC | 963.639 | 181.4833 | 35 | 0.56 |
| pHILIC | 963.639 | 117.08 | 35 | 0.56 |
| pHILIC | 963.639 | 416.5532 | 34.9 | 0.56 |
| pHILIC | 963.639 | 311.7002 | 34.9 | 0.56 |
| pHILIC | 963.639 | 118.0746 | 34.9 | 0.56 |
| pHILIC | 963.639 | 642.3795 | 34.9 | 0.56 |
| pHILIC | 963.639 | 154.8491 | 34.9 | 0.56 |
| pHILIC | 963.639 | 876.0741 | 34.9 | 0.56 |
| pHILIC | 963.639 | 292.9408 | 34.9 | 0.56 |
| pHILIC | 963.639 | 135.705 | 34.9 | 0.56 |
| pHILIC | 963.639 | 271.6296 | 34.9 | 0.56 |

| | | | | |
|---|---|---|---|---|
| pHILIC | 963.639 | 185.0371 | 34.9 | 0.56 |
| pHILIC | 963.639 | 105.4702 | 34.9 | 0.56 |
| pHILIC | 963.639 | 120.4803 | 34.9 | 0.56 |
| pHILIC | 963.639 | 103.1794 | 34.9 | 0.56 |
| pHILIC | 963.639 | 124.1972 | 34.9 | 0.56 |
| pHILIC | 963.639 | 178.9396 | 34.9 | 0.56 |
| pHILIC | 963.639 | 305.2832 | 34.9 | 0.56 |
| pHILIC | 963.639 | 155.8625 | 34.9 | 0.56 |
| pHILIC | 963.639 | 166.6558 | 34.9 | 0.56 |
| pHILIC | 963.639 | 192.3952 | 34.8 | 0.56 |
| pHILIC | 963.639 | 111.9134 | 34.8 | 0.56 |
| pHILIC | 963.639 | 191.702 | 34.8 | 0.56 |
| pHILIC | 963.639 | 675.3439 | 34.8 | 0.56 |
| pHILIC | 963.639 | 301.1203 | 34.8 | 0.56 |
| pHILIC | 963.639 | 123.8262 | 34.8 | 0.56 |
| pHILIC | 963.639 | 182.0016 | 34.8 | 0.56 |
| pHILIC | 963.639 | 180.3446 | 34.7 | 0.56 |
| pHILIC | 963.639 | 719.291 | 34.7 | 0.55 |
| pHILIC | 963.639 | 120.8183 | 34.7 | 0.55 |
| pHILIC | 963.639 | 327.7962 | 34.7 | 0.55 |
| pHILIC | 963.639 | 139.6472 | 34.7 | 0.55 |
| pHILIC | 963.639 | 114.7588 | 34.7 | 0.55 |
| pHILIC | 963.639 | 107.0962 | 34.7 | 0.55 |
| pHILIC | 963.639 | 180.3319 | 34.7 | 0.55 |
| pHILIC | 963.639 | 103.6431 | 34.7 | 0.55 |
| pHILIC | 963.639 | 618.1264 | 34.7 | 0.55 |
| pHILIC | 963.639 | 143.8689 | 34.6 | 0.55 |
| pHILIC | 963.639 | 148.0556 | 34.6 | 0.55 |
| pHILIC | 963.639 | 260.2055 | 34.6 | 0.55 |
| pHILIC | 963.639 | 121.0816 | 34.6 | 0.55 |
| pHILIC | 963.639 | 208.4865 | 34.6 | 0.55 |
| pHILIC | 963.639 | 175.0139 | 34.5 | 0.55 |
| pHILIC | 963.639 | 410.416 | 34.5 | 0.55 |
| pHILIC | 963.639 | 183.1972 | 34.5 | 0.55 |
| pHILIC | 963.639 | 778.0105 | 34.5 | 0.55 |
| pHILIC | 963.639 | 101.7229 | 34.5 | 0.55 |
| pHILIC | 963.639 | 205.0676 | 34.5 | 0.55 |
| pHILIC | 963.639 | 139.5989 | 34.5 | 0.55 |
| pHILIC | 963.639 | 151.2438 | 34.5 | 0.55 |
| pHILIC | 963.639 | 660.1592 | 34.5 | 0.55 |
| pHILIC | 963.639 | 164.3718 | 34.5 | 0.55 |
| pHILIC | 963.639 | 158.1794 | 34.5 | 0.55 |

| | | | | |
|---|---|---|---|---|
| pHILIC | 963.639 | 152.3329 | 34.5 | 0.55 |
| pHILIC | 963.639 | 312.9923 | 34.5 | 0.55 |
| pHILIC | 963.639 | 740.901 | 34.5 | 0.55 |
| pHILIC | 963.639 | 165.5004 | 34.5 | 0.55 |
| pHILIC | 963.639 | 139.9798 | 34.5 | 0.55 |
| pHILIC | 963.639 | 781.3313 | 34.4 | 0.55 |
| pHILIC | 963.639 | 483.0786 | 34.4 | 0.55 |
| pHILIC | 963.639 | 128.3337 | 34.4 | 0.55 |
| pHILIC | 963.639 | 335.8641 | 34.4 | 0.55 |
| pHILIC | 963.639 | 431.2929 | 34.4 | 0.55 |
| pHILIC | 963.639 | 275.0025 | 34.4 | 0.55 |
| pHILIC | 963.639 | 183.7441 | 34.4 | 0.55 |
| pHILIC | 963.639 | 109.6153 | 34.4 | 0.55 |
| pHILIC | 963.639 | 744.2678 | 34.4 | 0.55 |
| pHILIC | 963.639 | 112.8398 | 34.4 | 0.55 |
| pHILIC | 963.639 | 291.1305 | 34.4 | 0.55 |
| pHILIC | 963.639 | 106.6771 | 34.4 | 0.55 |
| pHILIC | 963.639 | 216.3722 | 34.4 | 0.55 |
| pHILIC | 963.639 | 108.1183 | 34.4 | 0.55 |
| pHILIC | 963.639 | 118.9299 | 34.3 | 0.55 |
| pHILIC | 963.639 | 104.0143 | 34.3 | 0.55 |
| pHILIC | 963.639 | 806.3568 | 34.3 | 0.55 |
| pHILIC | 963.639 | 138.9077 | 34.3 | 0.55 |
| pHILIC | 963.639 | 153.7424 | 34.3 | 0.55 |
| pHILIC | 963.639 | 343.642 | 34.3 | 0.55 |
| pHILIC | 963.639 | 179.753 | 34.3 | 0.55 |
| pHILIC | 963.639 | 289.4413 | 34.2 | 0.55 |
| pHILIC | 963.639 | 155.8791 | 34.2 | 0.55 |
| pHILIC | 963.639 | 125.7928 | 34.2 | 0.55 |
| pHILIC | 963.639 | 264.2746 | 34.2 | 0.55 |
| pHILIC | 963.639 | 109.9381 | 34.2 | 0.55 |
| pHILIC | 963.639 | 170.0708 | 34.2 | 0.55 |
| pHILIC | 963.639 | 190.1531 | 34.2 | 0.55 |
| pHILIC | 963.639 | 592.2023 | 34.2 | 0.55 |
| pHILIC | 963.639 | 765.5565 | 34.2 | 0.55 |
| pHILIC | 963.639 | 176.683 | 34.2 | 0.55 |
| pHILIC | 963.639 | 135.9521 | 34.1 | 0.55 |
| pHILIC | 963.639 | 594.5674 | 34.1 | 0.55 |
| pHILIC | 963.639 | 134.6498 | 34.1 | 0.55 |
| pHILIC | 963.639 | 180.3038 | 34.1 | 0.55 |
| pHILIC | 963.639 | 140.5638 | 34.1 | 0.55 |
| pHILIC | 963.639 | 240.3538 | 34.1 | 0.55 |

| | | | | |
|---|---|---|---|---|
| pHILIC | 963.639 | 127.5716 | 34.1 | 0.55 |
| pHILIC | 963.639 | 794.6005 | 34.1 | 0.55 |
| pHILIC | 963.639 | 178.0299 | 34.1 | 0.55 |
| pHILIC | 963.639 | 339.0888 | 34.1 | 0.54 |
| pHILIC | 963.639 | 187.9774 | 34.1 | 0.54 |
| pHILIC | 963.639 | 112.5895 | 34.1 | 0.54 |
| pHILIC | 963.639 | 135.1922 | 34.1 | 0.54 |
| pHILIC | 963.639 | 196.8416 | 34.1 | 0.54 |
| pHILIC | 963.639 | 121.7177 | 34.1 | 0.54 |
| pHILIC | 963.639 | 122.903 | 34.1 | 0.54 |
| pHILIC | 963.639 | 482.4509 | 34.1 | 0.54 |
| pHILIC | 963.639 | 587.566 | 34 | 0.54 |
| pHILIC | 963.639 | 467.972 | 34 | 0.54 |
| pHILIC | 963.639 | 298.92 | 34 | 0.54 |
| pHILIC | 963.639 | 454.4209 | 34 | 0.54 |
| pHILIC | 963.639 | 237.5154 | 34 | 0.54 |
| pHILIC | 963.639 | 126.3483 | 34 | 0.54 |
| pHILIC | 963.639 | 130.803 | 34 | 0.54 |
| pHILIC | 963.639 | 364.6177 | 34 | 0.54 |
| pHILIC | 963.639 | 401.3005 | 34 | 0.54 |
| pHILIC | 963.639 | 536.1924 | 34 | 0.54 |
| pHILIC | 963.639 | 111.0481 | 34 | 0.54 |
| pHILIC | 963.639 | 228.7261 | 34 | 0.54 |
| pHILIC | 963.639 | 137.1762 | 33.9 | 0.54 |
| pHILIC | 963.639 | 161.3106 | 33.9 | 0.54 |
| pHILIC | 963.639 | 123.7811 | 33.9 | 0.54 |
| pHILIC | 963.639 | 224.7245 | 33.9 | 0.54 |
| pHILIC | 963.639 | 207.3878 | 33.9 | 0.54 |
| pHILIC | 963.639 | 202.4433 | 33.9 | 0.54 |
| pHILIC | 963.639 | 214.1758 | 33.9 | 0.54 |
| pHILIC | 963.639 | 142.5769 | 33.9 | 0.54 |
| pHILIC | 963.639 | 248.1327 | 33.8 | 0.54 |
| pHILIC | 963.639 | 169.0423 | 33.8 | 0.54 |
| pHILIC | 963.639 | 541.7047 | 33.8 | 0.54 |
| pHILIC | 963.639 | 144.9962 | 33.8 | 0.54 |
| pHILIC | 963.639 | 250.9708 | 33.8 | 0.54 |
| pHILIC | 963.639 | 258.681 | 33.8 | 0.54 |
| pHILIC | 963.639 | 110.6424 | 33.8 | 0.54 |
| pHILIC | 963.639 | 280.4405 | 33.8 | 0.54 |
| pHILIC | 963.639 | 181.2902 | 33.8 | 0.54 |
| pHILIC | 963.639 | 777.1312 | 33.8 | 0.54 |
| pHILIC | 963.639 | 525.3336 | 33.8 | 0.54 |

| | | | | |
|---|---|---|---|---|
| pHILIC | 963.639 | 514.4731 | 33.8 | 0.54 |
| pHILIC | 963.639 | 350.2427 | 33.8 | 0.54 |
| pHILIC | 963.639 | 137.9596 | 33.8 | 0.54 |
| pHILIC | 963.639 | 148.7462 | 33.7 | 0.54 |
| pHILIC | 963.639 | 119.0749 | 33.7 | 0.54 |
| pHILIC | 963.639 | 162.0728 | 33.7 | 0.54 |
| pHILIC | 963.639 | 105.3065 | 33.7 | 0.54 |
| pHILIC | 963.639 | 159.4702 | 33.7 | 0.54 |
| pHILIC | 963.639 | 114.9235 | 33.7 | 0.54 |
| pHILIC | 963.639 | 129.0363 | 33.7 | 0.54 |
| pHILIC | 963.639 | 171.1954 | 33.7 | 0.54 |
| pHILIC | 963.639 | 309.9578 | 33.7 | 0.54 |
| pHILIC | 963.639 | 428.1251 | 33.6 | 0.54 |
| pHILIC | 963.639 | 152.6696 | 33.6 | 0.54 |
| pHILIC | 963.639 | 158.3925 | 33.6 | 0.54 |
| pHILIC | 963.639 | 508.4889 | 33.6 | 0.54 |
| pHILIC | 963.639 | 163.7629 | 33.6 | 0.54 |
| pHILIC | 963.639 | 1001.8193 | 33.6 | 0.54 |
| pHILIC | 963.639 | 123.0685 | 33.6 | 0.54 |
| pHILIC | 963.639 | 186.6634 | 33.6 | 0.54 |
| pHILIC | 963.639 | 731.4286 | 33.6 | 0.54 |
| pHILIC | 963.639 | 485.3119 | 33.6 | 0.54 |
| pHILIC | 963.639 | 185.5389 | 33.6 | 0.54 |
| pHILIC | 963.639 | 236.1433 | 33.6 | 0.54 |
| pHILIC | 963.639 | 161.6661 | 33.5 | 0.54 |
| pHILIC | 963.639 | 446.7703 | 33.5 | 0.54 |
| pHILIC | 963.639 | 277.1739 | 33.5 | 0.54 |
| pHILIC | 963.639 | 156.7417 | 33.5 | 0.53 |
| pHILIC | 963.639 | 125.942 | 33.4 | 0.53 |
| pHILIC | 963.639 | 628.8146 | 33.4 | 0.53 |
| pHILIC | 963.639 | 150.2935 | 33.4 | 0.53 |
| pHILIC | 963.639 | 140.0364 | 33.4 | 0.53 |
| pHILIC | 963.639 | 474.1865 | 33.4 | 0.53 |
| pHILIC | 963.639 | 609.2435 | 33.4 | 0.53 |
| pHILIC | 963.639 | 242.8729 | 33.4 | 0.53 |
| pHILIC | 963.639 | 554.4543 | 33.4 | 0.53 |
| pHILIC | 963.639 | 194.8242 | 33.4 | 0.53 |
| pHILIC | 963.639 | 395.5282 | 33.4 | 0.53 |
| pHILIC | 963.639 | 933.0649 | 33.4 | 0.53 |
| pHILIC | 963.639 | 449.1996 | 33.3 | 0.53 |
| pHILIC | 963.639 | 119.01 | 33.3 | 0.53 |
| pHILIC | 963.639 | 218.6154 | 33.3 | 0.53 |

| | | | | |
|---|---|---|---|---|
| pHILIC | 963.639 | 160.5892 | 33.3 | 0.53 |
| pHILIC | 963.639 | 750.4003 | 33.3 | 0.53 |
| pHILIC | 963.639 | 564.2621 | 33.2 | 0.53 |
| pHILIC | 963.639 | 113.7008 | 33.2 | 0.53 |
| pHILIC | 963.639 | 568.8847 | 33.2 | 0.53 |
| pHILIC | 963.639 | 530.357 | 33.2 | 0.53 |
| pHILIC | 963.639 | 216.7908 | 33.2 | 0.53 |
| pHILIC | 963.639 | 166.3113 | 33.2 | 0.53 |
| pHILIC | 963.639 | 450.2419 | 33.2 | 0.53 |
| pHILIC | 963.639 | 209.9357 | 33.2 | 0.53 |
| pHILIC | 963.639 | 126.1068 | 33.2 | 0.53 |
| pHILIC | 963.639 | 196.0429 | 33.2 | 0.53 |
| pHILIC | 963.639 | 151.487 | 33.2 | 0.53 |
| pHILIC | 963.639 | 515.3257 | 33.2 | 0.53 |
| pHILIC | 963.639 | 610.8207 | 33.2 | 0.53 |
| pHILIC | 963.639 | 409.2077 | 33.2 | 0.53 |
| pHILIC | 963.639 | 246.8127 | 33.2 | 0.53 |
| pHILIC | 963.639 | 111.8238 | 33.1 | 0.53 |
| pHILIC | 963.639 | 230.1619 | 33.1 | 0.53 |
| pHILIC | 963.639 | 253.1862 | 33.1 | 0.53 |
| pHILIC | 963.639 | 103.9424 | 33.1 | 0.53 |
| pHILIC | 963.639 | 549.6118 | 33.1 | 0.53 |
| pHILIC | 963.639 | 189.7007 | 33 | 0.53 |
| pHILIC | 963.639 | 395.6276 | 33 | 0.53 |
| pHILIC | 963.639 | 379.6912 | 33 | 0.53 |
| pHILIC | 963.639 | 111.9014 | 33 | 0.53 |
| pHILIC | 963.639 | 315.7827 | 33 | 0.53 |
| pHILIC | 963.639 | 572.5817 | 33 | 0.53 |
| pHILIC | 963.639 | 121.0264 | 33 | 0.53 |
| pHILIC | 963.639 | 129.2293 | 33 | 0.53 |
| pHILIC | 963.639 | 596.4142 | 33 | 0.53 |
| pHILIC | 963.639 | 234.4892 | 33 | 0.53 |
| pHILIC | 963.639 | 200.5311 | 33 | 0.53 |
| pHILIC | 963.639 | 327.1979 | 32.9 | 0.53 |
| pHILIC | 963.639 | 366.0616 | 32.9 | 0.53 |
| pHILIC | 963.639 | 599.6288 | 32.9 | 0.53 |
| pHILIC | 963.639 | 160.1719 | 32.9 | 0.53 |
| pHILIC | 963.639 | 604.2701 | 32.9 | 0.53 |
| pHILIC | 963.639 | 114.5464 | 32.9 | 0.53 |
| pHILIC | 963.639 | 382.0465 | 32.9 | 0.53 |
| pHILIC | 963.639 | 178.6416 | 32.8 | 0.52 |
| pHILIC | 963.639 | 113.3458 | 32.8 | 0.52 |

| | | | | |
|---|---|---|---|---|
| pHILIC | 963.639 | 543.4392 | 32.8 | 0.52 |
| pHILIC | 963.639 | 622.5739 | 32.8 | 0.52 |
| pHILIC | 963.639 | 278.55 | 32.8 | 0.52 |
| pHILIC | 963.639 | 146.2214 | 32.8 | 0.52 |
| pHILIC | 963.639 | 111.9973 | 32.8 | 0.52 |
| pHILIC | 963.639 | 144.1783 | 32.8 | 0.52 |
| pHILIC | 963.639 | 979.3525 | 32.8 | 0.52 |
| pHILIC | 963.639 | 338.5284 | 32.8 | 0.52 |
| pHILIC | 963.639 | 117.4499 | 32.7 | 0.52 |
| pHILIC | 963.639 | 197.4767 | 32.7 | 0.52 |
| pHILIC | 963.639 | 118.0943 | 32.7 | 0.52 |
| pHILIC | 963.639 | 194.3916 | 32.7 | 0.52 |
| pHILIC | 963.639 | 204.7464 | 32.7 | 0.52 |
| pHILIC | 963.639 | 817.2704 | 32.7 | 0.52 |
| pHILIC | 963.639 | 178.198 | 32.7 | 0.52 |
| pHILIC | 963.639 | 145.0863 | 32.7 | 0.52 |
| pHILIC | 963.639 | 782.1953 | 32.7 | 0.52 |
| pHILIC | 963.639 | 264.0536 | 32.6 | 0.52 |
| pHILIC | 963.639 | 143.6745 | 32.6 | 0.52 |
| pHILIC | 963.639 | 167.6632 | 32.6 | 0.52 |
| pHILIC | 963.639 | 133.8168 | 32.6 | 0.52 |
| pHILIC | 963.639 | 181.8508 | 32.6 | 0.52 |
| pHILIC | 963.639 | 343.7878 | 32.6 | 0.52 |
| pHILIC | 963.639 | 125.6033 | 32.6 | 0.52 |
| pHILIC | 963.639 | 224.2508 | 32.6 | 0.52 |
| pHILIC | 963.639 | 166.9227 | 32.6 | 0.52 |
| pHILIC | 963.639 | 146.4899 | 32.6 | 0.52 |
| pHILIC | 963.639 | 176.8504 | 32.6 | 0.52 |
| pHILIC | 963.639 | 213.6017 | 32.6 | 0.52 |
| pHILIC | 963.639 | 725.8105 | 32.6 | 0.52 |
| pHILIC | 963.639 | 134.5834 | 32.5 | 0.52 |
| pHILIC | 963.639 | 184.2861 | 32.5 | 0.52 |
| pHILIC | 963.639 | 430.0479 | 32.5 | 0.52 |
| pHILIC | 963.639 | 145.7084 | 32.5 | 0.52 |
| pHILIC | 963.639 | 768.8965 | 32.5 | 0.52 |
| pHILIC | 963.639 | 196.2875 | 32.5 | 0.52 |
| pHILIC | 963.639 | 127.5531 | 32.5 | 0.52 |
| pHILIC | 963.639 | 213.2226 | 32.5 | 0.52 |
| pHILIC | 963.639 | 169.2536 | 32.5 | 0.52 |
| pHILIC | 963.639 | 450.0402 | 32.5 | 0.52 |
| pHILIC | 963.639 | 106.5289 | 32.4 | 0.52 |
| pHILIC | 963.639 | 262.1637 | 32.4 | 0.52 |

| | | | | |
|---|---|---|---|---|
| pHILIC | 963.639 | 207.0982 | 32.4 | 0.52 |
| pHILIC | 963.639 | 790.5135 | 32.4 | 0.52 |
| pHILIC | 963.639 | 226.2049 | 32.3 | 0.52 |
| pHILIC | 963.639 | 337.4237 | 32.3 | 0.52 |
| pHILIC | 963.639 | 349.5069 | 32.3 | 0.52 |
| pHILIC | 963.639 | 149.0247 | 32.3 | 0.52 |
| pHILIC | 963.639 | 111.1367 | 32.3 | 0.52 |
| pHILIC | 963.639 | 159.662 | 32.3 | 0.52 |
| pHILIC | 963.639 | 123.3729 | 32.3 | 0.52 |
| pHILIC | 963.639 | 132.1742 | 32.3 | 0.52 |
| pHILIC | 963.639 | 268.5464 | 32.3 | 0.52 |
| pHILIC | 963.639 | 862.9946 | 32.3 | 0.52 |
| pHILIC | 963.639 | 497.4376 | 32.3 | 0.52 |
| pHILIC | 963.639 | 268.4326 | 32.2 | 0.51 |
| pHILIC | 963.639 | 516.6314 | 32.2 | 0.51 |
| pHILIC | 963.639 | 767.2427 | 32.2 | 0.51 |
| pHILIC | 963.639 | 362.479 | 32.2 | 0.51 |
| pHILIC | 963.639 | 115.3608 | 32.2 | 0.51 |
| pHILIC | 963.639 | 289.7281 | 32.2 | 0.51 |
| pHILIC | 963.639 | 285.7199 | 32.1 | 0.51 |
| pHILIC | 963.639 | 131.3797 | 32.1 | 0.51 |
| pHILIC | 963.639 | 112.9825 | 32.1 | 0.51 |
| pHILIC | 963.639 | 574.5178 | 32.1 | 0.51 |
| pHILIC | 963.639 | 192.9133 | 32.1 | 0.51 |
| pHILIC | 963.639 | 254.6266 | 32 | 0.51 |
| pHILIC | 963.639 | 494.059 | 32 | 0.51 |
| pHILIC | 963.639 | 261.6784 | 32 | 0.51 |
| pHILIC | 963.639 | 134.0041 | 32 | 0.51 |
| pHILIC | 963.639 | 617.4461 | 32 | 0.51 |
| pHILIC | 963.639 | 153.5011 | 32 | 0.51 |
| pHILIC | 963.639 | 130.9371 | 32 | 0.51 |
| pHILIC | 963.639 | 120.858 | 32 | 0.51 |
| pHILIC | 963.639 | 113.4383 | 32 | 0.51 |
| pHILIC | 963.639 | 405.725 | 32 | 0.51 |
| pHILIC | 963.639 | 149.8782 | 31.9 | 0.51 |
| pHILIC | 963.639 | 126.7409 | 31.9 | 0.51 |
| pHILIC | 963.639 | 121.3095 | 31.9 | 0.51 |
| pHILIC | 963.639 | 239.1105 | 31.9 | 0.51 |
| pHILIC | 963.639 | 123.4231 | 31.9 | 0.51 |
| pHILIC | 963.639 | 402.1013 | 31.9 | 0.51 |
| pHILIC | 963.639 | 321.8768 | 31.9 | 0.51 |
| pHILIC | 963.639 | 153.4171 | 31.9 | 0.51 |

| | | | | |
|---|---|---|---|---|
| pHILIC | 963.639 | 125.6586 | 31.9 | 0.51 |
| pHILIC | 963.639 | 334.2752 | 31.8 | 0.51 |
| pHILIC | 963.639 | 223.132 | 31.8 | 0.51 |
| pHILIC | 963.639 | 124.7594 | 31.8 | 0.51 |
| pHILIC | 963.639 | 110.6378 | 31.8 | 0.51 |
| pHILIC | 963.639 | 116.05 | 31.8 | 0.51 |
| pHILIC | 963.639 | 391.4376 | 31.8 | 0.51 |
| pHILIC | 963.639 | 184.1188 | 31.7 | 0.51 |
| pHILIC | 963.639 | 466.6016 | 31.7 | 0.51 |
| pHILIC | 963.639 | 149.5482 | 31.7 | 0.51 |
| pHILIC | 963.639 | 199.2469 | 31.7 | 0.51 |
| pHILIC | 963.639 | 710.7642 | 31.7 | 0.51 |
| pHILIC | 963.639 | 250.633 | 31.7 | 0.51 |
| pHILIC | 963.639 | 558.316 | 31.7 | 0.51 |
| pHILIC | 963.639 | 178.181 | 31.7 | 0.51 |
| pHILIC | 963.639 | 142.2318 | 31.7 | 0.51 |
| pHILIC | 963.639 | 369.7692 | 31.7 | 0.51 |
| pHILIC | 963.639 | 298.781 | 31.7 | 0.51 |
| pHILIC | 963.639 | 324.2993 | 31.7 | 0.51 |
| pHILIC | 963.639 | 116.8613 | 31.7 | 0.51 |
| pHILIC | 963.639 | 107.2037 | 31.7 | 0.51 |
| pHILIC | 963.639 | 336.6821 | 31.7 | 0.51 |
| pHILIC | 963.639 | 101.4709 | 31.7 | 0.51 |
| pHILIC | 963.639 | 133.7285 | 31.7 | 0.51 |
| pHILIC | 963.639 | 462.2982 | 31.7 | 0.51 |
| pHILIC | 963.639 | 290.0948 | 31.6 | 0.51 |
| pHILIC | 963.639 | 132.7393 | 31.6 | 0.51 |
| pHILIC | 963.639 | 163.4833 | 31.6 | 0.51 |
| pHILIC | 963.639 | 186.5543 | 31.6 | 0.51 |
| pHILIC | 963.639 | 146.1626 | 31.6 | 0.5 |
| pHILIC | 963.639 | 162.2347 | 31.6 | 0.5 |
| pHILIC | 963.639 | 137.9838 | 31.6 | 0.5 |
| pHILIC | 963.639 | 740.0475 | 31.6 | 0.5 |
| pHILIC | 963.639 | 102.1576 | 31.5 | 0.5 |
| pHILIC | 963.639 | 296.0506 | 31.5 | 0.5 |
| pHILIC | 963.639 | 114.2241 | 31.5 | 0.5 |
| pHILIC | 963.639 | 733.0981 | 31.5 | 0.5 |
| pHILIC | 963.639 | 126.0349 | 31.5 | 0.5 |
| pHILIC | 963.639 | 141.6765 | 31.5 | 0.5 |
| pHILIC | 963.639 | 160.0451 | 31.5 | 0.5 |
| pHILIC | 963.639 | 167.0274 | 31.5 | 0.5 |
| pHILIC | 963.639 | 195.4704 | 31.5 | 0.5 |

| | | | | |
|---|---|---|---|---|
| pHILIC | 963.639 | 141.2903 | 31.5 | 0.5 |
| pHILIC | 963.639 | 270.324 | 31.5 | 0.5 |
| pHILIC | 963.639 | 421.2076 | 31.5 | 0.5 |
| pHILIC | 963.639 | 224.6888 | 31.5 | 0.5 |
| pHILIC | 963.639 | 191.0471 | 31.4 | 0.5 |
| pHILIC | 963.639 | 124.1024 | 31.4 | 0.5 |
| pHILIC | 963.639 | 237.8748 | 31.4 | 0.5 |
| pHILIC | 963.639 | 237.4055 | 31.4 | 0.5 |
| pHILIC | 963.639 | 816.4188 | 31.4 | 0.5 |
| pHILIC | 963.639 | 258.7926 | 31.4 | 0.5 |
| pHILIC | 963.639 | 232.711 | 31.4 | 0.5 |
| pHILIC | 963.639 | 981.1394 | 31.4 | 0.5 |
| pHILIC | 963.639 | 111.4456 | 31.3 | 0.5 |
| pHILIC | 963.639 | 171.1374 | 31.3 | 0.5 |
| pHILIC | 963.639 | 114.1171 | 31.3 | 0.5 |
| pHILIC | 963.639 | 161.3722 | 31.3 | 0.5 |
| pHILIC | 963.639 | 569.2299 | 31.2 | 0.5 |
| pHILIC | 963.639 | 647.0023 | 31.2 | 0.5 |
| pHILIC | 963.639 | 111.3028 | 31.2 | 0.5 |
| pHILIC | 963.639 | 167.7132 | 31.2 | 0.5 |
| pHILIC | 963.639 | 157.2592 | 31.2 | 0.5 |
| pHILIC | 963.639 | 102.9938 | 31.2 | 0.5 |
| pHILIC | 963.639 | 126.97 | 31.2 | 0.5 |
| pHILIC | 963.639 | 580.92 | 31.1 | 0.5 |
| pHILIC | 963.639 | 131.1835 | 31.1 | 0.5 |
| pHILIC | 963.639 | 133.4222 | 31.1 | 0.5 |
| pHILIC | 963.639 | 124.1862 | 31.1 | 0.5 |
| pHILIC | 963.639 | 121.399 | 31.1 | 0.5 |
| pHILIC | 963.639 | 104.1284 | 31.1 | 0.5 |
| pHILIC | 963.639 | 606.5469 | 31.1 | 0.5 |
| pHILIC | 963.639 | 251.3555 | 31.1 | 0.5 |
| pHILIC | 963.639 | 171.7283 | 31 | 0.5 |
| pHILIC | 963.639 | 501.688 | 31 | 0.5 |
| pHILIC | 963.639 | 116.3848 | 31 | 0.5 |
| pHILIC | 963.639 | 140.7232 | 31 | 0.5 |
| pHILIC | 963.639 | 142.5871 | 31 | 0.5 |
| pHILIC | 963.639 | 555.1303 | 31 | 0.5 |
| pHILIC | 963.639 | 221.1788 | 31 | 0.5 |
| pHILIC | 963.639 | 122.4348 | 31 | 0.5 |
| pHILIC | 963.639 | 250.8995 | 31 | 0.49 |
| pHILIC | 963.639 | 288.3872 | 31 | 0.49 |
| pHILIC | 963.639 | 927.4877 | 31 | 0.49 |

| | | | | |
|---|---|---|---|---|
| pHILIC | 963.639 | 277.6792 | 31 | 0.49 |
| pHILIC | 963.639 | 113.4798 | 31 | 0.49 |
| pHILIC | 963.639 | 884.7968 | 30.9 | 0.49 |
| pHILIC | 963.639 | 118.2147 | 30.9 | 0.49 |
| pHILIC | 963.639 | 936.0561 | 30.9 | 0.49 |
| pHILIC | 963.639 | 208.4394 | 30.9 | 0.49 |
| pHILIC | 963.639 | 279.8751 | 30.9 | 0.49 |
| pHILIC | 963.639 | 884.5383 | 30.9 | 0.49 |
| pHILIC | 963.639 | 455.1099 | 30.9 | 0.49 |
| pHILIC | 963.639 | 430.129 | 30.9 | 0.49 |
| pHILIC | 963.639 | 312.746 | 30.9 | 0.49 |
| pHILIC | 963.639 | 180.629 | 30.9 | 0.49 |
| pHILIC | 963.639 | 130.8248 | 30.9 | 0.49 |
| pHILIC | 963.639 | 416.9202 | 30.8 | 0.49 |
| pHILIC | 963.639 | 115.337 | 30.8 | 0.49 |
| pHILIC | 963.639 | 123.1985 | 30.8 | 0.49 |
| pHILIC | 963.639 | 198.8605 | 30.8 | 0.49 |
| pHILIC | 963.639 | 340.7485 | 30.8 | 0.49 |
| pHILIC | 963.639 | 560.6349 | 30.8 | 0.49 |
| pHILIC | 963.639 | 415.9339 | 30.8 | 0.49 |
| pHILIC | 963.639 | 125.3903 | 30.8 | 0.49 |
| pHILIC | 963.639 | 292.7104 | 30.8 | 0.49 |
| pHILIC | 963.639 | 599.2717 | 30.8 | 0.49 |
| pHILIC | 963.639 | 150.6562 | 30.8 | 0.49 |
| pHILIC | 963.639 | 159.0256 | 30.8 | 0.49 |
| pHILIC | 963.639 | 817.751 | 30.7 | 0.49 |
| pHILIC | 963.639 | 293.6272 | 30.7 | 0.49 |
| pHILIC | 963.639 | 106.0617 | 30.7 | 0.49 |
| pHILIC | 963.639 | 412.649 | 30.7 | 0.49 |
| pHILIC | 963.639 | 271.3298 | 30.7 | 0.49 |
| pHILIC | 963.639 | 346.7332 | 30.7 | 0.49 |
| pHILIC | 963.639 | 359.5338 | 30.7 | 0.49 |
| pHILIC | 963.639 | 993.0956 | 30.7 | 0.49 |
| pHILIC | 963.639 | 183.8507 | 30.6 | 0.49 |
| pHILIC | 963.639 | 772.8279 | 30.6 | 0.49 |
| pHILIC | 963.639 | 111.0094 | 30.6 | 0.49 |
| pHILIC | 963.639 | 143.4895 | 30.6 | 0.49 |
| pHILIC | 963.639 | 191.2543 | 30.6 | 0.49 |
| pHILIC | 963.639 | 223.5876 | 30.6 | 0.49 |
| pHILIC | 963.639 | 101.8432 | 30.6 | 0.49 |
| pHILIC | 963.639 | 103.4671 | 30.6 | 0.49 |
| pHILIC | 963.639 | 177.1358 | 30.6 | 0.49 |

| | | | | |
|---|---|---|---|---|
| pHILIC | 963.639 | 115.5078 | 30.6 | 0.49 |
| pHILIC | 963.639 | 134.6999 | 30.5 | 0.49 |
| pHILIC | 963.639 | 358.0304 | 30.5 | 0.49 |
| pHILIC | 963.639 | 109.353 | 30.5 | 0.49 |
| pHILIC | 963.639 | 289.1615 | 30.5 | 0.49 |
| pHILIC | 963.639 | 200.7407 | 30.5 | 0.49 |
| pHILIC | 963.639 | 108.849 | 30.5 | 0.49 |
| pHILIC | 963.639 | 122.4805 | 30.5 | 0.49 |
| pHILIC | 963.639 | 148.5587 | 30.5 | 0.49 |
| pHILIC | 963.639 | 191.1303 | 30.5 | 0.49 |
| pHILIC | 963.639 | 123.6554 | 30.5 | 0.49 |
| pHILIC | 963.639 | 106.9221 | 30.5 | 0.49 |
| pHILIC | 963.639 | 376.0193 | 30.5 | 0.49 |
| pHILIC | 963.639 | 122.634 | 30.5 | 0.49 |
| pHILIC | 963.639 | 132.6064 | 30.4 | 0.49 |
| pHILIC | 963.639 | 386.2473 | 30.4 | 0.49 |
| pHILIC | 963.639 | 325.7076 | 30.4 | 0.49 |
| pHILIC | 963.639 | 126.3291 | 30.4 | 0.49 |
| pHILIC | 963.639 | 104.1802 | 30.4 | 0.49 |
| pHILIC | 963.639 | 138.0647 | 30.4 | 0.49 |
| pHILIC | 963.639 | 113.1109 | 30.4 | 0.49 |
| pHILIC | 963.639 | 141.9257 | 30.4 | 0.49 |
| pHILIC | 963.639 | 160.2673 | 30.4 | 0.49 |
| pHILIC | 963.639 | 111.3431 | 30.4 | 0.49 |
| pHILIC | 963.639 | 134.759 | 30.4 | 0.49 |
| pHILIC | 963.639 | 579.7757 | 30.4 | 0.49 |
| pHILIC | 963.639 | 207.133 | 30.4 | 0.49 |
| pHILIC | 963.639 | 532.341 | 30.4 | 0.49 |
| pHILIC | 963.639 | 235.0377 | 30.4 | 0.49 |
| pHILIC | 963.639 | 289.2006 | 30.3 | 0.48 |
| pHILIC | 963.639 | 345.1177 | 30.3 | 0.48 |
| pHILIC | 963.639 | 152.2005 | 30.3 | 0.48 |
| pHILIC | 963.639 | 661.2603 | 30.3 | 0.48 |
| pHILIC | 963.639 | 168.5848 | 30.3 | 0.48 |
| pHILIC | 963.639 | 124.1745 | 30.3 | 0.48 |
| pHILIC | 963.639 | 137.9729 | 30.2 | 0.48 |
| pHILIC | 963.639 | 207.8244 | 30.2 | 0.48 |
| pHILIC | 963.639 | 407.1326 | 30.2 | 0.48 |
| pHILIC | 963.639 | 160.453 | 30.2 | 0.48 |
| pHILIC | 963.639 | 128.7363 | 30.2 | 0.48 |
| pHILIC | 963.639 | 359.0829 | 30.2 | 0.48 |
| pHILIC | 963.639 | 688.0969 | 30.2 | 0.48 |

| | | | | |
|---|---|---|---|---|
| pHILIC | 963.639 | 120.5108 | 30.2 | 0.48 |
| pHILIC | 963.639 | 727.0992 | 30.2 | 0.48 |
| pHILIC | 963.639 | 359.631 | 30.2 | 0.48 |
| pHILIC | 963.639 | 296.8736 | 30.2 | 0.48 |
| pHILIC | 963.639 | 263.0011 | 30.2 | 0.48 |
| pHILIC | 963.639 | 105.6983 | 30.2 | 0.48 |
| pHILIC | 963.639 | 349.8236 | 30.2 | 0.48 |
| pHILIC | 963.639 | 696.7581 | 30.2 | 0.48 |
| pHILIC | 963.639 | 213.4915 | 30.1 | 0.48 |
| pHILIC | 963.639 | 143.2641 | 30.1 | 0.48 |
| pHILIC | 963.639 | 504.9004 | 30.1 | 0.48 |
| pHILIC | 963.639 | 126.4306 | 30.1 | 0.48 |
| pHILIC | 963.639 | 107.9082 | 30.1 | 0.48 |
| pHILIC | 963.639 | 567.8761 | 30.1 | 0.48 |
| pHILIC | 963.639 | 114.6709 | 30.1 | 0.48 |
| pHILIC | 963.639 | 869.9739 | 30.1 | 0.48 |
| pHILIC | 963.639 | 184.5294 | 30.1 | 0.48 |
| pHILIC | 963.639 | 106.3067 | 30.1 | 0.48 |
| pHILIC | 963.639 | 338.1313 | 30.1 | 0.48 |
| pHILIC | 963.639 | 235.1073 | 30.1 | 0.48 |
| pHILIC | 963.639 | 106.6339 | 30.1 | 0.48 |
| pHILIC | 963.639 | 410.0867 | 30.1 | 0.48 |
| pHILIC | 963.639 | 316.2225 | 30.1 | 0.48 |
| pHILIC | 963.639 | 134.8313 | 30.1 | 0.48 |
| pHILIC | 963.639 | 113.473 | 30.1 | 0.48 |
| pHILIC | 963.639 | 261.8821 | 30 | 0.48 |
| pHILIC | 963.639 | 177.535 | 30 | 0.48 |
| pHILIC | 963.639 | 209.8142 | 30 | 0.48 |
| pHILIC | 963.639 | 952.1373 | 30 | 0.48 |
| pHILIC | 963.639 | 291.2835 | 30 | 0.48 |
| pHILIC | 963.639 | 206.8145 | 30 | 0.48 |
| pHILIC | 963.639 | 723.8378 | 30 | 0.48 |
| pHILIC | 963.639 | 440.4246 | 30 | 0.48 |
| pHILIC | 963.639 | 389.1231 | 30 | 0.48 |
| pHILIC | 963.639 | 134.9086 | 30 | 0.48 |
| pHILIC | 963.639 | 339.4625 | 30 | 0.48 |
| pHILIC | 963.639 | 180.9503 | 30 | 0.48 |
| pHILIC | 963.639 | 233.6236 | 29.9 | 0.48 |
| pHILIC | 963.639 | 261.0166 | 29.9 | 0.48 |
| pHILIC | 963.639 | 116.8941 | 29.9 | 0.48 |
| pHILIC | 963.639 | 122.7598 | 29.9 | 0.48 |
| pHILIC | 963.639 | 270.7333 | 29.9 | 0.48 |

| | | | | |
|---|---|---|---|---|
| pHILIC | 963.639 | 233.5574 | 29.9 | 0.48 |
| pHILIC | 963.639 | 237.2605 | 29.9 | 0.48 |
| pHILIC | 963.639 | 466.8444 | 29.9 | 0.48 |
| pHILIC | 963.639 | 107.229 | 29.9 | 0.48 |
| pHILIC | 963.639 | 386.7099 | 29.9 | 0.48 |
| pHILIC | 963.639 | 424.3891 | 29.9 | 0.48 |
| pHILIC | 963.639 | 440.849 | 29.9 | 0.48 |
| pHILIC | 963.639 | 207.2938 | 29.8 | 0.48 |
| pHILIC | 963.639 | 381.0493 | 29.8 | 0.48 |
| pHILIC | 963.639 | 863.3595 | 29.8 | 0.48 |
| pHILIC | 963.639 | 210.9455 | 29.8 | 0.48 |
| pHILIC | 963.639 | 100.8813 | 29.8 | 0.48 |
| pHILIC | 963.639 | 198.452 | 29.8 | 0.48 |
| pHILIC | 963.639 | 235.2347 | 29.8 | 0.48 |
| pHILIC | 963.639 | 279.2136 | 29.7 | 0.48 |
| pHILIC | 963.639 | 384.8201 | 29.7 | 0.47 |
| pHILIC | 963.639 | 128.9925 | 29.7 | 0.47 |
| pHILIC | 963.639 | 213.5744 | 29.7 | 0.47 |
| pHILIC | 963.639 | 260.4208 | 29.7 | 0.47 |
| pHILIC | 963.639 | 167.4267 | 29.7 | 0.47 |
| pHILIC | 963.639 | 322.2691 | 29.7 | 0.47 |
| pHILIC | 963.639 | 131.868 | 29.7 | 0.47 |
| pHILIC | 963.639 | 282.5172 | 29.7 | 0.47 |
| pHILIC | 963.639 | 165.2893 | 29.7 | 0.47 |
| pHILIC | 963.639 | 112.4667 | 29.6 | 0.47 |
| pHILIC | 963.639 | 111.6264 | 29.6 | 0.47 |
| pHILIC | 963.639 | 104.3449 | 29.6 | 0.47 |
| pHILIC | 963.639 | 243.912 | 29.6 | 0.47 |
| pHILIC | 963.639 | 100.8703 | 29.6 | 0.47 |
| pHILIC | 963.639 | 101.5788 | 29.6 | 0.47 |
| pHILIC | 963.639 | 554.7247 | 29.6 | 0.47 |
| pHILIC | 963.639 | 109.146 | 29.6 | 0.47 |
| pHILIC | 963.639 | 203.6911 | 29.6 | 0.47 |
| pHILIC | 963.639 | 124.771 | 29.6 | 0.47 |
| pHILIC | 963.639 | 613.3848 | 29.5 | 0.47 |
| pHILIC | 963.639 | 168.8634 | 29.5 | 0.47 |
| pHILIC | 963.639 | 249.2343 | 29.5 | 0.47 |
| pHILIC | 963.639 | 206.1067 | 29.5 | 0.47 |
| pHILIC | 963.639 | 529.4039 | 29.5 | 0.47 |
| pHILIC | 963.639 | 112.7964 | 29.5 | 0.47 |
| pHILIC | 963.639 | 242.4828 | 29.5 | 0.47 |
| pHILIC | 963.639 | 411.6652 | 29.5 | 0.47 |

| | | | | |
|---|---|---|---|---|
| pHILIC | 963.639 | 149.1628 | 29.5 | 0.47 |
| pHILIC | 963.639 | 149.3992 | 29.5 | 0.47 |
| pHILIC | 963.639 | 151.8265 | 29.5 | 0.47 |
| pHILIC | 963.639 | 196.3203 | 29.5 | 0.47 |
| pHILIC | 963.639 | 132.2744 | 29.4 | 0.47 |
| pHILIC | 963.639 | 170.616 | 29.4 | 0.47 |
| pHILIC | 963.639 | 412.0497 | 29.4 | 0.47 |
| pHILIC | 963.639 | 287.796 | 29.4 | 0.47 |
| pHILIC | 963.639 | 144.33 | 29.4 | 0.47 |
| pHILIC | 963.639 | 176.5061 | 29.4 | 0.47 |
| pHILIC | 963.639 | 295.8737 | 29.3 | 0.47 |
| pHILIC | 963.639 | 180.0214 | 29.3 | 0.47 |
| pHILIC | 963.639 | 125.5629 | 29.3 | 0.47 |
| pHILIC | 963.639 | 200.5523 | 29.3 | 0.47 |
| pHILIC | 963.639 | 458.4465 | 29.3 | 0.47 |
| pHILIC | 963.639 | 351.3259 | 29.3 | 0.47 |
| pHILIC | 963.639 | 489.2869 | 29.3 | 0.47 |
| pHILIC | 963.639 | 134.9001 | 29.3 | 0.47 |
| pHILIC | 963.639 | 304.2783 | 29.3 | 0.47 |
| pHILIC | 963.639 | 395.2102 | 29.3 | 0.47 |
| pHILIC | 963.639 | 397.8388 | 29.3 | 0.47 |
| pHILIC | 963.639 | 283.9471 | 29.3 | 0.47 |
| pHILIC | 963.639 | 384.4283 | 29.3 | 0.47 |
| pHILIC | 963.639 | 287.2452 | 29.3 | 0.47 |
| pHILIC | 963.639 | 438.2693 | 29.2 | 0.47 |
| pHILIC | 963.639 | 373.339 | 29.2 | 0.47 |
| pHILIC | 963.639 | 185.212 | 29.2 | 0.47 |
| pHILIC | 963.639 | 142.6956 | 29.2 | 0.47 |
| pHILIC | 963.639 | 259.5176 | 29.2 | 0.47 |
| pHILIC | 963.639 | 132.3455 | 29.2 | 0.47 |
| pHILIC | 963.639 | 577.3755 | 29.2 | 0.47 |
| pHILIC | 963.639 | 460.397 | 29.2 | 0.47 |
| pHILIC | 963.639 | 285.3382 | 29.2 | 0.47 |
| pHILIC | 963.639 | 769.9515 | 29.2 | 0.47 |
| pHILIC | 963.639 | 392.1268 | 29.2 | 0.47 |
| pHILIC | 963.639 | 122.4593 | 29.1 | 0.47 |
| pHILIC | 963.639 | 124.6355 | 29.1 | 0.47 |
| pHILIC | 963.639 | 103.8188 | 29.1 | 0.47 |
| pHILIC | 963.639 | 173.03 | 29.1 | 0.47 |
| pHILIC | 963.639 | 220.6123 | 29.1 | 0.47 |
| pHILIC | 963.639 | 602.3895 | 29.1 | 0.47 |
| pHILIC | 963.639 | 160.3715 | 29.1 | 0.46 |

| | | | | |
|---|---|---|---|---|
| pHILIC | 963.639 | 122.0686 | 29.1 | 0.46 |
| pHILIC | 963.639 | 133.2988 | 29.1 | 0.46 |
| pHILIC | 963.639 | 561.3075 | 29.1 | 0.46 |
| pHILIC | 963.639 | 198.4784 | 29.1 | 0.46 |
| pHILIC | 963.639 | 109.5603 | 29 | 0.46 |
| pHILIC | 963.639 | 113.3282 | 29 | 0.46 |
| pHILIC | 963.639 | 174.8471 | 29 | 0.46 |
| pHILIC | 963.639 | 466.492 | 28.9 | 0.46 |
| pHILIC | 963.639 | 202.8734 | 28.9 | 0.46 |
| pHILIC | 963.639 | 233.1596 | 28.9 | 0.46 |
| pHILIC | 963.639 | 127.2224 | 28.8 | 0.46 |
| pHILIC | 963.639 | 202.5925 | 28.8 | 0.46 |
| pHILIC | 963.639 | 302.3006 | 28.8 | 0.46 |
| pHILIC | 963.639 | 171.1788 | 28.8 | 0.46 |
| pHILIC | 963.639 | 247.8888 | 28.8 | 0.46 |
| pHILIC | 963.639 | 220.9867 | 28.8 | 0.46 |
| pHILIC | 963.639 | 507.8419 | 28.8 | 0.46 |
| pHILIC | 963.639 | 110.2436 | 28.8 | 0.46 |
| pHILIC | 963.639 | 117.5788 | 28.8 | 0.46 |
| pHILIC | 963.639 | 427.5335 | 28.7 | 0.46 |
| pHILIC | 963.639 | 903.3493 | 28.7 | 0.46 |
| pHILIC | 963.639 | 413.589 | 28.7 | 0.46 |
| pHILIC | 963.639 | 186.0231 | 28.7 | 0.46 |
| pHILIC | 963.639 | 136.2444 | 28.7 | 0.46 |
| pHILIC | 963.639 | 647.1923 | 28.7 | 0.46 |
| pHILIC | 963.639 | 166.0899 | 28.7 | 0.46 |
| pHILIC | 963.639 | 101.3616 | 28.7 | 0.46 |
| pHILIC | 963.639 | 105.6891 | 28.6 | 0.46 |
| pHILIC | 963.639 | 167.5127 | 28.6 | 0.46 |
| pHILIC | 963.639 | 372.6566 | 28.6 | 0.46 |
| pHILIC | 963.639 | 189.9917 | 28.6 | 0.46 |
| pHILIC | 963.639 | 948.5888 | 28.5 | 0.46 |
| pHILIC | 963.639 | 217.8495 | 28.5 | 0.46 |
| pHILIC | 963.639 | 125.7059 | 28.5 | 0.46 |
| pHILIC | 963.639 | 119.3918 | 28.5 | 0.46 |
| pHILIC | 963.639 | 107.2362 | 28.5 | 0.46 |
| pHILIC | 963.639 | 659.2213 | 28.5 | 0.46 |
| pHILIC | 963.639 | 132.9248 | 28.5 | 0.46 |
| pHILIC | 963.639 | 203.8901 | 28.5 | 0.46 |
| pHILIC | 963.639 | 375.9321 | 28.5 | 0.45 |
| pHILIC | 963.639 | 526.6196 | 28.5 | 0.45 |
| pHILIC | 963.639 | 353.1065 | 28.5 | 0.45 |

| | | | | |
|---|---|---|---|---|
| pHILIC | 963.639 | 105.3353 | 28.4 | 0.45 |
| pHILIC | 963.639 | 101.2323 | 28.4 | 0.45 |
| pHILIC | 963.639 | 268.8568 | 28.4 | 0.45 |
| pHILIC | 963.639 | 105.5523 | 28.4 | 0.45 |
| pHILIC | 963.639 | 788.9123 | 28.4 | 0.45 |
| pHILIC | 963.639 | 101.5151 | 28.4 | 0.45 |
| pHILIC | 963.639 | 220.4947 | 28.4 | 0.45 |
| pHILIC | 963.639 | 179.7001 | 28.4 | 0.45 |
| pHILIC | 963.639 | 162.2768 | 28.4 | 0.45 |
| pHILIC | 963.639 | 128.0985 | 28.4 | 0.45 |
| pHILIC | 963.639 | 102.8847 | 28.4 | 0.45 |
| pHILIC | 963.639 | 429.8665 | 28.4 | 0.45 |
| pHILIC | 963.639 | 111.8337 | 28.4 | 0.45 |
| pHILIC | 963.639 | 113.2397 | 28.4 | 0.45 |
| pHILIC | 963.639 | 177.3299 | 28.3 | 0.45 |
| pHILIC | 963.639 | 840.5007 | 28.3 | 0.45 |
| pHILIC | 963.639 | 709.0541 | 28.3 | 0.45 |
| pHILIC | 963.639 | 136.7984 | 28.3 | 0.45 |
| pHILIC | 963.639 | 115.7618 | 28.3 | 0.45 |
| pHILIC | 963.639 | 122.0227 | 28.3 | 0.45 |
| pHILIC | 963.639 | 293.2171 | 28.3 | 0.45 |
| pHILIC | 963.639 | 228.1108 | 28.3 | 0.45 |
| pHILIC | 963.639 | 262.8976 | 28.2 | 0.45 |
| pHILIC | 963.639 | 184.7107 | 28.2 | 0.45 |
| pHILIC | 963.639 | 153.5186 | 28.2 | 0.45 |
| pHILIC | 963.639 | 711.2132 | 28.2 | 0.45 |
| pHILIC | 963.639 | 138.015 | 28.2 | 0.45 |
| pHILIC | 963.639 | 177.0816 | 28.2 | 0.45 |
| pHILIC | 963.639 | 104.8899 | 28.2 | 0.45 |
| pHILIC | 963.639 | 471.1617 | 28.2 | 0.45 |
| pHILIC | 963.639 | 126.875 | 28.1 | 0.45 |
| pHILIC | 963.639 | 467.0264 | 28.1 | 0.45 |
| pHILIC | 963.639 | 155.1336 | 28.1 | 0.45 |
| pHILIC | 963.639 | 343.5834 | 28.1 | 0.45 |
| pHILIC | 963.639 | 365.5292 | 28 | 0.45 |
| pHILIC | 963.639 | 110.7328 | 28 | 0.45 |
| pHILIC | 963.639 | 324.0767 | 27.9 | 0.45 |
| pHILIC | 963.639 | 256.4326 | 27.9 | 0.45 |
| pHILIC | 963.639 | 186.5847 | 27.9 | 0.45 |
| pHILIC | 963.639 | 116.2689 | 27.9 | 0.45 |
| pHILIC | 963.639 | 121.234 | 27.9 | 0.45 |
| pHILIC | 963.639 | 116.2989 | 27.9 | 0.45 |

| | | | | |
|---|---|---|---|---|
| pHILIC | 963.639 | 110.1796 | 27.9 | 0.45 |
| pHILIC | 963.639 | 215.6667 | 27.9 | 0.45 |
| pHILIC | 963.639 | 101.4942 | 27.9 | 0.45 |
| pHILIC | 963.639 | 164.9268 | 27.8 | 0.44 |
| pHILIC | 963.639 | 170.68 | 27.8 | 0.44 |
| pHILIC | 963.639 | 102.9757 | 27.8 | 0.44 |
| pHILIC | 963.639 | 231.5235 | 27.8 | 0.44 |
| pHILIC | 963.639 | 143.8583 | 27.7 | 0.44 |
| pHILIC | 963.639 | 522.4957 | 27.7 | 0.44 |
| pHILIC | 963.639 | 292.8533 | 27.7 | 0.44 |
| pHILIC | 963.639 | 106.3181 | 27.7 | 0.44 |
| pHILIC | 963.639 | 251.1146 | 27.7 | 0.44 |
| pHILIC | 963.639 | 150.7652 | 27.7 | 0.44 |
| pHILIC | 963.639 | 177.9667 | 27.6 | 0.44 |
| pHILIC | 963.639 | 299.5425 | 27.6 | 0.44 |
| pHILIC | 963.639 | 220.8407 | 27.6 | 0.44 |
| pHILIC | 963.639 | 162.8346 | 27.6 | 0.44 |
| pHILIC | 963.639 | 286.4174 | 27.6 | 0.44 |
| pHILIC | 963.639 | 637.6749 | 27.5 | 0.44 |
| pHILIC | 963.639 | 451.4223 | 27.5 | 0.44 |
| pHILIC | 963.639 | 534.0821 | 27.5 | 0.44 |
| pHILIC | 963.639 | 157.1812 | 27.5 | 0.44 |
| pHILIC | 963.639 | 159.7384 | 27.5 | 0.44 |
| pHILIC | 963.639 | 442.9174 | 27.5 | 0.44 |
| pHILIC | 963.639 | 115.6947 | 27.5 | 0.44 |
| pHILIC | 963.639 | 127.5056 | 27.5 | 0.44 |
| pHILIC | 963.639 | 134.2295 | 27.4 | 0.44 |
| pHILIC | 963.639 | 151.6425 | 27.4 | 0.44 |
| pHILIC | 963.639 | 239.8394 | 27.3 | 0.44 |
| pHILIC | 963.639 | 103.1895 | 27.3 | 0.44 |
| pHILIC | 963.639 | 635.7861 | 27.3 | 0.44 |
| pHILIC | 963.639 | 142.0633 | 27.3 | 0.44 |
| pHILIC | 963.639 | 187.688 | 27.3 | 0.44 |
| pHILIC | 963.639 | 246.9345 | 27.3 | 0.44 |
| pHILIC | 963.639 | 284.0977 | 27.2 | 0.43 |
| pHILIC | 963.639 | 419.7658 | 27.1 | 0.43 |
| pHILIC | 963.639 | 152.192 | 27.1 | 0.43 |
| pHILIC | 963.639 | 310.7913 | 27.1 | 0.43 |
| pHILIC | 963.639 | 152.2598 | 27.1 | 0.43 |
| pHILIC | 963.639 | 134.7125 | 27 | 0.43 |
| pHILIC | 963.639 | 127.4948 | 27 | 0.43 |
| pHILIC | 963.639 | 176.4038 | 26.9 | 0.43 |

| | | | | |
|---|---|---|---|---|
| pHILIC | 963.639 | 199.9333 | 26.9 | 0.43 |
| pHILIC | 963.639 | 100.707 | 26.9 | 0.43 |
| pHILIC | 963.639 | 178.2888 | 26.8 | 0.43 |
| pHILIC | 963.639 | 171.4 | 26.8 | 0.43 |
| pHILIC | 963.639 | 263.3435 | 26.8 | 0.43 |
| pHILIC | 963.639 | 151.6565 | 26.8 | 0.43 |
| pHILIC | 963.639 | 383.77 | 26.7 | 0.43 |
| pHILIC | 963.639 | 518.5049 | 26.7 | 0.43 |
| pHILIC | 963.639 | 151.1847 | 26.7 | 0.43 |
| pHILIC | 963.639 | 738.0997 | 26.6 | 0.43 |
| pHILIC | 963.639 | 219.7133 | 26.6 | 0.43 |
| pHILIC | 963.639 | 804.0789 | 26.5 | 0.42 |
| pHILIC | 963.639 | 452.6236 | 26.5 | 0.42 |
| pHILIC | 963.639 | 154.2263 | 26.5 | 0.42 |
| pHILIC | 963.639 | 161.2174 | 26.4 | 0.42 |
| pHILIC | 963.639 | 154.2661 | 26.4 | 0.42 |
| pHILIC | 963.639 | 345.0022 | 26.4 | 0.42 |
| pHILIC | 963.639 | 104.3696 | 26.3 | 0.42 |
| pHILIC | 963.639 | 244.9172 | 26.3 | 0.42 |
| pHILIC | 963.639 | 106.054 | 26.2 | 0.42 |
| pHILIC | 963.639 | 207.6997 | 26.2 | 0.42 |
| pHILIC | 963.639 | 174.9672 | 26.2 | 0.42 |
| pHILIC | 963.639 | 144.5845 | 26.1 | 0.42 |
| pHILIC | 963.639 | 128.5592 | 26 | 0.42 |
| pHILIC | 963.639 | 129.2867 | 26 | 0.42 |
| pHILIC | 963.639 | 111.2469 | 26 | 0.41 |
| pHILIC | 963.639 | 102.9445 | 25.9 | 0.41 |
| pHILIC | 963.639 | 233.3869 | 25.9 | 0.41 |
| pHILIC | 963.639 | 133.5918 | 25.8 | 0.41 |
| pHILIC | 963.639 | 126.5619 | 25.8 | 0.41 |
| pHILIC | 963.639 | 120.7852 | 25.7 | 0.41 |
| pHILIC | 963.639 | 286.7788 | 25.7 | 0.41 |
| pHILIC | 963.639 | 136.8376 | 25.6 | 0.41 |
| pHILIC | 963.639 | 155.0599 | 25.6 | 0.41 |
| pHILIC | 963.639 | 189.3746 | 25.5 | 0.41 |
| pHILIC | 963.639 | 144.3592 | 25.5 | 0.41 |
| pHILIC | 963.639 | 111.2359 | 25.3 | 0.4 |
| pHILIC | 963.639 | 159.7168 | 25.3 | 0.4 |
| pHILIC | 963.639 | 104.4927 | 25.2 | 0.4 |
| pHILIC | 963.639 | 123.9635 | 25.1 | 0.4 |
| pHILIC | 963.639 | 120.6558 | 25.1 | 0.4 |
| pHILIC | 963.639 | 164.4579 | 24.9 | 0.4 |

| | | | | |
|---|---|---|---|---|
| pHILIC | 963.639 | 185.2607 | 24.8 | 0.4 |
| pHILIC | 963.639 | 166.1981 | 24.8 | 0.4 |
| pHILIC | 963.639 | 112.9743 | 24.7 | 0.4 |
| pHILIC | 963.639 | 113.7882 | 24.4 | 0.39 |
| LC–MS Metho | Precursor m/z | Fragment m/z | Absolute Inter | Relative Intensity (%) |
| polarC18 | 134.019 | 74.0236 | 10718.4 | 100 |
| polarC18 | 134.019 | 134.0414 | 10542.3 | 98.36 |
| polarC18 | 134.019 | 92.9001 | 10242.9 | 95.56 |
| polarC18 | 134.019 | 133.9282 | 9941.5 | 92.75 |
| polarC18 | 134.019 | 88.0392 | 9372.9 | 87.45 |
| polarC18 | 134.019 | 74.9309 | 9275.7 | 86.54 |
| polarC18 | 134.019 | 120.0236 | 5874.2 | 54.8 |
| polarC18 | 134.019 | 111.0233 | 5199.6 | 48.51 |
| polarC18 | 134.019 | 151.9384 | 3905.2 | 36.43 |
| polarC18 | 134.019 | 134.0813 | 3533.7 | 32.97 |
| polarC18 | 134.019 | 116.0342 | 1989.5 | 18.56 |
| polarC18 | 134.019 | 115.9643 | 1773.7 | 16.55 |
| polarC18 | 134.019 | 86.0964 | 1698.9 | 15.85 |
| polarC18 | 134.019 | 102.0131 | 1371.8 | 12.8 |
| polarC18 | 134.019 | 93.0127 | 1194 | 11.14 |
| polarC18 | 134.019 | 88.0755 | 1171 | 10.92 |
| polarC18 | 134.019 | 92.0593 | 849.8 | 7.93 |
| polarC18 | 134.019 | 46.0287 | 810.2 | 7.56 |
| polarC18 | 134.019 | 92.9038 | 642.4 | 5.99 |
| polarC18 | 134.019 | 70.0648 | 532.5 | 4.97 |
| polarC18 | 134.019 | 116.9719 | 524.4 | 4.89 |
| polarC18 | 134.019 | 78.9969 | 474.2 | 4.42 |
| polarC18 | 134.019 | 134.06 | 351.7 | 3.28 |
| polarC18 | 134.019 | 110.9118 | 266.4 | 2.49 |
| polarC18 | 134.019 | 113.9638 | 209.9 | 1.96 |
| polarC18 | 134.019 | 73.028 | 180.7 | 1.69 |
| polarC18 | 134.019 | 74.937 | 166.1 | 1.55 |
| polarC18 | 134.019 | 51.109 | 155.2 | 1.45 |
| polarC18 | 134.019 | 93.46 | 151.4 | 1.41 |
| polarC18 | 134.019 | 87.0435 | 149.5 | 1.39 |
| polarC18 | 134.019 | 134.0707 | 147 | 1.37 |
| polarC18 | 134.019 | 74.0287 | 146.8 | 1.37 |
| polarC18 | 134.019 | 43.6726 | 143.2 | 1.34 |
| polarC18 | 134.019 | 74.0184 | 141.9 | 1.32 |
| polarC18 | 134.019 | 109.0409 | 141.7 | 1.32 |
| polarC18 | 134.019 | 93.7388 | 139.7 | 1.3 |
| polarC18 | 134.019 | 87.1269 | 139.5 | 1.3 |

| | | | | |
|---|---|---|---|---|
| polarC18 | 134.019 | 109.9011 | 139.1 | 1.3 |
| polarC18 | 134.019 | 64.1611 | 139 | 1.3 |
| polarC18 | 134.019 | 116.2489 | 135.9 | 1.27 |
| polarC18 | 134.019 | 57.2805 | 134.4 | 1.25 |
| polarC18 | 134.019 | 85.4876 | 132.2 | 1.23 |
| polarC18 | 134.019 | 88.1031 | 129.1 | 1.2 |
| polarC18 | 134.019 | 45.2172 | 128.7 | 1.2 |
| polarC18 | 134.019 | 126.8343 | 128.4 | 1.2 |
| polarC18 | 134.019 | 76.5563 | 127 | 1.18 |
| polarC18 | 134.019 | 145.4288 | 126.8 | 1.18 |
| polarC18 | 134.019 | 41.3247 | 126.2 | 1.18 |
| polarC18 | 134.019 | 64.0585 | 125.1 | 1.17 |
| polarC18 | 134.019 | 43.135 | 124.2 | 1.16 |
| polarC18 | 134.019 | 87.0998 | 115.6 | 1.08 |
| polarC18 | 134.019 | 65.2919 | 94.3 | 0.88 |
| polarC18 | 134.019 | 91.0586 | 93.6 | 0.87 |
| polarC18 | 134.019 | 57.4991 | 93.5 | 0.87 |
| polarC18 | 134.019 | 122.5378 | 93.4 | 0.87 |
| polarC18 | 134.019 | 71.049 | 92.3 | 0.86 |
| polarC18 | 134.019 | 56.1791 | 91 | 0.85 |
| polarC18 | 134.019 | 52.1053 | 90.6 | 0.85 |
| polarC18 | 134.019 | 46.4219 | 89.4 | 0.83 |
| polarC18 | 134.019 | 123.4231 | 88.7 | 0.83 |
| polarC18 | 134.019 | 60.9128 | 88.6 | 0.83 |
| polarC18 | 134.019 | 85.8212 | 87.6 | 0.82 |
| polarC18 | 134.019 | 49.3374 | 87.6 | 0.82 |
| polarC18 | 134.019 | 94.1373 | 87 | 0.81 |
| polarC18 | 134.019 | 46.865 | 86.6 | 0.81 |
| polarC18 | 134.019 | 83.5035 | 86.5 | 0.81 |
| polarC18 | 134.019 | 94.1513 | 86.4 | 0.81 |
| polarC18 | 134.019 | 82.9087 | 86.3 | 0.81 |
| polarC18 | 134.019 | 119.7056 | 86.2 | 0.8 |
| polarC18 | 134.019 | 112.8318 | 86.1 | 0.8 |
| polarC18 | 134.019 | 67.2226 | 86 | 0.8 |
| polarC18 | 134.019 | 40.7053 | 85.3 | 0.8 |
| polarC18 | 134.019 | 94.2562 | 85.3 | 0.8 |
| polarC18 | 134.019 | 80.372 | 84.4 | 0.79 |
| polarC18 | 134.019 | 115.9566 | 84.3 | 0.79 |
| polarC18 | 134.019 | 58.7549 | 83.9 | 0.78 |
| polarC18 | 134.019 | 76.6841 | 83.7 | 0.78 |
| polarC18 | 134.019 | 42.008 | 83.5 | 0.78 |
| polarC18 | 134.019 | 88.4242 | 83.4 | 0.78 |

| | | | | |
|---|---|---|---|---|
| polarC18 | 134.019 | 55.8301 | 83.4 | 0.78 |
| polarC18 | 134.019 | 43.8447 | 83.3 | 0.78 |
| polarC18 | 134.019 | 114.9987 | 82.7 | 0.77 |
| polarC18 | 134.019 | 120.7424 | 82.3 | 0.77 |
| polarC18 | 134.019 | 63.1718 | 82.2 | 0.77 |
| polarC18 | 134.019 | 47.0701 | 82.2 | 0.77 |
| polarC18 | 134.019 | 66.023 | 82.2 | 0.77 |
| polarC18 | 134.019 | 145.954 | 82 | 0.77 |
| polarC18 | 134.019 | 155.5349 | 81.8 | 0.76 |
| polarC18 | 134.019 | 63.3712 | 81.8 | 0.76 |
| polarC18 | 134.019 | 74.3265 | 81.7 | 0.76 |
| polarC18 | 134.019 | 91.3658 | 81.6 | 0.76 |
| polarC18 | 134.019 | 42.3669 | 81.4 | 0.76 |
| polarC18 | 134.019 | 114.0646 | 81.4 | 0.76 |
| polarC18 | 134.019 | 94.7247 | 81.3 | 0.76 |
| polarC18 | 134.019 | 93.0187 | 81.1 | 0.76 |
| polarC18 | 134.019 | 99.4203 | 80.8 | 0.75 |
| polarC18 | 134.019 | 43.51 | 80.7 | 0.75 |
| polarC18 | 134.019 | 80.5155 | 80.7 | 0.75 |
| polarC18 | 134.019 | 85.8099 | 80.5 | 0.75 |
| polarC18 | 134.019 | 42.8076 | 80.5 | 0.75 |
| polarC18 | 134.019 | 154.0462 | 80.3 | 0.75 |
| polarC18 | 134.019 | 99.8378 | 80 | 0.75 |
| polarC18 | 134.019 | 131.1119 | 80 | 0.75 |
| polarC18 | 134.019 | 101.8845 | 80 | 0.75 |
| polarC18 | 134.019 | 56.7441 | 79.9 | 0.75 |
| polarC18 | 134.019 | 84.6814 | 79.8 | 0.74 |
| polarC18 | 134.019 | 43.8799 | 79.8 | 0.74 |
| polarC18 | 134.019 | 55.5046 | 79.7 | 0.74 |
| polarC18 | 134.019 | 131.8291 | 79.7 | 0.74 |
| polarC18 | 134.019 | 88.0936 | 79.6 | 0.74 |
| polarC18 | 134.019 | 57.7112 | 79.6 | 0.74 |
| polarC18 | 134.019 | 59.913 | 79.6 | 0.74 |
| polarC18 | 134.019 | 41.3014 | 79.6 | 0.74 |
| polarC18 | 134.019 | 123.53 | 79.6 | 0.74 |
| polarC18 | 134.019 | 57.949 | 79.4 | 0.74 |
| polarC18 | 134.019 | 56.4061 | 79.2 | 0.74 |
| polarC18 | 134.019 | 148.0561 | 79.1 | 0.74 |
| polarC18 | 134.019 | 60.227 | 79.1 | 0.74 |
| polarC18 | 134.019 | 91.6449 | 78.9 | 0.74 |
| polarC18 | 134.019 | 79.0952 | 78.8 | 0.73 |
| polarC18 | 134.019 | 75.9362 | 78.4 | 0.73 |

| | | | | |
|---|---|---|---|---|
| polarC18 | 134.019 | 41.5718 | 78.4 | 0.73 |
| polarC18 | 134.019 | 79.6914 | 78.3 | 0.73 |
| polarC18 | 134.019 | 88.58 | 78.2 | 0.73 |
| polarC18 | 134.019 | 45.8793 | 78.1 | 0.73 |
| polarC18 | 134.019 | 93.6878 | 78 | 0.73 |
| polarC18 | 134.019 | 74.4355 | 78 | 0.73 |
| polarC18 | 134.019 | 119.329 | 77.9 | 0.73 |
| polarC18 | 134.019 | 113.0231 | 77.7 | 0.73 |
| polarC18 | 134.019 | 81.2886 | 77.7 | 0.72 |
| polarC18 | 134.019 | 118.0325 | 77.7 | 0.72 |
| polarC18 | 134.019 | 81.3154 | 77.6 | 0.72 |
| polarC18 | 134.019 | 67.1017 | 77.5 | 0.72 |
| polarC18 | 134.019 | 73.3496 | 77.4 | 0.72 |
| polarC18 | 134.019 | 141.9218 | 77.3 | 0.72 |
| polarC18 | 134.019 | 73.52 | 77.2 | 0.72 |
| polarC18 | 134.019 | 40.0986 | 77 | 0.72 |
| polarC18 | 134.019 | 130.4791 | 76.8 | 0.72 |
| polarC18 | 134.019 | 115.2269 | 76.8 | 0.72 |
| polarC18 | 134.019 | 86.9524 | 76.7 | 0.72 |
| polarC18 | 134.019 | 118.2435 | 76.6 | 0.72 |
| polarC18 | 134.019 | 60.9862 | 76.6 | 0.71 |
| polarC18 | 134.019 | 45.3138 | 76.4 | 0.71 |
| polarC18 | 134.019 | 97.9907 | 76.4 | 0.71 |
| polarC18 | 134.019 | 142.1943 | 76.4 | 0.71 |
| polarC18 | 134.019 | 105.8816 | 76.4 | 0.71 |
| polarC18 | 134.019 | 59.5827 | 76.3 | 0.71 |
| polarC18 | 134.019 | 66.4407 | 76.2 | 0.71 |
| polarC18 | 134.019 | 47.8915 | 76.2 | 0.71 |
| polarC18 | 134.019 | 46.0911 | 76.1 | 0.71 |
| polarC18 | 134.019 | 47.2409 | 76 | 0.71 |
| polarC18 | 134.019 | 65.1331 | 76 | 0.71 |
| polarC18 | 134.019 | 107.4844 | 76 | 0.71 |
| polarC18 | 134.019 | 55.3114 | 75.9 | 0.71 |
| polarC18 | 134.019 | 42.1412 | 75.9 | 0.71 |
| polarC18 | 134.019 | 83.6936 | 75.9 | 0.71 |
| polarC18 | 134.019 | 93.3631 | 75.8 | 0.71 |
| polarC18 | 134.019 | 52.0947 | 75.8 | 0.71 |
| polarC18 | 134.019 | 80.7525 | 75.8 | 0.71 |
| polarC18 | 134.019 | 125.0155 | 75.7 | 0.71 |
| polarC18 | 134.019 | 99.6632 | 75.7 | 0.71 |
| polarC18 | 134.019 | 78.3994 | 75.7 | 0.71 |
| polarC18 | 134.019 | 63.2639 | 75.7 | 0.71 |

| | | | | |
|---|---|---|---|---|
| polarC18 | 134.019 | 68.1235 | 75.7 | 0.71 |
| polarC18 | 134.019 | 101.1802 | 75.6 | 0.71 |
| polarC18 | 134.019 | 51.1324 | 75.5 | 0.7 |
| polarC18 | 134.019 | 55.4045 | 75.4 | 0.7 |
| polarC18 | 134.019 | 44.5163 | 75.3 | 0.7 |
| polarC18 | 134.019 | 46.295 | 75.3 | 0.7 |
| polarC18 | 134.019 | 42.1129 | 75 | 0.7 |
| polarC18 | 134.019 | 56.7999 | 75 | 0.7 |
| polarC18 | 134.019 | 63.5927 | 75 | 0.7 |
| polarC18 | 134.019 | 110.1351 | 75 | 0.7 |
| polarC18 | 134.019 | 112.2915 | 74.8 | 0.7 |
| polarC18 | 134.019 | 47.7303 | 74.8 | 0.7 |
| polarC18 | 134.019 | 152.8407 | 74.7 | 0.7 |
| polarC18 | 134.019 | 85.0944 | 74.3 | 0.69 |
| polarC18 | 134.019 | 97.0291 | 74.2 | 0.69 |
| polarC18 | 134.019 | 64.4425 | 74.2 | 0.69 |
| polarC18 | 134.019 | 133.0999 | 74.2 | 0.69 |
| polarC18 | 134.019 | 65.4638 | 74.1 | 0.69 |
| polarC18 | 134.019 | 86.6636 | 74 | 0.69 |
| polarC18 | 134.019 | 72.9725 | 74 | 0.69 |
| polarC18 | 134.019 | 86.1041 | 73.9 | 0.69 |
| polarC18 | 134.019 | 122.9867 | 73.8 | 0.69 |
| polarC18 | 134.019 | 94.1237 | 73.7 | 0.69 |
| polarC18 | 134.019 | 115.5004 | 73.7 | 0.69 |
| polarC18 | 134.019 | 51.2319 | 73.7 | 0.69 |
| polarC18 | 134.019 | 41.6712 | 73.6 | 0.69 |
| polarC18 | 134.019 | 60.3851 | 73.6 | 0.69 |
| polarC18 | 134.019 | 41.0988 | 73.6 | 0.69 |
| polarC18 | 134.019 | 40.3344 | 73.6 | 0.69 |
| polarC18 | 134.019 | 87.4861 | 73.6 | 0.69 |
| polarC18 | 134.019 | 89.5868 | 73.4 | 0.69 |
| polarC18 | 134.019 | 101.9816 | 73.4 | 0.68 |
| polarC18 | 134.019 | 110.4466 | 73.4 | 0.68 |
| polarC18 | 134.019 | 80.2164 | 73.4 | 0.68 |
| polarC18 | 134.019 | 117.9933 | 73.4 | 0.68 |
| polarC18 | 134.019 | 58.1497 | 73.3 | 0.68 |
| polarC18 | 134.019 | 135.8855 | 73.3 | 0.68 |
| polarC18 | 134.019 | 82.4786 | 73.2 | 0.68 |
| polarC18 | 134.019 | 151.1779 | 73.1 | 0.68 |
| polarC18 | 134.019 | 63.9274 | 73.1 | 0.68 |
| polarC18 | 134.019 | 55.9504 | 73.1 | 0.68 |
| polarC18 | 134.019 | 87.9033 | 73 | 0.68 |

| | | | | |
|---|---|---|---|---|
| polarC18 | 134.019 | 55.1627 | 73 | 0.68 |
| polarC18 | 134.019 | 44.7979 | 73 | 0.68 |
| polarC18 | 134.019 | 67.0155 | 72.9 | 0.68 |
| polarC18 | 134.019 | 45.0395 | 72.8 | 0.68 |
| polarC18 | 134.019 | 149.5291 | 72.8 | 0.68 |
| polarC18 | 134.019 | 45.7569 | 72.8 | 0.68 |
| polarC18 | 134.019 | 43.9159 | 72.8 | 0.68 |
| polarC18 | 134.019 | 43.6932 | 72.7 | 0.68 |
| polarC18 | 134.019 | 79.4846 | 72.7 | 0.68 |
| polarC18 | 134.019 | 49.5963 | 72.7 | 0.68 |
| polarC18 | 134.019 | 58.8678 | 72.7 | 0.68 |
| polarC18 | 134.019 | 86.1937 | 72.7 | 0.68 |
| polarC18 | 134.019 | 153.2202 | 72.7 | 0.68 |
| polarC18 | 134.019 | 60.9465 | 72.7 | 0.68 |
| polarC18 | 134.019 | 145.2947 | 72.6 | 0.68 |
| polarC18 | 134.019 | 116.4842 | 72.4 | 0.68 |
| polarC18 | 134.019 | 70.1457 | 72.4 | 0.68 |
| polarC18 | 134.019 | 44.4579 | 72.4 | 0.68 |
| polarC18 | 134.019 | 57.8723 | 72.3 | 0.67 |
| polarC18 | 134.019 | 52.4838 | 72.3 | 0.67 |
| polarC18 | 134.019 | 87.5302 | 72.2 | 0.67 |
| polarC18 | 134.019 | 87.1204 | 72.2 | 0.67 |
| polarC18 | 134.019 | 131.9253 | 72.2 | 0.67 |
| polarC18 | 134.019 | 47.8579 | 72.1 | 0.67 |
| polarC18 | 134.019 | 104.9495 | 72.1 | 0.67 |
| polarC18 | 134.019 | 46.3203 | 72.1 | 0.67 |
| polarC18 | 134.019 | 135.7241 | 72 | 0.67 |
| polarC18 | 134.019 | 46.9748 | 71.9 | 0.67 |
| polarC18 | 134.019 | 50.0533 | 71.8 | 0.67 |
| polarC18 | 134.019 | 114.1838 | 71.7 | 0.67 |
| polarC18 | 134.019 | 58.3873 | 71.7 | 0.67 |
| polarC18 | 134.019 | 44.5813 | 71.7 | 0.67 |
| polarC18 | 134.019 | 51.0834 | 71.6 | 0.67 |
| polarC18 | 134.019 | 60.2742 | 71.6 | 0.67 |
| polarC18 | 134.019 | 89.0825 | 71.6 | 0.67 |
| polarC18 | 134.019 | 152.9218 | 71.5 | 0.67 |
| polarC18 | 134.019 | 42.2748 | 71.5 | 0.67 |
| polarC18 | 134.019 | 60.4431 | 71.5 | 0.67 |
| polarC18 | 134.019 | 93.8848 | 71.3 | 0.67 |
| polarC18 | 134.019 | 143.5404 | 71.3 | 0.67 |
| polarC18 | 134.019 | 118.5998 | 71.3 | 0.67 |
| polarC18 | 134.019 | 44.877 | 71.2 | 0.66 |

| | | | | |
|---|---|---|---|---|
| polarC18 | 134.019 | 64.2671 | 71.2 | 0.66 |
| polarC18 | 134.019 | 84.0771 | 71.2 | 0.66 |
| polarC18 | 134.019 | 131.4319 | 71.2 | 0.66 |
| polarC18 | 134.019 | 134.7123 | 71.2 | 0.66 |
| polarC18 | 134.019 | 145.5424 | 71.1 | 0.66 |
| polarC18 | 134.019 | 99.2916 | 71.1 | 0.66 |
| polarC18 | 134.019 | 98.6043 | 71 | 0.66 |
| polarC18 | 134.019 | 82.8866 | 71 | 0.66 |
| polarC18 | 134.019 | 57.3066 | 70.9 | 0.66 |
| polarC18 | 134.019 | 88.5968 | 70.9 | 0.66 |
| polarC18 | 134.019 | 94.646 | 70.9 | 0.66 |
| polarC18 | 134.019 | 67.8291 | 70.9 | 0.66 |
| polarC18 | 134.019 | 41.8915 | 70.8 | 0.66 |
| polarC18 | 134.019 | 52.8737 | 70.7 | 0.66 |
| polarC18 | 134.019 | 105.5473 | 70.7 | 0.66 |
| polarC18 | 134.019 | 75.2018 | 70.7 | 0.66 |
| polarC18 | 134.019 | 139.053 | 70.7 | 0.66 |
| polarC18 | 134.019 | 46.0027 | 70.6 | 0.66 |
| polarC18 | 134.019 | 71.2059 | 70.6 | 0.66 |
| polarC18 | 134.019 | 78.8732 | 70.6 | 0.66 |
| polarC18 | 134.019 | 92.6608 | 70.5 | 0.66 |
| polarC18 | 134.019 | 56.5595 | 70.5 | 0.66 |
| polarC18 | 134.019 | 140.179 | 70.5 | 0.66 |
| polarC18 | 134.019 | 52.0559 | 70.5 | 0.66 |
| polarC18 | 134.019 | 117.1765 | 70.5 | 0.66 |
| polarC18 | 134.019 | 84.4115 | 70.5 | 0.66 |
| polarC18 | 134.019 | 120.2162 | 70.5 | 0.66 |
| polarC18 | 134.019 | 41.3086 | 70.5 | 0.66 |
| polarC18 | 134.019 | 113.9283 | 70.4 | 0.66 |
| polarC18 | 134.019 | 48.1181 | 70.4 | 0.66 |
| polarC18 | 134.019 | 103.4946 | 70.4 | 0.66 |
| polarC18 | 134.019 | 138.8913 | 70.3 | 0.66 |
| polarC18 | 134.019 | 46.8283 | 70.3 | 0.66 |
| polarC18 | 134.019 | 121.1433 | 70.3 | 0.66 |
| polarC18 | 134.019 | 83.0976 | 70.3 | 0.66 |
| polarC18 | 134.019 | 44.6528 | 70.3 | 0.66 |
| polarC18 | 134.019 | 56.6266 | 70.3 | 0.66 |
| polarC18 | 134.019 | 153.5519 | 70.3 | 0.66 |
| polarC18 | 134.019 | 50.6366 | 70.3 | 0.66 |
| polarC18 | 134.019 | 105.8181 | 70.3 | 0.66 |
| polarC18 | 134.019 | 58.3069 | 70.3 | 0.66 |
| polarC18 | 134.019 | 60.5357 | 70.2 | 0.66 |

| | | | | |
|---|---|---|---|---|
| polarC18 | 134.019 | 93.5714 | 70.2 | 0.65 |
| polarC18 | 134.019 | 142.1236 | 70.2 | 0.65 |
| polarC18 | 134.019 | 47.5901 | 70.1 | 0.65 |
| polarC18 | 134.019 | 48.3136 | 70.1 | 0.65 |
| polarC18 | 134.019 | 49.0855 | 70.1 | 0.65 |
| polarC18 | 134.019 | 97.3085 | 70.1 | 0.65 |
| polarC18 | 134.019 | 60.4058 | 70.1 | 0.65 |
| polarC18 | 134.019 | 62.8774 | 70.1 | 0.65 |
| polarC18 | 134.019 | 70.3058 | 70.1 | 0.65 |
| polarC18 | 134.019 | 71.6226 | 70 | 0.65 |
| polarC18 | 134.019 | 114.3982 | 70 | 0.65 |
| polarC18 | 134.019 | 54.3789 | 70 | 0.65 |
| polarC18 | 134.019 | 51.8432 | 70 | 0.65 |
| polarC18 | 134.019 | 92.8592 | 70 | 0.65 |
| polarC18 | 134.019 | 63.3437 | 69.9 | 0.65 |
| polarC18 | 134.019 | 120.9121 | 69.9 | 0.65 |
| polarC18 | 134.019 | 106.152 | 69.9 | 0.65 |
| polarC18 | 134.019 | 50.2224 | 69.8 | 0.65 |
| polarC18 | 134.019 | 79.8256 | 69.8 | 0.65 |
| polarC18 | 134.019 | 156.0397 | 69.7 | 0.65 |
| polarC18 | 134.019 | 110.5538 | 69.7 | 0.65 |
| polarC18 | 134.019 | 51.4648 | 69.7 | 0.65 |
| polarC18 | 134.019 | 64.9976 | 69.7 | 0.65 |
| polarC18 | 134.019 | 56.3375 | 69.6 | 0.65 |
| polarC18 | 134.019 | 95.8694 | 69.6 | 0.65 |
| polarC18 | 134.019 | 57.4188 | 69.6 | 0.65 |
| polarC18 | 134.019 | 89.7693 | 69.6 | 0.65 |
| polarC18 | 134.019 | 48.3237 | 69.6 | 0.65 |
| polarC18 | 134.019 | 106.5247 | 69.5 | 0.65 |
| polarC18 | 134.019 | 59.628 | 69.5 | 0.65 |
| polarC18 | 134.019 | 86.3676 | 69.5 | 0.65 |
| polarC18 | 134.019 | 111.3213 | 69.5 | 0.65 |
| polarC18 | 134.019 | 44.7988 | 69.5 | 0.65 |
| polarC18 | 134.019 | 131.4447 | 69.4 | 0.65 |
| polarC18 | 134.019 | 42.7997 | 69.4 | 0.65 |
| polarC18 | 134.019 | 66.3805 | 69.4 | 0.65 |
| polarC18 | 134.019 | 97.4644 | 69.4 | 0.65 |
| polarC18 | 134.019 | 63.8524 | 69.4 | 0.65 |
| polarC18 | 134.019 | 85.893 | 69.4 | 0.65 |
| polarC18 | 134.019 | 123.0227 | 69.4 | 0.65 |
| polarC18 | 134.019 | 88.0453 | 69.4 | 0.65 |
| polarC18 | 134.019 | 130.0592 | 69.4 | 0.65 |

| | | | | |
|---|---|---|---|---|
| polarC18 | 134.019 | 41.0897 | 69.3 | 0.65 |
| polarC18 | 134.019 | 92.6176 | 69.3 | 0.65 |
| polarC18 | 134.019 | 70.2597 | 69.3 | 0.65 |
| polarC18 | 134.019 | 128.1225 | 69.3 | 0.65 |
| polarC18 | 134.019 | 101.9742 | 69.3 | 0.65 |
| polarC18 | 134.019 | 122.945 | 69.3 | 0.65 |
| polarC18 | 134.019 | 53.9168 | 69.2 | 0.65 |
| polarC18 | 134.019 | 48.9561 | 69.2 | 0.65 |
| polarC18 | 134.019 | 93.445 | 69.2 | 0.65 |
| polarC18 | 134.019 | 44.7726 | 69.2 | 0.65 |
| polarC18 | 134.019 | 62.0041 | 69.2 | 0.65 |
| polarC18 | 134.019 | 66.108 | 69.2 | 0.65 |
| polarC18 | 134.019 | 125.8068 | 69.1 | 0.65 |
| polarC18 | 134.019 | 93.8696 | 69.1 | 0.64 |
| polarC18 | 134.019 | 143.4972 | 69.1 | 0.64 |
| polarC18 | 134.019 | 50.7577 | 69 | 0.64 |
| polarC18 | 134.019 | 89.9463 | 69 | 0.64 |
| polarC18 | 134.019 | 50.2619 | 69 | 0.64 |
| polarC18 | 134.019 | 42.6279 | 69 | 0.64 |
| polarC18 | 134.019 | 62.4433 | 68.9 | 0.64 |
| polarC18 | 134.019 | 151.9566 | 68.9 | 0.64 |
| polarC18 | 134.019 | 58.1023 | 68.9 | 0.64 |
| polarC18 | 134.019 | 90.4139 | 68.9 | 0.64 |
| polarC18 | 134.019 | 50.1017 | 68.9 | 0.64 |
| polarC18 | 134.019 | 134.002 | 68.8 | 0.64 |
| polarC18 | 134.019 | 41.165 | 68.7 | 0.64 |
| polarC18 | 134.019 | 123.0408 | 68.7 | 0.64 |
| polarC18 | 134.019 | 51.2871 | 68.7 | 0.64 |
| polarC18 | 134.019 | 108.833 | 68.7 | 0.64 |
| polarC18 | 134.019 | 100.0623 | 68.7 | 0.64 |
| polarC18 | 134.019 | 152.3519 | 68.7 | 0.64 |
| polarC18 | 134.019 | 52.5311 | 68.6 | 0.64 |
| polarC18 | 134.019 | 51.9151 | 68.6 | 0.64 |
| polarC18 | 134.019 | 47.8503 | 68.6 | 0.64 |
| polarC18 | 134.019 | 138.982 | 68.6 | 0.64 |
| polarC18 | 134.019 | 95.7252 | 68.5 | 0.64 |
| polarC18 | 134.019 | 56.4507 | 68.5 | 0.64 |
| polarC18 | 134.019 | 78.9872 | 68.5 | 0.64 |
| polarC18 | 134.019 | 80.8684 | 68.5 | 0.64 |
| polarC18 | 134.019 | 42.2363 | 68.5 | 0.64 |
| polarC18 | 134.019 | 48.4171 | 68.5 | 0.64 |
| polarC18 | 134.019 | 83.3518 | 68.5 | 0.64 |

| | | | | |
|---|---|---|---|---|
| polarC18 | 134.019 | 105.068 | 68.5 | 0.64 |
| polarC18 | 134.019 | 83.9281 | 68.5 | 0.64 |
| polarC18 | 134.019 | 49.5981 | 68.5 | 0.64 |
| polarC18 | 134.019 | 86.659 | 68.5 | 0.64 |
| polarC18 | 134.019 | 53.7432 | 68.4 | 0.64 |
| polarC18 | 134.019 | 80.1294 | 68.4 | 0.64 |
| polarC18 | 134.019 | 95.7366 | 68.4 | 0.64 |
| polarC18 | 134.019 | 120.5679 | 68.4 | 0.64 |
| polarC18 | 134.019 | 115.392 | 68.4 | 0.64 |
| polarC18 | 134.019 | 85.2477 | 68.4 | 0.64 |
| polarC18 | 134.019 | 40.939 | 68.4 | 0.64 |
| polarC18 | 134.019 | 128.0267 | 68.3 | 0.64 |
| polarC18 | 134.019 | 62.7675 | 68.3 | 0.64 |
| polarC18 | 134.019 | 117.5916 | 68.3 | 0.64 |
| polarC18 | 134.019 | 132.0368 | 68.3 | 0.64 |
| polarC18 | 134.019 | 40.4531 | 68.3 | 0.64 |
| polarC18 | 134.019 | 74.1766 | 68.3 | 0.64 |
| polarC18 | 134.019 | 94.5521 | 68.3 | 0.64 |
| polarC18 | 134.019 | 70.7694 | 68.2 | 0.64 |
| polarC18 | 134.019 | 149.7154 | 68.2 | 0.64 |
| polarC18 | 134.019 | 47.9733 | 68.2 | 0.64 |
| polarC18 | 134.019 | 103.5751 | 68.2 | 0.64 |
| polarC18 | 134.019 | 136.7928 | 68.2 | 0.64 |
| polarC18 | 134.019 | 65.9528 | 68.2 | 0.64 |
| polarC18 | 134.019 | 83.7304 | 68.2 | 0.64 |
| polarC18 | 134.019 | 47.6258 | 68.2 | 0.64 |
| polarC18 | 134.019 | 147.5205 | 68.2 | 0.64 |
| polarC18 | 134.019 | 44.4689 | 68.2 | 0.64 |
| polarC18 | 134.019 | 40.3463 | 68.1 | 0.64 |
| polarC18 | 134.019 | 97.6087 | 68.1 | 0.64 |
| polarC18 | 134.019 | 60.8228 | 68.1 | 0.64 |
| polarC18 | 134.019 | 52.9028 | 68 | 0.63 |
| polarC18 | 134.019 | 94.0544 | 68 | 0.63 |
| polarC18 | 134.019 | 148.7897 | 68 | 0.63 |
| polarC18 | 134.019 | 43.1957 | 68 | 0.63 |
| polarC18 | 134.019 | 93.4 | 68 | 0.63 |
| polarC18 | 134.019 | 65.0789 | 68 | 0.63 |
| polarC18 | 134.019 | 48.1525 | 68 | 0.63 |
| polarC18 | 134.019 | 41.4182 | 67.9 | 0.63 |
| polarC18 | 134.019 | 150.6054 | 67.9 | 0.63 |
| polarC18 | 134.019 | 54.8124 | 67.9 | 0.63 |
| polarC18 | 134.019 | 60.6905 | 67.9 | 0.63 |

| | | | | |
|---|---|---|---|---|
| polarC18 | 134.019 | 67.6843 | 67.9 | 0.63 |
| polarC18 | 134.019 | 92.234 | 67.9 | 0.63 |
| polarC18 | 134.019 | 70.0283 | 67.9 | 0.63 |
| polarC18 | 134.019 | 88.9241 | 67.9 | 0.63 |
| polarC18 | 134.019 | 72.9068 | 67.9 | 0.63 |
| polarC18 | 134.019 | 92.0905 | 67.8 | 0.63 |
| polarC18 | 134.019 | 104.0731 | 67.8 | 0.63 |
| polarC18 | 134.019 | 63.1511 | 67.8 | 0.63 |
| polarC18 | 134.019 | 94.6824 | 67.8 | 0.63 |
| polarC18 | 134.019 | 105.4156 | 67.8 | 0.63 |
| polarC18 | 134.019 | 55.939 | 67.8 | 0.63 |
| polarC18 | 134.019 | 105.9267 | 67.7 | 0.63 |
| polarC18 | 134.019 | 47.2257 | 67.7 | 0.63 |
| polarC18 | 134.019 | 41.4894 | 67.7 | 0.63 |
| polarC18 | 134.019 | 81.1227 | 67.6 | 0.63 |
| polarC18 | 134.019 | 78.4386 | 67.6 | 0.63 |
| polarC18 | 134.019 | 74.0348 | 67.6 | 0.63 |
| polarC18 | 134.019 | 145.6498 | 67.5 | 0.63 |
| polarC18 | 134.019 | 43.2181 | 67.5 | 0.63 |
| polarC18 | 134.019 | 49.6324 | 67.5 | 0.63 |
| polarC18 | 134.019 | 43.1852 | 67.5 | 0.63 |
| polarC18 | 134.019 | 56.9582 | 67.5 | 0.63 |
| polarC18 | 134.019 | 116.6779 | 67.5 | 0.63 |
| polarC18 | 134.019 | 40.9606 | 67.5 | 0.63 |
| polarC18 | 134.019 | 71.372 | 67.4 | 0.63 |
| polarC18 | 134.019 | 122.8557 | 67.4 | 0.63 |
| polarC18 | 134.019 | 61.9587 | 67.4 | 0.63 |
| polarC18 | 134.019 | 117.5845 | 67.4 | 0.63 |
| polarC18 | 134.019 | 138.3331 | 67.4 | 0.63 |
| polarC18 | 134.019 | 58.8382 | 67.4 | 0.63 |
| polarC18 | 134.019 | 133.8958 | 67.3 | 0.63 |
| polarC18 | 134.019 | 120.3084 | 67.3 | 0.63 |
| polarC18 | 134.019 | 132.3065 | 67.3 | 0.63 |
| polarC18 | 134.019 | 73.1023 | 67.3 | 0.63 |
| polarC18 | 134.019 | 48.1465 | 67.3 | 0.63 |
| polarC18 | 134.019 | 66.5153 | 67.3 | 0.63 |
| polarC18 | 134.019 | 47.6139 | 67.3 | 0.63 |
| polarC18 | 134.019 | 48.8803 | 67.3 | 0.63 |
| polarC18 | 134.019 | 54.3764 | 67.3 | 0.63 |
| polarC18 | 134.019 | 78.2821 | 67.2 | 0.63 |
| polarC18 | 134.019 | 106.6726 | 67.2 | 0.63 |
| polarC18 | 134.019 | 71.2764 | 67.2 | 0.63 |

| | | | | |
|---|---|---|---|---|
| polarC18 | 134.019 | 99.9659 | 67.2 | 0.63 |
| polarC18 | 134.019 | 94.9674 | 67.2 | 0.63 |
| polarC18 | 134.019 | 48.4811 | 67.2 | 0.63 |
| polarC18 | 134.019 | 125.8724 | 67.2 | 0.63 |
| polarC18 | 134.019 | 89.0967 | 67.2 | 0.63 |
| polarC18 | 134.019 | 58.8974 | 67.1 | 0.63 |
| polarC18 | 134.019 | 107.5457 | 67.1 | 0.63 |
| polarC18 | 134.019 | 74.1373 | 67.1 | 0.63 |
| polarC18 | 134.019 | 41.2306 | 67.1 | 0.63 |
| polarC18 | 134.019 | 42.6744 | 67 | 0.63 |
| polarC18 | 134.019 | 63.577 | 67 | 0.63 |
| polarC18 | 134.019 | 103.6866 | 67 | 0.62 |
| polarC18 | 134.019 | 60.0291 | 66.9 | 0.62 |
| polarC18 | 134.019 | 138.5444 | 66.9 | 0.62 |
| polarC18 | 134.019 | 133.6962 | 66.9 | 0.62 |
| polarC18 | 134.019 | 44.3087 | 66.8 | 0.62 |
| polarC18 | 134.019 | 72.1891 | 66.8 | 0.62 |
| polarC18 | 134.019 | 61.9476 | 66.8 | 0.62 |
| polarC18 | 134.019 | 88.286 | 66.8 | 0.62 |
| polarC18 | 134.019 | 116.3771 | 66.8 | 0.62 |
| polarC18 | 134.019 | 59.7741 | 66.8 | 0.62 |
| polarC18 | 134.019 | 67.3764 | 66.7 | 0.62 |
| polarC18 | 134.019 | 51.9327 | 66.7 | 0.62 |
| polarC18 | 134.019 | 67.7757 | 66.7 | 0.62 |
| polarC18 | 134.019 | 55.7577 | 66.7 | 0.62 |
| polarC18 | 134.019 | 111.8549 | 66.7 | 0.62 |
| polarC18 | 134.019 | 61.9781 | 66.6 | 0.62 |
| polarC18 | 134.019 | 133.4655 | 66.6 | 0.62 |
| polarC18 | 134.019 | 43.7931 | 66.6 | 0.62 |
| polarC18 | 134.019 | 142.3707 | 66.6 | 0.62 |
| polarC18 | 134.019 | 49.9145 | 66.6 | 0.62 |
| polarC18 | 134.019 | 43.5501 | 66.6 | 0.62 |
| polarC18 | 134.019 | 57.108 | 66.6 | 0.62 |
| polarC18 | 134.019 | 42.8859 | 66.6 | 0.62 |
| polarC18 | 134.019 | 51.8201 | 66.5 | 0.62 |
| polarC18 | 134.019 | 59.4278 | 66.5 | 0.62 |
| polarC18 | 134.019 | 146.4169 | 66.4 | 0.62 |
| polarC18 | 134.019 | 47.1961 | 66.4 | 0.62 |
| polarC18 | 134.019 | 41.1988 | 66.4 | 0.62 |
| polarC18 | 134.019 | 49.54 | 66.4 | 0.62 |
| polarC18 | 134.019 | 58.6746 | 66.4 | 0.62 |
| polarC18 | 134.019 | 132.739 | 66.4 | 0.62 |

| | | | | |
|---|---|---|---|---|
| polarC18 | 134.019 | 129.6067 | 66.4 | 0.62 |
| polarC18 | 134.019 | 64.3569 | 66.4 | 0.62 |
| polarC18 | 134.019 | 44.8144 | 66.3 | 0.62 |
| polarC18 | 134.019 | 59.8968 | 66.3 | 0.62 |
| polarC18 | 134.019 | 55.7473 | 66.3 | 0.62 |
| polarC18 | 134.019 | 151.1305 | 66.3 | 0.62 |
| polarC18 | 134.019 | 40.1583 | 66.3 | 0.62 |
| polarC18 | 134.019 | 58.5537 | 66.3 | 0.62 |
| polarC18 | 134.019 | 59.9348 | 66.3 | 0.62 |
| polarC18 | 134.019 | 66.703 | 66.3 | 0.62 |
| polarC18 | 134.019 | 86.9413 | 66.2 | 0.62 |
| polarC18 | 134.019 | 56.9043 | 66.2 | 0.62 |
| polarC18 | 134.019 | 51.5754 | 66.2 | 0.62 |
| polarC18 | 134.019 | 52.9075 | 66.2 | 0.62 |
| polarC18 | 134.019 | 84.8528 | 66.1 | 0.62 |
| polarC18 | 134.019 | 60.4178 | 66.1 | 0.62 |
| polarC18 | 134.019 | 72.5333 | 66.1 | 0.62 |
| polarC18 | 134.019 | 57.7629 | 66.1 | 0.62 |
| polarC18 | 134.019 | 75.6947 | 66.1 | 0.62 |
| polarC18 | 134.019 | 47.7388 | 66.1 | 0.62 |
| polarC18 | 134.019 | 43.8078 | 66.1 | 0.62 |
| polarC18 | 134.019 | 61.7571 | 66 | 0.62 |
| polarC18 | 134.019 | 47.7649 | 66 | 0.62 |
| polarC18 | 134.019 | 142.4132 | 66 | 0.62 |
| polarC18 | 134.019 | 122.1655 | 66 | 0.62 |
| polarC18 | 134.019 | 48.1551 | 66 | 0.62 |
| polarC18 | 134.019 | 47.1989 | 65.9 | 0.61 |
| polarC18 | 134.019 | 151.3761 | 65.9 | 0.61 |
| polarC18 | 134.019 | 68.1019 | 65.9 | 0.61 |
| polarC18 | 134.019 | 106.5759 | 65.9 | 0.61 |
| polarC18 | 134.019 | 82.7093 | 65.9 | 0.61 |
| polarC18 | 134.019 | 40.9305 | 65.8 | 0.61 |
| polarC18 | 134.019 | 73.1992 | 65.8 | 0.61 |
| polarC18 | 134.019 | 48.6348 | 65.8 | 0.61 |
| polarC18 | 134.019 | 70.9241 | 65.8 | 0.61 |
| polarC18 | 134.019 | 43.4158 | 65.8 | 0.61 |
| polarC18 | 134.019 | 45.4759 | 65.8 | 0.61 |
| polarC18 | 134.019 | 40.6301 | 65.8 | 0.61 |
| polarC18 | 134.019 | 86.8763 | 65.7 | 0.61 |
| polarC18 | 134.019 | 50.5977 | 65.7 | 0.61 |
| polarC18 | 134.019 | 73.9014 | 65.7 | 0.61 |
| polarC18 | 134.019 | 128.5186 | 65.7 | 0.61 |

| | | | | |
|---|---|---|---|---|
| polarC18 | 134.019 | 50.949 | 65.7 | 0.61 |
| polarC18 | 134.019 | 77.1418 | 65.7 | 0.61 |
| polarC18 | 134.019 | 59.6582 | 65.7 | 0.61 |
| polarC18 | 134.019 | 59.2447 | 65.6 | 0.61 |
| polarC18 | 134.019 | 58.9615 | 65.6 | 0.61 |
| polarC18 | 134.019 | 43.6619 | 65.6 | 0.61 |
| polarC18 | 134.019 | 55.7362 | 65.6 | 0.61 |
| polarC18 | 134.019 | 80.822 | 65.6 | 0.61 |
| polarC18 | 134.019 | 122.6076 | 65.6 | 0.61 |
| polarC18 | 134.019 | 93.9712 | 65.6 | 0.61 |
| polarC18 | 134.019 | 45.0479 | 65.6 | 0.61 |
| polarC18 | 134.019 | 77.3787 | 65.5 | 0.61 |
| polarC18 | 134.019 | 115.1735 | 65.5 | 0.61 |
| polarC18 | 134.019 | 124.7573 | 65.4 | 0.61 |
| polarC18 | 134.019 | 99.4477 | 65.4 | 0.61 |
| polarC18 | 134.019 | 73.5078 | 65.4 | 0.61 |
| polarC18 | 134.019 | 63.3861 | 65.4 | 0.61 |
| polarC18 | 134.019 | 78.844 | 65.3 | 0.61 |
| polarC18 | 134.019 | 122.391 | 65.3 | 0.61 |
| polarC18 | 134.019 | 68.525 | 65.3 | 0.61 |
| polarC18 | 134.019 | 110.8673 | 65.3 | 0.61 |
| polarC18 | 134.019 | 107.6753 | 65.3 | 0.61 |
| polarC18 | 134.019 | 40.7722 | 65.2 | 0.61 |
| polarC18 | 134.019 | 42.6689 | 65.2 | 0.61 |
| polarC18 | 134.019 | 54.8961 | 65.2 | 0.61 |
| polarC18 | 134.019 | 59.6223 | 65.1 | 0.61 |
| polarC18 | 134.019 | 146.0703 | 65.1 | 0.61 |
| polarC18 | 134.019 | 40.215 | 65 | 0.61 |
| polarC18 | 134.019 | 60.9948 | 65 | 0.61 |
| polarC18 | 134.019 | 45.5871 | 65 | 0.61 |
| polarC18 | 134.019 | 62.4582 | 65 | 0.61 |
| polarC18 | 134.019 | 74.6711 | 65 | 0.61 |
| polarC18 | 134.019 | 56.0578 | 65 | 0.61 |
| polarC18 | 134.019 | 64.1714 | 64.9 | 0.61 |
| polarC18 | 134.019 | 59.3779 | 64.9 | 0.61 |
| polarC18 | 134.019 | 56.6942 | 64.9 | 0.61 |
| polarC18 | 134.019 | 65.1243 | 64.9 | 0.61 |
| polarC18 | 134.019 | 40.0282 | 64.9 | 0.61 |
| polarC18 | 134.019 | 43.0849 | 64.9 | 0.61 |
| polarC18 | 134.019 | 48.0283 | 64.8 | 0.6 |
| polarC18 | 134.019 | 84.32 | 64.8 | 0.6 |
| polarC18 | 134.019 | 127.5466 | 64.8 | 0.6 |

| | | | | |
|---|---|---|---|---|
| polarC18 | 134.019 | 81.9027 | 64.8 | 0.6 |
| polarC18 | 134.019 | 119.817 | 64.8 | 0.6 |
| polarC18 | 134.019 | 41.3206 | 64.6 | 0.6 |
| polarC18 | 134.019 | 42.5847 | 64.6 | 0.6 |
| polarC18 | 134.019 | 54.2818 | 64.6 | 0.6 |
| polarC18 | 134.019 | 53.4878 | 64.6 | 0.6 |
| polarC18 | 134.019 | 54.8094 | 64.5 | 0.6 |
| polarC18 | 134.019 | 79.211 | 64.5 | 0.6 |
| polarC18 | 134.019 | 45.833 | 64.5 | 0.6 |
| polarC18 | 134.019 | 48.6312 | 64.5 | 0.6 |
| polarC18 | 134.019 | 121.6436 | 64.5 | 0.6 |
| polarC18 | 134.019 | 41.7054 | 64.5 | 0.6 |
| polarC18 | 134.019 | 80.8803 | 64.4 | 0.6 |
| polarC18 | 134.019 | 136.2278 | 64.4 | 0.6 |
| polarC18 | 134.019 | 56.2484 | 64.4 | 0.6 |
| polarC18 | 134.019 | 59.4592 | 64.4 | 0.6 |
| polarC18 | 134.019 | 40.6471 | 64.4 | 0.6 |
| polarC18 | 134.019 | 73.3922 | 64.4 | 0.6 |
| polarC18 | 134.019 | 74.7559 | 64.4 | 0.6 |
| polarC18 | 134.019 | 40.325 | 64.4 | 0.6 |
| polarC18 | 134.019 | 126.1299 | 64.3 | 0.6 |
| polarC18 | 134.019 | 53.1216 | 64.3 | 0.6 |
| polarC18 | 134.019 | 78.6685 | 64.3 | 0.6 |
| polarC18 | 134.019 | 143.5258 | 64.3 | 0.6 |
| polarC18 | 134.019 | 156.9872 | 64.3 | 0.6 |
| polarC18 | 134.019 | 61.0662 | 64.3 | 0.6 |
| polarC18 | 134.019 | 75.0846 | 64.2 | 0.6 |
| polarC18 | 134.019 | 111.5216 | 64.2 | 0.6 |
| polarC18 | 134.019 | 54.0231 | 64.2 | 0.6 |
| polarC18 | 134.019 | 71.0379 | 64.2 | 0.6 |
| polarC18 | 134.019 | 84.2354 | 64.2 | 0.6 |
| polarC18 | 134.019 | 69.1607 | 64.1 | 0.6 |
| polarC18 | 134.019 | 66.1054 | 64.1 | 0.6 |
| polarC18 | 134.019 | 62.2985 | 64.1 | 0.6 |
| polarC18 | 134.019 | 137.2556 | 64.1 | 0.6 |
| polarC18 | 134.019 | 124.2902 | 64.1 | 0.6 |
| polarC18 | 134.019 | 61.131 | 64.1 | 0.6 |
| polarC18 | 134.019 | 65.2658 | 64.1 | 0.6 |
| polarC18 | 134.019 | 40.5147 | 64.1 | 0.6 |
| polarC18 | 134.019 | 64.781 | 64.1 | 0.6 |
| polarC18 | 134.019 | 123.738 | 64 | 0.6 |
| polarC18 | 134.019 | 131.2934 | 64 | 0.6 |

| | | | | |
|---|---|---|---|---|
| polarC18 | 134.019 | 48.055 | 64 | 0.6 |
| polarC18 | 134.019 | 57.3771 | 63.9 | 0.6 |
| polarC18 | 134.019 | 94.167 | 63.9 | 0.6 |
| polarC18 | 134.019 | 90.7318 | 63.9 | 0.6 |
| polarC18 | 134.019 | 51.8266 | 63.9 | 0.6 |
| polarC18 | 134.019 | 80.0458 | 63.8 | 0.6 |
| polarC18 | 134.019 | 65.2574 | 63.8 | 0.6 |
| polarC18 | 134.019 | 103.5096 | 63.8 | 0.59 |
| polarC18 | 134.019 | 89.0257 | 63.7 | 0.59 |
| polarC18 | 134.019 | 60.0235 | 63.7 | 0.59 |
| polarC18 | 134.019 | 130.8087 | 63.7 | 0.59 |
| polarC18 | 134.019 | 60.1319 | 63.7 | 0.59 |
| polarC18 | 134.019 | 43.0505 | 63.7 | 0.59 |
| polarC18 | 134.019 | 47.4856 | 63.7 | 0.59 |
| polarC18 | 134.019 | 44.0929 | 63.7 | 0.59 |
| polarC18 | 134.019 | 133.0795 | 63.6 | 0.59 |
| polarC18 | 134.019 | 48.5082 | 63.6 | 0.59 |
| polarC18 | 134.019 | 78.9702 | 63.6 | 0.59 |
| polarC18 | 134.019 | 141.1088 | 63.5 | 0.59 |
| polarC18 | 134.019 | 78.4978 | 63.5 | 0.59 |
| polarC18 | 134.019 | 45.3225 | 63.5 | 0.59 |
| polarC18 | 134.019 | 55.4571 | 63.5 | 0.59 |
| polarC18 | 134.019 | 111.7071 | 63.5 | 0.59 |
| polarC18 | 134.019 | 141.9795 | 63.5 | 0.59 |
| polarC18 | 134.019 | 66.0689 | 63.4 | 0.59 |
| polarC18 | 134.019 | 96.6767 | 63.4 | 0.59 |
| polarC18 | 134.019 | 150.9763 | 63.3 | 0.59 |
| polarC18 | 134.019 | 65.5266 | 63.3 | 0.59 |
| polarC18 | 134.019 | 137.4078 | 63.3 | 0.59 |
| polarC18 | 134.019 | 43.6145 | 63.3 | 0.59 |
| polarC18 | 134.019 | 125.4512 | 63.3 | 0.59 |
| polarC18 | 134.019 | 73.5115 | 63.3 | 0.59 |
| polarC18 | 134.019 | 82.7404 | 63.3 | 0.59 |
| polarC18 | 134.019 | 63.5501 | 63.3 | 0.59 |
| polarC18 | 134.019 | 55.7968 | 63.3 | 0.59 |
| polarC18 | 134.019 | 56.3722 | 63.2 | 0.59 |
| polarC18 | 134.019 | 124.0416 | 63.2 | 0.59 |
| polarC18 | 134.019 | 73.8861 | 63.2 | 0.59 |
| polarC18 | 134.019 | 44.3449 | 63.2 | 0.59 |
| polarC18 | 134.019 | 56.1745 | 63.1 | 0.59 |
| polarC18 | 134.019 | 41.0026 | 63.1 | 0.59 |
| polarC18 | 134.019 | 85.5711 | 63 | 0.59 |

| | | | | |
|---|---|---|---|---|
| polarC18 | 134.019 | 129.8974 | 62.9 | 0.59 |
| polarC18 | 134.019 | 77.3218 | 62.9 | 0.59 |
| polarC18 | 134.019 | 113.4814 | 62.9 | 0.59 |
| polarC18 | 134.019 | 72.6111 | 62.8 | 0.59 |
| polarC18 | 134.019 | 50.7131 | 62.8 | 0.59 |
| polarC18 | 134.019 | 65.7051 | 62.8 | 0.59 |
| polarC18 | 134.019 | 41.5103 | 62.8 | 0.59 |
| polarC18 | 134.019 | 45.5749 | 62.7 | 0.58 |
| polarC18 | 134.019 | 50.6879 | 62.7 | 0.58 |
| polarC18 | 134.019 | 74.119 | 62.6 | 0.58 |
| polarC18 | 134.019 | 43.4597 | 62.6 | 0.58 |
| polarC18 | 134.019 | 90.9302 | 62.6 | 0.58 |
| polarC18 | 134.019 | 46.3097 | 62.6 | 0.58 |
| polarC18 | 134.019 | 133.8008 | 62.6 | 0.58 |
| polarC18 | 134.019 | 46.7118 | 62.6 | 0.58 |
| polarC18 | 134.019 | 42.4267 | 62.5 | 0.58 |
| polarC18 | 134.019 | 86.6356 | 62.5 | 0.58 |
| polarC18 | 134.019 | 104.3118 | 62.5 | 0.58 |
| polarC18 | 134.019 | 41.5074 | 62.4 | 0.58 |
| polarC18 | 134.019 | 45.7134 | 62.4 | 0.58 |
| polarC18 | 134.019 | 45.0887 | 62.4 | 0.58 |
| polarC18 | 134.019 | 58.1636 | 62.4 | 0.58 |
| polarC18 | 134.019 | 45.6469 | 62.3 | 0.58 |
| polarC18 | 134.019 | 84.2163 | 62.3 | 0.58 |
| polarC18 | 134.019 | 75.0019 | 62.3 | 0.58 |
| polarC18 | 134.019 | 110.0272 | 62.2 | 0.58 |
| polarC18 | 134.019 | 94.1429 | 62.2 | 0.58 |
| polarC18 | 134.019 | 42.7645 | 62.2 | 0.58 |
| polarC18 | 134.019 | 76.61 | 62.2 | 0.58 |
| polarC18 | 134.019 | 133.5914 | 62.1 | 0.58 |
| polarC18 | 134.019 | 41.3266 | 62.1 | 0.58 |
| polarC18 | 134.019 | 50.4836 | 62 | 0.58 |
| polarC18 | 134.019 | 104.1121 | 62 | 0.58 |
| polarC18 | 134.019 | 74.0684 | 61.9 | 0.58 |
| polarC18 | 134.019 | 61.0628 | 61.9 | 0.58 |
| polarC18 | 134.019 | 92.6039 | 61.9 | 0.58 |
| polarC18 | 134.019 | 43.9666 | 61.8 | 0.58 |
| polarC18 | 134.019 | 48.2452 | 61.8 | 0.58 |
| polarC18 | 134.019 | 45.1075 | 61.8 | 0.58 |
| polarC18 | 134.019 | 117.6816 | 61.7 | 0.58 |
| polarC18 | 134.019 | 112.1764 | 61.7 | 0.58 |
| polarC18 | 134.019 | 49.1103 | 61.7 | 0.58 |

| | | | | |
|---|---|---|---|---|
| polarC18 | 134.019 | 138.5556 | 61.6 | 0.58 |
| polarC18 | 134.019 | 87.3976 | 61.6 | 0.58 |
| polarC18 | 134.019 | 45.2051 | 61.6 | 0.57 |
| polarC18 | 134.019 | 53.8944 | 61.5 | 0.57 |
| polarC18 | 134.019 | 53.9657 | 61.5 | 0.57 |
| polarC18 | 134.019 | 121.1951 | 61.5 | 0.57 |
| polarC18 | 134.019 | 98.0619 | 61.4 | 0.57 |
| polarC18 | 134.019 | 78.4629 | 61.4 | 0.57 |
| polarC18 | 134.019 | 61.534 | 61.4 | 0.57 |
| polarC18 | 134.019 | 97.2151 | 61.3 | 0.57 |
| polarC18 | 134.019 | 69.9544 | 61.1 | 0.57 |
| polarC18 | 134.019 | 51.4409 | 61.1 | 0.57 |
| polarC18 | 134.019 | 133.3936 | 61.1 | 0.57 |
| polarC18 | 134.019 | 57.9399 | 61.1 | 0.57 |
| polarC18 | 134.019 | 131.5262 | 61 | 0.57 |
| polarC18 | 134.019 | 136.9491 | 61 | 0.57 |
| polarC18 | 134.019 | 43.8478 | 61 | 0.57 |
| polarC18 | 134.019 | 49.9717 | 60.9 | 0.57 |
| polarC18 | 134.019 | 126.7689 | 60.9 | 0.57 |
| polarC18 | 134.019 | 48.8897 | 60.8 | 0.57 |
| polarC18 | 134.019 | 43.8722 | 60.8 | 0.57 |
| polarC18 | 134.019 | 49.5705 | 60.8 | 0.57 |
| polarC18 | 134.019 | 40.481 | 60.7 | 0.57 |
| polarC18 | 134.019 | 51.1732 | 60.7 | 0.57 |
| polarC18 | 134.019 | 48.5962 | 60.7 | 0.57 |
| polarC18 | 134.019 | 62.4701 | 60.7 | 0.57 |
| polarC18 | 134.019 | 41.4027 | 60.7 | 0.57 |
| polarC18 | 134.019 | 130.9021 | 60.7 | 0.57 |
| polarC18 | 134.019 | 46.7557 | 60.6 | 0.57 |
| polarC18 | 134.019 | 48.8363 | 60.6 | 0.57 |
| polarC18 | 134.019 | 57.2677 | 60.5 | 0.56 |
| polarC18 | 134.019 | 59.1603 | 60.5 | 0.56 |
| polarC18 | 134.019 | 45.6091 | 60.3 | 0.56 |
| polarC18 | 134.019 | 47.2713 | 60.3 | 0.56 |
| polarC18 | 134.019 | 46.5914 | 60.2 | 0.56 |
| polarC18 | 134.019 | 51.3141 | 60.2 | 0.56 |
| polarC18 | 134.019 | 41.1889 | 60.1 | 0.56 |
| polarC18 | 134.019 | 74.797 | 60 | 0.56 |
| polarC18 | 134.019 | 41.3409 | 60 | 0.56 |
| polarC18 | 134.019 | 42.032 | 60 | 0.56 |
| polarC18 | 134.019 | 49.4927 | 59.9 | 0.56 |
| polarC18 | 134.019 | 104.9311 | 59.9 | 0.56 |

| | | | | |
|---|---|---|---|---|
| polarC18 | 134.019 | 56.332 | 59.8 | 0.56 |
| polarC18 | 134.019 | 73.8776 | 59.8 | 0.56 |
| polarC18 | 134.019 | 95.9427 | 59.7 | 0.56 |
| polarC18 | 134.019 | 81.8397 | 59.7 | 0.56 |
| polarC18 | 134.019 | 42.5711 | 59.7 | 0.56 |
| polarC18 | 134.019 | 125.371 | 59.6 | 0.56 |
| polarC18 | 134.019 | 45.2972 | 59.5 | 0.56 |
| polarC18 | 134.019 | 42.8225 | 59.5 | 0.56 |
| polarC18 | 134.019 | 90.2393 | 59.4 | 0.55 |
| polarC18 | 134.019 | 69.5274 | 59.4 | 0.55 |
| polarC18 | 134.019 | 40.3742 | 59.4 | 0.55 |
| polarC18 | 134.019 | 58.2876 | 59.3 | 0.55 |
| polarC18 | 134.019 | 105.8297 | 59.3 | 0.55 |
| polarC18 | 134.019 | 112.3493 | 59.3 | 0.55 |
| polarC18 | 134.019 | 78.3504 | 59.3 | 0.55 |
| polarC18 | 134.019 | 59.5666 | 59.1 | 0.55 |
| polarC18 | 134.019 | 78.6058 | 59.1 | 0.55 |
| polarC18 | 134.019 | 91.4298 | 58.9 | 0.55 |
| polarC18 | 134.019 | 86.8693 | 58.9 | 0.55 |
| polarC18 | 134.019 | 43.3763 | 58.8 | 0.55 |
| polarC18 | 134.019 | 77.0481 | 58.7 | 0.55 |
| polarC18 | 134.019 | 59.5132 | 58.7 | 0.55 |
| polarC18 | 134.019 | 92.169 | 58.6 | 0.55 |
| polarC18 | 134.019 | 58.6356 | 58.6 | 0.55 |
| polarC18 | 134.019 | 57.3459 | 58.5 | 0.55 |
| polarC18 | 134.019 | 52.4955 | 58.5 | 0.55 |
| polarC18 | 134.019 | 76.4548 | 58.5 | 0.55 |
| polarC18 | 134.019 | 72.6666 | 58.5 | 0.55 |
| polarC18 | 134.019 | 87.3703 | 58.4 | 0.54 |
| polarC18 | 134.019 | 49.7676 | 58.2 | 0.54 |
| polarC18 | 134.019 | 71.5257 | 58 | 0.54 |
| polarC18 | 134.019 | 41.9018 | 57.9 | 0.54 |
| polarC18 | 134.019 | 46.9825 | 57.7 | 0.54 |
| polarC18 | 134.019 | 66.9963 | 57.7 | 0.54 |
| polarC18 | 134.019 | 90.1311 | 57.6 | 0.54 |
| polarC18 | 134.019 | 110.182 | 57.6 | 0.54 |
| polarC18 | 134.019 | 103.3289 | 57 | 0.53 |
| polarC18 | 134.019 | 42.4651 | 56.3 | 0.53 |
| polarC18 | 134.019 | 94.346 | 56.2 | 0.52 |
| polarC18 | 134.019 | 98.3678 | 56.1 | 0.52 |
| polarC18 | 134.019 | 65.8894 | 56.1 | 0.52 |
| polarC18 | 134.019 | 145.244 | 56 | 0.52 |

| | | | | |
|---|---|---|---|---|
| polarC18 | 134.019 | 82.1384 | 55.9 | 0.52 |
| polarC18 | 134.019 | 43.1706 | 55.8 | 0.52 |
| polarC18 | 134.019 | 144.9708 | 55.8 | 0.52 |
| polarC18 | 134.019 | 81.4953 | 55.7 | 0.52 |
| polarC18 | 134.019 | 90.6858 | 55.6 | 0.52 |
| polarC18 | 134.019 | 104.2732 | 54.9 | 0.51 |
| polarC18 | 134.019 | 115.259 | 54.8 | 0.51 |
| polarC18 | 134.019 | 72.6883 | 54.8 | 0.51 |
| polarC18 | 134.019 | 94.267 | 54.6 | 0.51 |
| polarC18 | 134.019 | 156.5554 | 54.6 | 0.51 |
| polarC18 | 134.019 | 54.8027 | 54.4 | 0.51 |
| polarC18 | 134.019 | 51.1465 | 54.1 | 0.5 |
| polarC18 | 134.019 | 72.8629 | 54 | 0.5 |
| polarC18 | 134.019 | 106.3938 | 53.7 | 0.5 |
| polarC18 | 134.019 | 91.8544 | 53.6 | 0.5 |
| polarC18 | 134.019 | 52.0023 | 53.5 | 0.5 |
| polarC18 | 134.019 | 67.5854 | 53.4 | 0.5 |
| polarC18 | 134.019 | 109.7989 | 53 | 0.49 |
| polarC18 | 134.019 | 139.5044 | 52.2 | 0.49 |
| polarC18 | 134.019 | 63.1066 | 52.2 | 0.49 |
| polarC18 | 134.019 | 79.309 | 51.5 | 0.48 |
| polarC18 | 134.019 | 57.0732 | 51.5 | 0.48 |
| polarC18 | 134.019 | 134.654 | 51.5 | 0.48 |
| polarC18 | 134.019 | 90.2987 | 51.3 | 0.48 |
| polarC18 | 134.019 | 97.637 | 50.9 | 0.47 |
| polarC18 | 134.019 | 62.7288 | 50.7 | 0.47 |
| polarC18 | 134.019 | 61.2437 | 50.7 | 0.47 |
| polarC18 | 134.019 | 67.1256 | 50.5 | 0.47 |
| polarC18 | 134.019 | 132.1717 | 50.4 | 0.47 |
| polarC18 | 134.019 | 75.8018 | 50.3 | 0.47 |
| polarC18 | 134.019 | 150.9875 | 50.3 | 0.47 |
| polarC18 | 134.019 | 43.4442 | 50.2 | 0.47 |
| polarC18 | 134.019 | 124.6133 | 50.1 | 0.47 |
| polarC18 | 134.019 | 85.4107 | 49.5 | 0.46 |
| polarC18 | 134.019 | 62.9871 | 48.4 | 0.45 |
| polarC18 | 134.019 | 41.0402 | 48.3 | 0.45 |
| polarC18 | 134.019 | 46.9759 | 48.3 | 0.45 |
| polarC18 | 134.019 | 57.8205 | 48.2 | 0.45 |
| polarC18 | 134.019 | 92.572 | 47.9 | 0.45 |
| polarC18 | 134.019 | 41.747 | 47.7 | 0.44 |
| polarC18 | 134.019 | 63.2551 | 47.2 | 0.44 |
| polarC18 | 134.019 | 41.1552 | 46.9 | 0.44 |

| | | | | |
|---|---|---|---|---|
| polarC18 | 134.019 | 40.7191 | 45.5 | 0.42 |
| polarC18 | 218.139 | 194.9435 | 1409.6 | 100 |
| polarC18 | 218.139 | 135.9265 | 626.2 | 44.42 |
| polarC18 | 218.139 | 176.9323 | 448.8 | 31.84 |
| polarC18 | 218.139 | 128.9505 | 349.4 | 24.79 |
| polarC18 | 218.139 | 85.0284 | 262.1 | 18.6 |
| polarC18 | 218.139 | 176.9715 | 213.4 | 15.14 |
| polarC18 | 218.139 | 218.1034 | 206.9 | 14.68 |
| polarC18 | 218.139 | 218.213 | 166 | 11.78 |
| polarC18 | 218.139 | 218.1396 | 138.9 | 9.85 |
| polarC18 | 218.139 | 217.9601 | 103.4 | 7.34 |
| polarC18 | 218.139 | 228.2182 | 98.1 | 6.96 |
| polarC18 | 218.139 | 190.0901 | 83.9 | 5.95 |
| polarC18 | 218.139 | 72.0803 | 60.9 | 4.32 |
| polarC18 | 218.139 | 93.9567 | 53.1 | 3.77 |
| polarC18 | 218.139 | 51.5646 | 51.1 | 3.63 |
| polarC18 | 218.139 | 201.0418 | 50.4 | 3.58 |
| polarC18 | 218.139 | 93.4796 | 49.3 | 3.5 |
| polarC18 | 218.139 | 93.9672 | 49.2 | 3.49 |
| polarC18 | 218.139 | 102.0545 | 46.4 | 3.29 |
| polarC18 | 218.139 | 172.1959 | 45.3 | 3.21 |
| polarC18 | 218.139 | 214.0993 | 41.6 | 2.95 |
| polarC18 | 218.139 | 186.6991 | 41 | 2.91 |
| polarC18 | 218.139 | 233.5972 | 40.2 | 2.85 |
| polarC18 | 218.139 | 51.5445 | 38.8 | 2.75 |
| polarC18 | 218.139 | 90.418 | 38 | 2.7 |
| polarC18 | 218.139 | 119.1922 | 37.1 | 2.63 |
| polarC18 | 218.139 | 176.3173 | 37 | 2.62 |
| polarC18 | 218.139 | 180.5659 | 36.7 | 2.6 |
| polarC18 | 218.139 | 217.9938 | 36.4 | 2.58 |
| polarC18 | 218.139 | 204.7402 | 35.9 | 2.54 |
| polarC18 | 218.139 | 94.1373 | 35.6 | 2.53 |
| polarC18 | 218.139 | 194.9778 | 35.5 | 2.52 |
| polarC18 | 218.139 | 93.4997 | 35.1 | 2.49 |
| polarC18 | 218.139 | 242.5782 | 34.3 | 2.43 |
| polarC18 | 218.139 | 217.7781 | 33.8 | 2.4 |
| polarC18 | 218.139 | 162.7759 | 33.8 | 2.4 |
| polarC18 | 218.139 | 51.568 | 33.7 | 2.39 |
| polarC18 | 218.139 | 199.3579 | 33.6 | 2.39 |
| polarC18 | 218.139 | 61.2133 | 33.2 | 2.36 |
| polarC18 | 218.139 | 94.0857 | 32.6 | 2.31 |
| polarC18 | 218.139 | 94.2719 | 32.5 | 2.31 |

| | | | | |
|---|---|---|---|---|
| polarC18 | 218.139 | 135.0492 | 32.4 | 2.3 |
| polarC18 | 218.139 | 163.3845 | 32.4 | 2.3 |
| polarC18 | 218.139 | 234.8143 | 32.3 | 2.29 |
| polarC18 | 218.139 | 105.1212 | 32.1 | 2.28 |
| polarC18 | 218.139 | 207.8756 | 32 | 2.27 |
| polarC18 | 218.139 | 117.2596 | 31.9 | 2.26 |
| polarC18 | 218.139 | 51.5414 | 31.5 | 2.24 |
| polarC18 | 218.139 | 51.5782 | 31.5 | 2.23 |
| polarC18 | 218.139 | 85.4086 | 31.5 | 2.23 |
| polarC18 | 218.139 | 221.2046 | 31.5 | 2.23 |
| polarC18 | 218.139 | 67.4821 | 31.4 | 2.23 |
| polarC18 | 218.139 | 233.4839 | 31.3 | 2.22 |
| polarC18 | 218.139 | 112.8464 | 31.3 | 2.22 |
| polarC18 | 218.139 | 94.5584 | 31.1 | 2.21 |
| polarC18 | 218.139 | 190.8672 | 31 | 2.2 |
| polarC18 | 218.139 | 137.1487 | 31 | 2.2 |
| polarC18 | 218.139 | 55.4761 | 30.9 | 2.19 |
| polarC18 | 218.139 | 129.5287 | 30.4 | 2.16 |
| polarC18 | 218.139 | 154.2259 | 30.4 | 2.15 |
| polarC18 | 218.139 | 170.8247 | 30.3 | 2.15 |
| polarC18 | 218.139 | 200.3539 | 30.1 | 2.13 |
| polarC18 | 218.139 | 167.1047 | 29.9 | 2.12 |
| polarC18 | 218.139 | 121.8804 | 29.7 | 2.11 |
| polarC18 | 218.139 | 155.9747 | 29.4 | 2.08 |
| polarC18 | 218.139 | 168.6228 | 29.3 | 2.08 |
| polarC18 | 218.139 | 94.3248 | 28.7 | 2.03 |
| polarC18 | 218.139 | 153.9375 | 28.6 | 2.03 |
| polarC18 | 218.139 | 153.7274 | 28.5 | 2.02 |
| polarC18 | 218.139 | 93.9449 | 28.3 | 2.01 |
| polarC18 | 218.139 | 163.1124 | 28.3 | 2.01 |
| polarC18 | 218.139 | 166.9863 | 28.3 | 2 |
| polarC18 | 218.139 | 69.9928 | 28.2 | 2 |
| polarC18 | 218.139 | 94.6271 | 28.2 | 2 |
| polarC18 | 218.139 | 103.4523 | 28 | 1.99 |
| polarC18 | 218.139 | 93.9213 | 28 | 1.99 |
| polarC18 | 218.139 | 135.2265 | 28 | 1.99 |
| polarC18 | 218.139 | 189.411 | 27.9 | 1.98 |
| polarC18 | 218.139 | 94.1087 | 27.9 | 1.98 |
| polarC18 | 218.139 | 203.6442 | 27.9 | 1.98 |
| polarC18 | 218.139 | 204.5807 | 27.8 | 1.97 |
| polarC18 | 218.139 | 124.8966 | 27.8 | 1.97 |
| polarC18 | 218.139 | 66.2077 | 27.7 | 1.97 |

| | | | | |
|---|---|---|---|---|
| polarC18 | 218.139 | 98.5509 | 27.7 | 1.96 |
| polarC18 | 218.139 | 139.6345 | 27.7 | 1.96 |
| polarC18 | 218.139 | 99.3374 | 27.6 | 1.96 |
| polarC18 | 218.139 | 94.4673 | 27.6 | 1.96 |
| polarC18 | 218.139 | 196.6552 | 27.6 | 1.96 |
| polarC18 | 218.139 | 92.417 | 27.4 | 1.95 |
| polarC18 | 218.139 | 48.872 | 27.4 | 1.95 |
| polarC18 | 218.139 | 159.5924 | 27.4 | 1.94 |
| polarC18 | 218.139 | 130.6043 | 27.3 | 1.94 |
| polarC18 | 218.139 | 99.0066 | 27.3 | 1.94 |
| polarC18 | 218.139 | 120.6313 | 27.3 | 1.94 |
| polarC18 | 218.139 | 56.9736 | 27.2 | 1.93 |
| polarC18 | 218.139 | 217.3818 | 27.2 | 1.93 |
| polarC18 | 218.139 | 81.1962 | 27.1 | 1.93 |
| polarC18 | 218.139 | 131.2653 | 27.1 | 1.93 |
| polarC18 | 218.139 | 75.4026 | 27.1 | 1.92 |
| polarC18 | 218.139 | 204.3185 | 27.1 | 1.92 |
| polarC18 | 218.139 | 70.0551 | 27.1 | 1.92 |
| polarC18 | 218.139 | 103.7325 | 27.1 | 1.92 |
| polarC18 | 218.139 | 75.6736 | 27 | 1.91 |
| polarC18 | 218.139 | 88.0305 | 27 | 1.91 |
| polarC18 | 218.139 | 93.9077 | 26.9 | 1.91 |
| polarC18 | 218.139 | 93.6496 | 26.8 | 1.9 |
| polarC18 | 218.139 | 116.8602 | 26.7 | 1.89 |
| polarC18 | 218.139 | 91.6626 | 26.6 | 1.89 |
| polarC18 | 218.139 | 49.119 | 26.6 | 1.89 |
| polarC18 | 218.139 | 105.4089 | 26.5 | 1.88 |
| polarC18 | 218.139 | 201.3148 | 26.4 | 1.87 |
| polarC18 | 218.139 | 209.864 | 26.4 | 1.87 |
| polarC18 | 218.139 | 158.9718 | 26.4 | 1.87 |
| polarC18 | 218.139 | 56.9642 | 26.4 | 1.87 |
| polarC18 | 218.139 | 49.5851 | 26.3 | 1.87 |
| polarC18 | 218.139 | 206.1453 | 26.3 | 1.86 |
| polarC18 | 218.139 | 232.452 | 26.3 | 1.86 |
| polarC18 | 218.139 | 73.9628 | 26.3 | 1.86 |
| polarC18 | 218.139 | 226.3178 | 26.2 | 1.86 |
| polarC18 | 218.139 | 93.5444 | 26.2 | 1.86 |
| polarC18 | 218.139 | 97.367 | 26.2 | 1.86 |
| polarC18 | 218.139 | 241.1839 | 26.1 | 1.85 |
| polarC18 | 218.139 | 145.0211 | 26.1 | 1.85 |
| polarC18 | 218.139 | 171.223 | 26.1 | 1.85 |
| polarC18 | 218.139 | 165.2387 | 26 | 1.85 |

| | | | | |
|---|---|---|---|---|
| polarC18 | 218.139 | 65.6121 | 26 | 1.85 |
| polarC18 | 218.139 | 132.3333 | 26 | 1.84 |
| polarC18 | 218.139 | 118.7477 | 25.9 | 1.84 |
| polarC18 | 218.139 | 166.6603 | 25.9 | 1.84 |
| polarC18 | 218.139 | 102.3638 | 25.9 | 1.83 |
| polarC18 | 218.139 | 91.4918 | 25.8 | 1.83 |
| polarC18 | 218.139 | 188.9181 | 25.7 | 1.83 |
| polarC18 | 218.139 | 93.7724 | 25.7 | 1.83 |
| polarC18 | 218.139 | 93.8985 | 25.7 | 1.83 |
| polarC18 | 218.139 | 133.6187 | 25.7 | 1.82 |
| polarC18 | 218.139 | 100.4599 | 25.7 | 1.82 |
| polarC18 | 218.139 | 143.2845 | 25.7 | 1.82 |
| polarC18 | 218.139 | 186.5839 | 25.7 | 1.82 |
| polarC18 | 218.139 | 113.8642 | 25.6 | 1.82 |
| polarC18 | 218.139 | 77.1173 | 25.6 | 1.82 |
| polarC18 | 218.139 | 238.6496 | 25.5 | 1.81 |
| polarC18 | 218.139 | 59.7598 | 25.5 | 1.81 |
| polarC18 | 218.139 | 51.8747 | 25.4 | 1.8 |
| polarC18 | 218.139 | 102.5247 | 25.4 | 1.8 |
| polarC18 | 218.139 | 90.6456 | 25.4 | 1.8 |
| polarC18 | 218.139 | 55.3993 | 25.4 | 1.8 |
| polarC18 | 218.139 | 139.2983 | 25.4 | 1.8 |
| polarC18 | 218.139 | 177.6159 | 25.3 | 1.79 |
| polarC18 | 218.139 | 159.3164 | 25.3 | 1.79 |
| polarC18 | 218.139 | 118.5844 | 25.3 | 1.79 |
| polarC18 | 218.139 | 110.7618 | 25.3 | 1.79 |
| polarC18 | 218.139 | 57.0041 | 25.2 | 1.79 |
| polarC18 | 218.139 | 75.1289 | 25.2 | 1.79 |
| polarC18 | 218.139 | 219.4369 | 25.2 | 1.79 |
| polarC18 | 218.139 | 93.9831 | 25.2 | 1.78 |
| polarC18 | 218.139 | 88.9095 | 25.1 | 1.78 |
| polarC18 | 218.139 | 94.0254 | 25.1 | 1.78 |
| polarC18 | 218.139 | 214.3928 | 25 | 1.77 |
| polarC18 | 218.139 | 174.3279 | 25 | 1.77 |
| polarC18 | 218.139 | 120.1466 | 25 | 1.77 |
| polarC18 | 218.139 | 51.2671 | 25 | 1.77 |
| polarC18 | 218.139 | 188.6009 | 24.9 | 1.77 |
| polarC18 | 218.139 | 180.8939 | 24.9 | 1.77 |
| polarC18 | 218.139 | 147.7552 | 24.9 | 1.77 |
| polarC18 | 218.139 | 136.1921 | 24.9 | 1.76 |
| polarC18 | 218.139 | 169.733 | 24.9 | 1.76 |
| polarC18 | 218.139 | 86.8298 | 24.9 | 1.76 |

| | | | | |
|---|---|---|---|---|
| polarC18 | 218.139 | 210.7224 | 24.9 | 1.76 |
| polarC18 | 218.139 | 52.905 | 24.8 | 1.76 |
| polarC18 | 218.139 | 208.8797 | 24.8 | 1.76 |
| polarC18 | 218.139 | 58.5325 | 24.8 | 1.76 |
| polarC18 | 218.139 | 67.6462 | 24.7 | 1.76 |
| polarC18 | 218.139 | 81.2209 | 24.7 | 1.75 |
| polarC18 | 218.139 | 58.0821 | 24.7 | 1.75 |
| polarC18 | 218.139 | 219.3528 | 24.7 | 1.75 |
| polarC18 | 218.139 | 154.4825 | 24.6 | 1.75 |
| polarC18 | 218.139 | 58.6175 | 24.6 | 1.75 |
| polarC18 | 218.139 | 209.7917 | 24.6 | 1.74 |
| polarC18 | 218.139 | 139.5539 | 24.5 | 1.74 |
| polarC18 | 218.139 | 140.6609 | 24.4 | 1.73 |
| polarC18 | 218.139 | 98.0294 | 24.4 | 1.73 |
| polarC18 | 218.139 | 164.9827 | 24.3 | 1.72 |
| polarC18 | 218.139 | 149.4866 | 24.3 | 1.72 |
| polarC18 | 218.139 | 129.3569 | 24.2 | 1.72 |
| polarC18 | 218.139 | 184.5264 | 24.2 | 1.72 |
| polarC18 | 218.139 | 111.482 | 24.2 | 1.71 |
| polarC18 | 218.139 | 112.9219 | 24.1 | 1.71 |
| polarC18 | 218.139 | 212.8989 | 24.1 | 1.71 |
| polarC18 | 218.139 | 72.9381 | 24.1 | 1.71 |
| polarC18 | 218.139 | 68.0275 | 24 | 1.7 |
| polarC18 | 218.139 | 71.1758 | 23.9 | 1.7 |
| polarC18 | 218.139 | 117.4028 | 23.9 | 1.69 |
| polarC18 | 218.139 | 123.4782 | 23.7 | 1.68 |
| polarC18 | 218.139 | 199.6676 | 23.7 | 1.68 |
| polarC18 | 218.139 | 166.3693 | 23.7 | 1.68 |
| polarC18 | 218.139 | 64.3894 | 23.6 | 1.68 |
| polarC18 | 218.139 | 52.6978 | 23.6 | 1.68 |
| polarC18 | 218.139 | 94.0449 | 23.6 | 1.67 |
| polarC18 | 218.139 | 82.8002 | 23.4 | 1.66 |
| polarC18 | 218.139 | 240.9291 | 23.4 | 1.66 |
| polarC18 | 218.139 | 214.5663 | 23.4 | 1.66 |
| polarC18 | 218.139 | 155.7446 | 23.4 | 1.66 |
| polarC18 | 218.139 | 152.3469 | 23.3 | 1.66 |
| polarC18 | 218.139 | 171.9263 | 23.3 | 1.65 |
| polarC18 | 218.139 | 234.4399 | 23.2 | 1.64 |
| polarC18 | 218.139 | 123.8137 | 23 | 1.63 |
| polarC18 | 218.139 | 119.4532 | 22.9 | 1.63 |
| polarC18 | 218.139 | 162.3394 | 22.9 | 1.62 |
| polarC18 | 218.139 | 63.7265 | 22.8 | 1.62 |

| | | | | |
|---|---|---|---|---|
| polarC18 | 218.139 | 145.426 | 22.7 | 1.61 |
| polarC18 | 218.139 | 160.323 | 22.7 | 1.61 |
| polarC18 | 218.139 | 217.7027 | 22.6 | 1.6 |
| polarC18 | 218.139 | 208.4155 | 22.6 | 1.6 |
| polarC18 | 218.139 | 185.8958 | 22.5 | 1.6 |
| polarC18 | 218.139 | 49.0948 | 22.5 | 1.59 |
| polarC18 | 218.139 | 94.8736 | 22.4 | 1.59 |
| polarC18 | 218.139 | 51.1451 | 22.4 | 1.59 |
| polarC18 | 218.139 | 149.9472 | 22.3 | 1.58 |
| polarC18 | 218.139 | 95.9891 | 22.3 | 1.58 |
| polarC18 | 218.139 | 78.5671 | 22.2 | 1.57 |
| polarC18 | 218.139 | 74.1118 | 22.2 | 1.57 |
| polarC18 | 218.139 | 178.0025 | 22.1 | 1.57 |
| polarC18 | 218.139 | 111.9892 | 22.1 | 1.57 |
| polarC18 | 218.139 | 106.1024 | 22 | 1.56 |
| polarC18 | 218.139 | 66.8607 | 21.9 | 1.55 |
| polarC18 | 218.139 | 94.8457 | 21.8 | 1.55 |
| polarC18 | 218.139 | 120.9076 | 21.8 | 1.55 |
| polarC18 | 218.139 | 136.3613 | 21.7 | 1.54 |
| polarC18 | 218.139 | 60.1408 | 21.7 | 1.54 |
| polarC18 | 218.139 | 204.8099 | 21.7 | 1.54 |
| polarC18 | 218.139 | 94.1803 | 21.7 | 1.54 |
| polarC18 | 218.139 | 160.5628 | 21.7 | 1.54 |
| polarC18 | 218.139 | 237.0889 | 21.7 | 1.54 |
| polarC18 | 218.139 | 91.4058 | 21.7 | 1.54 |
| polarC18 | 218.139 | 107.1546 | 21.6 | 1.53 |
| polarC18 | 218.139 | 225.89 | 21.6 | 1.53 |
| polarC18 | 218.139 | 128.5638 | 21.6 | 1.53 |
| polarC18 | 218.139 | 53.6749 | 21.6 | 1.53 |
| polarC18 | 218.139 | 117.1656 | 21.5 | 1.53 |
| polarC18 | 218.139 | 213.8523 | 21.5 | 1.53 |
| polarC18 | 218.139 | 188.4919 | 21.5 | 1.53 |
| polarC18 | 218.139 | 97.1451 | 21.5 | 1.53 |
| polarC18 | 218.139 | 198.033 | 21.5 | 1.53 |
| polarC18 | 218.139 | 163.6524 | 21.5 | 1.53 |
| polarC18 | 218.139 | 81.4353 | 21.5 | 1.53 |
| polarC18 | 218.139 | 162.5503 | 21.5 | 1.53 |
| polarC18 | 218.139 | 187.6591 | 21.5 | 1.52 |
| polarC18 | 218.139 | 108.6323 | 21.5 | 1.52 |
| polarC18 | 218.139 | 103.691 | 21.4 | 1.52 |
| polarC18 | 218.139 | 77.0897 | 21.4 | 1.52 |
| polarC18 | 218.139 | 55.5153 | 21.4 | 1.52 |

| | | | | |
|---|---|---|---|---|
| polarC18 | 218.139 | 147.8379 | 21.4 | 1.52 |
| polarC18 | 218.139 | 187.1517 | 21.3 | 1.51 |
| polarC18 | 218.139 | 93.9748 | 21.3 | 1.51 |
| polarC18 | 218.139 | 94.9558 | 21.3 | 1.51 |
| polarC18 | 218.139 | 172.6723 | 21.3 | 1.51 |
| polarC18 | 218.139 | 59.9419 | 21.3 | 1.51 |
| polarC18 | 218.139 | 178.2163 | 21.2 | 1.51 |
| polarC18 | 218.139 | 112.28 | 21.2 | 1.5 |
| polarC18 | 218.139 | 123.0035 | 21.2 | 1.5 |
| polarC18 | 218.139 | 126.4503 | 21.2 | 1.5 |
| polarC18 | 218.139 | 60.1966 | 21.1 | 1.5 |
| polarC18 | 218.139 | 144.4818 | 21.1 | 1.5 |
| polarC18 | 218.139 | 117.59 | 21.1 | 1.5 |
| polarC18 | 218.139 | 56.9466 | 21.1 | 1.5 |
| polarC18 | 218.139 | 176.9537 | 21.1 | 1.5 |
| polarC18 | 218.139 | 182.9742 | 21.1 | 1.49 |
| polarC18 | 218.139 | 154.2522 | 21 | 1.49 |
| polarC18 | 218.139 | 165.5478 | 21 | 1.49 |
| polarC18 | 218.139 | 79.5708 | 21 | 1.49 |
| polarC18 | 218.139 | 103.2924 | 21 | 1.49 |
| polarC18 | 218.139 | 234.0889 | 21 | 1.49 |
| polarC18 | 218.139 | 55.2221 | 21 | 1.49 |
| polarC18 | 218.139 | 153.3099 | 21 | 1.49 |
| polarC18 | 218.139 | 117.7516 | 21 | 1.49 |
| polarC18 | 218.139 | 148.082 | 21 | 1.49 |
| polarC18 | 218.139 | 94.4972 | 20.9 | 1.49 |
| polarC18 | 218.139 | 58.2945 | 20.9 | 1.49 |
| polarC18 | 218.139 | 86.2673 | 20.9 | 1.48 |
| polarC18 | 218.139 | 155.9935 | 20.9 | 1.48 |
| polarC18 | 218.139 | 143.605 | 20.9 | 1.48 |
| polarC18 | 218.139 | 114.57 | 20.9 | 1.48 |
| polarC18 | 218.139 | 98.4579 | 20.8 | 1.48 |
| polarC18 | 218.139 | 57.81 | 20.8 | 1.48 |
| polarC18 | 218.139 | 69.0302 | 20.8 | 1.47 |
| polarC18 | 218.139 | 59.3483 | 20.7 | 1.47 |
| polarC18 | 218.139 | 141.0273 | 20.7 | 1.47 |
| polarC18 | 218.139 | 94.0005 | 20.7 | 1.47 |
| polarC18 | 218.139 | 241.0835 | 20.7 | 1.47 |
| polarC18 | 218.139 | 99.7449 | 20.7 | 1.47 |
| polarC18 | 218.139 | 150.9607 | 20.7 | 1.47 |
| polarC18 | 218.139 | 106.1216 | 20.6 | 1.46 |
| polarC18 | 218.139 | 179.9991 | 20.6 | 1.46 |

| | | | | |
|---|---|---|---|---|
| polarC18 | 218.139 | 183.4876 | 20.6 | 1.46 |
| polarC18 | 218.139 | 178.0268 | 20.6 | 1.46 |
| polarC18 | 218.139 | 65.3488 | 20.6 | 1.46 |
| polarC18 | 218.139 | 49.0459 | 20.6 | 1.46 |
| polarC18 | 218.139 | 133.4938 | 20.6 | 1.46 |
| polarC18 | 218.139 | 150.1356 | 20.6 | 1.46 |
| polarC18 | 218.139 | 212.4744 | 20.6 | 1.46 |
| polarC18 | 218.139 | 71.3618 | 20.6 | 1.46 |
| polarC18 | 218.139 | 192.1137 | 20.5 | 1.46 |
| polarC18 | 218.139 | 163.912 | 20.5 | 1.46 |
| polarC18 | 218.139 | 97.5611 | 20.5 | 1.46 |
| polarC18 | 218.139 | 146.3873 | 20.5 | 1.46 |
| polarC18 | 218.139 | 205.0275 | 20.5 | 1.45 |
| polarC18 | 218.139 | 237.6229 | 20.5 | 1.45 |
| polarC18 | 218.139 | 102.7357 | 20.5 | 1.45 |
| polarC18 | 218.139 | 116.608 | 20.5 | 1.45 |
| polarC18 | 218.139 | 162.0279 | 20.5 | 1.45 |
| polarC18 | 218.139 | 93.7013 | 20.4 | 1.45 |
| polarC18 | 218.139 | 135.7266 | 20.4 | 1.45 |
| polarC18 | 218.139 | 228.3994 | 20.4 | 1.45 |
| polarC18 | 218.139 | 122.5161 | 20.4 | 1.45 |
| polarC18 | 218.139 | 238.2152 | 20.4 | 1.45 |
| polarC18 | 218.139 | 84.3819 | 20.4 | 1.45 |
| polarC18 | 218.139 | 186.3569 | 20.4 | 1.45 |
| polarC18 | 218.139 | 164.1816 | 20.4 | 1.45 |
| polarC18 | 218.139 | 55.6607 | 20.4 | 1.45 |
| polarC18 | 218.139 | 101.1073 | 20.4 | 1.45 |
| polarC18 | 218.139 | 68.8939 | 20.4 | 1.44 |
| polarC18 | 218.139 | 179.1334 | 20.4 | 1.44 |
| polarC18 | 218.139 | 53.0796 | 20.3 | 1.44 |
| polarC18 | 218.139 | 77.067 | 20.3 | 1.44 |
| polarC18 | 218.139 | 97.8119 | 20.3 | 1.44 |
| polarC18 | 218.139 | 74.9909 | 20.3 | 1.44 |
| polarC18 | 218.139 | 52.3657 | 20.3 | 1.44 |
| polarC18 | 218.139 | 57.2211 | 20.3 | 1.44 |
| polarC18 | 218.139 | 77.1024 | 20.3 | 1.44 |
| polarC18 | 218.139 | 113.9659 | 20.3 | 1.44 |
| polarC18 | 218.139 | 177.5293 | 20.3 | 1.44 |
| polarC18 | 218.139 | 230.8256 | 20.3 | 1.44 |
| polarC18 | 218.139 | 172.9681 | 20.3 | 1.44 |
| polarC18 | 218.139 | 137.3864 | 20.3 | 1.44 |
| polarC18 | 218.139 | 135.5797 | 20.2 | 1.43 |

| | | | | |
|---|---|---|---|---|
| polarC18 | 218.139 | 68.6733 | 20.2 | 1.43 |
| polarC18 | 218.139 | 151.1911 | 20.2 | 1.43 |
| polarC18 | 218.139 | 226.8011 | 20.2 | 1.43 |
| polarC18 | 218.139 | 67.991 | 20.2 | 1.43 |
| polarC18 | 218.139 | 203.6786 | 20.2 | 1.43 |
| polarC18 | 218.139 | 114.0518 | 20.2 | 1.43 |
| polarC18 | 218.139 | 59.849 | 20.2 | 1.43 |
| polarC18 | 218.139 | 53.2993 | 20.2 | 1.43 |
| polarC18 | 218.139 | 78.6918 | 20.2 | 1.43 |
| polarC18 | 218.139 | 73.6328 | 20.2 | 1.43 |
| polarC18 | 218.139 | 55.7529 | 20.2 | 1.43 |
| polarC18 | 218.139 | 196.2063 | 20.2 | 1.43 |
| polarC18 | 218.139 | 144.5993 | 20.1 | 1.43 |
| polarC18 | 218.139 | 52.4812 | 20.1 | 1.43 |
| polarC18 | 218.139 | 101.2805 | 20.1 | 1.43 |
| polarC18 | 218.139 | 62.7347 | 20.1 | 1.43 |
| polarC18 | 218.139 | 229.2653 | 20.1 | 1.43 |
| polarC18 | 218.139 | 90.6553 | 20.1 | 1.43 |
| polarC18 | 218.139 | 181.8571 | 20.1 | 1.43 |
| polarC18 | 218.139 | 237.7966 | 20.1 | 1.43 |
| polarC18 | 218.139 | 49.1454 | 20.1 | 1.43 |
| polarC18 | 218.139 | 52.4407 | 20.1 | 1.43 |
| polarC18 | 218.139 | 170.5491 | 20.1 | 1.43 |
| polarC18 | 218.139 | 212.0099 | 20.1 | 1.42 |
| polarC18 | 218.139 | 123.4969 | 20.1 | 1.42 |
| polarC18 | 218.139 | 60.4706 | 20.1 | 1.42 |
| polarC18 | 218.139 | 65.6976 | 20 | 1.42 |
| polarC18 | 218.139 | 103.5149 | 20 | 1.42 |
| polarC18 | 218.139 | 188.9565 | 20 | 1.42 |
| polarC18 | 218.139 | 96.8922 | 20 | 1.42 |
| polarC18 | 218.139 | 113.6226 | 20 | 1.42 |
| polarC18 | 218.139 | 182.2576 | 20 | 1.42 |
| polarC18 | 218.139 | 238.876 | 20 | 1.42 |
| polarC18 | 218.139 | 74.3049 | 20 | 1.42 |
| polarC18 | 218.139 | 82.0977 | 20 | 1.42 |
| polarC18 | 218.139 | 95.5448 | 20 | 1.42 |
| polarC18 | 218.139 | 185.3633 | 19.9 | 1.41 |
| polarC18 | 218.139 | 78.4591 | 19.9 | 1.41 |
| polarC18 | 218.139 | 70.4434 | 19.9 | 1.41 |
| polarC18 | 218.139 | 119.4325 | 19.9 | 1.41 |
| polarC18 | 218.139 | 94.2291 | 19.9 | 1.41 |
| polarC18 | 218.139 | 92.5503 | 19.9 | 1.41 |

| | | | | |
|---|---|---|---|---|
| polarC18 | 218.139 | 116.3667 | 19.9 | 1.41 |
| polarC18 | 218.139 | 102.9418 | 19.9 | 1.41 |
| polarC18 | 218.139 | 236.6592 | 19.9 | 1.41 |
| polarC18 | 218.139 | 50.6105 | 19.9 | 1.41 |
| polarC18 | 218.139 | 74.0927 | 19.9 | 1.41 |
| polarC18 | 218.139 | 156.5691 | 19.9 | 1.41 |
| polarC18 | 218.139 | 74.2011 | 19.9 | 1.41 |
| polarC18 | 218.139 | 236.4871 | 19.9 | 1.41 |
| polarC18 | 218.139 | 58.7855 | 19.9 | 1.41 |
| polarC18 | 218.139 | 58.3992 | 19.9 | 1.41 |
| polarC18 | 218.139 | 222.0842 | 19.9 | 1.41 |
| polarC18 | 218.139 | 131.3448 | 19.8 | 1.41 |
| polarC18 | 218.139 | 167.4422 | 19.8 | 1.41 |
| polarC18 | 218.139 | 51.2338 | 19.8 | 1.41 |
| polarC18 | 218.139 | 172.3987 | 19.8 | 1.41 |
| polarC18 | 218.139 | 156.4043 | 19.8 | 1.41 |
| polarC18 | 218.139 | 93.2831 | 19.8 | 1.41 |
| polarC18 | 218.139 | 120.4134 | 19.8 | 1.41 |
| polarC18 | 218.139 | 140.4712 | 19.8 | 1.41 |
| polarC18 | 218.139 | 142.5605 | 19.8 | 1.4 |
| polarC18 | 218.139 | 168.3227 | 19.8 | 1.4 |
| polarC18 | 218.139 | 117.232 | 19.8 | 1.4 |
| polarC18 | 218.139 | 95.6384 | 19.8 | 1.4 |
| polarC18 | 218.139 | 101.7939 | 19.8 | 1.4 |
| polarC18 | 218.139 | 69.8085 | 19.8 | 1.4 |
| polarC18 | 218.139 | 68.0719 | 19.8 | 1.4 |
| polarC18 | 218.139 | 53.7175 | 19.8 | 1.4 |
| polarC18 | 218.139 | 55.6662 | 19.8 | 1.4 |
| polarC18 | 218.139 | 84.7468 | 19.7 | 1.4 |
| polarC18 | 218.139 | 65.3916 | 19.7 | 1.4 |
| polarC18 | 218.139 | 177.2058 | 19.7 | 1.4 |
| polarC18 | 218.139 | 139.8775 | 19.7 | 1.4 |
| polarC18 | 218.139 | 83.8323 | 19.7 | 1.4 |
| polarC18 | 218.139 | 118.1706 | 19.7 | 1.4 |
| polarC18 | 218.139 | 171.9928 | 19.7 | 1.4 |
| polarC18 | 218.139 | 115.8586 | 19.7 | 1.39 |
| polarC18 | 218.139 | 111.1021 | 19.7 | 1.39 |
| polarC18 | 218.139 | 51.5324 | 19.6 | 1.39 |
| polarC18 | 218.139 | 94.6167 | 19.6 | 1.39 |
| polarC18 | 218.139 | 81.9338 | 19.6 | 1.39 |
| polarC18 | 218.139 | 81.1151 | 19.6 | 1.39 |
| polarC18 | 218.139 | 73.1832 | 19.6 | 1.39 |

| | | | | |
|---|---|---|---|---|
| polarC18 | 218.139 | 56.9848 | 19.6 | 1.39 |
| polarC18 | 218.139 | 165.3766 | 19.6 | 1.39 |
| polarC18 | 218.139 | 214.6308 | 19.6 | 1.39 |
| polarC18 | 218.139 | 59.1891 | 19.6 | 1.39 |
| polarC18 | 218.139 | 145.3916 | 19.6 | 1.39 |
| polarC18 | 218.139 | 84.0136 | 19.6 | 1.39 |
| polarC18 | 218.139 | 177.7567 | 19.6 | 1.39 |
| polarC18 | 218.139 | 137.2236 | 19.6 | 1.39 |
| polarC18 | 218.139 | 196.3749 | 19.6 | 1.39 |
| polarC18 | 218.139 | 158.7035 | 19.6 | 1.39 |
| polarC18 | 218.139 | 226.4701 | 19.5 | 1.39 |
| polarC18 | 218.139 | 121.3654 | 19.5 | 1.38 |
| polarC18 | 218.139 | 203.3962 | 19.5 | 1.38 |
| polarC18 | 218.139 | 166.8903 | 19.5 | 1.38 |
| polarC18 | 218.139 | 135.5239 | 19.5 | 1.38 |
| polarC18 | 218.139 | 57.0949 | 19.5 | 1.38 |
| polarC18 | 218.139 | 95.0137 | 19.5 | 1.38 |
| polarC18 | 218.139 | 90.5937 | 19.5 | 1.38 |
| polarC18 | 218.139 | 99.8505 | 19.5 | 1.38 |
| polarC18 | 218.139 | 119.6078 | 19.5 | 1.38 |
| polarC18 | 218.139 | 123.2384 | 19.5 | 1.38 |
| polarC18 | 218.139 | 102.9245 | 19.5 | 1.38 |
| polarC18 | 218.139 | 49.1073 | 19.4 | 1.38 |
| polarC18 | 218.139 | 239.9369 | 19.4 | 1.38 |
| polarC18 | 218.139 | 141.242 | 19.4 | 1.38 |
| polarC18 | 218.139 | 152.2987 | 19.4 | 1.38 |
| polarC18 | 218.139 | 71.2378 | 19.4 | 1.38 |
| polarC18 | 218.139 | 240.6823 | 19.4 | 1.38 |
| polarC18 | 218.139 | 161.4469 | 19.4 | 1.38 |
| polarC18 | 218.139 | 50.8317 | 19.4 | 1.38 |
| polarC18 | 218.139 | 63.4261 | 19.4 | 1.38 |
| polarC18 | 218.139 | 64.0663 | 19.4 | 1.38 |
| polarC18 | 218.139 | 158.858 | 19.4 | 1.37 |
| polarC18 | 218.139 | 84.7033 | 19.4 | 1.37 |
| polarC18 | 218.139 | 77.1831 | 19.4 | 1.37 |
| polarC18 | 218.139 | 63.5317 | 19.4 | 1.37 |
| polarC18 | 218.139 | 229.7728 | 19.4 | 1.37 |
| polarC18 | 218.139 | 89.2112 | 19.4 | 1.37 |
| polarC18 | 218.139 | 220.776 | 19.3 | 1.37 |
| polarC18 | 218.139 | 170.7447 | 19.3 | 1.37 |
| polarC18 | 218.139 | 146.8087 | 19.3 | 1.37 |
| polarC18 | 218.139 | 83.9247 | 19.3 | 1.37 |

| | | | | |
|---|---|---|---|---|
| polarC18 | 218.139 | 152.6119 | 19.3 | 1.37 |
| polarC18 | 218.139 | 162.9619 | 19.3 | 1.37 |
| polarC18 | 218.139 | 151.2775 | 19.3 | 1.37 |
| polarC18 | 218.139 | 70.7326 | 19.3 | 1.37 |
| polarC18 | 218.139 | 94.335 | 19.3 | 1.37 |
| polarC18 | 218.139 | 156.0509 | 19.3 | 1.37 |
| polarC18 | 218.139 | 132.4989 | 19.3 | 1.37 |
| polarC18 | 218.139 | 219.0321 | 19.3 | 1.37 |
| polarC18 | 218.139 | 196.7778 | 19.3 | 1.37 |
| polarC18 | 218.139 | 131.05 | 19.3 | 1.37 |
| polarC18 | 218.139 | 226.8878 | 19.2 | 1.37 |
| polarC18 | 218.139 | 141.8251 | 19.2 | 1.36 |
| polarC18 | 218.139 | 231.9768 | 19.2 | 1.36 |
| polarC18 | 218.139 | 91.7137 | 19.2 | 1.36 |
| polarC18 | 218.139 | 195.0468 | 19.2 | 1.36 |
| polarC18 | 218.139 | 88.4636 | 19.2 | 1.36 |
| polarC18 | 218.139 | 219.8051 | 19.2 | 1.36 |
| polarC18 | 218.139 | 137.6082 | 19.2 | 1.36 |
| polarC18 | 218.139 | 94.1545 | 19.2 | 1.36 |
| polarC18 | 218.139 | 225.934 | 19.2 | 1.36 |
| polarC18 | 218.139 | 86.9061 | 19.2 | 1.36 |
| polarC18 | 218.139 | 76.1324 | 19.2 | 1.36 |
| polarC18 | 218.139 | 202.1217 | 19.2 | 1.36 |
| polarC18 | 218.139 | 84.5488 | 19.2 | 1.36 |
| polarC18 | 218.139 | 103.0574 | 19.2 | 1.36 |
| polarC18 | 218.139 | 186.9447 | 19.2 | 1.36 |
| polarC18 | 218.139 | 65.4157 | 19.2 | 1.36 |
| polarC18 | 218.139 | 126.5606 | 19.2 | 1.36 |
| polarC18 | 218.139 | 57.7727 | 19.1 | 1.36 |
| polarC18 | 218.139 | 134.0352 | 19.1 | 1.36 |
| polarC18 | 218.139 | 79.9832 | 19.1 | 1.36 |
| polarC18 | 218.139 | 167.7421 | 19.1 | 1.35 |
| polarC18 | 218.139 | 174.1347 | 19.1 | 1.35 |
| polarC18 | 218.139 | 65.1485 | 19.1 | 1.35 |
| polarC18 | 218.139 | 111.3203 | 19.1 | 1.35 |
| polarC18 | 218.139 | 52.3981 | 19.1 | 1.35 |
| polarC18 | 218.139 | 158.1255 | 19.1 | 1.35 |
| polarC18 | 218.139 | 100.2246 | 19.1 | 1.35 |
| polarC18 | 218.139 | 207.667 | 19 | 1.35 |
| polarC18 | 218.139 | 94.2426 | 19 | 1.35 |
| polarC18 | 218.139 | 67.9139 | 19 | 1.35 |
| polarC18 | 218.139 | 67.7109 | 19 | 1.35 |

| | | | | |
|---|---|---|---|---|
| polarC18 | 218.139 | 95.6738 | 19 | 1.35 |
| polarC18 | 218.139 | 80.7054 | 19 | 1.35 |
| polarC18 | 218.139 | 220.8844 | 19 | 1.35 |
| polarC18 | 218.139 | 157.9699 | 19 | 1.35 |
| polarC18 | 218.139 | 165.5164 | 19 | 1.35 |
| polarC18 | 218.139 | 109.2357 | 19 | 1.35 |
| polarC18 | 218.139 | 56.6933 | 19 | 1.35 |
| polarC18 | 218.139 | 135.5607 | 19 | 1.35 |
| polarC18 | 218.139 | 145.8906 | 18.9 | 1.34 |
| polarC18 | 218.139 | 53.1169 | 18.9 | 1.34 |
| polarC18 | 218.139 | 186.3843 | 18.9 | 1.34 |
| polarC18 | 218.139 | 63.7399 | 18.9 | 1.34 |
| polarC18 | 218.139 | 130.8199 | 18.9 | 1.34 |
| polarC18 | 218.139 | 95.2324 | 18.9 | 1.34 |
| polarC18 | 218.139 | 65.4464 | 18.9 | 1.34 |
| polarC18 | 218.139 | 54.0601 | 18.9 | 1.34 |
| polarC18 | 218.139 | 68.393 | 18.9 | 1.34 |
| polarC18 | 218.139 | 167.5205 | 18.9 | 1.34 |
| polarC18 | 218.139 | 81.5124 | 18.9 | 1.34 |
| polarC18 | 218.139 | 84.0808 | 18.9 | 1.34 |
| polarC18 | 218.139 | 75.4513 | 18.9 | 1.34 |
| polarC18 | 218.139 | 110.9377 | 18.9 | 1.34 |
| polarC18 | 218.139 | 66.9012 | 18.9 | 1.34 |
| polarC18 | 218.139 | 104.1203 | 18.9 | 1.34 |
| polarC18 | 218.139 | 119.2302 | 18.9 | 1.34 |
| polarC18 | 218.139 | 94.344 | 18.9 | 1.34 |
| polarC18 | 218.139 | 238.7169 | 18.9 | 1.34 |
| polarC18 | 218.139 | 59.4117 | 18.8 | 1.34 |
| polarC18 | 218.139 | 197.8569 | 18.8 | 1.34 |
| polarC18 | 218.139 | 144.5176 | 18.8 | 1.34 |
| polarC18 | 218.139 | 165.33 | 18.8 | 1.34 |
| polarC18 | 218.139 | 110.2383 | 18.8 | 1.34 |
| polarC18 | 218.139 | 133.8741 | 18.8 | 1.34 |
| polarC18 | 218.139 | 53.853 | 18.8 | 1.34 |
| polarC18 | 218.139 | 75.1875 | 18.8 | 1.34 |
| polarC18 | 218.139 | 61.7064 | 18.8 | 1.33 |
| polarC18 | 218.139 | 151.2164 | 18.8 | 1.33 |
| polarC18 | 218.139 | 88.9226 | 18.8 | 1.33 |
| polarC18 | 218.139 | 164.0705 | 18.7 | 1.33 |
| polarC18 | 218.139 | 175.477 | 18.7 | 1.33 |
| polarC18 | 218.139 | 77.936 | 18.7 | 1.33 |
| polarC18 | 218.139 | 124.9452 | 18.7 | 1.33 |

| | | | | |
|---|---|---|---|---|
| polarC18 | 218.139 | 197.496 | 18.7 | 1.33 |
| polarC18 | 218.139 | 74.668 | 18.7 | 1.33 |
| polarC18 | 218.139 | 62.1848 | 18.7 | 1.33 |
| polarC18 | 218.139 | 48.7026 | 18.7 | 1.33 |
| polarC18 | 218.139 | 146.6713 | 18.7 | 1.33 |
| polarC18 | 218.139 | 176.2402 | 18.7 | 1.32 |
| polarC18 | 218.139 | 132.5138 | 18.7 | 1.32 |
| polarC18 | 218.139 | 54.4707 | 18.7 | 1.32 |
| polarC18 | 218.139 | 54.9518 | 18.7 | 1.32 |
| polarC18 | 218.139 | 63.8496 | 18.7 | 1.32 |
| polarC18 | 218.139 | 77.8957 | 18.7 | 1.32 |
| polarC18 | 218.139 | 104.3022 | 18.7 | 1.32 |
| polarC18 | 218.139 | 130.1498 | 18.7 | 1.32 |
| polarC18 | 218.139 | 100.6779 | 18.7 | 1.32 |
| polarC18 | 218.139 | 90.7745 | 18.7 | 1.32 |
| polarC18 | 218.139 | 226.504 | 18.7 | 1.32 |
| polarC18 | 218.139 | 102.1182 | 18.6 | 1.32 |
| polarC18 | 218.139 | 105.275 | 18.6 | 1.32 |
| polarC18 | 218.139 | 176.4344 | 18.6 | 1.32 |
| polarC18 | 218.139 | 193.84 | 18.6 | 1.32 |
| polarC18 | 218.139 | 62.8662 | 18.6 | 1.32 |
| polarC18 | 218.139 | 238.0601 | 18.6 | 1.32 |
| polarC18 | 218.139 | 185.6093 | 18.6 | 1.32 |
| polarC18 | 218.139 | 68.679 | 18.6 | 1.32 |
| polarC18 | 218.139 | 57.0678 | 18.6 | 1.32 |
| polarC18 | 218.139 | 94.0135 | 18.6 | 1.32 |
| polarC18 | 218.139 | 81.6716 | 18.6 | 1.32 |
| polarC18 | 218.139 | 78.8193 | 18.6 | 1.32 |
| polarC18 | 218.139 | 96.3877 | 18.6 | 1.32 |
| polarC18 | 218.139 | 72.3841 | 18.6 | 1.32 |
| polarC18 | 218.139 | 108.1399 | 18.5 | 1.31 |
| polarC18 | 218.139 | 148.7774 | 18.5 | 1.31 |
| polarC18 | 218.139 | 55.4604 | 18.5 | 1.31 |
| polarC18 | 218.139 | 158.1871 | 18.5 | 1.31 |
| polarC18 | 218.139 | 239.8076 | 18.5 | 1.31 |
| polarC18 | 218.139 | 72.6222 | 18.5 | 1.31 |
| polarC18 | 218.139 | 49.1884 | 18.5 | 1.31 |
| polarC18 | 218.139 | 174.7253 | 18.5 | 1.31 |
| polarC18 | 218.139 | 114.2813 | 18.5 | 1.31 |
| polarC18 | 218.139 | 51.752 | 18.5 | 1.31 |
| polarC18 | 218.139 | 180.1208 | 18.5 | 1.31 |
| polarC18 | 218.139 | 103.6395 | 18.5 | 1.31 |

| | | | | |
|---|---|---|---|---|
| polarC18 | 218.139 | 72.5435 | 18.5 | 1.31 |
| polarC18 | 218.139 | 69.1054 | 18.5 | 1.31 |
| polarC18 | 218.139 | 86.2066 | 18.4 | 1.31 |
| polarC18 | 218.139 | 103.5617 | 18.4 | 1.31 |
| polarC18 | 218.139 | 86.6114 | 18.4 | 1.31 |
| polarC18 | 218.139 | 176.0914 | 18.4 | 1.31 |
| polarC18 | 218.139 | 128.3506 | 18.4 | 1.31 |
| polarC18 | 218.139 | 66.0887 | 18.4 | 1.31 |
| polarC18 | 218.139 | 67.6265 | 18.4 | 1.31 |
| polarC18 | 218.139 | 234.1858 | 18.4 | 1.31 |
| polarC18 | 218.139 | 56.7384 | 18.4 | 1.31 |
| polarC18 | 218.139 | 171.167 | 18.4 | 1.31 |
| polarC18 | 218.139 | 202.564 | 18.4 | 1.3 |
| polarC18 | 218.139 | 217.2524 | 18.4 | 1.3 |
| polarC18 | 218.139 | 121.3508 | 18.4 | 1.3 |
| polarC18 | 218.139 | 88.1777 | 18.4 | 1.3 |
| polarC18 | 218.139 | 143.6809 | 18.4 | 1.3 |
| polarC18 | 218.139 | 136.2415 | 18.4 | 1.3 |
| polarC18 | 218.139 | 184.1089 | 18.4 | 1.3 |
| polarC18 | 218.139 | 67 | 18.4 | 1.3 |
| polarC18 | 218.139 | 119.4016 | 18.3 | 1.3 |
| polarC18 | 218.139 | 76.2807 | 18.3 | 1.3 |
| polarC18 | 218.139 | 169.961 | 18.3 | 1.3 |
| polarC18 | 218.139 | 93.6386 | 18.3 | 1.3 |
| polarC18 | 218.139 | 225.7705 | 18.3 | 1.3 |
| polarC18 | 218.139 | 69.522 | 18.3 | 1.3 |
| polarC18 | 218.139 | 142.2631 | 18.3 | 1.3 |
| polarC18 | 218.139 | 121.5467 | 18.3 | 1.3 |
| polarC18 | 218.139 | 118.4182 | 18.3 | 1.3 |
| polarC18 | 218.139 | 213.7235 | 18.3 | 1.3 |
| polarC18 | 218.139 | 148.5754 | 18.3 | 1.3 |
| polarC18 | 218.139 | 168.7117 | 18.3 | 1.3 |
| polarC18 | 218.139 | 148.2096 | 18.3 | 1.3 |
| polarC18 | 218.139 | 48.7359 | 18.3 | 1.3 |
| polarC18 | 218.139 | 143.1139 | 18.3 | 1.29 |
| polarC18 | 218.139 | 90.1455 | 18.2 | 1.29 |
| polarC18 | 218.139 | 205.7112 | 18.2 | 1.29 |
| polarC18 | 218.139 | 146.2835 | 18.2 | 1.29 |
| polarC18 | 218.139 | 198.9105 | 18.2 | 1.29 |
| polarC18 | 218.139 | 192.5143 | 18.2 | 1.29 |
| polarC18 | 218.139 | 93.4375 | 18.2 | 1.29 |
| polarC18 | 218.139 | 221.8161 | 18.2 | 1.29 |

| | | | | |
|---|---|---|---|---|
| polarC18 | 218.139 | 53.0244 | 18.2 | 1.29 |
| polarC18 | 218.139 | 158.6162 | 18.2 | 1.29 |
| polarC18 | 218.139 | 126.5389 | 18.2 | 1.29 |
| polarC18 | 218.139 | 223.9225 | 18.2 | 1.29 |
| polarC18 | 218.139 | 156.8024 | 18.2 | 1.29 |
| polarC18 | 218.139 | 180.8252 | 18.2 | 1.29 |
| polarC18 | 218.139 | 202.6124 | 18.2 | 1.29 |
| polarC18 | 218.139 | 118.9036 | 18.2 | 1.29 |
| polarC18 | 218.139 | 158.2426 | 18.1 | 1.29 |
| polarC18 | 218.139 | 119.1055 | 18.1 | 1.29 |
| polarC18 | 218.139 | 63.8 | 18.1 | 1.29 |
| polarC18 | 218.139 | 88.8084 | 18.1 | 1.29 |
| polarC18 | 218.139 | 68.1068 | 18.1 | 1.29 |
| polarC18 | 218.139 | 153.1964 | 18.1 | 1.28 |
| polarC18 | 218.139 | 199.5675 | 18.1 | 1.28 |
| polarC18 | 218.139 | 137.9657 | 18.1 | 1.28 |
| polarC18 | 218.139 | 51.384 | 18.1 | 1.28 |
| polarC18 | 218.139 | 80.9682 | 18.1 | 1.28 |
| polarC18 | 218.139 | 157.1008 | 18.1 | 1.28 |
| polarC18 | 218.139 | 50.7758 | 18.1 | 1.28 |
| polarC18 | 218.139 | 113.6111 | 18.1 | 1.28 |
| polarC18 | 218.139 | 121.5806 | 18.1 | 1.28 |
| polarC18 | 218.139 | 49.6565 | 18.1 | 1.28 |
| polarC18 | 218.139 | 179.3199 | 18 | 1.28 |
| polarC18 | 218.139 | 118.3794 | 18 | 1.28 |
| polarC18 | 218.139 | 52.0374 | 18 | 1.28 |
| polarC18 | 218.139 | 191.5802 | 18 | 1.28 |
| polarC18 | 218.139 | 153.5905 | 18 | 1.28 |
| polarC18 | 218.139 | 127.7525 | 18 | 1.28 |
| polarC18 | 218.139 | 232.1418 | 18 | 1.28 |
| polarC18 | 218.139 | 103.9872 | 18 | 1.28 |
| polarC18 | 218.139 | 71.4003 | 18 | 1.28 |
| polarC18 | 218.139 | 116.5927 | 18 | 1.28 |
| polarC18 | 218.139 | 108.1681 | 18 | 1.27 |
| polarC18 | 218.139 | 94.99 | 18 | 1.27 |
| polarC18 | 218.139 | 52.2199 | 17.9 | 1.27 |
| polarC18 | 218.139 | 73.0057 | 17.9 | 1.27 |
| polarC18 | 218.139 | 113.6426 | 17.9 | 1.27 |
| polarC18 | 218.139 | 163.7518 | 17.9 | 1.27 |
| polarC18 | 218.139 | 75.2672 | 17.9 | 1.27 |
| polarC18 | 218.139 | 69.8455 | 17.9 | 1.27 |
| polarC18 | 218.139 | 108.284 | 17.9 | 1.27 |

| | | | | |
|---|---|---|---|---|
| polarC18 | 218.139 | 97.1574 | 17.9 | 1.27 |
| polarC18 | 218.139 | 75.3037 | 17.9 | 1.27 |
| polarC18 | 218.139 | 139.1823 | 17.9 | 1.27 |
| polarC18 | 218.139 | 95.1474 | 17.9 | 1.27 |
| polarC18 | 218.139 | 198.7514 | 17.9 | 1.27 |
| polarC18 | 218.139 | 119.9546 | 17.9 | 1.27 |
| polarC18 | 218.139 | 79.6341 | 17.9 | 1.27 |
| polarC18 | 218.139 | 163.1874 | 17.9 | 1.27 |
| polarC18 | 218.139 | 136.1147 | 17.8 | 1.27 |
| polarC18 | 218.139 | 175.2979 | 17.8 | 1.26 |
| polarC18 | 218.139 | 158.9282 | 17.8 | 1.26 |
| polarC18 | 218.139 | 212.7431 | 17.8 | 1.26 |
| polarC18 | 218.139 | 50.2526 | 17.8 | 1.26 |
| polarC18 | 218.139 | 68.0021 | 17.8 | 1.26 |
| polarC18 | 218.139 | 50.1336 | 17.8 | 1.26 |
| polarC18 | 218.139 | 101.965 | 17.8 | 1.26 |
| polarC18 | 218.139 | 170.4334 | 17.8 | 1.26 |
| polarC18 | 218.139 | 181.2681 | 17.8 | 1.26 |
| polarC18 | 218.139 | 93.4481 | 17.8 | 1.26 |
| polarC18 | 218.139 | 236.8193 | 17.7 | 1.26 |
| polarC18 | 218.139 | 51.6366 | 17.7 | 1.26 |
| polarC18 | 218.139 | 172.546 | 17.7 | 1.26 |
| polarC18 | 218.139 | 181.8991 | 17.7 | 1.26 |
| polarC18 | 218.139 | 87.8399 | 17.7 | 1.26 |
| polarC18 | 218.139 | 237.5154 | 17.7 | 1.26 |
| polarC18 | 218.139 | 116.7624 | 17.7 | 1.26 |
| polarC18 | 218.139 | 184.5543 | 17.7 | 1.26 |
| polarC18 | 218.139 | 53.9146 | 17.7 | 1.25 |
| polarC18 | 218.139 | 159.2305 | 17.6 | 1.25 |
| polarC18 | 218.139 | 78.6035 | 17.6 | 1.25 |
| polarC18 | 218.139 | 219.8688 | 17.6 | 1.25 |
| polarC18 | 218.139 | 236.7527 | 17.6 | 1.25 |
| polarC18 | 218.139 | 69.4741 | 17.6 | 1.25 |
| polarC18 | 218.139 | 136.6637 | 17.6 | 1.25 |
| polarC18 | 218.139 | 79.7532 | 17.6 | 1.25 |
| polarC18 | 218.139 | 224.7299 | 17.5 | 1.24 |
| polarC18 | 218.139 | 52.844 | 17.5 | 1.24 |
| polarC18 | 218.139 | 74.4365 | 17.5 | 1.24 |
| polarC18 | 218.139 | 72.6565 | 17.5 | 1.24 |
| polarC18 | 218.139 | 146.7711 | 17.5 | 1.24 |
| polarC18 | 218.139 | 58.1807 | 17.5 | 1.24 |
| polarC18 | 218.139 | 84.8705 | 17.4 | 1.24 |

| | | | | |
|---|---|---|---|---|
| polarC18 | 218.139 | 56.5023 | 17.4 | 1.24 |
| polarC18 | 218.139 | 140.0736 | 17.4 | 1.24 |
| polarC18 | 218.139 | 163.3086 | 17.4 | 1.23 |
| polarC18 | 218.139 | 68.9742 | 17.4 | 1.23 |
| polarC18 | 218.139 | 175.0877 | 17.3 | 1.23 |
| polarC18 | 218.139 | 176.4865 | 17.3 | 1.23 |
| polarC18 | 218.139 | 86.7983 | 17.3 | 1.23 |
| polarC18 | 218.139 | 176.7485 | 17.3 | 1.23 |
| polarC18 | 218.139 | 49.3501 | 17.3 | 1.23 |
| polarC18 | 218.139 | 110.1341 | 17.3 | 1.23 |
| polarC18 | 218.139 | 89.1535 | 17.3 | 1.23 |
| polarC18 | 218.139 | 79.149 | 17.3 | 1.23 |
| polarC18 | 218.139 | 164.8177 | 17.3 | 1.22 |
| polarC18 | 218.139 | 62.586 | 17.2 | 1.22 |
| polarC18 | 218.139 | 170.345 | 17.2 | 1.22 |
| polarC18 | 218.139 | 68.7735 | 17.2 | 1.22 |
| polarC18 | 218.139 | 110.5792 | 17.2 | 1.22 |
| polarC18 | 218.139 | 211.5981 | 17.1 | 1.22 |
| polarC18 | 218.139 | 83.0683 | 17.1 | 1.22 |
| polarC18 | 218.139 | 224.9958 | 17.1 | 1.21 |
| polarC18 | 218.139 | 53.401 | 17.1 | 1.21 |
| polarC18 | 218.139 | 157.3082 | 17.1 | 1.21 |
| polarC18 | 218.139 | 224.1069 | 17.1 | 1.21 |
| polarC18 | 218.139 | 127.9962 | 17 | 1.21 |
| polarC18 | 218.139 | 184.7315 | 17 | 1.21 |
| polarC18 | 218.139 | 90.8097 | 17 | 1.2 |
| polarC18 | 218.139 | 56.1557 | 16.9 | 1.2 |
| polarC18 | 218.139 | 111.936 | 16.9 | 1.2 |
| polarC18 | 218.139 | 121.3819 | 16.9 | 1.2 |
| polarC18 | 218.139 | 162.475 | 16.9 | 1.2 |
| polarC18 | 218.139 | 85.0634 | 16.9 | 1.2 |
| polarC18 | 218.139 | 163.8033 | 16.9 | 1.2 |
| polarC18 | 218.139 | 183.3109 | 16.9 | 1.2 |
| polarC18 | 218.139 | 71.2538 | 16.9 | 1.2 |
| polarC18 | 218.139 | 94.3658 | 16.8 | 1.19 |
| polarC18 | 218.139 | 121.6431 | 16.8 | 1.19 |
| polarC18 | 218.139 | 55.937 | 16.7 | 1.19 |
| polarC18 | 218.139 | 183.0087 | 16.7 | 1.18 |
| polarC18 | 218.139 | 201.6487 | 16.7 | 1.18 |
| polarC18 | 218.139 | 75.5312 | 16.6 | 1.18 |
| polarC18 | 218.139 | 168.3909 | 16.6 | 1.18 |
| polarC18 | 218.139 | 81.4133 | 16.6 | 1.18 |

| | | | | |
|---|---|---|---|---|
| polarC18 | 218.139 | 108.1245 | 16.6 | 1.17 |
| polarC18 | 218.139 | 70.0901 | 16.5 | 1.17 |
| polarC18 | 218.139 | 172.7952 | 16.5 | 1.17 |
| polarC18 | 218.139 | 161.5071 | 16.5 | 1.17 |
| polarC18 | 218.139 | 99.9073 | 16.4 | 1.16 |
| polarC18 | 218.139 | 94.7748 | 16.4 | 1.16 |
| polarC18 | 218.139 | 57.5779 | 16.3 | 1.16 |
| polarC18 | 218.139 | 55.2349 | 16.3 | 1.16 |
| polarC18 | 218.139 | 113.0932 | 16.3 | 1.16 |
| polarC18 | 218.139 | 78.3058 | 16.2 | 1.15 |
| polarC18 | 218.139 | 123.8367 | 16.1 | 1.15 |
| polarC18 | 218.139 | 59.523 | 16.1 | 1.14 |
| polarC18 | 218.139 | 50.3878 | 16 | 1.13 |
| polarC18 | 218.139 | 93.4608 | 16 | 1.13 |
| polarC18 | 218.139 | 204.8519 | 15.9 | 1.13 |
| polarC18 | 218.139 | 51.8145 | 15.9 | 1.13 |
| polarC18 | 218.139 | 133.9187 | 15.9 | 1.13 |
| polarC18 | 218.139 | 94.1217 | 15.9 | 1.13 |
| polarC18 | 218.139 | 130.9176 | 15.9 | 1.13 |
| polarC18 | 218.139 | 48.8019 | 15.9 | 1.13 |
| polarC18 | 218.139 | 99.9868 | 15.9 | 1.13 |
| polarC18 | 218.139 | 126.5152 | 15.9 | 1.13 |
| polarC18 | 218.139 | 229.7088 | 15.9 | 1.12 |
| polarC18 | 218.139 | 61.4231 | 15.9 | 1.12 |
| polarC18 | 218.139 | 164.378 | 15.8 | 1.12 |
| polarC18 | 218.139 | 105.7891 | 15.7 | 1.12 |
| polarC18 | 218.139 | 94.0309 | 15.7 | 1.12 |
| polarC18 | 218.139 | 223.4284 | 15.7 | 1.12 |
| polarC18 | 218.139 | 73.112 | 15.7 | 1.11 |
| polarC18 | 218.139 | 50.2299 | 15.7 | 1.11 |
| polarC18 | 218.139 | 120.7195 | 15.6 | 1.11 |
| polarC18 | 218.139 | 83.2766 | 15.6 | 1.11 |
| polarC18 | 218.139 | 227.892 | 15.6 | 1.11 |
| polarC18 | 218.139 | 54.5037 | 15.6 | 1.11 |
| polarC18 | 218.139 | 51.7379 | 15.5 | 1.1 |
| polarC18 | 218.139 | 108.2123 | 15.5 | 1.1 |
| polarC18 | 218.139 | 145.2263 | 15.5 | 1.1 |
| polarC18 | 218.139 | 68.4391 | 15.5 | 1.1 |
| polarC18 | 218.139 | 90.3671 | 15.5 | 1.1 |
| polarC18 | 218.139 | 212.8205 | 15.5 | 1.1 |
| polarC18 | 218.139 | 120.4484 | 15.4 | 1.1 |
| polarC18 | 218.139 | 93.5064 | 15.4 | 1.09 |

| | | | | |
|---|---|---|---|---|
| polarC18 | 218.139 | 74.6808 | 15.4 | 1.09 |
| polarC18 | 218.139 | 67.8409 | 15.4 | 1.09 |
| polarC18 | 218.139 | 163.0449 | 15.4 | 1.09 |
| polarC18 | 218.139 | 175.4159 | 15.3 | 1.09 |
| polarC18 | 218.139 | 187.4649 | 15.3 | 1.09 |
| polarC18 | 218.139 | 133.5304 | 15.3 | 1.09 |
| polarC18 | 218.139 | 191.6136 | 15.3 | 1.09 |
| polarC18 | 218.139 | 82.9484 | 15.3 | 1.08 |
| polarC18 | 218.139 | 120.95 | 15.2 | 1.08 |
| polarC18 | 218.139 | 83.7655 | 15.2 | 1.08 |
| polarC18 | 218.139 | 218.0518 | 15.2 | 1.08 |
| polarC18 | 218.139 | 94.6826 | 15.2 | 1.08 |
| polarC18 | 218.139 | 81.9883 | 15.1 | 1.07 |
| polarC18 | 218.139 | 163.5379 | 15.1 | 1.07 |
| polarC18 | 218.139 | 225.0994 | 15.1 | 1.07 |
| polarC18 | 218.139 | 116.3046 | 15.1 | 1.07 |
| polarC18 | 218.139 | 164.7232 | 15.1 | 1.07 |
| polarC18 | 218.139 | 68.4472 | 15.1 | 1.07 |
| polarC18 | 218.139 | 221.0405 | 15.1 | 1.07 |
| polarC18 | 218.139 | 94.2629 | 15.1 | 1.07 |
| polarC18 | 218.139 | 185.501 | 15.1 | 1.07 |
| polarC18 | 218.139 | 171.7324 | 15 | 1.07 |
| polarC18 | 218.139 | 239.5545 | 15 | 1.07 |
| polarC18 | 218.139 | 232.9677 | 15 | 1.06 |
| polarC18 | 218.139 | 94.8981 | 15 | 1.06 |
| polarC18 | 218.139 | 56.9617 | 15 | 1.06 |
| polarC18 | 218.139 | 203.0937 | 15 | 1.06 |
| polarC18 | 218.139 | 96.3425 | 15 | 1.06 |
| polarC18 | 218.139 | 179.0266 | 15 | 1.06 |
| polarC18 | 218.139 | 184.7008 | 15 | 1.06 |
| polarC18 | 218.139 | 61.4325 | 14.9 | 1.06 |
| polarC18 | 218.139 | 224.0611 | 14.9 | 1.06 |
| polarC18 | 218.139 | 51.3075 | 14.9 | 1.06 |
| polarC18 | 218.139 | 51.5507 | 14.9 | 1.06 |
| polarC18 | 218.139 | 50.8831 | 14.9 | 1.06 |
| polarC18 | 218.139 | 76.7568 | 14.9 | 1.06 |
| polarC18 | 218.139 | 165.8547 | 14.9 | 1.06 |
| polarC18 | 218.139 | 60.8894 | 14.9 | 1.05 |
| polarC18 | 218.139 | 178.675 | 14.8 | 1.05 |
| polarC18 | 218.139 | 138.3751 | 14.8 | 1.05 |
| polarC18 | 218.139 | 150.8715 | 14.8 | 1.05 |
| polarC18 | 218.139 | 52.7256 | 14.8 | 1.05 |

| | | | | |
|---|---|---|---|---|
| polarC18 | 218.139 | 192.6265 | 14.8 | 1.05 |
| polarC18 | 218.139 | 55.5828 | 14.8 | 1.05 |
| polarC18 | 218.139 | 121.8107 | 14.8 | 1.05 |
| polarC18 | 218.139 | 53.3681 | 14.8 | 1.05 |
| polarC18 | 218.139 | 90.662 | 14.8 | 1.05 |
| polarC18 | 218.139 | 138.9717 | 14.8 | 1.05 |
| polarC18 | 218.139 | 130.2557 | 14.7 | 1.05 |
| polarC18 | 218.139 | 94.0083 | 14.7 | 1.05 |
| polarC18 | 218.139 | 155.4809 | 14.7 | 1.05 |
| polarC18 | 218.139 | 215.6484 | 14.7 | 1.04 |
| polarC18 | 218.139 | 133.1323 | 14.7 | 1.04 |
| polarC18 | 218.139 | 74.1624 | 14.7 | 1.04 |
| polarC18 | 218.139 | 54.7234 | 14.7 | 1.04 |
| polarC18 | 218.139 | 61.3417 | 14.7 | 1.04 |
| polarC18 | 218.139 | 210.1152 | 14.7 | 1.04 |
| polarC18 | 218.139 | 236.3474 | 14.7 | 1.04 |
| polarC18 | 218.139 | 77.6423 | 14.7 | 1.04 |
| polarC18 | 218.139 | 154.9206 | 14.7 | 1.04 |
| polarC18 | 218.139 | 119.3659 | 14.7 | 1.04 |
| polarC18 | 218.139 | 65.956 | 14.7 | 1.04 |
| polarC18 | 218.139 | 49.8722 | 14.7 | 1.04 |
| polarC18 | 218.139 | 55.7206 | 14.7 | 1.04 |
| polarC18 | 218.139 | 91.633 | 14.7 | 1.04 |
| polarC18 | 218.139 | 191.9933 | 14.7 | 1.04 |
| polarC18 | 218.139 | 236.8738 | 14.6 | 1.04 |
| polarC18 | 218.139 | 49.0023 | 14.6 | 1.04 |
| polarC18 | 218.139 | 61.9107 | 14.6 | 1.04 |
| polarC18 | 218.139 | 68.8535 | 14.6 | 1.04 |
| polarC18 | 218.139 | 110.9661 | 14.6 | 1.04 |
| polarC18 | 218.139 | 59.7712 | 14.6 | 1.04 |
| polarC18 | 218.139 | 66.7584 | 14.6 | 1.04 |
| polarC18 | 218.139 | 206.3832 | 14.6 | 1.04 |
| polarC18 | 218.139 | 223.1172 | 14.6 | 1.04 |
| polarC18 | 218.139 | 149.5381 | 14.6 | 1.04 |
| polarC18 | 218.139 | 120.1243 | 14.6 | 1.03 |
| polarC18 | 218.139 | 192.7225 | 14.6 | 1.03 |
| polarC18 | 218.139 | 74.0144 | 14.6 | 1.03 |
| polarC18 | 218.139 | 162.5719 | 14.6 | 1.03 |
| polarC18 | 218.139 | 81.3729 | 14.6 | 1.03 |
| polarC18 | 218.139 | 170.5711 | 14.6 | 1.03 |
| polarC18 | 218.139 | 51.7249 | 14.6 | 1.03 |
| polarC18 | 218.139 | 83.2849 | 14.6 | 1.03 |

| | | | | |
|---|---|---|---|---|
| polarC18 | 218.139 | 175.2393 | 14.6 | 1.03 |
| polarC18 | 218.139 | 71.4785 | 14.5 | 1.03 |
| polarC18 | 218.139 | 103.3632 | 14.5 | 1.03 |
| polarC18 | 218.139 | 110.2041 | 14.5 | 1.03 |
| polarC18 | 218.139 | 56.1465 | 14.5 | 1.03 |
| polarC18 | 218.139 | 101.505 | 14.5 | 1.03 |
| polarC18 | 218.139 | 195.3261 | 14.5 | 1.03 |
| polarC18 | 218.139 | 225.1512 | 14.5 | 1.03 |
| polarC18 | 218.139 | 68.6383 | 14.5 | 1.03 |
| polarC18 | 218.139 | 83.2423 | 14.5 | 1.03 |
| polarC18 | 218.139 | 52.972 | 14.5 | 1.03 |
| polarC18 | 218.139 | 69.0363 | 14.5 | 1.03 |
| polarC18 | 218.139 | 141.4527 | 14.5 | 1.03 |
| polarC18 | 218.139 | 56.1812 | 14.5 | 1.03 |
| polarC18 | 218.139 | 48.7848 | 14.5 | 1.03 |
| polarC18 | 218.139 | 72.8799 | 14.5 | 1.03 |
| polarC18 | 218.139 | 101.998 | 14.5 | 1.03 |
| polarC18 | 218.139 | 67.2921 | 14.5 | 1.03 |
| polarC18 | 218.139 | 94.0564 | 14.5 | 1.03 |
| polarC18 | 218.139 | 119.6431 | 14.5 | 1.03 |
| polarC18 | 218.139 | 57.0918 | 14.5 | 1.03 |
| polarC18 | 218.139 | 64.2605 | 14.5 | 1.03 |
| polarC18 | 218.139 | 76.7425 | 14.5 | 1.03 |
| polarC18 | 218.139 | 147.9733 | 14.5 | 1.03 |
| polarC18 | 218.139 | 125.1721 | 14.5 | 1.03 |
| polarC18 | 218.139 | 52.2438 | 14.5 | 1.03 |
| polarC18 | 218.139 | 193.7729 | 14.5 | 1.03 |
| polarC18 | 218.139 | 167.9455 | 14.5 | 1.03 |
| polarC18 | 218.139 | 98.5263 | 14.5 | 1.03 |
| polarC18 | 218.139 | 95.8413 | 14.5 | 1.03 |
| polarC18 | 218.139 | 101.4754 | 14.5 | 1.03 |
| polarC18 | 218.139 | 104.7396 | 14.5 | 1.03 |
| polarC18 | 218.139 | 132.7363 | 14.4 | 1.02 |
| polarC18 | 218.139 | 105.6747 | 14.4 | 1.02 |
| polarC18 | 218.139 | 158.3847 | 14.4 | 1.02 |
| polarC18 | 218.139 | 92.9798 | 14.4 | 1.02 |
| polarC18 | 218.139 | 66.5606 | 14.4 | 1.02 |
| polarC18 | 218.139 | 86.337 | 14.4 | 1.02 |
| polarC18 | 218.139 | 74.9551 | 14.4 | 1.02 |
| polarC18 | 218.139 | 241.4486 | 14.4 | 1.02 |
| polarC18 | 218.139 | 171.1169 | 14.4 | 1.02 |
| polarC18 | 218.139 | 81.944 | 14.4 | 1.02 |

| | | | | |
|---|---|---|---|---|
| polarC18 | 218.139 | 134.8045 | 14.4 | 1.02 |
| polarC18 | 218.139 | 60.1921 | 14.4 | 1.02 |
| polarC18 | 218.139 | 116.2916 | 14.4 | 1.02 |
| polarC18 | 218.139 | 100.21 | 14.4 | 1.02 |
| polarC18 | 218.139 | 95.1557 | 14.4 | 1.02 |
| polarC18 | 218.139 | 174.0218 | 14.4 | 1.02 |
| polarC18 | 218.139 | 125.6698 | 14.4 | 1.02 |
| polarC18 | 218.139 | 228.7075 | 14.4 | 1.02 |
| polarC18 | 218.139 | 179.6879 | 14.4 | 1.02 |
| polarC18 | 218.139 | 147.5958 | 14.4 | 1.02 |
| polarC18 | 218.139 | 67.2303 | 14.4 | 1.02 |
| polarC18 | 218.139 | 186.6231 | 14.4 | 1.02 |
| polarC18 | 218.139 | 105.5668 | 14.4 | 1.02 |
| polarC18 | 218.139 | 82.7673 | 14.4 | 1.02 |
| polarC18 | 218.139 | 65.0012 | 14.4 | 1.02 |
| polarC18 | 218.139 | 120.3348 | 14.4 | 1.02 |
| polarC18 | 218.139 | 92.6339 | 14.4 | 1.02 |
| polarC18 | 218.139 | 195.9005 | 14.3 | 1.02 |
| polarC18 | 218.139 | 61.2914 | 14.3 | 1.02 |
| polarC18 | 218.139 | 105.5198 | 14.3 | 1.02 |
| polarC18 | 218.139 | 225.8311 | 14.3 | 1.02 |
| polarC18 | 218.139 | 52.9846 | 14.3 | 1.02 |
| polarC18 | 218.139 | 62.2339 | 14.3 | 1.02 |
| polarC18 | 218.139 | 109.4878 | 14.3 | 1.02 |
| polarC18 | 218.139 | 55.7748 | 14.3 | 1.01 |
| polarC18 | 218.139 | 240.0562 | 14.3 | 1.01 |
| polarC18 | 218.139 | 71.8835 | 14.3 | 1.01 |
| polarC18 | 218.139 | 82.5405 | 14.3 | 1.01 |
| polarC18 | 218.139 | 93.2387 | 14.3 | 1.01 |
| polarC18 | 218.139 | 116.479 | 14.3 | 1.01 |
| polarC18 | 218.139 | 170.1095 | 14.3 | 1.01 |
| polarC18 | 218.139 | 59.4066 | 14.3 | 1.01 |
| polarC18 | 218.139 | 83.105 | 14.3 | 1.01 |
| polarC18 | 218.139 | 206.4356 | 14.3 | 1.01 |
| polarC18 | 218.139 | 52.5057 | 14.3 | 1.01 |
| polarC18 | 218.139 | 209.7198 | 14.3 | 1.01 |
| polarC18 | 218.139 | 96.0399 | 14.3 | 1.01 |
| polarC18 | 218.139 | 137.5245 | 14.3 | 1.01 |
| polarC18 | 218.139 | 112.0402 | 14.3 | 1.01 |
| polarC18 | 218.139 | 73.8497 | 14.2 | 1.01 |
| polarC18 | 218.139 | 93.8351 | 14.2 | 1.01 |
| polarC18 | 218.139 | 57.3289 | 14.2 | 1.01 |

| | | | | |
|---|---|---|---|---|
| polarC18 | 218.139 | 230.9515 | 14.2 | 1.01 |
| polarC18 | 218.139 | 204.1198 | 14.2 | 1.01 |
| polarC18 | 218.139 | 88.7005 | 14.2 | 1.01 |
| polarC18 | 218.139 | 100.0069 | 14.2 | 1.01 |
| polarC18 | 218.139 | 51.8975 | 14.2 | 1.01 |
| polarC18 | 218.139 | 129.4623 | 14.2 | 1.01 |
| polarC18 | 218.139 | 94.4357 | 14.2 | 1.01 |
| polarC18 | 218.139 | 80.0704 | 14.2 | 1.01 |
| polarC18 | 218.139 | 51.7722 | 14.2 | 1.01 |
| polarC18 | 218.139 | 55.8299 | 14.2 | 1.01 |
| polarC18 | 218.139 | 91.1334 | 14.2 | 1.01 |
| polarC18 | 218.139 | 88.4248 | 14.2 | 1.01 |
| polarC18 | 218.139 | 100.7379 | 14.2 | 1.01 |
| polarC18 | 218.139 | 62.0473 | 14.2 | 1.01 |
| polarC18 | 218.139 | 111.5448 | 14.2 | 1.01 |
| polarC18 | 218.139 | 127.534 | 14.2 | 1.01 |
| polarC18 | 218.139 | 199.9214 | 14.2 | 1.01 |
| polarC18 | 218.139 | 66.7503 | 14.2 | 1.01 |
| polarC18 | 218.139 | 94.7894 | 14.2 | 1.01 |
| polarC18 | 218.139 | 63.2686 | 14.2 | 1.01 |
| polarC18 | 218.139 | 101.2655 | 14.2 | 1.01 |
| polarC18 | 218.139 | 117.8464 | 14.2 | 1 |
| polarC18 | 218.139 | 56.8098 | 14.2 | 1 |
| polarC18 | 218.139 | 135.1793 | 14.2 | 1 |
| polarC18 | 218.139 | 81.2524 | 14.1 | 1 |
| polarC18 | 218.139 | 73.2544 | 14.1 | 1 |
| polarC18 | 218.139 | 122.3754 | 14.1 | 1 |
| polarC18 | 218.139 | 58.162 | 14.1 | 1 |
| polarC18 | 218.139 | 56.2231 | 14.1 | 1 |
| polarC18 | 218.139 | 66.8405 | 14.1 | 1 |
| polarC18 | 218.139 | 195.0859 | 14.1 | 1 |
| polarC18 | 218.139 | 84.3672 | 14.1 | 1 |
| polarC18 | 218.139 | 84.577 | 14.1 | 1 |
| polarC18 | 218.139 | 98.2227 | 14.1 | 1 |
| polarC18 | 218.139 | 129.7032 | 14.1 | 1 |
| polarC18 | 218.139 | 197.3781 | 14.1 | 1 |
| polarC18 | 218.139 | 132.6204 | 14.1 | 1 |
| polarC18 | 218.139 | 60.0376 | 14.1 | 1 |
| polarC18 | 218.139 | 107.6688 | 14.1 | 1 |
| polarC18 | 218.139 | 125.9185 | 14.1 | 1 |
| polarC18 | 218.139 | 97.1868 | 14.1 | 1 |
| polarC18 | 218.139 | 54.3311 | 14.1 | 1 |

| | | | | |
|---|---|---|---|---|
| polarC18 | 218.139 | 71.7337 | 14.1 | 1 |
| polarC18 | 218.139 | 215.9923 | 14.1 | 1 |
| polarC18 | 218.139 | 69.4634 | 14.1 | 1 |
| polarC18 | 218.139 | 54.3085 | 14.1 | 1 |
| polarC18 | 218.139 | 74.9373 | 14.1 | 1 |
| polarC18 | 218.139 | 80.6022 | 14.1 | 1 |
| polarC18 | 218.139 | 130.9763 | 14.1 | 1 |
| polarC18 | 218.139 | 72.097 | 14.1 | 1 |
| polarC18 | 218.139 | 144.9193 | 14.1 | 1 |
| polarC18 | 218.139 | 61.9435 | 14.1 | 1 |
| polarC18 | 218.139 | 174.2839 | 14.1 | 1 |
| polarC18 | 218.139 | 130.8381 | 14.1 | 1 |
| polarC18 | 218.139 | 96.7905 | 14.1 | 1 |
| polarC18 | 218.139 | 73.7489 | 14.1 | 1 |
| polarC18 | 218.139 | 155.8761 | 14.1 | 1 |
| polarC18 | 218.139 | 130.4343 | 14.1 | 1 |
| polarC18 | 218.139 | 81.8922 | 14.1 | 1 |
| polarC18 | 218.139 | 74.2567 | 14.1 | 1 |
| polarC18 | 218.139 | 53.9119 | 14 | 1 |
| polarC18 | 218.139 | 93.7145 | 14 | 1 |
| polarC18 | 218.139 | 93.4892 | 14 | 1 |
| polarC18 | 218.139 | 62.0771 | 14 | 1 |
| polarC18 | 218.139 | 76.7086 | 14 | 1 |
| polarC18 | 218.139 | 146.2394 | 14 | 1 |
| polarC18 | 218.139 | 96.2116 | 14 | 1 |
| polarC18 | 218.139 | 167.3756 | 14 | 1 |
| polarC18 | 218.139 | 105.0954 | 14 | 1 |
| polarC18 | 218.139 | 148.5093 | 14 | 1 |
| polarC18 | 218.139 | 133.1595 | 14 | 1 |
| polarC18 | 218.139 | 50.2477 | 14 | 1 |
| polarC18 | 218.139 | 198.6384 | 14 | 0.99 |
| polarC18 | 218.139 | 82.4698 | 14 | 0.99 |
| polarC18 | 218.139 | 59.8649 | 14 | 0.99 |
| polarC18 | 218.139 | 50.1175 | 14 | 0.99 |
| polarC18 | 218.139 | 61.8811 | 14 | 0.99 |
| polarC18 | 218.139 | 233.892 | 14 | 0.99 |
| polarC18 | 218.139 | 144.0949 | 14 | 0.99 |
| polarC18 | 218.139 | 98.9845 | 14 | 0.99 |
| polarC18 | 218.139 | 75.1389 | 14 | 0.99 |
| polarC18 | 218.139 | 129.9595 | 14 | 0.99 |
| polarC18 | 218.139 | 200.2123 | 14 | 0.99 |
| polarC18 | 218.139 | 51.5544 | 14 | 0.99 |

| | | | | |
|---|---|---|---|---|
| polarC18 | 218.139 | 100.2929 | 14 | 0.99 |
| polarC18 | 218.139 | 55.3262 | 14 | 0.99 |
| polarC18 | 218.139 | 115.6433 | 14 | 0.99 |
| polarC18 | 218.139 | 200.6688 | 14 | 0.99 |
| polarC18 | 218.139 | 85.2906 | 14 | 0.99 |
| polarC18 | 218.139 | 227.6639 | 14 | 0.99 |
| polarC18 | 218.139 | 93.676 | 14 | 0.99 |
| polarC18 | 218.139 | 56.4926 | 14 | 0.99 |
| polarC18 | 218.139 | 52.7729 | 14 | 0.99 |
| polarC18 | 218.139 | 195.4376 | 14 | 0.99 |
| polarC18 | 218.139 | 240.4377 | 14 | 0.99 |
| polarC18 | 218.139 | 64.714 | 14 | 0.99 |
| polarC18 | 218.139 | 217.0996 | 14 | 0.99 |
| polarC18 | 218.139 | 66.1124 | 14 | 0.99 |
| polarC18 | 218.139 | 101.6632 | 14 | 0.99 |
| polarC18 | 218.139 | 93.8249 | 14 | 0.99 |
| polarC18 | 218.139 | 206.2058 | 14 | 0.99 |
| polarC18 | 218.139 | 227.2506 | 14 | 0.99 |
| polarC18 | 218.139 | 55.5687 | 14 | 0.99 |
| polarC18 | 218.139 | 94.6552 | 14 | 0.99 |
| polarC18 | 218.139 | 94.2009 | 14 | 0.99 |
| polarC18 | 218.139 | 49.9459 | 13.9 | 0.99 |
| polarC18 | 218.139 | 48.9254 | 13.9 | 0.99 |
| polarC18 | 218.139 | 111.2133 | 13.9 | 0.99 |
| polarC18 | 218.139 | 129.6213 | 13.9 | 0.99 |
| polarC18 | 218.139 | 220.6485 | 13.9 | 0.99 |
| polarC18 | 218.139 | 100.376 | 13.9 | 0.99 |
| polarC18 | 218.139 | 135.7551 | 13.9 | 0.99 |
| polarC18 | 218.139 | 63.5782 | 13.9 | 0.99 |
| polarC18 | 218.139 | 121.9204 | 13.9 | 0.99 |
| polarC18 | 218.139 | 204.6871 | 13.9 | 0.99 |
| polarC18 | 218.139 | 171.6468 | 13.9 | 0.99 |
| polarC18 | 218.139 | 194.4847 | 13.9 | 0.99 |
| polarC18 | 218.139 | 199.9487 | 13.9 | 0.99 |
| polarC18 | 218.139 | 69.1299 | 13.9 | 0.99 |
| polarC18 | 218.139 | 55.1674 | 13.9 | 0.99 |
| polarC18 | 218.139 | 102.8233 | 13.9 | 0.99 |
| polarC18 | 218.139 | 149.5894 | 13.9 | 0.99 |
| polarC18 | 218.139 | 129.1158 | 13.9 | 0.99 |
| polarC18 | 218.139 | 134.4329 | 13.9 | 0.99 |
| polarC18 | 218.139 | 58.072 | 13.9 | 0.99 |
| polarC18 | 218.139 | 203.8762 | 13.9 | 0.99 |

| | | | | |
|---|---|---|---|---|
| polarC18 | 218.139 | 167.894 | 13.9 | 0.99 |
| polarC18 | 218.139 | 71.2966 | 13.9 | 0.98 |
| polarC18 | 218.139 | 119.3242 | 13.9 | 0.98 |
| polarC18 | 218.139 | 167.8541 | 13.9 | 0.98 |
| polarC18 | 218.139 | 150.2798 | 13.9 | 0.98 |
| polarC18 | 218.139 | 82.513 | 13.9 | 0.98 |
| polarC18 | 218.139 | 60.3785 | 13.9 | 0.98 |
| polarC18 | 218.139 | 78.3193 | 13.9 | 0.98 |
| polarC18 | 218.139 | 65.9839 | 13.9 | 0.98 |
| polarC18 | 218.139 | 63.4515 | 13.9 | 0.98 |
| polarC18 | 218.139 | 182.2336 | 13.9 | 0.98 |
| polarC18 | 218.139 | 69.9663 | 13.9 | 0.98 |
| polarC18 | 218.139 | 167.9891 | 13.9 | 0.98 |
| polarC18 | 218.139 | 189.5702 | 13.9 | 0.98 |
| polarC18 | 218.139 | 174.2602 | 13.9 | 0.98 |
| polarC18 | 218.139 | 141.678 | 13.9 | 0.98 |
| polarC18 | 218.139 | 96.7844 | 13.9 | 0.98 |
| polarC18 | 218.139 | 232.039 | 13.8 | 0.98 |
| polarC18 | 218.139 | 71.8274 | 13.8 | 0.98 |
| polarC18 | 218.139 | 52.4037 | 13.8 | 0.98 |
| polarC18 | 218.139 | 56.3564 | 13.8 | 0.98 |
| polarC18 | 218.139 | 53.2937 | 13.8 | 0.98 |
| polarC18 | 218.139 | 56.0857 | 13.8 | 0.98 |
| polarC18 | 218.139 | 217.8396 | 13.8 | 0.98 |
| polarC18 | 218.139 | 90.866 | 13.8 | 0.98 |
| polarC18 | 218.139 | 77.3755 | 13.8 | 0.98 |
| polarC18 | 218.139 | 91.213 | 13.8 | 0.98 |
| polarC18 | 218.139 | 56.1599 | 13.8 | 0.98 |
| polarC18 | 218.139 | 59.2067 | 13.8 | 0.98 |
| polarC18 | 218.139 | 86.0452 | 13.8 | 0.98 |
| polarC18 | 218.139 | 89.3532 | 13.8 | 0.98 |
| polarC18 | 218.139 | 127.4903 | 13.8 | 0.98 |
| polarC18 | 218.139 | 133.6604 | 13.8 | 0.98 |
| polarC18 | 218.139 | 144.8964 | 13.8 | 0.98 |
| polarC18 | 218.139 | 240.7212 | 13.8 | 0.98 |
| polarC18 | 218.139 | 63.97 | 13.8 | 0.98 |
| polarC18 | 218.139 | 103.4389 | 13.8 | 0.98 |
| polarC18 | 218.139 | 232.6484 | 13.8 | 0.98 |
| polarC18 | 218.139 | 117.4208 | 13.8 | 0.98 |
| polarC18 | 218.139 | 86.7592 | 13.8 | 0.98 |
| polarC18 | 218.139 | 205.8641 | 13.8 | 0.98 |
| polarC18 | 218.139 | 159.6726 | 13.8 | 0.98 |

| | | | | |
|---|---|---|---|---|
| polarC18 | 218.139 | 59.4165 | 13.8 | 0.98 |
| polarC18 | 218.139 | 72.7851 | 13.8 | 0.98 |
| polarC18 | 218.139 | 143.8958 | 13.8 | 0.98 |
| polarC18 | 218.139 | 109.0585 | 13.8 | 0.98 |
| polarC18 | 218.139 | 79.7974 | 13.8 | 0.98 |
| polarC18 | 218.139 | 166.7979 | 13.8 | 0.98 |
| polarC18 | 218.139 | 117.6065 | 13.8 | 0.98 |
| polarC18 | 218.139 | 51.0133 | 13.8 | 0.98 |
| polarC18 | 218.139 | 69.3732 | 13.8 | 0.98 |
| polarC18 | 218.139 | 219.7381 | 13.7 | 0.98 |
| polarC18 | 218.139 | 105.0217 | 13.7 | 0.98 |
| polarC18 | 218.139 | 77.5721 | 13.7 | 0.98 |
| polarC18 | 218.139 | 173.7851 | 13.7 | 0.98 |
| polarC18 | 218.139 | 123.0479 | 13.7 | 0.98 |
| polarC18 | 218.139 | 173.0519 | 13.7 | 0.97 |
| polarC18 | 218.139 | 128.2599 | 13.7 | 0.97 |
| polarC18 | 218.139 | 104.3753 | 13.7 | 0.97 |
| polarC18 | 218.139 | 70.1769 | 13.7 | 0.97 |
| polarC18 | 218.139 | 171.8317 | 13.7 | 0.97 |
| polarC18 | 218.139 | 192.1566 | 13.7 | 0.97 |
| polarC18 | 218.139 | 82.7962 | 13.7 | 0.97 |
| polarC18 | 218.139 | 66.6498 | 13.7 | 0.97 |
| polarC18 | 218.139 | 66.6214 | 13.7 | 0.97 |
| polarC18 | 218.139 | 52.4745 | 13.7 | 0.97 |
| polarC18 | 218.139 | 207.1265 | 13.7 | 0.97 |
| polarC18 | 218.139 | 58.5948 | 13.7 | 0.97 |
| polarC18 | 218.139 | 90.7052 | 13.7 | 0.97 |
| polarC18 | 218.139 | 109.8989 | 13.7 | 0.97 |
| polarC18 | 218.139 | 80.1475 | 13.7 | 0.97 |
| polarC18 | 218.139 | 52.9558 | 13.7 | 0.97 |
| polarC18 | 218.139 | 134.7691 | 13.7 | 0.97 |
| polarC18 | 218.139 | 169.5113 | 13.7 | 0.97 |
| polarC18 | 218.139 | 165.7278 | 13.7 | 0.97 |
| polarC18 | 218.139 | 64.978 | 13.7 | 0.97 |
| polarC18 | 218.139 | 69.4904 | 13.7 | 0.97 |
| polarC18 | 218.139 | 91.9389 | 13.7 | 0.97 |
| polarC18 | 218.139 | 74.3847 | 13.7 | 0.97 |
| polarC18 | 218.139 | 51.0349 | 13.7 | 0.97 |
| polarC18 | 218.139 | 123.4192 | 13.7 | 0.97 |
| polarC18 | 218.139 | 51.8351 | 13.7 | 0.97 |
| polarC18 | 218.139 | 69.7168 | 13.7 | 0.97 |
| polarC18 | 218.139 | 78.9054 | 13.7 | 0.97 |

| | | | | |
|---|---|---|---|---|
| polarC18 | 218.139 | 239.083 | 13.7 | 0.97 |
| polarC18 | 218.139 | 57.2529 | 13.7 | 0.97 |
| polarC18 | 218.139 | 141.097 | 13.7 | 0.97 |
| polarC18 | 218.139 | 118.5622 | 13.7 | 0.97 |
| polarC18 | 218.139 | 88.186 | 13.7 | 0.97 |
| polarC18 | 218.139 | 93.5838 | 13.7 | 0.97 |
| polarC18 | 218.139 | 77.204 | 13.7 | 0.97 |
| polarC18 | 218.139 | 67.7268 | 13.7 | 0.97 |
| polarC18 | 218.139 | 176.2838 | 13.7 | 0.97 |
| polarC18 | 218.139 | 107.71 | 13.7 | 0.97 |
| polarC18 | 218.139 | 138.7635 | 13.7 | 0.97 |
| polarC18 | 218.139 | 109.7077 | 13.7 | 0.97 |
| polarC18 | 218.139 | 109.7524 | 13.7 | 0.97 |
| polarC18 | 218.139 | 68.4739 | 13.6 | 0.97 |
| polarC18 | 218.139 | 195.4659 | 13.6 | 0.97 |
| polarC18 | 218.139 | 90.7665 | 13.6 | 0.97 |
| polarC18 | 218.139 | 151.7451 | 13.6 | 0.97 |
| polarC18 | 218.139 | 75.7375 | 13.6 | 0.97 |
| polarC18 | 218.139 | 142.8107 | 13.6 | 0.97 |
| polarC18 | 218.139 | 58.2805 | 13.6 | 0.97 |
| polarC18 | 218.139 | 87.3043 | 13.6 | 0.97 |
| polarC18 | 218.139 | 57.5973 | 13.6 | 0.97 |
| polarC18 | 218.139 | 174.1082 | 13.6 | 0.97 |
| polarC18 | 218.139 | 59.6305 | 13.6 | 0.97 |
| polarC18 | 218.139 | 71.1429 | 13.6 | 0.97 |
| polarC18 | 218.139 | 59.0691 | 13.6 | 0.97 |
| polarC18 | 218.139 | 150.9195 | 13.6 | 0.97 |
| polarC18 | 218.139 | 61.8071 | 13.6 | 0.97 |
| polarC18 | 218.139 | 62.5922 | 13.6 | 0.97 |
| polarC18 | 218.139 | 134.5553 | 13.6 | 0.97 |
| polarC18 | 218.139 | 87.9303 | 13.6 | 0.97 |
| polarC18 | 218.139 | 87.3294 | 13.6 | 0.97 |
| polarC18 | 218.139 | 125.6301 | 13.6 | 0.97 |
| polarC18 | 218.139 | 117.6478 | 13.6 | 0.97 |
| polarC18 | 218.139 | 50.3953 | 13.6 | 0.97 |
| polarC18 | 218.139 | 93.3711 | 13.6 | 0.97 |
| polarC18 | 218.139 | 123.1412 | 13.6 | 0.97 |
| polarC18 | 218.139 | 219.492 | 13.6 | 0.97 |
| polarC18 | 218.139 | 94.1013 | 13.6 | 0.97 |
| polarC18 | 218.139 | 49.139 | 13.6 | 0.97 |
| polarC18 | 218.139 | 139.6067 | 13.6 | 0.96 |
| polarC18 | 218.139 | 155.4108 | 13.6 | 0.96 |

| | | | | |
|---|---|---|---|---|
| polarC18 | 218.139 | 206.7906 | 13.6 | 0.96 |
| polarC18 | 218.139 | 163.3295 | 13.6 | 0.96 |
| polarC18 | 218.139 | 79.69 | 13.6 | 0.96 |
| polarC18 | 218.139 | 66.4393 | 13.6 | 0.96 |
| polarC18 | 218.139 | 143.4208 | 13.6 | 0.96 |
| polarC18 | 218.139 | 58.7728 | 13.6 | 0.96 |
| polarC18 | 218.139 | 222.0014 | 13.6 | 0.96 |
| polarC18 | 218.139 | 78.2443 | 13.6 | 0.96 |
| polarC18 | 218.139 | 194.6964 | 13.6 | 0.96 |
| polarC18 | 218.139 | 60.1188 | 13.6 | 0.96 |
| polarC18 | 218.139 | 61.7492 | 13.6 | 0.96 |
| polarC18 | 218.139 | 187.4092 | 13.6 | 0.96 |
| polarC18 | 218.139 | 58.8688 | 13.6 | 0.96 |
| polarC18 | 218.139 | 59.558 | 13.6 | 0.96 |
| polarC18 | 218.139 | 211.2767 | 13.6 | 0.96 |
| polarC18 | 218.139 | 66.8697 | 13.6 | 0.96 |
| polarC18 | 218.139 | 201.9539 | 13.6 | 0.96 |
| polarC18 | 218.139 | 170.7127 | 13.6 | 0.96 |
| polarC18 | 218.139 | 181.0616 | 13.6 | 0.96 |
| polarC18 | 218.139 | 88.574 | 13.6 | 0.96 |
| polarC18 | 218.139 | 165.7491 | 13.5 | 0.96 |
| polarC18 | 218.139 | 102.5842 | 13.5 | 0.96 |
| polarC18 | 218.139 | 49.4246 | 13.5 | 0.96 |
| polarC18 | 218.139 | 50.4684 | 13.5 | 0.96 |
| polarC18 | 218.139 | 75.2272 | 13.5 | 0.96 |
| polarC18 | 218.139 | 96.6609 | 13.5 | 0.96 |
| polarC18 | 218.139 | 90.732 | 13.5 | 0.96 |
| polarC18 | 218.139 | 136.0434 | 13.5 | 0.96 |
| polarC18 | 218.139 | 102.5757 | 13.5 | 0.96 |
| polarC18 | 218.139 | 93.5236 | 13.5 | 0.96 |
| polarC18 | 218.139 | 193.6526 | 13.5 | 0.96 |
| polarC18 | 218.139 | 89.2203 | 13.5 | 0.96 |
| polarC18 | 218.139 | 50.2941 | 13.5 | 0.96 |
| polarC18 | 218.139 | 69.5868 | 13.5 | 0.96 |
| polarC18 | 218.139 | 52.6754 | 13.5 | 0.96 |
| polarC18 | 218.139 | 67.7037 | 13.5 | 0.96 |
| polarC18 | 218.139 | 86.2441 | 13.5 | 0.96 |
| polarC18 | 218.139 | 69.2653 | 13.5 | 0.96 |
| polarC18 | 218.139 | 165.6702 | 13.5 | 0.96 |
| polarC18 | 218.139 | 54.0123 | 13.5 | 0.96 |
| polarC18 | 218.139 | 201.6104 | 13.5 | 0.96 |
| polarC18 | 218.139 | 58.976 | 13.5 | 0.96 |

| | | | | |
|---|---|---|---|---|
| polarC18 | 218.139 | 167.2251 | 13.5 | 0.96 |
| polarC18 | 218.139 | 85.337 | 13.5 | 0.96 |
| polarC18 | 218.139 | 63.8116 | 13.5 | 0.96 |
| polarC18 | 218.139 | 61.159 | 13.5 | 0.96 |
| polarC18 | 218.139 | 51.4369 | 13.5 | 0.96 |
| polarC18 | 218.139 | 95.1104 | 13.5 | 0.96 |
| polarC18 | 218.139 | 203.0081 | 13.5 | 0.96 |
| polarC18 | 218.139 | 75.0757 | 13.5 | 0.96 |
| polarC18 | 218.139 | 67.8374 | 13.5 | 0.96 |
| polarC18 | 218.139 | 78.4764 | 13.5 | 0.96 |
| polarC18 | 218.139 | 206.4903 | 13.5 | 0.96 |
| polarC18 | 218.139 | 99.5211 | 13.5 | 0.96 |
| polarC18 | 218.139 | 157.3504 | 13.5 | 0.96 |
| polarC18 | 218.139 | 73.9448 | 13.5 | 0.96 |
| polarC18 | 218.139 | 223.9726 | 13.5 | 0.96 |
| polarC18 | 218.139 | 136.9413 | 13.5 | 0.96 |
| polarC18 | 218.139 | 123.8492 | 13.5 | 0.96 |
| polarC18 | 218.139 | 118.5452 | 13.5 | 0.96 |
| polarC18 | 218.139 | 125.0871 | 13.5 | 0.95 |
| polarC18 | 218.139 | 64.8336 | 13.5 | 0.95 |
| polarC18 | 218.139 | 226.9769 | 13.5 | 0.95 |
| polarC18 | 218.139 | 54.1295 | 13.5 | 0.95 |
| polarC18 | 218.139 | 115.4864 | 13.5 | 0.95 |
| polarC18 | 218.139 | 97.3827 | 13.5 | 0.95 |
| polarC18 | 218.139 | 130.5265 | 13.5 | 0.95 |
| polarC18 | 218.139 | 51.3028 | 13.5 | 0.95 |
| polarC18 | 218.139 | 191.2181 | 13.4 | 0.95 |
| polarC18 | 218.139 | 176.8482 | 13.4 | 0.95 |
| polarC18 | 218.139 | 95.0777 | 13.4 | 0.95 |
| polarC18 | 218.139 | 68.2867 | 13.4 | 0.95 |
| polarC18 | 218.139 | 126.0446 | 13.4 | 0.95 |
| polarC18 | 218.139 | 77.0824 | 13.4 | 0.95 |
| polarC18 | 218.139 | 64.1026 | 13.4 | 0.95 |
| polarC18 | 218.139 | 204.4392 | 13.4 | 0.95 |
| polarC18 | 218.139 | 154.7684 | 13.4 | 0.95 |
| polarC18 | 218.139 | 172.4585 | 13.4 | 0.95 |
| polarC18 | 218.139 | 49.0123 | 13.4 | 0.95 |
| polarC18 | 218.139 | 105.8209 | 13.4 | 0.95 |
| polarC18 | 218.139 | 177.8243 | 13.4 | 0.95 |
| polarC18 | 218.139 | 232.3738 | 13.4 | 0.95 |
| polarC18 | 218.139 | 149.3552 | 13.4 | 0.95 |
| polarC18 | 218.139 | 72.3249 | 13.4 | 0.95 |

| | | | | |
|---|---|---|---|---|
| polarC18 | 218.139 | 109.4784 | 13.4 | 0.95 |
| polarC18 | 218.139 | 69.1741 | 13.4 | 0.95 |
| polarC18 | 218.139 | 158.5616 | 13.4 | 0.95 |
| polarC18 | 218.139 | 91.3478 | 13.4 | 0.95 |
| polarC18 | 218.139 | 79.5866 | 13.4 | 0.95 |
| polarC18 | 218.139 | 90.0055 | 13.4 | 0.95 |
| polarC18 | 218.139 | 134.6422 | 13.4 | 0.95 |
| polarC18 | 218.139 | 113.5761 | 13.4 | 0.95 |
| polarC18 | 218.139 | 108.9478 | 13.4 | 0.95 |
| polarC18 | 218.139 | 82.5238 | 13.4 | 0.95 |
| polarC18 | 218.139 | 60.9871 | 13.4 | 0.95 |
| polarC18 | 218.139 | 215.7963 | 13.4 | 0.95 |
| polarC18 | 218.139 | 66.1429 | 13.4 | 0.95 |
| polarC18 | 218.139 | 85.1252 | 13.4 | 0.95 |
| polarC18 | 218.139 | 78.3251 | 13.4 | 0.95 |
| polarC18 | 218.139 | 103.2267 | 13.4 | 0.95 |
| polarC18 | 218.139 | 87.2726 | 13.4 | 0.95 |
| polarC18 | 218.139 | 158.0677 | 13.4 | 0.95 |
| polarC18 | 218.139 | 49.9874 | 13.4 | 0.95 |
| polarC18 | 218.139 | 59.9885 | 13.4 | 0.95 |
| polarC18 | 218.139 | 110.4687 | 13.4 | 0.95 |
| polarC18 | 218.139 | 59.5139 | 13.4 | 0.95 |
| polarC18 | 218.139 | 137.7982 | 13.4 | 0.95 |
| polarC18 | 218.139 | 81.111 | 13.4 | 0.95 |
| polarC18 | 218.139 | 111.7665 | 13.4 | 0.95 |
| polarC18 | 218.139 | 138.3909 | 13.4 | 0.95 |
| polarC18 | 218.139 | 141.4766 | 13.4 | 0.95 |
| polarC18 | 218.139 | 58.8216 | 13.4 | 0.95 |
| polarC18 | 218.139 | 53.8979 | 13.3 | 0.95 |
| polarC18 | 218.139 | 223.8536 | 13.3 | 0.95 |
| polarC18 | 218.139 | 84.6076 | 13.3 | 0.95 |
| polarC18 | 218.139 | 91.5255 | 13.3 | 0.95 |
| polarC18 | 218.139 | 58.2657 | 13.3 | 0.95 |
| polarC18 | 218.139 | 65.5362 | 13.3 | 0.95 |
| polarC18 | 218.139 | 134.7177 | 13.3 | 0.95 |
| polarC18 | 218.139 | 191.2729 | 13.3 | 0.95 |
| polarC18 | 218.139 | 76.777 | 13.3 | 0.95 |
| polarC18 | 218.139 | 63.7908 | 13.3 | 0.95 |
| polarC18 | 218.139 | 70.7877 | 13.3 | 0.95 |
| polarC18 | 218.139 | 189.939 | 13.3 | 0.95 |
| polarC18 | 218.139 | 87.5224 | 13.3 | 0.94 |
| polarC18 | 218.139 | 95.5876 | 13.3 | 0.94 |

| | | | | |
|---|---|---|---|---|
| polarC18 | 218.139 | 62.2452 | 13.3 | 0.94 |
| polarC18 | 218.139 | 96.1471 | 13.3 | 0.94 |
| polarC18 | 218.139 | 123.5978 | 13.3 | 0.94 |
| polarC18 | 218.139 | 62.7436 | 13.3 | 0.94 |
| polarC18 | 218.139 | 61.1359 | 13.3 | 0.94 |
| polarC18 | 218.139 | 158.588 | 13.3 | 0.94 |
| polarC18 | 218.139 | 229.4822 | 13.3 | 0.94 |
| polarC18 | 218.139 | 73.7075 | 13.3 | 0.94 |
| polarC18 | 218.139 | 52.9661 | 13.3 | 0.94 |
| polarC18 | 218.139 | 55.8412 | 13.3 | 0.94 |
| polarC18 | 218.139 | 195.0182 | 13.3 | 0.94 |
| polarC18 | 218.139 | 151.1082 | 13.3 | 0.94 |
| polarC18 | 218.139 | 210.5797 | 13.3 | 0.94 |
| polarC18 | 218.139 | 84.0062 | 13.3 | 0.94 |
| polarC18 | 218.139 | 174.7465 | 13.3 | 0.94 |
| polarC18 | 218.139 | 49.4443 | 13.3 | 0.94 |
| polarC18 | 218.139 | 125.2375 | 13.3 | 0.94 |
| polarC18 | 218.139 | 53.5701 | 13.3 | 0.94 |
| polarC18 | 218.139 | 67.4774 | 13.3 | 0.94 |
| polarC18 | 218.139 | 148.9729 | 13.3 | 0.94 |
| polarC18 | 218.139 | 224.8158 | 13.3 | 0.94 |
| polarC18 | 218.139 | 200.1004 | 13.3 | 0.94 |
| polarC18 | 218.139 | 63.9352 | 13.3 | 0.94 |
| polarC18 | 218.139 | 112.0696 | 13.3 | 0.94 |
| polarC18 | 218.139 | 227.1303 | 13.3 | 0.94 |
| polarC18 | 218.139 | 51.5601 | 13.2 | 0.94 |
| polarC18 | 218.139 | 58.3116 | 13.2 | 0.94 |
| polarC18 | 218.139 | 138.476 | 13.2 | 0.94 |
| polarC18 | 218.139 | 62.4659 | 13.2 | 0.94 |
| polarC18 | 218.139 | 141.2699 | 13.2 | 0.94 |
| polarC18 | 218.139 | 239.2945 | 13.2 | 0.94 |
| polarC18 | 218.139 | 68.6015 | 13.2 | 0.94 |
| polarC18 | 218.139 | 147.5262 | 13.2 | 0.94 |
| polarC18 | 218.139 | 139.7833 | 13.2 | 0.94 |
| polarC18 | 218.139 | 224.1549 | 13.2 | 0.94 |
| polarC18 | 218.139 | 83.7984 | 13.2 | 0.94 |
| polarC18 | 218.139 | 205.9844 | 13.2 | 0.94 |
| polarC18 | 218.139 | 109.7304 | 13.2 | 0.94 |
| polarC18 | 218.139 | 170.2367 | 13.2 | 0.94 |
| polarC18 | 218.139 | 140.1597 | 13.2 | 0.94 |
| polarC18 | 218.139 | 98.1877 | 13.2 | 0.94 |
| polarC18 | 218.139 | 67.6715 | 13.2 | 0.94 |

| | | | | |
|---|---|---|---|---|
| polarC18 | 218.139 | 104.9111 | 13.2 | 0.94 |
| polarC18 | 218.139 | 93.3325 | 13.2 | 0.94 |
| polarC18 | 218.139 | 91.3551 | 13.2 | 0.94 |
| polarC18 | 218.139 | 76.4619 | 13.2 | 0.94 |
| polarC18 | 218.139 | 83.5172 | 13.2 | 0.94 |
| polarC18 | 218.139 | 111.367 | 13.2 | 0.94 |
| polarC18 | 218.139 | 63.9009 | 13.2 | 0.94 |
| polarC18 | 218.139 | 146.1002 | 13.2 | 0.94 |
| polarC18 | 218.139 | 127.7887 | 13.2 | 0.94 |
| polarC18 | 218.139 | 150.2436 | 13.2 | 0.94 |
| polarC18 | 218.139 | 77.2181 | 13.2 | 0.94 |
| polarC18 | 218.139 | 112.3372 | 13.2 | 0.94 |
| polarC18 | 218.139 | 115.671 | 13.2 | 0.94 |
| polarC18 | 218.139 | 71.5028 | 13.2 | 0.94 |
| polarC18 | 218.139 | 109.133 | 13.2 | 0.94 |
| polarC18 | 218.139 | 77.7642 | 13.2 | 0.94 |
| polarC18 | 218.139 | 114.3772 | 13.2 | 0.94 |
| polarC18 | 218.139 | 121.1 | 13.2 | 0.94 |
| polarC18 | 218.139 | 89.0284 | 13.2 | 0.94 |
| polarC18 | 218.139 | 208.5011 | 13.2 | 0.94 |
| polarC18 | 218.139 | 214.3323 | 13.2 | 0.94 |
| polarC18 | 218.139 | 149.913 | 13.2 | 0.94 |
| polarC18 | 218.139 | 71.9775 | 13.2 | 0.94 |
| polarC18 | 218.139 | 75.923 | 13.2 | 0.94 |
| polarC18 | 218.139 | 63.7076 | 13.2 | 0.94 |
| polarC18 | 218.139 | 71.4447 | 13.2 | 0.93 |
| polarC18 | 218.139 | 50.122 | 13.2 | 0.93 |
| polarC18 | 218.139 | 80.9476 | 13.2 | 0.93 |
| polarC18 | 218.139 | 50.2054 | 13.2 | 0.93 |
| polarC18 | 218.139 | 93.5944 | 13.2 | 0.93 |
| polarC18 | 218.139 | 121.4956 | 13.2 | 0.93 |
| polarC18 | 218.139 | 49.2937 | 13.2 | 0.93 |
| polarC18 | 218.139 | 48.8792 | 13.2 | 0.93 |
| polarC18 | 218.139 | 70.4502 | 13.2 | 0.93 |
| polarC18 | 218.139 | 51.5139 | 13.2 | 0.93 |
| polarC18 | 218.139 | 52.1222 | 13.2 | 0.93 |
| polarC18 | 218.139 | 100.8655 | 13.2 | 0.93 |
| polarC18 | 218.139 | 157.5856 | 13.2 | 0.93 |
| polarC18 | 218.139 | 222.7988 | 13.2 | 0.93 |
| polarC18 | 218.139 | 146.5252 | 13.2 | 0.93 |
| polarC18 | 218.139 | 132.543 | 13.2 | 0.93 |
| polarC18 | 218.139 | 181.3144 | 13.2 | 0.93 |

| | | | | |
|---|---|---|---|---|
| polarC18 | 218.139 | 58.8843 | 13.1 | 0.93 |
| polarC18 | 218.139 | 130.9505 | 13.1 | 0.93 |
| polarC18 | 218.139 | 99.8267 | 13.1 | 0.93 |
| polarC18 | 218.139 | 61.1882 | 13.1 | 0.93 |
| polarC18 | 218.139 | 105.1627 | 13.1 | 0.93 |
| polarC18 | 218.139 | 50.7131 | 13.1 | 0.93 |
| polarC18 | 218.139 | 184.9274 | 13.1 | 0.93 |
| polarC18 | 218.139 | 48.9833 | 13.1 | 0.93 |
| polarC18 | 218.139 | 62.0544 | 13.1 | 0.93 |
| polarC18 | 218.139 | 70.5745 | 13.1 | 0.93 |
| polarC18 | 218.139 | 154.1251 | 13.1 | 0.93 |
| polarC18 | 218.139 | 196.4236 | 13.1 | 0.93 |
| polarC18 | 218.139 | 139.4084 | 13.1 | 0.93 |
| polarC18 | 218.139 | 77.9502 | 13.1 | 0.93 |
| polarC18 | 218.139 | 68.8292 | 13.1 | 0.93 |
| polarC18 | 218.139 | 57.8869 | 13.1 | 0.93 |
| polarC18 | 218.139 | 220.4144 | 13.1 | 0.93 |
| polarC18 | 218.139 | 91.7196 | 13.1 | 0.93 |
| polarC18 | 218.139 | 212.3848 | 13.1 | 0.93 |
| polarC18 | 218.139 | 117.3819 | 13.1 | 0.93 |
| polarC18 | 218.139 | 105.2953 | 13.1 | 0.93 |
| polarC18 | 218.139 | 129.1518 | 13.1 | 0.93 |
| polarC18 | 218.139 | 203.3248 | 13.1 | 0.93 |
| polarC18 | 218.139 | 59.0908 | 13.1 | 0.93 |
| polarC18 | 218.139 | 66.8665 | 13.1 | 0.93 |
| polarC18 | 218.139 | 233.0551 | 13.1 | 0.93 |
| polarC18 | 218.139 | 138.0133 | 13.1 | 0.93 |
| polarC18 | 218.139 | 66.6159 | 13.1 | 0.93 |
| polarC18 | 218.139 | 78.26 | 13.1 | 0.93 |
| polarC18 | 218.139 | 115.7116 | 13.1 | 0.93 |
| polarC18 | 218.139 | 102.6283 | 13.1 | 0.93 |
| polarC18 | 218.139 | 101.5294 | 13.1 | 0.93 |
| polarC18 | 218.139 | 57.1091 | 13.1 | 0.93 |
| polarC18 | 218.139 | 65.9335 | 13.1 | 0.93 |
| polarC18 | 218.139 | 199.6302 | 13.1 | 0.93 |
| polarC18 | 218.139 | 142.2184 | 13.1 | 0.93 |
| polarC18 | 218.139 | 66.6636 | 13.1 | 0.93 |
| polarC18 | 218.139 | 61.0166 | 13.1 | 0.93 |
| polarC18 | 218.139 | 63.4453 | 13.1 | 0.93 |
| polarC18 | 218.139 | 65.3581 | 13.1 | 0.93 |
| polarC18 | 218.139 | 155.9199 | 13.1 | 0.93 |
| polarC18 | 218.139 | 128.2383 | 13.1 | 0.93 |

| | | | | |
|---|---|---|---|---|
| polarC18 | 218.139 | 93.7269 | 13.1 | 0.93 |
| polarC18 | 218.139 | 59.7112 | 13.1 | 0.93 |
| polarC18 | 218.139 | 74.5719 | 13.1 | 0.93 |
| polarC18 | 218.139 | 50.4305 | 13.1 | 0.93 |
| polarC18 | 218.139 | 90.9401 | 13.1 | 0.93 |
| polarC18 | 218.139 | 54.7572 | 13.1 | 0.93 |
| polarC18 | 218.139 | 97.3963 | 13.1 | 0.93 |
| polarC18 | 218.139 | 66.0714 | 13.1 | 0.93 |
| polarC18 | 218.139 | 112.8958 | 13.1 | 0.93 |
| polarC18 | 218.139 | 73.3998 | 13.1 | 0.93 |
| polarC18 | 218.139 | 190.5916 | 13.1 | 0.93 |
| polarC18 | 218.139 | 52.6159 | 13.1 | 0.93 |
| polarC18 | 218.139 | 104.0736 | 13.1 | 0.93 |
| polarC18 | 218.139 | 148.268 | 13.1 | 0.93 |
| polarC18 | 218.139 | 102.7609 | 13.1 | 0.93 |
| polarC18 | 218.139 | 114.395 | 13.1 | 0.93 |
| polarC18 | 218.139 | 70.9168 | 13.1 | 0.93 |
| polarC18 | 218.139 | 66.0058 | 13.1 | 0.93 |
| polarC18 | 218.139 | 171.9546 | 13.1 | 0.93 |
| polarC18 | 218.139 | 60.7487 | 13.1 | 0.93 |
| polarC18 | 218.139 | 64.9071 | 13.1 | 0.93 |
| polarC18 | 218.139 | 87.2277 | 13 | 0.93 |
| polarC18 | 218.139 | 219.102 | 13 | 0.93 |
| polarC18 | 218.139 | 229.9477 | 13 | 0.93 |
| polarC18 | 218.139 | 189.9721 | 13 | 0.93 |
| polarC18 | 218.139 | 194.0532 | 13 | 0.93 |
| polarC18 | 218.139 | 67.0347 | 13 | 0.93 |
| polarC18 | 218.139 | 102.9796 | 13 | 0.93 |
| polarC18 | 218.139 | 62.2309 | 13 | 0.93 |
| polarC18 | 218.139 | 131.1215 | 13 | 0.93 |
| polarC18 | 218.139 | 95.2865 | 13 | 0.92 |
| polarC18 | 218.139 | 50.4752 | 13 | 0.92 |
| polarC18 | 218.139 | 104.0983 | 13 | 0.92 |
| polarC18 | 218.139 | 62.9147 | 13 | 0.92 |
| polarC18 | 218.139 | 131.2427 | 13 | 0.92 |
| polarC18 | 218.139 | 81.7857 | 13 | 0.92 |
| polarC18 | 218.139 | 125.7535 | 13 | 0.92 |
| polarC18 | 218.139 | 201.8558 | 13 | 0.92 |
| polarC18 | 218.139 | 79.912 | 13 | 0.92 |
| polarC18 | 218.139 | 148.8175 | 13 | 0.92 |
| polarC18 | 218.139 | 77.0054 | 13 | 0.92 |
| polarC18 | 218.139 | 94.1606 | 13 | 0.92 |

| | | | | |
|---|---|---|---|---|
| polarC18 | 218.139 | 89.9872 | 13 | 0.92 |
| polarC18 | 218.139 | 83.943 | 13 | 0.92 |
| polarC18 | 218.139 | 113.0431 | 13 | 0.92 |
| polarC18 | 218.139 | 117.1158 | 13 | 0.92 |
| polarC18 | 218.139 | 93.3778 | 13 | 0.92 |
| polarC18 | 218.139 | 166.4117 | 13 | 0.92 |
| polarC18 | 218.139 | 164.2638 | 13 | 0.92 |
| polarC18 | 218.139 | 167.3348 | 13 | 0.92 |
| polarC18 | 218.139 | 90.0513 | 13 | 0.92 |
| polarC18 | 218.139 | 130.0505 | 13 | 0.92 |
| polarC18 | 218.139 | 147.3617 | 13 | 0.92 |
| polarC18 | 218.139 | 71.4714 | 13 | 0.92 |
| polarC18 | 218.139 | 114.3402 | 13 | 0.92 |
| polarC18 | 218.139 | 52.463 | 13 | 0.92 |
| polarC18 | 218.139 | 60.6648 | 13 | 0.92 |
| polarC18 | 218.139 | 108.4228 | 13 | 0.92 |
| polarC18 | 218.139 | 137.3049 | 13 | 0.92 |
| polarC18 | 218.139 | 58.6869 | 13 | 0.92 |
| polarC18 | 218.139 | 98.9234 | 13 | 0.92 |
| polarC18 | 218.139 | 61.5029 | 13 | 0.92 |
| polarC18 | 218.139 | 128.0785 | 13 | 0.92 |
| polarC18 | 218.139 | 140.296 | 13 | 0.92 |
| polarC18 | 218.139 | 50.4494 | 13 | 0.92 |
| polarC18 | 218.139 | 115.4747 | 13 | 0.92 |
| polarC18 | 218.139 | 96.7305 | 13 | 0.92 |
| polarC18 | 218.139 | 143.2695 | 13 | 0.92 |
| polarC18 | 218.139 | 78.2394 | 13 | 0.92 |
| polarC18 | 218.139 | 55.7816 | 13 | 0.92 |
| polarC18 | 218.139 | 222.867 | 13 | 0.92 |
| polarC18 | 218.139 | 70.3874 | 13 | 0.92 |
| polarC18 | 218.139 | 53.1766 | 13 | 0.92 |
| polarC18 | 218.139 | 78.3387 | 13 | 0.92 |
| polarC18 | 218.139 | 76.8935 | 13 | 0.92 |
| polarC18 | 218.139 | 60.5392 | 13 | 0.92 |
| polarC18 | 218.139 | 93.6142 | 13 | 0.92 |
| polarC18 | 218.139 | 86.454 | 13 | 0.92 |
| polarC18 | 218.139 | 191.4805 | 13 | 0.92 |
| polarC18 | 218.139 | 98.01 | 13 | 0.92 |
| polarC18 | 218.139 | 82.2286 | 13 | 0.92 |
| polarC18 | 218.139 | 105.7413 | 13 | 0.92 |
| polarC18 | 218.139 | 83.7485 | 12.9 | 0.92 |
| polarC18 | 218.139 | 127.6688 | 12.9 | 0.92 |

| | | | | |
|---|---|---|---|---|
| polarC18 | 218.139 | 98.3942 | 12.9 | 0.92 |
| polarC18 | 218.139 | 205.2218 | 12.9 | 0.92 |
| polarC18 | 218.139 | 54.6732 | 12.9 | 0.92 |
| polarC18 | 218.139 | 72.9122 | 12.9 | 0.92 |
| polarC18 | 218.139 | 169.6306 | 12.9 | 0.92 |
| polarC18 | 218.139 | 49.9479 | 12.9 | 0.92 |
| polarC18 | 218.139 | 103.9638 | 12.9 | 0.92 |
| polarC18 | 218.139 | 183.0345 | 12.9 | 0.92 |
| polarC18 | 218.139 | 104.5096 | 12.9 | 0.92 |
| polarC18 | 218.139 | 81.2434 | 12.9 | 0.92 |
| polarC18 | 218.139 | 87.9137 | 12.9 | 0.92 |
| polarC18 | 218.139 | 150.4799 | 12.9 | 0.92 |
| polarC18 | 218.139 | 93.2176 | 12.9 | 0.92 |
| polarC18 | 218.139 | 220.3724 | 12.9 | 0.92 |
| polarC18 | 218.139 | 134.1912 | 12.9 | 0.92 |
| polarC18 | 218.139 | 162.0912 | 12.9 | 0.92 |
| polarC18 | 218.139 | 79.3228 | 12.9 | 0.92 |
| polarC18 | 218.139 | 54.9831 | 12.9 | 0.92 |
| polarC18 | 218.139 | 94.5942 | 12.9 | 0.92 |
| polarC18 | 218.139 | 110.063 | 12.9 | 0.92 |
| polarC18 | 218.139 | 63.6755 | 12.9 | 0.92 |
| polarC18 | 218.139 | 79.5935 | 12.9 | 0.92 |
| polarC18 | 218.139 | 62.2698 | 12.9 | 0.92 |
| polarC18 | 218.139 | 84.5288 | 12.9 | 0.92 |
| polarC18 | 218.139 | 183.8435 | 12.9 | 0.92 |
| polarC18 | 218.139 | 138.2109 | 12.9 | 0.91 |
| polarC18 | 218.139 | 85.0112 | 12.9 | 0.91 |
| polarC18 | 218.139 | 49.3761 | 12.9 | 0.91 |
| polarC18 | 218.139 | 94.5346 | 12.9 | 0.91 |
| polarC18 | 218.139 | 206.7544 | 12.9 | 0.91 |
| polarC18 | 218.139 | 106.1665 | 12.9 | 0.91 |
| polarC18 | 218.139 | 68.0444 | 12.9 | 0.91 |
| polarC18 | 218.139 | 195.8132 | 12.9 | 0.91 |
| polarC18 | 218.139 | 78.6523 | 12.9 | 0.91 |
| polarC18 | 218.139 | 111.6707 | 12.9 | 0.91 |
| polarC18 | 218.139 | 137.8559 | 12.9 | 0.91 |
| polarC18 | 218.139 | 118.9871 | 12.9 | 0.91 |
| polarC18 | 218.139 | 216.1967 | 12.9 | 0.91 |
| polarC18 | 218.139 | 75.5874 | 12.9 | 0.91 |
| polarC18 | 218.139 | 120.5432 | 12.9 | 0.91 |
| polarC18 | 218.139 | 87.1036 | 12.9 | 0.91 |
| polarC18 | 218.139 | 122.0693 | 12.9 | 0.91 |

| | | | | |
|---|---|---|---|---|
| polarC18 | 218.139 | 55.4281 | 12.9 | 0.91 |
| polarC18 | 218.139 | 210.6888 | 12.9 | 0.91 |
| polarC18 | 218.139 | 48.8536 | 12.9 | 0.91 |
| polarC18 | 218.139 | 120.992 | 12.9 | 0.91 |
| polarC18 | 218.139 | 53.1557 | 12.9 | 0.91 |
| polarC18 | 218.139 | 82.2201 | 12.9 | 0.91 |
| polarC18 | 218.139 | 203.7677 | 12.9 | 0.91 |
| polarC18 | 218.139 | 93.1629 | 12.9 | 0.91 |
| polarC18 | 218.139 | 196.0458 | 12.9 | 0.91 |
| polarC18 | 218.139 | 59.9559 | 12.9 | 0.91 |
| polarC18 | 218.139 | 50.2352 | 12.9 | 0.91 |
| polarC18 | 218.139 | 90.8581 | 12.9 | 0.91 |
| polarC18 | 218.139 | 120.1735 | 12.9 | 0.91 |
| polarC18 | 218.139 | 96.6886 | 12.9 | 0.91 |
| polarC18 | 218.139 | 234.9781 | 12.9 | 0.91 |
| polarC18 | 218.139 | 216.8685 | 12.9 | 0.91 |
| polarC18 | 218.139 | 184.9744 | 12.9 | 0.91 |
| polarC18 | 218.139 | 174.5139 | 12.9 | 0.91 |
| polarC18 | 218.139 | 197.1852 | 12.9 | 0.91 |
| polarC18 | 218.139 | 149.5701 | 12.9 | 0.91 |
| polarC18 | 218.139 | 54.3163 | 12.9 | 0.91 |
| polarC18 | 218.139 | 50.264 | 12.9 | 0.91 |
| polarC18 | 218.139 | 130.7881 | 12.9 | 0.91 |
| polarC18 | 218.139 | 131.894 | 12.9 | 0.91 |
| polarC18 | 218.139 | 95.8058 | 12.9 | 0.91 |
| polarC18 | 218.139 | 48.8345 | 12.9 | 0.91 |
| polarC18 | 218.139 | 71.1831 | 12.9 | 0.91 |
| polarC18 | 218.139 | 61.9015 | 12.8 | 0.91 |
| polarC18 | 218.139 | 64.4647 | 12.8 | 0.91 |
| polarC18 | 218.139 | 97.4615 | 12.8 | 0.91 |
| polarC18 | 218.139 | 62.5729 | 12.8 | 0.91 |
| polarC18 | 218.139 | 117.7324 | 12.8 | 0.91 |
| polarC18 | 218.139 | 109.0935 | 12.8 | 0.91 |
| polarC18 | 218.139 | 117.576 | 12.8 | 0.91 |
| polarC18 | 218.139 | 54.1111 | 12.8 | 0.91 |
| polarC18 | 218.139 | 78.6692 | 12.8 | 0.91 |
| polarC18 | 218.139 | 92.5621 | 12.8 | 0.91 |
| polarC18 | 218.139 | 108.7216 | 12.8 | 0.91 |
| polarC18 | 218.139 | 141.8409 | 12.8 | 0.91 |
| polarC18 | 218.139 | 60.4138 | 12.8 | 0.91 |
| polarC18 | 218.139 | 119.8075 | 12.8 | 0.91 |
| polarC18 | 218.139 | 96.9297 | 12.8 | 0.91 |

| | | | | |
|---|---|---|---|---|
| polarC18 | 218.139 | 79.2126 | 12.8 | 0.91 |
| polarC18 | 218.139 | 217.5957 | 12.8 | 0.91 |
| polarC18 | 218.139 | 66.2405 | 12.8 | 0.91 |
| polarC18 | 218.139 | 51.5475 | 12.8 | 0.91 |
| polarC18 | 218.139 | 106.6864 | 12.8 | 0.91 |
| polarC18 | 218.139 | 175.7252 | 12.8 | 0.91 |
| polarC18 | 218.139 | 136.8068 | 12.8 | 0.91 |
| polarC18 | 218.139 | 108.5366 | 12.8 | 0.91 |
| polarC18 | 218.139 | 60.5351 | 12.8 | 0.91 |
| polarC18 | 218.139 | 88.8954 | 12.8 | 0.91 |
| polarC18 | 218.139 | 142.3208 | 12.8 | 0.91 |
| polarC18 | 218.139 | 69.9451 | 12.8 | 0.91 |
| polarC18 | 218.139 | 223.6098 | 12.8 | 0.91 |
| polarC18 | 218.139 | 178.6111 | 12.8 | 0.91 |
| polarC18 | 218.139 | 63.0509 | 12.8 | 0.91 |
| polarC18 | 218.139 | 76.0432 | 12.8 | 0.91 |
| polarC18 | 218.139 | 86.0264 | 12.8 | 0.91 |
| polarC18 | 218.139 | 101.7116 | 12.8 | 0.91 |
| polarC18 | 218.139 | 170.2573 | 12.8 | 0.91 |
| polarC18 | 218.139 | 223.8832 | 12.8 | 0.91 |
| polarC18 | 218.139 | 160.8847 | 12.8 | 0.91 |
| polarC18 | 218.139 | 55.8014 | 12.8 | 0.91 |
| polarC18 | 218.139 | 116.6363 | 12.8 | 0.91 |
| polarC18 | 218.139 | 148.0033 | 12.8 | 0.91 |
| polarC18 | 218.139 | 56.8555 | 12.8 | 0.91 |
| polarC18 | 218.139 | 51.9749 | 12.8 | 0.91 |
| polarC18 | 218.139 | 207.3586 | 12.8 | 0.91 |
| polarC18 | 218.139 | 73.0371 | 12.8 | 0.91 |
| polarC18 | 218.139 | 59.1553 | 12.8 | 0.91 |
| polarC18 | 218.139 | 181.1735 | 12.8 | 0.91 |
| polarC18 | 218.139 | 90.2796 | 12.8 | 0.91 |
| polarC18 | 218.139 | 115.2353 | 12.8 | 0.91 |
| polarC18 | 218.139 | 145.7871 | 12.8 | 0.91 |
| polarC18 | 218.139 | 142.237 | 12.8 | 0.91 |
| polarC18 | 218.139 | 124.1926 | 12.8 | 0.91 |
| polarC18 | 218.139 | 143.1897 | 12.8 | 0.91 |
| polarC18 | 218.139 | 173.349 | 12.8 | 0.91 |
| polarC18 | 218.139 | 49.8146 | 12.8 | 0.91 |
| polarC18 | 218.139 | 166.848 | 12.8 | 0.9 |
| polarC18 | 218.139 | 67.4966 | 12.8 | 0.9 |
| polarC18 | 218.139 | 218.5943 | 12.8 | 0.9 |
| polarC18 | 218.139 | 146.8306 | 12.8 | 0.9 |

| | | | | |
|---|---|---|---|---|
| polarC18 | 218.139 | 136.8763 | 12.7 | 0.9 |
| polarC18 | 218.139 | 135.7414 | 12.7 | 0.9 |
| polarC18 | 218.139 | 62.349 | 12.7 | 0.9 |
| polarC18 | 218.139 | 136.0818 | 12.7 | 0.9 |
| polarC18 | 218.139 | 57.0325 | 12.7 | 0.9 |
| polarC18 | 218.139 | 63.562 | 12.7 | 0.9 |
| polarC18 | 218.139 | 91.8111 | 12.7 | 0.9 |
| polarC18 | 218.139 | 50.879 | 12.7 | 0.9 |
| polarC18 | 218.139 | 70.4531 | 12.7 | 0.9 |
| polarC18 | 218.139 | 106.1556 | 12.7 | 0.9 |
| polarC18 | 218.139 | 71.9645 | 12.7 | 0.9 |
| polarC18 | 218.139 | 149.6562 | 12.7 | 0.9 |
| polarC18 | 218.139 | 103.8648 | 12.7 | 0.9 |
| polarC18 | 218.139 | 60.4373 | 12.7 | 0.9 |
| polarC18 | 218.139 | 65.0638 | 12.7 | 0.9 |
| polarC18 | 218.139 | 54.3352 | 12.7 | 0.9 |
| polarC18 | 218.139 | 107.6418 | 12.7 | 0.9 |
| polarC18 | 218.139 | 125.0461 | 12.7 | 0.9 |
| polarC18 | 218.139 | 117.3651 | 12.7 | 0.9 |
| polarC18 | 218.139 | 50.6469 | 12.7 | 0.9 |
| polarC18 | 218.139 | 241.3485 | 12.7 | 0.9 |
| polarC18 | 218.139 | 174.7739 | 12.7 | 0.9 |
| polarC18 | 218.139 | 49.4839 | 12.7 | 0.9 |
| polarC18 | 218.139 | 138.7499 | 12.7 | 0.9 |
| polarC18 | 218.139 | 76.4891 | 12.7 | 0.9 |
| polarC18 | 218.139 | 59.8677 | 12.7 | 0.9 |
| polarC18 | 218.139 | 162.6284 | 12.7 | 0.9 |
| polarC18 | 218.139 | 65.8497 | 12.7 | 0.9 |
| polarC18 | 218.139 | 53.541 | 12.7 | 0.9 |
| polarC18 | 218.139 | 68.1471 | 12.7 | 0.9 |
| polarC18 | 218.139 | 160.0061 | 12.7 | 0.9 |
| polarC18 | 218.139 | 118.6564 | 12.7 | 0.9 |
| polarC18 | 218.139 | 99.6969 | 12.7 | 0.9 |
| polarC18 | 218.139 | 66.2012 | 12.7 | 0.9 |
| polarC18 | 218.139 | 60.5061 | 12.7 | 0.9 |
| polarC18 | 218.139 | 137.6851 | 12.7 | 0.9 |
| polarC18 | 218.139 | 157.1854 | 12.7 | 0.9 |
| polarC18 | 218.139 | 87.6689 | 12.7 | 0.9 |
| polarC18 | 218.139 | 60.3555 | 12.7 | 0.9 |
| polarC18 | 218.139 | 62.8538 | 12.7 | 0.9 |
| polarC18 | 218.139 | 55.8108 | 12.7 | 0.9 |
| polarC18 | 218.139 | 76.7034 | 12.7 | 0.9 |

| | | | | |
|---|---|---|---|---|
| polarC18 | 218.139 | 62.6005 | 12.7 | 0.9 |
| polarC18 | 218.139 | 97.4969 | 12.7 | 0.9 |
| polarC18 | 218.139 | 68.8197 | 12.7 | 0.9 |
| polarC18 | 218.139 | 222.6637 | 12.7 | 0.9 |
| polarC18 | 218.139 | 70.0414 | 12.7 | 0.9 |
| polarC18 | 218.139 | 71.0283 | 12.7 | 0.9 |
| polarC18 | 218.139 | 120.2993 | 12.7 | 0.9 |
| polarC18 | 218.139 | 126.3898 | 12.7 | 0.9 |
| polarC18 | 218.139 | 52.3293 | 12.7 | 0.9 |
| polarC18 | 218.139 | 129.409 | 12.7 | 0.9 |
| polarC18 | 218.139 | 53.5681 | 12.7 | 0.9 |
| polarC18 | 218.139 | 117.9186 | 12.7 | 0.9 |
| polarC18 | 218.139 | 225.6549 | 12.7 | 0.9 |
| polarC18 | 218.139 | 174.6639 | 12.7 | 0.9 |
| polarC18 | 218.139 | 64.219 | 12.7 | 0.9 |
| polarC18 | 218.139 | 116.8755 | 12.7 | 0.9 |
| polarC18 | 218.139 | 50.1975 | 12.7 | 0.9 |
| polarC18 | 218.139 | 58.9037 | 12.7 | 0.9 |
| polarC18 | 218.139 | 71.3084 | 12.7 | 0.9 |
| polarC18 | 218.139 | 53.3802 | 12.7 | 0.9 |
| polarC18 | 218.139 | 85.8689 | 12.7 | 0.9 |
| polarC18 | 218.139 | 58.0176 | 12.7 | 0.9 |
| polarC18 | 218.139 | 105.3908 | 12.7 | 0.9 |
| polarC18 | 218.139 | 186.0832 | 12.7 | 0.9 |
| polarC18 | 218.139 | 63.6283 | 12.7 | 0.9 |
| polarC18 | 218.139 | 53.6436 | 12.6 | 0.9 |
| polarC18 | 218.139 | 53.4333 | 12.6 | 0.9 |
| polarC18 | 218.139 | 99.6751 | 12.6 | 0.9 |
| polarC18 | 218.139 | 56.4323 | 12.6 | 0.9 |
| polarC18 | 218.139 | 199.737 | 12.6 | 0.9 |
| polarC18 | 218.139 | 134.0511 | 12.6 | 0.9 |
| polarC18 | 218.139 | 66.3779 | 12.6 | 0.9 |
| polarC18 | 218.139 | 62.0379 | 12.6 | 0.9 |
| polarC18 | 218.139 | 70.252 | 12.6 | 0.9 |
| polarC18 | 218.139 | 94.4775 | 12.6 | 0.9 |
| polarC18 | 218.139 | 51.5583 | 12.6 | 0.9 |
| polarC18 | 218.139 | 65.4548 | 12.6 | 0.9 |
| polarC18 | 218.139 | 66.5944 | 12.6 | 0.9 |
| polarC18 | 218.139 | 238.3925 | 12.6 | 0.9 |
| polarC18 | 218.139 | 67.8326 | 12.6 | 0.9 |
| polarC18 | 218.139 | 165.4715 | 12.6 | 0.9 |
| polarC18 | 218.139 | 63.3048 | 12.6 | 0.9 |

| | | | | |
|---|---|---|---|---|
| polarC18 | 218.139 | 70.937 | 12.6 | 0.9 |
| polarC18 | 218.139 | 125.8312 | 12.6 | 0.9 |
| polarC18 | 218.139 | 120.4998 | 12.6 | 0.9 |
| polarC18 | 218.139 | 71.8345 | 12.6 | 0.9 |
| polarC18 | 218.139 | 67.9226 | 12.6 | 0.9 |
| polarC18 | 218.139 | 70.1016 | 12.6 | 0.9 |
| polarC18 | 218.139 | 140.6156 | 12.6 | 0.9 |
| polarC18 | 218.139 | 112.6981 | 12.6 | 0.9 |
| polarC18 | 218.139 | 69.8615 | 12.6 | 0.89 |
| polarC18 | 218.139 | 67.1496 | 12.6 | 0.89 |
| polarC18 | 218.139 | 216.1124 | 12.6 | 0.89 |
| polarC18 | 218.139 | 134.4467 | 12.6 | 0.89 |
| polarC18 | 218.139 | 101.9172 | 12.6 | 0.89 |
| polarC18 | 218.139 | 63.5469 | 12.6 | 0.89 |
| polarC18 | 218.139 | 48.6879 | 12.6 | 0.89 |
| polarC18 | 218.139 | 181.7623 | 12.6 | 0.89 |
| polarC18 | 218.139 | 75.3516 | 12.6 | 0.89 |
| polarC18 | 218.139 | 125.4588 | 12.6 | 0.89 |
| polarC18 | 218.139 | 48.6112 | 12.6 | 0.89 |
| polarC18 | 218.139 | 241.567 | 12.6 | 0.89 |
| polarC18 | 218.139 | 146.5706 | 12.6 | 0.89 |
| polarC18 | 218.139 | 79.6788 | 12.6 | 0.89 |
| polarC18 | 218.139 | 206.9607 | 12.6 | 0.89 |
| polarC18 | 218.139 | 88.8525 | 12.6 | 0.89 |
| polarC18 | 218.139 | 75.6572 | 12.6 | 0.89 |
| polarC18 | 218.139 | 80.2494 | 12.6 | 0.89 |
| polarC18 | 218.139 | 72.2922 | 12.6 | 0.89 |
| polarC18 | 218.139 | 50.5701 | 12.6 | 0.89 |
| polarC18 | 218.139 | 80.4163 | 12.6 | 0.89 |
| polarC18 | 218.139 | 114.8351 | 12.6 | 0.89 |
| polarC18 | 218.139 | 168.6547 | 12.6 | 0.89 |
| polarC18 | 218.139 | 59.841 | 12.6 | 0.89 |
| polarC18 | 218.139 | 67.8656 | 12.6 | 0.89 |
| polarC18 | 218.139 | 146.1641 | 12.6 | 0.89 |
| polarC18 | 218.139 | 133.2461 | 12.6 | 0.89 |
| polarC18 | 218.139 | 117.4529 | 12.6 | 0.89 |
| polarC18 | 218.139 | 121.4091 | 12.6 | 0.89 |
| polarC18 | 218.139 | 75.5966 | 12.6 | 0.89 |
| polarC18 | 218.139 | 60.7208 | 12.6 | 0.89 |
| polarC18 | 218.139 | 77.7712 | 12.6 | 0.89 |
| polarC18 | 218.139 | 117.3332 | 12.6 | 0.89 |
| polarC18 | 218.139 | 240.7963 | 12.6 | 0.89 |

| | | | | |
|---|---|---|---|---|
| polarC18 | 218.139 | 51.4664 | 12.6 | 0.89 |
| polarC18 | 218.139 | 152.5887 | 12.6 | 0.89 |
| polarC18 | 218.139 | 83.0355 | 12.6 | 0.89 |
| polarC18 | 218.139 | 58.738 | 12.6 | 0.89 |
| polarC18 | 218.139 | 56.7066 | 12.6 | 0.89 |
| polarC18 | 218.139 | 55.5397 | 12.6 | 0.89 |
| polarC18 | 218.139 | 59.4716 | 12.5 | 0.89 |
| polarC18 | 218.139 | 95.8212 | 12.5 | 0.89 |
| polarC18 | 218.139 | 161.1801 | 12.5 | 0.89 |
| polarC18 | 218.139 | 96.9613 | 12.5 | 0.89 |
| polarC18 | 218.139 | 135.7987 | 12.5 | 0.89 |
| polarC18 | 218.139 | 65.5586 | 12.5 | 0.89 |
| polarC18 | 218.139 | 92.9432 | 12.5 | 0.89 |
| polarC18 | 218.139 | 171.528 | 12.5 | 0.89 |
| polarC18 | 218.139 | 111.2998 | 12.5 | 0.89 |
| polarC18 | 218.139 | 79.6216 | 12.5 | 0.89 |
| polarC18 | 218.139 | 62.9069 | 12.5 | 0.89 |
| polarC18 | 218.139 | 55.1111 | 12.5 | 0.89 |
| polarC18 | 218.139 | 67.1391 | 12.5 | 0.89 |
| polarC18 | 218.139 | 145.6582 | 12.5 | 0.89 |
| polarC18 | 218.139 | 57.1117 | 12.5 | 0.89 |
| polarC18 | 218.139 | 114.4726 | 12.5 | 0.89 |
| polarC18 | 218.139 | 103.8766 | 12.5 | 0.89 |
| polarC18 | 218.139 | 108.4957 | 12.5 | 0.89 |
| polarC18 | 218.139 | 135.6686 | 12.5 | 0.89 |
| polarC18 | 218.139 | 231.8239 | 12.5 | 0.89 |
| polarC18 | 218.139 | 169.0846 | 12.5 | 0.89 |
| polarC18 | 218.139 | 83.4141 | 12.5 | 0.89 |
| polarC18 | 218.139 | 227.0255 | 12.5 | 0.89 |
| polarC18 | 218.139 | 72.7554 | 12.5 | 0.89 |
| polarC18 | 218.139 | 150.0111 | 12.5 | 0.89 |
| polarC18 | 218.139 | 66.3494 | 12.5 | 0.89 |
| polarC18 | 218.139 | 101.6256 | 12.5 | 0.89 |
| polarC18 | 218.139 | 54.0941 | 12.5 | 0.89 |
| polarC18 | 218.139 | 75.7858 | 12.5 | 0.89 |
| polarC18 | 218.139 | 193.7349 | 12.5 | 0.89 |
| polarC18 | 218.139 | 201.1908 | 12.5 | 0.89 |
| polarC18 | 218.139 | 57.7291 | 12.5 | 0.89 |
| polarC18 | 218.139 | 101.3085 | 12.5 | 0.89 |
| polarC18 | 218.139 | 100.4399 | 12.5 | 0.89 |
| polarC18 | 218.139 | 59.3441 | 12.5 | 0.89 |
| polarC18 | 218.139 | 54.595 | 12.5 | 0.89 |

| | | | | |
|---|---|---|---|---|
| polarC18 | 218.139 | 59.1856 | 12.5 | 0.89 |
| polarC18 | 218.139 | 81.6161 | 12.5 | 0.89 |
| polarC18 | 218.139 | 51.7966 | 12.5 | 0.89 |
| polarC18 | 218.139 | 149.2589 | 12.5 | 0.89 |
| polarC18 | 218.139 | 59.2509 | 12.5 | 0.89 |
| polarC18 | 218.139 | 152.8268 | 12.5 | 0.89 |
| polarC18 | 218.139 | 180.7197 | 12.5 | 0.89 |
| polarC18 | 218.139 | 53.2367 | 12.5 | 0.88 |
| polarC18 | 218.139 | 53.56 | 12.5 | 0.88 |
| polarC18 | 218.139 | 62.2648 | 12.5 | 0.88 |
| polarC18 | 218.139 | 173.5144 | 12.5 | 0.88 |
| polarC18 | 218.139 | 156.9576 | 12.5 | 0.88 |
| polarC18 | 218.139 | 97.2119 | 12.5 | 0.88 |
| polarC18 | 218.139 | 179.3704 | 12.5 | 0.88 |
| polarC18 | 218.139 | 149.1278 | 12.5 | 0.88 |
| polarC18 | 218.139 | 223.5281 | 12.5 | 0.88 |
| polarC18 | 218.139 | 91.6882 | 12.4 | 0.88 |
| polarC18 | 218.139 | 111.885 | 12.4 | 0.88 |
| polarC18 | 218.139 | 123.3358 | 12.4 | 0.88 |
| polarC18 | 218.139 | 100.1753 | 12.4 | 0.88 |
| polarC18 | 218.139 | 54.6355 | 12.4 | 0.88 |
| polarC18 | 218.139 | 67.7754 | 12.4 | 0.88 |
| polarC18 | 218.139 | 60.3922 | 12.4 | 0.88 |
| polarC18 | 218.139 | 98.325 | 12.4 | 0.88 |
| polarC18 | 218.139 | 61.6897 | 12.4 | 0.88 |
| polarC18 | 218.139 | 101.4091 | 12.4 | 0.88 |
| polarC18 | 218.139 | 110.2276 | 12.4 | 0.88 |
| polarC18 | 218.139 | 190.1583 | 12.4 | 0.88 |
| polarC18 | 218.139 | 49.1312 | 12.4 | 0.88 |
| polarC18 | 218.139 | 52.8869 | 12.4 | 0.88 |
| polarC18 | 218.139 | 68.0117 | 12.4 | 0.88 |
| polarC18 | 218.139 | 104.9007 | 12.4 | 0.88 |
| polarC18 | 218.139 | 58.8631 | 12.4 | 0.88 |
| polarC18 | 218.139 | 242.8613 | 12.4 | 0.88 |
| polarC18 | 218.139 | 166.7707 | 12.4 | 0.88 |
| polarC18 | 218.139 | 96.0109 | 12.4 | 0.88 |
| polarC18 | 218.139 | 48.917 | 12.4 | 0.88 |
| polarC18 | 218.139 | 107.8628 | 12.4 | 0.88 |
| polarC18 | 218.139 | 140.2043 | 12.4 | 0.88 |
| polarC18 | 218.139 | 102.6614 | 12.4 | 0.88 |
| polarC18 | 218.139 | 221.0925 | 12.4 | 0.88 |
| polarC18 | 218.139 | 69.6056 | 12.4 | 0.88 |

| | | | | |
|---|---|---|---|---|
| polarC18 | 218.139 | 136.9124 | 12.4 | 0.88 |
| polarC18 | 218.139 | 51.0702 | 12.4 | 0.88 |
| polarC18 | 218.139 | 78.9563 | 12.4 | 0.88 |
| polarC18 | 218.139 | 119.8928 | 12.4 | 0.88 |
| polarC18 | 218.139 | 60.7308 | 12.4 | 0.88 |
| polarC18 | 218.139 | 114.4633 | 12.4 | 0.88 |
| polarC18 | 218.139 | 73.0149 | 12.4 | 0.88 |
| polarC18 | 218.139 | 112.6146 | 12.4 | 0.88 |
| polarC18 | 218.139 | 194.3258 | 12.4 | 0.88 |
| polarC18 | 218.139 | 100.7929 | 12.4 | 0.88 |
| polarC18 | 218.139 | 102.9106 | 12.4 | 0.88 |
| polarC18 | 218.139 | 99.3235 | 12.4 | 0.88 |
| polarC18 | 218.139 | 188.1105 | 12.4 | 0.88 |
| polarC18 | 218.139 | 231.0916 | 12.4 | 0.88 |
| polarC18 | 218.139 | 54.1178 | 12.4 | 0.88 |
| polarC18 | 218.139 | 94.5258 | 12.4 | 0.88 |
| polarC18 | 218.139 | 176.8678 | 12.4 | 0.88 |
| polarC18 | 218.139 | 143.5728 | 12.4 | 0.88 |
| polarC18 | 218.139 | 94.211 | 12.4 | 0.88 |
| polarC18 | 218.139 | 51.9357 | 12.4 | 0.88 |
| polarC18 | 218.139 | 81.9388 | 12.4 | 0.88 |
| polarC18 | 218.139 | 163.3656 | 12.4 | 0.88 |
| polarC18 | 218.139 | 70.6174 | 12.4 | 0.88 |
| polarC18 | 218.139 | 148.4331 | 12.4 | 0.88 |
| polarC18 | 218.139 | 55.321 | 12.4 | 0.88 |
| polarC18 | 218.139 | 81.9743 | 12.4 | 0.88 |
| polarC18 | 218.139 | 84.3063 | 12.4 | 0.88 |
| polarC18 | 218.139 | 121.7715 | 12.4 | 0.88 |
| polarC18 | 218.139 | 64.1833 | 12.4 | 0.88 |
| polarC18 | 218.139 | 92.538 | 12.4 | 0.88 |
| polarC18 | 218.139 | 208.0001 | 12.4 | 0.88 |
| polarC18 | 218.139 | 124.5264 | 12.3 | 0.88 |
| polarC18 | 218.139 | 60.0146 | 12.3 | 0.88 |
| polarC18 | 218.139 | 191.7464 | 12.3 | 0.88 |
| polarC18 | 218.139 | 57.2023 | 12.3 | 0.88 |
| polarC18 | 218.139 | 194.5405 | 12.3 | 0.88 |
| polarC18 | 218.139 | 62.6966 | 12.3 | 0.88 |
| polarC18 | 218.139 | 222.1932 | 12.3 | 0.88 |
| polarC18 | 218.139 | 61.3388 | 12.3 | 0.88 |
| polarC18 | 218.139 | 64.6252 | 12.3 | 0.87 |
| polarC18 | 218.139 | 90.2175 | 12.3 | 0.87 |
| polarC18 | 218.139 | 242.4213 | 12.3 | 0.87 |

| | | | | |
|---|---|---|---|---|
| polarC18 | 218.139 | 77.6459 | 12.3 | 0.87 |
| polarC18 | 218.139 | 169.6951 | 12.3 | 0.87 |
| polarC18 | 218.139 | 161.5801 | 12.3 | 0.87 |
| polarC18 | 218.139 | 75.5926 | 12.3 | 0.87 |
| polarC18 | 218.139 | 100.8088 | 12.3 | 0.87 |
| polarC18 | 218.139 | 121.4431 | 12.3 | 0.87 |
| polarC18 | 218.139 | 107.404 | 12.3 | 0.87 |
| polarC18 | 218.139 | 69.2043 | 12.3 | 0.87 |
| polarC18 | 218.139 | 83.5717 | 12.3 | 0.87 |
| polarC18 | 218.139 | 91.7958 | 12.3 | 0.87 |
| polarC18 | 218.139 | 73.6407 | 12.3 | 0.87 |
| polarC18 | 218.139 | 88.5432 | 12.3 | 0.87 |
| polarC18 | 218.139 | 53.353 | 12.3 | 0.87 |
| polarC18 | 218.139 | 97.5917 | 12.3 | 0.87 |
| polarC18 | 218.139 | 199.8558 | 12.3 | 0.87 |
| polarC18 | 218.139 | 103.4126 | 12.3 | 0.87 |
| polarC18 | 218.139 | 81.813 | 12.3 | 0.87 |
| polarC18 | 218.139 | 99.9596 | 12.3 | 0.87 |
| polarC18 | 218.139 | 91.1953 | 12.3 | 0.87 |
| polarC18 | 218.139 | 60.1579 | 12.3 | 0.87 |
| polarC18 | 218.139 | 57.4174 | 12.3 | 0.87 |
| polarC18 | 218.139 | 161.6949 | 12.3 | 0.87 |
| polarC18 | 218.139 | 89.6373 | 12.3 | 0.87 |
| polarC18 | 218.139 | 61.7993 | 12.3 | 0.87 |
| polarC18 | 218.139 | 52.1032 | 12.3 | 0.87 |
| polarC18 | 218.139 | 88.0652 | 12.3 | 0.87 |
| polarC18 | 218.139 | 57.9768 | 12.3 | 0.87 |
| polarC18 | 218.139 | 128.2989 | 12.3 | 0.87 |
| polarC18 | 218.139 | 83.619 | 12.3 | 0.87 |
| polarC18 | 218.139 | 66.1634 | 12.3 | 0.87 |
| polarC18 | 218.139 | 120.5989 | 12.3 | 0.87 |
| polarC18 | 218.139 | 167.5689 | 12.3 | 0.87 |
| polarC18 | 218.139 | 97.4361 | 12.3 | 0.87 |
| polarC18 | 218.139 | 141.9814 | 12.3 | 0.87 |
| polarC18 | 218.139 | 150.309 | 12.3 | 0.87 |
| polarC18 | 218.139 | 134.4868 | 12.3 | 0.87 |
| polarC18 | 218.139 | 64.5558 | 12.3 | 0.87 |
| polarC18 | 218.139 | 49.2148 | 12.3 | 0.87 |
| polarC18 | 218.139 | 93.7898 | 12.3 | 0.87 |
| polarC18 | 218.139 | 55.7393 | 12.3 | 0.87 |
| polarC18 | 218.139 | 52.9221 | 12.3 | 0.87 |
| polarC18 | 218.139 | 125.697 | 12.3 | 0.87 |

| | | | | |
|---|---|---|---|---|
| polarC18 | 218.139 | 220.8415 | 12.3 | 0.87 |
| polarC18 | 218.139 | 82.2358 | 12.3 | 0.87 |
| polarC18 | 218.139 | 66.7713 | 12.3 | 0.87 |
| polarC18 | 218.139 | 52.6464 | 12.3 | 0.87 |
| polarC18 | 218.139 | 55.4936 | 12.3 | 0.87 |
| polarC18 | 218.139 | 52.0501 | 12.3 | 0.87 |
| polarC18 | 218.139 | 76.4152 | 12.3 | 0.87 |
| polarC18 | 218.139 | 120.1904 | 12.3 | 0.87 |
| polarC18 | 218.139 | 154.8723 | 12.3 | 0.87 |
| polarC18 | 218.139 | 160.2666 | 12.3 | 0.87 |
| polarC18 | 218.139 | 68.5127 | 12.3 | 0.87 |
| polarC18 | 218.139 | 62.9386 | 12.2 | 0.87 |
| polarC18 | 218.139 | 50.7104 | 12.2 | 0.87 |
| polarC18 | 218.139 | 49.303 | 12.2 | 0.87 |
| polarC18 | 218.139 | 220.1511 | 12.2 | 0.87 |
| polarC18 | 218.139 | 81.3508 | 12.2 | 0.87 |
| polarC18 | 218.139 | 93.8725 | 12.2 | 0.87 |
| polarC18 | 218.139 | 62.0003 | 12.2 | 0.87 |
| polarC18 | 218.139 | 70.6615 | 12.2 | 0.87 |
| polarC18 | 218.139 | 178.7685 | 12.2 | 0.87 |
| polarC18 | 218.139 | 89.6977 | 12.2 | 0.87 |
| polarC18 | 218.139 | 82.8168 | 12.2 | 0.87 |
| polarC18 | 218.139 | 100.8923 | 12.2 | 0.87 |
| polarC18 | 218.139 | 79.537 | 12.2 | 0.87 |
| polarC18 | 218.139 | 52.9893 | 12.2 | 0.87 |
| polarC18 | 218.139 | 161.2824 | 12.2 | 0.87 |
| polarC18 | 218.139 | 73.4605 | 12.2 | 0.87 |
| polarC18 | 218.139 | 179.9419 | 12.2 | 0.87 |
| polarC18 | 218.139 | 61.4469 | 12.2 | 0.87 |
| polarC18 | 218.139 | 120.2017 | 12.2 | 0.87 |
| polarC18 | 218.139 | 63.0974 | 12.2 | 0.87 |
| polarC18 | 218.139 | 237.2608 | 12.2 | 0.87 |
| polarC18 | 218.139 | 157.6673 | 12.2 | 0.87 |
| polarC18 | 218.139 | 99.2562 | 12.2 | 0.87 |
| polarC18 | 218.139 | 152.9845 | 12.2 | 0.87 |
| polarC18 | 218.139 | 203.0528 | 12.2 | 0.87 |
| polarC18 | 218.139 | 58.2527 | 12.2 | 0.87 |
| polarC18 | 218.139 | 159.2007 | 12.2 | 0.87 |
| polarC18 | 218.139 | 199.0785 | 12.2 | 0.87 |
| polarC18 | 218.139 | 48.6036 | 12.2 | 0.87 |
| polarC18 | 218.139 | 65.4729 | 12.2 | 0.87 |
| polarC18 | 218.139 | 51.7214 | 12.2 | 0.87 |

| | | | | |
|---|---|---|---|---|
| polarC18 | 218.139 | 109.0111 | 12.2 | 0.87 |
| polarC18 | 218.139 | 211.8407 | 12.2 | 0.87 |
| polarC18 | 218.139 | 90.895 | 12.2 | 0.87 |
| polarC18 | 218.139 | 229.1566 | 12.2 | 0.86 |
| polarC18 | 218.139 | 137.2051 | 12.2 | 0.86 |
| polarC18 | 218.139 | 100.2579 | 12.2 | 0.86 |
| polarC18 | 218.139 | 131.593 | 12.2 | 0.86 |
| polarC18 | 218.139 | 152.4701 | 12.2 | 0.86 |
| polarC18 | 218.139 | 196.957 | 12.2 | 0.86 |
| polarC18 | 218.139 | 76.2116 | 12.2 | 0.86 |
| polarC18 | 218.139 | 100.875 | 12.2 | 0.86 |
| polarC18 | 218.139 | 74.3436 | 12.2 | 0.86 |
| polarC18 | 218.139 | 80.33 | 12.2 | 0.86 |
| polarC18 | 218.139 | 150.6648 | 12.2 | 0.86 |
| polarC18 | 218.139 | 51.9886 | 12.2 | 0.86 |
| polarC18 | 218.139 | 61.0121 | 12.2 | 0.86 |
| polarC18 | 218.139 | 137.0908 | 12.2 | 0.86 |
| polarC18 | 218.139 | 53.0348 | 12.2 | 0.86 |
| polarC18 | 218.139 | 101.0958 | 12.2 | 0.86 |
| polarC18 | 218.139 | 138.0804 | 12.2 | 0.86 |
| polarC18 | 218.139 | 107.3405 | 12.2 | 0.86 |
| polarC18 | 218.139 | 210.2911 | 12.2 | 0.86 |
| polarC18 | 218.139 | 52.4131 | 12.1 | 0.86 |
| polarC18 | 218.139 | 108.3234 | 12.1 | 0.86 |
| polarC18 | 218.139 | 83.2162 | 12.1 | 0.86 |
| polarC18 | 218.139 | 52.9945 | 12.1 | 0.86 |
| polarC18 | 218.139 | 49.9603 | 12.1 | 0.86 |
| polarC18 | 218.139 | 61.916 | 12.1 | 0.86 |
| polarC18 | 218.139 | 114.2521 | 12.1 | 0.86 |
| polarC18 | 218.139 | 57.946 | 12.1 | 0.86 |
| polarC18 | 218.139 | 57.2865 | 12.1 | 0.86 |
| polarC18 | 218.139 | 69.1877 | 12.1 | 0.86 |
| polarC18 | 218.139 | 167.6855 | 12.1 | 0.86 |
| polarC18 | 218.139 | 49.1417 | 12.1 | 0.86 |
| polarC18 | 218.139 | 73.0443 | 12.1 | 0.86 |
| polarC18 | 218.139 | 97.6481 | 12.1 | 0.86 |
| polarC18 | 218.139 | 164.6377 | 12.1 | 0.86 |
| polarC18 | 218.139 | 51.5222 | 12.1 | 0.86 |
| polarC18 | 218.139 | 57.5297 | 12.1 | 0.86 |
| polarC18 | 218.139 | 118.1269 | 12.1 | 0.86 |
| polarC18 | 218.139 | 183.1192 | 12.1 | 0.86 |
| polarC18 | 218.139 | 111.5784 | 12.1 | 0.86 |

| | | | | |
|---|---|---|---|---|
| polarC18 | 218.139 | 53.6167 | 12.1 | 0.86 |
| polarC18 | 218.139 | 209.6127 | 12.1 | 0.86 |
| polarC18 | 218.139 | 52.4078 | 12.1 | 0.86 |
| polarC18 | 218.139 | 152.6681 | 12.1 | 0.86 |
| polarC18 | 218.139 | 63.1137 | 12.1 | 0.86 |
| polarC18 | 218.139 | 148.7553 | 12.1 | 0.86 |
| polarC18 | 218.139 | 106.8628 | 12.1 | 0.86 |
| polarC18 | 218.139 | 145.006 | 12.1 | 0.86 |
| polarC18 | 218.139 | 55.1542 | 12.1 | 0.86 |
| polarC18 | 218.139 | 62.0345 | 12.1 | 0.86 |
| polarC18 | 218.139 | 104.1443 | 12.1 | 0.86 |
| polarC18 | 218.139 | 170.3248 | 12.1 | 0.86 |
| polarC18 | 218.139 | 99.401 | 12.1 | 0.86 |
| polarC18 | 218.139 | 99.5698 | 12.1 | 0.86 |
| polarC18 | 218.139 | 55.0914 | 12.1 | 0.86 |
| polarC18 | 218.139 | 110.0041 | 12.1 | 0.86 |
| polarC18 | 218.139 | 82.1889 | 12.1 | 0.86 |
| polarC18 | 218.139 | 84.3958 | 12.1 | 0.86 |
| polarC18 | 218.139 | 77.0162 | 12.1 | 0.86 |
| polarC18 | 218.139 | 75.4177 | 12.1 | 0.86 |
| polarC18 | 218.139 | 72.9275 | 12.1 | 0.86 |
| polarC18 | 218.139 | 54.6891 | 12.1 | 0.86 |
| polarC18 | 218.139 | 58.3079 | 12.1 | 0.86 |
| polarC18 | 218.139 | 99.2777 | 12.1 | 0.86 |
| polarC18 | 218.139 | 57.2178 | 12.1 | 0.85 |
| polarC18 | 218.139 | 85.5468 | 12 | 0.85 |
| polarC18 | 218.139 | 147.0532 | 12 | 0.85 |
| polarC18 | 218.139 | 94.8582 | 12 | 0.85 |
| polarC18 | 218.139 | 74.2635 | 12 | 0.85 |
| polarC18 | 218.139 | 88.2671 | 12 | 0.85 |
| polarC18 | 218.139 | 120.6536 | 12 | 0.85 |
| polarC18 | 218.139 | 72.1394 | 12 | 0.85 |
| polarC18 | 218.139 | 230.7192 | 12 | 0.85 |
| polarC18 | 218.139 | 131.4489 | 12 | 0.85 |
| polarC18 | 218.139 | 142.9127 | 12 | 0.85 |
| polarC18 | 218.139 | 219.6219 | 12 | 0.85 |
| polarC18 | 218.139 | 102.8737 | 12 | 0.85 |
| polarC18 | 218.139 | 124.4287 | 12 | 0.85 |
| polarC18 | 218.139 | 112.478 | 12 | 0.85 |
| polarC18 | 218.139 | 87.7369 | 12 | 0.85 |
| polarC18 | 218.139 | 117.7943 | 12 | 0.85 |
| polarC18 | 218.139 | 61.2296 | 12 | 0.85 |

| | | | | |
|---|---|---|---|---|
| polarC18 | 218.139 | 73.9396 | 12 | 0.85 |
| polarC18 | 218.139 | 94.6002 | 12 | 0.85 |
| polarC18 | 218.139 | 106.889 | 12 | 0.85 |
| polarC18 | 218.139 | 110.6695 | 12 | 0.85 |
| polarC18 | 218.139 | 119.5783 | 12 | 0.85 |
| polarC18 | 218.139 | 81.3929 | 12 | 0.85 |
| polarC18 | 218.139 | 80.1279 | 12 | 0.85 |
| polarC18 | 218.139 | 64.6457 | 12 | 0.85 |
| polarC18 | 218.139 | 61.9317 | 12 | 0.85 |
| polarC18 | 218.139 | 211.1583 | 12 | 0.85 |
| polarC18 | 218.139 | 68.3337 | 12 | 0.85 |
| polarC18 | 218.139 | 87.4545 | 12 | 0.85 |
| polarC18 | 218.139 | 103.9979 | 12 | 0.85 |
| polarC18 | 218.139 | 60.5532 | 12 | 0.85 |
| polarC18 | 218.139 | 161.6487 | 12 | 0.85 |
| polarC18 | 218.139 | 115.4519 | 12 | 0.85 |
| polarC18 | 218.139 | 206.5896 | 12 | 0.85 |
| polarC18 | 218.139 | 96.517 | 12 | 0.85 |
| polarC18 | 218.139 | 95.7573 | 12 | 0.85 |
| polarC18 | 218.139 | 129.2171 | 12 | 0.85 |
| polarC18 | 218.139 | 142.5873 | 12 | 0.85 |
| polarC18 | 218.139 | 132.256 | 12 | 0.85 |
| polarC18 | 218.139 | 123.3179 | 12 | 0.85 |
| polarC18 | 218.139 | 71.0673 | 12 | 0.85 |
| polarC18 | 218.139 | 118.698 | 12 | 0.85 |
| polarC18 | 218.139 | 51.5076 | 12 | 0.85 |
| polarC18 | 218.139 | 222.4252 | 12 | 0.85 |
| polarC18 | 218.139 | 58.7139 | 12 | 0.85 |
| polarC18 | 218.139 | 135.0835 | 12 | 0.85 |
| polarC18 | 218.139 | 78.5427 | 12 | 0.85 |
| polarC18 | 218.139 | 88.5651 | 12 | 0.85 |
| polarC18 | 218.139 | 58.6665 | 12 | 0.85 |
| polarC18 | 218.139 | 69.867 | 12 | 0.85 |
| polarC18 | 218.139 | 50.7353 | 12 | 0.85 |
| polarC18 | 218.139 | 199.2151 | 12 | 0.85 |
| polarC18 | 218.139 | 227.4862 | 12 | 0.85 |
| polarC18 | 218.139 | 121.8318 | 12 | 0.85 |
| polarC18 | 218.139 | 131.3715 | 12 | 0.85 |
| polarC18 | 218.139 | 121.7216 | 12 | 0.85 |
| polarC18 | 218.139 | 65.6721 | 12 | 0.85 |
| polarC18 | 218.139 | 51.8425 | 11.9 | 0.85 |
| polarC18 | 218.139 | 127.0335 | 11.9 | 0.85 |

| | | | | |
|---|---|---|---|---|
| polarC18 | 218.139 | 52.589 | 11.9 | 0.85 |
| polarC18 | 218.139 | 145.9991 | 11.9 | 0.85 |
| polarC18 | 218.139 | 57.7794 | 11.9 | 0.85 |
| polarC18 | 218.139 | 87.6749 | 11.9 | 0.85 |
| polarC18 | 218.139 | 153.8141 | 11.9 | 0.85 |
| polarC18 | 218.139 | 64.3122 | 11.9 | 0.85 |
| polarC18 | 218.139 | 48.6793 | 11.9 | 0.85 |
| polarC18 | 218.139 | 170.6393 | 11.9 | 0.85 |
| polarC18 | 218.139 | 164.2142 | 11.9 | 0.85 |
| polarC18 | 218.139 | 173.7666 | 11.9 | 0.85 |
| polarC18 | 218.139 | 73.629 | 11.9 | 0.85 |
| polarC18 | 218.139 | 99.5453 | 11.9 | 0.85 |
| polarC18 | 218.139 | 107.3889 | 11.9 | 0.85 |
| polarC18 | 218.139 | 60.268 | 11.9 | 0.85 |
| polarC18 | 218.139 | 165.9117 | 11.9 | 0.85 |
| polarC18 | 218.139 | 125.8938 | 11.9 | 0.85 |
| polarC18 | 218.139 | 69.2626 | 11.9 | 0.85 |
| polarC18 | 218.139 | 147.3949 | 11.9 | 0.85 |
| polarC18 | 218.139 | 75.21 | 11.9 | 0.85 |
| polarC18 | 218.139 | 210.3409 | 11.9 | 0.85 |
| polarC18 | 218.139 | 100.1562 | 11.9 | 0.85 |
| polarC18 | 218.139 | 152.716 | 11.9 | 0.85 |
| polarC18 | 218.139 | 68.9158 | 11.9 | 0.85 |
| polarC18 | 218.139 | 78.8578 | 11.9 | 0.85 |
| polarC18 | 218.139 | 136.2034 | 11.9 | 0.84 |
| polarC18 | 218.139 | 78.0257 | 11.9 | 0.84 |
| polarC18 | 218.139 | 101.2968 | 11.9 | 0.84 |
| polarC18 | 218.139 | 229.5607 | 11.9 | 0.84 |
| polarC18 | 218.139 | 104.7306 | 11.9 | 0.84 |
| polarC18 | 218.139 | 91.7644 | 11.9 | 0.84 |
| polarC18 | 218.139 | 69.221 | 11.9 | 0.84 |
| polarC18 | 218.139 | 64.5274 | 11.9 | 0.84 |
| polarC18 | 218.139 | 58.1137 | 11.9 | 0.84 |
| polarC18 | 218.139 | 64.7514 | 11.9 | 0.84 |
| polarC18 | 218.139 | 73.2778 | 11.9 | 0.84 |
| polarC18 | 218.139 | 186.1045 | 11.9 | 0.84 |
| polarC18 | 218.139 | 70.8006 | 11.9 | 0.84 |
| polarC18 | 218.139 | 98.7111 | 11.9 | 0.84 |
| polarC18 | 218.139 | 74.0234 | 11.9 | 0.84 |
| polarC18 | 218.139 | 202.3749 | 11.9 | 0.84 |
| polarC18 | 218.139 | 152.0178 | 11.9 | 0.84 |
| polarC18 | 218.139 | 181.4292 | 11.9 | 0.84 |

| | | | | |
|---|---|---|---|---|
| polarC18 | 218.139 | 191.5112 | 11.9 | 0.84 |
| polarC18 | 218.139 | 63.0687 | 11.9 | 0.84 |
| polarC18 | 218.139 | 52.8334 | 11.9 | 0.84 |
| polarC18 | 218.139 | 105.1897 | 11.9 | 0.84 |
| polarC18 | 218.139 | 107.651 | 11.9 | 0.84 |
| polarC18 | 218.139 | 49.7258 | 11.9 | 0.84 |
| polarC18 | 218.139 | 79.2794 | 11.9 | 0.84 |
| polarC18 | 218.139 | 147.2722 | 11.9 | 0.84 |
| polarC18 | 218.139 | 49.1355 | 11.9 | 0.84 |
| polarC18 | 218.139 | 91.3409 | 11.8 | 0.84 |
| polarC18 | 218.139 | 57.4481 | 11.8 | 0.84 |
| polarC18 | 218.139 | 241.2901 | 11.8 | 0.84 |
| polarC18 | 218.139 | 79.0347 | 11.8 | 0.84 |
| polarC18 | 218.139 | 50.7997 | 11.8 | 0.84 |
| polarC18 | 218.139 | 230.5368 | 11.8 | 0.84 |
| polarC18 | 218.139 | 70.5709 | 11.8 | 0.84 |
| polarC18 | 218.139 | 121.6272 | 11.8 | 0.84 |
| polarC18 | 218.139 | 142.9739 | 11.8 | 0.84 |
| polarC18 | 218.139 | 98.1978 | 11.8 | 0.84 |
| polarC18 | 218.139 | 59.4519 | 11.8 | 0.84 |
| polarC18 | 218.139 | 224.5758 | 11.8 | 0.84 |
| polarC18 | 218.139 | 113.5067 | 11.8 | 0.84 |
| polarC18 | 218.139 | 163.9756 | 11.8 | 0.84 |
| polarC18 | 218.139 | 127.2463 | 11.8 | 0.84 |
| polarC18 | 218.139 | 49.1004 | 11.8 | 0.84 |
| polarC18 | 218.139 | 218.3545 | 11.8 | 0.84 |
| polarC18 | 218.139 | 89.6028 | 11.8 | 0.84 |
| polarC18 | 218.139 | 83.6098 | 11.8 | 0.84 |
| polarC18 | 218.139 | 95.3019 | 11.8 | 0.84 |
| polarC18 | 218.139 | 199.1577 | 11.8 | 0.84 |
| polarC18 | 218.139 | 88.5914 | 11.8 | 0.84 |
| polarC18 | 218.139 | 55.8271 | 11.8 | 0.84 |
| polarC18 | 218.139 | 50.9418 | 11.8 | 0.84 |
| polarC18 | 218.139 | 60.5467 | 11.8 | 0.84 |
| polarC18 | 218.139 | 69.1982 | 11.8 | 0.84 |
| polarC18 | 218.139 | 153.0206 | 11.8 | 0.84 |
| polarC18 | 218.139 | 88.0734 | 11.8 | 0.84 |
| polarC18 | 218.139 | 214.7985 | 11.8 | 0.84 |
| polarC18 | 218.139 | 137.7481 | 11.8 | 0.84 |
| polarC18 | 218.139 | 126.3033 | 11.8 | 0.84 |
| polarC18 | 218.139 | 192.8368 | 11.8 | 0.84 |
| polarC18 | 218.139 | 119.1638 | 11.8 | 0.84 |

| | | | | |
|---|---|---|---|---|
| polarC18 | 218.139 | 127.8832 | 11.8 | 0.84 |
| polarC18 | 218.139 | 49.8954 | 11.8 | 0.83 |
| polarC18 | 218.139 | 94.0756 | 11.8 | 0.83 |
| polarC18 | 218.139 | 186.7773 | 11.8 | 0.83 |
| polarC18 | 218.139 | 105.0706 | 11.8 | 0.83 |
| polarC18 | 218.139 | 147.8219 | 11.8 | 0.83 |
| polarC18 | 218.139 | 90.751 | 11.8 | 0.83 |
| polarC18 | 218.139 | 56.2744 | 11.8 | 0.83 |
| polarC18 | 218.139 | 201.0732 | 11.7 | 0.83 |
| polarC18 | 218.139 | 168.1201 | 11.7 | 0.83 |
| polarC18 | 218.139 | 108.5057 | 11.7 | 0.83 |
| polarC18 | 218.139 | 176.5294 | 11.7 | 0.83 |
| polarC18 | 218.139 | 58.4346 | 11.7 | 0.83 |
| polarC18 | 218.139 | 204.3915 | 11.7 | 0.83 |
| polarC18 | 218.139 | 97.8773 | 11.7 | 0.83 |
| polarC18 | 218.139 | 49.9624 | 11.7 | 0.83 |
| polarC18 | 218.139 | 162.6656 | 11.7 | 0.83 |
| polarC18 | 218.139 | 156.9001 | 11.7 | 0.83 |
| polarC18 | 218.139 | 134.866 | 11.7 | 0.83 |
| polarC18 | 218.139 | 53.9452 | 11.7 | 0.83 |
| polarC18 | 218.139 | 91.5136 | 11.7 | 0.83 |
| polarC18 | 218.139 | 149.3276 | 11.7 | 0.83 |
| polarC18 | 218.139 | 93.5182 | 11.7 | 0.83 |
| polarC18 | 218.139 | 86.6896 | 11.7 | 0.83 |
| polarC18 | 218.139 | 84.1789 | 11.7 | 0.83 |
| polarC18 | 218.139 | 143.9569 | 11.7 | 0.83 |
| polarC18 | 218.139 | 167.1906 | 11.7 | 0.83 |
| polarC18 | 218.139 | 67.1001 | 11.7 | 0.83 |
| polarC18 | 218.139 | 107.2982 | 11.7 | 0.83 |
| polarC18 | 218.139 | 147.4632 | 11.7 | 0.83 |
| polarC18 | 218.139 | 75.3754 | 11.7 | 0.83 |
| polarC18 | 218.139 | 179.7968 | 11.7 | 0.83 |
| polarC18 | 218.139 | 219.2169 | 11.7 | 0.83 |
| polarC18 | 218.139 | 67.0236 | 11.7 | 0.83 |
| polarC18 | 218.139 | 85.7077 | 11.7 | 0.83 |
| polarC18 | 218.139 | 57.5403 | 11.7 | 0.83 |
| polarC18 | 218.139 | 189.1393 | 11.7 | 0.83 |
| polarC18 | 218.139 | 136.6376 | 11.7 | 0.83 |
| polarC18 | 218.139 | 80.842 | 11.7 | 0.83 |
| polarC18 | 218.139 | 215.2872 | 11.7 | 0.83 |
| polarC18 | 218.139 | 70.3631 | 11.7 | 0.83 |
| polarC18 | 218.139 | 71.9978 | 11.7 | 0.83 |

| | | | | |
|---|---|---|---|---|
| polarC18 | 218.139 | 56.208 | 11.7 | 0.83 |
| polarC18 | 218.139 | 109.5674 | 11.7 | 0.83 |
| polarC18 | 218.139 | 143.8441 | 11.7 | 0.83 |
| polarC18 | 218.139 | 126.5724 | 11.7 | 0.83 |
| polarC18 | 218.139 | 104.2202 | 11.7 | 0.83 |
| polarC18 | 218.139 | 99.2881 | 11.7 | 0.83 |
| polarC18 | 218.139 | 49.393 | 11.7 | 0.83 |
| polarC18 | 218.139 | 197.8217 | 11.7 | 0.83 |
| polarC18 | 218.139 | 145.535 | 11.7 | 0.83 |
| polarC18 | 218.139 | 85.8109 | 11.7 | 0.83 |
| polarC18 | 218.139 | 102.3566 | 11.7 | 0.83 |
| polarC18 | 218.139 | 98.6295 | 11.7 | 0.83 |
| polarC18 | 218.139 | 64.3473 | 11.7 | 0.83 |
| polarC18 | 218.139 | 111.7806 | 11.7 | 0.83 |
| polarC18 | 218.139 | 196.2443 | 11.6 | 0.83 |
| polarC18 | 218.139 | 188.5223 | 11.6 | 0.83 |
| polarC18 | 218.139 | 50.9787 | 11.6 | 0.83 |
| polarC18 | 218.139 | 227.2866 | 11.6 | 0.83 |
| polarC18 | 218.139 | 53.9313 | 11.6 | 0.83 |
| polarC18 | 218.139 | 120.6839 | 11.6 | 0.83 |
| polarC18 | 218.139 | 71.3197 | 11.6 | 0.83 |
| polarC18 | 218.139 | 52.4661 | 11.6 | 0.83 |
| polarC18 | 218.139 | 106.8822 | 11.6 | 0.83 |
| polarC18 | 218.139 | 82.0475 | 11.6 | 0.83 |
| polarC18 | 218.139 | 98.239 | 11.6 | 0.83 |
| polarC18 | 218.139 | 82.0273 | 11.6 | 0.82 |
| polarC18 | 218.139 | 58.9856 | 11.6 | 0.82 |
| polarC18 | 218.139 | 74.7849 | 11.6 | 0.82 |
| polarC18 | 218.139 | 56.9309 | 11.6 | 0.82 |
| polarC18 | 218.139 | 58.5146 | 11.6 | 0.82 |
| polarC18 | 218.139 | 151.4086 | 11.6 | 0.82 |
| polarC18 | 218.139 | 51.5346 | 11.6 | 0.82 |
| polarC18 | 218.139 | 146.7483 | 11.6 | 0.82 |
| polarC18 | 218.139 | 109.7597 | 11.6 | 0.82 |
| polarC18 | 218.139 | 175.3485 | 11.6 | 0.82 |
| polarC18 | 218.139 | 53.2655 | 11.6 | 0.82 |
| polarC18 | 218.139 | 124.9696 | 11.6 | 0.82 |
| polarC18 | 218.139 | 155.0139 | 11.6 | 0.82 |
| polarC18 | 218.139 | 146.7884 | 11.6 | 0.82 |
| polarC18 | 218.139 | 98.8621 | 11.6 | 0.82 |
| polarC18 | 218.139 | 130.5113 | 11.6 | 0.82 |
| polarC18 | 218.139 | 120.2251 | 11.6 | 0.82 |

| | | | | |
|---|---|---|---|---|
| polarC18 | 218.139 | 125.805 | 11.6 | 0.82 |
| polarC18 | 218.139 | 51.2431 | 11.6 | 0.82 |
| polarC18 | 218.139 | 70.4639 | 11.6 | 0.82 |
| polarC18 | 218.139 | 89.4228 | 11.6 | 0.82 |
| polarC18 | 218.139 | 168.873 | 11.6 | 0.82 |
| polarC18 | 218.139 | 155.1636 | 11.6 | 0.82 |
| polarC18 | 218.139 | 52.8528 | 11.6 | 0.82 |
| polarC18 | 218.139 | 68.2951 | 11.6 | 0.82 |
| polarC18 | 218.139 | 210.8145 | 11.6 | 0.82 |
| polarC18 | 218.139 | 83.5562 | 11.6 | 0.82 |
| polarC18 | 218.139 | 114.2995 | 11.6 | 0.82 |
| polarC18 | 218.139 | 228.5677 | 11.5 | 0.82 |
| polarC18 | 218.139 | 62.4808 | 11.5 | 0.82 |
| polarC18 | 218.139 | 88.9716 | 11.5 | 0.82 |
| polarC18 | 218.139 | 90.915 | 11.5 | 0.82 |
| polarC18 | 218.139 | 185.2055 | 11.5 | 0.82 |
| polarC18 | 218.139 | 82.1288 | 11.5 | 0.82 |
| polarC18 | 218.139 | 71.5138 | 11.5 | 0.82 |
| polarC18 | 218.139 | 138.3134 | 11.5 | 0.82 |
| polarC18 | 218.139 | 57.2429 | 11.5 | 0.82 |
| polarC18 | 218.139 | 54.8349 | 11.5 | 0.82 |
| polarC18 | 218.139 | 204.9089 | 11.5 | 0.82 |
| polarC18 | 218.139 | 58.88 | 11.5 | 0.82 |
| polarC18 | 218.139 | 53.6925 | 11.5 | 0.82 |
| polarC18 | 218.139 | 146.3158 | 11.5 | 0.82 |
| polarC18 | 218.139 | 53.2199 | 11.5 | 0.82 |
| polarC18 | 218.139 | 104.1733 | 11.5 | 0.82 |
| polarC18 | 218.139 | 164.8537 | 11.5 | 0.82 |
| polarC18 | 218.139 | 130.4605 | 11.5 | 0.82 |
| polarC18 | 218.139 | 88.832 | 11.5 | 0.82 |
| polarC18 | 218.139 | 60.495 | 11.5 | 0.82 |
| polarC18 | 218.139 | 81.9234 | 11.5 | 0.82 |
| polarC18 | 218.139 | 157.0039 | 11.5 | 0.82 |
| polarC18 | 218.139 | 181.8281 | 11.5 | 0.81 |
| polarC18 | 218.139 | 160.086 | 11.5 | 0.81 |
| polarC18 | 218.139 | 106.4051 | 11.5 | 0.81 |
| polarC18 | 218.139 | 119.4206 | 11.5 | 0.81 |
| polarC18 | 218.139 | 176.1944 | 11.5 | 0.81 |
| polarC18 | 218.139 | 56.1881 | 11.5 | 0.81 |
| polarC18 | 218.139 | 60.6134 | 11.5 | 0.81 |
| polarC18 | 218.139 | 91.8795 | 11.5 | 0.81 |
| polarC18 | 218.139 | 106.5314 | 11.5 | 0.81 |

| | | | | |
|---|---|---|---|---|
| polarC18 | 218.139 | 63.1172 | 11.5 | 0.81 |
| polarC18 | 218.139 | 58.5665 | 11.5 | 0.81 |
| polarC18 | 218.139 | 153.614 | 11.5 | 0.81 |
| polarC18 | 218.139 | 183.2136 | 11.5 | 0.81 |
| polarC18 | 218.139 | 124.85 | 11.5 | 0.81 |
| polarC18 | 218.139 | 242.3542 | 11.4 | 0.81 |
| polarC18 | 218.139 | 156.0305 | 11.4 | 0.81 |
| polarC18 | 218.139 | 99.427 | 11.4 | 0.81 |
| polarC18 | 218.139 | 72.2221 | 11.4 | 0.81 |
| polarC18 | 218.139 | 219.1587 | 11.4 | 0.81 |
| polarC18 | 218.139 | 147.4879 | 11.4 | 0.81 |
| polarC18 | 218.139 | 62.7069 | 11.4 | 0.81 |
| polarC18 | 218.139 | 196.8234 | 11.4 | 0.81 |
| polarC18 | 218.139 | 132.581 | 11.4 | 0.81 |
| polarC18 | 218.139 | 92.515 | 11.4 | 0.81 |
| polarC18 | 218.139 | 218.6573 | 11.4 | 0.81 |
| polarC18 | 218.139 | 164.9242 | 11.4 | 0.81 |
| polarC18 | 218.139 | 89.2313 | 11.4 | 0.81 |
| polarC18 | 218.139 | 118.512 | 11.4 | 0.81 |
| polarC18 | 218.139 | 123.0263 | 11.4 | 0.81 |
| polarC18 | 218.139 | 199.2886 | 11.4 | 0.81 |
| polarC18 | 218.139 | 197.0583 | 11.4 | 0.81 |
| polarC18 | 218.139 | 73.02 | 11.4 | 0.81 |
| polarC18 | 218.139 | 79.3335 | 11.4 | 0.81 |
| polarC18 | 218.139 | 182.3757 | 11.4 | 0.81 |
| polarC18 | 218.139 | 51.5287 | 11.4 | 0.81 |
| polarC18 | 218.139 | 167.7715 | 11.4 | 0.81 |
| polarC18 | 218.139 | 69.9791 | 11.4 | 0.81 |
| polarC18 | 218.139 | 88 | 11.4 | 0.81 |
| polarC18 | 218.139 | 118.2858 | 11.4 | 0.81 |
| polarC18 | 218.139 | 74.5425 | 11.4 | 0.81 |
| polarC18 | 218.139 | 212.0403 | 11.4 | 0.81 |
| polarC18 | 218.139 | 105.9886 | 11.4 | 0.81 |
| polarC18 | 218.139 | 79.3697 | 11.4 | 0.81 |
| polarC18 | 218.139 | 132.7049 | 11.4 | 0.81 |
| polarC18 | 218.139 | 63.4691 | 11.4 | 0.81 |
| polarC18 | 218.139 | 176.7926 | 11.4 | 0.81 |
| polarC18 | 218.139 | 61.5861 | 11.4 | 0.81 |
| polarC18 | 218.139 | 126.6991 | 11.4 | 0.81 |
| polarC18 | 218.139 | 80.4807 | 11.4 | 0.81 |
| polarC18 | 218.139 | 75.1043 | 11.4 | 0.81 |
| polarC18 | 218.139 | 102.026 | 11.4 | 0.81 |

| | | | | |
|---|---|---|---|---|
| polarC18 | 218.139 | 66.5475 | 11.4 | 0.81 |
| polarC18 | 218.139 | 70.8475 | 11.3 | 0.8 |
| polarC18 | 218.139 | 70.8966 | 11.3 | 0.8 |
| polarC18 | 218.139 | 57.0875 | 11.3 | 0.8 |
| polarC18 | 218.139 | 126.162 | 11.3 | 0.8 |
| polarC18 | 218.139 | 61.0376 | 11.3 | 0.8 |
| polarC18 | 218.139 | 169.4598 | 11.3 | 0.8 |
| polarC18 | 218.139 | 110.6951 | 11.3 | 0.8 |
| polarC18 | 218.139 | 122.7601 | 11.3 | 0.8 |
| polarC18 | 218.139 | 94.3047 | 11.3 | 0.8 |
| polarC18 | 218.139 | 49.3121 | 11.3 | 0.8 |
| polarC18 | 218.139 | 51.9666 | 11.3 | 0.8 |
| polarC18 | 218.139 | 57.0408 | 11.3 | 0.8 |
| polarC18 | 218.139 | 54.4837 | 11.3 | 0.8 |
| polarC18 | 218.139 | 54.4519 | 11.3 | 0.8 |
| polarC18 | 218.139 | 142.6496 | 11.3 | 0.8 |
| polarC18 | 218.139 | 190.0063 | 11.3 | 0.8 |
| polarC18 | 218.139 | 50.7225 | 11.3 | 0.8 |
| polarC18 | 218.139 | 50.2074 | 11.3 | 0.8 |
| polarC18 | 218.139 | 58.4756 | 11.3 | 0.8 |
| polarC18 | 218.139 | 66.3119 | 11.3 | 0.8 |
| polarC18 | 218.139 | 171.3749 | 11.3 | 0.8 |
| polarC18 | 218.139 | 97.5404 | 11.3 | 0.8 |
| polarC18 | 218.139 | 65.7148 | 11.3 | 0.8 |
| polarC18 | 218.139 | 95.058 | 11.3 | 0.8 |
| polarC18 | 218.139 | 239.6718 | 11.3 | 0.8 |
| polarC18 | 218.139 | 52.7327 | 11.3 | 0.8 |
| polarC18 | 218.139 | 127.626 | 11.3 | 0.8 |
| polarC18 | 218.139 | 151.6711 | 11.3 | 0.8 |
| polarC18 | 218.139 | 168.2996 | 11.3 | 0.8 |
| polarC18 | 218.139 | 88.0487 | 11.3 | 0.8 |
| polarC18 | 218.139 | 72.265 | 11.3 | 0.8 |
| polarC18 | 218.139 | 58.3859 | 11.3 | 0.8 |
| polarC18 | 218.139 | 169.2886 | 11.3 | 0.8 |
| polarC18 | 218.139 | 66.9146 | 11.3 | 0.8 |
| polarC18 | 218.139 | 128.5482 | 11.3 | 0.8 |
| polarC18 | 218.139 | 175.1476 | 11.3 | 0.8 |
| polarC18 | 218.139 | 50.0027 | 11.3 | 0.8 |
| polarC18 | 218.139 | 228.9106 | 11.3 | 0.8 |
| polarC18 | 218.139 | 55.0304 | 11.3 | 0.8 |
| polarC18 | 218.139 | 160.9565 | 11.3 | 0.8 |
| polarC18 | 218.139 | 53.8591 | 11.3 | 0.8 |

| | | | | |
|---|---|---|---|---|
| polarC18 | 218.139 | 177.4104 | 11.3 | 0.8 |
| polarC18 | 218.139 | 54.9257 | 11.3 | 0.8 |
| polarC18 | 218.139 | 168.8033 | 11.3 | 0.8 |
| polarC18 | 218.139 | 86.565 | 11.2 | 0.8 |
| polarC18 | 218.139 | 68.4 | 11.2 | 0.8 |
| polarC18 | 218.139 | 167.1733 | 11.2 | 0.8 |
| polarC18 | 218.139 | 70.9641 | 11.2 | 0.8 |
| polarC18 | 218.139 | 82.5798 | 11.2 | 0.8 |
| polarC18 | 218.139 | 94.4874 | 11.2 | 0.8 |
| polarC18 | 218.139 | 74.8066 | 11.2 | 0.8 |
| polarC18 | 218.139 | 64.0885 | 11.2 | 0.8 |
| polarC18 | 218.139 | 51.9318 | 11.2 | 0.8 |
| polarC18 | 218.139 | 77.4375 | 11.2 | 0.8 |
| polarC18 | 218.139 | 156.2978 | 11.2 | 0.8 |
| polarC18 | 218.139 | 48.8886 | 11.2 | 0.8 |
| polarC18 | 218.139 | 55.0402 | 11.2 | 0.8 |
| polarC18 | 218.139 | 81.0717 | 11.2 | 0.8 |
| polarC18 | 218.139 | 61.9582 | 11.2 | 0.79 |
| polarC18 | 218.139 | 99.0315 | 11.2 | 0.79 |
| polarC18 | 218.139 | 68.0776 | 11.2 | 0.79 |
| polarC18 | 218.139 | 157.6273 | 11.2 | 0.79 |
| polarC18 | 218.139 | 64.9107 | 11.2 | 0.79 |
| polarC18 | 218.139 | 65.7716 | 11.2 | 0.79 |
| polarC18 | 218.139 | 69.3991 | 11.2 | 0.79 |
| polarC18 | 218.139 | 69.5108 | 11.2 | 0.79 |
| polarC18 | 218.139 | 68.7357 | 11.2 | 0.79 |
| polarC18 | 218.139 | 145.6309 | 11.2 | 0.79 |
| polarC18 | 218.139 | 78.3692 | 11.2 | 0.79 |
| polarC18 | 218.139 | 113.3681 | 11.2 | 0.79 |
| polarC18 | 218.139 | 88.302 | 11.2 | 0.79 |
| polarC18 | 218.139 | 231.5918 | 11.2 | 0.79 |
| polarC18 | 218.139 | 54.1612 | 11.2 | 0.79 |
| polarC18 | 218.139 | 139.4931 | 11.2 | 0.79 |
| polarC18 | 218.139 | 73.664 | 11.2 | 0.79 |
| polarC18 | 218.139 | 85.1462 | 11.2 | 0.79 |
| polarC18 | 218.139 | 66.3338 | 11.2 | 0.79 |
| polarC18 | 218.139 | 101.456 | 11.2 | 0.79 |
| polarC18 | 218.139 | 78.5961 | 11.1 | 0.79 |
| polarC18 | 218.139 | 57.5937 | 11.1 | 0.79 |
| polarC18 | 218.139 | 89.6247 | 11.1 | 0.79 |
| polarC18 | 218.139 | 53.4528 | 11.1 | 0.79 |
| polarC18 | 218.139 | 188.3717 | 11.1 | 0.79 |

| | | | | |
|---|---|---|---|---|
| polarC18 | 218.139 | 114.1182 | 11.1 | 0.79 |
| polarC18 | 218.139 | 73.6879 | 11.1 | 0.79 |
| polarC18 | 218.139 | 102.6791 | 11.1 | 0.79 |
| polarC18 | 218.139 | 58.4508 | 11.1 | 0.79 |
| polarC18 | 218.139 | 54.7845 | 11.1 | 0.79 |
| polarC18 | 218.139 | 87.8457 | 11.1 | 0.79 |
| polarC18 | 218.139 | 156.1993 | 11.1 | 0.79 |
| polarC18 | 218.139 | 103.1748 | 11.1 | 0.79 |
| polarC18 | 218.139 | 119.5973 | 11.1 | 0.79 |
| polarC18 | 218.139 | 80.2031 | 11.1 | 0.79 |
| polarC18 | 218.139 | 138.1204 | 11.1 | 0.79 |
| polarC18 | 218.139 | 208.2514 | 11.1 | 0.79 |
| polarC18 | 218.139 | 51.167 | 11.1 | 0.79 |
| polarC18 | 218.139 | 64.711 | 11.1 | 0.79 |
| polarC18 | 218.139 | 68.4852 | 11.1 | 0.79 |
| polarC18 | 218.139 | 66.9473 | 11.1 | 0.79 |
| polarC18 | 218.139 | 93.385 | 11.1 | 0.79 |
| polarC18 | 218.139 | 61.0684 | 11.1 | 0.79 |
| polarC18 | 218.139 | 70.9675 | 11.1 | 0.79 |
| polarC18 | 218.139 | 101.0243 | 11.1 | 0.78 |
| polarC18 | 218.139 | 69.4815 | 11.1 | 0.78 |
| polarC18 | 218.139 | 196.7018 | 11.1 | 0.78 |
| polarC18 | 218.139 | 102.6355 | 11.1 | 0.78 |
| polarC18 | 218.139 | 144.2686 | 11 | 0.78 |
| polarC18 | 218.139 | 93.7826 | 11 | 0.78 |
| polarC18 | 218.139 | 75.197 | 11 | 0.78 |
| polarC18 | 218.139 | 118.1996 | 11 | 0.78 |
| polarC18 | 218.139 | 84.8486 | 11 | 0.78 |
| polarC18 | 218.139 | 71.6481 | 11 | 0.78 |
| polarC18 | 218.139 | 61.7153 | 11 | 0.78 |
| polarC18 | 218.139 | 107.8085 | 11 | 0.78 |
| polarC18 | 218.139 | 224.2468 | 11 | 0.78 |
| polarC18 | 218.139 | 58.2722 | 11 | 0.78 |
| polarC18 | 218.139 | 175.6562 | 11 | 0.78 |
| polarC18 | 218.139 | 79.9382 | 11 | 0.78 |
| polarC18 | 218.139 | 102.2433 | 11 | 0.78 |
| polarC18 | 218.139 | 198.8697 | 11 | 0.78 |
| polarC18 | 218.139 | 58.7327 | 11 | 0.78 |
| polarC18 | 218.139 | 85.8292 | 11 | 0.78 |
| polarC18 | 218.139 | 85.9196 | 11 | 0.78 |
| polarC18 | 218.139 | 68.9364 | 10.9 | 0.78 |
| polarC18 | 218.139 | 72.996 | 10.9 | 0.78 |

| | | | | |
|---|---|---|---|---|
| polarC18 | 218.139 | 92.2094 | 10.9 | 0.77 |
| polarC18 | 218.139 | 72.454 | 10.9 | 0.77 |
| polarC18 | 218.139 | 88.3104 | 10.9 | 0.77 |
| polarC18 | 218.139 | 116.4056 | 10.9 | 0.77 |
| polarC18 | 218.139 | 62.2968 | 10.9 | 0.77 |
| polarC18 | 218.139 | 61.84 | 10.9 | 0.77 |
| polarC18 | 218.139 | 116.5242 | 10.9 | 0.77 |
| polarC18 | 218.139 | 97.453 | 10.9 | 0.77 |
| polarC18 | 218.139 | 164.576 | 10.9 | 0.77 |
| polarC18 | 218.139 | 52.0419 | 10.9 | 0.77 |
| polarC18 | 218.139 | 68.119 | 10.9 | 0.77 |
| polarC18 | 218.139 | 104.3531 | 10.9 | 0.77 |
| polarC18 | 218.139 | 64.732 | 10.9 | 0.77 |
| polarC18 | 218.139 | 70.5094 | 10.9 | 0.77 |
| polarC18 | 218.139 | 132.5663 | 10.9 | 0.77 |
| polarC18 | 218.139 | 75.8352 | 10.8 | 0.77 |
| polarC18 | 218.139 | 84.0235 | 10.8 | 0.77 |
| polarC18 | 218.139 | 50.047 | 10.8 | 0.77 |
| polarC18 | 218.139 | 52.555 | 10.8 | 0.77 |
| polarC18 | 218.139 | 132.3523 | 10.8 | 0.77 |
| polarC18 | 218.139 | 168.5752 | 10.8 | 0.77 |
| polarC18 | 218.139 | 136.4418 | 10.8 | 0.77 |
| polarC18 | 218.139 | 187.6314 | 10.8 | 0.77 |
| polarC18 | 218.139 | 109.0737 | 10.8 | 0.77 |
| polarC18 | 218.139 | 49.3528 | 10.8 | 0.76 |
| polarC18 | 218.139 | 67.7975 | 10.8 | 0.76 |
| polarC18 | 218.139 | 222.354 | 10.8 | 0.76 |
| polarC18 | 218.139 | 51.4172 | 10.8 | 0.76 |
| polarC18 | 218.139 | 53.8011 | 10.8 | 0.76 |
| polarC18 | 218.139 | 85.1032 | 10.8 | 0.76 |
| polarC18 | 218.139 | 54.7597 | 10.7 | 0.76 |
| polarC18 | 218.139 | 91.0689 | 10.7 | 0.76 |
| polarC18 | 218.139 | 64.8747 | 10.7 | 0.76 |
| polarC18 | 218.139 | 156.228 | 10.7 | 0.76 |
| polarC18 | 218.139 | 87.9088 | 10.7 | 0.76 |
| polarC18 | 218.139 | 58.6419 | 10.7 | 0.76 |
| polarC18 | 218.139 | 150.7648 | 10.7 | 0.76 |
| polarC18 | 218.139 | 50.7812 | 10.7 | 0.76 |
| polarC18 | 218.139 | 111.9178 | 10.7 | 0.76 |
| polarC18 | 218.139 | 94.5688 | 10.7 | 0.76 |
| polarC18 | 218.139 | 73.4828 | 10.7 | 0.76 |
| polarC18 | 218.139 | 111.5223 | 10.6 | 0.76 |

| | | | | |
|---|---|---|---|---|
| polarC18 | 218.139 | 71.5529 | 10.6 | 0.76 |
| polarC18 | 218.139 | 168.7541 | 10.6 | 0.75 |
| polarC18 | 218.139 | 93.2573 | 10.6 | 0.75 |
| polarC18 | 218.139 | 97.3113 | 10.6 | 0.75 |
| polarC18 | 218.139 | 51.587 | 10.6 | 0.75 |
| polarC18 | 218.139 | 129.1966 | 10.6 | 0.75 |
| polarC18 | 218.139 | 82.7802 | 10.6 | 0.75 |
| polarC18 | 218.139 | 158.1639 | 10.6 | 0.75 |
| polarC18 | 218.139 | 82.3232 | 10.6 | 0.75 |
| polarC18 | 218.139 | 85.7028 | 10.6 | 0.75 |
| polarC18 | 218.139 | 73.6946 | 10.5 | 0.75 |
| polarC18 | 218.139 | 107.6331 | 10.5 | 0.75 |
| polarC18 | 218.139 | 226.5447 | 10.5 | 0.75 |
| polarC18 | 218.139 | 176.3688 | 10.5 | 0.75 |
| polarC18 | 218.139 | 114.4178 | 10.5 | 0.75 |
| polarC18 | 218.139 | 76.4048 | 10.5 | 0.74 |
| polarC18 | 218.139 | 88.6446 | 10.5 | 0.74 |
| polarC18 | 218.139 | 66.0095 | 10.5 | 0.74 |
| polarC18 | 218.139 | 48.8684 | 10.5 | 0.74 |
| polarC18 | 218.139 | 83.9066 | 10.5 | 0.74 |
| polarC18 | 218.139 | 55.9735 | 10.5 | 0.74 |
| polarC18 | 218.139 | 210.0799 | 10.5 | 0.74 |
| polarC18 | 218.139 | 52.141 | 10.4 | 0.74 |
| polarC18 | 218.139 | 89.3949 | 10.4 | 0.74 |
| polarC18 | 218.139 | 78.656 | 10.4 | 0.74 |
| polarC18 | 218.139 | 63.9226 | 10.4 | 0.74 |
| polarC18 | 218.139 | 79.6179 | 10.4 | 0.74 |
| polarC18 | 218.139 | 136.7879 | 10.4 | 0.74 |
| polarC18 | 218.139 | 117.9091 | 10.3 | 0.73 |
| polarC18 | 218.139 | 49.8175 | 10.3 | 0.73 |
| polarC18 | 218.139 | 117.1462 | 10.3 | 0.73 |
| polarC18 | 218.139 | 62.6832 | 10.3 | 0.73 |
| polarC18 | 218.139 | 158.6532 | 10.3 | 0.73 |
| polarC18 | 218.139 | 88.7695 | 10.3 | 0.73 |
| polarC18 | 218.139 | 56.7317 | 10.2 | 0.73 |
| polarC18 | 218.139 | 127.8961 | 10.2 | 0.72 |
| polarC18 | 218.139 | 218.9151 | 10.2 | 0.72 |
| polarC18 | 218.139 | 49.776 | 10.2 | 0.72 |
| polarC18 | 218.139 | 177.1104 | 10.2 | 0.72 |
| polarC18 | 218.139 | 60.1316 | 10.2 | 0.72 |
| polarC18 | 218.139 | 63.2234 | 10.2 | 0.72 |
| polarC18 | 218.139 | 61.4193 | 10.1 | 0.72 |

| | | | | |
|---|---|---|---|---|
| polarC18 | 218.139 | 123.4018 | 10.1 | 0.72 |
| polarC18 | 218.139 | 156.7483 | 10.1 | 0.72 |
| polarC18 | 218.139 | 66.4292 | 10.1 | 0.72 |
| polarC18 | 218.139 | 98.1086 | 10.1 | 0.72 |
| polarC18 | 218.139 | 239.4028 | 10.1 | 0.72 |
| polarC18 | 218.139 | 152.5243 | 10.1 | 0.71 |
| polarC18 | 218.139 | 161.3634 | 10 | 0.71 |
| polarC18 | 218.139 | 59.5612 | 10 | 0.71 |
| polarC18 | 218.139 | 85.1814 | 10 | 0.71 |
| polarC18 | 218.139 | 59.8589 | 10 | 0.71 |
| polarC18 | 218.139 | 88.3675 | 10 | 0.71 |
| polarC18 | 218.139 | 177.2561 | 10 | 0.71 |
| polarC18 | 218.139 | 53.8324 | 9.9 | 0.71 |
| polarC18 | 218.139 | 57.5331 | 9.9 | 0.7 |
| polarC18 | 218.139 | 124.9315 | 9.8 | 0.7 |
| polarC18 | 218.139 | 213.5598 | 9.8 | 0.7 |
| polarC18 | 218.139 | 154.8485 | 9.8 | 0.69 |
| polarC18 | 218.139 | 126.1068 | 9.7 | 0.69 |
| polarC18 | 218.139 | 55.4019 | 9.7 | 0.69 |
| polarC18 | 218.139 | 81.7324 | 9.7 | 0.69 |
| polarC18 | 218.139 | 49.22 | 9.7 | 0.69 |
| polarC18 | 218.139 | 50.6692 | 9.7 | 0.69 |
| polarC18 | 218.139 | 138.1405 | 9.5 | 0.68 |
| polarC18 | 218.139 | 94.4485 | 9.5 | 0.67 |
| polarC18 | 218.139 | 54.1659 | 9.5 | 0.67 |
| polarC18 | 218.139 | 140.7941 | 9.5 | 0.67 |
| polarC18 | 218.139 | 169.3584 | 9.3 | 0.66 |
| polarC18 | 218.139 | 71.5998 | 9.3 | 0.66 |
| polarC18 | 218.139 | 153.8488 | 9.3 | 0.66 |
| polarC18 | 218.139 | 120.2489 | 9.2 | 0.66 |
| polarC18 | 218.139 | 148.0644 | 9.2 | 0.65 |
| polarC18 | 218.139 | 215.4471 | 9.2 | 0.65 |
| polarC18 | 218.139 | 68.1432 | 9 | 0.64 |
| polarC18 | 218.139 | 68.8016 | 9 | 0.64 |
| polarC18 | 218.139 | 77.2088 | 9 | 0.64 |
| polarC18 | 218.139 | 195.8684 | 9 | 0.64 |
| polarC18 | 218.139 | 75.8152 | 8.9 | 0.63 |
| polarC18 | 218.139 | 127.1687 | 8.9 | 0.63 |
| polarC18 | 218.139 | 74.7093 | 8.8 | 0.62 |
| polarC18 | 218.139 | 82.8818 | 8.8 | 0.62 |
| polarC18 | 218.139 | 113.9843 | 8.7 | 0.62 |
| polarC18 | 218.139 | 81.8583 | 8.7 | 0.62 |

| | | | | |
|---|---|---|---|---|
| polarC18 | 218.139 | 60.4739 | 8.7 | 0.62 |
| polarC18 | 218.139 | 80.6987 | 8.7 | 0.61 |
| polarC18 | 218.139 | 53.2412 | 8.7 | 0.61 |
| polarC18 | 218.139 | 51.2171 | 8.7 | 0.61 |
| polarC18 | 218.139 | 151.8945 | 8.7 | 0.61 |
| polarC18 | 218.139 | 132.7187 | 8.6 | 0.61 |
| polarC18 | 218.139 | 108.4511 | 8.6 | 0.61 |
| polarC18 | 218.139 | 76.9957 | 8.6 | 0.61 |
| polarC18 | 218.139 | 175.1835 | 8.6 | 0.61 |
| polarC18 | 218.139 | 189.5043 | 8.6 | 0.61 |
| polarC18 | 218.139 | 65.6013 | 8.6 | 0.61 |
| polarC18 | 218.139 | 237.5748 | 8.5 | 0.61 |
| polarC18 | 218.139 | 111.647 | 8.5 | 0.61 |
| polarC18 | 218.139 | 55.8487 | 8.5 | 0.6 |
| polarC18 | 218.139 | 58.027 | 8.5 | 0.6 |
| polarC18 | 218.139 | 152.4178 | 8.5 | 0.6 |
| polarC18 | 218.139 | 205.924 | 8.5 | 0.6 |
| polarC18 | 218.139 | 158.818 | 8.5 | 0.6 |
| polarC18 | 218.139 | 103.9103 | 8.4 | 0.6 |
| polarC18 | 218.139 | 138.2963 | 8.4 | 0.6 |
| polarC18 | 218.139 | 78.1562 | 8.4 | 0.6 |
| polarC18 | 218.139 | 90.9231 | 8.4 | 0.6 |
| polarC18 | 218.139 | 102.1749 | 8.4 | 0.6 |
| polarC18 | 218.139 | 90.7975 | 8.4 | 0.6 |
| polarC18 | 218.139 | 97.7883 | 8.4 | 0.6 |
| polarC18 | 218.139 | 71.211 | 8.4 | 0.6 |
| polarC18 | 218.139 | 202.9825 | 8.4 | 0.6 |
| polarC18 | 218.139 | 207.0773 | 8.4 | 0.59 |
| polarC18 | 218.139 | 90.0469 | 8.3 | 0.59 |
| polarC18 | 218.139 | 77.914 | 8.3 | 0.59 |
| polarC18 | 218.139 | 71.1082 | 8.3 | 0.59 |
| polarC18 | 218.139 | 60.592 | 8.3 | 0.59 |
| polarC18 | 218.139 | 135.9723 | 8.3 | 0.59 |
| polarC18 | 218.139 | 109.9928 | 8.3 | 0.59 |
| polarC18 | 218.139 | 65.3356 | 8.3 | 0.59 |
| polarC18 | 218.139 | 51.8776 | 8.3 | 0.59 |
| polarC18 | 218.139 | 165.1458 | 8.3 | 0.59 |
| polarC18 | 218.139 | 68.1777 | 8.3 | 0.59 |
| polarC18 | 218.139 | 210.2559 | 8.3 | 0.59 |
| polarC18 | 218.139 | 78.924 | 8.3 | 0.59 |
| polarC18 | 218.139 | 52.5642 | 8.3 | 0.59 |
| polarC18 | 218.139 | 129.795 | 8.3 | 0.59 |

| | | | | |
|---|---|---|---|---|
| polarC18 | 218.139 | 72.7273 | 8.2 | 0.59 |
| polarC18 | 218.139 | 145.2027 | 8.2 | 0.58 |
| polarC18 | 218.139 | 126.7577 | 8.2 | 0.58 |
| polarC18 | 218.139 | 197.5869 | 8.2 | 0.58 |
| polarC18 | 218.139 | 65.292 | 8.2 | 0.58 |
| polarC18 | 218.139 | 110.22 | 8.2 | 0.58 |
| polarC18 | 218.139 | 93.5305 | 8.2 | 0.58 |
| polarC18 | 218.139 | 180.1541 | 8.2 | 0.58 |
| polarC18 | 218.139 | 105.8453 | 8.2 | 0.58 |
| polarC18 | 218.139 | 56.955 | 8.2 | 0.58 |
| polarC18 | 218.139 | 49.9439 | 8.2 | 0.58 |
| polarC18 | 218.139 | 54.2983 | 8.2 | 0.58 |
| polarC18 | 218.139 | 55.2748 | 8.2 | 0.58 |
| polarC18 | 218.139 | 88.1192 | 8.2 | 0.58 |
| polarC18 | 218.139 | 75.6157 | 8.2 | 0.58 |
| polarC18 | 218.139 | 114.4352 | 8.2 | 0.58 |
| polarC18 | 218.139 | 59.3943 | 8.2 | 0.58 |
| polarC18 | 218.139 | 55.7667 | 8.2 | 0.58 |
| polarC18 | 218.139 | 49.7544 | 8.1 | 0.58 |
| polarC18 | 218.139 | 114.7316 | 8.1 | 0.58 |
| polarC18 | 218.139 | 125.1266 | 8.1 | 0.58 |
| polarC18 | 218.139 | 93.4238 | 8.1 | 0.58 |
| polarC18 | 218.139 | 157.8487 | 8.1 | 0.58 |
| polarC18 | 218.139 | 52.1523 | 8.1 | 0.58 |
| polarC18 | 218.139 | 70.8899 | 8.1 | 0.57 |
| polarC18 | 218.139 | 50.3193 | 8.1 | 0.57 |
| polarC18 | 218.139 | 85.043 | 8.1 | 0.57 |
| polarC18 | 218.139 | 134.7522 | 8.1 | 0.57 |
| polarC18 | 218.139 | 150.349 | 8.1 | 0.57 |
| polarC18 | 218.139 | 71.4957 | 8.1 | 0.57 |
| polarC18 | 218.139 | 67.0042 | 8.1 | 0.57 |
| polarC18 | 218.139 | 73.3851 | 8.1 | 0.57 |
| polarC18 | 218.139 | 228.6674 | 8.1 | 0.57 |
| polarC18 | 218.139 | 113.3147 | 8.1 | 0.57 |
| polarC18 | 218.139 | 103.6555 | 8.1 | 0.57 |
| polarC18 | 218.139 | 61.4389 | 8.1 | 0.57 |
| polarC18 | 218.139 | 59.994 | 8.1 | 0.57 |
| polarC18 | 218.139 | 109.145 | 8.1 | 0.57 |
| polarC18 | 218.139 | 59.1463 | 8.1 | 0.57 |
| polarC18 | 218.139 | 114.6399 | 8.1 | 0.57 |
| polarC18 | 218.139 | 232.8255 | 8 | 0.57 |
| polarC18 | 218.139 | 48.9314 | 8 | 0.57 |

| | | | | |
|---|---|---|---|---|
| polarC18 | 218.139 | 60.8153 | 8 | 0.57 |
| polarC18 | 218.139 | 86.3892 | 8 | 0.57 |
| polarC18 | 218.139 | 91.5616 | 8 | 0.57 |
| polarC18 | 218.139 | 205.6095 | 8 | 0.57 |
| polarC18 | 218.139 | 53.2139 | 8 | 0.57 |
| polarC18 | 218.139 | 235.176 | 8 | 0.57 |
| polarC18 | 218.139 | 163.5936 | 8 | 0.57 |
| polarC18 | 218.139 | 133.2874 | 8 | 0.57 |
| polarC18 | 218.139 | 69.3657 | 8 | 0.57 |
| polarC18 | 218.139 | 97.5135 | 8 | 0.57 |
| polarC18 | 218.139 | 58.1095 | 8 | 0.57 |
| polarC18 | 218.139 | 129.3429 | 8 | 0.57 |
| polarC18 | 218.139 | 54.2617 | 8 | 0.57 |
| polarC18 | 218.139 | 106.7575 | 8 | 0.57 |
| polarC18 | 218.139 | 73.2033 | 8 | 0.57 |
| polarC18 | 218.139 | 49.6054 | 8 | 0.57 |
| polarC18 | 218.139 | 56.5848 | 8 | 0.57 |
| polarC18 | 218.139 | 74.2761 | 8 | 0.57 |
| polarC18 | 218.139 | 173.5933 | 8 | 0.57 |
| polarC18 | 218.139 | 116.4421 | 8 | 0.57 |
| polarC18 | 218.139 | 76.3721 | 8 | 0.56 |
| polarC18 | 218.139 | 58.7801 | 8 | 0.56 |
| polarC18 | 218.139 | 58.3707 | 8 | 0.56 |
| polarC18 | 218.139 | 220.1216 | 7.9 | 0.56 |
| polarC18 | 218.139 | 125.0075 | 7.9 | 0.56 |
| polarC18 | 218.139 | 62.6421 | 7.9 | 0.56 |
| polarC18 | 218.139 | 109.8357 | 7.9 | 0.56 |
| polarC18 | 218.139 | 127.5641 | 7.9 | 0.56 |
| polarC18 | 218.139 | 236.5941 | 7.9 | 0.56 |
| polarC18 | 218.139 | 48.8413 | 7.9 | 0.56 |
| polarC18 | 218.139 | 59.8336 | 7.9 | 0.56 |
| polarC18 | 218.139 | 70.9272 | 7.9 | 0.56 |
| polarC18 | 218.139 | 71.1127 | 7.9 | 0.56 |
| polarC18 | 218.139 | 81.3569 | 7.9 | 0.56 |
| polarC18 | 218.139 | 150.8008 | 7.9 | 0.56 |
| polarC18 | 218.139 | 152.8062 | 7.9 | 0.56 |
| polarC18 | 218.139 | 65.7624 | 7.9 | 0.56 |
| polarC18 | 218.139 | 136.2242 | 7.9 | 0.56 |
| polarC18 | 218.139 | 73.3067 | 7.9 | 0.56 |
| polarC18 | 218.139 | 52.7988 | 7.9 | 0.56 |
| polarC18 | 218.139 | 100.7807 | 7.9 | 0.56 |
| polarC18 | 218.139 | 104.2003 | 7.9 | 0.56 |

| | | | | |
|---|---|---|---|---|
| polarC18 | 218.139 | 53.6324 | 7.9 | 0.56 |
| polarC18 | 218.139 | 183.6946 | 7.9 | 0.56 |
| polarC18 | 218.139 | 128.0602 | 7.9 | 0.56 |
| polarC18 | 218.139 | 190.994 | 7.9 | 0.56 |
| polarC18 | 218.139 | 69.4216 | 7.9 | 0.56 |
| polarC18 | 218.139 | 68.1678 | 7.9 | 0.56 |
| polarC18 | 218.139 | 71.52 | 7.9 | 0.56 |
| polarC18 | 218.139 | 62.0029 | 7.9 | 0.56 |
| polarC18 | 218.139 | 151.1314 | 7.9 | 0.56 |
| polarC18 | 218.139 | 101.1761 | 7.9 | 0.56 |
| polarC18 | 218.139 | 75.2574 | 7.9 | 0.56 |
| polarC18 | 218.139 | 108.2469 | 7.9 | 0.56 |
| polarC18 | 218.139 | 142.1385 | 7.9 | 0.56 |
| polarC18 | 218.139 | 68.7431 | 7.9 | 0.56 |
| polarC18 | 218.139 | 73.6158 | 7.9 | 0.56 |
| polarC18 | 218.139 | 49.7315 | 7.9 | 0.56 |
| polarC18 | 218.139 | 82.5487 | 7.9 | 0.56 |
| polarC18 | 218.139 | 51.5157 | 7.9 | 0.56 |
| polarC18 | 218.139 | 92.1214 | 7.9 | 0.56 |
| polarC18 | 218.139 | 84.0554 | 7.9 | 0.56 |
| polarC18 | 218.139 | 98.6383 | 7.9 | 0.56 |
| polarC18 | 218.139 | 49.5251 | 7.9 | 0.56 |
| polarC18 | 218.139 | 96.7614 | 7.9 | 0.56 |
| polarC18 | 218.139 | 81.9951 | 7.8 | 0.56 |
| polarC18 | 218.139 | 49.321 | 7.8 | 0.56 |
| polarC18 | 218.139 | 121.109 | 7.8 | 0.56 |
| polarC18 | 218.139 | 146.441 | 7.8 | 0.56 |
| polarC18 | 218.139 | 94.1722 | 7.8 | 0.56 |
| polarC18 | 218.139 | 81.4294 | 7.8 | 0.56 |
| polarC18 | 218.139 | 99.0642 | 7.8 | 0.56 |
| polarC18 | 218.139 | 210.7027 | 7.8 | 0.56 |
| polarC18 | 218.139 | 80.0851 | 7.8 | 0.56 |
| polarC18 | 218.139 | 60.2545 | 7.8 | 0.56 |
| polarC18 | 218.139 | 68.0192 | 7.8 | 0.56 |
| polarC18 | 218.139 | 57.6496 | 7.8 | 0.56 |
| polarC18 | 218.139 | 132.0925 | 7.8 | 0.56 |
| polarC18 | 218.139 | 82.0216 | 7.8 | 0.55 |
| polarC18 | 218.139 | 74.8462 | 7.8 | 0.55 |
| polarC18 | 218.139 | 57.7358 | 7.8 | 0.55 |
| polarC18 | 218.139 | 198.2204 | 7.8 | 0.55 |
| polarC18 | 218.139 | 148.536 | 7.8 | 0.55 |
| polarC18 | 218.139 | 146.4653 | 7.8 | 0.55 |

| | | | | |
|---|---|---|---|---|
| polarC18 | 218.139 | 81.8089 | 7.8 | 0.55 |
| polarC18 | 218.139 | 56.7237 | 7.8 | 0.55 |
| polarC18 | 218.139 | 106.0867 | 7.8 | 0.55 |
| polarC18 | 218.139 | 108.9867 | 7.8 | 0.55 |
| polarC18 | 218.139 | 73.9198 | 7.8 | 0.55 |
| polarC18 | 218.139 | 62.3579 | 7.8 | 0.55 |
| polarC18 | 218.139 | 53.799 | 7.8 | 0.55 |
| polarC18 | 218.139 | 59.9622 | 7.8 | 0.55 |
| polarC18 | 218.139 | 104.2836 | 7.8 | 0.55 |
| polarC18 | 218.139 | 144.6942 | 7.8 | 0.55 |
| polarC18 | 218.139 | 69.5689 | 7.8 | 0.55 |
| polarC18 | 218.139 | 83.8253 | 7.8 | 0.55 |
| polarC18 | 218.139 | 66.9634 | 7.8 | 0.55 |
| polarC18 | 218.139 | 121.5307 | 7.8 | 0.55 |
| polarC18 | 218.139 | 64.0502 | 7.8 | 0.55 |
| polarC18 | 218.139 | 143.4835 | 7.8 | 0.55 |
| polarC18 | 218.139 | 200.4639 | 7.8 | 0.55 |
| polarC18 | 218.139 | 52.4706 | 7.8 | 0.55 |
| polarC18 | 218.139 | 72.1511 | 7.8 | 0.55 |
| polarC18 | 218.139 | 199.4102 | 7.8 | 0.55 |
| polarC18 | 218.139 | 96.2033 | 7.8 | 0.55 |
| polarC18 | 218.139 | 69.194 | 7.8 | 0.55 |
| polarC18 | 218.139 | 87.0187 | 7.8 | 0.55 |
| polarC18 | 218.139 | 60.2405 | 7.8 | 0.55 |
| polarC18 | 218.139 | 116.124 | 7.8 | 0.55 |
| polarC18 | 218.139 | 51.6808 | 7.8 | 0.55 |
| polarC18 | 218.139 | 239.1604 | 7.8 | 0.55 |
| polarC18 | 218.139 | 128.6284 | 7.8 | 0.55 |
| polarC18 | 218.139 | 57.6018 | 7.8 | 0.55 |
| polarC18 | 218.139 | 123.5748 | 7.8 | 0.55 |
| polarC18 | 218.139 | 49.6175 | 7.8 | 0.55 |
| polarC18 | 218.139 | 73.3779 | 7.8 | 0.55 |
| polarC18 | 218.139 | 54.8218 | 7.8 | 0.55 |
| polarC18 | 218.139 | 65.1359 | 7.8 | 0.55 |
| polarC18 | 218.139 | 53.5863 | 7.8 | 0.55 |
| polarC18 | 218.139 | 177.5733 | 7.8 | 0.55 |
| polarC18 | 218.139 | 68.4641 | 7.7 | 0.55 |
| polarC18 | 218.139 | 112.7865 | 7.7 | 0.55 |
| polarC18 | 218.139 | 140.0537 | 7.7 | 0.55 |
| polarC18 | 218.139 | 65.5456 | 7.7 | 0.55 |
| polarC18 | 218.139 | 81.7631 | 7.7 | 0.55 |
| polarC18 | 218.139 | 83.7771 | 7.7 | 0.55 |

| | | | | |
|---|---|---|---|---|
| polarC18 | 218.139 | 92.0067 | 7.7 | 0.55 |
| polarC18 | 218.139 | 82.9416 | 7.7 | 0.55 |
| polarC18 | 218.139 | 125.819 | 7.7 | 0.55 |
| polarC18 | 218.139 | 59.9338 | 7.7 | 0.55 |
| polarC18 | 218.139 | 215.6116 | 7.7 | 0.55 |
| polarC18 | 218.139 | 118.7978 | 7.7 | 0.55 |
| polarC18 | 218.139 | 92.3317 | 7.7 | 0.55 |
| polarC18 | 218.139 | 115.4332 | 7.7 | 0.55 |
| polarC18 | 218.139 | 200.6961 | 7.7 | 0.55 |
| polarC18 | 218.139 | 58.8922 | 7.7 | 0.55 |
| polarC18 | 218.139 | 107.0705 | 7.7 | 0.55 |
| polarC18 | 218.139 | 54.1159 | 7.7 | 0.55 |
| polarC18 | 218.139 | 166.1494 | 7.7 | 0.55 |
| polarC18 | 218.139 | 122.9794 | 7.7 | 0.55 |
| polarC18 | 218.139 | 65.8302 | 7.7 | 0.55 |
| polarC18 | 218.139 | 60.3875 | 7.7 | 0.55 |
| polarC18 | 218.139 | 60.9099 | 7.7 | 0.55 |
| polarC18 | 218.139 | 234.2255 | 7.7 | 0.55 |
| polarC18 | 218.139 | 116.6223 | 7.7 | 0.55 |
| polarC18 | 218.139 | 67.1662 | 7.7 | 0.55 |
| polarC18 | 218.139 | 71.7953 | 7.7 | 0.55 |
| polarC18 | 218.139 | 69.1086 | 7.7 | 0.55 |
| polarC18 | 218.139 | 94.094 | 7.7 | 0.55 |
| polarC18 | 218.139 | 155.5035 | 7.7 | 0.55 |
| polarC18 | 218.139 | 131.7923 | 7.7 | 0.55 |
| polarC18 | 218.139 | 78.4343 | 7.7 | 0.55 |
| polarC18 | 218.139 | 63.5426 | 7.7 | 0.55 |
| polarC18 | 218.139 | 77.109 | 7.7 | 0.55 |
| polarC18 | 218.139 | 91.0904 | 7.7 | 0.55 |
| polarC18 | 218.139 | 90.6981 | 7.7 | 0.55 |
| polarC18 | 218.139 | 55.9811 | 7.7 | 0.55 |
| polarC18 | 218.139 | 67.245 | 7.7 | 0.55 |
| polarC18 | 218.139 | 90.8328 | 7.7 | 0.55 |
| polarC18 | 218.139 | 50.4624 | 7.7 | 0.55 |
| polarC18 | 218.139 | 51.0795 | 7.7 | 0.54 |
| polarC18 | 218.139 | 110.3298 | 7.7 | 0.54 |
| polarC18 | 218.139 | 146.208 | 7.7 | 0.54 |
| polarC18 | 218.139 | 67.4377 | 7.7 | 0.54 |
| polarC18 | 218.139 | 48.7284 | 7.7 | 0.54 |
| polarC18 | 218.139 | 56.7052 | 7.7 | 0.54 |
| polarC18 | 218.139 | 61.2976 | 7.7 | 0.54 |
| polarC18 | 218.139 | 184.4323 | 7.7 | 0.54 |

| | | | | |
|---|---|---|---|---|
| polarC18 | 218.139 | 64.8993 | 7.7 | 0.54 |
| polarC18 | 218.139 | 54.2835 | 7.7 | 0.54 |
| polarC18 | 218.139 | 62.9574 | 7.7 | 0.54 |
| polarC18 | 218.139 | 88.1979 | 7.7 | 0.54 |
| polarC18 | 218.139 | 60.2481 | 7.7 | 0.54 |
| polarC18 | 218.139 | 65.2827 | 7.7 | 0.54 |
| polarC18 | 218.139 | 57.0486 | 7.7 | 0.54 |
| polarC18 | 218.139 | 146.0313 | 7.7 | 0.54 |
| polarC18 | 218.139 | 125.2886 | 7.7 | 0.54 |
| polarC18 | 218.139 | 171.898 | 7.7 | 0.54 |
| polarC18 | 218.139 | 81.5974 | 7.7 | 0.54 |
| polarC18 | 218.139 | 49.6239 | 7.7 | 0.54 |
| polarC18 | 218.139 | 143.0024 | 7.7 | 0.54 |
| polarC18 | 218.139 | 224.8646 | 7.7 | 0.54 |
| polarC18 | 218.139 | 184.2418 | 7.7 | 0.54 |
| polarC18 | 218.139 | 55.7424 | 7.7 | 0.54 |
| polarC18 | 218.139 | 218.6861 | 7.7 | 0.54 |
| polarC18 | 218.139 | 63.8894 | 7.7 | 0.54 |
| polarC18 | 218.139 | 104.7508 | 7.7 | 0.54 |
| polarC18 | 218.139 | 134.4145 | 7.6 | 0.54 |
| polarC18 | 218.139 | 50.4903 | 7.6 | 0.54 |
| polarC18 | 218.139 | 62.8483 | 7.6 | 0.54 |
| polarC18 | 218.139 | 111.1919 | 7.6 | 0.54 |
| polarC18 | 218.139 | 105.1057 | 7.6 | 0.54 |
| polarC18 | 218.139 | 76.9898 | 7.6 | 0.54 |
| polarC18 | 218.139 | 89.4523 | 7.6 | 0.54 |
| polarC18 | 218.139 | 51.4298 | 7.6 | 0.54 |
| polarC18 | 218.139 | 142.4212 | 7.6 | 0.54 |
| polarC18 | 218.139 | 108.273 | 7.6 | 0.54 |
| polarC18 | 218.139 | 85.1719 | 7.6 | 0.54 |
| polarC18 | 218.139 | 68.4968 | 7.6 | 0.54 |
| polarC18 | 218.139 | 73.0727 | 7.6 | 0.54 |
| polarC18 | 218.139 | 230.8741 | 7.6 | 0.54 |
| polarC18 | 218.139 | 63.0186 | 7.6 | 0.54 |
| polarC18 | 218.139 | 65.6545 | 7.6 | 0.54 |
| polarC18 | 218.139 | 59.0227 | 7.6 | 0.54 |
| polarC18 | 218.139 | 61.8283 | 7.6 | 0.54 |
| polarC18 | 218.139 | 65.9658 | 7.6 | 0.54 |
| polarC18 | 218.139 | 143.5547 | 7.6 | 0.54 |
| polarC18 | 218.139 | 163.9365 | 7.6 | 0.54 |
| polarC18 | 218.139 | 55.6682 | 7.6 | 0.54 |
| polarC18 | 218.139 | 111.2502 | 7.6 | 0.54 |

| | | | | |
|---|---|---|---|---|
| polarC18 | 218.139 | 58.24 | 7.6 | 0.54 |
| polarC18 | 218.139 | 121.3194 | 7.6 | 0.54 |
| polarC18 | 218.139 | 77.8333 | 7.6 | 0.54 |
| polarC18 | 218.139 | 146.0777 | 7.6 | 0.54 |
| polarC18 | 218.139 | 119.7056 | 7.6 | 0.54 |
| polarC18 | 218.139 | 128.148 | 7.6 | 0.54 |
| polarC18 | 218.139 | 82.2494 | 7.6 | 0.54 |
| polarC18 | 218.139 | 117.3926 | 7.6 | 0.54 |
| polarC18 | 218.139 | 195.6692 | 7.6 | 0.54 |
| polarC18 | 218.139 | 53.822 | 7.6 | 0.54 |
| polarC18 | 218.139 | 98.1476 | 7.6 | 0.54 |
| polarC18 | 218.139 | 57.1359 | 7.6 | 0.54 |
| polarC18 | 218.139 | 70.1438 | 7.6 | 0.54 |
| polarC18 | 218.139 | 50.8548 | 7.6 | 0.54 |
| polarC18 | 218.139 | 49.8099 | 7.6 | 0.54 |
| polarC18 | 218.139 | 49.8457 | 7.6 | 0.54 |
| polarC18 | 218.139 | 51.4546 | 7.6 | 0.54 |
| polarC18 | 218.139 | 50.5381 | 7.6 | 0.54 |
| polarC18 | 218.139 | 78.6979 | 7.6 | 0.54 |
| polarC18 | 218.139 | 70.1732 | 7.6 | 0.54 |
| polarC18 | 218.139 | 49.1538 | 7.6 | 0.54 |
| polarC18 | 218.139 | 113.9511 | 7.6 | 0.54 |
| polarC18 | 218.139 | 89.8621 | 7.6 | 0.54 |
| polarC18 | 218.139 | 101.1689 | 7.6 | 0.54 |
| polarC18 | 218.139 | 49.3856 | 7.6 | 0.54 |
| polarC18 | 218.139 | 74.0976 | 7.6 | 0.54 |
| polarC18 | 218.139 | 125.8757 | 7.6 | 0.54 |
| polarC18 | 218.139 | 94.7306 | 7.6 | 0.54 |
| polarC18 | 218.139 | 142.0354 | 7.6 | 0.54 |
| polarC18 | 218.139 | 108.0629 | 7.6 | 0.54 |
| polarC18 | 218.139 | 101.8298 | 7.6 | 0.54 |
| polarC18 | 218.139 | 175.4557 | 7.6 | 0.54 |
| polarC18 | 218.139 | 57.9055 | 7.6 | 0.54 |
| polarC18 | 218.139 | 104.8908 | 7.6 | 0.54 |
| polarC18 | 218.139 | 74.4807 | 7.6 | 0.54 |
| polarC18 | 218.139 | 150.4654 | 7.6 | 0.54 |
| polarC18 | 218.139 | 63.3582 | 7.6 | 0.54 |
| polarC18 | 218.139 | 57.1887 | 7.6 | 0.54 |
| polarC18 | 218.139 | 85.2278 | 7.6 | 0.54 |
| polarC18 | 218.139 | 96.5938 | 7.6 | 0.54 |
| polarC18 | 218.139 | 49.1504 | 7.6 | 0.54 |
| polarC18 | 218.139 | 93.8153 | 7.6 | 0.54 |

| | | | | |
|---|---|---|---|---|
| polarC18 | 218.139 | 53.9186 | 7.6 | 0.54 |
| polarC18 | 218.139 | 147.3259 | 7.6 | 0.54 |
| polarC18 | 218.139 | 185.2485 | 7.6 | 0.54 |
| polarC18 | 218.139 | 127.3002 | 7.6 | 0.54 |
| polarC18 | 218.139 | 70.7481 | 7.6 | 0.54 |
| polarC18 | 218.139 | 142.5064 | 7.6 | 0.54 |
| polarC18 | 218.139 | 110.9054 | 7.6 | 0.54 |
| polarC18 | 218.139 | 150.3767 | 7.6 | 0.54 |
| polarC18 | 218.139 | 50.323 | 7.6 | 0.54 |
| polarC18 | 218.139 | 97.7564 | 7.6 | 0.54 |
| polarC18 | 218.139 | 94.2808 | 7.6 | 0.54 |
| polarC18 | 218.139 | 94.8238 | 7.6 | 0.54 |
| polarC18 | 218.139 | 95.7651 | 7.6 | 0.54 |
| polarC18 | 218.139 | 78.3568 | 7.6 | 0.54 |
| polarC18 | 218.139 | 102.9686 | 7.6 | 0.54 |
| polarC18 | 218.139 | 53.9392 | 7.6 | 0.54 |
| polarC18 | 218.139 | 76.3469 | 7.6 | 0.54 |
| polarC18 | 218.139 | 143.8769 | 7.6 | 0.54 |
| polarC18 | 218.139 | 50.0673 | 7.6 | 0.54 |
| polarC18 | 218.139 | 81.9621 | 7.5 | 0.54 |
| polarC18 | 218.139 | 209.1799 | 7.5 | 0.54 |
| polarC18 | 218.139 | 113.6661 | 7.5 | 0.54 |
| polarC18 | 218.139 | 102.6718 | 7.5 | 0.54 |
| polarC18 | 218.139 | 118.8628 | 7.5 | 0.54 |
| polarC18 | 218.139 | 165.6863 | 7.5 | 0.54 |
| polarC18 | 218.139 | 121.5926 | 7.5 | 0.54 |
| polarC18 | 218.139 | 54.1569 | 7.5 | 0.53 |
| polarC18 | 218.139 | 74.6602 | 7.5 | 0.53 |
| polarC18 | 218.139 | 49.5217 | 7.5 | 0.53 |
| polarC18 | 218.139 | 51.1806 | 7.5 | 0.53 |
| polarC18 | 218.139 | 86.6664 | 7.5 | 0.53 |
| polarC18 | 218.139 | 101.6491 | 7.5 | 0.53 |
| polarC18 | 218.139 | 81.1694 | 7.5 | 0.53 |
| polarC18 | 218.139 | 110.7068 | 7.5 | 0.53 |
| polarC18 | 218.139 | 94.2542 | 7.5 | 0.53 |
| polarC18 | 218.139 | 124.3761 | 7.5 | 0.53 |
| polarC18 | 218.139 | 49.9244 | 7.5 | 0.53 |
| polarC18 | 218.139 | 68.2742 | 7.5 | 0.53 |
| polarC18 | 218.139 | 76.029 | 7.5 | 0.53 |
| polarC18 | 218.139 | 95.37 | 7.5 | 0.53 |
| polarC18 | 218.139 | 79.1849 | 7.5 | 0.53 |
| polarC18 | 218.139 | 81.7788 | 7.5 | 0.53 |

| | | | | |
|---|---|---|---|---|
| polarC18 | 218.139 | 78.3915 | 7.5 | 0.53 |
| polarC18 | 218.139 | 188.2716 | 7.5 | 0.53 |
| polarC18 | 218.139 | 48.8807 | 7.5 | 0.53 |
| polarC18 | 218.139 | 110.1866 | 7.5 | 0.53 |
| polarC18 | 218.139 | 56.3011 | 7.5 | 0.53 |
| polarC18 | 218.139 | 63.5712 | 7.5 | 0.53 |
| polarC18 | 218.139 | 81.1059 | 7.5 | 0.53 |
| polarC18 | 218.139 | 70.3773 | 7.5 | 0.53 |
| polarC18 | 218.139 | 65.1198 | 7.5 | 0.53 |
| polarC18 | 218.139 | 93.6196 | 7.5 | 0.53 |
| polarC18 | 218.139 | 49.6898 | 7.5 | 0.53 |
| polarC18 | 218.139 | 112.9332 | 7.5 | 0.53 |
| polarC18 | 218.139 | 100.4305 | 7.5 | 0.53 |
| polarC18 | 218.139 | 234.5113 | 7.5 | 0.53 |
| polarC18 | 218.139 | 135.9481 | 7.5 | 0.53 |
| polarC18 | 218.139 | 178.9859 | 7.5 | 0.53 |
| polarC18 | 218.139 | 192.9977 | 7.5 | 0.53 |
| polarC18 | 218.139 | 148.4796 | 7.5 | 0.53 |
| polarC18 | 218.139 | 64.3241 | 7.5 | 0.53 |
| polarC18 | 218.139 | 68.2419 | 7.5 | 0.53 |
| polarC18 | 218.139 | 82.6231 | 7.5 | 0.53 |
| polarC18 | 218.139 | 158.1077 | 7.5 | 0.53 |
| polarC18 | 218.139 | 130.2031 | 7.5 | 0.53 |
| polarC18 | 218.139 | 82.3107 | 7.5 | 0.53 |
| polarC18 | 218.139 | 189.4562 | 7.5 | 0.53 |
| polarC18 | 218.139 | 71.0944 | 7.5 | 0.53 |
| polarC18 | 218.139 | 95.7733 | 7.5 | 0.53 |
| polarC18 | 218.139 | 114.0115 | 7.5 | 0.53 |
| polarC18 | 218.139 | 129.6038 | 7.5 | 0.53 |
| polarC18 | 218.139 | 186.6742 | 7.5 | 0.53 |
| polarC18 | 218.139 | 107.9329 | 7.5 | 0.53 |
| polarC18 | 218.139 | 50.6311 | 7.5 | 0.53 |
| polarC18 | 218.139 | 52.3735 | 7.5 | 0.53 |
| polarC18 | 218.139 | 64.6749 | 7.5 | 0.53 |
| polarC18 | 218.139 | 64.4245 | 7.5 | 0.53 |
| polarC18 | 218.139 | 78.2247 | 7.5 | 0.53 |
| polarC18 | 218.139 | 68.595 | 7.5 | 0.53 |
| polarC18 | 218.139 | 78.0934 | 7.5 | 0.53 |
| polarC18 | 218.139 | 68.0851 | 7.5 | 0.53 |
| polarC18 | 218.139 | 119.9027 | 7.5 | 0.53 |
| polarC18 | 218.139 | 93.466 | 7.5 | 0.53 |
| polarC18 | 218.139 | 207.1703 | 7.5 | 0.53 |

| | | | | |
|---|---|---|---|---|
| polarC18 | 218.139 | 160.7152 | 7.5 | 0.53 |
| polarC18 | 218.139 | 132.2397 | 7.5 | 0.53 |
| polarC18 | 218.139 | 119.9227 | 7.5 | 0.53 |
| polarC18 | 218.139 | 150.99 | 7.5 | 0.53 |
| polarC18 | 218.139 | 62.2513 | 7.5 | 0.53 |
| polarC18 | 218.139 | 59.1719 | 7.5 | 0.53 |
| polarC18 | 218.139 | 64.9448 | 7.5 | 0.53 |
| polarC18 | 218.139 | 70.0791 | 7.5 | 0.53 |
| polarC18 | 218.139 | 108.1553 | 7.5 | 0.53 |
| polarC18 | 218.139 | 159.5716 | 7.5 | 0.53 |
| polarC18 | 218.139 | 49.6455 | 7.5 | 0.53 |
| polarC18 | 218.139 | 62.2371 | 7.5 | 0.53 |
| polarC18 | 218.139 | 96.6516 | 7.5 | 0.53 |
| polarC18 | 218.139 | 109.6966 | 7.5 | 0.53 |
| polarC18 | 218.139 | 134.0931 | 7.5 | 0.53 |
| polarC18 | 218.139 | 231.1725 | 7.5 | 0.53 |
| polarC18 | 218.139 | 134.0205 | 7.5 | 0.53 |
| polarC18 | 218.139 | 112.0279 | 7.5 | 0.53 |
| polarC18 | 218.139 | 132.632 | 7.5 | 0.53 |
| polarC18 | 218.139 | 75.6648 | 7.5 | 0.53 |
| polarC18 | 218.139 | 88.0105 | 7.5 | 0.53 |
| polarC18 | 218.139 | 90.5855 | 7.5 | 0.53 |
| polarC18 | 218.139 | 98.3114 | 7.5 | 0.53 |
| polarC18 | 218.139 | 54.0634 | 7.4 | 0.53 |
| polarC18 | 218.139 | 109.7728 | 7.4 | 0.53 |
| polarC18 | 218.139 | 61.5961 | 7.4 | 0.53 |
| polarC18 | 218.139 | 134.8875 | 7.4 | 0.53 |
| polarC18 | 218.139 | 90.48 | 7.4 | 0.53 |
| polarC18 | 218.139 | 87.6 | 7.4 | 0.53 |
| polarC18 | 218.139 | 171.5915 | 7.4 | 0.53 |
| polarC18 | 218.139 | 197.2187 | 7.4 | 0.53 |
| polarC18 | 218.139 | 190.7896 | 7.4 | 0.53 |
| polarC18 | 218.139 | 60.4466 | 7.4 | 0.53 |
| polarC18 | 218.139 | 82.9088 | 7.4 | 0.53 |
| polarC18 | 218.139 | 53.816 | 7.4 | 0.53 |
| polarC18 | 218.139 | 139.0223 | 7.4 | 0.53 |
| polarC18 | 218.139 | 137.9977 | 7.4 | 0.53 |
| polarC18 | 218.139 | 56.7157 | 7.4 | 0.53 |
| polarC18 | 218.139 | 56.633 | 7.4 | 0.53 |
| polarC18 | 218.139 | 236.3791 | 7.4 | 0.53 |
| polarC18 | 218.139 | 114.7839 | 7.4 | 0.53 |
| polarC18 | 218.139 | 99.6616 | 7.4 | 0.53 |

| | | | | |
|---|---|---|---|---|
| polarC18 | 218.139 | 66.2927 | 7.4 | 0.53 |
| polarC18 | 218.139 | 190.812 | 7.4 | 0.53 |
| polarC18 | 218.139 | 128.9923 | 7.4 | 0.53 |
| polarC18 | 218.139 | 74.1009 | 7.4 | 0.53 |
| polarC18 | 218.139 | 49.3945 | 7.4 | 0.53 |
| polarC18 | 218.139 | 129.0361 | 7.4 | 0.53 |
| polarC18 | 218.139 | 98.7931 | 7.4 | 0.53 |
| polarC18 | 218.139 | 50.6938 | 7.4 | 0.53 |
| polarC18 | 218.139 | 100.0783 | 7.4 | 0.53 |
| polarC18 | 218.139 | 133.4709 | 7.4 | 0.53 |
| polarC18 | 218.139 | 72.242 | 7.4 | 0.53 |
| polarC18 | 218.139 | 61.3093 | 7.4 | 0.53 |
| polarC18 | 218.139 | 83.8676 | 7.4 | 0.53 |
| polarC18 | 218.139 | 150.034 | 7.4 | 0.53 |
| polarC18 | 218.139 | 70.6887 | 7.4 | 0.53 |
| polarC18 | 218.139 | 98.0356 | 7.4 | 0.53 |
| polarC18 | 218.139 | 80.8921 | 7.4 | 0.53 |
| polarC18 | 218.139 | 50.7063 | 7.4 | 0.53 |
| polarC18 | 218.139 | 65.1007 | 7.4 | 0.53 |
| polarC18 | 218.139 | 103.2086 | 7.4 | 0.53 |
| polarC18 | 218.139 | 63.361 | 7.4 | 0.53 |
| polarC18 | 218.139 | 72.7619 | 7.4 | 0.53 |
| polarC18 | 218.139 | 157.2695 | 7.4 | 0.53 |
| polarC18 | 218.139 | 107.1022 | 7.4 | 0.52 |
| polarC18 | 218.139 | 66.5019 | 7.4 | 0.52 |
| polarC18 | 218.139 | 94.8911 | 7.4 | 0.52 |
| polarC18 | 218.139 | 103.6696 | 7.4 | 0.52 |
| polarC18 | 218.139 | 84.5176 | 7.4 | 0.52 |
| polarC18 | 218.139 | 79.4452 | 7.4 | 0.52 |
| polarC18 | 218.139 | 75.3446 | 7.4 | 0.52 |
| polarC18 | 218.139 | 64.0437 | 7.4 | 0.52 |
| polarC18 | 218.139 | 80.863 | 7.4 | 0.52 |
| polarC18 | 218.139 | 186.4473 | 7.4 | 0.52 |
| polarC18 | 218.139 | 80.8237 | 7.4 | 0.52 |
| polarC18 | 218.139 | 98.583 | 7.4 | 0.52 |
| polarC18 | 218.139 | 159.7658 | 7.4 | 0.52 |
| polarC18 | 218.139 | 58.8527 | 7.4 | 0.52 |
| polarC18 | 218.139 | 193.7122 | 7.4 | 0.52 |
| polarC18 | 218.139 | 189.6968 | 7.4 | 0.52 |
| polarC18 | 218.139 | 71.9821 | 7.4 | 0.52 |
| polarC18 | 218.139 | 52.2926 | 7.4 | 0.52 |
| polarC18 | 218.139 | 124.3481 | 7.4 | 0.52 |

| | | | | |
|---|---|---|---|---|
| polarC18 | 218.139 | 201.7549 | 7.4 | 0.52 |
| polarC18 | 218.139 | 196.0866 | 7.4 | 0.52 |
| polarC18 | 218.139 | 171.6647 | 7.4 | 0.52 |
| polarC18 | 218.139 | 108.8146 | 7.4 | 0.52 |
| polarC18 | 218.139 | 60.5884 | 7.4 | 0.52 |
| polarC18 | 218.139 | 207.8085 | 7.4 | 0.52 |
| polarC18 | 218.139 | 179.2968 | 7.4 | 0.52 |
| polarC18 | 218.139 | 128.2737 | 7.4 | 0.52 |
| polarC18 | 218.139 | 80.7217 | 7.4 | 0.52 |
| polarC18 | 218.139 | 79.5293 | 7.4 | 0.52 |
| polarC18 | 218.139 | 80.0651 | 7.4 | 0.52 |
| polarC18 | 218.139 | 161.9043 | 7.4 | 0.52 |
| polarC18 | 218.139 | 173.2039 | 7.4 | 0.52 |
| polarC18 | 218.139 | 128.5736 | 7.4 | 0.52 |
| polarC18 | 218.139 | 213.9881 | 7.4 | 0.52 |
| polarC18 | 218.139 | 55.2622 | 7.4 | 0.52 |
| polarC18 | 218.139 | 218.8153 | 7.4 | 0.52 |
| polarC18 | 218.139 | 83.0558 | 7.4 | 0.52 |
| polarC18 | 218.139 | 56.7611 | 7.4 | 0.52 |
| polarC18 | 218.139 | 90.9768 | 7.4 | 0.52 |
| polarC18 | 218.139 | 71.8023 | 7.4 | 0.52 |
| polarC18 | 218.139 | 61.3592 | 7.4 | 0.52 |
| polarC18 | 218.139 | 56.2855 | 7.4 | 0.52 |
| polarC18 | 218.139 | 229.5995 | 7.4 | 0.52 |
| polarC18 | 218.139 | 95.604 | 7.4 | 0.52 |
| polarC18 | 218.139 | 186.7209 | 7.4 | 0.52 |
| polarC18 | 218.139 | 65.1704 | 7.4 | 0.52 |
| polarC18 | 218.139 | 84.8889 | 7.4 | 0.52 |
| polarC18 | 218.139 | 86.409 | 7.4 | 0.52 |
| polarC18 | 218.139 | 116.7029 | 7.4 | 0.52 |
| polarC18 | 218.139 | 69.9566 | 7.4 | 0.52 |
| polarC18 | 218.139 | 187.1199 | 7.4 | 0.52 |
| polarC18 | 218.139 | 165.4342 | 7.4 | 0.52 |
| polarC18 | 218.139 | 51.4627 | 7.4 | 0.52 |
| polarC18 | 218.139 | 120.0985 | 7.4 | 0.52 |
| polarC18 | 218.139 | 149.6746 | 7.4 | 0.52 |
| polarC18 | 218.139 | 119.5471 | 7.4 | 0.52 |
| polarC18 | 218.139 | 78.9673 | 7.4 | 0.52 |
| polarC18 | 218.139 | 237.4656 | 7.4 | 0.52 |
| polarC18 | 218.139 | 52.8825 | 7.4 | 0.52 |
| polarC18 | 218.139 | 69.0173 | 7.4 | 0.52 |
| polarC18 | 218.139 | 97.992 | 7.4 | 0.52 |

| | | | | |
|---|---|---|---|---|
| polarC18 | 218.139 | 58.4832 | 7.4 | 0.52 |
| polarC18 | 218.139 | 196.9262 | 7.4 | 0.52 |
| polarC18 | 218.139 | 210.0468 | 7.4 | 0.52 |
| polarC18 | 218.139 | 105.5578 | 7.3 | 0.52 |
| polarC18 | 218.139 | 70.7369 | 7.3 | 0.52 |
| polarC18 | 218.139 | 58.6327 | 7.3 | 0.52 |
| polarC18 | 218.139 | 173.8646 | 7.3 | 0.52 |
| polarC18 | 218.139 | 178.9487 | 7.3 | 0.52 |
| polarC18 | 218.139 | 77.3213 | 7.3 | 0.52 |
| polarC18 | 218.139 | 91.915 | 7.3 | 0.52 |
| polarC18 | 218.139 | 74.2472 | 7.3 | 0.52 |
| polarC18 | 218.139 | 152.4838 | 7.3 | 0.52 |
| polarC18 | 218.139 | 52.5601 | 7.3 | 0.52 |
| polarC18 | 218.139 | 51.9529 | 7.3 | 0.52 |
| polarC18 | 218.139 | 179.6175 | 7.3 | 0.52 |
| polarC18 | 218.139 | 198.3206 | 7.3 | 0.52 |
| polarC18 | 218.139 | 118.8737 | 7.3 | 0.52 |
| polarC18 | 218.139 | 88.5112 | 7.3 | 0.52 |
| polarC18 | 218.139 | 58.7581 | 7.3 | 0.52 |
| polarC18 | 218.139 | 84.6878 | 7.3 | 0.52 |
| polarC18 | 218.139 | 84.5714 | 7.3 | 0.52 |
| polarC18 | 218.139 | 65.9796 | 7.3 | 0.52 |
| polarC18 | 218.139 | 157.9338 | 7.3 | 0.52 |
| polarC18 | 218.139 | 59.8087 | 7.3 | 0.52 |
| polarC18 | 218.139 | 123.8695 | 7.3 | 0.52 |
| polarC18 | 218.139 | 50.1426 | 7.3 | 0.52 |
| polarC18 | 218.139 | 71.6361 | 7.3 | 0.52 |
| polarC18 | 218.139 | 91.7443 | 7.3 | 0.52 |
| polarC18 | 218.139 | 60.8689 | 7.3 | 0.52 |
| polarC18 | 218.139 | 48.6098 | 7.3 | 0.52 |
| polarC18 | 218.139 | 90.3143 | 7.3 | 0.52 |
| polarC18 | 218.139 | 73.4987 | 7.3 | 0.52 |
| polarC18 | 218.139 | 148.3852 | 7.3 | 0.52 |
| polarC18 | 218.139 | 69.8512 | 7.3 | 0.52 |
| polarC18 | 218.139 | 73.7649 | 7.3 | 0.52 |
| polarC18 | 218.139 | 115.0476 | 7.3 | 0.52 |
| polarC18 | 218.139 | 127.4486 | 7.3 | 0.52 |
| polarC18 | 218.139 | 182.6028 | 7.3 | 0.52 |
| polarC18 | 218.139 | 69.4303 | 7.3 | 0.52 |
| polarC18 | 218.139 | 87.3899 | 7.3 | 0.52 |
| polarC18 | 218.139 | 102.7854 | 7.3 | 0.52 |
| polarC18 | 218.139 | 59.6591 | 7.3 | 0.52 |

| | | | | |
|---|---|---|---|---|
| polarC18 | 218.139 | 50.0914 | 7.3 | 0.52 |
| polarC18 | 218.139 | 119.2124 | 7.3 | 0.52 |
| polarC18 | 218.139 | 151.3463 | 7.3 | 0.52 |
| polarC18 | 218.139 | 54.37 | 7.3 | 0.52 |
| polarC18 | 218.139 | 76.7326 | 7.3 | 0.52 |
| polarC18 | 218.139 | 58.7086 | 7.3 | 0.52 |
| polarC18 | 218.139 | 208.8154 | 7.3 | 0.52 |
| polarC18 | 218.139 | 145.2912 | 7.3 | 0.52 |
| polarC18 | 218.139 | 122.2368 | 7.3 | 0.52 |
| polarC18 | 218.139 | 83.7603 | 7.3 | 0.52 |
| polarC18 | 218.139 | 81.0478 | 7.3 | 0.52 |
| polarC18 | 218.139 | 79.5245 | 7.3 | 0.52 |
| polarC18 | 218.139 | 215.7238 | 7.3 | 0.52 |
| polarC18 | 218.139 | 78.0428 | 7.3 | 0.52 |
| polarC18 | 218.139 | 57.5236 | 7.3 | 0.52 |
| polarC18 | 218.139 | 145.69 | 7.3 | 0.52 |
| polarC18 | 218.139 | 115.9993 | 7.3 | 0.52 |
| polarC18 | 218.139 | 73.4499 | 7.3 | 0.52 |
| polarC18 | 218.139 | 151.3865 | 7.3 | 0.52 |
| polarC18 | 218.139 | 74.2073 | 7.3 | 0.52 |
| polarC18 | 218.139 | 76.7846 | 7.3 | 0.52 |
| polarC18 | 218.139 | 103.1651 | 7.3 | 0.52 |
| polarC18 | 218.139 | 113.1128 | 7.3 | 0.52 |
| polarC18 | 218.139 | 154.5368 | 7.3 | 0.52 |
| polarC18 | 218.139 | 51.2632 | 7.3 | 0.52 |
| polarC18 | 218.139 | 63.9051 | 7.3 | 0.52 |
| polarC18 | 218.139 | 51.2928 | 7.3 | 0.52 |
| polarC18 | 218.139 | 212.5013 | 7.3 | 0.52 |
| polarC18 | 218.139 | 140.916 | 7.3 | 0.52 |
| polarC18 | 218.139 | 179.7184 | 7.3 | 0.52 |
| polarC18 | 218.139 | 69.3503 | 7.3 | 0.52 |
| polarC18 | 218.139 | 99.4804 | 7.3 | 0.52 |
| polarC18 | 218.139 | 160.6741 | 7.3 | 0.52 |
| polarC18 | 218.139 | 122.9304 | 7.3 | 0.52 |
| polarC18 | 218.139 | 49.3732 | 7.3 | 0.52 |
| polarC18 | 218.139 | 66.2689 | 7.3 | 0.52 |
| polarC18 | 218.139 | 105.0338 | 7.3 | 0.52 |
| polarC18 | 218.139 | 95.4085 | 7.3 | 0.52 |
| polarC18 | 218.139 | 146.8522 | 7.3 | 0.52 |
| polarC18 | 218.139 | 81.4668 | 7.3 | 0.52 |
| polarC18 | 218.139 | 232.6984 | 7.3 | 0.52 |
| polarC18 | 218.139 | 127.4106 | 7.3 | 0.52 |

| | | | | |
|---|---|---|---|---|
| polarC18 | 218.139 | 91.6731 | 7.3 | 0.52 |
| polarC18 | 218.139 | 162.1121 | 7.3 | 0.52 |
| polarC18 | 218.139 | 155.5915 | 7.3 | 0.52 |
| polarC18 | 218.139 | 140.8699 | 7.3 | 0.52 |
| polarC18 | 218.139 | 75.4759 | 7.3 | 0.52 |
| polarC18 | 218.139 | 193.5894 | 7.3 | 0.52 |
| polarC18 | 218.139 | 48.9962 | 7.3 | 0.52 |
| polarC18 | 218.139 | 158.3333 | 7.3 | 0.52 |
| polarC18 | 218.139 | 61.6605 | 7.3 | 0.52 |
| polarC18 | 218.139 | 69.2762 | 7.3 | 0.52 |
| polarC18 | 218.139 | 65.2553 | 7.3 | 0.52 |
| polarC18 | 218.139 | 113.4192 | 7.3 | 0.52 |
| polarC18 | 218.139 | 127.9632 | 7.3 | 0.52 |
| polarC18 | 218.139 | 155.9581 | 7.3 | 0.52 |
| polarC18 | 218.139 | 53.8671 | 7.3 | 0.52 |
| polarC18 | 218.139 | 156.8697 | 7.3 | 0.52 |
| polarC18 | 218.139 | 109.9275 | 7.3 | 0.52 |
| polarC18 | 218.139 | 122.2209 | 7.3 | 0.52 |
| polarC18 | 218.139 | 149.0629 | 7.3 | 0.52 |
| polarC18 | 218.139 | 86.5387 | 7.3 | 0.52 |
| polarC18 | 218.139 | 49.1658 | 7.3 | 0.52 |
| polarC18 | 218.139 | 86.1421 | 7.3 | 0.52 |
| polarC18 | 218.139 | 156.3253 | 7.3 | 0.52 |
| polarC18 | 218.139 | 189.6045 | 7.3 | 0.52 |
| polarC18 | 218.139 | 51.0653 | 7.3 | 0.52 |
| polarC18 | 218.139 | 51.9843 | 7.3 | 0.52 |
| polarC18 | 218.139 | 70.2197 | 7.3 | 0.52 |
| polarC18 | 218.139 | 72.1365 | 7.3 | 0.52 |
| polarC18 | 218.139 | 229.0324 | 7.3 | 0.52 |
| polarC18 | 218.139 | 109.9684 | 7.3 | 0.52 |
| polarC18 | 218.139 | 97.8261 | 7.3 | 0.52 |
| polarC18 | 218.139 | 90.3468 | 7.3 | 0.52 |
| polarC18 | 218.139 | 64.1603 | 7.3 | 0.52 |
| polarC18 | 218.139 | 54.522 | 7.3 | 0.52 |
| polarC18 | 218.139 | 55.852 | 7.3 | 0.52 |
| polarC18 | 218.139 | 71.3367 | 7.3 | 0.51 |
| polarC18 | 218.139 | 135.6198 | 7.3 | 0.51 |
| polarC18 | 218.139 | 75.414 | 7.3 | 0.51 |
| polarC18 | 218.139 | 71.6731 | 7.3 | 0.51 |
| polarC18 | 218.139 | 58.165 | 7.3 | 0.51 |
| polarC18 | 218.139 | 75.7957 | 7.3 | 0.51 |
| polarC18 | 218.139 | 96.4455 | 7.3 | 0.51 |

| | | | | |
|---|---|---|---|---|
| polarC18 | 218.139 | 63.3253 | 7.3 | 0.51 |
| polarC18 | 218.139 | 108.8777 | 7.3 | 0.51 |
| polarC18 | 218.139 | 182.3132 | 7.3 | 0.51 |
| polarC18 | 218.139 | 61.4601 | 7.3 | 0.51 |
| polarC18 | 218.139 | 136.4095 | 7.3 | 0.51 |
| polarC18 | 218.139 | 102.7004 | 7.2 | 0.51 |
| polarC18 | 218.139 | 92.7952 | 7.2 | 0.51 |
| polarC18 | 218.139 | 61.4763 | 7.2 | 0.51 |
| polarC18 | 218.139 | 52.202 | 7.2 | 0.51 |
| polarC18 | 218.139 | 89.0857 | 7.2 | 0.51 |
| polarC18 | 218.139 | 101.785 | 7.2 | 0.51 |
| polarC18 | 218.139 | 53.4483 | 7.2 | 0.51 |
| polarC18 | 218.139 | 90.7563 | 7.2 | 0.51 |
| polarC18 | 218.139 | 97.6772 | 7.2 | 0.51 |
| polarC18 | 218.139 | 133.757 | 7.2 | 0.51 |
| polarC18 | 218.139 | 67.0839 | 7.2 | 0.51 |
| polarC18 | 218.139 | 115.5883 | 7.2 | 0.51 |
| polarC18 | 218.139 | 199.7032 | 7.2 | 0.51 |
| polarC18 | 218.139 | 129.6379 | 7.2 | 0.51 |
| polarC18 | 218.139 | 64.4113 | 7.2 | 0.51 |
| polarC18 | 218.139 | 63.786 | 7.2 | 0.51 |
| polarC18 | 218.139 | 98.1791 | 7.2 | 0.51 |
| polarC18 | 218.139 | 151.8665 | 7.2 | 0.51 |
| polarC18 | 218.139 | 169.1422 | 7.2 | 0.51 |
| polarC18 | 218.139 | 69.2429 | 7.2 | 0.51 |
| polarC18 | 218.139 | 51.4242 | 7.2 | 0.51 |
| polarC18 | 218.139 | 172.6286 | 7.2 | 0.51 |
| polarC18 | 218.139 | 58.8881 | 7.2 | 0.51 |
| polarC18 | 218.139 | 57.2647 | 7.2 | 0.51 |
| polarC18 | 218.139 | 67.5049 | 7.2 | 0.51 |
| polarC18 | 218.139 | 49.3708 | 7.2 | 0.51 |
| polarC18 | 218.139 | 58.3598 | 7.2 | 0.51 |
| polarC18 | 218.139 | 242.2092 | 7.2 | 0.51 |
| polarC18 | 218.139 | 85.6334 | 7.2 | 0.51 |
| polarC18 | 218.139 | 52.0628 | 7.2 | 0.51 |
| polarC18 | 218.139 | 116.1483 | 7.2 | 0.51 |
| polarC18 | 218.139 | 78.2896 | 7.2 | 0.51 |
| polarC18 | 218.139 | 72.5741 | 7.2 | 0.51 |
| polarC18 | 218.139 | 80.2454 | 7.2 | 0.51 |
| polarC18 | 218.139 | 66.7781 | 7.2 | 0.51 |
| polarC18 | 218.139 | 91.1525 | 7.2 | 0.51 |
| polarC18 | 218.139 | 66.4034 | 7.2 | 0.51 |

| | | | | |
|---|---|---|---|---|
| polarC18 | 218.139 | 81.2075 | 7.2 | 0.51 |
| polarC18 | 218.139 | 60.7824 | 7.2 | 0.51 |
| polarC18 | 218.139 | 57.3451 | 7.2 | 0.51 |
| polarC18 | 218.139 | 154.4624 | 7.2 | 0.51 |
| polarC18 | 218.139 | 77.2465 | 7.2 | 0.51 |
| polarC18 | 218.139 | 121.1687 | 7.2 | 0.51 |
| polarC18 | 218.139 | 59.7171 | 7.2 | 0.51 |
| polarC18 | 218.139 | 88.7638 | 7.2 | 0.51 |
| polarC18 | 218.139 | 65.9179 | 7.2 | 0.51 |
| polarC18 | 218.139 | 119.4731 | 7.2 | 0.51 |
| polarC18 | 218.139 | 90.1955 | 7.2 | 0.51 |
| polarC18 | 218.139 | 77.4139 | 7.2 | 0.51 |
| polarC18 | 218.139 | 62.1104 | 7.2 | 0.51 |
| polarC18 | 218.139 | 51.1165 | 7.2 | 0.51 |
| polarC18 | 218.139 | 57.8307 | 7.2 | 0.51 |
| polarC18 | 218.139 | 83.0434 | 7.2 | 0.51 |
| polarC18 | 218.139 | 167.3138 | 7.2 | 0.51 |
| polarC18 | 218.139 | 64.8533 | 7.2 | 0.51 |
| polarC18 | 218.139 | 80.345 | 7.2 | 0.51 |
| polarC18 | 218.139 | 65.0784 | 7.2 | 0.51 |
| polarC18 | 218.139 | 140.4161 | 7.2 | 0.51 |
| polarC18 | 218.139 | 99.8881 | 7.2 | 0.51 |
| polarC18 | 218.139 | 60.6222 | 7.2 | 0.51 |
| polarC18 | 218.139 | 54.8152 | 7.2 | 0.51 |
| polarC18 | 218.139 | 129.138 | 7.2 | 0.51 |
| polarC18 | 218.139 | 52.0712 | 7.2 | 0.51 |
| polarC18 | 218.139 | 74.3223 | 7.2 | 0.51 |
| polarC18 | 218.139 | 80.6086 | 7.2 | 0.51 |
| polarC18 | 218.139 | 228.4294 | 7.2 | 0.51 |
| polarC18 | 218.139 | 166.5892 | 7.2 | 0.51 |
| polarC18 | 218.139 | 64.2547 | 7.2 | 0.51 |
| polarC18 | 218.139 | 51.3404 | 7.2 | 0.51 |
| polarC18 | 218.139 | 211.4487 | 7.2 | 0.51 |
| polarC18 | 218.139 | 171.4587 | 7.2 | 0.51 |
| polarC18 | 218.139 | 53.6544 | 7.2 | 0.51 |
| polarC18 | 218.139 | 99.2004 | 7.2 | 0.51 |
| polarC18 | 218.139 | 78.4123 | 7.2 | 0.51 |
| polarC18 | 218.139 | 103.0786 | 7.2 | 0.51 |
| polarC18 | 218.139 | 79.8409 | 7.2 | 0.51 |
| polarC18 | 218.139 | 77.6342 | 7.2 | 0.51 |
| polarC18 | 218.139 | 151.9814 | 7.2 | 0.51 |
| polarC18 | 218.139 | 95.8003 | 7.2 | 0.51 |

| | | | | |
|---|---|---|---|---|
| polarC18 | 218.139 | 99.1396 | 7.2 | 0.51 |
| polarC18 | 218.139 | 70.7733 | 7.2 | 0.51 |
| polarC18 | 218.139 | 90.1172 | 7.2 | 0.51 |
| polarC18 | 218.139 | 50.4928 | 7.2 | 0.51 |
| polarC18 | 218.139 | 73.2881 | 7.2 | 0.51 |
| polarC18 | 218.139 | 141.0798 | 7.2 | 0.51 |
| polarC18 | 218.139 | 85.9913 | 7.2 | 0.51 |
| polarC18 | 218.139 | 55.6993 | 7.2 | 0.51 |
| polarC18 | 218.139 | 50.3467 | 7.2 | 0.51 |
| polarC18 | 218.139 | 104.3419 | 7.2 | 0.51 |
| polarC18 | 218.139 | 92.4286 | 7.2 | 0.51 |
| polarC18 | 218.139 | 242.9911 | 7.2 | 0.51 |
| polarC18 | 218.139 | 180.2343 | 7.2 | 0.51 |
| polarC18 | 218.139 | 91.1718 | 7.2 | 0.51 |
| polarC18 | 218.139 | 144.7388 | 7.2 | 0.51 |
| polarC18 | 218.139 | 62.0876 | 7.2 | 0.51 |
| polarC18 | 218.139 | 173.7395 | 7.2 | 0.51 |
| polarC18 | 218.139 | 101.9841 | 7.2 | 0.51 |
| polarC18 | 218.139 | 66.3257 | 7.2 | 0.51 |
| polarC18 | 218.139 | 74.1793 | 7.2 | 0.51 |
| polarC18 | 218.139 | 182.1906 | 7.2 | 0.51 |
| polarC18 | 218.139 | 113.6537 | 7.2 | 0.51 |
| polarC18 | 218.139 | 115.1467 | 7.2 | 0.51 |
| polarC18 | 218.139 | 76.7391 | 7.2 | 0.51 |
| polarC18 | 218.139 | 59.8121 | 7.2 | 0.51 |
| polarC18 | 218.139 | 90.6051 | 7.2 | 0.51 |
| polarC18 | 218.139 | 148.1159 | 7.2 | 0.51 |
| polarC18 | 218.139 | 190.9465 | 7.2 | 0.51 |
| polarC18 | 218.139 | 127.8234 | 7.2 | 0.51 |
| polarC18 | 218.139 | 65.5844 | 7.2 | 0.51 |
| polarC18 | 218.139 | 234.6771 | 7.2 | 0.51 |
| polarC18 | 218.139 | 81.0629 | 7.2 | 0.51 |
| polarC18 | 218.139 | 91.825 | 7.2 | 0.51 |
| polarC18 | 218.139 | 74.883 | 7.2 | 0.51 |
| polarC18 | 218.139 | 51.8011 | 7.2 | 0.51 |
| polarC18 | 218.139 | 227.6314 | 7.2 | 0.51 |
| polarC18 | 218.139 | 51.3141 | 7.2 | 0.51 |
| polarC18 | 218.139 | 112.5684 | 7.2 | 0.51 |
| polarC18 | 218.139 | 66.4076 | 7.2 | 0.51 |
| polarC18 | 218.139 | 115.7369 | 7.2 | 0.51 |
| polarC18 | 218.139 | 49.3359 | 7.2 | 0.51 |
| polarC18 | 218.139 | 59.32 | 7.2 | 0.51 |

| | | | | |
|---|---|---|---|---|
| polarC18 | 218.139 | 167.063 | 7.2 | 0.51 |
| polarC18 | 218.139 | 65.4829 | 7.2 | 0.51 |
| polarC18 | 218.139 | 123.2192 | 7.2 | 0.51 |
| polarC18 | 218.139 | 82.1777 | 7.2 | 0.51 |
| polarC18 | 218.139 | 51.287 | 7.2 | 0.51 |
| polarC18 | 218.139 | 105.3664 | 7.2 | 0.51 |
| polarC18 | 218.139 | 59.0363 | 7.2 | 0.51 |
| polarC18 | 218.139 | 140.8064 | 7.2 | 0.51 |
| polarC18 | 218.139 | 157.2434 | 7.2 | 0.51 |
| polarC18 | 218.139 | 67.2549 | 7.2 | 0.51 |
| polarC18 | 218.139 | 96.9375 | 7.2 | 0.51 |
| polarC18 | 218.139 | 52.5842 | 7.2 | 0.51 |
| polarC18 | 218.139 | 89.9009 | 7.2 | 0.51 |
| polarC18 | 218.139 | 62.2022 | 7.2 | 0.51 |
| polarC18 | 218.139 | 51.6341 | 7.2 | 0.51 |
| polarC18 | 218.139 | 110.7156 | 7.2 | 0.51 |
| polarC18 | 218.139 | 70.7612 | 7.2 | 0.51 |
| polarC18 | 218.139 | 49.7963 | 7.2 | 0.51 |
| polarC18 | 218.139 | 166.4513 | 7.1 | 0.51 |
| polarC18 | 218.139 | 69.0926 | 7.1 | 0.51 |
| polarC18 | 218.139 | 109.5927 | 7.1 | 0.51 |
| polarC18 | 218.139 | 66.2659 | 7.1 | 0.51 |
| polarC18 | 218.139 | 50.609 | 7.1 | 0.51 |
| polarC18 | 218.139 | 162.8244 | 7.1 | 0.51 |
| polarC18 | 218.139 | 144.4151 | 7.1 | 0.51 |
| polarC18 | 218.139 | 226.0648 | 7.1 | 0.51 |
| polarC18 | 218.139 | 86.2493 | 7.1 | 0.51 |
| polarC18 | 218.139 | 64.4269 | 7.1 | 0.51 |
| polarC18 | 218.139 | 60.2323 | 7.1 | 0.51 |
| polarC18 | 218.139 | 108.4145 | 7.1 | 0.51 |
| polarC18 | 218.139 | 65.1053 | 7.1 | 0.51 |
| polarC18 | 218.139 | 141.0067 | 7.1 | 0.51 |
| polarC18 | 218.139 | 57.8446 | 7.1 | 0.51 |
| polarC18 | 218.139 | 165.1217 | 7.1 | 0.51 |
| polarC18 | 218.139 | 114.1952 | 7.1 | 0.51 |
| polarC18 | 218.139 | 67.1963 | 7.1 | 0.51 |
| polarC18 | 218.139 | 49.2478 | 7.1 | 0.51 |
| polarC18 | 218.139 | 139.097 | 7.1 | 0.51 |
| polarC18 | 218.139 | 52.3134 | 7.1 | 0.51 |
| polarC18 | 218.139 | 144.7915 | 7.1 | 0.51 |
| polarC18 | 218.139 | 79.9522 | 7.1 | 0.51 |
| polarC18 | 218.139 | 188.0361 | 7.1 | 0.51 |

| | | | | |
|---|---|---|---|---|
| polarC18 | 218.139 | 145.4108 | 7.1 | 0.51 |
| polarC18 | 218.139 | 55.9574 | 7.1 | 0.51 |
| polarC18 | 218.139 | 86.1641 | 7.1 | 0.51 |
| polarC18 | 218.139 | 104.775 | 7.1 | 0.51 |
| polarC18 | 218.139 | 86.8463 | 7.1 | 0.51 |
| polarC18 | 218.139 | 58.7879 | 7.1 | 0.51 |
| polarC18 | 218.139 | 205.0874 | 7.1 | 0.51 |
| polarC18 | 218.139 | 53.1517 | 7.1 | 0.51 |
| polarC18 | 218.139 | 117.6684 | 7.1 | 0.51 |
| polarC18 | 218.139 | 195.617 | 7.1 | 0.51 |
| polarC18 | 218.139 | 58.0026 | 7.1 | 0.51 |
| polarC18 | 218.139 | 68.173 | 7.1 | 0.51 |
| polarC18 | 218.139 | 52.7535 | 7.1 | 0.51 |
| polarC18 | 218.139 | 89.6585 | 7.1 | 0.51 |
| polarC18 | 218.139 | 53.9516 | 7.1 | 0.51 |
| polarC18 | 218.139 | 77.9204 | 7.1 | 0.51 |
| polarC18 | 218.139 | 235.1447 | 7.1 | 0.51 |
| polarC18 | 218.139 | 97.0771 | 7.1 | 0.51 |
| polarC18 | 218.139 | 172.6047 | 7.1 | 0.51 |
| polarC18 | 218.139 | 66.8885 | 7.1 | 0.51 |
| polarC18 | 218.139 | 88.1421 | 7.1 | 0.51 |
| polarC18 | 218.139 | 125.5173 | 7.1 | 0.51 |
| polarC18 | 218.139 | 122.4117 | 7.1 | 0.51 |
| polarC18 | 218.139 | 87.7486 | 7.1 | 0.51 |
| polarC18 | 218.139 | 65.5499 | 7.1 | 0.51 |
| polarC18 | 218.139 | 137.1711 | 7.1 | 0.51 |
| polarC18 | 218.139 | 52.2856 | 7.1 | 0.5 |
| polarC18 | 218.139 | 88.2907 | 7.1 | 0.5 |
| polarC18 | 218.139 | 123.6265 | 7.1 | 0.5 |
| polarC18 | 218.139 | 60.6686 | 7.1 | 0.5 |
| polarC18 | 218.139 | 57.9273 | 7.1 | 0.5 |
| polarC18 | 218.139 | 51.5366 | 7.1 | 0.5 |
| polarC18 | 218.139 | 181.1279 | 7.1 | 0.5 |
| polarC18 | 218.139 | 147.4138 | 7.1 | 0.5 |
| polarC18 | 218.139 | 49.5341 | 7.1 | 0.5 |
| polarC18 | 218.139 | 89.7246 | 7.1 | 0.5 |
| polarC18 | 218.139 | 71.7783 | 7.1 | 0.5 |
| polarC18 | 218.139 | 75.6353 | 7.1 | 0.5 |
| polarC18 | 218.139 | 95.0458 | 7.1 | 0.5 |
| polarC18 | 218.139 | 126.7997 | 7.1 | 0.5 |
| polarC18 | 218.139 | 178.405 | 7.1 | 0.5 |
| polarC18 | 218.139 | 58.6944 | 7.1 | 0.5 |

| | | | | |
|---|---|---|---|---|
| polarC18 | 218.139 | 148.6392 | 7.1 | 0.5 |
| polarC18 | 218.139 | 104.9642 | 7.1 | 0.5 |
| polarC18 | 218.139 | 131.067 | 7.1 | 0.5 |
| polarC18 | 218.139 | 50.8013 | 7.1 | 0.5 |
| polarC18 | 218.139 | 163.4128 | 7.1 | 0.5 |
| polarC18 | 218.139 | 113.8761 | 7.1 | 0.5 |
| polarC18 | 218.139 | 55.9192 | 7.1 | 0.5 |
| polarC18 | 218.139 | 48.7821 | 7.1 | 0.5 |
| polarC18 | 218.139 | 85.8513 | 7.1 | 0.5 |
| polarC18 | 218.139 | 211.8758 | 7.1 | 0.5 |
| polarC18 | 218.139 | 81.3136 | 7.1 | 0.5 |
| polarC18 | 218.139 | 102.6132 | 7.1 | 0.5 |
| polarC18 | 218.139 | 88.7333 | 7.1 | 0.5 |
| polarC18 | 218.139 | 65.2303 | 7.1 | 0.5 |
| polarC18 | 218.139 | 82.9139 | 7.1 | 0.5 |
| polarC18 | 218.139 | 187.289 | 7.1 | 0.5 |
| polarC18 | 218.139 | 62.304 | 7.1 | 0.5 |
| polarC18 | 218.139 | 172.2697 | 7.1 | 0.5 |
| polarC18 | 218.139 | 88.6671 | 7.1 | 0.5 |
| polarC18 | 218.139 | 82.568 | 7.1 | 0.5 |
| polarC18 | 218.139 | 90.9865 | 7.1 | 0.5 |
| polarC18 | 218.139 | 50.916 | 7.1 | 0.5 |
| polarC18 | 218.139 | 166.01 | 7.1 | 0.5 |
| polarC18 | 218.139 | 51.6396 | 7.1 | 0.5 |
| polarC18 | 218.139 | 120.9246 | 7.1 | 0.5 |
| polarC18 | 218.139 | 128.3771 | 7.1 | 0.5 |
| polarC18 | 218.139 | 87.3784 | 7.1 | 0.5 |
| polarC18 | 218.139 | 143.356 | 7.1 | 0.5 |
| polarC18 | 218.139 | 206.6299 | 7.1 | 0.5 |
| polarC18 | 218.139 | 51.2044 | 7.1 | 0.5 |
| polarC18 | 218.139 | 137.0099 | 7.1 | 0.5 |
| polarC18 | 218.139 | 82.2586 | 7.1 | 0.5 |
| polarC18 | 218.139 | 85.441 | 7.1 | 0.5 |
| polarC18 | 218.139 | 58.9637 | 7.1 | 0.5 |
| polarC18 | 218.139 | 63.6897 | 7.1 | 0.5 |
| polarC18 | 218.139 | 209.3907 | 7.1 | 0.5 |
| polarC18 | 218.139 | 125.2167 | 7.1 | 0.5 |
| polarC18 | 218.139 | 190.7658 | 7.1 | 0.5 |
| polarC18 | 218.139 | 222.9906 | 7.1 | 0.5 |
| polarC18 | 218.139 | 106.3805 | 7.1 | 0.5 |
| polarC18 | 218.139 | 72.5194 | 7.1 | 0.5 |
| polarC18 | 218.139 | 58.8968 | 7.1 | 0.5 |

| | | | | |
|---|---|---|---|---|
| polarC18 | 218.139 | 52.5027 | 7.1 | 0.5 |
| polarC18 | 218.139 | 109.5558 | 7.1 | 0.5 |
| polarC18 | 218.139 | 69.3545 | 7.1 | 0.5 |
| polarC18 | 218.139 | 116.0926 | 7.1 | 0.5 |
| polarC18 | 218.139 | 74.3908 | 7.1 | 0.5 |
| polarC18 | 218.139 | 114.6598 | 7.1 | 0.5 |
| polarC18 | 218.139 | 176.0574 | 7.1 | 0.5 |
| polarC18 | 218.139 | 49.4145 | 7.1 | 0.5 |
| polarC18 | 218.139 | 145.1768 | 7.1 | 0.5 |
| polarC18 | 218.139 | 71.1538 | 7.1 | 0.5 |
| polarC18 | 218.139 | 154.4106 | 7.1 | 0.5 |
| polarC18 | 218.139 | 48.8654 | 7.1 | 0.5 |
| polarC18 | 218.139 | 144.3814 | 7.1 | 0.5 |
| polarC18 | 218.139 | 72.4383 | 7.1 | 0.5 |
| polarC18 | 218.139 | 116.0309 | 7.1 | 0.5 |
| polarC18 | 218.139 | 83.7051 | 7.1 | 0.5 |
| polarC18 | 218.139 | 161.3176 | 7.1 | 0.5 |
| polarC18 | 218.139 | 75.4468 | 7.1 | 0.5 |
| polarC18 | 218.139 | 97.5778 | 7.1 | 0.5 |
| polarC18 | 218.139 | 118.4011 | 7.1 | 0.5 |
| polarC18 | 218.139 | 202.8325 | 7.1 | 0.5 |
| polarC18 | 218.139 | 169.9372 | 7.1 | 0.5 |
| polarC18 | 218.139 | 111.926 | 7.1 | 0.5 |
| polarC18 | 218.139 | 77.5542 | 7.1 | 0.5 |
| polarC18 | 218.139 | 57.451 | 7.1 | 0.5 |
| polarC18 | 218.139 | 57.8489 | 7.1 | 0.5 |
| polarC18 | 218.139 | 68.3688 | 7.1 | 0.5 |
| polarC18 | 218.139 | 68.5373 | 7.1 | 0.5 |
| polarC18 | 218.139 | 56.4619 | 7.1 | 0.5 |
| polarC18 | 218.139 | 187.3313 | 7.1 | 0.5 |
| polarC18 | 218.139 | 126.8904 | 7.1 | 0.5 |
| polarC18 | 218.139 | 130.0368 | 7.1 | 0.5 |
| polarC18 | 218.139 | 98.4984 | 7.1 | 0.5 |
| polarC18 | 218.139 | 83.5522 | 7.1 | 0.5 |
| polarC18 | 218.139 | 113.3071 | 7.1 | 0.5 |
| polarC18 | 218.139 | 63.995 | 7.1 | 0.5 |
| polarC18 | 218.139 | 122.7295 | 7.1 | 0.5 |
| polarC18 | 218.139 | 93.4006 | 7.1 | 0.5 |
| polarC18 | 218.139 | 51.3325 | 7.1 | 0.5 |
| polarC18 | 218.139 | 115.2968 | 7.1 | 0.5 |
| polarC18 | 218.139 | 76.1909 | 7.1 | 0.5 |
| polarC18 | 218.139 | 113.7447 | 7.1 | 0.5 |

| | | | | |
|---|---|---|---|---|
| polarC18 | 218.139 | 77.735 | 7.1 | 0.5 |
| polarC18 | 218.139 | 50.3823 | 7.1 | 0.5 |
| polarC18 | 218.139 | 50.4091 | 7.1 | 0.5 |
| polarC18 | 218.139 | 58.9358 | 7.1 | 0.5 |
| polarC18 | 218.139 | 133.0589 | 7.1 | 0.5 |
| polarC18 | 218.139 | 102.0052 | 7.1 | 0.5 |
| polarC18 | 218.139 | 171.0465 | 7.1 | 0.5 |
| polarC18 | 218.139 | 150.5971 | 7.1 | 0.5 |
| polarC18 | 218.139 | 123.7419 | 7.1 | 0.5 |
| polarC18 | 218.139 | 49.5581 | 7.1 | 0.5 |
| polarC18 | 218.139 | 106.2477 | 7.1 | 0.5 |
| polarC18 | 218.139 | 148.5537 | 7.1 | 0.5 |
| polarC18 | 218.139 | 52.3091 | 7.1 | 0.5 |
| polarC18 | 218.139 | 83.1097 | 7 | 0.5 |
| polarC18 | 218.139 | 105.2307 | 7 | 0.5 |
| polarC18 | 218.139 | 69.2476 | 7 | 0.5 |
| polarC18 | 218.139 | 55.3402 | 7 | 0.5 |
| polarC18 | 218.139 | 105.3489 | 7 | 0.5 |
| polarC18 | 218.139 | 55.5961 | 7 | 0.5 |
| polarC18 | 218.139 | 73.0298 | 7 | 0.5 |
| polarC18 | 218.139 | 162.1712 | 7 | 0.5 |
| polarC18 | 218.139 | 145.2752 | 7 | 0.5 |
| polarC18 | 218.139 | 237.3956 | 7 | 0.5 |
| polarC18 | 218.139 | 196.8768 | 7 | 0.5 |
| polarC18 | 218.139 | 76.5623 | 7 | 0.5 |
| polarC18 | 218.139 | 60.0451 | 7 | 0.5 |
| polarC18 | 218.139 | 157.4107 | 7 | 0.5 |
| polarC18 | 218.139 | 87.0703 | 7 | 0.5 |
| polarC18 | 218.139 | 187.1956 | 7 | 0.5 |
| polarC18 | 218.139 | 48.797 | 7 | 0.5 |
| polarC18 | 218.139 | 105.8702 | 7 | 0.5 |
| polarC18 | 218.139 | 68.1125 | 7 | 0.5 |
| polarC18 | 218.139 | 79.0677 | 7 | 0.5 |
| polarC18 | 218.139 | 63.3413 | 7 | 0.5 |
| polarC18 | 218.139 | 176.8267 | 7 | 0.5 |
| polarC18 | 218.139 | 79.8909 | 7 | 0.5 |
| polarC18 | 218.139 | 52.1954 | 7 | 0.5 |
| polarC18 | 218.139 | 159.4306 | 7 | 0.5 |
| polarC18 | 218.139 | 77.5271 | 7 | 0.5 |
| polarC18 | 218.139 | 69.9153 | 7 | 0.5 |
| polarC18 | 218.139 | 60.3035 | 7 | 0.5 |
| polarC18 | 218.139 | 169.6008 | 7 | 0.5 |

| | | | | |
|---|---|---|---|---|
| polarC18 | 218.139 | 105.0102 | 7 | 0.5 |
| polarC18 | 218.139 | 96.8069 | 7 | 0.5 |
| polarC18 | 218.139 | 130.6378 | 7 | 0.5 |
| polarC18 | 218.139 | 190.7431 | 7 | 0.5 |
| polarC18 | 218.139 | 121.128 | 7 | 0.5 |
| polarC18 | 218.139 | 81.6301 | 7 | 0.5 |
| polarC18 | 218.139 | 131.6898 | 7 | 0.5 |
| polarC18 | 218.139 | 105.9248 | 7 | 0.5 |
| polarC18 | 218.139 | 52.3843 | 7 | 0.5 |
| polarC18 | 218.139 | 50.3596 | 7 | 0.5 |
| polarC18 | 218.139 | 196.9866 | 7 | 0.5 |
| polarC18 | 218.139 | 143.7398 | 7 | 0.5 |
| polarC18 | 218.139 | 52.1158 | 7 | 0.5 |
| polarC18 | 218.139 | 56.1281 | 7 | 0.5 |
| polarC18 | 218.139 | 128.6003 | 7 | 0.5 |
| polarC18 | 218.139 | 49.9165 | 7 | 0.5 |
| polarC18 | 218.139 | 62.9971 | 7 | 0.5 |
| polarC18 | 218.139 | 72.4229 | 7 | 0.5 |
| polarC18 | 218.139 | 53.0222 | 7 | 0.5 |
| polarC18 | 218.139 | 67.7596 | 7 | 0.5 |
| polarC18 | 218.139 | 100.5934 | 7 | 0.5 |
| polarC18 | 218.139 | 69.1228 | 7 | 0.5 |
| polarC18 | 218.139 | 113.5879 | 7 | 0.5 |
| polarC18 | 218.139 | 82.7591 | 7 | 0.5 |
| polarC18 | 218.139 | 220.0683 | 7 | 0.5 |
| polarC18 | 218.139 | 60.937 | 7 | 0.5 |
| polarC18 | 218.139 | 147.891 | 7 | 0.5 |
| polarC18 | 218.139 | 85.4186 | 7 | 0.5 |
| polarC18 | 218.139 | 119.2823 | 7 | 0.5 |
| polarC18 | 218.139 | 107.4762 | 7 | 0.5 |
| polarC18 | 218.139 | 116.806 | 7 | 0.5 |
| polarC18 | 218.139 | 50.4245 | 7 | 0.5 |
| polarC18 | 218.139 | 59.4625 | 7 | 0.5 |
| polarC18 | 218.139 | 212.9544 | 7 | 0.5 |
| polarC18 | 218.139 | 49.5187 | 7 | 0.5 |
| polarC18 | 218.139 | 96.4683 | 7 | 0.5 |
| polarC18 | 218.139 | 55.7083 | 7 | 0.5 |
| polarC18 | 218.139 | 140.6736 | 7 | 0.5 |
| polarC18 | 218.139 | 224.6214 | 7 | 0.5 |
| polarC18 | 218.139 | 103.5483 | 7 | 0.5 |
| polarC18 | 218.139 | 95.2781 | 7 | 0.5 |
| polarC18 | 218.139 | 52.1887 | 7 | 0.5 |

| | | | | |
|---|---|---|---|---|
| polarC18 | 218.139 | 57.7207 | 7 | 0.5 |
| polarC18 | 218.139 | 202.06 | 7 | 0.5 |
| polarC18 | 218.139 | 193.3455 | 7 | 0.5 |
| polarC18 | 218.139 | 75.0506 | 7 | 0.5 |
| polarC18 | 218.139 | 107.2105 | 7 | 0.5 |
| polarC18 | 218.139 | 78.9333 | 7 | 0.5 |
| polarC18 | 218.139 | 78.1292 | 7 | 0.5 |
| polarC18 | 218.139 | 78.2182 | 7 | 0.5 |
| polarC18 | 218.139 | 134.815 | 7 | 0.5 |
| polarC18 | 218.139 | 55.934 | 7 | 0.5 |
| polarC18 | 218.139 | 51.0067 | 7 | 0.5 |
| polarC18 | 218.139 | 179.6494 | 7 | 0.5 |
| polarC18 | 218.139 | 61.4892 | 7 | 0.5 |
| polarC18 | 218.139 | 195.9851 | 7 | 0.5 |
| polarC18 | 218.139 | 51.9062 | 7 | 0.5 |
| polarC18 | 218.139 | 115.9458 | 7 | 0.5 |
| polarC18 | 218.139 | 118.6056 | 7 | 0.5 |
| polarC18 | 218.139 | 56.9102 | 7 | 0.5 |
| polarC18 | 218.139 | 89.1924 | 7 | 0.5 |
| polarC18 | 218.139 | 103.4855 | 7 | 0.5 |
| polarC18 | 218.139 | 138.3385 | 7 | 0.5 |
| polarC18 | 218.139 | 74.7902 | 7 | 0.5 |
| polarC18 | 218.139 | 91.4437 | 7 | 0.5 |
| polarC18 | 218.139 | 143.9444 | 7 | 0.5 |
| polarC18 | 218.139 | 66.1041 | 7 | 0.5 |
| polarC18 | 218.139 | 79.286 | 7 | 0.5 |
| polarC18 | 218.139 | 121.8632 | 7 | 0.5 |
| polarC18 | 218.139 | 155.8945 | 7 | 0.5 |
| polarC18 | 218.139 | 83.6817 | 7 | 0.5 |
| polarC18 | 218.139 | 155.2033 | 7 | 0.5 |
| polarC18 | 218.139 | 52.5577 | 7 | 0.5 |
| polarC18 | 218.139 | 74.5329 | 7 | 0.5 |
| polarC18 | 218.139 | 145.6142 | 7 | 0.5 |
| polarC18 | 218.139 | 107.1467 | 7 | 0.5 |
| polarC18 | 218.139 | 86.0205 | 7 | 0.5 |
| polarC18 | 218.139 | 127.6481 | 7 | 0.5 |
| polarC18 | 218.139 | 141.1552 | 7 | 0.5 |
| polarC18 | 218.139 | 53.1674 | 7 | 0.5 |
| polarC18 | 218.139 | 122.8961 | 7 | 0.5 |
| polarC18 | 218.139 | 233.7534 | 7 | 0.5 |
| polarC18 | 218.139 | 66.5917 | 7 | 0.5 |
| polarC18 | 218.139 | 137.664 | 7 | 0.5 |

| | | | | |
|---|---|---|---|---|
| polarC18 | 218.139 | 108.8654 | 7 | 0.5 |
| polarC18 | 218.139 | 49.7095 | 7 | 0.5 |
| polarC18 | 218.139 | 50.8809 | 7 | 0.5 |
| polarC18 | 218.139 | 51.1342 | 7 | 0.5 |
| polarC18 | 218.139 | 179.8478 | 7 | 0.5 |
| polarC18 | 218.139 | 107.1273 | 7 | 0.5 |
| polarC18 | 218.139 | 102.8337 | 7 | 0.5 |
| polarC18 | 218.139 | 76.861 | 7 | 0.5 |
| polarC18 | 218.139 | 60.19 | 7 | 0.5 |
| polarC18 | 218.139 | 70.5992 | 7 | 0.5 |
| polarC18 | 218.139 | 129.9013 | 7 | 0.5 |
| polarC18 | 218.139 | 65.22 | 7 | 0.5 |
| polarC18 | 218.139 | 76.2708 | 7 | 0.5 |
| polarC18 | 218.139 | 58.9571 | 7 | 0.5 |
| polarC18 | 218.139 | 88.6296 | 7 | 0.5 |
| polarC18 | 218.139 | 138.5528 | 7 | 0.49 |
| polarC18 | 218.139 | 69.9027 | 7 | 0.49 |
| polarC18 | 218.139 | 72.3202 | 7 | 0.49 |
| polarC18 | 218.139 | 175.7659 | 7 | 0.49 |
| polarC18 | 218.139 | 155.0475 | 7 | 0.49 |
| polarC18 | 218.139 | 133.701 | 7 | 0.49 |
| polarC18 | 218.139 | 67.1027 | 7 | 0.49 |
| polarC18 | 218.139 | 52.2412 | 7 | 0.49 |
| polarC18 | 218.139 | 50.1809 | 7 | 0.49 |
| polarC18 | 218.139 | 65.286 | 7 | 0.49 |
| polarC18 | 218.139 | 72.6409 | 7 | 0.49 |
| polarC18 | 218.139 | 48.7688 | 7 | 0.49 |
| polarC18 | 218.139 | 123.9397 | 7 | 0.49 |
| polarC18 | 218.139 | 96.7975 | 7 | 0.49 |
| polarC18 | 218.139 | 54.6038 | 7 | 0.49 |
| polarC18 | 218.139 | 113.2302 | 7 | 0.49 |
| polarC18 | 218.139 | 49.0266 | 7 | 0.49 |
| polarC18 | 218.139 | 83.9337 | 7 | 0.49 |
| polarC18 | 218.139 | 137.8676 | 7 | 0.49 |
| polarC18 | 218.139 | 66.3063 | 7 | 0.49 |
| polarC18 | 218.139 | 49.547 | 7 | 0.49 |
| polarC18 | 218.139 | 77.3095 | 7 | 0.49 |
| polarC18 | 218.139 | 69.2592 | 7 | 0.49 |
| polarC18 | 218.139 | 63.6467 | 7 | 0.49 |
| polarC18 | 218.139 | 51.745 | 7 | 0.49 |
| polarC18 | 218.139 | 122.1544 | 7 | 0.49 |
| polarC18 | 218.139 | 64.1688 | 7 | 0.49 |

| | | | | |
|---|---|---|---|---|
| polarC18 | 218.139 | 74.9494 | 7 | 0.49 |
| polarC18 | 218.139 | 70.1677 | 7 | 0.49 |
| polarC18 | 218.139 | 93.8073 | 7 | 0.49 |
| polarC18 | 218.139 | 71.1295 | 7 | 0.49 |
| polarC18 | 218.139 | 67.1751 | 7 | 0.49 |
| polarC18 | 218.139 | 52.5148 | 7 | 0.49 |
| polarC18 | 218.139 | 79.6048 | 7 | 0.49 |
| polarC18 | 218.139 | 76.8793 | 7 | 0.49 |
| polarC18 | 218.139 | 61.7664 | 7 | 0.49 |
| polarC18 | 218.139 | 136.5679 | 7 | 0.49 |
| polarC18 | 218.139 | 92.8686 | 7 | 0.49 |
| polarC18 | 218.139 | 80.8852 | 7 | 0.49 |
| polarC18 | 218.139 | 143.9741 | 7 | 0.49 |
| polarC18 | 218.139 | 202.8833 | 7 | 0.49 |
| polarC18 | 218.139 | 189.3233 | 7 | 0.49 |
| polarC18 | 218.139 | 125.3949 | 7 | 0.49 |
| polarC18 | 218.139 | 71.2895 | 7 | 0.49 |
| polarC18 | 218.139 | 72.1992 | 7 | 0.49 |
| polarC18 | 218.139 | 78.8406 | 7 | 0.49 |
| polarC18 | 218.139 | 131.6244 | 7 | 0.49 |
| polarC18 | 218.139 | 71.2144 | 7 | 0.49 |
| polarC18 | 218.139 | 169.3248 | 7 | 0.49 |
| polarC18 | 218.139 | 120.8518 | 6.9 | 0.49 |
| polarC18 | 218.139 | 119.8184 | 6.9 | 0.49 |
| polarC18 | 218.139 | 79.9239 | 6.9 | 0.49 |
| polarC18 | 218.139 | 50.0634 | 6.9 | 0.49 |
| polarC18 | 218.139 | 59.3613 | 6.9 | 0.49 |
| polarC18 | 218.139 | 72.692 | 6.9 | 0.49 |
| polarC18 | 218.139 | 74.755 | 6.9 | 0.49 |
| polarC18 | 218.139 | 142.3443 | 6.9 | 0.49 |
| polarC18 | 218.139 | 82.6491 | 6.9 | 0.49 |
| polarC18 | 218.139 | 80.2777 | 6.9 | 0.49 |
| polarC18 | 218.139 | 81.6261 | 6.9 | 0.49 |
| polarC18 | 218.139 | 145.8311 | 6.9 | 0.49 |
| polarC18 | 218.139 | 117.9401 | 6.9 | 0.49 |
| polarC18 | 218.139 | 91.3971 | 6.9 | 0.49 |
| polarC18 | 218.139 | 51.2021 | 6.9 | 0.49 |
| polarC18 | 218.139 | 110.4966 | 6.9 | 0.49 |
| polarC18 | 218.139 | 84.4218 | 6.9 | 0.49 |
| polarC18 | 218.139 | 111.6578 | 6.9 | 0.49 |
| polarC18 | 218.139 | 75.8667 | 6.9 | 0.49 |
| polarC18 | 218.139 | 88.6948 | 6.9 | 0.49 |

| | | | | |
|---|---|---|---|---|
| polarC18 | 218.139 | 71.2198 | 6.9 | 0.49 |
| polarC18 | 218.139 | 51.8726 | 6.9 | 0.49 |
| polarC18 | 218.139 | 54.9653 | 6.9 | 0.49 |
| polarC18 | 218.139 | 208.7773 | 6.9 | 0.49 |
| polarC18 | 218.139 | 55.785 | 6.9 | 0.49 |
| polarC18 | 218.139 | 131.5503 | 6.9 | 0.49 |
| polarC18 | 218.139 | 63.9792 | 6.9 | 0.49 |
| polarC18 | 218.139 | 55.4805 | 6.9 | 0.49 |
| polarC18 | 218.139 | 80.4925 | 6.9 | 0.49 |
| polarC18 | 218.139 | 75.9861 | 6.9 | 0.49 |
| polarC18 | 218.139 | 49.8018 | 6.9 | 0.49 |
| polarC18 | 218.139 | 98.62 | 6.9 | 0.49 |
| polarC18 | 218.139 | 64.3842 | 6.9 | 0.49 |
| polarC18 | 218.139 | 69.2344 | 6.9 | 0.49 |
| polarC18 | 218.139 | 79.5767 | 6.9 | 0.49 |
| polarC18 | 218.139 | 95.0717 | 6.9 | 0.49 |
| polarC18 | 218.139 | 54.3588 | 6.9 | 0.49 |
| polarC18 | 218.139 | 58.3546 | 6.9 | 0.49 |
| polarC18 | 218.139 | 209.945 | 6.9 | 0.49 |
| polarC18 | 218.139 | 102.8803 | 6.9 | 0.49 |
| polarC18 | 218.139 | 90.5808 | 6.9 | 0.49 |
| polarC18 | 218.139 | 107.064 | 6.9 | 0.49 |
| polarC18 | 218.139 | 126.9022 | 6.9 | 0.49 |
| polarC18 | 218.139 | 64.47 | 6.9 | 0.49 |
| polarC18 | 218.139 | 99.4362 | 6.9 | 0.49 |
| polarC18 | 218.139 | 186.4842 | 6.9 | 0.49 |
| polarC18 | 218.139 | 50.4604 | 6.9 | 0.49 |
| polarC18 | 218.139 | 71.043 | 6.9 | 0.49 |
| polarC18 | 218.139 | 87.6622 | 6.9 | 0.49 |
| polarC18 | 218.139 | 87.8288 | 6.9 | 0.49 |
| polarC18 | 218.139 | 50.2092 | 6.9 | 0.49 |
| polarC18 | 218.139 | 89.5657 | 6.9 | 0.49 |
| polarC18 | 218.139 | 65.4357 | 6.9 | 0.49 |
| polarC18 | 218.139 | 202.6832 | 6.9 | 0.49 |
| polarC18 | 218.139 | 52.8307 | 6.9 | 0.49 |
| polarC18 | 218.139 | 60.7376 | 6.9 | 0.49 |
| polarC18 | 218.139 | 146.9413 | 6.9 | 0.49 |
| polarC18 | 218.139 | 215.9139 | 6.9 | 0.49 |
| polarC18 | 218.139 | 91.9254 | 6.9 | 0.49 |
| polarC18 | 218.139 | 184.3681 | 6.9 | 0.49 |
| polarC18 | 218.139 | 78.6245 | 6.9 | 0.49 |
| polarC18 | 218.139 | 83.4455 | 6.9 | 0.49 |

| | | | | |
|---|---|---|---|---|
| polarC18 | 218.139 | 53.0685 | 6.9 | 0.49 |
| polarC18 | 218.139 | 104.5965 | 6.9 | 0.49 |
| polarC18 | 218.139 | 123.9227 | 6.9 | 0.49 |
| polarC18 | 218.139 | 192.272 | 6.9 | 0.49 |
| polarC18 | 218.139 | 49.4071 | 6.9 | 0.49 |
| polarC18 | 218.139 | 130.3893 | 6.9 | 0.49 |
| polarC18 | 218.139 | 82.334 | 6.9 | 0.49 |
| polarC18 | 218.139 | 73.3238 | 6.9 | 0.49 |
| polarC18 | 218.139 | 89.1034 | 6.9 | 0.49 |
| polarC18 | 218.139 | 52.3798 | 6.9 | 0.49 |
| polarC18 | 218.139 | 73.2251 | 6.9 | 0.49 |
| polarC18 | 218.139 | 70.9453 | 6.9 | 0.49 |
| polarC18 | 218.139 | 83.644 | 6.9 | 0.49 |
| polarC18 | 218.139 | 115.7768 | 6.9 | 0.49 |
| polarC18 | 218.139 | 64.479 | 6.9 | 0.49 |
| polarC18 | 218.139 | 64.6895 | 6.9 | 0.49 |
| polarC18 | 218.139 | 52.5809 | 6.9 | 0.49 |
| polarC18 | 218.139 | 90.1568 | 6.9 | 0.49 |
| polarC18 | 218.139 | 75.6909 | 6.9 | 0.49 |
| polarC18 | 218.139 | 53.0053 | 6.9 | 0.49 |
| polarC18 | 218.139 | 96.7204 | 6.9 | 0.49 |
| polarC18 | 218.139 | 88.7561 | 6.9 | 0.49 |
| polarC18 | 218.139 | 59.097 | 6.9 | 0.49 |
| polarC18 | 218.139 | 105.4821 | 6.9 | 0.49 |
| polarC18 | 218.139 | 143.3171 | 6.9 | 0.49 |
| polarC18 | 218.139 | 60.9039 | 6.9 | 0.49 |
| polarC18 | 218.139 | 98.5098 | 6.9 | 0.49 |
| polarC18 | 218.139 | 56.734 | 6.9 | 0.49 |
| polarC18 | 218.139 | 61.2673 | 6.9 | 0.49 |
| polarC18 | 218.139 | 168.8448 | 6.9 | 0.49 |
| polarC18 | 218.139 | 215.6842 | 6.9 | 0.49 |
| polarC18 | 218.139 | 101.9283 | 6.9 | 0.49 |
| polarC18 | 218.139 | 64.9598 | 6.9 | 0.49 |
| polarC18 | 218.139 | 54.1716 | 6.9 | 0.49 |
| polarC18 | 218.139 | 127.2287 | 6.9 | 0.49 |
| polarC18 | 218.139 | 103.5357 | 6.9 | 0.49 |
| polarC18 | 218.139 | 232.506 | 6.9 | 0.49 |
| polarC18 | 218.139 | 92.5891 | 6.9 | 0.49 |
| polarC18 | 218.139 | 57.7748 | 6.9 | 0.49 |
| polarC18 | 218.139 | 170.7944 | 6.9 | 0.49 |
| polarC18 | 218.139 | 82.0696 | 6.9 | 0.49 |
| polarC18 | 218.139 | 52.3953 | 6.9 | 0.49 |

| | | | | |
|---|---|---|---|---|
| polarC18 | 218.139 | 139.9024 | 6.9 | 0.49 |
| polarC18 | 218.139 | 147.1435 | 6.9 | 0.49 |
| polarC18 | 218.139 | 148.7951 | 6.9 | 0.49 |
| polarC18 | 218.139 | 59.7309 | 6.9 | 0.49 |
| polarC18 | 218.139 | 179.1814 | 6.9 | 0.49 |
| polarC18 | 218.139 | 64.62 | 6.9 | 0.49 |
| polarC18 | 218.139 | 136.1743 | 6.9 | 0.49 |
| polarC18 | 218.139 | 78.2931 | 6.9 | 0.49 |
| polarC18 | 218.139 | 51.923 | 6.9 | 0.49 |
| polarC18 | 218.139 | 97.8905 | 6.9 | 0.49 |
| polarC18 | 218.139 | 61.531 | 6.9 | 0.49 |
| polarC18 | 218.139 | 68.3286 | 6.9 | 0.49 |
| polarC18 | 218.139 | 49.3318 | 6.9 | 0.49 |
| polarC18 | 218.139 | 197.9089 | 6.9 | 0.49 |
| polarC18 | 218.139 | 215.3587 | 6.9 | 0.49 |
| polarC18 | 218.139 | 55.6753 | 6.9 | 0.49 |
| polarC18 | 218.139 | 131.993 | 6.9 | 0.49 |
| polarC18 | 218.139 | 72.5068 | 6.9 | 0.49 |
| polarC18 | 218.139 | 62.8058 | 6.9 | 0.49 |
| polarC18 | 218.139 | 52.8673 | 6.9 | 0.49 |
| polarC18 | 218.139 | 106.4622 | 6.9 | 0.49 |
| polarC18 | 218.139 | 59.1222 | 6.9 | 0.49 |
| polarC18 | 218.139 | 183.7279 | 6.9 | 0.49 |
| polarC18 | 218.139 | 69.745 | 6.9 | 0.49 |
| polarC18 | 218.139 | 51.7665 | 6.9 | 0.49 |
| polarC18 | 218.139 | 145.7094 | 6.9 | 0.49 |
| polarC18 | 218.139 | 50.5877 | 6.9 | 0.49 |
| polarC18 | 218.139 | 53.6619 | 6.9 | 0.49 |
| polarC18 | 218.139 | 148.3088 | 6.9 | 0.49 |
| polarC18 | 218.139 | 55.8174 | 6.9 | 0.49 |
| polarC18 | 218.139 | 93.6055 | 6.9 | 0.49 |
| polarC18 | 218.139 | 100.474 | 6.9 | 0.49 |
| polarC18 | 218.139 | 90.434 | 6.9 | 0.49 |
| polarC18 | 218.139 | 71.7004 | 6.9 | 0.49 |
| polarC18 | 218.139 | 55.0283 | 6.9 | 0.49 |
| polarC18 | 218.139 | 144.074 | 6.9 | 0.49 |
| polarC18 | 218.139 | 86.3651 | 6.9 | 0.49 |
| polarC18 | 218.139 | 116.3977 | 6.9 | 0.49 |
| polarC18 | 218.139 | 111.1296 | 6.9 | 0.49 |
| polarC18 | 218.139 | 104.5602 | 6.9 | 0.49 |
| polarC18 | 218.139 | 49.0674 | 6.9 | 0.49 |
| polarC18 | 218.139 | 103.6156 | 6.9 | 0.49 |

| | | | | |
|---|---|---|---|---|
| polarC18 | 218.139 | 93.1218 | 6.9 | 0.49 |
| polarC18 | 218.139 | 81.0282 | 6.9 | 0.49 |
| polarC18 | 218.139 | 49.8687 | 6.9 | 0.49 |
| polarC18 | 218.139 | 48.8958 | 6.9 | 0.49 |
| polarC18 | 218.139 | 58.8988 | 6.9 | 0.49 |
| polarC18 | 218.139 | 96.2723 | 6.9 | 0.49 |
| polarC18 | 218.139 | 87.7736 | 6.9 | 0.49 |
| polarC18 | 218.139 | 108.7309 | 6.9 | 0.49 |
| polarC18 | 218.139 | 75.1815 | 6.9 | 0.49 |
| polarC18 | 218.139 | 127.1167 | 6.9 | 0.49 |
| polarC18 | 218.139 | 55.0042 | 6.9 | 0.49 |
| polarC18 | 218.139 | 230.0216 | 6.9 | 0.49 |
| polarC18 | 218.139 | 155.3922 | 6.9 | 0.49 |
| polarC18 | 218.139 | 100.2353 | 6.9 | 0.49 |
| polarC18 | 218.139 | 54.3773 | 6.9 | 0.49 |
| polarC18 | 218.139 | 93.9951 | 6.9 | 0.49 |
| polarC18 | 218.139 | 94.5806 | 6.9 | 0.49 |
| polarC18 | 218.139 | 63.5866 | 6.9 | 0.49 |
| polarC18 | 218.139 | 64.1264 | 6.9 | 0.49 |
| polarC18 | 218.139 | 78.7124 | 6.9 | 0.49 |
| polarC18 | 218.139 | 134.661 | 6.9 | 0.49 |
| polarC18 | 218.139 | 89.8802 | 6.9 | 0.49 |
| polarC18 | 218.139 | 106.0664 | 6.9 | 0.49 |
| polarC18 | 218.139 | 126.3238 | 6.9 | 0.49 |
| polarC18 | 218.139 | 217.4607 | 6.9 | 0.49 |
| polarC18 | 218.139 | 86.9352 | 6.9 | 0.49 |
| polarC18 | 218.139 | 64.8116 | 6.9 | 0.49 |
| polarC18 | 218.139 | 185.0494 | 6.9 | 0.49 |
| polarC18 | 218.139 | 146.6194 | 6.9 | 0.49 |
| polarC18 | 218.139 | 71.6164 | 6.9 | 0.49 |
| polarC18 | 218.139 | 168.9947 | 6.9 | 0.49 |
| polarC18 | 218.139 | 137.9076 | 6.9 | 0.49 |
| polarC18 | 218.139 | 49.8215 | 6.9 | 0.49 |
| polarC18 | 218.139 | 49.7292 | 6.9 | 0.49 |
| polarC18 | 218.139 | 57.1311 | 6.9 | 0.49 |
| polarC18 | 218.139 | 94.2228 | 6.9 | 0.49 |
| polarC18 | 218.139 | 54.1906 | 6.9 | 0.49 |
| polarC18 | 218.139 | 94.42 | 6.9 | 0.49 |
| polarC18 | 218.139 | 93.6884 | 6.9 | 0.49 |
| polarC18 | 218.139 | 225.3354 | 6.9 | 0.49 |
| polarC18 | 218.139 | 64.4571 | 6.9 | 0.49 |
| polarC18 | 218.139 | 227.3305 | 6.9 | 0.49 |

| | | | | |
|---|---|---|---|---|
| polarC18 | 218.139 | 107.8797 | 6.9 | 0.49 |
| polarC18 | 218.139 | 115.0593 | 6.9 | 0.49 |
| polarC18 | 218.139 | 141.4102 | 6.9 | 0.49 |
| polarC18 | 218.139 | 167.5937 | 6.9 | 0.49 |
| polarC18 | 218.139 | 66.3916 | 6.9 | 0.49 |
| polarC18 | 218.139 | 157.6915 | 6.9 | 0.49 |
| polarC18 | 218.139 | 65.5967 | 6.9 | 0.49 |
| polarC18 | 218.139 | 64.5107 | 6.9 | 0.49 |
| polarC18 | 218.139 | 223.7923 | 6.9 | 0.49 |
| polarC18 | 218.139 | 195.4969 | 6.9 | 0.49 |
| polarC18 | 218.139 | 57.5821 | 6.9 | 0.49 |
| polarC18 | 218.139 | 72.9528 | 6.9 | 0.49 |
| polarC18 | 218.139 | 222.3174 | 6.9 | 0.49 |
| polarC18 | 218.139 | 96.6297 | 6.9 | 0.49 |
| polarC18 | 218.139 | 103.7691 | 6.9 | 0.49 |
| polarC18 | 218.139 | 132.2287 | 6.9 | 0.49 |
| polarC18 | 218.139 | 104.9867 | 6.9 | 0.49 |
| polarC18 | 218.139 | 86.9939 | 6.9 | 0.49 |
| polarC18 | 218.139 | 53.3225 | 6.9 | 0.49 |
| polarC18 | 218.139 | 48.7748 | 6.9 | 0.49 |
| polarC18 | 218.139 | 100.749 | 6.9 | 0.49 |
| polarC18 | 218.139 | 87.1489 | 6.9 | 0.49 |
| polarC18 | 218.139 | 51.8366 | 6.9 | 0.49 |
| polarC18 | 218.139 | 139.3203 | 6.9 | 0.49 |
| polarC18 | 218.139 | 61.3537 | 6.8 | 0.49 |
| polarC18 | 218.139 | 55.7308 | 6.8 | 0.49 |
| polarC18 | 218.139 | 55.6838 | 6.8 | 0.49 |
| polarC18 | 218.139 | 54.7726 | 6.8 | 0.49 |
| polarC18 | 218.139 | 67.8143 | 6.8 | 0.49 |
| polarC18 | 218.139 | 234.7798 | 6.8 | 0.49 |
| polarC18 | 218.139 | 148.354 | 6.8 | 0.49 |
| polarC18 | 218.139 | 53.8374 | 6.8 | 0.49 |
| polarC18 | 218.139 | 157.7309 | 6.8 | 0.49 |
| polarC18 | 218.139 | 49.7803 | 6.8 | 0.49 |
| polarC18 | 218.139 | 86.6714 | 6.8 | 0.49 |
| polarC18 | 218.139 | 183.6211 | 6.8 | 0.49 |
| polarC18 | 218.139 | 64.9823 | 6.8 | 0.49 |
| polarC18 | 218.139 | 94.1955 | 6.8 | 0.49 |
| polarC18 | 218.139 | 107.0314 | 6.8 | 0.49 |
| polarC18 | 218.139 | 107.9781 | 6.8 | 0.49 |
| polarC18 | 218.139 | 99.7573 | 6.8 | 0.49 |
| polarC18 | 218.139 | 225.5953 | 6.8 | 0.49 |

| | | | | |
|---|---|---|---|---|
| polarC18 | 218.139 | 91.3084 | 6.8 | 0.49 |
| polarC18 | 218.139 | 63.7713 | 6.8 | 0.49 |
| polarC18 | 218.139 | 60.0212 | 6.8 | 0.49 |
| polarC18 | 218.139 | 71.3674 | 6.8 | 0.49 |
| polarC18 | 218.139 | 141.7858 | 6.8 | 0.49 |
| polarC18 | 218.139 | 121.0665 | 6.8 | 0.49 |
| polarC18 | 218.139 | 128.3342 | 6.8 | 0.49 |
| polarC18 | 218.139 | 182.0569 | 6.8 | 0.49 |
| polarC18 | 218.139 | 130.6215 | 6.8 | 0.49 |
| polarC18 | 218.139 | 93.1756 | 6.8 | 0.49 |
| polarC18 | 218.139 | 96.1581 | 6.8 | 0.49 |
| polarC18 | 218.139 | 84.6957 | 6.8 | 0.49 |
| polarC18 | 218.139 | 92.2878 | 6.8 | 0.49 |
| polarC18 | 218.139 | 115.8862 | 6.8 | 0.48 |
| polarC18 | 218.139 | 187.4932 | 6.8 | 0.48 |
| polarC18 | 218.139 | 49.9028 | 6.8 | 0.48 |
| polarC18 | 218.139 | 93.0633 | 6.8 | 0.48 |
| polarC18 | 218.139 | 120.4765 | 6.8 | 0.48 |
| polarC18 | 218.139 | 105.3807 | 6.8 | 0.48 |
| polarC18 | 218.139 | 82.5183 | 6.8 | 0.48 |
| polarC18 | 218.139 | 119.8805 | 6.8 | 0.48 |
| polarC18 | 218.139 | 58.087 | 6.8 | 0.48 |
| polarC18 | 218.139 | 88.635 | 6.8 | 0.48 |
| polarC18 | 218.139 | 119.2629 | 6.8 | 0.48 |
| polarC18 | 218.139 | 51.7807 | 6.8 | 0.48 |
| polarC18 | 218.139 | 199.3261 | 6.8 | 0.48 |
| polarC18 | 218.139 | 235.0349 | 6.8 | 0.48 |
| polarC18 | 218.139 | 78.2815 | 6.8 | 0.48 |
| polarC18 | 218.139 | 89.3287 | 6.8 | 0.48 |
| polarC18 | 218.139 | 88.3347 | 6.8 | 0.48 |
| polarC18 | 218.139 | 54.3561 | 6.8 | 0.48 |
| polarC18 | 218.139 | 226.1168 | 6.8 | 0.48 |
| polarC18 | 218.139 | 131.1482 | 6.8 | 0.48 |
| polarC18 | 218.139 | 132.3829 | 6.8 | 0.48 |
| polarC18 | 218.139 | 175.8343 | 6.8 | 0.48 |
| polarC18 | 218.139 | 66.0639 | 6.8 | 0.48 |
| polarC18 | 218.139 | 186.233 | 6.8 | 0.48 |
| polarC18 | 218.139 | 66.4572 | 6.8 | 0.48 |
| polarC18 | 218.139 | 60.9422 | 6.8 | 0.48 |
| polarC18 | 218.139 | 51.0936 | 6.8 | 0.48 |
| polarC18 | 218.139 | 169.5305 | 6.8 | 0.48 |
| polarC18 | 218.139 | 224.2993 | 6.8 | 0.48 |

| | | | | |
|---|---|---|---|---|
| polarC18 | 218.139 | 57.1951 | 6.8 | 0.48 |
| polarC18 | 218.139 | 77.7221 | 6.8 | 0.48 |
| polarC18 | 218.139 | 55.2908 | 6.8 | 0.48 |
| polarC18 | 218.139 | 141.1327 | 6.8 | 0.48 |
| polarC18 | 218.139 | 66.0169 | 6.8 | 0.48 |
| polarC18 | 218.139 | 77.3632 | 6.8 | 0.48 |
| polarC18 | 218.139 | 112.0841 | 6.8 | 0.48 |
| polarC18 | 218.139 | 59.49 | 6.8 | 0.48 |
| polarC18 | 218.139 | 67.0754 | 6.8 | 0.48 |
| polarC18 | 218.139 | 50.2432 | 6.8 | 0.48 |
| polarC18 | 218.139 | 178.3694 | 6.8 | 0.48 |
| polarC18 | 218.139 | 67.5715 | 6.8 | 0.48 |
| polarC18 | 218.139 | 66.6104 | 6.8 | 0.48 |
| polarC18 | 218.139 | 192.8897 | 6.8 | 0.48 |
| polarC18 | 218.139 | 55.8347 | 6.8 | 0.48 |
| polarC18 | 218.139 | 135.2839 | 6.8 | 0.48 |
| polarC18 | 218.139 | 52.0347 | 6.8 | 0.48 |
| polarC18 | 218.139 | 76.3204 | 6.8 | 0.48 |
| polarC18 | 218.139 | 197.7641 | 6.8 | 0.48 |
| polarC18 | 218.139 | 114.8432 | 6.8 | 0.48 |
| polarC18 | 218.139 | 63.6047 | 6.8 | 0.48 |
| polarC18 | 218.139 | 151.4785 | 6.8 | 0.48 |
| polarC18 | 218.139 | 145.5997 | 6.8 | 0.48 |
| polarC18 | 218.139 | 74.3303 | 6.8 | 0.48 |
| polarC18 | 218.139 | 67.3671 | 6.8 | 0.48 |
| polarC18 | 218.139 | 148.9 | 6.8 | 0.48 |
| polarC18 | 218.139 | 217.663 | 6.8 | 0.48 |
| polarC18 | 218.139 | 122.3872 | 6.8 | 0.48 |
| polarC18 | 218.139 | 66.6306 | 6.8 | 0.48 |
| polarC18 | 218.139 | 81.7082 | 6.8 | 0.48 |
| polarC18 | 218.139 | 126.1303 | 6.8 | 0.48 |
| polarC18 | 218.139 | 85.7615 | 6.8 | 0.48 |
| polarC18 | 218.139 | 55.247 | 6.8 | 0.48 |
| polarC18 | 218.139 | 150.1851 | 6.8 | 0.48 |
| polarC18 | 218.139 | 70.0327 | 6.8 | 0.48 |
| polarC18 | 218.139 | 71.9259 | 6.8 | 0.48 |
| polarC18 | 218.139 | 94.1485 | 6.8 | 0.48 |
| polarC18 | 218.139 | 156.2745 | 6.8 | 0.48 |
| polarC18 | 218.139 | 115.0765 | 6.8 | 0.48 |
| polarC18 | 218.139 | 120.1087 | 6.8 | 0.48 |
| polarC18 | 218.139 | 81.6054 | 6.8 | 0.48 |
| polarC18 | 218.139 | 51.9641 | 6.8 | 0.48 |

| | | | | |
|---|---|---|---|---|
| polarC18 | 218.139 | 168.5208 | 6.8 | 0.48 |
| polarC18 | 218.139 | 77.5769 | 6.8 | 0.48 |
| polarC18 | 218.139 | 121.7558 | 6.8 | 0.48 |
| polarC18 | 218.139 | 132.4644 | 6.8 | 0.48 |
| polarC18 | 218.139 | 65.2422 | 6.8 | 0.48 |
| polarC18 | 218.139 | 103.7454 | 6.8 | 0.48 |
| polarC18 | 218.139 | 103.8117 | 6.8 | 0.48 |
| polarC18 | 218.139 | 71.2266 | 6.8 | 0.48 |
| polarC18 | 218.139 | 65.6491 | 6.8 | 0.48 |
| polarC18 | 218.139 | 193.9672 | 6.8 | 0.48 |
| polarC18 | 218.139 | 115.5417 | 6.8 | 0.48 |
| polarC18 | 218.139 | 207.4446 | 6.8 | 0.48 |
| polarC18 | 218.139 | 179.8776 | 6.8 | 0.48 |
| polarC18 | 218.139 | 75.7319 | 6.8 | 0.48 |
| polarC18 | 218.139 | 216.3374 | 6.8 | 0.48 |
| polarC18 | 218.139 | 117.0568 | 6.8 | 0.48 |
| polarC18 | 218.139 | 70.9052 | 6.8 | 0.48 |
| polarC18 | 218.139 | 68.645 | 6.8 | 0.48 |
| polarC18 | 218.139 | 91.4239 | 6.8 | 0.48 |
| polarC18 | 218.139 | 65.4231 | 6.8 | 0.48 |
| polarC18 | 218.139 | 60.6761 | 6.8 | 0.48 |
| polarC18 | 218.139 | 63.8355 | 6.8 | 0.48 |
| polarC18 | 218.139 | 106.9623 | 6.8 | 0.48 |
| polarC18 | 218.139 | 94.389 | 6.8 | 0.48 |
| polarC18 | 218.139 | 55.6229 | 6.8 | 0.48 |
| polarC18 | 218.139 | 113.8464 | 6.8 | 0.48 |
| polarC18 | 218.139 | 70.4095 | 6.8 | 0.48 |
| polarC18 | 218.139 | 120.2142 | 6.8 | 0.48 |
| polarC18 | 218.139 | 56.1356 | 6.8 | 0.48 |
| polarC18 | 218.139 | 88.7397 | 6.8 | 0.48 |
| polarC18 | 218.139 | 188.7225 | 6.8 | 0.48 |
| polarC18 | 218.139 | 58.6096 | 6.8 | 0.48 |
| polarC18 | 218.139 | 99.7837 | 6.8 | 0.48 |
| polarC18 | 218.139 | 53.2253 | 6.8 | 0.48 |
| polarC18 | 218.139 | 65.1664 | 6.8 | 0.48 |
| polarC18 | 218.139 | 174.6866 | 6.8 | 0.48 |
| polarC18 | 218.139 | 51.3689 | 6.8 | 0.48 |
| polarC18 | 218.139 | 134.699 | 6.8 | 0.48 |
| polarC18 | 218.139 | 55.5483 | 6.8 | 0.48 |
| polarC18 | 218.139 | 182.7963 | 6.8 | 0.48 |
| polarC18 | 218.139 | 75.5255 | 6.8 | 0.48 |
| polarC18 | 218.139 | 227.0556 | 6.8 | 0.48 |

| | | | | |
|---|---|---|---|---|
| polarC18 | 218.139 | 108.3087 | 6.8 | 0.48 |
| polarC18 | 218.139 | 51.7136 | 6.8 | 0.48 |
| polarC18 | 218.139 | 53.3019 | 6.8 | 0.48 |
| polarC18 | 218.139 | 153.9814 | 6.8 | 0.48 |
| polarC18 | 218.139 | 241.638 | 6.8 | 0.48 |
| polarC18 | 218.139 | 108.9154 | 6.8 | 0.48 |
| polarC18 | 218.139 | 70.2908 | 6.8 | 0.48 |
| polarC18 | 218.139 | 52.0763 | 6.8 | 0.48 |
| polarC18 | 218.139 | 56.9347 | 6.8 | 0.48 |
| polarC18 | 218.139 | 70.9136 | 6.8 | 0.48 |
| polarC18 | 218.139 | 51.9508 | 6.8 | 0.48 |
| polarC18 | 218.139 | 85.6948 | 6.8 | 0.48 |
| polarC18 | 218.139 | 57.1793 | 6.8 | 0.48 |
| polarC18 | 218.139 | 61.4683 | 6.8 | 0.48 |
| polarC18 | 218.139 | 209.3232 | 6.8 | 0.48 |
| polarC18 | 218.139 | 59.4199 | 6.8 | 0.48 |
| polarC18 | 218.139 | 48.8487 | 6.8 | 0.48 |
| polarC18 | 218.139 | 135.6582 | 6.8 | 0.48 |
| polarC18 | 218.139 | 173.2435 | 6.8 | 0.48 |
| polarC18 | 218.139 | 139.9502 | 6.8 | 0.48 |
| polarC18 | 218.139 | 74.4017 | 6.8 | 0.48 |
| polarC18 | 218.139 | 61.1798 | 6.8 | 0.48 |
| polarC18 | 218.139 | 67.8267 | 6.8 | 0.48 |
| polarC18 | 218.139 | 94.8665 | 6.8 | 0.48 |
| polarC18 | 218.139 | 104.7158 | 6.8 | 0.48 |
| polarC18 | 218.139 | 49.7401 | 6.8 | 0.48 |
| polarC18 | 218.139 | 74.5962 | 6.8 | 0.48 |
| polarC18 | 218.139 | 65.9283 | 6.8 | 0.48 |
| polarC18 | 218.139 | 182.687 | 6.8 | 0.48 |
| polarC18 | 218.139 | 161.2078 | 6.8 | 0.48 |
| polarC18 | 218.139 | 92.9718 | 6.8 | 0.48 |
| polarC18 | 218.139 | 65.3846 | 6.8 | 0.48 |
| polarC18 | 218.139 | 218.77 | 6.8 | 0.48 |
| polarC18 | 218.139 | 74.6939 | 6.8 | 0.48 |
| polarC18 | 218.139 | 66.0024 | 6.8 | 0.48 |
| polarC18 | 218.139 | 83.4006 | 6.8 | 0.48 |
| polarC18 | 218.139 | 102.5384 | 6.8 | 0.48 |
| polarC18 | 218.139 | 53.3863 | 6.8 | 0.48 |
| polarC18 | 218.139 | 222.5351 | 6.8 | 0.48 |
| polarC18 | 218.139 | 59.2221 | 6.8 | 0.48 |
| polarC18 | 218.139 | 238.7932 | 6.8 | 0.48 |
| polarC18 | 218.139 | 63.3183 | 6.8 | 0.48 |

| | | | | |
|---|---|---|---|---|
| polarC18 | 218.139 | 59.2774 | 6.8 | 0.48 |
| polarC18 | 218.139 | 92.8545 | 6.8 | 0.48 |
| polarC18 | 218.139 | 61.7777 | 6.8 | 0.48 |
| polarC18 | 218.139 | 203.9425 | 6.8 | 0.48 |
| polarC18 | 218.139 | 56.2443 | 6.8 | 0.48 |
| polarC18 | 218.139 | 56.529 | 6.8 | 0.48 |
| polarC18 | 218.139 | 138.4486 | 6.8 | 0.48 |
| polarC18 | 218.139 | 49.0978 | 6.8 | 0.48 |
| polarC18 | 218.139 | 54.817 | 6.8 | 0.48 |
| polarC18 | 218.139 | 91.3292 | 6.8 | 0.48 |
| polarC18 | 218.139 | 61.7302 | 6.8 | 0.48 |
| polarC18 | 218.139 | 92.8766 | 6.8 | 0.48 |
| polarC18 | 218.139 | 48.9503 | 6.8 | 0.48 |
| polarC18 | 218.139 | 87.1892 | 6.8 | 0.48 |
| polarC18 | 218.139 | 80.2963 | 6.8 | 0.48 |
| polarC18 | 218.139 | 52.1542 | 6.8 | 0.48 |
| polarC18 | 218.139 | 180.9503 | 6.8 | 0.48 |
| polarC18 | 218.139 | 49.6985 | 6.8 | 0.48 |
| polarC18 | 218.139 | 87.717 | 6.8 | 0.48 |
| polarC18 | 218.139 | 72.234 | 6.8 | 0.48 |
| polarC18 | 218.139 | 60.7908 | 6.8 | 0.48 |
| polarC18 | 218.139 | 82.4869 | 6.8 | 0.48 |
| polarC18 | 218.139 | 68.8393 | 6.8 | 0.48 |
| polarC18 | 218.139 | 56.7004 | 6.8 | 0.48 |
| polarC18 | 218.139 | 76.5096 | 6.8 | 0.48 |
| polarC18 | 218.139 | 70.0686 | 6.8 | 0.48 |
| polarC18 | 218.139 | 70.1354 | 6.8 | 0.48 |
| polarC18 | 218.139 | 61.0347 | 6.8 | 0.48 |
| polarC18 | 218.139 | 53.8722 | 6.8 | 0.48 |
| polarC18 | 218.139 | 49.1613 | 6.8 | 0.48 |
| polarC18 | 218.139 | 56.7838 | 6.8 | 0.48 |
| polarC18 | 218.139 | 83.9892 | 6.8 | 0.48 |
| polarC18 | 218.139 | 161.0672 | 6.8 | 0.48 |
| polarC18 | 218.139 | 49.9641 | 6.8 | 0.48 |
| polarC18 | 218.139 | 191.4509 | 6.8 | 0.48 |
| polarC18 | 218.139 | 64.0554 | 6.8 | 0.48 |
| polarC18 | 218.139 | 71.4073 | 6.8 | 0.48 |
| polarC18 | 218.139 | 76.0072 | 6.8 | 0.48 |
| polarC18 | 218.139 | 52.721 | 6.8 | 0.48 |
| polarC18 | 218.139 | 84.7919 | 6.8 | 0.48 |
| polarC18 | 218.139 | 50.5036 | 6.8 | 0.48 |
| polarC18 | 218.139 | 58.67 | 6.8 | 0.48 |

| | | | | |
|---|---|---|---|---|
| polarC18 | 218.139 | 80.1911 | 6.8 | 0.48 |
| polarC18 | 218.139 | 60.6798 | 6.8 | 0.48 |
| polarC18 | 218.139 | 61.1545 | 6.8 | 0.48 |
| polarC18 | 218.139 | 52.7287 | 6.8 | 0.48 |
| polarC18 | 218.139 | 63.9258 | 6.8 | 0.48 |
| polarC18 | 218.139 | 59.8793 | 6.8 | 0.48 |
| polarC18 | 218.139 | 76.3909 | 6.8 | 0.48 |
| polarC18 | 218.139 | 53.9949 | 6.8 | 0.48 |
| polarC18 | 218.139 | 81.3961 | 6.8 | 0.48 |
| polarC18 | 218.139 | 142.4648 | 6.8 | 0.48 |
| polarC18 | 218.139 | 119.3766 | 6.8 | 0.48 |
| polarC18 | 218.139 | 78.9513 | 6.7 | 0.48 |
| polarC18 | 218.139 | 122.4706 | 6.7 | 0.48 |
| polarC18 | 218.139 | 57.5858 | 6.7 | 0.48 |
| polarC18 | 218.139 | 97.6875 | 6.7 | 0.48 |
| polarC18 | 218.139 | 61.985 | 6.7 | 0.48 |
| polarC18 | 218.139 | 87.0014 | 6.7 | 0.48 |
| polarC18 | 218.139 | 61.3919 | 6.7 | 0.48 |
| polarC18 | 218.139 | 61.3062 | 6.7 | 0.48 |
| polarC18 | 218.139 | 51.6595 | 6.7 | 0.48 |
| polarC18 | 218.139 | 71.9474 | 6.7 | 0.48 |
| polarC18 | 218.139 | 75.7517 | 6.7 | 0.48 |
| polarC18 | 218.139 | 64.7072 | 6.7 | 0.48 |
| polarC18 | 218.139 | 108.6643 | 6.7 | 0.48 |
| polarC18 | 218.139 | 97.4704 | 6.7 | 0.48 |
| polarC18 | 218.139 | 51.858 | 6.7 | 0.48 |
| polarC18 | 218.139 | 112.4616 | 6.7 | 0.48 |
| polarC18 | 218.139 | 62.2878 | 6.7 | 0.48 |
| polarC18 | 218.139 | 143.8609 | 6.7 | 0.48 |
| polarC18 | 218.139 | 76.3669 | 6.7 | 0.48 |
| polarC18 | 218.139 | 240.6355 | 6.7 | 0.48 |
| polarC18 | 218.139 | 156.3773 | 6.7 | 0.48 |
| polarC18 | 218.139 | 51.2415 | 6.7 | 0.48 |
| polarC18 | 218.139 | 64.3935 | 6.7 | 0.48 |
| polarC18 | 218.139 | 63.9757 | 6.7 | 0.48 |
| polarC18 | 218.139 | 241.3838 | 6.7 | 0.48 |
| polarC18 | 218.139 | 129.6759 | 6.7 | 0.48 |
| polarC18 | 218.139 | 76.7251 | 6.7 | 0.48 |
| polarC18 | 218.139 | 85.4002 | 6.7 | 0.48 |
| polarC18 | 218.139 | 74.2531 | 6.7 | 0.48 |
| polarC18 | 218.139 | 196.6019 | 6.7 | 0.48 |
| polarC18 | 218.139 | 81.8031 | 6.7 | 0.48 |

| | | | | |
|---|---|---|---|---|
| polarC18 | 218.139 | 152.2223 | 6.7 | 0.48 |
| polarC18 | 218.139 | 81.2326 | 6.7 | 0.48 |
| polarC18 | 218.139 | 209.2349 | 6.7 | 0.48 |
| polarC18 | 218.139 | 160.9033 | 6.7 | 0.48 |
| polarC18 | 218.139 | 66.9587 | 6.7 | 0.48 |
| polarC18 | 218.139 | 59.1427 | 6.7 | 0.48 |
| polarC18 | 218.139 | 92.6041 | 6.7 | 0.48 |
| polarC18 | 218.139 | 56.0735 | 6.7 | 0.48 |
| polarC18 | 218.139 | 182.6462 | 6.7 | 0.48 |
| polarC18 | 218.139 | 74.34 | 6.7 | 0.48 |
| polarC18 | 218.139 | 61.6367 | 6.7 | 0.48 |
| polarC18 | 218.139 | 142.7404 | 6.7 | 0.48 |
| polarC18 | 218.139 | 83.7172 | 6.7 | 0.48 |
| polarC18 | 218.139 | 69.2795 | 6.7 | 0.48 |
| polarC18 | 218.139 | 54.949 | 6.7 | 0.48 |
| polarC18 | 218.139 | 73.6598 | 6.7 | 0.48 |
| polarC18 | 218.139 | 56.2984 | 6.7 | 0.48 |
| polarC18 | 218.139 | 214.9133 | 6.7 | 0.48 |
| polarC18 | 218.139 | 103.1858 | 6.7 | 0.48 |
| polarC18 | 218.139 | 50.2674 | 6.7 | 0.48 |
| polarC18 | 218.139 | 50.6212 | 6.7 | 0.48 |
| polarC18 | 218.139 | 211.1245 | 6.7 | 0.48 |
| polarC18 | 218.139 | 61.0406 | 6.7 | 0.48 |
| polarC18 | 218.139 | 53.9636 | 6.7 | 0.48 |
| polarC18 | 218.139 | 94.8318 | 6.7 | 0.48 |
| polarC18 | 218.139 | 55.89 | 6.7 | 0.48 |
| polarC18 | 218.139 | 70.1811 | 6.7 | 0.48 |
| polarC18 | 218.139 | 102.1539 | 6.7 | 0.48 |
| polarC18 | 218.139 | 52.1106 | 6.7 | 0.48 |
| polarC18 | 218.139 | 106.2631 | 6.7 | 0.48 |
| polarC18 | 218.139 | 51.5386 | 6.7 | 0.48 |
| polarC18 | 218.139 | 61.8424 | 6.7 | 0.48 |
| polarC18 | 218.139 | 54.4189 | 6.7 | 0.48 |
| polarC18 | 218.139 | 60.0724 | 6.7 | 0.48 |
| polarC18 | 218.139 | 55.217 | 6.7 | 0.48 |
| polarC18 | 218.139 | 242.175 | 6.7 | 0.48 |
| polarC18 | 218.139 | 113.9009 | 6.7 | 0.48 |
| polarC18 | 218.139 | 57.4914 | 6.7 | 0.48 |
| polarC18 | 218.139 | 118.9524 | 6.7 | 0.48 |
| polarC18 | 218.139 | 73.2322 | 6.7 | 0.48 |
| polarC18 | 218.139 | 51.4261 | 6.7 | 0.48 |
| polarC18 | 218.139 | 164.4211 | 6.7 | 0.48 |

| | | | | |
|---|---|---|---|---|
| polarC18 | 218.139 | 198.706 | 6.7 | 0.48 |
| polarC18 | 218.139 | 50.0545 | 6.7 | 0.48 |
| polarC18 | 218.139 | 103.7005 | 6.7 | 0.48 |
| polarC18 | 218.139 | 104.44 | 6.7 | 0.48 |
| polarC18 | 218.139 | 128.1714 | 6.7 | 0.48 |
| polarC18 | 218.139 | 60.2788 | 6.7 | 0.48 |
| polarC18 | 218.139 | 66.8202 | 6.7 | 0.48 |
| polarC18 | 218.139 | 51.9255 | 6.7 | 0.48 |
| polarC18 | 218.139 | 49.9547 | 6.7 | 0.48 |
| polarC18 | 218.139 | 50.4221 | 6.7 | 0.48 |
| polarC18 | 218.139 | 112.0055 | 6.7 | 0.48 |
| polarC18 | 218.139 | 51.583 | 6.7 | 0.48 |
| polarC18 | 218.139 | 124.6987 | 6.7 | 0.48 |
| polarC18 | 218.139 | 57.2458 | 6.7 | 0.48 |
| polarC18 | 218.139 | 78.21 | 6.7 | 0.48 |
| polarC18 | 218.139 | 62.5526 | 6.7 | 0.48 |
| polarC18 | 218.139 | 101.8653 | 6.7 | 0.48 |
| polarC18 | 218.139 | 213.35 | 6.7 | 0.48 |
| polarC18 | 218.139 | 49.7707 | 6.7 | 0.48 |
| polarC18 | 218.139 | 103.2422 | 6.7 | 0.48 |
| polarC18 | 218.139 | 98.0791 | 6.7 | 0.48 |
| polarC18 | 218.139 | 82.2841 | 6.7 | 0.48 |
| polarC18 | 218.139 | 116.5718 | 6.7 | 0.48 |
| polarC18 | 218.139 | 68.204 | 6.7 | 0.48 |
| polarC18 | 218.139 | 90.0656 | 6.7 | 0.48 |
| polarC18 | 218.139 | 93.5722 | 6.7 | 0.48 |
| polarC18 | 218.139 | 76.5736 | 6.7 | 0.48 |
| polarC18 | 218.139 | 120.7644 | 6.7 | 0.48 |
| polarC18 | 218.139 | 169.9936 | 6.7 | 0.48 |
| polarC18 | 218.139 | 49.6098 | 6.7 | 0.48 |
| polarC18 | 218.139 | 112.5075 | 6.7 | 0.48 |
| polarC18 | 218.139 | 188.2322 | 6.7 | 0.48 |
| polarC18 | 218.139 | 172.7593 | 6.7 | 0.48 |
| polarC18 | 218.139 | 105.2207 | 6.7 | 0.48 |
| polarC18 | 218.139 | 57.9899 | 6.7 | 0.48 |
| polarC18 | 218.139 | 108.7648 | 6.7 | 0.48 |
| polarC18 | 218.139 | 62.9215 | 6.7 | 0.48 |
| polarC18 | 218.139 | 78.791 | 6.7 | 0.48 |
| polarC18 | 218.139 | 127.1309 | 6.7 | 0.48 |
| polarC18 | 218.139 | 83.9526 | 6.7 | 0.48 |
| polarC18 | 218.139 | 51.6118 | 6.7 | 0.48 |
| polarC18 | 218.139 | 55.9079 | 6.7 | 0.48 |

| | | | | |
|---|---|---|---|---|
| polarC18 | 218.139 | 59.8201 | 6.7 | 0.48 |
| polarC18 | 218.139 | 191.7065 | 6.7 | 0.48 |
| polarC18 | 218.139 | 53.2436 | 6.7 | 0.48 |
| polarC18 | 218.139 | 193.288 | 6.7 | 0.48 |
| polarC18 | 218.139 | 68.6954 | 6.7 | 0.48 |
| polarC18 | 218.139 | 66.6892 | 6.7 | 0.48 |
| polarC18 | 218.139 | 104.8369 | 6.7 | 0.48 |
| polarC18 | 218.139 | 143.5211 | 6.7 | 0.48 |
| polarC18 | 218.139 | 86.3037 | 6.7 | 0.48 |
| polarC18 | 218.139 | 164.4377 | 6.7 | 0.48 |
| polarC18 | 218.139 | 151.9261 | 6.7 | 0.48 |
| polarC18 | 218.139 | 162.9961 | 6.7 | 0.47 |
| polarC18 | 218.139 | 114.2721 | 6.7 | 0.47 |
| polarC18 | 218.139 | 56.362 | 6.7 | 0.47 |
| polarC18 | 218.139 | 50.8735 | 6.7 | 0.47 |
| polarC18 | 218.139 | 69.6751 | 6.7 | 0.47 |
| polarC18 | 218.139 | 108.2596 | 6.7 | 0.47 |
| polarC18 | 218.139 | 60.7529 | 6.7 | 0.47 |
| polarC18 | 218.139 | 58.8423 | 6.7 | 0.47 |
| polarC18 | 218.139 | 141.3021 | 6.7 | 0.47 |
| polarC18 | 218.139 | 170.8775 | 6.7 | 0.47 |
| polarC18 | 218.139 | 64.5663 | 6.7 | 0.47 |
| polarC18 | 218.139 | 51.8885 | 6.7 | 0.47 |
| polarC18 | 218.139 | 61.5693 | 6.7 | 0.47 |
| polarC18 | 218.139 | 60.3137 | 6.7 | 0.47 |
| polarC18 | 218.139 | 58.9985 | 6.7 | 0.47 |
| polarC18 | 218.139 | 58.7672 | 6.7 | 0.47 |
| polarC18 | 218.139 | 215.0515 | 6.7 | 0.47 |
| polarC18 | 218.139 | 49.7478 | 6.7 | 0.47 |
| polarC18 | 218.139 | 77.6114 | 6.7 | 0.47 |
| polarC18 | 218.139 | 73.0478 | 6.7 | 0.47 |
| polarC18 | 218.139 | 56.5188 | 6.7 | 0.47 |
| polarC18 | 218.139 | 171.7506 | 6.7 | 0.47 |
| polarC18 | 218.139 | 131.0963 | 6.7 | 0.47 |
| polarC18 | 218.139 | 113.7637 | 6.7 | 0.47 |
| polarC18 | 218.139 | 84.7157 | 6.7 | 0.47 |
| polarC18 | 218.139 | 183.5978 | 6.7 | 0.47 |
| polarC18 | 218.139 | 170.0673 | 6.7 | 0.47 |
| polarC18 | 218.139 | 71.3508 | 6.7 | 0.47 |
| polarC18 | 218.139 | 71.715 | 6.7 | 0.47 |
| polarC18 | 218.139 | 101.231 | 6.7 | 0.47 |
| polarC18 | 218.139 | 83.6326 | 6.7 | 0.47 |

| | | | | |
|---|---|---|---|---|
| polarC18 | 218.139 | 115.605 | 6.7 | 0.47 |
| polarC18 | 218.139 | 64.7667 | 6.7 | 0.47 |
| polarC18 | 218.139 | 54.8095 | 6.7 | 0.47 |
| polarC18 | 218.139 | 58.6507 | 6.7 | 0.47 |
| polarC18 | 218.139 | 174.6112 | 6.7 | 0.47 |
| polarC18 | 218.139 | 136.8565 | 6.7 | 0.47 |
| polarC18 | 218.139 | 81.141 | 6.7 | 0.47 |
| polarC18 | 218.139 | 57.1929 | 6.7 | 0.47 |
| polarC18 | 218.139 | 204.8788 | 6.7 | 0.47 |
| polarC18 | 218.139 | 87.5433 | 6.7 | 0.47 |
| polarC18 | 218.139 | 68.5199 | 6.7 | 0.47 |
| polarC18 | 218.139 | 85.5888 | 6.7 | 0.47 |
| polarC18 | 218.139 | 224.5265 | 6.7 | 0.47 |
| polarC18 | 218.139 | 107.692 | 6.7 | 0.47 |
| polarC18 | 218.139 | 101.4024 | 6.7 | 0.47 |
| polarC18 | 218.139 | 128.1602 | 6.7 | 0.47 |
| polarC18 | 218.139 | 64.0775 | 6.7 | 0.47 |
| polarC18 | 218.139 | 85.3689 | 6.7 | 0.47 |
| polarC18 | 218.139 | 154.8347 | 6.7 | 0.47 |
| polarC18 | 218.139 | 50.7843 | 6.7 | 0.47 |
| polarC18 | 218.139 | 114.1781 | 6.7 | 0.47 |
| polarC18 | 218.139 | 226.0034 | 6.7 | 0.47 |
| polarC18 | 218.139 | 125.6556 | 6.7 | 0.47 |
| polarC18 | 218.139 | 49.3913 | 6.7 | 0.47 |
| polarC18 | 218.139 | 226.2238 | 6.7 | 0.47 |
| polarC18 | 218.139 | 104.93 | 6.7 | 0.47 |
| polarC18 | 218.139 | 162.1375 | 6.7 | 0.47 |
| polarC18 | 218.139 | 79.4184 | 6.7 | 0.47 |
| polarC18 | 218.139 | 72.6034 | 6.7 | 0.47 |
| polarC18 | 218.139 | 63.2537 | 6.7 | 0.47 |
| polarC18 | 218.139 | 130.1775 | 6.7 | 0.47 |
| polarC18 | 218.139 | 131.7432 | 6.7 | 0.47 |
| polarC18 | 218.139 | 160.8292 | 6.7 | 0.47 |
| polarC18 | 218.139 | 49.3092 | 6.7 | 0.47 |
| polarC18 | 218.139 | 189.788 | 6.7 | 0.47 |
| polarC18 | 218.139 | 129.8477 | 6.7 | 0.47 |
| polarC18 | 218.139 | 109.3872 | 6.7 | 0.47 |
| polarC18 | 218.139 | 74.8311 | 6.7 | 0.47 |
| polarC18 | 218.139 | 65.3155 | 6.7 | 0.47 |
| polarC18 | 218.139 | 89.5298 | 6.7 | 0.47 |
| polarC18 | 218.139 | 53.0953 | 6.7 | 0.47 |
| polarC18 | 218.139 | 235.2846 | 6.7 | 0.47 |

| | | | | |
|---|---|---|---|---|
| polarC18 | 218.139 | 150.4105 | 6.7 | 0.47 |
| polarC18 | 218.139 | 48.7548 | 6.7 | 0.47 |
| polarC18 | 218.139 | 57.8369 | 6.7 | 0.47 |
| polarC18 | 218.139 | 149.1006 | 6.7 | 0.47 |
| polarC18 | 218.139 | 147.077 | 6.7 | 0.47 |
| polarC18 | 218.139 | 186.1518 | 6.7 | 0.47 |
| polarC18 | 218.139 | 50.2617 | 6.7 | 0.47 |
| polarC18 | 218.139 | 128.2023 | 6.7 | 0.47 |
| polarC18 | 218.139 | 87.6425 | 6.7 | 0.47 |
| polarC18 | 218.139 | 75.158 | 6.7 | 0.47 |
| polarC18 | 218.139 | 49.5266 | 6.7 | 0.47 |
| polarC18 | 218.139 | 50.6127 | 6.7 | 0.47 |
| polarC18 | 218.139 | 59.7979 | 6.7 | 0.47 |
| polarC18 | 218.139 | 74.7978 | 6.7 | 0.47 |
| polarC18 | 218.139 | 50.4433 | 6.7 | 0.47 |
| polarC18 | 218.139 | 92.1827 | 6.7 | 0.47 |
| polarC18 | 218.139 | 126.6283 | 6.7 | 0.47 |
| polarC18 | 218.139 | 148.8658 | 6.7 | 0.47 |
| polarC18 | 218.139 | 65.1128 | 6.7 | 0.47 |
| polarC18 | 218.139 | 79.2283 | 6.7 | 0.47 |
| polarC18 | 218.139 | 131.9565 | 6.7 | 0.47 |
| polarC18 | 218.139 | 242.2552 | 6.7 | 0.47 |
| polarC18 | 218.139 | 53.509 | 6.7 | 0.47 |
| polarC18 | 218.139 | 74.0518 | 6.7 | 0.47 |
| polarC18 | 218.139 | 109.1117 | 6.7 | 0.47 |
| polarC18 | 218.139 | 61.0789 | 6.7 | 0.47 |
| polarC18 | 218.139 | 197.4525 | 6.7 | 0.47 |
| polarC18 | 218.139 | 61.4866 | 6.7 | 0.47 |
| polarC18 | 218.139 | 50.1588 | 6.7 | 0.47 |
| polarC18 | 218.139 | 50.1945 | 6.7 | 0.47 |
| polarC18 | 218.139 | 55.5629 | 6.7 | 0.47 |
| polarC18 | 218.139 | 59.2533 | 6.7 | 0.47 |
| polarC18 | 218.139 | 135.3132 | 6.7 | 0.47 |
| polarC18 | 218.139 | 200.4354 | 6.7 | 0.47 |
| polarC18 | 218.139 | 137.5663 | 6.7 | 0.47 |
| polarC18 | 218.139 | 220.5912 | 6.7 | 0.47 |
| polarC18 | 218.139 | 57.5659 | 6.7 | 0.47 |
| polarC18 | 218.139 | 151.1489 | 6.7 | 0.47 |
| polarC18 | 218.139 | 57.3166 | 6.7 | 0.47 |
| polarC18 | 218.139 | 140.5686 | 6.7 | 0.47 |
| polarC18 | 218.139 | 49.9749 | 6.7 | 0.47 |
| polarC18 | 218.139 | 72.5955 | 6.7 | 0.47 |

| | | | | |
|---|---|---|---|---|
| polarC18 | 218.139 | 54.4125 | 6.7 | 0.47 |
| polarC18 | 218.139 | 70.3341 | 6.7 | 0.47 |
| polarC18 | 218.139 | 64.4406 | 6.7 | 0.47 |
| polarC18 | 218.139 | 49.7452 | 6.7 | 0.47 |
| polarC18 | 218.139 | 236.1378 | 6.7 | 0.47 |
| polarC18 | 218.139 | 102.6854 | 6.7 | 0.47 |
| polarC18 | 218.139 | 51.8026 | 6.7 | 0.47 |
| polarC18 | 218.139 | 86.0858 | 6.7 | 0.47 |
| polarC18 | 218.139 | 224.7586 | 6.7 | 0.47 |
| polarC18 | 218.139 | 79.3958 | 6.7 | 0.47 |
| polarC18 | 218.139 | 77.5811 | 6.7 | 0.47 |
| polarC18 | 218.139 | 64.2485 | 6.7 | 0.47 |
| polarC18 | 218.139 | 136.3273 | 6.7 | 0.47 |
| polarC18 | 218.139 | 53.0296 | 6.7 | 0.47 |
| polarC18 | 218.139 | 75.1322 | 6.6 | 0.47 |
| polarC18 | 218.139 | 52.9808 | 6.6 | 0.47 |
| polarC18 | 218.139 | 102.7511 | 6.6 | 0.47 |
| polarC18 | 218.139 | 57.6141 | 6.6 | 0.47 |
| polarC18 | 218.139 | 84.2322 | 6.6 | 0.47 |
| polarC18 | 218.139 | 80.1406 | 6.6 | 0.47 |
| polarC18 | 218.139 | 65.7513 | 6.6 | 0.47 |
| polarC18 | 218.139 | 57.7615 | 6.6 | 0.47 |
| polarC18 | 218.139 | 138.5796 | 6.6 | 0.47 |
| polarC18 | 218.139 | 105.9419 | 6.6 | 0.47 |
| polarC18 | 218.139 | 148.0516 | 6.6 | 0.47 |
| polarC18 | 218.139 | 177.0656 | 6.6 | 0.47 |
| polarC18 | 218.139 | 75.8088 | 6.6 | 0.47 |
| polarC18 | 218.139 | 69.5056 | 6.6 | 0.47 |
| polarC18 | 218.139 | 76.7534 | 6.6 | 0.47 |
| polarC18 | 218.139 | 70.7762 | 6.6 | 0.47 |
| polarC18 | 218.139 | 50.0524 | 6.6 | 0.47 |
| polarC18 | 218.139 | 102.0983 | 6.6 | 0.47 |
| polarC18 | 218.139 | 54.2305 | 6.6 | 0.47 |
| polarC18 | 218.139 | 135.3986 | 6.6 | 0.47 |
| polarC18 | 218.139 | 160.5164 | 6.6 | 0.47 |
| polarC18 | 218.139 | 212.3461 | 6.6 | 0.47 |
| polarC18 | 218.139 | 120.8677 | 6.6 | 0.47 |
| polarC18 | 218.139 | 242.0385 | 6.6 | 0.47 |
| polarC18 | 218.139 | 156.7043 | 6.6 | 0.47 |
| polarC18 | 218.139 | 72.8718 | 6.6 | 0.47 |
| polarC18 | 218.139 | 88.9321 | 6.6 | 0.47 |
| polarC18 | 218.139 | 48.9421 | 6.6 | 0.47 |

| | | | | |
|---|---|---|---|---|
| polarC18 | 218.139 | 73.3897 | 6.6 | 0.47 |
| polarC18 | 218.139 | 140.2582 | 6.6 | 0.47 |
| polarC18 | 218.139 | 133.303 | 6.6 | 0.47 |
| polarC18 | 218.139 | 56.4256 | 6.6 | 0.47 |
| polarC18 | 218.139 | 194.1818 | 6.6 | 0.47 |
| polarC18 | 218.139 | 67.9053 | 6.6 | 0.47 |
| polarC18 | 218.139 | 147.1022 | 6.6 | 0.47 |
| polarC18 | 218.139 | 62.9305 | 6.6 | 0.47 |
| polarC18 | 218.139 | 52.3328 | 6.6 | 0.47 |
| polarC18 | 218.139 | 86.0579 | 6.6 | 0.47 |
| polarC18 | 218.139 | 216.58 | 6.6 | 0.47 |
| polarC18 | 218.139 | 89.6075 | 6.6 | 0.47 |
| polarC18 | 218.139 | 141.4356 | 6.6 | 0.47 |
| polarC18 | 218.139 | 103.1487 | 6.6 | 0.47 |
| polarC18 | 218.139 | 60.1747 | 6.6 | 0.47 |
| polarC18 | 218.139 | 200.0611 | 6.6 | 0.47 |
| polarC18 | 218.139 | 170.6799 | 6.6 | 0.47 |
| polarC18 | 218.139 | 213.2466 | 6.6 | 0.47 |
| polarC18 | 218.139 | 89.1856 | 6.6 | 0.47 |
| polarC18 | 218.139 | 63.8442 | 6.6 | 0.47 |
| polarC18 | 218.139 | 124.5816 | 6.6 | 0.47 |
| polarC18 | 218.139 | 55.2239 | 6.6 | 0.47 |
| polarC18 | 218.139 | 53.5345 | 6.6 | 0.47 |
| polarC18 | 218.139 | 124.7737 | 6.6 | 0.47 |
| polarC18 | 218.139 | 57.4659 | 6.6 | 0.47 |
| polarC18 | 218.139 | 60.8114 | 6.6 | 0.47 |
| polarC18 | 218.139 | 115.3987 | 6.6 | 0.47 |
| polarC18 | 218.139 | 161.8385 | 6.6 | 0.47 |
| polarC18 | 218.139 | 109.8538 | 6.6 | 0.47 |
| polarC18 | 218.139 | 54.3501 | 6.6 | 0.47 |
| polarC18 | 218.139 | 50.2554 | 6.6 | 0.47 |
| polarC18 | 218.139 | 63.5688 | 6.6 | 0.47 |
| polarC18 | 218.139 | 155.7803 | 6.6 | 0.47 |
| polarC18 | 218.139 | 137.4562 | 6.6 | 0.47 |
| polarC18 | 218.139 | 88.1599 | 6.6 | 0.47 |
| polarC18 | 218.139 | 50.5582 | 6.6 | 0.47 |
| polarC18 | 218.139 | 147.2178 | 6.6 | 0.47 |
| polarC18 | 218.139 | 57.2058 | 6.6 | 0.47 |
| polarC18 | 218.139 | 49.9807 | 6.6 | 0.47 |
| polarC18 | 218.139 | 94.8174 | 6.6 | 0.47 |
| polarC18 | 218.139 | 83.4404 | 6.6 | 0.47 |
| polarC18 | 218.139 | 67.5983 | 6.6 | 0.47 |

| | | | | |
|---|---|---|---|---|
| polarC18 | 218.139 | 191.1357 | 6.6 | 0.47 |
| polarC18 | 218.139 | 90.2706 | 6.6 | 0.47 |
| polarC18 | 218.139 | 54.0703 | 6.6 | 0.47 |
| polarC18 | 218.139 | 111.4453 | 6.6 | 0.47 |
| polarC18 | 218.139 | 133.6896 | 6.6 | 0.47 |
| polarC18 | 218.139 | 174.3561 | 6.6 | 0.47 |
| polarC18 | 218.139 | 175.6268 | 6.6 | 0.47 |
| polarC18 | 218.139 | 82.8633 | 6.6 | 0.47 |
| polarC18 | 218.139 | 105.7621 | 6.6 | 0.47 |
| polarC18 | 218.139 | 60.6353 | 6.6 | 0.47 |
| polarC18 | 218.139 | 172.5855 | 6.6 | 0.47 |
| polarC18 | 218.139 | 77.5391 | 6.6 | 0.47 |
| polarC18 | 218.139 | 101.3306 | 6.6 | 0.47 |
| polarC18 | 218.139 | 90.2492 | 6.6 | 0.47 |
| polarC18 | 218.139 | 52.2544 | 6.6 | 0.47 |
| polarC18 | 218.139 | 88.6889 | 6.6 | 0.47 |
| polarC18 | 218.139 | 227.8293 | 6.6 | 0.47 |
| polarC18 | 218.139 | 63.5587 | 6.6 | 0.47 |
| polarC18 | 218.139 | 74.4442 | 6.6 | 0.47 |
| polarC18 | 218.139 | 86.0513 | 6.6 | 0.47 |
| polarC18 | 218.139 | 65.2948 | 6.6 | 0.47 |
| polarC18 | 218.139 | 99.3734 | 6.6 | 0.47 |
| polarC18 | 218.139 | 161.404 | 6.6 | 0.47 |
| polarC18 | 218.139 | 73.9973 | 6.6 | 0.47 |
| polarC18 | 218.139 | 166.8682 | 6.6 | 0.47 |
| polarC18 | 218.139 | 147.0343 | 6.6 | 0.47 |
| polarC18 | 218.139 | 166.1286 | 6.6 | 0.47 |
| polarC18 | 218.139 | 92.8055 | 6.6 | 0.47 |
| polarC18 | 218.139 | 110.4441 | 6.6 | 0.47 |
| polarC18 | 218.139 | 151.6259 | 6.6 | 0.47 |
| polarC18 | 218.139 | 49.4725 | 6.6 | 0.47 |
| polarC18 | 218.139 | 104.5002 | 6.6 | 0.47 |
| polarC18 | 218.139 | 64.4935 | 6.6 | 0.47 |
| polarC18 | 218.139 | 60.352 | 6.6 | 0.47 |
| polarC18 | 218.139 | 73.6547 | 6.6 | 0.47 |
| polarC18 | 218.139 | 124.0956 | 6.6 | 0.47 |
| polarC18 | 218.139 | 65.6348 | 6.6 | 0.47 |
| polarC18 | 218.139 | 55.2962 | 6.6 | 0.47 |
| polarC18 | 218.139 | 223.688 | 6.6 | 0.47 |
| polarC18 | 218.139 | 173.9213 | 6.6 | 0.47 |
| polarC18 | 218.139 | 161.6114 | 6.6 | 0.47 |
| polarC18 | 218.139 | 70.1932 | 6.6 | 0.47 |

| | | | | |
|---|---|---|---|---|
| polarC18 | 218.139 | 73.9703 | 6.6 | 0.47 |
| polarC18 | 218.139 | 70.8409 | 6.6 | 0.47 |
| polarC18 | 218.139 | 52.0984 | 6.6 | 0.47 |
| polarC18 | 218.139 | 83.1701 | 6.6 | 0.47 |
| polarC18 | 218.139 | 226.3957 | 6.6 | 0.47 |
| polarC18 | 218.139 | 94.2371 | 6.6 | 0.47 |
| polarC18 | 218.139 | 55.2392 | 6.6 | 0.47 |
| polarC18 | 218.139 | 109.4352 | 6.6 | 0.47 |
| polarC18 | 218.139 | 231.6879 | 6.6 | 0.47 |
| polarC18 | 218.139 | 224.9216 | 6.6 | 0.47 |
| polarC18 | 218.139 | 50.6292 | 6.6 | 0.47 |
| polarC18 | 218.139 | 89.3901 | 6.6 | 0.47 |
| polarC18 | 218.139 | 81.5412 | 6.6 | 0.47 |
| polarC18 | 218.139 | 55.8324 | 6.6 | 0.47 |
| polarC18 | 218.139 | 108.6055 | 6.6 | 0.47 |
| polarC18 | 218.139 | 48.815 | 6.6 | 0.47 |
| polarC18 | 218.139 | 49.7831 | 6.6 | 0.47 |
| polarC18 | 218.139 | 66.5066 | 6.6 | 0.47 |
| polarC18 | 218.139 | 60.0055 | 6.6 | 0.47 |
| polarC18 | 218.139 | 121.4552 | 6.6 | 0.47 |
| polarC18 | 218.139 | 194.2173 | 6.6 | 0.47 |
| polarC18 | 218.139 | 231.8792 | 6.6 | 0.47 |
| polarC18 | 218.139 | 191.7803 | 6.6 | 0.47 |
| polarC18 | 218.139 | 91.457 | 6.6 | 0.47 |
| polarC18 | 218.139 | 101.3662 | 6.6 | 0.47 |
| polarC18 | 218.139 | 113.4784 | 6.6 | 0.47 |
| polarC18 | 218.139 | 67.2955 | 6.6 | 0.47 |
| polarC18 | 218.139 | 59.6554 | 6.6 | 0.47 |
| polarC18 | 218.139 | 93.0713 | 6.6 | 0.47 |
| polarC18 | 218.139 | 87.1801 | 6.6 | 0.47 |
| polarC18 | 218.139 | 65.3754 | 6.6 | 0.47 |
| polarC18 | 218.139 | 81.3037 | 6.6 | 0.47 |
| polarC18 | 218.139 | 67.2996 | 6.6 | 0.47 |
| polarC18 | 218.139 | 70.8521 | 6.6 | 0.47 |
| polarC18 | 218.139 | 51.3043 | 6.6 | 0.47 |
| polarC18 | 218.139 | 92.6154 | 6.6 | 0.47 |
| polarC18 | 218.139 | 67.1914 | 6.6 | 0.47 |
| polarC18 | 218.139 | 52.9587 | 6.6 | 0.47 |
| polarC18 | 218.139 | 150.4366 | 6.6 | 0.47 |
| polarC18 | 218.139 | 93.8549 | 6.6 | 0.47 |
| polarC18 | 218.139 | 188.4493 | 6.6 | 0.47 |
| polarC18 | 218.139 | 58.5573 | 6.6 | 0.47 |

| | | | | |
|---|---|---|---|---|
| polarC18 | 218.139 | 211.481 | 6.6 | 0.47 |
| polarC18 | 218.139 | 53.6404 | 6.6 | 0.47 |
| polarC18 | 218.139 | 77.3566 | 6.6 | 0.47 |
| polarC18 | 218.139 | 78.7245 | 6.6 | 0.47 |
| polarC18 | 218.139 | 94.7458 | 6.6 | 0.47 |
| polarC18 | 218.139 | 115.3249 | 6.6 | 0.47 |
| polarC18 | 218.139 | 152.0871 | 6.6 | 0.47 |
| polarC18 | 218.139 | 57.0292 | 6.6 | 0.47 |
| polarC18 | 218.139 | 64.2369 | 6.6 | 0.47 |
| polarC18 | 218.139 | 51.647 | 6.6 | 0.47 |
| polarC18 | 218.139 | 56.6878 | 6.6 | 0.47 |
| polarC18 | 218.139 | 80.9179 | 6.6 | 0.47 |
| polarC18 | 218.139 | 75.4897 | 6.6 | 0.47 |
| polarC18 | 218.139 | 75.5758 | 6.6 | 0.47 |
| polarC18 | 218.139 | 139.0672 | 6.6 | 0.47 |
| polarC18 | 218.139 | 102.8166 | 6.6 | 0.47 |
| polarC18 | 218.139 | 58.7652 | 6.6 | 0.47 |
| polarC18 | 218.139 | 128.7601 | 6.6 | 0.47 |
| polarC18 | 218.139 | 111.8975 | 6.6 | 0.47 |
| polarC18 | 218.139 | 61.6462 | 6.6 | 0.47 |
| polarC18 | 218.139 | 70.3169 | 6.6 | 0.47 |
| polarC18 | 218.139 | 64.0261 | 6.6 | 0.47 |
| polarC18 | 218.139 | 49.1786 | 6.6 | 0.47 |
| polarC18 | 218.139 | 81.451 | 6.6 | 0.47 |
| polarC18 | 218.139 | 65.208 | 6.6 | 0.47 |
| polarC18 | 218.139 | 180.7003 | 6.6 | 0.47 |
| polarC18 | 218.139 | 53.0618 | 6.6 | 0.47 |
| polarC18 | 218.139 | 222.0417 | 6.6 | 0.47 |
| polarC18 | 218.139 | 63.0541 | 6.6 | 0.47 |
| polarC18 | 218.139 | 72.147 | 6.6 | 0.47 |
| polarC18 | 218.139 | 65.042 | 6.6 | 0.47 |
| polarC18 | 218.139 | 92.199 | 6.6 | 0.47 |
| polarC18 | 218.139 | 106.9269 | 6.6 | 0.47 |
| polarC18 | 218.139 | 90.9719 | 6.6 | 0.47 |
| polarC18 | 218.139 | 169.8373 | 6.6 | 0.47 |
| polarC18 | 218.139 | 108.0744 | 6.6 | 0.47 |
| polarC18 | 218.139 | 49.2073 | 6.6 | 0.47 |
| polarC18 | 218.139 | 107.851 | 6.6 | 0.47 |
| polarC18 | 218.139 | 59.946 | 6.6 | 0.47 |
| polarC18 | 218.139 | 77.4771 | 6.6 | 0.47 |
| polarC18 | 218.139 | 184.5986 | 6.6 | 0.47 |
| polarC18 | 218.139 | 114.0612 | 6.6 | 0.47 |

| | | | | |
|---|---|---|---|---|
| polarC18 | 218.139 | 100.3273 | 6.6 | 0.47 |
| polarC18 | 218.139 | 222.9505 | 6.6 | 0.47 |
| polarC18 | 218.139 | 57.3922 | 6.6 | 0.47 |
| polarC18 | 218.139 | 62.7918 | 6.6 | 0.47 |
| polarC18 | 218.139 | 62.2128 | 6.6 | 0.47 |
| polarC18 | 218.139 | 83.4883 | 6.6 | 0.47 |
| polarC18 | 218.139 | 68.9039 | 6.6 | 0.47 |
| polarC18 | 218.139 | 70.9488 | 6.6 | 0.47 |
| polarC18 | 218.139 | 56.6234 | 6.6 | 0.47 |
| polarC18 | 218.139 | 59.8041 | 6.6 | 0.47 |
| polarC18 | 218.139 | 76.6775 | 6.6 | 0.47 |
| polarC18 | 218.139 | 169.8658 | 6.6 | 0.47 |
| polarC18 | 218.139 | 64.4026 | 6.6 | 0.47 |
| polarC18 | 218.139 | 208.7456 | 6.6 | 0.47 |
| polarC18 | 218.139 | 51.479 | 6.6 | 0.47 |
| polarC18 | 218.139 | 84.6603 | 6.6 | 0.47 |
| polarC18 | 218.139 | 56.823 | 6.6 | 0.47 |
| polarC18 | 218.139 | 94.4429 | 6.6 | 0.47 |
| polarC18 | 218.139 | 70.7235 | 6.6 | 0.47 |
| polarC18 | 218.139 | 103.3278 | 6.6 | 0.47 |
| polarC18 | 218.139 | 116.7156 | 6.6 | 0.47 |
| polarC18 | 218.139 | 151.6472 | 6.6 | 0.47 |
| polarC18 | 218.139 | 76.185 | 6.6 | 0.47 |
| polarC18 | 218.139 | 163.6777 | 6.6 | 0.47 |
| polarC18 | 218.139 | 52.851 | 6.6 | 0.47 |
| polarC18 | 218.139 | 62.9625 | 6.6 | 0.47 |
| polarC18 | 218.139 | 161.3029 | 6.6 | 0.47 |
| polarC18 | 218.139 | 149.5196 | 6.6 | 0.47 |
| polarC18 | 218.139 | 94.7037 | 6.6 | 0.47 |
| polarC18 | 218.139 | 88.8707 | 6.6 | 0.47 |
| polarC18 | 218.139 | 51.7091 | 6.6 | 0.47 |
| polarC18 | 218.139 | 120.884 | 6.6 | 0.47 |
| polarC18 | 218.139 | 52.8619 | 6.6 | 0.47 |
| polarC18 | 218.139 | 84.4065 | 6.6 | 0.47 |
| polarC18 | 218.139 | 81.496 | 6.6 | 0.47 |
| polarC18 | 218.139 | 64.9345 | 6.6 | 0.47 |
| polarC18 | 218.139 | 90.0171 | 6.6 | 0.47 |
| polarC18 | 218.139 | 173.7156 | 6.6 | 0.47 |
| polarC18 | 218.139 | 187.7943 | 6.6 | 0.47 |
| polarC18 | 218.139 | 62.4154 | 6.6 | 0.47 |
| polarC18 | 218.139 | 100.6532 | 6.6 | 0.47 |
| polarC18 | 218.139 | 75.5668 | 6.6 | 0.47 |

| | | | | |
|---|---|---|---|---|
| polarC18 | 218.139 | 52.0037 | 6.6 | 0.47 |
| polarC18 | 218.139 | 70.6651 | 6.6 | 0.47 |
| polarC18 | 218.139 | 49.9301 | 6.6 | 0.47 |
| polarC18 | 218.139 | 133.5623 | 6.6 | 0.47 |
| polarC18 | 218.139 | 88.9977 | 6.6 | 0.47 |
| polarC18 | 218.139 | 49.4551 | 6.6 | 0.47 |
| polarC18 | 218.139 | 50.3298 | 6.6 | 0.47 |
| polarC18 | 218.139 | 52.5663 | 6.6 | 0.47 |
| polarC18 | 218.139 | 77.9241 | 6.6 | 0.47 |
| polarC18 | 218.139 | 49.4617 | 6.6 | 0.47 |
| polarC18 | 218.139 | 52.143 | 6.6 | 0.47 |
| polarC18 | 218.139 | 101.6562 | 6.6 | 0.47 |
| polarC18 | 218.139 | 222.3885 | 6.6 | 0.47 |
| polarC18 | 218.139 | 62.612 | 6.6 | 0.47 |
| polarC18 | 218.139 | 136.9542 | 6.6 | 0.47 |
| polarC18 | 218.139 | 51.1904 | 6.6 | 0.47 |
| polarC18 | 218.139 | 49.9103 | 6.6 | 0.47 |
| polarC18 | 218.139 | 50.0487 | 6.6 | 0.47 |
| polarC18 | 218.139 | 60.6177 | 6.6 | 0.47 |
| polarC18 | 218.139 | 72.4058 | 6.6 | 0.47 |
| polarC18 | 218.139 | 56.2386 | 6.6 | 0.47 |
| polarC18 | 218.139 | 56.4163 | 6.6 | 0.47 |
| polarC18 | 218.139 | 54.2126 | 6.6 | 0.47 |
| polarC18 | 218.139 | 76.3849 | 6.6 | 0.47 |
| polarC18 | 218.139 | 70.1403 | 6.6 | 0.47 |
| polarC18 | 218.139 | 82.3412 | 6.6 | 0.47 |
| polarC18 | 218.139 | 72.4903 | 6.6 | 0.47 |
| polarC18 | 218.139 | 62.3414 | 6.6 | 0.47 |
| polarC18 | 218.139 | 92.4663 | 6.6 | 0.47 |
| polarC18 | 218.139 | 183.3399 | 6.6 | 0.47 |
| polarC18 | 218.139 | 174.4181 | 6.6 | 0.47 |
| polarC18 | 218.139 | 189.7664 | 6.6 | 0.47 |
| polarC18 | 218.139 | 62.0196 | 6.6 | 0.47 |
| polarC18 | 218.139 | 84.8028 | 6.6 | 0.47 |
| polarC18 | 218.139 | 71.6923 | 6.6 | 0.47 |
| polarC18 | 218.139 | 68.553 | 6.6 | 0.47 |
| polarC18 | 218.139 | 225.5311 | 6.6 | 0.46 |
| polarC18 | 218.139 | 111.0723 | 6.6 | 0.46 |
| polarC18 | 218.139 | 109.2165 | 6.6 | 0.46 |
| polarC18 | 218.139 | 49.1238 | 6.6 | 0.46 |
| polarC18 | 218.139 | 138.8361 | 6.6 | 0.46 |
| polarC18 | 218.139 | 84.1354 | 6.6 | 0.46 |

| | | | | |
|---|---|---|---|---|
| polarC18 | 218.139 | 58.2766 | 6.6 | 0.46 |
| polarC18 | 218.139 | 231.7265 | 6.6 | 0.46 |
| polarC18 | 218.139 | 103.763 | 6.6 | 0.46 |
| polarC18 | 218.139 | 236.2372 | 6.6 | 0.46 |
| polarC18 | 218.139 | 157.998 | 6.6 | 0.46 |
| polarC18 | 218.139 | 139.3936 | 6.5 | 0.46 |
| polarC18 | 218.139 | 61.1977 | 6.5 | 0.46 |
| polarC18 | 218.139 | 54.3269 | 6.5 | 0.46 |
| polarC18 | 218.139 | 110.6213 | 6.5 | 0.46 |
| polarC18 | 218.139 | 57.6869 | 6.5 | 0.46 |
| polarC18 | 218.139 | 87.2484 | 6.5 | 0.46 |
| polarC18 | 218.139 | 110.5996 | 6.5 | 0.46 |
| polarC18 | 218.139 | 102.5017 | 6.5 | 0.46 |
| polarC18 | 218.139 | 75.4717 | 6.5 | 0.46 |
| polarC18 | 218.139 | 139.1999 | 6.5 | 0.46 |
| polarC18 | 218.139 | 113.054 | 6.5 | 0.46 |
| polarC18 | 218.139 | 242.9094 | 6.5 | 0.46 |
| polarC18 | 218.139 | 111.4614 | 6.5 | 0.46 |
| polarC18 | 218.139 | 146.8772 | 6.5 | 0.46 |
| polarC18 | 218.139 | 239.1263 | 6.5 | 0.46 |
| polarC18 | 218.139 | 54.4224 | 6.5 | 0.46 |
| polarC18 | 218.139 | 76.1182 | 6.5 | 0.46 |
| polarC18 | 218.139 | 166.6287 | 6.5 | 0.46 |
| polarC18 | 218.139 | 61.8342 | 6.5 | 0.46 |
| polarC18 | 218.139 | 56.061 | 6.5 | 0.46 |
| polarC18 | 218.139 | 60.2613 | 6.5 | 0.46 |
| polarC18 | 218.139 | 56.2311 | 6.5 | 0.46 |
| polarC18 | 218.139 | 112.5567 | 6.5 | 0.46 |
| polarC18 | 218.139 | 143.7888 | 6.5 | 0.46 |
| polarC18 | 218.139 | 57.9342 | 6.5 | 0.46 |
| polarC18 | 218.139 | 172.525 | 6.5 | 0.46 |
| polarC18 | 218.139 | 93.1692 | 6.5 | 0.46 |
| polarC18 | 218.139 | 62.9811 | 6.5 | 0.46 |
| polarC18 | 218.139 | 49.6132 | 6.5 | 0.46 |
| polarC18 | 218.139 | 90.6227 | 6.5 | 0.46 |
| polarC18 | 218.139 | 178.5813 | 6.5 | 0.46 |
| polarC18 | 218.139 | 126.1907 | 6.5 | 0.46 |
| polarC18 | 218.139 | 73.7787 | 6.5 | 0.46 |
| polarC18 | 218.139 | 50.924 | 6.5 | 0.46 |
| polarC18 | 218.139 | 53.7268 | 6.5 | 0.46 |
| polarC18 | 218.139 | 89.054 | 6.5 | 0.46 |
| polarC18 | 218.139 | 127.1891 | 6.5 | 0.46 |

| | | | | |
|---|---|---|---|---|
| polarC18 | 218.139 | 136.3909 | 6.5 | 0.46 |
| polarC18 | 218.139 | 50.3543 | 6.5 | 0.46 |
| polarC18 | 218.139 | 72.3134 | 6.5 | 0.46 |
| polarC18 | 218.139 | 108.437 | 6.5 | 0.46 |
| polarC18 | 218.139 | 109.8621 | 6.5 | 0.46 |
| polarC18 | 218.139 | 227.3837 | 6.5 | 0.46 |
| polarC18 | 218.139 | 80.9331 | 6.5 | 0.46 |
| polarC18 | 218.139 | 78.5808 | 6.5 | 0.46 |
| polarC18 | 218.139 | 80.5543 | 6.5 | 0.46 |
| polarC18 | 218.139 | 51.6561 | 6.5 | 0.46 |
| polarC18 | 218.139 | 83.3016 | 6.5 | 0.46 |
| polarC18 | 218.139 | 61.7811 | 6.5 | 0.46 |
| polarC18 | 218.139 | 59.2598 | 6.5 | 0.46 |
| polarC18 | 218.139 | 110.2963 | 6.5 | 0.46 |
| polarC18 | 218.139 | 50.9833 | 6.5 | 0.46 |
| polarC18 | 218.139 | 71.0173 | 6.5 | 0.46 |
| polarC18 | 218.139 | 80.6252 | 6.5 | 0.46 |
| polarC18 | 218.139 | 50.317 | 6.5 | 0.46 |
| polarC18 | 218.139 | 65.3513 | 6.5 | 0.46 |
| polarC18 | 218.139 | 78.2731 | 6.5 | 0.46 |
| polarC18 | 218.139 | 64.5133 | 6.5 | 0.46 |
| polarC18 | 218.139 | 155.5239 | 6.5 | 0.46 |
| polarC18 | 218.139 | 128.8978 | 6.5 | 0.46 |
| polarC18 | 218.139 | 66.4358 | 6.5 | 0.46 |
| polarC18 | 218.139 | 50.6771 | 6.5 | 0.46 |
| polarC18 | 218.139 | 71.5069 | 6.5 | 0.46 |
| polarC18 | 218.139 | 70.6364 | 6.5 | 0.46 |
| polarC18 | 218.139 | 168.4944 | 6.5 | 0.46 |
| polarC18 | 218.139 | 193.4267 | 6.5 | 0.46 |
| polarC18 | 218.139 | 90.4987 | 6.5 | 0.46 |
| polarC18 | 218.139 | 115.9266 | 6.5 | 0.46 |
| polarC18 | 218.139 | 75.3908 | 6.5 | 0.46 |
| polarC18 | 218.139 | 67.4115 | 6.5 | 0.46 |
| polarC18 | 218.139 | 80.9606 | 6.5 | 0.46 |
| polarC18 | 218.139 | 89.246 | 6.5 | 0.46 |
| polarC18 | 218.139 | 121.5634 | 6.5 | 0.46 |
| polarC18 | 218.139 | 93.9273 | 6.5 | 0.46 |
| polarC18 | 218.139 | 86.1716 | 6.5 | 0.46 |
| polarC18 | 218.139 | 55.7342 | 6.5 | 0.46 |
| polarC18 | 218.139 | 130.3141 | 6.5 | 0.46 |
| polarC18 | 218.139 | 87.7437 | 6.5 | 0.46 |
| polarC18 | 218.139 | 72.5986 | 6.5 | 0.46 |

| | | | | |
|---|---|---|---|---|
| polarC18 | 218.139 | 154.5006 | 6.5 | 0.46 |
| polarC18 | 218.139 | 74.1385 | 6.5 | 0.46 |
| polarC18 | 218.139 | 82.1561 | 6.5 | 0.46 |
| polarC18 | 218.139 | 122.1137 | 6.5 | 0.46 |
| polarC18 | 218.139 | 73.1887 | 6.5 | 0.46 |
| polarC18 | 218.139 | 49.4872 | 6.5 | 0.46 |
| polarC18 | 218.139 | 96.0602 | 6.5 | 0.46 |
| polarC18 | 218.139 | 78.5906 | 6.5 | 0.46 |
| polarC18 | 218.139 | 92.6423 | 6.5 | 0.46 |
| polarC18 | 218.139 | 85.9335 | 6.5 | 0.46 |
| polarC18 | 218.139 | 102.9576 | 6.5 | 0.46 |
| polarC18 | 218.139 | 63.872 | 6.5 | 0.46 |
| polarC18 | 218.139 | 54.6597 | 6.5 | 0.46 |
| polarC18 | 218.139 | 235.6031 | 6.5 | 0.46 |
| polarC18 | 218.139 | 85.7231 | 6.5 | 0.46 |
| polarC18 | 218.139 | 63.4347 | 6.5 | 0.46 |
| polarC18 | 218.139 | 86.4353 | 6.5 | 0.46 |
| polarC18 | 218.139 | 170.0372 | 6.5 | 0.46 |
| polarC18 | 218.139 | 70.4269 | 6.5 | 0.46 |
| polarC18 | 218.139 | 172.2208 | 6.5 | 0.46 |
| polarC18 | 218.139 | 88.1687 | 6.5 | 0.46 |
| polarC18 | 218.139 | 58.2371 | 6.5 | 0.46 |
| polarC18 | 218.139 | 68.6348 | 6.5 | 0.46 |
| polarC18 | 218.139 | 78.1753 | 6.5 | 0.46 |
| polarC18 | 218.139 | 85.6527 | 6.5 | 0.46 |
| polarC18 | 218.139 | 211.7693 | 6.5 | 0.46 |
| polarC18 | 218.139 | 140.9475 | 6.5 | 0.46 |
| polarC18 | 218.139 | 107.2862 | 6.5 | 0.46 |
| polarC18 | 218.139 | 203.4222 | 6.5 | 0.46 |
| polarC18 | 218.139 | 104.3871 | 6.5 | 0.46 |
| polarC18 | 218.139 | 195.1475 | 6.5 | 0.46 |
| polarC18 | 218.139 | 213.0055 | 6.5 | 0.46 |
| polarC18 | 218.139 | 70.6458 | 6.5 | 0.46 |
| polarC18 | 218.139 | 59.854 | 6.5 | 0.46 |
| polarC18 | 218.139 | 104.1306 | 6.5 | 0.46 |
| polarC18 | 218.139 | 194.7114 | 6.5 | 0.46 |
| polarC18 | 218.139 | 48.7091 | 6.5 | 0.46 |
| polarC18 | 218.139 | 50.3037 | 6.5 | 0.46 |
| polarC18 | 218.139 | 83.2595 | 6.5 | 0.46 |
| polarC18 | 218.139 | 113.8839 | 6.5 | 0.46 |
| polarC18 | 218.139 | 51.7862 | 6.5 | 0.46 |
| polarC18 | 218.139 | 60.7956 | 6.5 | 0.46 |

| | | | | |
|---|---|---|---|---|
| polarC18 | 218.139 | 143.1411 | 6.5 | 0.46 |
| polarC18 | 218.139 | 77.4328 | 6.5 | 0.46 |
| polarC18 | 218.139 | 49.0647 | 6.5 | 0.46 |
| polarC18 | 218.139 | 122.0972 | 6.5 | 0.46 |
| polarC18 | 218.139 | 96.2866 | 6.5 | 0.46 |
| polarC18 | 218.139 | 57.1824 | 6.5 | 0.46 |
| polarC18 | 218.139 | 52.9394 | 6.5 | 0.46 |
| polarC18 | 218.139 | 65.153 | 6.5 | 0.46 |
| polarC18 | 218.139 | 71.8537 | 6.5 | 0.46 |
| polarC18 | 218.139 | 78.5747 | 6.5 | 0.46 |
| polarC18 | 218.139 | 99.717 | 6.5 | 0.46 |
| polarC18 | 218.139 | 89.9358 | 6.5 | 0.46 |
| polarC18 | 218.139 | 63.6656 | 6.5 | 0.46 |
| polarC18 | 218.139 | 205.3725 | 6.5 | 0.46 |
| polarC18 | 218.139 | 55.5779 | 6.5 | 0.46 |
| polarC18 | 218.139 | 77.3999 | 6.5 | 0.46 |
| polarC18 | 218.139 | 49.8259 | 6.5 | 0.46 |
| polarC18 | 218.139 | 51.821 | 6.5 | 0.46 |
| polarC18 | 218.139 | 50.1141 | 6.5 | 0.46 |
| polarC18 | 218.139 | 122.79 | 6.5 | 0.46 |
| polarC18 | 218.139 | 154.0538 | 6.5 | 0.46 |
| polarC18 | 218.139 | 146.1227 | 6.5 | 0.46 |
| polarC18 | 218.139 | 119.8438 | 6.5 | 0.46 |
| polarC18 | 218.139 | 107.5211 | 6.5 | 0.46 |
| polarC18 | 218.139 | 56.2365 | 6.5 | 0.46 |
| polarC18 | 218.139 | 167.6243 | 6.5 | 0.46 |
| polarC18 | 218.139 | 206.464 | 6.5 | 0.46 |
| polarC18 | 218.139 | 53.2227 | 6.5 | 0.46 |
| polarC18 | 218.139 | 127.8068 | 6.5 | 0.46 |
| polarC18 | 218.139 | 77.3459 | 6.5 | 0.46 |
| polarC18 | 218.139 | 60.7709 | 6.5 | 0.46 |
| polarC18 | 218.139 | 58.4645 | 6.5 | 0.46 |
| polarC18 | 218.139 | 146.179 | 6.5 | 0.46 |
| polarC18 | 218.139 | 48.9467 | 6.5 | 0.46 |
| polarC18 | 218.139 | 139.3803 | 6.5 | 0.46 |
| polarC18 | 218.139 | 88.2433 | 6.5 | 0.46 |
| polarC18 | 218.139 | 85.5671 | 6.5 | 0.46 |
| polarC18 | 218.139 | 208.1821 | 6.5 | 0.46 |
| polarC18 | 218.139 | 52.1011 | 6.5 | 0.46 |
| polarC18 | 218.139 | 90.1267 | 6.5 | 0.46 |
| polarC18 | 218.139 | 110.9166 | 6.5 | 0.46 |
| polarC18 | 218.139 | 189.7218 | 6.5 | 0.46 |

| | | | | |
|---|---|---|---|---|
| polarC18 | 218.139 | 58.8587 | 6.5 | 0.46 |
| polarC18 | 218.139 | 130.7473 | 6.5 | 0.46 |
| polarC18 | 218.139 | 186.2908 | 6.5 | 0.46 |
| polarC18 | 218.139 | 188.0795 | 6.5 | 0.46 |
| polarC18 | 218.139 | 132.6641 | 6.5 | 0.46 |
| polarC18 | 218.139 | 98.3781 | 6.5 | 0.46 |
| polarC18 | 218.139 | 55.1299 | 6.5 | 0.46 |
| polarC18 | 218.139 | 71.5751 | 6.5 | 0.46 |
| polarC18 | 218.139 | 86.1192 | 6.5 | 0.46 |
| polarC18 | 218.139 | 115.136 | 6.5 | 0.46 |
| polarC18 | 218.139 | 106.5155 | 6.5 | 0.46 |
| polarC18 | 218.139 | 58.5299 | 6.5 | 0.46 |
| polarC18 | 218.139 | 120.8407 | 6.5 | 0.46 |
| polarC18 | 218.139 | 96.2952 | 6.5 | 0.46 |
| polarC18 | 218.139 | 58.6765 | 6.5 | 0.46 |
| polarC18 | 218.139 | 70.3423 | 6.5 | 0.46 |
| polarC18 | 218.139 | 145.9039 | 6.5 | 0.46 |
| polarC18 | 218.139 | 60.8753 | 6.5 | 0.46 |
| polarC18 | 218.139 | 187.9286 | 6.5 | 0.46 |
| polarC18 | 218.139 | 124.2586 | 6.5 | 0.46 |
| polarC18 | 218.139 | 54.4255 | 6.5 | 0.46 |
| polarC18 | 218.139 | 70.4201 | 6.5 | 0.46 |
| polarC18 | 218.139 | 62.737 | 6.5 | 0.46 |
| polarC18 | 218.139 | 154.9024 | 6.5 | 0.46 |
| polarC18 | 218.139 | 107.1739 | 6.5 | 0.46 |
| polarC18 | 218.139 | 49.0336 | 6.5 | 0.46 |
| polarC18 | 218.139 | 92.9319 | 6.5 | 0.46 |
| polarC18 | 218.139 | 83.02 | 6.5 | 0.46 |
| polarC18 | 218.139 | 54.0908 | 6.5 | 0.46 |
| polarC18 | 218.139 | 107.552 | 6.5 | 0.46 |
| polarC18 | 218.139 | 233.2798 | 6.5 | 0.46 |
| polarC18 | 218.139 | 60.4677 | 6.5 | 0.46 |
| polarC18 | 218.139 | 148.2805 | 6.5 | 0.46 |
| polarC18 | 218.139 | 59.5858 | 6.5 | 0.46 |
| polarC18 | 218.139 | 65.9037 | 6.5 | 0.46 |
| polarC18 | 218.139 | 216.7721 | 6.5 | 0.46 |
| polarC18 | 218.139 | 53.7235 | 6.5 | 0.46 |
| polarC18 | 218.139 | 70.5054 | 6.5 | 0.46 |
| polarC18 | 218.139 | 236.9445 | 6.5 | 0.46 |
| polarC18 | 218.139 | 60.2277 | 6.5 | 0.46 |
| polarC18 | 218.139 | 95.6645 | 6.5 | 0.46 |
| polarC18 | 218.139 | 58.5412 | 6.5 | 0.46 |

| | | | | |
|---|---|---|---|---|
| polarC18 | 218.139 | 58.3408 | 6.5 | 0.46 |
| polarC18 | 218.139 | 126.0094 | 6.5 | 0.46 |
| polarC18 | 218.139 | 48.7455 | 6.5 | 0.46 |
| polarC18 | 218.139 | 88.7861 | 6.5 | 0.46 |
| polarC18 | 218.139 | 87.6376 | 6.5 | 0.46 |
| polarC18 | 218.139 | 69.6602 | 6.5 | 0.46 |
| polarC18 | 218.139 | 49.3055 | 6.5 | 0.46 |
| polarC18 | 218.139 | 58.4951 | 6.5 | 0.46 |
| polarC18 | 218.139 | 62.5115 | 6.5 | 0.46 |
| polarC18 | 218.139 | 61.3352 | 6.5 | 0.46 |
| polarC18 | 218.139 | 140.028 | 6.5 | 0.46 |
| polarC18 | 218.139 | 95.9 | 6.5 | 0.46 |
| polarC18 | 218.139 | 68.2101 | 6.5 | 0.46 |
| polarC18 | 218.139 | 50.7623 | 6.5 | 0.46 |
| polarC18 | 218.139 | 194.8759 | 6.5 | 0.46 |
| polarC18 | 218.139 | 84.1653 | 6.5 | 0.46 |
| polarC18 | 218.139 | 51.4459 | 6.5 | 0.46 |
| polarC18 | 218.139 | 154.2692 | 6.5 | 0.46 |
| polarC18 | 218.139 | 104.2303 | 6.5 | 0.46 |
| polarC18 | 218.139 | 146.3641 | 6.5 | 0.46 |
| polarC18 | 218.139 | 175.557 | 6.5 | 0.46 |
| polarC18 | 218.139 | 147.7268 | 6.5 | 0.46 |
| polarC18 | 218.139 | 58.4436 | 6.5 | 0.46 |
| polarC18 | 218.139 | 122.191 | 6.5 | 0.46 |
| polarC18 | 218.139 | 178.8524 | 6.5 | 0.46 |
| polarC18 | 218.139 | 82.2733 | 6.5 | 0.46 |
| polarC18 | 218.139 | 86.3153 | 6.5 | 0.46 |
| polarC18 | 218.139 | 58.6545 | 6.5 | 0.46 |
| polarC18 | 218.139 | 221.543 | 6.5 | 0.46 |
| polarC18 | 218.139 | 52.6045 | 6.5 | 0.46 |
| polarC18 | 218.139 | 86.2188 | 6.5 | 0.46 |
| polarC18 | 218.139 | 160.7516 | 6.5 | 0.46 |
| polarC18 | 218.139 | 62.6918 | 6.5 | 0.46 |
| polarC18 | 218.139 | 72.7725 | 6.5 | 0.46 |
| polarC18 | 218.139 | 124.1618 | 6.5 | 0.46 |
| polarC18 | 218.139 | 49.985 | 6.5 | 0.46 |
| polarC18 | 218.139 | 189.1962 | 6.5 | 0.46 |
| polarC18 | 218.139 | 84.6334 | 6.5 | 0.46 |
| polarC18 | 218.139 | 64.7952 | 6.5 | 0.46 |
| polarC18 | 218.139 | 92.4051 | 6.5 | 0.46 |
| polarC18 | 218.139 | 141.1792 | 6.5 | 0.46 |
| polarC18 | 218.139 | 67.3101 | 6.5 | 0.46 |

| | | | | |
|---|---|---|---|---|
| polarC18 | 218.139 | 129.4368 | 6.5 | 0.46 |
| polarC18 | 218.139 | 176.5081 | 6.5 | 0.46 |
| polarC18 | 218.139 | 68.7982 | 6.5 | 0.46 |
| polarC18 | 218.139 | 52.0455 | 6.5 | 0.46 |
| polarC18 | 218.139 | 106.0195 | 6.5 | 0.46 |
| polarC18 | 218.139 | 123.1157 | 6.5 | 0.46 |
| polarC18 | 218.139 | 71.0314 | 6.5 | 0.46 |
| polarC18 | 218.139 | 66.5732 | 6.5 | 0.46 |
| polarC18 | 218.139 | 81.1638 | 6.5 | 0.46 |
| polarC18 | 218.139 | 77.2833 | 6.5 | 0.46 |
| polarC18 | 218.139 | 50.6421 | 6.5 | 0.46 |
| polarC18 | 218.139 | 122.5849 | 6.5 | 0.46 |
| polarC18 | 218.139 | 77.4518 | 6.5 | 0.46 |
| polarC18 | 218.139 | 64.8424 | 6.5 | 0.46 |
| polarC18 | 218.139 | 71.2853 | 6.5 | 0.46 |
| polarC18 | 218.139 | 173.3139 | 6.5 | 0.46 |
| polarC18 | 218.139 | 71.1908 | 6.5 | 0.46 |
| polarC18 | 218.139 | 62.4185 | 6.5 | 0.46 |
| polarC18 | 218.139 | 71.3704 | 6.5 | 0.46 |
| polarC18 | 218.139 | 53.1997 | 6.5 | 0.46 |
| polarC18 | 218.139 | 51.8065 | 6.5 | 0.46 |
| polarC18 | 218.139 | 98.4325 | 6.5 | 0.46 |
| polarC18 | 218.139 | 67.4439 | 6.5 | 0.46 |
| polarC18 | 218.139 | 72.8085 | 6.5 | 0.46 |
| polarC18 | 218.139 | 167.3972 | 6.5 | 0.46 |
| polarC18 | 218.139 | 140.4363 | 6.5 | 0.46 |
| polarC18 | 218.139 | 79.2198 | 6.5 | 0.46 |
| polarC18 | 218.139 | 106.7152 | 6.5 | 0.46 |
| polarC18 | 218.139 | 105.7192 | 6.5 | 0.46 |
| polarC18 | 218.139 | 133.7376 | 6.5 | 0.46 |
| polarC18 | 218.139 | 58.6301 | 6.5 | 0.46 |
| polarC18 | 218.139 | 49.2623 | 6.5 | 0.46 |
| polarC18 | 218.139 | 69.7234 | 6.5 | 0.46 |
| polarC18 | 218.139 | 215.1709 | 6.5 | 0.46 |
| polarC18 | 218.139 | 152.8578 | 6.5 | 0.46 |
| polarC18 | 218.139 | 122.5049 | 6.5 | 0.46 |
| polarC18 | 218.139 | 69.0005 | 6.5 | 0.46 |
| polarC18 | 218.139 | 240.1309 | 6.5 | 0.46 |
| polarC18 | 218.139 | 103.3548 | 6.5 | 0.46 |
| polarC18 | 218.139 | 79.4901 | 6.5 | 0.46 |
| polarC18 | 218.139 | 68.5697 | 6.5 | 0.46 |
| polarC18 | 218.139 | 190.702 | 6.5 | 0.46 |

| | | | | |
|---|---|---|---|---|
| polarC18 | 218.139 | 69.7717 | 6.5 | 0.46 |
| polarC18 | 218.139 | 208.1175 | 6.5 | 0.46 |
| polarC18 | 218.139 | 102.4707 | 6.5 | 0.46 |
| polarC18 | 218.139 | 101.3924 | 6.5 | 0.46 |
| polarC18 | 218.139 | 127.5038 | 6.5 | 0.46 |
| polarC18 | 218.139 | 54.1797 | 6.5 | 0.46 |
| polarC18 | 218.139 | 51.471 | 6.5 | 0.46 |
| polarC18 | 218.139 | 49.0875 | 6.5 | 0.46 |
| polarC18 | 218.139 | 161.1569 | 6.5 | 0.46 |
| polarC18 | 218.139 | 79.1092 | 6.5 | 0.46 |
| polarC18 | 218.139 | 49.6015 | 6.5 | 0.46 |
| polarC18 | 218.139 | 232.8573 | 6.4 | 0.46 |
| polarC18 | 218.139 | 50.4841 | 6.4 | 0.46 |
| polarC18 | 218.139 | 85.7301 | 6.4 | 0.46 |
| polarC18 | 218.139 | 92.3476 | 6.4 | 0.46 |
| polarC18 | 218.139 | 50.7028 | 6.4 | 0.46 |
| polarC18 | 218.139 | 49.8866 | 6.4 | 0.46 |
| polarC18 | 218.139 | 228.6317 | 6.4 | 0.46 |
| polarC18 | 218.139 | 86.6345 | 6.4 | 0.46 |
| polarC18 | 218.139 | 202.5125 | 6.4 | 0.46 |
| polarC18 | 218.139 | 158.4525 | 6.4 | 0.46 |
| polarC18 | 218.139 | 48.8584 | 6.4 | 0.46 |
| polarC18 | 218.139 | 80.1132 | 6.4 | 0.46 |
| polarC18 | 218.139 | 50.4156 | 6.4 | 0.46 |
| polarC18 | 218.139 | 96.6777 | 6.4 | 0.46 |
| polarC18 | 218.139 | 64.9662 | 6.4 | 0.46 |
| polarC18 | 218.139 | 58.3622 | 6.4 | 0.46 |
| polarC18 | 218.139 | 75.0678 | 6.4 | 0.46 |
| polarC18 | 218.139 | 63.7215 | 6.4 | 0.46 |
| polarC18 | 218.139 | 61.367 | 6.4 | 0.46 |
| polarC18 | 218.139 | 48.7888 | 6.4 | 0.46 |
| polarC18 | 218.139 | 61.374 | 6.4 | 0.46 |
| polarC18 | 218.139 | 69.9613 | 6.4 | 0.46 |
| polarC18 | 218.139 | 177.7818 | 6.4 | 0.46 |
| polarC18 | 218.139 | 115.7558 | 6.4 | 0.46 |
| polarC18 | 218.139 | 105.4702 | 6.4 | 0.46 |
| polarC18 | 218.139 | 93.3185 | 6.4 | 0.46 |
| polarC18 | 218.139 | 100.4939 | 6.4 | 0.46 |
| polarC18 | 218.139 | 166.5584 | 6.4 | 0.46 |
| polarC18 | 218.139 | 102.7166 | 6.4 | 0.46 |
| polarC18 | 218.139 | 121.6608 | 6.4 | 0.46 |
| polarC18 | 218.139 | 54.3947 | 6.4 | 0.46 |

| | | | | |
|---|---|---|---|---|
| polarC18 | 218.139 | 159.3392 | 6.4 | 0.46 |
| polarC18 | 218.139 | 55.5429 | 6.4 | 0.46 |
| polarC18 | 218.139 | 67.2812 | 6.4 | 0.46 |
| polarC18 | 218.139 | 61.92 | 6.4 | 0.46 |
| polarC18 | 218.139 | 51.0298 | 6.4 | 0.46 |
| polarC18 | 218.139 | 112.2044 | 6.4 | 0.46 |
| polarC18 | 218.139 | 61.8247 | 6.4 | 0.46 |
| polarC18 | 218.139 | 199.8323 | 6.4 | 0.46 |
| polarC18 | 218.139 | 70.5403 | 6.4 | 0.46 |
| polarC18 | 218.139 | 197.1357 | 6.4 | 0.46 |
| polarC18 | 218.139 | 150.0638 | 6.4 | 0.46 |
| polarC18 | 218.139 | 70.4833 | 6.4 | 0.46 |
| polarC18 | 218.139 | 75.7563 | 6.4 | 0.46 |
| polarC18 | 218.139 | 80.5425 | 6.4 | 0.46 |
| polarC18 | 218.139 | 80.7713 | 6.4 | 0.46 |
| polarC18 | 218.139 | 128.3667 | 6.4 | 0.46 |
| polarC18 | 218.139 | 69.1434 | 6.4 | 0.46 |
| polarC18 | 218.139 | 54.0319 | 6.4 | 0.46 |
| polarC18 | 218.139 | 82.2975 | 6.4 | 0.46 |
| polarC18 | 218.139 | 57.8333 | 6.4 | 0.46 |
| polarC18 | 218.139 | 56.7417 | 6.4 | 0.46 |
| polarC18 | 218.139 | 81.0798 | 6.4 | 0.46 |
| polarC18 | 218.139 | 89.0358 | 6.4 | 0.46 |
| polarC18 | 218.139 | 160.2466 | 6.4 | 0.46 |
| polarC18 | 218.139 | 59.6473 | 6.4 | 0.46 |
| polarC18 | 218.139 | 130.3337 | 6.4 | 0.46 |
| polarC18 | 218.139 | 117.6243 | 6.4 | 0.46 |
| polarC18 | 218.139 | 67.8597 | 6.4 | 0.46 |
| polarC18 | 218.139 | 86.9697 | 6.4 | 0.46 |
| polarC18 | 218.139 | 128.7181 | 6.4 | 0.46 |
| polarC18 | 218.139 | 93.2505 | 6.4 | 0.46 |
| polarC18 | 218.139 | 50.7887 | 6.4 | 0.46 |
| polarC18 | 218.139 | 52.0058 | 6.4 | 0.46 |
| polarC18 | 218.139 | 55.3705 | 6.4 | 0.46 |
| polarC18 | 218.139 | 54.1395 | 6.4 | 0.46 |
| polarC18 | 218.139 | 61.6418 | 6.4 | 0.46 |
| polarC18 | 218.139 | 63.3107 | 6.4 | 0.46 |
| polarC18 | 218.139 | 48.8986 | 6.4 | 0.46 |
| polarC18 | 218.139 | 56.0673 | 6.4 | 0.46 |
| polarC18 | 218.139 | 56.5937 | 6.4 | 0.46 |
| polarC18 | 218.139 | 100.4527 | 6.4 | 0.46 |
| polarC18 | 218.139 | 115.5554 | 6.4 | 0.46 |

| | | | | |
|---|---|---|---|---|
| polarC18 | 218.139 | 163.4294 | 6.4 | 0.46 |
| polarC18 | 218.139 | 77.0571 | 6.4 | 0.46 |
| polarC18 | 218.139 | 61.1501 | 6.4 | 0.46 |
| polarC18 | 218.139 | 90.7877 | 6.4 | 0.46 |
| polarC18 | 218.139 | 132.9879 | 6.4 | 0.46 |
| polarC18 | 218.139 | 54.2698 | 6.4 | 0.46 |
| polarC18 | 218.139 | 89.6797 | 6.4 | 0.46 |
| polarC18 | 218.139 | 60.8964 | 6.4 | 0.46 |
| polarC18 | 218.139 | 63.8204 | 6.4 | 0.46 |
| polarC18 | 218.139 | 70.3557 | 6.4 | 0.46 |
| polarC18 | 218.139 | 195.1881 | 6.4 | 0.46 |
| polarC18 | 218.139 | 54.4355 | 6.4 | 0.46 |
| polarC18 | 218.139 | 96.4275 | 6.4 | 0.46 |
| polarC18 | 218.139 | 97.1109 | 6.4 | 0.46 |
| polarC18 | 218.139 | 58.552 | 6.4 | 0.46 |
| polarC18 | 218.139 | 109.508 | 6.4 | 0.46 |
| polarC18 | 218.139 | 185.9944 | 6.4 | 0.46 |
| polarC18 | 218.139 | 64.8449 | 6.4 | 0.46 |
| polarC18 | 218.139 | 59.4392 | 6.4 | 0.46 |
| polarC18 | 218.139 | 58.3471 | 6.4 | 0.46 |
| polarC18 | 218.139 | 50.6372 | 6.4 | 0.46 |
| polarC18 | 218.139 | 185.4261 | 6.4 | 0.46 |
| polarC18 | 218.139 | 50.9546 | 6.4 | 0.46 |
| polarC18 | 218.139 | 72.0661 | 6.4 | 0.46 |
| polarC18 | 218.139 | 53.8742 | 6.4 | 0.46 |
| polarC18 | 218.139 | 54.9103 | 6.4 | 0.46 |
| polarC18 | 218.139 | 71.547 | 6.4 | 0.46 |
| polarC18 | 218.139 | 125.0704 | 6.4 | 0.46 |
| polarC18 | 218.139 | 159.4716 | 6.4 | 0.46 |
| polarC18 | 218.139 | 162.6965 | 6.4 | 0.46 |
| polarC18 | 218.139 | 62.7716 | 6.4 | 0.46 |
| polarC18 | 218.139 | 76.0551 | 6.4 | 0.46 |
| polarC18 | 218.139 | 71.293 | 6.4 | 0.46 |
| polarC18 | 218.139 | 58.3879 | 6.4 | 0.46 |
| polarC18 | 218.139 | 137.1185 | 6.4 | 0.46 |
| polarC18 | 218.139 | 191.3928 | 6.4 | 0.46 |
| polarC18 | 218.139 | 67.7376 | 6.4 | 0.45 |
| polarC18 | 218.139 | 66.8288 | 6.4 | 0.45 |
| polarC18 | 218.139 | 198.4918 | 6.4 | 0.45 |
| polarC18 | 218.139 | 65.7931 | 6.4 | 0.45 |
| polarC18 | 218.139 | 110.2701 | 6.4 | 0.45 |
| polarC18 | 218.139 | 68.315 | 6.4 | 0.45 |

| | | | | |
|---|---|---|---|---|
| polarC18 | 218.139 | 89.075 | 6.4 | 0.45 |
| polarC18 | 218.139 | 132.8127 | 6.4 | 0.45 |
| polarC18 | 218.139 | 215.49 | 6.4 | 0.45 |
| polarC18 | 218.139 | 50.8039 | 6.4 | 0.45 |
| polarC18 | 218.139 | 160.4999 | 6.4 | 0.45 |
| polarC18 | 218.139 | 77.2709 | 6.4 | 0.45 |
| polarC18 | 218.139 | 79.4337 | 6.4 | 0.45 |
| polarC18 | 218.139 | 91.9476 | 6.4 | 0.45 |
| polarC18 | 218.139 | 144.8813 | 6.4 | 0.45 |
| polarC18 | 218.139 | 174.8395 | 6.4 | 0.45 |
| polarC18 | 218.139 | 216.4509 | 6.4 | 0.45 |
| polarC18 | 218.139 | 74.1582 | 6.4 | 0.45 |
| polarC18 | 218.139 | 121.6535 | 6.4 | 0.45 |
| polarC18 | 218.139 | 53.6016 | 6.4 | 0.45 |
| polarC18 | 218.139 | 156.4218 | 6.4 | 0.45 |
| polarC18 | 218.139 | 63.1453 | 6.4 | 0.45 |
| polarC18 | 218.139 | 63.3334 | 6.4 | 0.45 |
| polarC18 | 218.139 | 231.5175 | 6.4 | 0.45 |
| polarC18 | 218.139 | 52.2303 | 6.4 | 0.45 |
| polarC18 | 218.139 | 49.4571 | 6.4 | 0.45 |
| polarC18 | 218.139 | 76.5675 | 6.4 | 0.45 |
| polarC18 | 218.139 | 105.9118 | 6.4 | 0.45 |
| polarC18 | 218.139 | 76.0179 | 6.4 | 0.45 |
| polarC18 | 218.139 | 213.6562 | 6.4 | 0.45 |
| polarC18 | 218.139 | 87.4751 | 6.4 | 0.45 |
| polarC18 | 218.139 | 114.4482 | 6.4 | 0.45 |
| polarC18 | 218.139 | 65.2251 | 6.4 | 0.45 |
| polarC18 | 218.139 | 213.454 | 6.4 | 0.45 |
| polarC18 | 218.139 | 122.4824 | 6.4 | 0.45 |
| polarC18 | 218.139 | 96.3298 | 6.4 | 0.45 |
| polarC18 | 218.139 | 82.3466 | 6.4 | 0.45 |
| polarC18 | 218.139 | 59.623 | 6.4 | 0.45 |
| polarC18 | 218.139 | 52.9504 | 6.4 | 0.45 |
| polarC18 | 218.139 | 145.7583 | 6.4 | 0.45 |
| polarC18 | 218.139 | 102.5121 | 6.4 | 0.45 |
| polarC18 | 218.139 | 187.0133 | 6.4 | 0.45 |
| polarC18 | 218.139 | 86.9308 | 6.4 | 0.45 |
| polarC18 | 218.139 | 213.1326 | 6.4 | 0.45 |
| polarC18 | 218.139 | 52.7169 | 6.4 | 0.45 |
| polarC18 | 218.139 | 223.2043 | 6.4 | 0.45 |
| polarC18 | 218.139 | 94.7964 | 6.4 | 0.45 |
| polarC18 | 218.139 | 49.9226 | 6.4 | 0.45 |

| | | | | |
|---|---|---|---|---|
| polarC18 | 218.139 | 66.8551 | 6.4 | 0.45 |
| polarC18 | 218.139 | 108.6873 | 6.4 | 0.45 |
| polarC18 | 218.139 | 127.7788 | 6.4 | 0.45 |
| polarC18 | 218.139 | 76.5361 | 6.4 | 0.45 |
| polarC18 | 218.139 | 55.3501 | 6.4 | 0.45 |
| polarC18 | 218.139 | 162.0657 | 6.4 | 0.45 |
| polarC18 | 218.139 | 128.7312 | 6.4 | 0.45 |
| polarC18 | 218.139 | 166.2019 | 6.4 | 0.45 |
| polarC18 | 218.139 | 49.816 | 6.4 | 0.45 |
| polarC18 | 218.139 | 91.4489 | 6.4 | 0.45 |
| polarC18 | 218.139 | 66.6847 | 6.4 | 0.45 |
| polarC18 | 218.139 | 126.1761 | 6.4 | 0.45 |
| polarC18 | 218.139 | 144.6823 | 6.4 | 0.45 |
| polarC18 | 218.139 | 205.798 | 6.4 | 0.45 |
| polarC18 | 218.139 | 229.2265 | 6.4 | 0.45 |
| polarC18 | 218.139 | 64.4973 | 6.4 | 0.45 |
| polarC18 | 218.139 | 115.0356 | 6.4 | 0.45 |
| polarC18 | 218.139 | 106.3714 | 6.4 | 0.45 |
| polarC18 | 218.139 | 58.9802 | 6.4 | 0.45 |
| polarC18 | 218.139 | 56.4744 | 6.4 | 0.45 |
| polarC18 | 218.139 | 111.4312 | 6.4 | 0.45 |
| polarC18 | 218.139 | 52.2983 | 6.4 | 0.45 |
| polarC18 | 218.139 | 78.1966 | 6.4 | 0.45 |
| polarC18 | 218.139 | 61.0724 | 6.4 | 0.45 |
| polarC18 | 218.139 | 53.8765 | 6.4 | 0.45 |
| polarC18 | 218.139 | 138.8895 | 6.4 | 0.45 |
| polarC18 | 218.139 | 82.8255 | 6.4 | 0.45 |
| polarC18 | 218.139 | 58.6811 | 6.4 | 0.45 |
| polarC18 | 218.139 | 154.1435 | 6.4 | 0.45 |
| polarC18 | 218.139 | 49.2416 | 6.4 | 0.45 |
| polarC18 | 218.139 | 67.2092 | 6.4 | 0.45 |
| polarC18 | 218.139 | 107.2293 | 6.4 | 0.45 |
| polarC18 | 218.139 | 137.7235 | 6.4 | 0.45 |
| polarC18 | 218.139 | 134.7346 | 6.4 | 0.45 |
| polarC18 | 218.139 | 54.3433 | 6.4 | 0.45 |
| polarC18 | 218.139 | 62.0295 | 6.4 | 0.45 |
| polarC18 | 218.139 | 72.2971 | 6.4 | 0.45 |
| polarC18 | 218.139 | 131.2159 | 6.4 | 0.45 |
| polarC18 | 218.139 | 117.3023 | 6.4 | 0.45 |
| polarC18 | 218.139 | 86.4876 | 6.4 | 0.45 |
| polarC18 | 218.139 | 62.2276 | 6.4 | 0.45 |
| polarC18 | 218.139 | 82.9739 | 6.4 | 0.45 |

| | | | | |
|---|---|---|---|---|
| polarC18 | 218.139 | 120.5145 | 6.4 | 0.45 |
| polarC18 | 218.139 | 50.3935 | 6.4 | 0.45 |
| polarC18 | 218.139 | 109.7993 | 6.4 | 0.45 |
| polarC18 | 218.139 | 59.6965 | 6.4 | 0.45 |
| polarC18 | 218.139 | 54.5172 | 6.4 | 0.45 |
| polarC18 | 218.139 | 84.4626 | 6.4 | 0.45 |
| polarC18 | 218.139 | 67.4683 | 6.4 | 0.45 |
| polarC18 | 218.139 | 52.1458 | 6.4 | 0.45 |
| polarC18 | 218.139 | 56.3955 | 6.4 | 0.45 |
| polarC18 | 218.139 | 54.6753 | 6.4 | 0.45 |
| polarC18 | 218.139 | 60.5659 | 6.4 | 0.45 |
| polarC18 | 218.139 | 111.093 | 6.4 | 0.45 |
| polarC18 | 218.139 | 128.1299 | 6.4 | 0.45 |
| polarC18 | 218.139 | 98.1026 | 6.4 | 0.45 |
| polarC18 | 218.139 | 50.987 | 6.4 | 0.45 |
| polarC18 | 218.139 | 86.7044 | 6.4 | 0.45 |
| polarC18 | 218.139 | 124.5019 | 6.4 | 0.45 |
| polarC18 | 218.139 | 99.0863 | 6.4 | 0.45 |
| polarC18 | 218.139 | 105.2645 | 6.4 | 0.45 |
| polarC18 | 218.139 | 69.6394 | 6.4 | 0.45 |
| polarC18 | 218.139 | 54.5561 | 6.4 | 0.45 |
| polarC18 | 218.139 | 77.9047 | 6.4 | 0.45 |
| polarC18 | 218.139 | 174.8696 | 6.4 | 0.45 |
| polarC18 | 218.139 | 85.9656 | 6.4 | 0.45 |
| polarC18 | 218.139 | 110.8921 | 6.4 | 0.45 |
| polarC18 | 218.139 | 211.4103 | 6.4 | 0.45 |
| polarC18 | 218.139 | 49.0889 | 6.4 | 0.45 |
| polarC18 | 218.139 | 66.515 | 6.4 | 0.45 |
| polarC18 | 218.139 | 55.4041 | 6.4 | 0.45 |
| polarC18 | 218.139 | 68.3461 | 6.4 | 0.45 |
| polarC18 | 218.139 | 123.1734 | 6.4 | 0.45 |
| polarC18 | 218.139 | 107.1344 | 6.4 | 0.45 |
| polarC18 | 218.139 | 145.8087 | 6.4 | 0.45 |
| polarC18 | 218.139 | 53.9597 | 6.4 | 0.45 |
| polarC18 | 218.139 | 51.6155 | 6.4 | 0.45 |
| polarC18 | 218.139 | 93.5585 | 6.4 | 0.45 |
| polarC18 | 218.139 | 73.5151 | 6.4 | 0.45 |
| polarC18 | 218.139 | 173.6568 | 6.4 | 0.45 |
| polarC18 | 218.139 | 81.5054 | 6.4 | 0.45 |
| polarC18 | 218.139 | 49.6758 | 6.4 | 0.45 |
| polarC18 | 218.139 | 66.1158 | 6.4 | 0.45 |
| polarC18 | 218.139 | 65.8572 | 6.4 | 0.45 |

| | | | | |
|---|---|---|---|---|
| polarC18 | 218.139 | 67.2677 | 6.4 | 0.45 |
| polarC18 | 218.139 | 113.8139 | 6.4 | 0.45 |
| polarC18 | 218.139 | 104.9415 | 6.4 | 0.45 |
| polarC18 | 218.139 | 94.9984 | 6.4 | 0.45 |
| polarC18 | 218.139 | 49.8414 | 6.4 | 0.45 |
| polarC18 | 218.139 | 160.3568 | 6.4 | 0.45 |
| polarC18 | 218.139 | 125.5896 | 6.4 | 0.45 |
| polarC18 | 218.139 | 147.1619 | 6.4 | 0.45 |
| polarC18 | 218.139 | 122.2627 | 6.4 | 0.45 |
| polarC18 | 218.139 | 85.2555 | 6.4 | 0.45 |
| polarC18 | 218.139 | 68.907 | 6.4 | 0.45 |
| polarC18 | 218.139 | 81.5861 | 6.4 | 0.45 |
| polarC18 | 218.139 | 56.8822 | 6.4 | 0.45 |
| polarC18 | 218.139 | 62.0736 | 6.4 | 0.45 |
| polarC18 | 218.139 | 120.071 | 6.4 | 0.45 |
| polarC18 | 218.139 | 52.5618 | 6.4 | 0.45 |
| polarC18 | 218.139 | 61.4098 | 6.4 | 0.45 |
| polarC18 | 218.139 | 181.9717 | 6.4 | 0.45 |
| polarC18 | 218.139 | 165.5894 | 6.4 | 0.45 |
| polarC18 | 218.139 | 59.0468 | 6.4 | 0.45 |
| polarC18 | 218.139 | 93.0064 | 6.4 | 0.45 |
| polarC18 | 218.139 | 89.2917 | 6.4 | 0.45 |
| polarC18 | 218.139 | 51.1402 | 6.4 | 0.45 |
| polarC18 | 218.139 | 149.0308 | 6.4 | 0.45 |
| polarC18 | 218.139 | 79.7633 | 6.4 | 0.45 |
| polarC18 | 218.139 | 86.3586 | 6.4 | 0.45 |
| polarC18 | 218.139 | 65.8025 | 6.4 | 0.45 |
| polarC18 | 218.139 | 73.7589 | 6.4 | 0.45 |
| polarC18 | 218.139 | 114.7499 | 6.4 | 0.45 |
| polarC18 | 218.139 | 64.6403 | 6.4 | 0.45 |
| polarC18 | 218.139 | 69.1186 | 6.4 | 0.45 |
| polarC18 | 218.139 | 206.3515 | 6.4 | 0.45 |
| polarC18 | 218.139 | 59.692 | 6.4 | 0.45 |
| polarC18 | 218.139 | 54.8689 | 6.4 | 0.45 |
| polarC18 | 218.139 | 57.653 | 6.4 | 0.45 |
| polarC18 | 218.139 | 55.6953 | 6.4 | 0.45 |
| polarC18 | 218.139 | 71.4385 | 6.4 | 0.45 |
| polarC18 | 218.139 | 106.6225 | 6.4 | 0.45 |
| polarC18 | 218.139 | 131.3566 | 6.4 | 0.45 |
| polarC18 | 218.139 | 127.2634 | 6.4 | 0.45 |
| polarC18 | 218.139 | 67.082 | 6.4 | 0.45 |
| polarC18 | 218.139 | 62.5764 | 6.4 | 0.45 |

| | | | | |
|---|---|---|---|---|
| polarC18 | 218.139 | 104.3095 | 6.4 | 0.45 |
| polarC18 | 218.139 | 79.8014 | 6.4 | 0.45 |
| polarC18 | 218.139 | 93.1279 | 6.4 | 0.45 |
| polarC18 | 218.139 | 61.5115 | 6.4 | 0.45 |
| polarC18 | 218.139 | 58.6075 | 6.4 | 0.45 |
| polarC18 | 218.139 | 174.6448 | 6.4 | 0.45 |
| polarC18 | 218.139 | 94.9387 | 6.4 | 0.45 |
| polarC18 | 218.139 | 100.9939 | 6.4 | 0.45 |
| polarC18 | 218.139 | 81.456 | 6.4 | 0.45 |
| polarC18 | 218.139 | 184.6344 | 6.4 | 0.45 |
| polarC18 | 218.139 | 88.6588 | 6.4 | 0.45 |
| polarC18 | 218.139 | 74.734 | 6.4 | 0.45 |
| polarC18 | 218.139 | 159.8929 | 6.4 | 0.45 |
| polarC18 | 218.139 | 75.3701 | 6.4 | 0.45 |
| polarC18 | 218.139 | 148.2417 | 6.4 | 0.45 |
| polarC18 | 218.139 | 48.6232 | 6.4 | 0.45 |
| polarC18 | 218.139 | 80.2852 | 6.4 | 0.45 |
| polarC18 | 218.139 | 128.2137 | 6.3 | 0.45 |
| polarC18 | 218.139 | 78.4471 | 6.3 | 0.45 |
| polarC18 | 218.139 | 67.1129 | 6.3 | 0.45 |
| polarC18 | 218.139 | 108.0036 | 6.3 | 0.45 |
| polarC18 | 218.139 | 230.2665 | 6.3 | 0.45 |
| polarC18 | 218.139 | 168.1616 | 6.3 | 0.45 |
| polarC18 | 218.139 | 100.7581 | 6.3 | 0.45 |
| polarC18 | 218.139 | 72.6087 | 6.3 | 0.45 |
| polarC18 | 218.139 | 69.8781 | 6.3 | 0.45 |
| polarC18 | 218.139 | 201.1059 | 6.3 | 0.45 |
| polarC18 | 218.139 | 197.5243 | 6.3 | 0.45 |
| polarC18 | 218.139 | 66.3184 | 6.3 | 0.45 |
| polarC18 | 218.139 | 138.2411 | 6.3 | 0.45 |
| polarC18 | 218.139 | 88.6535 | 6.3 | 0.45 |
| polarC18 | 218.139 | 56.6653 | 6.3 | 0.45 |
| polarC18 | 218.139 | 165.8054 | 6.3 | 0.45 |
| polarC18 | 218.139 | 74.6125 | 6.3 | 0.45 |
| polarC18 | 218.139 | 180.2769 | 6.3 | 0.45 |
| polarC18 | 218.139 | 55.1603 | 6.3 | 0.45 |
| polarC18 | 218.139 | 70.7047 | 6.3 | 0.45 |
| polarC18 | 218.139 | 79.4697 | 6.3 | 0.45 |
| polarC18 | 218.139 | 177.0189 | 6.3 | 0.45 |
| polarC18 | 218.139 | 56.0818 | 6.3 | 0.45 |
| polarC18 | 218.139 | 173.3797 | 6.3 | 0.45 |
| polarC18 | 218.139 | 55.5195 | 6.3 | 0.45 |

| | | | | |
|---|---|---|---|---|
| polarC18 | 218.139 | 58.5629 | 6.3 | 0.45 |
| polarC18 | 218.139 | 122.0327 | 6.3 | 0.45 |
| polarC18 | 218.139 | 50.9764 | 6.3 | 0.45 |
| polarC18 | 218.139 | 111.3564 | 6.3 | 0.45 |
| polarC18 | 218.139 | 66.1763 | 6.3 | 0.45 |
| polarC18 | 218.139 | 68.0985 | 6.3 | 0.45 |
| polarC18 | 218.139 | 142.857 | 6.3 | 0.45 |
| polarC18 | 218.139 | 241.9989 | 6.3 | 0.45 |
| polarC18 | 218.139 | 120.5293 | 6.3 | 0.45 |
| polarC18 | 218.139 | 100.098 | 6.3 | 0.45 |
| polarC18 | 218.139 | 78.5332 | 6.3 | 0.45 |
| polarC18 | 218.139 | 66.6594 | 6.3 | 0.45 |
| polarC18 | 218.139 | 49.5523 | 6.3 | 0.45 |
| polarC18 | 218.139 | 135.8552 | 6.3 | 0.45 |
| polarC18 | 218.139 | 142.9285 | 6.3 | 0.45 |
| polarC18 | 218.139 | 53.6672 | 6.3 | 0.45 |
| polarC18 | 218.139 | 200.7666 | 6.3 | 0.45 |
| polarC18 | 218.139 | 51.1868 | 6.3 | 0.45 |
| polarC18 | 218.139 | 66.194 | 6.3 | 0.45 |
| polarC18 | 218.139 | 170.3919 | 6.3 | 0.45 |
| polarC18 | 218.139 | 71.4161 | 6.3 | 0.45 |
| polarC18 | 218.139 | 79.7858 | 6.3 | 0.45 |
| polarC18 | 218.139 | 50.6906 | 6.3 | 0.45 |
| polarC18 | 218.139 | 104.9232 | 6.3 | 0.45 |
| polarC18 | 218.139 | 50.1832 | 6.3 | 0.45 |
| polarC18 | 218.139 | 57.4057 | 6.3 | 0.45 |
| polarC18 | 218.139 | 80.9868 | 6.3 | 0.45 |
| polarC18 | 218.139 | 115.0043 | 6.3 | 0.45 |
| polarC18 | 218.139 | 69.0577 | 6.3 | 0.45 |
| polarC18 | 218.139 | 100.0286 | 6.3 | 0.45 |
| polarC18 | 218.139 | 78.4143 | 6.3 | 0.45 |
| polarC18 | 218.139 | 91.5558 | 6.3 | 0.45 |
| polarC18 | 218.139 | 96.1666 | 6.3 | 0.45 |
| polarC18 | 218.139 | 53.4949 | 6.3 | 0.45 |
| polarC18 | 218.139 | 122.3243 | 6.3 | 0.45 |
| polarC18 | 218.139 | 53.13 | 6.3 | 0.45 |
| polarC18 | 218.139 | 134.0001 | 6.3 | 0.45 |
| polarC18 | 218.139 | 117.0019 | 6.3 | 0.45 |
| polarC18 | 218.139 | 84.1205 | 6.3 | 0.45 |
| polarC18 | 218.139 | 74.7797 | 6.3 | 0.45 |
| polarC18 | 218.139 | 52.8227 | 6.3 | 0.45 |
| polarC18 | 218.139 | 76.6301 | 6.3 | 0.45 |

| | | | | |
|---|---|---|---|---|
| polarC18 | 218.139 | 185.5667 | 6.3 | 0.45 |
| polarC18 | 218.139 | 53.8832 | 6.3 | 0.45 |
| polarC18 | 218.139 | 86.0368 | 6.3 | 0.45 |
| polarC18 | 218.139 | 65.0478 | 6.3 | 0.45 |
| polarC18 | 218.139 | 61.8915 | 6.3 | 0.45 |
| polarC18 | 218.139 | 68.429 | 6.3 | 0.45 |
| polarC18 | 218.139 | 79.3073 | 6.3 | 0.45 |
| polarC18 | 218.139 | 56.3122 | 6.3 | 0.45 |
| polarC18 | 218.139 | 88.8893 | 6.3 | 0.45 |
| polarC18 | 218.139 | 51.7986 | 6.3 | 0.45 |
| polarC18 | 218.139 | 90.0344 | 6.3 | 0.45 |
| polarC18 | 218.139 | 160.4662 | 6.3 | 0.45 |
| polarC18 | 218.139 | 140.7306 | 6.3 | 0.45 |
| polarC18 | 218.139 | 129.4504 | 6.3 | 0.45 |
| polarC18 | 218.139 | 70.9316 | 6.3 | 0.45 |
| polarC18 | 218.139 | 88.793 | 6.3 | 0.45 |
| polarC18 | 218.139 | 59.2859 | 6.3 | 0.45 |
| polarC18 | 218.139 | 66.7234 | 6.3 | 0.45 |
| polarC18 | 218.139 | 78.7963 | 6.3 | 0.45 |
| polarC18 | 218.139 | 73.3272 | 6.3 | 0.45 |
| polarC18 | 218.139 | 125.7689 | 6.3 | 0.45 |
| polarC18 | 218.139 | 83.3279 | 6.3 | 0.45 |
| polarC18 | 218.139 | 76.2643 | 6.3 | 0.45 |
| polarC18 | 218.139 | 59.3087 | 6.3 | 0.45 |
| polarC18 | 218.139 | 90.7255 | 6.3 | 0.45 |
| polarC18 | 218.139 | 48.8182 | 6.3 | 0.45 |
| polarC18 | 218.139 | 55.0176 | 6.3 | 0.45 |
| polarC18 | 218.139 | 50.4807 | 6.3 | 0.45 |
| polarC18 | 218.139 | 55.3932 | 6.3 | 0.45 |
| polarC18 | 218.139 | 48.6306 | 6.3 | 0.45 |
| polarC18 | 218.139 | 61.5648 | 6.3 | 0.45 |
| polarC18 | 218.139 | 157.2072 | 6.3 | 0.45 |
| polarC18 | 218.139 | 79.7382 | 6.3 | 0.45 |
| polarC18 | 218.139 | 83.0818 | 6.3 | 0.45 |
| polarC18 | 218.139 | 78.982 | 6.3 | 0.45 |
| polarC18 | 218.139 | 164.1088 | 6.3 | 0.45 |
| polarC18 | 218.139 | 52.1129 | 6.3 | 0.45 |
| polarC18 | 218.139 | 60.7636 | 6.3 | 0.45 |
| polarC18 | 218.139 | 77.8459 | 6.3 | 0.45 |
| polarC18 | 218.139 | 199.1277 | 6.3 | 0.45 |
| polarC18 | 218.139 | 85.7679 | 6.3 | 0.45 |
| polarC18 | 218.139 | 100.2487 | 6.3 | 0.45 |

| | | | | |
|---|---|---|---|---|
| polarC18 | 218.139 | 73.5592 | 6.3 | 0.45 |
| polarC18 | 218.139 | 120.8963 | 6.3 | 0.45 |
| polarC18 | 218.139 | 67.4217 | 6.3 | 0.45 |
| polarC18 | 218.139 | 221.4612 | 6.3 | 0.45 |
| polarC18 | 218.139 | 137.3267 | 6.3 | 0.45 |
| polarC18 | 218.139 | 52.7847 | 6.3 | 0.45 |
| polarC18 | 218.139 | 49.9938 | 6.3 | 0.45 |
| polarC18 | 218.139 | 58.901 | 6.3 | 0.45 |
| polarC18 | 218.139 | 129.3065 | 6.3 | 0.45 |
| polarC18 | 218.139 | 133.4175 | 6.3 | 0.45 |
| polarC18 | 218.139 | 48.7305 | 6.3 | 0.45 |
| polarC18 | 218.139 | 97.1337 | 6.3 | 0.45 |
| polarC18 | 218.139 | 112.3581 | 6.3 | 0.45 |
| polarC18 | 218.139 | 169.9143 | 6.3 | 0.45 |
| polarC18 | 218.139 | 98.8476 | 6.3 | 0.45 |
| polarC18 | 218.139 | 55.8548 | 6.3 | 0.45 |
| polarC18 | 218.139 | 79.0005 | 6.3 | 0.45 |
| polarC18 | 218.139 | 49.0828 | 6.3 | 0.45 |
| polarC18 | 218.139 | 235.8057 | 6.3 | 0.45 |
| polarC18 | 218.139 | 153.6389 | 6.3 | 0.45 |
| polarC18 | 218.139 | 118.0619 | 6.3 | 0.45 |
| polarC18 | 218.139 | 97.12 | 6.3 | 0.45 |
| polarC18 | 218.139 | 95.812 | 6.3 | 0.45 |
| polarC18 | 218.139 | 49.8518 | 6.3 | 0.45 |
| polarC18 | 218.139 | 61.2196 | 6.3 | 0.45 |
| polarC18 | 218.139 | 206.9041 | 6.3 | 0.45 |
| polarC18 | 218.139 | 87.6117 | 6.3 | 0.45 |
| polarC18 | 218.139 | 55.4463 | 6.3 | 0.45 |
| polarC18 | 218.139 | 129.7535 | 6.3 | 0.45 |
| polarC18 | 218.139 | 221.7646 | 6.3 | 0.45 |
| polarC18 | 218.139 | 49.2519 | 6.3 | 0.45 |
| polarC18 | 218.139 | 89.7785 | 6.3 | 0.45 |
| polarC18 | 218.139 | 55.8202 | 6.3 | 0.45 |
| polarC18 | 218.139 | 89.8429 | 6.3 | 0.45 |
| polarC18 | 218.139 | 131.4998 | 6.3 | 0.45 |
| polarC18 | 218.139 | 75.1139 | 6.3 | 0.45 |
| polarC18 | 218.139 | 102.4368 | 6.3 | 0.45 |
| polarC18 | 218.139 | 115.3765 | 6.3 | 0.45 |
| polarC18 | 218.139 | 51.6289 | 6.3 | 0.45 |
| polarC18 | 218.139 | 108.3448 | 6.3 | 0.45 |
| polarC18 | 218.139 | 59.534 | 6.3 | 0.45 |
| polarC18 | 218.139 | 142.7251 | 6.3 | 0.45 |

| | | | | |
|---|---|---|---|---|
| polarC18 | 218.139 | 48.8223 | 6.3 | 0.45 |
| polarC18 | 218.139 | 111.138 | 6.3 | 0.45 |
| polarC18 | 218.139 | 49.4759 | 6.3 | 0.45 |
| polarC18 | 218.139 | 69.7881 | 6.3 | 0.45 |
| polarC18 | 218.139 | 173.4099 | 6.3 | 0.45 |
| polarC18 | 218.139 | 204.6333 | 6.3 | 0.45 |
| polarC18 | 218.139 | 87.2947 | 6.3 | 0.45 |
| polarC18 | 218.139 | 61.1011 | 6.3 | 0.45 |
| polarC18 | 218.139 | 119.0368 | 6.3 | 0.45 |
| polarC18 | 218.139 | 150.6227 | 6.3 | 0.45 |
| polarC18 | 218.139 | 65.0077 | 6.3 | 0.45 |
| polarC18 | 218.139 | 50.5548 | 6.3 | 0.45 |
| polarC18 | 218.139 | 122.8375 | 6.3 | 0.45 |
| polarC18 | 218.139 | 55.6809 | 6.3 | 0.45 |
| polarC18 | 218.139 | 50.2781 | 6.3 | 0.45 |
| polarC18 | 218.139 | 53.6301 | 6.3 | 0.45 |
| polarC18 | 218.139 | 72.5674 | 6.3 | 0.45 |
| polarC18 | 218.139 | 59.151 | 6.3 | 0.45 |
| polarC18 | 218.139 | 68.9231 | 6.3 | 0.45 |
| polarC18 | 218.139 | 72.3904 | 6.3 | 0.45 |
| polarC18 | 218.139 | 68.649 | 6.3 | 0.45 |
| polarC18 | 218.139 | 220.7332 | 6.3 | 0.45 |
| polarC18 | 218.139 | 51.0863 | 6.3 | 0.45 |
| polarC18 | 218.139 | 165.2759 | 6.3 | 0.45 |
| polarC18 | 218.139 | 104.6697 | 6.3 | 0.45 |
| polarC18 | 218.139 | 154.9531 | 6.3 | 0.45 |
| polarC18 | 218.139 | 51.4209 | 6.3 | 0.45 |
| polarC18 | 218.139 | 109.4118 | 6.3 | 0.45 |
| polarC18 | 218.139 | 57.1437 | 6.3 | 0.45 |
| polarC18 | 218.139 | 59.6443 | 6.3 | 0.45 |
| polarC18 | 218.139 | 101.4639 | 6.3 | 0.45 |
| polarC18 | 218.139 | 164.1438 | 6.3 | 0.45 |
| polarC18 | 218.139 | 105.0409 | 6.3 | 0.45 |
| polarC18 | 218.139 | 73.5236 | 6.3 | 0.45 |
| polarC18 | 218.139 | 101.5567 | 6.3 | 0.45 |
| polarC18 | 218.139 | 126.8168 | 6.3 | 0.45 |
| polarC18 | 218.139 | 159.715 | 6.3 | 0.45 |
| polarC18 | 218.139 | 58.429 | 6.3 | 0.45 |
| polarC18 | 218.139 | 125.5507 | 6.3 | 0.45 |
| polarC18 | 218.139 | 49.2855 | 6.3 | 0.45 |
| polarC18 | 218.139 | 48.6395 | 6.3 | 0.45 |
| polarC18 | 218.139 | 226.8498 | 6.3 | 0.45 |

| | | | | |
|---|---|---|---|---|
| polarC18 | 218.139 | 122.3063 | 6.3 | 0.45 |
| polarC18 | 218.139 | 69.3031 | 6.3 | 0.45 |
| polarC18 | 218.139 | 121.2308 | 6.3 | 0.45 |
| polarC18 | 218.139 | 70.4353 | 6.3 | 0.45 |
| polarC18 | 218.139 | 50.4464 | 6.3 | 0.45 |
| polarC18 | 218.139 | 179.1577 | 6.3 | 0.45 |
| polarC18 | 218.139 | 49.6804 | 6.3 | 0.45 |
| polarC18 | 218.139 | 92.9137 | 6.3 | 0.45 |
| polarC18 | 218.139 | 93.0518 | 6.3 | 0.45 |
| polarC18 | 218.139 | 53.849 | 6.3 | 0.45 |
| polarC18 | 218.139 | 73.6374 | 6.3 | 0.45 |
| polarC18 | 218.139 | 63.2593 | 6.3 | 0.45 |
| polarC18 | 218.139 | 72.3615 | 6.3 | 0.45 |
| polarC18 | 218.139 | 175.8733 | 6.3 | 0.45 |
| polarC18 | 218.139 | 53.8633 | 6.3 | 0.45 |
| polarC18 | 218.139 | 100.9059 | 6.3 | 0.45 |
| polarC18 | 218.139 | 92.2275 | 6.3 | 0.45 |
| polarC18 | 218.139 | 216.2471 | 6.3 | 0.45 |
| polarC18 | 218.139 | 54.276 | 6.3 | 0.45 |
| polarC18 | 218.139 | 192.0611 | 6.3 | 0.45 |
| polarC18 | 218.139 | 91.8975 | 6.3 | 0.45 |
| polarC18 | 218.139 | 49.3406 | 6.3 | 0.45 |
| polarC18 | 218.139 | 51.0917 | 6.3 | 0.45 |
| polarC18 | 218.139 | 152.9418 | 6.3 | 0.45 |
| polarC18 | 218.139 | 126.8406 | 6.3 | 0.45 |
| polarC18 | 218.139 | 72.2478 | 6.3 | 0.44 |
| polarC18 | 218.139 | 56.0643 | 6.3 | 0.44 |
| polarC18 | 218.139 | 78.0647 | 6.3 | 0.44 |
| polarC18 | 218.139 | 49.7333 | 6.3 | 0.44 |
| polarC18 | 218.139 | 62.1599 | 6.3 | 0.44 |
| polarC18 | 218.139 | 153.1126 | 6.3 | 0.44 |
| polarC18 | 218.139 | 75.684 | 6.3 | 0.44 |
| polarC18 | 218.139 | 55.7613 | 6.3 | 0.44 |
| polarC18 | 218.139 | 121.7078 | 6.3 | 0.44 |
| polarC18 | 218.139 | 119.6893 | 6.3 | 0.44 |
| polarC18 | 218.139 | 108.8976 | 6.3 | 0.44 |
| polarC18 | 218.139 | 240.2091 | 6.3 | 0.44 |
| polarC18 | 218.139 | 191.8706 | 6.3 | 0.44 |
| polarC18 | 218.139 | 237.1639 | 6.3 | 0.44 |
| polarC18 | 218.139 | 57.792 | 6.3 | 0.44 |
| polarC18 | 218.139 | 59.2815 | 6.3 | 0.44 |
| polarC18 | 218.139 | 193.9413 | 6.3 | 0.44 |

| | | | | |
|---|---|---|---|---|
| polarC18 | 218.139 | 104.7032 | 6.3 | 0.44 |
| polarC18 | 218.139 | 71.8237 | 6.3 | 0.44 |
| polarC18 | 218.139 | 58.6226 | 6.3 | 0.44 |
| polarC18 | 218.139 | 79.5127 | 6.3 | 0.44 |
| polarC18 | 218.139 | 62.9756 | 6.3 | 0.44 |
| polarC18 | 218.139 | 63.3669 | 6.3 | 0.44 |
| polarC18 | 218.139 | 90.1804 | 6.3 | 0.44 |
| polarC18 | 218.139 | 94.5455 | 6.3 | 0.44 |
| polarC18 | 218.139 | 55.8152 | 6.3 | 0.44 |
| polarC18 | 218.139 | 49.021 | 6.3 | 0.44 |
| polarC18 | 218.139 | 77.1741 | 6.3 | 0.44 |
| polarC18 | 218.139 | 124.3064 | 6.3 | 0.44 |
| polarC18 | 218.139 | 70.8229 | 6.3 | 0.44 |
| polarC18 | 218.139 | 80.1369 | 6.3 | 0.44 |
| polarC18 | 218.139 | 174.5643 | 6.3 | 0.44 |
| polarC18 | 218.139 | 56.4437 | 6.3 | 0.44 |
| polarC18 | 218.139 | 62.1722 | 6.3 | 0.44 |
| polarC18 | 218.139 | 76.6099 | 6.3 | 0.44 |
| polarC18 | 218.139 | 110.0272 | 6.3 | 0.44 |
| polarC18 | 218.139 | 104.6273 | 6.3 | 0.44 |
| polarC18 | 218.139 | 62.3747 | 6.3 | 0.44 |
| polarC18 | 218.139 | 56.9987 | 6.3 | 0.44 |
| polarC18 | 218.139 | 87.9495 | 6.3 | 0.44 |
| polarC18 | 218.139 | 69.4098 | 6.3 | 0.44 |
| polarC18 | 218.139 | 89.8748 | 6.3 | 0.44 |
| polarC18 | 218.139 | 196.1367 | 6.3 | 0.44 |
| polarC18 | 218.139 | 104.4743 | 6.3 | 0.44 |
| polarC18 | 218.139 | 139.2733 | 6.3 | 0.44 |
| polarC18 | 218.139 | 57.5476 | 6.3 | 0.44 |
| polarC18 | 218.139 | 77.5147 | 6.3 | 0.44 |
| polarC18 | 218.139 | 136.2646 | 6.3 | 0.44 |
| polarC18 | 218.139 | 104.3171 | 6.3 | 0.44 |
| polarC18 | 218.139 | 70.7077 | 6.3 | 0.44 |
| polarC18 | 218.139 | 86.0147 | 6.3 | 0.44 |
| polarC18 | 218.139 | 109.4547 | 6.3 | 0.44 |
| polarC18 | 218.139 | 90.1875 | 6.3 | 0.44 |
| polarC18 | 218.139 | 55.3046 | 6.3 | 0.44 |
| polarC18 | 218.139 | 104.4116 | 6.3 | 0.44 |
| polarC18 | 218.139 | 77.779 | 6.3 | 0.44 |
| polarC18 | 218.139 | 91.3679 | 6.3 | 0.44 |
| polarC18 | 218.139 | 60.1051 | 6.3 | 0.44 |
| polarC18 | 218.139 | 116.5838 | 6.2 | 0.44 |

| | | | | |
|---|---|---|---|---|
| polarC18 | 218.139 | 60.5605 | 6.2 | 0.44 |
| polarC18 | 218.139 | 51.0503 | 6.2 | 0.44 |
| polarC18 | 218.139 | 49.278 | 6.2 | 0.44 |
| polarC18 | 218.139 | 202.3367 | 6.2 | 0.44 |
| polarC18 | 218.139 | 143.3379 | 6.2 | 0.44 |
| polarC18 | 218.139 | 141.9253 | 6.2 | 0.44 |
| polarC18 | 218.139 | 67.3147 | 6.2 | 0.44 |
| polarC18 | 218.139 | 196.5382 | 6.2 | 0.44 |
| polarC18 | 218.139 | 193.0502 | 6.2 | 0.44 |
| polarC18 | 218.139 | 124.4085 | 6.2 | 0.44 |
| polarC18 | 218.139 | 59.8002 | 6.2 | 0.44 |
| polarC18 | 218.139 | 63.2206 | 6.2 | 0.44 |
| polarC18 | 218.139 | 214.2425 | 6.2 | 0.44 |
| polarC18 | 218.139 | 53.9819 | 6.2 | 0.44 |
| polarC18 | 218.139 | 84.8129 | 6.2 | 0.44 |
| polarC18 | 218.139 | 146.0189 | 6.2 | 0.44 |
| polarC18 | 218.139 | 83.1621 | 6.2 | 0.44 |
| polarC18 | 218.139 | 140.1419 | 6.2 | 0.44 |
| polarC18 | 218.139 | 165.023 | 6.2 | 0.44 |
| polarC18 | 218.139 | 144.5733 | 6.2 | 0.44 |
| polarC18 | 218.139 | 117.9758 | 6.2 | 0.44 |
| polarC18 | 218.139 | 137.0301 | 6.2 | 0.44 |
| polarC18 | 218.139 | 65.3084 | 6.2 | 0.44 |
| polarC18 | 218.139 | 97.6275 | 6.2 | 0.44 |
| polarC18 | 218.139 | 131.0372 | 6.2 | 0.44 |
| polarC18 | 218.139 | 78.3521 | 6.2 | 0.44 |
| polarC18 | 218.139 | 147.7063 | 6.2 | 0.44 |
| polarC18 | 218.139 | 138.6498 | 6.2 | 0.44 |
| polarC18 | 218.139 | 68.1337 | 6.2 | 0.44 |
| polarC18 | 218.139 | 53.1583 | 6.2 | 0.44 |
| polarC18 | 218.139 | 129.019 | 6.2 | 0.44 |
| polarC18 | 218.139 | 80.9783 | 6.2 | 0.44 |
| polarC18 | 218.139 | 144.766 | 6.2 | 0.44 |
| polarC18 | 218.139 | 65.8 | 6.2 | 0.44 |
| polarC18 | 218.139 | 115.1899 | 6.2 | 0.44 |
| polarC18 | 218.139 | 58.7419 | 6.2 | 0.44 |
| polarC18 | 218.139 | 80.2173 | 6.2 | 0.44 |
| polarC18 | 218.139 | 66.5844 | 6.2 | 0.44 |
| polarC18 | 218.139 | 111.2352 | 6.2 | 0.44 |
| polarC18 | 218.139 | 148.0388 | 6.2 | 0.44 |
| polarC18 | 218.139 | 87.8603 | 6.2 | 0.44 |
| polarC18 | 218.139 | 99.294 | 6.2 | 0.44 |

| | | | | |
|---|---|---|---|---|
| polarC18 | 218.139 | 98.8227 | 6.2 | 0.44 |
| polarC18 | 218.139 | 185.1328 | 6.2 | 0.44 |
| polarC18 | 218.139 | 64.1963 | 6.2 | 0.44 |
| polarC18 | 218.139 | 99.0733 | 6.2 | 0.44 |
| polarC18 | 218.139 | 74.8566 | 6.2 | 0.44 |
| polarC18 | 218.139 | 99.408 | 6.2 | 0.44 |
| polarC18 | 218.139 | 74.2304 | 6.2 | 0.44 |
| polarC18 | 218.139 | 61.116 | 6.2 | 0.44 |
| polarC18 | 218.139 | 58.8101 | 6.2 | 0.44 |
| polarC18 | 218.139 | 143.5876 | 6.2 | 0.44 |
| polarC18 | 218.139 | 55.6414 | 6.2 | 0.44 |
| polarC18 | 218.139 | 143.0888 | 6.2 | 0.44 |
| polarC18 | 218.139 | 55.9863 | 6.2 | 0.44 |
| polarC18 | 218.139 | 60.9643 | 6.2 | 0.44 |
| polarC18 | 218.139 | 93.566 | 6.2 | 0.44 |
| polarC18 | 218.139 | 70.1967 | 6.2 | 0.44 |
| polarC18 | 218.139 | 96.2332 | 6.2 | 0.44 |
| polarC18 | 218.139 | 57.3396 | 6.2 | 0.44 |
| polarC18 | 218.139 | 113.6948 | 6.2 | 0.44 |
| polarC18 | 218.139 | 191.6598 | 6.2 | 0.44 |
| polarC18 | 218.139 | 54.6311 | 6.2 | 0.44 |
| polarC18 | 218.139 | 168.9001 | 6.2 | 0.44 |
| polarC18 | 218.139 | 85.5254 | 6.2 | 0.44 |
| polarC18 | 218.139 | 50.0651 | 6.2 | 0.44 |
| polarC18 | 218.139 | 60.1229 | 6.2 | 0.44 |
| polarC18 | 218.139 | 121.7367 | 6.2 | 0.44 |
| polarC18 | 218.139 | 146.4191 | 6.2 | 0.44 |
| polarC18 | 218.139 | 59.6893 | 6.2 | 0.44 |
| polarC18 | 218.139 | 100.3146 | 6.2 | 0.44 |
| polarC18 | 218.139 | 62.8766 | 6.2 | 0.44 |
| polarC18 | 218.139 | 69.8949 | 6.2 | 0.44 |
| polarC18 | 218.139 | 66.9306 | 6.2 | 0.44 |
| polarC18 | 218.139 | 104.0162 | 6.2 | 0.44 |
| polarC18 | 218.139 | 56.1384 | 6.2 | 0.44 |
| polarC18 | 218.139 | 198.9996 | 6.2 | 0.44 |
| polarC18 | 218.139 | 111.4147 | 6.2 | 0.44 |
| polarC18 | 218.139 | 88.0991 | 6.2 | 0.44 |
| polarC18 | 218.139 | 114.6513 | 6.2 | 0.44 |
| polarC18 | 218.139 | 59.5391 | 6.2 | 0.44 |
| polarC18 | 218.139 | 237.8494 | 6.2 | 0.44 |
| polarC18 | 218.139 | 109.4443 | 6.2 | 0.44 |
| polarC18 | 218.139 | 129.7218 | 6.2 | 0.44 |

| | | | | |
|---|---|---|---|---|
| polarC18 | 218.139 | 155.6174 | 6.2 | 0.44 |
| polarC18 | 218.139 | 239.8613 | 6.2 | 0.44 |
| polarC18 | 218.139 | 48.637 | 6.2 | 0.44 |
| polarC18 | 218.139 | 55.1073 | 6.2 | 0.44 |
| polarC18 | 218.139 | 91.9876 | 6.2 | 0.44 |
| polarC18 | 218.139 | 224.3424 | 6.2 | 0.44 |
| polarC18 | 218.139 | 52.2815 | 6.2 | 0.44 |
| polarC18 | 218.139 | 64.9918 | 6.2 | 0.44 |
| polarC18 | 218.139 | 53.252 | 6.2 | 0.44 |
| polarC18 | 218.139 | 98.1416 | 6.2 | 0.44 |
| polarC18 | 218.139 | 188.8128 | 6.2 | 0.44 |
| polarC18 | 218.139 | 168.0628 | 6.2 | 0.44 |
| polarC18 | 218.139 | 83.5658 | 6.2 | 0.44 |
| polarC18 | 218.139 | 195.3587 | 6.2 | 0.44 |
| polarC18 | 218.139 | 238.2472 | 6.2 | 0.44 |
| polarC18 | 218.139 | 82.505 | 6.2 | 0.44 |
| polarC18 | 218.139 | 63.862 | 6.2 | 0.44 |
| polarC18 | 218.139 | 148.0166 | 6.2 | 0.44 |
| polarC18 | 218.139 | 167.1463 | 6.2 | 0.44 |
| polarC18 | 218.139 | 61.8651 | 6.2 | 0.44 |
| polarC18 | 218.139 | 73.3681 | 6.2 | 0.44 |
| polarC18 | 218.139 | 149.1999 | 6.2 | 0.44 |
| polarC18 | 218.139 | 217.6258 | 6.2 | 0.44 |
| polarC18 | 218.139 | 110.1617 | 6.2 | 0.44 |
| polarC18 | 218.139 | 216.9065 | 6.2 | 0.44 |
| polarC18 | 218.139 | 102.3177 | 6.2 | 0.44 |
| polarC18 | 218.139 | 64.8839 | 6.2 | 0.44 |
| polarC18 | 218.139 | 144.6313 | 6.2 | 0.44 |
| polarC18 | 218.139 | 69.6178 | 6.2 | 0.44 |
| polarC18 | 218.139 | 82.4583 | 6.2 | 0.44 |
| polarC18 | 218.139 | 55.8812 | 6.2 | 0.44 |
| polarC18 | 218.139 | 52.8595 | 6.2 | 0.44 |
| polarC18 | 218.139 | 80.0443 | 6.2 | 0.44 |
| polarC18 | 218.139 | 167.1305 | 6.2 | 0.44 |
| polarC18 | 218.139 | 74.8357 | 6.2 | 0.44 |
| polarC18 | 218.139 | 181.0153 | 6.2 | 0.44 |
| polarC18 | 218.139 | 72.7682 | 6.2 | 0.44 |
| polarC18 | 218.139 | 56.4652 | 6.2 | 0.44 |
| polarC18 | 218.139 | 57.8998 | 6.2 | 0.44 |
| polarC18 | 218.139 | 76.4097 | 6.2 | 0.44 |
| polarC18 | 218.139 | 113.1353 | 6.2 | 0.44 |
| polarC18 | 218.139 | 90.8378 | 6.2 | 0.44 |

| | | | | |
|---|---|---|---|---|
| polarC18 | 218.139 | 58.5596 | 6.2 | 0.44 |
| polarC18 | 218.139 | 158.8921 | 6.2 | 0.44 |
| polarC18 | 218.139 | 152.0016 | 6.2 | 0.44 |
| polarC18 | 218.139 | 123.8046 | 6.2 | 0.44 |
| polarC18 | 218.139 | 61.0508 | 6.2 | 0.44 |
| polarC18 | 218.139 | 93.8488 | 6.2 | 0.44 |
| polarC18 | 218.139 | 67.0128 | 6.2 | 0.44 |
| polarC18 | 218.139 | 74.2879 | 6.2 | 0.44 |
| polarC18 | 218.139 | 104.1078 | 6.2 | 0.44 |
| polarC18 | 218.139 | 55.9482 | 6.2 | 0.44 |
| polarC18 | 218.139 | 179.7469 | 6.2 | 0.44 |
| polarC18 | 218.139 | 56.0258 | 6.2 | 0.44 |
| polarC18 | 218.139 | 93.6002 | 6.2 | 0.44 |
| polarC18 | 218.139 | 64.2121 | 6.2 | 0.44 |
| polarC18 | 218.139 | 174.9554 | 6.2 | 0.44 |
| polarC18 | 218.139 | 140.775 | 6.2 | 0.44 |
| polarC18 | 218.139 | 64.8576 | 6.2 | 0.44 |
| polarC18 | 218.139 | 155.266 | 6.2 | 0.44 |
| polarC18 | 218.139 | 230.1711 | 6.2 | 0.44 |
| polarC18 | 218.139 | 51.3371 | 6.2 | 0.44 |
| polarC18 | 218.139 | 195.5932 | 6.2 | 0.44 |
| polarC18 | 218.139 | 59.7518 | 6.2 | 0.44 |
| polarC18 | 218.139 | 54.5778 | 6.2 | 0.44 |
| polarC18 | 218.139 | 53.3266 | 6.2 | 0.44 |
| polarC18 | 218.139 | 214.1857 | 6.2 | 0.44 |
| polarC18 | 218.139 | 53.0928 | 6.2 | 0.44 |
| polarC18 | 218.139 | 48.9557 | 6.2 | 0.44 |
| polarC18 | 218.139 | 155.0896 | 6.2 | 0.44 |
| polarC18 | 218.139 | 51.9407 | 6.2 | 0.44 |
| polarC18 | 218.139 | 85.875 | 6.2 | 0.44 |
| polarC18 | 218.139 | 92.2637 | 6.2 | 0.44 |
| polarC18 | 218.139 | 54.7184 | 6.2 | 0.44 |
| polarC18 | 218.139 | 103.0282 | 6.2 | 0.44 |
| polarC18 | 218.139 | 117.0344 | 6.2 | 0.44 |
| polarC18 | 218.139 | 54.693 | 6.2 | 0.44 |
| polarC18 | 218.139 | 64.6605 | 6.2 | 0.44 |
| polarC18 | 218.139 | 93.5119 | 6.2 | 0.44 |
| polarC18 | 218.139 | 91.1115 | 6.2 | 0.44 |
| polarC18 | 218.139 | 56.3868 | 6.2 | 0.44 |
| polarC18 | 218.139 | 48.7593 | 6.2 | 0.44 |
| polarC18 | 218.139 | 78.2142 | 6.2 | 0.44 |
| polarC18 | 218.139 | 55.0877 | 6.2 | 0.44 |

| | | | | |
|---|---|---|---|---|
| polarC18 | 218.139 | 74.9431 | 6.2 | 0.44 |
| polarC18 | 218.139 | 68.0618 | 6.2 | 0.44 |
| polarC18 | 218.139 | 88.3538 | 6.2 | 0.44 |
| polarC18 | 218.139 | 51.903 | 6.2 | 0.44 |
| polarC18 | 218.139 | 207.2406 | 6.2 | 0.44 |
| polarC18 | 218.139 | 131.3831 | 6.2 | 0.44 |
| polarC18 | 218.139 | 64.0605 | 6.2 | 0.44 |
| polarC18 | 218.139 | 96.0205 | 6.2 | 0.44 |
| polarC18 | 218.139 | 73.1468 | 6.2 | 0.44 |
| polarC18 | 218.139 | 80.761 | 6.2 | 0.44 |
| polarC18 | 218.139 | 72.0447 | 6.2 | 0.44 |
| polarC18 | 218.139 | 178.1575 | 6.2 | 0.44 |
| polarC18 | 218.139 | 65.9765 | 6.2 | 0.44 |
| polarC18 | 218.139 | 58.1007 | 6.2 | 0.44 |
| polarC18 | 218.139 | 57.5597 | 6.2 | 0.44 |
| polarC18 | 218.139 | 165.569 | 6.2 | 0.44 |
| polarC18 | 218.139 | 62.2393 | 6.2 | 0.44 |
| polarC18 | 218.139 | 121.2561 | 6.2 | 0.44 |
| polarC18 | 218.139 | 180.9764 | 6.2 | 0.44 |
| polarC18 | 218.139 | 163.462 | 6.2 | 0.44 |
| polarC18 | 218.139 | 68.7536 | 6.2 | 0.44 |
| polarC18 | 218.139 | 58.4474 | 6.2 | 0.44 |
| polarC18 | 218.139 | 94.3826 | 6.2 | 0.44 |
| polarC18 | 218.139 | 68.8805 | 6.2 | 0.44 |
| polarC18 | 218.139 | 113.0327 | 6.2 | 0.44 |
| polarC18 | 218.139 | 180.2072 | 6.2 | 0.44 |
| polarC18 | 218.139 | 116.8314 | 6.2 | 0.44 |
| polarC18 | 218.139 | 119.124 | 6.2 | 0.44 |
| polarC18 | 218.139 | 66.4821 | 6.2 | 0.44 |
| polarC18 | 218.139 | 83.6963 | 6.2 | 0.44 |
| polarC18 | 218.139 | 65.3194 | 6.2 | 0.44 |
| polarC18 | 218.139 | 136.8252 | 6.2 | 0.44 |
| polarC18 | 218.139 | 49.5831 | 6.2 | 0.44 |
| polarC18 | 218.139 | 86.1077 | 6.2 | 0.44 |
| polarC18 | 218.139 | 198.2433 | 6.2 | 0.44 |
| polarC18 | 218.139 | 125.4862 | 6.2 | 0.44 |
| polarC18 | 218.139 | 83.3656 | 6.2 | 0.44 |
| polarC18 | 218.139 | 118.4743 | 6.2 | 0.44 |
| polarC18 | 218.139 | 69.3597 | 6.2 | 0.44 |
| polarC18 | 218.139 | 84.5996 | 6.2 | 0.44 |
| polarC18 | 218.139 | 218.4395 | 6.2 | 0.44 |
| polarC18 | 218.139 | 241.7545 | 6.2 | 0.44 |

| | | | | |
|---|---|---|---|---|
| polarC18 | 218.139 | 164.3032 | 6.2 | 0.44 |
| polarC18 | 218.139 | 145.9742 | 6.2 | 0.44 |
| polarC18 | 218.139 | 123.6883 | 6.2 | 0.44 |
| polarC18 | 218.139 | 112.812 | 6.2 | 0.44 |
| polarC18 | 218.139 | 120.0186 | 6.2 | 0.44 |
| polarC18 | 218.139 | 48.6615 | 6.2 | 0.44 |
| polarC18 | 218.139 | 57.6053 | 6.2 | 0.44 |
| polarC18 | 218.139 | 80.3683 | 6.2 | 0.44 |
| polarC18 | 218.139 | 64.1404 | 6.2 | 0.44 |
| polarC18 | 218.139 | 163.2269 | 6.2 | 0.44 |
| polarC18 | 218.139 | 51.1236 | 6.2 | 0.44 |
| polarC18 | 218.139 | 97.913 | 6.2 | 0.44 |
| polarC18 | 218.139 | 242.7277 | 6.2 | 0.44 |
| polarC18 | 218.139 | 108.9623 | 6.2 | 0.44 |
| polarC18 | 218.139 | 221.6103 | 6.2 | 0.44 |
| polarC18 | 218.139 | 65.6804 | 6.2 | 0.44 |
| polarC18 | 218.139 | 126.5494 | 6.2 | 0.44 |
| polarC18 | 218.139 | 54.1864 | 6.2 | 0.44 |
| polarC18 | 218.139 | 118.9648 | 6.2 | 0.44 |
| polarC18 | 218.139 | 57.1907 | 6.2 | 0.44 |
| polarC18 | 218.139 | 73.098 | 6.2 | 0.44 |
| polarC18 | 218.139 | 105.3063 | 6.2 | 0.44 |
| polarC18 | 218.139 | 50.432 | 6.2 | 0.44 |
| polarC18 | 218.139 | 127.8338 | 6.2 | 0.44 |
| polarC18 | 218.139 | 57.1135 | 6.2 | 0.44 |
| polarC18 | 218.139 | 64.073 | 6.2 | 0.44 |
| polarC18 | 218.139 | 59.3783 | 6.2 | 0.44 |
| polarC18 | 218.139 | 73.2437 | 6.2 | 0.44 |
| polarC18 | 218.139 | 49.3378 | 6.2 | 0.44 |
| polarC18 | 218.139 | 87.7813 | 6.2 | 0.44 |
| polarC18 | 218.139 | 59.3108 | 6.2 | 0.44 |
| polarC18 | 218.139 | 58.031 | 6.2 | 0.44 |
| polarC18 | 218.139 | 150.2574 | 6.2 | 0.44 |
| polarC18 | 218.139 | 181.4541 | 6.2 | 0.44 |
| polarC18 | 218.139 | 131.5371 | 6.2 | 0.44 |
| polarC18 | 218.139 | 61.9346 | 6.2 | 0.44 |
| polarC18 | 218.139 | 72.5893 | 6.2 | 0.44 |
| polarC18 | 218.139 | 50.2404 | 6.2 | 0.44 |
| polarC18 | 218.139 | 66.341 | 6.2 | 0.44 |
| polarC18 | 218.139 | 162.9469 | 6.2 | 0.44 |
| polarC18 | 218.139 | 129.7767 | 6.2 | 0.44 |
| polarC18 | 218.139 | 67.951 | 6.2 | 0.44 |

| | | | | |
|---|---|---|---|---|
| polarC18 | 218.139 | 94.8819 | 6.2 | 0.44 |
| polarC18 | 218.139 | 57.423 | 6.2 | 0.44 |
| polarC18 | 218.139 | 57.0767 | 6.2 | 0.44 |
| polarC18 | 218.139 | 50.8596 | 6.2 | 0.44 |
| polarC18 | 218.139 | 175.5073 | 6.2 | 0.44 |
| polarC18 | 218.139 | 85.186 | 6.2 | 0.44 |
| polarC18 | 218.139 | 104.5722 | 6.2 | 0.44 |
| polarC18 | 218.139 | 101.4965 | 6.2 | 0.44 |
| polarC18 | 218.139 | 91.0281 | 6.2 | 0.44 |
| polarC18 | 218.139 | 63.9159 | 6.2 | 0.44 |
| polarC18 | 218.139 | 62.3122 | 6.2 | 0.44 |
| polarC18 | 218.139 | 148.7216 | 6.2 | 0.44 |
| polarC18 | 218.139 | 49.1339 | 6.2 | 0.44 |
| polarC18 | 218.139 | 69.0524 | 6.2 | 0.44 |
| polarC18 | 218.139 | 61.7455 | 6.2 | 0.44 |
| polarC18 | 218.139 | 87.4347 | 6.2 | 0.44 |
| polarC18 | 218.139 | 125.1426 | 6.2 | 0.44 |
| polarC18 | 218.139 | 68.3501 | 6.2 | 0.44 |
| polarC18 | 218.139 | 67.9835 | 6.2 | 0.44 |
| polarC18 | 218.139 | 56.4066 | 6.2 | 0.44 |
| polarC18 | 218.139 | 98.6605 | 6.2 | 0.44 |
| polarC18 | 218.139 | 66.5378 | 6.2 | 0.44 |
| polarC18 | 218.139 | 133.6396 | 6.2 | 0.44 |
| polarC18 | 218.139 | 55.7023 | 6.2 | 0.44 |
| polarC18 | 218.139 | 121.6883 | 6.2 | 0.44 |
| polarC18 | 218.139 | 50.8221 | 6.2 | 0.44 |
| polarC18 | 218.139 | 76.26 | 6.2 | 0.44 |
| polarC18 | 218.139 | 60.3226 | 6.2 | 0.44 |
| polarC18 | 218.139 | 63.5259 | 6.2 | 0.44 |
| polarC18 | 218.139 | 87.6798 | 6.2 | 0.44 |
| polarC18 | 218.139 | 61.4302 | 6.2 | 0.44 |
| polarC18 | 218.139 | 175.9318 | 6.2 | 0.44 |
| polarC18 | 218.139 | 80.9939 | 6.1 | 0.44 |
| polarC18 | 218.139 | 180.0651 | 6.1 | 0.44 |
| polarC18 | 218.139 | 49.8063 | 6.1 | 0.44 |
| polarC18 | 218.139 | 62.5213 | 6.1 | 0.44 |
| polarC18 | 218.139 | 116.4556 | 6.1 | 0.44 |
| polarC18 | 218.139 | 54.9387 | 6.1 | 0.44 |
| polarC18 | 218.139 | 71.2657 | 6.1 | 0.44 |
| polarC18 | 218.139 | 84.0289 | 6.1 | 0.44 |
| polarC18 | 218.139 | 74.6435 | 6.1 | 0.44 |
| polarC18 | 218.139 | 107.7961 | 6.1 | 0.44 |

| | | | | |
|---|---|---|---|---|
| polarC18 | 218.139 | 88.344 | 6.1 | 0.44 |
| polarC18 | 218.139 | 49.2897 | 6.1 | 0.44 |
| polarC18 | 218.139 | 52.1067 | 6.1 | 0.44 |
| polarC18 | 218.139 | 56.1993 | 6.1 | 0.44 |
| polarC18 | 218.139 | 57.3126 | 6.1 | 0.44 |
| polarC18 | 218.139 | 67.0313 | 6.1 | 0.44 |
| polarC18 | 218.139 | 70.9849 | 6.1 | 0.44 |
| polarC18 | 218.139 | 83.4055 | 6.1 | 0.44 |
| polarC18 | 218.139 | 65.5622 | 6.1 | 0.44 |
| polarC18 | 218.139 | 187.6934 | 6.1 | 0.44 |
| polarC18 | 218.139 | 124.8394 | 6.1 | 0.44 |
| polarC18 | 218.139 | 51.6181 | 6.1 | 0.44 |
| polarC18 | 218.139 | 66.8156 | 6.1 | 0.44 |
| polarC18 | 218.139 | 124.9814 | 6.1 | 0.44 |
| polarC18 | 218.139 | 152.7604 | 6.1 | 0.44 |
| polarC18 | 218.139 | 96.9799 | 6.1 | 0.44 |
| polarC18 | 218.139 | 92.13 | 6.1 | 0.44 |
| polarC18 | 218.139 | 53.6781 | 6.1 | 0.44 |
| polarC18 | 218.139 | 87.9625 | 6.1 | 0.44 |
| polarC18 | 218.139 | 53.4724 | 6.1 | 0.44 |
| polarC18 | 218.139 | 53.2861 | 6.1 | 0.44 |
| polarC18 | 218.139 | 70.7677 | 6.1 | 0.44 |
| polarC18 | 218.139 | 54.0522 | 6.1 | 0.44 |
| polarC18 | 218.139 | 67.1461 | 6.1 | 0.44 |
| polarC18 | 218.139 | 89.8163 | 6.1 | 0.44 |
| polarC18 | 218.139 | 89.7836 | 6.1 | 0.44 |
| polarC18 | 218.139 | 66.224 | 6.1 | 0.44 |
| polarC18 | 218.139 | 108.8217 | 6.1 | 0.44 |
| polarC18 | 218.139 | 54.9756 | 6.1 | 0.44 |
| polarC18 | 218.139 | 175.2165 | 6.1 | 0.44 |
| polarC18 | 218.139 | 84.5639 | 6.1 | 0.44 |
| polarC18 | 218.139 | 131.203 | 6.1 | 0.44 |
| polarC18 | 218.139 | 72.8285 | 6.1 | 0.44 |
| polarC18 | 218.139 | 50.921 | 6.1 | 0.44 |
| polarC18 | 218.139 | 52.176 | 6.1 | 0.43 |
| polarC18 | 218.139 | 102.2815 | 6.1 | 0.43 |
| polarC18 | 218.139 | 159.252 | 6.1 | 0.43 |
| polarC18 | 218.139 | 53.8404 | 6.1 | 0.43 |
| polarC18 | 218.139 | 66.7994 | 6.1 | 0.43 |
| polarC18 | 218.139 | 103.7921 | 6.1 | 0.43 |
| polarC18 | 218.139 | 68.1823 | 6.1 | 0.43 |
| polarC18 | 218.139 | 110.8801 | 6.1 | 0.43 |

| | | | | |
|---|---|---|---|---|
| polarC18 | 218.139 | 55.3764 | 6.1 | 0.43 |
| polarC18 | 218.139 | 103.4793 | 6.1 | 0.43 |
| polarC18 | 218.139 | 132.365 | 6.1 | 0.43 |
| polarC18 | 218.139 | 54.3224 | 6.1 | 0.43 |
| polarC18 | 218.139 | 232.7417 | 6.1 | 0.43 |
| polarC18 | 218.139 | 80.9535 | 6.1 | 0.43 |
| polarC18 | 218.139 | 49.5487 | 6.1 | 0.43 |
| polarC18 | 218.139 | 150.2114 | 6.1 | 0.43 |
| polarC18 | 218.139 | 69.8415 | 6.1 | 0.43 |
| polarC18 | 218.139 | 154.1847 | 6.1 | 0.43 |
| polarC18 | 218.139 | 107.0143 | 6.1 | 0.43 |
| polarC18 | 218.139 | 91.2307 | 6.1 | 0.43 |
| polarC18 | 218.139 | 92.151 | 6.1 | 0.43 |
| polarC18 | 218.139 | 81.3421 | 6.1 | 0.43 |
| polarC18 | 218.139 | 48.602 | 6.1 | 0.43 |
| polarC18 | 218.139 | 81.3865 | 6.1 | 0.43 |
| polarC18 | 218.139 | 101.9039 | 6.1 | 0.43 |
| polarC18 | 218.139 | 118.6339 | 6.1 | 0.43 |
| polarC18 | 218.139 | 80.2675 | 6.1 | 0.43 |
| polarC18 | 218.139 | 63.7807 | 6.1 | 0.43 |
| polarC18 | 218.139 | 66.5529 | 6.1 | 0.43 |
| polarC18 | 218.139 | 101.3502 | 6.1 | 0.43 |
| polarC18 | 218.139 | 72.8346 | 6.1 | 0.43 |
| polarC18 | 218.139 | 101.0719 | 6.1 | 0.43 |
| polarC18 | 218.139 | 66.0243 | 6.1 | 0.43 |
| polarC18 | 218.139 | 141.7629 | 6.1 | 0.43 |
| polarC18 | 218.139 | 144.4609 | 6.1 | 0.43 |
| polarC18 | 218.139 | 158.2129 | 6.1 | 0.43 |
| polarC18 | 218.139 | 199.3864 | 6.1 | 0.43 |
| polarC18 | 218.139 | 83.8851 | 6.1 | 0.43 |
| polarC18 | 218.139 | 50.4179 | 6.1 | 0.43 |
| polarC18 | 218.139 | 194.9117 | 6.1 | 0.43 |
| polarC18 | 218.139 | 75.3239 | 6.1 | 0.43 |
| polarC18 | 218.139 | 89.2256 | 6.1 | 0.43 |
| polarC18 | 218.139 | 108.0385 | 6.1 | 0.43 |
| polarC18 | 218.139 | 166.7382 | 6.1 | 0.43 |
| polarC18 | 218.139 | 80.1706 | 6.1 | 0.43 |
| polarC18 | 218.139 | 60.8845 | 6.1 | 0.43 |
| polarC18 | 218.139 | 60.7057 | 6.1 | 0.43 |
| polarC18 | 218.139 | 62.3965 | 6.1 | 0.43 |
| polarC18 | 218.139 | 51.1631 | 6.1 | 0.43 |
| polarC18 | 218.139 | 68.81 | 6.1 | 0.43 |

| | | | | |
|---|---|---|---|---|
| polarC18 | 218.139 | 56.5135 | 6.1 | 0.43 |
| polarC18 | 218.139 | 68.9818 | 6.1 | 0.43 |
| polarC18 | 218.139 | 187.372 | 6.1 | 0.43 |
| polarC18 | 218.139 | 91.7286 | 6.1 | 0.43 |
| polarC18 | 218.139 | 54.2445 | 6.1 | 0.43 |
| polarC18 | 218.139 | 73.4699 | 6.1 | 0.43 |
| polarC18 | 218.139 | 85.2141 | 6.1 | 0.43 |
| polarC18 | 218.139 | 71.7901 | 6.1 | 0.43 |
| polarC18 | 218.139 | 51.8514 | 6.1 | 0.43 |
| polarC18 | 218.139 | 59.4367 | 6.1 | 0.43 |
| polarC18 | 218.139 | 228.8598 | 6.1 | 0.43 |
| polarC18 | 218.139 | 64.2435 | 6.1 | 0.43 |
| polarC18 | 218.139 | 69.6298 | 6.1 | 0.43 |
| polarC18 | 218.139 | 75.6072 | 6.1 | 0.43 |
| polarC18 | 218.139 | 103.3767 | 6.1 | 0.43 |
| polarC18 | 218.139 | 126.9366 | 6.1 | 0.43 |
| polarC18 | 218.139 | 114.9315 | 6.1 | 0.43 |
| polarC18 | 218.139 | 209.9095 | 6.1 | 0.43 |
| polarC18 | 218.139 | 92.7656 | 6.1 | 0.43 |
| polarC18 | 218.139 | 118.6168 | 6.1 | 0.43 |
| polarC18 | 218.139 | 133.107 | 6.1 | 0.43 |
| polarC18 | 218.139 | 127.3857 | 6.1 | 0.43 |
| polarC18 | 218.139 | 104.9492 | 6.1 | 0.43 |
| polarC18 | 218.139 | 117.2179 | 6.1 | 0.43 |
| polarC18 | 218.139 | 104.5236 | 6.1 | 0.43 |
| polarC18 | 218.139 | 60.1543 | 6.1 | 0.43 |
| polarC18 | 218.139 | 60.7258 | 6.1 | 0.43 |
| polarC18 | 218.139 | 63.8687 | 6.1 | 0.43 |
| polarC18 | 218.139 | 193.1753 | 6.1 | 0.43 |
| polarC18 | 218.139 | 132.4221 | 6.1 | 0.43 |
| polarC18 | 218.139 | 132.609 | 6.1 | 0.43 |
| polarC18 | 218.139 | 81.5322 | 6.1 | 0.43 |
| polarC18 | 218.139 | 51.4441 | 6.1 | 0.43 |
| polarC18 | 218.139 | 117.9884 | 6.1 | 0.43 |
| polarC18 | 218.139 | 58.8125 | 6.1 | 0.43 |
| polarC18 | 218.139 | 97.5097 | 6.1 | 0.43 |
| polarC18 | 218.139 | 73.866 | 6.1 | 0.43 |
| polarC18 | 218.139 | 52.6122 | 6.1 | 0.43 |
| polarC18 | 218.139 | 56.078 | 6.1 | 0.43 |
| polarC18 | 218.139 | 128.4366 | 6.1 | 0.43 |
| polarC18 | 218.139 | 117.7056 | 6.1 | 0.43 |
| polarC18 | 218.139 | 68.1248 | 6.1 | 0.43 |

| | | | | |
|---|---|---|---|---|
| polarC18 | 218.139 | 74.0309 | 6.1 | 0.43 |
| polarC18 | 218.139 | 53.7892 | 6.1 | 0.43 |
| polarC18 | 218.139 | 74.4681 | 6.1 | 0.43 |
| polarC18 | 218.139 | 152.1186 | 6.1 | 0.43 |
| polarC18 | 218.139 | 132.6451 | 6.1 | 0.43 |
| polarC18 | 218.139 | 71.135 | 6.1 | 0.43 |
| polarC18 | 218.139 | 75.7215 | 6.1 | 0.43 |
| polarC18 | 218.139 | 55.6326 | 6.1 | 0.43 |
| polarC18 | 218.139 | 53.7097 | 6.1 | 0.43 |
| polarC18 | 218.139 | 241.8718 | 6.1 | 0.43 |
| polarC18 | 218.139 | 118.183 | 6.1 | 0.43 |
| polarC18 | 218.139 | 62.7565 | 6.1 | 0.43 |
| polarC18 | 218.139 | 49.3183 | 6.1 | 0.43 |
| polarC18 | 218.139 | 48.7385 | 6.1 | 0.43 |
| polarC18 | 218.139 | 176.6821 | 6.1 | 0.43 |
| polarC18 | 218.139 | 63.0789 | 6.1 | 0.43 |
| polarC18 | 218.139 | 110.116 | 6.1 | 0.43 |
| polarC18 | 218.139 | 145.6731 | 6.1 | 0.43 |
| polarC18 | 218.139 | 129.5485 | 6.1 | 0.43 |
| polarC18 | 218.139 | 91.28 | 6.1 | 0.43 |
| polarC18 | 218.139 | 69.822 | 6.1 | 0.43 |
| polarC18 | 218.139 | 66.0775 | 6.1 | 0.43 |
| polarC18 | 218.139 | 186.6485 | 6.1 | 0.43 |
| polarC18 | 218.139 | 83.0999 | 6.1 | 0.43 |
| polarC18 | 218.139 | 72.2752 | 6.1 | 0.43 |
| polarC18 | 218.139 | 48.696 | 6.1 | 0.43 |
| polarC18 | 218.139 | 94.0618 | 6.1 | 0.43 |
| polarC18 | 218.139 | 59.4766 | 6.1 | 0.43 |
| polarC18 | 218.139 | 231.3156 | 6.1 | 0.43 |
| polarC18 | 218.139 | 54.5817 | 6.1 | 0.43 |
| polarC18 | 218.139 | 66.6986 | 6.1 | 0.43 |
| polarC18 | 218.139 | 209.0862 | 6.1 | 0.43 |
| polarC18 | 218.139 | 54.1837 | 6.1 | 0.43 |
| polarC18 | 218.139 | 79.7108 | 6.1 | 0.43 |
| polarC18 | 218.139 | 53.9216 | 6.1 | 0.43 |
| polarC18 | 218.139 | 58.4624 | 6.1 | 0.43 |
| polarC18 | 218.139 | 169.2538 | 6.1 | 0.43 |
| polarC18 | 218.139 | 75.7652 | 6.1 | 0.43 |
| polarC18 | 218.139 | 65.8655 | 6.1 | 0.43 |
| polarC18 | 218.139 | 74.7239 | 6.1 | 0.43 |
| polarC18 | 218.139 | 69.1698 | 6.1 | 0.43 |
| polarC18 | 218.139 | 51.1047 | 6.1 | 0.43 |

| | | | | |
|---|---|---|---|---|
| polarC18 | 218.139 | 48.9089 | 6.1 | 0.43 |
| polarC18 | 218.139 | 159.5327 | 6.1 | 0.43 |
| polarC18 | 218.139 | 179.5468 | 6.1 | 0.43 |
| polarC18 | 218.139 | 111.8178 | 6.1 | 0.43 |
| polarC18 | 218.139 | 102.7939 | 6.1 | 0.43 |
| polarC18 | 218.139 | 86.9729 | 6.1 | 0.43 |
| polarC18 | 218.139 | 79.668 | 6.1 | 0.43 |
| polarC18 | 218.139 | 97.6009 | 6.1 | 0.43 |
| polarC18 | 218.139 | 241.0119 | 6.1 | 0.43 |
| polarC18 | 218.139 | 67.9625 | 6.1 | 0.43 |
| polarC18 | 218.139 | 48.7214 | 6.1 | 0.43 |
| polarC18 | 218.139 | 113.2504 | 6.1 | 0.43 |
| polarC18 | 218.139 | 90.6333 | 6.1 | 0.43 |
| polarC18 | 218.139 | 60.1351 | 6.1 | 0.43 |
| polarC18 | 218.139 | 59.3333 | 6.1 | 0.43 |
| polarC18 | 218.139 | 81.2113 | 6.1 | 0.43 |
| polarC18 | 218.139 | 110.1481 | 6.1 | 0.43 |
| polarC18 | 218.139 | 74.9721 | 6.1 | 0.43 |
| polarC18 | 218.139 | 82.1941 | 6.1 | 0.43 |
| polarC18 | 218.139 | 53.0548 | 6.1 | 0.43 |
| polarC18 | 218.139 | 55.5609 | 6.1 | 0.43 |
| polarC18 | 218.139 | 66.8024 | 6.1 | 0.43 |
| polarC18 | 218.139 | 80.6572 | 6.1 | 0.43 |
| polarC18 | 218.139 | 65.7561 | 6.1 | 0.43 |
| polarC18 | 218.139 | 124.6053 | 6.1 | 0.43 |
| polarC18 | 218.139 | 62.1926 | 6.1 | 0.43 |
| polarC18 | 218.139 | 59.3278 | 6.1 | 0.43 |
| polarC18 | 218.139 | 65.7314 | 6.1 | 0.43 |
| polarC18 | 218.139 | 204.6611 | 6.1 | 0.43 |
| polarC18 | 218.139 | 75.8262 | 6.1 | 0.43 |
| polarC18 | 218.139 | 78.6727 | 6.1 | 0.43 |
| polarC18 | 218.139 | 50.0147 | 6.1 | 0.43 |
| polarC18 | 218.139 | 89.5066 | 6.1 | 0.43 |
| polarC18 | 218.139 | 66.4689 | 6.1 | 0.43 |
| polarC18 | 218.139 | 64.2293 | 6.1 | 0.43 |
| polarC18 | 218.139 | 55.7168 | 6.1 | 0.43 |
| polarC18 | 218.139 | 143.4326 | 6.1 | 0.43 |
| polarC18 | 218.139 | 95.865 | 6.1 | 0.43 |
| polarC18 | 218.139 | 77.982 | 6.1 | 0.43 |
| polarC18 | 218.139 | 51.849 | 6.1 | 0.43 |
| polarC18 | 218.139 | 75.5801 | 6.1 | 0.43 |
| polarC18 | 218.139 | 109.0387 | 6.1 | 0.43 |

| | | | | |
|---|---|---|---|---|
| polarC18 | 218.139 | 54.2003 | 6.1 | 0.43 |
| polarC18 | 218.139 | 71.9399 | 6.1 | 0.43 |
| polarC18 | 218.139 | 93.6296 | 6.1 | 0.43 |
| polarC18 | 218.139 | 53.5477 | 6.1 | 0.43 |
| polarC18 | 218.139 | 49.7688 | 6.1 | 0.43 |
| polarC18 | 218.139 | 96.0941 | 6.1 | 0.43 |
| polarC18 | 218.139 | 56.4978 | 6.1 | 0.43 |
| polarC18 | 218.139 | 54.4102 | 6.1 | 0.43 |
| polarC18 | 218.139 | 133.5739 | 6.1 | 0.43 |
| polarC18 | 218.139 | 76.241 | 6.1 | 0.43 |
| polarC18 | 218.139 | 73.5895 | 6.1 | 0.43 |
| polarC18 | 218.139 | 135.6364 | 6.1 | 0.43 |
| polarC18 | 218.139 | 136.5058 | 6.1 | 0.43 |
| polarC18 | 218.139 | 56.5232 | 6.1 | 0.43 |
| polarC18 | 218.139 | 72.919 | 6.1 | 0.43 |
| polarC18 | 218.139 | 114.6705 | 6.1 | 0.43 |
| polarC18 | 218.139 | 91.2465 | 6.1 | 0.43 |
| polarC18 | 218.139 | 82.1442 | 6.1 | 0.43 |
| polarC18 | 218.139 | 240.3809 | 6.1 | 0.43 |
| polarC18 | 218.139 | 55.4194 | 6.1 | 0.43 |
| polarC18 | 218.139 | 62.4854 | 6.1 | 0.43 |
| polarC18 | 218.139 | 63.7165 | 6.1 | 0.43 |
| polarC18 | 218.139 | 63.7946 | 6.1 | 0.43 |
| polarC18 | 218.139 | 74.5392 | 6.1 | 0.43 |
| polarC18 | 218.139 | 104.052 | 6.1 | 0.43 |
| polarC18 | 218.139 | 207.0355 | 6.1 | 0.43 |
| polarC18 | 218.139 | 68.9883 | 6.1 | 0.43 |
| polarC18 | 218.139 | 145.072 | 6.1 | 0.43 |
| polarC18 | 218.139 | 83.7429 | 6.1 | 0.43 |
| polarC18 | 218.139 | 58.9522 | 6.1 | 0.43 |
| polarC18 | 218.139 | 60.7025 | 6.1 | 0.43 |
| polarC18 | 218.139 | 89.0237 | 6.1 | 0.43 |
| polarC18 | 218.139 | 154.5944 | 6 | 0.43 |
| polarC18 | 218.139 | 66.7188 | 6 | 0.43 |
| polarC18 | 218.139 | 190.2881 | 6 | 0.43 |
| polarC18 | 218.139 | 57.3266 | 6 | 0.43 |
| polarC18 | 218.139 | 50.4416 | 6 | 0.43 |
| polarC18 | 218.139 | 173.5377 | 6 | 0.43 |
| polarC18 | 218.139 | 62.9895 | 6 | 0.43 |
| polarC18 | 218.139 | 151.1619 | 6 | 0.43 |
| polarC18 | 218.139 | 55.0022 | 6 | 0.43 |
| polarC18 | 218.139 | 96.776 | 6 | 0.43 |

| | | | | |
|---|---|---|---|---|
| polarC18 | 218.139 | 56.6546 | 6 | 0.43 |
| polarC18 | 218.139 | 124.3582 | 6 | 0.43 |
| polarC18 | 218.139 | 60.6497 | 6 | 0.43 |
| polarC18 | 218.139 | 71.4654 | 6 | 0.43 |
| polarC18 | 218.139 | 61.4935 | 6 | 0.43 |
| polarC18 | 218.139 | 55.9627 | 6 | 0.43 |
| polarC18 | 218.139 | 56.2959 | 6 | 0.43 |
| polarC18 | 218.139 | 68.8227 | 6 | 0.43 |
| polarC18 | 218.139 | 67.5568 | 6 | 0.43 |
| polarC18 | 218.139 | 60.1117 | 6 | 0.43 |
| polarC18 | 218.139 | 56.0917 | 6 | 0.43 |
| polarC18 | 218.139 | 57.2157 | 6 | 0.43 |
| polarC18 | 218.139 | 117.999 | 6 | 0.43 |
| polarC18 | 218.139 | 59.9063 | 6 | 0.43 |
| polarC18 | 218.139 | 132.8682 | 6 | 0.43 |
| polarC18 | 218.139 | 106.3109 | 6 | 0.43 |
| polarC18 | 218.139 | 72.1736 | 6 | 0.43 |
| polarC18 | 218.139 | 59.8962 | 6 | 0.43 |
| polarC18 | 218.139 | 225.258 | 6 | 0.43 |
| polarC18 | 218.139 | 183.8844 | 6 | 0.43 |
| polarC18 | 218.139 | 58.457 | 6 | 0.43 |
| polarC18 | 218.139 | 188.6797 | 6 | 0.43 |
| polarC18 | 218.139 | 109.5435 | 6 | 0.43 |
| polarC18 | 218.139 | 63.0765 | 6 | 0.43 |
| polarC18 | 218.139 | 96.0316 | 6 | 0.43 |
| polarC18 | 218.139 | 208.3264 | 6 | 0.43 |
| polarC18 | 218.139 | 124.7159 | 6 | 0.43 |
| polarC18 | 218.139 | 76.1644 | 6 | 0.43 |
| polarC18 | 218.139 | 85.5359 | 6 | 0.43 |
| polarC18 | 218.139 | 48.9216 | 6 | 0.43 |
| polarC18 | 218.139 | 56.2893 | 6 | 0.43 |
| polarC18 | 218.139 | 65.005 | 6 | 0.43 |
| polarC18 | 218.139 | 69.3819 | 6 | 0.43 |
| polarC18 | 218.139 | 84.9971 | 6 | 0.43 |
| polarC18 | 218.139 | 150.5167 | 6 | 0.43 |
| polarC18 | 218.139 | 55.1004 | 6 | 0.43 |
| polarC18 | 218.139 | 124.6638 | 6 | 0.43 |
| polarC18 | 218.139 | 79.3116 | 6 | 0.43 |
| polarC18 | 218.139 | 73.6794 | 6 | 0.43 |
| polarC18 | 218.139 | 170.8539 | 6 | 0.43 |
| polarC18 | 218.139 | 50.5289 | 6 | 0.43 |
| polarC18 | 218.139 | 71.5387 | 6 | 0.43 |

| | | | | |
|---|---|---|---|---|
| polarC18 | 218.139 | 90.2235 | 6 | 0.43 |
| polarC18 | 218.139 | 79.8779 | 6 | 0.43 |
| polarC18 | 218.139 | 84.6239 | 6 | 0.43 |
| polarC18 | 218.139 | 57.9107 | 6 | 0.43 |
| polarC18 | 218.139 | 75.8833 | 6 | 0.43 |
| polarC18 | 218.139 | 169.8155 | 6 | 0.43 |
| polarC18 | 218.139 | 69.9887 | 6 | 0.43 |
| polarC18 | 218.139 | 115.9143 | 6 | 0.43 |
| polarC18 | 218.139 | 161.8082 | 6 | 0.43 |
| polarC18 | 218.139 | 63.4202 | 6 | 0.43 |
| polarC18 | 218.139 | 50.7308 | 6 | 0.43 |
| polarC18 | 218.139 | 98.471 | 6 | 0.43 |
| polarC18 | 218.139 | 87.6322 | 6 | 0.43 |
| polarC18 | 218.139 | 109.5753 | 6 | 0.43 |
| polarC18 | 218.139 | 141.647 | 6 | 0.43 |
| polarC18 | 218.139 | 53.5919 | 6 | 0.43 |
| polarC18 | 218.139 | 79.6971 | 6 | 0.43 |
| polarC18 | 218.139 | 201.4882 | 6 | 0.43 |
| polarC18 | 218.139 | 84.939 | 6 | 0.43 |
| polarC18 | 218.139 | 197.7979 | 6 | 0.43 |
| polarC18 | 218.139 | 84.348 | 6 | 0.43 |
| polarC18 | 218.139 | 109.6292 | 6 | 0.43 |
| polarC18 | 218.139 | 115.8695 | 6 | 0.43 |
| polarC18 | 218.139 | 79.1549 | 6 | 0.43 |
| polarC18 | 218.139 | 87.0339 | 6 | 0.43 |
| polarC18 | 218.139 | 50.6144 | 6 | 0.43 |
| polarC18 | 218.139 | 53.3884 | 6 | 0.43 |
| polarC18 | 218.139 | 61.2327 | 6 | 0.43 |
| polarC18 | 218.139 | 52.2644 | 6 | 0.43 |
| polarC18 | 218.139 | 59.0016 | 6 | 0.43 |
| polarC18 | 218.139 | 56.2463 | 6 | 0.43 |
| polarC18 | 218.139 | 112.4073 | 6 | 0.43 |
| polarC18 | 218.139 | 112.3891 | 6 | 0.43 |
| polarC18 | 218.139 | 104.4825 | 6 | 0.43 |
| polarC18 | 218.139 | 68.8351 | 6 | 0.43 |
| polarC18 | 218.139 | 58.3039 | 6 | 0.43 |
| polarC18 | 218.139 | 193.3825 | 6 | 0.43 |
| polarC18 | 218.139 | 116.8969 | 6 | 0.43 |
| polarC18 | 218.139 | 67.22 | 6 | 0.43 |
| polarC18 | 218.139 | 84.2755 | 6 | 0.43 |
| polarC18 | 218.139 | 121.8467 | 6 | 0.43 |
| polarC18 | 218.139 | 80.2613 | 6 | 0.43 |

| | | | | |
|---|---|---|---|---|
| polarC18 | 218.139 | 108.8376 | 6 | 0.43 |
| polarC18 | 218.139 | 54.8011 | 6 | 0.43 |
| polarC18 | 218.139 | 201.5552 | 6 | 0.43 |
| polarC18 | 218.139 | 57.4877 | 6 | 0.43 |
| polarC18 | 218.139 | 99.9776 | 6 | 0.43 |
| polarC18 | 218.139 | 59.818 | 6 | 0.43 |
| polarC18 | 218.139 | 97.2482 | 6 | 0.43 |
| polarC18 | 218.139 | 53.2787 | 6 | 0.43 |
| polarC18 | 218.139 | 66.0583 | 6 | 0.43 |
| polarC18 | 218.139 | 106.9961 | 6 | 0.43 |
| polarC18 | 218.139 | 73.4184 | 6 | 0.43 |
| polarC18 | 218.139 | 49.7174 | 6 | 0.43 |
| polarC18 | 218.139 | 63.515 | 6 | 0.43 |
| polarC18 | 218.139 | 65.6298 | 6 | 0.43 |
| polarC18 | 218.139 | 50.0825 | 6 | 0.43 |
| polarC18 | 218.139 | 129.819 | 6 | 0.43 |
| polarC18 | 218.139 | 141.2239 | 6 | 0.42 |
| polarC18 | 218.139 | 183.7578 | 6 | 0.42 |
| polarC18 | 218.139 | 61.995 | 6 | 0.42 |
| polarC18 | 218.139 | 62.8277 | 6 | 0.42 |
| polarC18 | 218.139 | 128.4122 | 6 | 0.42 |
| polarC18 | 218.139 | 90.8803 | 6 | 0.42 |
| polarC18 | 218.139 | 134.2107 | 6 | 0.42 |
| polarC18 | 218.139 | 73.6041 | 6 | 0.42 |
| polarC18 | 218.139 | 70.0635 | 6 | 0.42 |
| polarC18 | 218.139 | 183.1675 | 6 | 0.42 |
| polarC18 | 218.139 | 64.7476 | 6 | 0.42 |
| polarC18 | 218.139 | 55.2603 | 6 | 0.42 |
| polarC18 | 218.139 | 80.777 | 6 | 0.42 |
| polarC18 | 218.139 | 83.9698 | 6 | 0.42 |
| polarC18 | 218.139 | 230.2223 | 6 | 0.42 |
| polarC18 | 218.139 | 73.6202 | 6 | 0.42 |
| polarC18 | 218.139 | 55.9985 | 6 | 0.42 |
| polarC18 | 218.139 | 84.7362 | 6 | 0.42 |
| polarC18 | 218.139 | 75.1698 | 6 | 0.42 |
| polarC18 | 218.139 | 239.5937 | 6 | 0.42 |
| polarC18 | 218.139 | 182.149 | 6 | 0.42 |
| polarC18 | 218.139 | 63.0236 | 6 | 0.42 |
| polarC18 | 218.139 | 116.0204 | 6 | 0.42 |
| polarC18 | 218.139 | 66.4196 | 6 | 0.42 |
| polarC18 | 218.139 | 153.6984 | 6 | 0.42 |
| polarC18 | 218.139 | 105.4569 | 6 | 0.42 |

| | | | | |
|---|---|---|---|---|
| polarC18 | 218.139 | 219.9841 | 6 | 0.42 |
| polarC18 | 218.139 | 95.1747 | 6 | 0.42 |
| polarC18 | 218.139 | 91.2989 | 6 | 0.42 |
| polarC18 | 218.139 | 62.7798 | 6 | 0.42 |
| polarC18 | 218.139 | 57.8248 | 6 | 0.42 |
| polarC18 | 218.139 | 54.7165 | 6 | 0.42 |
| polarC18 | 218.139 | 157.5227 | 6 | 0.42 |
| polarC18 | 218.139 | 113.0645 | 6 | 0.42 |
| polarC18 | 218.139 | 92.1021 | 6 | 0.42 |
| polarC18 | 218.139 | 50.2322 | 6 | 0.42 |
| polarC18 | 218.139 | 58.3917 | 6 | 0.42 |
| polarC18 | 218.139 | 144.5342 | 6 | 0.42 |
| polarC18 | 218.139 | 56.9944 | 6 | 0.42 |
| polarC18 | 218.139 | 83.7902 | 6 | 0.42 |
| polarC18 | 218.139 | 83.8591 | 6 | 0.42 |
| polarC18 | 218.139 | 201.687 | 6 | 0.42 |
| polarC18 | 218.139 | 54.1515 | 6 | 0.42 |
| polarC18 | 218.139 | 136.7069 | 6 | 0.42 |
| polarC18 | 218.139 | 114.623 | 6 | 0.42 |
| polarC18 | 218.139 | 97.8687 | 6 | 0.42 |
| polarC18 | 218.139 | 54.7685 | 6 | 0.42 |
| polarC18 | 218.139 | 64.7707 | 6 | 0.42 |
| polarC18 | 218.139 | 75.4411 | 6 | 0.42 |
| polarC18 | 218.139 | 173.9404 | 6 | 0.42 |
| polarC18 | 218.139 | 119.0965 | 6 | 0.42 |
| polarC18 | 218.139 | 116.2538 | 6 | 0.42 |
| polarC18 | 218.139 | 205.4949 | 6 | 0.42 |
| polarC18 | 218.139 | 62.3381 | 6 | 0.42 |
| polarC18 | 218.139 | 57.4618 | 6 | 0.42 |
| polarC18 | 218.139 | 141.7352 | 6 | 0.42 |
| polarC18 | 218.139 | 85.9431 | 6 | 0.42 |
| polarC18 | 218.139 | 79.9619 | 6 | 0.42 |
| polarC18 | 218.139 | 51.8294 | 6 | 0.42 |
| polarC18 | 218.139 | 108.2027 | 6 | 0.42 |
| polarC18 | 218.139 | 214.4501 | 6 | 0.42 |
| polarC18 | 218.139 | 77.2534 | 6 | 0.42 |
| polarC18 | 218.139 | 238.9098 | 6 | 0.42 |
| polarC18 | 218.139 | 216.0535 | 6 | 0.42 |
| polarC18 | 218.139 | 49.269 | 6 | 0.42 |
| polarC18 | 218.139 | 53.3084 | 6 | 0.42 |
| polarC18 | 218.139 | 72.4641 | 6 | 0.42 |
| polarC18 | 218.139 | 139.5936 | 6 | 0.42 |

| | | | | |
|---|---|---|---|---|
| polarC18 | 218.139 | 176.5971 | 6 | 0.42 |
| polarC18 | 218.139 | 58.0967 | 5.9 | 0.42 |
| polarC18 | 218.139 | 189.1003 | 5.9 | 0.42 |
| polarC18 | 218.139 | 51.8399 | 5.9 | 0.42 |
| polarC18 | 218.139 | 52.2321 | 5.9 | 0.42 |
| polarC18 | 218.139 | 169.3916 | 5.9 | 0.42 |
| polarC18 | 218.139 | 139.6727 | 5.9 | 0.42 |
| polarC18 | 218.139 | 70.9906 | 5.9 | 0.42 |
| polarC18 | 218.139 | 154.7192 | 5.9 | 0.42 |
| polarC18 | 218.139 | 63.1873 | 5.9 | 0.42 |
| polarC18 | 218.139 | 57.6625 | 5.9 | 0.42 |
| polarC18 | 218.139 | 62.3091 | 5.9 | 0.42 |
| polarC18 | 218.139 | 88.1093 | 5.9 | 0.42 |
| polarC18 | 218.139 | 69.7603 | 5.9 | 0.42 |
| polarC18 | 218.139 | 68.1599 | 5.9 | 0.42 |
| polarC18 | 218.139 | 67.8686 | 5.9 | 0.42 |
| polarC18 | 218.139 | 179.5263 | 5.9 | 0.42 |
| polarC18 | 218.139 | 76.1068 | 5.9 | 0.42 |
| polarC18 | 218.139 | 185.4482 | 5.9 | 0.42 |
| polarC18 | 218.139 | 241.8253 | 5.9 | 0.42 |
| polarC18 | 218.139 | 63.4761 | 5.9 | 0.42 |
| polarC18 | 218.139 | 214.0322 | 5.9 | 0.42 |
| polarC18 | 218.139 | 161.0234 | 5.9 | 0.42 |
| polarC18 | 218.139 | 51.2982 | 5.9 | 0.42 |
| polarC18 | 218.139 | 207.6395 | 5.9 | 0.42 |
| polarC18 | 218.139 | 89.6642 | 5.9 | 0.42 |
| polarC18 | 218.139 | 69.4675 | 5.9 | 0.42 |
| polarC18 | 218.139 | 54.7048 | 5.9 | 0.42 |
| polarC18 | 218.139 | 68.9581 | 5.9 | 0.42 |
| polarC18 | 218.139 | 67.5306 | 5.9 | 0.42 |
| polarC18 | 218.139 | 101.5736 | 5.9 | 0.42 |
| polarC18 | 218.139 | 60.1839 | 5.9 | 0.42 |
| polarC18 | 218.139 | 115.9871 | 5.9 | 0.42 |
| polarC18 | 218.139 | 48.8253 | 5.9 | 0.42 |
| polarC18 | 218.139 | 82.8414 | 5.9 | 0.42 |
| polarC18 | 218.139 | 58.5059 | 5.9 | 0.42 |
| polarC18 | 218.139 | 69.391 | 5.9 | 0.42 |
| polarC18 | 218.139 | 91.5383 | 5.9 | 0.42 |
| polarC18 | 218.139 | 191.5504 | 5.9 | 0.42 |
| polarC18 | 218.139 | 133.4577 | 5.9 | 0.42 |
| polarC18 | 218.139 | 113.1812 | 5.9 | 0.42 |
| polarC18 | 218.139 | 85.1581 | 5.9 | 0.42 |

| | | | | |
|---|---|---|---|---|
| polarC18 | 218.139 | 66.6032 | 5.9 | 0.42 |
| polarC18 | 218.139 | 49.8499 | 5.9 | 0.42 |
| polarC18 | 218.139 | 158.2892 | 5.9 | 0.42 |
| polarC18 | 218.139 | 107.6606 | 5.9 | 0.42 |
| polarC18 | 218.139 | 103.9364 | 5.9 | 0.42 |
| polarC18 | 218.139 | 82.2073 | 5.9 | 0.42 |
| polarC18 | 218.139 | 49.3262 | 5.9 | 0.42 |
| polarC18 | 218.139 | 101.2537 | 5.9 | 0.42 |
| polarC18 | 218.139 | 64.6667 | 5.9 | 0.42 |
| polarC18 | 218.139 | 166.8149 | 5.9 | 0.42 |
| polarC18 | 218.139 | 115.3464 | 5.9 | 0.42 |
| polarC18 | 218.139 | 205.1776 | 5.9 | 0.42 |
| polarC18 | 218.139 | 61.1274 | 5.9 | 0.42 |
| polarC18 | 218.139 | 80.1831 | 5.9 | 0.42 |
| polarC18 | 218.139 | 78.5218 | 5.9 | 0.42 |
| polarC18 | 218.139 | 60.3348 | 5.9 | 0.42 |
| polarC18 | 218.139 | 71.329 | 5.9 | 0.42 |
| polarC18 | 218.139 | 50.9988 | 5.9 | 0.42 |
| polarC18 | 218.139 | 171.5665 | 5.9 | 0.42 |
| polarC18 | 218.139 | 89.6853 | 5.9 | 0.42 |
| polarC18 | 218.139 | 96.7463 | 5.9 | 0.42 |
| polarC18 | 218.139 | 139.7199 | 5.9 | 0.42 |
| polarC18 | 218.139 | 194.781 | 5.9 | 0.42 |
| polarC18 | 218.139 | 80.1199 | 5.9 | 0.42 |
| polarC18 | 218.139 | 55.8381 | 5.9 | 0.42 |
| polarC18 | 218.139 | 231.3708 | 5.9 | 0.42 |
| polarC18 | 218.139 | 52.3556 | 5.9 | 0.42 |
| polarC18 | 218.139 | 100.5847 | 5.9 | 0.42 |
| polarC18 | 218.139 | 228.1067 | 5.9 | 0.42 |
| polarC18 | 218.139 | 49.0183 | 5.9 | 0.42 |
| polarC18 | 218.139 | 61.0751 | 5.9 | 0.42 |
| polarC18 | 218.139 | 168.1932 | 5.9 | 0.42 |
| polarC18 | 218.139 | 168.9301 | 5.9 | 0.42 |
| polarC18 | 218.139 | 66.3958 | 5.9 | 0.42 |
| polarC18 | 218.139 | 51.9913 | 5.9 | 0.42 |
| polarC18 | 218.139 | 73.6084 | 5.9 | 0.42 |
| polarC18 | 218.139 | 57.0538 | 5.9 | 0.42 |
| polarC18 | 218.139 | 70.9017 | 5.9 | 0.42 |
| polarC18 | 218.139 | 134.2884 | 5.9 | 0.42 |
| polarC18 | 218.139 | 170.3677 | 5.9 | 0.42 |
| polarC18 | 218.139 | 92.0576 | 5.9 | 0.42 |
| polarC18 | 218.139 | 184.5036 | 5.9 | 0.42 |

| | | | | |
|---|---|---|---|---|
| polarC18 | 218.139 | 78.117 | 5.9 | 0.42 |
| polarC18 | 218.139 | 97.2224 | 5.9 | 0.42 |
| polarC18 | 218.139 | 80.5246 | 5.9 | 0.42 |
| polarC18 | 218.139 | 92.2551 | 5.9 | 0.42 |
| polarC18 | 218.139 | 100.1054 | 5.9 | 0.42 |
| polarC18 | 218.139 | 74.2979 | 5.9 | 0.42 |
| polarC18 | 218.139 | 112.8588 | 5.9 | 0.42 |
| polarC18 | 218.139 | 165.7871 | 5.9 | 0.42 |
| polarC18 | 218.139 | 126.7171 | 5.9 | 0.42 |
| polarC18 | 218.139 | 48.9013 | 5.9 | 0.42 |
| polarC18 | 218.139 | 112.1368 | 5.9 | 0.42 |
| polarC18 | 218.139 | 58.5364 | 5.9 | 0.42 |
| polarC18 | 218.139 | 109.2525 | 5.9 | 0.42 |
| polarC18 | 218.139 | 88.5058 | 5.9 | 0.42 |
| polarC18 | 218.139 | 75.1934 | 5.9 | 0.42 |
| polarC18 | 218.139 | 93.0301 | 5.9 | 0.42 |
| polarC18 | 218.139 | 77.3306 | 5.9 | 0.42 |
| polarC18 | 218.139 | 94.3516 | 5.9 | 0.42 |
| polarC18 | 218.139 | 118.0124 | 5.9 | 0.42 |
| polarC18 | 218.139 | 111.7299 | 5.9 | 0.42 |
| polarC18 | 218.139 | 48.712 | 5.9 | 0.42 |
| polarC18 | 218.139 | 75.7599 | 5.9 | 0.42 |
| polarC18 | 218.139 | 105.8024 | 5.9 | 0.42 |
| polarC18 | 218.139 | 50.5822 | 5.9 | 0.42 |
| polarC18 | 218.139 | 62.633 | 5.9 | 0.42 |
| polarC18 | 218.139 | 68.3387 | 5.9 | 0.42 |
| polarC18 | 218.139 | 98.4802 | 5.9 | 0.42 |
| polarC18 | 218.139 | 59.6721 | 5.9 | 0.42 |
| polarC18 | 218.139 | 63.8308 | 5.9 | 0.42 |
| polarC18 | 218.139 | 49.0812 | 5.9 | 0.42 |
| polarC18 | 218.139 | 50.937 | 5.9 | 0.42 |
| polarC18 | 218.139 | 101.4267 | 5.9 | 0.42 |
| polarC18 | 218.139 | 55.8723 | 5.9 | 0.42 |
| polarC18 | 218.139 | 99.6155 | 5.9 | 0.42 |
| polarC18 | 218.139 | 197.6728 | 5.9 | 0.42 |
| polarC18 | 218.139 | 139.9144 | 5.9 | 0.42 |
| polarC18 | 218.139 | 63.1953 | 5.9 | 0.42 |
| polarC18 | 218.139 | 168.7198 | 5.9 | 0.42 |
| polarC18 | 218.139 | 58.206 | 5.9 | 0.42 |
| polarC18 | 218.139 | 54.0504 | 5.9 | 0.42 |
| polarC18 | 218.139 | 59.2472 | 5.9 | 0.42 |
| polarC18 | 218.139 | 108.7933 | 5.9 | 0.42 |

| | | | | |
|---|---|---|---|---|
| polarC18 | 218.139 | 92.2173 | 5.9 | 0.42 |
| polarC18 | 218.139 | 83.7841 | 5.9 | 0.42 |
| polarC18 | 218.139 | 49.9531 | 5.9 | 0.42 |
| polarC18 | 218.139 | 89.0447 | 5.9 | 0.42 |
| polarC18 | 218.139 | 53.9344 | 5.9 | 0.42 |
| polarC18 | 218.139 | 51.2396 | 5.9 | 0.42 |
| polarC18 | 218.139 | 52.2262 | 5.9 | 0.42 |
| polarC18 | 218.139 | 142.0614 | 5.9 | 0.42 |
| polarC18 | 218.139 | 69.6011 | 5.9 | 0.42 |
| polarC18 | 218.139 | 70.124 | 5.9 | 0.42 |
| polarC18 | 218.139 | 177.9092 | 5.9 | 0.42 |
| polarC18 | 218.139 | 96.1215 | 5.9 | 0.42 |
| polarC18 | 218.139 | 68.193 | 5.9 | 0.42 |
| polarC18 | 218.139 | 54.289 | 5.9 | 0.42 |
| polarC18 | 218.139 | 93.7407 | 5.9 | 0.42 |
| polarC18 | 218.139 | 61.5984 | 5.9 | 0.42 |
| polarC18 | 218.139 | 120.9681 | 5.9 | 0.42 |
| polarC18 | 218.139 | 147.2017 | 5.9 | 0.42 |
| polarC18 | 218.139 | 59.3026 | 5.9 | 0.42 |
| polarC18 | 218.139 | 94.2923 | 5.9 | 0.42 |
| polarC18 | 218.139 | 60.8052 | 5.9 | 0.42 |
| polarC18 | 218.139 | 226.706 | 5.9 | 0.42 |
| polarC18 | 218.139 | 56.1496 | 5.9 | 0.42 |
| polarC18 | 218.139 | 81.6479 | 5.8 | 0.42 |
| polarC18 | 218.139 | 136.0185 | 5.8 | 0.41 |
| polarC18 | 218.139 | 165.8331 | 5.8 | 0.41 |
| polarC18 | 218.139 | 101.9753 | 5.8 | 0.41 |
| polarC18 | 218.139 | 88.2095 | 5.8 | 0.41 |
| polarC18 | 218.139 | 49.6867 | 5.8 | 0.41 |
| polarC18 | 218.139 | 52.6551 | 5.8 | 0.41 |
| polarC18 | 218.139 | 74.8648 | 5.8 | 0.41 |
| polarC18 | 218.139 | 49.3827 | 5.8 | 0.41 |
| polarC18 | 218.139 | 108.332 | 5.8 | 0.41 |
| polarC18 | 218.139 | 117.2746 | 5.8 | 0.41 |
| polarC18 | 218.139 | 141.7042 | 5.8 | 0.41 |
| polarC18 | 218.139 | 67.2872 | 5.8 | 0.41 |
| polarC18 | 218.139 | 52.538 | 5.8 | 0.41 |
| polarC18 | 218.139 | 65.8367 | 5.8 | 0.41 |
| polarC18 | 218.139 | 126.529 | 5.8 | 0.41 |
| polarC18 | 218.139 | 100.899 | 5.8 | 0.41 |
| polarC18 | 218.139 | 92.7219 | 5.8 | 0.41 |
| polarC18 | 218.139 | 50.1759 | 5.8 | 0.41 |

| | | | | |
|---|---|---|---|---|
| polarC18 | 218.139 | 53.1406 | 5.8 | 0.41 |
| polarC18 | 218.139 | 102.1845 | 5.8 | 0.41 |
| polarC18 | 218.139 | 147.579 | 5.8 | 0.41 |
| polarC18 | 218.139 | 126.4949 | 5.8 | 0.41 |
| polarC18 | 218.139 | 139.5747 | 5.8 | 0.41 |
| polarC18 | 218.139 | 55.2828 | 5.8 | 0.41 |
| polarC18 | 218.139 | 197.2754 | 5.8 | 0.41 |
| polarC18 | 218.139 | 136.6524 | 5.8 | 0.41 |
| polarC18 | 218.139 | 56.5538 | 5.8 | 0.41 |
| polarC18 | 218.139 | 120.2874 | 5.8 | 0.41 |
| polarC18 | 218.139 | 54.287 | 5.8 | 0.41 |
| polarC18 | 218.139 | 202.3029 | 5.8 | 0.41 |
| polarC18 | 218.139 | 66.2275 | 5.8 | 0.41 |
| polarC18 | 218.139 | 81.5663 | 5.8 | 0.41 |
| polarC18 | 218.139 | 54.7419 | 5.8 | 0.41 |
| polarC18 | 218.139 | 127.1022 | 5.8 | 0.41 |
| polarC18 | 218.139 | 79.6434 | 5.8 | 0.41 |
| polarC18 | 218.139 | 107.5938 | 5.8 | 0.41 |
| polarC18 | 218.139 | 117.1815 | 5.8 | 0.41 |
| polarC18 | 218.139 | 65.7366 | 5.8 | 0.41 |
| polarC18 | 218.139 | 57.4365 | 5.8 | 0.41 |
| polarC18 | 218.139 | 92.1673 | 5.8 | 0.41 |
| polarC18 | 218.139 | 78.8975 | 5.8 | 0.41 |
| polarC18 | 218.139 | 52.9996 | 5.8 | 0.41 |
| polarC18 | 218.139 | 54.3719 | 5.8 | 0.41 |
| polarC18 | 218.139 | 60.7349 | 5.8 | 0.41 |
| polarC18 | 218.139 | 98.091 | 5.8 | 0.41 |
| polarC18 | 218.139 | 189.2399 | 5.8 | 0.41 |
| polarC18 | 218.139 | 51.8907 | 5.8 | 0.41 |
| polarC18 | 218.139 | 218.539 | 5.8 | 0.41 |
| polarC18 | 218.139 | 65.2665 | 5.8 | 0.41 |
| polarC18 | 218.139 | 108.0216 | 5.8 | 0.41 |
| polarC18 | 218.139 | 78.0591 | 5.8 | 0.41 |
| polarC18 | 218.139 | 72.3763 | 5.8 | 0.41 |
| polarC18 | 218.139 | 55.8955 | 5.8 | 0.41 |
| polarC18 | 218.139 | 82.2448 | 5.8 | 0.41 |
| polarC18 | 218.139 | 172.1456 | 5.8 | 0.41 |
| polarC18 | 218.139 | 150.3952 | 5.8 | 0.41 |
| polarC18 | 218.139 | 84.2932 | 5.8 | 0.41 |
| polarC18 | 218.139 | 61.2828 | 5.8 | 0.41 |
| polarC18 | 218.139 | 72.8662 | 5.8 | 0.41 |
| polarC18 | 218.139 | 65.8141 | 5.8 | 0.41 |

| | | | | |
|---|---|---|---|---|
| polarC18 | 218.139 | 52.5864 | 5.8 | 0.41 |
| polarC18 | 218.139 | 49.2457 | 5.8 | 0.41 |
| polarC18 | 218.139 | 49.4463 | 5.8 | 0.41 |
| polarC18 | 218.139 | 86.2978 | 5.8 | 0.41 |
| polarC18 | 218.139 | 112.2705 | 5.8 | 0.41 |
| polarC18 | 218.139 | 175.0473 | 5.8 | 0.41 |
| polarC18 | 218.139 | 51.5705 | 5.8 | 0.41 |
| polarC18 | 218.139 | 100.9618 | 5.8 | 0.41 |
| polarC18 | 218.139 | 124.6314 | 5.8 | 0.41 |
| polarC18 | 218.139 | 71.2001 | 5.8 | 0.41 |
| polarC18 | 218.139 | 52.0299 | 5.8 | 0.41 |
| polarC18 | 218.139 | 66.044 | 5.8 | 0.41 |
| polarC18 | 218.139 | 129.7338 | 5.8 | 0.41 |
| polarC18 | 218.139 | 50.7493 | 5.8 | 0.41 |
| polarC18 | 218.139 | 214.9558 | 5.8 | 0.41 |
| polarC18 | 218.139 | 55.8623 | 5.8 | 0.41 |
| polarC18 | 218.139 | 150.8365 | 5.8 | 0.41 |
| polarC18 | 218.139 | 178.3085 | 5.8 | 0.41 |
| polarC18 | 218.139 | 62.4013 | 5.8 | 0.41 |
| polarC18 | 218.139 | 137.134 | 5.8 | 0.41 |
| polarC18 | 218.139 | 145.1212 | 5.8 | 0.41 |
| polarC18 | 218.139 | 63.2773 | 5.8 | 0.41 |
| polarC18 | 218.139 | 49.4321 | 5.8 | 0.41 |
| polarC18 | 218.139 | 112.5819 | 5.8 | 0.41 |
| polarC18 | 218.139 | 152.5384 | 5.8 | 0.41 |
| polarC18 | 218.139 | 143.447 | 5.8 | 0.41 |
| polarC18 | 218.139 | 49.8296 | 5.8 | 0.41 |
| polarC18 | 218.139 | 74.6169 | 5.8 | 0.41 |
| polarC18 | 218.139 | 128.4261 | 5.8 | 0.41 |
| polarC18 | 218.139 | 157.5553 | 5.8 | 0.41 |
| polarC18 | 218.139 | 133.6712 | 5.8 | 0.41 |
| polarC18 | 218.139 | 52.4886 | 5.8 | 0.41 |
| polarC18 | 218.139 | 68.048 | 5.8 | 0.41 |
| polarC18 | 218.139 | 50.3914 | 5.8 | 0.41 |
| polarC18 | 218.139 | 101.2146 | 5.8 | 0.41 |
| polarC18 | 218.139 | 59.2046 | 5.8 | 0.41 |
| polarC18 | 218.139 | 53.6588 | 5.8 | 0.41 |
| polarC18 | 218.139 | 174.4755 | 5.8 | 0.41 |
| polarC18 | 218.139 | 101.5885 | 5.8 | 0.41 |
| polarC18 | 218.139 | 135.0226 | 5.8 | 0.41 |
| polarC18 | 218.139 | 75.6996 | 5.8 | 0.41 |
| polarC18 | 218.139 | 56.3492 | 5.8 | 0.41 |

| | | | | |
|---|---|---|---|---|
| polarC18 | 218.139 | 59.3414 | 5.8 | 0.41 |
| polarC18 | 218.139 | 65.9958 | 5.8 | 0.41 |
| polarC18 | 218.139 | 110.7424 | 5.8 | 0.41 |
| polarC18 | 218.139 | 73.6497 | 5.8 | 0.41 |
| polarC18 | 218.139 | 134.5996 | 5.8 | 0.41 |
| polarC18 | 218.139 | 51.8327 | 5.8 | 0.41 |
| polarC18 | 218.139 | 184.8613 | 5.8 | 0.41 |
| polarC18 | 218.139 | 159.0504 | 5.8 | 0.41 |
| polarC18 | 218.139 | 143.7552 | 5.8 | 0.41 |
| polarC18 | 218.139 | 236.5264 | 5.8 | 0.41 |
| polarC18 | 218.139 | 96.612 | 5.8 | 0.41 |
| polarC18 | 218.139 | 64.5766 | 5.8 | 0.41 |
| polarC18 | 218.139 | 77.1278 | 5.8 | 0.41 |
| polarC18 | 218.139 | 86.3254 | 5.8 | 0.41 |
| polarC18 | 218.139 | 112.2591 | 5.8 | 0.41 |
| polarC18 | 218.139 | 59.1086 | 5.8 | 0.41 |
| polarC18 | 218.139 | 58.5031 | 5.8 | 0.41 |
| polarC18 | 218.139 | 146.5536 | 5.8 | 0.41 |
| polarC18 | 218.139 | 96.945 | 5.8 | 0.41 |
| polarC18 | 218.139 | 67.6205 | 5.8 | 0.41 |
| polarC18 | 218.139 | 142.2882 | 5.8 | 0.41 |
| polarC18 | 218.139 | 66.062 | 5.8 | 0.41 |
| polarC18 | 218.139 | 59.1999 | 5.8 | 0.41 |
| polarC18 | 218.139 | 50.1003 | 5.8 | 0.41 |
| polarC18 | 218.139 | 64.0991 | 5.8 | 0.41 |
| polarC18 | 218.139 | 189.2675 | 5.8 | 0.41 |
| polarC18 | 218.139 | 66.5884 | 5.8 | 0.41 |
| polarC18 | 218.139 | 190.3827 | 5.7 | 0.41 |
| polarC18 | 218.139 | 155.8515 | 5.7 | 0.41 |
| polarC18 | 218.139 | 48.9858 | 5.7 | 0.41 |
| polarC18 | 218.139 | 58.0694 | 5.7 | 0.41 |
| polarC18 | 218.139 | 114.9575 | 5.7 | 0.41 |
| polarC18 | 218.139 | 228.7744 | 5.7 | 0.41 |
| polarC18 | 218.139 | 117.6924 | 5.7 | 0.41 |
| polarC18 | 218.139 | 99.5795 | 5.7 | 0.41 |
| polarC18 | 218.139 | 53.7041 | 5.7 | 0.41 |
| polarC18 | 218.139 | 130.8681 | 5.7 | 0.41 |
| polarC18 | 218.139 | 65.2695 | 5.7 | 0.41 |
| polarC18 | 218.139 | 162.841 | 5.7 | 0.41 |
| polarC18 | 218.139 | 120.138 | 5.7 | 0.41 |
| polarC18 | 218.139 | 68.9549 | 5.7 | 0.41 |
| polarC18 | 218.139 | 52.8706 | 5.7 | 0.41 |

| | | | | |
|---|---|---|---|---|
| polarC18 | 218.139 | 50.6325 | 5.7 | 0.41 |
| polarC18 | 218.139 | 87.5577 | 5.7 | 0.41 |
| polarC18 | 218.139 | 89.2379 | 5.7 | 0.41 |
| polarC18 | 218.139 | 81.288 | 5.7 | 0.41 |
| polarC18 | 218.139 | 209.7451 | 5.7 | 0.41 |
| polarC18 | 218.139 | 82.6039 | 5.7 | 0.41 |
| polarC18 | 218.139 | 67.6295 | 5.7 | 0.41 |
| polarC18 | 218.139 | 57.9999 | 5.7 | 0.41 |
| polarC18 | 218.139 | 149.6909 | 5.7 | 0.41 |
| polarC18 | 218.139 | 149.0154 | 5.7 | 0.41 |
| polarC18 | 218.139 | 55.5927 | 5.7 | 0.41 |
| polarC18 | 218.139 | 74.9768 | 5.7 | 0.41 |
| polarC18 | 218.139 | 130.1928 | 5.7 | 0.41 |
| polarC18 | 218.139 | 232.2594 | 5.7 | 0.41 |
| polarC18 | 218.139 | 66.7357 | 5.7 | 0.41 |
| polarC18 | 218.139 | 51.4188 | 5.7 | 0.41 |
| polarC18 | 218.139 | 78.7568 | 5.7 | 0.41 |
| polarC18 | 218.139 | 51.705 | 5.7 | 0.41 |
| polarC18 | 218.139 | 92.238 | 5.7 | 0.41 |
| polarC18 | 218.139 | 99.8737 | 5.7 | 0.41 |
| polarC18 | 218.139 | 210.5464 | 5.7 | 0.41 |
| polarC18 | 218.139 | 62.1622 | 5.7 | 0.41 |
| polarC18 | 218.139 | 79.1008 | 5.7 | 0.41 |
| polarC18 | 218.139 | 48.9577 | 5.7 | 0.41 |
| polarC18 | 218.139 | 126.9817 | 5.7 | 0.41 |
| polarC18 | 218.139 | 86.7356 | 5.7 | 0.41 |
| polarC18 | 218.139 | 49.6325 | 5.7 | 0.41 |
| polarC18 | 218.139 | 112.9951 | 5.7 | 0.41 |
| polarC18 | 218.139 | 120.5724 | 5.7 | 0.41 |
| polarC18 | 218.139 | 226.7596 | 5.7 | 0.41 |
| polarC18 | 218.139 | 237.8939 | 5.7 | 0.41 |
| polarC18 | 218.139 | 87.5165 | 5.7 | 0.41 |
| polarC18 | 218.139 | 49.5085 | 5.7 | 0.41 |
| polarC18 | 218.139 | 86.5911 | 5.7 | 0.41 |
| polarC18 | 218.139 | 165.1884 | 5.7 | 0.41 |
| polarC18 | 218.139 | 169.0329 | 5.7 | 0.41 |
| polarC18 | 218.139 | 62.0641 | 5.7 | 0.41 |
| polarC18 | 218.139 | 163.6931 | 5.7 | 0.41 |
| polarC18 | 218.139 | 102.1485 | 5.7 | 0.41 |
| polarC18 | 218.139 | 58.8194 | 5.7 | 0.41 |
| polarC18 | 218.139 | 56.4579 | 5.7 | 0.4 |
| polarC18 | 218.139 | 89.2425 | 5.7 | 0.4 |

| | | | | |
|---|---|---|---|---|
| polarC18 | 218.139 | 94.5857 | 5.7 | 0.4 |
| polarC18 | 218.139 | 54.3013 | 5.7 | 0.4 |
| polarC18 | 218.139 | 131.9327 | 5.7 | 0.4 |
| polarC18 | 218.139 | 62.5139 | 5.7 | 0.4 |
| polarC18 | 218.139 | 67.8958 | 5.7 | 0.4 |
| polarC18 | 218.139 | 76.4858 | 5.7 | 0.4 |
| polarC18 | 218.139 | 112.0549 | 5.7 | 0.4 |
| polarC18 | 218.139 | 103.5973 | 5.7 | 0.4 |
| polarC18 | 218.139 | 67.3871 | 5.7 | 0.4 |
| polarC18 | 218.139 | 105.7004 | 5.7 | 0.4 |
| polarC18 | 218.139 | 101.5496 | 5.7 | 0.4 |
| polarC18 | 218.139 | 49.8384 | 5.7 | 0.4 |
| polarC18 | 218.139 | 129.1705 | 5.7 | 0.4 |
| polarC18 | 218.139 | 106.6145 | 5.7 | 0.4 |
| polarC18 | 218.139 | 91.6274 | 5.7 | 0.4 |
| polarC18 | 218.139 | 97.2379 | 5.7 | 0.4 |
| polarC18 | 218.139 | 61.5715 | 5.7 | 0.4 |
| polarC18 | 218.139 | 160.2 | 5.7 | 0.4 |
| polarC18 | 218.139 | 53.8931 | 5.7 | 0.4 |
| polarC18 | 218.139 | 230.7645 | 5.7 | 0.4 |
| polarC18 | 218.139 | 73.271 | 5.7 | 0.4 |
| polarC18 | 218.139 | 80.9995 | 5.7 | 0.4 |
| polarC18 | 218.139 | 208.3015 | 5.7 | 0.4 |
| polarC18 | 218.139 | 75.4584 | 5.7 | 0.4 |
| polarC18 | 218.139 | 70.5352 | 5.7 | 0.4 |
| polarC18 | 218.139 | 58.8034 | 5.7 | 0.4 |
| polarC18 | 218.139 | 53.3155 | 5.7 | 0.4 |
| polarC18 | 218.139 | 63.4166 | 5.7 | 0.4 |
| polarC18 | 218.139 | 77.351 | 5.7 | 0.4 |
| polarC18 | 218.139 | 113.1686 | 5.7 | 0.4 |
| polarC18 | 218.139 | 106.2387 | 5.7 | 0.4 |
| polarC18 | 218.139 | 66.5278 | 5.7 | 0.4 |
| polarC18 | 218.139 | 53.5063 | 5.7 | 0.4 |
| polarC18 | 218.139 | 188.4055 | 5.7 | 0.4 |
| polarC18 | 218.139 | 78.6868 | 5.7 | 0.4 |
| polarC18 | 218.139 | 177.3326 | 5.7 | 0.4 |
| polarC18 | 218.139 | 114.1561 | 5.7 | 0.4 |
| polarC18 | 218.139 | 49.3006 | 5.7 | 0.4 |
| polarC18 | 218.139 | 53.2027 | 5.7 | 0.4 |
| polarC18 | 218.139 | 156.9321 | 5.7 | 0.4 |
| polarC18 | 218.139 | 85.1385 | 5.7 | 0.4 |
| polarC18 | 218.139 | 104.4215 | 5.7 | 0.4 |

| | | | | |
|---|---|---|---|---|
| polarC18 | 218.139 | 113.981 | 5.7 | 0.4 |
| polarC18 | 218.139 | 54.4576 | 5.7 | 0.4 |
| polarC18 | 218.139 | 74.8923 | 5.7 | 0.4 |
| polarC18 | 218.139 | 75.7916 | 5.7 | 0.4 |
| polarC18 | 218.139 | 147.9461 | 5.7 | 0.4 |
| polarC18 | 218.139 | 86.7264 | 5.7 | 0.4 |
| polarC18 | 218.139 | 100.3399 | 5.7 | 0.4 |
| polarC18 | 218.139 | 87.0827 | 5.7 | 0.4 |
| polarC18 | 218.139 | 78.1508 | 5.7 | 0.4 |
| polarC18 | 218.139 | 99.3518 | 5.7 | 0.4 |
| polarC18 | 218.139 | 65.9894 | 5.7 | 0.4 |
| polarC18 | 218.139 | 68.4351 | 5.7 | 0.4 |
| polarC18 | 218.139 | 98.7369 | 5.7 | 0.4 |
| polarC18 | 218.139 | 182.3377 | 5.7 | 0.4 |
| polarC18 | 218.139 | 126.6004 | 5.7 | 0.4 |
| polarC18 | 218.139 | 51.028 | 5.7 | 0.4 |
| polarC18 | 218.139 | 48.6631 | 5.7 | 0.4 |
| polarC18 | 218.139 | 67.6517 | 5.7 | 0.4 |
| polarC18 | 218.139 | 124.9157 | 5.7 | 0.4 |
| polarC18 | 218.139 | 117.9616 | 5.7 | 0.4 |
| polarC18 | 218.139 | 73.0889 | 5.7 | 0.4 |
| polarC18 | 218.139 | 66.0472 | 5.7 | 0.4 |
| polarC18 | 218.139 | 127.2015 | 5.7 | 0.4 |
| polarC18 | 218.139 | 76.1721 | 5.7 | 0.4 |
| polarC18 | 218.139 | 74.127 | 5.7 | 0.4 |
| polarC18 | 218.139 | 50.0307 | 5.6 | 0.4 |
| polarC18 | 218.139 | 234.3603 | 5.6 | 0.4 |
| polarC18 | 218.139 | 87.2779 | 5.6 | 0.4 |
| polarC18 | 218.139 | 75.9136 | 5.6 | 0.4 |
| polarC18 | 218.139 | 61.4736 | 5.6 | 0.4 |
| polarC18 | 218.139 | 110.8718 | 5.6 | 0.4 |
| polarC18 | 218.139 | 51.0956 | 5.6 | 0.4 |
| polarC18 | 218.139 | 111.8402 | 5.6 | 0.4 |
| polarC18 | 218.139 | 73.4067 | 5.6 | 0.4 |
| polarC18 | 218.139 | 52.9975 | 5.6 | 0.4 |
| polarC18 | 218.139 | 186.8847 | 5.6 | 0.4 |
| polarC18 | 218.139 | 172.3077 | 5.6 | 0.4 |
| polarC18 | 218.139 | 98.0501 | 5.6 | 0.4 |
| polarC18 | 218.139 | 74.6071 | 5.6 | 0.4 |
| polarC18 | 218.139 | 98.906 | 5.6 | 0.4 |
| polarC18 | 218.139 | 110.2783 | 5.6 | 0.4 |
| polarC18 | 218.139 | 58.0439 | 5.6 | 0.4 |

| | | | | |
|---|---|---|---|---|
| polarC18 | 218.139 | 144.0177 | 5.6 | 0.4 |
| polarC18 | 218.139 | 66.833 | 5.6 | 0.4 |
| polarC18 | 218.139 | 63.9453 | 5.6 | 0.4 |
| polarC18 | 218.139 | 50.0074 | 5.6 | 0.4 |
| polarC18 | 218.139 | 97.2637 | 5.6 | 0.4 |
| polarC18 | 218.139 | 49.1288 | 5.6 | 0.4 |
| polarC18 | 218.139 | 108.8872 | 5.6 | 0.4 |
| polarC18 | 218.139 | 197.4185 | 5.6 | 0.4 |
| polarC18 | 218.139 | 67.849 | 5.6 | 0.4 |
| polarC18 | 218.139 | 98.3837 | 5.6 | 0.4 |
| polarC18 | 218.139 | 64.9424 | 5.6 | 0.4 |
| polarC18 | 218.139 | 49.0492 | 5.6 | 0.4 |
| polarC18 | 218.139 | 51.9117 | 5.6 | 0.4 |
| polarC18 | 218.139 | 206.0275 | 5.6 | 0.4 |
| polarC18 | 218.139 | 73.7244 | 5.6 | 0.4 |
| polarC18 | 218.139 | 49.0536 | 5.6 | 0.4 |
| polarC18 | 218.139 | 200.2961 | 5.6 | 0.4 |
| polarC18 | 218.139 | 175.5931 | 5.6 | 0.4 |
| polarC18 | 218.139 | 59.2374 | 5.6 | 0.4 |
| polarC18 | 218.139 | 99.3013 | 5.6 | 0.4 |
| polarC18 | 218.139 | 110.9819 | 5.6 | 0.4 |
| polarC18 | 218.139 | 133.5433 | 5.6 | 0.4 |
| polarC18 | 218.139 | 58.2017 | 5.6 | 0.4 |
| polarC18 | 218.139 | 51.7545 | 5.6 | 0.4 |
| polarC18 | 218.139 | 59.7492 | 5.6 | 0.4 |
| polarC18 | 218.139 | 60.4997 | 5.6 | 0.4 |
| polarC18 | 218.139 | 53.4216 | 5.6 | 0.4 |
| polarC18 | 218.139 | 165.0511 | 5.6 | 0.4 |
| polarC18 | 218.139 | 60.0341 | 5.6 | 0.4 |
| polarC18 | 218.139 | 80.3371 | 5.6 | 0.4 |
| polarC18 | 218.139 | 63.8851 | 5.6 | 0.4 |
| polarC18 | 218.139 | 109.6697 | 5.6 | 0.4 |
| polarC18 | 218.139 | 201.4352 | 5.6 | 0.4 |
| polarC18 | 218.139 | 54.4798 | 5.6 | 0.4 |
| polarC18 | 218.139 | 62.0272 | 5.6 | 0.4 |
| polarC18 | 218.139 | 139.5216 | 5.6 | 0.4 |
| polarC18 | 218.139 | 62.6364 | 5.6 | 0.4 |
| polarC18 | 218.139 | 56.6414 | 5.6 | 0.4 |
| polarC18 | 218.139 | 49.2652 | 5.6 | 0.4 |
| polarC18 | 218.139 | 126.6095 | 5.6 | 0.4 |
| polarC18 | 218.139 | 227.4323 | 5.6 | 0.4 |
| polarC18 | 218.139 | 111.6868 | 5.6 | 0.4 |

| | | | | |
|---|---|---|---|---|
| polarC18 | 218.139 | 166.9282 | 5.6 | 0.4 |
| polarC18 | 218.139 | 223.3604 | 5.6 | 0.4 |
| polarC18 | 218.139 | 160.3073 | 5.6 | 0.4 |
| polarC18 | 218.139 | 53.4466 | 5.6 | 0.4 |
| polarC18 | 218.139 | 180.4769 | 5.6 | 0.4 |
| polarC18 | 218.139 | 87.8009 | 5.6 | 0.4 |
| polarC18 | 218.139 | 235.65 | 5.6 | 0.4 |
| polarC18 | 218.139 | 185.7565 | 5.6 | 0.4 |
| polarC18 | 218.139 | 53.7616 | 5.6 | 0.4 |
| polarC18 | 218.139 | 49.7983 | 5.6 | 0.4 |
| polarC18 | 218.139 | 52.2789 | 5.6 | 0.4 |
| polarC18 | 218.139 | 63.6096 | 5.6 | 0.4 |
| polarC18 | 218.139 | 161.9479 | 5.6 | 0.4 |
| polarC18 | 218.139 | 210.3987 | 5.6 | 0.4 |
| polarC18 | 218.139 | 107.2533 | 5.6 | 0.4 |
| polarC18 | 218.139 | 71.7833 | 5.6 | 0.4 |
| polarC18 | 218.139 | 112.7337 | 5.6 | 0.4 |
| polarC18 | 218.139 | 72.8442 | 5.6 | 0.4 |
| polarC18 | 218.139 | 68.2625 | 5.6 | 0.4 |
| polarC18 | 218.139 | 64.5019 | 5.6 | 0.4 |
| polarC18 | 218.139 | 50.1613 | 5.6 | 0.4 |
| polarC18 | 218.139 | 57.2104 | 5.6 | 0.4 |
| polarC18 | 218.139 | 51.1521 | 5.6 | 0.4 |
| polarC18 | 218.139 | 77.3357 | 5.6 | 0.4 |
| polarC18 | 218.139 | 60.2714 | 5.6 | 0.4 |
| polarC18 | 218.139 | 51.5889 | 5.6 | 0.4 |
| polarC18 | 218.139 | 152.1409 | 5.6 | 0.4 |
| polarC18 | 218.139 | 70.0454 | 5.6 | 0.4 |
| polarC18 | 218.139 | 55.4783 | 5.6 | 0.4 |
| polarC18 | 218.139 | 50.7675 | 5.6 | 0.4 |
| polarC18 | 218.139 | 53.5002 | 5.6 | 0.4 |
| polarC18 | 218.139 | 91.7593 | 5.6 | 0.4 |
| polarC18 | 218.139 | 74.4513 | 5.6 | 0.4 |
| polarC18 | 218.139 | 57.7146 | 5.6 | 0.4 |
| polarC18 | 218.139 | 66.4136 | 5.6 | 0.4 |
| polarC18 | 218.139 | 179.0592 | 5.6 | 0.4 |
| polarC18 | 218.139 | 49.0563 | 5.6 | 0.4 |
| polarC18 | 218.139 | 53.9727 | 5.6 | 0.4 |
| polarC18 | 218.139 | 57.0211 | 5.6 | 0.39 |
| polarC18 | 218.139 | 112.6876 | 5.6 | 0.39 |
| polarC18 | 218.139 | 57.4799 | 5.6 | 0.39 |
| polarC18 | 218.139 | 73.3167 | 5.6 | 0.39 |

| | | | | |
|---|---|---|---|---|
| polarC18 | 218.139 | 165.299 | 5.6 | 0.39 |
| polarC18 | 218.139 | 54.8977 | 5.6 | 0.39 |
| polarC18 | 218.139 | 69.1577 | 5.6 | 0.39 |
| polarC18 | 218.139 | 78.8849 | 5.6 | 0.39 |
| polarC18 | 218.139 | 49.8482 | 5.6 | 0.39 |
| polarC18 | 218.139 | 148.4598 | 5.6 | 0.39 |
| polarC18 | 218.139 | 63.2857 | 5.6 | 0.39 |
| polarC18 | 218.139 | 99.6522 | 5.6 | 0.39 |
| polarC18 | 218.139 | 126.0692 | 5.6 | 0.39 |
| polarC18 | 218.139 | 56.1697 | 5.6 | 0.39 |
| polarC18 | 218.139 | 119.7478 | 5.6 | 0.39 |
| polarC18 | 218.139 | 192.2196 | 5.6 | 0.39 |
| polarC18 | 218.139 | 49.4684 | 5.6 | 0.39 |
| polarC18 | 218.139 | 61.3841 | 5.6 | 0.39 |
| polarC18 | 218.139 | 61.4554 | 5.6 | 0.39 |
| polarC18 | 218.139 | 59.2954 | 5.6 | 0.39 |
| polarC18 | 218.139 | 129.9871 | 5.6 | 0.39 |
| polarC18 | 218.139 | 52.7891 | 5.6 | 0.39 |
| polarC18 | 218.139 | 95.8813 | 5.6 | 0.39 |
| polarC18 | 218.139 | 172.7206 | 5.6 | 0.39 |
| polarC18 | 218.139 | 64.7797 | 5.6 | 0.39 |
| polarC18 | 218.139 | 88.4857 | 5.6 | 0.39 |
| polarC18 | 218.139 | 91.9314 | 5.6 | 0.39 |
| polarC18 | 218.139 | 49.5658 | 5.5 | 0.39 |
| polarC18 | 218.139 | 67.0503 | 5.5 | 0.39 |
| polarC18 | 218.139 | 77.6873 | 5.5 | 0.39 |
| polarC18 | 218.139 | 59.5311 | 5.5 | 0.39 |
| polarC18 | 218.139 | 55.6882 | 5.5 | 0.39 |
| polarC18 | 218.139 | 88.804 | 5.5 | 0.39 |
| polarC18 | 218.139 | 61.1653 | 5.5 | 0.39 |
| polarC18 | 218.139 | 148.4118 | 5.5 | 0.39 |
| polarC18 | 218.139 | 50.6819 | 5.5 | 0.39 |
| polarC18 | 218.139 | 83.1494 | 5.5 | 0.39 |
| polarC18 | 218.139 | 103.9474 | 5.5 | 0.39 |
| polarC18 | 218.139 | 58.9221 | 5.5 | 0.39 |
| polarC18 | 218.139 | 77.9971 | 5.5 | 0.39 |
| polarC18 | 218.139 | 53.6527 | 5.5 | 0.39 |
| polarC18 | 218.139 | 120.0593 | 5.5 | 0.39 |
| polarC18 | 218.139 | 96.002 | 5.5 | 0.39 |
| polarC18 | 218.139 | 103.6045 | 5.5 | 0.39 |
| polarC18 | 218.139 | 69.8854 | 5.5 | 0.39 |
| polarC18 | 218.139 | 98.9457 | 5.5 | 0.39 |

| | | | | |
|---|---|---|---|---|
| polarC18 | 218.139 | 66.0953 | 5.5 | 0.39 |
| polarC18 | 218.139 | 113.3245 | 5.5 | 0.39 |
| polarC18 | 218.139 | 50.2602 | 5.5 | 0.39 |
| polarC18 | 218.139 | 53.1017 | 5.5 | 0.39 |
| polarC18 | 218.139 | 58.9266 | 5.5 | 0.39 |
| polarC18 | 218.139 | 146.341 | 5.5 | 0.39 |
| polarC18 | 218.139 | 72.9564 | 5.5 | 0.39 |
| polarC18 | 218.139 | 51.5987 | 5.5 | 0.39 |
| polarC18 | 218.139 | 69.665 | 5.5 | 0.39 |
| polarC18 | 218.139 | 73.2123 | 5.5 | 0.39 |
| polarC18 | 218.139 | 152.3289 | 5.5 | 0.39 |
| polarC18 | 218.139 | 67.5676 | 5.5 | 0.39 |
| polarC18 | 218.139 | 77.605 | 5.5 | 0.39 |
| polarC18 | 218.139 | 95.6121 | 5.5 | 0.39 |
| polarC18 | 218.139 | 99.4714 | 5.5 | 0.39 |
| polarC18 | 218.139 | 96.2446 | 5.5 | 0.39 |
| polarC18 | 218.139 | 82.715 | 5.5 | 0.39 |
| polarC18 | 218.139 | 59.269 | 5.5 | 0.39 |
| polarC18 | 218.139 | 234.2716 | 5.5 | 0.39 |
| polarC18 | 218.139 | 62.3347 | 5.5 | 0.39 |
| polarC18 | 218.139 | 111.6248 | 5.5 | 0.39 |
| polarC18 | 218.139 | 90.6728 | 5.5 | 0.39 |
| polarC18 | 218.139 | 59.7903 | 5.5 | 0.39 |
| polarC18 | 218.139 | 67.2136 | 5.5 | 0.39 |
| polarC18 | 218.139 | 68.6142 | 5.5 | 0.39 |
| polarC18 | 218.139 | 53.347 | 5.5 | 0.39 |
| polarC18 | 218.139 | 52.3902 | 5.5 | 0.39 |
| polarC18 | 218.139 | 49.5913 | 5.5 | 0.39 |
| polarC18 | 218.139 | 65.8906 | 5.5 | 0.39 |
| polarC18 | 218.139 | 77.5656 | 5.5 | 0.39 |
| polarC18 | 218.139 | 76.6441 | 5.5 | 0.39 |
| polarC18 | 218.139 | 63.3777 | 5.5 | 0.39 |
| polarC18 | 218.139 | 56.165 | 5.5 | 0.39 |
| polarC18 | 218.139 | 79.1413 | 5.5 | 0.39 |
| polarC18 | 218.139 | 65.8063 | 5.5 | 0.39 |
| polarC18 | 218.139 | 76.6847 | 5.5 | 0.39 |
| polarC18 | 218.139 | 102.9155 | 5.5 | 0.39 |
| polarC18 | 218.139 | 187.7263 | 5.5 | 0.39 |
| polarC18 | 218.139 | 193.1051 | 5.5 | 0.39 |
| polarC18 | 218.139 | 63.0744 | 5.5 | 0.39 |
| polarC18 | 218.139 | 53.7286 | 5.5 | 0.39 |
| polarC18 | 218.139 | 97.1929 | 5.5 | 0.39 |

| | | | | |
|---|---|---|---|---|
| polarC18 | 218.139 | 177.4358 | 5.5 | 0.39 |
| polarC18 | 218.139 | 95.8522 | 5.5 | 0.39 |
| polarC18 | 218.139 | 228.9803 | 5.5 | 0.39 |
| polarC18 | 218.139 | 99.8091 | 5.5 | 0.39 |
| polarC18 | 218.139 | 237.3258 | 5.5 | 0.39 |
| polarC18 | 218.139 | 201.9303 | 5.5 | 0.39 |
| polarC18 | 218.139 | 106.481 | 5.5 | 0.39 |
| polarC18 | 218.139 | 120.372 | 5.5 | 0.39 |
| polarC18 | 218.139 | 138.4063 | 5.5 | 0.39 |
| polarC18 | 218.139 | 86.2629 | 5.5 | 0.39 |
| polarC18 | 218.139 | 224.4896 | 5.5 | 0.39 |
| polarC18 | 218.139 | 153.1809 | 5.5 | 0.39 |
| polarC18 | 218.139 | 118.6458 | 5.5 | 0.39 |
| polarC18 | 218.139 | 151.8003 | 5.5 | 0.39 |
| polarC18 | 218.139 | 142.6133 | 5.5 | 0.39 |
| polarC18 | 218.139 | 110.7915 | 5.5 | 0.39 |
| polarC18 | 218.139 | 113.8375 | 5.5 | 0.39 |
| polarC18 | 218.139 | 106.9707 | 5.5 | 0.39 |
| polarC18 | 218.139 | 156.6547 | 5.5 | 0.39 |
| polarC18 | 218.139 | 50.3807 | 5.5 | 0.39 |
| polarC18 | 218.139 | 148.1808 | 5.5 | 0.39 |
| polarC18 | 218.139 | 105.6285 | 5.5 | 0.39 |
| polarC18 | 218.139 | 132.0435 | 5.5 | 0.39 |
| polarC18 | 218.139 | 173.8195 | 5.5 | 0.39 |
| polarC18 | 218.139 | 80.8085 | 5.5 | 0.39 |
| polarC18 | 218.139 | 108.0977 | 5.5 | 0.39 |
| polarC18 | 218.139 | 51.0193 | 5.5 | 0.39 |
| polarC18 | 218.139 | 54.5515 | 5.5 | 0.39 |
| polarC18 | 218.139 | 197.6286 | 5.5 | 0.39 |
| polarC18 | 218.139 | 61.5568 | 5.5 | 0.39 |
| polarC18 | 218.139 | 122.8607 | 5.5 | 0.39 |
| polarC18 | 218.139 | 240.8643 | 5.5 | 0.39 |
| polarC18 | 218.139 | 174.4562 | 5.5 | 0.39 |
| polarC18 | 218.139 | 50.498 | 5.5 | 0.39 |
| polarC18 | 218.139 | 56.6635 | 5.5 | 0.39 |
| polarC18 | 218.139 | 171.0218 | 5.5 | 0.39 |
| polarC18 | 218.139 | 58.6459 | 5.5 | 0.39 |
| polarC18 | 218.139 | 61.9276 | 5.5 | 0.39 |
| polarC18 | 218.139 | 60.079 | 5.5 | 0.39 |
| polarC18 | 218.139 | 92.3943 | 5.5 | 0.39 |
| polarC18 | 218.139 | 64.3098 | 5.4 | 0.39 |
| polarC18 | 218.139 | 147.8694 | 5.4 | 0.39 |

| | | | | |
|---|---|---|---|---|
| polarC18 | 218.139 | 61.8605 | 5.4 | 0.39 |
| polarC18 | 218.139 | 52.4846 | 5.4 | 0.39 |
| polarC18 | 218.139 | 62.8856 | 5.4 | 0.39 |
| polarC18 | 218.139 | 153.3627 | 5.4 | 0.39 |
| polarC18 | 218.139 | 109.8152 | 5.4 | 0.39 |
| polarC18 | 218.139 | 118.8133 | 5.4 | 0.39 |
| polarC18 | 218.139 | 49.7502 | 5.4 | 0.39 |
| polarC18 | 218.139 | 161.5382 | 5.4 | 0.39 |
| polarC18 | 218.139 | 52.4377 | 5.4 | 0.39 |
| polarC18 | 218.139 | 69.3343 | 5.4 | 0.39 |
| polarC18 | 218.139 | 58.8392 | 5.4 | 0.39 |
| polarC18 | 218.139 | 106.1399 | 5.4 | 0.39 |
| polarC18 | 218.139 | 119.352 | 5.4 | 0.39 |
| polarC18 | 218.139 | 151.6048 | 5.4 | 0.39 |
| polarC18 | 218.139 | 99.6835 | 5.4 | 0.39 |
| polarC18 | 218.139 | 50.593 | 5.4 | 0.39 |
| polarC18 | 218.139 | 62.8825 | 5.4 | 0.39 |
| polarC18 | 218.139 | 64.5318 | 5.4 | 0.39 |
| polarC18 | 218.139 | 98.5428 | 5.4 | 0.39 |
| polarC18 | 218.139 | 94.0377 | 5.4 | 0.39 |
| polarC18 | 218.139 | 201.5124 | 5.4 | 0.39 |
| polarC18 | 218.139 | 56.253 | 5.4 | 0.39 |
| polarC18 | 218.139 | 217.8899 | 5.4 | 0.39 |
| polarC18 | 218.139 | 80.5614 | 5.4 | 0.39 |
| polarC18 | 218.139 | 79.5812 | 5.4 | 0.39 |
| polarC18 | 218.139 | 73.8241 | 5.4 | 0.39 |
| polarC18 | 218.139 | 134.6319 | 5.4 | 0.39 |
| polarC18 | 218.139 | 69.8328 | 5.4 | 0.39 |
| polarC18 | 218.139 | 77.5016 | 5.4 | 0.38 |
| polarC18 | 218.139 | 119.7837 | 5.4 | 0.38 |
| polarC18 | 218.139 | 61.2443 | 5.4 | 0.38 |
| polarC18 | 218.139 | 69.9108 | 5.4 | 0.38 |
| polarC18 | 218.139 | 240.2447 | 5.4 | 0.38 |
| polarC18 | 218.139 | 76.6657 | 5.4 | 0.38 |
| polarC18 | 218.139 | 57.4768 | 5.4 | 0.38 |
| polarC18 | 218.139 | 90.5362 | 5.4 | 0.38 |
| polarC18 | 218.139 | 96.9994 | 5.4 | 0.38 |
| polarC18 | 218.139 | 60.7684 | 5.4 | 0.38 |
| polarC18 | 218.139 | 123.4596 | 5.4 | 0.38 |
| polarC18 | 218.139 | 83.5062 | 5.4 | 0.38 |
| polarC18 | 218.139 | 62.0589 | 5.4 | 0.38 |
| polarC18 | 218.139 | 101.4335 | 5.4 | 0.38 |

| | | | | |
|---|---|---|---|---|
| polarC18 | 218.139 | 140.4911 | 5.4 | 0.38 |
| polarC18 | 218.139 | 50.237 | 5.4 | 0.38 |
| polarC18 | 218.139 | 66.8057 | 5.4 | 0.38 |
| polarC18 | 218.139 | 57.571 | 5.4 | 0.38 |
| polarC18 | 218.139 | 53.6234 | 5.4 | 0.38 |
| polarC18 | 218.139 | 173.5747 | 5.4 | 0.38 |
| polarC18 | 218.139 | 142.5219 | 5.4 | 0.38 |
| polarC18 | 218.139 | 65.1826 | 5.4 | 0.38 |
| polarC18 | 218.139 | 49.6851 | 5.4 | 0.38 |
| polarC18 | 218.139 | 157.1513 | 5.4 | 0.38 |
| polarC18 | 218.139 | 139.7677 | 5.4 | 0.38 |
| polarC18 | 218.139 | 178.2605 | 5.4 | 0.38 |
| polarC18 | 218.139 | 51.9799 | 5.4 | 0.38 |
| polarC18 | 218.139 | 143.0425 | 5.4 | 0.38 |
| polarC18 | 218.139 | 59.5187 | 5.4 | 0.38 |
| polarC18 | 218.139 | 130.8058 | 5.4 | 0.38 |
| polarC18 | 218.139 | 84.5537 | 5.4 | 0.38 |
| polarC18 | 218.139 | 101.0539 | 5.4 | 0.38 |
| polarC18 | 218.139 | 168.774 | 5.4 | 0.38 |
| polarC18 | 218.139 | 83.5925 | 5.4 | 0.38 |
| polarC18 | 218.139 | 50.9093 | 5.4 | 0.38 |
| polarC18 | 218.139 | 52.9293 | 5.4 | 0.38 |
| polarC18 | 218.139 | 59.2118 | 5.4 | 0.38 |
| polarC18 | 218.139 | 181.3771 | 5.4 | 0.38 |
| polarC18 | 218.139 | 73.8117 | 5.4 | 0.38 |
| polarC18 | 218.139 | 137.9873 | 5.4 | 0.38 |
| polarC18 | 218.139 | 181.8801 | 5.4 | 0.38 |
| polarC18 | 218.139 | 58.9327 | 5.4 | 0.38 |
| polarC18 | 218.139 | 63.0409 | 5.4 | 0.38 |
| polarC18 | 218.139 | 89.3345 | 5.4 | 0.38 |
| polarC18 | 218.139 | 118.497 | 5.4 | 0.38 |
| polarC18 | 218.139 | 159.4981 | 5.4 | 0.38 |
| polarC18 | 218.139 | 164.3359 | 5.4 | 0.38 |
| polarC18 | 218.139 | 61.0638 | 5.4 | 0.38 |
| polarC18 | 218.139 | 75.0578 | 5.4 | 0.38 |
| polarC18 | 218.139 | 84.1993 | 5.4 | 0.38 |
| polarC18 | 218.139 | 235.5534 | 5.4 | 0.38 |
| polarC18 | 218.139 | 49.3687 | 5.4 | 0.38 |
| polarC18 | 218.139 | 158.3642 | 5.4 | 0.38 |
| polarC18 | 218.139 | 142.361 | 5.4 | 0.38 |
| polarC18 | 218.139 | 207.294 | 5.4 | 0.38 |
| polarC18 | 218.139 | 88.329 | 5.4 | 0.38 |

| | | | | |
|---|---|---|---|---|
| polarC18 | 218.139 | 57.3019 | 5.4 | 0.38 |
| polarC18 | 218.139 | 50.786 | 5.4 | 0.38 |
| polarC18 | 218.139 | 71.5655 | 5.4 | 0.38 |
| polarC18 | 218.139 | 59.6165 | 5.4 | 0.38 |
| polarC18 | 218.139 | 69.3204 | 5.4 | 0.38 |
| polarC18 | 218.139 | 70.2291 | 5.4 | 0.38 |
| polarC18 | 218.139 | 105.7296 | 5.4 | 0.38 |
| polarC18 | 218.139 | 61.5822 | 5.4 | 0.38 |
| polarC18 | 218.139 | 158.0275 | 5.4 | 0.38 |
| polarC18 | 218.139 | 49.3247 | 5.4 | 0.38 |
| polarC18 | 218.139 | 50.5845 | 5.4 | 0.38 |
| polarC18 | 218.139 | 143.7701 | 5.4 | 0.38 |
| polarC18 | 218.139 | 91.7373 | 5.4 | 0.38 |
| polarC18 | 218.139 | 75.386 | 5.4 | 0.38 |
| polarC18 | 218.139 | 62.7025 | 5.4 | 0.38 |
| polarC18 | 218.139 | 65.4636 | 5.4 | 0.38 |
| polarC18 | 218.139 | 219.0669 | 5.4 | 0.38 |
| polarC18 | 218.139 | 81.0409 | 5.4 | 0.38 |
| polarC18 | 218.139 | 89.7959 | 5.4 | 0.38 |
| polarC18 | 218.139 | 122.8731 | 5.4 | 0.38 |
| polarC18 | 218.139 | 50.1537 | 5.4 | 0.38 |
| polarC18 | 218.139 | 59.0772 | 5.4 | 0.38 |
| polarC18 | 218.139 | 124.7251 | 5.4 | 0.38 |
| polarC18 | 218.139 | 71.2561 | 5.3 | 0.38 |
| polarC18 | 218.139 | 69.54 | 5.3 | 0.38 |
| polarC18 | 218.139 | 93.3485 | 5.3 | 0.38 |
| polarC18 | 218.139 | 62.7255 | 5.3 | 0.38 |
| polarC18 | 218.139 | 187.5843 | 5.3 | 0.38 |
| polarC18 | 218.139 | 162.448 | 5.3 | 0.38 |
| polarC18 | 218.139 | 52.3449 | 5.3 | 0.38 |
| polarC18 | 218.139 | 105.9748 | 5.3 | 0.38 |
| polarC18 | 218.139 | 197.9863 | 5.3 | 0.38 |
| polarC18 | 218.139 | 56.5993 | 5.3 | 0.38 |
| polarC18 | 218.139 | 87.499 | 5.3 | 0.38 |
| polarC18 | 218.139 | 137.0702 | 5.3 | 0.38 |
| polarC18 | 218.139 | 179.9318 | 5.3 | 0.38 |
| polarC18 | 218.139 | 51.2116 | 5.3 | 0.38 |
| polarC18 | 218.139 | 125.5672 | 5.3 | 0.38 |
| polarC18 | 218.139 | 188.2065 | 5.3 | 0.38 |
| polarC18 | 218.139 | 93.2688 | 5.3 | 0.38 |
| polarC18 | 218.139 | 113.3321 | 5.3 | 0.38 |
| polarC18 | 218.139 | 154.3738 | 5.3 | 0.38 |

| | | | | |
|---|---|---|---|---|
| polarC18 | 218.139 | 58.9086 | 5.3 | 0.38 |
| polarC18 | 218.139 | 123.3541 | 5.3 | 0.38 |
| polarC18 | 218.139 | 100.8213 | 5.3 | 0.38 |
| polarC18 | 218.139 | 57.1468 | 5.3 | 0.38 |
| polarC18 | 218.139 | 99.2134 | 5.3 | 0.38 |
| polarC18 | 218.139 | 127.6797 | 5.3 | 0.38 |
| polarC18 | 218.139 | 64.9573 | 5.3 | 0.38 |
| polarC18 | 218.139 | 62.9836 | 5.3 | 0.38 |
| polarC18 | 218.139 | 93.1825 | 5.3 | 0.38 |
| polarC18 | 218.139 | 64.4893 | 5.3 | 0.38 |
| polarC18 | 218.139 | 126.0925 | 5.3 | 0.38 |
| polarC18 | 218.139 | 87.0125 | 5.3 | 0.38 |
| polarC18 | 218.139 | 52.4907 | 5.3 | 0.38 |
| polarC18 | 218.139 | 63.6499 | 5.3 | 0.38 |
| polarC18 | 218.139 | 68.3564 | 5.3 | 0.38 |
| polarC18 | 218.139 | 69.7524 | 5.3 | 0.38 |
| polarC18 | 218.139 | 163.0672 | 5.3 | 0.38 |
| polarC18 | 218.139 | 68.2775 | 5.3 | 0.38 |
| polarC18 | 218.139 | 84.7266 | 5.3 | 0.38 |
| polarC18 | 218.139 | 141.3137 | 5.3 | 0.38 |
| polarC18 | 218.139 | 111.4947 | 5.3 | 0.38 |
| polarC18 | 218.139 | 50.3493 | 5.3 | 0.38 |
| polarC18 | 218.139 | 57.2568 | 5.3 | 0.38 |
| polarC18 | 218.139 | 56.6955 | 5.3 | 0.38 |
| polarC18 | 218.139 | 92.9654 | 5.3 | 0.38 |
| polarC18 | 218.139 | 182.9264 | 5.3 | 0.38 |
| polarC18 | 218.139 | 147.4278 | 5.3 | 0.38 |
| polarC18 | 218.139 | 118.1541 | 5.3 | 0.38 |
| polarC18 | 218.139 | 80.1623 | 5.3 | 0.38 |
| polarC18 | 218.139 | 63.9584 | 5.3 | 0.38 |
| polarC18 | 218.139 | 81.8273 | 5.3 | 0.38 |
| polarC18 | 218.139 | 67.5519 | 5.3 | 0.38 |
| polarC18 | 218.139 | 67.1308 | 5.3 | 0.38 |
| polarC18 | 218.139 | 145.1639 | 5.3 | 0.38 |
| polarC18 | 218.139 | 48.8606 | 5.3 | 0.38 |
| polarC18 | 218.139 | 153.1621 | 5.3 | 0.38 |
| polarC18 | 218.139 | 79.7322 | 5.3 | 0.38 |
| polarC18 | 218.139 | 122.656 | 5.3 | 0.38 |
| polarC18 | 218.139 | 122.6646 | 5.3 | 0.38 |
| polarC18 | 218.139 | 99.6372 | 5.3 | 0.38 |
| polarC18 | 218.139 | 54.1736 | 5.3 | 0.38 |
| polarC18 | 218.139 | 85.1169 | 5.3 | 0.38 |

| | | | | |
|---|---|---|---|---|
| polarC18 | 218.139 | 48.6692 | 5.3 | 0.38 |
| polarC18 | 218.139 | 84.6829 | 5.3 | 0.38 |
| polarC18 | 218.139 | 71.899 | 5.3 | 0.38 |
| polarC18 | 218.139 | 151.254 | 5.3 | 0.38 |
| polarC18 | 218.139 | 138.6102 | 5.3 | 0.38 |
| polarC18 | 218.139 | 49.1116 | 5.3 | 0.38 |
| polarC18 | 218.139 | 111.9463 | 5.3 | 0.38 |
| polarC18 | 218.139 | 213.9582 | 5.3 | 0.38 |
| polarC18 | 218.139 | 142.7629 | 5.3 | 0.38 |
| polarC18 | 218.139 | 128.8449 | 5.3 | 0.38 |
| polarC18 | 218.139 | 58.3193 | 5.3 | 0.38 |
| polarC18 | 218.139 | 85.3131 | 5.3 | 0.37 |
| polarC18 | 218.139 | 102.7743 | 5.3 | 0.37 |
| polarC18 | 218.139 | 215.8397 | 5.3 | 0.37 |
| polarC18 | 218.139 | 112.3808 | 5.3 | 0.37 |
| polarC18 | 218.139 | 73.6733 | 5.3 | 0.37 |
| polarC18 | 218.139 | 50.1496 | 5.3 | 0.37 |
| polarC18 | 218.139 | 75.431 | 5.3 | 0.37 |
| polarC18 | 218.139 | 53.5574 | 5.3 | 0.37 |
| polarC18 | 218.139 | 163.1657 | 5.3 | 0.37 |
| polarC18 | 218.139 | 58.4408 | 5.3 | 0.37 |
| polarC18 | 218.139 | 85.7346 | 5.3 | 0.37 |
| polarC18 | 218.139 | 85.332 | 5.3 | 0.37 |
| polarC18 | 218.139 | 115.7661 | 5.3 | 0.37 |
| polarC18 | 218.139 | 60.9905 | 5.3 | 0.37 |
| polarC18 | 218.139 | 54.0225 | 5.3 | 0.37 |
| polarC18 | 218.139 | 119.3886 | 5.3 | 0.37 |
| polarC18 | 218.139 | 131.2879 | 5.3 | 0.37 |
| polarC18 | 218.139 | 111.0074 | 5.3 | 0.37 |
| polarC18 | 218.139 | 50.9722 | 5.3 | 0.37 |
| polarC18 | 218.139 | 234.0079 | 5.3 | 0.37 |
| polarC18 | 218.139 | 89.3716 | 5.3 | 0.37 |
| polarC18 | 218.139 | 152.7897 | 5.3 | 0.37 |
| polarC18 | 218.139 | 98.2884 | 5.3 | 0.37 |
| polarC18 | 218.139 | 51.0477 | 5.3 | 0.37 |
| polarC18 | 218.139 | 70.8159 | 5.3 | 0.37 |
| polarC18 | 218.139 | 52.159 | 5.3 | 0.37 |
| polarC18 | 218.139 | 193.6305 | 5.3 | 0.37 |
| polarC18 | 218.139 | 128.0337 | 5.3 | 0.37 |
| polarC18 | 218.139 | 71.6844 | 5.3 | 0.37 |
| polarC18 | 218.139 | 61.208 | 5.3 | 0.37 |
| polarC18 | 218.139 | 97.9638 | 5.3 | 0.37 |

| | | | | |
|---|---|---|---|---|
| polarC18 | 218.139 | 70.5483 | 5.3 | 0.37 |
| polarC18 | 218.139 | 201.996 | 5.3 | 0.37 |
| polarC18 | 218.139 | 178.0976 | 5.3 | 0.37 |
| polarC18 | 218.139 | 63.2006 | 5.3 | 0.37 |
| polarC18 | 218.139 | 63.94 | 5.3 | 0.37 |
| polarC18 | 218.139 | 163.2519 | 5.3 | 0.37 |
| polarC18 | 218.139 | 106.0012 | 5.3 | 0.37 |
| polarC18 | 218.139 | 82.6878 | 5.3 | 0.37 |
| polarC18 | 218.139 | 73.8584 | 5.3 | 0.37 |
| polarC18 | 218.139 | 101.5248 | 5.3 | 0.37 |
| polarC18 | 218.139 | 169.7759 | 5.3 | 0.37 |
| polarC18 | 218.139 | 168.3605 | 5.3 | 0.37 |
| polarC18 | 218.139 | 80.6633 | 5.3 | 0.37 |
| polarC18 | 218.139 | 179.5963 | 5.2 | 0.37 |
| polarC18 | 218.139 | 70.0719 | 5.2 | 0.37 |
| polarC18 | 218.139 | 70.1531 | 5.2 | 0.37 |
| polarC18 | 218.139 | 85.0896 | 5.2 | 0.37 |
| polarC18 | 218.139 | 85.0066 | 5.2 | 0.37 |
| polarC18 | 218.139 | 171.5023 | 5.2 | 0.37 |
| polarC18 | 218.139 | 117.7778 | 5.2 | 0.37 |
| polarC18 | 218.139 | 64.4535 | 5.2 | 0.37 |
| polarC18 | 218.139 | 117.6347 | 5.2 | 0.37 |
| polarC18 | 218.139 | 115.8455 | 5.2 | 0.37 |
| polarC18 | 218.139 | 111.8545 | 5.2 | 0.37 |
| polarC18 | 218.139 | 59.3535 | 5.2 | 0.37 |
| polarC18 | 218.139 | 64.0918 | 5.2 | 0.37 |
| polarC18 | 218.139 | 89.0952 | 5.2 | 0.37 |
| polarC18 | 218.139 | 80.1955 | 5.2 | 0.37 |
| polarC18 | 218.139 | 65.9503 | 5.2 | 0.37 |
| polarC18 | 218.139 | 71.7464 | 5.2 | 0.37 |
| polarC18 | 218.139 | 78.482 | 5.2 | 0.37 |
| polarC18 | 218.139 | 137.5439 | 5.2 | 0.37 |
| polarC18 | 218.139 | 49.3297 | 5.2 | 0.37 |
| polarC18 | 218.139 | 80.2917 | 5.2 | 0.37 |
| polarC18 | 218.139 | 62.2903 | 5.2 | 0.37 |
| polarC18 | 218.139 | 63.4642 | 5.2 | 0.37 |
| polarC18 | 218.139 | 156.0709 | 5.2 | 0.37 |
| polarC18 | 218.139 | 101.5401 | 5.2 | 0.37 |
| polarC18 | 218.139 | 50.0864 | 5.2 | 0.37 |
| polarC18 | 218.139 | 57.3498 | 5.2 | 0.37 |
| polarC18 | 218.139 | 60.0403 | 5.2 | 0.37 |
| polarC18 | 218.139 | 52.7678 | 5.2 | 0.37 |

| | | | | |
|---|---|---|---|---|
| polarC18 | 218.139 | 59.5477 | 5.2 | 0.37 |
| polarC18 | 218.139 | 190.2373 | 5.2 | 0.37 |
| polarC18 | 218.139 | 83.9986 | 5.2 | 0.37 |
| polarC18 | 218.139 | 84.4935 | 5.2 | 0.37 |
| polarC18 | 218.139 | 127.4616 | 5.2 | 0.37 |
| polarC18 | 218.139 | 49.3961 | 5.2 | 0.37 |
| polarC18 | 218.139 | 123.6551 | 5.2 | 0.37 |
| polarC18 | 218.139 | 80.1569 | 5.2 | 0.37 |
| polarC18 | 218.139 | 204.0371 | 5.2 | 0.37 |
| polarC18 | 218.139 | 103.5007 | 5.2 | 0.37 |
| polarC18 | 218.139 | 60.6923 | 5.2 | 0.37 |
| polarC18 | 218.139 | 85.0961 | 5.2 | 0.37 |
| polarC18 | 218.139 | 57.5166 | 5.2 | 0.37 |
| polarC18 | 218.139 | 140.5625 | 5.2 | 0.37 |
| polarC18 | 218.139 | 95.5768 | 5.2 | 0.37 |
| polarC18 | 218.139 | 53.4984 | 5.2 | 0.37 |
| polarC18 | 218.139 | 73.3734 | 5.2 | 0.37 |
| polarC18 | 218.139 | 50.8952 | 5.2 | 0.37 |
| polarC18 | 218.139 | 48.7147 | 5.2 | 0.37 |
| polarC18 | 218.139 | 57.1289 | 5.2 | 0.37 |
| polarC18 | 218.139 | 52.6312 | 5.2 | 0.37 |
| polarC18 | 218.139 | 50.3332 | 5.2 | 0.37 |
| polarC18 | 218.139 | 62.0412 | 5.2 | 0.37 |
| polarC18 | 218.139 | 85.4561 | 5.2 | 0.37 |
| polarC18 | 218.139 | 123.609 | 5.2 | 0.37 |
| polarC18 | 218.139 | 52.7435 | 5.2 | 0.37 |
| polarC18 | 218.139 | 95.939 | 5.2 | 0.37 |
| polarC18 | 218.139 | 80.6182 | 5.2 | 0.37 |
| polarC18 | 218.139 | 52.415 | 5.2 | 0.37 |
| polarC18 | 218.139 | 217.1713 | 5.2 | 0.37 |
| polarC18 | 218.139 | 194.4162 | 5.2 | 0.37 |
| polarC18 | 218.139 | 123.6984 | 5.2 | 0.37 |
| polarC18 | 218.139 | 238.1054 | 5.2 | 0.37 |
| polarC18 | 218.139 | 76.9177 | 5.2 | 0.37 |
| polarC18 | 218.139 | 58.7028 | 5.2 | 0.37 |
| polarC18 | 218.139 | 139.4647 | 5.2 | 0.37 |
| polarC18 | 218.139 | 159.3735 | 5.2 | 0.37 |
| polarC18 | 218.139 | 85.8987 | 5.2 | 0.37 |
| polarC18 | 218.139 | 160.595 | 5.2 | 0.37 |
| polarC18 | 218.139 | 77.9436 | 5.2 | 0.37 |
| polarC18 | 218.139 | 130.4031 | 5.2 | 0.37 |
| polarC18 | 218.139 | 70.0275 | 5.2 | 0.37 |

| | | | | |
|---|---|---|---|---|
| polarC18 | 218.139 | 62.4492 | 5.2 | 0.37 |
| polarC18 | 218.139 | 79.5078 | 5.2 | 0.37 |
| polarC18 | 218.139 | 84.2375 | 5.2 | 0.37 |
| polarC18 | 218.139 | 114.1433 | 5.2 | 0.37 |
| polarC18 | 218.139 | 121.192 | 5.2 | 0.37 |
| polarC18 | 218.139 | 109.2761 | 5.2 | 0.37 |
| polarC18 | 218.139 | 127.5218 | 5.2 | 0.37 |
| polarC18 | 218.139 | 124.018 | 5.2 | 0.37 |
| polarC18 | 218.139 | 67.9451 | 5.2 | 0.37 |
| polarC18 | 218.139 | 133.1804 | 5.2 | 0.37 |
| polarC18 | 218.139 | 53.0635 | 5.2 | 0.37 |
| polarC18 | 218.139 | 120.325 | 5.2 | 0.37 |
| polarC18 | 218.139 | 63.6596 | 5.2 | 0.37 |
| polarC18 | 218.139 | 168.2556 | 5.2 | 0.37 |
| polarC18 | 218.139 | 100.2824 | 5.2 | 0.37 |
| polarC18 | 218.139 | 171.0651 | 5.2 | 0.37 |
| polarC18 | 218.139 | 61.802 | 5.2 | 0.37 |
| polarC18 | 218.139 | 95.7918 | 5.2 | 0.37 |
| polarC18 | 218.139 | 69.0824 | 5.2 | 0.37 |
| polarC18 | 218.139 | 100.972 | 5.2 | 0.37 |
| polarC18 | 218.139 | 139.3409 | 5.2 | 0.37 |
| polarC18 | 218.139 | 167.2735 | 5.2 | 0.37 |
| polarC18 | 218.139 | 87.7121 | 5.2 | 0.37 |
| polarC18 | 218.139 | 55.8919 | 5.2 | 0.37 |
| polarC18 | 218.139 | 86.3982 | 5.2 | 0.37 |
| polarC18 | 218.139 | 105.1763 | 5.2 | 0.37 |
| polarC18 | 218.139 | 241.4933 | 5.2 | 0.37 |
| polarC18 | 218.139 | 54.8314 | 5.2 | 0.37 |
| polarC18 | 218.139 | 105.9646 | 5.2 | 0.37 |
| polarC18 | 218.139 | 57.5078 | 5.2 | 0.37 |
| polarC18 | 218.139 | 77.8615 | 5.2 | 0.37 |
| polarC18 | 218.139 | 81.1315 | 5.2 | 0.37 |
| polarC18 | 218.139 | 217.4941 | 5.2 | 0.37 |
| polarC18 | 218.139 | 51.0104 | 5.2 | 0.37 |
| polarC18 | 218.139 | 213.6271 | 5.2 | 0.37 |
| polarC18 | 218.139 | 208.0444 | 5.2 | 0.37 |
| polarC18 | 218.139 | 117.0144 | 5.2 | 0.37 |
| polarC18 | 218.139 | 55.5565 | 5.2 | 0.37 |
| polarC18 | 218.139 | 92.1351 | 5.2 | 0.37 |
| polarC18 | 218.139 | 87.0934 | 5.2 | 0.37 |
| polarC18 | 218.139 | 49.6515 | 5.2 | 0.37 |
| polarC18 | 218.139 | 86.5208 | 5.2 | 0.37 |

| | | | | |
|---|---|---|---|---|
| polarC18 | 218.139 | 74.3716 | 5.2 | 0.37 |
| polarC18 | 218.139 | 157.5368 | 5.2 | 0.37 |
| polarC18 | 218.139 | 94.6493 | 5.2 | 0.37 |
| polarC18 | 218.139 | 84.2234 | 5.2 | 0.37 |
| polarC18 | 218.139 | 62.4831 | 5.2 | 0.37 |
| polarC18 | 218.139 | 71.6209 | 5.2 | 0.37 |
| polarC18 | 218.139 | 225.3923 | 5.2 | 0.37 |
| polarC18 | 218.139 | 113.173 | 5.2 | 0.37 |
| polarC18 | 218.139 | 81.7943 | 5.2 | 0.37 |
| polarC18 | 218.139 | 59.0141 | 5.2 | 0.37 |
| polarC18 | 218.139 | 71.8582 | 5.2 | 0.37 |
| polarC18 | 218.139 | 101.3752 | 5.2 | 0.37 |
| polarC18 | 218.139 | 73.8625 | 5.2 | 0.37 |
| polarC18 | 218.139 | 219.5533 | 5.2 | 0.37 |
| polarC18 | 218.139 | 152.5031 | 5.2 | 0.37 |
| polarC18 | 218.139 | 110.4111 | 5.1 | 0.37 |
| polarC18 | 218.139 | 84.6522 | 5.1 | 0.37 |
| polarC18 | 218.139 | 95.5031 | 5.1 | 0.37 |
| polarC18 | 218.139 | 123.0651 | 5.1 | 0.37 |
| polarC18 | 218.139 | 88.9872 | 5.1 | 0.37 |
| polarC18 | 218.139 | 174.3908 | 5.1 | 0.37 |
| polarC18 | 218.139 | 63.7672 | 5.1 | 0.37 |
| polarC18 | 218.139 | 148.1011 | 5.1 | 0.37 |
| polarC18 | 218.139 | 89.261 | 5.1 | 0.37 |
| polarC18 | 218.139 | 79.8332 | 5.1 | 0.36 |
| polarC18 | 218.139 | 60.9957 | 5.1 | 0.36 |
| polarC18 | 218.139 | 164.7898 | 5.1 | 0.36 |
| polarC18 | 218.139 | 80.6502 | 5.1 | 0.36 |
| polarC18 | 218.139 | 56.5053 | 5.1 | 0.36 |
| polarC18 | 218.139 | 138.9125 | 5.1 | 0.36 |
| polarC18 | 218.139 | 49.8609 | 5.1 | 0.36 |
| polarC18 | 218.139 | 75.6232 | 5.1 | 0.36 |
| polarC18 | 218.139 | 107.7517 | 5.1 | 0.36 |
| polarC18 | 218.139 | 111.7102 | 5.1 | 0.36 |
| polarC18 | 218.139 | 210.9095 | 5.1 | 0.36 |
| polarC18 | 218.139 | 59.9237 | 5.1 | 0.36 |
| polarC18 | 218.139 | 130.6847 | 5.1 | 0.36 |
| polarC18 | 218.139 | 63.4594 | 5.1 | 0.36 |
| polarC18 | 218.139 | 59.1254 | 5.1 | 0.36 |
| polarC18 | 218.139 | 77.3688 | 5.1 | 0.36 |
| polarC18 | 218.139 | 50.0351 | 5.1 | 0.36 |
| polarC18 | 218.139 | 82.5321 | 5.1 | 0.36 |

| | | | | |
|---|---|---|---|---|
| polarC18 | 218.139 | 137.7059 | 5.1 | 0.36 |
| polarC18 | 218.139 | 50.6643 | 5.1 | 0.36 |
| polarC18 | 218.139 | 48.9336 | 5.1 | 0.36 |
| polarC18 | 218.139 | 56.0161 | 5.1 | 0.36 |
| polarC18 | 218.139 | 138.5248 | 5.1 | 0.36 |
| polarC18 | 218.139 | 151.717 | 5.1 | 0.36 |
| polarC18 | 218.139 | 131.3008 | 5.1 | 0.36 |
| polarC18 | 218.139 | 98.4257 | 5.1 | 0.36 |
| polarC18 | 218.139 | 239.9057 | 5.1 | 0.36 |
| polarC18 | 218.139 | 155.3466 | 5.1 | 0.36 |
| polarC18 | 218.139 | 51.6653 | 5.1 | 0.36 |
| polarC18 | 218.139 | 55.4011 | 5.1 | 0.36 |
| polarC18 | 218.139 | 57.6673 | 5.1 | 0.36 |
| polarC18 | 218.139 | 62.502 | 5.1 | 0.36 |
| polarC18 | 218.139 | 88.5218 | 5.1 | 0.36 |
| polarC18 | 218.139 | 127.343 | 5.1 | 0.36 |
| polarC18 | 218.139 | 72.1249 | 5.1 | 0.36 |
| polarC18 | 218.139 | 78.7348 | 5.1 | 0.36 |
| polarC18 | 218.139 | 131.6028 | 5.1 | 0.36 |
| polarC18 | 218.139 | 76.1526 | 5.1 | 0.36 |
| polarC18 | 218.139 | 69.4418 | 5.1 | 0.36 |
| polarC18 | 218.139 | 206.8331 | 5.1 | 0.36 |
| polarC18 | 218.139 | 96.5101 | 5.1 | 0.36 |
| polarC18 | 218.139 | 103.2811 | 5.1 | 0.36 |
| polarC18 | 218.139 | 229.9905 | 5.1 | 0.36 |
| polarC18 | 218.139 | 53.4241 | 5.1 | 0.36 |
| polarC18 | 218.139 | 184.3333 | 5.1 | 0.36 |
| polarC18 | 218.139 | 49.3746 | 5.1 | 0.36 |
| polarC18 | 218.139 | 204.5309 | 5.1 | 0.36 |
| polarC18 | 218.139 | 173.4268 | 5.1 | 0.36 |
| polarC18 | 218.139 | 89.1477 | 5.1 | 0.36 |
| polarC18 | 218.139 | 53.1537 | 5.1 | 0.36 |
| polarC18 | 218.139 | 91.4131 | 5.1 | 0.36 |
| polarC18 | 218.139 | 64.0007 | 5.1 | 0.36 |
| polarC18 | 218.139 | 53.1067 | 5.1 | 0.36 |
| polarC18 | 218.139 | 120.9359 | 5.1 | 0.36 |
| polarC18 | 218.139 | 66.7547 | 5.1 | 0.36 |
| polarC18 | 218.139 | 74.0419 | 5.1 | 0.36 |
| polarC18 | 218.139 | 72.1954 | 5.1 | 0.36 |
| polarC18 | 218.139 | 95.5545 | 5.1 | 0.36 |
| polarC18 | 218.139 | 142.7011 | 5.1 | 0.36 |
| polarC18 | 218.139 | 102.3848 | 5.1 | 0.36 |

| | | | | |
|---|---|---|---|---|
| polarC18 | 218.139 | 65.4331 | 5.1 | 0.36 |
| polarC18 | 218.139 | 72.7069 | 5.1 | 0.36 |
| polarC18 | 218.139 | 162.6028 | 5.1 | 0.36 |
| polarC18 | 218.139 | 73.9337 | 5.1 | 0.36 |
| polarC18 | 218.139 | 58.6842 | 5.1 | 0.36 |
| polarC18 | 218.139 | 66.0669 | 5.1 | 0.36 |
| polarC18 | 218.139 | 172.0228 | 5.1 | 0.36 |
| polarC18 | 218.139 | 57.0231 | 5.1 | 0.36 |
| polarC18 | 218.139 | 67.3558 | 5.1 | 0.36 |
| polarC18 | 218.139 | 85.6708 | 5.1 | 0.36 |
| polarC18 | 218.139 | 93.4085 | 5.1 | 0.36 |
| polarC18 | 218.139 | 141.3645 | 5.1 | 0.36 |
| polarC18 | 218.139 | 110.546 | 5.1 | 0.36 |
| polarC18 | 218.139 | 52.8397 | 5.1 | 0.36 |
| polarC18 | 218.139 | 105.2527 | 5.1 | 0.36 |
| polarC18 | 218.139 | 55.3131 | 5.1 | 0.36 |
| polarC18 | 218.139 | 122.6336 | 5.1 | 0.36 |
| polarC18 | 218.139 | 51.8047 | 5.1 | 0.36 |
| polarC18 | 218.139 | 53.8955 | 5.1 | 0.36 |
| polarC18 | 218.139 | 52.444 | 5.1 | 0.36 |
| polarC18 | 218.139 | 56.9233 | 5.1 | 0.36 |
| polarC18 | 218.139 | 67.6536 | 5.1 | 0.36 |
| polarC18 | 218.139 | 181.6065 | 5.1 | 0.36 |
| polarC18 | 218.139 | 87.196 | 5.1 | 0.36 |
| polarC18 | 218.139 | 50.7149 | 5.1 | 0.36 |
| polarC18 | 218.139 | 126.6679 | 5.1 | 0.36 |
| polarC18 | 218.139 | 126.2657 | 5.1 | 0.36 |
| polarC18 | 218.139 | 78.2288 | 5.1 | 0.36 |
| polarC18 | 218.139 | 106.1115 | 5.1 | 0.36 |
| polarC18 | 218.139 | 62.7669 | 5.1 | 0.36 |
| polarC18 | 218.139 | 92.9105 | 5.1 | 0.36 |
| polarC18 | 218.139 | 119.7921 | 5.1 | 0.36 |
| polarC18 | 218.139 | 153.056 | 5.1 | 0.36 |
| polarC18 | 218.139 | 80.5922 | 5.1 | 0.36 |
| polarC18 | 218.139 | 205.6459 | 5.1 | 0.36 |
| polarC18 | 218.139 | 197.7412 | 5.1 | 0.36 |
| polarC18 | 218.139 | 53.0366 | 5.1 | 0.36 |
| polarC18 | 218.139 | 154.5733 | 5.1 | 0.36 |
| polarC18 | 218.139 | 102.3908 | 5.1 | 0.36 |
| polarC18 | 218.139 | 169.1756 | 5.1 | 0.36 |
| polarC18 | 218.139 | 93.8424 | 5.1 | 0.36 |
| polarC18 | 218.139 | 166.0617 | 5.1 | 0.36 |

| | | | | |
|---|---|---|---|---|
| polarC18 | 218.139 | 114.7072 | 5.1 | 0.36 |
| polarC18 | 218.139 | 139.8467 | 5.1 | 0.36 |
| polarC18 | 218.139 | 55.9784 | 5.1 | 0.36 |
| polarC18 | 218.139 | 104.3963 | 5.1 | 0.36 |
| polarC18 | 218.139 | 74.6349 | 5.1 | 0.36 |
| polarC18 | 218.139 | 61.5387 | 5.1 | 0.36 |
| polarC18 | 218.139 | 56.5897 | 5.1 | 0.36 |
| polarC18 | 218.139 | 215.5546 | 5.1 | 0.36 |
| polarC18 | 218.139 | 199.4526 | 5.1 | 0.36 |
| polarC18 | 218.139 | 142.0469 | 5.1 | 0.36 |
| polarC18 | 218.139 | 74.6474 | 5.1 | 0.36 |
| polarC18 | 218.139 | 53.8352 | 5.1 | 0.36 |
| polarC18 | 218.139 | 56.7289 | 5.1 | 0.36 |
| polarC18 | 218.139 | 58.9884 | 5.1 | 0.36 |
| polarC18 | 218.139 | 51.1608 | 5.1 | 0.36 |
| polarC18 | 218.139 | 79.8659 | 5.1 | 0.36 |
| polarC18 | 218.139 | 225.198 | 5.1 | 0.36 |
| polarC18 | 218.139 | 83.0766 | 5.1 | 0.36 |
| polarC18 | 218.139 | 132.1296 | 5.1 | 0.36 |
| polarC18 | 218.139 | 49.7217 | 5.1 | 0.36 |
| polarC18 | 218.139 | 70.164 | 5.1 | 0.36 |
| polarC18 | 218.139 | 49.4293 | 5.1 | 0.36 |
| polarC18 | 218.139 | 227.5451 | 5.1 | 0.36 |
| polarC18 | 218.139 | 50.3114 | 5.1 | 0.36 |
| polarC18 | 218.139 | 145.7392 | 5.1 | 0.36 |
| polarC18 | 218.139 | 228.1718 | 5.1 | 0.36 |
| polarC18 | 218.139 | 153.8639 | 5.1 | 0.36 |
| polarC18 | 218.139 | 54.8738 | 5.1 | 0.36 |
| polarC18 | 218.139 | 82.1024 | 5 | 0.36 |
| polarC18 | 218.139 | 56.2174 | 5 | 0.36 |
| polarC18 | 218.139 | 88.4568 | 5 | 0.36 |
| polarC18 | 218.139 | 71.1003 | 5 | 0.36 |
| polarC18 | 218.139 | 178.4924 | 5 | 0.36 |
| polarC18 | 218.139 | 68.7612 | 5 | 0.36 |
| polarC18 | 218.139 | 79.0859 | 5 | 0.36 |
| polarC18 | 218.139 | 117.0746 | 5 | 0.36 |
| polarC18 | 218.139 | 201.0161 | 5 | 0.36 |
| polarC18 | 218.139 | 56.1613 | 5 | 0.36 |
| polarC18 | 218.139 | 153.0694 | 5 | 0.36 |
| polarC18 | 218.139 | 145.0843 | 5 | 0.36 |
| polarC18 | 218.139 | 85.3516 | 5 | 0.36 |
| polarC18 | 218.139 | 55.7584 | 5 | 0.36 |

| | | | | |
|---|---|---|---|---|
| polarC18 | 218.139 | 94.5123 | 5 | 0.36 |
| polarC18 | 218.139 | 126.4662 | 5 | 0.36 |
| polarC18 | 218.139 | 73.8304 | 5 | 0.36 |
| polarC18 | 218.139 | 60.5992 | 5 | 0.36 |
| polarC18 | 218.139 | 50.87 | 5 | 0.36 |
| polarC18 | 218.139 | 77.3247 | 5 | 0.36 |
| polarC18 | 218.139 | 130.2935 | 5 | 0.36 |
| polarC18 | 218.139 | 95.4548 | 5 | 0.36 |
| polarC18 | 218.139 | 60.4428 | 5 | 0.36 |
| polarC18 | 218.139 | 130.9995 | 5 | 0.36 |
| polarC18 | 218.139 | 56.9125 | 5 | 0.36 |
| polarC18 | 218.139 | 69.9283 | 5 | 0.36 |
| polarC18 | 218.139 | 97.2786 | 5 | 0.36 |
| polarC18 | 218.139 | 78.6627 | 5 | 0.36 |
| polarC18 | 218.139 | 100.9814 | 5 | 0.36 |
| polarC18 | 218.139 | 53.5649 | 5 | 0.36 |
| polarC18 | 218.139 | 112.7204 | 5 | 0.36 |
| polarC18 | 218.139 | 196.4625 | 5 | 0.36 |
| polarC18 | 218.139 | 98.6712 | 5 | 0.36 |
| polarC18 | 218.139 | 159.6533 | 5 | 0.36 |
| polarC18 | 218.139 | 49.3657 | 5 | 0.36 |
| polarC18 | 218.139 | 150.4235 | 5 | 0.36 |
| polarC18 | 218.139 | 109.9753 | 5 | 0.36 |
| polarC18 | 218.139 | 71.0077 | 5 | 0.36 |
| polarC18 | 218.139 | 70.5852 | 5 | 0.36 |
| polarC18 | 218.139 | 84.2849 | 5 | 0.36 |
| polarC18 | 218.139 | 96.9521 | 5 | 0.36 |
| polarC18 | 218.139 | 101.8902 | 5 | 0.35 |
| polarC18 | 218.139 | 74.7459 | 5 | 0.35 |
| polarC18 | 218.139 | 79.6576 | 5 | 0.35 |
| polarC18 | 218.139 | 117.8547 | 5 | 0.35 |
| polarC18 | 218.139 | 71.9677 | 5 | 0.35 |
| polarC18 | 218.139 | 71.5322 | 5 | 0.35 |
| polarC18 | 218.139 | 79.1128 | 5 | 0.35 |
| polarC18 | 218.139 | 50.9506 | 5 | 0.35 |
| polarC18 | 218.139 | 86.512 | 5 | 0.35 |
| polarC18 | 218.139 | 72.2818 | 5 | 0.35 |
| polarC18 | 218.139 | 124.5433 | 5 | 0.35 |
| polarC18 | 218.139 | 154.0882 | 5 | 0.35 |
| polarC18 | 218.139 | 141.8959 | 5 | 0.35 |
| polarC18 | 218.139 | 203.2305 | 5 | 0.35 |
| polarC18 | 218.139 | 50.6356 | 5 | 0.35 |

| | | | | |
|---|---|---|---|---|
| polarC18 | 218.139 | 107.7626 | 5 | 0.35 |
| polarC18 | 218.139 | 50.9293 | 5 | 0.35 |
| polarC18 | 218.139 | 74.525 | 5 | 0.35 |
| polarC18 | 218.139 | 64.2331 | 5 | 0.35 |
| polarC18 | 218.139 | 68.7572 | 5 | 0.35 |
| polarC18 | 218.139 | 226.5828 | 5 | 0.35 |
| polarC18 | 218.139 | 70.6224 | 5 | 0.35 |
| polarC18 | 218.139 | 69.742 | 5 | 0.35 |
| polarC18 | 218.139 | 106.7269 | 5 | 0.35 |
| polarC18 | 218.139 | 86.2804 | 5 | 0.35 |
| polarC18 | 218.139 | 49.3583 | 5 | 0.35 |
| polarC18 | 218.139 | 74.5685 | 5 | 0.35 |
| polarC18 | 218.139 | 85.6756 | 5 | 0.35 |
| polarC18 | 218.139 | 84.3119 | 5 | 0.35 |
| polarC18 | 218.139 | 92.9914 | 5 | 0.35 |
| polarC18 | 218.139 | 73.1616 | 5 | 0.35 |
| polarC18 | 218.139 | 181.2344 | 5 | 0.35 |
| polarC18 | 218.139 | 116.9427 | 5 | 0.35 |
| polarC18 | 218.139 | 49.8333 | 5 | 0.35 |
| polarC18 | 218.139 | 102.2642 | 5 | 0.35 |
| polarC18 | 218.139 | 169.2144 | 5 | 0.35 |
| polarC18 | 218.139 | 107.3648 | 5 | 0.35 |
| polarC18 | 218.139 | 92.4937 | 5 | 0.35 |
| polarC18 | 218.139 | 142.8306 | 5 | 0.35 |
| polarC18 | 218.139 | 51.8656 | 5 | 0.35 |
| polarC18 | 218.139 | 180.3131 | 5 | 0.35 |
| polarC18 | 218.139 | 170.1612 | 5 | 0.35 |
| polarC18 | 218.139 | 132.0208 | 5 | 0.35 |
| polarC18 | 218.139 | 51.7779 | 5 | 0.35 |
| polarC18 | 218.139 | 69.5009 | 5 | 0.35 |
| polarC18 | 218.139 | 57.7533 | 5 | 0.35 |
| polarC18 | 218.139 | 131.5791 | 5 | 0.35 |
| polarC18 | 218.139 | 51.9698 | 5 | 0.35 |
| polarC18 | 218.139 | 111.3439 | 5 | 0.35 |
| polarC18 | 218.139 | 95.1422 | 5 | 0.35 |
| polarC18 | 218.139 | 62.4932 | 5 | 0.35 |
| polarC18 | 218.139 | 78.8089 | 5 | 0.35 |
| polarC18 | 218.139 | 90.849 | 5 | 0.35 |
| polarC18 | 218.139 | 162.3681 | 5 | 0.35 |
| polarC18 | 218.139 | 60.9262 | 5 | 0.35 |
| polarC18 | 218.139 | 116.6547 | 5 | 0.35 |
| polarC18 | 218.139 | 235.0902 | 5 | 0.35 |

| | | | | |
|---|---|---|---|---|
| polarC18 | 218.139 | 54.3829 | 5 | 0.35 |
| polarC18 | 218.139 | 53.9896 | 5 | 0.35 |
| polarC18 | 218.139 | 60.8649 | 5 | 0.35 |
| polarC18 | 218.139 | 139.8276 | 5 | 0.35 |
| polarC18 | 218.139 | 91.8701 | 5 | 0.35 |
| polarC18 | 218.139 | 115.7022 | 5 | 0.35 |
| polarC18 | 218.139 | 95.5118 | 5 | 0.35 |
| polarC18 | 218.139 | 84.9341 | 5 | 0.35 |
| polarC18 | 218.139 | 57.5623 | 4.9 | 0.35 |
| polarC18 | 218.139 | 51.432 | 4.9 | 0.35 |
| polarC18 | 218.139 | 135.4964 | 4.9 | 0.35 |
| polarC18 | 218.139 | 195.7698 | 4.9 | 0.35 |
| polarC18 | 218.139 | 77.2612 | 4.9 | 0.35 |
| polarC18 | 218.139 | 124.5687 | 4.9 | 0.35 |
| polarC18 | 218.139 | 198.1326 | 4.9 | 0.35 |
| polarC18 | 218.139 | 155.634 | 4.9 | 0.35 |
| polarC18 | 218.139 | 52.8935 | 4.9 | 0.35 |
| polarC18 | 218.139 | 203.5197 | 4.9 | 0.35 |
| polarC18 | 218.139 | 69.8261 | 4.9 | 0.35 |
| polarC18 | 218.139 | 106.4216 | 4.9 | 0.35 |
| polarC18 | 218.139 | 141.9651 | 4.9 | 0.35 |
| polarC18 | 218.139 | 70.0197 | 4.9 | 0.35 |
| polarC18 | 218.139 | 155.9439 | 4.9 | 0.35 |
| polarC18 | 218.139 | 73.0011 | 4.9 | 0.35 |
| polarC18 | 218.139 | 93.0829 | 4.9 | 0.35 |
| polarC18 | 218.139 | 212.0675 | 4.9 | 0.35 |
| polarC18 | 218.139 | 179.2439 | 4.9 | 0.35 |
| polarC18 | 218.139 | 230.6499 | 4.9 | 0.35 |
| polarC18 | 218.139 | 67.1851 | 4.9 | 0.35 |
| polarC18 | 218.139 | 60.1151 | 4.9 | 0.35 |
| polarC18 | 218.139 | 64.2399 | 4.9 | 0.35 |
| polarC18 | 218.139 | 59.3162 | 4.9 | 0.35 |
| polarC18 | 218.139 | 183.522 | 4.9 | 0.35 |
| polarC18 | 218.139 | 59.8702 | 4.9 | 0.35 |
| polarC18 | 218.139 | 219.3151 | 4.9 | 0.35 |
| polarC18 | 218.139 | 147.299 | 4.9 | 0.35 |
| polarC18 | 218.139 | 242.4682 | 4.9 | 0.35 |
| polarC18 | 218.139 | 61.6096 | 4.9 | 0.35 |
| polarC18 | 218.139 | 144.1555 | 4.9 | 0.35 |
| polarC18 | 218.139 | 57.84 | 4.9 | 0.35 |
| polarC18 | 218.139 | 175.9064 | 4.9 | 0.35 |
| polarC18 | 218.139 | 52.6402 | 4.9 | 0.35 |

| | | | | |
|---|---|---|---|---|
| polarC18 | 218.139 | 109.9455 | 4.9 | 0.35 |
| polarC18 | 218.139 | 76.6147 | 4.9 | 0.35 |
| polarC18 | 218.139 | 227.7511 | 4.9 | 0.35 |
| polarC18 | 218.139 | 120.3812 | 4.9 | 0.35 |
| polarC18 | 218.139 | 104.3257 | 4.9 | 0.35 |
| polarC18 | 218.139 | 65.475 | 4.9 | 0.35 |
| polarC18 | 218.139 | 60.8456 | 4.9 | 0.35 |
| polarC18 | 218.139 | 49.2598 | 4.9 | 0.35 |
| polarC18 | 218.139 | 54.4377 | 4.9 | 0.35 |
| polarC18 | 218.139 | 56.3395 | 4.9 | 0.35 |
| polarC18 | 218.139 | 54.0559 | 4.9 | 0.35 |
| polarC18 | 218.139 | 73.5349 | 4.9 | 0.35 |
| polarC18 | 218.139 | 50.4528 | 4.9 | 0.35 |
| polarC18 | 218.139 | 60.7091 | 4.9 | 0.35 |
| polarC18 | 218.139 | 61.254 | 4.9 | 0.35 |
| polarC18 | 218.139 | 64.6936 | 4.9 | 0.35 |
| polarC18 | 218.139 | 48.7189 | 4.9 | 0.35 |
| polarC18 | 218.139 | 51.259 | 4.9 | 0.35 |
| polarC18 | 218.139 | 149.5522 | 4.9 | 0.35 |
| polarC18 | 218.139 | 56.3078 | 4.9 | 0.35 |
| polarC18 | 218.139 | 239.2155 | 4.9 | 0.35 |
| polarC18 | 218.139 | 127.0551 | 4.9 | 0.35 |
| polarC18 | 218.139 | 59.4684 | 4.9 | 0.35 |
| polarC18 | 218.139 | 140.5409 | 4.9 | 0.35 |
| polarC18 | 218.139 | 164.7721 | 4.9 | 0.35 |
| polarC18 | 218.139 | 86.7692 | 4.9 | 0.35 |
| polarC18 | 218.139 | 53.0406 | 4.9 | 0.35 |
| polarC18 | 218.139 | 49.2087 | 4.9 | 0.35 |
| polarC18 | 218.139 | 141.5762 | 4.9 | 0.35 |
| polarC18 | 218.139 | 87.2402 | 4.9 | 0.35 |
| polarC18 | 218.139 | 102.2118 | 4.9 | 0.35 |
| polarC18 | 218.139 | 211.3845 | 4.9 | 0.35 |
| polarC18 | 218.139 | 70.7127 | 4.9 | 0.35 |
| polarC18 | 218.139 | 138.9853 | 4.9 | 0.35 |
| polarC18 | 218.139 | 68.7666 | 4.9 | 0.35 |
| polarC18 | 218.139 | 59.359 | 4.9 | 0.35 |
| polarC18 | 218.139 | 82.846 | 4.9 | 0.35 |
| polarC18 | 218.139 | 118.2702 | 4.9 | 0.35 |
| polarC18 | 218.139 | 203.3592 | 4.9 | 0.35 |
| polarC18 | 218.139 | 52.6952 | 4.9 | 0.35 |
| polarC18 | 218.139 | 124.762 | 4.9 | 0.35 |
| polarC18 | 218.139 | 48.6324 | 4.9 | 0.35 |

| | | | | |
|---|---|---|---|---|
| polarC18 | 218.139 | 53.1895 | 4.9 | 0.35 |
| polarC18 | 218.139 | 77.2956 | 4.9 | 0.35 |
| polarC18 | 218.139 | 212.1052 | 4.9 | 0.35 |
| polarC18 | 218.139 | 233.1898 | 4.9 | 0.35 |
| polarC18 | 218.139 | 50.0282 | 4.9 | 0.35 |
| polarC18 | 218.139 | 67.01 | 4.9 | 0.35 |
| polarC18 | 218.139 | 50.7922 | 4.9 | 0.35 |
| polarC18 | 218.139 | 49.4801 | 4.9 | 0.35 |
| polarC18 | 218.139 | 119.6562 | 4.9 | 0.35 |
| polarC18 | 218.139 | 95.4426 | 4.9 | 0.35 |
| polarC18 | 218.139 | 52.3998 | 4.9 | 0.35 |
| polarC18 | 218.139 | 134.1566 | 4.9 | 0.35 |
| polarC18 | 218.139 | 84.247 | 4.9 | 0.35 |
| polarC18 | 218.139 | 152.904 | 4.9 | 0.35 |
| polarC18 | 218.139 | 51.3609 | 4.9 | 0.35 |
| polarC18 | 218.139 | 60.1256 | 4.9 | 0.35 |
| polarC18 | 218.139 | 84.2512 | 4.9 | 0.35 |
| polarC18 | 218.139 | 77.3795 | 4.9 | 0.35 |
| polarC18 | 218.139 | 74.5166 | 4.9 | 0.35 |
| polarC18 | 218.139 | 73.2376 | 4.9 | 0.35 |
| polarC18 | 218.139 | 220.203 | 4.9 | 0.35 |
| polarC18 | 218.139 | 78.049 | 4.9 | 0.35 |
| polarC18 | 218.139 | 51.5102 | 4.9 | 0.35 |
| polarC18 | 218.139 | 63.4973 | 4.9 | 0.35 |
| polarC18 | 218.139 | 222.7592 | 4.9 | 0.34 |
| polarC18 | 218.139 | 67.8753 | 4.9 | 0.34 |
| polarC18 | 218.139 | 64.8606 | 4.9 | 0.34 |
| polarC18 | 218.139 | 78.786 | 4.9 | 0.34 |
| polarC18 | 218.139 | 93.4135 | 4.9 | 0.34 |
| polarC18 | 218.139 | 104.2491 | 4.9 | 0.34 |
| polarC18 | 218.139 | 49.8829 | 4.9 | 0.34 |
| polarC18 | 218.139 | 70.78 | 4.9 | 0.34 |
| polarC18 | 218.139 | 50.3423 | 4.9 | 0.34 |
| polarC18 | 218.139 | 68.9451 | 4.9 | 0.34 |
| polarC18 | 218.139 | 82.8332 | 4.9 | 0.34 |
| polarC18 | 218.139 | 63.8148 | 4.9 | 0.34 |
| polarC18 | 218.139 | 242.0888 | 4.8 | 0.34 |
| polarC18 | 218.139 | 57.6841 | 4.8 | 0.34 |
| polarC18 | 218.139 | 158.6338 | 4.8 | 0.34 |
| polarC18 | 218.139 | 74.8203 | 4.8 | 0.34 |
| polarC18 | 218.139 | 208.1458 | 4.8 | 0.34 |
| polarC18 | 218.139 | 77.0966 | 4.8 | 0.34 |

| | | | | |
|---|---|---|---|---|
| polarC18 | 218.139 | 221.3173 | 4.8 | 0.34 |
| polarC18 | 218.139 | 55.3146 | 4.8 | 0.34 |
| polarC18 | 218.139 | 59.5257 | 4.8 | 0.34 |
| polarC18 | 218.139 | 67.8531 | 4.8 | 0.34 |
| polarC18 | 218.139 | 80.0397 | 4.8 | 0.34 |
| polarC18 | 218.139 | 71.9289 | 4.8 | 0.34 |
| polarC18 | 218.139 | 151.5533 | 4.8 | 0.34 |
| polarC18 | 218.139 | 53.4363 | 4.8 | 0.34 |
| polarC18 | 218.139 | 64.3066 | 4.8 | 0.34 |
| polarC18 | 218.139 | 74.4308 | 4.8 | 0.34 |
| polarC18 | 218.139 | 186.9215 | 4.8 | 0.34 |
| polarC18 | 218.139 | 49.308 | 4.8 | 0.34 |
| polarC18 | 218.139 | 101.0451 | 4.8 | 0.34 |
| polarC18 | 218.139 | 69.9721 | 4.8 | 0.34 |
| polarC18 | 218.139 | 51.4951 | 4.8 | 0.34 |
| polarC18 | 218.139 | 69.4265 | 4.8 | 0.34 |
| polarC18 | 218.139 | 53.5758 | 4.8 | 0.34 |
| polarC18 | 218.139 | 103.0404 | 4.8 | 0.34 |
| polarC18 | 218.139 | 183.4154 | 4.8 | 0.34 |
| polarC18 | 218.139 | 55.0622 | 4.8 | 0.34 |
| polarC18 | 218.139 | 117.3507 | 4.8 | 0.34 |
| polarC18 | 218.139 | 128.664 | 4.8 | 0.34 |
| polarC18 | 218.139 | 145.4588 | 4.8 | 0.34 |
| polarC18 | 218.139 | 190.6605 | 4.8 | 0.34 |
| polarC18 | 218.139 | 117.0874 | 4.8 | 0.34 |
| polarC18 | 218.139 | 53.9021 | 4.8 | 0.34 |
| polarC18 | 218.139 | 56.1244 | 4.8 | 0.34 |
| polarC18 | 218.139 | 95.6806 | 4.8 | 0.34 |
| polarC18 | 218.139 | 84.5062 | 4.8 | 0.34 |
| polarC18 | 218.139 | 117.8312 | 4.8 | 0.34 |
| polarC18 | 218.139 | 53.9763 | 4.8 | 0.34 |
| polarC18 | 218.139 | 65.7207 | 4.8 | 0.34 |
| polarC18 | 218.139 | 188.6356 | 4.8 | 0.34 |
| polarC18 | 218.139 | 184.2714 | 4.8 | 0.34 |
| polarC18 | 218.139 | 81.1542 | 4.8 | 0.34 |
| polarC18 | 218.139 | 130.0808 | 4.8 | 0.34 |
| polarC18 | 218.139 | 170.5085 | 4.8 | 0.34 |
| polarC18 | 218.139 | 49.0748 | 4.8 | 0.34 |
| polarC18 | 218.139 | 145.3195 | 4.8 | 0.34 |
| polarC18 | 218.139 | 91.8372 | 4.8 | 0.34 |
| polarC18 | 218.139 | 107.4597 | 4.8 | 0.34 |
| polarC18 | 218.139 | 64.4493 | 4.8 | 0.34 |

| | | | | |
|---|---|---|---|---|
| polarC18 | 218.139 | 96.4624 | 4.8 | 0.34 |
| polarC18 | 218.139 | 106.2254 | 4.8 | 0.34 |
| polarC18 | 218.139 | 59.5425 | 4.8 | 0.34 |
| polarC18 | 218.139 | 57.9082 | 4.8 | 0.34 |
| polarC18 | 218.139 | 127.1483 | 4.8 | 0.34 |
| polarC18 | 218.139 | 70.2257 | 4.8 | 0.34 |
| polarC18 | 218.139 | 229.6449 | 4.8 | 0.34 |
| polarC18 | 218.139 | 126.9574 | 4.8 | 0.34 |
| polarC18 | 218.139 | 204.2478 | 4.8 | 0.34 |
| polarC18 | 218.139 | 139.4496 | 4.8 | 0.34 |
| polarC18 | 218.139 | 76.0106 | 4.8 | 0.34 |
| polarC18 | 218.139 | 56.3544 | 4.8 | 0.34 |
| polarC18 | 218.139 | 70.8063 | 4.8 | 0.34 |
| polarC18 | 218.139 | 179.201 | 4.8 | 0.34 |
| polarC18 | 218.139 | 87.2349 | 4.7 | 0.34 |
| polarC18 | 218.139 | 79.4865 | 4.7 | 0.34 |
| polarC18 | 218.139 | 98.353 | 4.7 | 0.34 |
| polarC18 | 218.139 | 49.3547 | 4.7 | 0.34 |
| polarC18 | 218.139 | 117.5263 | 4.7 | 0.34 |
| polarC18 | 218.139 | 101.4203 | 4.7 | 0.34 |
| polarC18 | 218.139 | 80.4333 | 4.7 | 0.34 |
| polarC18 | 218.139 | 102.7065 | 4.7 | 0.34 |
| polarC18 | 218.139 | 180.0196 | 4.7 | 0.34 |
| polarC18 | 218.139 | 60.058 | 4.7 | 0.34 |
| polarC18 | 218.139 | 198.5594 | 4.7 | 0.34 |
| polarC18 | 218.139 | 128.9094 | 4.7 | 0.33 |
| polarC18 | 218.139 | 67.2367 | 4.7 | 0.33 |
| polarC18 | 218.139 | 76.6357 | 4.7 | 0.33 |
| polarC18 | 218.139 | 79.559 | 4.7 | 0.33 |
| polarC18 | 218.139 | 50.6266 | 4.7 | 0.33 |
| polarC18 | 218.139 | 70.5303 | 4.7 | 0.33 |
| polarC18 | 218.139 | 208.6772 | 4.7 | 0.33 |
| polarC18 | 218.139 | 171.7043 | 4.7 | 0.33 |
| polarC18 | 218.139 | 64.9534 | 4.7 | 0.33 |
| polarC18 | 218.139 | 163.8661 | 4.7 | 0.33 |
| polarC18 | 218.139 | 109.7216 | 4.7 | 0.33 |
| polarC18 | 218.139 | 55.3092 | 4.7 | 0.33 |
| polarC18 | 218.139 | 109.244 | 4.7 | 0.33 |
| polarC18 | 218.139 | 57.0633 | 4.7 | 0.33 |
| polarC18 | 218.139 | 98.217 | 4.7 | 0.33 |
| polarC18 | 218.139 | 232.5458 | 4.7 | 0.33 |
| polarC18 | 218.139 | 104.9995 | 4.7 | 0.33 |

| | | | | |
|---|---|---|---|---|
| polarC18 | 218.139 | 117.2881 | 4.7 | 0.33 |
| polarC18 | 218.139 | 91.0022 | 4.7 | 0.33 |
| polarC18 | 218.139 | 130.1236 | 4.7 | 0.33 |
| polarC18 | 218.139 | 56.3929 | 4.7 | 0.33 |
| polarC18 | 218.139 | 66.9411 | 4.7 | 0.33 |
| polarC18 | 218.139 | 75.8486 | 4.7 | 0.33 |
| polarC18 | 218.139 | 88.1043 | 4.7 | 0.33 |
| polarC18 | 218.139 | 196.5052 | 4.7 | 0.33 |
| polarC18 | 218.139 | 147.6283 | 4.7 | 0.33 |
| polarC18 | 218.139 | 223.4717 | 4.7 | 0.33 |
| polarC18 | 218.139 | 72.0154 | 4.7 | 0.33 |
| polarC18 | 218.139 | 59.8251 | 4.7 | 0.33 |
| polarC18 | 218.139 | 68.3197 | 4.7 | 0.33 |
| polarC18 | 218.139 | 53.3599 | 4.7 | 0.33 |
| polarC18 | 218.139 | 58.3336 | 4.7 | 0.33 |
| polarC18 | 218.139 | 120.0056 | 4.7 | 0.33 |
| polarC18 | 218.139 | 55.3003 | 4.7 | 0.33 |
| polarC18 | 218.139 | 50.9049 | 4.7 | 0.33 |
| polarC18 | 218.139 | 207.5957 | 4.7 | 0.33 |
| polarC18 | 218.139 | 131.5609 | 4.7 | 0.33 |
| polarC18 | 218.139 | 87.5745 | 4.7 | 0.33 |
| polarC18 | 218.139 | 73.2964 | 4.7 | 0.33 |
| polarC18 | 218.139 | 49.0162 | 4.7 | 0.33 |
| polarC18 | 218.139 | 79.4618 | 4.6 | 0.33 |
| polarC18 | 218.139 | 165.1711 | 4.6 | 0.33 |
| polarC18 | 218.139 | 105.2414 | 4.6 | 0.33 |
| polarC18 | 218.139 | 91.4647 | 4.6 | 0.33 |
| polarC18 | 218.139 | 50.9223 | 4.6 | 0.33 |
| polarC18 | 218.139 | 57.8026 | 4.6 | 0.33 |
| polarC18 | 218.139 | 81.9836 | 4.6 | 0.33 |
| polarC18 | 218.139 | 67.978 | 4.6 | 0.33 |
| polarC18 | 218.139 | 63.6129 | 4.6 | 0.33 |
| polarC18 | 218.139 | 79.3837 | 4.6 | 0.33 |
| polarC18 | 218.139 | 50.5117 | 4.6 | 0.33 |
| polarC18 | 218.139 | 87.7243 | 4.6 | 0.33 |
| polarC18 | 218.139 | 62.508 | 4.6 | 0.33 |
| polarC18 | 218.139 | 160.4339 | 4.6 | 0.33 |
| polarC18 | 218.139 | 101.4819 | 4.6 | 0.33 |
| polarC18 | 218.139 | 59.3996 | 4.6 | 0.33 |
| polarC18 | 218.139 | 57.0142 | 4.6 | 0.33 |
| polarC18 | 218.139 | 50.9114 | 4.6 | 0.33 |
| polarC18 | 218.139 | 205.4429 | 4.6 | 0.33 |

| | | | | |
|---|---|---|---|---|
| polarC18 | 218.139 | 70.2158 | 4.6 | 0.33 |
| polarC18 | 218.139 | 153.0402 | 4.6 | 0.33 |
| polarC18 | 218.139 | 113.8232 | 4.6 | 0.33 |
| polarC18 | 218.139 | 135.2619 | 4.6 | 0.33 |
| polarC18 | 218.139 | 48.7929 | 4.6 | 0.33 |
| polarC18 | 218.139 | 57.3547 | 4.6 | 0.33 |
| polarC18 | 218.139 | 58.1875 | 4.6 | 0.33 |
| polarC18 | 218.139 | 54.44 | 4.6 | 0.33 |
| polarC18 | 218.139 | 56.3781 | 4.6 | 0.33 |
| polarC18 | 218.139 | 50.1481 | 4.6 | 0.33 |
| polarC18 | 218.139 | 137.8924 | 4.6 | 0.33 |
| polarC18 | 218.139 | 104.0069 | 4.6 | 0.33 |
| polarC18 | 218.139 | 90.0425 | 4.6 | 0.33 |
| polarC18 | 218.139 | 53.7466 | 4.6 | 0.32 |
| polarC18 | 218.139 | 60.2383 | 4.6 | 0.32 |
| polarC18 | 218.139 | 60.3851 | 4.6 | 0.32 |
| polarC18 | 218.139 | 207.9503 | 4.6 | 0.32 |
| polarC18 | 218.139 | 50.5617 | 4.6 | 0.32 |
| polarC18 | 218.139 | 116.2218 | 4.6 | 0.32 |
| polarC18 | 218.139 | 128.4557 | 4.6 | 0.32 |
| polarC18 | 218.139 | 75.3136 | 4.6 | 0.32 |
| polarC18 | 218.139 | 126.3139 | 4.6 | 0.32 |
| polarC18 | 218.139 | 82.8941 | 4.6 | 0.32 |
| polarC18 | 218.139 | 54.9172 | 4.6 | 0.32 |
| polarC18 | 218.139 | 105.3296 | 4.6 | 0.32 |
| polarC18 | 218.139 | 98.2627 | 4.6 | 0.32 |
| polarC18 | 218.139 | 153.5634 | 4.5 | 0.32 |
| polarC18 | 218.139 | 54.1538 | 4.5 | 0.32 |
| polarC18 | 218.139 | 68.0561 | 4.5 | 0.32 |
| polarC18 | 218.139 | 69.0639 | 4.5 | 0.32 |
| polarC18 | 218.139 | 137.8809 | 4.5 | 0.32 |
| polarC18 | 218.139 | 132.0095 | 4.5 | 0.32 |
| polarC18 | 218.139 | 57.2558 | 4.5 | 0.32 |
| polarC18 | 218.139 | 176.395 | 4.5 | 0.32 |
| polarC18 | 218.139 | 179.2697 | 4.5 | 0.32 |
| polarC18 | 218.139 | 109.658 | 4.5 | 0.32 |
| polarC18 | 218.139 | 69.7764 | 4.5 | 0.32 |
| polarC18 | 218.139 | 75.5554 | 4.5 | 0.32 |
| polarC18 | 218.139 | 52.3583 | 4.5 | 0.32 |
| polarC18 | 218.139 | 71.1501 | 4.5 | 0.32 |
| polarC18 | 218.139 | 163.5174 | 4.5 | 0.32 |
| polarC18 | 218.139 | 53.2683 | 4.5 | 0.32 |

| | | | | |
|---|---|---|---|---|
| polarC18 | 218.139 | 89.6187 | 4.5 | 0.32 |
| polarC18 | 218.139 | 106.7931 | 4.5 | 0.32 |
| polarC18 | 218.139 | 67.0668 | 4.5 | 0.32 |
| polarC18 | 218.139 | 112.2161 | 4.5 | 0.32 |
| polarC18 | 218.139 | 220.969 | 4.5 | 0.32 |
| polarC18 | 218.139 | 124.9549 | 4.5 | 0.32 |
| polarC18 | 218.139 | 56.2113 | 4.5 | 0.32 |
| polarC18 | 218.139 | 68.899 | 4.5 | 0.32 |
| polarC18 | 218.139 | 183.3826 | 4.5 | 0.32 |
| polarC18 | 218.139 | 95.122 | 4.4 | 0.32 |
| polarC18 | 218.139 | 97.4851 | 4.4 | 0.31 |
| polarC18 | 218.139 | 169.0162 | 4.4 | 0.31 |
| polarC18 | 218.139 | 77.1904 | 4.4 | 0.31 |
| polarC18 | 218.139 | 215.251 | 4.4 | 0.31 |
| polarC18 | 218.139 | 56.0991 | 4.4 | 0.31 |
| polarC18 | 218.139 | 99.2299 | 4.4 | 0.31 |
| polarC18 | 218.139 | 77.1669 | 4.4 | 0.31 |
| polarC18 | 218.139 | 51.1319 | 4.4 | 0.31 |
| polarC18 | 218.139 | 68.9965 | 4.4 | 0.31 |
| polarC18 | 218.139 | 116.3257 | 4.4 | 0.31 |
| polarC18 | 218.139 | 62.8696 | 4.3 | 0.31 |
| polarC18 | 218.139 | 118.9432 | 4.3 | 0.31 |
| polarC18 | 218.139 | 59.6086 | 4.3 | 0.31 |
| polarC18 | 218.139 | 80.7135 | 4.3 | 0.3 |
| polarC18 | 218.139 | 66.036 | 4.3 | 0.3 |
| polarC18 | 218.139 | 50.8617 | 4.2 | 0.3 |
| polarC18 | 218.139 | 59.0277 | 4.2 | 0.3 |
| polarC18 | 276.144 | 252.8727 | 5200.5 | 100 |
| polarC18 | 276.144 | 208.8831 | 3869.1 | 74.4 |
| polarC18 | 276.144 | 190.8725 | 742.3 | 14.27 |
| polarC18 | 276.144 | 276.2323 | 275.4 | 5.29 |
| polarC18 | 276.144 | 174.9692 | 247.5 | 4.76 |
| polarC18 | 276.144 | 130.0499 | 189.6 | 3.65 |
| polarC18 | 276.144 | 276.1359 | 172.1 | 3.31 |
| polarC18 | 276.144 | 84.0443 | 160.3 | 3.08 |
| polarC18 | 276.144 | 116.0706 | 151.4 | 2.91 |
| polarC18 | 276.144 | 147.0764 | 145.1 | 2.79 |
| polarC18 | 276.144 | 99.9869 | 129.6 | 2.49 |
| polarC18 | 276.144 | 226.8959 | 125.7 | 2.42 |
| polarC18 | 276.144 | 180.8882 | 120.2 | 2.31 |
| polarC18 | 276.144 | 228.2173 | 111.1 | 2.14 |
| polarC18 | 276.144 | 155.9505 | 95.7 | 1.84 |

| | | | | |
|---|---|---|---|---|
| polarC18 | 276.144 | 179.8791 | 84.5 | 1.62 |
| polarC18 | 276.144 | 132.9589 | 82.5 | 1.59 |
| polarC18 | 276.144 | 143.9508 | 69.9 | 1.34 |
| polarC18 | 276.144 | 161.093 | 68.4 | 1.32 |
| polarC18 | 276.144 | 197.8912 | 67.6 | 1.3 |
| polarC18 | 276.144 | 144.9602 | 51.4 | 0.99 |
| polarC18 | 276.144 | 275.9058 | 45.5 | 0.87 |
| polarC18 | 276.144 | 241.4507 | 42.5 | 0.82 |
| polarC18 | 276.144 | 173.961 | 39.5 | 0.76 |
| polarC18 | 276.144 | 196.8603 | 38.4 | 0.74 |
| polarC18 | 276.144 | 70.065 | 37.5 | 0.72 |
| polarC18 | 276.144 | 93.49 | 37.3 | 0.72 |
| polarC18 | 276.144 | 259.1985 | 36.7 | 0.71 |
| polarC18 | 276.144 | 93.9849 | 36 | 0.69 |
| polarC18 | 276.144 | 276.8965 | 35.7 | 0.69 |
| polarC18 | 276.144 | 242.2095 | 35.6 | 0.68 |
| polarC18 | 276.144 | 125.0795 | 35.3 | 0.68 |
| polarC18 | 276.144 | 234.9946 | 35.1 | 0.67 |
| polarC18 | 276.144 | 266.5792 | 35 | 0.67 |
| polarC18 | 276.144 | 295.1317 | 34.6 | 0.67 |
| polarC18 | 276.144 | 93.9249 | 34.5 | 0.66 |
| polarC18 | 276.144 | 85.0283 | 34.2 | 0.66 |
| polarC18 | 276.144 | 148.7131 | 33.9 | 0.65 |
| polarC18 | 276.144 | 276.2896 | 33.9 | 0.65 |
| polarC18 | 276.144 | 153.3967 | 33.9 | 0.65 |
| polarC18 | 276.144 | 199.5234 | 33.8 | 0.65 |
| polarC18 | 276.144 | 69.3173 | 33 | 0.64 |
| polarC18 | 276.144 | 148.8901 | 33 | 0.63 |
| polarC18 | 276.144 | 253.8756 | 32.6 | 0.63 |
| polarC18 | 276.144 | 58.9604 | 32.6 | 0.63 |
| polarC18 | 276.144 | 142.6787 | 32.5 | 0.63 |
| polarC18 | 276.144 | 220.1981 | 32.1 | 0.62 |
| polarC18 | 276.144 | 94.0097 | 32.1 | 0.62 |
| polarC18 | 276.144 | 270.8673 | 31.3 | 0.6 |
| polarC18 | 276.144 | 93.4197 | 31 | 0.6 |
| polarC18 | 276.144 | 133.3932 | 30.5 | 0.59 |
| polarC18 | 276.144 | 205.4331 | 30 | 0.58 |
| polarC18 | 276.144 | 147.4249 | 29.5 | 0.57 |
| polarC18 | 276.144 | 94.0519 | 29.4 | 0.57 |
| polarC18 | 276.144 | 167.4473 | 29.4 | 0.57 |
| polarC18 | 276.144 | 257.7515 | 29.2 | 0.56 |
| polarC18 | 276.144 | 94.0586 | 29.2 | 0.56 |

| | | | | |
|---|---|---|---|---|
| polarC18 | 276.144 | 258.1738 | 28.9 | 0.56 |
| polarC18 | 276.144 | 107.6054 | 28.9 | 0.56 |
| polarC18 | 276.144 | 77.004 | 28.9 | 0.55 |
| polarC18 | 276.144 | 264.4519 | 28.7 | 0.55 |
| polarC18 | 276.144 | 267.616 | 28.6 | 0.55 |
| polarC18 | 276.144 | 95.5115 | 28.6 | 0.55 |
| polarC18 | 276.144 | 288.7415 | 28.4 | 0.55 |
| polarC18 | 276.144 | 77.4167 | 28.2 | 0.54 |
| polarC18 | 276.144 | 237.8785 | 28.2 | 0.54 |
| polarC18 | 276.144 | 159.2767 | 28 | 0.54 |
| polarC18 | 276.144 | 96.526 | 28 | 0.54 |
| polarC18 | 276.144 | 79.7019 | 27.9 | 0.54 |
| polarC18 | 276.144 | 231.388 | 27.9 | 0.54 |
| polarC18 | 276.144 | 116.1446 | 27.9 | 0.54 |
| polarC18 | 276.144 | 234.8651 | 27.8 | 0.54 |
| polarC18 | 276.144 | 128.5981 | 27.8 | 0.53 |
| polarC18 | 276.144 | 130.6043 | 27.5 | 0.53 |
| polarC18 | 276.144 | 174.5485 | 27.5 | 0.53 |
| polarC18 | 276.144 | 243.6265 | 27.5 | 0.53 |
| polarC18 | 276.144 | 300.2434 | 27.3 | 0.52 |
| polarC18 | 276.144 | 170.487 | 27.2 | 0.52 |
| polarC18 | 276.144 | 261.5875 | 27.2 | 0.52 |
| polarC18 | 276.144 | 93.9748 | 27.1 | 0.52 |
| polarC18 | 276.144 | 298.2146 | 27.1 | 0.52 |
| polarC18 | 276.144 | 164.2538 | 27.1 | 0.52 |
| polarC18 | 276.144 | 111.8037 | 26.9 | 0.52 |
| polarC18 | 276.144 | 269.3732 | 26.9 | 0.52 |
| polarC18 | 276.144 | 130.0261 | 26.9 | 0.52 |
| polarC18 | 276.144 | 51.5623 | 26.8 | 0.52 |
| polarC18 | 276.144 | 56.0372 | 26.8 | 0.52 |
| polarC18 | 276.144 | 133.4847 | 26.8 | 0.51 |
| polarC18 | 276.144 | 184.0871 | 26.8 | 0.51 |
| polarC18 | 276.144 | 89.2466 | 26.8 | 0.51 |
| polarC18 | 276.144 | 267.7271 | 26.7 | 0.51 |
| polarC18 | 276.144 | 301.5271 | 26.7 | 0.51 |
| polarC18 | 276.144 | 51.5474 | 26.7 | 0.51 |
| polarC18 | 276.144 | 121.5453 | 26.7 | 0.51 |
| polarC18 | 276.144 | 61.2263 | 26.6 | 0.51 |
| polarC18 | 276.144 | 79.0882 | 26.5 | 0.51 |
| polarC18 | 276.144 | 301.7569 | 26.5 | 0.51 |
| polarC18 | 276.144 | 99.0649 | 26.5 | 0.51 |
| polarC18 | 276.144 | 131.0467 | 26.5 | 0.51 |

| | | | | |
|---|---|---|---|---|
| polarC18 | 276.144 | 79.6071 | 26.4 | 0.51 |
| polarC18 | 276.144 | 67.3847 | 26.4 | 0.51 |
| polarC18 | 276.144 | 269.4531 | 26.4 | 0.51 |
| polarC18 | 276.144 | 106.0318 | 26.3 | 0.51 |
| polarC18 | 276.144 | 66.1204 | 26.3 | 0.51 |
| polarC18 | 276.144 | 226.7887 | 26.3 | 0.51 |
| polarC18 | 276.144 | 56.948 | 26.3 | 0.51 |
| polarC18 | 276.144 | 174.3541 | 26.1 | 0.5 |
| polarC18 | 276.144 | 80.4768 | 26.1 | 0.5 |
| polarC18 | 276.144 | 110.0235 | 25.9 | 0.5 |
| polarC18 | 276.144 | 257.5741 | 25.8 | 0.5 |
| polarC18 | 276.144 | 51.5675 | 25.7 | 0.49 |
| polarC18 | 276.144 | 118.1964 | 25.6 | 0.49 |
| polarC18 | 276.144 | 56.6174 | 25.5 | 0.49 |
| polarC18 | 276.144 | 301.1813 | 25.5 | 0.49 |
| polarC18 | 276.144 | 140.2766 | 25.5 | 0.49 |
| polarC18 | 276.144 | 203.5369 | 25.4 | 0.49 |
| polarC18 | 276.144 | 91.1411 | 25.4 | 0.49 |
| polarC18 | 276.144 | 74.1801 | 25.4 | 0.49 |
| polarC18 | 276.144 | 202.7538 | 25.4 | 0.49 |
| polarC18 | 276.144 | 152.741 | 25.3 | 0.49 |
| polarC18 | 276.144 | 67.2738 | 25.3 | 0.49 |
| polarC18 | 276.144 | 297.9363 | 25.3 | 0.49 |
| polarC18 | 276.144 | 139.2562 | 25.2 | 0.48 |
| polarC18 | 276.144 | 216.4268 | 25.1 | 0.48 |
| polarC18 | 276.144 | 139.3243 | 25 | 0.48 |
| polarC18 | 276.144 | 127.8054 | 24.9 | 0.48 |
| polarC18 | 276.144 | 279.559 | 24.9 | 0.48 |
| polarC18 | 276.144 | 127.8493 | 24.8 | 0.48 |
| polarC18 | 276.144 | 161.5632 | 24.8 | 0.48 |
| polarC18 | 276.144 | 56.9671 | 24.7 | 0.47 |
| polarC18 | 276.144 | 140.859 | 24.7 | 0.47 |
| polarC18 | 276.144 | 229.8423 | 24.7 | 0.47 |
| polarC18 | 276.144 | 54.9658 | 24.6 | 0.47 |
| polarC18 | 276.144 | 208.3011 | 24.6 | 0.47 |
| polarC18 | 276.144 | 127.3193 | 24.5 | 0.47 |
| polarC18 | 276.144 | 102.3133 | 24.4 | 0.47 |
| polarC18 | 276.144 | 298.5244 | 24.3 | 0.47 |
| polarC18 | 276.144 | 236.1143 | 24.3 | 0.47 |
| polarC18 | 276.144 | 225.2477 | 24.2 | 0.47 |
| polarC18 | 276.144 | 94.6372 | 24.1 | 0.46 |
| polarC18 | 276.144 | 121.4981 | 23.9 | 0.46 |

| | | | | |
|---|---|---|---|---|
| polarC18 | 276.144 | 74.49 | 23.8 | 0.46 |
| polarC18 | 276.144 | 199.6607 | 23.5 | 0.45 |
| polarC18 | 276.144 | 84.5939 | 23.5 | 0.45 |
| polarC18 | 276.144 | 59.1905 | 23.4 | 0.45 |
| polarC18 | 276.144 | 290.1332 | 23.3 | 0.45 |
| polarC18 | 276.144 | 140.3716 | 23.1 | 0.44 |
| polarC18 | 276.144 | 93.9507 | 22.9 | 0.44 |
| polarC18 | 276.144 | 51.56 | 22.9 | 0.44 |
| polarC18 | 276.144 | 224.6933 | 22.8 | 0.44 |
| polarC18 | 276.144 | 56.3093 | 22.8 | 0.44 |
| polarC18 | 276.144 | 139.2268 | 22.8 | 0.44 |
| polarC18 | 276.144 | 93.9605 | 22.7 | 0.44 |
| polarC18 | 276.144 | 83.6173 | 22.7 | 0.44 |
| polarC18 | 276.144 | 105.614 | 22.7 | 0.44 |
| polarC18 | 276.144 | 74.8002 | 22.6 | 0.43 |
| polarC18 | 276.144 | 94.114 | 22.6 | 0.43 |
| polarC18 | 276.144 | 94.0365 | 22.5 | 0.43 |
| polarC18 | 276.144 | 176.1017 | 22.5 | 0.43 |
| polarC18 | 276.144 | 195.4437 | 22.5 | 0.43 |
| polarC18 | 276.144 | 93.8759 | 22.4 | 0.43 |
| polarC18 | 276.144 | 72.937 | 22.4 | 0.43 |
| polarC18 | 276.144 | 292.5289 | 22.4 | 0.43 |
| polarC18 | 276.144 | 126.4686 | 22.4 | 0.43 |
| polarC18 | 276.144 | 100.8018 | 22.4 | 0.43 |
| polarC18 | 276.144 | 124.8947 | 22.3 | 0.43 |
| polarC18 | 276.144 | 200.8519 | 22.3 | 0.43 |
| polarC18 | 276.144 | 79.6882 | 22.3 | 0.43 |
| polarC18 | 276.144 | 187.8188 | 22.2 | 0.43 |
| polarC18 | 276.144 | 272.4593 | 22.2 | 0.43 |
| polarC18 | 276.144 | 196.202 | 22.2 | 0.43 |
| polarC18 | 276.144 | 139.7788 | 22.2 | 0.43 |
| polarC18 | 276.144 | 276.1888 | 22.1 | 0.43 |
| polarC18 | 276.144 | 80.9483 | 22.1 | 0.43 |
| polarC18 | 276.144 | 190.1543 | 22.1 | 0.43 |
| polarC18 | 276.144 | 288.5432 | 22.1 | 0.42 |
| polarC18 | 276.144 | 257.3148 | 22.1 | 0.42 |
| polarC18 | 276.144 | 210.9291 | 22.1 | 0.42 |
| polarC18 | 276.144 | 93.8547 | 22.1 | 0.42 |
| polarC18 | 276.144 | 295.6949 | 22 | 0.42 |
| polarC18 | 276.144 | 67.3012 | 22 | 0.42 |
| polarC18 | 276.144 | 111.0158 | 22 | 0.42 |
| polarC18 | 276.144 | 139.9274 | 21.9 | 0.42 |

| | | | | |
|---|---|---|---|---|
| polarC18 | 276.144 | 279.8138 | 21.9 | 0.42 |
| polarC18 | 276.144 | 129.2368 | 21.9 | 0.42 |
| polarC18 | 276.144 | 67.975 | 21.9 | 0.42 |
| polarC18 | 276.144 | 94.2125 | 21.8 | 0.42 |
| polarC18 | 276.144 | 77.0231 | 21.8 | 0.42 |
| polarC18 | 276.144 | 191.9666 | 21.7 | 0.42 |
| polarC18 | 276.144 | 254.7523 | 21.7 | 0.42 |
| polarC18 | 276.144 | 255.469 | 21.7 | 0.42 |
| polarC18 | 276.144 | 119.4809 | 21.7 | 0.42 |
| polarC18 | 276.144 | 99.8796 | 21.7 | 0.42 |
| polarC18 | 276.144 | 139.098 | 21.6 | 0.42 |
| polarC18 | 276.144 | 94.7609 | 21.6 | 0.42 |
| polarC18 | 276.144 | 299.862 | 21.6 | 0.42 |
| polarC18 | 276.144 | 62.3315 | 21.6 | 0.42 |
| polarC18 | 276.144 | 94.3093 | 21.6 | 0.41 |
| polarC18 | 276.144 | 271.9167 | 21.6 | 0.41 |
| polarC18 | 276.144 | 159.7558 | 21.6 | 0.41 |
| polarC18 | 276.144 | 92.8826 | 21.6 | 0.41 |
| polarC18 | 276.144 | 143.1731 | 21.5 | 0.41 |
| polarC18 | 276.144 | 299.6921 | 21.5 | 0.41 |
| polarC18 | 276.144 | 294.0289 | 21.4 | 0.41 |
| polarC18 | 276.144 | 94.7756 | 21.4 | 0.41 |
| polarC18 | 276.144 | 177.5106 | 21.4 | 0.41 |
| polarC18 | 276.144 | 301.9189 | 21.4 | 0.41 |
| polarC18 | 276.144 | 152.4246 | 21.4 | 0.41 |
| polarC18 | 276.144 | 180.301 | 21.4 | 0.41 |
| polarC18 | 276.144 | 179.087 | 21.4 | 0.41 |
| polarC18 | 276.144 | 51.3796 | 21.4 | 0.41 |
| polarC18 | 276.144 | 183.1386 | 21.4 | 0.41 |
| polarC18 | 276.144 | 277.577 | 21.4 | 0.41 |
| polarC18 | 276.144 | 192.1102 | 21.3 | 0.41 |
| polarC18 | 276.144 | 195.9764 | 21.3 | 0.41 |
| polarC18 | 276.144 | 244.3889 | 21.3 | 0.41 |
| polarC18 | 276.144 | 167.797 | 21.3 | 0.41 |
| polarC18 | 276.144 | 55.2113 | 21.3 | 0.41 |
| polarC18 | 276.144 | 217.4043 | 21.3 | 0.41 |
| polarC18 | 276.144 | 182.6305 | 21.3 | 0.41 |
| polarC18 | 276.144 | 70.9816 | 21.3 | 0.41 |
| polarC18 | 276.144 | 65.0189 | 21.2 | 0.41 |
| polarC18 | 276.144 | 51.5757 | 21.2 | 0.41 |
| polarC18 | 276.144 | 247.3972 | 21.2 | 0.41 |
| polarC18 | 276.144 | 94.4978 | 21.2 | 0.41 |

| | | | | |
|---|---|---|---|---|
| polarC18 | 276.144 | 63.8651 | 21.2 | 0.41 |
| polarC18 | 276.144 | 79.3162 | 21.2 | 0.41 |
| polarC18 | 276.144 | 237.3101 | 21.1 | 0.41 |
| polarC18 | 276.144 | 94.0261 | 21.1 | 0.41 |
| polarC18 | 276.144 | 85.8075 | 21.1 | 0.41 |
| polarC18 | 276.144 | 177.6898 | 21.1 | 0.41 |
| polarC18 | 276.144 | 268.4483 | 21.1 | 0.41 |
| polarC18 | 276.144 | 91.4668 | 21.1 | 0.41 |
| polarC18 | 276.144 | 192.1539 | 21.1 | 0.41 |
| polarC18 | 276.144 | 80.9713 | 21.1 | 0.41 |
| polarC18 | 276.144 | 87.3224 | 21.1 | 0.41 |
| polarC18 | 276.144 | 274.5911 | 21 | 0.4 |
| polarC18 | 276.144 | 152.0224 | 21 | 0.4 |
| polarC18 | 276.144 | 101.7395 | 21 | 0.4 |
| polarC18 | 276.144 | 65.6759 | 21 | 0.4 |
| polarC18 | 276.144 | 170.6708 | 21 | 0.4 |
| polarC18 | 276.144 | 263.101 | 20.9 | 0.4 |
| polarC18 | 276.144 | 82.8646 | 20.9 | 0.4 |
| polarC18 | 276.144 | 124.4189 | 20.9 | 0.4 |
| polarC18 | 276.144 | 292.7328 | 20.9 | 0.4 |
| polarC18 | 276.144 | 135.7441 | 20.9 | 0.4 |
| polarC18 | 276.144 | 69.7031 | 20.9 | 0.4 |
| polarC18 | 276.144 | 52.2661 | 20.9 | 0.4 |
| polarC18 | 276.144 | 89.6852 | 20.9 | 0.4 |
| polarC18 | 276.144 | 168.7139 | 20.8 | 0.4 |
| polarC18 | 276.144 | 216.9253 | 20.8 | 0.4 |
| polarC18 | 276.144 | 93.1901 | 20.8 | 0.4 |
| polarC18 | 276.144 | 102.4148 | 20.8 | 0.4 |
| polarC18 | 276.144 | 181.6537 | 20.8 | 0.4 |
| polarC18 | 276.144 | 55.5361 | 20.8 | 0.4 |
| polarC18 | 276.144 | 263.2989 | 20.8 | 0.4 |
| polarC18 | 276.144 | 239.2367 | 20.8 | 0.4 |
| polarC18 | 276.144 | 68.739 | 20.8 | 0.4 |
| polarC18 | 276.144 | 290.9075 | 20.8 | 0.4 |
| polarC18 | 276.144 | 87.393 | 20.8 | 0.4 |
| polarC18 | 276.144 | 206.9223 | 20.7 | 0.4 |
| polarC18 | 276.144 | 150.9826 | 20.7 | 0.4 |
| polarC18 | 276.144 | 159.3915 | 20.7 | 0.4 |
| polarC18 | 276.144 | 78.0561 | 20.7 | 0.4 |
| polarC18 | 276.144 | 222.1663 | 20.7 | 0.4 |
| polarC18 | 276.144 | 301.9916 | 20.7 | 0.4 |
| polarC18 | 276.144 | 127.038 | 20.7 | 0.4 |

| | | | | |
|---|---|---|---|---|
| polarC18 | 276.144 | 146.6555 | 20.7 | 0.4 |
| polarC18 | 276.144 | 249.1622 | 20.7 | 0.4 |
| polarC18 | 276.144 | 198.7356 | 20.7 | 0.4 |
| polarC18 | 276.144 | 258.2457 | 20.7 | 0.4 |
| polarC18 | 276.144 | 81.9209 | 20.7 | 0.4 |
| polarC18 | 276.144 | 153.3598 | 20.7 | 0.4 |
| polarC18 | 276.144 | 89.0439 | 20.7 | 0.4 |
| polarC18 | 276.144 | 130.5108 | 20.7 | 0.4 |
| polarC18 | 276.144 | 110.3048 | 20.7 | 0.4 |
| polarC18 | 276.144 | 59.5499 | 20.7 | 0.4 |
| polarC18 | 276.144 | 150.1847 | 20.6 | 0.4 |
| polarC18 | 276.144 | 86.3294 | 20.6 | 0.4 |
| polarC18 | 276.144 | 63.8384 | 20.6 | 0.4 |
| polarC18 | 276.144 | 251.2195 | 20.6 | 0.4 |
| polarC18 | 276.144 | 59.4823 | 20.6 | 0.4 |
| polarC18 | 276.144 | 296.1972 | 20.6 | 0.4 |
| polarC18 | 276.144 | 65.3687 | 20.6 | 0.4 |
| polarC18 | 276.144 | 211.7021 | 20.6 | 0.4 |
| polarC18 | 276.144 | 62.8798 | 20.6 | 0.4 |
| polarC18 | 276.144 | 185.5576 | 20.6 | 0.4 |
| polarC18 | 276.144 | 149.235 | 20.6 | 0.4 |
| polarC18 | 276.144 | 120.9868 | 20.6 | 0.4 |
| polarC18 | 276.144 | 124.5167 | 20.6 | 0.4 |
| polarC18 | 276.144 | 97.7223 | 20.5 | 0.4 |
| polarC18 | 276.144 | 86.3052 | 20.5 | 0.4 |
| polarC18 | 276.144 | 171.6564 | 20.5 | 0.39 |
| polarC18 | 276.144 | 94.6566 | 20.5 | 0.39 |
| polarC18 | 276.144 | 108.3387 | 20.5 | 0.39 |
| polarC18 | 276.144 | 71.1666 | 20.5 | 0.39 |
| polarC18 | 276.144 | 74.7894 | 20.4 | 0.39 |
| polarC18 | 276.144 | 200.3951 | 20.4 | 0.39 |
| polarC18 | 276.144 | 239.652 | 20.4 | 0.39 |
| polarC18 | 276.144 | 94.137 | 20.4 | 0.39 |
| polarC18 | 276.144 | 204.8407 | 20.4 | 0.39 |
| polarC18 | 276.144 | 78.7539 | 20.4 | 0.39 |
| polarC18 | 276.144 | 53.1185 | 20.4 | 0.39 |
| polarC18 | 276.144 | 188.1194 | 20.4 | 0.39 |
| polarC18 | 276.144 | 82.0302 | 20.4 | 0.39 |
| polarC18 | 276.144 | 230.55 | 20.4 | 0.39 |
| polarC18 | 276.144 | 88.3603 | 20.4 | 0.39 |
| polarC18 | 276.144 | 93.1064 | 20.4 | 0.39 |
| polarC18 | 276.144 | 127.1752 | 20.4 | 0.39 |

| | | | | |
|---|---|---|---|---|
| polarC18 | 276.144 | 146.1303 | 20.4 | 0.39 |
| polarC18 | 276.144 | 151.2582 | 20.3 | 0.39 |
| polarC18 | 276.144 | 66.7975 | 20.3 | 0.39 |
| polarC18 | 276.144 | 147.176 | 20.3 | 0.39 |
| polarC18 | 276.144 | 114.1325 | 20.3 | 0.39 |
| polarC18 | 276.144 | 99.8247 | 20.3 | 0.39 |
| polarC18 | 276.144 | 129.0442 | 20.3 | 0.39 |
| polarC18 | 276.144 | 190.8494 | 20.3 | 0.39 |
| polarC18 | 276.144 | 264.6915 | 20.3 | 0.39 |
| polarC18 | 276.144 | 105.6352 | 20.3 | 0.39 |
| polarC18 | 276.144 | 151.1265 | 20.3 | 0.39 |
| polarC18 | 276.144 | 53.802 | 20.3 | 0.39 |
| polarC18 | 276.144 | 283.4718 | 20.3 | 0.39 |
| polarC18 | 276.144 | 135.5945 | 20.3 | 0.39 |
| polarC18 | 276.144 | 56.9576 | 20.3 | 0.39 |
| polarC18 | 276.144 | 235.8694 | 20.3 | 0.39 |
| polarC18 | 276.144 | 198.7102 | 20.3 | 0.39 |
| polarC18 | 276.144 | 202.8859 | 20.3 | 0.39 |
| polarC18 | 276.144 | 205.5929 | 20.3 | 0.39 |
| polarC18 | 276.144 | 272.8251 | 20.3 | 0.39 |
| polarC18 | 276.144 | 156.0973 | 20.3 | 0.39 |
| polarC18 | 276.144 | 199.9184 | 20.2 | 0.39 |
| polarC18 | 276.144 | 242.6284 | 20.2 | 0.39 |
| polarC18 | 276.144 | 286.5434 | 20.2 | 0.39 |
| polarC18 | 276.144 | 236.4974 | 20.2 | 0.39 |
| polarC18 | 276.144 | 66.6964 | 20.2 | 0.39 |
| polarC18 | 276.144 | 101.5513 | 20.2 | 0.39 |
| polarC18 | 276.144 | 203.9654 | 20.2 | 0.39 |
| polarC18 | 276.144 | 229.9324 | 20.2 | 0.39 |
| polarC18 | 276.144 | 50.6334 | 20.2 | 0.39 |
| polarC18 | 276.144 | 290.7876 | 20.2 | 0.39 |
| polarC18 | 276.144 | 186.3517 | 20.2 | 0.39 |
| polarC18 | 276.144 | 161.2329 | 20.2 | 0.39 |
| polarC18 | 276.144 | 150.4619 | 20.1 | 0.39 |
| polarC18 | 276.144 | 258.9975 | 20.1 | 0.39 |
| polarC18 | 276.144 | 51.5297 | 20.1 | 0.39 |
| polarC18 | 276.144 | 162.5723 | 20.1 | 0.39 |
| polarC18 | 276.144 | 294.4864 | 20.1 | 0.39 |
| polarC18 | 276.144 | 193.6076 | 20.1 | 0.39 |
| polarC18 | 276.144 | 57.6175 | 20.1 | 0.39 |
| polarC18 | 276.144 | 126.3246 | 20.1 | 0.39 |
| polarC18 | 276.144 | 115.1621 | 20 | 0.39 |

| | | | | |
|---|---|---|---|---|
| polarC18 | 276.144 | 229.1836 | 20 | 0.39 |
| polarC18 | 276.144 | 101.3305 | 20 | 0.39 |
| polarC18 | 276.144 | 275.1344 | 20 | 0.39 |
| polarC18 | 276.144 | 180.3738 | 20 | 0.39 |
| polarC18 | 276.144 | 133.6722 | 20 | 0.38 |
| polarC18 | 276.144 | 90.0127 | 20 | 0.38 |
| polarC18 | 276.144 | 150.6058 | 20 | 0.38 |
| polarC18 | 276.144 | 79.5612 | 19.9 | 0.38 |
| polarC18 | 276.144 | 95.1812 | 19.9 | 0.38 |
| polarC18 | 276.144 | 203.8113 | 19.9 | 0.38 |
| polarC18 | 276.144 | 98.9275 | 19.9 | 0.38 |
| polarC18 | 276.144 | 172.7079 | 19.9 | 0.38 |
| polarC18 | 276.144 | 84.3851 | 19.9 | 0.38 |
| polarC18 | 276.144 | 55.0178 | 19.9 | 0.38 |
| polarC18 | 276.144 | 89.0784 | 19.9 | 0.38 |
| polarC18 | 276.144 | 93.945 | 19.9 | 0.38 |
| polarC18 | 276.144 | 65.1803 | 19.9 | 0.38 |
| polarC18 | 276.144 | 233.0775 | 19.9 | 0.38 |
| polarC18 | 276.144 | 170.1869 | 19.9 | 0.38 |
| polarC18 | 276.144 | 235.4556 | 19.8 | 0.38 |
| polarC18 | 276.144 | 212.2927 | 19.8 | 0.38 |
| polarC18 | 276.144 | 259.6583 | 19.8 | 0.38 |
| polarC18 | 276.144 | 67.2776 | 19.8 | 0.38 |
| polarC18 | 276.144 | 154.5431 | 19.8 | 0.38 |
| polarC18 | 276.144 | 75.1596 | 19.8 | 0.38 |
| polarC18 | 276.144 | 53.7285 | 19.8 | 0.38 |
| polarC18 | 276.144 | 86.7494 | 19.8 | 0.38 |
| polarC18 | 276.144 | 246.4968 | 19.8 | 0.38 |
| polarC18 | 276.144 | 109.6198 | 19.8 | 0.38 |
| polarC18 | 276.144 | 117.4023 | 19.8 | 0.38 |
| polarC18 | 276.144 | 70.9651 | 19.8 | 0.38 |
| polarC18 | 276.144 | 78.9617 | 19.8 | 0.38 |
| polarC18 | 276.144 | 85.3297 | 19.8 | 0.38 |
| polarC18 | 276.144 | 116.5162 | 19.7 | 0.38 |
| polarC18 | 276.144 | 112.3511 | 19.7 | 0.38 |
| polarC18 | 276.144 | 81.9665 | 19.7 | 0.38 |
| polarC18 | 276.144 | 163.6218 | 19.7 | 0.38 |
| polarC18 | 276.144 | 276.5042 | 19.7 | 0.38 |
| polarC18 | 276.144 | 53.6316 | 19.7 | 0.38 |
| polarC18 | 276.144 | 73.8934 | 19.7 | 0.38 |
| polarC18 | 276.144 | 64.6938 | 19.7 | 0.38 |
| polarC18 | 276.144 | 261.4577 | 19.7 | 0.38 |

| | | | | |
|---|---|---|---|---|
| polarC18 | 276.144 | 163.7832 | 19.7 | 0.38 |
| polarC18 | 276.144 | 289.1753 | 19.6 | 0.38 |
| polarC18 | 276.144 | 67.0081 | 19.6 | 0.38 |
| polarC18 | 276.144 | 130.98 | 19.6 | 0.38 |
| polarC18 | 276.144 | 78.3296 | 19.6 | 0.38 |
| polarC18 | 276.144 | 87.8559 | 19.6 | 0.38 |
| polarC18 | 276.144 | 132.699 | 19.6 | 0.38 |
| polarC18 | 276.144 | 50.1972 | 19.6 | 0.38 |
| polarC18 | 276.144 | 232.0893 | 19.6 | 0.38 |
| polarC18 | 276.144 | 98.9751 | 19.5 | 0.38 |
| polarC18 | 276.144 | 70.8494 | 19.5 | 0.38 |
| polarC18 | 276.144 | 62.6283 | 19.5 | 0.38 |
| polarC18 | 276.144 | 115.5935 | 19.5 | 0.38 |
| polarC18 | 276.144 | 175.6354 | 19.5 | 0.38 |
| polarC18 | 276.144 | 93.3597 | 19.5 | 0.38 |
| polarC18 | 276.144 | 149.6352 | 19.5 | 0.38 |
| polarC18 | 276.144 | 178.636 | 19.5 | 0.37 |
| polarC18 | 276.144 | 88.2721 | 19.5 | 0.37 |
| polarC18 | 276.144 | 184.327 | 19.5 | 0.37 |
| polarC18 | 276.144 | 254.3106 | 19.5 | 0.37 |
| polarC18 | 276.144 | 62.4659 | 19.5 | 0.37 |
| polarC18 | 276.144 | 273.1152 | 19.5 | 0.37 |
| polarC18 | 276.144 | 62.1101 | 19.5 | 0.37 |
| polarC18 | 276.144 | 59.344 | 19.5 | 0.37 |
| polarC18 | 276.144 | 56.6536 | 19.4 | 0.37 |
| polarC18 | 276.144 | 213.6893 | 19.4 | 0.37 |
| polarC18 | 276.144 | 174.886 | 19.4 | 0.37 |
| polarC18 | 276.144 | 86.0729 | 19.4 | 0.37 |
| polarC18 | 276.144 | 191.2253 | 19.4 | 0.37 |
| polarC18 | 276.144 | 96.7145 | 19.4 | 0.37 |
| polarC18 | 276.144 | 84.6306 | 19.4 | 0.37 |
| polarC18 | 276.144 | 61.0079 | 19.4 | 0.37 |
| polarC18 | 276.144 | 267.2034 | 19.3 | 0.37 |
| polarC18 | 276.144 | 159.6613 | 19.3 | 0.37 |
| polarC18 | 276.144 | 84.4124 | 19.3 | 0.37 |
| polarC18 | 276.144 | 94.2401 | 19.3 | 0.37 |
| polarC18 | 276.144 | 155.0084 | 19.3 | 0.37 |
| polarC18 | 276.144 | 247.9702 | 19.3 | 0.37 |
| polarC18 | 276.144 | 110.2356 | 19.3 | 0.37 |
| polarC18 | 276.144 | 89.5131 | 19.3 | 0.37 |
| polarC18 | 276.144 | 85.2438 | 19.3 | 0.37 |
| polarC18 | 276.144 | 154.313 | 19.3 | 0.37 |

| | | | | |
|---|---|---|---|---|
| polarC18 | 276.144 | 133.2577 | 19.3 | 0.37 |
| polarC18 | 276.144 | 152.848 | 19.2 | 0.37 |
| polarC18 | 276.144 | 93.9055 | 19.2 | 0.37 |
| polarC18 | 276.144 | 89.4927 | 19.2 | 0.37 |
| polarC18 | 276.144 | 215.077 | 19.2 | 0.37 |
| polarC18 | 276.144 | 179.4982 | 19.2 | 0.37 |
| polarC18 | 276.144 | 216.8979 | 19.2 | 0.37 |
| polarC18 | 276.144 | 188.5436 | 19.2 | 0.37 |
| polarC18 | 276.144 | 288.3276 | 19.2 | 0.37 |
| polarC18 | 276.144 | 260.3663 | 19.2 | 0.37 |
| polarC18 | 276.144 | 50.2756 | 19.1 | 0.37 |
| polarC18 | 276.144 | 81.8725 | 19.1 | 0.37 |
| polarC18 | 276.144 | 164.7733 | 19.1 | 0.37 |
| polarC18 | 276.144 | 235.9923 | 19.1 | 0.37 |
| polarC18 | 276.144 | 249.5261 | 19.1 | 0.37 |
| polarC18 | 276.144 | 192.4615 | 19.1 | 0.37 |
| polarC18 | 276.144 | 263.5098 | 19.1 | 0.37 |
| polarC18 | 276.144 | 99.8874 | 19.1 | 0.37 |
| polarC18 | 276.144 | 84.037 | 19.1 | 0.37 |
| polarC18 | 276.144 | 153.9924 | 19.1 | 0.37 |
| polarC18 | 276.144 | 75.4809 | 19.1 | 0.37 |
| polarC18 | 276.144 | 119.8684 | 19.1 | 0.37 |
| polarC18 | 276.144 | 79.7847 | 19.1 | 0.37 |
| polarC18 | 276.144 | 171.9751 | 19 | 0.37 |
| polarC18 | 276.144 | 52.6848 | 19 | 0.37 |
| polarC18 | 276.144 | 185.853 | 19 | 0.37 |
| polarC18 | 276.144 | 133.8995 | 19 | 0.37 |
| polarC18 | 276.144 | 276.8176 | 19 | 0.37 |
| polarC18 | 276.144 | 231.6467 | 19 | 0.37 |
| polarC18 | 276.144 | 179.1211 | 19 | 0.36 |
| polarC18 | 276.144 | 83.323 | 19 | 0.36 |
| polarC18 | 276.144 | 121.1971 | 19 | 0.36 |
| polarC18 | 276.144 | 115.6589 | 19 | 0.36 |
| polarC18 | 276.144 | 109.8421 | 18.9 | 0.36 |
| polarC18 | 276.144 | 174.6121 | 18.9 | 0.36 |
| polarC18 | 276.144 | 227.0661 | 18.9 | 0.36 |
| polarC18 | 276.144 | 134.4934 | 18.9 | 0.36 |
| polarC18 | 276.144 | 181.1157 | 18.9 | 0.36 |
| polarC18 | 276.144 | 52.7215 | 18.9 | 0.36 |
| polarC18 | 276.144 | 50.7028 | 18.9 | 0.36 |
| polarC18 | 276.144 | 259.6154 | 18.9 | 0.36 |
| polarC18 | 276.144 | 286.3061 | 18.9 | 0.36 |

| | | | | |
|---|---|---|---|---|
| polarC18 | 276.144 | 224.7723 | 18.9 | 0.36 |
| polarC18 | 276.144 | 84.2553 | 18.8 | 0.36 |
| polarC18 | 276.144 | 154.2865 | 18.8 | 0.36 |
| polarC18 | 276.144 | 75.1141 | 18.8 | 0.36 |
| polarC18 | 276.144 | 51.5443 | 18.8 | 0.36 |
| polarC18 | 276.144 | 50.2531 | 18.8 | 0.36 |
| polarC18 | 276.144 | 73.4733 | 18.8 | 0.36 |
| polarC18 | 276.144 | 204.1088 | 18.8 | 0.36 |
| polarC18 | 276.144 | 178.3349 | 18.8 | 0.36 |
| polarC18 | 276.144 | 215.653 | 18.8 | 0.36 |
| polarC18 | 276.144 | 153.6944 | 18.8 | 0.36 |
| polarC18 | 276.144 | 100.711 | 18.7 | 0.36 |
| polarC18 | 276.144 | 292.0153 | 18.7 | 0.36 |
| polarC18 | 276.144 | 237.2639 | 18.7 | 0.36 |
| polarC18 | 276.144 | 127.4244 | 18.7 | 0.36 |
| polarC18 | 276.144 | 238.462 | 18.7 | 0.36 |
| polarC18 | 276.144 | 168.0838 | 18.7 | 0.36 |
| polarC18 | 276.144 | 51.1822 | 18.7 | 0.36 |
| polarC18 | 276.144 | 62.954 | 18.7 | 0.36 |
| polarC18 | 276.144 | 148.6944 | 18.6 | 0.36 |
| polarC18 | 276.144 | 220.522 | 18.6 | 0.36 |
| polarC18 | 276.144 | 51.2235 | 18.6 | 0.36 |
| polarC18 | 276.144 | 185.6751 | 18.6 | 0.36 |
| polarC18 | 276.144 | 85.4382 | 18.6 | 0.36 |
| polarC18 | 276.144 | 63.9258 | 18.6 | 0.36 |
| polarC18 | 276.144 | 165.7694 | 18.6 | 0.36 |
| polarC18 | 276.144 | 128.2185 | 18.5 | 0.36 |
| polarC18 | 276.144 | 105.2662 | 18.5 | 0.36 |
| polarC18 | 276.144 | 100.4336 | 18.5 | 0.36 |
| polarC18 | 276.144 | 128.1328 | 18.5 | 0.36 |
| polarC18 | 276.144 | 287.919 | 18.5 | 0.36 |
| polarC18 | 276.144 | 272.6279 | 18.5 | 0.35 |
| polarC18 | 276.144 | 118.5778 | 18.5 | 0.35 |
| polarC18 | 276.144 | 258.6911 | 18.4 | 0.35 |
| polarC18 | 276.144 | 61.4114 | 18.4 | 0.35 |
| polarC18 | 276.144 | 156.3871 | 18.4 | 0.35 |
| polarC18 | 276.144 | 137.2813 | 18.4 | 0.35 |
| polarC18 | 276.144 | 64.6896 | 18.4 | 0.35 |
| polarC18 | 276.144 | 61.8183 | 18.4 | 0.35 |
| polarC18 | 276.144 | 80.6269 | 18.4 | 0.35 |
| polarC18 | 276.144 | 104.7084 | 18.3 | 0.35 |
| polarC18 | 276.144 | 93.4144 | 18.3 | 0.35 |

| | | | | |
|---|---|---|---|---|
| polarC18 | 276.144 | 183.8727 | 18.3 | 0.35 |
| polarC18 | 276.144 | 66.8548 | 18.3 | 0.35 |
| polarC18 | 276.144 | 63.6812 | 18.3 | 0.35 |
| polarC18 | 276.144 | 58.6875 | 18.3 | 0.35 |
| polarC18 | 276.144 | 274.4162 | 18.2 | 0.35 |
| polarC18 | 276.144 | 94.7058 | 18.2 | 0.35 |
| polarC18 | 276.144 | 80.2405 | 18.2 | 0.35 |
| polarC18 | 276.144 | 165.607 | 18.2 | 0.35 |
| polarC18 | 276.144 | 219.7029 | 18.2 | 0.35 |
| polarC18 | 276.144 | 56.5621 | 18.2 | 0.35 |
| polarC18 | 276.144 | 243.0787 | 18.2 | 0.35 |
| polarC18 | 276.144 | 60.8552 | 18.2 | 0.35 |
| polarC18 | 276.144 | 153.9631 | 18.2 | 0.35 |
| polarC18 | 276.144 | 92.9145 | 18.1 | 0.35 |
| polarC18 | 276.144 | 140.9186 | 18.1 | 0.35 |
| polarC18 | 276.144 | 100.3701 | 18.1 | 0.35 |
| polarC18 | 276.144 | 153.257 | 18.1 | 0.35 |
| polarC18 | 276.144 | 139.0836 | 18.1 | 0.35 |
| polarC18 | 276.144 | 112.0028 | 18.1 | 0.35 |
| polarC18 | 276.144 | 76.5545 | 18 | 0.35 |
| polarC18 | 276.144 | 57.6704 | 18 | 0.35 |
| polarC18 | 276.144 | 50.9282 | 18 | 0.35 |
| polarC18 | 276.144 | 204.3116 | 18 | 0.35 |
| polarC18 | 276.144 | 176.2746 | 18 | 0.35 |
| polarC18 | 276.144 | 156.9582 | 18 | 0.35 |
| polarC18 | 276.144 | 94.0948 | 17.9 | 0.34 |
| polarC18 | 276.144 | 89.1945 | 17.9 | 0.34 |
| polarC18 | 276.144 | 149.4609 | 17.9 | 0.34 |
| polarC18 | 276.144 | 54.2357 | 17.8 | 0.34 |
| polarC18 | 276.144 | 94.1889 | 17.8 | 0.34 |
| polarC18 | 276.144 | 152.6255 | 17.8 | 0.34 |
| polarC18 | 276.144 | 130.4312 | 17.8 | 0.34 |
| polarC18 | 276.144 | 89.9435 | 17.7 | 0.34 |
| polarC18 | 276.144 | 69.1434 | 17.7 | 0.34 |
| polarC18 | 276.144 | 134.2323 | 17.7 | 0.34 |
| polarC18 | 276.144 | 185.0244 | 17.6 | 0.34 |
| polarC18 | 276.144 | 202.571 | 17.6 | 0.34 |
| polarC18 | 276.144 | 73.5528 | 17.5 | 0.34 |
| polarC18 | 276.144 | 114.6419 | 17.5 | 0.34 |
| polarC18 | 276.144 | 144.7465 | 17.4 | 0.34 |
| polarC18 | 276.144 | 65.6064 | 17.4 | 0.33 |
| polarC18 | 276.144 | 156.1328 | 17.4 | 0.33 |

| | | | | |
|---|---|---|---|---|
| polarC18 | 276.144 | 145.3601 | 17.4 | 0.33 |
| polarC18 | 276.144 | 134.0292 | 17.4 | 0.33 |
| polarC18 | 276.144 | 69.1164 | 17.4 | 0.33 |
| polarC18 | 276.144 | 221.8275 | 17.3 | 0.33 |
| polarC18 | 276.144 | 55.4304 | 17.3 | 0.33 |
| polarC18 | 276.144 | 98.9453 | 17.3 | 0.33 |
| polarC18 | 276.144 | 70.3423 | 17.2 | 0.33 |
| polarC18 | 276.144 | 65.4368 | 17.2 | 0.33 |
| polarC18 | 276.144 | 72.7036 | 17.2 | 0.33 |
| polarC18 | 276.144 | 106.2788 | 17.1 | 0.33 |
| polarC18 | 276.144 | 103.0316 | 17.1 | 0.33 |
| polarC18 | 276.144 | 191.0479 | 17 | 0.33 |
| polarC18 | 276.144 | 182.0801 | 17 | 0.33 |
| polarC18 | 276.144 | 63.7969 | 17 | 0.33 |
| polarC18 | 276.144 | 153.1509 | 17 | 0.33 |
| polarC18 | 276.144 | 74.4703 | 17 | 0.33 |
| polarC18 | 276.144 | 54.9377 | 16.9 | 0.33 |
| polarC18 | 276.144 | 78.1464 | 16.9 | 0.32 |
| polarC18 | 276.144 | 138.7525 | 16.6 | 0.32 |
| polarC18 | 276.144 | 100.2776 | 16.6 | 0.32 |
| polarC18 | 276.144 | 60.0099 | 16.5 | 0.32 |
| polarC18 | 276.144 | 224.8443 | 16.5 | 0.32 |
| polarC18 | 276.144 | 103.0698 | 16.5 | 0.32 |
| polarC18 | 276.144 | 124.2514 | 16.4 | 0.32 |
| polarC18 | 276.144 | 96.1859 | 16.4 | 0.32 |
| polarC18 | 276.144 | 177.3612 | 16.3 | 0.31 |
| polarC18 | 276.144 | 164.2993 | 16.3 | 0.31 |
| polarC18 | 276.144 | 117.4294 | 16.3 | 0.31 |
| polarC18 | 276.144 | 186.9688 | 16.2 | 0.31 |
| polarC18 | 276.144 | 94.3176 | 16.2 | 0.31 |
| polarC18 | 276.144 | 99.6545 | 16.2 | 0.31 |
| polarC18 | 276.144 | 188.3431 | 16.2 | 0.31 |
| polarC18 | 276.144 | 159.8596 | 16.2 | 0.31 |
| polarC18 | 276.144 | 55.938 | 16.1 | 0.31 |
| polarC18 | 276.144 | 102.3884 | 16.1 | 0.31 |
| polarC18 | 276.144 | 114.9482 | 16.1 | 0.31 |
| polarC18 | 276.144 | 96.344 | 16.1 | 0.31 |
| polarC18 | 276.144 | 56.7323 | 16.1 | 0.31 |
| polarC18 | 276.144 | 142.4638 | 16 | 0.31 |
| polarC18 | 276.144 | 204.6731 | 16 | 0.31 |
| polarC18 | 276.144 | 63.7559 | 16 | 0.31 |
| polarC18 | 276.144 | 295.9268 | 16 | 0.31 |

| | | | | |
|---|---|---|---|---|
| polarC18 | 276.144 | 57.07 | 16 | 0.31 |
| polarC18 | 276.144 | 68.7611 | 15.9 | 0.31 |
| polarC18 | 276.144 | 75.4977 | 15.9 | 0.31 |
| polarC18 | 276.144 | 68.1472 | 15.9 | 0.31 |
| polarC18 | 276.144 | 87.1168 | 15.9 | 0.31 |
| polarC18 | 276.144 | 62.5045 | 15.9 | 0.31 |
| polarC18 | 276.144 | 94.5985 | 15.8 | 0.3 |
| polarC18 | 276.144 | 174.7157 | 15.8 | 0.3 |
| polarC18 | 276.144 | 174.3115 | 15.8 | 0.3 |
| polarC18 | 276.144 | 300.8654 | 15.7 | 0.3 |
| polarC18 | 276.144 | 144.1431 | 15.7 | 0.3 |
| polarC18 | 276.144 | 104.2996 | 15.7 | 0.3 |
| polarC18 | 276.144 | 81.1848 | 15.7 | 0.3 |
| polarC18 | 276.144 | 107.1814 | 15.7 | 0.3 |
| polarC18 | 276.144 | 94.5287 | 15.7 | 0.3 |
| polarC18 | 276.144 | 248.8122 | 15.7 | 0.3 |
| polarC18 | 276.144 | 95.5837 | 15.6 | 0.3 |
| polarC18 | 276.144 | 63.5873 | 15.6 | 0.3 |
| polarC18 | 276.144 | 94.627 | 15.6 | 0.3 |
| polarC18 | 276.144 | 221.7053 | 15.6 | 0.3 |
| polarC18 | 276.144 | 233.8759 | 15.6 | 0.3 |
| polarC18 | 276.144 | 59.2301 | 15.6 | 0.3 |
| polarC18 | 276.144 | 152.4755 | 15.6 | 0.3 |
| polarC18 | 276.144 | 201.9605 | 15.5 | 0.3 |
| polarC18 | 276.144 | 164.4023 | 15.5 | 0.3 |
| polarC18 | 276.144 | 275.6801 | 15.5 | 0.3 |
| polarC18 | 276.144 | 172.7857 | 15.5 | 0.3 |
| polarC18 | 276.144 | 249.9124 | 15.5 | 0.3 |
| polarC18 | 276.144 | 216.7625 | 15.5 | 0.3 |
| polarC18 | 276.144 | 157.6275 | 15.5 | 0.3 |
| polarC18 | 276.144 | 56.9644 | 15.5 | 0.3 |
| polarC18 | 276.144 | 145.8356 | 15.5 | 0.3 |
| polarC18 | 276.144 | 76.8652 | 15.5 | 0.3 |
| polarC18 | 276.144 | 105.1693 | 15.5 | 0.3 |
| polarC18 | 276.144 | 164.5859 | 15.5 | 0.3 |
| polarC18 | 276.144 | 88.8959 | 15.5 | 0.3 |
| polarC18 | 276.144 | 178.5433 | 15.5 | 0.3 |
| polarC18 | 276.144 | 238.8715 | 15.4 | 0.3 |
| polarC18 | 276.144 | 58.8923 | 15.4 | 0.3 |
| polarC18 | 276.144 | 183.2736 | 15.4 | 0.3 |
| polarC18 | 276.144 | 161.9606 | 15.4 | 0.3 |
| polarC18 | 276.144 | 207.0691 | 15.4 | 0.3 |

| | | | | |
|---|---|---|---|---|
| polarC18 | 276.144 | 158.8216 | 15.4 | 0.3 |
| polarC18 | 276.144 | 217.8739 | 15.3 | 0.3 |
| polarC18 | 276.144 | 64.3299 | 15.3 | 0.29 |
| polarC18 | 276.144 | 65.272 | 15.3 | 0.29 |
| polarC18 | 276.144 | 71.3987 | 15.3 | 0.29 |
| polarC18 | 276.144 | 51.6001 | 15.3 | 0.29 |
| polarC18 | 276.144 | 111.1864 | 15.3 | 0.29 |
| polarC18 | 276.144 | 275.7575 | 15.3 | 0.29 |
| polarC18 | 276.144 | 245.8567 | 15.3 | 0.29 |
| polarC18 | 276.144 | 107.0874 | 15.3 | 0.29 |
| polarC18 | 276.144 | 64.4357 | 15.2 | 0.29 |
| polarC18 | 276.144 | 245.9587 | 15.2 | 0.29 |
| polarC18 | 276.144 | 149.9748 | 15.2 | 0.29 |
| polarC18 | 276.144 | 150.1125 | 15.2 | 0.29 |
| polarC18 | 276.144 | 201.211 | 15.2 | 0.29 |
| polarC18 | 276.144 | 244.5171 | 15.2 | 0.29 |
| polarC18 | 276.144 | 108.6429 | 15.2 | 0.29 |
| polarC18 | 276.144 | 193.1933 | 15.2 | 0.29 |
| polarC18 | 276.144 | 278.24 | 15.2 | 0.29 |
| polarC18 | 276.144 | 155.5919 | 15.1 | 0.29 |
| polarC18 | 276.144 | 82.3266 | 15.1 | 0.29 |
| polarC18 | 276.144 | 131.3349 | 15.1 | 0.29 |
| polarC18 | 276.144 | 217.9776 | 15.1 | 0.29 |
| polarC18 | 276.144 | 128.5553 | 15.1 | 0.29 |
| polarC18 | 276.144 | 256.3237 | 15.1 | 0.29 |
| polarC18 | 276.144 | 77.4672 | 15.1 | 0.29 |
| polarC18 | 276.144 | 240.2314 | 15.1 | 0.29 |
| polarC18 | 276.144 | 164.6114 | 15.1 | 0.29 |
| polarC18 | 276.144 | 136.1176 | 15.1 | 0.29 |
| polarC18 | 276.144 | 207.485 | 15 | 0.29 |
| polarC18 | 276.144 | 141.3464 | 15 | 0.29 |
| polarC18 | 276.144 | 77.3366 | 15 | 0.29 |
| polarC18 | 276.144 | 101.0004 | 15 | 0.29 |
| polarC18 | 276.144 | 224.2708 | 15 | 0.29 |
| polarC18 | 276.144 | 197.0906 | 15 | 0.29 |
| polarC18 | 276.144 | 246.7172 | 15 | 0.29 |
| polarC18 | 276.144 | 130.6968 | 15 | 0.29 |
| polarC18 | 276.144 | 94.2694 | 15 | 0.29 |
| polarC18 | 276.144 | 59.1081 | 15 | 0.29 |
| polarC18 | 276.144 | 158.3633 | 15 | 0.29 |
| polarC18 | 276.144 | 172.135 | 15 | 0.29 |
| polarC18 | 276.144 | 100.8847 | 15 | 0.29 |

| | | | | |
|---|---|---|---|---|
| polarC18 | 276.144 | 55.7917 | 15 | 0.29 |
| polarC18 | 276.144 | 57.9265 | 15 | 0.29 |
| polarC18 | 276.144 | 77.7066 | 15 | 0.29 |
| polarC18 | 276.144 | 65.3654 | 15 | 0.29 |
| polarC18 | 276.144 | 223.0995 | 15 | 0.29 |
| polarC18 | 276.144 | 149.7239 | 15 | 0.29 |
| polarC18 | 276.144 | 187.7589 | 15 | 0.29 |
| polarC18 | 276.144 | 50.5898 | 15 | 0.29 |
| polarC18 | 276.144 | 78.4384 | 15 | 0.29 |
| polarC18 | 276.144 | 60.8506 | 15 | 0.29 |
| polarC18 | 276.144 | 235.8069 | 15 | 0.29 |
| polarC18 | 276.144 | 134.4122 | 14.9 | 0.29 |
| polarC18 | 276.144 | 85.6994 | 14.9 | 0.29 |
| polarC18 | 276.144 | 71.6481 | 14.9 | 0.29 |
| polarC18 | 276.144 | 53.4599 | 14.9 | 0.29 |
| polarC18 | 276.144 | 79.6388 | 14.9 | 0.29 |
| polarC18 | 276.144 | 179.4229 | 14.9 | 0.29 |
| polarC18 | 276.144 | 202.952 | 14.9 | 0.29 |
| polarC18 | 276.144 | 68.3614 | 14.9 | 0.29 |
| polarC18 | 276.144 | 63.3741 | 14.9 | 0.29 |
| polarC18 | 276.144 | 183.2359 | 14.9 | 0.29 |
| polarC18 | 276.144 | 80.0492 | 14.9 | 0.29 |
| polarC18 | 276.144 | 86.819 | 14.9 | 0.29 |
| polarC18 | 276.144 | 91.5573 | 14.9 | 0.29 |
| polarC18 | 276.144 | 175.4049 | 14.9 | 0.29 |
| polarC18 | 276.144 | 156.5636 | 14.9 | 0.29 |
| polarC18 | 276.144 | 91.5308 | 14.9 | 0.29 |
| polarC18 | 276.144 | 70.4357 | 14.9 | 0.29 |
| polarC18 | 276.144 | 61.8432 | 14.9 | 0.29 |
| polarC18 | 276.144 | 75.7185 | 14.9 | 0.29 |
| polarC18 | 276.144 | 219.3668 | 14.8 | 0.29 |
| polarC18 | 276.144 | 74.3138 | 14.8 | 0.29 |
| polarC18 | 276.144 | 94.7491 | 14.8 | 0.29 |
| polarC18 | 276.144 | 229.5871 | 14.8 | 0.29 |
| polarC18 | 276.144 | 299.7767 | 14.8 | 0.29 |
| polarC18 | 276.144 | 99.4583 | 14.8 | 0.28 |
| polarC18 | 276.144 | 134.4406 | 14.8 | 0.28 |
| polarC18 | 276.144 | 52.8934 | 14.8 | 0.28 |
| polarC18 | 276.144 | 274.7489 | 14.8 | 0.28 |
| polarC18 | 276.144 | 193.0177 | 14.8 | 0.28 |
| polarC18 | 276.144 | 141.7224 | 14.8 | 0.28 |
| polarC18 | 276.144 | 195.1456 | 14.8 | 0.28 |

| | | | | |
|---|---|---|---|---|
| polarC18 | 276.144 | 54.6721 | 14.8 | 0.28 |
| polarC18 | 276.144 | 98.4465 | 14.8 | 0.28 |
| polarC18 | 276.144 | 204.895 | 14.8 | 0.28 |
| polarC18 | 276.144 | 159.1187 | 14.8 | 0.28 |
| polarC18 | 276.144 | 63.2312 | 14.8 | 0.28 |
| polarC18 | 276.144 | 163.7375 | 14.8 | 0.28 |
| polarC18 | 276.144 | 83.1371 | 14.7 | 0.28 |
| polarC18 | 276.144 | 53.91 | 14.7 | 0.28 |
| polarC18 | 276.144 | 79.4977 | 14.7 | 0.28 |
| polarC18 | 276.144 | 228.4065 | 14.7 | 0.28 |
| polarC18 | 276.144 | 57.0621 | 14.7 | 0.28 |
| polarC18 | 276.144 | 80.6094 | 14.7 | 0.28 |
| polarC18 | 276.144 | 62.1633 | 14.7 | 0.28 |
| polarC18 | 276.144 | 91.7767 | 14.7 | 0.28 |
| polarC18 | 276.144 | 116.8669 | 14.7 | 0.28 |
| polarC18 | 276.144 | 102.7012 | 14.7 | 0.28 |
| polarC18 | 276.144 | 217.2895 | 14.7 | 0.28 |
| polarC18 | 276.144 | 202.5186 | 14.7 | 0.28 |
| polarC18 | 276.144 | 148.5374 | 14.7 | 0.28 |
| polarC18 | 276.144 | 290.3593 | 14.7 | 0.28 |
| polarC18 | 276.144 | 83.7073 | 14.7 | 0.28 |
| polarC18 | 276.144 | 70.4122 | 14.7 | 0.28 |
| polarC18 | 276.144 | 53.219 | 14.7 | 0.28 |
| polarC18 | 276.144 | 111.8814 | 14.7 | 0.28 |
| polarC18 | 276.144 | 81.4412 | 14.7 | 0.28 |
| polarC18 | 276.144 | 109.8277 | 14.7 | 0.28 |
| polarC18 | 276.144 | 251.7759 | 14.7 | 0.28 |
| polarC18 | 276.144 | 137.6462 | 14.7 | 0.28 |
| polarC18 | 276.144 | 197.6533 | 14.7 | 0.28 |
| polarC18 | 276.144 | 117.8511 | 14.6 | 0.28 |
| polarC18 | 276.144 | 113.7611 | 14.6 | 0.28 |
| polarC18 | 276.144 | 165.1196 | 14.6 | 0.28 |
| polarC18 | 276.144 | 67.3785 | 14.6 | 0.28 |
| polarC18 | 276.144 | 129.6602 | 14.6 | 0.28 |
| polarC18 | 276.144 | 151.1123 | 14.6 | 0.28 |
| polarC18 | 276.144 | 64.6842 | 14.6 | 0.28 |
| polarC18 | 276.144 | 278.1584 | 14.6 | 0.28 |
| polarC18 | 276.144 | 76.9191 | 14.6 | 0.28 |
| polarC18 | 276.144 | 159.6217 | 14.6 | 0.28 |
| polarC18 | 276.144 | 100.9039 | 14.6 | 0.28 |
| polarC18 | 276.144 | 65.1653 | 14.6 | 0.28 |
| polarC18 | 276.144 | 98.633 | 14.6 | 0.28 |

| | | | | |
|---|---|---|---|---|
| polarC18 | 276.144 | 211.4087 | 14.6 | 0.28 |
| polarC18 | 276.144 | 262.3869 | 14.6 | 0.28 |
| polarC18 | 276.144 | 65.9199 | 14.6 | 0.28 |
| polarC18 | 276.144 | 56.9547 | 14.6 | 0.28 |
| polarC18 | 276.144 | 220.339 | 14.6 | 0.28 |
| polarC18 | 276.144 | 256.0993 | 14.6 | 0.28 |
| polarC18 | 276.144 | 120.973 | 14.6 | 0.28 |
| polarC18 | 276.144 | 78.25 | 14.6 | 0.28 |
| polarC18 | 276.144 | 191.7984 | 14.6 | 0.28 |
| polarC18 | 276.144 | 72.8491 | 14.5 | 0.28 |
| polarC18 | 276.144 | 83.5299 | 14.5 | 0.28 |
| polarC18 | 276.144 | 111.4472 | 14.5 | 0.28 |
| polarC18 | 276.144 | 125.2677 | 14.5 | 0.28 |
| polarC18 | 276.144 | 62.4035 | 14.5 | 0.28 |
| polarC18 | 276.144 | 80.6185 | 14.5 | 0.28 |
| polarC18 | 276.144 | 115.3095 | 14.5 | 0.28 |
| polarC18 | 276.144 | 58.8444 | 14.5 | 0.28 |
| polarC18 | 276.144 | 173.4113 | 14.5 | 0.28 |
| polarC18 | 276.144 | 52.3257 | 14.5 | 0.28 |
| polarC18 | 276.144 | 52.3856 | 14.5 | 0.28 |
| polarC18 | 276.144 | 197.4406 | 14.5 | 0.28 |
| polarC18 | 276.144 | 214.3111 | 14.5 | 0.28 |
| polarC18 | 276.144 | 56.7832 | 14.5 | 0.28 |
| polarC18 | 276.144 | 94.6879 | 14.5 | 0.28 |
| polarC18 | 276.144 | 266.9072 | 14.5 | 0.28 |
| polarC18 | 276.144 | 138.119 | 14.5 | 0.28 |
| polarC18 | 276.144 | 52.6923 | 14.5 | 0.28 |
| polarC18 | 276.144 | 63.4004 | 14.5 | 0.28 |
| polarC18 | 276.144 | 212.0408 | 14.5 | 0.28 |
| polarC18 | 276.144 | 77.5638 | 14.5 | 0.28 |
| polarC18 | 276.144 | 78.0301 | 14.5 | 0.28 |
| polarC18 | 276.144 | 91.7254 | 14.5 | 0.28 |
| polarC18 | 276.144 | 94.3036 | 14.5 | 0.28 |
| polarC18 | 276.144 | 159.646 | 14.5 | 0.28 |
| polarC18 | 276.144 | 68.5198 | 14.5 | 0.28 |
| polarC18 | 276.144 | 58.0076 | 14.5 | 0.28 |
| polarC18 | 276.144 | 57.1952 | 14.5 | 0.28 |
| polarC18 | 276.144 | 54.5588 | 14.5 | 0.28 |
| polarC18 | 276.144 | 297.3288 | 14.5 | 0.28 |
| polarC18 | 276.144 | 248.1064 | 14.5 | 0.28 |
| polarC18 | 276.144 | 195.8365 | 14.5 | 0.28 |
| polarC18 | 276.144 | 136.8888 | 14.5 | 0.28 |

| | | | | |
|---|---|---|---|---|
| polarC18 | 276.144 | 146.6131 | 14.5 | 0.28 |
| polarC18 | 276.144 | 125.7854 | 14.5 | 0.28 |
| polarC18 | 276.144 | 94.3321 | 14.5 | 0.28 |
| polarC18 | 276.144 | 204.0711 | 14.5 | 0.28 |
| polarC18 | 276.144 | 102.6371 | 14.5 | 0.28 |
| polarC18 | 276.144 | 109.2197 | 14.5 | 0.28 |
| polarC18 | 276.144 | 96.9846 | 14.5 | 0.28 |
| polarC18 | 276.144 | 143.3974 | 14.5 | 0.28 |
| polarC18 | 276.144 | 222.8474 | 14.5 | 0.28 |
| polarC18 | 276.144 | 69.85 | 14.5 | 0.28 |
| polarC18 | 276.144 | 52.6332 | 14.5 | 0.28 |
| polarC18 | 276.144 | 90.0905 | 14.4 | 0.28 |
| polarC18 | 276.144 | 151.2882 | 14.4 | 0.28 |
| polarC18 | 276.144 | 129.4242 | 14.4 | 0.28 |
| polarC18 | 276.144 | 180.4197 | 14.4 | 0.28 |
| polarC18 | 276.144 | 53.0804 | 14.4 | 0.28 |
| polarC18 | 276.144 | 71.0627 | 14.4 | 0.28 |
| polarC18 | 276.144 | 122.6428 | 14.4 | 0.28 |
| polarC18 | 276.144 | 137.3885 | 14.4 | 0.28 |
| polarC18 | 276.144 | 67.3469 | 14.4 | 0.28 |
| polarC18 | 276.144 | 189.5972 | 14.4 | 0.28 |
| polarC18 | 276.144 | 90.0523 | 14.4 | 0.28 |
| polarC18 | 276.144 | 64.99 | 14.4 | 0.28 |
| polarC18 | 276.144 | 235.0597 | 14.4 | 0.28 |
| polarC18 | 276.144 | 221.2638 | 14.4 | 0.28 |
| polarC18 | 276.144 | 187.2721 | 14.4 | 0.28 |
| polarC18 | 276.144 | 66.0457 | 14.4 | 0.28 |
| polarC18 | 276.144 | 206.5152 | 14.4 | 0.28 |
| polarC18 | 276.144 | 91.5933 | 14.4 | 0.28 |
| polarC18 | 276.144 | 111.2491 | 14.4 | 0.28 |
| polarC18 | 276.144 | 79.8653 | 14.4 | 0.28 |
| polarC18 | 276.144 | 55.7146 | 14.4 | 0.28 |
| polarC18 | 276.144 | 68.642 | 14.4 | 0.28 |
| polarC18 | 276.144 | 199.4794 | 14.4 | 0.28 |
| polarC18 | 276.144 | 93.6823 | 14.4 | 0.28 |
| polarC18 | 276.144 | 255.8278 | 14.4 | 0.28 |
| polarC18 | 276.144 | 154.8552 | 14.4 | 0.28 |
| polarC18 | 276.144 | 92.1118 | 14.4 | 0.28 |
| polarC18 | 276.144 | 78.2724 | 14.4 | 0.28 |
| polarC18 | 276.144 | 149.1365 | 14.4 | 0.28 |
| polarC18 | 276.144 | 273.4602 | 14.4 | 0.28 |
| polarC18 | 276.144 | 106.5354 | 14.4 | 0.28 |

| | | | | |
|---|---|---|---|---|
| polarC18 | 276.144 | 62.6045 | 14.4 | 0.28 |
| polarC18 | 276.144 | 135.957 | 14.4 | 0.28 |
| polarC18 | 276.144 | 121.1697 | 14.3 | 0.28 |
| polarC18 | 276.144 | 65.7352 | 14.3 | 0.28 |
| polarC18 | 276.144 | 71.6429 | 14.3 | 0.28 |
| polarC18 | 276.144 | 265.9325 | 14.3 | 0.28 |
| polarC18 | 276.144 | 119.7609 | 14.3 | 0.28 |
| polarC18 | 276.144 | 120.8476 | 14.3 | 0.28 |
| polarC18 | 276.144 | 168.1358 | 14.3 | 0.28 |
| polarC18 | 276.144 | 95.9638 | 14.3 | 0.28 |
| polarC18 | 276.144 | 107.3082 | 14.3 | 0.28 |
| polarC18 | 276.144 | 270.1612 | 14.3 | 0.28 |
| polarC18 | 276.144 | 62.1389 | 14.3 | 0.28 |
| polarC18 | 276.144 | 136.5167 | 14.3 | 0.28 |
| polarC18 | 276.144 | 51.1368 | 14.3 | 0.28 |
| polarC18 | 276.144 | 70.2673 | 14.3 | 0.28 |
| polarC18 | 276.144 | 175.3635 | 14.3 | 0.28 |
| polarC18 | 276.144 | 72.7403 | 14.3 | 0.28 |
| polarC18 | 276.144 | 94.003 | 14.3 | 0.28 |
| polarC18 | 276.144 | 150.3131 | 14.3 | 0.27 |
| polarC18 | 276.144 | 258.4057 | 14.3 | 0.27 |
| polarC18 | 276.144 | 72.4218 | 14.3 | 0.27 |
| polarC18 | 276.144 | 88.929 | 14.3 | 0.27 |
| polarC18 | 276.144 | 50.5218 | 14.3 | 0.27 |
| polarC18 | 276.144 | 94.8655 | 14.3 | 0.27 |
| polarC18 | 276.144 | 83.5244 | 14.3 | 0.27 |
| polarC18 | 276.144 | 226.1565 | 14.3 | 0.27 |
| polarC18 | 276.144 | 87.4525 | 14.3 | 0.27 |
| polarC18 | 276.144 | 73.1663 | 14.3 | 0.27 |
| polarC18 | 276.144 | 51.7425 | 14.3 | 0.27 |
| polarC18 | 276.144 | 94.2768 | 14.3 | 0.27 |
| polarC18 | 276.144 | 68.9397 | 14.3 | 0.27 |
| polarC18 | 276.144 | 172.6555 | 14.3 | 0.27 |
| polarC18 | 276.144 | 201.072 | 14.3 | 0.27 |
| polarC18 | 276.144 | 90.4412 | 14.3 | 0.27 |
| polarC18 | 276.144 | 85.755 | 14.3 | 0.27 |
| polarC18 | 276.144 | 180.8265 | 14.3 | 0.27 |
| polarC18 | 276.144 | 96.6192 | 14.3 | 0.27 |
| polarC18 | 276.144 | 216.2714 | 14.3 | 0.27 |
| polarC18 | 276.144 | 120.897 | 14.3 | 0.27 |
| polarC18 | 276.144 | 90.3205 | 14.3 | 0.27 |
| polarC18 | 276.144 | 164.4778 | 14.2 | 0.27 |

| | | | | |
|---|---|---|---|---|
| polarC18 | 276.144 | 60.6325 | 14.2 | 0.27 |
| polarC18 | 276.144 | 148.3849 | 14.2 | 0.27 |
| polarC18 | 276.144 | 82.7211 | 14.2 | 0.27 |
| polarC18 | 276.144 | 135.8773 | 14.2 | 0.27 |
| polarC18 | 276.144 | 76.7885 | 14.2 | 0.27 |
| polarC18 | 276.144 | 156.9227 | 14.2 | 0.27 |
| polarC18 | 276.144 | 51.5021 | 14.2 | 0.27 |
| polarC18 | 276.144 | 298.572 | 14.2 | 0.27 |
| polarC18 | 276.144 | 60.3763 | 14.2 | 0.27 |
| polarC18 | 276.144 | 107.7423 | 14.2 | 0.27 |
| polarC18 | 276.144 | 149.0804 | 14.2 | 0.27 |
| polarC18 | 276.144 | 109.2968 | 14.2 | 0.27 |
| polarC18 | 276.144 | 109.97 | 14.2 | 0.27 |
| polarC18 | 276.144 | 174.4931 | 14.2 | 0.27 |
| polarC18 | 276.144 | 138.9289 | 14.2 | 0.27 |
| polarC18 | 276.144 | 194.6545 | 14.2 | 0.27 |
| polarC18 | 276.144 | 170.7532 | 14.2 | 0.27 |
| polarC18 | 276.144 | 244.5743 | 14.2 | 0.27 |
| polarC18 | 276.144 | 246.2816 | 14.2 | 0.27 |
| polarC18 | 276.144 | 94.9549 | 14.2 | 0.27 |
| polarC18 | 276.144 | 123.3517 | 14.2 | 0.27 |
| polarC18 | 276.144 | 130.3104 | 14.2 | 0.27 |
| polarC18 | 276.144 | 249.9634 | 14.2 | 0.27 |
| polarC18 | 276.144 | 235.2919 | 14.2 | 0.27 |
| polarC18 | 276.144 | 137.8128 | 14.2 | 0.27 |
| polarC18 | 276.144 | 211.9851 | 14.2 | 0.27 |
| polarC18 | 276.144 | 185.1129 | 14.2 | 0.27 |
| polarC18 | 276.144 | 132.7325 | 14.2 | 0.27 |
| polarC18 | 276.144 | 300.337 | 14.2 | 0.27 |
| polarC18 | 276.144 | 187.6763 | 14.2 | 0.27 |
| polarC18 | 276.144 | 102.395 | 14.2 | 0.27 |
| polarC18 | 276.144 | 108.0465 | 14.2 | 0.27 |
| polarC18 | 276.144 | 186.0472 | 14.2 | 0.27 |
| polarC18 | 276.144 | 61.6965 | 14.2 | 0.27 |
| polarC18 | 276.144 | 210.8062 | 14.2 | 0.27 |
| polarC18 | 276.144 | 247.055 | 14.2 | 0.27 |
| polarC18 | 276.144 | 55.4937 | 14.2 | 0.27 |
| polarC18 | 276.144 | 256.5226 | 14.2 | 0.27 |
| polarC18 | 276.144 | 260.7335 | 14.2 | 0.27 |
| polarC18 | 276.144 | 95.4319 | 14.2 | 0.27 |
| polarC18 | 276.144 | 278.3008 | 14.2 | 0.27 |
| polarC18 | 276.144 | 76.5316 | 14.2 | 0.27 |

| | | | | |
|---|---|---|---|---|
| polarC18 | 276.144 | 120.7374 | 14.2 | 0.27 |
| polarC18 | 276.144 | 130.8735 | 14.2 | 0.27 |
| polarC18 | 276.144 | 52.3228 | 14.2 | 0.27 |
| polarC18 | 276.144 | 283.6616 | 14.2 | 0.27 |
| polarC18 | 276.144 | 118.4777 | 14.2 | 0.27 |
| polarC18 | 276.144 | 241.1458 | 14.2 | 0.27 |
| polarC18 | 276.144 | 167.2578 | 14.1 | 0.27 |
| polarC18 | 276.144 | 160.3762 | 14.1 | 0.27 |
| polarC18 | 276.144 | 60.6607 | 14.1 | 0.27 |
| polarC18 | 276.144 | 95.3656 | 14.1 | 0.27 |
| polarC18 | 276.144 | 52.3538 | 14.1 | 0.27 |
| polarC18 | 276.144 | 68.3198 | 14.1 | 0.27 |
| polarC18 | 276.144 | 129.2029 | 14.1 | 0.27 |
| polarC18 | 276.144 | 81.1005 | 14.1 | 0.27 |
| polarC18 | 276.144 | 103.7252 | 14.1 | 0.27 |
| polarC18 | 276.144 | 187.8958 | 14.1 | 0.27 |
| polarC18 | 276.144 | 50.7339 | 14.1 | 0.27 |
| polarC18 | 276.144 | 118.083 | 14.1 | 0.27 |
| polarC18 | 276.144 | 229.6525 | 14.1 | 0.27 |
| polarC18 | 276.144 | 60.2043 | 14.1 | 0.27 |
| polarC18 | 276.144 | 118.7121 | 14.1 | 0.27 |
| polarC18 | 276.144 | 142.0083 | 14.1 | 0.27 |
| polarC18 | 276.144 | 127.6237 | 14.1 | 0.27 |
| polarC18 | 276.144 | 77.5307 | 14.1 | 0.27 |
| polarC18 | 276.144 | 184.8378 | 14.1 | 0.27 |
| polarC18 | 276.144 | 224.3702 | 14.1 | 0.27 |
| polarC18 | 276.144 | 118.2918 | 14.1 | 0.27 |
| polarC18 | 276.144 | 51.4636 | 14.1 | 0.27 |
| polarC18 | 276.144 | 64.5891 | 14.1 | 0.27 |
| polarC18 | 276.144 | 229.3236 | 14.1 | 0.27 |
| polarC18 | 276.144 | 134.7984 | 14.1 | 0.27 |
| polarC18 | 276.144 | 188.5001 | 14.1 | 0.27 |
| polarC18 | 276.144 | 56.9421 | 14.1 | 0.27 |
| polarC18 | 276.144 | 108.2533 | 14.1 | 0.27 |
| polarC18 | 276.144 | 147.4587 | 14.1 | 0.27 |
| polarC18 | 276.144 | 67.9388 | 14.1 | 0.27 |
| polarC18 | 276.144 | 199.0738 | 14.1 | 0.27 |
| polarC18 | 276.144 | 100.9335 | 14.1 | 0.27 |
| polarC18 | 276.144 | 63.424 | 14.1 | 0.27 |
| polarC18 | 276.144 | 283.3523 | 14.1 | 0.27 |
| polarC18 | 276.144 | 100.0894 | 14.1 | 0.27 |
| polarC18 | 276.144 | 119.9104 | 14.1 | 0.27 |

| | | | | |
|---|---|---|---|---|
| polarC18 | 276.144 | 272.2834 | 14.1 | 0.27 |
| polarC18 | 276.144 | 59.161 | 14.1 | 0.27 |
| polarC18 | 276.144 | 189.7356 | 14.1 | 0.27 |
| polarC18 | 276.144 | 105.1778 | 14.1 | 0.27 |
| polarC18 | 276.144 | 52.093 | 14.1 | 0.27 |
| polarC18 | 276.144 | 145.2026 | 14.1 | 0.27 |
| polarC18 | 276.144 | 215.5261 | 14.1 | 0.27 |
| polarC18 | 276.144 | 163.3692 | 14.1 | 0.27 |
| polarC18 | 276.144 | 231.3053 | 14.1 | 0.27 |
| polarC18 | 276.144 | 54.9741 | 14.1 | 0.27 |
| polarC18 | 276.144 | 89.9802 | 14 | 0.27 |
| polarC18 | 276.144 | 200.5817 | 14 | 0.27 |
| polarC18 | 276.144 | 97.9289 | 14 | 0.27 |
| polarC18 | 276.144 | 294.1423 | 14 | 0.27 |
| polarC18 | 276.144 | 281.7046 | 14 | 0.27 |
| polarC18 | 276.144 | 147.2929 | 14 | 0.27 |
| polarC18 | 276.144 | 77.1836 | 14 | 0.27 |
| polarC18 | 276.144 | 111.792 | 14 | 0.27 |
| polarC18 | 276.144 | 185.7394 | 14 | 0.27 |
| polarC18 | 276.144 | 53.0721 | 14 | 0.27 |
| polarC18 | 276.144 | 153.2918 | 14 | 0.27 |
| polarC18 | 276.144 | 53.0761 | 14 | 0.27 |
| polarC18 | 276.144 | 141.2802 | 14 | 0.27 |
| polarC18 | 276.144 | 161.8116 | 14 | 0.27 |
| polarC18 | 276.144 | 93.7518 | 14 | 0.27 |
| polarC18 | 276.144 | 170.0873 | 14 | 0.27 |
| polarC18 | 276.144 | 89.8542 | 14 | 0.27 |
| polarC18 | 276.144 | 252.7166 | 14 | 0.27 |
| polarC18 | 276.144 | 111.9294 | 14 | 0.27 |
| polarC18 | 276.144 | 175.8352 | 14 | 0.27 |
| polarC18 | 276.144 | 273.3661 | 14 | 0.27 |
| polarC18 | 276.144 | 94.4424 | 14 | 0.27 |
| polarC18 | 276.144 | 68.4276 | 14 | 0.27 |
| polarC18 | 276.144 | 112.3667 | 14 | 0.27 |
| polarC18 | 276.144 | 297.373 | 14 | 0.27 |
| polarC18 | 276.144 | 113.3413 | 14 | 0.27 |
| polarC18 | 276.144 | 118.9873 | 14 | 0.27 |
| polarC18 | 276.144 | 67.9526 | 14 | 0.27 |
| polarC18 | 276.144 | 69.7863 | 14 | 0.27 |
| polarC18 | 276.144 | 288.1011 | 14 | 0.27 |
| polarC18 | 276.144 | 109.4555 | 14 | 0.27 |
| polarC18 | 276.144 | 74.8033 | 14 | 0.27 |

| | | | | |
|---|---|---|---|---|
| polarC18 | 276.144 | 226.2985 | 14 | 0.27 |
| polarC18 | 276.144 | 123.9157 | 14 | 0.27 |
| polarC18 | 276.144 | 66.3818 | 14 | 0.27 |
| polarC18 | 276.144 | 117.6952 | 14 | 0.27 |
| polarC18 | 276.144 | 67.5869 | 14 | 0.27 |
| polarC18 | 276.144 | 75.2976 | 14 | 0.27 |
| polarC18 | 276.144 | 60.2222 | 13.9 | 0.27 |
| polarC18 | 276.144 | 227.5208 | 13.9 | 0.27 |
| polarC18 | 276.144 | 66.8631 | 13.9 | 0.27 |
| polarC18 | 276.144 | 53.5454 | 13.9 | 0.27 |
| polarC18 | 276.144 | 63.3318 | 13.9 | 0.27 |
| polarC18 | 276.144 | 206.4205 | 13.9 | 0.27 |
| polarC18 | 276.144 | 280.7319 | 13.9 | 0.27 |
| polarC18 | 276.144 | 166.0156 | 13.9 | 0.27 |
| polarC18 | 276.144 | 57.3562 | 13.9 | 0.27 |
| polarC18 | 276.144 | 106.2614 | 13.9 | 0.27 |
| polarC18 | 276.144 | 166.7687 | 13.9 | 0.27 |
| polarC18 | 276.144 | 94.5101 | 13.9 | 0.27 |
| polarC18 | 276.144 | 108.0868 | 13.9 | 0.27 |
| polarC18 | 276.144 | 74.947 | 13.9 | 0.27 |
| polarC18 | 276.144 | 118.0187 | 13.9 | 0.27 |
| polarC18 | 276.144 | 121.9475 | 13.9 | 0.27 |
| polarC18 | 276.144 | 198.4412 | 13.9 | 0.27 |
| polarC18 | 276.144 | 122.114 | 13.9 | 0.27 |
| polarC18 | 276.144 | 59.0639 | 13.9 | 0.27 |
| polarC18 | 276.144 | 94.5717 | 13.9 | 0.27 |
| polarC18 | 276.144 | 90.6877 | 13.9 | 0.27 |
| polarC18 | 276.144 | 232.8577 | 13.9 | 0.27 |
| polarC18 | 276.144 | 77.5849 | 13.9 | 0.27 |
| polarC18 | 276.144 | 94.2566 | 13.9 | 0.27 |
| polarC18 | 276.144 | 73.5751 | 13.9 | 0.27 |
| polarC18 | 276.144 | 179.3912 | 13.9 | 0.27 |
| polarC18 | 276.144 | 61.3303 | 13.9 | 0.27 |
| polarC18 | 276.144 | 148.3662 | 13.9 | 0.27 |
| polarC18 | 276.144 | 108.9948 | 13.9 | 0.27 |
| polarC18 | 276.144 | 125.0093 | 13.9 | 0.27 |
| polarC18 | 276.144 | 99.3985 | 13.9 | 0.27 |
| polarC18 | 276.144 | 251.5379 | 13.9 | 0.27 |
| polarC18 | 276.144 | 81.6907 | 13.9 | 0.27 |
| polarC18 | 276.144 | 155.9122 | 13.9 | 0.27 |
| polarC18 | 276.144 | 78.5047 | 13.9 | 0.27 |
| polarC18 | 276.144 | 74.4129 | 13.9 | 0.27 |

| | | | | |
|---|---|---|---|---|
| polarC18 | 276.144 | 89.9193 | 13.9 | 0.27 |
| polarC18 | 276.144 | 284.0104 | 13.9 | 0.27 |
| polarC18 | 276.144 | 82.5207 | 13.9 | 0.27 |
| polarC18 | 276.144 | 106.9081 | 13.9 | 0.27 |
| polarC18 | 276.144 | 130.7087 | 13.9 | 0.27 |
| polarC18 | 276.144 | 122.233 | 13.9 | 0.27 |
| polarC18 | 276.144 | 63.1424 | 13.9 | 0.27 |
| polarC18 | 276.144 | 182.7957 | 13.9 | 0.27 |
| polarC18 | 276.144 | 51.7755 | 13.9 | 0.27 |
| polarC18 | 276.144 | 119.5361 | 13.9 | 0.27 |
| polarC18 | 276.144 | 98.4934 | 13.9 | 0.27 |
| polarC18 | 276.144 | 300.9483 | 13.9 | 0.27 |
| polarC18 | 276.144 | 52.4757 | 13.9 | 0.27 |
| polarC18 | 276.144 | 153.172 | 13.9 | 0.27 |
| polarC18 | 276.144 | 58.4016 | 13.9 | 0.27 |
| polarC18 | 276.144 | 217.6471 | 13.9 | 0.27 |
| polarC18 | 276.144 | 60.5934 | 13.9 | 0.27 |
| polarC18 | 276.144 | 289.5253 | 13.9 | 0.27 |
| polarC18 | 276.144 | 172.0438 | 13.9 | 0.27 |
| polarC18 | 276.144 | 123.7164 | 13.9 | 0.27 |
| polarC18 | 276.144 | 104.9454 | 13.9 | 0.27 |
| polarC18 | 276.144 | 222.2974 | 13.9 | 0.27 |
| polarC18 | 276.144 | 89.1667 | 13.9 | 0.27 |
| polarC18 | 276.144 | 50.5185 | 13.9 | 0.27 |
| polarC18 | 276.144 | 108.8682 | 13.9 | 0.27 |
| polarC18 | 276.144 | 235.3477 | 13.9 | 0.27 |
| polarC18 | 276.144 | 63.1298 | 13.9 | 0.27 |
| polarC18 | 276.144 | 115.0209 | 13.9 | 0.27 |
| polarC18 | 276.144 | 94.4289 | 13.9 | 0.27 |
| polarC18 | 276.144 | 155.4034 | 13.9 | 0.27 |
| polarC18 | 276.144 | 134.7484 | 13.9 | 0.27 |
| polarC18 | 276.144 | 179.3562 | 13.9 | 0.27 |
| polarC18 | 276.144 | 157.4883 | 13.9 | 0.27 |
| polarC18 | 276.144 | 235.9244 | 13.9 | 0.27 |
| polarC18 | 276.144 | 130.5482 | 13.9 | 0.27 |
| polarC18 | 276.144 | 68.6512 | 13.9 | 0.27 |
| polarC18 | 276.144 | 76.4674 | 13.9 | 0.27 |
| polarC18 | 276.144 | 59.7616 | 13.9 | 0.27 |
| polarC18 | 276.144 | 138.003 | 13.8 | 0.27 |
| polarC18 | 276.144 | 166.5318 | 13.8 | 0.27 |
| polarC18 | 276.144 | 68.1279 | 13.8 | 0.27 |
| polarC18 | 276.144 | 226.4537 | 13.8 | 0.27 |

| | | | | |
|---|---|---|---|---|
| polarC18 | 276.144 | 125.4863 | 13.8 | 0.27 |
| polarC18 | 276.144 | 139.1609 | 13.8 | 0.27 |
| polarC18 | 276.144 | 279.2866 | 13.8 | 0.27 |
| polarC18 | 276.144 | 274.9155 | 13.8 | 0.27 |
| polarC18 | 276.144 | 106.9369 | 13.8 | 0.27 |
| polarC18 | 276.144 | 77.756 | 13.8 | 0.27 |
| polarC18 | 276.144 | 255.7855 | 13.8 | 0.27 |
| polarC18 | 276.144 | 63.6839 | 13.8 | 0.27 |
| polarC18 | 276.144 | 60.2959 | 13.8 | 0.27 |
| polarC18 | 276.144 | 102.271 | 13.8 | 0.27 |
| polarC18 | 276.144 | 195.054 | 13.8 | 0.27 |
| polarC18 | 276.144 | 168.6229 | 13.8 | 0.27 |
| polarC18 | 276.144 | 95.191 | 13.8 | 0.27 |
| polarC18 | 276.144 | 131.4126 | 13.8 | 0.27 |
| polarC18 | 276.144 | 117.7282 | 13.8 | 0.27 |
| polarC18 | 276.144 | 102.7506 | 13.8 | 0.27 |
| polarC18 | 276.144 | 161.1656 | 13.8 | 0.27 |
| polarC18 | 276.144 | 84.9702 | 13.8 | 0.27 |
| polarC18 | 276.144 | 99.0899 | 13.8 | 0.27 |
| polarC18 | 276.144 | 189.046 | 13.8 | 0.27 |
| polarC18 | 276.144 | 139.3438 | 13.8 | 0.27 |
| polarC18 | 276.144 | 71.1536 | 13.8 | 0.27 |
| polarC18 | 276.144 | 134.0584 | 13.8 | 0.27 |
| polarC18 | 276.144 | 50.3083 | 13.8 | 0.27 |
| polarC18 | 276.144 | 65.306 | 13.8 | 0.27 |
| polarC18 | 276.144 | 133.0175 | 13.8 | 0.27 |
| polarC18 | 276.144 | 81.3922 | 13.8 | 0.27 |
| polarC18 | 276.144 | 97.5802 | 13.8 | 0.27 |
| polarC18 | 276.144 | 62.4493 | 13.8 | 0.27 |
| polarC18 | 276.144 | 177.3413 | 13.8 | 0.27 |
| polarC18 | 276.144 | 92.019 | 13.8 | 0.27 |
| polarC18 | 276.144 | 81.312 | 13.8 | 0.27 |
| polarC18 | 276.144 | 122.9897 | 13.8 | 0.26 |
| polarC18 | 276.144 | 168.6496 | 13.8 | 0.26 |
| polarC18 | 276.144 | 63.5086 | 13.8 | 0.26 |
| polarC18 | 276.144 | 59.1835 | 13.8 | 0.26 |
| polarC18 | 276.144 | 257.9536 | 13.8 | 0.26 |
| polarC18 | 276.144 | 177.8921 | 13.8 | 0.26 |
| polarC18 | 276.144 | 72.1081 | 13.8 | 0.26 |
| polarC18 | 276.144 | 57.9792 | 13.8 | 0.26 |
| polarC18 | 276.144 | 225.0254 | 13.8 | 0.26 |
| polarC18 | 276.144 | 172.2858 | 13.8 | 0.26 |

| | | | | |
|---|---|---|---|---|
| polarC18 | 276.144 | 82.1299 | 13.8 | 0.26 |
| polarC18 | 276.144 | 51.0702 | 13.8 | 0.26 |
| polarC18 | 276.144 | 104.354 | 13.8 | 0.26 |
| polarC18 | 276.144 | 55.2692 | 13.8 | 0.26 |
| polarC18 | 276.144 | 270.067 | 13.8 | 0.26 |
| polarC18 | 276.144 | 69.2727 | 13.8 | 0.26 |
| polarC18 | 276.144 | 101.3378 | 13.8 | 0.26 |
| polarC18 | 276.144 | 82.7814 | 13.8 | 0.26 |
| polarC18 | 276.144 | 199.7811 | 13.7 | 0.26 |
| polarC18 | 276.144 | 83.9107 | 13.7 | 0.26 |
| polarC18 | 276.144 | 176.0718 | 13.7 | 0.26 |
| polarC18 | 276.144 | 206.6649 | 13.7 | 0.26 |
| polarC18 | 276.144 | 53.8076 | 13.7 | 0.26 |
| polarC18 | 276.144 | 214.6009 | 13.7 | 0.26 |
| polarC18 | 276.144 | 158.0838 | 13.7 | 0.26 |
| polarC18 | 276.144 | 58.9083 | 13.7 | 0.26 |
| polarC18 | 276.144 | 92.6245 | 13.7 | 0.26 |
| polarC18 | 276.144 | 98.59 | 13.7 | 0.26 |
| polarC18 | 276.144 | 85.2634 | 13.7 | 0.26 |
| polarC18 | 276.144 | 100.4667 | 13.7 | 0.26 |
| polarC18 | 276.144 | 52.373 | 13.7 | 0.26 |
| polarC18 | 276.144 | 98.5788 | 13.7 | 0.26 |
| polarC18 | 276.144 | 102.5199 | 13.7 | 0.26 |
| polarC18 | 276.144 | 197.193 | 13.7 | 0.26 |
| polarC18 | 276.144 | 55.105 | 13.7 | 0.26 |
| polarC18 | 276.144 | 122.3263 | 13.7 | 0.26 |
| polarC18 | 276.144 | 162.1532 | 13.7 | 0.26 |
| polarC18 | 276.144 | 90.1647 | 13.7 | 0.26 |
| polarC18 | 276.144 | 66.5964 | 13.7 | 0.26 |
| polarC18 | 276.144 | 80.6365 | 13.7 | 0.26 |
| polarC18 | 276.144 | 98.51 | 13.7 | 0.26 |
| polarC18 | 276.144 | 71.6394 | 13.7 | 0.26 |
| polarC18 | 276.144 | 154.1106 | 13.7 | 0.26 |
| polarC18 | 276.144 | 267.572 | 13.7 | 0.26 |
| polarC18 | 276.144 | 63.6772 | 13.7 | 0.26 |
| polarC18 | 276.144 | 83.4339 | 13.7 | 0.26 |
| polarC18 | 276.144 | 91.6499 | 13.7 | 0.26 |
| polarC18 | 276.144 | 198.9147 | 13.7 | 0.26 |
| polarC18 | 276.144 | 138.0697 | 13.7 | 0.26 |
| polarC18 | 276.144 | 53.9515 | 13.7 | 0.26 |
| polarC18 | 276.144 | 60.8349 | 13.7 | 0.26 |
| polarC18 | 276.144 | 170.4349 | 13.7 | 0.26 |

| | | | | |
|---|---|---|---|---|
| polarC18 | 276.144 | 51.6391 | 13.7 | 0.26 |
| polarC18 | 276.144 | 73.6395 | 13.7 | 0.26 |
| polarC18 | 276.144 | 135.0015 | 13.7 | 0.26 |
| polarC18 | 276.144 | 66.0001 | 13.7 | 0.26 |
| polarC18 | 276.144 | 74.0312 | 13.7 | 0.26 |
| polarC18 | 276.144 | 225.6467 | 13.7 | 0.26 |
| polarC18 | 276.144 | 163.5773 | 13.7 | 0.26 |
| polarC18 | 276.144 | 62.3847 | 13.7 | 0.26 |
| polarC18 | 276.144 | 214.3991 | 13.7 | 0.26 |
| polarC18 | 276.144 | 158.7924 | 13.7 | 0.26 |
| polarC18 | 276.144 | 68.8659 | 13.7 | 0.26 |
| polarC18 | 276.144 | 57.963 | 13.7 | 0.26 |
| polarC18 | 276.144 | 122.8049 | 13.7 | 0.26 |
| polarC18 | 276.144 | 61.7951 | 13.7 | 0.26 |
| polarC18 | 276.144 | 64.5127 | 13.7 | 0.26 |
| polarC18 | 276.144 | 136.4261 | 13.7 | 0.26 |
| polarC18 | 276.144 | 121.6922 | 13.7 | 0.26 |
| polarC18 | 276.144 | 118.6245 | 13.7 | 0.26 |
| polarC18 | 276.144 | 125.0969 | 13.7 | 0.26 |
| polarC18 | 276.144 | 251.0601 | 13.7 | 0.26 |
| polarC18 | 276.144 | 270.5147 | 13.7 | 0.26 |
| polarC18 | 276.144 | 253.2062 | 13.7 | 0.26 |
| polarC18 | 276.144 | 86.4751 | 13.7 | 0.26 |
| polarC18 | 276.144 | 64.6278 | 13.7 | 0.26 |
| polarC18 | 276.144 | 54.2975 | 13.7 | 0.26 |
| polarC18 | 276.144 | 125.8614 | 13.7 | 0.26 |
| polarC18 | 276.144 | 172.165 | 13.7 | 0.26 |
| polarC18 | 276.144 | 263.8035 | 13.7 | 0.26 |
| polarC18 | 276.144 | 106.9253 | 13.7 | 0.26 |
| polarC18 | 276.144 | 100.6758 | 13.7 | 0.26 |
| polarC18 | 276.144 | 82.634 | 13.6 | 0.26 |
| polarC18 | 276.144 | 264.8855 | 13.6 | 0.26 |
| polarC18 | 276.144 | 266.4888 | 13.6 | 0.26 |
| polarC18 | 276.144 | 225.0719 | 13.6 | 0.26 |
| polarC18 | 276.144 | 224.6675 | 13.6 | 0.26 |
| polarC18 | 276.144 | 67.2429 | 13.6 | 0.26 |
| polarC18 | 276.144 | 56.7935 | 13.6 | 0.26 |
| polarC18 | 276.144 | 182.818 | 13.6 | 0.26 |
| polarC18 | 276.144 | 127.5057 | 13.6 | 0.26 |
| polarC18 | 276.144 | 87.3509 | 13.6 | 0.26 |
| polarC18 | 276.144 | 60.8689 | 13.6 | 0.26 |
| polarC18 | 276.144 | 281.2415 | 13.6 | 0.26 |

| | | | | |
|---|---|---|---|---|
| polarC18 | 276.144 | 54.4159 | 13.6 | 0.26 |
| polarC18 | 276.144 | 113.1338 | 13.6 | 0.26 |
| polarC18 | 276.144 | 57.2715 | 13.6 | 0.26 |
| polarC18 | 276.144 | 54.6309 | 13.6 | 0.26 |
| polarC18 | 276.144 | 162.4979 | 13.6 | 0.26 |
| polarC18 | 276.144 | 154.1792 | 13.6 | 0.26 |
| polarC18 | 276.144 | 55.0211 | 13.6 | 0.26 |
| polarC18 | 276.144 | 71.3848 | 13.6 | 0.26 |
| polarC18 | 276.144 | 59.2571 | 13.6 | 0.26 |
| polarC18 | 276.144 | 80.7729 | 13.6 | 0.26 |
| polarC18 | 276.144 | 70.4834 | 13.6 | 0.26 |
| polarC18 | 276.144 | 145.9296 | 13.6 | 0.26 |
| polarC18 | 276.144 | 261.6925 | 13.6 | 0.26 |
| polarC18 | 276.144 | 62.6653 | 13.6 | 0.26 |
| polarC18 | 276.144 | 162.9638 | 13.6 | 0.26 |
| polarC18 | 276.144 | 83.0428 | 13.6 | 0.26 |
| polarC18 | 276.144 | 157.9594 | 13.6 | 0.26 |
| polarC18 | 276.144 | 71.881 | 13.6 | 0.26 |
| polarC18 | 276.144 | 228.492 | 13.6 | 0.26 |
| polarC18 | 276.144 | 60.5671 | 13.6 | 0.26 |
| polarC18 | 276.144 | 228.877 | 13.6 | 0.26 |
| polarC18 | 276.144 | 67.6282 | 13.6 | 0.26 |
| polarC18 | 276.144 | 66.2706 | 13.6 | 0.26 |
| polarC18 | 276.144 | 119.8257 | 13.6 | 0.26 |
| polarC18 | 276.144 | 93.2421 | 13.6 | 0.26 |
| polarC18 | 276.144 | 180.6953 | 13.6 | 0.26 |
| polarC18 | 276.144 | 294.0919 | 13.6 | 0.26 |
| polarC18 | 276.144 | 135.7897 | 13.6 | 0.26 |
| polarC18 | 276.144 | 147.8134 | 13.6 | 0.26 |
| polarC18 | 276.144 | 67.1812 | 13.6 | 0.26 |
| polarC18 | 276.144 | 78.5733 | 13.6 | 0.26 |
| polarC18 | 276.144 | 62.3126 | 13.6 | 0.26 |
| polarC18 | 276.144 | 56.3399 | 13.5 | 0.26 |
| polarC18 | 276.144 | 53.4516 | 13.5 | 0.26 |
| polarC18 | 276.144 | 165.5435 | 13.5 | 0.26 |
| polarC18 | 276.144 | 298.757 | 13.5 | 0.26 |
| polarC18 | 276.144 | 216.4928 | 13.5 | 0.26 |
| polarC18 | 276.144 | 143.0723 | 13.5 | 0.26 |
| polarC18 | 276.144 | 74.7554 | 13.5 | 0.26 |
| polarC18 | 276.144 | 76.7165 | 13.5 | 0.26 |
| polarC18 | 276.144 | 56.072 | 13.5 | 0.26 |
| polarC18 | 276.144 | 103.6899 | 13.5 | 0.26 |

| | | | | |
|---|---|---|---|---|
| polarC18 | 276.144 | 117.6795 | 13.5 | 0.26 |
| polarC18 | 276.144 | 149.6516 | 13.5 | 0.26 |
| polarC18 | 276.144 | 57.3041 | 13.5 | 0.26 |
| polarC18 | 276.144 | 156.3595 | 13.5 | 0.26 |
| polarC18 | 276.144 | 118.0432 | 13.5 | 0.26 |
| polarC18 | 276.144 | 84.4391 | 13.5 | 0.26 |
| polarC18 | 276.144 | 300.0859 | 13.5 | 0.26 |
| polarC18 | 276.144 | 63.612 | 13.5 | 0.26 |
| polarC18 | 276.144 | 63.2151 | 13.5 | 0.26 |
| polarC18 | 276.144 | 155.0594 | 13.5 | 0.26 |
| polarC18 | 276.144 | 297.6403 | 13.5 | 0.26 |
| polarC18 | 276.144 | 66.5354 | 13.5 | 0.26 |
| polarC18 | 276.144 | 56.1232 | 13.5 | 0.26 |
| polarC18 | 276.144 | 166.4553 | 13.5 | 0.26 |
| polarC18 | 276.144 | 217.7035 | 13.5 | 0.26 |
| polarC18 | 276.144 | 175.7798 | 13.5 | 0.26 |
| polarC18 | 276.144 | 219.3223 | 13.5 | 0.26 |
| polarC18 | 276.144 | 140.5549 | 13.5 | 0.26 |
| polarC18 | 276.144 | 61.93 | 13.5 | 0.26 |
| polarC18 | 276.144 | 242.4971 | 13.5 | 0.26 |
| polarC18 | 276.144 | 224.9852 | 13.5 | 0.26 |
| polarC18 | 276.144 | 146.4547 | 13.5 | 0.26 |
| polarC18 | 276.144 | 97.9376 | 13.5 | 0.26 |
| polarC18 | 276.144 | 57.7552 | 13.5 | 0.26 |
| polarC18 | 276.144 | 211.4614 | 13.5 | 0.26 |
| polarC18 | 276.144 | 146.1561 | 13.5 | 0.26 |
| polarC18 | 276.144 | 56.2356 | 13.5 | 0.26 |
| polarC18 | 276.144 | 54.6129 | 13.5 | 0.26 |
| polarC18 | 276.144 | 99.4231 | 13.5 | 0.26 |
| polarC18 | 276.144 | 163.9765 | 13.5 | 0.26 |
| polarC18 | 276.144 | 183.6334 | 13.5 | 0.26 |
| polarC18 | 276.144 | 138.1431 | 13.5 | 0.26 |
| polarC18 | 276.144 | 192.2648 | 13.5 | 0.26 |
| polarC18 | 276.144 | 93.3742 | 13.5 | 0.26 |
| polarC18 | 276.144 | 57.4698 | 13.5 | 0.26 |
| polarC18 | 276.144 | 152.3963 | 13.5 | 0.26 |
| polarC18 | 276.144 | 104.2187 | 13.5 | 0.26 |
| polarC18 | 276.144 | 82.3437 | 13.4 | 0.26 |
| polarC18 | 276.144 | 99.2799 | 13.4 | 0.26 |
| polarC18 | 276.144 | 82.8325 | 13.4 | 0.26 |
| polarC18 | 276.144 | 144.2132 | 13.4 | 0.26 |
| polarC18 | 276.144 | 82.4181 | 13.4 | 0.26 |

| | | | | |
|---|---|---|---|---|
| polarC18 | 276.144 | 89.9708 | 13.4 | 0.26 |
| polarC18 | 276.144 | 78.4713 | 13.4 | 0.26 |
| polarC18 | 276.144 | 116.7035 | 13.4 | 0.26 |
| polarC18 | 276.144 | 287.6707 | 13.4 | 0.26 |
| polarC18 | 276.144 | 188.3784 | 13.4 | 0.26 |
| polarC18 | 276.144 | 92.3261 | 13.4 | 0.26 |
| polarC18 | 276.144 | 111.5224 | 13.4 | 0.26 |
| polarC18 | 276.144 | 93.1255 | 13.4 | 0.26 |
| polarC18 | 276.144 | 133.3357 | 13.4 | 0.26 |
| polarC18 | 276.144 | 135.3411 | 13.4 | 0.26 |
| polarC18 | 276.144 | 148.1077 | 13.4 | 0.26 |
| polarC18 | 276.144 | 70.0033 | 13.4 | 0.26 |
| polarC18 | 276.144 | 123.2006 | 13.4 | 0.26 |
| polarC18 | 276.144 | 274.1957 | 13.4 | 0.26 |
| polarC18 | 276.144 | 65.5271 | 13.4 | 0.26 |
| polarC18 | 276.144 | 61.4342 | 13.4 | 0.26 |
| polarC18 | 276.144 | 55.0131 | 13.4 | 0.26 |
| polarC18 | 276.144 | 196.2391 | 13.4 | 0.26 |
| polarC18 | 276.144 | 60.6576 | 13.4 | 0.26 |
| polarC18 | 276.144 | 53.5666 | 13.4 | 0.26 |
| polarC18 | 276.144 | 120.3917 | 13.4 | 0.26 |
| polarC18 | 276.144 | 186.4293 | 13.4 | 0.26 |
| polarC18 | 276.144 | 61.0778 | 13.4 | 0.26 |
| polarC18 | 276.144 | 150.1684 | 13.4 | 0.26 |
| polarC18 | 276.144 | 50.9537 | 13.4 | 0.26 |
| polarC18 | 276.144 | 97.1582 | 13.4 | 0.26 |
| polarC18 | 276.144 | 64.7896 | 13.4 | 0.26 |
| polarC18 | 276.144 | 59.0842 | 13.4 | 0.26 |
| polarC18 | 276.144 | 144.9083 | 13.4 | 0.26 |
| polarC18 | 276.144 | 177.807 | 13.4 | 0.26 |
| polarC18 | 276.144 | 58.2733 | 13.4 | 0.26 |
| polarC18 | 276.144 | 203.5155 | 13.4 | 0.26 |
| polarC18 | 276.144 | 173.0463 | 13.4 | 0.26 |
| polarC18 | 276.144 | 126.3829 | 13.4 | 0.26 |
| polarC18 | 276.144 | 165.9856 | 13.4 | 0.26 |
| polarC18 | 276.144 | 128.2993 | 13.4 | 0.26 |
| polarC18 | 276.144 | 85.1723 | 13.4 | 0.26 |
| polarC18 | 276.144 | 141.9599 | 13.4 | 0.26 |
| polarC18 | 276.144 | 265.9695 | 13.4 | 0.26 |
| polarC18 | 276.144 | 215.9142 | 13.4 | 0.26 |
| polarC18 | 276.144 | 92.4011 | 13.4 | 0.26 |
| polarC18 | 276.144 | 60.301 | 13.4 | 0.26 |

| | | | | |
|---|---|---|---|---|
| polarC18 | 276.144 | 51.7149 | 13.4 | 0.26 |
| polarC18 | 276.144 | 67.8124 | 13.4 | 0.26 |
| polarC18 | 276.144 | 209.7373 | 13.4 | 0.26 |
| polarC18 | 276.144 | 62.712 | 13.4 | 0.26 |
| polarC18 | 276.144 | 256.0022 | 13.4 | 0.26 |
| polarC18 | 276.144 | 211.0888 | 13.4 | 0.26 |
| polarC18 | 276.144 | 110.043 | 13.4 | 0.26 |
| polarC18 | 276.144 | 51.6071 | 13.4 | 0.26 |
| polarC18 | 276.144 | 182.5495 | 13.4 | 0.26 |
| polarC18 | 276.144 | 138.8534 | 13.4 | 0.26 |
| polarC18 | 276.144 | 137.8987 | 13.4 | 0.26 |
| polarC18 | 276.144 | 92.9391 | 13.4 | 0.26 |
| polarC18 | 276.144 | 210.6798 | 13.3 | 0.26 |
| polarC18 | 276.144 | 147.8689 | 13.3 | 0.26 |
| polarC18 | 276.144 | 93.4732 | 13.3 | 0.26 |
| polarC18 | 276.144 | 223.6416 | 13.3 | 0.26 |
| polarC18 | 276.144 | 81.2768 | 13.3 | 0.26 |
| polarC18 | 276.144 | 53.8144 | 13.3 | 0.26 |
| polarC18 | 276.144 | 234.3477 | 13.3 | 0.26 |
| polarC18 | 276.144 | 155.2196 | 13.3 | 0.26 |
| polarC18 | 276.144 | 62.5656 | 13.3 | 0.26 |
| polarC18 | 276.144 | 92.6583 | 13.3 | 0.26 |
| polarC18 | 276.144 | 139.366 | 13.3 | 0.26 |
| polarC18 | 276.144 | 130.0991 | 13.3 | 0.26 |
| polarC18 | 276.144 | 230.0144 | 13.3 | 0.26 |
| polarC18 | 276.144 | 227.023 | 13.3 | 0.26 |
| polarC18 | 276.144 | 213.2853 | 13.3 | 0.26 |
| polarC18 | 276.144 | 199.6219 | 13.3 | 0.26 |
| polarC18 | 276.144 | 113.0891 | 13.3 | 0.26 |
| polarC18 | 276.144 | 122.1979 | 13.3 | 0.26 |
| polarC18 | 276.144 | 155.1471 | 13.3 | 0.26 |
| polarC18 | 276.144 | 57.1112 | 13.3 | 0.26 |
| polarC18 | 276.144 | 147.9373 | 13.3 | 0.26 |
| polarC18 | 276.144 | 85.2779 | 13.3 | 0.26 |
| polarC18 | 276.144 | 235.2198 | 13.3 | 0.26 |
| polarC18 | 276.144 | 58.3387 | 13.3 | 0.26 |
| polarC18 | 276.144 | 60.1912 | 13.3 | 0.26 |
| polarC18 | 276.144 | 70.2273 | 13.3 | 0.26 |
| polarC18 | 276.144 | 87.8239 | 13.3 | 0.26 |
| polarC18 | 276.144 | 70.6943 | 13.3 | 0.26 |
| polarC18 | 276.144 | 270.277 | 13.3 | 0.26 |
| polarC18 | 276.144 | 106.7081 | 13.3 | 0.26 |

| | | | | |
|---|---|---|---|---|
| polarC18 | 276.144 | 75.1783 | 13.3 | 0.26 |
| polarC18 | 276.144 | 89.1141 | 13.3 | 0.26 |
| polarC18 | 276.144 | 82.9595 | 13.3 | 0.26 |
| polarC18 | 276.144 | 281.0407 | 13.3 | 0.26 |
| polarC18 | 276.144 | 113.0113 | 13.3 | 0.26 |
| polarC18 | 276.144 | 236.4486 | 13.3 | 0.26 |
| polarC18 | 276.144 | 160.892 | 13.3 | 0.26 |
| polarC18 | 276.144 | 121.729 | 13.3 | 0.26 |
| polarC18 | 276.144 | 141.5844 | 13.3 | 0.26 |
| polarC18 | 276.144 | 243.3221 | 13.3 | 0.26 |
| polarC18 | 276.144 | 63.3659 | 13.3 | 0.26 |
| polarC18 | 276.144 | 85.6326 | 13.3 | 0.26 |
| polarC18 | 276.144 | 73.6236 | 13.3 | 0.26 |
| polarC18 | 276.144 | 74.2436 | 13.3 | 0.26 |
| polarC18 | 276.144 | 228.742 | 13.3 | 0.26 |
| polarC18 | 276.144 | 268.5063 | 13.3 | 0.26 |
| polarC18 | 276.144 | 159.7347 | 13.3 | 0.26 |
| polarC18 | 276.144 | 144.3002 | 13.3 | 0.26 |
| polarC18 | 276.144 | 71.0417 | 13.3 | 0.26 |
| polarC18 | 276.144 | 171.4148 | 13.3 | 0.26 |
| polarC18 | 276.144 | 55.3043 | 13.3 | 0.26 |
| polarC18 | 276.144 | 52.7872 | 13.3 | 0.25 |
| polarC18 | 276.144 | 126.9007 | 13.3 | 0.25 |
| polarC18 | 276.144 | 74.4418 | 13.3 | 0.25 |
| polarC18 | 276.144 | 280.0856 | 13.3 | 0.25 |
| polarC18 | 276.144 | 52.9274 | 13.3 | 0.25 |
| polarC18 | 276.144 | 58.4308 | 13.2 | 0.25 |
| polarC18 | 276.144 | 57.4184 | 13.2 | 0.25 |
| polarC18 | 276.144 | 70.7516 | 13.2 | 0.25 |
| polarC18 | 276.144 | 171.5638 | 13.2 | 0.25 |
| polarC18 | 276.144 | 63.6618 | 13.2 | 0.25 |
| polarC18 | 276.144 | 149.1923 | 13.2 | 0.25 |
| polarC18 | 276.144 | 232.1582 | 13.2 | 0.25 |
| polarC18 | 276.144 | 92.8945 | 13.2 | 0.25 |
| polarC18 | 276.144 | 178.4813 | 13.2 | 0.25 |
| polarC18 | 276.144 | 77.4934 | 13.2 | 0.25 |
| polarC18 | 276.144 | 70.9311 | 13.2 | 0.25 |
| polarC18 | 276.144 | 81.4349 | 13.2 | 0.25 |
| polarC18 | 276.144 | 280.1301 | 13.2 | 0.25 |
| polarC18 | 276.144 | 115.2913 | 13.2 | 0.25 |
| polarC18 | 276.144 | 124.5061 | 13.2 | 0.25 |
| polarC18 | 276.144 | 146.9494 | 13.2 | 0.25 |

| | | | | |
|---|---|---|---|---|
| polarC18 | 276.144 | 109.9288 | 13.2 | 0.25 |
| polarC18 | 276.144 | 85.8169 | 13.2 | 0.25 |
| polarC18 | 276.144 | 55.3287 | 13.2 | 0.25 |
| polarC18 | 276.144 | 125.9689 | 13.2 | 0.25 |
| polarC18 | 276.144 | 76.9496 | 13.2 | 0.25 |
| polarC18 | 276.144 | 105.57 | 13.2 | 0.25 |
| polarC18 | 276.144 | 143.76 | 13.2 | 0.25 |
| polarC18 | 276.144 | 198.6441 | 13.2 | 0.25 |
| polarC18 | 276.144 | 65.303 | 13.2 | 0.25 |
| polarC18 | 276.144 | 177.3855 | 13.2 | 0.25 |
| polarC18 | 276.144 | 131.5633 | 13.2 | 0.25 |
| polarC18 | 276.144 | 54.6689 | 13.2 | 0.25 |
| polarC18 | 276.144 | 91.857 | 13.2 | 0.25 |
| polarC18 | 276.144 | 92.6141 | 13.2 | 0.25 |
| polarC18 | 276.144 | 275.4412 | 13.2 | 0.25 |
| polarC18 | 276.144 | 97.2021 | 13.2 | 0.25 |
| polarC18 | 276.144 | 123.0617 | 13.2 | 0.25 |
| polarC18 | 276.144 | 236.6424 | 13.2 | 0.25 |
| polarC18 | 276.144 | 136.2688 | 13.2 | 0.25 |
| polarC18 | 276.144 | 263.5924 | 13.2 | 0.25 |
| polarC18 | 276.144 | 71.6186 | 13.2 | 0.25 |
| polarC18 | 276.144 | 98.0497 | 13.2 | 0.25 |
| polarC18 | 276.144 | 197.2288 | 13.2 | 0.25 |
| polarC18 | 276.144 | 94.1708 | 13.2 | 0.25 |
| polarC18 | 276.144 | 94.5155 | 13.2 | 0.25 |
| polarC18 | 276.144 | 135.6614 | 13.2 | 0.25 |
| polarC18 | 276.144 | 53.4664 | 13.2 | 0.25 |
| polarC18 | 276.144 | 70.4442 | 13.2 | 0.25 |
| polarC18 | 276.144 | 225.2927 | 13.2 | 0.25 |
| polarC18 | 276.144 | 61.1233 | 13.2 | 0.25 |
| polarC18 | 276.144 | 150.841 | 13.2 | 0.25 |
| polarC18 | 276.144 | 108.3051 | 13.2 | 0.25 |
| polarC18 | 276.144 | 290.5993 | 13.2 | 0.25 |
| polarC18 | 276.144 | 153.8761 | 13.2 | 0.25 |
| polarC18 | 276.144 | 53.3475 | 13.2 | 0.25 |
| polarC18 | 276.144 | 76.6062 | 13.2 | 0.25 |
| polarC18 | 276.144 | 86.1803 | 13.2 | 0.25 |
| polarC18 | 276.144 | 69.4271 | 13.2 | 0.25 |
| polarC18 | 276.144 | 132.1025 | 13.2 | 0.25 |
| polarC18 | 276.144 | 227.3208 | 13.2 | 0.25 |
| polarC18 | 276.144 | 59.9507 | 13.2 | 0.25 |
| polarC18 | 276.144 | 82.5978 | 13.1 | 0.25 |

| | | | | |
|---|---|---|---|---|
| polarC18 | 276.144 | 163.0999 | 13.1 | 0.25 |
| polarC18 | 276.144 | 289.5987 | 13.1 | 0.25 |
| polarC18 | 276.144 | 78.1013 | 13.1 | 0.25 |
| polarC18 | 276.144 | 106.4956 | 13.1 | 0.25 |
| polarC18 | 276.144 | 196.9291 | 13.1 | 0.25 |
| polarC18 | 276.144 | 82.0993 | 13.1 | 0.25 |
| polarC18 | 276.144 | 54.1417 | 13.1 | 0.25 |
| polarC18 | 276.144 | 113.4412 | 13.1 | 0.25 |
| polarC18 | 276.144 | 104.4244 | 13.1 | 0.25 |
| polarC18 | 276.144 | 72.3581 | 13.1 | 0.25 |
| polarC18 | 276.144 | 199.6981 | 13.1 | 0.25 |
| polarC18 | 276.144 | 199.8261 | 13.1 | 0.25 |
| polarC18 | 276.144 | 155.4184 | 13.1 | 0.25 |
| polarC18 | 276.144 | 51.3818 | 13.1 | 0.25 |
| polarC18 | 276.144 | 98.1874 | 13.1 | 0.25 |
| polarC18 | 276.144 | 104.1949 | 13.1 | 0.25 |
| polarC18 | 276.144 | 86.0064 | 13.1 | 0.25 |
| polarC18 | 276.144 | 60.4178 | 13.1 | 0.25 |
| polarC18 | 276.144 | 276.0405 | 13.1 | 0.25 |
| polarC18 | 276.144 | 127.3418 | 13.1 | 0.25 |
| polarC18 | 276.144 | 83.3872 | 13.1 | 0.25 |
| polarC18 | 276.144 | 94.35 | 13.1 | 0.25 |
| polarC18 | 276.144 | 221.9687 | 13.1 | 0.25 |
| polarC18 | 276.144 | 89.8782 | 13.1 | 0.25 |
| polarC18 | 276.144 | 249.8104 | 13.1 | 0.25 |
| polarC18 | 276.144 | 89.2712 | 13.1 | 0.25 |
| polarC18 | 276.144 | 276.4397 | 13.1 | 0.25 |
| polarC18 | 276.144 | 61.1322 | 13.1 | 0.25 |
| polarC18 | 276.144 | 106.2986 | 13.1 | 0.25 |
| polarC18 | 276.144 | 89.0291 | 13.1 | 0.25 |
| polarC18 | 276.144 | 219.4667 | 13.1 | 0.25 |
| polarC18 | 276.144 | 132.1645 | 13.1 | 0.25 |
| polarC18 | 276.144 | 240.5518 | 13.1 | 0.25 |
| polarC18 | 276.144 | 165.497 | 13.1 | 0.25 |
| polarC18 | 276.144 | 227.8849 | 13.1 | 0.25 |
| polarC18 | 276.144 | 165.4758 | 13.1 | 0.25 |
| polarC18 | 276.144 | 149.4177 | 13.1 | 0.25 |
| polarC18 | 276.144 | 196.4067 | 13.1 | 0.25 |
| polarC18 | 276.144 | 74.6197 | 13.1 | 0.25 |
| polarC18 | 276.144 | 117.7929 | 13.1 | 0.25 |
| polarC18 | 276.144 | 181.7026 | 13.1 | 0.25 |
| polarC18 | 276.144 | 213.5633 | 13.1 | 0.25 |

| | | | | |
|---|---|---|---|---|
| polarC18 | 276.144 | 74.0007 | 13.1 | 0.25 |
| polarC18 | 276.144 | 162.3146 | 13.1 | 0.25 |
| polarC18 | 276.144 | 110.9406 | 13.1 | 0.25 |
| polarC18 | 276.144 | 91.7955 | 13.1 | 0.25 |
| polarC18 | 276.144 | 260.9761 | 13.1 | 0.25 |
| polarC18 | 276.144 | 106.2286 | 13 | 0.25 |
| polarC18 | 276.144 | 86.4603 | 13 | 0.25 |
| polarC18 | 276.144 | 169.5595 | 13 | 0.25 |
| polarC18 | 276.144 | 52.2748 | 13 | 0.25 |
| polarC18 | 276.144 | 118.052 | 13 | 0.25 |
| polarC18 | 276.144 | 302.0792 | 13 | 0.25 |
| polarC18 | 276.144 | 145.6693 | 13 | 0.25 |
| polarC18 | 276.144 | 115.2713 | 13 | 0.25 |
| polarC18 | 276.144 | 123.3943 | 13 | 0.25 |
| polarC18 | 276.144 | 71.7773 | 13 | 0.25 |
| polarC18 | 276.144 | 297.1685 | 13 | 0.25 |
| polarC18 | 276.144 | 139.7253 | 13 | 0.25 |
| polarC18 | 276.144 | 127.5243 | 13 | 0.25 |
| polarC18 | 276.144 | 130.0674 | 13 | 0.25 |
| polarC18 | 276.144 | 91.0736 | 13 | 0.25 |
| polarC18 | 276.144 | 142.5024 | 13 | 0.25 |
| polarC18 | 276.144 | 154.648 | 13 | 0.25 |
| polarC18 | 276.144 | 202.4259 | 13 | 0.25 |
| polarC18 | 276.144 | 93.4793 | 13 | 0.25 |
| polarC18 | 276.144 | 178.3733 | 13 | 0.25 |
| polarC18 | 276.144 | 62.1197 | 13 | 0.25 |
| polarC18 | 276.144 | 83.5924 | 13 | 0.25 |
| polarC18 | 276.144 | 232.3199 | 13 | 0.25 |
| polarC18 | 276.144 | 172.0874 | 13 | 0.25 |
| polarC18 | 276.144 | 264.2688 | 13 | 0.25 |
| polarC18 | 276.144 | 166.8069 | 13 | 0.25 |
| polarC18 | 276.144 | 67.0279 | 13 | 0.25 |
| polarC18 | 276.144 | 65.2683 | 13 | 0.25 |
| polarC18 | 276.144 | 195.2385 | 13 | 0.25 |
| polarC18 | 276.144 | 84.8511 | 13 | 0.25 |
| polarC18 | 276.144 | 110.7573 | 13 | 0.25 |
| polarC18 | 276.144 | 165.7349 | 13 | 0.25 |
| polarC18 | 276.144 | 76.9735 | 13 | 0.25 |
| polarC18 | 276.144 | 237.4051 | 13 | 0.25 |
| polarC18 | 276.144 | 255.6571 | 13 | 0.25 |
| polarC18 | 276.144 | 82.7899 | 13 | 0.25 |
| polarC18 | 276.144 | 124.8498 | 13 | 0.25 |

| | | | | |
|---|---|---|---|---|
| polarC18 | 276.144 | 50.3462 | 13 | 0.25 |
| polarC18 | 276.144 | 118.7285 | 13 | 0.25 |
| polarC18 | 276.144 | 133.0645 | 13 | 0.25 |
| polarC18 | 276.144 | 50.8912 | 13 | 0.25 |
| polarC18 | 276.144 | 85.5564 | 13 | 0.25 |
| polarC18 | 276.144 | 222.7656 | 13 | 0.25 |
| polarC18 | 276.144 | 214.6719 | 13 | 0.25 |
| polarC18 | 276.144 | 133.6946 | 13 | 0.25 |
| polarC18 | 276.144 | 93.8977 | 13 | 0.25 |
| polarC18 | 276.144 | 195.1683 | 13 | 0.25 |
| polarC18 | 276.144 | 54.5414 | 13 | 0.25 |
| polarC18 | 276.144 | 83.353 | 13 | 0.25 |
| polarC18 | 276.144 | 61.6741 | 13 | 0.25 |
| polarC18 | 276.144 | 96.1437 | 13 | 0.25 |
| polarC18 | 276.144 | 203.2657 | 13 | 0.25 |
| polarC18 | 276.144 | 93.6564 | 13 | 0.25 |
| polarC18 | 276.144 | 83.2111 | 13 | 0.25 |
| polarC18 | 276.144 | 126.4962 | 13 | 0.25 |
| polarC18 | 276.144 | 222.0591 | 13 | 0.25 |
| polarC18 | 276.144 | 186.6051 | 13 | 0.25 |
| polarC18 | 276.144 | 168.469 | 13 | 0.25 |
| polarC18 | 276.144 | 105.2749 | 13 | 0.25 |
| polarC18 | 276.144 | 58.3156 | 13 | 0.25 |
| polarC18 | 276.144 | 71.7509 | 13 | 0.25 |
| polarC18 | 276.144 | 57.2232 | 13 | 0.25 |
| polarC18 | 276.144 | 65.2535 | 13 | 0.25 |
| polarC18 | 276.144 | 73.1264 | 13 | 0.25 |
| polarC18 | 276.144 | 64.1838 | 13 | 0.25 |
| polarC18 | 276.144 | 51.5401 | 13 | 0.25 |
| polarC18 | 276.144 | 76.1138 | 12.9 | 0.25 |
| polarC18 | 276.144 | 109.7468 | 12.9 | 0.25 |
| polarC18 | 276.144 | 76.9908 | 12.9 | 0.25 |
| polarC18 | 276.144 | 94.2479 | 12.9 | 0.25 |
| polarC18 | 276.144 | 66.1112 | 12.9 | 0.25 |
| polarC18 | 276.144 | 93.032 | 12.9 | 0.25 |
| polarC18 | 276.144 | 120.0472 | 12.9 | 0.25 |
| polarC18 | 276.144 | 127.2729 | 12.9 | 0.25 |
| polarC18 | 276.144 | 78.3068 | 12.9 | 0.25 |
| polarC18 | 276.144 | 66.8397 | 12.9 | 0.25 |
| polarC18 | 276.144 | 215.9492 | 12.9 | 0.25 |
| polarC18 | 276.144 | 118.6649 | 12.9 | 0.25 |
| polarC18 | 276.144 | 157.0439 | 12.9 | 0.25 |

| | | | | |
|---|---|---|---|---|
| polarC18 | 276.144 | 50.6827 | 12.9 | 0.25 |
| polarC18 | 276.144 | 129.8427 | 12.9 | 0.25 |
| polarC18 | 276.144 | 110.3832 | 12.9 | 0.25 |
| polarC18 | 276.144 | 72.536 | 12.9 | 0.25 |
| polarC18 | 276.144 | 135.9875 | 12.9 | 0.25 |
| polarC18 | 276.144 | 128.6808 | 12.9 | 0.25 |
| polarC18 | 276.144 | 132.5097 | 12.9 | 0.25 |
| polarC18 | 276.144 | 288.8656 | 12.9 | 0.25 |
| polarC18 | 276.144 | 141.1244 | 12.9 | 0.25 |
| polarC18 | 276.144 | 62.5477 | 12.9 | 0.25 |
| polarC18 | 276.144 | 197.3419 | 12.9 | 0.25 |
| polarC18 | 276.144 | 77.213 | 12.9 | 0.25 |
| polarC18 | 276.144 | 264.751 | 12.9 | 0.25 |
| polarC18 | 276.144 | 69.0519 | 12.9 | 0.25 |
| polarC18 | 276.144 | 94.6119 | 12.9 | 0.25 |
| polarC18 | 276.144 | 74.888 | 12.9 | 0.25 |
| polarC18 | 276.144 | 66.7941 | 12.9 | 0.25 |
| polarC18 | 276.144 | 221.0176 | 12.9 | 0.25 |
| polarC18 | 276.144 | 145.3869 | 12.9 | 0.25 |
| polarC18 | 276.144 | 137.0377 | 12.9 | 0.25 |
| polarC18 | 276.144 | 189.1183 | 12.9 | 0.25 |
| polarC18 | 276.144 | 154.8704 | 12.9 | 0.25 |
| polarC18 | 276.144 | 115.9269 | 12.9 | 0.25 |
| polarC18 | 276.144 | 53.3233 | 12.9 | 0.25 |
| polarC18 | 276.144 | 103.7572 | 12.9 | 0.25 |
| polarC18 | 276.144 | 197.4106 | 12.9 | 0.25 |
| polarC18 | 276.144 | 251.5972 | 12.9 | 0.25 |
| polarC18 | 276.144 | 69.3257 | 12.9 | 0.25 |
| polarC18 | 276.144 | 58.2782 | 12.9 | 0.25 |
| polarC18 | 276.144 | 94.3442 | 12.9 | 0.25 |
| polarC18 | 276.144 | 68.4579 | 12.9 | 0.25 |
| polarC18 | 276.144 | 118.5463 | 12.9 | 0.25 |
| polarC18 | 276.144 | 291.8464 | 12.9 | 0.25 |
| polarC18 | 276.144 | 160.3182 | 12.9 | 0.25 |
| polarC18 | 276.144 | 275.5498 | 12.9 | 0.25 |
| polarC18 | 276.144 | 61.0882 | 12.9 | 0.25 |
| polarC18 | 276.144 | 224.5201 | 12.9 | 0.25 |
| polarC18 | 276.144 | 71.7094 | 12.9 | 0.25 |
| polarC18 | 276.144 | 70.8015 | 12.9 | 0.25 |
| polarC18 | 276.144 | 248.6324 | 12.9 | 0.25 |
| polarC18 | 276.144 | 125.0614 | 12.9 | 0.25 |
| polarC18 | 276.144 | 129.8152 | 12.9 | 0.25 |

| | | | | |
|---|---|---|---|---|
| polarC18 | 276.144 | 163.177 | 12.8 | 0.25 |
| polarC18 | 276.144 | 84.8958 | 12.8 | 0.25 |
| polarC18 | 276.144 | 121.6073 | 12.8 | 0.25 |
| polarC18 | 276.144 | 61.1772 | 12.8 | 0.25 |
| polarC18 | 276.144 | 72.7146 | 12.8 | 0.25 |
| polarC18 | 276.144 | 50.6366 | 12.8 | 0.25 |
| polarC18 | 276.144 | 82.392 | 12.8 | 0.25 |
| polarC18 | 276.144 | 92.1705 | 12.8 | 0.25 |
| polarC18 | 276.144 | 59.3158 | 12.8 | 0.25 |
| polarC18 | 276.144 | 94.941 | 12.8 | 0.25 |
| polarC18 | 276.144 | 71.4432 | 12.8 | 0.25 |
| polarC18 | 276.144 | 52.1988 | 12.8 | 0.25 |
| polarC18 | 276.144 | 63.4448 | 12.8 | 0.25 |
| polarC18 | 276.144 | 71.4137 | 12.8 | 0.25 |
| polarC18 | 276.144 | 70.0966 | 12.8 | 0.25 |
| polarC18 | 276.144 | 148.2206 | 12.8 | 0.25 |
| polarC18 | 276.144 | 80.8938 | 12.8 | 0.25 |
| polarC18 | 276.144 | 170.1172 | 12.8 | 0.25 |
| polarC18 | 276.144 | 121.9275 | 12.8 | 0.25 |
| polarC18 | 276.144 | 105.6274 | 12.8 | 0.25 |
| polarC18 | 276.144 | 107.9946 | 12.8 | 0.25 |
| polarC18 | 276.144 | 76.5007 | 12.8 | 0.25 |
| polarC18 | 276.144 | 141.7742 | 12.8 | 0.25 |
| polarC18 | 276.144 | 59.9976 | 12.8 | 0.25 |
| polarC18 | 276.144 | 68.4344 | 12.8 | 0.25 |
| polarC18 | 276.144 | 88.8159 | 12.8 | 0.25 |
| polarC18 | 276.144 | 52.9938 | 12.8 | 0.25 |
| polarC18 | 276.144 | 213.0715 | 12.8 | 0.25 |
| polarC18 | 276.144 | 197.384 | 12.8 | 0.25 |
| polarC18 | 276.144 | 87.4752 | 12.8 | 0.25 |
| polarC18 | 276.144 | 67.3071 | 12.8 | 0.25 |
| polarC18 | 276.144 | 170.7313 | 12.8 | 0.25 |
| polarC18 | 276.144 | 178.1722 | 12.8 | 0.25 |
| polarC18 | 276.144 | 301.6359 | 12.8 | 0.25 |
| polarC18 | 276.144 | 87.7383 | 12.8 | 0.25 |
| polarC18 | 276.144 | 50.5756 | 12.8 | 0.25 |
| polarC18 | 276.144 | 68.712 | 12.8 | 0.25 |
| polarC18 | 276.144 | 67.1057 | 12.8 | 0.25 |
| polarC18 | 276.144 | 63.5444 | 12.8 | 0.25 |
| polarC18 | 276.144 | 65.8343 | 12.8 | 0.25 |
| polarC18 | 276.144 | 64.9127 | 12.8 | 0.25 |
| polarC18 | 276.144 | 86.6932 | 12.8 | 0.25 |

| | | | | |
|---|---|---|---|---|
| polarC18 | 276.144 | 226.6061 | 12.8 | 0.25 |
| polarC18 | 276.144 | 56.2313 | 12.8 | 0.25 |
| polarC18 | 276.144 | 58.9006 | 12.8 | 0.25 |
| polarC18 | 276.144 | 98.2081 | 12.8 | 0.25 |
| polarC18 | 276.144 | 75.7943 | 12.8 | 0.25 |
| polarC18 | 276.144 | 87.912 | 12.8 | 0.25 |
| polarC18 | 276.144 | 136.1368 | 12.7 | 0.25 |
| polarC18 | 276.144 | 99.5179 | 12.7 | 0.25 |
| polarC18 | 276.144 | 76.6435 | 12.7 | 0.25 |
| polarC18 | 276.144 | 154.0159 | 12.7 | 0.25 |
| polarC18 | 276.144 | 58.1665 | 12.7 | 0.24 |
| polarC18 | 276.144 | 188.4082 | 12.7 | 0.24 |
| polarC18 | 276.144 | 75.1255 | 12.7 | 0.24 |
| polarC18 | 276.144 | 61.0305 | 12.7 | 0.24 |
| polarC18 | 276.144 | 56.6037 | 12.7 | 0.24 |
| polarC18 | 276.144 | 93.8919 | 12.7 | 0.24 |
| polarC18 | 276.144 | 191.6646 | 12.7 | 0.24 |
| polarC18 | 276.144 | 129.9999 | 12.7 | 0.24 |
| polarC18 | 276.144 | 100.0336 | 12.7 | 0.24 |
| polarC18 | 276.144 | 112.7209 | 12.7 | 0.24 |
| polarC18 | 276.144 | 55.2463 | 12.7 | 0.24 |
| polarC18 | 276.144 | 112.1535 | 12.7 | 0.24 |
| polarC18 | 276.144 | 71.7541 | 12.7 | 0.24 |
| polarC18 | 276.144 | 62.5885 | 12.7 | 0.24 |
| polarC18 | 276.144 | 242.5555 | 12.7 | 0.24 |
| polarC18 | 276.144 | 55.4452 | 12.7 | 0.24 |
| polarC18 | 276.144 | 64.2361 | 12.7 | 0.24 |
| polarC18 | 276.144 | 265.0348 | 12.7 | 0.24 |
| polarC18 | 276.144 | 60.7174 | 12.7 | 0.24 |
| polarC18 | 276.144 | 100.4422 | 12.7 | 0.24 |
| polarC18 | 276.144 | 232.7812 | 12.7 | 0.24 |
| polarC18 | 276.144 | 68.9485 | 12.7 | 0.24 |
| polarC18 | 276.144 | 51.8379 | 12.7 | 0.24 |
| polarC18 | 276.144 | 63.2383 | 12.7 | 0.24 |
| polarC18 | 276.144 | 58.5218 | 12.7 | 0.24 |
| polarC18 | 276.144 | 196.7208 | 12.7 | 0.24 |
| polarC18 | 276.144 | 187.4482 | 12.7 | 0.24 |
| polarC18 | 276.144 | 157.7794 | 12.7 | 0.24 |
| polarC18 | 276.144 | 62.0562 | 12.7 | 0.24 |
| polarC18 | 276.144 | 136.6231 | 12.7 | 0.24 |
| polarC18 | 276.144 | 65.9704 | 12.7 | 0.24 |
| polarC18 | 276.144 | 56.796 | 12.7 | 0.24 |

| | | | | |
|---|---|---|---|---|
| polarC18 | 276.144 | 217.0792 | 12.7 | 0.24 |
| polarC18 | 276.144 | 157.823 | 12.7 | 0.24 |
| polarC18 | 276.144 | 56.3699 | 12.7 | 0.24 |
| polarC18 | 276.144 | 58.6496 | 12.7 | 0.24 |
| polarC18 | 276.144 | 50.4392 | 12.7 | 0.24 |
| polarC18 | 276.144 | 80.088 | 12.7 | 0.24 |
| polarC18 | 276.144 | 162.2414 | 12.7 | 0.24 |
| polarC18 | 276.144 | 273.0551 | 12.7 | 0.24 |
| polarC18 | 276.144 | 75.1356 | 12.7 | 0.24 |
| polarC18 | 276.144 | 213.3672 | 12.7 | 0.24 |
| polarC18 | 276.144 | 98.2957 | 12.7 | 0.24 |
| polarC18 | 276.144 | 155.2015 | 12.7 | 0.24 |
| polarC18 | 276.144 | 54.1893 | 12.6 | 0.24 |
| polarC18 | 276.144 | 181.1935 | 12.6 | 0.24 |
| polarC18 | 276.144 | 91.8698 | 12.6 | 0.24 |
| polarC18 | 276.144 | 50.3234 | 12.6 | 0.24 |
| polarC18 | 276.144 | 56.151 | 12.6 | 0.24 |
| polarC18 | 276.144 | 68.9939 | 12.6 | 0.24 |
| polarC18 | 276.144 | 112.8641 | 12.6 | 0.24 |
| polarC18 | 276.144 | 137.5231 | 12.6 | 0.24 |
| polarC18 | 276.144 | 280.3349 | 12.6 | 0.24 |
| polarC18 | 276.144 | 158.8894 | 12.6 | 0.24 |
| polarC18 | 276.144 | 217.7867 | 12.6 | 0.24 |
| polarC18 | 276.144 | 58.7059 | 12.6 | 0.24 |
| polarC18 | 276.144 | 156.4841 | 12.6 | 0.24 |
| polarC18 | 276.144 | 187.2307 | 12.6 | 0.24 |
| polarC18 | 276.144 | 123.1776 | 12.6 | 0.24 |
| polarC18 | 276.144 | 151.515 | 12.6 | 0.24 |
| polarC18 | 276.144 | 85.0127 | 12.6 | 0.24 |
| polarC18 | 276.144 | 51.7782 | 12.6 | 0.24 |
| polarC18 | 276.144 | 119.9858 | 12.6 | 0.24 |
| polarC18 | 276.144 | 56.2463 | 12.6 | 0.24 |
| polarC18 | 276.144 | 145.7513 | 12.6 | 0.24 |
| polarC18 | 276.144 | 65.1339 | 12.6 | 0.24 |
| polarC18 | 276.144 | 128.2301 | 12.6 | 0.24 |
| polarC18 | 276.144 | 53.9679 | 12.6 | 0.24 |
| polarC18 | 276.144 | 108.455 | 12.6 | 0.24 |
| polarC18 | 276.144 | 137.1576 | 12.6 | 0.24 |
| polarC18 | 276.144 | 175.5681 | 12.6 | 0.24 |
| polarC18 | 276.144 | 87.9341 | 12.6 | 0.24 |
| polarC18 | 276.144 | 54.9226 | 12.5 | 0.24 |
| polarC18 | 276.144 | 62.0286 | 12.5 | 0.24 |

| | | | | |
|---|---|---|---|---|
| polarC18 | 276.144 | 128.9292 | 12.5 | 0.24 |
| polarC18 | 276.144 | 238.1043 | 12.5 | 0.24 |
| polarC18 | 276.144 | 196.6754 | 12.5 | 0.24 |
| polarC18 | 276.144 | 59.0533 | 12.5 | 0.24 |
| polarC18 | 276.144 | 84.1489 | 12.5 | 0.24 |
| polarC18 | 276.144 | 116.8448 | 12.5 | 0.24 |
| polarC18 | 276.144 | 250.0134 | 12.5 | 0.24 |
| polarC18 | 276.144 | 154.2535 | 12.5 | 0.24 |
| polarC18 | 276.144 | 160.3504 | 12.5 | 0.24 |
| polarC18 | 276.144 | 134.0972 | 12.5 | 0.24 |
| polarC18 | 276.144 | 62.7713 | 12.5 | 0.24 |
| polarC18 | 276.144 | 58.6903 | 12.5 | 0.24 |
| polarC18 | 276.144 | 273.264 | 12.5 | 0.24 |
| polarC18 | 276.144 | 101.46 | 12.5 | 0.24 |
| polarC18 | 276.144 | 160.5118 | 12.5 | 0.24 |
| polarC18 | 276.144 | 57.266 | 12.5 | 0.24 |
| polarC18 | 276.144 | 300.6871 | 12.5 | 0.24 |
| polarC18 | 276.144 | 116.4174 | 12.5 | 0.24 |
| polarC18 | 276.144 | 57.1824 | 12.5 | 0.24 |
| polarC18 | 276.144 | 115.9893 | 12.5 | 0.24 |
| polarC18 | 276.144 | 61.9591 | 12.5 | 0.24 |
| polarC18 | 276.144 | 110.4556 | 12.5 | 0.24 |
| polarC18 | 276.144 | 54.0301 | 12.5 | 0.24 |
| polarC18 | 276.144 | 144.3666 | 12.5 | 0.24 |
| polarC18 | 276.144 | 50.8524 | 12.5 | 0.24 |
| polarC18 | 276.144 | 136.3339 | 12.5 | 0.24 |
| polarC18 | 276.144 | 82.7392 | 12.5 | 0.24 |
| polarC18 | 276.144 | 286.8518 | 12.5 | 0.24 |
| polarC18 | 276.144 | 187.9582 | 12.5 | 0.24 |
| polarC18 | 276.144 | 69.6673 | 12.5 | 0.24 |
| polarC18 | 276.144 | 129.6421 | 12.5 | 0.24 |
| polarC18 | 276.144 | 87.9902 | 12.5 | 0.24 |
| polarC18 | 276.144 | 221.3301 | 12.5 | 0.24 |
| polarC18 | 276.144 | 136.4572 | 12.5 | 0.24 |
| polarC18 | 276.144 | 90.3586 | 12.5 | 0.24 |
| polarC18 | 276.144 | 132.5509 | 12.5 | 0.24 |
| polarC18 | 276.144 | 72.1286 | 12.5 | 0.24 |
| polarC18 | 276.144 | 57.5312 | 12.5 | 0.24 |
| polarC18 | 276.144 | 161.2972 | 12.5 | 0.24 |
| polarC18 | 276.144 | 57.1931 | 12.4 | 0.24 |
| polarC18 | 276.144 | 151.0557 | 12.4 | 0.24 |
| polarC18 | 276.144 | 147.3439 | 12.4 | 0.24 |

| | | | | |
|---|---|---|---|---|
| polarC18 | 276.144 | 167.9754 | 12.4 | 0.24 |
| polarC18 | 276.144 | 77.8897 | 12.4 | 0.24 |
| polarC18 | 276.144 | 228.7983 | 12.4 | 0.24 |
| polarC18 | 276.144 | 58.2123 | 12.4 | 0.24 |
| polarC18 | 276.144 | 165.5234 | 12.4 | 0.24 |
| polarC18 | 276.144 | 266.3104 | 12.4 | 0.24 |
| polarC18 | 276.144 | 223.4349 | 12.4 | 0.24 |
| polarC18 | 276.144 | 88.0913 | 12.4 | 0.24 |
| polarC18 | 276.144 | 118.1589 | 12.4 | 0.24 |
| polarC18 | 276.144 | 117.1144 | 12.4 | 0.24 |
| polarC18 | 276.144 | 89.6467 | 12.4 | 0.24 |
| polarC18 | 276.144 | 61.9923 | 12.4 | 0.24 |
| polarC18 | 276.144 | 69.044 | 12.4 | 0.24 |
| polarC18 | 276.144 | 93.9931 | 12.4 | 0.24 |
| polarC18 | 276.144 | 167.1484 | 12.4 | 0.24 |
| polarC18 | 276.144 | 71.0155 | 12.4 | 0.24 |
| polarC18 | 276.144 | 118.849 | 12.4 | 0.24 |
| polarC18 | 276.144 | 74.5028 | 12.4 | 0.24 |
| polarC18 | 276.144 | 72.6089 | 12.4 | 0.24 |
| polarC18 | 276.144 | 125.5431 | 12.4 | 0.24 |
| polarC18 | 276.144 | 111.5615 | 12.4 | 0.24 |
| polarC18 | 276.144 | 50.1488 | 12.4 | 0.24 |
| polarC18 | 276.144 | 67.7379 | 12.4 | 0.24 |
| polarC18 | 276.144 | 110.659 | 12.4 | 0.24 |
| polarC18 | 276.144 | 169.2156 | 12.4 | 0.24 |
| polarC18 | 276.144 | 138.5228 | 12.4 | 0.24 |
| polarC18 | 276.144 | 52.6147 | 12.4 | 0.24 |
| polarC18 | 276.144 | 194.0727 | 12.4 | 0.24 |
| polarC18 | 276.144 | 179.9186 | 12.4 | 0.24 |
| polarC18 | 276.144 | 141.3081 | 12.4 | 0.24 |
| polarC18 | 276.144 | 151.8996 | 12.4 | 0.24 |
| polarC18 | 276.144 | 173.3861 | 12.4 | 0.24 |
| polarC18 | 276.144 | 79.2598 | 12.4 | 0.24 |
| polarC18 | 276.144 | 157.6576 | 12.3 | 0.24 |
| polarC18 | 276.144 | 93.6368 | 12.3 | 0.24 |
| polarC18 | 276.144 | 71.5907 | 12.3 | 0.24 |
| polarC18 | 276.144 | 55.0847 | 12.3 | 0.24 |
| polarC18 | 276.144 | 87.8756 | 12.3 | 0.24 |
| polarC18 | 276.144 | 59.6851 | 12.3 | 0.24 |
| polarC18 | 276.144 | 274.4859 | 12.3 | 0.24 |
| polarC18 | 276.144 | 241.0433 | 12.3 | 0.24 |
| polarC18 | 276.144 | 60.0201 | 12.3 | 0.24 |

| | | | | |
|---|---|---|---|---|
| polarC18 | 276.144 | 89.9343 | 12.3 | 0.24 |
| polarC18 | 276.144 | 180.0015 | 12.3 | 0.24 |
| polarC18 | 276.144 | 88.4694 | 12.3 | 0.24 |
| polarC18 | 276.144 | 54.5214 | 12.3 | 0.24 |
| polarC18 | 276.144 | 211.2516 | 12.3 | 0.24 |
| polarC18 | 276.144 | 52.7952 | 12.3 | 0.24 |
| polarC18 | 276.144 | 289.7258 | 12.3 | 0.24 |
| polarC18 | 276.144 | 82.7156 | 12.3 | 0.24 |
| polarC18 | 276.144 | 57 | 12.3 | 0.24 |
| polarC18 | 276.144 | 56.4076 | 12.3 | 0.24 |
| polarC18 | 276.144 | 140.5337 | 12.3 | 0.24 |
| polarC18 | 276.144 | 251.1554 | 12.3 | 0.24 |
| polarC18 | 276.144 | 93.5921 | 12.3 | 0.24 |
| polarC18 | 276.144 | 150.7551 | 12.3 | 0.24 |
| polarC18 | 276.144 | 93.5656 | 12.3 | 0.24 |
| polarC18 | 276.144 | 82.8112 | 12.3 | 0.24 |
| polarC18 | 276.144 | 123.6344 | 12.3 | 0.24 |
| polarC18 | 276.144 | 67.6668 | 12.3 | 0.24 |
| polarC18 | 276.144 | 73.5172 | 12.3 | 0.24 |
| polarC18 | 276.144 | 90.1454 | 12.3 | 0.24 |
| polarC18 | 276.144 | 218.9031 | 12.2 | 0.24 |
| polarC18 | 276.144 | 141.7892 | 12.2 | 0.24 |
| polarC18 | 276.144 | 59.6689 | 12.2 | 0.24 |
| polarC18 | 276.144 | 98.0271 | 12.2 | 0.24 |
| polarC18 | 276.144 | 79.8403 | 12.2 | 0.24 |
| polarC18 | 276.144 | 178.5076 | 12.2 | 0.24 |
| polarC18 | 276.144 | 156.9831 | 12.2 | 0.23 |
| polarC18 | 276.144 | 135.8416 | 12.2 | 0.23 |
| polarC18 | 276.144 | 210.7593 | 12.2 | 0.23 |
| polarC18 | 276.144 | 127.1962 | 12.2 | 0.23 |
| polarC18 | 276.144 | 213.1556 | 12.2 | 0.23 |
| polarC18 | 276.144 | 104.7017 | 12.2 | 0.23 |
| polarC18 | 276.144 | 103.3492 | 12.2 | 0.23 |
| polarC18 | 276.144 | 117.584 | 12.2 | 0.23 |
| polarC18 | 276.144 | 99.8411 | 12.2 | 0.23 |
| polarC18 | 276.144 | 66.3239 | 12.2 | 0.23 |
| polarC18 | 276.144 | 171.2692 | 12.2 | 0.23 |
| polarC18 | 276.144 | 56.0308 | 12.2 | 0.23 |
| polarC18 | 276.144 | 69.2931 | 12.2 | 0.23 |
| polarC18 | 276.144 | 53.899 | 12.2 | 0.23 |
| polarC18 | 276.144 | 245.0402 | 12.2 | 0.23 |
| polarC18 | 276.144 | 75.1652 | 12.2 | 0.23 |

| | | | | |
|---|---|---|---|---|
| polarC18 | 276.144 | 224.431 | 12.2 | 0.23 |
| polarC18 | 276.144 | 185.6039 | 12.2 | 0.23 |
| polarC18 | 276.144 | 271.0856 | 12.2 | 0.23 |
| polarC18 | 276.144 | 222.4057 | 12.2 | 0.23 |
| polarC18 | 276.144 | 83.9903 | 12.2 | 0.23 |
| polarC18 | 276.144 | 58.7915 | 12.2 | 0.23 |
| polarC18 | 276.144 | 55.0307 | 12.2 | 0.23 |
| polarC18 | 276.144 | 87.7226 | 12.2 | 0.23 |
| polarC18 | 276.144 | 165.3882 | 12.2 | 0.23 |
| polarC18 | 276.144 | 128.2656 | 12.2 | 0.23 |
| polarC18 | 276.144 | 210.539 | 12.2 | 0.23 |
| polarC18 | 276.144 | 81.6813 | 12.1 | 0.23 |
| polarC18 | 276.144 | 80.8415 | 12.1 | 0.23 |
| polarC18 | 276.144 | 93.8199 | 12.1 | 0.23 |
| polarC18 | 276.144 | 53.2696 | 12.1 | 0.23 |
| polarC18 | 276.144 | 67.4048 | 12.1 | 0.23 |
| polarC18 | 276.144 | 64.2229 | 12.1 | 0.23 |
| polarC18 | 276.144 | 68.4758 | 12.1 | 0.23 |
| polarC18 | 276.144 | 52.9552 | 12.1 | 0.23 |
| polarC18 | 276.144 | 99.3592 | 12.1 | 0.23 |
| polarC18 | 276.144 | 61.2439 | 12.1 | 0.23 |
| polarC18 | 276.144 | 98.1299 | 12.1 | 0.23 |
| polarC18 | 276.144 | 68.0429 | 12.1 | 0.23 |
| polarC18 | 276.144 | 162.0511 | 12.1 | 0.23 |
| polarC18 | 276.144 | 117.919 | 12.1 | 0.23 |
| polarC18 | 276.144 | 129.9852 | 12.1 | 0.23 |
| polarC18 | 276.144 | 71.9386 | 12.1 | 0.23 |
| polarC18 | 276.144 | 75.7301 | 12.1 | 0.23 |
| polarC18 | 276.144 | 74.6934 | 12.1 | 0.23 |
| polarC18 | 276.144 | 101.719 | 12.1 | 0.23 |
| polarC18 | 276.144 | 51.4407 | 12.1 | 0.23 |
| polarC18 | 276.144 | 133.5826 | 12.1 | 0.23 |
| polarC18 | 276.144 | 134.7094 | 12.1 | 0.23 |
| polarC18 | 276.144 | 192.5694 | 12.1 | 0.23 |
| polarC18 | 276.144 | 65.4316 | 12.1 | 0.23 |
| polarC18 | 276.144 | 61.1667 | 12.1 | 0.23 |
| polarC18 | 276.144 | 266.8022 | 12.1 | 0.23 |
| polarC18 | 276.144 | 62.7179 | 12.1 | 0.23 |
| polarC18 | 276.144 | 156.6413 | 12.1 | 0.23 |
| polarC18 | 276.144 | 137.8577 | 12.1 | 0.23 |
| polarC18 | 276.144 | 53.6371 | 12 | 0.23 |
| polarC18 | 276.144 | 90.5065 | 12 | 0.23 |

| | | | | |
|---|---|---|---|---|
| polarC18 | 276.144 | 93.6763 | 12 | 0.23 |
| polarC18 | 276.144 | 84.0632 | 12 | 0.23 |
| polarC18 | 276.144 | 215.176 | 12 | 0.23 |
| polarC18 | 276.144 | 75.8379 | 12 | 0.23 |
| polarC18 | 276.144 | 54.4967 | 12 | 0.23 |
| polarC18 | 276.144 | 51.453 | 12 | 0.23 |
| polarC18 | 276.144 | 64.0049 | 12 | 0.23 |
| polarC18 | 276.144 | 86.6333 | 12 | 0.23 |
| polarC18 | 276.144 | 280.8447 | 12 | 0.23 |
| polarC18 | 276.144 | 83.7413 | 12 | 0.23 |
| polarC18 | 276.144 | 95.0027 | 12 | 0.23 |
| polarC18 | 276.144 | 80.3473 | 12 | 0.23 |
| polarC18 | 276.144 | 216.3066 | 12 | 0.23 |
| polarC18 | 276.144 | 156.8754 | 12 | 0.23 |
| polarC18 | 276.144 | 240.1238 | 12 | 0.23 |
| polarC18 | 276.144 | 75.8969 | 12 | 0.23 |
| polarC18 | 276.144 | 75.0734 | 12 | 0.23 |
| polarC18 | 276.144 | 78.9463 | 12 | 0.23 |
| polarC18 | 276.144 | 136.1562 | 12 | 0.23 |
| polarC18 | 276.144 | 81.2608 | 12 | 0.23 |
| polarC18 | 276.144 | 287.6235 | 12 | 0.23 |
| polarC18 | 276.144 | 82.3485 | 12 | 0.23 |
| polarC18 | 276.144 | 95.2385 | 12 | 0.23 |
| polarC18 | 276.144 | 138.5834 | 12 | 0.23 |
| polarC18 | 276.144 | 106.2386 | 12 | 0.23 |
| polarC18 | 276.144 | 84.4184 | 12 | 0.23 |
| polarC18 | 276.144 | 102.2498 | 12 | 0.23 |
| polarC18 | 276.144 | 205.3213 | 11.9 | 0.23 |
| polarC18 | 276.144 | 53.1993 | 11.9 | 0.23 |
| polarC18 | 276.144 | 296.4446 | 11.9 | 0.23 |
| polarC18 | 276.144 | 128.2055 | 11.9 | 0.23 |
| polarC18 | 276.144 | 78.0156 | 11.9 | 0.23 |
| polarC18 | 276.144 | 75.2134 | 11.9 | 0.23 |
| polarC18 | 276.144 | 83.372 | 11.9 | 0.23 |
| polarC18 | 276.144 | 291.0177 | 11.9 | 0.23 |
| polarC18 | 276.144 | 176.1935 | 11.9 | 0.23 |
| polarC18 | 276.144 | 193.9846 | 11.9 | 0.23 |
| polarC18 | 276.144 | 83.0291 | 11.9 | 0.23 |
| polarC18 | 276.144 | 75.8494 | 11.9 | 0.23 |
| polarC18 | 276.144 | 111.08 | 11.9 | 0.23 |
| polarC18 | 276.144 | 93.6883 | 11.9 | 0.23 |
| polarC18 | 276.144 | 201.4077 | 11.9 | 0.23 |

| | | | | |
|---|---|---|---|---|
| polarC18 | 276.144 | 88.0208 | 11.9 | 0.23 |
| polarC18 | 276.144 | 101.7129 | 11.9 | 0.23 |
| polarC18 | 276.144 | 58.8313 | 11.9 | 0.23 |
| polarC18 | 276.144 | 62.7538 | 11.9 | 0.23 |
| polarC18 | 276.144 | 84.1531 | 11.9 | 0.23 |
| polarC18 | 276.144 | 253.9073 | 11.9 | 0.23 |
| polarC18 | 276.144 | 123.66 | 11.9 | 0.23 |
| polarC18 | 276.144 | 167.689 | 11.9 | 0.23 |
| polarC18 | 276.144 | 53.3898 | 11.8 | 0.23 |
| polarC18 | 276.144 | 71.2494 | 11.8 | 0.23 |
| polarC18 | 276.144 | 164.049 | 11.8 | 0.23 |
| polarC18 | 276.144 | 167.876 | 11.8 | 0.23 |
| polarC18 | 276.144 | 150.7894 | 11.8 | 0.23 |
| polarC18 | 276.144 | 201.015 | 11.8 | 0.23 |
| polarC18 | 276.144 | 167.6376 | 11.8 | 0.23 |
| polarC18 | 276.144 | 52.4533 | 11.8 | 0.23 |
| polarC18 | 276.144 | 146.3131 | 11.8 | 0.23 |
| polarC18 | 276.144 | 109.7318 | 11.8 | 0.23 |
| polarC18 | 276.144 | 95.3306 | 11.8 | 0.23 |
| polarC18 | 276.144 | 216.9729 | 11.8 | 0.23 |
| polarC18 | 276.144 | 108.5004 | 11.8 | 0.23 |
| polarC18 | 276.144 | 140.7381 | 11.8 | 0.23 |
| polarC18 | 276.144 | 51.5163 | 11.8 | 0.23 |
| polarC18 | 276.144 | 152.7144 | 11.8 | 0.23 |
| polarC18 | 276.144 | 126.6909 | 11.8 | 0.23 |
| polarC18 | 276.144 | 59.7587 | 11.8 | 0.23 |
| polarC18 | 276.144 | 54.0266 | 11.8 | 0.23 |
| polarC18 | 276.144 | 69.6218 | 11.8 | 0.23 |
| polarC18 | 276.144 | 64.4993 | 11.8 | 0.23 |
| polarC18 | 276.144 | 225.1763 | 11.8 | 0.23 |
| polarC18 | 276.144 | 96.6396 | 11.8 | 0.23 |
| polarC18 | 276.144 | 101.4694 | 11.8 | 0.23 |
| polarC18 | 276.144 | 63.9083 | 11.8 | 0.23 |
| polarC18 | 276.144 | 168.1186 | 11.8 | 0.23 |
| polarC18 | 276.144 | 124.3606 | 11.8 | 0.23 |
| polarC18 | 276.144 | 83.4885 | 11.8 | 0.23 |
| polarC18 | 276.144 | 67.3556 | 11.8 | 0.23 |
| polarC18 | 276.144 | 53.4387 | 11.8 | 0.23 |
| polarC18 | 276.144 | 156.8229 | 11.8 | 0.23 |
| polarC18 | 276.144 | 80.0963 | 11.8 | 0.23 |
| polarC18 | 276.144 | 51.4783 | 11.8 | 0.23 |
| polarC18 | 276.144 | 114.8079 | 11.8 | 0.23 |

| | | | | |
|---|---|---|---|---|
| polarC18 | 276.144 | 205.69 | 11.8 | 0.23 |
| polarC18 | 276.144 | 119.0027 | 11.7 | 0.23 |
| polarC18 | 276.144 | 223.8152 | 11.7 | 0.23 |
| polarC18 | 276.144 | 60.1167 | 11.7 | 0.23 |
| polarC18 | 276.144 | 53.4266 | 11.7 | 0.23 |
| polarC18 | 276.144 | 100.0236 | 11.7 | 0.23 |
| polarC18 | 276.144 | 93.1594 | 11.7 | 0.23 |
| polarC18 | 276.144 | 152.5156 | 11.7 | 0.23 |
| polarC18 | 276.144 | 74.1034 | 11.7 | 0.23 |
| polarC18 | 276.144 | 284.8555 | 11.7 | 0.23 |
| polarC18 | 276.144 | 132.3533 | 11.7 | 0.23 |
| polarC18 | 276.144 | 266.3719 | 11.7 | 0.23 |
| polarC18 | 276.144 | 69.7464 | 11.7 | 0.23 |
| polarC18 | 276.144 | 223.8853 | 11.7 | 0.23 |
| polarC18 | 276.144 | 69.2316 | 11.7 | 0.23 |
| polarC18 | 276.144 | 75.64 | 11.7 | 0.22 |
| polarC18 | 276.144 | 180.8445 | 11.7 | 0.22 |
| polarC18 | 276.144 | 57.3949 | 11.7 | 0.22 |
| polarC18 | 276.144 | 185.2495 | 11.7 | 0.22 |
| polarC18 | 276.144 | 62.1262 | 11.7 | 0.22 |
| polarC18 | 276.144 | 286.3977 | 11.7 | 0.22 |
| polarC18 | 276.144 | 154.0392 | 11.7 | 0.22 |
| polarC18 | 276.144 | 68.6019 | 11.7 | 0.22 |
| polarC18 | 276.144 | 170.1503 | 11.7 | 0.22 |
| polarC18 | 276.144 | 112.9584 | 11.7 | 0.22 |
| polarC18 | 276.144 | 99.314 | 11.7 | 0.22 |
| polarC18 | 276.144 | 50.3574 | 11.7 | 0.22 |
| polarC18 | 276.144 | 129.3318 | 11.7 | 0.22 |
| polarC18 | 276.144 | 83.3637 | 11.6 | 0.22 |
| polarC18 | 276.144 | 54.102 | 11.6 | 0.22 |
| polarC18 | 276.144 | 96.6887 | 11.6 | 0.22 |
| polarC18 | 276.144 | 103.5781 | 11.6 | 0.22 |
| polarC18 | 276.144 | 56.9223 | 11.6 | 0.22 |
| polarC18 | 276.144 | 241.991 | 11.6 | 0.22 |
| polarC18 | 276.144 | 107.265 | 11.6 | 0.22 |
| polarC18 | 276.144 | 101.5112 | 11.6 | 0.22 |
| polarC18 | 276.144 | 93.4533 | 11.6 | 0.22 |
| polarC18 | 276.144 | 158.9996 | 11.6 | 0.22 |
| polarC18 | 276.144 | 132.9101 | 11.6 | 0.22 |
| polarC18 | 276.144 | 55.0819 | 11.6 | 0.22 |
| polarC18 | 276.144 | 229.8757 | 11.6 | 0.22 |
| polarC18 | 276.144 | 197.5793 | 11.6 | 0.22 |

| | | | | |
|---|---|---|---|---|
| polarC18 | 276.144 | 80.8645 | 11.6 | 0.22 |
| polarC18 | 276.144 | 56.5586 | 11.6 | 0.22 |
| polarC18 | 276.144 | 64.147 | 11.6 | 0.22 |
| polarC18 | 276.144 | 94.388 | 11.6 | 0.22 |
| polarC18 | 276.144 | 296.3769 | 11.6 | 0.22 |
| polarC18 | 276.144 | 50.018 | 11.6 | 0.22 |
| polarC18 | 276.144 | 89.0961 | 11.6 | 0.22 |
| polarC18 | 276.144 | 58.2684 | 11.6 | 0.22 |
| polarC18 | 276.144 | 191.2751 | 11.6 | 0.22 |
| polarC18 | 276.144 | 70.0537 | 11.6 | 0.22 |
| polarC18 | 276.144 | 116.0797 | 11.5 | 0.22 |
| polarC18 | 276.144 | 240.1704 | 11.5 | 0.22 |
| polarC18 | 276.144 | 216.8166 | 11.5 | 0.22 |
| polarC18 | 276.144 | 60.5256 | 11.5 | 0.22 |
| polarC18 | 276.144 | 79.815 | 11.5 | 0.22 |
| polarC18 | 276.144 | 73.9576 | 11.5 | 0.22 |
| polarC18 | 276.144 | 175.7181 | 11.5 | 0.22 |
| polarC18 | 276.144 | 259.3622 | 11.5 | 0.22 |
| polarC18 | 276.144 | 65.517 | 11.5 | 0.22 |
| polarC18 | 276.144 | 90.0669 | 11.5 | 0.22 |
| polarC18 | 276.144 | 81.6472 | 11.5 | 0.22 |
| polarC18 | 276.144 | 52.4971 | 11.5 | 0.22 |
| polarC18 | 276.144 | 107.6504 | 11.5 | 0.22 |
| polarC18 | 276.144 | 62.9211 | 11.5 | 0.22 |
| polarC18 | 276.144 | 110.6106 | 11.5 | 0.22 |
| polarC18 | 276.144 | 87.6757 | 11.5 | 0.22 |
| polarC18 | 276.144 | 209.7092 | 11.5 | 0.22 |
| polarC18 | 276.144 | 240.6276 | 11.5 | 0.22 |
| polarC18 | 276.144 | 153.3156 | 11.5 | 0.22 |
| polarC18 | 276.144 | 269.6326 | 11.4 | 0.22 |
| polarC18 | 276.144 | 89.5521 | 11.4 | 0.22 |
| polarC18 | 276.144 | 53.7773 | 11.4 | 0.22 |
| polarC18 | 276.144 | 117.8291 | 11.4 | 0.22 |
| polarC18 | 276.144 | 50.1267 | 11.4 | 0.22 |
| polarC18 | 276.144 | 132.9742 | 11.4 | 0.22 |
| polarC18 | 276.144 | 122.5183 | 11.4 | 0.22 |
| polarC18 | 276.144 | 103.1615 | 11.4 | 0.22 |
| polarC18 | 276.144 | 166.0662 | 11.4 | 0.22 |
| polarC18 | 276.144 | 66.9338 | 11.4 | 0.22 |
| polarC18 | 276.144 | 89.3604 | 11.4 | 0.22 |
| polarC18 | 276.144 | 78.9773 | 11.4 | 0.22 |
| polarC18 | 276.144 | 53.5417 | 11.4 | 0.22 |

| | | | | |
|---|---|---|---|---|
| polarC18 | 276.144 | 54.1806 | 11.4 | 0.22 |
| polarC18 | 276.144 | 141.5649 | 11.4 | 0.22 |
| polarC18 | 276.144 | 90.1219 | 11.4 | 0.22 |
| polarC18 | 276.144 | 154.933 | 11.4 | 0.22 |
| polarC18 | 276.144 | 121.2975 | 11.3 | 0.22 |
| polarC18 | 276.144 | 137.1433 | 11.3 | 0.22 |
| polarC18 | 276.144 | 56.8818 | 11.3 | 0.22 |
| polarC18 | 276.144 | 244.7025 | 11.3 | 0.22 |
| polarC18 | 276.144 | 67.6044 | 11.3 | 0.22 |
| polarC18 | 276.144 | 66.3194 | 11.3 | 0.22 |
| polarC18 | 276.144 | 85.7602 | 11.3 | 0.22 |
| polarC18 | 276.144 | 203.4081 | 11.3 | 0.22 |
| polarC18 | 276.144 | 170.9605 | 11.3 | 0.22 |
| polarC18 | 276.144 | 117.3406 | 11.3 | 0.22 |
| polarC18 | 276.144 | 130.5257 | 11.3 | 0.22 |
| polarC18 | 276.144 | 162.4341 | 11.3 | 0.22 |
| polarC18 | 276.144 | 105.4905 | 11.3 | 0.22 |
| polarC18 | 276.144 | 55.9845 | 11.3 | 0.22 |
| polarC18 | 276.144 | 126.745 | 11.3 | 0.22 |
| polarC18 | 276.144 | 100.7399 | 11.3 | 0.22 |
| polarC18 | 276.144 | 59.5248 | 11.3 | 0.22 |
| polarC18 | 276.144 | 233.7714 | 11.3 | 0.22 |
| polarC18 | 276.144 | 58.591 | 11.3 | 0.22 |
| polarC18 | 276.144 | 157.5686 | 11.2 | 0.22 |
| polarC18 | 276.144 | 108.1276 | 11.2 | 0.22 |
| polarC18 | 276.144 | 55.6541 | 11.2 | 0.22 |
| polarC18 | 276.144 | 88.1063 | 11.2 | 0.21 |
| polarC18 | 276.144 | 88.56 | 11.2 | 0.21 |
| polarC18 | 276.144 | 57.0569 | 11.2 | 0.21 |
| polarC18 | 276.144 | 194.9471 | 11.2 | 0.21 |
| polarC18 | 276.144 | 57.5135 | 11.2 | 0.21 |
| polarC18 | 276.144 | 79.4068 | 11.1 | 0.21 |
| polarC18 | 276.144 | 78.6784 | 11.1 | 0.21 |
| polarC18 | 276.144 | 197.9403 | 11.1 | 0.21 |
| polarC18 | 276.144 | 113.9419 | 11.1 | 0.21 |
| polarC18 | 276.144 | 103.8907 | 11.1 | 0.21 |
| polarC18 | 276.144 | 57.7781 | 11.1 | 0.21 |
| polarC18 | 276.144 | 112.6296 | 11.1 | 0.21 |
| polarC18 | 276.144 | 95.0992 | 11.1 | 0.21 |
| polarC18 | 276.144 | 80.8094 | 11.1 | 0.21 |
| polarC18 | 276.144 | 68.7502 | 11.1 | 0.21 |
| polarC18 | 276.144 | 155.6471 | 11.1 | 0.21 |

| | | | | |
|---|---|---|---|---|
| polarC18 | 276.144 | 52.2121 | 11.1 | 0.21 |
| polarC18 | 276.144 | 154.083 | 11.1 | 0.21 |
| polarC18 | 276.144 | 204.1461 | 11.1 | 0.21 |
| polarC18 | 276.144 | 139.863 | 11.1 | 0.21 |
| polarC18 | 276.144 | 209.8416 | 11.1 | 0.21 |
| polarC18 | 276.144 | 59.5674 | 11.1 | 0.21 |
| polarC18 | 276.144 | 86.7619 | 11.1 | 0.21 |
| polarC18 | 276.144 | 67.2652 | 11.1 | 0.21 |
| polarC18 | 276.144 | 86.8516 | 11.1 | 0.21 |
| polarC18 | 276.144 | 76.1702 | 11 | 0.21 |
| polarC18 | 276.144 | 171.5038 | 11 | 0.21 |
| polarC18 | 276.144 | 84.5134 | 11 | 0.21 |
| polarC18 | 276.144 | 155.5549 | 11 | 0.21 |
| polarC18 | 276.144 | 208.396 | 11 | 0.21 |
| polarC18 | 276.144 | 216.064 | 11 | 0.21 |
| polarC18 | 276.144 | 74.5858 | 11 | 0.21 |
| polarC18 | 276.144 | 132.1343 | 11 | 0.21 |
| polarC18 | 276.144 | 82.6517 | 10.9 | 0.21 |
| polarC18 | 276.144 | 65.1021 | 10.9 | 0.21 |
| polarC18 | 276.144 | 250.0794 | 10.9 | 0.21 |
| polarC18 | 276.144 | 226.3816 | 10.9 | 0.21 |
| polarC18 | 276.144 | 86.6155 | 10.9 | 0.21 |
| polarC18 | 276.144 | 132.0818 | 10.8 | 0.21 |
| polarC18 | 276.144 | 139.1452 | 10.8 | 0.21 |
| polarC18 | 276.144 | 215.0102 | 10.8 | 0.21 |
| polarC18 | 276.144 | 63.3535 | 10.8 | 0.21 |
| polarC18 | 276.144 | 131.4844 | 10.8 | 0.21 |
| polarC18 | 276.144 | 58.7472 | 10.8 | 0.21 |
| polarC18 | 276.144 | 261.2759 | 10.8 | 0.21 |
| polarC18 | 276.144 | 56.2154 | 10.7 | 0.21 |
| polarC18 | 276.144 | 77.9263 | 10.6 | 0.2 |
| polarC18 | 276.144 | 69.5039 | 10.6 | 0.2 |
| polarC18 | 276.144 | 81.6415 | 10.6 | 0.2 |
| polarC18 | 276.144 | 55.7255 | 10.6 | 0.2 |
| polarC18 | 276.144 | 69.8278 | 10.5 | 0.2 |
| polarC18 | 276.144 | 72.7824 | 10.5 | 0.2 |
| polarC18 | 276.144 | 162.8781 | 10.4 | 0.2 |
| polarC18 | 276.144 | 131.1229 | 10.4 | 0.2 |
| polarC18 | 276.144 | 93.5336 | 10.4 | 0.2 |
| polarC18 | 276.144 | 78.4098 | 10.4 | 0.2 |
| polarC18 | 276.144 | 258.1311 | 10.3 | 0.2 |
| polarC18 | 276.144 | 50.097 | 10.3 | 0.2 |

| | | | | |
|---|---|---|---|---|
| polarC18 | 276.144 | 82.1069 | 10.3 | 0.2 |
| polarC18 | 276.144 | 247.6148 | 10.2 | 0.2 |
| polarC18 | 276.144 | 133.2018 | 10.2 | 0.2 |
| polarC18 | 276.144 | 116.7668 | 10.1 | 0.2 |
| polarC18 | 276.144 | 194.7758 | 10.1 | 0.19 |
| polarC18 | 276.144 | 229.986 | 10 | 0.19 |
| polarC18 | 276.144 | 187.6522 | 10 | 0.19 |
| polarC18 | 276.144 | 233.1652 | 10 | 0.19 |
| polarC18 | 276.144 | 83.5429 | 9.9 | 0.19 |
| polarC18 | 276.144 | 101.3878 | 9.9 | 0.19 |
| polarC18 | 276.144 | 51.4743 | 9.9 | 0.19 |
| polarC18 | 276.144 | 74.085 | 9.9 | 0.19 |
| polarC18 | 276.144 | 115.2543 | 9.8 | 0.19 |
| polarC18 | 276.144 | 56.3732 | 9.6 | 0.19 |
| polarC18 | 276.144 | 214.5244 | 9.6 | 0.18 |
| polarC18 | 276.144 | 78.7596 | 9.6 | 0.18 |
| polarC18 | 276.144 | 54.099 | 9.6 | 0.18 |
| polarC18 | 276.144 | 190.9887 | 9.5 | 0.18 |
| polarC18 | 276.144 | 53.8903 | 9.4 | 0.18 |
| polarC18 | 276.144 | 82.0845 | 9.4 | 0.18 |
| polarC18 | 276.144 | 195.9184 | 9.3 | 0.18 |
| polarC18 | 276.144 | 275.2131 | 9.3 | 0.18 |
| polarC18 | 276.144 | 86.094 | 9.3 | 0.18 |
| polarC18 | 276.144 | 77.5765 | 9.2 | 0.18 |
| polarC18 | 276.144 | 133.9364 | 9.2 | 0.18 |
| polarC18 | 276.144 | 56.9832 | 9.2 | 0.18 |
| polarC18 | 276.144 | 275.9905 | 9.2 | 0.18 |
| polarC18 | 276.144 | 195.7595 | 9.1 | 0.18 |
| polarC18 | 276.144 | 68.9351 | 9 | 0.17 |
| polarC18 | 276.144 | 68.2422 | 9 | 0.17 |
| polarC18 | 276.144 | 223.9417 | 9 | 0.17 |
| polarC18 | 276.144 | 66.7873 | 9 | 0.17 |
| polarC18 | 276.144 | 54.9469 | 9 | 0.17 |
| polarC18 | 276.144 | 169.9655 | 9 | 0.17 |
| polarC18 | 276.144 | 255.4054 | 9 | 0.17 |
| polarC18 | 276.144 | 61.2913 | 9 | 0.17 |
| polarC18 | 276.144 | 217.1779 | 8.9 | 0.17 |
| polarC18 | 276.144 | 103.5542 | 8.9 | 0.17 |
| polarC18 | 276.144 | 141.8454 | 8.9 | 0.17 |
| polarC18 | 276.144 | 58.9795 | 8.9 | 0.17 |
| polarC18 | 276.144 | 124.8084 | 8.9 | 0.17 |
| polarC18 | 276.144 | 79.5143 | 8.9 | 0.17 |

| | | | | |
|---|---|---|---|---|
| polarC18 | 276.144 | 209.186 | 8.9 | 0.17 |
| polarC18 | 276.144 | 233.8033 | 8.9 | 0.17 |
| polarC18 | 276.144 | 266.2329 | 8.8 | 0.17 |
| polarC18 | 276.144 | 161.8695 | 8.8 | 0.17 |
| polarC18 | 276.144 | 69.94 | 8.8 | 0.17 |
| polarC18 | 276.144 | 128.879 | 8.8 | 0.17 |
| polarC18 | 276.144 | 80.7856 | 8.8 | 0.17 |
| polarC18 | 276.144 | 300.1366 | 8.8 | 0.17 |
| polarC18 | 276.144 | 95.0853 | 8.8 | 0.17 |
| polarC18 | 276.144 | 222.7166 | 8.8 | 0.17 |
| polarC18 | 276.144 | 133.866 | 8.8 | 0.17 |
| polarC18 | 276.144 | 71.9614 | 8.8 | 0.17 |
| polarC18 | 276.144 | 55.0476 | 8.7 | 0.17 |
| polarC18 | 276.144 | 148.5228 | 8.7 | 0.17 |
| polarC18 | 276.144 | 131.9594 | 8.7 | 0.17 |
| polarC18 | 276.144 | 66.9002 | 8.7 | 0.17 |
| polarC18 | 276.144 | 53.6856 | 8.7 | 0.17 |
| polarC18 | 276.144 | 52.8968 | 8.7 | 0.17 |
| polarC18 | 276.144 | 252.254 | 8.7 | 0.17 |
| polarC18 | 276.144 | 68.5779 | 8.7 | 0.17 |
| polarC18 | 276.144 | 177.039 | 8.7 | 0.17 |
| polarC18 | 276.144 | 282.537 | 8.7 | 0.17 |
| polarC18 | 276.144 | 101.0325 | 8.7 | 0.17 |
| polarC18 | 276.144 | 274.0776 | 8.7 | 0.17 |
| polarC18 | 276.144 | 96.0911 | 8.7 | 0.17 |
| polarC18 | 276.144 | 115.1508 | 8.6 | 0.17 |
| polarC18 | 276.144 | 202.1742 | 8.6 | 0.17 |
| polarC18 | 276.144 | 74.0981 | 8.6 | 0.17 |
| polarC18 | 276.144 | 163.6065 | 8.6 | 0.17 |
| polarC18 | 276.144 | 70.5127 | 8.6 | 0.17 |
| polarC18 | 276.144 | 198.4081 | 8.6 | 0.17 |
| polarC18 | 276.144 | 79.5401 | 8.6 | 0.17 |
| polarC18 | 276.144 | 89.9757 | 8.6 | 0.17 |
| polarC18 | 276.144 | 175.4267 | 8.6 | 0.17 |
| polarC18 | 276.144 | 92.778 | 8.6 | 0.17 |
| polarC18 | 276.144 | 212.8348 | 8.6 | 0.17 |
| polarC18 | 276.144 | 94.0893 | 8.6 | 0.17 |
| polarC18 | 276.144 | 95.6359 | 8.6 | 0.16 |
| polarC18 | 276.144 | 133.7645 | 8.6 | 0.16 |
| polarC18 | 276.144 | 98.2425 | 8.6 | 0.16 |
| polarC18 | 276.144 | 72.7746 | 8.6 | 0.16 |
| polarC18 | 276.144 | 93.3314 | 8.6 | 0.16 |

| | | | | |
|---|---|---|---|---|
| polarC18 | 276.144 | 155.3629 | 8.6 | 0.16 |
| polarC18 | 276.144 | 91.4807 | 8.6 | 0.16 |
| polarC18 | 276.144 | 189.4332 | 8.6 | 0.16 |
| polarC18 | 276.144 | 59.1152 | 8.5 | 0.16 |
| polarC18 | 276.144 | 69.4144 | 8.5 | 0.16 |
| polarC18 | 276.144 | 146.8973 | 8.5 | 0.16 |
| polarC18 | 276.144 | 79.5801 | 8.5 | 0.16 |
| polarC18 | 276.144 | 81.3373 | 8.5 | 0.16 |
| polarC18 | 276.144 | 71.8073 | 8.5 | 0.16 |
| polarC18 | 276.144 | 127.2575 | 8.5 | 0.16 |
| polarC18 | 276.144 | 64.9547 | 8.5 | 0.16 |
| polarC18 | 276.144 | 52.7414 | 8.5 | 0.16 |
| polarC18 | 276.144 | 68.6869 | 8.5 | 0.16 |
| polarC18 | 276.144 | 73.2333 | 8.4 | 0.16 |
| polarC18 | 276.144 | 78.6613 | 8.4 | 0.16 |
| polarC18 | 276.144 | 53.7691 | 8.4 | 0.16 |
| polarC18 | 276.144 | 72.6822 | 8.4 | 0.16 |
| polarC18 | 276.144 | 57.8801 | 8.4 | 0.16 |
| polarC18 | 276.144 | 101.6875 | 8.4 | 0.16 |
| polarC18 | 276.144 | 80.0756 | 8.4 | 0.16 |
| polarC18 | 276.144 | 209.6703 | 8.4 | 0.16 |
| polarC18 | 276.144 | 198.5293 | 8.4 | 0.16 |
| polarC18 | 276.144 | 57.4001 | 8.4 | 0.16 |
| polarC18 | 276.144 | 221.625 | 8.4 | 0.16 |
| polarC18 | 276.144 | 184.1599 | 8.4 | 0.16 |
| polarC18 | 276.144 | 184.6205 | 8.4 | 0.16 |
| polarC18 | 276.144 | 142.986 | 8.4 | 0.16 |
| polarC18 | 276.144 | 147.6252 | 8.4 | 0.16 |
| polarC18 | 276.144 | 213.1828 | 8.4 | 0.16 |
| polarC18 | 276.144 | 158.3031 | 8.4 | 0.16 |
| polarC18 | 276.144 | 137.6075 | 8.4 | 0.16 |
| polarC18 | 276.144 | 102.0998 | 8.4 | 0.16 |
| polarC18 | 276.144 | 51.7646 | 8.4 | 0.16 |
| polarC18 | 276.144 | 273.697 | 8.4 | 0.16 |
| polarC18 | 276.144 | 109.4024 | 8.4 | 0.16 |
| polarC18 | 276.144 | 93.7331 | 8.3 | 0.16 |
| polarC18 | 276.144 | 246.9917 | 8.3 | 0.16 |
| polarC18 | 276.144 | 84.3684 | 8.3 | 0.16 |
| polarC18 | 276.144 | 177.983 | 8.3 | 0.16 |
| polarC18 | 276.144 | 82.8178 | 8.3 | 0.16 |
| polarC18 | 276.144 | 96.9171 | 8.3 | 0.16 |
| polarC18 | 276.144 | 56.9325 | 8.3 | 0.16 |

| | | | | |
|---|---|---|---|---|
| polarC18 | 276.144 | 81.9863 | 8.3 | 0.16 |
| polarC18 | 276.144 | 55.4551 | 8.3 | 0.16 |
| polarC18 | 276.144 | 116.5351 | 8.3 | 0.16 |
| polarC18 | 276.144 | 54.5329 | 8.3 | 0.16 |
| polarC18 | 276.144 | 88.2004 | 8.3 | 0.16 |
| polarC18 | 276.144 | 205.0485 | 8.3 | 0.16 |
| polarC18 | 276.144 | 257.8128 | 8.3 | 0.16 |
| polarC18 | 276.144 | 221.5663 | 8.3 | 0.16 |
| polarC18 | 276.144 | 50.8994 | 8.3 | 0.16 |
| polarC18 | 276.144 | 240.8574 | 8.3 | 0.16 |
| polarC18 | 276.144 | 89.7403 | 8.3 | 0.16 |
| polarC18 | 276.144 | 131.3769 | 8.3 | 0.16 |
| polarC18 | 276.144 | 82.9023 | 8.3 | 0.16 |
| polarC18 | 276.144 | 53.2152 | 8.3 | 0.16 |
| polarC18 | 276.144 | 238.7605 | 8.3 | 0.16 |
| polarC18 | 276.144 | 166.3119 | 8.2 | 0.16 |
| polarC18 | 276.144 | 188.9997 | 8.2 | 0.16 |
| polarC18 | 276.144 | 52.7456 | 8.2 | 0.16 |
| polarC18 | 276.144 | 67.9346 | 8.2 | 0.16 |
| polarC18 | 276.144 | 138.6564 | 8.2 | 0.16 |
| polarC18 | 276.144 | 96.5508 | 8.2 | 0.16 |
| polarC18 | 276.144 | 108.1418 | 8.2 | 0.16 |
| polarC18 | 276.144 | 93.2123 | 8.2 | 0.16 |
| polarC18 | 276.144 | 240.7936 | 8.2 | 0.16 |
| polarC18 | 276.144 | 109.0848 | 8.2 | 0.16 |
| polarC18 | 276.144 | 75.1899 | 8.2 | 0.16 |
| polarC18 | 276.144 | 74.6132 | 8.2 | 0.16 |
| polarC18 | 276.144 | 101.5854 | 8.2 | 0.16 |
| polarC18 | 276.144 | 192.9394 | 8.2 | 0.16 |
| polarC18 | 276.144 | 57.2814 | 8.2 | 0.16 |
| polarC18 | 276.144 | 105.5528 | 8.2 | 0.16 |
| polarC18 | 276.144 | 57.3101 | 8.2 | 0.16 |
| polarC18 | 276.144 | 81.9474 | 8.2 | 0.16 |
| polarC18 | 276.144 | 86.9943 | 8.2 | 0.16 |
| polarC18 | 276.144 | 102.9588 | 8.2 | 0.16 |
| polarC18 | 276.144 | 182.1645 | 8.2 | 0.16 |
| polarC18 | 276.144 | 84.2497 | 8.2 | 0.16 |
| polarC18 | 276.144 | 250.2972 | 8.2 | 0.16 |
| polarC18 | 276.144 | 56.7369 | 8.2 | 0.16 |
| polarC18 | 276.144 | 122.1743 | 8.2 | 0.16 |
| polarC18 | 276.144 | 52.7327 | 8.2 | 0.16 |
| polarC18 | 276.144 | 79.0674 | 8.2 | 0.16 |

| | | | | |
|---|---|---|---|---|
| polarC18 | 276.144 | 54.2048 | 8.2 | 0.16 |
| polarC18 | 276.144 | 54.4562 | 8.2 | 0.16 |
| polarC18 | 276.144 | 106.8847 | 8.2 | 0.16 |
| polarC18 | 276.144 | 134.9861 | 8.2 | 0.16 |
| polarC18 | 276.144 | 166.6382 | 8.2 | 0.16 |
| polarC18 | 276.144 | 79.4196 | 8.2 | 0.16 |
| polarC18 | 276.144 | 120.9331 | 8.2 | 0.16 |
| polarC18 | 276.144 | 62.6591 | 8.1 | 0.16 |
| polarC18 | 276.144 | 70.5796 | 8.1 | 0.16 |
| polarC18 | 276.144 | 64.5984 | 8.1 | 0.16 |
| polarC18 | 276.144 | 50.5611 | 8.1 | 0.16 |
| polarC18 | 276.144 | 136.5864 | 8.1 | 0.16 |
| polarC18 | 276.144 | 83.6294 | 8.1 | 0.16 |
| polarC18 | 276.144 | 50.3408 | 8.1 | 0.16 |
| polarC18 | 276.144 | 61.1735 | 8.1 | 0.16 |
| polarC18 | 276.144 | 179.2086 | 8.1 | 0.16 |
| polarC18 | 276.144 | 56.476 | 8.1 | 0.16 |
| polarC18 | 276.144 | 112.0986 | 8.1 | 0.16 |
| polarC18 | 276.144 | 89.9946 | 8.1 | 0.16 |
| polarC18 | 276.144 | 129.0173 | 8.1 | 0.16 |
| polarC18 | 276.144 | 77.7719 | 8.1 | 0.16 |
| polarC18 | 276.144 | 91.1014 | 8.1 | 0.16 |
| polarC18 | 276.144 | 114.9029 | 8.1 | 0.16 |
| polarC18 | 276.144 | 263.2424 | 8.1 | 0.16 |
| polarC18 | 276.144 | 50.9563 | 8.1 | 0.16 |
| polarC18 | 276.144 | 77.7886 | 8.1 | 0.16 |
| polarC18 | 276.144 | 238.8195 | 8.1 | 0.16 |
| polarC18 | 276.144 | 184.3992 | 8.1 | 0.16 |
| polarC18 | 276.144 | 53.4061 | 8.1 | 0.16 |
| polarC18 | 276.144 | 154.6896 | 8.1 | 0.16 |
| polarC18 | 276.144 | 180.1568 | 8.1 | 0.16 |
| polarC18 | 276.144 | 280.524 | 8.1 | 0.16 |
| polarC18 | 276.144 | 84.081 | 8.1 | 0.16 |
| polarC18 | 276.144 | 83.7796 | 8.1 | 0.16 |
| polarC18 | 276.144 | 253.731 | 8.1 | 0.16 |
| polarC18 | 276.144 | 151.3357 | 8.1 | 0.16 |
| polarC18 | 276.144 | 146.7708 | 8.1 | 0.16 |
| polarC18 | 276.144 | 70.3737 | 8.1 | 0.16 |
| polarC18 | 276.144 | 66.5494 | 8.1 | 0.16 |
| polarC18 | 276.144 | 232.6629 | 8.1 | 0.16 |
| polarC18 | 276.144 | 109.1836 | 8.1 | 0.16 |
| polarC18 | 276.144 | 107.7146 | 8.1 | 0.16 |

| | | | | |
|---|---|---|---|---|
| polarC18 | 276.144 | 204.4377 | 8.1 | 0.16 |
| polarC18 | 276.144 | 65.5621 | 8.1 | 0.16 |
| polarC18 | 276.144 | 170.2278 | 8.1 | 0.16 |
| polarC18 | 276.144 | 113.5405 | 8.1 | 0.15 |
| polarC18 | 276.144 | 83.5544 | 8.1 | 0.15 |
| polarC18 | 276.144 | 84.6818 | 8.1 | 0.15 |
| polarC18 | 276.144 | 57.3804 | 8 | 0.15 |
| polarC18 | 276.144 | 51.9537 | 8 | 0.15 |
| polarC18 | 276.144 | 75.819 | 8 | 0.15 |
| polarC18 | 276.144 | 144.3856 | 8 | 0.15 |
| polarC18 | 276.144 | 98.0842 | 8 | 0.15 |
| polarC18 | 276.144 | 73.6808 | 8 | 0.15 |
| polarC18 | 276.144 | 73.0735 | 8 | 0.15 |
| polarC18 | 276.144 | 165.3515 | 8 | 0.15 |
| polarC18 | 276.144 | 88.4874 | 8 | 0.15 |
| polarC18 | 276.144 | 93.6945 | 8 | 0.15 |
| polarC18 | 276.144 | 211.4938 | 8 | 0.15 |
| polarC18 | 276.144 | 229.085 | 8 | 0.15 |
| polarC18 | 276.144 | 68.928 | 8 | 0.15 |
| polarC18 | 276.144 | 277.4533 | 8 | 0.15 |
| polarC18 | 276.144 | 55.3502 | 8 | 0.15 |
| polarC18 | 276.144 | 62.9395 | 8 | 0.15 |
| polarC18 | 276.144 | 91.0678 | 8 | 0.15 |
| polarC18 | 276.144 | 215.2141 | 8 | 0.15 |
| polarC18 | 276.144 | 259.4911 | 8 | 0.15 |
| polarC18 | 276.144 | 51.0075 | 8 | 0.15 |
| polarC18 | 276.144 | 242.8855 | 8 | 0.15 |
| polarC18 | 276.144 | 60.8309 | 8 | 0.15 |
| polarC18 | 276.144 | 251.4613 | 8 | 0.15 |
| polarC18 | 276.144 | 79.9724 | 8 | 0.15 |
| polarC18 | 276.144 | 167.3135 | 8 | 0.15 |
| polarC18 | 276.144 | 214.2784 | 8 | 0.15 |
| polarC18 | 276.144 | 80.6951 | 8 | 0.15 |
| polarC18 | 276.144 | 151.4138 | 8 | 0.15 |
| polarC18 | 276.144 | 63.7949 | 8 | 0.15 |
| polarC18 | 276.144 | 145.8094 | 8 | 0.15 |
| polarC18 | 276.144 | 103.7639 | 8 | 0.15 |
| polarC18 | 276.144 | 70.2201 | 8 | 0.15 |
| polarC18 | 276.144 | 100.7907 | 8 | 0.15 |
| polarC18 | 276.144 | 63.4735 | 8 | 0.15 |
| polarC18 | 276.144 | 66.9226 | 8 | 0.15 |
| polarC18 | 276.144 | 114.8326 | 8 | 0.15 |

| | | | | |
|---|---|---|---|---|
| polarC18 | 276.144 | 156.7145 | 8 | 0.15 |
| polarC18 | 276.144 | 155.0939 | 8 | 0.15 |
| polarC18 | 276.144 | 116.0881 | 8 | 0.15 |
| polarC18 | 276.144 | 51.0929 | 8 | 0.15 |
| polarC18 | 276.144 | 139.9829 | 8 | 0.15 |
| polarC18 | 276.144 | 117.5689 | 8 | 0.15 |
| polarC18 | 276.144 | 104.3866 | 8 | 0.15 |
| polarC18 | 276.144 | 59.7246 | 8 | 0.15 |
| polarC18 | 276.144 | 106.6777 | 8 | 0.15 |
| polarC18 | 276.144 | 264.0522 | 8 | 0.15 |
| polarC18 | 276.144 | 51.1722 | 8 | 0.15 |
| polarC18 | 276.144 | 84.5305 | 8 | 0.15 |
| polarC18 | 276.144 | 100.1332 | 8 | 0.15 |
| polarC18 | 276.144 | 95.9903 | 8 | 0.15 |
| polarC18 | 276.144 | 269.5722 | 8 | 0.15 |
| polarC18 | 276.144 | 100.1768 | 8 | 0.15 |
| polarC18 | 276.144 | 90.2454 | 7.9 | 0.15 |
| polarC18 | 276.144 | 160.9208 | 7.9 | 0.15 |
| polarC18 | 276.144 | 81.4784 | 7.9 | 0.15 |
| polarC18 | 276.144 | 143.8493 | 7.9 | 0.15 |
| polarC18 | 276.144 | 81.6133 | 7.9 | 0.15 |
| polarC18 | 276.144 | 143.6575 | 7.9 | 0.15 |
| polarC18 | 276.144 | 62.5238 | 7.9 | 0.15 |
| polarC18 | 276.144 | 94.8001 | 7.9 | 0.15 |
| polarC18 | 276.144 | 103.2376 | 7.9 | 0.15 |
| polarC18 | 276.144 | 92.4846 | 7.9 | 0.15 |
| polarC18 | 276.144 | 197.5031 | 7.9 | 0.15 |
| polarC18 | 276.144 | 105.8372 | 7.9 | 0.15 |
| polarC18 | 276.144 | 50.9726 | 7.9 | 0.15 |
| polarC18 | 276.144 | 238.2075 | 7.9 | 0.15 |
| polarC18 | 276.144 | 174.7417 | 7.9 | 0.15 |
| polarC18 | 276.144 | 91.2124 | 7.9 | 0.15 |
| polarC18 | 276.144 | 213.514 | 7.9 | 0.15 |
| polarC18 | 276.144 | 293.5181 | 7.9 | 0.15 |
| polarC18 | 276.144 | 66.0182 | 7.9 | 0.15 |
| polarC18 | 276.144 | 90.1781 | 7.9 | 0.15 |
| polarC18 | 276.144 | 68.5038 | 7.9 | 0.15 |
| polarC18 | 276.144 | 57.4734 | 7.9 | 0.15 |
| polarC18 | 276.144 | 277.0146 | 7.9 | 0.15 |
| polarC18 | 276.144 | 78.6044 | 7.9 | 0.15 |
| polarC18 | 276.144 | 81.6522 | 7.9 | 0.15 |
| polarC18 | 276.144 | 57.014 | 7.9 | 0.15 |

| | | | | |
|---|---|---|---|---|
| polarC18 | 276.144 | 158.6039 | 7.9 | 0.15 |
| polarC18 | 276.144 | 231.011 | 7.9 | 0.15 |
| polarC18 | 276.144 | 221.1743 | 7.9 | 0.15 |
| polarC18 | 276.144 | 114.2391 | 7.9 | 0.15 |
| polarC18 | 276.144 | 64.3398 | 7.9 | 0.15 |
| polarC18 | 276.144 | 281.9659 | 7.9 | 0.15 |
| polarC18 | 276.144 | 79.3451 | 7.9 | 0.15 |
| polarC18 | 276.144 | 273.5779 | 7.9 | 0.15 |
| polarC18 | 276.144 | 81.7651 | 7.9 | 0.15 |
| polarC18 | 276.144 | 268.229 | 7.9 | 0.15 |
| polarC18 | 276.144 | 243.462 | 7.9 | 0.15 |
| polarC18 | 276.144 | 92.5792 | 7.9 | 0.15 |
| polarC18 | 276.144 | 261.4989 | 7.9 | 0.15 |
| polarC18 | 276.144 | 134.9014 | 7.9 | 0.15 |
| polarC18 | 276.144 | 200.4261 | 7.9 | 0.15 |
| polarC18 | 276.144 | 163.9246 | 7.9 | 0.15 |
| polarC18 | 276.144 | 50.9327 | 7.9 | 0.15 |
| polarC18 | 276.144 | 243.417 | 7.9 | 0.15 |
| polarC18 | 276.144 | 55.5276 | 7.9 | 0.15 |
| polarC18 | 276.144 | 50.4495 | 7.9 | 0.15 |
| polarC18 | 276.144 | 51.8264 | 7.9 | 0.15 |
| polarC18 | 276.144 | 58.1432 | 7.9 | 0.15 |
| polarC18 | 276.144 | 100.5719 | 7.9 | 0.15 |
| polarC18 | 276.144 | 55.2401 | 7.9 | 0.15 |
| polarC18 | 276.144 | 136.9849 | 7.9 | 0.15 |
| polarC18 | 276.144 | 75.7428 | 7.9 | 0.15 |
| polarC18 | 276.144 | 104.5957 | 7.9 | 0.15 |
| polarC18 | 276.144 | 83.0649 | 7.9 | 0.15 |
| polarC18 | 276.144 | 101.4424 | 7.9 | 0.15 |
| polarC18 | 276.144 | 201.1593 | 7.9 | 0.15 |
| polarC18 | 276.144 | 59.0998 | 7.9 | 0.15 |
| polarC18 | 276.144 | 59.0911 | 7.9 | 0.15 |
| polarC18 | 276.144 | 206.7207 | 7.9 | 0.15 |
| polarC18 | 276.144 | 217.4288 | 7.9 | 0.15 |
| polarC18 | 276.144 | 72.6971 | 7.9 | 0.15 |
| polarC18 | 276.144 | 122.6748 | 7.9 | 0.15 |
| polarC18 | 276.144 | 79.588 | 7.9 | 0.15 |
| polarC18 | 276.144 | 66.2599 | 7.9 | 0.15 |
| polarC18 | 276.144 | 262.2185 | 7.9 | 0.15 |
| polarC18 | 276.144 | 50.9238 | 7.9 | 0.15 |
| polarC18 | 276.144 | 74.2496 | 7.9 | 0.15 |
| polarC18 | 276.144 | 103.9016 | 7.9 | 0.15 |

| | | | | |
|---|---|---|---|---|
| polarC18 | 276.144 | 74.8617 | 7.9 | 0.15 |
| polarC18 | 276.144 | 51.0953 | 7.9 | 0.15 |
| polarC18 | 276.144 | 199.0208 | 7.9 | 0.15 |
| polarC18 | 276.144 | 52.6827 | 7.9 | 0.15 |
| polarC18 | 276.144 | 67.5392 | 7.9 | 0.15 |
| polarC18 | 276.144 | 77.9842 | 7.8 | 0.15 |
| polarC18 | 276.144 | 207.0152 | 7.8 | 0.15 |
| polarC18 | 276.144 | 144.1816 | 7.8 | 0.15 |
| polarC18 | 276.144 | 174.2499 | 7.8 | 0.15 |
| polarC18 | 276.144 | 90.0268 | 7.8 | 0.15 |
| polarC18 | 276.144 | 57.1809 | 7.8 | 0.15 |
| polarC18 | 276.144 | 62.7028 | 7.8 | 0.15 |
| polarC18 | 276.144 | 140.6174 | 7.8 | 0.15 |
| polarC18 | 276.144 | 53.8854 | 7.8 | 0.15 |
| polarC18 | 276.144 | 71.5658 | 7.8 | 0.15 |
| polarC18 | 276.144 | 51.9729 | 7.8 | 0.15 |
| polarC18 | 276.144 | 113.4227 | 7.8 | 0.15 |
| polarC18 | 276.144 | 129.2199 | 7.8 | 0.15 |
| polarC18 | 276.144 | 68.4308 | 7.8 | 0.15 |
| polarC18 | 276.144 | 66.5657 | 7.8 | 0.15 |
| polarC18 | 276.144 | 88.5849 | 7.8 | 0.15 |
| polarC18 | 276.144 | 150.2978 | 7.8 | 0.15 |
| polarC18 | 276.144 | 53.0845 | 7.8 | 0.15 |
| polarC18 | 276.144 | 89.7305 | 7.8 | 0.15 |
| polarC18 | 276.144 | 63.7434 | 7.8 | 0.15 |
| polarC18 | 276.144 | 136.7217 | 7.8 | 0.15 |
| polarC18 | 276.144 | 188.1621 | 7.8 | 0.15 |
| polarC18 | 276.144 | 236.9044 | 7.8 | 0.15 |
| polarC18 | 276.144 | 88.8407 | 7.8 | 0.15 |
| polarC18 | 276.144 | 93.8636 | 7.8 | 0.15 |
| polarC18 | 276.144 | 286.4989 | 7.8 | 0.15 |
| polarC18 | 276.144 | 178.5738 | 7.8 | 0.15 |
| polarC18 | 276.144 | 73.5254 | 7.8 | 0.15 |
| polarC18 | 276.144 | 109.3871 | 7.8 | 0.15 |
| polarC18 | 276.144 | 63.4322 | 7.8 | 0.15 |
| polarC18 | 276.144 | 90.084 | 7.8 | 0.15 |
| polarC18 | 276.144 | 70.3064 | 7.8 | 0.15 |
| polarC18 | 276.144 | 59.9525 | 7.8 | 0.15 |
| polarC18 | 276.144 | 137.0985 | 7.8 | 0.15 |
| polarC18 | 276.144 | 95.9444 | 7.8 | 0.15 |
| polarC18 | 276.144 | 63.7183 | 7.8 | 0.15 |
| polarC18 | 276.144 | 98.199 | 7.8 | 0.15 |

| | | | | |
|---|---|---|---|---|
| polarC18 | 276.144 | 105.7814 | 7.8 | 0.15 |
| polarC18 | 276.144 | 109.1708 | 7.8 | 0.15 |
| polarC18 | 276.144 | 207.0423 | 7.8 | 0.15 |
| polarC18 | 276.144 | 236.6952 | 7.8 | 0.15 |
| polarC18 | 276.144 | 61.437 | 7.8 | 0.15 |
| polarC18 | 276.144 | 138.9953 | 7.8 | 0.15 |
| polarC18 | 276.144 | 96.2975 | 7.8 | 0.15 |
| polarC18 | 276.144 | 56.3239 | 7.8 | 0.15 |
| polarC18 | 276.144 | 57.7378 | 7.8 | 0.15 |
| polarC18 | 276.144 | 119.2215 | 7.8 | 0.15 |
| polarC18 | 276.144 | 223.6719 | 7.8 | 0.15 |
| polarC18 | 276.144 | 57.8922 | 7.8 | 0.15 |
| polarC18 | 276.144 | 77.4503 | 7.8 | 0.15 |
| polarC18 | 276.144 | 64.1925 | 7.8 | 0.15 |
| polarC18 | 276.144 | 108.5334 | 7.8 | 0.15 |
| polarC18 | 276.144 | 277.9908 | 7.8 | 0.15 |
| polarC18 | 276.144 | 159.9459 | 7.8 | 0.15 |
| polarC18 | 276.144 | 67.9464 | 7.8 | 0.15 |
| polarC18 | 276.144 | 59.8692 | 7.8 | 0.15 |
| polarC18 | 276.144 | 54.1973 | 7.8 | 0.15 |
| polarC18 | 276.144 | 190.4763 | 7.8 | 0.15 |
| polarC18 | 276.144 | 74.5193 | 7.8 | 0.15 |
| polarC18 | 276.144 | 99.3309 | 7.8 | 0.15 |
| polarC18 | 276.144 | 88.3286 | 7.8 | 0.15 |
| polarC18 | 276.144 | 131.4496 | 7.8 | 0.15 |
| polarC18 | 276.144 | 112.4539 | 7.8 | 0.15 |
| polarC18 | 276.144 | 94.5793 | 7.8 | 0.15 |
| polarC18 | 276.144 | 162.2202 | 7.8 | 0.15 |
| polarC18 | 276.144 | 230.965 | 7.8 | 0.15 |
| polarC18 | 276.144 | 177.2133 | 7.8 | 0.15 |
| polarC18 | 276.144 | 117.644 | 7.8 | 0.15 |
| polarC18 | 276.144 | 290.2966 | 7.8 | 0.15 |
| polarC18 | 276.144 | 65.4737 | 7.8 | 0.15 |
| polarC18 | 276.144 | 139.5042 | 7.8 | 0.15 |
| polarC18 | 276.144 | 134.3991 | 7.8 | 0.15 |
| polarC18 | 276.144 | 73.3215 | 7.8 | 0.15 |
| polarC18 | 276.144 | 161.3709 | 7.8 | 0.15 |
| polarC18 | 276.144 | 139.6412 | 7.8 | 0.15 |
| polarC18 | 276.144 | 56.8237 | 7.8 | 0.15 |
| polarC18 | 276.144 | 283.7858 | 7.8 | 0.15 |
| polarC18 | 276.144 | 73.2831 | 7.8 | 0.15 |
| polarC18 | 276.144 | 92.7959 | 7.7 | 0.15 |

| | | | | |
|---|---|---|---|---|
| polarC18 | 276.144 | 58.3281 | 7.7 | 0.15 |
| polarC18 | 276.144 | 226.246 | 7.7 | 0.15 |
| polarC18 | 276.144 | 86.4809 | 7.7 | 0.15 |
| polarC18 | 276.144 | 64.1785 | 7.7 | 0.15 |
| polarC18 | 276.144 | 124.0045 | 7.7 | 0.15 |
| polarC18 | 276.144 | 50.9902 | 7.7 | 0.15 |
| polarC18 | 276.144 | 61.4289 | 7.7 | 0.15 |
| polarC18 | 276.144 | 287.1054 | 7.7 | 0.15 |
| polarC18 | 276.144 | 86.4323 | 7.7 | 0.15 |
| polarC18 | 276.144 | 258.7484 | 7.7 | 0.15 |
| polarC18 | 276.144 | 268.2916 | 7.7 | 0.15 |
| polarC18 | 276.144 | 57.8838 | 7.7 | 0.15 |
| polarC18 | 276.144 | 172.1042 | 7.7 | 0.15 |
| polarC18 | 276.144 | 84.1201 | 7.7 | 0.15 |
| polarC18 | 276.144 | 226.0352 | 7.7 | 0.15 |
| polarC18 | 276.144 | 94.7122 | 7.7 | 0.15 |
| polarC18 | 276.144 | 191.4135 | 7.7 | 0.15 |
| polarC18 | 276.144 | 220.1631 | 7.7 | 0.15 |
| polarC18 | 276.144 | 102.5634 | 7.7 | 0.15 |
| polarC18 | 276.144 | 84.2845 | 7.7 | 0.15 |
| polarC18 | 276.144 | 58.3341 | 7.7 | 0.15 |
| polarC18 | 276.144 | 246.3782 | 7.7 | 0.15 |
| polarC18 | 276.144 | 77.6608 | 7.7 | 0.15 |
| polarC18 | 276.144 | 53.7558 | 7.7 | 0.15 |
| polarC18 | 276.144 | 95.2021 | 7.7 | 0.15 |
| polarC18 | 276.144 | 84.5438 | 7.7 | 0.15 |
| polarC18 | 276.144 | 60.4445 | 7.7 | 0.15 |
| polarC18 | 276.144 | 283.9559 | 7.7 | 0.15 |
| polarC18 | 276.144 | 68.7962 | 7.7 | 0.15 |
| polarC18 | 276.144 | 75.2901 | 7.7 | 0.15 |
| polarC18 | 276.144 | 139.5516 | 7.7 | 0.15 |
| polarC18 | 276.144 | 130.1299 | 7.7 | 0.15 |
| polarC18 | 276.144 | 148.726 | 7.7 | 0.15 |
| polarC18 | 276.144 | 147.1332 | 7.7 | 0.15 |
| polarC18 | 276.144 | 280.4442 | 7.7 | 0.15 |
| polarC18 | 276.144 | 77.3212 | 7.7 | 0.15 |
| polarC18 | 276.144 | 66.1166 | 7.7 | 0.15 |
| polarC18 | 276.144 | 177.6102 | 7.7 | 0.15 |
| polarC18 | 276.144 | 147.3954 | 7.7 | 0.15 |
| polarC18 | 276.144 | 108.9461 | 7.7 | 0.15 |
| polarC18 | 276.144 | 52.2038 | 7.7 | 0.15 |
| polarC18 | 276.144 | 79.243 | 7.7 | 0.15 |

| | | | | |
|---|---|---|---|---|
| polarC18 | 276.144 | 194.1533 | 7.7 | 0.15 |
| polarC18 | 276.144 | 254.1991 | 7.7 | 0.15 |
| polarC18 | 276.144 | 73.9821 | 7.7 | 0.15 |
| polarC18 | 276.144 | 169.7058 | 7.7 | 0.15 |
| polarC18 | 276.144 | 120.9445 | 7.7 | 0.15 |
| polarC18 | 276.144 | 119.2303 | 7.7 | 0.15 |
| polarC18 | 276.144 | 279.0264 | 7.7 | 0.15 |
| polarC18 | 276.144 | 94.6066 | 7.7 | 0.15 |
| polarC18 | 276.144 | 132.9275 | 7.7 | 0.15 |
| polarC18 | 276.144 | 228.3478 | 7.7 | 0.15 |
| polarC18 | 276.144 | 72.1915 | 7.7 | 0.15 |
| polarC18 | 276.144 | 134.9618 | 7.7 | 0.15 |
| polarC18 | 276.144 | 74.9753 | 7.7 | 0.15 |
| polarC18 | 276.144 | 52.6966 | 7.7 | 0.15 |
| polarC18 | 276.144 | 66.6428 | 7.7 | 0.15 |
| polarC18 | 276.144 | 210.3446 | 7.7 | 0.15 |
| polarC18 | 276.144 | 136.8309 | 7.7 | 0.15 |
| polarC18 | 276.144 | 78.736 | 7.7 | 0.15 |
| polarC18 | 276.144 | 75.4856 | 7.7 | 0.15 |
| polarC18 | 276.144 | 289.4652 | 7.7 | 0.15 |
| polarC18 | 276.144 | 53.1393 | 7.7 | 0.15 |
| polarC18 | 276.144 | 82.6105 | 7.7 | 0.15 |
| polarC18 | 276.144 | 271.8587 | 7.7 | 0.15 |
| polarC18 | 276.144 | 62.0516 | 7.7 | 0.15 |
| polarC18 | 276.144 | 246.0662 | 7.7 | 0.15 |
| polarC18 | 276.144 | 194.7532 | 7.7 | 0.15 |
| polarC18 | 276.144 | 195.0879 | 7.7 | 0.15 |
| polarC18 | 276.144 | 85.2866 | 7.7 | 0.15 |
| polarC18 | 276.144 | 63.9467 | 7.7 | 0.15 |
| polarC18 | 276.144 | 121.6248 | 7.7 | 0.15 |
| polarC18 | 276.144 | 137.5047 | 7.7 | 0.15 |
| polarC18 | 276.144 | 155.4812 | 7.7 | 0.15 |
| polarC18 | 276.144 | 113.6347 | 7.7 | 0.15 |
| polarC18 | 276.144 | 57.3149 | 7.7 | 0.15 |
| polarC18 | 276.144 | 61.6368 | 7.7 | 0.15 |
| polarC18 | 276.144 | 235.6162 | 7.7 | 0.15 |
| polarC18 | 276.144 | 76.0953 | 7.7 | 0.15 |
| polarC18 | 276.144 | 53.4765 | 7.7 | 0.15 |
| polarC18 | 276.144 | 53.6835 | 7.7 | 0.15 |
| polarC18 | 276.144 | 62.8075 | 7.7 | 0.15 |
| polarC18 | 276.144 | 69.6606 | 7.7 | 0.15 |
| polarC18 | 276.144 | 143.0463 | 7.7 | 0.15 |

| | | | | |
|---|---|---|---|---|
| polarC18 | 276.144 | 66.8782 | 7.7 | 0.15 |
| polarC18 | 276.144 | 132.6354 | 7.7 | 0.15 |
| polarC18 | 276.144 | 90.3385 | 7.7 | 0.15 |
| polarC18 | 276.144 | 50.6065 | 7.7 | 0.15 |
| polarC18 | 276.144 | 60.269 | 7.7 | 0.15 |
| polarC18 | 276.144 | 73.11 | 7.7 | 0.15 |
| polarC18 | 276.144 | 218.2071 | 7.7 | 0.15 |
| polarC18 | 276.144 | 178.108 | 7.7 | 0.15 |
| polarC18 | 276.144 | 93.0608 | 7.7 | 0.15 |
| polarC18 | 276.144 | 163.1908 | 7.7 | 0.15 |
| polarC18 | 276.144 | 240.9966 | 7.7 | 0.15 |
| polarC18 | 276.144 | 272.2227 | 7.7 | 0.15 |
| polarC18 | 276.144 | 83.8808 | 7.7 | 0.15 |
| polarC18 | 276.144 | 240.0272 | 7.7 | 0.15 |
| polarC18 | 276.144 | 73.4048 | 7.7 | 0.15 |
| polarC18 | 276.144 | 52.7898 | 7.7 | 0.15 |
| polarC18 | 276.144 | 198.2439 | 7.7 | 0.15 |
| polarC18 | 276.144 | 96.4351 | 7.7 | 0.15 |
| polarC18 | 276.144 | 94.1007 | 7.7 | 0.15 |
| polarC18 | 276.144 | 90.0213 | 7.6 | 0.15 |
| polarC18 | 276.144 | 120.1612 | 7.6 | 0.15 |
| polarC18 | 276.144 | 148.2408 | 7.6 | 0.15 |
| polarC18 | 276.144 | 58.2651 | 7.6 | 0.15 |
| polarC18 | 276.144 | 232.0155 | 7.6 | 0.15 |
| polarC18 | 276.144 | 93.4395 | 7.6 | 0.15 |
| polarC18 | 276.144 | 70.4236 | 7.6 | 0.15 |
| polarC18 | 276.144 | 82.3171 | 7.6 | 0.15 |
| polarC18 | 276.144 | 123.0126 | 7.6 | 0.15 |
| polarC18 | 276.144 | 78.563 | 7.6 | 0.15 |
| polarC18 | 276.144 | 240.7356 | 7.6 | 0.15 |
| polarC18 | 276.144 | 173.7416 | 7.6 | 0.15 |
| polarC18 | 276.144 | 213.1042 | 7.6 | 0.15 |
| polarC18 | 276.144 | 57.574 | 7.6 | 0.15 |
| polarC18 | 276.144 | 227.2901 | 7.6 | 0.15 |
| polarC18 | 276.144 | 286.1918 | 7.6 | 0.15 |
| polarC18 | 276.144 | 56.8665 | 7.6 | 0.15 |
| polarC18 | 276.144 | 65.5953 | 7.6 | 0.15 |
| polarC18 | 276.144 | 65.9233 | 7.6 | 0.15 |
| polarC18 | 276.144 | 71.3309 | 7.6 | 0.15 |
| polarC18 | 276.144 | 69.8684 | 7.6 | 0.15 |
| polarC18 | 276.144 | 77.6662 | 7.6 | 0.15 |
| polarC18 | 276.144 | 51.5494 | 7.6 | 0.15 |

| | | | | |
|---|---|---|---|---|
| polarC18 | 276.144 | 145.0336 | 7.6 | 0.15 |
| polarC18 | 276.144 | 52.2719 | 7.6 | 0.15 |
| polarC18 | 276.144 | 188.0809 | 7.6 | 0.15 |
| polarC18 | 276.144 | 146.0714 | 7.6 | 0.15 |
| polarC18 | 276.144 | 73.8286 | 7.6 | 0.15 |
| polarC18 | 276.144 | 183.9731 | 7.6 | 0.15 |
| polarC18 | 276.144 | 86.7559 | 7.6 | 0.15 |
| polarC18 | 276.144 | 147.3836 | 7.6 | 0.15 |
| polarC18 | 276.144 | 139.9049 | 7.6 | 0.15 |
| polarC18 | 276.144 | 111.6736 | 7.6 | 0.15 |
| polarC18 | 276.144 | 110.9853 | 7.6 | 0.15 |
| polarC18 | 276.144 | 64.5677 | 7.6 | 0.15 |
| polarC18 | 276.144 | 54.3649 | 7.6 | 0.15 |
| polarC18 | 276.144 | 258.5549 | 7.6 | 0.15 |
| polarC18 | 276.144 | 147.0119 | 7.6 | 0.15 |
| polarC18 | 276.144 | 125.6901 | 7.6 | 0.15 |
| polarC18 | 276.144 | 119.7893 | 7.6 | 0.15 |
| polarC18 | 276.144 | 254.8743 | 7.6 | 0.15 |
| polarC18 | 276.144 | 51.6929 | 7.6 | 0.15 |
| polarC18 | 276.144 | 88.5927 | 7.6 | 0.15 |
| polarC18 | 276.144 | 68.6626 | 7.6 | 0.15 |
| polarC18 | 276.144 | 106.8362 | 7.6 | 0.15 |
| polarC18 | 276.144 | 254.6155 | 7.6 | 0.15 |
| polarC18 | 276.144 | 84.9594 | 7.6 | 0.15 |
| polarC18 | 276.144 | 53.575 | 7.6 | 0.15 |
| polarC18 | 276.144 | 71.537 | 7.6 | 0.15 |
| polarC18 | 276.144 | 86.9671 | 7.6 | 0.15 |
| polarC18 | 276.144 | 51.8619 | 7.6 | 0.15 |
| polarC18 | 276.144 | 86.4509 | 7.6 | 0.15 |
| polarC18 | 276.144 | 293.2318 | 7.6 | 0.15 |
| polarC18 | 276.144 | 51.755 | 7.6 | 0.15 |
| polarC18 | 276.144 | 258.8414 | 7.6 | 0.15 |
| polarC18 | 276.144 | 63.9113 | 7.6 | 0.15 |
| polarC18 | 276.144 | 75.3692 | 7.6 | 0.15 |
| polarC18 | 276.144 | 67.6981 | 7.6 | 0.15 |
| polarC18 | 276.144 | 75.058 | 7.6 | 0.15 |
| polarC18 | 276.144 | 62.1164 | 7.6 | 0.15 |
| polarC18 | 276.144 | 54.9771 | 7.6 | 0.15 |
| polarC18 | 276.144 | 51.0904 | 7.6 | 0.15 |
| polarC18 | 276.144 | 66.1978 | 7.6 | 0.15 |
| polarC18 | 276.144 | 58.9349 | 7.6 | 0.15 |
| polarC18 | 276.144 | 55.2278 | 7.6 | 0.15 |

| | | | | |
|---|---|---|---|---|
| polarC18 | 276.144 | 50.7846 | 7.6 | 0.15 |
| polarC18 | 276.144 | 296.5781 | 7.6 | 0.15 |
| polarC18 | 276.144 | 101.1588 | 7.6 | 0.15 |
| polarC18 | 276.144 | 220.6348 | 7.6 | 0.15 |
| polarC18 | 276.144 | 60.2846 | 7.6 | 0.15 |
| polarC18 | 276.144 | 84.1275 | 7.6 | 0.15 |
| polarC18 | 276.144 | 69.8793 | 7.6 | 0.15 |
| polarC18 | 276.144 | 222.1353 | 7.6 | 0.15 |
| polarC18 | 276.144 | 192.7718 | 7.6 | 0.15 |
| polarC18 | 276.144 | 107.8519 | 7.6 | 0.15 |
| polarC18 | 276.144 | 200.4579 | 7.6 | 0.15 |
| polarC18 | 276.144 | 60.4222 | 7.6 | 0.15 |
| polarC18 | 276.144 | 50.0796 | 7.6 | 0.15 |
| polarC18 | 276.144 | 63.807 | 7.6 | 0.15 |
| polarC18 | 276.144 | 67.0543 | 7.6 | 0.15 |
| polarC18 | 276.144 | 177.4117 | 7.6 | 0.15 |
| polarC18 | 276.144 | 107.3344 | 7.6 | 0.15 |
| polarC18 | 276.144 | 107.4322 | 7.6 | 0.15 |
| polarC18 | 276.144 | 56.8427 | 7.6 | 0.15 |
| polarC18 | 276.144 | 51.8606 | 7.6 | 0.15 |
| polarC18 | 276.144 | 53.9366 | 7.6 | 0.15 |
| polarC18 | 276.144 | 128.3078 | 7.6 | 0.15 |
| polarC18 | 276.144 | 67.6645 | 7.6 | 0.15 |
| polarC18 | 276.144 | 89.379 | 7.6 | 0.15 |
| polarC18 | 276.144 | 106.1393 | 7.6 | 0.15 |
| polarC18 | 276.144 | 102.5474 | 7.6 | 0.15 |
| polarC18 | 276.144 | 188.755 | 7.6 | 0.15 |
| polarC18 | 276.144 | 88.9211 | 7.6 | 0.15 |
| polarC18 | 276.144 | 94.818 | 7.6 | 0.15 |
| polarC18 | 276.144 | 273.9136 | 7.6 | 0.15 |
| polarC18 | 276.144 | 252.2171 | 7.6 | 0.15 |
| polarC18 | 276.144 | 50.0503 | 7.6 | 0.15 |
| polarC18 | 276.144 | 59.2908 | 7.6 | 0.15 |
| polarC18 | 276.144 | 56.3166 | 7.6 | 0.15 |
| polarC18 | 276.144 | 241.9206 | 7.6 | 0.15 |
| polarC18 | 276.144 | 95.8065 | 7.6 | 0.15 |
| polarC18 | 276.144 | 260.4665 | 7.6 | 0.15 |
| polarC18 | 276.144 | 61.4597 | 7.6 | 0.15 |
| polarC18 | 276.144 | 81.0708 | 7.6 | 0.15 |
| polarC18 | 276.144 | 202.3617 | 7.6 | 0.15 |
| polarC18 | 276.144 | 149.5127 | 7.6 | 0.15 |
| polarC18 | 276.144 | 56.9776 | 7.6 | 0.15 |

| | | | | |
|---|---|---|---|---|
| polarC18 | 276.144 | 51.8232 | 7.6 | 0.15 |
| polarC18 | 276.144 | 232.63 | 7.6 | 0.15 |
| polarC18 | 276.144 | 65.0275 | 7.6 | 0.15 |
| polarC18 | 276.144 | 169.2421 | 7.6 | 0.15 |
| polarC18 | 276.144 | 229.2526 | 7.6 | 0.15 |
| polarC18 | 276.144 | 72.2092 | 7.5 | 0.15 |
| polarC18 | 276.144 | 157.525 | 7.5 | 0.15 |
| polarC18 | 276.144 | 103.6612 | 7.5 | 0.15 |
| polarC18 | 276.144 | 220.888 | 7.5 | 0.15 |
| polarC18 | 276.144 | 112.2848 | 7.5 | 0.15 |
| polarC18 | 276.144 | 103.7063 | 7.5 | 0.15 |
| polarC18 | 276.144 | 251.693 | 7.5 | 0.15 |
| polarC18 | 276.144 | 59.3575 | 7.5 | 0.15 |
| polarC18 | 276.144 | 82.134 | 7.5 | 0.15 |
| polarC18 | 276.144 | 75.0002 | 7.5 | 0.15 |
| polarC18 | 276.144 | 83.9728 | 7.5 | 0.15 |
| polarC18 | 276.144 | 60.7897 | 7.5 | 0.14 |
| polarC18 | 276.144 | 158.2133 | 7.5 | 0.14 |
| polarC18 | 276.144 | 215.0432 | 7.5 | 0.14 |
| polarC18 | 276.144 | 73.7346 | 7.5 | 0.14 |
| polarC18 | 276.144 | 52.2023 | 7.5 | 0.14 |
| polarC18 | 276.144 | 83.2762 | 7.5 | 0.14 |
| polarC18 | 276.144 | 173.0865 | 7.5 | 0.14 |
| polarC18 | 276.144 | 116.2305 | 7.5 | 0.14 |
| polarC18 | 276.144 | 57.809 | 7.5 | 0.14 |
| polarC18 | 276.144 | 82.3341 | 7.5 | 0.14 |
| polarC18 | 276.144 | 66.0524 | 7.5 | 0.14 |
| polarC18 | 276.144 | 263.4601 | 7.5 | 0.14 |
| polarC18 | 276.144 | 91.8167 | 7.5 | 0.14 |
| polarC18 | 276.144 | 112.6884 | 7.5 | 0.14 |
| polarC18 | 276.144 | 73.8038 | 7.5 | 0.14 |
| polarC18 | 276.144 | 121.2412 | 7.5 | 0.14 |
| polarC18 | 276.144 | 91.2188 | 7.5 | 0.14 |
| polarC18 | 276.144 | 95.0343 | 7.5 | 0.14 |
| polarC18 | 276.144 | 53.1639 | 7.5 | 0.14 |
| polarC18 | 276.144 | 136.929 | 7.5 | 0.14 |
| polarC18 | 276.144 | 117.1942 | 7.5 | 0.14 |
| polarC18 | 276.144 | 98.1518 | 7.5 | 0.14 |
| polarC18 | 276.144 | 73.6306 | 7.5 | 0.14 |
| polarC18 | 276.144 | 156.4506 | 7.5 | 0.14 |
| polarC18 | 276.144 | 58.7247 | 7.5 | 0.14 |
| polarC18 | 276.144 | 78.6205 | 7.5 | 0.14 |

| | | | | |
|---|---|---|---|---|
| polarC18 | 276.144 | 117.6016 | 7.5 | 0.14 |
| polarC18 | 276.144 | 67.9236 | 7.5 | 0.14 |
| polarC18 | 276.144 | 183.7551 | 7.5 | 0.14 |
| polarC18 | 276.144 | 169.4118 | 7.5 | 0.14 |
| polarC18 | 276.144 | 81.5348 | 7.5 | 0.14 |
| polarC18 | 276.144 | 128.8177 | 7.5 | 0.14 |
| polarC18 | 276.144 | 173.9781 | 7.5 | 0.14 |
| polarC18 | 276.144 | 106.6556 | 7.5 | 0.14 |
| polarC18 | 276.144 | 58.8184 | 7.5 | 0.14 |
| polarC18 | 276.144 | 211.146 | 7.5 | 0.14 |
| polarC18 | 276.144 | 96.2583 | 7.5 | 0.14 |
| polarC18 | 276.144 | 56.3536 | 7.5 | 0.14 |
| polarC18 | 276.144 | 55.8831 | 7.5 | 0.14 |
| polarC18 | 276.144 | 191.1384 | 7.5 | 0.14 |
| polarC18 | 276.144 | 119.7095 | 7.5 | 0.14 |
| polarC18 | 276.144 | 73.6888 | 7.5 | 0.14 |
| polarC18 | 276.144 | 142.8944 | 7.5 | 0.14 |
| polarC18 | 276.144 | 125.3083 | 7.5 | 0.14 |
| polarC18 | 276.144 | 63.1544 | 7.5 | 0.14 |
| polarC18 | 276.144 | 107.0375 | 7.5 | 0.14 |
| polarC18 | 276.144 | 166.0518 | 7.5 | 0.14 |
| polarC18 | 276.144 | 300.8084 | 7.5 | 0.14 |
| polarC18 | 276.144 | 186.2765 | 7.5 | 0.14 |
| polarC18 | 276.144 | 51.2379 | 7.5 | 0.14 |
| polarC18 | 276.144 | 69.8465 | 7.5 | 0.14 |
| polarC18 | 276.144 | 73.028 | 7.5 | 0.14 |
| polarC18 | 276.144 | 107.1583 | 7.5 | 0.14 |
| polarC18 | 276.144 | 222.004 | 7.5 | 0.14 |
| polarC18 | 276.144 | 57.5513 | 7.5 | 0.14 |
| polarC18 | 276.144 | 168.5536 | 7.5 | 0.14 |
| polarC18 | 276.144 | 82.4341 | 7.5 | 0.14 |
| polarC18 | 276.144 | 80.9798 | 7.5 | 0.14 |
| polarC18 | 276.144 | 197.3003 | 7.5 | 0.14 |
| polarC18 | 276.144 | 57.0834 | 7.5 | 0.14 |
| polarC18 | 276.144 | 85.3499 | 7.5 | 0.14 |
| polarC18 | 276.144 | 55.1991 | 7.5 | 0.14 |
| polarC18 | 276.144 | 79.7957 | 7.5 | 0.14 |
| polarC18 | 276.144 | 105.5371 | 7.5 | 0.14 |
| polarC18 | 276.144 | 60.1574 | 7.5 | 0.14 |
| polarC18 | 276.144 | 64.0775 | 7.5 | 0.14 |
| polarC18 | 276.144 | 51.7041 | 7.5 | 0.14 |
| polarC18 | 276.144 | 68.0609 | 7.5 | 0.14 |

| | | | | |
|---|---|---|---|---|
| polarC18 | 276.144 | 68.1017 | 7.5 | 0.14 |
| polarC18 | 276.144 | 190.2699 | 7.5 | 0.14 |
| polarC18 | 276.144 | 65.9517 | 7.5 | 0.14 |
| polarC18 | 276.144 | 74.2324 | 7.5 | 0.14 |
| polarC18 | 276.144 | 80.2504 | 7.5 | 0.14 |
| polarC18 | 276.144 | 113.627 | 7.5 | 0.14 |
| polarC18 | 276.144 | 200.5085 | 7.5 | 0.14 |
| polarC18 | 276.144 | 53.9037 | 7.5 | 0.14 |
| polarC18 | 276.144 | 153.2793 | 7.5 | 0.14 |
| polarC18 | 276.144 | 52.0993 | 7.5 | 0.14 |
| polarC18 | 276.144 | 141.492 | 7.5 | 0.14 |
| polarC18 | 276.144 | 59.936 | 7.5 | 0.14 |
| polarC18 | 276.144 | 55.6883 | 7.5 | 0.14 |
| polarC18 | 276.144 | 68.3837 | 7.5 | 0.14 |
| polarC18 | 276.144 | 99.7525 | 7.5 | 0.14 |
| polarC18 | 276.144 | 169.1564 | 7.5 | 0.14 |
| polarC18 | 276.144 | 167.7344 | 7.5 | 0.14 |
| polarC18 | 276.144 | 139.7411 | 7.5 | 0.14 |
| polarC18 | 276.144 | 52.0858 | 7.5 | 0.14 |
| polarC18 | 276.144 | 56.1327 | 7.5 | 0.14 |
| polarC18 | 276.144 | 55.4579 | 7.5 | 0.14 |
| polarC18 | 276.144 | 156.5278 | 7.5 | 0.14 |
| polarC18 | 276.144 | 129.7364 | 7.5 | 0.14 |
| polarC18 | 276.144 | 76.8507 | 7.5 | 0.14 |
| polarC18 | 276.144 | 57.5042 | 7.5 | 0.14 |
| polarC18 | 276.144 | 139.6136 | 7.5 | 0.14 |
| polarC18 | 276.144 | 55.5193 | 7.5 | 0.14 |
| polarC18 | 276.144 | 252.4632 | 7.5 | 0.14 |
| polarC18 | 276.144 | 88.0383 | 7.5 | 0.14 |
| polarC18 | 276.144 | 165.4091 | 7.5 | 0.14 |
| polarC18 | 276.144 | 175.4877 | 7.5 | 0.14 |
| polarC18 | 276.144 | 69.6012 | 7.5 | 0.14 |
| polarC18 | 276.144 | 207.5152 | 7.5 | 0.14 |
| polarC18 | 276.144 | 191.7359 | 7.5 | 0.14 |
| polarC18 | 276.144 | 139.2009 | 7.5 | 0.14 |
| polarC18 | 276.144 | 110.8396 | 7.5 | 0.14 |
| polarC18 | 276.144 | 233.5633 | 7.5 | 0.14 |
| polarC18 | 276.144 | 253.3354 | 7.5 | 0.14 |
| polarC18 | 276.144 | 55.5707 | 7.5 | 0.14 |
| polarC18 | 276.144 | 84.2364 | 7.5 | 0.14 |
| polarC18 | 276.144 | 57.1029 | 7.5 | 0.14 |
| polarC18 | 276.144 | 89.4165 | 7.5 | 0.14 |

| | | | | |
|---|---|---|---|---|
| polarC18 | 276.144 | 51.0541 | 7.5 | 0.14 |
| polarC18 | 276.144 | 71.0465 | 7.5 | 0.14 |
| polarC18 | 276.144 | 60.2284 | 7.5 | 0.14 |
| polarC18 | 276.144 | 168.9097 | 7.5 | 0.14 |
| polarC18 | 276.144 | 282.4433 | 7.5 | 0.14 |
| polarC18 | 276.144 | 101.2629 | 7.5 | 0.14 |
| polarC18 | 276.144 | 232.3665 | 7.5 | 0.14 |
| polarC18 | 276.144 | 67.2539 | 7.5 | 0.14 |
| polarC18 | 276.144 | 51.2758 | 7.5 | 0.14 |
| polarC18 | 276.144 | 70.7014 | 7.5 | 0.14 |
| polarC18 | 276.144 | 104.3729 | 7.5 | 0.14 |
| polarC18 | 276.144 | 187.5697 | 7.5 | 0.14 |
| polarC18 | 276.144 | 108.8385 | 7.5 | 0.14 |
| polarC18 | 276.144 | 117.6529 | 7.5 | 0.14 |
| polarC18 | 276.144 | 88.38 | 7.5 | 0.14 |
| polarC18 | 276.144 | 112.0831 | 7.5 | 0.14 |
| polarC18 | 276.144 | 85.4729 | 7.5 | 0.14 |
| polarC18 | 276.144 | 113.7435 | 7.5 | 0.14 |
| polarC18 | 276.144 | 200.1915 | 7.5 | 0.14 |
| polarC18 | 276.144 | 73.1453 | 7.5 | 0.14 |
| polarC18 | 276.144 | 83.2889 | 7.4 | 0.14 |
| polarC18 | 276.144 | 206.6442 | 7.4 | 0.14 |
| polarC18 | 276.144 | 61.5004 | 7.4 | 0.14 |
| polarC18 | 276.144 | 233.2248 | 7.4 | 0.14 |
| polarC18 | 276.144 | 66.3435 | 7.4 | 0.14 |
| polarC18 | 276.144 | 165.016 | 7.4 | 0.14 |
| polarC18 | 276.144 | 81.4567 | 7.4 | 0.14 |
| polarC18 | 276.144 | 82.559 | 7.4 | 0.14 |
| polarC18 | 276.144 | 65.7768 | 7.4 | 0.14 |
| polarC18 | 276.144 | 145.1588 | 7.4 | 0.14 |
| polarC18 | 276.144 | 121.1851 | 7.4 | 0.14 |
| polarC18 | 276.144 | 195.5144 | 7.4 | 0.14 |
| polarC18 | 276.144 | 94.0322 | 7.4 | 0.14 |
| polarC18 | 276.144 | 60.883 | 7.4 | 0.14 |
| polarC18 | 276.144 | 55.1689 | 7.4 | 0.14 |
| polarC18 | 276.144 | 63.19 | 7.4 | 0.14 |
| polarC18 | 276.144 | 231.0738 | 7.4 | 0.14 |
| polarC18 | 276.144 | 50.4574 | 7.4 | 0.14 |
| polarC18 | 276.144 | 81.0039 | 7.4 | 0.14 |
| polarC18 | 276.144 | 143.2103 | 7.4 | 0.14 |
| polarC18 | 276.144 | 139.0137 | 7.4 | 0.14 |
| polarC18 | 276.144 | 76.4359 | 7.4 | 0.14 |

| | | | | |
|---|---|---|---|---|
| polarC18 | 276.144 | 59.7277 | 7.4 | 0.14 |
| polarC18 | 276.144 | 53.294 | 7.4 | 0.14 |
| polarC18 | 276.144 | 170.5554 | 7.4 | 0.14 |
| polarC18 | 276.144 | 51.3395 | 7.4 | 0.14 |
| polarC18 | 276.144 | 95.422 | 7.4 | 0.14 |
| polarC18 | 276.144 | 80.2959 | 7.4 | 0.14 |
| polarC18 | 276.144 | 109.2811 | 7.4 | 0.14 |
| polarC18 | 276.144 | 206.3418 | 7.4 | 0.14 |
| polarC18 | 276.144 | 61.5604 | 7.4 | 0.14 |
| polarC18 | 276.144 | 181.3785 | 7.4 | 0.14 |
| polarC18 | 276.144 | 124.3742 | 7.4 | 0.14 |
| polarC18 | 276.144 | 60.2408 | 7.4 | 0.14 |
| polarC18 | 276.144 | 81.0555 | 7.4 | 0.14 |
| polarC18 | 276.144 | 103.972 | 7.4 | 0.14 |
| polarC18 | 276.144 | 52.6075 | 7.4 | 0.14 |
| polarC18 | 276.144 | 56.9885 | 7.4 | 0.14 |
| polarC18 | 276.144 | 244.6498 | 7.4 | 0.14 |
| polarC18 | 276.144 | 54.2216 | 7.4 | 0.14 |
| polarC18 | 276.144 | 115.404 | 7.4 | 0.14 |
| polarC18 | 276.144 | 177.7392 | 7.4 | 0.14 |
| polarC18 | 276.144 | 85.3005 | 7.4 | 0.14 |
| polarC18 | 276.144 | 211.3361 | 7.4 | 0.14 |
| polarC18 | 276.144 | 72.0021 | 7.4 | 0.14 |
| polarC18 | 276.144 | 110.3594 | 7.4 | 0.14 |
| polarC18 | 276.144 | 78.783 | 7.4 | 0.14 |
| polarC18 | 276.144 | 294.3932 | 7.4 | 0.14 |
| polarC18 | 276.144 | 283.1327 | 7.4 | 0.14 |
| polarC18 | 276.144 | 79.0083 | 7.4 | 0.14 |
| polarC18 | 276.144 | 56.6479 | 7.4 | 0.14 |
| polarC18 | 276.144 | 107.706 | 7.4 | 0.14 |
| polarC18 | 276.144 | 71.4881 | 7.4 | 0.14 |
| polarC18 | 276.144 | 218.5526 | 7.4 | 0.14 |
| polarC18 | 276.144 | 93.919 | 7.4 | 0.14 |
| polarC18 | 276.144 | 55.6375 | 7.4 | 0.14 |
| polarC18 | 276.144 | 252.0472 | 7.4 | 0.14 |
| polarC18 | 276.144 | 182.2336 | 7.4 | 0.14 |
| polarC18 | 276.144 | 133.5232 | 7.4 | 0.14 |
| polarC18 | 276.144 | 115.7215 | 7.4 | 0.14 |
| polarC18 | 276.144 | 82.6633 | 7.4 | 0.14 |
| polarC18 | 276.144 | 78.672 | 7.4 | 0.14 |
| polarC18 | 276.144 | 277.0935 | 7.4 | 0.14 |
| polarC18 | 276.144 | 54.6328 | 7.4 | 0.14 |

| | | | | |
|---|---|---|---|---|
| polarC18 | 276.144 | 88.0754 | 7.4 | 0.14 |
| polarC18 | 276.144 | 60.4359 | 7.4 | 0.14 |
| polarC18 | 276.144 | 111.4337 | 7.4 | 0.14 |
| polarC18 | 276.144 | 61.8356 | 7.4 | 0.14 |
| polarC18 | 276.144 | 74.6235 | 7.4 | 0.14 |
| polarC18 | 276.144 | 141.8175 | 7.4 | 0.14 |
| polarC18 | 276.144 | 257.2191 | 7.4 | 0.14 |
| polarC18 | 276.144 | 93.3417 | 7.4 | 0.14 |
| polarC18 | 276.144 | 108.0722 | 7.4 | 0.14 |
| polarC18 | 276.144 | 240.4523 | 7.4 | 0.14 |
| polarC18 | 276.144 | 260.5075 | 7.4 | 0.14 |
| polarC18 | 276.144 | 73.3519 | 7.4 | 0.14 |
| polarC18 | 276.144 | 114.6152 | 7.4 | 0.14 |
| polarC18 | 276.144 | 246.6469 | 7.4 | 0.14 |
| polarC18 | 276.144 | 84.181 | 7.4 | 0.14 |
| polarC18 | 276.144 | 194.222 | 7.4 | 0.14 |
| polarC18 | 276.144 | 112.743 | 7.4 | 0.14 |
| polarC18 | 276.144 | 109.6359 | 7.4 | 0.14 |
| polarC18 | 276.144 | 285.3098 | 7.4 | 0.14 |
| polarC18 | 276.144 | 177.5394 | 7.4 | 0.14 |
| polarC18 | 276.144 | 113.581 | 7.4 | 0.14 |
| polarC18 | 276.144 | 63.4832 | 7.4 | 0.14 |
| polarC18 | 276.144 | 81.4096 | 7.4 | 0.14 |
| polarC18 | 276.144 | 93.8685 | 7.4 | 0.14 |
| polarC18 | 276.144 | 50.7724 | 7.4 | 0.14 |
| polarC18 | 276.144 | 68.3675 | 7.4 | 0.14 |
| polarC18 | 276.144 | 182.2762 | 7.4 | 0.14 |
| polarC18 | 276.144 | 101.2997 | 7.4 | 0.14 |
| polarC18 | 276.144 | 190.7083 | 7.4 | 0.14 |
| polarC18 | 276.144 | 110.5858 | 7.4 | 0.14 |
| polarC18 | 276.144 | 295.7739 | 7.4 | 0.14 |
| polarC18 | 276.144 | 76.1611 | 7.4 | 0.14 |
| polarC18 | 276.144 | 146.0919 | 7.4 | 0.14 |
| polarC18 | 276.144 | 50.3949 | 7.4 | 0.14 |
| polarC18 | 276.144 | 52.059 | 7.4 | 0.14 |
| polarC18 | 276.144 | 156.0703 | 7.4 | 0.14 |
| polarC18 | 276.144 | 109.0127 | 7.4 | 0.14 |
| polarC18 | 276.144 | 56.5025 | 7.4 | 0.14 |
| polarC18 | 276.144 | 74.8158 | 7.4 | 0.14 |
| polarC18 | 276.144 | 69.9998 | 7.4 | 0.14 |
| polarC18 | 276.144 | 299.4792 | 7.4 | 0.14 |
| polarC18 | 276.144 | 97.6046 | 7.4 | 0.14 |

| | | | | |
|---|---|---|---|---|
| polarC18 | 276.144 | 88.8105 | 7.4 | 0.14 |
| polarC18 | 276.144 | 153.3384 | 7.4 | 0.14 |
| polarC18 | 276.144 | 214.9245 | 7.4 | 0.14 |
| polarC18 | 276.144 | 80.5134 | 7.4 | 0.14 |
| polarC18 | 276.144 | 54.4655 | 7.4 | 0.14 |
| polarC18 | 276.144 | 71.417 | 7.4 | 0.14 |
| polarC18 | 276.144 | 208.1676 | 7.4 | 0.14 |
| polarC18 | 276.144 | 143.6049 | 7.4 | 0.14 |
| polarC18 | 276.144 | 78.464 | 7.4 | 0.14 |
| polarC18 | 276.144 | 94.5349 | 7.4 | 0.14 |
| polarC18 | 276.144 | 53.9953 | 7.4 | 0.14 |
| polarC18 | 276.144 | 124.6579 | 7.4 | 0.14 |
| polarC18 | 276.144 | 63.6431 | 7.4 | 0.14 |
| polarC18 | 276.144 | 69.0909 | 7.4 | 0.14 |
| polarC18 | 276.144 | 67.0752 | 7.4 | 0.14 |
| polarC18 | 276.144 | 111.8606 | 7.4 | 0.14 |
| polarC18 | 276.144 | 97.4871 | 7.4 | 0.14 |
| polarC18 | 276.144 | 154.6707 | 7.4 | 0.14 |
| polarC18 | 276.144 | 90.5294 | 7.4 | 0.14 |
| polarC18 | 276.144 | 92.7626 | 7.4 | 0.14 |
| polarC18 | 276.144 | 95.6126 | 7.4 | 0.14 |
| polarC18 | 276.144 | 284.2457 | 7.4 | 0.14 |
| polarC18 | 276.144 | 98.0168 | 7.4 | 0.14 |
| polarC18 | 276.144 | 177.488 | 7.4 | 0.14 |
| polarC18 | 276.144 | 177.9302 | 7.4 | 0.14 |
| polarC18 | 276.144 | 67.1551 | 7.4 | 0.14 |
| polarC18 | 276.144 | 57.9437 | 7.4 | 0.14 |
| polarC18 | 276.144 | 55.8883 | 7.4 | 0.14 |
| polarC18 | 276.144 | 161.0184 | 7.4 | 0.14 |
| polarC18 | 276.144 | 54.915 | 7.4 | 0.14 |
| polarC18 | 276.144 | 149.4481 | 7.4 | 0.14 |
| polarC18 | 276.144 | 166.4277 | 7.4 | 0.14 |
| polarC18 | 276.144 | 73.9777 | 7.4 | 0.14 |
| polarC18 | 276.144 | 62.1076 | 7.4 | 0.14 |
| polarC18 | 276.144 | 54.5966 | 7.4 | 0.14 |
| polarC18 | 276.144 | 163.6511 | 7.4 | 0.14 |
| polarC18 | 276.144 | 149.1039 | 7.4 | 0.14 |
| polarC18 | 276.144 | 121.362 | 7.4 | 0.14 |
| polarC18 | 276.144 | 171.8574 | 7.4 | 0.14 |
| polarC18 | 276.144 | 165.8458 | 7.4 | 0.14 |
| polarC18 | 276.144 | 271.384 | 7.4 | 0.14 |
| polarC18 | 276.144 | 51.021 | 7.4 | 0.14 |

| | | | | |
|---|---|---|---|---|
| polarC18 | 276.144 | 189.9982 | 7.4 | 0.14 |
| polarC18 | 276.144 | 84.9935 | 7.4 | 0.14 |
| polarC18 | 276.144 | 133.3447 | 7.4 | 0.14 |
| polarC18 | 276.144 | 59.6617 | 7.4 | 0.14 |
| polarC18 | 276.144 | 227.6996 | 7.4 | 0.14 |
| polarC18 | 276.144 | 81.0905 | 7.4 | 0.14 |
| polarC18 | 276.144 | 62.2277 | 7.4 | 0.14 |
| polarC18 | 276.144 | 79.7152 | 7.4 | 0.14 |
| polarC18 | 276.144 | 54.6193 | 7.4 | 0.14 |
| polarC18 | 276.144 | 53.8181 | 7.4 | 0.14 |
| polarC18 | 276.144 | 144.3182 | 7.4 | 0.14 |
| polarC18 | 276.144 | 103.1941 | 7.4 | 0.14 |
| polarC18 | 276.144 | 96.8724 | 7.4 | 0.14 |
| polarC18 | 276.144 | 75.5655 | 7.4 | 0.14 |
| polarC18 | 276.144 | 90.2799 | 7.4 | 0.14 |
| polarC18 | 276.144 | 199.129 | 7.3 | 0.14 |
| polarC18 | 276.144 | 114.9587 | 7.3 | 0.14 |
| polarC18 | 276.144 | 99.4942 | 7.3 | 0.14 |
| polarC18 | 276.144 | 123.5135 | 7.3 | 0.14 |
| polarC18 | 276.144 | 283.4178 | 7.3 | 0.14 |
| polarC18 | 276.144 | 107.3841 | 7.3 | 0.14 |
| polarC18 | 276.144 | 100.8698 | 7.3 | 0.14 |
| polarC18 | 276.144 | 95.0218 | 7.3 | 0.14 |
| polarC18 | 276.144 | 89.8436 | 7.3 | 0.14 |
| polarC18 | 276.144 | 103.0902 | 7.3 | 0.14 |
| polarC18 | 276.144 | 181.6251 | 7.3 | 0.14 |
| polarC18 | 276.144 | 85.834 | 7.3 | 0.14 |
| polarC18 | 276.144 | 53.2814 | 7.3 | 0.14 |
| polarC18 | 276.144 | 103.8517 | 7.3 | 0.14 |
| polarC18 | 276.144 | 97.0887 | 7.3 | 0.14 |
| polarC18 | 276.144 | 50.7513 | 7.3 | 0.14 |
| polarC18 | 276.144 | 67.1255 | 7.3 | 0.14 |
| polarC18 | 276.144 | 118.691 | 7.3 | 0.14 |
| polarC18 | 276.144 | 62.9727 | 7.3 | 0.14 |
| polarC18 | 276.144 | 202.4882 | 7.3 | 0.14 |
| polarC18 | 276.144 | 52.9466 | 7.3 | 0.14 |
| polarC18 | 276.144 | 93.4063 | 7.3 | 0.14 |
| polarC18 | 276.144 | 52.9976 | 7.3 | 0.14 |
| polarC18 | 276.144 | 150.2814 | 7.3 | 0.14 |
| polarC18 | 276.144 | 182.8613 | 7.3 | 0.14 |
| polarC18 | 276.144 | 151.4801 | 7.3 | 0.14 |
| polarC18 | 276.144 | 64.7924 | 7.3 | 0.14 |

| | | | | |
|---|---|---|---|---|
| polarC18 | 276.144 | 57.4957 | 7.3 | 0.14 |
| polarC18 | 276.144 | 52.7584 | 7.3 | 0.14 |
| polarC18 | 276.144 | 77.0419 | 7.3 | 0.14 |
| polarC18 | 276.144 | 50.5709 | 7.3 | 0.14 |
| polarC18 | 276.144 | 70.5827 | 7.3 | 0.14 |
| polarC18 | 276.144 | 50.9978 | 7.3 | 0.14 |
| polarC18 | 276.144 | 53.7964 | 7.3 | 0.14 |
| polarC18 | 276.144 | 195.274 | 7.3 | 0.14 |
| polarC18 | 276.144 | 68.3299 | 7.3 | 0.14 |
| polarC18 | 276.144 | 281.291 | 7.3 | 0.14 |
| polarC18 | 276.144 | 182.5317 | 7.3 | 0.14 |
| polarC18 | 276.144 | 70.4907 | 7.3 | 0.14 |
| polarC18 | 276.144 | 118.4325 | 7.3 | 0.14 |
| polarC18 | 276.144 | 69.6906 | 7.3 | 0.14 |
| polarC18 | 276.144 | 145.0716 | 7.3 | 0.14 |
| polarC18 | 276.144 | 56.5745 | 7.3 | 0.14 |
| polarC18 | 276.144 | 51.2006 | 7.3 | 0.14 |
| polarC18 | 276.144 | 90.4205 | 7.3 | 0.14 |
| polarC18 | 276.144 | 94.9777 | 7.3 | 0.14 |
| polarC18 | 276.144 | 188.2083 | 7.3 | 0.14 |
| polarC18 | 276.144 | 126.181 | 7.3 | 0.14 |
| polarC18 | 276.144 | 104.1065 | 7.3 | 0.14 |
| polarC18 | 276.144 | 281.0003 | 7.3 | 0.14 |
| polarC18 | 276.144 | 209.5669 | 7.3 | 0.14 |
| polarC18 | 276.144 | 151.932 | 7.3 | 0.14 |
| polarC18 | 276.144 | 55.5098 | 7.3 | 0.14 |
| polarC18 | 276.144 | 84.6762 | 7.3 | 0.14 |
| polarC18 | 276.144 | 78.9711 | 7.3 | 0.14 |
| polarC18 | 276.144 | 233.3002 | 7.3 | 0.14 |
| polarC18 | 276.144 | 61.6795 | 7.3 | 0.14 |
| polarC18 | 276.144 | 161.624 | 7.3 | 0.14 |
| polarC18 | 276.144 | 61.8761 | 7.3 | 0.14 |
| polarC18 | 276.144 | 104.2783 | 7.3 | 0.14 |
| polarC18 | 276.144 | 103.3286 | 7.3 | 0.14 |
| polarC18 | 276.144 | 119.7315 | 7.3 | 0.14 |
| polarC18 | 276.144 | 64.4079 | 7.3 | 0.14 |
| polarC18 | 276.144 | 60.4477 | 7.3 | 0.14 |
| polarC18 | 276.144 | 50.7365 | 7.3 | 0.14 |
| polarC18 | 276.144 | 227.9246 | 7.3 | 0.14 |
| polarC18 | 276.144 | 109.707 | 7.3 | 0.14 |
| polarC18 | 276.144 | 123.5861 | 7.3 | 0.14 |
| polarC18 | 276.144 | 60.829 | 7.3 | 0.14 |

| | | | | |
|---|---|---|---|---|
| polarC18 | 276.144 | 193.5626 | 7.3 | 0.14 |
| polarC18 | 276.144 | 61.9689 | 7.3 | 0.14 |
| polarC18 | 276.144 | 197.125 | 7.3 | 0.14 |
| polarC18 | 276.144 | 51.2576 | 7.3 | 0.14 |
| polarC18 | 276.144 | 58.336 | 7.3 | 0.14 |
| polarC18 | 276.144 | 259.1628 | 7.3 | 0.14 |
| polarC18 | 276.144 | 87.2765 | 7.3 | 0.14 |
| polarC18 | 276.144 | 51.7865 | 7.3 | 0.14 |
| polarC18 | 276.144 | 50.217 | 7.3 | 0.14 |
| polarC18 | 276.144 | 51.8536 | 7.3 | 0.14 |
| polarC18 | 276.144 | 60.3356 | 7.3 | 0.14 |
| polarC18 | 276.144 | 73.0483 | 7.3 | 0.14 |
| polarC18 | 276.144 | 102.125 | 7.3 | 0.14 |
| polarC18 | 276.144 | 50.4134 | 7.3 | 0.14 |
| polarC18 | 276.144 | 160.9932 | 7.3 | 0.14 |
| polarC18 | 276.144 | 189.8087 | 7.3 | 0.14 |
| polarC18 | 276.144 | 84.693 | 7.3 | 0.14 |
| polarC18 | 276.144 | 263.1476 | 7.3 | 0.14 |
| polarC18 | 276.144 | 87.2847 | 7.3 | 0.14 |
| polarC18 | 276.144 | 291.2471 | 7.3 | 0.14 |
| polarC18 | 276.144 | 64.8306 | 7.3 | 0.14 |
| polarC18 | 276.144 | 53.6094 | 7.3 | 0.14 |
| polarC18 | 276.144 | 102.2166 | 7.3 | 0.14 |
| polarC18 | 276.144 | 51.745 | 7.3 | 0.14 |
| polarC18 | 276.144 | 165.5828 | 7.3 | 0.14 |
| polarC18 | 276.144 | 105.046 | 7.3 | 0.14 |
| polarC18 | 276.144 | 80.7318 | 7.3 | 0.14 |
| polarC18 | 276.144 | 152.207 | 7.3 | 0.14 |
| polarC18 | 276.144 | 155.9305 | 7.3 | 0.14 |
| polarC18 | 276.144 | 109.8736 | 7.3 | 0.14 |
| polarC18 | 276.144 | 142.3452 | 7.3 | 0.14 |
| polarC18 | 276.144 | 74.5698 | 7.3 | 0.14 |
| polarC18 | 276.144 | 77.4238 | 7.3 | 0.14 |
| polarC18 | 276.144 | 147.2311 | 7.3 | 0.14 |
| polarC18 | 276.144 | 61.7189 | 7.3 | 0.14 |
| polarC18 | 276.144 | 67.2231 | 7.3 | 0.14 |
| polarC18 | 276.144 | 51.4916 | 7.3 | 0.14 |
| polarC18 | 276.144 | 145.6527 | 7.3 | 0.14 |
| polarC18 | 276.144 | 166.9123 | 7.3 | 0.14 |
| polarC18 | 276.144 | 57.5679 | 7.3 | 0.14 |
| polarC18 | 276.144 | 74.0145 | 7.3 | 0.14 |
| polarC18 | 276.144 | 171.1823 | 7.3 | 0.14 |

| | | | | |
|---|---|---|---|---|
| polarC18 | 276.144 | 124.3214 | 7.3 | 0.14 |
| polarC18 | 276.144 | 67.1224 | 7.3 | 0.14 |
| polarC18 | 276.144 | 109.4837 | 7.3 | 0.14 |
| polarC18 | 276.144 | 84.7629 | 7.3 | 0.14 |
| polarC18 | 276.144 | 111.1626 | 7.3 | 0.14 |
| polarC18 | 276.144 | 64.7583 | 7.3 | 0.14 |
| polarC18 | 276.144 | 119.1549 | 7.3 | 0.14 |
| polarC18 | 276.144 | 67.2256 | 7.3 | 0.14 |
| polarC18 | 276.144 | 57.4901 | 7.3 | 0.14 |
| polarC18 | 276.144 | 60.5737 | 7.3 | 0.14 |
| polarC18 | 276.144 | 64.1875 | 7.3 | 0.14 |
| polarC18 | 276.144 | 148.9856 | 7.3 | 0.14 |
| polarC18 | 276.144 | 67.2026 | 7.3 | 0.14 |
| polarC18 | 276.144 | 86.3848 | 7.3 | 0.14 |
| polarC18 | 276.144 | 292.3623 | 7.3 | 0.14 |
| polarC18 | 276.144 | 50.0748 | 7.3 | 0.14 |
| polarC18 | 276.144 | 143.9234 | 7.3 | 0.14 |
| polarC18 | 276.144 | 163.0163 | 7.3 | 0.14 |
| polarC18 | 276.144 | 55.147 | 7.3 | 0.14 |
| polarC18 | 276.144 | 73.5858 | 7.3 | 0.14 |
| polarC18 | 276.144 | 63.5387 | 7.3 | 0.14 |
| polarC18 | 276.144 | 60.3977 | 7.3 | 0.14 |
| polarC18 | 276.144 | 248.546 | 7.3 | 0.14 |
| polarC18 | 276.144 | 107.7563 | 7.3 | 0.14 |
| polarC18 | 276.144 | 216.5198 | 7.3 | 0.14 |
| polarC18 | 276.144 | 72.54 | 7.3 | 0.14 |
| polarC18 | 276.144 | 175.0052 | 7.3 | 0.14 |
| polarC18 | 276.144 | 102.8918 | 7.3 | 0.14 |
| polarC18 | 276.144 | 79.1188 | 7.3 | 0.14 |
| polarC18 | 276.144 | 124.966 | 7.3 | 0.14 |
| polarC18 | 276.144 | 130.768 | 7.3 | 0.14 |
| polarC18 | 276.144 | 54.4948 | 7.3 | 0.14 |
| polarC18 | 276.144 | 165.9448 | 7.3 | 0.14 |
| polarC18 | 276.144 | 253.2716 | 7.3 | 0.14 |
| polarC18 | 276.144 | 82.3544 | 7.3 | 0.14 |
| polarC18 | 276.144 | 160.5872 | 7.3 | 0.14 |
| polarC18 | 276.144 | 103.9404 | 7.3 | 0.14 |
| polarC18 | 276.144 | 142.7915 | 7.3 | 0.14 |
| polarC18 | 276.144 | 116.818 | 7.3 | 0.14 |
| polarC18 | 276.144 | 121.6343 | 7.3 | 0.14 |
| polarC18 | 276.144 | 93.6677 | 7.3 | 0.14 |
| polarC18 | 276.144 | 133.2824 | 7.3 | 0.14 |

| | | | | |
|---|---|---|---|---|
| polarC18 | 276.144 | 163.9403 | 7.3 | 0.14 |
| polarC18 | 276.144 | 114.1798 | 7.3 | 0.14 |
| polarC18 | 276.144 | 57.6279 | 7.3 | 0.14 |
| polarC18 | 276.144 | 61.6854 | 7.3 | 0.14 |
| polarC18 | 276.144 | 69.0482 | 7.3 | 0.14 |
| polarC18 | 276.144 | 52.3101 | 7.3 | 0.14 |
| polarC18 | 276.144 | 137.229 | 7.3 | 0.14 |
| polarC18 | 276.144 | 83.6883 | 7.3 | 0.14 |
| polarC18 | 276.144 | 50.9212 | 7.3 | 0.14 |
| polarC18 | 276.144 | 76.2216 | 7.3 | 0.14 |
| polarC18 | 276.144 | 116.9444 | 7.3 | 0.14 |
| polarC18 | 276.144 | 71.4305 | 7.3 | 0.14 |
| polarC18 | 276.144 | 95.1493 | 7.3 | 0.14 |
| polarC18 | 276.144 | 109.2892 | 7.3 | 0.14 |
| polarC18 | 276.144 | 170.0514 | 7.3 | 0.14 |
| polarC18 | 276.144 | 103.4917 | 7.3 | 0.14 |
| polarC18 | 276.144 | 116.3863 | 7.3 | 0.14 |
| polarC18 | 276.144 | 183.4571 | 7.3 | 0.14 |
| polarC18 | 276.144 | 105.519 | 7.3 | 0.14 |
| polarC18 | 276.144 | 129.2584 | 7.3 | 0.14 |
| polarC18 | 276.144 | 58.5382 | 7.3 | 0.14 |
| polarC18 | 276.144 | 110.1571 | 7.3 | 0.14 |
| polarC18 | 276.144 | 283.8663 | 7.3 | 0.14 |
| polarC18 | 276.144 | 82.0511 | 7.3 | 0.14 |
| polarC18 | 276.144 | 144.5572 | 7.3 | 0.14 |
| polarC18 | 276.144 | 62.841 | 7.3 | 0.14 |
| polarC18 | 276.144 | 243.867 | 7.3 | 0.14 |
| polarC18 | 276.144 | 144.7673 | 7.2 | 0.14 |
| polarC18 | 276.144 | 192.177 | 7.2 | 0.14 |
| polarC18 | 276.144 | 70.1443 | 7.2 | 0.14 |
| polarC18 | 276.144 | 163.3273 | 7.2 | 0.14 |
| polarC18 | 276.144 | 60.1302 | 7.2 | 0.14 |
| polarC18 | 276.144 | 296.3037 | 7.2 | 0.14 |
| polarC18 | 276.144 | 74.8273 | 7.2 | 0.14 |
| polarC18 | 276.144 | 233.5113 | 7.2 | 0.14 |
| polarC18 | 276.144 | 194.7003 | 7.2 | 0.14 |
| polarC18 | 276.144 | 118.6987 | 7.2 | 0.14 |
| polarC18 | 276.144 | 68.267 | 7.2 | 0.14 |
| polarC18 | 276.144 | 165.2241 | 7.2 | 0.14 |
| polarC18 | 276.144 | 56.2083 | 7.2 | 0.14 |
| polarC18 | 276.144 | 149.1743 | 7.2 | 0.14 |
| polarC18 | 276.144 | 178.9464 | 7.2 | 0.14 |

| | | | | |
|---|---|---|---|---|
| polarC18 | 276.144 | 53.6792 | 7.2 | 0.14 |
| polarC18 | 276.144 | 112.0155 | 7.2 | 0.14 |
| polarC18 | 276.144 | 64.4773 | 7.2 | 0.14 |
| polarC18 | 276.144 | 201.7414 | 7.2 | 0.14 |
| polarC18 | 276.144 | 56.1367 | 7.2 | 0.14 |
| polarC18 | 276.144 | 76.9801 | 7.2 | 0.14 |
| polarC18 | 276.144 | 60.1807 | 7.2 | 0.14 |
| polarC18 | 276.144 | 50.8106 | 7.2 | 0.14 |
| polarC18 | 276.144 | 50.0907 | 7.2 | 0.14 |
| polarC18 | 276.144 | 159.2364 | 7.2 | 0.14 |
| polarC18 | 276.144 | 262.082 | 7.2 | 0.14 |
| polarC18 | 276.144 | 190.431 | 7.2 | 0.14 |
| polarC18 | 276.144 | 60.9049 | 7.2 | 0.14 |
| polarC18 | 276.144 | 91.982 | 7.2 | 0.14 |
| polarC18 | 276.144 | 59.5692 | 7.2 | 0.14 |
| polarC18 | 276.144 | 142.1268 | 7.2 | 0.14 |
| polarC18 | 276.144 | 68.4534 | 7.2 | 0.14 |
| polarC18 | 276.144 | 101.6747 | 7.2 | 0.14 |
| polarC18 | 276.144 | 57.9151 | 7.2 | 0.14 |
| polarC18 | 276.144 | 199.2224 | 7.2 | 0.14 |
| polarC18 | 276.144 | 61.989 | 7.2 | 0.14 |
| polarC18 | 276.144 | 66.7744 | 7.2 | 0.14 |
| polarC18 | 276.144 | 98.1605 | 7.2 | 0.14 |
| polarC18 | 276.144 | 181.9508 | 7.2 | 0.14 |
| polarC18 | 276.144 | 98.8007 | 7.2 | 0.14 |
| polarC18 | 276.144 | 58.1251 | 7.2 | 0.14 |
| polarC18 | 276.144 | 106.38 | 7.2 | 0.14 |
| polarC18 | 276.144 | 83.3176 | 7.2 | 0.14 |
| polarC18 | 276.144 | 221.6551 | 7.2 | 0.14 |
| polarC18 | 276.144 | 88.4071 | 7.2 | 0.14 |
| polarC18 | 276.144 | 145.4143 | 7.2 | 0.14 |
| polarC18 | 276.144 | 54.6164 | 7.2 | 0.14 |
| polarC18 | 276.144 | 226.7268 | 7.2 | 0.14 |
| polarC18 | 276.144 | 67.6851 | 7.2 | 0.14 |
| polarC18 | 276.144 | 176.7728 | 7.2 | 0.14 |
| polarC18 | 276.144 | 55.5441 | 7.2 | 0.14 |
| polarC18 | 276.144 | 137.0506 | 7.2 | 0.14 |
| polarC18 | 276.144 | 175.75 | 7.2 | 0.14 |
| polarC18 | 276.144 | 52.8042 | 7.2 | 0.14 |
| polarC18 | 276.144 | 230.7519 | 7.2 | 0.14 |
| polarC18 | 276.144 | 161.6517 | 7.2 | 0.14 |
| polarC18 | 276.144 | 199.9428 | 7.2 | 0.14 |

| | | | | |
|---|---|---|---|---|
| polarC18 | 276.144 | 78.349 | 7.2 | 0.14 |
| polarC18 | 276.144 | 74.845 | 7.2 | 0.14 |
| polarC18 | 276.144 | 76.268 | 7.2 | 0.14 |
| polarC18 | 276.144 | 76.6911 | 7.2 | 0.14 |
| polarC18 | 276.144 | 100.162 | 7.2 | 0.14 |
| polarC18 | 276.144 | 105.5282 | 7.2 | 0.14 |
| polarC18 | 276.144 | 188.016 | 7.2 | 0.14 |
| polarC18 | 276.144 | 141.9131 | 7.2 | 0.14 |
| polarC18 | 276.144 | 156.3427 | 7.2 | 0.14 |
| polarC18 | 276.144 | 76.3083 | 7.2 | 0.14 |
| polarC18 | 276.144 | 52.8872 | 7.2 | 0.14 |
| polarC18 | 276.144 | 217.5184 | 7.2 | 0.14 |
| polarC18 | 276.144 | 107.2812 | 7.2 | 0.14 |
| polarC18 | 276.144 | 126.2895 | 7.2 | 0.14 |
| polarC18 | 276.144 | 148.7704 | 7.2 | 0.14 |
| polarC18 | 276.144 | 114.1074 | 7.2 | 0.14 |
| polarC18 | 276.144 | 258.8954 | 7.2 | 0.14 |
| polarC18 | 276.144 | 50.6914 | 7.2 | 0.14 |
| polarC18 | 276.144 | 158.7354 | 7.2 | 0.14 |
| polarC18 | 276.144 | 110.0754 | 7.2 | 0.14 |
| polarC18 | 276.144 | 214.7205 | 7.2 | 0.14 |
| polarC18 | 276.144 | 65.8852 | 7.2 | 0.14 |
| polarC18 | 276.144 | 176.8124 | 7.2 | 0.14 |
| polarC18 | 276.144 | 80.3127 | 7.2 | 0.14 |
| polarC18 | 276.144 | 58.6228 | 7.2 | 0.14 |
| polarC18 | 276.144 | 66.4687 | 7.2 | 0.14 |
| polarC18 | 276.144 | 82.3757 | 7.2 | 0.14 |
| polarC18 | 276.144 | 94.899 | 7.2 | 0.14 |
| polarC18 | 276.144 | 115.789 | 7.2 | 0.14 |
| polarC18 | 276.144 | 70.5167 | 7.2 | 0.14 |
| polarC18 | 276.144 | 50.8397 | 7.2 | 0.14 |
| polarC18 | 276.144 | 102.199 | 7.2 | 0.14 |
| polarC18 | 276.144 | 91.0804 | 7.2 | 0.14 |
| polarC18 | 276.144 | 162.4008 | 7.2 | 0.14 |
| polarC18 | 276.144 | 112.4232 | 7.2 | 0.14 |
| polarC18 | 276.144 | 60.1455 | 7.2 | 0.14 |
| polarC18 | 276.144 | 144.0703 | 7.2 | 0.14 |
| polarC18 | 276.144 | 68.5629 | 7.2 | 0.14 |
| polarC18 | 276.144 | 62.1528 | 7.2 | 0.14 |
| polarC18 | 276.144 | 66.346 | 7.2 | 0.14 |
| polarC18 | 276.144 | 73.1489 | 7.2 | 0.14 |
| polarC18 | 276.144 | 108.8938 | 7.2 | 0.14 |

| | | | | |
|---|---|---|---|---|
| polarC18 | 276.144 | 83.1086 | 7.2 | 0.14 |
| polarC18 | 276.144 | 107.8727 | 7.2 | 0.14 |
| polarC18 | 276.144 | 91.1078 | 7.2 | 0.14 |
| polarC18 | 276.144 | 59.1693 | 7.2 | 0.14 |
| polarC18 | 276.144 | 126.6227 | 7.2 | 0.14 |
| polarC18 | 276.144 | 66.6245 | 7.2 | 0.14 |
| polarC18 | 276.144 | 116.2113 | 7.2 | 0.14 |
| polarC18 | 276.144 | 107.7631 | 7.2 | 0.14 |
| polarC18 | 276.144 | 186.4553 | 7.2 | 0.14 |
| polarC18 | 276.144 | 212.4064 | 7.2 | 0.14 |
| polarC18 | 276.144 | 51.0309 | 7.2 | 0.14 |
| polarC18 | 276.144 | 123.7559 | 7.2 | 0.14 |
| polarC18 | 276.144 | 60.4396 | 7.2 | 0.14 |
| polarC18 | 276.144 | 71.553 | 7.2 | 0.14 |
| polarC18 | 276.144 | 114.9299 | 7.2 | 0.14 |
| polarC18 | 276.144 | 301.2592 | 7.2 | 0.14 |
| polarC18 | 276.144 | 66.2542 | 7.2 | 0.14 |
| polarC18 | 276.144 | 158.0189 | 7.2 | 0.14 |
| polarC18 | 276.144 | 62.4576 | 7.2 | 0.14 |
| polarC18 | 276.144 | 76.7589 | 7.2 | 0.14 |
| polarC18 | 276.144 | 92.1655 | 7.2 | 0.14 |
| polarC18 | 276.144 | 121.3764 | 7.2 | 0.14 |
| polarC18 | 276.144 | 108.728 | 7.2 | 0.14 |
| polarC18 | 276.144 | 50.3189 | 7.2 | 0.14 |
| polarC18 | 276.144 | 99.2258 | 7.2 | 0.14 |
| polarC18 | 276.144 | 243.3699 | 7.2 | 0.14 |
| polarC18 | 276.144 | 61.0002 | 7.2 | 0.14 |
| polarC18 | 276.144 | 233.0115 | 7.2 | 0.14 |
| polarC18 | 276.144 | 71.1998 | 7.2 | 0.14 |
| polarC18 | 276.144 | 246.4153 | 7.2 | 0.14 |
| polarC18 | 276.144 | 62.5335 | 7.2 | 0.14 |
| polarC18 | 276.144 | 52.8616 | 7.2 | 0.14 |
| polarC18 | 276.144 | 56.0405 | 7.2 | 0.14 |
| polarC18 | 276.144 | 140.893 | 7.2 | 0.14 |
| polarC18 | 276.144 | 98.5196 | 7.2 | 0.14 |
| polarC18 | 276.144 | 116.4255 | 7.2 | 0.14 |
| polarC18 | 276.144 | 97.8071 | 7.2 | 0.14 |
| polarC18 | 276.144 | 81.3247 | 7.2 | 0.14 |
| polarC18 | 276.144 | 222.9272 | 7.2 | 0.14 |
| polarC18 | 276.144 | 157.993 | 7.2 | 0.14 |
| polarC18 | 276.144 | 297.4678 | 7.2 | 0.14 |
| polarC18 | 276.144 | 77.2069 | 7.2 | 0.14 |

| | | | | |
|---|---|---|---|---|
| polarC18 | 276.144 | 94.8498 | 7.2 | 0.14 |
| polarC18 | 276.144 | 52.1922 | 7.2 | 0.14 |
| polarC18 | 276.144 | 87.6672 | 7.2 | 0.14 |
| polarC18 | 276.144 | 235.111 | 7.2 | 0.14 |
| polarC18 | 276.144 | 60.9574 | 7.2 | 0.14 |
| polarC18 | 276.144 | 163.5517 | 7.2 | 0.14 |
| polarC18 | 276.144 | 95.1625 | 7.2 | 0.14 |
| polarC18 | 276.144 | 207.3391 | 7.2 | 0.14 |
| polarC18 | 276.144 | 51.3515 | 7.2 | 0.14 |
| polarC18 | 276.144 | 98.6898 | 7.2 | 0.14 |
| polarC18 | 276.144 | 234.8918 | 7.2 | 0.14 |
| polarC18 | 276.144 | 89.1724 | 7.2 | 0.14 |
| polarC18 | 276.144 | 95.2913 | 7.2 | 0.14 |
| polarC18 | 276.144 | 78.4432 | 7.2 | 0.14 |
| polarC18 | 276.144 | 128.6288 | 7.2 | 0.14 |
| polarC18 | 276.144 | 63.5618 | 7.2 | 0.14 |
| polarC18 | 276.144 | 174.1639 | 7.2 | 0.14 |
| polarC18 | 276.144 | 81.8953 | 7.2 | 0.14 |
| polarC18 | 276.144 | 62.7807 | 7.2 | 0.14 |
| polarC18 | 276.144 | 51.5138 | 7.2 | 0.14 |
| polarC18 | 276.144 | 114.1925 | 7.2 | 0.14 |
| polarC18 | 276.144 | 61.4519 | 7.2 | 0.14 |
| polarC18 | 276.144 | 85.6203 | 7.1 | 0.14 |
| polarC18 | 276.144 | 149.6018 | 7.1 | 0.14 |
| polarC18 | 276.144 | 99.6092 | 7.1 | 0.14 |
| polarC18 | 276.144 | 59.0241 | 7.1 | 0.14 |
| polarC18 | 276.144 | 75.4441 | 7.1 | 0.14 |
| polarC18 | 276.144 | 232.2193 | 7.1 | 0.14 |
| polarC18 | 276.144 | 64.9729 | 7.1 | 0.14 |
| polarC18 | 276.144 | 50.1869 | 7.1 | 0.14 |
| polarC18 | 276.144 | 58.2283 | 7.1 | 0.14 |
| polarC18 | 276.144 | 112.5873 | 7.1 | 0.14 |
| polarC18 | 276.144 | 87.1463 | 7.1 | 0.14 |
| polarC18 | 276.144 | 56.5351 | 7.1 | 0.14 |
| polarC18 | 276.144 | 51.4758 | 7.1 | 0.14 |
| polarC18 | 276.144 | 78.8111 | 7.1 | 0.14 |
| polarC18 | 276.144 | 59.1222 | 7.1 | 0.14 |
| polarC18 | 276.144 | 85.5419 | 7.1 | 0.14 |
| polarC18 | 276.144 | 73.8157 | 7.1 | 0.14 |
| polarC18 | 276.144 | 213.4503 | 7.1 | 0.14 |
| polarC18 | 276.144 | 51.939 | 7.1 | 0.14 |
| polarC18 | 276.144 | 125.1904 | 7.1 | 0.14 |

| | | | | |
|---|---|---|---|---|
| polarC18 | 276.144 | 94.559 | 7.1 | 0.14 |
| polarC18 | 276.144 | 52.4209 | 7.1 | 0.14 |
| polarC18 | 276.144 | 181.1414 | 7.1 | 0.14 |
| polarC18 | 276.144 | 192.354 | 7.1 | 0.14 |
| polarC18 | 276.144 | 59.0866 | 7.1 | 0.14 |
| polarC18 | 276.144 | 68.8564 | 7.1 | 0.14 |
| polarC18 | 276.144 | 131.4394 | 7.1 | 0.14 |
| polarC18 | 276.144 | 74.1227 | 7.1 | 0.14 |
| polarC18 | 276.144 | 101.7322 | 7.1 | 0.14 |
| polarC18 | 276.144 | 262.7289 | 7.1 | 0.14 |
| polarC18 | 276.144 | 53.2218 | 7.1 | 0.14 |
| polarC18 | 276.144 | 50.3503 | 7.1 | 0.14 |
| polarC18 | 276.144 | 92.4141 | 7.1 | 0.14 |
| polarC18 | 276.144 | 141.1565 | 7.1 | 0.14 |
| polarC18 | 276.144 | 113.8176 | 7.1 | 0.14 |
| polarC18 | 276.144 | 70.0089 | 7.1 | 0.14 |
| polarC18 | 276.144 | 93.0218 | 7.1 | 0.14 |
| polarC18 | 276.144 | 272.7346 | 7.1 | 0.14 |
| polarC18 | 276.144 | 123.4774 | 7.1 | 0.14 |
| polarC18 | 276.144 | 72.8917 | 7.1 | 0.14 |
| polarC18 | 276.144 | 52.9245 | 7.1 | 0.14 |
| polarC18 | 276.144 | 71.0075 | 7.1 | 0.14 |
| polarC18 | 276.144 | 75.8673 | 7.1 | 0.14 |
| polarC18 | 276.144 | 109.7861 | 7.1 | 0.14 |
| polarC18 | 276.144 | 83.1912 | 7.1 | 0.14 |
| polarC18 | 276.144 | 75.3878 | 7.1 | 0.14 |
| polarC18 | 276.144 | 56.6277 | 7.1 | 0.14 |
| polarC18 | 276.144 | 125.9006 | 7.1 | 0.14 |
| polarC18 | 276.144 | 141.4592 | 7.1 | 0.14 |
| polarC18 | 276.144 | 76.9311 | 7.1 | 0.14 |
| polarC18 | 276.144 | 50.9061 | 7.1 | 0.14 |
| polarC18 | 276.144 | 147.7645 | 7.1 | 0.14 |
| polarC18 | 276.144 | 131.8934 | 7.1 | 0.14 |
| polarC18 | 276.144 | 64.1428 | 7.1 | 0.14 |
| polarC18 | 276.144 | 140.9826 | 7.1 | 0.14 |
| polarC18 | 276.144 | 75.0201 | 7.1 | 0.14 |
| polarC18 | 276.144 | 68.7914 | 7.1 | 0.14 |
| polarC18 | 276.144 | 117.7436 | 7.1 | 0.14 |
| polarC18 | 276.144 | 243.9112 | 7.1 | 0.14 |
| polarC18 | 276.144 | 137.7569 | 7.1 | 0.14 |
| polarC18 | 276.144 | 60.356 | 7.1 | 0.14 |
| polarC18 | 276.144 | 84.3423 | 7.1 | 0.14 |

| | | | | |
|---|---|---|---|---|
| polarC18 | 276.144 | 77.0946 | 7.1 | 0.14 |
| polarC18 | 276.144 | 88.9805 | 7.1 | 0.14 |
| polarC18 | 276.144 | 52.9068 | 7.1 | 0.14 |
| polarC18 | 276.144 | 71.0848 | 7.1 | 0.14 |
| polarC18 | 276.144 | 62.0372 | 7.1 | 0.14 |
| polarC18 | 276.144 | 55.1436 | 7.1 | 0.14 |
| polarC18 | 276.144 | 87.8299 | 7.1 | 0.14 |
| polarC18 | 276.144 | 64.8023 | 7.1 | 0.14 |
| polarC18 | 276.144 | 64.0751 | 7.1 | 0.14 |
| polarC18 | 276.144 | 74.3936 | 7.1 | 0.14 |
| polarC18 | 276.144 | 222.5022 | 7.1 | 0.14 |
| polarC18 | 276.144 | 69.9966 | 7.1 | 0.14 |
| polarC18 | 276.144 | 57.7289 | 7.1 | 0.14 |
| polarC18 | 276.144 | 92.1486 | 7.1 | 0.14 |
| polarC18 | 276.144 | 63.1121 | 7.1 | 0.14 |
| polarC18 | 276.144 | 65.7908 | 7.1 | 0.14 |
| polarC18 | 276.144 | 87.3792 | 7.1 | 0.14 |
| polarC18 | 276.144 | 171.0223 | 7.1 | 0.14 |
| polarC18 | 276.144 | 108.0037 | 7.1 | 0.14 |
| polarC18 | 276.144 | 54.4183 | 7.1 | 0.14 |
| polarC18 | 276.144 | 62.8006 | 7.1 | 0.14 |
| polarC18 | 276.144 | 156.4219 | 7.1 | 0.14 |
| polarC18 | 276.144 | 74.2804 | 7.1 | 0.14 |
| polarC18 | 276.144 | 183.1856 | 7.1 | 0.14 |
| polarC18 | 276.144 | 61.3495 | 7.1 | 0.14 |
| polarC18 | 276.144 | 64.3464 | 7.1 | 0.14 |
| polarC18 | 276.144 | 153.6532 | 7.1 | 0.14 |
| polarC18 | 276.144 | 96.4038 | 7.1 | 0.14 |
| polarC18 | 276.144 | 165.826 | 7.1 | 0.14 |
| polarC18 | 276.144 | 56.1003 | 7.1 | 0.14 |
| polarC18 | 276.144 | 201.3118 | 7.1 | 0.14 |
| polarC18 | 276.144 | 58.8248 | 7.1 | 0.14 |
| polarC18 | 276.144 | 74.5614 | 7.1 | 0.14 |
| polarC18 | 276.144 | 124.5414 | 7.1 | 0.14 |
| polarC18 | 276.144 | 131.2084 | 7.1 | 0.14 |
| polarC18 | 276.144 | 54.0807 | 7.1 | 0.14 |
| polarC18 | 276.144 | 70.4594 | 7.1 | 0.14 |
| polarC18 | 276.144 | 64.1947 | 7.1 | 0.14 |
| polarC18 | 276.144 | 97.7635 | 7.1 | 0.14 |
| polarC18 | 276.144 | 159.0638 | 7.1 | 0.14 |
| polarC18 | 276.144 | 152.8291 | 7.1 | 0.14 |
| polarC18 | 276.144 | 170.5797 | 7.1 | 0.14 |

| | | | | |
|---|---|---|---|---|
| polarC18 | 276.144 | 140.1777 | 7.1 | 0.14 |
| polarC18 | 276.144 | 52.3879 | 7.1 | 0.14 |
| polarC18 | 276.144 | 69.2282 | 7.1 | 0.14 |
| polarC18 | 276.144 | 102.7897 | 7.1 | 0.14 |
| polarC18 | 276.144 | 253.6268 | 7.1 | 0.14 |
| polarC18 | 276.144 | 110.5928 | 7.1 | 0.14 |
| polarC18 | 276.144 | 64.3137 | 7.1 | 0.14 |
| polarC18 | 276.144 | 170.636 | 7.1 | 0.14 |
| polarC18 | 276.144 | 122.4474 | 7.1 | 0.14 |
| polarC18 | 276.144 | 126.116 | 7.1 | 0.14 |
| polarC18 | 276.144 | 80.1825 | 7.1 | 0.14 |
| polarC18 | 276.144 | 81.9372 | 7.1 | 0.14 |
| polarC18 | 276.144 | 139.7543 | 7.1 | 0.14 |
| polarC18 | 276.144 | 96.2025 | 7.1 | 0.14 |
| polarC18 | 276.144 | 128.7867 | 7.1 | 0.14 |
| polarC18 | 276.144 | 60.5326 | 7.1 | 0.14 |
| polarC18 | 276.144 | 238.3701 | 7.1 | 0.14 |
| polarC18 | 276.144 | 205.4865 | 7.1 | 0.14 |
| polarC18 | 276.144 | 167.057 | 7.1 | 0.14 |
| polarC18 | 276.144 | 52.1007 | 7.1 | 0.14 |
| polarC18 | 276.144 | 102.6493 | 7.1 | 0.14 |
| polarC18 | 276.144 | 66.5385 | 7.1 | 0.14 |
| polarC18 | 276.144 | 62.5722 | 7.1 | 0.14 |
| polarC18 | 276.144 | 63.5652 | 7.1 | 0.14 |
| polarC18 | 276.144 | 248.2577 | 7.1 | 0.14 |
| polarC18 | 276.144 | 59.6797 | 7.1 | 0.14 |
| polarC18 | 276.144 | 185.4516 | 7.1 | 0.14 |
| polarC18 | 276.144 | 144.2394 | 7.1 | 0.14 |
| polarC18 | 276.144 | 296.6493 | 7.1 | 0.14 |
| polarC18 | 276.144 | 151.3153 | 7.1 | 0.14 |
| polarC18 | 276.144 | 52.2478 | 7.1 | 0.14 |
| polarC18 | 276.144 | 272.8869 | 7.1 | 0.14 |
| polarC18 | 276.144 | 159.0237 | 7.1 | 0.14 |
| polarC18 | 276.144 | 152.4891 | 7.1 | 0.14 |
| polarC18 | 276.144 | 94.5542 | 7.1 | 0.14 |
| polarC18 | 276.144 | 77.1065 | 7.1 | 0.14 |
| polarC18 | 276.144 | 76.8125 | 7.1 | 0.14 |
| polarC18 | 276.144 | 248.9083 | 7.1 | 0.14 |
| polarC18 | 276.144 | 175.5876 | 7.1 | 0.14 |
| polarC18 | 276.144 | 205.2135 | 7.1 | 0.14 |
| polarC18 | 276.144 | 264.1434 | 7.1 | 0.14 |
| polarC18 | 276.144 | 64.9237 | 7.1 | 0.14 |

| | | | | |
|---|---|---|---|---|
| polarC18 | 276.144 | 183.3897 | 7.1 | 0.14 |
| polarC18 | 276.144 | 67.7854 | 7.1 | 0.14 |
| polarC18 | 276.144 | 94.7014 | 7.1 | 0.14 |
| polarC18 | 276.144 | 286.942 | 7.1 | 0.14 |
| polarC18 | 276.144 | 51.5647 | 7.1 | 0.14 |
| polarC18 | 276.144 | 54.4199 | 7.1 | 0.14 |
| polarC18 | 276.144 | 126.0495 | 7.1 | 0.14 |
| polarC18 | 276.144 | 166.965 | 7.1 | 0.14 |
| polarC18 | 276.144 | 54.3441 | 7.1 | 0.14 |
| polarC18 | 276.144 | 126.8401 | 7.1 | 0.14 |
| polarC18 | 276.144 | 98.8184 | 7.1 | 0.14 |
| polarC18 | 276.144 | 222.2206 | 7.1 | 0.14 |
| polarC18 | 276.144 | 120.3832 | 7.1 | 0.14 |
| polarC18 | 276.144 | 242.817 | 7.1 | 0.14 |
| polarC18 | 276.144 | 70.4736 | 7.1 | 0.14 |
| polarC18 | 276.144 | 112.5519 | 7.1 | 0.14 |
| polarC18 | 276.144 | 300.4017 | 7.1 | 0.14 |
| polarC18 | 276.144 | 92.0451 | 7.1 | 0.14 |
| polarC18 | 276.144 | 197.8323 | 7.1 | 0.14 |
| polarC18 | 276.144 | 94.1257 | 7.1 | 0.14 |
| polarC18 | 276.144 | 129.1151 | 7.1 | 0.14 |
| polarC18 | 276.144 | 184.7981 | 7.1 | 0.14 |
| polarC18 | 276.144 | 64.7108 | 7.1 | 0.14 |
| polarC18 | 276.144 | 55.2312 | 7.1 | 0.14 |
| polarC18 | 276.144 | 57.262 | 7.1 | 0.14 |
| polarC18 | 276.144 | 103.4035 | 7.1 | 0.14 |
| polarC18 | 276.144 | 84.3473 | 7.1 | 0.14 |
| polarC18 | 276.144 | 55.6827 | 7.1 | 0.14 |
| polarC18 | 276.144 | 183.8959 | 7.1 | 0.14 |
| polarC18 | 276.144 | 111.3211 | 7.1 | 0.14 |
| polarC18 | 276.144 | 169.5003 | 7.1 | 0.14 |
| polarC18 | 276.144 | 101.129 | 7.1 | 0.14 |
| polarC18 | 276.144 | 105.7343 | 7.1 | 0.14 |
| polarC18 | 276.144 | 88.3869 | 7.1 | 0.14 |
| polarC18 | 276.144 | 68.55 | 7.1 | 0.14 |
| polarC18 | 276.144 | 61.6319 | 7.1 | 0.14 |
| polarC18 | 276.144 | 53.2987 | 7.1 | 0.14 |
| polarC18 | 276.144 | 92.0849 | 7.1 | 0.14 |
| polarC18 | 276.144 | 108.762 | 7.1 | 0.14 |
| polarC18 | 276.144 | 94.3989 | 7.1 | 0.14 |
| polarC18 | 276.144 | 118.8651 | 7.1 | 0.14 |
| polarC18 | 276.144 | 90.6749 | 7.1 | 0.14 |

| | | | | |
|---|---|---|---|---|
| polarC18 | 276.144 | 52.945 | 7.1 | 0.14 |
| polarC18 | 276.144 | 119.1339 | 7.1 | 0.14 |
| polarC18 | 276.144 | 121.2199 | 7.1 | 0.14 |
| polarC18 | 276.144 | 73.0579 | 7.1 | 0.14 |
| polarC18 | 276.144 | 108.7711 | 7.1 | 0.14 |
| polarC18 | 276.144 | 149.9051 | 7.1 | 0.14 |
| polarC18 | 276.144 | 74.0398 | 7.1 | 0.14 |
| polarC18 | 276.144 | 64.562 | 7.1 | 0.14 |
| polarC18 | 276.144 | 112.4994 | 7.1 | 0.14 |
| polarC18 | 276.144 | 174.9217 | 7.1 | 0.14 |
| polarC18 | 276.144 | 174.8339 | 7.1 | 0.14 |
| polarC18 | 276.144 | 74.0783 | 7.1 | 0.14 |
| polarC18 | 276.144 | 139.4445 | 7.1 | 0.14 |
| polarC18 | 276.144 | 87.1581 | 7.1 | 0.14 |
| polarC18 | 276.144 | 138.2432 | 7.1 | 0.14 |
| polarC18 | 276.144 | 62.9819 | 7.1 | 0.14 |
| polarC18 | 276.144 | 52.145 | 7.1 | 0.14 |
| polarC18 | 276.144 | 91.5358 | 7.1 | 0.14 |
| polarC18 | 276.144 | 179.8482 | 7.1 | 0.14 |
| polarC18 | 276.144 | 105.9971 | 7.1 | 0.14 |
| polarC18 | 276.144 | 117.0221 | 7.1 | 0.14 |
| polarC18 | 276.144 | 118.8053 | 7.1 | 0.14 |
| polarC18 | 276.144 | 194.358 | 7.1 | 0.14 |
| polarC18 | 276.144 | 184.5964 | 7.1 | 0.14 |
| polarC18 | 276.144 | 291.9695 | 7.1 | 0.14 |
| polarC18 | 276.144 | 83.1271 | 7.1 | 0.14 |
| polarC18 | 276.144 | 260.2773 | 7.1 | 0.14 |
| polarC18 | 276.144 | 243.5241 | 7.1 | 0.14 |
| polarC18 | 276.144 | 101.8603 | 7.1 | 0.14 |
| polarC18 | 276.144 | 55.0456 | 7.1 | 0.14 |
| polarC18 | 276.144 | 151.4331 | 7.1 | 0.14 |
| polarC18 | 276.144 | 231.1643 | 7.1 | 0.14 |
| polarC18 | 276.144 | 76.189 | 7.1 | 0.14 |
| polarC18 | 276.144 | 55.7188 | 7.1 | 0.14 |
| polarC18 | 276.144 | 90.8956 | 7.1 | 0.14 |
| polarC18 | 276.144 | 192.9051 | 7.1 | 0.14 |
| polarC18 | 276.144 | 54.3307 | 7.1 | 0.14 |
| polarC18 | 276.144 | 55.3674 | 7.1 | 0.14 |
| polarC18 | 276.144 | 120.5352 | 7.1 | 0.14 |
| polarC18 | 276.144 | 121.8711 | 7.1 | 0.14 |
| polarC18 | 276.144 | 200.3308 | 7 | 0.14 |
| polarC18 | 276.144 | 172.007 | 7 | 0.14 |

| | | | | |
|---|---|---|---|---|
| polarC18 | 276.144 | 282.8028 | 7 | 0.14 |
| polarC18 | 276.144 | 85.9726 | 7 | 0.14 |
| polarC18 | 276.144 | 133.8048 | 7 | 0.14 |
| polarC18 | 276.144 | 106.2214 | 7 | 0.14 |
| polarC18 | 276.144 | 151.4974 | 7 | 0.14 |
| polarC18 | 276.144 | 165.9018 | 7 | 0.14 |
| polarC18 | 276.144 | 144.5233 | 7 | 0.14 |
| polarC18 | 276.144 | 270.386 | 7 | 0.14 |
| polarC18 | 276.144 | 100.6664 | 7 | 0.14 |
| polarC18 | 276.144 | 66.6385 | 7 | 0.14 |
| polarC18 | 276.144 | 204.376 | 7 | 0.14 |
| polarC18 | 276.144 | 88.8834 | 7 | 0.14 |
| polarC18 | 276.144 | 151.9791 | 7 | 0.14 |
| polarC18 | 276.144 | 135.6326 | 7 | 0.14 |
| polarC18 | 276.144 | 81.9119 | 7 | 0.14 |
| polarC18 | 276.144 | 92.7721 | 7 | 0.14 |
| polarC18 | 276.144 | 56.0527 | 7 | 0.14 |
| polarC18 | 276.144 | 141.1706 | 7 | 0.14 |
| polarC18 | 276.144 | 127.7651 | 7 | 0.14 |
| polarC18 | 276.144 | 97.9721 | 7 | 0.14 |
| polarC18 | 276.144 | 92.9837 | 7 | 0.14 |
| polarC18 | 276.144 | 110.1114 | 7 | 0.14 |
| polarC18 | 276.144 | 108.1605 | 7 | 0.14 |
| polarC18 | 276.144 | 67.1441 | 7 | 0.14 |
| polarC18 | 276.144 | 60.9375 | 7 | 0.14 |
| polarC18 | 276.144 | 50.0726 | 7 | 0.14 |
| polarC18 | 276.144 | 86.8141 | 7 | 0.14 |
| polarC18 | 276.144 | 142.2102 | 7 | 0.14 |
| polarC18 | 276.144 | 72.2923 | 7 | 0.14 |
| polarC18 | 276.144 | 94.7544 | 7 | 0.14 |
| polarC18 | 276.144 | 260.8486 | 7 | 0.14 |
| polarC18 | 276.144 | 73.4773 | 7 | 0.14 |
| polarC18 | 276.144 | 54.4289 | 7 | 0.14 |
| polarC18 | 276.144 | 110.9664 | 7 | 0.14 |
| polarC18 | 276.144 | 182.695 | 7 | 0.14 |
| polarC18 | 276.144 | 186.1232 | 7 | 0.14 |
| polarC18 | 276.144 | 149.3089 | 7 | 0.14 |
| polarC18 | 276.144 | 68.2959 | 7 | 0.14 |
| polarC18 | 276.144 | 50.4849 | 7 | 0.14 |
| polarC18 | 276.144 | 85.044 | 7 | 0.14 |
| polarC18 | 276.144 | 58.0239 | 7 | 0.14 |
| polarC18 | 276.144 | 94.4494 | 7 | 0.14 |

| | | | | |
|---|---|---|---|---|
| polarC18 | 276.144 | 122.059 | 7 | 0.14 |
| polarC18 | 276.144 | 87.5456 | 7 | 0.14 |
| polarC18 | 276.144 | 151.4483 | 7 | 0.14 |
| polarC18 | 276.144 | 69.2616 | 7 | 0.14 |
| polarC18 | 276.144 | 231.9579 | 7 | 0.14 |
| polarC18 | 276.144 | 59.5065 | 7 | 0.14 |
| polarC18 | 276.144 | 70.6837 | 7 | 0.14 |
| polarC18 | 276.144 | 78.537 | 7 | 0.14 |
| polarC18 | 276.144 | 169.7758 | 7 | 0.14 |
| polarC18 | 276.144 | 70.7558 | 7 | 0.14 |
| polarC18 | 276.144 | 114.0694 | 7 | 0.14 |
| polarC18 | 276.144 | 137.7182 | 7 | 0.14 |
| polarC18 | 276.144 | 65.8462 | 7 | 0.14 |
| polarC18 | 276.144 | 254.4193 | 7 | 0.14 |
| polarC18 | 276.144 | 67.6427 | 7 | 0.14 |
| polarC18 | 276.144 | 115.5038 | 7 | 0.14 |
| polarC18 | 276.144 | 140.0592 | 7 | 0.14 |
| polarC18 | 276.144 | 246.1212 | 7 | 0.14 |
| polarC18 | 276.144 | 105.8727 | 7 | 0.14 |
| polarC18 | 276.144 | 126.8102 | 7 | 0.14 |
| polarC18 | 276.144 | 73.4253 | 7 | 0.14 |
| polarC18 | 276.144 | 66.8242 | 7 | 0.14 |
| polarC18 | 276.144 | 92.4562 | 7 | 0.14 |
| polarC18 | 276.144 | 156.2128 | 7 | 0.14 |
| polarC18 | 276.144 | 189.6256 | 7 | 0.14 |
| polarC18 | 276.144 | 255.0791 | 7 | 0.14 |
| polarC18 | 276.144 | 260.414 | 7 | 0.14 |
| polarC18 | 276.144 | 188.9089 | 7 | 0.14 |
| polarC18 | 276.144 | 73.4824 | 7 | 0.14 |
| polarC18 | 276.144 | 164.0002 | 7 | 0.14 |
| polarC18 | 276.144 | 64.8049 | 7 | 0.14 |
| polarC18 | 276.144 | 111.8451 | 7 | 0.14 |
| polarC18 | 276.144 | 254.6722 | 7 | 0.14 |
| polarC18 | 276.144 | 166.8859 | 7 | 0.14 |
| polarC18 | 276.144 | 65.7033 | 7 | 0.13 |
| polarC18 | 276.144 | 109.9506 | 7 | 0.13 |
| polarC18 | 276.144 | 94.1767 | 7 | 0.13 |
| polarC18 | 276.144 | 149.5345 | 7 | 0.13 |
| polarC18 | 276.144 | 148.4004 | 7 | 0.13 |
| polarC18 | 276.144 | 62.6808 | 7 | 0.13 |
| polarC18 | 276.144 | 70.5348 | 7 | 0.13 |
| polarC18 | 276.144 | 76.2813 | 7 | 0.13 |

| | | | | |
|---|---|---|---|---|
| polarC18 | 276.144 | 211.2817 | 7 | 0.13 |
| polarC18 | 276.144 | 134.6715 | 7 | 0.13 |
| polarC18 | 276.144 | 95.2434 | 7 | 0.13 |
| polarC18 | 276.144 | 70.3211 | 7 | 0.13 |
| polarC18 | 276.144 | 131.6585 | 7 | 0.13 |
| polarC18 | 276.144 | 85.764 | 7 | 0.13 |
| polarC18 | 276.144 | 157.0623 | 7 | 0.13 |
| polarC18 | 276.144 | 266.0535 | 7 | 0.13 |
| polarC18 | 276.144 | 64.7148 | 7 | 0.13 |
| polarC18 | 276.144 | 59.9419 | 7 | 0.13 |
| polarC18 | 276.144 | 75.3975 | 7 | 0.13 |
| polarC18 | 276.144 | 71.4211 | 7 | 0.13 |
| polarC18 | 276.144 | 55.1347 | 7 | 0.13 |
| polarC18 | 276.144 | 96.8812 | 7 | 0.13 |
| polarC18 | 276.144 | 59.0148 | 7 | 0.13 |
| polarC18 | 276.144 | 52.2438 | 7 | 0.13 |
| polarC18 | 276.144 | 218.2943 | 7 | 0.13 |
| polarC18 | 276.144 | 122.212 | 7 | 0.13 |
| polarC18 | 276.144 | 97.3568 | 7 | 0.13 |
| polarC18 | 276.144 | 196.023 | 7 | 0.13 |
| polarC18 | 276.144 | 95.6668 | 7 | 0.13 |
| polarC18 | 276.144 | 198.5894 | 7 | 0.13 |
| polarC18 | 276.144 | 204.2292 | 7 | 0.13 |
| polarC18 | 276.144 | 64.6144 | 7 | 0.13 |
| polarC18 | 276.144 | 55.32 | 7 | 0.13 |
| polarC18 | 276.144 | 117.3245 | 7 | 0.13 |
| polarC18 | 276.144 | 73.7792 | 7 | 0.13 |
| polarC18 | 276.144 | 57.4582 | 7 | 0.13 |
| polarC18 | 276.144 | 132.667 | 7 | 0.13 |
| polarC18 | 276.144 | 83.225 | 7 | 0.13 |
| polarC18 | 276.144 | 255.2597 | 7 | 0.13 |
| polarC18 | 276.144 | 61.1363 | 7 | 0.13 |
| polarC18 | 276.144 | 104.0565 | 7 | 0.13 |
| polarC18 | 276.144 | 271.7251 | 7 | 0.13 |
| polarC18 | 276.144 | 173.5152 | 7 | 0.13 |
| polarC18 | 276.144 | 203.3082 | 7 | 0.13 |
| polarC18 | 276.144 | 70.0423 | 7 | 0.13 |
| polarC18 | 276.144 | 69.0959 | 7 | 0.13 |
| polarC18 | 276.144 | 247.4849 | 7 | 0.13 |
| polarC18 | 276.144 | 194.4459 | 7 | 0.13 |
| polarC18 | 276.144 | 252.3167 | 7 | 0.13 |
| polarC18 | 276.144 | 62.3006 | 7 | 0.13 |

| | | | | |
|---|---|---|---|---|
| polarC18 | 276.144 | 85.4913 | 7 | 0.13 |
| polarC18 | 276.144 | 55.4837 | 7 | 0.13 |
| polarC18 | 276.144 | 80.6737 | 7 | 0.13 |
| polarC18 | 276.144 | 107.6711 | 7 | 0.13 |
| polarC18 | 276.144 | 80.5036 | 7 | 0.13 |
| polarC18 | 276.144 | 87.0166 | 7 | 0.13 |
| polarC18 | 276.144 | 245.4295 | 7 | 0.13 |
| polarC18 | 276.144 | 50.0997 | 7 | 0.13 |
| polarC18 | 276.144 | 71.1971 | 7 | 0.13 |
| polarC18 | 276.144 | 54.8298 | 7 | 0.13 |
| polarC18 | 276.144 | 85.6911 | 7 | 0.13 |
| polarC18 | 276.144 | 107.962 | 7 | 0.13 |
| polarC18 | 276.144 | 234.1709 | 7 | 0.13 |
| polarC18 | 276.144 | 233.7366 | 7 | 0.13 |
| polarC18 | 276.144 | 93.573 | 7 | 0.13 |
| polarC18 | 276.144 | 72.2783 | 7 | 0.13 |
| polarC18 | 276.144 | 89.5777 | 7 | 0.13 |
| polarC18 | 276.144 | 66.663 | 7 | 0.13 |
| polarC18 | 276.144 | 121.3444 | 7 | 0.13 |
| polarC18 | 276.144 | 271.2169 | 7 | 0.13 |
| polarC18 | 276.144 | 213.9556 | 7 | 0.13 |
| polarC18 | 276.144 | 130.9461 | 7 | 0.13 |
| polarC18 | 276.144 | 50.2489 | 7 | 0.13 |
| polarC18 | 276.144 | 130.1419 | 7 | 0.13 |
| polarC18 | 276.144 | 61.2115 | 7 | 0.13 |
| polarC18 | 276.144 | 63.2357 | 7 | 0.13 |
| polarC18 | 276.144 | 225.9222 | 7 | 0.13 |
| polarC18 | 276.144 | 115.4329 | 7 | 0.13 |
| polarC18 | 276.144 | 198.5594 | 7 | 0.13 |
| polarC18 | 276.144 | 67.703 | 7 | 0.13 |
| polarC18 | 276.144 | 92.4236 | 7 | 0.13 |
| polarC18 | 276.144 | 268.6966 | 7 | 0.13 |
| polarC18 | 276.144 | 92.3966 | 7 | 0.13 |
| polarC18 | 276.144 | 279.9753 | 7 | 0.13 |
| polarC18 | 276.144 | 108.7148 | 7 | 0.13 |
| polarC18 | 276.144 | 149.7794 | 7 | 0.13 |
| polarC18 | 276.144 | 282.9227 | 7 | 0.13 |
| polarC18 | 276.144 | 72.0192 | 7 | 0.13 |
| polarC18 | 276.144 | 140.3403 | 7 | 0.13 |
| polarC18 | 276.144 | 95.9018 | 7 | 0.13 |
| polarC18 | 276.144 | 148.9669 | 7 | 0.13 |
| polarC18 | 276.144 | 57.251 | 7 | 0.13 |

| | | | | |
|---|---|---|---|---|
| polarC18 | 276.144 | 82.8867 | 7 | 0.13 |
| polarC18 | 276.144 | 63.2429 | 7 | 0.13 |
| polarC18 | 276.144 | 136.9425 | 7 | 0.13 |
| polarC18 | 276.144 | 61.9416 | 7 | 0.13 |
| polarC18 | 276.144 | 121.0495 | 7 | 0.13 |
| polarC18 | 276.144 | 54.8653 | 7 | 0.13 |
| polarC18 | 276.144 | 230.2347 | 7 | 0.13 |
| polarC18 | 276.144 | 239.7819 | 7 | 0.13 |
| polarC18 | 276.144 | 108.0967 | 7 | 0.13 |
| polarC18 | 276.144 | 91.1584 | 7 | 0.13 |
| polarC18 | 276.144 | 167.7156 | 7 | 0.13 |
| polarC18 | 276.144 | 87.1335 | 7 | 0.13 |
| polarC18 | 276.144 | 79.0809 | 7 | 0.13 |
| polarC18 | 276.144 | 130.7562 | 7 | 0.13 |
| polarC18 | 276.144 | 259.8701 | 7 | 0.13 |
| polarC18 | 276.144 | 205.8826 | 7 | 0.13 |
| polarC18 | 276.144 | 298.3646 | 7 | 0.13 |
| polarC18 | 276.144 | 66.1492 | 7 | 0.13 |
| polarC18 | 276.144 | 168.9978 | 7 | 0.13 |
| polarC18 | 276.144 | 77.9474 | 7 | 0.13 |
| polarC18 | 276.144 | 57.4923 | 7 | 0.13 |
| polarC18 | 276.144 | 52.4567 | 7 | 0.13 |
| polarC18 | 276.144 | 105.7524 | 7 | 0.13 |
| polarC18 | 276.144 | 51.8111 | 7 | 0.13 |
| polarC18 | 276.144 | 65.4707 | 7 | 0.13 |
| polarC18 | 276.144 | 86.5874 | 7 | 0.13 |
| polarC18 | 276.144 | 109.0499 | 7 | 0.13 |
| polarC18 | 276.144 | 57.0748 | 7 | 0.13 |
| polarC18 | 276.144 | 116.2985 | 7 | 0.13 |
| polarC18 | 276.144 | 92.8214 | 7 | 0.13 |
| polarC18 | 276.144 | 165.1811 | 7 | 0.13 |
| polarC18 | 276.144 | 75.5325 | 7 | 0.13 |
| polarC18 | 276.144 | 127.0205 | 7 | 0.13 |
| polarC18 | 276.144 | 162.3795 | 7 | 0.13 |
| polarC18 | 276.144 | 96.4419 | 7 | 0.13 |
| polarC18 | 276.144 | 82.2999 | 7 | 0.13 |
| polarC18 | 276.144 | 54.3588 | 7 | 0.13 |
| polarC18 | 276.144 | 51.8127 | 7 | 0.13 |
| polarC18 | 276.144 | 91.3063 | 7 | 0.13 |
| polarC18 | 276.144 | 262.0097 | 7 | 0.13 |
| polarC18 | 276.144 | 79.8228 | 7 | 0.13 |
| polarC18 | 276.144 | 209.9246 | 7 | 0.13 |

| | | | | |
|---|---|---|---|---|
| polarC18 | 276.144 | 138.6749 | 7 | 0.13 |
| polarC18 | 276.144 | 237.7995 | 7 | 0.13 |
| polarC18 | 276.144 | 50.5461 | 7 | 0.13 |
| polarC18 | 276.144 | 159.2911 | 7 | 0.13 |
| polarC18 | 276.144 | 79.9173 | 7 | 0.13 |
| polarC18 | 276.144 | 51.2067 | 7 | 0.13 |
| polarC18 | 276.144 | 156.0046 | 7 | 0.13 |
| polarC18 | 276.144 | 117.5293 | 7 | 0.13 |
| polarC18 | 276.144 | 126.7066 | 7 | 0.13 |
| polarC18 | 276.144 | 122.0197 | 7 | 0.13 |
| polarC18 | 276.144 | 80.4414 | 7 | 0.13 |
| polarC18 | 276.144 | 178.2495 | 7 | 0.13 |
| polarC18 | 276.144 | 75.5801 | 7 | 0.13 |
| polarC18 | 276.144 | 254.3559 | 7 | 0.13 |
| polarC18 | 276.144 | 50.0019 | 7 | 0.13 |
| polarC18 | 276.144 | 53.5495 | 7 | 0.13 |
| polarC18 | 276.144 | 88.7456 | 7 | 0.13 |
| polarC18 | 276.144 | 62.1834 | 7 | 0.13 |
| polarC18 | 276.144 | 56.2175 | 7 | 0.13 |
| polarC18 | 276.144 | 65.6498 | 7 | 0.13 |
| polarC18 | 276.144 | 130.2252 | 7 | 0.13 |
| polarC18 | 276.144 | 175.8587 | 7 | 0.13 |
| polarC18 | 276.144 | 233.6605 | 7 | 0.13 |
| polarC18 | 276.144 | 123.242 | 7 | 0.13 |
| polarC18 | 276.144 | 193.2156 | 7 | 0.13 |
| polarC18 | 276.144 | 94.8583 | 7 | 0.13 |
| polarC18 | 276.144 | 134.6901 | 7 | 0.13 |
| polarC18 | 276.144 | 100.2644 | 7 | 0.13 |
| polarC18 | 276.144 | 75.7473 | 7 | 0.13 |
| polarC18 | 276.144 | 55.9312 | 7 | 0.13 |
| polarC18 | 276.144 | 69.8228 | 7 | 0.13 |
| polarC18 | 276.144 | 57.3396 | 7 | 0.13 |
| polarC18 | 276.144 | 129.5379 | 7 | 0.13 |
| polarC18 | 276.144 | 175.5403 | 7 | 0.13 |
| polarC18 | 276.144 | 67.8063 | 7 | 0.13 |
| polarC18 | 276.144 | 74.3563 | 7 | 0.13 |
| polarC18 | 276.144 | 220.7734 | 7 | 0.13 |
| polarC18 | 276.144 | 149.6981 | 6.9 | 0.13 |
| polarC18 | 276.144 | 279.4137 | 6.9 | 0.13 |
| polarC18 | 276.144 | 99.6329 | 6.9 | 0.13 |
| polarC18 | 276.144 | 103.8259 | 6.9 | 0.13 |
| polarC18 | 276.144 | 90.4134 | 6.9 | 0.13 |

| | | | | |
|---|---|---|---|---|
| polarC18 | 276.144 | 237.0187 | 6.9 | 0.13 |
| polarC18 | 276.144 | 74.3856 | 6.9 | 0.13 |
| polarC18 | 276.144 | 149.8269 | 6.9 | 0.13 |
| polarC18 | 276.144 | 77.3539 | 6.9 | 0.13 |
| polarC18 | 276.144 | 100.1179 | 6.9 | 0.13 |
| polarC18 | 276.144 | 73.2883 | 6.9 | 0.13 |
| polarC18 | 276.144 | 280.5836 | 6.9 | 0.13 |
| polarC18 | 276.144 | 157.4177 | 6.9 | 0.13 |
| polarC18 | 276.144 | 132.119 | 6.9 | 0.13 |
| polarC18 | 276.144 | 54.2131 | 6.9 | 0.13 |
| polarC18 | 276.144 | 68.6666 | 6.9 | 0.13 |
| polarC18 | 276.144 | 118.7911 | 6.9 | 0.13 |
| polarC18 | 276.144 | 60.7098 | 6.9 | 0.13 |
| polarC18 | 276.144 | 140.5227 | 6.9 | 0.13 |
| polarC18 | 276.144 | 52.3151 | 6.9 | 0.13 |
| polarC18 | 276.144 | 204.7703 | 6.9 | 0.13 |
| polarC18 | 276.144 | 158.9347 | 6.9 | 0.13 |
| polarC18 | 276.144 | 122.2596 | 6.9 | 0.13 |
| polarC18 | 276.144 | 263.051 | 6.9 | 0.13 |
| polarC18 | 276.144 | 113.5599 | 6.9 | 0.13 |
| polarC18 | 276.144 | 64.9195 | 6.9 | 0.13 |
| polarC18 | 276.144 | 57.0497 | 6.9 | 0.13 |
| polarC18 | 276.144 | 130.3659 | 6.9 | 0.13 |
| polarC18 | 276.144 | 55.2156 | 6.9 | 0.13 |
| polarC18 | 276.144 | 117.5363 | 6.9 | 0.13 |
| polarC18 | 276.144 | 74.7033 | 6.9 | 0.13 |
| polarC18 | 276.144 | 88.6808 | 6.9 | 0.13 |
| polarC18 | 276.144 | 103.0394 | 6.9 | 0.13 |
| polarC18 | 276.144 | 218.2663 | 6.9 | 0.13 |
| polarC18 | 276.144 | 129.5185 | 6.9 | 0.13 |
| polarC18 | 276.144 | 79.4626 | 6.9 | 0.13 |
| polarC18 | 276.144 | 54.3276 | 6.9 | 0.13 |
| polarC18 | 276.144 | 154.4056 | 6.9 | 0.13 |
| polarC18 | 276.144 | 69.6553 | 6.9 | 0.13 |
| polarC18 | 276.144 | 54.905 | 6.9 | 0.13 |
| polarC18 | 276.144 | 98.1363 | 6.9 | 0.13 |
| polarC18 | 276.144 | 104.977 | 6.9 | 0.13 |
| polarC18 | 276.144 | 64.6007 | 6.9 | 0.13 |
| polarC18 | 276.144 | 156.7936 | 6.9 | 0.13 |
| polarC18 | 276.144 | 90.7341 | 6.9 | 0.13 |
| polarC18 | 276.144 | 74.656 | 6.9 | 0.13 |
| polarC18 | 276.144 | 52.443 | 6.9 | 0.13 |

| | | | | |
|---|---|---|---|---|
| polarC18 | 276.144 | 65.0562 | 6.9 | 0.13 |
| polarC18 | 276.144 | 263.5552 | 6.9 | 0.13 |
| polarC18 | 276.144 | 95.6049 | 6.9 | 0.13 |
| polarC18 | 276.144 | 58.1483 | 6.9 | 0.13 |
| polarC18 | 276.144 | 219.1237 | 6.9 | 0.13 |
| polarC18 | 276.144 | 102.1197 | 6.9 | 0.13 |
| polarC18 | 276.144 | 299.2424 | 6.9 | 0.13 |
| polarC18 | 276.144 | 86.6105 | 6.9 | 0.13 |
| polarC18 | 276.144 | 87.8808 | 6.9 | 0.13 |
| polarC18 | 276.144 | 57.7607 | 6.9 | 0.13 |
| polarC18 | 276.144 | 93.4443 | 6.9 | 0.13 |
| polarC18 | 276.144 | 116.8911 | 6.9 | 0.13 |
| polarC18 | 276.144 | 145.0949 | 6.9 | 0.13 |
| polarC18 | 276.144 | 51.5262 | 6.9 | 0.13 |
| polarC18 | 276.144 | 73.7419 | 6.9 | 0.13 |
| polarC18 | 276.144 | 56.6299 | 6.9 | 0.13 |
| polarC18 | 276.144 | 233.6268 | 6.9 | 0.13 |
| polarC18 | 276.144 | 105.1432 | 6.9 | 0.13 |
| polarC18 | 276.144 | 61.9832 | 6.9 | 0.13 |
| polarC18 | 276.144 | 113.5151 | 6.9 | 0.13 |
| polarC18 | 276.144 | 153.7863 | 6.9 | 0.13 |
| polarC18 | 276.144 | 161.5098 | 6.9 | 0.13 |
| polarC18 | 276.144 | 290.0698 | 6.9 | 0.13 |
| polarC18 | 276.144 | 161.4926 | 6.9 | 0.13 |
| polarC18 | 276.144 | 65.6948 | 6.9 | 0.13 |
| polarC18 | 276.144 | 97.0738 | 6.9 | 0.13 |
| polarC18 | 276.144 | 75.8094 | 6.9 | 0.13 |
| polarC18 | 276.144 | 80.4671 | 6.9 | 0.13 |
| polarC18 | 276.144 | 104.2904 | 6.9 | 0.13 |
| polarC18 | 276.144 | 151.9655 | 6.9 | 0.13 |
| polarC18 | 276.144 | 60.9913 | 6.9 | 0.13 |
| polarC18 | 276.144 | 185.3178 | 6.9 | 0.13 |
| polarC18 | 276.144 | 253.777 | 6.9 | 0.13 |
| polarC18 | 276.144 | 249.0666 | 6.9 | 0.13 |
| polarC18 | 276.144 | 60.3728 | 6.9 | 0.13 |
| polarC18 | 276.144 | 50.6783 | 6.9 | 0.13 |
| polarC18 | 276.144 | 73.4687 | 6.9 | 0.13 |
| polarC18 | 276.144 | 109.4185 | 6.9 | 0.13 |
| polarC18 | 276.144 | 197.7529 | 6.9 | 0.13 |
| polarC18 | 276.144 | 83.8574 | 6.9 | 0.13 |
| polarC18 | 276.144 | 99.783 | 6.9 | 0.13 |
| polarC18 | 276.144 | 99.082 | 6.9 | 0.13 |

| | | | | |
|---|---|---|---|---|
| polarC18 | 276.144 | 67.3121 | 6.9 | 0.13 |
| polarC18 | 276.144 | 181.8604 | 6.9 | 0.13 |
| polarC18 | 276.144 | 92.7505 | 6.9 | 0.13 |
| polarC18 | 276.144 | 252.5634 | 6.9 | 0.13 |
| polarC18 | 276.144 | 82.2384 | 6.9 | 0.13 |
| polarC18 | 276.144 | 154.7728 | 6.9 | 0.13 |
| polarC18 | 276.144 | 219.0382 | 6.9 | 0.13 |
| polarC18 | 276.144 | 50.4083 | 6.9 | 0.13 |
| polarC18 | 276.144 | 123.5763 | 6.9 | 0.13 |
| polarC18 | 276.144 | 94.0171 | 6.9 | 0.13 |
| polarC18 | 276.144 | 62.3913 | 6.9 | 0.13 |
| polarC18 | 276.144 | 57.6194 | 6.9 | 0.13 |
| polarC18 | 276.144 | 157.3 | 6.9 | 0.13 |
| polarC18 | 276.144 | 57.4606 | 6.9 | 0.13 |
| polarC18 | 276.144 | 115.5256 | 6.9 | 0.13 |
| polarC18 | 276.144 | 136.7523 | 6.9 | 0.13 |
| polarC18 | 276.144 | 143.8193 | 6.9 | 0.13 |
| polarC18 | 276.144 | 195.8105 | 6.9 | 0.13 |
| polarC18 | 276.144 | 204.7275 | 6.9 | 0.13 |
| polarC18 | 276.144 | 224.2124 | 6.9 | 0.13 |
| polarC18 | 276.144 | 59.9959 | 6.9 | 0.13 |
| polarC18 | 276.144 | 167.9185 | 6.9 | 0.13 |
| polarC18 | 276.144 | 117.3141 | 6.9 | 0.13 |
| polarC18 | 276.144 | 53.9425 | 6.9 | 0.13 |
| polarC18 | 276.144 | 116.9562 | 6.9 | 0.13 |
| polarC18 | 276.144 | 108.1055 | 6.9 | 0.13 |
| polarC18 | 276.144 | 288.8164 | 6.9 | 0.13 |
| polarC18 | 276.144 | 52.9297 | 6.9 | 0.13 |
| polarC18 | 276.144 | 62.8307 | 6.9 | 0.13 |
| polarC18 | 276.144 | 243.0309 | 6.9 | 0.13 |
| polarC18 | 276.144 | 69.8935 | 6.9 | 0.13 |
| polarC18 | 276.144 | 114.3707 | 6.9 | 0.13 |
| polarC18 | 276.144 | 97.5653 | 6.9 | 0.13 |
| polarC18 | 276.144 | 71.7735 | 6.9 | 0.13 |
| polarC18 | 276.144 | 133.5721 | 6.9 | 0.13 |
| polarC18 | 276.144 | 182.3149 | 6.9 | 0.13 |
| polarC18 | 276.144 | 52.2634 | 6.9 | 0.13 |
| polarC18 | 276.144 | 72.2641 | 6.9 | 0.13 |
| polarC18 | 276.144 | 114.3276 | 6.9 | 0.13 |
| polarC18 | 276.144 | 107.1292 | 6.9 | 0.13 |
| polarC18 | 276.144 | 100.1829 | 6.9 | 0.13 |
| polarC18 | 276.144 | 111.601 | 6.9 | 0.13 |

| | | | | |
|---|---|---|---|---|
| polarC18 | 276.144 | 56.1632 | 6.9 | 0.13 |
| polarC18 | 276.144 | 59.9295 | 6.9 | 0.13 |
| polarC18 | 276.144 | 115.6792 | 6.9 | 0.13 |
| polarC18 | 276.144 | 100.6221 | 6.9 | 0.13 |
| polarC18 | 276.144 | 54.0095 | 6.9 | 0.13 |
| polarC18 | 276.144 | 193.0701 | 6.9 | 0.13 |
| polarC18 | 276.144 | 132.2706 | 6.9 | 0.13 |
| polarC18 | 276.144 | 68.1612 | 6.9 | 0.13 |
| polarC18 | 276.144 | 174.181 | 6.9 | 0.13 |
| polarC18 | 276.144 | 162.6409 | 6.9 | 0.13 |
| polarC18 | 276.144 | 234.0065 | 6.9 | 0.13 |
| polarC18 | 276.144 | 65.9947 | 6.9 | 0.13 |
| polarC18 | 276.144 | 151.8608 | 6.9 | 0.13 |
| polarC18 | 276.144 | 104.557 | 6.9 | 0.13 |
| polarC18 | 276.144 | 55.8909 | 6.9 | 0.13 |
| polarC18 | 276.144 | 129.1794 | 6.9 | 0.13 |
| polarC18 | 276.144 | 131.7118 | 6.9 | 0.13 |
| polarC18 | 276.144 | 93.0753 | 6.9 | 0.13 |
| polarC18 | 276.144 | 62.9998 | 6.9 | 0.13 |
| polarC18 | 276.144 | 299.4278 | 6.9 | 0.13 |
| polarC18 | 276.144 | 239.5675 | 6.9 | 0.13 |
| polarC18 | 276.144 | 64.0191 | 6.9 | 0.13 |
| polarC18 | 276.144 | 62.9633 | 6.9 | 0.13 |
| polarC18 | 276.144 | 60.318 | 6.9 | 0.13 |
| polarC18 | 276.144 | 73.4452 | 6.9 | 0.13 |
| polarC18 | 276.144 | 89.8953 | 6.9 | 0.13 |
| polarC18 | 276.144 | 80.8775 | 6.9 | 0.13 |
| polarC18 | 276.144 | 60.0291 | 6.9 | 0.13 |
| polarC18 | 276.144 | 113.3853 | 6.9 | 0.13 |
| polarC18 | 276.144 | 55.0732 | 6.9 | 0.13 |
| polarC18 | 276.144 | 139.1318 | 6.9 | 0.13 |
| polarC18 | 276.144 | 84.2294 | 6.9 | 0.13 |
| polarC18 | 276.144 | 209.7746 | 6.9 | 0.13 |
| polarC18 | 276.144 | 120.4834 | 6.9 | 0.13 |
| polarC18 | 276.144 | 66.7703 | 6.9 | 0.13 |
| polarC18 | 276.144 | 152.6498 | 6.9 | 0.13 |
| polarC18 | 276.144 | 159.5143 | 6.9 | 0.13 |
| polarC18 | 276.144 | 127.2133 | 6.9 | 0.13 |
| polarC18 | 276.144 | 196.8105 | 6.9 | 0.13 |
| polarC18 | 276.144 | 181.7861 | 6.9 | 0.13 |
| polarC18 | 276.144 | 69.9538 | 6.9 | 0.13 |
| polarC18 | 276.144 | 210.4804 | 6.9 | 0.13 |

| | | | | |
|---|---|---|---|---|
| polarC18 | 276.144 | 116.3124 | 6.9 | 0.13 |
| polarC18 | 276.144 | 110.7829 | 6.9 | 0.13 |
| polarC18 | 276.144 | 59.1308 | 6.9 | 0.13 |
| polarC18 | 276.144 | 131.7417 | 6.9 | 0.13 |
| polarC18 | 276.144 | 120.8692 | 6.9 | 0.13 |
| polarC18 | 276.144 | 72.1185 | 6.9 | 0.13 |
| polarC18 | 276.144 | 148.4256 | 6.9 | 0.13 |
| polarC18 | 276.144 | 82.8788 | 6.9 | 0.13 |
| polarC18 | 276.144 | 98.7647 | 6.9 | 0.13 |
| polarC18 | 276.144 | 89.2137 | 6.9 | 0.13 |
| polarC18 | 276.144 | 66.0023 | 6.9 | 0.13 |
| polarC18 | 276.144 | 58.8148 | 6.9 | 0.13 |
| polarC18 | 276.144 | 54.2568 | 6.9 | 0.13 |
| polarC18 | 276.144 | 109.9981 | 6.9 | 0.13 |
| polarC18 | 276.144 | 57.7521 | 6.9 | 0.13 |
| polarC18 | 276.144 | 219.2343 | 6.9 | 0.13 |
| polarC18 | 276.144 | 59.3836 | 6.9 | 0.13 |
| polarC18 | 276.144 | 72.8971 | 6.9 | 0.13 |
| polarC18 | 276.144 | 147.8502 | 6.9 | 0.13 |
| polarC18 | 276.144 | 297.9933 | 6.9 | 0.13 |
| polarC18 | 276.144 | 176.9792 | 6.9 | 0.13 |
| polarC18 | 276.144 | 193.9319 | 6.9 | 0.13 |
| polarC18 | 276.144 | 59.3111 | 6.9 | 0.13 |
| polarC18 | 276.144 | 56.1594 | 6.9 | 0.13 |
| polarC18 | 276.144 | 184.3763 | 6.9 | 0.13 |
| polarC18 | 276.144 | 87.1998 | 6.9 | 0.13 |
| polarC18 | 276.144 | 120.0994 | 6.9 | 0.13 |
| polarC18 | 276.144 | 262.136 | 6.9 | 0.13 |
| polarC18 | 276.144 | 76.3137 | 6.9 | 0.13 |
| polarC18 | 276.144 | 50.3749 | 6.9 | 0.13 |
| polarC18 | 276.144 | 120.5558 | 6.9 | 0.13 |
| polarC18 | 276.144 | 289.2665 | 6.9 | 0.13 |
| polarC18 | 276.144 | 52.0536 | 6.9 | 0.13 |
| polarC18 | 276.144 | 64.7521 | 6.9 | 0.13 |
| polarC18 | 276.144 | 57.6913 | 6.9 | 0.13 |
| polarC18 | 276.144 | 60.3491 | 6.9 | 0.13 |
| polarC18 | 276.144 | 86.6223 | 6.9 | 0.13 |
| polarC18 | 276.144 | 93.2775 | 6.9 | 0.13 |
| polarC18 | 276.144 | 122.998 | 6.9 | 0.13 |
| polarC18 | 276.144 | 100.686 | 6.9 | 0.13 |
| polarC18 | 276.144 | 83.735 | 6.9 | 0.13 |
| polarC18 | 276.144 | 193.5054 | 6.9 | 0.13 |

| | | | | |
|---|---|---|---|---|
| polarC18 | 276.144 | 82.51 | 6.9 | 0.13 |
| polarC18 | 276.144 | 172.4516 | 6.9 | 0.13 |
| polarC18 | 276.144 | 61.7617 | 6.9 | 0.13 |
| polarC18 | 276.144 | 162.6716 | 6.9 | 0.13 |
| polarC18 | 276.144 | 97.8812 | 6.9 | 0.13 |
| polarC18 | 276.144 | 286.0634 | 6.9 | 0.13 |
| polarC18 | 276.144 | 146.2828 | 6.9 | 0.13 |
| polarC18 | 276.144 | 64.0972 | 6.9 | 0.13 |
| polarC18 | 276.144 | 217.1261 | 6.9 | 0.13 |
| polarC18 | 276.144 | 268.8859 | 6.9 | 0.13 |
| polarC18 | 276.144 | 190.0934 | 6.9 | 0.13 |
| polarC18 | 276.144 | 129.629 | 6.9 | 0.13 |
| polarC18 | 276.144 | 70.1636 | 6.9 | 0.13 |
| polarC18 | 276.144 | 66.0611 | 6.9 | 0.13 |
| polarC18 | 276.144 | 98.0084 | 6.9 | 0.13 |
| polarC18 | 276.144 | 181.8305 | 6.9 | 0.13 |
| polarC18 | 276.144 | 120.1936 | 6.9 | 0.13 |
| polarC18 | 276.144 | 141.8784 | 6.9 | 0.13 |
| polarC18 | 276.144 | 78.7012 | 6.9 | 0.13 |
| polarC18 | 276.144 | 173.6167 | 6.9 | 0.13 |
| polarC18 | 276.144 | 175.2331 | 6.9 | 0.13 |
| polarC18 | 276.144 | 111.8298 | 6.9 | 0.13 |
| polarC18 | 276.144 | 138.5025 | 6.9 | 0.13 |
| polarC18 | 276.144 | 70.5092 | 6.9 | 0.13 |
| polarC18 | 276.144 | 99.6813 | 6.9 | 0.13 |
| polarC18 | 276.144 | 54.6005 | 6.9 | 0.13 |
| polarC18 | 276.144 | 67.1621 | 6.9 | 0.13 |
| polarC18 | 276.144 | 59.6717 | 6.9 | 0.13 |
| polarC18 | 276.144 | 59.9921 | 6.9 | 0.13 |
| polarC18 | 276.144 | 59.8668 | 6.9 | 0.13 |
| polarC18 | 276.144 | 59.0277 | 6.9 | 0.13 |
| polarC18 | 276.144 | 63.3596 | 6.9 | 0.13 |
| polarC18 | 276.144 | 73.0854 | 6.9 | 0.13 |
| polarC18 | 276.144 | 88.001 | 6.9 | 0.13 |
| polarC18 | 276.144 | 88.1244 | 6.9 | 0.13 |
| polarC18 | 276.144 | 59.3082 | 6.9 | 0.13 |
| polarC18 | 276.144 | 74.0486 | 6.9 | 0.13 |
| polarC18 | 276.144 | 109.5433 | 6.9 | 0.13 |
| polarC18 | 276.144 | 146.4366 | 6.9 | 0.13 |
| polarC18 | 276.144 | 186.2513 | 6.9 | 0.13 |
| polarC18 | 276.144 | 90.2181 | 6.9 | 0.13 |
| polarC18 | 276.144 | 234.378 | 6.9 | 0.13 |

| | | | | |
|---|---|---|---|---|
| polarC18 | 276.144 | 63.8438 | 6.9 | 0.13 |
| polarC18 | 276.144 | 239.4428 | 6.9 | 0.13 |
| polarC18 | 276.144 | 209.4137 | 6.9 | 0.13 |
| polarC18 | 276.144 | 52.168 | 6.9 | 0.13 |
| polarC18 | 276.144 | 70.2065 | 6.9 | 0.13 |
| polarC18 | 276.144 | 70.3624 | 6.9 | 0.13 |
| polarC18 | 276.144 | 63.73 | 6.9 | 0.13 |
| polarC18 | 276.144 | 56.6392 | 6.9 | 0.13 |
| polarC18 | 276.144 | 234.6983 | 6.9 | 0.13 |
| polarC18 | 276.144 | 105.7053 | 6.9 | 0.13 |
| polarC18 | 276.144 | 74.775 | 6.9 | 0.13 |
| polarC18 | 276.144 | 54.0831 | 6.9 | 0.13 |
| polarC18 | 276.144 | 65.7474 | 6.9 | 0.13 |
| polarC18 | 276.144 | 58.9369 | 6.9 | 0.13 |
| polarC18 | 276.144 | 71.0594 | 6.9 | 0.13 |
| polarC18 | 276.144 | 69.2158 | 6.9 | 0.13 |
| polarC18 | 276.144 | 160.8373 | 6.9 | 0.13 |
| polarC18 | 276.144 | 71.9654 | 6.9 | 0.13 |
| polarC18 | 276.144 | 78.2299 | 6.9 | 0.13 |
| polarC18 | 276.144 | 50.4745 | 6.9 | 0.13 |
| polarC18 | 276.144 | 115.2641 | 6.9 | 0.13 |
| polarC18 | 276.144 | 89.2804 | 6.9 | 0.13 |
| polarC18 | 276.144 | 154.2301 | 6.9 | 0.13 |
| polarC18 | 276.144 | 57.6526 | 6.9 | 0.13 |
| polarC18 | 276.144 | 105.2465 | 6.9 | 0.13 |
| polarC18 | 276.144 | 57.7314 | 6.9 | 0.13 |
| polarC18 | 276.144 | 136.3966 | 6.9 | 0.13 |
| polarC18 | 276.144 | 73.1956 | 6.9 | 0.13 |
| polarC18 | 276.144 | 273.8249 | 6.9 | 0.13 |
| polarC18 | 276.144 | 84.2257 | 6.9 | 0.13 |
| polarC18 | 276.144 | 227.4118 | 6.9 | 0.13 |
| polarC18 | 276.144 | 199.5984 | 6.9 | 0.13 |
| polarC18 | 276.144 | 149.5676 | 6.9 | 0.13 |
| polarC18 | 276.144 | 150.7201 | 6.9 | 0.13 |
| polarC18 | 276.144 | 55.3179 | 6.9 | 0.13 |
| polarC18 | 276.144 | 144.2762 | 6.9 | 0.13 |
| polarC18 | 276.144 | 112.9476 | 6.9 | 0.13 |
| polarC18 | 276.144 | 67.0602 | 6.9 | 0.13 |
| polarC18 | 276.144 | 152.9677 | 6.9 | 0.13 |
| polarC18 | 276.144 | 67.1118 | 6.9 | 0.13 |
| polarC18 | 276.144 | 299.3318 | 6.9 | 0.13 |
| polarC18 | 276.144 | 69.5128 | 6.9 | 0.13 |

| | | | | |
|---|---|---|---|---|
| polarC18 | 276.144 | 73.2529 | 6.9 | 0.13 |
| polarC18 | 276.144 | 93.4632 | 6.9 | 0.13 |
| polarC18 | 276.144 | 153.4457 | 6.9 | 0.13 |
| polarC18 | 276.144 | 144.6225 | 6.9 | 0.13 |
| polarC18 | 276.144 | 207.1264 | 6.9 | 0.13 |
| polarC18 | 276.144 | 92.6329 | 6.9 | 0.13 |
| polarC18 | 276.144 | 56.424 | 6.9 | 0.13 |
| polarC18 | 276.144 | 82.2939 | 6.9 | 0.13 |
| polarC18 | 276.144 | 105.2593 | 6.9 | 0.13 |
| polarC18 | 276.144 | 60.8157 | 6.9 | 0.13 |
| polarC18 | 276.144 | 65.4496 | 6.9 | 0.13 |
| polarC18 | 276.144 | 121.6563 | 6.9 | 0.13 |
| polarC18 | 276.144 | 73.9415 | 6.8 | 0.13 |
| polarC18 | 276.144 | 95.7604 | 6.8 | 0.13 |
| polarC18 | 276.144 | 71.7621 | 6.8 | 0.13 |
| polarC18 | 276.144 | 257.1207 | 6.8 | 0.13 |
| polarC18 | 276.144 | 130.7892 | 6.8 | 0.13 |
| polarC18 | 276.144 | 229.0011 | 6.8 | 0.13 |
| polarC18 | 276.144 | 171.8989 | 6.8 | 0.13 |
| polarC18 | 276.144 | 53.7151 | 6.8 | 0.13 |
| polarC18 | 276.144 | 67.8314 | 6.8 | 0.13 |
| polarC18 | 276.144 | 57.673 | 6.8 | 0.13 |
| polarC18 | 276.144 | 87.5037 | 6.8 | 0.13 |
| polarC18 | 276.144 | 66.968 | 6.8 | 0.13 |
| polarC18 | 276.144 | 75.6222 | 6.8 | 0.13 |
| polarC18 | 276.144 | 126.2632 | 6.8 | 0.13 |
| polarC18 | 276.144 | 86.1018 | 6.8 | 0.13 |
| polarC18 | 276.144 | 100.0161 | 6.8 | 0.13 |
| polarC18 | 276.144 | 95.7243 | 6.8 | 0.13 |
| polarC18 | 276.144 | 89.1884 | 6.8 | 0.13 |
| polarC18 | 276.144 | 287.4207 | 6.8 | 0.13 |
| polarC18 | 276.144 | 121.0079 | 6.8 | 0.13 |
| polarC18 | 276.144 | 52.878 | 6.8 | 0.13 |
| polarC18 | 276.144 | 126.0929 | 6.8 | 0.13 |
| polarC18 | 276.144 | 58.368 | 6.8 | 0.13 |
| polarC18 | 276.144 | 288.4702 | 6.8 | 0.13 |
| polarC18 | 276.144 | 143.0214 | 6.8 | 0.13 |
| polarC18 | 276.144 | 70.7908 | 6.8 | 0.13 |
| polarC18 | 276.144 | 144.2561 | 6.8 | 0.13 |
| polarC18 | 276.144 | 56.5499 | 6.8 | 0.13 |
| polarC18 | 276.144 | 225.2009 | 6.8 | 0.13 |
| polarC18 | 276.144 | 189.7072 | 6.8 | 0.13 |

| | | | | |
|---|---|---|---|---|
| polarC18 | 276.144 | 107.9187 | 6.8 | 0.13 |
| polarC18 | 276.144 | 50.5639 | 6.8 | 0.13 |
| polarC18 | 276.144 | 67.8996 | 6.8 | 0.13 |
| polarC18 | 276.144 | 54.6665 | 6.8 | 0.13 |
| polarC18 | 276.144 | 96.4738 | 6.8 | 0.13 |
| polarC18 | 276.144 | 65.3788 | 6.8 | 0.13 |
| polarC18 | 276.144 | 132.0481 | 6.8 | 0.13 |
| polarC18 | 276.144 | 160.085 | 6.8 | 0.13 |
| polarC18 | 276.144 | 55.7964 | 6.8 | 0.13 |
| polarC18 | 276.144 | 108.6915 | 6.8 | 0.13 |
| polarC18 | 276.144 | 77.3469 | 6.8 | 0.13 |
| polarC18 | 276.144 | 51.8945 | 6.8 | 0.13 |
| polarC18 | 276.144 | 56.0567 | 6.8 | 0.13 |
| polarC18 | 276.144 | 50.1908 | 6.8 | 0.13 |
| polarC18 | 276.144 | 64.5655 | 6.8 | 0.13 |
| polarC18 | 276.144 | 96.9386 | 6.8 | 0.13 |
| polarC18 | 276.144 | 163.2583 | 6.8 | 0.13 |
| polarC18 | 276.144 | 62.8218 | 6.8 | 0.13 |
| polarC18 | 276.144 | 92.2437 | 6.8 | 0.13 |
| polarC18 | 276.144 | 117.4456 | 6.8 | 0.13 |
| polarC18 | 276.144 | 124.0611 | 6.8 | 0.13 |
| polarC18 | 276.144 | 177.2933 | 6.8 | 0.13 |
| polarC18 | 276.144 | 93.9133 | 6.8 | 0.13 |
| polarC18 | 276.144 | 238.9138 | 6.8 | 0.13 |
| polarC18 | 276.144 | 123.296 | 6.8 | 0.13 |
| polarC18 | 276.144 | 57.146 | 6.8 | 0.13 |
| polarC18 | 276.144 | 51.3411 | 6.8 | 0.13 |
| polarC18 | 276.144 | 239.5995 | 6.8 | 0.13 |
| polarC18 | 276.144 | 138.0343 | 6.8 | 0.13 |
| polarC18 | 276.144 | 258.6271 | 6.8 | 0.13 |
| polarC18 | 276.144 | 75.7059 | 6.8 | 0.13 |
| polarC18 | 276.144 | 68.281 | 6.8 | 0.13 |
| polarC18 | 276.144 | 124.0921 | 6.8 | 0.13 |
| polarC18 | 276.144 | 249.5732 | 6.8 | 0.13 |
| polarC18 | 276.144 | 67.8368 | 6.8 | 0.13 |
| polarC18 | 276.144 | 168.0409 | 6.8 | 0.13 |
| polarC18 | 276.144 | 105.0916 | 6.8 | 0.13 |
| polarC18 | 276.144 | 225.5792 | 6.8 | 0.13 |
| polarC18 | 276.144 | 65.1928 | 6.8 | 0.13 |
| polarC18 | 276.144 | 63.9577 | 6.8 | 0.13 |
| polarC18 | 276.144 | 126.0014 | 6.8 | 0.13 |
| polarC18 | 276.144 | 97.039 | 6.8 | 0.13 |

| | | | | |
|---|---|---|---|---|
| polarC18 | 276.144 | 172.3731 | 6.8 | 0.13 |
| polarC18 | 276.144 | 133.1231 | 6.8 | 0.13 |
| polarC18 | 276.144 | 227.8422 | 6.8 | 0.13 |
| polarC18 | 276.144 | 50.1041 | 6.8 | 0.13 |
| polarC18 | 276.144 | 198.194 | 6.8 | 0.13 |
| polarC18 | 276.144 | 257.8822 | 6.8 | 0.13 |
| polarC18 | 276.144 | 64.3551 | 6.8 | 0.13 |
| polarC18 | 276.144 | 78.1268 | 6.8 | 0.13 |
| polarC18 | 276.144 | 94.4355 | 6.8 | 0.13 |
| polarC18 | 276.144 | 54.7987 | 6.8 | 0.13 |
| polarC18 | 276.144 | 164.9755 | 6.8 | 0.13 |
| polarC18 | 276.144 | 70.4083 | 6.8 | 0.13 |
| polarC18 | 276.144 | 51.5791 | 6.8 | 0.13 |
| polarC18 | 276.144 | 68.5663 | 6.8 | 0.13 |
| polarC18 | 276.144 | 122.8312 | 6.8 | 0.13 |
| polarC18 | 276.144 | 56.3608 | 6.8 | 0.13 |
| polarC18 | 276.144 | 93.3185 | 6.8 | 0.13 |
| polarC18 | 276.144 | 121.118 | 6.8 | 0.13 |
| polarC18 | 276.144 | 60.2896 | 6.8 | 0.13 |
| polarC18 | 276.144 | 122.8505 | 6.8 | 0.13 |
| polarC18 | 276.144 | 215.7912 | 6.8 | 0.13 |
| polarC18 | 276.144 | 100.6339 | 6.8 | 0.13 |
| polarC18 | 276.144 | 50.7092 | 6.8 | 0.13 |
| polarC18 | 276.144 | 52.51 | 6.8 | 0.13 |
| polarC18 | 276.144 | 51.969 | 6.8 | 0.13 |
| polarC18 | 276.144 | 83.0725 | 6.8 | 0.13 |
| polarC18 | 276.144 | 118.8822 | 6.8 | 0.13 |
| polarC18 | 276.144 | 64.2806 | 6.8 | 0.13 |
| polarC18 | 276.144 | 51.8356 | 6.8 | 0.13 |
| polarC18 | 276.144 | 149.3837 | 6.8 | 0.13 |
| polarC18 | 276.144 | 98.229 | 6.8 | 0.13 |
| polarC18 | 276.144 | 59.5919 | 6.8 | 0.13 |
| polarC18 | 276.144 | 161.6948 | 6.8 | 0.13 |
| polarC18 | 276.144 | 55.1119 | 6.8 | 0.13 |
| polarC18 | 276.144 | 243.6692 | 6.8 | 0.13 |
| polarC18 | 276.144 | 160.602 | 6.8 | 0.13 |
| polarC18 | 276.144 | 123.2831 | 6.8 | 0.13 |
| polarC18 | 276.144 | 284.4254 | 6.8 | 0.13 |
| polarC18 | 276.144 | 65.4103 | 6.8 | 0.13 |
| polarC18 | 276.144 | 120.7573 | 6.8 | 0.13 |
| polarC18 | 276.144 | 54.3484 | 6.8 | 0.13 |
| polarC18 | 276.144 | 187.7832 | 6.8 | 0.13 |

| | | | | |
|---|---|---|---|---|
| polarC18 | 276.144 | 207.6031 | 6.8 | 0.13 |
| polarC18 | 276.144 | 241.3076 | 6.8 | 0.13 |
| polarC18 | 276.144 | 82.4538 | 6.8 | 0.13 |
| polarC18 | 276.144 | 56.8719 | 6.8 | 0.13 |
| polarC18 | 276.144 | 211.5756 | 6.8 | 0.13 |
| polarC18 | 276.144 | 115.5498 | 6.8 | 0.13 |
| polarC18 | 276.144 | 104.5809 | 6.8 | 0.13 |
| polarC18 | 276.144 | 51.1153 | 6.8 | 0.13 |
| polarC18 | 276.144 | 80.0031 | 6.8 | 0.13 |
| polarC18 | 276.144 | 86.5746 | 6.8 | 0.13 |
| polarC18 | 276.144 | 58.9175 | 6.8 | 0.13 |
| polarC18 | 276.144 | 105.6429 | 6.8 | 0.13 |
| polarC18 | 276.144 | 65.5914 | 6.8 | 0.13 |
| polarC18 | 276.144 | 92.1562 | 6.8 | 0.13 |
| polarC18 | 276.144 | 51.8151 | 6.8 | 0.13 |
| polarC18 | 276.144 | 61.0535 | 6.8 | 0.13 |
| polarC18 | 276.144 | 52.9875 | 6.8 | 0.13 |
| polarC18 | 276.144 | 54.1619 | 6.8 | 0.13 |
| polarC18 | 276.144 | 124.7858 | 6.8 | 0.13 |
| polarC18 | 276.144 | 64.3036 | 6.8 | 0.13 |
| polarC18 | 276.144 | 93.0095 | 6.8 | 0.13 |
| polarC18 | 276.144 | 72.9509 | 6.8 | 0.13 |
| polarC18 | 276.144 | 97.0978 | 6.8 | 0.13 |
| polarC18 | 276.144 | 149.3334 | 6.8 | 0.13 |
| polarC18 | 276.144 | 51.3636 | 6.8 | 0.13 |
| polarC18 | 276.144 | 67.3202 | 6.8 | 0.13 |
| polarC18 | 276.144 | 94.9339 | 6.8 | 0.13 |
| polarC18 | 276.144 | 52.0667 | 6.8 | 0.13 |
| polarC18 | 276.144 | 61.4182 | 6.8 | 0.13 |
| polarC18 | 276.144 | 71.0962 | 6.8 | 0.13 |
| polarC18 | 276.144 | 84.4614 | 6.8 | 0.13 |
| polarC18 | 276.144 | 55.2584 | 6.8 | 0.13 |
| polarC18 | 276.144 | 85.2329 | 6.8 | 0.13 |
| polarC18 | 276.144 | 57.3485 | 6.8 | 0.13 |
| polarC18 | 276.144 | 280.8876 | 6.8 | 0.13 |
| polarC18 | 276.144 | 51.5316 | 6.8 | 0.13 |
| polarC18 | 276.144 | 71.5054 | 6.8 | 0.13 |
| polarC18 | 276.144 | 205.9221 | 6.8 | 0.13 |
| polarC18 | 276.144 | 176.6547 | 6.8 | 0.13 |
| polarC18 | 276.144 | 142.7099 | 6.8 | 0.13 |
| polarC18 | 276.144 | 273.7417 | 6.8 | 0.13 |
| polarC18 | 276.144 | 156.8409 | 6.8 | 0.13 |

| | | | | |
|---|---|---|---|---|
| polarC18 | 276.144 | 90.4756 | 6.8 | 0.13 |
| polarC18 | 276.144 | 56.2523 | 6.8 | 0.13 |
| polarC18 | 276.144 | 57.7572 | 6.8 | 0.13 |
| polarC18 | 276.144 | 57.3782 | 6.8 | 0.13 |
| polarC18 | 276.144 | 127.1671 | 6.8 | 0.13 |
| polarC18 | 276.144 | 50.9887 | 6.8 | 0.13 |
| polarC18 | 276.144 | 237.6475 | 6.8 | 0.13 |
| polarC18 | 276.144 | 72.0618 | 6.8 | 0.13 |
| polarC18 | 276.144 | 74.9136 | 6.8 | 0.13 |
| polarC18 | 276.144 | 186.894 | 6.8 | 0.13 |
| polarC18 | 276.144 | 62.4785 | 6.8 | 0.13 |
| polarC18 | 276.144 | 120.3686 | 6.8 | 0.13 |
| polarC18 | 276.144 | 114.9229 | 6.8 | 0.13 |
| polarC18 | 276.144 | 54.9451 | 6.8 | 0.13 |
| polarC18 | 276.144 | 50.4963 | 6.8 | 0.13 |
| polarC18 | 276.144 | 92.1436 | 6.8 | 0.13 |
| polarC18 | 276.144 | 83.9586 | 6.8 | 0.13 |
| polarC18 | 276.144 | 264.5692 | 6.8 | 0.13 |
| polarC18 | 276.144 | 75.2297 | 6.8 | 0.13 |
| polarC18 | 276.144 | 196.3136 | 6.8 | 0.13 |
| polarC18 | 276.144 | 90.6093 | 6.8 | 0.13 |
| polarC18 | 276.144 | 168.6888 | 6.8 | 0.13 |
| polarC18 | 276.144 | 112.8498 | 6.8 | 0.13 |
| polarC18 | 276.144 | 272.0151 | 6.8 | 0.13 |
| polarC18 | 276.144 | 52.111 | 6.8 | 0.13 |
| polarC18 | 276.144 | 116.664 | 6.8 | 0.13 |
| polarC18 | 276.144 | 66.8209 | 6.8 | 0.13 |
| polarC18 | 276.144 | 52.9768 | 6.8 | 0.13 |
| polarC18 | 276.144 | 54.2418 | 6.8 | 0.13 |
| polarC18 | 276.144 | 74.476 | 6.8 | 0.13 |
| polarC18 | 276.144 | 106.1822 | 6.8 | 0.13 |
| polarC18 | 276.144 | 55.3453 | 6.8 | 0.13 |
| polarC18 | 276.144 | 132.0236 | 6.8 | 0.13 |
| polarC18 | 276.144 | 82.223 | 6.8 | 0.13 |
| polarC18 | 276.144 | 61.0627 | 6.8 | 0.13 |
| polarC18 | 276.144 | 93.2295 | 6.8 | 0.13 |
| polarC18 | 276.144 | 52.042 | 6.8 | 0.13 |
| polarC18 | 276.144 | 124.195 | 6.8 | 0.13 |
| polarC18 | 276.144 | 90.1262 | 6.8 | 0.13 |
| polarC18 | 276.144 | 223.9984 | 6.8 | 0.13 |
| polarC18 | 276.144 | 51.2449 | 6.8 | 0.13 |
| polarC18 | 276.144 | 183.4277 | 6.8 | 0.13 |

| | | | | |
|---|---|---|---|---|
| polarC18 | 276.144 | 188.4422 | 6.8 | 0.13 |
| polarC18 | 276.144 | 81.376 | 6.8 | 0.13 |
| polarC18 | 276.144 | 211.9162 | 6.8 | 0.13 |
| polarC18 | 276.144 | 56.2925 | 6.8 | 0.13 |
| polarC18 | 276.144 | 91.6243 | 6.8 | 0.13 |
| polarC18 | 276.144 | 103.5241 | 6.8 | 0.13 |
| polarC18 | 276.144 | 140.7225 | 6.8 | 0.13 |
| polarC18 | 276.144 | 71.2435 | 6.8 | 0.13 |
| polarC18 | 276.144 | 166.3912 | 6.8 | 0.13 |
| polarC18 | 276.144 | 122.6207 | 6.8 | 0.13 |
| polarC18 | 276.144 | 58.8422 | 6.8 | 0.13 |
| polarC18 | 276.144 | 128.6382 | 6.8 | 0.13 |
| polarC18 | 276.144 | 252.8036 | 6.8 | 0.13 |
| polarC18 | 276.144 | 152.1097 | 6.8 | 0.13 |
| polarC18 | 276.144 | 180.1904 | 6.8 | 0.13 |
| polarC18 | 276.144 | 107.9035 | 6.8 | 0.13 |
| polarC18 | 276.144 | 60.3828 | 6.8 | 0.13 |
| polarC18 | 276.144 | 89.7624 | 6.8 | 0.13 |
| polarC18 | 276.144 | 82.6141 | 6.8 | 0.13 |
| polarC18 | 276.144 | 128.7553 | 6.8 | 0.13 |
| polarC18 | 276.144 | 113.1482 | 6.8 | 0.13 |
| polarC18 | 276.144 | 122.0679 | 6.8 | 0.13 |
| polarC18 | 276.144 | 86.2757 | 6.8 | 0.13 |
| polarC18 | 276.144 | 297.857 | 6.8 | 0.13 |
| polarC18 | 276.144 | 143.7918 | 6.8 | 0.13 |
| polarC18 | 276.144 | 63.3358 | 6.8 | 0.13 |
| polarC18 | 276.144 | 77.9316 | 6.8 | 0.13 |
| polarC18 | 276.144 | 159.7775 | 6.8 | 0.13 |
| polarC18 | 276.144 | 109.5098 | 6.8 | 0.13 |
| polarC18 | 276.144 | 57.3702 | 6.8 | 0.13 |
| polarC18 | 276.144 | 123.4623 | 6.8 | 0.13 |
| polarC18 | 276.144 | 89.2245 | 6.8 | 0.13 |
| polarC18 | 276.144 | 60.5647 | 6.8 | 0.13 |
| polarC18 | 276.144 | 153.6111 | 6.8 | 0.13 |
| polarC18 | 276.144 | 128.1077 | 6.8 | 0.13 |
| polarC18 | 276.144 | 80.7434 | 6.8 | 0.13 |
| polarC18 | 276.144 | 285.377 | 6.8 | 0.13 |
| polarC18 | 276.144 | 87.6355 | 6.8 | 0.13 |
| polarC18 | 276.144 | 52.3688 | 6.8 | 0.13 |
| polarC18 | 276.144 | 72.2222 | 6.8 | 0.13 |
| polarC18 | 276.144 | 93.4246 | 6.8 | 0.13 |
| polarC18 | 276.144 | 75.8525 | 6.8 | 0.13 |

| | | | | |
|---|---|---|---|---|
| polarC18 | 276.144 | 98.4081 | 6.8 | 0.13 |
| polarC18 | 276.144 | 138.7773 | 6.8 | 0.13 |
| polarC18 | 276.144 | 239.137 | 6.8 | 0.13 |
| polarC18 | 276.144 | 89.1803 | 6.8 | 0.13 |
| polarC18 | 276.144 | 131.8544 | 6.8 | 0.13 |
| polarC18 | 276.144 | 91.7612 | 6.8 | 0.13 |
| polarC18 | 276.144 | 142.2274 | 6.8 | 0.13 |
| polarC18 | 276.144 | 143.7346 | 6.8 | 0.13 |
| polarC18 | 276.144 | 81.0772 | 6.8 | 0.13 |
| polarC18 | 276.144 | 79.6648 | 6.8 | 0.13 |
| polarC18 | 276.144 | 72.2874 | 6.8 | 0.13 |
| polarC18 | 276.144 | 125.5949 | 6.8 | 0.13 |
| polarC18 | 276.144 | 58.7679 | 6.8 | 0.13 |
| polarC18 | 276.144 | 60.4973 | 6.8 | 0.13 |
| polarC18 | 276.144 | 111.3027 | 6.8 | 0.13 |
| polarC18 | 276.144 | 136.1822 | 6.8 | 0.13 |
| polarC18 | 276.144 | 95.2848 | 6.8 | 0.13 |
| polarC18 | 276.144 | 60.0356 | 6.8 | 0.13 |
| polarC18 | 276.144 | 249.4057 | 6.8 | 0.13 |
| polarC18 | 276.144 | 65.484 | 6.8 | 0.13 |
| polarC18 | 276.144 | 179.6691 | 6.8 | 0.13 |
| polarC18 | 276.144 | 86.0856 | 6.8 | 0.13 |
| polarC18 | 276.144 | 61.1403 | 6.8 | 0.13 |
| polarC18 | 276.144 | 60.0422 | 6.8 | 0.13 |
| polarC18 | 276.144 | 156.4062 | 6.8 | 0.13 |
| polarC18 | 276.144 | 136.2233 | 6.8 | 0.13 |
| polarC18 | 276.144 | 57.3629 | 6.8 | 0.13 |
| polarC18 | 276.144 | 68.7215 | 6.8 | 0.13 |
| polarC18 | 276.144 | 248.6884 | 6.8 | 0.13 |
| polarC18 | 276.144 | 52.4883 | 6.8 | 0.13 |
| polarC18 | 276.144 | 59.1035 | 6.8 | 0.13 |
| polarC18 | 276.144 | 54.3187 | 6.8 | 0.13 |
| polarC18 | 276.144 | 74.8309 | 6.8 | 0.13 |
| polarC18 | 276.144 | 214.4808 | 6.8 | 0.13 |
| polarC18 | 276.144 | 121.7562 | 6.8 | 0.13 |
| polarC18 | 276.144 | 71.78 | 6.8 | 0.13 |
| polarC18 | 276.144 | 110.0321 | 6.8 | 0.13 |
| polarC18 | 276.144 | 80.4344 | 6.8 | 0.13 |
| polarC18 | 276.144 | 232.4771 | 6.8 | 0.13 |
| polarC18 | 276.144 | 199.9964 | 6.8 | 0.13 |
| polarC18 | 276.144 | 158.7162 | 6.8 | 0.13 |
| polarC18 | 276.144 | 241.3762 | 6.8 | 0.13 |

| | | | | |
|---|---|---|---|---|
| polarC18 | 276.144 | 58.5995 | 6.8 | 0.13 |
| polarC18 | 276.144 | 260.6301 | 6.8 | 0.13 |
| polarC18 | 276.144 | 85.9847 | 6.8 | 0.13 |
| polarC18 | 276.144 | 112.1834 | 6.8 | 0.13 |
| polarC18 | 276.144 | 50.0541 | 6.8 | 0.13 |
| polarC18 | 276.144 | 51.1784 | 6.8 | 0.13 |
| polarC18 | 276.144 | 125.5012 | 6.8 | 0.13 |
| polarC18 | 276.144 | 58.7112 | 6.8 | 0.13 |
| polarC18 | 276.144 | 150.221 | 6.8 | 0.13 |
| polarC18 | 276.144 | 56.515 | 6.8 | 0.13 |
| polarC18 | 276.144 | 61.3075 | 6.8 | 0.13 |
| polarC18 | 276.144 | 231.5022 | 6.8 | 0.13 |
| polarC18 | 276.144 | 52.0486 | 6.8 | 0.13 |
| polarC18 | 276.144 | 58.6422 | 6.8 | 0.13 |
| polarC18 | 276.144 | 87.5775 | 6.8 | 0.13 |
| polarC18 | 276.144 | 269.1394 | 6.8 | 0.13 |
| polarC18 | 276.144 | 82.4833 | 6.8 | 0.13 |
| polarC18 | 276.144 | 81.8406 | 6.8 | 0.13 |
| polarC18 | 276.144 | 62.911 | 6.8 | 0.13 |
| polarC18 | 276.144 | 61.7015 | 6.8 | 0.13 |
| polarC18 | 276.144 | 141.5158 | 6.8 | 0.13 |
| polarC18 | 276.144 | 91.9516 | 6.8 | 0.13 |
| polarC18 | 276.144 | 118.6765 | 6.8 | 0.13 |
| polarC18 | 276.144 | 87.9955 | 6.8 | 0.13 |
| polarC18 | 276.144 | 285.7939 | 6.8 | 0.13 |
| polarC18 | 276.144 | 262.8879 | 6.8 | 0.13 |
| polarC18 | 276.144 | 172.5537 | 6.8 | 0.13 |
| polarC18 | 276.144 | 65.582 | 6.8 | 0.13 |
| polarC18 | 276.144 | 136.527 | 6.8 | 0.13 |
| polarC18 | 276.144 | 176.951 | 6.8 | 0.13 |
| polarC18 | 276.144 | 64.5576 | 6.8 | 0.13 |
| polarC18 | 276.144 | 53.74 | 6.8 | 0.13 |
| polarC18 | 276.144 | 61.1051 | 6.8 | 0.13 |
| polarC18 | 276.144 | 72.2486 | 6.8 | 0.13 |
| polarC18 | 276.144 | 138.3343 | 6.8 | 0.13 |
| polarC18 | 276.144 | 78.6643 | 6.8 | 0.13 |
| polarC18 | 276.144 | 106.757 | 6.8 | 0.13 |
| polarC18 | 276.144 | 160.6922 | 6.8 | 0.13 |
| polarC18 | 276.144 | 98.4807 | 6.7 | 0.13 |
| polarC18 | 276.144 | 78.2895 | 6.7 | 0.13 |
| polarC18 | 276.144 | 52.3993 | 6.7 | 0.13 |
| polarC18 | 276.144 | 191.6409 | 6.7 | 0.13 |

| | | | | |
|---|---|---|---|---|
| polarC18 | 276.144 | 71.0493 | 6.7 | 0.13 |
| polarC18 | 276.144 | 53.7664 | 6.7 | 0.13 |
| polarC18 | 276.144 | 119.3956 | 6.7 | 0.13 |
| polarC18 | 276.144 | 98.5852 | 6.7 | 0.13 |
| polarC18 | 276.144 | 52.1661 | 6.7 | 0.13 |
| polarC18 | 276.144 | 292.6258 | 6.7 | 0.13 |
| polarC18 | 276.144 | 110.5439 | 6.7 | 0.13 |
| polarC18 | 276.144 | 76.8854 | 6.7 | 0.13 |
| polarC18 | 276.144 | 58.7609 | 6.7 | 0.13 |
| polarC18 | 276.144 | 66.8808 | 6.7 | 0.13 |
| polarC18 | 276.144 | 66.6835 | 6.7 | 0.13 |
| polarC18 | 276.144 | 69.3801 | 6.7 | 0.13 |
| polarC18 | 276.144 | 198.15 | 6.7 | 0.13 |
| polarC18 | 276.144 | 72.6366 | 6.7 | 0.13 |
| polarC18 | 276.144 | 179.7327 | 6.7 | 0.13 |
| polarC18 | 276.144 | 79.9843 | 6.7 | 0.13 |
| polarC18 | 276.144 | 57.5989 | 6.7 | 0.13 |
| polarC18 | 276.144 | 78.3202 | 6.7 | 0.13 |
| polarC18 | 276.144 | 61.7381 | 6.7 | 0.13 |
| polarC18 | 276.144 | 104.1551 | 6.7 | 0.13 |
| polarC18 | 276.144 | 66.1409 | 6.7 | 0.13 |
| polarC18 | 276.144 | 72.3938 | 6.7 | 0.13 |
| polarC18 | 276.144 | 119.6878 | 6.7 | 0.13 |
| polarC18 | 276.144 | 153.5348 | 6.7 | 0.13 |
| polarC18 | 276.144 | 113.352 | 6.7 | 0.13 |
| polarC18 | 276.144 | 53.5071 | 6.7 | 0.13 |
| polarC18 | 276.144 | 218.9763 | 6.7 | 0.13 |
| polarC18 | 276.144 | 66.1943 | 6.7 | 0.13 |
| polarC18 | 276.144 | 84.2404 | 6.7 | 0.13 |
| polarC18 | 276.144 | 110.1653 | 6.7 | 0.13 |
| polarC18 | 276.144 | 184.1212 | 6.7 | 0.13 |
| polarC18 | 276.144 | 85.8548 | 6.7 | 0.13 |
| polarC18 | 276.144 | 150.512 | 6.7 | 0.13 |
| polarC18 | 276.144 | 82.009 | 6.7 | 0.13 |
| polarC18 | 276.144 | 215.7061 | 6.7 | 0.13 |
| polarC18 | 276.144 | 284.1998 | 6.7 | 0.13 |
| polarC18 | 276.144 | 66.9631 | 6.7 | 0.13 |
| polarC18 | 276.144 | 234.6466 | 6.7 | 0.13 |
| polarC18 | 276.144 | 177.6303 | 6.7 | 0.13 |
| polarC18 | 276.144 | 62.5187 | 6.7 | 0.13 |
| polarC18 | 276.144 | 69.1686 | 6.7 | 0.13 |
| polarC18 | 276.144 | 131.2375 | 6.7 | 0.13 |

| | | | | |
|---|---|---|---|---|
| polarC18 | 276.144 | 59.5429 | 6.7 | 0.13 |
| polarC18 | 276.144 | 102.833 | 6.7 | 0.13 |
| polarC18 | 276.144 | 204.7002 | 6.7 | 0.13 |
| polarC18 | 276.144 | 57.1316 | 6.7 | 0.13 |
| polarC18 | 276.144 | 164.1922 | 6.7 | 0.13 |
| polarC18 | 276.144 | 62.1701 | 6.7 | 0.13 |
| polarC18 | 276.144 | 272.9454 | 6.7 | 0.13 |
| polarC18 | 276.144 | 69.2255 | 6.7 | 0.13 |
| polarC18 | 276.144 | 78.5122 | 6.7 | 0.13 |
| polarC18 | 276.144 | 160.6779 | 6.7 | 0.13 |
| polarC18 | 276.144 | 65.0609 | 6.7 | 0.13 |
| polarC18 | 276.144 | 148.6167 | 6.7 | 0.13 |
| polarC18 | 276.144 | 100.9441 | 6.7 | 0.13 |
| polarC18 | 276.144 | 54.6593 | 6.7 | 0.13 |
| polarC18 | 276.144 | 77.0596 | 6.7 | 0.13 |
| polarC18 | 276.144 | 160.9385 | 6.7 | 0.13 |
| polarC18 | 276.144 | 93.7046 | 6.7 | 0.13 |
| polarC18 | 276.144 | 229.7572 | 6.7 | 0.13 |
| polarC18 | 276.144 | 82.2475 | 6.7 | 0.13 |
| polarC18 | 276.144 | 112.6743 | 6.7 | 0.13 |
| polarC18 | 276.144 | 53.9764 | 6.7 | 0.13 |
| polarC18 | 276.144 | 56.6669 | 6.7 | 0.13 |
| polarC18 | 276.144 | 84.8918 | 6.7 | 0.13 |
| polarC18 | 276.144 | 146.9178 | 6.7 | 0.13 |
| polarC18 | 276.144 | 66.0863 | 6.7 | 0.13 |
| polarC18 | 276.144 | 77.868 | 6.7 | 0.13 |
| polarC18 | 276.144 | 126.1493 | 6.7 | 0.13 |
| polarC18 | 276.144 | 50.321 | 6.7 | 0.13 |
| polarC18 | 276.144 | 174.4232 | 6.7 | 0.13 |
| polarC18 | 276.144 | 134.6804 | 6.7 | 0.13 |
| polarC18 | 276.144 | 70.8236 | 6.7 | 0.13 |
| polarC18 | 276.144 | 187.8572 | 6.7 | 0.13 |
| polarC18 | 276.144 | 57.6495 | 6.7 | 0.13 |
| polarC18 | 276.144 | 89.8606 | 6.7 | 0.13 |
| polarC18 | 276.144 | 64.9624 | 6.7 | 0.13 |
| polarC18 | 276.144 | 212.1693 | 6.7 | 0.13 |
| polarC18 | 276.144 | 84.1854 | 6.7 | 0.13 |
| polarC18 | 276.144 | 206.2197 | 6.7 | 0.13 |
| polarC18 | 276.144 | 57.865 | 6.7 | 0.13 |
| polarC18 | 276.144 | 90.9905 | 6.7 | 0.13 |
| polarC18 | 276.144 | 191.9178 | 6.7 | 0.13 |
| polarC18 | 276.144 | 181.2558 | 6.7 | 0.13 |

| | | | | |
|---|---|---|---|---|
| polarC18 | 276.144 | 87.5192 | 6.7 | 0.13 |
| polarC18 | 276.144 | 186.5663 | 6.7 | 0.13 |
| polarC18 | 276.144 | 95.2179 | 6.7 | 0.13 |
| polarC18 | 276.144 | 55.3607 | 6.7 | 0.13 |
| polarC18 | 276.144 | 80.5326 | 6.7 | 0.13 |
| polarC18 | 276.144 | 119.8896 | 6.7 | 0.13 |
| polarC18 | 276.144 | 54.663 | 6.7 | 0.13 |
| polarC18 | 276.144 | 71.3683 | 6.7 | 0.13 |
| polarC18 | 276.144 | 208.6427 | 6.7 | 0.13 |
| polarC18 | 276.144 | 146.0036 | 6.7 | 0.13 |
| polarC18 | 276.144 | 220.0792 | 6.7 | 0.13 |
| polarC18 | 276.144 | 249.337 | 6.7 | 0.13 |
| polarC18 | 276.144 | 54.8137 | 6.7 | 0.13 |
| polarC18 | 276.144 | 112.5214 | 6.7 | 0.13 |
| polarC18 | 276.144 | 58.6648 | 6.7 | 0.13 |
| polarC18 | 276.144 | 158.966 | 6.7 | 0.13 |
| polarC18 | 276.144 | 85.6691 | 6.7 | 0.13 |
| polarC18 | 276.144 | 67.4649 | 6.7 | 0.13 |
| polarC18 | 276.144 | 65.1379 | 6.7 | 0.13 |
| polarC18 | 276.144 | 69.4854 | 6.7 | 0.13 |
| polarC18 | 276.144 | 80.4143 | 6.7 | 0.13 |
| polarC18 | 276.144 | 56.9077 | 6.7 | 0.13 |
| polarC18 | 276.144 | 176.6366 | 6.7 | 0.13 |
| polarC18 | 276.144 | 107.9108 | 6.7 | 0.13 |
| polarC18 | 276.144 | 171.384 | 6.7 | 0.13 |
| polarC18 | 276.144 | 72.7682 | 6.7 | 0.13 |
| polarC18 | 276.144 | 106.169 | 6.7 | 0.13 |
| polarC18 | 276.144 | 113.161 | 6.7 | 0.13 |
| polarC18 | 276.144 | 133.9865 | 6.7 | 0.13 |
| polarC18 | 276.144 | 63.1469 | 6.7 | 0.13 |
| polarC18 | 276.144 | 104.537 | 6.7 | 0.13 |
| polarC18 | 276.144 | 77.5583 | 6.7 | 0.13 |
| polarC18 | 276.144 | 169.0712 | 6.7 | 0.13 |
| polarC18 | 276.144 | 61.7293 | 6.7 | 0.13 |
| polarC18 | 276.144 | 158.2835 | 6.7 | 0.13 |
| polarC18 | 276.144 | 93.2352 | 6.7 | 0.13 |
| polarC18 | 276.144 | 52.1391 | 6.7 | 0.13 |
| polarC18 | 276.144 | 62.025 | 6.7 | 0.13 |
| polarC18 | 276.144 | 60.5274 | 6.7 | 0.13 |
| polarC18 | 276.144 | 73.7594 | 6.7 | 0.13 |
| polarC18 | 276.144 | 137.1874 | 6.7 | 0.13 |
| polarC18 | 276.144 | 86.4666 | 6.7 | 0.13 |

| | | | | |
|---|---|---|---|---|
| polarC18 | 276.144 | 60.6444 | 6.7 | 0.13 |
| polarC18 | 276.144 | 234.2536 | 6.7 | 0.13 |
| polarC18 | 276.144 | 166.2371 | 6.7 | 0.13 |
| polarC18 | 276.144 | 186.0259 | 6.7 | 0.13 |
| polarC18 | 276.144 | 61.8831 | 6.7 | 0.13 |
| polarC18 | 276.144 | 51.8756 | 6.7 | 0.13 |
| polarC18 | 276.144 | 166.1696 | 6.7 | 0.13 |
| polarC18 | 276.144 | 70.7865 | 6.7 | 0.13 |
| polarC18 | 276.144 | 110.6014 | 6.7 | 0.13 |
| polarC18 | 276.144 | 83.8684 | 6.7 | 0.13 |
| polarC18 | 276.144 | 265.6369 | 6.7 | 0.13 |
| polarC18 | 276.144 | 52.6011 | 6.7 | 0.13 |
| polarC18 | 276.144 | 111.7704 | 6.7 | 0.13 |
| polarC18 | 276.144 | 102.2582 | 6.7 | 0.13 |
| polarC18 | 276.144 | 128.7708 | 6.7 | 0.13 |
| polarC18 | 276.144 | 140.7526 | 6.7 | 0.13 |
| polarC18 | 276.144 | 65.229 | 6.7 | 0.13 |
| polarC18 | 276.144 | 279.665 | 6.7 | 0.13 |
| polarC18 | 276.144 | 122.8757 | 6.7 | 0.13 |
| polarC18 | 276.144 | 265.8222 | 6.7 | 0.13 |
| polarC18 | 276.144 | 67.651 | 6.7 | 0.13 |
| polarC18 | 276.144 | 69.3073 | 6.7 | 0.13 |
| polarC18 | 276.144 | 92.9611 | 6.7 | 0.13 |
| polarC18 | 276.144 | 61.5085 | 6.7 | 0.13 |
| polarC18 | 276.144 | 79.6152 | 6.7 | 0.13 |
| polarC18 | 276.144 | 64.2529 | 6.7 | 0.13 |
| polarC18 | 276.144 | 54.5472 | 6.7 | 0.13 |
| polarC18 | 276.144 | 64.4655 | 6.7 | 0.13 |
| polarC18 | 276.144 | 89.3068 | 6.7 | 0.13 |
| polarC18 | 276.144 | 64.2638 | 6.7 | 0.13 |
| polarC18 | 276.144 | 66.0636 | 6.7 | 0.13 |
| polarC18 | 276.144 | 71.0245 | 6.7 | 0.13 |
| polarC18 | 276.144 | 72.7474 | 6.7 | 0.13 |
| polarC18 | 276.144 | 62.6566 | 6.7 | 0.13 |
| polarC18 | 276.144 | 127.4465 | 6.7 | 0.13 |
| polarC18 | 276.144 | 82.4224 | 6.7 | 0.13 |
| polarC18 | 276.144 | 212.3203 | 6.7 | 0.13 |
| polarC18 | 276.144 | 65.5544 | 6.7 | 0.13 |
| polarC18 | 276.144 | 134.9262 | 6.7 | 0.13 |
| polarC18 | 276.144 | 73.3083 | 6.7 | 0.13 |
| polarC18 | 276.144 | 117.6354 | 6.7 | 0.13 |
| polarC18 | 276.144 | 270.6409 | 6.7 | 0.13 |

| | | | | |
|---|---|---|---|---|
| polarC18 | 276.144 | 67.4502 | 6.7 | 0.13 |
| polarC18 | 276.144 | 57.9323 | 6.7 | 0.13 |
| polarC18 | 276.144 | 99.639 | 6.7 | 0.13 |
| polarC18 | 276.144 | 98.6547 | 6.7 | 0.13 |
| polarC18 | 276.144 | 152.2852 | 6.7 | 0.13 |
| polarC18 | 276.144 | 61.755 | 6.7 | 0.13 |
| polarC18 | 276.144 | 105.0139 | 6.7 | 0.13 |
| polarC18 | 276.144 | 62.0095 | 6.7 | 0.13 |
| polarC18 | 276.144 | 95.0427 | 6.7 | 0.13 |
| polarC18 | 276.144 | 66.8354 | 6.7 | 0.13 |
| polarC18 | 276.144 | 63.0802 | 6.7 | 0.13 |
| polarC18 | 276.144 | 110.6837 | 6.7 | 0.13 |
| polarC18 | 276.144 | 120.1543 | 6.7 | 0.13 |
| polarC18 | 276.144 | 126.0397 | 6.7 | 0.13 |
| polarC18 | 276.144 | 50.5786 | 6.7 | 0.13 |
| polarC18 | 276.144 | 107.0526 | 6.7 | 0.13 |
| polarC18 | 276.144 | 152.9446 | 6.7 | 0.13 |
| polarC18 | 276.144 | 144.0082 | 6.7 | 0.13 |
| polarC18 | 276.144 | 116.2815 | 6.7 | 0.13 |
| polarC18 | 276.144 | 65.9649 | 6.7 | 0.13 |
| polarC18 | 276.144 | 103.9533 | 6.7 | 0.13 |
| polarC18 | 276.144 | 76.6261 | 6.7 | 0.13 |
| polarC18 | 276.144 | 224.5709 | 6.7 | 0.13 |
| polarC18 | 276.144 | 98.1713 | 6.7 | 0.13 |
| polarC18 | 276.144 | 51.6641 | 6.7 | 0.13 |
| polarC18 | 276.144 | 66.2333 | 6.7 | 0.13 |
| polarC18 | 276.144 | 265.5694 | 6.7 | 0.13 |
| polarC18 | 276.144 | 71.6083 | 6.7 | 0.13 |
| polarC18 | 276.144 | 277.878 | 6.7 | 0.13 |
| polarC18 | 276.144 | 82.5146 | 6.7 | 0.13 |
| polarC18 | 276.144 | 93.2501 | 6.7 | 0.13 |
| polarC18 | 276.144 | 103.0183 | 6.7 | 0.13 |
| polarC18 | 276.144 | 61.6179 | 6.7 | 0.13 |
| polarC18 | 276.144 | 57.3668 | 6.7 | 0.13 |
| polarC18 | 276.144 | 76.3864 | 6.7 | 0.13 |
| polarC18 | 276.144 | 56.2662 | 6.7 | 0.13 |
| polarC18 | 276.144 | 70.6778 | 6.7 | 0.13 |
| polarC18 | 276.144 | 76.4108 | 6.7 | 0.13 |
| polarC18 | 276.144 | 77.1935 | 6.7 | 0.13 |
| polarC18 | 276.144 | 88.9922 | 6.7 | 0.13 |
| polarC18 | 276.144 | 204.9882 | 6.7 | 0.13 |
| polarC18 | 276.144 | 53.9163 | 6.7 | 0.13 |

| | | | | |
|---|---|---|---|---|
| polarC18 | 276.144 | 98.9537 | 6.7 | 0.13 |
| polarC18 | 276.144 | 63.6214 | 6.7 | 0.13 |
| polarC18 | 276.144 | 94.314 | 6.7 | 0.13 |
| polarC18 | 276.144 | 55.5027 | 6.7 | 0.13 |
| polarC18 | 276.144 | 116.3783 | 6.7 | 0.13 |
| polarC18 | 276.144 | 92.8269 | 6.7 | 0.13 |
| polarC18 | 276.144 | 220.4469 | 6.7 | 0.13 |
| polarC18 | 276.144 | 171.09 | 6.7 | 0.13 |
| polarC18 | 276.144 | 107.0248 | 6.7 | 0.13 |
| polarC18 | 276.144 | 131.5736 | 6.7 | 0.13 |
| polarC18 | 276.144 | 59.1199 | 6.7 | 0.13 |
| polarC18 | 276.144 | 116.189 | 6.7 | 0.13 |
| polarC18 | 276.144 | 99.0317 | 6.7 | 0.13 |
| polarC18 | 276.144 | 60.917 | 6.7 | 0.13 |
| polarC18 | 276.144 | 192.2917 | 6.7 | 0.13 |
| polarC18 | 276.144 | 195.3695 | 6.7 | 0.13 |
| polarC18 | 276.144 | 70.2003 | 6.7 | 0.13 |
| polarC18 | 276.144 | 178.9216 | 6.7 | 0.13 |
| polarC18 | 276.144 | 52.2236 | 6.7 | 0.13 |
| polarC18 | 276.144 | 140.6024 | 6.7 | 0.13 |
| polarC18 | 276.144 | 61.0484 | 6.7 | 0.13 |
| polarC18 | 276.144 | 85.768 | 6.7 | 0.13 |
| polarC18 | 276.144 | 75.6458 | 6.7 | 0.13 |
| polarC18 | 276.144 | 219.6203 | 6.7 | 0.13 |
| polarC18 | 276.144 | 72.9288 | 6.7 | 0.13 |
| polarC18 | 276.144 | 254.966 | 6.7 | 0.13 |
| polarC18 | 276.144 | 119.6465 | 6.7 | 0.13 |
| polarC18 | 276.144 | 68.826 | 6.7 | 0.13 |
| polarC18 | 276.144 | 125.2354 | 6.7 | 0.13 |
| polarC18 | 276.144 | 59.2416 | 6.7 | 0.13 |
| polarC18 | 276.144 | 54.868 | 6.7 | 0.13 |
| polarC18 | 276.144 | 64.1177 | 6.7 | 0.13 |
| polarC18 | 276.144 | 103.3818 | 6.7 | 0.13 |
| polarC18 | 276.144 | 84.5783 | 6.7 | 0.13 |
| polarC18 | 276.144 | 176.1257 | 6.7 | 0.13 |
| polarC18 | 276.144 | 67.4385 | 6.7 | 0.13 |
| polarC18 | 276.144 | 140.4088 | 6.7 | 0.13 |
| polarC18 | 276.144 | 190.8089 | 6.7 | 0.13 |
| polarC18 | 276.144 | 58.9642 | 6.7 | 0.13 |
| polarC18 | 276.144 | 118.233 | 6.7 | 0.13 |
| polarC18 | 276.144 | 54.509 | 6.7 | 0.13 |
| polarC18 | 276.144 | 50.7394 | 6.7 | 0.13 |

| | | | | |
|---|---|---|---|---|
| polarC18 | 276.144 | 76.9428 | 6.7 | 0.13 |
| polarC18 | 276.144 | 60.1087 | 6.7 | 0.13 |
| polarC18 | 276.144 | 52.341 | 6.7 | 0.13 |
| polarC18 | 276.144 | 111.3419 | 6.7 | 0.13 |
| polarC18 | 276.144 | 155.1216 | 6.7 | 0.13 |
| polarC18 | 276.144 | 59.8797 | 6.7 | 0.13 |
| polarC18 | 276.144 | 53.3038 | 6.7 | 0.13 |
| polarC18 | 276.144 | 64.0436 | 6.7 | 0.13 |
| polarC18 | 276.144 | 58.3767 | 6.7 | 0.13 |
| polarC18 | 276.144 | 195.6121 | 6.7 | 0.13 |
| polarC18 | 276.144 | 70.6654 | 6.7 | 0.13 |
| polarC18 | 276.144 | 68.5239 | 6.7 | 0.13 |
| polarC18 | 276.144 | 88.2904 | 6.7 | 0.13 |
| polarC18 | 276.144 | 202.3254 | 6.7 | 0.13 |
| polarC18 | 276.144 | 51.3838 | 6.7 | 0.13 |
| polarC18 | 276.144 | 134.6506 | 6.7 | 0.13 |
| polarC18 | 276.144 | 123.7909 | 6.7 | 0.13 |
| polarC18 | 276.144 | 96.4601 | 6.7 | 0.13 |
| polarC18 | 276.144 | 194.3802 | 6.7 | 0.13 |
| polarC18 | 276.144 | 127.9385 | 6.7 | 0.13 |
| polarC18 | 276.144 | 110.2772 | 6.7 | 0.13 |
| polarC18 | 276.144 | 93.7939 | 6.7 | 0.13 |
| polarC18 | 276.144 | 52.8577 | 6.7 | 0.13 |
| polarC18 | 276.144 | 233.6901 | 6.7 | 0.13 |
| polarC18 | 276.144 | 238.5338 | 6.7 | 0.13 |
| polarC18 | 276.144 | 179.2903 | 6.7 | 0.13 |
| polarC18 | 276.144 | 101.0702 | 6.7 | 0.13 |
| polarC18 | 276.144 | 88.8687 | 6.7 | 0.13 |
| polarC18 | 276.144 | 163.4216 | 6.7 | 0.13 |
| polarC18 | 276.144 | 164.0163 | 6.7 | 0.13 |
| polarC18 | 276.144 | 134.504 | 6.7 | 0.13 |
| polarC18 | 276.144 | 64.8613 | 6.7 | 0.13 |
| polarC18 | 276.144 | 107.468 | 6.7 | 0.13 |
| polarC18 | 276.144 | 50.2508 | 6.7 | 0.13 |
| polarC18 | 276.144 | 215.989 | 6.7 | 0.13 |
| polarC18 | 276.144 | 54.7524 | 6.7 | 0.13 |
| polarC18 | 276.144 | 71.3526 | 6.7 | 0.13 |
| polarC18 | 276.144 | 162.5943 | 6.7 | 0.13 |
| polarC18 | 276.144 | 55.3438 | 6.7 | 0.13 |
| polarC18 | 276.144 | 107.1356 | 6.7 | 0.13 |
| polarC18 | 276.144 | 221.227 | 6.7 | 0.13 |
| polarC18 | 276.144 | 78.8423 | 6.7 | 0.13 |

| | | | | |
|---|---|---|---|---|
| polarC18 | 276.144 | 208.0527 | 6.7 | 0.13 |
| polarC18 | 276.144 | 128.6662 | 6.7 | 0.13 |
| polarC18 | 276.144 | 166.4886 | 6.7 | 0.13 |
| polarC18 | 276.144 | 50.7006 | 6.7 | 0.13 |
| polarC18 | 276.144 | 78.0035 | 6.7 | 0.13 |
| polarC18 | 276.144 | 68.9623 | 6.7 | 0.13 |
| polarC18 | 276.144 | 53.0984 | 6.7 | 0.13 |
| polarC18 | 276.144 | 282.2892 | 6.7 | 0.13 |
| polarC18 | 276.144 | 92.2307 | 6.7 | 0.13 |
| polarC18 | 276.144 | 72.7985 | 6.7 | 0.13 |
| polarC18 | 276.144 | 56.1528 | 6.7 | 0.13 |
| polarC18 | 276.144 | 53.5381 | 6.7 | 0.13 |
| polarC18 | 276.144 | 53.3341 | 6.7 | 0.13 |
| polarC18 | 276.144 | 53.7841 | 6.7 | 0.13 |
| polarC18 | 276.144 | 62.5835 | 6.7 | 0.13 |
| polarC18 | 276.144 | 51.2696 | 6.7 | 0.13 |
| polarC18 | 276.144 | 80.7179 | 6.7 | 0.13 |
| polarC18 | 276.144 | 110.1032 | 6.7 | 0.13 |
| polarC18 | 276.144 | 186.9359 | 6.7 | 0.13 |
| polarC18 | 276.144 | 77.5994 | 6.7 | 0.13 |
| polarC18 | 276.144 | 105.4685 | 6.7 | 0.13 |
| polarC18 | 276.144 | 55.2518 | 6.7 | 0.13 |
| polarC18 | 276.144 | 150.8111 | 6.7 | 0.13 |
| polarC18 | 276.144 | 66.0693 | 6.7 | 0.13 |
| polarC18 | 276.144 | 73.3122 | 6.7 | 0.13 |
| polarC18 | 276.144 | 173.0141 | 6.7 | 0.13 |
| polarC18 | 276.144 | 220.1125 | 6.7 | 0.13 |
| polarC18 | 276.144 | 244.0499 | 6.7 | 0.13 |
| polarC18 | 276.144 | 60.0451 | 6.7 | 0.13 |
| polarC18 | 276.144 | 99.9203 | 6.7 | 0.13 |
| polarC18 | 276.144 | 137.9429 | 6.7 | 0.13 |
| polarC18 | 276.144 | 168.8622 | 6.7 | 0.13 |
| polarC18 | 276.144 | 189.5307 | 6.7 | 0.13 |
| polarC18 | 276.144 | 235.5108 | 6.7 | 0.13 |
| polarC18 | 276.144 | 51.7527 | 6.7 | 0.13 |
| polarC18 | 276.144 | 87.1734 | 6.7 | 0.13 |
| polarC18 | 276.144 | 59.9878 | 6.7 | 0.13 |
| polarC18 | 276.144 | 90.0365 | 6.7 | 0.13 |
| polarC18 | 276.144 | 224.1394 | 6.7 | 0.13 |
| polarC18 | 276.144 | 157.0863 | 6.7 | 0.13 |
| polarC18 | 276.144 | 60.8976 | 6.6 | 0.13 |
| polarC18 | 276.144 | 93.4974 | 6.6 | 0.13 |

| | | | | |
|---|---|---|---|---|
| polarC18 | 276.144 | 104.7652 | 6.6 | 0.13 |
| polarC18 | 276.144 | 139.1173 | 6.6 | 0.13 |
| polarC18 | 276.144 | 162.1088 | 6.6 | 0.13 |
| polarC18 | 276.144 | 63.3637 | 6.6 | 0.13 |
| polarC18 | 276.144 | 68.7995 | 6.6 | 0.13 |
| polarC18 | 276.144 | 128.9483 | 6.6 | 0.13 |
| polarC18 | 276.144 | 131.6846 | 6.6 | 0.13 |
| polarC18 | 276.144 | 293.3784 | 6.6 | 0.13 |
| polarC18 | 276.144 | 92.5539 | 6.6 | 0.13 |
| polarC18 | 276.144 | 137.2047 | 6.6 | 0.13 |
| polarC18 | 276.144 | 65.5431 | 6.6 | 0.13 |
| polarC18 | 276.144 | 112.9768 | 6.6 | 0.13 |
| polarC18 | 276.144 | 98.2217 | 6.6 | 0.13 |
| polarC18 | 276.144 | 137.9099 | 6.6 | 0.13 |
| polarC18 | 276.144 | 75.9485 | 6.6 | 0.13 |
| polarC18 | 276.144 | 61.3809 | 6.6 | 0.13 |
| polarC18 | 276.144 | 108.485 | 6.6 | 0.13 |
| polarC18 | 276.144 | 243.2494 | 6.6 | 0.13 |
| polarC18 | 276.144 | 178.1302 | 6.6 | 0.13 |
| polarC18 | 276.144 | 76.8457 | 6.6 | 0.13 |
| polarC18 | 276.144 | 242.9913 | 6.6 | 0.13 |
| polarC18 | 276.144 | 74.8397 | 6.6 | 0.13 |
| polarC18 | 276.144 | 56.3278 | 6.6 | 0.13 |
| polarC18 | 276.144 | 50.3778 | 6.6 | 0.13 |
| polarC18 | 276.144 | 173.2903 | 6.6 | 0.13 |
| polarC18 | 276.144 | 97.6727 | 6.6 | 0.13 |
| polarC18 | 276.144 | 61.1519 | 6.6 | 0.13 |
| polarC18 | 276.144 | 166.6954 | 6.6 | 0.13 |
| polarC18 | 276.144 | 68.137 | 6.6 | 0.13 |
| polarC18 | 276.144 | 73.651 | 6.6 | 0.13 |
| polarC18 | 276.144 | 66.9522 | 6.6 | 0.13 |
| polarC18 | 276.144 | 53.0468 | 6.6 | 0.13 |
| polarC18 | 276.144 | 80.2113 | 6.6 | 0.13 |
| polarC18 | 276.144 | 70.8673 | 6.6 | 0.13 |
| polarC18 | 276.144 | 51.4117 | 6.6 | 0.13 |
| polarC18 | 276.144 | 75.7271 | 6.6 | 0.13 |
| polarC18 | 276.144 | 96.7005 | 6.6 | 0.13 |
| polarC18 | 276.144 | 67.4155 | 6.6 | 0.13 |
| polarC18 | 276.144 | 119.1751 | 6.6 | 0.13 |
| polarC18 | 276.144 | 103.478 | 6.6 | 0.13 |
| polarC18 | 276.144 | 68.3089 | 6.6 | 0.13 |
| polarC18 | 276.144 | 279.3342 | 6.6 | 0.13 |

| | | | | |
|---|---|---|---|---|
| polarC18 | 276.144 | 77.8608 | 6.6 | 0.13 |
| polarC18 | 276.144 | 185.6485 | 6.6 | 0.13 |
| polarC18 | 276.144 | 57.3895 | 6.6 | 0.13 |
| polarC18 | 276.144 | 72.6031 | 6.6 | 0.13 |
| polarC18 | 276.144 | 100.043 | 6.6 | 0.13 |
| polarC18 | 276.144 | 129.2919 | 6.6 | 0.13 |
| polarC18 | 276.144 | 126.8529 | 6.6 | 0.13 |
| polarC18 | 276.144 | 144.7056 | 6.6 | 0.13 |
| polarC18 | 276.144 | 122.3408 | 6.6 | 0.13 |
| polarC18 | 276.144 | 92.1352 | 6.6 | 0.13 |
| polarC18 | 276.144 | 124.8843 | 6.6 | 0.13 |
| polarC18 | 276.144 | 52.8648 | 6.6 | 0.13 |
| polarC18 | 276.144 | 267.0974 | 6.6 | 0.13 |
| polarC18 | 276.144 | 206.5905 | 6.6 | 0.13 |
| polarC18 | 276.144 | 175.0616 | 6.6 | 0.13 |
| polarC18 | 276.144 | 135.0994 | 6.6 | 0.13 |
| polarC18 | 276.144 | 148.4605 | 6.6 | 0.13 |
| polarC18 | 276.144 | 97.5501 | 6.6 | 0.13 |
| polarC18 | 276.144 | 185.0057 | 6.6 | 0.13 |
| polarC18 | 276.144 | 159.3514 | 6.6 | 0.13 |
| polarC18 | 276.144 | 250.3336 | 6.6 | 0.13 |
| polarC18 | 276.144 | 119.1654 | 6.6 | 0.13 |
| polarC18 | 276.144 | 149.2546 | 6.6 | 0.13 |
| polarC18 | 276.144 | 88.7773 | 6.6 | 0.13 |
| polarC18 | 276.144 | 56.1038 | 6.6 | 0.13 |
| polarC18 | 276.144 | 51.8909 | 6.6 | 0.13 |
| polarC18 | 276.144 | 110.1503 | 6.6 | 0.13 |
| polarC18 | 276.144 | 165.0771 | 6.6 | 0.13 |
| polarC18 | 276.144 | 206.3964 | 6.6 | 0.13 |
| polarC18 | 276.144 | 52.0684 | 6.6 | 0.13 |
| polarC18 | 276.144 | 128.4001 | 6.6 | 0.13 |
| polarC18 | 276.144 | 74.0951 | 6.6 | 0.13 |
| polarC18 | 276.144 | 64.4169 | 6.6 | 0.13 |
| polarC18 | 276.144 | 59.9235 | 6.6 | 0.13 |
| polarC18 | 276.144 | 140.1531 | 6.6 | 0.13 |
| polarC18 | 276.144 | 56.7429 | 6.6 | 0.13 |
| polarC18 | 276.144 | 101.5257 | 6.6 | 0.13 |
| polarC18 | 276.144 | 163.3519 | 6.6 | 0.13 |
| polarC18 | 276.144 | 111.044 | 6.6 | 0.13 |
| polarC18 | 276.144 | 169.1912 | 6.6 | 0.13 |
| polarC18 | 276.144 | 83.6628 | 6.6 | 0.13 |
| polarC18 | 276.144 | 60.9091 | 6.6 | 0.13 |

| | | | | |
|---|---|---|---|---|
| polarC18 | 276.144 | 84.8475 | 6.6 | 0.13 |
| polarC18 | 276.144 | 50.9708 | 6.6 | 0.13 |
| polarC18 | 276.144 | 51.6684 | 6.6 | 0.13 |
| polarC18 | 276.144 | 190.3705 | 6.6 | 0.13 |
| polarC18 | 276.144 | 126.8884 | 6.6 | 0.13 |
| polarC18 | 276.144 | 111.6633 | 6.6 | 0.13 |
| polarC18 | 276.144 | 130.9045 | 6.6 | 0.13 |
| polarC18 | 276.144 | 203.7847 | 6.6 | 0.13 |
| polarC18 | 276.144 | 70.8407 | 6.6 | 0.13 |
| polarC18 | 276.144 | 225.8407 | 6.6 | 0.13 |
| polarC18 | 276.144 | 86.5542 | 6.6 | 0.13 |
| polarC18 | 276.144 | 75.3844 | 6.6 | 0.13 |
| polarC18 | 276.144 | 85.0706 | 6.6 | 0.13 |
| polarC18 | 276.144 | 61.3883 | 6.6 | 0.13 |
| polarC18 | 276.144 | 69.4231 | 6.6 | 0.13 |
| polarC18 | 276.144 | 50.0376 | 6.6 | 0.13 |
| polarC18 | 276.144 | 96.5343 | 6.6 | 0.13 |
| polarC18 | 276.144 | 69.7517 | 6.6 | 0.13 |
| polarC18 | 276.144 | 199.2798 | 6.6 | 0.13 |
| polarC18 | 276.144 | 57.4759 | 6.6 | 0.13 |
| polarC18 | 276.144 | 211.6245 | 6.6 | 0.13 |
| polarC18 | 276.144 | 51.8499 | 6.6 | 0.13 |
| polarC18 | 276.144 | 63.2994 | 6.6 | 0.13 |
| polarC18 | 276.144 | 60.3048 | 6.6 | 0.13 |
| polarC18 | 276.144 | 50.1698 | 6.6 | 0.13 |
| polarC18 | 276.144 | 54.5189 | 6.6 | 0.13 |
| polarC18 | 276.144 | 97.9956 | 6.6 | 0.13 |
| polarC18 | 276.144 | 138.6363 | 6.6 | 0.13 |
| polarC18 | 276.144 | 55.3229 | 6.6 | 0.13 |
| polarC18 | 276.144 | 137.6852 | 6.6 | 0.13 |
| polarC18 | 276.144 | 193.275 | 6.6 | 0.13 |
| polarC18 | 276.144 | 131.3438 | 6.6 | 0.13 |
| polarC18 | 276.144 | 51.5701 | 6.6 | 0.13 |
| polarC18 | 276.144 | 96.214 | 6.6 | 0.13 |
| polarC18 | 276.144 | 103.2146 | 6.6 | 0.13 |
| polarC18 | 276.144 | 103.001 | 6.6 | 0.13 |
| polarC18 | 276.144 | 56.7478 | 6.6 | 0.13 |
| polarC18 | 276.144 | 250.6843 | 6.6 | 0.13 |
| polarC18 | 276.144 | 63.1188 | 6.6 | 0.13 |
| polarC18 | 276.144 | 135.9028 | 6.6 | 0.13 |
| polarC18 | 276.144 | 60.9485 | 6.6 | 0.13 |
| polarC18 | 276.144 | 69.543 | 6.6 | 0.13 |

| | | | | |
|---|---|---|---|---|
| polarC18 | 276.144 | 90.8493 | 6.6 | 0.13 |
| polarC18 | 276.144 | 108.291 | 6.6 | 0.13 |
| polarC18 | 276.144 | 105.6493 | 6.6 | 0.13 |
| polarC18 | 276.144 | 69.7954 | 6.6 | 0.13 |
| polarC18 | 276.144 | 53.0937 | 6.6 | 0.13 |
| polarC18 | 276.144 | 82.6022 | 6.6 | 0.13 |
| polarC18 | 276.144 | 63.0898 | 6.6 | 0.13 |
| polarC18 | 276.144 | 75.5188 | 6.6 | 0.13 |
| polarC18 | 276.144 | 55.7753 | 6.6 | 0.13 |
| polarC18 | 276.144 | 102.5021 | 6.6 | 0.13 |
| polarC18 | 276.144 | 208.5186 | 6.6 | 0.13 |
| polarC18 | 276.144 | 166.8561 | 6.6 | 0.13 |
| polarC18 | 276.144 | 64.326 | 6.6 | 0.13 |
| polarC18 | 276.144 | 86.0895 | 6.6 | 0.13 |
| polarC18 | 276.144 | 104.1356 | 6.6 | 0.13 |
| polarC18 | 276.144 | 56.5874 | 6.6 | 0.13 |
| polarC18 | 276.144 | 182.6069 | 6.6 | 0.13 |
| polarC18 | 276.144 | 58.6797 | 6.6 | 0.13 |
| polarC18 | 276.144 | 237.5641 | 6.6 | 0.13 |
| polarC18 | 276.144 | 73.7018 | 6.6 | 0.13 |
| polarC18 | 276.144 | 65.6262 | 6.6 | 0.13 |
| polarC18 | 276.144 | 104.1462 | 6.6 | 0.13 |
| polarC18 | 276.144 | 82.4457 | 6.6 | 0.13 |
| polarC18 | 276.144 | 63.8584 | 6.6 | 0.13 |
| polarC18 | 276.144 | 118.507 | 6.6 | 0.13 |
| polarC18 | 276.144 | 90.0763 | 6.6 | 0.13 |
| polarC18 | 276.144 | 292.4682 | 6.6 | 0.13 |
| polarC18 | 276.144 | 150.2574 | 6.6 | 0.13 |
| polarC18 | 276.144 | 69.4057 | 6.6 | 0.13 |
| polarC18 | 276.144 | 94.8863 | 6.6 | 0.13 |
| polarC18 | 276.144 | 173.4685 | 6.6 | 0.13 |
| polarC18 | 276.144 | 99.6967 | 6.6 | 0.13 |
| polarC18 | 276.144 | 92.3309 | 6.6 | 0.13 |
| polarC18 | 276.144 | 257.0198 | 6.6 | 0.13 |
| polarC18 | 276.144 | 102.4969 | 6.6 | 0.13 |
| polarC18 | 276.144 | 140.3175 | 6.6 | 0.13 |
| polarC18 | 276.144 | 169.4732 | 6.6 | 0.13 |
| polarC18 | 276.144 | 53.3596 | 6.6 | 0.13 |
| polarC18 | 276.144 | 159.605 | 6.6 | 0.13 |
| polarC18 | 276.144 | 98.3448 | 6.6 | 0.13 |
| polarC18 | 276.144 | 81.2422 | 6.6 | 0.13 |
| polarC18 | 276.144 | 58.5165 | 6.6 | 0.13 |

| | | | | |
|---|---|---|---|---|
| polarC18 | 276.144 | 129.4045 | 6.6 | 0.13 |
| polarC18 | 276.144 | 105.5611 | 6.6 | 0.13 |
| polarC18 | 276.144 | 62.5756 | 6.6 | 0.13 |
| polarC18 | 276.144 | 78.4987 | 6.6 | 0.13 |
| polarC18 | 276.144 | 55.9166 | 6.6 | 0.13 |
| polarC18 | 276.144 | 114.5836 | 6.6 | 0.13 |
| polarC18 | 276.144 | 87.0659 | 6.6 | 0.13 |
| polarC18 | 276.144 | 76.8197 | 6.6 | 0.13 |
| polarC18 | 276.144 | 57.1376 | 6.6 | 0.13 |
| polarC18 | 276.144 | 88.8776 | 6.6 | 0.13 |
| polarC18 | 276.144 | 188.978 | 6.6 | 0.13 |
| polarC18 | 276.144 | 106.9907 | 6.6 | 0.13 |
| polarC18 | 276.144 | 153.7553 | 6.6 | 0.13 |
| polarC18 | 276.144 | 297.7582 | 6.6 | 0.13 |
| polarC18 | 276.144 | 122.9373 | 6.6 | 0.13 |
| polarC18 | 276.144 | 51.017 | 6.6 | 0.13 |
| polarC18 | 276.144 | 88.9439 | 6.6 | 0.13 |
| polarC18 | 276.144 | 55.8956 | 6.6 | 0.13 |
| polarC18 | 276.144 | 77.6265 | 6.6 | 0.13 |
| polarC18 | 276.144 | 59.7975 | 6.6 | 0.13 |
| polarC18 | 276.144 | 215.7381 | 6.6 | 0.13 |
| polarC18 | 276.144 | 62.672 | 6.6 | 0.13 |
| polarC18 | 276.144 | 58.6457 | 6.6 | 0.13 |
| polarC18 | 276.144 | 170.0179 | 6.6 | 0.13 |
| polarC18 | 276.144 | 234.125 | 6.6 | 0.13 |
| polarC18 | 276.144 | 68.5325 | 6.6 | 0.13 |
| polarC18 | 276.144 | 76.593 | 6.6 | 0.13 |
| polarC18 | 276.144 | 70.1488 | 6.6 | 0.13 |
| polarC18 | 276.144 | 65.3564 | 6.6 | 0.13 |
| polarC18 | 276.144 | 59.4079 | 6.6 | 0.13 |
| polarC18 | 276.144 | 102.5905 | 6.6 | 0.13 |
| polarC18 | 276.144 | 55.8078 | 6.6 | 0.13 |
| polarC18 | 276.144 | 72.9678 | 6.6 | 0.13 |
| polarC18 | 276.144 | 75.1212 | 6.6 | 0.13 |
| polarC18 | 276.144 | 186.7953 | 6.6 | 0.13 |
| polarC18 | 276.144 | 67.6101 | 6.6 | 0.13 |
| polarC18 | 276.144 | 54.2318 | 6.6 | 0.13 |
| polarC18 | 276.144 | 61.1831 | 6.6 | 0.13 |
| polarC18 | 276.144 | 138.5344 | 6.6 | 0.13 |
| polarC18 | 276.144 | 105.6051 | 6.6 | 0.13 |
| polarC18 | 276.144 | 237.186 | 6.6 | 0.13 |
| polarC18 | 276.144 | 79.1678 | 6.6 | 0.13 |

| | | | | |
|---|---|---|---|---|
| polarC18 | 276.144 | 108.9599 | 6.6 | 0.13 |
| polarC18 | 276.144 | 116.4661 | 6.6 | 0.13 |
| polarC18 | 276.144 | 199.1581 | 6.6 | 0.13 |
| polarC18 | 276.144 | 57.4434 | 6.6 | 0.13 |
| polarC18 | 276.144 | 228.6734 | 6.6 | 0.13 |
| polarC18 | 276.144 | 75.7905 | 6.6 | 0.13 |
| polarC18 | 276.144 | 98.7366 | 6.6 | 0.13 |
| polarC18 | 276.144 | 82.0641 | 6.6 | 0.13 |
| polarC18 | 276.144 | 86.3121 | 6.6 | 0.13 |
| polarC18 | 276.144 | 187.154 | 6.6 | 0.13 |
| polarC18 | 276.144 | 62.8936 | 6.6 | 0.13 |
| polarC18 | 276.144 | 104.7494 | 6.6 | 0.13 |
| polarC18 | 276.144 | 58.13 | 6.6 | 0.13 |
| polarC18 | 276.144 | 76.5456 | 6.6 | 0.13 |
| polarC18 | 276.144 | 64.0098 | 6.6 | 0.13 |
| polarC18 | 276.144 | 77.2367 | 6.6 | 0.13 |
| polarC18 | 276.144 | 75.8408 | 6.6 | 0.13 |
| polarC18 | 276.144 | 92.4054 | 6.6 | 0.13 |
| polarC18 | 276.144 | 131.1072 | 6.6 | 0.13 |
| polarC18 | 276.144 | 123.3607 | 6.6 | 0.13 |
| polarC18 | 276.144 | 92.0023 | 6.6 | 0.13 |
| polarC18 | 276.144 | 80.1475 | 6.6 | 0.13 |
| polarC18 | 276.144 | 97.898 | 6.6 | 0.13 |
| polarC18 | 276.144 | 263.1994 | 6.6 | 0.13 |
| polarC18 | 276.144 | 106.7213 | 6.6 | 0.13 |
| polarC18 | 276.144 | 60.2669 | 6.6 | 0.13 |
| polarC18 | 276.144 | 53.6545 | 6.6 | 0.13 |
| polarC18 | 276.144 | 289.413 | 6.6 | 0.13 |
| polarC18 | 276.144 | 73.5615 | 6.6 | 0.13 |
| polarC18 | 276.144 | 51.2304 | 6.6 | 0.13 |
| polarC18 | 276.144 | 71.6765 | 6.6 | 0.13 |
| polarC18 | 276.144 | 65.7864 | 6.6 | 0.13 |
| polarC18 | 276.144 | 185.7114 | 6.6 | 0.13 |
| polarC18 | 276.144 | 193.159 | 6.6 | 0.13 |
| polarC18 | 276.144 | 110.5314 | 6.6 | 0.13 |
| polarC18 | 276.144 | 65.1539 | 6.6 | 0.13 |
| polarC18 | 276.144 | 179.8162 | 6.6 | 0.13 |
| polarC18 | 276.144 | 105.6917 | 6.6 | 0.13 |
| polarC18 | 276.144 | 61.9557 | 6.6 | 0.13 |
| polarC18 | 276.144 | 79.6315 | 6.6 | 0.13 |
| polarC18 | 276.144 | 99.1887 | 6.6 | 0.13 |
| polarC18 | 276.144 | 119.0337 | 6.6 | 0.13 |

| | | | | |
|---|---|---|---|---|
| polarC18 | 276.144 | 248.9456 | 6.6 | 0.13 |
| polarC18 | 276.144 | 84.8103 | 6.6 | 0.13 |
| polarC18 | 276.144 | 62.4291 | 6.6 | 0.13 |
| polarC18 | 276.144 | 92.0334 | 6.6 | 0.13 |
| polarC18 | 276.144 | 78.5934 | 6.6 | 0.13 |
| polarC18 | 276.144 | 50.1227 | 6.6 | 0.13 |
| polarC18 | 276.144 | 94.6738 | 6.6 | 0.13 |
| polarC18 | 276.144 | 114.9751 | 6.6 | 0.13 |
| polarC18 | 276.144 | 64.2183 | 6.6 | 0.13 |
| polarC18 | 276.144 | 100.2942 | 6.6 | 0.13 |
| polarC18 | 276.144 | 68.4876 | 6.6 | 0.13 |
| polarC18 | 276.144 | 136.2802 | 6.6 | 0.13 |
| polarC18 | 276.144 | 66.4085 | 6.6 | 0.13 |
| polarC18 | 276.144 | 126.4302 | 6.6 | 0.13 |
| polarC18 | 276.144 | 86.886 | 6.6 | 0.13 |
| polarC18 | 276.144 | 52.4067 | 6.6 | 0.13 |
| polarC18 | 276.144 | 69.0784 | 6.6 | 0.13 |
| polarC18 | 276.144 | 105.4322 | 6.6 | 0.13 |
| polarC18 | 276.144 | 100.89 | 6.6 | 0.13 |
| polarC18 | 276.144 | 135.5233 | 6.6 | 0.13 |
| polarC18 | 276.144 | 96.2377 | 6.6 | 0.13 |
| polarC18 | 276.144 | 55.0906 | 6.6 | 0.13 |
| polarC18 | 276.144 | 56.1385 | 6.6 | 0.13 |
| polarC18 | 276.144 | 180.6086 | 6.6 | 0.13 |
| polarC18 | 276.144 | 75.3528 | 6.6 | 0.13 |
| polarC18 | 276.144 | 72.1342 | 6.6 | 0.13 |
| polarC18 | 276.144 | 285.2504 | 6.6 | 0.13 |
| polarC18 | 276.144 | 173.1171 | 6.6 | 0.13 |
| polarC18 | 276.144 | 58.3867 | 6.6 | 0.13 |
| polarC18 | 276.144 | 84.4885 | 6.6 | 0.13 |
| polarC18 | 276.144 | 71.7704 | 6.6 | 0.13 |
| polarC18 | 276.144 | 87.9286 | 6.6 | 0.13 |
| polarC18 | 276.144 | 71.5255 | 6.6 | 0.13 |
| polarC18 | 276.144 | 81.2835 | 6.6 | 0.13 |
| polarC18 | 276.144 | 65.0756 | 6.6 | 0.13 |
| polarC18 | 276.144 | 54.5832 | 6.6 | 0.13 |
| polarC18 | 276.144 | 76.869 | 6.6 | 0.13 |
| polarC18 | 276.144 | 79.0475 | 6.6 | 0.13 |
| polarC18 | 276.144 | 68.6369 | 6.6 | 0.13 |
| polarC18 | 276.144 | 114.7894 | 6.6 | 0.13 |
| polarC18 | 276.144 | 53.9063 | 6.6 | 0.13 |
| polarC18 | 276.144 | 108.5826 | 6.6 | 0.13 |

| | | | | |
|---|---|---|---|---|
| polarC18 | 276.144 | 181.7625 | 6.6 | 0.13 |
| polarC18 | 276.144 | 226.5314 | 6.6 | 0.13 |
| polarC18 | 276.144 | 92.08 | 6.6 | 0.13 |
| polarC18 | 276.144 | 96.7473 | 6.6 | 0.13 |
| polarC18 | 276.144 | 208.927 | 6.6 | 0.13 |
| polarC18 | 276.144 | 272.1457 | 6.6 | 0.13 |
| polarC18 | 276.144 | 124.43 | 6.6 | 0.13 |
| polarC18 | 276.144 | 66.8753 | 6.6 | 0.13 |
| polarC18 | 276.144 | 97.6392 | 6.6 | 0.13 |
| polarC18 | 276.144 | 110.9122 | 6.6 | 0.13 |
| polarC18 | 276.144 | 58.7333 | 6.6 | 0.13 |
| polarC18 | 276.144 | 61.2984 | 6.6 | 0.13 |
| polarC18 | 276.144 | 56.391 | 6.6 | 0.13 |
| polarC18 | 276.144 | 71.5576 | 6.6 | 0.13 |
| polarC18 | 276.144 | 91.0376 | 6.6 | 0.13 |
| polarC18 | 276.144 | 222.9792 | 6.6 | 0.13 |
| polarC18 | 276.144 | 54.5139 | 6.6 | 0.13 |
| polarC18 | 276.144 | 296.5096 | 6.6 | 0.13 |
| polarC18 | 276.144 | 99.6 | 6.6 | 0.13 |
| polarC18 | 276.144 | 84.3203 | 6.6 | 0.13 |
| polarC18 | 276.144 | 58.1188 | 6.6 | 0.13 |
| polarC18 | 276.144 | 106.8024 | 6.6 | 0.13 |
| polarC18 | 276.144 | 61.5308 | 6.6 | 0.13 |
| polarC18 | 276.144 | 163.0317 | 6.6 | 0.13 |
| polarC18 | 276.144 | 166.7394 | 6.6 | 0.13 |
| polarC18 | 276.144 | 117.3798 | 6.6 | 0.13 |
| polarC18 | 276.144 | 71.3416 | 6.6 | 0.13 |
| polarC18 | 276.144 | 86.1518 | 6.6 | 0.13 |
| polarC18 | 276.144 | 65.0648 | 6.6 | 0.13 |
| polarC18 | 276.144 | 65.0932 | 6.6 | 0.13 |
| polarC18 | 276.144 | 116.5232 | 6.6 | 0.13 |
| polarC18 | 276.144 | 73.2091 | 6.6 | 0.13 |
| polarC18 | 276.144 | 74.0663 | 6.6 | 0.13 |
| polarC18 | 276.144 | 101.8546 | 6.6 | 0.13 |
| polarC18 | 276.144 | 64.2849 | 6.6 | 0.13 |
| polarC18 | 276.144 | 127.9692 | 6.6 | 0.13 |
| polarC18 | 276.144 | 58.6669 | 6.6 | 0.13 |
| polarC18 | 276.144 | 105.9355 | 6.6 | 0.13 |
| polarC18 | 276.144 | 54.79 | 6.6 | 0.13 |
| polarC18 | 276.144 | 82.2833 | 6.6 | 0.13 |
| polarC18 | 276.144 | 164.8758 | 6.6 | 0.13 |
| polarC18 | 276.144 | 141.6513 | 6.6 | 0.13 |

| | | | | |
|---|---|---|---|---|
| polarC18 | 276.144 | 230.5233 | 6.6 | 0.13 |
| polarC18 | 276.144 | 50.0587 | 6.6 | 0.13 |
| polarC18 | 276.144 | 158.4611 | 6.6 | 0.13 |
| polarC18 | 276.144 | 53.8376 | 6.6 | 0.13 |
| polarC18 | 276.144 | 53.3613 | 6.6 | 0.13 |
| polarC18 | 276.144 | 129.0964 | 6.6 | 0.13 |
| polarC18 | 276.144 | 99.6741 | 6.6 | 0.13 |
| polarC18 | 276.144 | 115.0695 | 6.5 | 0.13 |
| polarC18 | 276.144 | 50.0639 | 6.5 | 0.13 |
| polarC18 | 276.144 | 73.9949 | 6.5 | 0.13 |
| polarC18 | 276.144 | 97.8477 | 6.5 | 0.13 |
| polarC18 | 276.144 | 93.2964 | 6.5 | 0.13 |
| polarC18 | 276.144 | 295.4438 | 6.5 | 0.13 |
| polarC18 | 276.144 | 64.4281 | 6.5 | 0.13 |
| polarC18 | 276.144 | 136.4886 | 6.5 | 0.13 |
| polarC18 | 276.144 | 74.8554 | 6.5 | 0.13 |
| polarC18 | 276.144 | 99.0131 | 6.5 | 0.13 |
| polarC18 | 276.144 | 117.0065 | 6.5 | 0.13 |
| polarC18 | 276.144 | 171.1651 | 6.5 | 0.13 |
| polarC18 | 276.144 | 55.334 | 6.5 | 0.13 |
| polarC18 | 276.144 | 225.7136 | 6.5 | 0.13 |
| polarC18 | 276.144 | 206.1734 | 6.5 | 0.13 |
| polarC18 | 276.144 | 270.7187 | 6.5 | 0.13 |
| polarC18 | 276.144 | 97.518 | 6.5 | 0.13 |
| polarC18 | 276.144 | 52.2077 | 6.5 | 0.13 |
| polarC18 | 276.144 | 208.4344 | 6.5 | 0.13 |
| polarC18 | 276.144 | 58.3316 | 6.5 | 0.13 |
| polarC18 | 276.144 | 119.6985 | 6.5 | 0.13 |
| polarC18 | 276.144 | 288.9352 | 6.5 | 0.13 |
| polarC18 | 276.144 | 54.9354 | 6.5 | 0.13 |
| polarC18 | 276.144 | 51.0417 | 6.5 | 0.13 |
| polarC18 | 276.144 | 287.5544 | 6.5 | 0.13 |
| polarC18 | 276.144 | 292.812 | 6.5 | 0.13 |
| polarC18 | 276.144 | 61.5978 | 6.5 | 0.13 |
| polarC18 | 276.144 | 53.4991 | 6.5 | 0.13 |
| polarC18 | 276.144 | 59.0968 | 6.5 | 0.13 |
| polarC18 | 276.144 | 68.0871 | 6.5 | 0.13 |
| polarC18 | 276.144 | 108.8066 | 6.5 | 0.13 |
| polarC18 | 276.144 | 87.6857 | 6.5 | 0.13 |
| polarC18 | 276.144 | 88.3009 | 6.5 | 0.13 |
| polarC18 | 276.144 | 73.0618 | 6.5 | 0.13 |
| polarC18 | 276.144 | 59.1339 | 6.5 | 0.13 |

| | | | | |
|---|---|---|---|---|
| polarC18 | 276.144 | 88.9862 | 6.5 | 0.13 |
| polarC18 | 276.144 | 73.7278 | 6.5 | 0.13 |
| polarC18 | 276.144 | 79.231 | 6.5 | 0.13 |
| polarC18 | 276.144 | 125.3535 | 6.5 | 0.13 |
| polarC18 | 276.144 | 65.9868 | 6.5 | 0.13 |
| polarC18 | 276.144 | 71.0329 | 6.5 | 0.13 |
| polarC18 | 276.144 | 91.2462 | 6.5 | 0.13 |
| polarC18 | 276.144 | 61.5751 | 6.5 | 0.13 |
| polarC18 | 276.144 | 207.7055 | 6.5 | 0.13 |
| polarC18 | 276.144 | 71.2859 | 6.5 | 0.13 |
| polarC18 | 276.144 | 90.2863 | 6.5 | 0.13 |
| polarC18 | 276.144 | 65.67 | 6.5 | 0.13 |
| polarC18 | 276.144 | 148.7391 | 6.5 | 0.13 |
| polarC18 | 276.144 | 137.3633 | 6.5 | 0.13 |
| polarC18 | 276.144 | 145.5678 | 6.5 | 0.13 |
| polarC18 | 276.144 | 130.7442 | 6.5 | 0.13 |
| polarC18 | 276.144 | 110.224 | 6.5 | 0.13 |
| polarC18 | 276.144 | 114.4634 | 6.5 | 0.13 |
| polarC18 | 276.144 | 68.2771 | 6.5 | 0.13 |
| polarC18 | 276.144 | 139.4297 | 6.5 | 0.13 |
| polarC18 | 276.144 | 140.9437 | 6.5 | 0.13 |
| polarC18 | 276.144 | 163.882 | 6.5 | 0.13 |
| polarC18 | 276.144 | 188.4651 | 6.5 | 0.13 |
| polarC18 | 276.144 | 173.2594 | 6.5 | 0.13 |
| polarC18 | 276.144 | 100.838 | 6.5 | 0.13 |
| polarC18 | 276.144 | 70.4862 | 6.5 | 0.13 |
| polarC18 | 276.144 | 72.4726 | 6.5 | 0.13 |
| polarC18 | 276.144 | 129.5836 | 6.5 | 0.13 |
| polarC18 | 276.144 | 233.4426 | 6.5 | 0.13 |
| polarC18 | 276.144 | 182.3604 | 6.5 | 0.13 |
| polarC18 | 276.144 | 87.8696 | 6.5 | 0.13 |
| polarC18 | 276.144 | 73.3913 | 6.5 | 0.13 |
| polarC18 | 276.144 | 68.2339 | 6.5 | 0.13 |
| polarC18 | 276.144 | 95.4789 | 6.5 | 0.13 |
| polarC18 | 276.144 | 91.9328 | 6.5 | 0.13 |
| polarC18 | 276.144 | 117.0705 | 6.5 | 0.13 |
| polarC18 | 276.144 | 63.15 | 6.5 | 0.13 |
| polarC18 | 276.144 | 107.1664 | 6.5 | 0.13 |
| polarC18 | 276.144 | 55.7048 | 6.5 | 0.13 |
| polarC18 | 276.144 | 91.8913 | 6.5 | 0.13 |
| polarC18 | 276.144 | 65.4629 | 6.5 | 0.13 |
| polarC18 | 276.144 | 65.3099 | 6.5 | 0.13 |

| | | | | |
|---|---|---|---|---|
| polarC18 | 276.144 | 62.3941 | 6.5 | 0.13 |
| polarC18 | 276.144 | 57.4226 | 6.5 | 0.13 |
| polarC18 | 276.144 | 66.4428 | 6.5 | 0.13 |
| polarC18 | 276.144 | 53.404 | 6.5 | 0.13 |
| polarC18 | 276.144 | 51.0133 | 6.5 | 0.13 |
| polarC18 | 276.144 | 54.4541 | 6.5 | 0.13 |
| polarC18 | 276.144 | 138.4539 | 6.5 | 0.13 |
| polarC18 | 276.144 | 97.2132 | 6.5 | 0.13 |
| polarC18 | 276.144 | 84.0559 | 6.5 | 0.13 |
| polarC18 | 276.144 | 64.0266 | 6.5 | 0.13 |
| polarC18 | 276.144 | 121.1601 | 6.5 | 0.13 |
| polarC18 | 276.144 | 126.774 | 6.5 | 0.13 |
| polarC18 | 276.144 | 90.2543 | 6.5 | 0.13 |
| polarC18 | 276.144 | 81.9713 | 6.5 | 0.13 |
| polarC18 | 276.144 | 156.197 | 6.5 | 0.13 |
| polarC18 | 276.144 | 171.0528 | 6.5 | 0.13 |
| polarC18 | 276.144 | 64.9442 | 6.5 | 0.13 |
| polarC18 | 276.144 | 121.5096 | 6.5 | 0.13 |
| polarC18 | 276.144 | 71.9256 | 6.5 | 0.13 |
| polarC18 | 276.144 | 72.733 | 6.5 | 0.13 |
| polarC18 | 276.144 | 127.7059 | 6.5 | 0.13 |
| polarC18 | 276.144 | 66.333 | 6.5 | 0.13 |
| polarC18 | 276.144 | 161.2047 | 6.5 | 0.13 |
| polarC18 | 276.144 | 117.985 | 6.5 | 0.13 |
| polarC18 | 276.144 | 81.8246 | 6.5 | 0.13 |
| polarC18 | 276.144 | 52.6191 | 6.5 | 0.13 |
| polarC18 | 276.144 | 53.3193 | 6.5 | 0.13 |
| polarC18 | 276.144 | 134.2496 | 6.5 | 0.13 |
| polarC18 | 276.144 | 76.2776 | 6.5 | 0.13 |
| polarC18 | 276.144 | 78.8242 | 6.5 | 0.13 |
| polarC18 | 276.144 | 68.3142 | 6.5 | 0.13 |
| polarC18 | 276.144 | 91.62 | 6.5 | 0.13 |
| polarC18 | 276.144 | 259.2926 | 6.5 | 0.13 |
| polarC18 | 276.144 | 97.9874 | 6.5 | 0.13 |
| polarC18 | 276.144 | 273.9874 | 6.5 | 0.13 |
| polarC18 | 276.144 | 220.0263 | 6.5 | 0.13 |
| polarC18 | 276.144 | 76.021 | 6.5 | 0.13 |
| polarC18 | 276.144 | 138.1608 | 6.5 | 0.13 |
| polarC18 | 276.144 | 179.585 | 6.5 | 0.13 |
| polarC18 | 276.144 | 256.2813 | 6.5 | 0.13 |
| polarC18 | 276.144 | 54.0592 | 6.5 | 0.13 |
| polarC18 | 276.144 | 61.0044 | 6.5 | 0.13 |

| | | | | |
|---|---|---|---|---|
| polarC18 | 276.144 | 87.4977 | 6.5 | 0.13 |
| polarC18 | 276.144 | 115.2442 | 6.5 | 0.13 |
| polarC18 | 276.144 | 55.7721 | 6.5 | 0.13 |
| polarC18 | 276.144 | 256.8878 | 6.5 | 0.13 |
| polarC18 | 276.144 | 53.6923 | 6.5 | 0.13 |
| polarC18 | 276.144 | 173.581 | 6.5 | 0.13 |
| polarC18 | 276.144 | 55.8548 | 6.5 | 0.13 |
| polarC18 | 276.144 | 56.6556 | 6.5 | 0.13 |
| polarC18 | 276.144 | 98.7451 | 6.5 | 0.13 |
| polarC18 | 276.144 | 87.1266 | 6.5 | 0.13 |
| polarC18 | 276.144 | 87.2401 | 6.5 | 0.13 |
| polarC18 | 276.144 | 64.0476 | 6.5 | 0.13 |
| polarC18 | 276.144 | 210.5706 | 6.5 | 0.13 |
| polarC18 | 276.144 | 181.3311 | 6.5 | 0.13 |
| polarC18 | 276.144 | 108.0154 | 6.5 | 0.13 |
| polarC18 | 276.144 | 104.9001 | 6.5 | 0.13 |
| polarC18 | 276.144 | 124.2067 | 6.5 | 0.13 |
| polarC18 | 276.144 | 286.1439 | 6.5 | 0.13 |
| polarC18 | 276.144 | 73.0086 | 6.5 | 0.13 |
| polarC18 | 276.144 | 137.7728 | 6.5 | 0.13 |
| polarC18 | 276.144 | 111.0552 | 6.5 | 0.13 |
| polarC18 | 276.144 | 58.0555 | 6.5 | 0.13 |
| polarC18 | 276.144 | 83.5678 | 6.5 | 0.13 |
| polarC18 | 276.144 | 221.0615 | 6.5 | 0.13 |
| polarC18 | 276.144 | 84.9476 | 6.5 | 0.13 |
| polarC18 | 276.144 | 84.2061 | 6.5 | 0.13 |
| polarC18 | 276.144 | 65.5578 | 6.5 | 0.13 |
| polarC18 | 276.144 | 206.3758 | 6.5 | 0.13 |
| polarC18 | 276.144 | 59.539 | 6.5 | 0.13 |
| polarC18 | 276.144 | 60.9148 | 6.5 | 0.13 |
| polarC18 | 276.144 | 244.9164 | 6.5 | 0.13 |
| polarC18 | 276.144 | 50.9105 | 6.5 | 0.13 |
| polarC18 | 276.144 | 257.6105 | 6.5 | 0.13 |
| polarC18 | 276.144 | 79.145 | 6.5 | 0.13 |
| polarC18 | 276.144 | 101.4141 | 6.5 | 0.13 |
| polarC18 | 276.144 | 70.1748 | 6.5 | 0.13 |
| polarC18 | 276.144 | 188.6071 | 6.5 | 0.13 |
| polarC18 | 276.144 | 183.7265 | 6.5 | 0.12 |
| polarC18 | 276.144 | 95.0273 | 6.5 | 0.12 |
| polarC18 | 276.144 | 121.0592 | 6.5 | 0.12 |
| polarC18 | 276.144 | 183.168 | 6.5 | 0.12 |
| polarC18 | 276.144 | 174.28 | 6.5 | 0.12 |

| | | | | |
|---|---|---|---|---|
| polarC18 | 276.144 | 108.923 | 6.5 | 0.12 |
| polarC18 | 276.144 | 153.5112 | 6.5 | 0.12 |
| polarC18 | 276.144 | 51.93 | 6.5 | 0.12 |
| polarC18 | 276.144 | 264.8214 | 6.5 | 0.12 |
| polarC18 | 276.144 | 89.0017 | 6.5 | 0.12 |
| polarC18 | 276.144 | 255.5383 | 6.5 | 0.12 |
| polarC18 | 276.144 | 57.5569 | 6.5 | 0.12 |
| polarC18 | 276.144 | 143.4514 | 6.5 | 0.12 |
| polarC18 | 276.144 | 235.4246 | 6.5 | 0.12 |
| polarC18 | 276.144 | 59.4955 | 6.5 | 0.12 |
| polarC18 | 276.144 | 62.0325 | 6.5 | 0.12 |
| polarC18 | 276.144 | 99.1728 | 6.5 | 0.12 |
| polarC18 | 276.144 | 170.2492 | 6.5 | 0.12 |
| polarC18 | 276.144 | 50.1145 | 6.5 | 0.12 |
| polarC18 | 276.144 | 281.803 | 6.5 | 0.12 |
| polarC18 | 276.144 | 74.4303 | 6.5 | 0.12 |
| polarC18 | 276.144 | 293.0396 | 6.5 | 0.12 |
| polarC18 | 276.144 | 53.1793 | 6.5 | 0.12 |
| polarC18 | 276.144 | 145.2731 | 6.5 | 0.12 |
| polarC18 | 276.144 | 145.8997 | 6.5 | 0.12 |
| polarC18 | 276.144 | 204.5348 | 6.5 | 0.12 |
| polarC18 | 276.144 | 68.9063 | 6.5 | 0.12 |
| polarC18 | 276.144 | 137.1161 | 6.5 | 0.12 |
| polarC18 | 276.144 | 118.8229 | 6.5 | 0.12 |
| polarC18 | 276.144 | 130.3399 | 6.5 | 0.12 |
| polarC18 | 276.144 | 54.3765 | 6.5 | 0.12 |
| polarC18 | 276.144 | 106.9179 | 6.5 | 0.12 |
| polarC18 | 276.144 | 155.3356 | 6.5 | 0.12 |
| polarC18 | 276.144 | 93.5256 | 6.5 | 0.12 |
| polarC18 | 276.144 | 76.1787 | 6.5 | 0.12 |
| polarC18 | 276.144 | 74.9393 | 6.5 | 0.12 |
| polarC18 | 276.144 | 54.7591 | 6.5 | 0.12 |
| polarC18 | 276.144 | 136.3202 | 6.5 | 0.12 |
| polarC18 | 276.144 | 143.2493 | 6.5 | 0.12 |
| polarC18 | 276.144 | 101.7696 | 6.5 | 0.12 |
| polarC18 | 276.144 | 200.9688 | 6.5 | 0.12 |
| polarC18 | 276.144 | 57.3303 | 6.5 | 0.12 |
| polarC18 | 276.144 | 240.2807 | 6.5 | 0.12 |
| polarC18 | 276.144 | 267.3598 | 6.5 | 0.12 |
| polarC18 | 276.144 | 58.926 | 6.5 | 0.12 |
| polarC18 | 276.144 | 210.6109 | 6.5 | 0.12 |
| polarC18 | 276.144 | 80.4839 | 6.5 | 0.12 |

| | | | | |
|---|---|---|---|---|
| polarC18 | 276.144 | 55.8488 | 6.5 | 0.12 |
| polarC18 | 276.144 | 289.8223 | 6.5 | 0.12 |
| polarC18 | 276.144 | 224.3298 | 6.5 | 0.12 |
| polarC18 | 276.144 | 103.6059 | 6.5 | 0.12 |
| polarC18 | 276.144 | 273.205 | 6.5 | 0.12 |
| polarC18 | 276.144 | 83.5376 | 6.5 | 0.12 |
| polarC18 | 276.144 | 251.3452 | 6.5 | 0.12 |
| polarC18 | 276.144 | 109.7712 | 6.5 | 0.12 |
| polarC18 | 276.144 | 61.3523 | 6.5 | 0.12 |
| polarC18 | 276.144 | 192.8472 | 6.5 | 0.12 |
| polarC18 | 276.144 | 75.3223 | 6.5 | 0.12 |
| polarC18 | 276.144 | 120.3288 | 6.5 | 0.12 |
| polarC18 | 276.144 | 113.3206 | 6.5 | 0.12 |
| polarC18 | 276.144 | 59.5992 | 6.5 | 0.12 |
| polarC18 | 276.144 | 88.1887 | 6.5 | 0.12 |
| polarC18 | 276.144 | 131.9918 | 6.5 | 0.12 |
| polarC18 | 276.144 | 130.1828 | 6.5 | 0.12 |
| polarC18 | 276.144 | 59.2614 | 6.5 | 0.12 |
| polarC18 | 276.144 | 200.65 | 6.5 | 0.12 |
| polarC18 | 276.144 | 79.5835 | 6.5 | 0.12 |
| polarC18 | 276.144 | 97.7001 | 6.5 | 0.12 |
| polarC18 | 276.144 | 53.1003 | 6.5 | 0.12 |
| polarC18 | 276.144 | 214.0778 | 6.5 | 0.12 |
| polarC18 | 276.144 | 73.7541 | 6.5 | 0.12 |
| polarC18 | 276.144 | 87.3396 | 6.5 | 0.12 |
| polarC18 | 276.144 | 139.5797 | 6.5 | 0.12 |
| polarC18 | 276.144 | 75.9717 | 6.5 | 0.12 |
| polarC18 | 276.144 | 76.0794 | 6.5 | 0.12 |
| polarC18 | 276.144 | 130.5806 | 6.5 | 0.12 |
| polarC18 | 276.144 | 179.6359 | 6.5 | 0.12 |
| polarC18 | 276.144 | 64.1672 | 6.5 | 0.12 |
| polarC18 | 276.144 | 103.6259 | 6.5 | 0.12 |
| polarC18 | 276.144 | 67.3427 | 6.5 | 0.12 |
| polarC18 | 276.144 | 114.0818 | 6.5 | 0.12 |
| polarC18 | 276.144 | 289.1236 | 6.5 | 0.12 |
| polarC18 | 276.144 | 68.6156 | 6.5 | 0.12 |
| polarC18 | 276.144 | 64.4922 | 6.5 | 0.12 |
| polarC18 | 276.144 | 103.4377 | 6.5 | 0.12 |
| polarC18 | 276.144 | 104.8498 | 6.5 | 0.12 |
| polarC18 | 276.144 | 54.6359 | 6.5 | 0.12 |
| polarC18 | 276.144 | 65.1879 | 6.5 | 0.12 |
| polarC18 | 276.144 | 275.3156 | 6.5 | 0.12 |

| | | | | |
|---|---|---|---|---|
| polarC18 | 276.144 | 203.6134 | 6.5 | 0.12 |
| polarC18 | 276.144 | 55.2722 | 6.5 | 0.12 |
| polarC18 | 276.144 | 58.0105 | 6.5 | 0.12 |
| polarC18 | 276.144 | 90.8046 | 6.5 | 0.12 |
| polarC18 | 276.144 | 54.107 | 6.5 | 0.12 |
| polarC18 | 276.144 | 78.2007 | 6.5 | 0.12 |
| polarC18 | 276.144 | 62.8362 | 6.5 | 0.12 |
| polarC18 | 276.144 | 114.2282 | 6.5 | 0.12 |
| polarC18 | 276.144 | 77.2877 | 6.5 | 0.12 |
| polarC18 | 276.144 | 143.1879 | 6.5 | 0.12 |
| polarC18 | 276.144 | 256.4326 | 6.5 | 0.12 |
| polarC18 | 276.144 | 274.8718 | 6.5 | 0.12 |
| polarC18 | 276.144 | 112.6818 | 6.5 | 0.12 |
| polarC18 | 276.144 | 75.1468 | 6.5 | 0.12 |
| polarC18 | 276.144 | 114.4423 | 6.5 | 0.12 |
| polarC18 | 276.144 | 72.8159 | 6.5 | 0.12 |
| polarC18 | 276.144 | 109.3593 | 6.5 | 0.12 |
| polarC18 | 276.144 | 219.8934 | 6.5 | 0.12 |
| polarC18 | 276.144 | 202.7203 | 6.5 | 0.12 |
| polarC18 | 276.144 | 111.4598 | 6.5 | 0.12 |
| polarC18 | 276.144 | 261.0533 | 6.5 | 0.12 |
| polarC18 | 276.144 | 90.1975 | 6.5 | 0.12 |
| polarC18 | 276.144 | 125.7234 | 6.5 | 0.12 |
| polarC18 | 276.144 | 198.6163 | 6.5 | 0.12 |
| polarC18 | 276.144 | 97.667 | 6.5 | 0.12 |
| polarC18 | 276.144 | 93.4318 | 6.5 | 0.12 |
| polarC18 | 276.144 | 106.8136 | 6.5 | 0.12 |
| polarC18 | 276.144 | 61.2517 | 6.5 | 0.12 |
| polarC18 | 276.144 | 288.6286 | 6.5 | 0.12 |
| polarC18 | 276.144 | 198.0899 | 6.5 | 0.12 |
| polarC18 | 276.144 | 56.5259 | 6.5 | 0.12 |
| polarC18 | 276.144 | 67.2473 | 6.5 | 0.12 |
| polarC18 | 276.144 | 62.8594 | 6.5 | 0.12 |
| polarC18 | 276.144 | 71.9122 | 6.5 | 0.12 |
| polarC18 | 276.144 | 55.2849 | 6.5 | 0.12 |
| polarC18 | 276.144 | 112.9979 | 6.5 | 0.12 |
| polarC18 | 276.144 | 183.6872 | 6.5 | 0.12 |
| polarC18 | 276.144 | 52.3206 | 6.5 | 0.12 |
| polarC18 | 276.144 | 142.8676 | 6.5 | 0.12 |
| polarC18 | 276.144 | 90.3141 | 6.5 | 0.12 |
| polarC18 | 276.144 | 189.2617 | 6.5 | 0.12 |
| polarC18 | 276.144 | 66.1763 | 6.5 | 0.12 |

| | | | | |
|---|---|---|---|---|
| polarC18 | 276.144 | 69.8611 | 6.5 | 0.12 |
| polarC18 | 276.144 | 120.8834 | 6.5 | 0.12 |
| polarC18 | 276.144 | 105.0782 | 6.5 | 0.12 |
| polarC18 | 276.144 | 72.7116 | 6.5 | 0.12 |
| polarC18 | 276.144 | 119.2836 | 6.5 | 0.12 |
| polarC18 | 276.144 | 241.0839 | 6.5 | 0.12 |
| polarC18 | 276.144 | 115.5182 | 6.5 | 0.12 |
| polarC18 | 276.144 | 52.7264 | 6.5 | 0.12 |
| polarC18 | 276.144 | 51.656 | 6.5 | 0.12 |
| polarC18 | 276.144 | 51.0001 | 6.5 | 0.12 |
| polarC18 | 276.144 | 72.2182 | 6.5 | 0.12 |
| polarC18 | 276.144 | 128.608 | 6.5 | 0.12 |
| polarC18 | 276.144 | 84.1706 | 6.5 | 0.12 |
| polarC18 | 276.144 | 164.3313 | 6.5 | 0.12 |
| polarC18 | 276.144 | 62.233 | 6.5 | 0.12 |
| polarC18 | 276.144 | 75.0415 | 6.5 | 0.12 |
| polarC18 | 276.144 | 103.8579 | 6.5 | 0.12 |
| polarC18 | 276.144 | 71.3154 | 6.5 | 0.12 |
| polarC18 | 276.144 | 117.591 | 6.5 | 0.12 |
| polarC18 | 276.144 | 50.4347 | 6.5 | 0.12 |
| polarC18 | 276.144 | 90.8118 | 6.5 | 0.12 |
| polarC18 | 276.144 | 87.0924 | 6.5 | 0.12 |
| polarC18 | 276.144 | 60.7206 | 6.5 | 0.12 |
| polarC18 | 276.144 | 126.8317 | 6.5 | 0.12 |
| polarC18 | 276.144 | 170.8932 | 6.5 | 0.12 |
| polarC18 | 276.144 | 262.3305 | 6.5 | 0.12 |
| polarC18 | 276.144 | 167.6008 | 6.5 | 0.12 |
| polarC18 | 276.144 | 55.9349 | 6.5 | 0.12 |
| polarC18 | 276.144 | 115.6254 | 6.5 | 0.12 |
| polarC18 | 276.144 | 55.0755 | 6.5 | 0.12 |
| polarC18 | 276.144 | 51.355 | 6.5 | 0.12 |
| polarC18 | 276.144 | 88.6035 | 6.5 | 0.12 |
| polarC18 | 276.144 | 54.3918 | 6.5 | 0.12 |
| polarC18 | 276.144 | 155.771 | 6.5 | 0.12 |
| polarC18 | 276.144 | 52.621 | 6.5 | 0.12 |
| polarC18 | 276.144 | 51.1227 | 6.5 | 0.12 |
| polarC18 | 276.144 | 50.6553 | 6.5 | 0.12 |
| polarC18 | 276.144 | 99.6229 | 6.5 | 0.12 |
| polarC18 | 276.144 | 56.2893 | 6.5 | 0.12 |
| polarC18 | 276.144 | 71.8177 | 6.5 | 0.12 |
| polarC18 | 276.144 | 51.0461 | 6.5 | 0.12 |
| polarC18 | 276.144 | 112.2419 | 6.5 | 0.12 |

| | | | | |
|---|---|---|---|---|
| polarC18 | 276.144 | 134.3221 | 6.5 | 0.12 |
| polarC18 | 276.144 | 120.4223 | 6.5 | 0.12 |
| polarC18 | 276.144 | 137.5335 | 6.5 | 0.12 |
| polarC18 | 276.144 | 158.3848 | 6.5 | 0.12 |
| polarC18 | 276.144 | 62.5958 | 6.5 | 0.12 |
| polarC18 | 276.144 | 132.6217 | 6.5 | 0.12 |
| polarC18 | 276.144 | 55.094 | 6.5 | 0.12 |
| polarC18 | 276.144 | 53.4799 | 6.5 | 0.12 |
| polarC18 | 276.144 | 91.5214 | 6.5 | 0.12 |
| polarC18 | 276.144 | 101.1402 | 6.5 | 0.12 |
| polarC18 | 276.144 | 79.6509 | 6.5 | 0.12 |
| polarC18 | 276.144 | 108.5408 | 6.5 | 0.12 |
| polarC18 | 276.144 | 118.5176 | 6.5 | 0.12 |
| polarC18 | 276.144 | 99.058 | 6.5 | 0.12 |
| polarC18 | 276.144 | 98.5583 | 6.5 | 0.12 |
| polarC18 | 276.144 | 85.2684 | 6.5 | 0.12 |
| polarC18 | 276.144 | 91.7188 | 6.5 | 0.12 |
| polarC18 | 276.144 | 60.3858 | 6.5 | 0.12 |
| polarC18 | 276.144 | 94.9629 | 6.5 | 0.12 |
| polarC18 | 276.144 | 192.0199 | 6.4 | 0.12 |
| polarC18 | 276.144 | 187.5015 | 6.4 | 0.12 |
| polarC18 | 276.144 | 212.7294 | 6.4 | 0.12 |
| polarC18 | 276.144 | 131.5205 | 6.4 | 0.12 |
| polarC18 | 276.144 | 75.5433 | 6.4 | 0.12 |
| polarC18 | 276.144 | 73.0206 | 6.4 | 0.12 |
| polarC18 | 276.144 | 80.493 | 6.4 | 0.12 |
| polarC18 | 276.144 | 154.3527 | 6.4 | 0.12 |
| polarC18 | 276.144 | 80.1647 | 6.4 | 0.12 |
| polarC18 | 276.144 | 238.6568 | 6.4 | 0.12 |
| polarC18 | 276.144 | 66.3745 | 6.4 | 0.12 |
| polarC18 | 276.144 | 204.482 | 6.4 | 0.12 |
| polarC18 | 276.144 | 293.2875 | 6.4 | 0.12 |
| polarC18 | 276.144 | 51.0021 | 6.4 | 0.12 |
| polarC18 | 276.144 | 62.0465 | 6.4 | 0.12 |
| polarC18 | 276.144 | 243.7522 | 6.4 | 0.12 |
| polarC18 | 276.144 | 50.4879 | 6.4 | 0.12 |
| polarC18 | 276.144 | 74.5313 | 6.4 | 0.12 |
| polarC18 | 276.144 | 80.3026 | 6.4 | 0.12 |
| polarC18 | 276.144 | 50.3797 | 6.4 | 0.12 |
| polarC18 | 276.144 | 55.9185 | 6.4 | 0.12 |
| polarC18 | 276.144 | 65.2823 | 6.4 | 0.12 |
| polarC18 | 276.144 | 54.7252 | 6.4 | 0.12 |

| | | | | |
|---|---|---|---|---|
| polarC18 | 276.144 | 171.216 | 6.4 | 0.12 |
| polarC18 | 276.144 | 221.1411 | 6.4 | 0.12 |
| polarC18 | 276.144 | 54.4767 | 6.4 | 0.12 |
| polarC18 | 276.144 | 96.9065 | 6.4 | 0.12 |
| polarC18 | 276.144 | 134.8304 | 6.4 | 0.12 |
| polarC18 | 276.144 | 55.6628 | 6.4 | 0.12 |
| polarC18 | 276.144 | 56.1148 | 6.4 | 0.12 |
| polarC18 | 276.144 | 75.5931 | 6.4 | 0.12 |
| polarC18 | 276.144 | 188.2655 | 6.4 | 0.12 |
| polarC18 | 276.144 | 237.2234 | 6.4 | 0.12 |
| polarC18 | 276.144 | 104.5206 | 6.4 | 0.12 |
| polarC18 | 276.144 | 89.4508 | 6.4 | 0.12 |
| polarC18 | 276.144 | 110.638 | 6.4 | 0.12 |
| polarC18 | 276.144 | 62.9583 | 6.4 | 0.12 |
| polarC18 | 276.144 | 66.8157 | 6.4 | 0.12 |
| polarC18 | 276.144 | 50.2828 | 6.4 | 0.12 |
| polarC18 | 276.144 | 52.3471 | 6.4 | 0.12 |
| polarC18 | 276.144 | 189.5531 | 6.4 | 0.12 |
| polarC18 | 276.144 | 52.5568 | 6.4 | 0.12 |
| polarC18 | 276.144 | 126.7906 | 6.4 | 0.12 |
| polarC18 | 276.144 | 119.4496 | 6.4 | 0.12 |
| polarC18 | 276.144 | 75.7692 | 6.4 | 0.12 |
| polarC18 | 276.144 | 244.4183 | 6.4 | 0.12 |
| polarC18 | 276.144 | 56.976 | 6.4 | 0.12 |
| polarC18 | 276.144 | 52.7668 | 6.4 | 0.12 |
| polarC18 | 276.144 | 183.8248 | 6.4 | 0.12 |
| polarC18 | 276.144 | 111.0668 | 6.4 | 0.12 |
| polarC18 | 276.144 | 59.2482 | 6.4 | 0.12 |
| polarC18 | 276.144 | 151.8399 | 6.4 | 0.12 |
| polarC18 | 276.144 | 55.5792 | 6.4 | 0.12 |
| polarC18 | 276.144 | 138.7188 | 6.4 | 0.12 |
| polarC18 | 276.144 | 155.4449 | 6.4 | 0.12 |
| polarC18 | 276.144 | 62.1952 | 6.4 | 0.12 |
| polarC18 | 276.144 | 51.5417 | 6.4 | 0.12 |
| polarC18 | 276.144 | 61.4749 | 6.4 | 0.12 |
| polarC18 | 276.144 | 101.4204 | 6.4 | 0.12 |
| polarC18 | 276.144 | 72.1248 | 6.4 | 0.12 |
| polarC18 | 276.144 | 214.021 | 6.4 | 0.12 |
| polarC18 | 276.144 | 53.0402 | 6.4 | 0.12 |
| polarC18 | 276.144 | 90.7391 | 6.4 | 0.12 |
| polarC18 | 276.144 | 50.2349 | 6.4 | 0.12 |
| polarC18 | 276.144 | 51.8989 | 6.4 | 0.12 |

| | | | | |
|---|---|---|---|---|
| polarC18 | 276.144 | 87.8991 | 6.4 | 0.12 |
| polarC18 | 276.144 | 50.5885 | 6.4 | 0.12 |
| polarC18 | 276.144 | 53.1305 | 6.4 | 0.12 |
| polarC18 | 276.144 | 51.6291 | 6.4 | 0.12 |
| polarC18 | 276.144 | 66.6775 | 6.4 | 0.12 |
| polarC18 | 276.144 | 108.2396 | 6.4 | 0.12 |
| polarC18 | 276.144 | 60.4312 | 6.4 | 0.12 |
| polarC18 | 276.144 | 251.2727 | 6.4 | 0.12 |
| polarC18 | 276.144 | 140.8817 | 6.4 | 0.12 |
| polarC18 | 276.144 | 67.5525 | 6.4 | 0.12 |
| polarC18 | 276.144 | 102.6006 | 6.4 | 0.12 |
| polarC18 | 276.144 | 55.19 | 6.4 | 0.12 |
| polarC18 | 276.144 | 93.6199 | 6.4 | 0.12 |
| polarC18 | 276.144 | 246.7961 | 6.4 | 0.12 |
| polarC18 | 276.144 | 82.7953 | 6.4 | 0.12 |
| polarC18 | 276.144 | 118.4652 | 6.4 | 0.12 |
| polarC18 | 276.144 | 84.2115 | 6.4 | 0.12 |
| polarC18 | 276.144 | 82.6562 | 6.4 | 0.12 |
| polarC18 | 276.144 | 75.47 | 6.4 | 0.12 |
| polarC18 | 276.144 | 180.2691 | 6.4 | 0.12 |
| polarC18 | 276.144 | 70.9613 | 6.4 | 0.12 |
| polarC18 | 276.144 | 80.4536 | 6.4 | 0.12 |
| polarC18 | 276.144 | 77.0777 | 6.4 | 0.12 |
| polarC18 | 276.144 | 208.9556 | 6.4 | 0.12 |
| polarC18 | 276.144 | 124.4717 | 6.4 | 0.12 |
| polarC18 | 276.144 | 63.9768 | 6.4 | 0.12 |
| polarC18 | 276.144 | 69.3459 | 6.4 | 0.12 |
| polarC18 | 276.144 | 75.3411 | 6.4 | 0.12 |
| polarC18 | 276.144 | 143.4825 | 6.4 | 0.12 |
| polarC18 | 276.144 | 113.7749 | 6.4 | 0.12 |
| polarC18 | 276.144 | 60.7638 | 6.4 | 0.12 |
| polarC18 | 276.144 | 82.1657 | 6.4 | 0.12 |
| polarC18 | 276.144 | 59.0065 | 6.4 | 0.12 |
| polarC18 | 276.144 | 192.6038 | 6.4 | 0.12 |
| polarC18 | 276.144 | 97.0786 | 6.4 | 0.12 |
| polarC18 | 276.144 | 75.4577 | 6.4 | 0.12 |
| polarC18 | 276.144 | 210.8577 | 6.4 | 0.12 |
| polarC18 | 276.144 | 50.472 | 6.4 | 0.12 |
| polarC18 | 276.144 | 101.9683 | 6.4 | 0.12 |
| polarC18 | 276.144 | 111.1186 | 6.4 | 0.12 |
| polarC18 | 276.144 | 74.6706 | 6.4 | 0.12 |
| polarC18 | 276.144 | 55.8273 | 6.4 | 0.12 |

| | | | | |
|---|---|---|---|---|
| polarC18 | 276.144 | 63.9651 | 6.4 | 0.12 |
| polarC18 | 276.144 | 161.3251 | 6.4 | 0.12 |
| polarC18 | 276.144 | 87.918 | 6.4 | 0.12 |
| polarC18 | 276.144 | 177.668 | 6.4 | 0.12 |
| polarC18 | 276.144 | 77.7842 | 6.4 | 0.12 |
| polarC18 | 276.144 | 59.7164 | 6.4 | 0.12 |
| polarC18 | 276.144 | 113.0703 | 6.4 | 0.12 |
| polarC18 | 276.144 | 62.5211 | 6.4 | 0.12 |
| polarC18 | 276.144 | 58.6334 | 6.4 | 0.12 |
| polarC18 | 276.144 | 61.2777 | 6.4 | 0.12 |
| polarC18 | 276.144 | 107.4061 | 6.4 | 0.12 |
| polarC18 | 276.144 | 274.9896 | 6.4 | 0.12 |
| polarC18 | 276.144 | 133.8211 | 6.4 | 0.12 |
| polarC18 | 276.144 | 163.8112 | 6.4 | 0.12 |
| polarC18 | 276.144 | 78.1692 | 6.4 | 0.12 |
| polarC18 | 276.144 | 87.9426 | 6.4 | 0.12 |
| polarC18 | 276.144 | 121.1028 | 6.4 | 0.12 |
| polarC18 | 276.144 | 88.4497 | 6.4 | 0.12 |
| polarC18 | 276.144 | 76.6382 | 6.4 | 0.12 |
| polarC18 | 276.144 | 69.0234 | 6.4 | 0.12 |
| polarC18 | 276.144 | 73.0019 | 6.4 | 0.12 |
| polarC18 | 276.144 | 59.4778 | 6.4 | 0.12 |
| polarC18 | 276.144 | 82.4034 | 6.4 | 0.12 |
| polarC18 | 276.144 | 52.9195 | 6.4 | 0.12 |
| polarC18 | 276.144 | 76.3194 | 6.4 | 0.12 |
| polarC18 | 276.144 | 80.7398 | 6.4 | 0.12 |
| polarC18 | 276.144 | 55.3904 | 6.4 | 0.12 |
| polarC18 | 276.144 | 202.6268 | 6.4 | 0.12 |
| polarC18 | 276.144 | 108.9791 | 6.4 | 0.12 |
| polarC18 | 276.144 | 141.026 | 6.4 | 0.12 |
| polarC18 | 276.144 | 60.1268 | 6.4 | 0.12 |
| polarC18 | 276.144 | 65.0472 | 6.4 | 0.12 |
| polarC18 | 276.144 | 64.1361 | 6.4 | 0.12 |
| polarC18 | 276.144 | 66.3895 | 6.4 | 0.12 |
| polarC18 | 276.144 | 60.7472 | 6.4 | 0.12 |
| polarC18 | 276.144 | 58.2569 | 6.4 | 0.12 |
| polarC18 | 276.144 | 65.034 | 6.4 | 0.12 |
| polarC18 | 276.144 | 76.4059 | 6.4 | 0.12 |
| polarC18 | 276.144 | 51.4077 | 6.4 | 0.12 |
| polarC18 | 276.144 | 278.457 | 6.4 | 0.12 |
| polarC18 | 276.144 | 263.7439 | 6.4 | 0.12 |
| polarC18 | 276.144 | 118.301 | 6.4 | 0.12 |

| | | | | |
|---|---|---|---|---|
| polarC18 | 276.144 | 162.5232 | 6.4 | 0.12 |
| polarC18 | 276.144 | 92.4791 | 6.4 | 0.12 |
| polarC18 | 276.144 | 55.874 | 6.4 | 0.12 |
| polarC18 | 276.144 | 51.1645 | 6.4 | 0.12 |
| polarC18 | 276.144 | 59.0494 | 6.4 | 0.12 |
| polarC18 | 276.144 | 78.4296 | 6.4 | 0.12 |
| polarC18 | 276.144 | 145.9519 | 6.4 | 0.12 |
| polarC18 | 276.144 | 204.2783 | 6.4 | 0.12 |
| polarC18 | 276.144 | 57.4142 | 6.4 | 0.12 |
| polarC18 | 276.144 | 121.7098 | 6.4 | 0.12 |
| polarC18 | 276.144 | 71.5435 | 6.4 | 0.12 |
| polarC18 | 276.144 | 159.9781 | 6.4 | 0.12 |
| polarC18 | 276.144 | 123.0816 | 6.4 | 0.12 |
| polarC18 | 276.144 | 130.8398 | 6.4 | 0.12 |
| polarC18 | 276.144 | 138.2155 | 6.4 | 0.12 |
| polarC18 | 276.144 | 167.9424 | 6.4 | 0.12 |
| polarC18 | 276.144 | 51.6899 | 6.4 | 0.12 |
| polarC18 | 276.144 | 77.1886 | 6.4 | 0.12 |
| polarC18 | 276.144 | 76.4251 | 6.4 | 0.12 |
| polarC18 | 276.144 | 55.7389 | 6.4 | 0.12 |
| polarC18 | 276.144 | 63.4603 | 6.4 | 0.12 |
| polarC18 | 276.144 | 119.7417 | 6.4 | 0.12 |
| polarC18 | 276.144 | 53.7864 | 6.4 | 0.12 |
| polarC18 | 276.144 | 78.9852 | 6.4 | 0.12 |
| polarC18 | 276.144 | 68.2188 | 6.4 | 0.12 |
| polarC18 | 276.144 | 295.0567 | 6.4 | 0.12 |
| polarC18 | 276.144 | 84.0096 | 6.4 | 0.12 |
| polarC18 | 276.144 | 185.9946 | 6.4 | 0.12 |
| polarC18 | 276.144 | 78.651 | 6.4 | 0.12 |
| polarC18 | 276.144 | 63.8125 | 6.4 | 0.12 |
| polarC18 | 276.144 | 83.8977 | 6.4 | 0.12 |
| polarC18 | 276.144 | 129.4554 | 6.4 | 0.12 |
| polarC18 | 276.144 | 66.2106 | 6.4 | 0.12 |
| polarC18 | 276.144 | 112.6975 | 6.4 | 0.12 |
| polarC18 | 276.144 | 72.0792 | 6.4 | 0.12 |
| polarC18 | 276.144 | 55.3135 | 6.4 | 0.12 |
| polarC18 | 276.144 | 167.1246 | 6.4 | 0.12 |
| polarC18 | 276.144 | 110.0826 | 6.4 | 0.12 |
| polarC18 | 276.144 | 57.7474 | 6.4 | 0.12 |
| polarC18 | 276.144 | 135.0758 | 6.4 | 0.12 |
| polarC18 | 276.144 | 104.8613 | 6.4 | 0.12 |
| polarC18 | 276.144 | 92.0564 | 6.4 | 0.12 |

| | | | | |
|---|---|---|---|---|
| polarC18 | 276.144 | 53.4202 | 6.4 | 0.12 |
| polarC18 | 276.144 | 60.8873 | 6.4 | 0.12 |
| polarC18 | 276.144 | 54.1251 | 6.4 | 0.12 |
| polarC18 | 276.144 | 155.1733 | 6.4 | 0.12 |
| polarC18 | 276.144 | 96.9281 | 6.4 | 0.12 |
| polarC18 | 276.144 | 155.2919 | 6.4 | 0.12 |
| polarC18 | 276.144 | 129.3928 | 6.4 | 0.12 |
| polarC18 | 276.144 | 95.5311 | 6.4 | 0.12 |
| polarC18 | 276.144 | 77.1176 | 6.4 | 0.12 |
| polarC18 | 276.144 | 291.1888 | 6.4 | 0.12 |
| polarC18 | 276.144 | 61.7687 | 6.4 | 0.12 |
| polarC18 | 276.144 | 93.016 | 6.4 | 0.12 |
| polarC18 | 276.144 | 85.3831 | 6.4 | 0.12 |
| polarC18 | 276.144 | 68.2607 | 6.4 | 0.12 |
| polarC18 | 276.144 | 93.1815 | 6.4 | 0.12 |
| polarC18 | 276.144 | 77.9976 | 6.4 | 0.12 |
| polarC18 | 276.144 | 81.8762 | 6.4 | 0.12 |
| polarC18 | 276.144 | 114.2572 | 6.4 | 0.12 |
| polarC18 | 276.144 | 65.8787 | 6.4 | 0.12 |
| polarC18 | 276.144 | 116.9957 | 6.4 | 0.12 |
| polarC18 | 276.144 | 87.107 | 6.4 | 0.12 |
| polarC18 | 276.144 | 301.3546 | 6.4 | 0.12 |
| polarC18 | 276.144 | 69.4326 | 6.4 | 0.12 |
| polarC18 | 276.144 | 57.1695 | 6.4 | 0.12 |
| polarC18 | 276.144 | 175.7334 | 6.4 | 0.12 |
| polarC18 | 276.144 | 114.8594 | 6.4 | 0.12 |
| polarC18 | 276.144 | 59.4288 | 6.4 | 0.12 |
| polarC18 | 276.144 | 81.8139 | 6.4 | 0.12 |
| polarC18 | 276.144 | 177.1858 | 6.4 | 0.12 |
| polarC18 | 276.144 | 88.8314 | 6.4 | 0.12 |
| polarC18 | 276.144 | 69.7688 | 6.4 | 0.12 |
| polarC18 | 276.144 | 86.5581 | 6.4 | 0.12 |
| polarC18 | 276.144 | 223.3675 | 6.4 | 0.12 |
| polarC18 | 276.144 | 57.1165 | 6.4 | 0.12 |
| polarC18 | 276.144 | 295.3604 | 6.4 | 0.12 |
| polarC18 | 276.144 | 78.1599 | 6.4 | 0.12 |
| polarC18 | 276.144 | 70.7805 | 6.4 | 0.12 |
| polarC18 | 276.144 | 109.3075 | 6.4 | 0.12 |
| polarC18 | 276.144 | 91.2954 | 6.4 | 0.12 |
| polarC18 | 276.144 | 107.0093 | 6.4 | 0.12 |
| polarC18 | 276.144 | 74.5755 | 6.4 | 0.12 |
| polarC18 | 276.144 | 98.3222 | 6.4 | 0.12 |

| | | | | |
|---|---|---|---|---|
| polarC18 | 276.144 | 79.523 | 6.4 | 0.12 |
| polarC18 | 276.144 | 132.0713 | 6.4 | 0.12 |
| polarC18 | 276.144 | 95.3497 | 6.4 | 0.12 |
| polarC18 | 276.144 | 133.974 | 6.4 | 0.12 |
| polarC18 | 276.144 | 64.6316 | 6.4 | 0.12 |
| polarC18 | 276.144 | 145.3341 | 6.4 | 0.12 |
| polarC18 | 276.144 | 51.6186 | 6.4 | 0.12 |
| polarC18 | 276.144 | 129.6777 | 6.4 | 0.12 |
| polarC18 | 276.144 | 102.38 | 6.4 | 0.12 |
| polarC18 | 276.144 | 55.0499 | 6.4 | 0.12 |
| polarC18 | 276.144 | 101.7573 | 6.4 | 0.12 |
| polarC18 | 276.144 | 71.3264 | 6.4 | 0.12 |
| polarC18 | 276.144 | 69.6402 | 6.4 | 0.12 |
| polarC18 | 276.144 | 153.4796 | 6.4 | 0.12 |
| polarC18 | 276.144 | 155.7198 | 6.4 | 0.12 |
| polarC18 | 276.144 | 52.2005 | 6.4 | 0.12 |
| polarC18 | 276.144 | 68.4665 | 6.4 | 0.12 |
| polarC18 | 276.144 | 256.7114 | 6.4 | 0.12 |
| polarC18 | 276.144 | 57.8243 | 6.4 | 0.12 |
| polarC18 | 276.144 | 73.3597 | 6.4 | 0.12 |
| polarC18 | 276.144 | 94.9097 | 6.4 | 0.12 |
| polarC18 | 276.144 | 95.4992 | 6.4 | 0.12 |
| polarC18 | 276.144 | 96.1225 | 6.4 | 0.12 |
| polarC18 | 276.144 | 236.5602 | 6.4 | 0.12 |
| polarC18 | 276.144 | 60.2081 | 6.4 | 0.12 |
| polarC18 | 276.144 | 168.3562 | 6.4 | 0.12 |
| polarC18 | 276.144 | 50.06 | 6.4 | 0.12 |
| polarC18 | 276.144 | 67.5434 | 6.4 | 0.12 |
| polarC18 | 276.144 | 289.3353 | 6.4 | 0.12 |
| polarC18 | 276.144 | 67.7805 | 6.4 | 0.12 |
| polarC18 | 276.144 | 65.3131 | 6.4 | 0.12 |
| polarC18 | 276.144 | 118.3197 | 6.4 | 0.12 |
| polarC18 | 276.144 | 68.7796 | 6.4 | 0.12 |
| polarC18 | 276.144 | 58.203 | 6.4 | 0.12 |
| polarC18 | 276.144 | 68.7065 | 6.4 | 0.12 |
| polarC18 | 276.144 | 84.7823 | 6.4 | 0.12 |
| polarC18 | 276.144 | 216.1147 | 6.4 | 0.12 |
| polarC18 | 276.144 | 55.0332 | 6.4 | 0.12 |
| polarC18 | 276.144 | 71.8676 | 6.4 | 0.12 |
| polarC18 | 276.144 | 95.4695 | 6.4 | 0.12 |
| polarC18 | 276.144 | 57.4361 | 6.4 | 0.12 |
| polarC18 | 276.144 | 68.8106 | 6.4 | 0.12 |

| | | | | |
|---|---|---|---|---|
| polarC18 | 276.144 | 57.8993 | 6.4 | 0.12 |
| polarC18 | 276.144 | 60.9757 | 6.4 | 0.12 |
| polarC18 | 276.144 | 96.4206 | 6.4 | 0.12 |
| polarC18 | 276.144 | 166.4135 | 6.4 | 0.12 |
| polarC18 | 276.144 | 134.5529 | 6.4 | 0.12 |
| polarC18 | 276.144 | 161.2716 | 6.4 | 0.12 |
| polarC18 | 276.144 | 96.015 | 6.4 | 0.12 |
| polarC18 | 276.144 | 132.42 | 6.4 | 0.12 |
| polarC18 | 276.144 | 70.326 | 6.4 | 0.12 |
| polarC18 | 276.144 | 253.0468 | 6.4 | 0.12 |
| polarC18 | 276.144 | 107.8847 | 6.4 | 0.12 |
| polarC18 | 276.144 | 59.4921 | 6.4 | 0.12 |
| polarC18 | 276.144 | 115.0795 | 6.4 | 0.12 |
| polarC18 | 276.144 | 66.4746 | 6.4 | 0.12 |
| polarC18 | 276.144 | 105.5076 | 6.4 | 0.12 |
| polarC18 | 276.144 | 73.1024 | 6.4 | 0.12 |
| polarC18 | 276.144 | 170.8194 | 6.4 | 0.12 |
| polarC18 | 276.144 | 63.2695 | 6.4 | 0.12 |
| polarC18 | 276.144 | 130.2734 | 6.4 | 0.12 |
| polarC18 | 276.144 | 83.0099 | 6.4 | 0.12 |
| polarC18 | 276.144 | 62.6982 | 6.4 | 0.12 |
| polarC18 | 276.144 | 87.2245 | 6.4 | 0.12 |
| polarC18 | 276.144 | 58.7997 | 6.4 | 0.12 |
| polarC18 | 276.144 | 130.6151 | 6.4 | 0.12 |
| polarC18 | 276.144 | 279.0652 | 6.4 | 0.12 |
| polarC18 | 276.144 | 172.8604 | 6.4 | 0.12 |
| polarC18 | 276.144 | 67.4588 | 6.4 | 0.12 |
| polarC18 | 276.144 | 72.5648 | 6.4 | 0.12 |
| polarC18 | 276.144 | 54.7724 | 6.4 | 0.12 |
| polarC18 | 276.144 | 81.2508 | 6.4 | 0.12 |
| polarC18 | 276.144 | 111.1494 | 6.4 | 0.12 |
| polarC18 | 276.144 | 262.8051 | 6.4 | 0.12 |
| polarC18 | 276.144 | 101.6531 | 6.4 | 0.12 |
| polarC18 | 276.144 | 86.5205 | 6.4 | 0.12 |
| polarC18 | 276.144 | 70.3834 | 6.4 | 0.12 |
| polarC18 | 276.144 | 55.4914 | 6.4 | 0.12 |
| polarC18 | 276.144 | 52.8909 | 6.4 | 0.12 |
| polarC18 | 276.144 | 129.8889 | 6.4 | 0.12 |
| polarC18 | 276.144 | 111.0918 | 6.4 | 0.12 |
| polarC18 | 276.144 | 145.515 | 6.4 | 0.12 |
| polarC18 | 276.144 | 50.2961 | 6.4 | 0.12 |
| polarC18 | 276.144 | 84.6165 | 6.3 | 0.12 |

| | | | | |
|---|---|---|---|---|
| polarC18 | 276.144 | 71.8522 | 6.3 | 0.12 |
| polarC18 | 276.144 | 183.5147 | 6.3 | 0.12 |
| polarC18 | 276.144 | 89.3869 | 6.3 | 0.12 |
| polarC18 | 276.144 | 56.5917 | 6.3 | 0.12 |
| polarC18 | 276.144 | 123.9962 | 6.3 | 0.12 |
| polarC18 | 276.144 | 93.743 | 6.3 | 0.12 |
| polarC18 | 276.144 | 178.8901 | 6.3 | 0.12 |
| polarC18 | 276.144 | 153.5571 | 6.3 | 0.12 |
| polarC18 | 276.144 | 98.3774 | 6.3 | 0.12 |
| polarC18 | 276.144 | 79.5267 | 6.3 | 0.12 |
| polarC18 | 276.144 | 95.1166 | 6.3 | 0.12 |
| polarC18 | 276.144 | 112.1953 | 6.3 | 0.12 |
| polarC18 | 276.144 | 87.0437 | 6.3 | 0.12 |
| polarC18 | 276.144 | 52.5513 | 6.3 | 0.12 |
| polarC18 | 276.144 | 65.0731 | 6.3 | 0.12 |
| polarC18 | 276.144 | 57.4665 | 6.3 | 0.12 |
| polarC18 | 276.144 | 76.6195 | 6.3 | 0.12 |
| polarC18 | 276.144 | 53.3529 | 6.3 | 0.12 |
| polarC18 | 276.144 | 213.2185 | 6.3 | 0.12 |
| polarC18 | 276.144 | 162.1257 | 6.3 | 0.12 |
| polarC18 | 276.144 | 99.9636 | 6.3 | 0.12 |
| polarC18 | 276.144 | 55.9422 | 6.3 | 0.12 |
| polarC18 | 276.144 | 56.9305 | 6.3 | 0.12 |
| polarC18 | 276.144 | 60.16 | 6.3 | 0.12 |
| polarC18 | 276.144 | 65.2214 | 6.3 | 0.12 |
| polarC18 | 276.144 | 151.6584 | 6.3 | 0.12 |
| polarC18 | 276.144 | 53.4714 | 6.3 | 0.12 |
| polarC18 | 276.144 | 58.2868 | 6.3 | 0.12 |
| polarC18 | 276.144 | 57.858 | 6.3 | 0.12 |
| polarC18 | 276.144 | 167.3716 | 6.3 | 0.12 |
| polarC18 | 276.144 | 62.6191 | 6.3 | 0.12 |
| polarC18 | 276.144 | 53.2669 | 6.3 | 0.12 |
| polarC18 | 276.144 | 66.188 | 6.3 | 0.12 |
| polarC18 | 276.144 | 86.7143 | 6.3 | 0.12 |
| polarC18 | 276.144 | 136.8775 | 6.3 | 0.12 |
| polarC18 | 276.144 | 71.2044 | 6.3 | 0.12 |
| polarC18 | 276.144 | 85.3897 | 6.3 | 0.12 |
| polarC18 | 276.144 | 88.1579 | 6.3 | 0.12 |
| polarC18 | 276.144 | 109.4279 | 6.3 | 0.12 |
| polarC18 | 276.144 | 51.7904 | 6.3 | 0.12 |
| polarC18 | 276.144 | 177.4521 | 6.3 | 0.12 |
| polarC18 | 276.144 | 60.1851 | 6.3 | 0.12 |

| | | | | |
|---|---|---|---|---|
| polarC18 | 276.144 | 59.823 | 6.3 | 0.12 |
| polarC18 | 276.144 | 65.2644 | 6.3 | 0.12 |
| polarC18 | 276.144 | 185.9464 | 6.3 | 0.12 |
| polarC18 | 276.144 | 68.1115 | 6.3 | 0.12 |
| polarC18 | 276.144 | 74.1098 | 6.3 | 0.12 |
| polarC18 | 276.144 | 65.3005 | 6.3 | 0.12 |
| polarC18 | 276.144 | 62.486 | 6.3 | 0.12 |
| polarC18 | 276.144 | 54.6233 | 6.3 | 0.12 |
| polarC18 | 276.144 | 56.8568 | 6.3 | 0.12 |
| polarC18 | 276.144 | 137.1278 | 6.3 | 0.12 |
| polarC18 | 276.144 | 156.1127 | 6.3 | 0.12 |
| polarC18 | 276.144 | 89.462 | 6.3 | 0.12 |
| polarC18 | 276.144 | 65.9464 | 6.3 | 0.12 |
| polarC18 | 276.144 | 72.8682 | 6.3 | 0.12 |
| polarC18 | 276.144 | 83.8351 | 6.3 | 0.12 |
| polarC18 | 276.144 | 243.9532 | 6.3 | 0.12 |
| polarC18 | 276.144 | 53.7178 | 6.3 | 0.12 |
| polarC18 | 276.144 | 127.949 | 6.3 | 0.12 |
| polarC18 | 276.144 | 52.327 | 6.3 | 0.12 |
| polarC18 | 276.144 | 70.5042 | 6.3 | 0.12 |
| polarC18 | 276.144 | 89.8838 | 6.3 | 0.12 |
| polarC18 | 276.144 | 268.4047 | 6.3 | 0.12 |
| polarC18 | 276.144 | 59.9319 | 6.3 | 0.12 |
| polarC18 | 276.144 | 62.4357 | 6.3 | 0.12 |
| polarC18 | 276.144 | 59.6886 | 6.3 | 0.12 |
| polarC18 | 276.144 | 67.6789 | 6.3 | 0.12 |
| polarC18 | 276.144 | 116.5792 | 6.3 | 0.12 |
| polarC18 | 276.144 | 71.4959 | 6.3 | 0.12 |
| polarC18 | 276.144 | 229.5313 | 6.3 | 0.12 |
| polarC18 | 276.144 | 76.5624 | 6.3 | 0.12 |
| polarC18 | 276.144 | 118.7781 | 6.3 | 0.12 |
| polarC18 | 276.144 | 52.5865 | 6.3 | 0.12 |
| polarC18 | 276.144 | 128.7319 | 6.3 | 0.12 |
| polarC18 | 276.144 | 72.0383 | 6.3 | 0.12 |
| polarC18 | 276.144 | 84.2793 | 6.3 | 0.12 |
| polarC18 | 276.144 | 52.543 | 6.3 | 0.12 |
| polarC18 | 276.144 | 72.4606 | 6.3 | 0.12 |
| polarC18 | 276.144 | 106.9811 | 6.3 | 0.12 |
| polarC18 | 276.144 | 75.9081 | 6.3 | 0.12 |
| polarC18 | 276.144 | 105.7228 | 6.3 | 0.12 |
| polarC18 | 276.144 | 62.6506 | 6.3 | 0.12 |
| polarC18 | 276.144 | 89.2359 | 6.3 | 0.12 |

| | | | | |
|---|---|---|---|---|
| polarC18 | 276.144 | 126.229 | 6.3 | 0.12 |
| polarC18 | 276.144 | 89.3382 | 6.3 | 0.12 |
| polarC18 | 276.144 | 101.6378 | 6.3 | 0.12 |
| polarC18 | 276.144 | 119.2397 | 6.3 | 0.12 |
| polarC18 | 276.144 | 52.7747 | 6.3 | 0.12 |
| polarC18 | 276.144 | 179.0261 | 6.3 | 0.12 |
| polarC18 | 276.144 | 63.3479 | 6.3 | 0.12 |
| polarC18 | 276.144 | 57.1203 | 6.3 | 0.12 |
| polarC18 | 276.144 | 57.5895 | 6.3 | 0.12 |
| polarC18 | 276.144 | 70.9885 | 6.3 | 0.12 |
| polarC18 | 276.144 | 92.2008 | 6.3 | 0.12 |
| polarC18 | 276.144 | 92.4638 | 6.3 | 0.12 |
| polarC18 | 276.144 | 63.2869 | 6.3 | 0.12 |
| polarC18 | 276.144 | 113.6807 | 6.3 | 0.12 |
| polarC18 | 276.144 | 67.4184 | 6.3 | 0.12 |
| polarC18 | 276.144 | 103.2548 | 6.3 | 0.12 |
| polarC18 | 276.144 | 64.7978 | 6.3 | 0.12 |
| polarC18 | 276.144 | 50.1768 | 6.3 | 0.12 |
| polarC18 | 276.144 | 120.576 | 6.3 | 0.12 |
| polarC18 | 276.144 | 66.7455 | 6.3 | 0.12 |
| polarC18 | 276.144 | 160.4826 | 6.3 | 0.12 |
| polarC18 | 276.144 | 139.6924 | 6.3 | 0.12 |
| polarC18 | 276.144 | 158.1745 | 6.3 | 0.12 |
| polarC18 | 276.144 | 61.3561 | 6.3 | 0.12 |
| polarC18 | 276.144 | 79.9652 | 6.3 | 0.12 |
| polarC18 | 276.144 | 51.4232 | 6.3 | 0.12 |
| polarC18 | 276.144 | 175.1394 | 6.3 | 0.12 |
| polarC18 | 276.144 | 154.6128 | 6.3 | 0.12 |
| polarC18 | 276.144 | 142.173 | 6.3 | 0.12 |
| polarC18 | 276.144 | 65.3817 | 6.3 | 0.12 |
| polarC18 | 276.144 | 101.0162 | 6.3 | 0.12 |
| polarC18 | 276.144 | 80.7108 | 6.3 | 0.12 |
| polarC18 | 276.144 | 215.5859 | 6.3 | 0.12 |
| polarC18 | 276.144 | 197.6115 | 6.3 | 0.12 |
| polarC18 | 276.144 | 66.6278 | 6.3 | 0.12 |
| polarC18 | 276.144 | 80.5426 | 6.3 | 0.12 |
| polarC18 | 276.144 | 96.1641 | 6.3 | 0.12 |
| polarC18 | 276.144 | 56.4494 | 6.3 | 0.12 |
| polarC18 | 276.144 | 112.0311 | 6.3 | 0.12 |
| polarC18 | 276.144 | 127.7229 | 6.3 | 0.12 |
| polarC18 | 276.144 | 220.981 | 6.3 | 0.12 |
| polarC18 | 276.144 | 88.7651 | 6.3 | 0.12 |

| | | | | |
|---|---|---|---|---|
| polarC18 | 276.144 | 55.1849 | 6.3 | 0.12 |
| polarC18 | 276.144 | 55.1874 | 6.3 | 0.12 |
| polarC18 | 276.144 | 122.7691 | 6.3 | 0.12 |
| polarC18 | 276.144 | 87.1881 | 6.3 | 0.12 |
| polarC18 | 276.144 | 186.1925 | 6.3 | 0.12 |
| polarC18 | 276.144 | 103.8803 | 6.3 | 0.12 |
| polarC18 | 276.144 | 56.8888 | 6.3 | 0.12 |
| polarC18 | 276.144 | 123.2605 | 6.3 | 0.12 |
| polarC18 | 276.144 | 171.6262 | 6.3 | 0.12 |
| polarC18 | 276.144 | 60.5449 | 6.3 | 0.12 |
| polarC18 | 276.144 | 147.2158 | 6.3 | 0.12 |
| polarC18 | 276.144 | 96.8276 | 6.3 | 0.12 |
| polarC18 | 276.144 | 162.3513 | 6.3 | 0.12 |
| polarC18 | 276.144 | 50.4687 | 6.3 | 0.12 |
| polarC18 | 276.144 | 114.6256 | 6.3 | 0.12 |
| polarC18 | 276.144 | 76.2002 | 6.3 | 0.12 |
| polarC18 | 276.144 | 52.9491 | 6.3 | 0.12 |
| polarC18 | 276.144 | 57.4038 | 6.3 | 0.12 |
| polarC18 | 276.144 | 89.8165 | 6.3 | 0.12 |
| polarC18 | 276.144 | 66.5139 | 6.3 | 0.12 |
| polarC18 | 276.144 | 81.0866 | 6.3 | 0.12 |
| polarC18 | 276.144 | 104.6311 | 6.3 | 0.12 |
| polarC18 | 276.144 | 54.1949 | 6.3 | 0.12 |
| polarC18 | 276.144 | 142.1055 | 6.3 | 0.12 |
| polarC18 | 276.144 | 143.6742 | 6.3 | 0.12 |
| polarC18 | 276.144 | 156.6778 | 6.3 | 0.12 |
| polarC18 | 276.144 | 51.4473 | 6.3 | 0.12 |
| polarC18 | 276.144 | 78.6889 | 6.3 | 0.12 |
| polarC18 | 276.144 | 103.649 | 6.3 | 0.12 |
| polarC18 | 276.144 | 235.1515 | 6.3 | 0.12 |
| polarC18 | 276.144 | 71.8291 | 6.3 | 0.12 |
| polarC18 | 276.144 | 180.0979 | 6.3 | 0.12 |
| polarC18 | 276.144 | 278.96 | 6.3 | 0.12 |
| polarC18 | 276.144 | 61.3999 | 6.3 | 0.12 |
| polarC18 | 276.144 | 69.9447 | 6.3 | 0.12 |
| polarC18 | 276.144 | 74.4579 | 6.3 | 0.12 |
| polarC18 | 276.144 | 183.2989 | 6.3 | 0.12 |
| polarC18 | 276.144 | 51.0097 | 6.3 | 0.12 |
| polarC18 | 276.144 | 241.7454 | 6.3 | 0.12 |
| polarC18 | 276.144 | 62.0928 | 6.3 | 0.12 |
| polarC18 | 276.144 | 138.8383 | 6.3 | 0.12 |
| polarC18 | 276.144 | 92.2381 | 6.3 | 0.12 |

| | | | | |
|---|---|---|---|---|
| polarC18 | 276.144 | 279.1216 | 6.3 | 0.12 |
| polarC18 | 276.144 | 95.269 | 6.3 | 0.12 |
| polarC18 | 276.144 | 72.1815 | 6.3 | 0.12 |
| polarC18 | 276.144 | 194.5789 | 6.3 | 0.12 |
| polarC18 | 276.144 | 153.5839 | 6.3 | 0.12 |
| polarC18 | 276.144 | 50.1025 | 6.3 | 0.12 |
| polarC18 | 276.144 | 71.948 | 6.3 | 0.12 |
| polarC18 | 276.144 | 64.5159 | 6.3 | 0.12 |
| polarC18 | 276.144 | 52.2313 | 6.3 | 0.12 |
| polarC18 | 276.144 | 81.4518 | 6.3 | 0.12 |
| polarC18 | 276.144 | 51.0719 | 6.3 | 0.12 |
| polarC18 | 276.144 | 52.2172 | 6.3 | 0.12 |
| polarC18 | 276.144 | 112.7086 | 6.3 | 0.12 |
| polarC18 | 276.144 | 67.6127 | 6.3 | 0.12 |
| polarC18 | 276.144 | 79.2878 | 6.3 | 0.12 |
| polarC18 | 276.144 | 157.326 | 6.3 | 0.12 |
| polarC18 | 276.144 | 73.1759 | 6.3 | 0.12 |
| polarC18 | 276.144 | 110.1919 | 6.3 | 0.12 |
| polarC18 | 276.144 | 50.3269 | 6.3 | 0.12 |
| polarC18 | 276.144 | 87.6216 | 6.3 | 0.12 |
| polarC18 | 276.144 | 113.894 | 6.3 | 0.12 |
| polarC18 | 276.144 | 91.3706 | 6.3 | 0.12 |
| polarC18 | 276.144 | 65.6919 | 6.3 | 0.12 |
| polarC18 | 276.144 | 177.7732 | 6.3 | 0.12 |
| polarC18 | 276.144 | 77.0546 | 6.3 | 0.12 |
| polarC18 | 276.144 | 67.841 | 6.3 | 0.12 |
| polarC18 | 276.144 | 84.5977 | 6.3 | 0.12 |
| polarC18 | 276.144 | 143.962 | 6.3 | 0.12 |
| polarC18 | 276.144 | 280.9286 | 6.3 | 0.12 |
| polarC18 | 276.144 | 62.7834 | 6.3 | 0.12 |
| polarC18 | 276.144 | 59.1575 | 6.3 | 0.12 |
| polarC18 | 276.144 | 116.1741 | 6.3 | 0.12 |
| polarC18 | 276.144 | 216.8527 | 6.3 | 0.12 |
| polarC18 | 276.144 | 65.4128 | 6.3 | 0.12 |
| polarC18 | 276.144 | 101.2711 | 6.3 | 0.12 |
| polarC18 | 276.144 | 149.1578 | 6.3 | 0.12 |
| polarC18 | 276.144 | 227.2022 | 6.3 | 0.12 |
| polarC18 | 276.144 | 56.8395 | 6.3 | 0.12 |
| polarC18 | 276.144 | 56.8547 | 6.3 | 0.12 |
| polarC18 | 276.144 | 54.702 | 6.3 | 0.12 |
| polarC18 | 276.144 | 73.8584 | 6.3 | 0.12 |
| polarC18 | 276.144 | 86.3781 | 6.3 | 0.12 |

| | | | | |
|---|---|---|---|---|
| polarC18 | 276.144 | 51.735 | 6.3 | 0.12 |
| polarC18 | 276.144 | 57.9284 | 6.3 | 0.12 |
| polarC18 | 276.144 | 64.2466 | 6.3 | 0.12 |
| polarC18 | 276.144 | 109.3267 | 6.3 | 0.12 |
| polarC18 | 276.144 | 66.7201 | 6.3 | 0.12 |
| polarC18 | 276.144 | 188.6415 | 6.3 | 0.12 |
| polarC18 | 276.144 | 106.0523 | 6.3 | 0.12 |
| polarC18 | 276.144 | 196.5089 | 6.3 | 0.12 |
| polarC18 | 276.144 | 282.7448 | 6.3 | 0.12 |
| polarC18 | 276.144 | 57.0768 | 6.3 | 0.12 |
| polarC18 | 276.144 | 71.2686 | 6.3 | 0.12 |
| polarC18 | 276.144 | 76.9994 | 6.3 | 0.12 |
| polarC18 | 276.144 | 185.8761 | 6.3 | 0.12 |
| polarC18 | 276.144 | 180.9702 | 6.3 | 0.12 |
| polarC18 | 276.144 | 260.9286 | 6.3 | 0.12 |
| polarC18 | 276.144 | 121.8836 | 6.3 | 0.12 |
| polarC18 | 276.144 | 218.8407 | 6.3 | 0.12 |
| polarC18 | 276.144 | 70.6408 | 6.3 | 0.12 |
| polarC18 | 276.144 | 76.7697 | 6.3 | 0.12 |
| polarC18 | 276.144 | 209.3037 | 6.3 | 0.12 |
| polarC18 | 276.144 | 65.3321 | 6.3 | 0.12 |
| polarC18 | 276.144 | 193.1198 | 6.3 | 0.12 |
| polarC18 | 276.144 | 131.5047 | 6.3 | 0.12 |
| polarC18 | 276.144 | 170.5326 | 6.3 | 0.12 |
| polarC18 | 276.144 | 55.9883 | 6.3 | 0.12 |
| polarC18 | 276.144 | 95.9247 | 6.3 | 0.12 |
| polarC18 | 276.144 | 54.1926 | 6.3 | 0.12 |
| polarC18 | 276.144 | 145.6311 | 6.3 | 0.12 |
| polarC18 | 276.144 | 169.7335 | 6.3 | 0.12 |
| polarC18 | 276.144 | 84.698 | 6.3 | 0.12 |
| polarC18 | 276.144 | 146.7898 | 6.3 | 0.12 |
| polarC18 | 276.144 | 154.9644 | 6.3 | 0.12 |
| polarC18 | 276.144 | 57.0431 | 6.3 | 0.12 |
| polarC18 | 276.144 | 69.1875 | 6.3 | 0.12 |
| polarC18 | 276.144 | 188.9564 | 6.3 | 0.12 |
| polarC18 | 276.144 | 109.2378 | 6.3 | 0.12 |
| polarC18 | 276.144 | 106.2907 | 6.3 | 0.12 |
| polarC18 | 276.144 | 72.058 | 6.3 | 0.12 |
| polarC18 | 276.144 | 61.5496 | 6.3 | 0.12 |
| polarC18 | 276.144 | 122.8863 | 6.3 | 0.12 |
| polarC18 | 276.144 | 81.5499 | 6.3 | 0.12 |
| polarC18 | 276.144 | 89.1093 | 6.3 | 0.12 |

| | | | | |
|---|---|---|---|---|
| polarC18 | 276.144 | 162.9339 | 6.3 | 0.12 |
| polarC18 | 276.144 | 54.6933 | 6.3 | 0.12 |
| polarC18 | 276.144 | 185.5001 | 6.3 | 0.12 |
| polarC18 | 276.144 | 68.8826 | 6.3 | 0.12 |
| polarC18 | 276.144 | 69.3113 | 6.3 | 0.12 |
| polarC18 | 276.144 | 138.1009 | 6.3 | 0.12 |
| polarC18 | 276.144 | 57.8115 | 6.3 | 0.12 |
| polarC18 | 276.144 | 85.8819 | 6.3 | 0.12 |
| polarC18 | 276.144 | 68.5365 | 6.3 | 0.12 |
| polarC18 | 276.144 | 62.4325 | 6.3 | 0.12 |
| polarC18 | 276.144 | 55.7292 | 6.3 | 0.12 |
| polarC18 | 276.144 | 67.9164 | 6.3 | 0.12 |
| polarC18 | 276.144 | 70.9857 | 6.3 | 0.12 |
| polarC18 | 276.144 | 126.3683 | 6.3 | 0.12 |
| polarC18 | 276.144 | 112.1393 | 6.3 | 0.12 |
| polarC18 | 276.144 | 247.1835 | 6.3 | 0.12 |
| polarC18 | 276.144 | 57.1892 | 6.3 | 0.12 |
| polarC18 | 276.144 | 63.3088 | 6.3 | 0.12 |
| polarC18 | 276.144 | 246.5583 | 6.3 | 0.12 |
| polarC18 | 276.144 | 51.8097 | 6.3 | 0.12 |
| polarC18 | 276.144 | 126.5975 | 6.3 | 0.12 |
| polarC18 | 276.144 | 239.0248 | 6.3 | 0.12 |
| polarC18 | 276.144 | 94.5892 | 6.3 | 0.12 |
| polarC18 | 276.144 | 61.2332 | 6.3 | 0.12 |
| polarC18 | 276.144 | 119.5855 | 6.3 | 0.12 |
| polarC18 | 276.144 | 51.7367 | 6.3 | 0.12 |
| polarC18 | 276.144 | 177.9603 | 6.3 | 0.12 |
| polarC18 | 276.144 | 103.1526 | 6.3 | 0.12 |
| polarC18 | 276.144 | 54.9867 | 6.3 | 0.12 |
| polarC18 | 276.144 | 96.507 | 6.3 | 0.12 |
| polarC18 | 276.144 | 69.8149 | 6.3 | 0.12 |
| polarC18 | 276.144 | 60.363 | 6.3 | 0.12 |
| polarC18 | 276.144 | 66.9466 | 6.3 | 0.12 |
| polarC18 | 276.144 | 135.5587 | 6.3 | 0.12 |
| polarC18 | 276.144 | 98.993 | 6.3 | 0.12 |
| polarC18 | 276.144 | 51.8925 | 6.3 | 0.12 |
| polarC18 | 276.144 | 58.4581 | 6.3 | 0.12 |
| polarC18 | 276.144 | 57.4058 | 6.3 | 0.12 |
| polarC18 | 276.144 | 63.0531 | 6.3 | 0.12 |
| polarC18 | 276.144 | 75.2715 | 6.2 | 0.12 |
| polarC18 | 276.144 | 50.8892 | 6.2 | 0.12 |
| polarC18 | 276.144 | 77.4793 | 6.2 | 0.12 |

| | | | | |
|---|---|---|---|---|
| polarC18 | 276.144 | 137.6634 | 6.2 | 0.12 |
| polarC18 | 276.144 | 64.4587 | 6.2 | 0.12 |
| polarC18 | 276.144 | 82.934 | 6.2 | 0.12 |
| polarC18 | 276.144 | 55.0626 | 6.2 | 0.12 |
| polarC18 | 276.144 | 89.9864 | 6.2 | 0.12 |
| polarC18 | 276.144 | 89.5899 | 6.2 | 0.12 |
| polarC18 | 276.144 | 156.9373 | 6.2 | 0.12 |
| polarC18 | 276.144 | 99.02 | 6.2 | 0.12 |
| polarC18 | 276.144 | 210.3004 | 6.2 | 0.12 |
| polarC18 | 276.144 | 62.4818 | 6.2 | 0.12 |
| polarC18 | 276.144 | 146.484 | 6.2 | 0.12 |
| polarC18 | 276.144 | 120.4302 | 6.2 | 0.12 |
| polarC18 | 276.144 | 54.3961 | 6.2 | 0.12 |
| polarC18 | 276.144 | 108.8152 | 6.2 | 0.12 |
| polarC18 | 276.144 | 152.6967 | 6.2 | 0.12 |
| polarC18 | 276.144 | 156.2786 | 6.2 | 0.12 |
| polarC18 | 276.144 | 150.5433 | 6.2 | 0.12 |
| polarC18 | 276.144 | 68.1945 | 6.2 | 0.12 |
| polarC18 | 276.144 | 56.1881 | 6.2 | 0.12 |
| polarC18 | 276.144 | 57.5114 | 6.2 | 0.12 |
| polarC18 | 276.144 | 72.3409 | 6.2 | 0.12 |
| polarC18 | 276.144 | 166.985 | 6.2 | 0.12 |
| polarC18 | 276.144 | 68.9256 | 6.2 | 0.12 |
| polarC18 | 276.144 | 72.3306 | 6.2 | 0.12 |
| polarC18 | 276.144 | 89.8686 | 6.2 | 0.12 |
| polarC18 | 276.144 | 175.0227 | 6.2 | 0.12 |
| polarC18 | 276.144 | 60.3137 | 6.2 | 0.12 |
| polarC18 | 276.144 | 74.1528 | 6.2 | 0.12 |
| polarC18 | 276.144 | 76.6019 | 6.2 | 0.12 |
| polarC18 | 276.144 | 53.0548 | 6.2 | 0.12 |
| polarC18 | 276.144 | 170.8619 | 6.2 | 0.12 |
| polarC18 | 276.144 | 148.6654 | 6.2 | 0.12 |
| polarC18 | 276.144 | 59.3984 | 6.2 | 0.12 |
| polarC18 | 276.144 | 68.8502 | 6.2 | 0.12 |
| polarC18 | 276.144 | 164.2798 | 6.2 | 0.12 |
| polarC18 | 276.144 | 183.0077 | 6.2 | 0.12 |
| polarC18 | 276.144 | 106.4796 | 6.2 | 0.12 |
| polarC18 | 276.144 | 53.3884 | 6.2 | 0.12 |
| polarC18 | 276.144 | 57.0224 | 6.2 | 0.12 |
| polarC18 | 276.144 | 54.3123 | 6.2 | 0.12 |
| polarC18 | 276.144 | 77.164 | 6.2 | 0.12 |
| polarC18 | 276.144 | 116.2552 | 6.2 | 0.12 |

| | | | | |
|---|---|---|---|---|
| polarC18 | 276.144 | 79.6714 | 6.2 | 0.12 |
| polarC18 | 276.144 | 152.3145 | 6.2 | 0.12 |
| polarC18 | 276.144 | 97.3499 | 6.2 | 0.12 |
| polarC18 | 276.144 | 158.8406 | 6.2 | 0.12 |
| polarC18 | 276.144 | 57.9374 | 6.2 | 0.12 |
| polarC18 | 276.144 | 63.851 | 6.2 | 0.12 |
| polarC18 | 276.144 | 122.2952 | 6.2 | 0.12 |
| polarC18 | 276.144 | 195.8634 | 6.2 | 0.12 |
| polarC18 | 276.144 | 102.7388 | 6.2 | 0.12 |
| polarC18 | 276.144 | 73.8647 | 6.2 | 0.12 |
| polarC18 | 276.144 | 102.1091 | 6.2 | 0.12 |
| polarC18 | 276.144 | 53.0599 | 6.2 | 0.12 |
| polarC18 | 276.144 | 297.259 | 6.2 | 0.12 |
| polarC18 | 276.144 | 50.285 | 6.2 | 0.12 |
| polarC18 | 276.144 | 164.9226 | 6.2 | 0.12 |
| polarC18 | 276.144 | 112.5292 | 6.2 | 0.12 |
| polarC18 | 276.144 | 51.433 | 6.2 | 0.12 |
| polarC18 | 276.144 | 78.9104 | 6.2 | 0.12 |
| polarC18 | 276.144 | 63.2019 | 6.2 | 0.12 |
| polarC18 | 276.144 | 63.7074 | 6.2 | 0.12 |
| polarC18 | 276.144 | 82.9248 | 6.2 | 0.12 |
| polarC18 | 276.144 | 94.0743 | 6.2 | 0.12 |
| polarC18 | 276.144 | 151.9997 | 6.2 | 0.12 |
| polarC18 | 276.144 | 64.4837 | 6.2 | 0.12 |
| polarC18 | 276.144 | 82.2429 | 6.2 | 0.12 |
| polarC18 | 276.144 | 73.9232 | 6.2 | 0.12 |
| polarC18 | 276.144 | 62.0882 | 6.2 | 0.12 |
| polarC18 | 276.144 | 65.3226 | 6.2 | 0.12 |
| polarC18 | 276.144 | 62.1995 | 6.2 | 0.12 |
| polarC18 | 276.144 | 97.6191 | 6.2 | 0.12 |
| polarC18 | 276.144 | 74.5545 | 6.2 | 0.12 |
| polarC18 | 276.144 | 189.876 | 6.2 | 0.12 |
| polarC18 | 276.144 | 162.8515 | 6.2 | 0.12 |
| polarC18 | 276.144 | 56.0065 | 6.2 | 0.12 |
| polarC18 | 276.144 | 98.2511 | 6.2 | 0.12 |
| polarC18 | 276.144 | 61.6485 | 6.2 | 0.12 |
| polarC18 | 276.144 | 182.9007 | 6.2 | 0.12 |
| polarC18 | 276.144 | 261.3987 | 6.2 | 0.12 |
| polarC18 | 276.144 | 66.5249 | 6.2 | 0.12 |
| polarC18 | 276.144 | 68.2493 | 6.2 | 0.12 |
| polarC18 | 276.144 | 102.0684 | 6.2 | 0.12 |
| polarC18 | 276.144 | 75.2636 | 6.2 | 0.12 |

| | | | | |
|---|---|---|---|---|
| polarC18 | 276.144 | 51.2832 | 6.2 | 0.12 |
| polarC18 | 276.144 | 64.2016 | 6.2 | 0.12 |
| polarC18 | 276.144 | 99.2465 | 6.2 | 0.12 |
| polarC18 | 276.144 | 242.0701 | 6.2 | 0.12 |
| polarC18 | 276.144 | 162.725 | 6.2 | 0.12 |
| polarC18 | 276.144 | 68.3021 | 6.2 | 0.12 |
| polarC18 | 276.144 | 161.5844 | 6.2 | 0.12 |
| polarC18 | 276.144 | 106.0076 | 6.2 | 0.12 |
| polarC18 | 276.144 | 93.153 | 6.2 | 0.12 |
| polarC18 | 276.144 | 52.853 | 6.2 | 0.12 |
| polarC18 | 276.144 | 108.7963 | 6.2 | 0.12 |
| polarC18 | 276.144 | 71.2131 | 6.2 | 0.12 |
| polarC18 | 276.144 | 64.0598 | 6.2 | 0.12 |
| polarC18 | 276.144 | 61.9252 | 6.2 | 0.12 |
| polarC18 | 276.144 | 53.8817 | 6.2 | 0.12 |
| polarC18 | 276.144 | 143.3402 | 6.2 | 0.12 |
| polarC18 | 276.144 | 96.8104 | 6.2 | 0.12 |
| polarC18 | 276.144 | 75.8724 | 6.2 | 0.12 |
| polarC18 | 276.144 | 104.0666 | 6.2 | 0.12 |
| polarC18 | 276.144 | 78.6275 | 6.2 | 0.12 |
| polarC18 | 276.144 | 64.0942 | 6.2 | 0.12 |
| polarC18 | 276.144 | 56.2593 | 6.2 | 0.12 |
| polarC18 | 276.144 | 55.921 | 6.2 | 0.12 |
| polarC18 | 276.144 | 281.8623 | 6.2 | 0.12 |
| polarC18 | 276.144 | 90.1321 | 6.2 | 0.12 |
| polarC18 | 276.144 | 77.6369 | 6.2 | 0.12 |
| polarC18 | 276.144 | 282.1124 | 6.2 | 0.12 |
| polarC18 | 276.144 | 139.6534 | 6.2 | 0.12 |
| polarC18 | 276.144 | 277.5094 | 6.2 | 0.12 |
| polarC18 | 276.144 | 64.9824 | 6.2 | 0.12 |
| polarC18 | 276.144 | 61.5688 | 6.2 | 0.12 |
| polarC18 | 276.144 | 80.5676 | 6.2 | 0.12 |
| polarC18 | 276.144 | 163.129 | 6.2 | 0.12 |
| polarC18 | 276.144 | 116.5522 | 6.2 | 0.12 |
| polarC18 | 276.144 | 105.3622 | 6.2 | 0.12 |
| polarC18 | 276.144 | 83.0854 | 6.2 | 0.12 |
| polarC18 | 276.144 | 80.9177 | 6.2 | 0.12 |
| polarC18 | 276.144 | 96.9649 | 6.2 | 0.12 |
| polarC18 | 276.144 | 138.8266 | 6.2 | 0.12 |
| polarC18 | 276.144 | 51.6212 | 6.2 | 0.12 |
| polarC18 | 276.144 | 71.1135 | 6.2 | 0.12 |
| polarC18 | 276.144 | 54.3355 | 6.2 | 0.12 |

| | | | | |
|---|---|---|---|---|
| polarC18 | 276.144 | 119.0743 | 6.2 | 0.12 |
| polarC18 | 276.144 | 225.677 | 6.2 | 0.12 |
| polarC18 | 276.144 | 87.9636 | 6.2 | 0.12 |
| polarC18 | 276.144 | 78.1867 | 6.2 | 0.12 |
| polarC18 | 276.144 | 141.9371 | 6.2 | 0.12 |
| polarC18 | 276.144 | 176.7195 | 6.2 | 0.12 |
| polarC18 | 276.144 | 57.0938 | 6.2 | 0.12 |
| polarC18 | 276.144 | 56.4052 | 6.2 | 0.12 |
| polarC18 | 276.144 | 86.4387 | 6.2 | 0.12 |
| polarC18 | 276.144 | 53.4284 | 6.2 | 0.12 |
| polarC18 | 276.144 | 59.7852 | 6.2 | 0.12 |
| polarC18 | 276.144 | 51.8782 | 6.2 | 0.12 |
| polarC18 | 276.144 | 62.3722 | 6.2 | 0.12 |
| polarC18 | 276.144 | 132.2445 | 6.2 | 0.12 |
| polarC18 | 276.144 | 137.9879 | 6.2 | 0.12 |
| polarC18 | 276.144 | 66.1921 | 6.2 | 0.12 |
| polarC18 | 276.144 | 128.008 | 6.2 | 0.12 |
| polarC18 | 276.144 | 253.0039 | 6.2 | 0.12 |
| polarC18 | 276.144 | 61.9382 | 6.2 | 0.12 |
| polarC18 | 276.144 | 50.7943 | 6.2 | 0.12 |
| polarC18 | 276.144 | 208.3228 | 6.2 | 0.12 |
| polarC18 | 276.144 | 180.0305 | 6.2 | 0.12 |
| polarC18 | 276.144 | 275.6136 | 6.2 | 0.12 |
| polarC18 | 276.144 | 119.8057 | 6.2 | 0.12 |
| polarC18 | 276.144 | 60.0983 | 6.2 | 0.12 |
| polarC18 | 276.144 | 148.0385 | 6.2 | 0.12 |
| polarC18 | 276.144 | 111.9044 | 6.2 | 0.12 |
| polarC18 | 276.144 | 64.6803 | 6.2 | 0.12 |
| polarC18 | 276.144 | 88.0596 | 6.2 | 0.12 |
| polarC18 | 276.144 | 87.9545 | 6.2 | 0.12 |
| polarC18 | 276.144 | 71.6323 | 6.2 | 0.12 |
| polarC18 | 276.144 | 202.0779 | 6.2 | 0.12 |
| polarC18 | 276.144 | 85.5045 | 6.2 | 0.12 |
| polarC18 | 276.144 | 67.0648 | 6.2 | 0.12 |
| polarC18 | 276.144 | 194.0197 | 6.2 | 0.12 |
| polarC18 | 276.144 | 139.4885 | 6.2 | 0.12 |
| polarC18 | 276.144 | 61.8008 | 6.2 | 0.12 |
| polarC18 | 276.144 | 77.8533 | 6.2 | 0.12 |
| polarC18 | 276.144 | 181.5775 | 6.2 | 0.12 |
| polarC18 | 276.144 | 68.376 | 6.2 | 0.12 |
| polarC18 | 276.144 | 111.7046 | 6.2 | 0.12 |
| polarC18 | 276.144 | 214.9751 | 6.2 | 0.12 |

| | | | | |
|---|---|---|---|---|
| polarC18 | 276.144 | 67.9696 | 6.2 | 0.12 |
| polarC18 | 276.144 | 82.233 | 6.2 | 0.12 |
| polarC18 | 276.144 | 73.3491 | 6.2 | 0.12 |
| polarC18 | 276.144 | 50.9781 | 6.2 | 0.12 |
| polarC18 | 276.144 | 119.321 | 6.2 | 0.12 |
| polarC18 | 276.144 | 73.1233 | 6.2 | 0.12 |
| polarC18 | 276.144 | 79.1906 | 6.2 | 0.12 |
| polarC18 | 276.144 | 55.6724 | 6.2 | 0.12 |
| polarC18 | 276.144 | 107.8227 | 6.2 | 0.12 |
| polarC18 | 276.144 | 108.8856 | 6.2 | 0.12 |
| polarC18 | 276.144 | 82.0596 | 6.2 | 0.12 |
| polarC18 | 276.144 | 56.5432 | 6.2 | 0.12 |
| polarC18 | 276.144 | 161.7469 | 6.2 | 0.12 |
| polarC18 | 276.144 | 65.1146 | 6.2 | 0.12 |
| polarC18 | 276.144 | 72.0279 | 6.2 | 0.12 |
| polarC18 | 276.144 | 127.3895 | 6.2 | 0.12 |
| polarC18 | 276.144 | 67.0945 | 6.2 | 0.12 |
| polarC18 | 276.144 | 104.4487 | 6.2 | 0.12 |
| polarC18 | 276.144 | 183.6548 | 6.2 | 0.12 |
| polarC18 | 276.144 | 104.8811 | 6.2 | 0.12 |
| polarC18 | 276.144 | 50.466 | 6.2 | 0.12 |
| polarC18 | 276.144 | 63.5902 | 6.2 | 0.12 |
| polarC18 | 276.144 | 64.1553 | 6.2 | 0.12 |
| polarC18 | 276.144 | 243.1233 | 6.2 | 0.12 |
| polarC18 | 276.144 | 131.1614 | 6.2 | 0.12 |
| polarC18 | 276.144 | 70.2149 | 6.2 | 0.12 |
| polarC18 | 276.144 | 140.3511 | 6.2 | 0.12 |
| polarC18 | 276.144 | 107.4747 | 6.2 | 0.12 |
| polarC18 | 276.144 | 51.0109 | 6.2 | 0.12 |
| polarC18 | 276.144 | 69.0275 | 6.2 | 0.12 |
| polarC18 | 276.144 | 168.4135 | 6.2 | 0.12 |
| polarC18 | 276.144 | 75.1418 | 6.2 | 0.12 |
| polarC18 | 276.144 | 108.3629 | 6.2 | 0.12 |
| polarC18 | 276.144 | 61.4257 | 6.2 | 0.12 |
| polarC18 | 276.144 | 164.1086 | 6.2 | 0.12 |
| polarC18 | 276.144 | 53.2095 | 6.2 | 0.12 |
| polarC18 | 276.144 | 51.652 | 6.2 | 0.12 |
| polarC18 | 276.144 | 87.5539 | 6.2 | 0.12 |
| polarC18 | 276.144 | 88.6979 | 6.2 | 0.12 |
| polarC18 | 276.144 | 222.8866 | 6.2 | 0.12 |
| polarC18 | 276.144 | 60.8243 | 6.2 | 0.12 |
| polarC18 | 276.144 | 68.469 | 6.2 | 0.12 |

| | | | | |
|---|---|---|---|---|
| polarC18 | 276.144 | 216.2214 | 6.2 | 0.12 |
| polarC18 | 276.144 | 169.5781 | 6.2 | 0.12 |
| polarC18 | 276.144 | 194.607 | 6.2 | 0.12 |
| polarC18 | 276.144 | 152.8102 | 6.2 | 0.12 |
| polarC18 | 276.144 | 124.5966 | 6.2 | 0.12 |
| polarC18 | 276.144 | 53.107 | 6.2 | 0.12 |
| polarC18 | 276.144 | 59.1383 | 6.2 | 0.12 |
| polarC18 | 276.144 | 50.7144 | 6.2 | 0.12 |
| polarC18 | 276.144 | 56.7197 | 6.2 | 0.12 |
| polarC18 | 276.144 | 143.0048 | 6.2 | 0.12 |
| polarC18 | 276.144 | 102.1613 | 6.2 | 0.12 |
| polarC18 | 276.144 | 68.756 | 6.2 | 0.12 |
| polarC18 | 276.144 | 105.188 | 6.2 | 0.12 |
| polarC18 | 276.144 | 53.6352 | 6.2 | 0.12 |
| polarC18 | 276.144 | 51.8483 | 6.2 | 0.12 |
| polarC18 | 276.144 | 113.9184 | 6.2 | 0.12 |
| polarC18 | 276.144 | 52.8185 | 6.2 | 0.12 |
| polarC18 | 276.144 | 73.8127 | 6.2 | 0.12 |
| polarC18 | 276.144 | 119.9631 | 6.2 | 0.12 |
| polarC18 | 276.144 | 54.6033 | 6.2 | 0.12 |
| polarC18 | 276.144 | 217.9141 | 6.2 | 0.12 |
| polarC18 | 276.144 | 50.9467 | 6.2 | 0.12 |
| polarC18 | 276.144 | 127.1517 | 6.2 | 0.12 |
| polarC18 | 276.144 | 53.925 | 6.1 | 0.12 |
| polarC18 | 276.144 | 56.8121 | 6.1 | 0.12 |
| polarC18 | 276.144 | 88.4737 | 6.1 | 0.12 |
| polarC18 | 276.144 | 146.722 | 6.1 | 0.12 |
| polarC18 | 276.144 | 278.7028 | 6.1 | 0.12 |
| polarC18 | 276.144 | 70.2998 | 6.1 | 0.12 |
| polarC18 | 276.144 | 59.8603 | 6.1 | 0.12 |
| polarC18 | 276.144 | 56.7104 | 6.1 | 0.12 |
| polarC18 | 276.144 | 69.3916 | 6.1 | 0.12 |
| polarC18 | 276.144 | 144.6093 | 6.1 | 0.12 |
| polarC18 | 276.144 | 62.4445 | 6.1 | 0.12 |
| polarC18 | 276.144 | 66.9121 | 6.1 | 0.12 |
| polarC18 | 276.144 | 235.5751 | 6.1 | 0.12 |
| polarC18 | 276.144 | 105.8951 | 6.1 | 0.12 |
| polarC18 | 276.144 | 121.8969 | 6.1 | 0.12 |
| polarC18 | 276.144 | 158.4738 | 6.1 | 0.12 |
| polarC18 | 276.144 | 104.8318 | 6.1 | 0.12 |
| polarC18 | 276.144 | 69.5225 | 6.1 | 0.12 |
| polarC18 | 276.144 | 67.3174 | 6.1 | 0.12 |

| | | | | |
|---|---|---|---|---|
| polarC18 | 276.144 | 148.785 | 6.1 | 0.12 |
| polarC18 | 276.144 | 162.1402 | 6.1 | 0.12 |
| polarC18 | 276.144 | 50.4269 | 6.1 | 0.12 |
| polarC18 | 276.144 | 57.2423 | 6.1 | 0.12 |
| polarC18 | 276.144 | 53.5194 | 6.1 | 0.12 |
| polarC18 | 276.144 | 65.7609 | 6.1 | 0.12 |
| polarC18 | 276.144 | 172.934 | 6.1 | 0.12 |
| polarC18 | 276.144 | 97.9083 | 6.1 | 0.12 |
| polarC18 | 276.144 | 100.4127 | 6.1 | 0.12 |
| polarC18 | 276.144 | 129.7109 | 6.1 | 0.12 |
| polarC18 | 276.144 | 61.3686 | 6.1 | 0.12 |
| polarC18 | 276.144 | 52.7379 | 6.1 | 0.12 |
| polarC18 | 276.144 | 60.6904 | 6.1 | 0.12 |
| polarC18 | 276.144 | 85.6489 | 6.1 | 0.12 |
| polarC18 | 276.144 | 61.621 | 6.1 | 0.12 |
| polarC18 | 276.144 | 69.457 | 6.1 | 0.12 |
| polarC18 | 276.144 | 67.0162 | 6.1 | 0.12 |
| polarC18 | 276.144 | 230.3511 | 6.1 | 0.12 |
| polarC18 | 276.144 | 52.3025 | 6.1 | 0.12 |
| polarC18 | 276.144 | 89.7468 | 6.1 | 0.12 |
| polarC18 | 276.144 | 120.6603 | 6.1 | 0.12 |
| polarC18 | 276.144 | 121.5563 | 6.1 | 0.12 |
| polarC18 | 276.144 | 67.9074 | 6.1 | 0.12 |
| polarC18 | 276.144 | 69.0171 | 6.1 | 0.12 |
| polarC18 | 276.144 | 60.4899 | 6.1 | 0.12 |
| polarC18 | 276.144 | 50.6672 | 6.1 | 0.12 |
| polarC18 | 276.144 | 93.7268 | 6.1 | 0.12 |
| polarC18 | 276.144 | 110.8001 | 6.1 | 0.12 |
| polarC18 | 276.144 | 67.7759 | 6.1 | 0.12 |
| polarC18 | 276.144 | 58.9662 | 6.1 | 0.12 |
| polarC18 | 276.144 | 123.1455 | 6.1 | 0.12 |
| polarC18 | 276.144 | 60.2445 | 6.1 | 0.12 |
| polarC18 | 276.144 | 178.7121 | 6.1 | 0.12 |
| polarC18 | 276.144 | 79.2277 | 6.1 | 0.12 |
| polarC18 | 276.144 | 56.7777 | 6.1 | 0.12 |
| polarC18 | 276.144 | 59.7936 | 6.1 | 0.12 |
| polarC18 | 276.144 | 130.2518 | 6.1 | 0.12 |
| polarC18 | 276.144 | 66.7776 | 6.1 | 0.12 |
| polarC18 | 276.144 | 60.6514 | 6.1 | 0.12 |
| polarC18 | 276.144 | 158.7647 | 6.1 | 0.12 |
| polarC18 | 276.144 | 71.2728 | 6.1 | 0.12 |
| polarC18 | 276.144 | 54.2663 | 6.1 | 0.12 |

| | | | | |
|---|---|---|---|---|
| polarC18 | 276.144 | 68.1255 | 6.1 | 0.12 |
| polarC18 | 276.144 | 182.4557 | 6.1 | 0.12 |
| polarC18 | 276.144 | 50.7049 | 6.1 | 0.12 |
| polarC18 | 276.144 | 55.7031 | 6.1 | 0.12 |
| polarC18 | 276.144 | 79.9284 | 6.1 | 0.12 |
| polarC18 | 276.144 | 102.0137 | 6.1 | 0.12 |
| polarC18 | 276.144 | 70.0715 | 6.1 | 0.12 |
| polarC18 | 276.144 | 80.0798 | 6.1 | 0.12 |
| polarC18 | 276.144 | 58.4083 | 6.1 | 0.12 |
| polarC18 | 276.144 | 60.9701 | 6.1 | 0.12 |
| polarC18 | 276.144 | 89.6522 | 6.1 | 0.12 |
| polarC18 | 276.144 | 55.9988 | 6.1 | 0.12 |
| polarC18 | 276.144 | 108.3485 | 6.1 | 0.12 |
| polarC18 | 276.144 | 67.2373 | 6.1 | 0.12 |
| polarC18 | 276.144 | 61.2809 | 6.1 | 0.12 |
| polarC18 | 276.144 | 54.5618 | 6.1 | 0.12 |
| polarC18 | 276.144 | 91.9209 | 6.1 | 0.12 |
| polarC18 | 276.144 | 55.0072 | 6.1 | 0.12 |
| polarC18 | 276.144 | 252.1238 | 6.1 | 0.12 |
| polarC18 | 276.144 | 118.4232 | 6.1 | 0.12 |
| polarC18 | 276.144 | 112.3235 | 6.1 | 0.12 |
| polarC18 | 276.144 | 291.3197 | 6.1 | 0.12 |
| polarC18 | 276.144 | 111.7289 | 6.1 | 0.12 |
| polarC18 | 276.144 | 200.8246 | 6.1 | 0.12 |
| polarC18 | 276.144 | 66.3162 | 6.1 | 0.12 |
| polarC18 | 276.144 | 82.6382 | 6.1 | 0.12 |
| polarC18 | 276.144 | 138.171 | 6.1 | 0.12 |
| polarC18 | 276.144 | 169.8899 | 6.1 | 0.12 |
| polarC18 | 276.144 | 102.819 | 6.1 | 0.12 |
| polarC18 | 276.144 | 84.5839 | 6.1 | 0.12 |
| polarC18 | 276.144 | 137.2438 | 6.1 | 0.12 |
| polarC18 | 276.144 | 167.1766 | 6.1 | 0.12 |
| polarC18 | 276.144 | 126.6385 | 6.1 | 0.12 |
| polarC18 | 276.144 | 110.3404 | 6.1 | 0.12 |
| polarC18 | 276.144 | 88.8061 | 6.1 | 0.12 |
| polarC18 | 276.144 | 65.3506 | 6.1 | 0.12 |
| polarC18 | 276.144 | 86.5351 | 6.1 | 0.12 |
| polarC18 | 276.144 | 62.6901 | 6.1 | 0.12 |
| polarC18 | 276.144 | 173.8171 | 6.1 | 0.12 |
| polarC18 | 276.144 | 108.0561 | 6.1 | 0.12 |
| polarC18 | 276.144 | 114.0493 | 6.1 | 0.12 |
| polarC18 | 276.144 | 182.5076 | 6.1 | 0.12 |

| | | | | |
|---|---|---|---|---|
| polarC18 | 276.144 | 129.8265 | 6.1 | 0.12 |
| polarC18 | 276.144 | 69.2815 | 6.1 | 0.12 |
| polarC18 | 276.144 | 56.8152 | 6.1 | 0.12 |
| polarC18 | 276.144 | 80.699 | 6.1 | 0.12 |
| polarC18 | 276.144 | 129.5744 | 6.1 | 0.12 |
| polarC18 | 276.144 | 194.8991 | 6.1 | 0.12 |
| polarC18 | 276.144 | 195.665 | 6.1 | 0.12 |
| polarC18 | 276.144 | 50.7479 | 6.1 | 0.12 |
| polarC18 | 276.144 | 89.8741 | 6.1 | 0.12 |
| polarC18 | 276.144 | 84.1995 | 6.1 | 0.12 |
| polarC18 | 276.144 | 86.3187 | 6.1 | 0.12 |
| polarC18 | 276.144 | 150.6269 | 6.1 | 0.12 |
| polarC18 | 276.144 | 58.7445 | 6.1 | 0.12 |
| polarC18 | 276.144 | 50.0195 | 6.1 | 0.12 |
| polarC18 | 276.144 | 119.6361 | 6.1 | 0.12 |
| polarC18 | 276.144 | 81.3843 | 6.1 | 0.12 |
| polarC18 | 276.144 | 199.2485 | 6.1 | 0.12 |
| polarC18 | 276.144 | 109.8655 | 6.1 | 0.12 |
| polarC18 | 276.144 | 63.4147 | 6.1 | 0.12 |
| polarC18 | 276.144 | 66.7253 | 6.1 | 0.12 |
| polarC18 | 276.144 | 124.9464 | 6.1 | 0.12 |
| polarC18 | 276.144 | 125.9875 | 6.1 | 0.12 |
| polarC18 | 276.144 | 59.6959 | 6.1 | 0.12 |
| polarC18 | 276.144 | 84.0216 | 6.1 | 0.12 |
| polarC18 | 276.144 | 60.5297 | 6.1 | 0.12 |
| polarC18 | 276.144 | 108.1141 | 6.1 | 0.12 |
| polarC18 | 276.144 | 193.8121 | 6.1 | 0.12 |
| polarC18 | 276.144 | 115.6378 | 6.1 | 0.12 |
| polarC18 | 276.144 | 64.5916 | 6.1 | 0.12 |
| polarC18 | 276.144 | 67.9115 | 6.1 | 0.12 |
| polarC18 | 276.144 | 283.0303 | 6.1 | 0.12 |
| polarC18 | 276.144 | 120.9172 | 6.1 | 0.12 |
| polarC18 | 276.144 | 105.9108 | 6.1 | 0.12 |
| polarC18 | 276.144 | 144.939 | 6.1 | 0.12 |
| polarC18 | 276.144 | 69.8886 | 6.1 | 0.12 |
| polarC18 | 276.144 | 55.8623 | 6.1 | 0.12 |
| polarC18 | 276.144 | 55.1204 | 6.1 | 0.12 |
| polarC18 | 276.144 | 151.5736 | 6.1 | 0.12 |
| polarC18 | 276.144 | 229.3691 | 6.1 | 0.12 |
| polarC18 | 276.144 | 77.3894 | 6.1 | 0.12 |
| polarC18 | 276.144 | 51.1969 | 6.1 | 0.12 |
| polarC18 | 276.144 | 51.8052 | 6.1 | 0.12 |

| | | | | |
|---|---|---|---|---|
| polarC18 | 276.144 | 74.2211 | 6.1 | 0.12 |
| polarC18 | 276.144 | 77.1562 | 6.1 | 0.12 |
| polarC18 | 276.144 | 64.6348 | 6.1 | 0.12 |
| polarC18 | 276.144 | 73.278 | 6.1 | 0.12 |
| polarC18 | 276.144 | 107.5875 | 6.1 | 0.12 |
| polarC18 | 276.144 | 106.9704 | 6.1 | 0.12 |
| polarC18 | 276.144 | 68.3244 | 6.1 | 0.12 |
| polarC18 | 276.144 | 105.4594 | 6.1 | 0.12 |
| polarC18 | 276.144 | 109.8878 | 6.1 | 0.12 |
| polarC18 | 276.144 | 114.1531 | 6.1 | 0.12 |
| polarC18 | 276.144 | 146.8349 | 6.1 | 0.12 |
| polarC18 | 276.144 | 156.5936 | 6.1 | 0.12 |
| polarC18 | 276.144 | 103.6172 | 6.1 | 0.12 |
| polarC18 | 276.144 | 51.4268 | 6.1 | 0.12 |
| polarC18 | 276.144 | 59.6091 | 6.1 | 0.12 |
| polarC18 | 276.144 | 52.3357 | 6.1 | 0.12 |
| polarC18 | 276.144 | 52.6498 | 6.1 | 0.12 |
| polarC18 | 276.144 | 91.4878 | 6.1 | 0.12 |
| polarC18 | 276.144 | 88.7334 | 6.1 | 0.12 |
| polarC18 | 276.144 | 169.1718 | 6.1 | 0.12 |
| polarC18 | 276.144 | 202.0541 | 6.1 | 0.12 |
| polarC18 | 276.144 | 184.2261 | 6.1 | 0.12 |
| polarC18 | 276.144 | 185.2021 | 6.1 | 0.12 |
| polarC18 | 276.144 | 57.6672 | 6.1 | 0.12 |
| polarC18 | 276.144 | 106.3703 | 6.1 | 0.12 |
| polarC18 | 276.144 | 54.1098 | 6.1 | 0.12 |
| polarC18 | 276.144 | 103.7134 | 6.1 | 0.12 |
| polarC18 | 276.144 | 79.6614 | 6.1 | 0.12 |
| polarC18 | 276.144 | 66.6344 | 6.1 | 0.12 |
| polarC18 | 276.144 | 55.2141 | 6.1 | 0.12 |
| polarC18 | 276.144 | 74.5979 | 6.1 | 0.12 |
| polarC18 | 276.144 | 103.4484 | 6.1 | 0.12 |
| polarC18 | 276.144 | 148.3502 | 6.1 | 0.12 |
| polarC18 | 276.144 | 86.8413 | 6.1 | 0.12 |
| polarC18 | 276.144 | 70.2892 | 6.1 | 0.12 |
| polarC18 | 276.144 | 165.2912 | 6.1 | 0.12 |
| polarC18 | 276.144 | 78.598 | 6.1 | 0.12 |
| polarC18 | 276.144 | 76.0919 | 6.1 | 0.12 |
| polarC18 | 276.144 | 118.3569 | 6.1 | 0.12 |
| polarC18 | 276.144 | 114.4218 | 6.1 | 0.12 |
| polarC18 | 276.144 | 181.5278 | 6.1 | 0.12 |
| polarC18 | 276.144 | 71.9429 | 6.1 | 0.12 |

| | | | | |
|---|---|---|---|---|
| polarC18 | 276.144 | 92.5473 | 6.1 | 0.12 |
| polarC18 | 276.144 | 77.8639 | 6.1 | 0.12 |
| polarC18 | 276.144 | 246.0204 | 6.1 | 0.12 |
| polarC18 | 276.144 | 60.865 | 6.1 | 0.12 |
| polarC18 | 276.144 | 62.9961 | 6.1 | 0.12 |
| polarC18 | 276.144 | 58.0609 | 6.1 | 0.12 |
| polarC18 | 276.144 | 61.8382 | 6.1 | 0.12 |
| polarC18 | 276.144 | 78.3748 | 6.1 | 0.12 |
| polarC18 | 276.144 | 151.7457 | 6.1 | 0.12 |
| polarC18 | 276.144 | 122.1383 | 6.1 | 0.12 |
| polarC18 | 276.144 | 122.3113 | 6.1 | 0.12 |
| polarC18 | 276.144 | 89.1347 | 6.1 | 0.12 |
| polarC18 | 276.144 | 62.1604 | 6.1 | 0.12 |
| polarC18 | 276.144 | 102.5081 | 6 | 0.12 |
| polarC18 | 276.144 | 60.5543 | 6 | 0.12 |
| polarC18 | 276.144 | 157.4727 | 6 | 0.12 |
| polarC18 | 276.144 | 56.5075 | 6 | 0.12 |
| polarC18 | 276.144 | 84.4009 | 6 | 0.12 |
| polarC18 | 276.144 | 202.8316 | 6 | 0.12 |
| polarC18 | 276.144 | 127.1016 | 6 | 0.12 |
| polarC18 | 276.144 | 110.3131 | 6 | 0.12 |
| polarC18 | 276.144 | 78.61 | 6 | 0.12 |
| polarC18 | 276.144 | 123.7827 | 6 | 0.12 |
| polarC18 | 276.144 | 120.5857 | 6 | 0.12 |
| polarC18 | 276.144 | 51.4596 | 6 | 0.12 |
| polarC18 | 276.144 | 85.6539 | 6 | 0.12 |
| polarC18 | 276.144 | 83.8509 | 6 | 0.12 |
| polarC18 | 276.144 | 58.9498 | 6 | 0.12 |
| polarC18 | 276.144 | 89.7919 | 6 | 0.12 |
| polarC18 | 276.144 | 204.1737 | 6 | 0.12 |
| polarC18 | 276.144 | 94.6472 | 6 | 0.12 |
| polarC18 | 276.144 | 184.8852 | 6 | 0.12 |
| polarC18 | 276.144 | 75.061 | 6 | 0.12 |
| polarC18 | 276.144 | 66.1234 | 6 | 0.12 |
| polarC18 | 276.144 | 80.6807 | 6 | 0.12 |
| polarC18 | 276.144 | 71.4502 | 6 | 0.12 |
| polarC18 | 276.144 | 193.4229 | 6 | 0.12 |
| polarC18 | 276.144 | 121.7353 | 6 | 0.12 |
| polarC18 | 276.144 | 85.2234 | 6 | 0.12 |
| polarC18 | 276.144 | 89.786 | 6 | 0.12 |
| polarC18 | 276.144 | 74.2354 | 6 | 0.12 |
| polarC18 | 276.144 | 74.58 | 6 | 0.12 |

| | | | | |
|---|---|---|---|---|
| polarC18 | 276.144 | 77.9912 | 6 | 0.12 |
| polarC18 | 276.144 | 60.0177 | 6 | 0.12 |
| polarC18 | 276.144 | 152.2225 | 6 | 0.12 |
| polarC18 | 276.144 | 128.1768 | 6 | 0.12 |
| polarC18 | 276.144 | 116.1589 | 6 | 0.12 |
| polarC18 | 276.144 | 65.4 | 6 | 0.12 |
| polarC18 | 276.144 | 211.1894 | 6 | 0.12 |
| polarC18 | 276.144 | 55.608 | 6 | 0.12 |
| polarC18 | 276.144 | 74.2939 | 6 | 0.12 |
| polarC18 | 276.144 | 50.1529 | 6 | 0.12 |
| polarC18 | 276.144 | 87.0514 | 6 | 0.12 |
| polarC18 | 276.144 | 260.0399 | 6 | 0.12 |
| polarC18 | 276.144 | 146.5005 | 6 | 0.12 |
| polarC18 | 276.144 | 55.8448 | 6 | 0.12 |
| polarC18 | 276.144 | 151.813 | 6 | 0.12 |
| polarC18 | 276.144 | 172.8314 | 6 | 0.12 |
| polarC18 | 276.144 | 88.5038 | 6 | 0.12 |
| polarC18 | 276.144 | 99.507 | 6 | 0.12 |
| polarC18 | 276.144 | 266.4109 | 6 | 0.12 |
| polarC18 | 276.144 | 53.3291 | 6 | 0.12 |
| polarC18 | 276.144 | 88.9662 | 6 | 0.12 |
| polarC18 | 276.144 | 69.6506 | 6 | 0.12 |
| polarC18 | 276.144 | 59.6014 | 6 | 0.12 |
| polarC18 | 276.144 | 113.7958 | 6 | 0.12 |
| polarC18 | 276.144 | 102.0586 | 6 | 0.12 |
| polarC18 | 276.144 | 120.8271 | 6 | 0.12 |
| polarC18 | 276.144 | 69.4884 | 6 | 0.12 |
| polarC18 | 276.144 | 177.0774 | 6 | 0.12 |
| polarC18 | 276.144 | 73.7899 | 6 | 0.12 |
| polarC18 | 276.144 | 105.3223 | 6 | 0.12 |
| polarC18 | 276.144 | 111.7213 | 6 | 0.12 |
| polarC18 | 276.144 | 74.4788 | 6 | 0.12 |
| polarC18 | 276.144 | 63.5538 | 6 | 0.12 |
| polarC18 | 276.144 | 78.2366 | 6 | 0.12 |
| polarC18 | 276.144 | 64.3644 | 6 | 0.12 |
| polarC18 | 276.144 | 52.3798 | 6 | 0.12 |
| polarC18 | 276.144 | 71.2166 | 6 | 0.12 |
| polarC18 | 276.144 | 171.2463 | 6 | 0.12 |
| polarC18 | 276.144 | 83.9849 | 6 | 0.12 |
| polarC18 | 276.144 | 75.5066 | 6 | 0.12 |
| polarC18 | 276.144 | 171.1223 | 6 | 0.12 |
| polarC18 | 276.144 | 53.2902 | 6 | 0.12 |

| | | | | |
|---|---|---|---|---|
| polarC18 | 276.144 | 68.655 | 6 | 0.12 |
| polarC18 | 276.144 | 55.6519 | 6 | 0.12 |
| polarC18 | 276.144 | 50.4218 | 6 | 0.12 |
| polarC18 | 276.144 | 126.758 | 6 | 0.12 |
| polarC18 | 276.144 | 78.3138 | 6 | 0.12 |
| polarC18 | 276.144 | 76.4017 | 6 | 0.12 |
| polarC18 | 276.144 | 84.711 | 6 | 0.12 |
| polarC18 | 276.144 | 144.3399 | 6 | 0.12 |
| polarC18 | 276.144 | 60.7335 | 6 | 0.12 |
| polarC18 | 276.144 | 50.441 | 6 | 0.12 |
| polarC18 | 276.144 | 118.7626 | 6 | 0.12 |
| polarC18 | 276.144 | 53.5312 | 6 | 0.12 |
| polarC18 | 276.144 | 168.9585 | 6 | 0.12 |
| polarC18 | 276.144 | 51.6092 | 6 | 0.12 |
| polarC18 | 276.144 | 103.4124 | 6 | 0.12 |
| polarC18 | 276.144 | 81.3884 | 6 | 0.12 |
| polarC18 | 276.144 | 66.0158 | 6 | 0.12 |
| polarC18 | 276.144 | 70.1847 | 6 | 0.12 |
| polarC18 | 276.144 | 121.7207 | 6 | 0.12 |
| polarC18 | 276.144 | 51.583 | 6 | 0.12 |
| polarC18 | 276.144 | 50.1956 | 6 | 0.12 |
| polarC18 | 276.144 | 53.7797 | 6 | 0.12 |
| polarC18 | 276.144 | 95.3361 | 6 | 0.12 |
| polarC18 | 276.144 | 103.5488 | 6 | 0.12 |
| polarC18 | 276.144 | 52.289 | 6 | 0.12 |
| polarC18 | 276.144 | 138.625 | 6 | 0.11 |
| polarC18 | 276.144 | 54.2069 | 6 | 0.11 |
| polarC18 | 276.144 | 167.856 | 6 | 0.11 |
| polarC18 | 276.144 | 93.3822 | 6 | 0.11 |
| polarC18 | 276.144 | 81.4273 | 6 | 0.11 |
| polarC18 | 276.144 | 246.1636 | 6 | 0.11 |
| polarC18 | 276.144 | 158.2268 | 6 | 0.11 |
| polarC18 | 276.144 | 66.736 | 6 | 0.11 |
| polarC18 | 276.144 | 124.1822 | 6 | 0.11 |
| polarC18 | 276.144 | 70.6143 | 6 | 0.11 |
| polarC18 | 276.144 | 51.6988 | 6 | 0.11 |
| polarC18 | 276.144 | 140.2531 | 6 | 0.11 |
| polarC18 | 276.144 | 52.0875 | 6 | 0.11 |
| polarC18 | 276.144 | 59.0753 | 6 | 0.11 |
| polarC18 | 276.144 | 261.3432 | 6 | 0.11 |
| polarC18 | 276.144 | 80.4451 | 6 | 0.11 |
| polarC18 | 276.144 | 63.0706 | 6 | 0.11 |

| | | | | |
|---|---|---|---|---|
| polarC18 | 276.144 | 99.7037 | 6 | 0.11 |
| polarC18 | 276.144 | 62.6753 | 6 | 0.11 |
| polarC18 | 276.144 | 227.7919 | 6 | 0.11 |
| polarC18 | 276.144 | 150.0759 | 6 | 0.11 |
| polarC18 | 276.144 | 128.3207 | 6 | 0.11 |
| polarC18 | 276.144 | 91.7677 | 6 | 0.11 |
| polarC18 | 276.144 | 54.0726 | 6 | 0.11 |
| polarC18 | 276.144 | 135.4021 | 6 | 0.11 |
| polarC18 | 276.144 | 113.6573 | 6 | 0.11 |
| polarC18 | 276.144 | 100.1246 | 6 | 0.11 |
| polarC18 | 276.144 | 179.4522 | 5.9 | 0.11 |
| polarC18 | 276.144 | 58.5781 | 5.9 | 0.11 |
| polarC18 | 276.144 | 116.9308 | 5.9 | 0.11 |
| polarC18 | 276.144 | 58.758 | 5.9 | 0.11 |
| polarC18 | 276.144 | 50.3286 | 5.9 | 0.11 |
| polarC18 | 276.144 | 76.9831 | 5.9 | 0.11 |
| polarC18 | 276.144 | 51.4619 | 5.9 | 0.11 |
| polarC18 | 276.144 | 66.466 | 5.9 | 0.11 |
| polarC18 | 276.144 | 66.5757 | 5.9 | 0.11 |
| polarC18 | 276.144 | 146.3978 | 5.9 | 0.11 |
| polarC18 | 276.144 | 73.0162 | 5.9 | 0.11 |
| polarC18 | 276.144 | 72.7004 | 5.9 | 0.11 |
| polarC18 | 276.144 | 127.6832 | 5.9 | 0.11 |
| polarC18 | 276.144 | 154.5904 | 5.9 | 0.11 |
| polarC18 | 276.144 | 103.2245 | 5.9 | 0.11 |
| polarC18 | 276.144 | 60.7791 | 5.9 | 0.11 |
| polarC18 | 276.144 | 61.8277 | 5.9 | 0.11 |
| polarC18 | 276.144 | 255.1255 | 5.9 | 0.11 |
| polarC18 | 276.144 | 118.5899 | 5.9 | 0.11 |
| polarC18 | 276.144 | 62.1574 | 5.9 | 0.11 |
| polarC18 | 276.144 | 61.1964 | 5.9 | 0.11 |
| polarC18 | 276.144 | 55.5617 | 5.9 | 0.11 |
| polarC18 | 276.144 | 103.5362 | 5.9 | 0.11 |
| polarC18 | 276.144 | 50.8148 | 5.9 | 0.11 |
| polarC18 | 276.144 | 71.716 | 5.9 | 0.11 |
| polarC18 | 276.144 | 51.7091 | 5.9 | 0.11 |
| polarC18 | 276.144 | 58.3246 | 5.9 | 0.11 |
| polarC18 | 276.144 | 140.7831 | 5.9 | 0.11 |
| polarC18 | 276.144 | 74.1337 | 5.9 | 0.11 |
| polarC18 | 276.144 | 168.3086 | 5.9 | 0.11 |
| polarC18 | 276.144 | 187.1207 | 5.9 | 0.11 |
| polarC18 | 276.144 | 86.6996 | 5.9 | 0.11 |

| | | | | |
|---|---|---|---|---|
| polarC18 | 276.144 | 59.7556 | 5.9 | 0.11 |
| polarC18 | 276.144 | 81.0616 | 5.9 | 0.11 |
| polarC18 | 276.144 | 151.0185 | 5.9 | 0.11 |
| polarC18 | 276.144 | 85.7935 | 5.9 | 0.11 |
| polarC18 | 276.144 | 62.3646 | 5.9 | 0.11 |
| polarC18 | 276.144 | 53.2781 | 5.9 | 0.11 |
| polarC18 | 276.144 | 70.9472 | 5.9 | 0.11 |
| polarC18 | 276.144 | 270.9909 | 5.9 | 0.11 |
| polarC18 | 276.144 | 263.3889 | 5.9 | 0.11 |
| polarC18 | 276.144 | 53.6968 | 5.9 | 0.11 |
| polarC18 | 276.144 | 92.4311 | 5.9 | 0.11 |
| polarC18 | 276.144 | 78.8499 | 5.9 | 0.11 |
| polarC18 | 276.144 | 102.4208 | 5.9 | 0.11 |
| polarC18 | 276.144 | 66.7658 | 5.9 | 0.11 |
| polarC18 | 276.144 | 69.6802 | 5.9 | 0.11 |
| polarC18 | 276.144 | 64.6198 | 5.9 | 0.11 |
| polarC18 | 276.144 | 51.4946 | 5.9 | 0.11 |
| polarC18 | 276.144 | 58.0319 | 5.9 | 0.11 |
| polarC18 | 276.144 | 199.3023 | 5.9 | 0.11 |
| polarC18 | 276.144 | 192.8767 | 5.9 | 0.11 |
| polarC18 | 276.144 | 58.4478 | 5.9 | 0.11 |
| polarC18 | 276.144 | 83.6368 | 5.9 | 0.11 |
| polarC18 | 276.144 | 105.3958 | 5.9 | 0.11 |
| polarC18 | 276.144 | 104.1737 | 5.9 | 0.11 |
| polarC18 | 276.144 | 141.5522 | 5.9 | 0.11 |
| polarC18 | 276.144 | 105.3773 | 5.9 | 0.11 |
| polarC18 | 276.144 | 66.8043 | 5.9 | 0.11 |
| polarC18 | 276.144 | 158.3322 | 5.9 | 0.11 |
| polarC18 | 276.144 | 152.9272 | 5.9 | 0.11 |
| polarC18 | 276.144 | 66.9033 | 5.9 | 0.11 |
| polarC18 | 276.144 | 266.9762 | 5.9 | 0.11 |
| polarC18 | 276.144 | 55.2016 | 5.9 | 0.11 |
| polarC18 | 276.144 | 53.2508 | 5.9 | 0.11 |
| polarC18 | 276.144 | 73.6272 | 5.9 | 0.11 |
| polarC18 | 276.144 | 67.1301 | 5.9 | 0.11 |
| polarC18 | 276.144 | 51.7467 | 5.9 | 0.11 |
| polarC18 | 276.144 | 58.9107 | 5.9 | 0.11 |
| polarC18 | 276.144 | 110.9753 | 5.9 | 0.11 |
| polarC18 | 276.144 | 85.3226 | 5.9 | 0.11 |
| polarC18 | 276.144 | 89.1488 | 5.9 | 0.11 |
| polarC18 | 276.144 | 55.0965 | 5.9 | 0.11 |
| polarC18 | 276.144 | 53.5044 | 5.9 | 0.11 |

| | | | | |
|---|---|---|---|---|
| polarC18 | 276.144 | 89.5605 | 5.9 | 0.11 |
| polarC18 | 276.144 | 88.014 | 5.9 | 0.11 |
| polarC18 | 276.144 | 111.8704 | 5.9 | 0.11 |
| polarC18 | 276.144 | 73.2434 | 5.9 | 0.11 |
| polarC18 | 276.144 | 76.4211 | 5.9 | 0.11 |
| polarC18 | 276.144 | 80.3368 | 5.9 | 0.11 |
| polarC18 | 276.144 | 120.2124 | 5.9 | 0.11 |
| polarC18 | 276.144 | 189.2934 | 5.9 | 0.11 |
| polarC18 | 276.144 | 143.1158 | 5.9 | 0.11 |
| polarC18 | 276.144 | 58.7645 | 5.9 | 0.11 |
| polarC18 | 276.144 | 98.0584 | 5.9 | 0.11 |
| polarC18 | 276.144 | 252.5193 | 5.9 | 0.11 |
| polarC18 | 276.144 | 53.5617 | 5.9 | 0.11 |
| polarC18 | 276.144 | 172.49 | 5.9 | 0.11 |
| polarC18 | 276.144 | 115.8513 | 5.9 | 0.11 |
| polarC18 | 276.144 | 131.2818 | 5.9 | 0.11 |
| polarC18 | 276.144 | 127.355 | 5.9 | 0.11 |
| polarC18 | 276.144 | 103.2862 | 5.9 | 0.11 |
| polarC18 | 276.144 | 119.0523 | 5.9 | 0.11 |
| polarC18 | 276.144 | 247.7697 | 5.9 | 0.11 |
| polarC18 | 276.144 | 262.9435 | 5.9 | 0.11 |
| polarC18 | 276.144 | 141.6096 | 5.9 | 0.11 |
| polarC18 | 276.144 | 50.6493 | 5.9 | 0.11 |
| polarC18 | 276.144 | 101.8903 | 5.9 | 0.11 |
| polarC18 | 276.144 | 97.0467 | 5.9 | 0.11 |
| polarC18 | 276.144 | 126.944 | 5.9 | 0.11 |
| polarC18 | 276.144 | 58.2948 | 5.9 | 0.11 |
| polarC18 | 276.144 | 78.6459 | 5.9 | 0.11 |
| polarC18 | 276.144 | 178.79 | 5.9 | 0.11 |
| polarC18 | 276.144 | 53.6073 | 5.9 | 0.11 |
| polarC18 | 276.144 | 181.4783 | 5.9 | 0.11 |
| polarC18 | 276.144 | 86.7818 | 5.9 | 0.11 |
| polarC18 | 276.144 | 131.5426 | 5.9 | 0.11 |
| polarC18 | 276.144 | 86.0314 | 5.8 | 0.11 |
| polarC18 | 276.144 | 65.1179 | 5.8 | 0.11 |
| polarC18 | 276.144 | 58.1557 | 5.8 | 0.11 |
| polarC18 | 276.144 | 60.0534 | 5.8 | 0.11 |
| polarC18 | 276.144 | 51.5191 | 5.8 | 0.11 |
| polarC18 | 276.144 | 298.6515 | 5.8 | 0.11 |
| polarC18 | 276.144 | 119.5668 | 5.8 | 0.11 |
| polarC18 | 276.144 | 88.8906 | 5.8 | 0.11 |
| polarC18 | 276.144 | 173.9162 | 5.8 | 0.11 |

| | | | | |
|---|---|---|---|---|
| polarC18 | 276.144 | 124.442 | 5.8 | 0.11 |
| polarC18 | 276.144 | 61.4673 | 5.8 | 0.11 |
| polarC18 | 276.144 | 52.5481 | 5.8 | 0.11 |
| polarC18 | 276.144 | 129.5019 | 5.8 | 0.11 |
| polarC18 | 276.144 | 160.1601 | 5.8 | 0.11 |
| polarC18 | 276.144 | 56.1786 | 5.8 | 0.11 |
| polarC18 | 276.144 | 102.5331 | 5.8 | 0.11 |
| polarC18 | 276.144 | 70.1347 | 5.8 | 0.11 |
| polarC18 | 276.144 | 289.9055 | 5.8 | 0.11 |
| polarC18 | 276.144 | 242.4548 | 5.8 | 0.11 |
| polarC18 | 276.144 | 72.96 | 5.8 | 0.11 |
| polarC18 | 276.144 | 70.1595 | 5.8 | 0.11 |
| polarC18 | 276.144 | 67.8626 | 5.8 | 0.11 |
| polarC18 | 276.144 | 83.4123 | 5.8 | 0.11 |
| polarC18 | 276.144 | 77.7594 | 5.8 | 0.11 |
| polarC18 | 276.144 | 105.6735 | 5.8 | 0.11 |
| polarC18 | 276.144 | 77.1412 | 5.8 | 0.11 |
| polarC18 | 276.144 | 58.3958 | 5.8 | 0.11 |
| polarC18 | 276.144 | 75.5134 | 5.8 | 0.11 |
| polarC18 | 276.144 | 58.5087 | 5.8 | 0.11 |
| polarC18 | 276.144 | 68.8147 | 5.8 | 0.11 |
| polarC18 | 276.144 | 67.5709 | 5.8 | 0.11 |
| polarC18 | 276.144 | 64.9963 | 5.8 | 0.11 |
| polarC18 | 276.144 | 194.3306 | 5.8 | 0.11 |
| polarC18 | 276.144 | 162.2658 | 5.8 | 0.11 |
| polarC18 | 276.144 | 78.4547 | 5.8 | 0.11 |
| polarC18 | 276.144 | 68.9119 | 5.8 | 0.11 |
| polarC18 | 276.144 | 100.3888 | 5.8 | 0.11 |
| polarC18 | 276.144 | 55.8294 | 5.8 | 0.11 |
| polarC18 | 276.144 | 144.7813 | 5.8 | 0.11 |
| polarC18 | 276.144 | 152.0686 | 5.8 | 0.11 |
| polarC18 | 276.144 | 111.1984 | 5.8 | 0.11 |
| polarC18 | 276.144 | 72.569 | 5.8 | 0.11 |
| polarC18 | 276.144 | 89.3168 | 5.8 | 0.11 |
| polarC18 | 276.144 | 50.3056 | 5.8 | 0.11 |
| polarC18 | 276.144 | 51.1087 | 5.8 | 0.11 |
| polarC18 | 276.144 | 54.3169 | 5.8 | 0.11 |
| polarC18 | 276.144 | 86.0633 | 5.8 | 0.11 |
| polarC18 | 276.144 | 164.4224 | 5.8 | 0.11 |
| polarC18 | 276.144 | 59.3199 | 5.8 | 0.11 |
| polarC18 | 276.144 | 54.5774 | 5.8 | 0.11 |
| polarC18 | 276.144 | 77.1308 | 5.8 | 0.11 |

| | | | | |
|---|---|---|---|---|
| polarC18 | 276.144 | 199.4334 | 5.8 | 0.11 |
| polarC18 | 276.144 | 71.8717 | 5.8 | 0.11 |
| polarC18 | 276.144 | 53.7079 | 5.8 | 0.11 |
| polarC18 | 276.144 | 275.0795 | 5.8 | 0.11 |
| polarC18 | 276.144 | 64.1714 | 5.8 | 0.11 |
| polarC18 | 276.144 | 54.2875 | 5.8 | 0.11 |
| polarC18 | 276.144 | 54.301 | 5.8 | 0.11 |
| polarC18 | 276.144 | 146.1175 | 5.8 | 0.11 |
| polarC18 | 276.144 | 207.9472 | 5.8 | 0.11 |
| polarC18 | 276.144 | 58.8626 | 5.8 | 0.11 |
| polarC18 | 276.144 | 60.3929 | 5.8 | 0.11 |
| polarC18 | 276.144 | 68.0252 | 5.8 | 0.11 |
| polarC18 | 276.144 | 152.4583 | 5.8 | 0.11 |
| polarC18 | 276.144 | 159.5606 | 5.8 | 0.11 |
| polarC18 | 276.144 | 79.3755 | 5.8 | 0.11 |
| polarC18 | 276.144 | 53.2357 | 5.8 | 0.11 |
| polarC18 | 276.144 | 68.1524 | 5.8 | 0.11 |
| polarC18 | 276.144 | 169.289 | 5.8 | 0.11 |
| polarC18 | 276.144 | 152.0963 | 5.8 | 0.11 |
| polarC18 | 276.144 | 282.3945 | 5.8 | 0.11 |
| polarC18 | 276.144 | 96.6028 | 5.8 | 0.11 |
| polarC18 | 276.144 | 64.1512 | 5.8 | 0.11 |
| polarC18 | 276.144 | 131.6979 | 5.8 | 0.11 |
| polarC18 | 276.144 | 76.8569 | 5.8 | 0.11 |
| polarC18 | 276.144 | 115.6107 | 5.8 | 0.11 |
| polarC18 | 276.144 | 154.632 | 5.8 | 0.11 |
| polarC18 | 276.144 | 224.6264 | 5.7 | 0.11 |
| polarC18 | 276.144 | 246.8875 | 5.7 | 0.11 |
| polarC18 | 276.144 | 71.0105 | 5.7 | 0.11 |
| polarC18 | 276.144 | 92.8107 | 5.7 | 0.11 |
| polarC18 | 276.144 | 54.0153 | 5.7 | 0.11 |
| polarC18 | 276.144 | 56.1083 | 5.7 | 0.11 |
| polarC18 | 276.144 | 119.9995 | 5.7 | 0.11 |
| polarC18 | 276.144 | 117.6661 | 5.7 | 0.11 |
| polarC18 | 276.144 | 81.35 | 5.7 | 0.11 |
| polarC18 | 276.144 | 58.4327 | 5.7 | 0.11 |
| polarC18 | 276.144 | 73.263 | 5.7 | 0.11 |
| polarC18 | 276.144 | 94.985 | 5.7 | 0.11 |
| polarC18 | 276.144 | 130.4538 | 5.7 | 0.11 |
| polarC18 | 276.144 | 130.1697 | 5.7 | 0.11 |
| polarC18 | 276.144 | 87.4249 | 5.7 | 0.11 |
| polarC18 | 276.144 | 134.3115 | 5.7 | 0.11 |

| | | | | |
|---|---|---|---|---|
| polarC18 | 276.144 | 74.0909 | 5.7 | 0.11 |
| polarC18 | 276.144 | 184.6431 | 5.7 | 0.11 |
| polarC18 | 276.144 | 277.7551 | 5.7 | 0.11 |
| polarC18 | 276.144 | 55.9034 | 5.7 | 0.11 |
| polarC18 | 276.144 | 109.3481 | 5.7 | 0.11 |
| polarC18 | 276.144 | 195.5572 | 5.7 | 0.11 |
| polarC18 | 276.144 | 64.6987 | 5.7 | 0.11 |
| polarC18 | 276.144 | 136.1026 | 5.7 | 0.11 |
| polarC18 | 276.144 | 134.5712 | 5.7 | 0.11 |
| polarC18 | 276.144 | 161.1391 | 5.7 | 0.11 |
| polarC18 | 276.144 | 90.3861 | 5.7 | 0.11 |
| polarC18 | 276.144 | 101.4792 | 5.7 | 0.11 |
| polarC18 | 276.144 | 219.2758 | 5.7 | 0.11 |
| polarC18 | 276.144 | 77.8225 | 5.7 | 0.11 |
| polarC18 | 276.144 | 274.309 | 5.7 | 0.11 |
| polarC18 | 276.144 | 72.2955 | 5.7 | 0.11 |
| polarC18 | 276.144 | 50.9196 | 5.7 | 0.11 |
| polarC18 | 276.144 | 56.8098 | 5.7 | 0.11 |
| polarC18 | 276.144 | 97.7396 | 5.7 | 0.11 |
| polarC18 | 276.144 | 71.3445 | 5.7 | 0.11 |
| polarC18 | 276.144 | 80.6678 | 5.7 | 0.11 |
| polarC18 | 276.144 | 72.1639 | 5.7 | 0.11 |
| polarC18 | 276.144 | 83.101 | 5.7 | 0.11 |
| polarC18 | 276.144 | 93.6056 | 5.7 | 0.11 |
| polarC18 | 276.144 | 116.7136 | 5.7 | 0.11 |
| polarC18 | 276.144 | 61.934 | 5.7 | 0.11 |
| polarC18 | 276.144 | 66.3138 | 5.7 | 0.11 |
| polarC18 | 276.144 | 97.6815 | 5.7 | 0.11 |
| polarC18 | 276.144 | 139.8276 | 5.7 | 0.11 |
| polarC18 | 276.144 | 73.206 | 5.7 | 0.11 |
| polarC18 | 276.144 | 266.6997 | 5.7 | 0.11 |
| polarC18 | 276.144 | 268.0602 | 5.7 | 0.11 |
| polarC18 | 276.144 | 181.7351 | 5.7 | 0.11 |
| polarC18 | 276.144 | 161.4642 | 5.7 | 0.11 |
| polarC18 | 276.144 | 74.2268 | 5.7 | 0.11 |
| polarC18 | 276.144 | 60.7732 | 5.7 | 0.11 |
| polarC18 | 276.144 | 154.4349 | 5.7 | 0.11 |
| polarC18 | 276.144 | 60.618 | 5.7 | 0.11 |
| polarC18 | 276.144 | 104.2657 | 5.7 | 0.11 |
| polarC18 | 276.144 | 55.9615 | 5.7 | 0.11 |
| polarC18 | 276.144 | 100.991 | 5.7 | 0.11 |
| polarC18 | 276.144 | 128.5455 | 5.7 | 0.11 |

| | | | | |
|---|---|---|---|---|
| polarC18 | 276.144 | 163.7967 | 5.7 | 0.11 |
| polarC18 | 276.144 | 193.3941 | 5.7 | 0.11 |
| polarC18 | 276.144 | 51.9602 | 5.7 | 0.11 |
| polarC18 | 276.144 | 107.2305 | 5.7 | 0.11 |
| polarC18 | 276.144 | 63.3936 | 5.7 | 0.11 |
| polarC18 | 276.144 | 174.1443 | 5.7 | 0.11 |
| polarC18 | 276.144 | 50.6031 | 5.7 | 0.11 |
| polarC18 | 276.144 | 117.4816 | 5.7 | 0.11 |
| polarC18 | 276.144 | 62.7358 | 5.7 | 0.11 |
| polarC18 | 276.144 | 256.4655 | 5.7 | 0.11 |
| polarC18 | 276.144 | 167.5798 | 5.7 | 0.11 |
| polarC18 | 276.144 | 60.2782 | 5.7 | 0.11 |
| polarC18 | 276.144 | 116.4514 | 5.7 | 0.11 |
| polarC18 | 276.144 | 132.4371 | 5.7 | 0.11 |
| polarC18 | 276.144 | 119.312 | 5.7 | 0.11 |
| polarC18 | 276.144 | 159.3246 | 5.7 | 0.11 |
| polarC18 | 276.144 | 155.7457 | 5.6 | 0.11 |
| polarC18 | 276.144 | 229.4798 | 5.6 | 0.11 |
| polarC18 | 276.144 | 81.7908 | 5.6 | 0.11 |
| polarC18 | 276.144 | 178.0759 | 5.6 | 0.11 |
| polarC18 | 276.144 | 125.682 | 5.6 | 0.11 |
| polarC18 | 276.144 | 268.1853 | 5.6 | 0.11 |
| polarC18 | 276.144 | 55.1161 | 5.6 | 0.11 |
| polarC18 | 276.144 | 66.6675 | 5.6 | 0.11 |
| polarC18 | 276.144 | 54.0475 | 5.6 | 0.11 |
| polarC18 | 276.144 | 86.6882 | 5.6 | 0.11 |
| polarC18 | 276.144 | 141.1917 | 5.6 | 0.11 |
| polarC18 | 276.144 | 71.6637 | 5.6 | 0.11 |
| polarC18 | 276.144 | 89.8891 | 5.6 | 0.11 |
| polarC18 | 276.144 | 172.6201 | 5.6 | 0.11 |
| polarC18 | 276.144 | 203.0779 | 5.6 | 0.11 |
| polarC18 | 276.144 | 139.3991 | 5.6 | 0.11 |
| polarC18 | 276.144 | 60.4657 | 5.6 | 0.11 |
| polarC18 | 276.144 | 102.9491 | 5.6 | 0.11 |
| polarC18 | 276.144 | 131.8764 | 5.6 | 0.11 |
| polarC18 | 276.144 | 83.4214 | 5.6 | 0.11 |
| polarC18 | 276.144 | 68.7022 | 5.6 | 0.11 |
| polarC18 | 276.144 | 85.3766 | 5.6 | 0.11 |
| polarC18 | 276.144 | 65.8613 | 5.6 | 0.11 |
| polarC18 | 276.144 | 137.0683 | 5.6 | 0.11 |
| polarC18 | 276.144 | 63.4962 | 5.6 | 0.11 |
| polarC18 | 276.144 | 67.7325 | 5.6 | 0.11 |

| | | | | |
|---|---|---|---|---|
| polarC18 | 276.144 | 56.1657 | 5.6 | 0.11 |
| polarC18 | 276.144 | 110.7365 | 5.6 | 0.11 |
| polarC18 | 276.144 | 132.8442 | 5.6 | 0.11 |
| polarC18 | 276.144 | 66.7402 | 5.6 | 0.11 |
| polarC18 | 276.144 | 80.2356 | 5.6 | 0.11 |
| polarC18 | 276.144 | 101.9793 | 5.6 | 0.11 |
| polarC18 | 276.144 | 86.2899 | 5.6 | 0.11 |
| polarC18 | 276.144 | 99.5925 | 5.6 | 0.11 |
| polarC18 | 276.144 | 55.8392 | 5.6 | 0.11 |
| polarC18 | 276.144 | 266.8478 | 5.6 | 0.11 |
| polarC18 | 276.144 | 63.2122 | 5.6 | 0.11 |
| polarC18 | 276.144 | 109.5182 | 5.6 | 0.11 |
| polarC18 | 276.144 | 121.4395 | 5.6 | 0.11 |
| polarC18 | 276.144 | 89.1232 | 5.6 | 0.11 |
| polarC18 | 276.144 | 52.1174 | 5.6 | 0.11 |
| polarC18 | 276.144 | 85.8023 | 5.6 | 0.11 |
| polarC18 | 276.144 | 87.033 | 5.6 | 0.11 |
| polarC18 | 276.144 | 56.0192 | 5.6 | 0.11 |
| polarC18 | 276.144 | 51.7269 | 5.6 | 0.11 |
| polarC18 | 276.144 | 50.9304 | 5.6 | 0.11 |
| polarC18 | 276.144 | 278.7529 | 5.6 | 0.11 |
| polarC18 | 276.144 | 166.6694 | 5.6 | 0.11 |
| polarC18 | 276.144 | 56.9701 | 5.6 | 0.11 |
| polarC18 | 276.144 | 58.1072 | 5.6 | 0.11 |
| polarC18 | 276.144 | 72.0339 | 5.6 | 0.11 |
| polarC18 | 276.144 | 63.5963 | 5.6 | 0.11 |
| polarC18 | 276.144 | 285.916 | 5.6 | 0.11 |
| polarC18 | 276.144 | 102.0338 | 5.6 | 0.11 |
| polarC18 | 276.144 | 64.761 | 5.6 | 0.11 |
| polarC18 | 276.144 | 81.4664 | 5.6 | 0.11 |
| polarC18 | 276.144 | 67.0379 | 5.6 | 0.11 |
| polarC18 | 276.144 | 70.7228 | 5.6 | 0.11 |
| polarC18 | 276.144 | 269.9861 | 5.6 | 0.11 |
| polarC18 | 276.144 | 54.8724 | 5.6 | 0.11 |
| polarC18 | 276.144 | 141.3969 | 5.6 | 0.11 |
| polarC18 | 276.144 | 54.3096 | 5.6 | 0.11 |
| polarC18 | 276.144 | 56.6873 | 5.6 | 0.11 |
| polarC18 | 276.144 | 76.5771 | 5.6 | 0.11 |
| polarC18 | 276.144 | 79.2022 | 5.6 | 0.11 |
| polarC18 | 276.144 | 98.7182 | 5.6 | 0.11 |
| polarC18 | 276.144 | 81.4602 | 5.6 | 0.11 |
| polarC18 | 276.144 | 74.6828 | 5.6 | 0.11 |

| | | | | |
|---|---|---|---|---|
| polarC18 | 276.144 | 116.879 | 5.6 | 0.11 |
| polarC18 | 276.144 | 53.8284 | 5.6 | 0.11 |
| polarC18 | 276.144 | 79.8857 | 5.6 | 0.11 |
| polarC18 | 276.144 | 134.8801 | 5.6 | 0.11 |
| polarC18 | 276.144 | 92.9997 | 5.6 | 0.11 |
| polarC18 | 276.144 | 55.678 | 5.6 | 0.11 |
| polarC18 | 276.144 | 60.1196 | 5.5 | 0.11 |
| polarC18 | 276.144 | 267.5311 | 5.5 | 0.11 |
| polarC18 | 276.144 | 60.1507 | 5.5 | 0.11 |
| polarC18 | 276.144 | 124.2949 | 5.5 | 0.11 |
| polarC18 | 276.144 | 51.9068 | 5.5 | 0.11 |
| polarC18 | 276.144 | 97.6612 | 5.5 | 0.11 |
| polarC18 | 276.144 | 76.1573 | 5.5 | 0.11 |
| polarC18 | 276.144 | 206.8229 | 5.5 | 0.11 |
| polarC18 | 276.144 | 101.1986 | 5.5 | 0.11 |
| polarC18 | 276.144 | 143.9835 | 5.5 | 0.11 |
| polarC18 | 276.144 | 144.0911 | 5.5 | 0.11 |
| polarC18 | 276.144 | 88.6627 | 5.5 | 0.11 |
| polarC18 | 276.144 | 54.1757 | 5.5 | 0.11 |
| polarC18 | 276.144 | 104.5457 | 5.5 | 0.11 |
| polarC18 | 276.144 | 296.9575 | 5.5 | 0.11 |
| polarC18 | 276.144 | 284.5215 | 5.5 | 0.11 |
| polarC18 | 276.144 | 86.681 | 5.5 | 0.11 |
| polarC18 | 276.144 | 62.6929 | 5.5 | 0.11 |
| polarC18 | 276.144 | 52.4491 | 5.5 | 0.11 |
| polarC18 | 276.144 | 111.1032 | 5.5 | 0.11 |
| polarC18 | 276.144 | 98.6752 | 5.5 | 0.11 |
| polarC18 | 276.144 | 193.8359 | 5.5 | 0.11 |
| polarC18 | 276.144 | 121.0715 | 5.5 | 0.11 |
| polarC18 | 276.144 | 112.1672 | 5.5 | 0.11 |
| polarC18 | 276.144 | 72.0729 | 5.5 | 0.11 |
| polarC18 | 276.144 | 119.9494 | 5.5 | 0.11 |
| polarC18 | 276.144 | 77.0677 | 5.5 | 0.11 |
| polarC18 | 276.144 | 68.2548 | 5.5 | 0.11 |
| polarC18 | 276.144 | 53.9936 | 5.5 | 0.11 |
| polarC18 | 276.144 | 79.6947 | 5.5 | 0.11 |
| polarC18 | 276.144 | 78.4897 | 5.5 | 0.11 |
| polarC18 | 276.144 | 81.7452 | 5.5 | 0.11 |
| polarC18 | 276.144 | 61.86 | 5.5 | 0.11 |
| polarC18 | 276.144 | 82.801 | 5.5 | 0.11 |
| polarC18 | 276.144 | 158.0394 | 5.5 | 0.11 |
| polarC18 | 276.144 | 100.9792 | 5.5 | 0.11 |

| | | | | |
|---|---|---|---|---|
| polarC18 | 276.144 | 66.1536 | 5.5 | 0.11 |
| polarC18 | 276.144 | 123.8076 | 5.5 | 0.11 |
| polarC18 | 276.144 | 234.5326 | 5.5 | 0.11 |
| polarC18 | 276.144 | 100.108 | 5.5 | 0.11 |
| polarC18 | 276.144 | 65.3174 | 5.5 | 0.11 |
| polarC18 | 276.144 | 63.9508 | 5.5 | 0.11 |
| polarC18 | 276.144 | 86.4433 | 5.5 | 0.11 |
| polarC18 | 276.144 | 66.4299 | 5.5 | 0.11 |
| polarC18 | 276.144 | 61.5542 | 5.5 | 0.11 |
| polarC18 | 276.144 | 79.7362 | 5.5 | 0.11 |
| polarC18 | 276.144 | 85.8255 | 5.5 | 0.11 |
| polarC18 | 276.144 | 135.4127 | 5.5 | 0.11 |
| polarC18 | 276.144 | 163.525 | 5.5 | 0.11 |
| polarC18 | 276.144 | 244.8634 | 5.5 | 0.11 |
| polarC18 | 276.144 | 71.2989 | 5.5 | 0.11 |
| polarC18 | 276.144 | 235.6857 | 5.5 | 0.11 |
| polarC18 | 276.144 | 239.9059 | 5.5 | 0.11 |
| polarC18 | 276.144 | 70.1001 | 5.5 | 0.11 |
| polarC18 | 276.144 | 83.6484 | 5.5 | 0.11 |
| polarC18 | 276.144 | 56.3843 | 5.5 | 0.11 |
| polarC18 | 276.144 | 253.143 | 5.5 | 0.11 |
| polarC18 | 276.144 | 278.3938 | 5.5 | 0.11 |
| polarC18 | 276.144 | 98.5447 | 5.5 | 0.11 |
| polarC18 | 276.144 | 103.3063 | 5.5 | 0.11 |
| polarC18 | 276.144 | 155.0339 | 5.5 | 0.11 |
| polarC18 | 276.144 | 60.2149 | 5.5 | 0.11 |
| polarC18 | 276.144 | 53.0354 | 5.5 | 0.11 |
| polarC18 | 276.144 | 55.7421 | 5.5 | 0.11 |
| polarC18 | 276.144 | 50.2653 | 5.5 | 0.11 |
| polarC18 | 276.144 | 114.5742 | 5.5 | 0.11 |
| polarC18 | 276.144 | 120.6461 | 5.5 | 0.11 |
| polarC18 | 276.144 | 85.8919 | 5.5 | 0.11 |
| polarC18 | 276.144 | 247.6913 | 5.5 | 0.11 |
| polarC18 | 276.144 | 58.3842 | 5.5 | 0.11 |
| polarC18 | 276.144 | 86.7996 | 5.5 | 0.11 |
| polarC18 | 276.144 | 144.4816 | 5.5 | 0.1 |
| polarC18 | 276.144 | 101.7856 | 5.5 | 0.1 |
| polarC18 | 276.144 | 70.5965 | 5.5 | 0.1 |
| polarC18 | 276.144 | 52.9427 | 5.5 | 0.1 |
| polarC18 | 276.144 | 97.145 | 5.5 | 0.1 |
| polarC18 | 276.144 | 75.69 | 5.5 | 0.1 |
| polarC18 | 276.144 | 99.5605 | 5.5 | 0.1 |

| | | | | |
|---|---|---|---|---|
| polarC18 | 276.144 | 116.4801 | 5.5 | 0.1 |
| polarC18 | 276.144 | 91.3457 | 5.4 | 0.1 |
| polarC18 | 276.144 | 82.9437 | 5.4 | 0.1 |
| polarC18 | 276.144 | 216.7158 | 5.4 | 0.1 |
| polarC18 | 276.144 | 117.2989 | 5.4 | 0.1 |
| polarC18 | 276.144 | 57.8542 | 5.4 | 0.1 |
| polarC18 | 276.144 | 69.0558 | 5.4 | 0.1 |
| polarC18 | 276.144 | 141.7529 | 5.4 | 0.1 |
| polarC18 | 276.144 | 57.775 | 5.4 | 0.1 |
| polarC18 | 276.144 | 51.8665 | 5.4 | 0.1 |
| polarC18 | 276.144 | 87.7986 | 5.4 | 0.1 |
| polarC18 | 276.144 | 111.4261 | 5.4 | 0.1 |
| polarC18 | 276.144 | 105.4196 | 5.4 | 0.1 |
| polarC18 | 276.144 | 55.3933 | 5.4 | 0.1 |
| polarC18 | 276.144 | 233.2659 | 5.4 | 0.1 |
| polarC18 | 276.144 | 62.4724 | 5.4 | 0.1 |
| polarC18 | 276.144 | 77.5438 | 5.4 | 0.1 |
| polarC18 | 276.144 | 85.2074 | 5.4 | 0.1 |
| polarC18 | 276.144 | 154.3926 | 5.4 | 0.1 |
| polarC18 | 276.144 | 70.9405 | 5.4 | 0.1 |
| polarC18 | 276.144 | 51.7387 | 5.4 | 0.1 |
| polarC18 | 276.144 | 58.1404 | 5.4 | 0.1 |
| polarC18 | 276.144 | 143.625 | 5.4 | 0.1 |
| polarC18 | 276.144 | 88.5377 | 5.4 | 0.1 |
| polarC18 | 276.144 | 70.5644 | 5.4 | 0.1 |
| polarC18 | 276.144 | 105.3014 | 5.4 | 0.1 |
| polarC18 | 276.144 | 69.4611 | 5.4 | 0.1 |
| polarC18 | 276.144 | 150.8571 | 5.4 | 0.1 |
| polarC18 | 276.144 | 88.7536 | 5.4 | 0.1 |
| polarC18 | 276.144 | 61.4894 | 5.4 | 0.1 |
| polarC18 | 276.144 | 83.5018 | 5.4 | 0.1 |
| polarC18 | 276.144 | 96.6593 | 5.4 | 0.1 |
| polarC18 | 276.144 | 293.814 | 5.4 | 0.1 |
| polarC18 | 276.144 | 58.0168 | 5.4 | 0.1 |
| polarC18 | 276.144 | 91.3342 | 5.4 | 0.1 |
| polarC18 | 276.144 | 158.6797 | 5.4 | 0.1 |
| polarC18 | 276.144 | 96.9544 | 5.4 | 0.1 |
| polarC18 | 276.144 | 78.5789 | 5.4 | 0.1 |
| polarC18 | 276.144 | 55.7462 | 5.4 | 0.1 |
| polarC18 | 276.144 | 130.9311 | 5.4 | 0.1 |
| polarC18 | 276.144 | 77.4581 | 5.4 | 0.1 |
| polarC18 | 276.144 | 201.0925 | 5.4 | 0.1 |

| | | | | |
|---|---|---|---|---|
| polarC18 | 276.144 | 237.361 | 5.4 | 0.1 |
| polarC18 | 276.144 | 105.7737 | 5.4 | 0.1 |
| polarC18 | 276.144 | 60.755 | 5.4 | 0.1 |
| polarC18 | 276.144 | 102.5567 | 5.4 | 0.1 |
| polarC18 | 276.144 | 74.6331 | 5.4 | 0.1 |
| polarC18 | 276.144 | 56.3001 | 5.4 | 0.1 |
| polarC18 | 276.144 | 50.0864 | 5.4 | 0.1 |
| polarC18 | 276.144 | 86.1431 | 5.4 | 0.1 |
| polarC18 | 276.144 | 80.8592 | 5.4 | 0.1 |
| polarC18 | 276.144 | 69.8968 | 5.4 | 0.1 |
| polarC18 | 276.144 | 101.1105 | 5.4 | 0.1 |
| polarC18 | 276.144 | 70.477 | 5.4 | 0.1 |
| polarC18 | 276.144 | 64.666 | 5.4 | 0.1 |
| polarC18 | 276.144 | 69.3718 | 5.4 | 0.1 |
| polarC18 | 276.144 | 54.2157 | 5.4 | 0.1 |
| polarC18 | 276.144 | 229.6984 | 5.4 | 0.1 |
| polarC18 | 276.144 | 80.8001 | 5.4 | 0.1 |
| polarC18 | 276.144 | 105.1259 | 5.4 | 0.1 |
| polarC18 | 276.144 | 93.1388 | 5.4 | 0.1 |
| polarC18 | 276.144 | 175.175 | 5.4 | 0.1 |
| polarC18 | 276.144 | 53.4108 | 5.4 | 0.1 |
| polarC18 | 276.144 | 70.3912 | 5.4 | 0.1 |
| polarC18 | 276.144 | 131.9277 | 5.4 | 0.1 |
| polarC18 | 276.144 | 54.4624 | 5.4 | 0.1 |
| polarC18 | 276.144 | 196.5444 | 5.4 | 0.1 |
| polarC18 | 276.144 | 60.142 | 5.4 | 0.1 |
| polarC18 | 276.144 | 122.3634 | 5.4 | 0.1 |
| polarC18 | 276.144 | 103.0573 | 5.4 | 0.1 |
| polarC18 | 276.144 | 65.4447 | 5.4 | 0.1 |
| polarC18 | 276.144 | 174.7889 | 5.4 | 0.1 |
| polarC18 | 276.144 | 109.5342 | 5.4 | 0.1 |
| polarC18 | 276.144 | 96.5182 | 5.4 | 0.1 |
| polarC18 | 276.144 | 155.4662 | 5.4 | 0.1 |
| polarC18 | 276.144 | 165.9175 | 5.4 | 0.1 |
| polarC18 | 276.144 | 75.8756 | 5.4 | 0.1 |
| polarC18 | 276.144 | 71.1735 | 5.4 | 0.1 |
| polarC18 | 276.144 | 85.6129 | 5.4 | 0.1 |
| polarC18 | 276.144 | 125.8181 | 5.4 | 0.1 |
| polarC18 | 276.144 | 63.2779 | 5.4 | 0.1 |
| polarC18 | 276.144 | 92.5893 | 5.4 | 0.1 |
| polarC18 | 276.144 | 52.9716 | 5.3 | 0.1 |
| polarC18 | 276.144 | 85.8397 | 5.3 | 0.1 |

| | | | | |
|---|---|---|---|---|
| polarC18 | 276.144 | 52.7075 | 5.3 | 0.1 |
| polarC18 | 276.144 | 69.9239 | 5.3 | 0.1 |
| polarC18 | 276.144 | 171.7177 | 5.3 | 0.1 |
| polarC18 | 276.144 | 282.9834 | 5.3 | 0.1 |
| polarC18 | 276.144 | 52.1358 | 5.3 | 0.1 |
| polarC18 | 276.144 | 72.4338 | 5.3 | 0.1 |
| polarC18 | 276.144 | 65.3869 | 5.3 | 0.1 |
| polarC18 | 276.144 | 66.2242 | 5.3 | 0.1 |
| polarC18 | 276.144 | 113.5232 | 5.3 | 0.1 |
| polarC18 | 276.144 | 93.7132 | 5.3 | 0.1 |
| polarC18 | 276.144 | 261.9117 | 5.3 | 0.1 |
| polarC18 | 276.144 | 60.5418 | 5.3 | 0.1 |
| polarC18 | 276.144 | 67.7712 | 5.3 | 0.1 |
| polarC18 | 276.144 | 51.5362 | 5.3 | 0.1 |
| polarC18 | 276.144 | 52.5006 | 5.3 | 0.1 |
| polarC18 | 276.144 | 81.6569 | 5.3 | 0.1 |
| polarC18 | 276.144 | 61.1603 | 5.3 | 0.1 |
| polarC18 | 276.144 | 67.8791 | 5.3 | 0.1 |
| polarC18 | 276.144 | 79.5307 | 5.3 | 0.1 |
| polarC18 | 276.144 | 61.3385 | 5.3 | 0.1 |
| polarC18 | 276.144 | 195.7 | 5.3 | 0.1 |
| polarC18 | 276.144 | 114.2503 | 5.3 | 0.1 |
| polarC18 | 276.144 | 123.7031 | 5.3 | 0.1 |
| polarC18 | 276.144 | 185.054 | 5.3 | 0.1 |
| polarC18 | 276.144 | 54.8006 | 5.3 | 0.1 |
| polarC18 | 276.144 | 133.4582 | 5.3 | 0.1 |
| polarC18 | 276.144 | 64.2917 | 5.3 | 0.1 |
| polarC18 | 276.144 | 96.6105 | 5.3 | 0.1 |
| polarC18 | 276.144 | 66.581 | 5.3 | 0.1 |
| polarC18 | 276.144 | 135.0436 | 5.3 | 0.1 |
| polarC18 | 276.144 | 79.9403 | 5.3 | 0.1 |
| polarC18 | 276.144 | 197.0657 | 5.3 | 0.1 |
| polarC18 | 276.144 | 240.513 | 5.3 | 0.1 |
| polarC18 | 276.144 | 186.5384 | 5.3 | 0.1 |
| polarC18 | 276.144 | 169.6298 | 5.3 | 0.1 |
| polarC18 | 276.144 | 61.0438 | 5.3 | 0.1 |
| polarC18 | 276.144 | 146.0317 | 5.3 | 0.1 |
| polarC18 | 276.144 | 73.2241 | 5.3 | 0.1 |
| polarC18 | 276.144 | 111.5476 | 5.3 | 0.1 |
| polarC18 | 276.144 | 144.5797 | 5.3 | 0.1 |
| polarC18 | 276.144 | 235.9538 | 5.3 | 0.1 |
| polarC18 | 276.144 | 187.0095 | 5.3 | 0.1 |

| | | | | |
|---|---|---|---|---|
| polarC18 | 276.144 | 77.9658 | 5.3 | 0.1 |
| polarC18 | 276.144 | 62.211 | 5.3 | 0.1 |
| polarC18 | 276.144 | 87.0282 | 5.3 | 0.1 |
| polarC18 | 276.144 | 50.8874 | 5.3 | 0.1 |
| polarC18 | 276.144 | 71.5008 | 5.3 | 0.1 |
| polarC18 | 276.144 | 160.229 | 5.3 | 0.1 |
| polarC18 | 276.144 | 114.7012 | 5.3 | 0.1 |
| polarC18 | 276.144 | 51.9277 | 5.3 | 0.1 |
| polarC18 | 276.144 | 61.7724 | 5.3 | 0.1 |
| polarC18 | 276.144 | 65.5366 | 5.3 | 0.1 |
| polarC18 | 276.144 | 277.6539 | 5.3 | 0.1 |
| polarC18 | 276.144 | 50.6175 | 5.3 | 0.1 |
| polarC18 | 276.144 | 69.2013 | 5.3 | 0.1 |
| polarC18 | 276.144 | 165.3263 | 5.3 | 0.1 |
| polarC18 | 276.144 | 87.5362 | 5.3 | 0.1 |
| polarC18 | 276.144 | 55.6445 | 5.3 | 0.1 |
| polarC18 | 276.144 | 62.7983 | 5.3 | 0.1 |
| polarC18 | 276.144 | 74.9191 | 5.3 | 0.1 |
| polarC18 | 276.144 | 72.5451 | 5.3 | 0.1 |
| polarC18 | 276.144 | 91.1637 | 5.3 | 0.1 |
| polarC18 | 276.144 | 208.6809 | 5.3 | 0.1 |
| polarC18 | 276.144 | 51.8321 | 5.3 | 0.1 |
| polarC18 | 276.144 | 115.5765 | 5.3 | 0.1 |
| polarC18 | 276.144 | 101.7769 | 5.3 | 0.1 |
| polarC18 | 276.144 | 74.4548 | 5.3 | 0.1 |
| polarC18 | 276.144 | 52.8254 | 5.3 | 0.1 |
| polarC18 | 276.144 | 146.9963 | 5.3 | 0.1 |
| polarC18 | 276.144 | 90.7753 | 5.3 | 0.1 |
| polarC18 | 276.144 | 93.1663 | 5.3 | 0.1 |
| polarC18 | 276.144 | 213.6011 | 5.3 | 0.1 |
| polarC18 | 276.144 | 164.7164 | 5.3 | 0.1 |
| polarC18 | 276.144 | 264.1032 | 5.3 | 0.1 |
| polarC18 | 276.144 | 67.2073 | 5.3 | 0.1 |
| polarC18 | 276.144 | 58.5727 | 5.3 | 0.1 |
| polarC18 | 276.144 | 106.3165 | 5.3 | 0.1 |
| polarC18 | 276.144 | 124.8365 | 5.3 | 0.1 |
| polarC18 | 276.144 | 65.798 | 5.3 | 0.1 |
| polarC18 | 276.144 | 68.6306 | 5.3 | 0.1 |
| polarC18 | 276.144 | 64.6479 | 5.3 | 0.1 |
| polarC18 | 276.144 | 114.3896 | 5.3 | 0.1 |
| polarC18 | 276.144 | 198.1128 | 5.3 | 0.1 |
| polarC18 | 276.144 | 143.3284 | 5.3 | 0.1 |

| | | | | |
|---|---|---|---|---|
| polarC18 | 276.144 | 255.7491 | 5.2 | 0.1 |
| polarC18 | 276.144 | 100.2269 | 5.2 | 0.1 |
| polarC18 | 276.144 | 285.6659 | 5.2 | 0.1 |
| polarC18 | 276.144 | 51.8732 | 5.2 | 0.1 |
| polarC18 | 276.144 | 80.5633 | 5.2 | 0.1 |
| polarC18 | 276.144 | 50.3632 | 5.2 | 0.1 |
| polarC18 | 276.144 | 101.6473 | 5.2 | 0.1 |
| polarC18 | 276.144 | 102.6752 | 5.2 | 0.1 |
| polarC18 | 276.144 | 166.6021 | 5.2 | 0.1 |
| polarC18 | 276.144 | 52.2094 | 5.2 | 0.1 |
| polarC18 | 276.144 | 51.917 | 5.2 | 0.1 |
| polarC18 | 276.144 | 99.5678 | 5.2 | 0.1 |
| polarC18 | 276.144 | 251.8385 | 5.2 | 0.1 |
| polarC18 | 276.144 | 70.6273 | 5.2 | 0.1 |
| polarC18 | 276.144 | 173.1716 | 5.2 | 0.1 |
| polarC18 | 276.144 | 175.4681 | 5.2 | 0.1 |
| polarC18 | 276.144 | 63.8168 | 5.2 | 0.1 |
| polarC18 | 276.144 | 63.4768 | 5.2 | 0.1 |
| polarC18 | 276.144 | 116.6471 | 5.2 | 0.1 |
| polarC18 | 276.144 | 73.6652 | 5.2 | 0.1 |
| polarC18 | 276.144 | 82.6772 | 5.2 | 0.1 |
| polarC18 | 276.144 | 126.3101 | 5.2 | 0.1 |
| polarC18 | 276.144 | 70.6523 | 5.2 | 0.1 |
| polarC18 | 276.144 | 79.9609 | 5.2 | 0.1 |
| polarC18 | 276.144 | 70.7059 | 5.2 | 0.1 |
| polarC18 | 276.144 | 240.37 | 5.2 | 0.1 |
| polarC18 | 276.144 | 88.6167 | 5.2 | 0.1 |
| polarC18 | 276.144 | 61.8528 | 5.2 | 0.1 |
| polarC18 | 276.144 | 267.4858 | 5.2 | 0.1 |
| polarC18 | 276.144 | 61.4726 | 5.2 | 0.1 |
| polarC18 | 276.144 | 172.4247 | 5.2 | 0.1 |
| polarC18 | 276.144 | 104.3674 | 5.2 | 0.1 |
| polarC18 | 276.144 | 74.7797 | 5.2 | 0.1 |
| polarC18 | 276.144 | 151.6175 | 5.2 | 0.1 |
| polarC18 | 276.144 | 79.7302 | 5.2 | 0.1 |
| polarC18 | 276.144 | 76.9078 | 5.2 | 0.1 |
| polarC18 | 276.144 | 127.9008 | 5.2 | 0.1 |
| polarC18 | 276.144 | 109.4663 | 5.2 | 0.1 |
| polarC18 | 276.144 | 58.8604 | 5.2 | 0.1 |
| polarC18 | 276.144 | 223.2662 | 5.2 | 0.1 |
| polarC18 | 276.144 | 51.6459 | 5.2 | 0.1 |
| polarC18 | 276.144 | 72.047 | 5.2 | 0.1 |

| | | | | |
|---|---|---|---|---|
| polarC18 | 276.144 | 62.8833 | 5.2 | 0.1 |
| polarC18 | 276.144 | 119.4289 | 5.2 | 0.1 |
| polarC18 | 276.144 | 219.7495 | 5.2 | 0.1 |
| polarC18 | 276.144 | 74.129 | 5.2 | 0.1 |
| polarC18 | 276.144 | 62.9007 | 5.2 | 0.1 |
| polarC18 | 276.144 | 80.1585 | 5.2 | 0.1 |
| polarC18 | 276.144 | 60.7849 | 5.2 | 0.1 |
| polarC18 | 276.144 | 51.2321 | 5.2 | 0.1 |
| polarC18 | 276.144 | 195.3997 | 5.2 | 0.1 |
| polarC18 | 276.144 | 116.2038 | 5.2 | 0.1 |
| polarC18 | 276.144 | 58.5348 | 5.2 | 0.1 |
| polarC18 | 276.144 | 83.1512 | 5.2 | 0.1 |
| polarC18 | 276.144 | 99.9149 | 5.2 | 0.1 |
| polarC18 | 276.144 | 96.7981 | 5.2 | 0.1 |
| polarC18 | 276.144 | 81.5398 | 5.2 | 0.1 |
| polarC18 | 276.144 | 154.4751 | 5.2 | 0.1 |
| polarC18 | 276.144 | 84.6873 | 5.2 | 0.1 |
| polarC18 | 276.144 | 98.8557 | 5.2 | 0.1 |
| polarC18 | 276.144 | 58.2062 | 5.2 | 0.1 |
| polarC18 | 276.144 | 58.7158 | 5.2 | 0.1 |
| polarC18 | 276.144 | 95.3713 | 5.2 | 0.1 |
| polarC18 | 276.144 | 128.803 | 5.2 | 0.1 |
| polarC18 | 276.144 | 116.8551 | 5.2 | 0.1 |
| polarC18 | 276.144 | 61.8628 | 5.2 | 0.1 |
| polarC18 | 276.144 | 92.9696 | 5.2 | 0.1 |
| polarC18 | 276.144 | 79.4492 | 5.2 | 0.1 |
| polarC18 | 276.144 | 110.677 | 5.2 | 0.1 |
| polarC18 | 276.144 | 114.2716 | 5.2 | 0.1 |
| polarC18 | 276.144 | 166.939 | 5.2 | 0.1 |
| polarC18 | 276.144 | 85.0639 | 5.2 | 0.1 |
| polarC18 | 276.144 | 105.8073 | 5.2 | 0.1 |
| polarC18 | 276.144 | 64.8844 | 5.2 | 0.1 |
| polarC18 | 276.144 | 79.9446 | 5.2 | 0.1 |
| polarC18 | 276.144 | 79.87 | 5.2 | 0.1 |
| polarC18 | 276.144 | 95.0692 | 5.2 | 0.1 |
| polarC18 | 276.144 | 219.9946 | 5.2 | 0.1 |
| polarC18 | 276.144 | 56.0467 | 5.2 | 0.1 |
| polarC18 | 276.144 | 108.7457 | 5.1 | 0.1 |
| polarC18 | 276.144 | 270.9308 | 5.1 | 0.1 |
| polarC18 | 276.144 | 73.2022 | 5.1 | 0.1 |
| polarC18 | 276.144 | 141.3238 | 5.1 | 0.1 |
| polarC18 | 276.144 | 77.035 | 5.1 | 0.1 |

| | | | | |
|---|---|---|---|---|
| polarC18 | 276.144 | 171.442 | 5.1 | 0.1 |
| polarC18 | 276.144 | 104.3453 | 5.1 | 0.1 |
| polarC18 | 276.144 | 81.1376 | 5.1 | 0.1 |
| polarC18 | 276.144 | 112.3936 | 5.1 | 0.1 |
| polarC18 | 276.144 | 84.7785 | 5.1 | 0.1 |
| polarC18 | 276.144 | 55.3719 | 5.1 | 0.1 |
| polarC18 | 276.144 | 64.4519 | 5.1 | 0.1 |
| polarC18 | 276.144 | 63.6053 | 5.1 | 0.1 |
| polarC18 | 276.144 | 93.6615 | 5.1 | 0.1 |
| polarC18 | 276.144 | 53.4887 | 5.1 | 0.1 |
| polarC18 | 276.144 | 86.9327 | 5.1 | 0.1 |
| polarC18 | 276.144 | 292.1646 | 5.1 | 0.1 |
| polarC18 | 276.144 | 68.0499 | 5.1 | 0.1 |
| polarC18 | 276.144 | 167.0111 | 5.1 | 0.1 |
| polarC18 | 276.144 | 84.6686 | 5.1 | 0.1 |
| polarC18 | 276.144 | 138.8149 | 5.1 | 0.1 |
| polarC18 | 276.144 | 68.8192 | 5.1 | 0.1 |
| polarC18 | 276.144 | 147.7462 | 5.1 | 0.1 |
| polarC18 | 276.144 | 68.529 | 5.1 | 0.1 |
| polarC18 | 276.144 | 96.7655 | 5.1 | 0.1 |
| polarC18 | 276.144 | 62.7097 | 5.1 | 0.1 |
| polarC18 | 276.144 | 222.349 | 5.1 | 0.1 |
| polarC18 | 276.144 | 121.2324 | 5.1 | 0.1 |
| polarC18 | 276.144 | 112.603 | 5.1 | 0.1 |
| polarC18 | 276.144 | 56.3446 | 5.1 | 0.1 |
| polarC18 | 276.144 | 190.9364 | 5.1 | 0.1 |
| polarC18 | 276.144 | 50.3725 | 5.1 | 0.1 |
| polarC18 | 276.144 | 58.5499 | 5.1 | 0.1 |
| polarC18 | 276.144 | 56.2623 | 5.1 | 0.1 |
| polarC18 | 276.144 | 167.1931 | 5.1 | 0.1 |
| polarC18 | 276.144 | 63.2803 | 5.1 | 0.1 |
| polarC18 | 276.144 | 56.1283 | 5.1 | 0.1 |
| polarC18 | 276.144 | 74.8233 | 5.1 | 0.1 |
| polarC18 | 276.144 | 151.6987 | 5.1 | 0.1 |
| polarC18 | 276.144 | 86.2838 | 5.1 | 0.1 |
| polarC18 | 276.144 | 108.6069 | 5.1 | 0.1 |
| polarC18 | 276.144 | 132.5979 | 5.1 | 0.1 |
| polarC18 | 276.144 | 119.1931 | 5.1 | 0.1 |
| polarC18 | 276.144 | 54.1438 | 5.1 | 0.1 |
| polarC18 | 276.144 | 76.8795 | 5.1 | 0.1 |
| polarC18 | 276.144 | 179.6059 | 5.1 | 0.1 |
| polarC18 | 276.144 | 79.6434 | 5.1 | 0.1 |

| | | | | |
|---|---|---|---|---|
| polarC18 | 276.144 | 245.2631 | 5.1 | 0.1 |
| polarC18 | 276.144 | 207.7393 | 5.1 | 0.1 |
| polarC18 | 276.144 | 58.0295 | 5.1 | 0.1 |
| polarC18 | 276.144 | 56.9177 | 5.1 | 0.1 |
| polarC18 | 276.144 | 141.0119 | 5.1 | 0.1 |
| polarC18 | 276.144 | 66.2785 | 5.1 | 0.1 |
| polarC18 | 276.144 | 198.7671 | 5.1 | 0.1 |
| polarC18 | 276.144 | 74.851 | 5.1 | 0.1 |
| polarC18 | 276.144 | 75.0672 | 5.1 | 0.1 |
| polarC18 | 276.144 | 72.5207 | 5.1 | 0.1 |
| polarC18 | 276.144 | 78.4595 | 5.1 | 0.1 |
| polarC18 | 276.144 | 52.6515 | 5.1 | 0.1 |
| polarC18 | 276.144 | 121.428 | 5.1 | 0.1 |
| polarC18 | 276.144 | 59.4408 | 5.1 | 0.1 |
| polarC18 | 276.144 | 58.7278 | 5.1 | 0.1 |
| polarC18 | 276.144 | 150.6877 | 5.1 | 0.1 |
| polarC18 | 276.144 | 101.8153 | 5.1 | 0.1 |
| polarC18 | 276.144 | 137.4852 | 5.1 | 0.1 |
| polarC18 | 276.144 | 119.5985 | 5.1 | 0.1 |
| polarC18 | 276.144 | 77.677 | 5.1 | 0.1 |
| polarC18 | 276.144 | 133.0387 | 5.1 | 0.1 |
| polarC18 | 276.144 | 75.0962 | 5.1 | 0.1 |
| polarC18 | 276.144 | 132.7798 | 5.1 | 0.1 |
| polarC18 | 276.144 | 64.4624 | 5.1 | 0.1 |
| polarC18 | 276.144 | 71.4646 | 5.1 | 0.1 |
| polarC18 | 276.144 | 190.2061 | 5.1 | 0.1 |
| polarC18 | 276.144 | 81.5656 | 5 | 0.1 |
| polarC18 | 276.144 | 120.1134 | 5 | 0.1 |
| polarC18 | 276.144 | 82.0159 | 5 | 0.1 |
| polarC18 | 276.144 | 132.5611 | 5 | 0.1 |
| polarC18 | 276.144 | 74.162 | 5 | 0.1 |
| polarC18 | 276.144 | 91.1806 | 5 | 0.1 |
| polarC18 | 276.144 | 159.9054 | 5 | 0.1 |
| polarC18 | 276.144 | 79.1151 | 5 | 0.1 |
| polarC18 | 276.144 | 93.0376 | 5 | 0.1 |
| polarC18 | 276.144 | 68.4112 | 5 | 0.1 |
| polarC18 | 276.144 | 54.7778 | 5 | 0.1 |
| polarC18 | 276.144 | 52.0212 | 5 | 0.1 |
| polarC18 | 276.144 | 127.0844 | 5 | 0.1 |
| polarC18 | 276.144 | 92.9322 | 5 | 0.1 |
| polarC18 | 276.144 | 210.2568 | 5 | 0.1 |
| polarC18 | 276.144 | 118.8377 | 5 | 0.1 |

| | | | | |
|---|---|---|---|---|
| polarC18 | 276.144 | 58.9957 | 5 | 0.1 |
| polarC18 | 276.144 | 239.9993 | 5 | 0.1 |
| polarC18 | 276.144 | 87.0565 | 5 | 0.1 |
| polarC18 | 276.144 | 74.9666 | 5 | 0.1 |
| polarC18 | 276.144 | 82.0248 | 5 | 0.1 |
| polarC18 | 276.144 | 81.3979 | 5 | 0.1 |
| polarC18 | 276.144 | 212.6488 | 5 | 0.1 |
| polarC18 | 276.144 | 105.3353 | 5 | 0.1 |
| polarC18 | 276.144 | 63.272 | 5 | 0.1 |
| polarC18 | 276.144 | 166.0967 | 5 | 0.1 |
| polarC18 | 276.144 | 60.5387 | 5 | 0.1 |
| polarC18 | 276.144 | 104.9309 | 5 | 0.1 |
| polarC18 | 276.144 | 63.8865 | 5 | 0.1 |
| polarC18 | 276.144 | 230.0456 | 5 | 0.1 |
| polarC18 | 276.144 | 79.955 | 5 | 0.1 |
| polarC18 | 276.144 | 61.5191 | 5 | 0.1 |
| polarC18 | 276.144 | 167.4815 | 5 | 0.1 |
| polarC18 | 276.144 | 269.7465 | 5 | 0.1 |
| polarC18 | 276.144 | 210.2237 | 5 | 0.1 |
| polarC18 | 276.144 | 85.6033 | 5 | 0.1 |
| polarC18 | 276.144 | 199.8773 | 5 | 0.1 |
| polarC18 | 276.144 | 64.7954 | 5 | 0.1 |
| polarC18 | 276.144 | 88.4259 | 5 | 0.1 |
| polarC18 | 276.144 | 80.9344 | 5 | 0.1 |
| polarC18 | 276.144 | 92.7892 | 5 | 0.1 |
| polarC18 | 276.144 | 71.5619 | 5 | 0.1 |
| polarC18 | 276.144 | 127.5916 | 5 | 0.1 |
| polarC18 | 276.144 | 53.7499 | 5 | 0.1 |
| polarC18 | 276.144 | 185.4324 | 5 | 0.1 |
| polarC18 | 276.144 | 104.0259 | 5 | 0.1 |
| polarC18 | 276.144 | 142.7751 | 5 | 0.1 |
| polarC18 | 276.144 | 84.7045 | 5 | 0.1 |
| polarC18 | 276.144 | 59.7421 | 5 | 0.1 |
| polarC18 | 276.144 | 245.3043 | 5 | 0.1 |
| polarC18 | 276.144 | 67.4102 | 5 | 0.1 |
| polarC18 | 276.144 | 173.3282 | 5 | 0.1 |
| polarC18 | 276.144 | 57.287 | 5 | 0.1 |
| polarC18 | 276.144 | 163.854 | 4.9 | 0.1 |
| polarC18 | 276.144 | 110.4031 | 4.9 | 0.1 |
| polarC18 | 276.144 | 134.0052 | 4.9 | 0.1 |
| polarC18 | 276.144 | 225.9909 | 4.9 | 0.1 |
| polarC18 | 276.144 | 62.2961 | 4.9 | 0.1 |

| | | | | |
|---|---|---|---|---|
| polarC18 | 276.144 | 69.0034 | 4.9 | 0.1 |
| polarC18 | 276.144 | 62.5102 | 4.9 | 0.1 |
| polarC18 | 276.144 | 73.1534 | 4.9 | 0.09 |
| polarC18 | 276.144 | 155.9836 | 4.9 | 0.09 |
| polarC18 | 276.144 | 91.1205 | 4.9 | 0.09 |
| polarC18 | 276.144 | 222.09 | 4.9 | 0.09 |
| polarC18 | 276.144 | 277.3452 | 4.9 | 0.09 |
| polarC18 | 276.144 | 94.1508 | 4.9 | 0.09 |
| polarC18 | 276.144 | 265.1897 | 4.9 | 0.09 |
| polarC18 | 276.144 | 56.0983 | 4.9 | 0.09 |
| polarC18 | 276.144 | 100.3223 | 4.9 | 0.09 |
| polarC18 | 276.144 | 104.0797 | 4.9 | 0.09 |
| polarC18 | 276.144 | 147.5662 | 4.9 | 0.09 |
| polarC18 | 276.144 | 56.0171 | 4.9 | 0.09 |
| polarC18 | 276.144 | 133.0035 | 4.9 | 0.09 |
| polarC18 | 276.144 | 50.1547 | 4.9 | 0.09 |
| polarC18 | 276.144 | 51.3323 | 4.9 | 0.09 |
| polarC18 | 276.144 | 95.0099 | 4.9 | 0.09 |
| polarC18 | 276.144 | 88.2215 | 4.9 | 0.09 |
| polarC18 | 276.144 | 92.0672 | 4.9 | 0.09 |
| polarC18 | 276.144 | 73.9195 | 4.9 | 0.09 |
| polarC18 | 276.144 | 54.7409 | 4.9 | 0.09 |
| polarC18 | 276.144 | 64.7729 | 4.9 | 0.09 |
| polarC18 | 276.144 | 252.1643 | 4.9 | 0.09 |
| polarC18 | 276.144 | 59.6412 | 4.9 | 0.09 |
| polarC18 | 276.144 | 96.7818 | 4.9 | 0.09 |
| polarC18 | 276.144 | 58.8827 | 4.9 | 0.09 |
| polarC18 | 276.144 | 105.7985 | 4.9 | 0.09 |
| polarC18 | 276.144 | 229.4233 | 4.9 | 0.09 |
| polarC18 | 276.144 | 55.7897 | 4.9 | 0.09 |
| polarC18 | 276.144 | 220.7408 | 4.9 | 0.09 |
| polarC18 | 276.144 | 127.1186 | 4.9 | 0.09 |
| polarC18 | 276.144 | 105.9764 | 4.9 | 0.09 |
| polarC18 | 276.144 | 123.3244 | 4.9 | 0.09 |
| polarC18 | 276.144 | 74.6296 | 4.9 | 0.09 |
| polarC18 | 276.144 | 50.6689 | 4.9 | 0.09 |
| polarC18 | 276.144 | 62.7285 | 4.9 | 0.09 |
| polarC18 | 276.144 | 50.8774 | 4.9 | 0.09 |
| polarC18 | 276.144 | 51.1856 | 4.9 | 0.09 |
| polarC18 | 276.144 | 54.4522 | 4.9 | 0.09 |
| polarC18 | 276.144 | 59.7336 | 4.9 | 0.09 |
| polarC18 | 276.144 | 107.3532 | 4.9 | 0.09 |

| | | | | |
|---|---|---|---|---|
| polarC18 | 276.144 | 65.3843 | 4.9 | 0.09 |
| polarC18 | 276.144 | 104.6581 | 4.9 | 0.09 |
| polarC18 | 276.144 | 71.1778 | 4.9 | 0.09 |
| polarC18 | 276.144 | 78.8205 | 4.9 | 0.09 |
| polarC18 | 276.144 | 78.4797 | 4.9 | 0.09 |
| polarC18 | 276.144 | 78.3988 | 4.9 | 0.09 |
| polarC18 | 276.144 | 87.7931 | 4.9 | 0.09 |
| polarC18 | 276.144 | 87.653 | 4.9 | 0.09 |
| polarC18 | 276.144 | 129.3167 | 4.9 | 0.09 |
| polarC18 | 276.144 | 124.3849 | 4.9 | 0.09 |
| polarC18 | 276.144 | 64.3822 | 4.9 | 0.09 |
| polarC18 | 276.144 | 60.1553 | 4.9 | 0.09 |
| polarC18 | 276.144 | 74.1712 | 4.9 | 0.09 |
| polarC18 | 276.144 | 213.8253 | 4.9 | 0.09 |
| polarC18 | 276.144 | 106.0601 | 4.9 | 0.09 |
| polarC18 | 276.144 | 116.433 | 4.9 | 0.09 |
| polarC18 | 276.144 | 276.6855 | 4.9 | 0.09 |
| polarC18 | 276.144 | 79.264 | 4.9 | 0.09 |
| polarC18 | 276.144 | 56.8013 | 4.8 | 0.09 |
| polarC18 | 276.144 | 164.6529 | 4.8 | 0.09 |
| polarC18 | 276.144 | 68.0379 | 4.8 | 0.09 |
| polarC18 | 276.144 | 70.0859 | 4.8 | 0.09 |
| polarC18 | 276.144 | 83.8295 | 4.8 | 0.09 |
| polarC18 | 276.144 | 55.9693 | 4.8 | 0.09 |
| polarC18 | 276.144 | 61.8575 | 4.8 | 0.09 |
| polarC18 | 276.144 | 240.9574 | 4.8 | 0.09 |
| polarC18 | 276.144 | 156.7582 | 4.8 | 0.09 |
| polarC18 | 276.144 | 242.5957 | 4.8 | 0.09 |
| polarC18 | 276.144 | 80.7225 | 4.8 | 0.09 |
| polarC18 | 276.144 | 117.8163 | 4.8 | 0.09 |
| polarC18 | 276.144 | 159.4182 | 4.8 | 0.09 |
| polarC18 | 276.144 | 106.4533 | 4.8 | 0.09 |
| polarC18 | 276.144 | 227.9946 | 4.8 | 0.09 |
| polarC18 | 276.144 | 232.5748 | 4.8 | 0.09 |
| polarC18 | 276.144 | 68.5598 | 4.8 | 0.09 |
| polarC18 | 276.144 | 147.521 | 4.8 | 0.09 |
| polarC18 | 276.144 | 80.7626 | 4.8 | 0.09 |
| polarC18 | 276.144 | 75.4536 | 4.8 | 0.09 |
| polarC18 | 276.144 | 103.6376 | 4.8 | 0.09 |
| polarC18 | 276.144 | 165.1505 | 4.8 | 0.09 |
| polarC18 | 276.144 | 54.4689 | 4.8 | 0.09 |
| polarC18 | 276.144 | 60.1666 | 4.8 | 0.09 |

| | | | | |
|---|---|---|---|---|
| polarC18 | 276.144 | 76.7107 | 4.8 | 0.09 |
| polarC18 | 276.144 | 90.767 | 4.8 | 0.09 |
| polarC18 | 276.144 | 73.4328 | 4.8 | 0.09 |
| polarC18 | 276.144 | 81.5738 | 4.8 | 0.09 |
| polarC18 | 276.144 | 60.4844 | 4.8 | 0.09 |
| polarC18 | 276.144 | 59.8296 | 4.8 | 0.09 |
| polarC18 | 276.144 | 70.2835 | 4.8 | 0.09 |
| polarC18 | 276.144 | 53.6493 | 4.8 | 0.09 |
| polarC18 | 276.144 | 75.247 | 4.8 | 0.09 |
| polarC18 | 276.144 | 51.004 | 4.8 | 0.09 |
| polarC18 | 276.144 | 82.8923 | 4.8 | 0.09 |
| polarC18 | 276.144 | 55.9799 | 4.8 | 0.09 |
| polarC18 | 276.144 | 79.1025 | 4.8 | 0.09 |
| polarC18 | 276.144 | 73.3565 | 4.7 | 0.09 |
| polarC18 | 276.144 | 50.7426 | 4.7 | 0.09 |
| polarC18 | 276.144 | 56.751 | 4.7 | 0.09 |
| polarC18 | 276.144 | 52.9367 | 4.7 | 0.09 |
| polarC18 | 276.144 | 59.8862 | 4.7 | 0.09 |
| polarC18 | 276.144 | 64.676 | 4.7 | 0.09 |
| polarC18 | 276.144 | 58.8848 | 4.7 | 0.09 |
| polarC18 | 276.144 | 71.1849 | 4.7 | 0.09 |
| polarC18 | 276.144 | 84.5035 | 4.7 | 0.09 |
| polarC18 | 276.144 | 108.8588 | 4.7 | 0.09 |
| polarC18 | 276.144 | 53.0009 | 4.7 | 0.09 |
| polarC18 | 276.144 | 232.5107 | 4.7 | 0.09 |
| polarC18 | 276.144 | 112.9333 | 4.7 | 0.09 |
| polarC18 | 276.144 | 67.5935 | 4.7 | 0.09 |
| polarC18 | 276.144 | 89.6116 | 4.7 | 0.09 |
| polarC18 | 276.144 | 69.1964 | 4.7 | 0.09 |
| polarC18 | 276.144 | 148.5803 | 4.7 | 0.09 |
| polarC18 | 276.144 | 285.9858 | 4.7 | 0.09 |
| polarC18 | 276.144 | 68.9718 | 4.7 | 0.09 |
| polarC18 | 276.144 | 161.9352 | 4.7 | 0.09 |
| polarC18 | 276.144 | 84.9288 | 4.7 | 0.09 |
| polarC18 | 276.144 | 55.1288 | 4.7 | 0.09 |
| polarC18 | 276.144 | 90.2081 | 4.6 | 0.09 |
| polarC18 | 276.144 | 117.6273 | 4.6 | 0.09 |
| polarC18 | 276.144 | 64.1614 | 4.6 | 0.09 |
| polarC18 | 276.144 | 130.325 | 4.6 | 0.09 |
| polarC18 | 276.144 | 117.7163 | 4.6 | 0.09 |
| polarC18 | 276.144 | 115.326 | 4.6 | 0.09 |
| polarC18 | 276.144 | 78.3527 | 4.6 | 0.09 |

| | | | | |
|---|---|---|---|---|
| polarC18 | 276.144 | 137.5819 | 4.6 | 0.09 |
| polarC18 | 276.144 | 51.4716 | 4.6 | 0.09 |
| polarC18 | 276.144 | 79.0301 | 4.6 | 0.09 |
| polarC18 | 276.144 | 77.8135 | 4.6 | 0.09 |
| polarC18 | 276.144 | 77.9176 | 4.6 | 0.09 |
| polarC18 | 276.144 | 118.1139 | 4.6 | 0.09 |
| polarC18 | 276.144 | 52.8166 | 4.6 | 0.09 |
| polarC18 | 276.144 | 50.5963 | 4.6 | 0.09 |
| polarC18 | 276.144 | 56.2833 | 4.6 | 0.09 |
| polarC18 | 276.144 | 95.8668 | 4.6 | 0.09 |
| polarC18 | 276.144 | 104.3115 | 4.5 | 0.09 |
| polarC18 | 276.144 | 72.409 | 4.5 | 0.09 |
| polarC18 | 276.144 | 60.699 | 4.5 | 0.09 |
| polarC18 | 276.144 | 68.4717 | 4.5 | 0.09 |
| polarC18 | 276.144 | 52.7007 | 4.5 | 0.09 |
| polarC18 | 276.144 | 109.0406 | 4.4 | 0.09 |
| polarC18 | 276.144 | 79.4875 | 4.3 | 0.08 |
| polarC18 | 276.144 | 106.786 | 4.3 | 0.08 |
| polarC18 | 276.144 | 80.2857 | 3.9 | 0.07 |
| polarC18 | 325.216 | 80.9478 | 19456.9 | 100 |
| polarC18 | 325.216 | 325.2195 | 14264.8 | 73.31 |
| polarC18 | 325.216 | 57.0698 | 3499.5 | 17.99 |
| polarC18 | 325.216 | 71.0855 | 2070.9 | 10.64 |
| polarC18 | 325.216 | 195.0837 | 1292.9 | 6.65 |
| polarC18 | 325.216 | 228.2204 | 994.1 | 5.11 |
| polarC18 | 325.216 | 139.0213 | 592.9 | 3.05 |
| polarC18 | 325.216 | 325.2717 | 495.2 | 2.55 |
| polarC18 | 325.216 | 213.0948 | 209.4 | 1.08 |
| polarC18 | 325.216 | 209.1177 | 179.5 | 0.92 |
| polarC18 | 325.216 | 308.2676 | 147.1 | 0.76 |
| polarC18 | 325.216 | 71.1225 | 87.8 | 0.45 |
| polarC18 | 325.216 | 166.1354 | 87.2 | 0.45 |
| polarC18 | 325.216 | 118.3219 | 86.6 | 0.45 |
| polarC18 | 325.216 | 82.9451 | 81.1 | 0.42 |
| polarC18 | 325.216 | 250.5333 | 73.8 | 0.38 |
| polarC18 | 325.216 | 93.4323 | 72 | 0.37 |
| polarC18 | 325.216 | 138.0216 | 69.1 | 0.36 |
| polarC18 | 325.216 | 94.1089 | 68.9 | 0.35 |
| polarC18 | 325.216 | 299.2988 | 68.4 | 0.35 |
| polarC18 | 325.216 | 224.0337 | 68.2 | 0.35 |
| polarC18 | 325.216 | 197.6492 | 67.2 | 0.35 |
| polarC18 | 325.216 | 218.5794 | 66.9 | 0.34 |

| | | | | |
|---|---|---|---|---|
| polarC18 | 325.216 | 113.219 | 66.6 | 0.34 |
| polarC18 | 325.216 | 349.1524 | 64.7 | 0.33 |
| polarC18 | 325.216 | 211.8781 | 64.4 | 0.33 |
| polarC18 | 325.216 | 79.6848 | 64 | 0.33 |
| polarC18 | 325.216 | 128.9526 | 56.8 | 0.29 |
| polarC18 | 325.216 | 243.2094 | 52.8 | 0.27 |
| polarC18 | 325.216 | 93.9298 | 52.3 | 0.27 |
| polarC18 | 325.216 | 139.5274 | 52.2 | 0.27 |
| polarC18 | 325.216 | 198.9735 | 51.3 | 0.26 |
| polarC18 | 325.216 | 92.3316 | 51 | 0.26 |
| polarC18 | 325.216 | 213.9813 | 50.9 | 0.26 |
| polarC18 | 325.216 | 52.1876 | 50.8 | 0.26 |
| polarC18 | 325.216 | 103.4469 | 50.7 | 0.26 |
| polarC18 | 325.216 | 93.4737 | 50.6 | 0.26 |
| polarC18 | 325.216 | 303.3717 | 50.6 | 0.26 |
| polarC18 | 325.216 | 67.5511 | 50.5 | 0.26 |
| polarC18 | 325.216 | 190.0147 | 50.3 | 0.26 |
| polarC18 | 325.216 | 94.0391 | 50 | 0.26 |
| polarC18 | 325.216 | 327.8323 | 49.9 | 0.26 |
| polarC18 | 325.216 | 198.8975 | 49.9 | 0.26 |
| polarC18 | 325.216 | 281.782 | 49.8 | 0.26 |
| polarC18 | 325.216 | 83.7605 | 49.6 | 0.26 |
| polarC18 | 325.216 | 264.8655 | 49.5 | 0.25 |
| polarC18 | 325.216 | 163.3196 | 49.4 | 0.25 |
| polarC18 | 325.216 | 154.7678 | 49.4 | 0.25 |
| polarC18 | 325.216 | 130.8461 | 49.2 | 0.25 |
| polarC18 | 325.216 | 288.7065 | 49.1 | 0.25 |
| polarC18 | 325.216 | 163.0204 | 49 | 0.25 |
| polarC18 | 325.216 | 225.3253 | 48.9 | 0.25 |
| polarC18 | 325.216 | 231.44 | 48.9 | 0.25 |
| polarC18 | 325.216 | 105.7572 | 48.9 | 0.25 |
| polarC18 | 325.216 | 148.7106 | 48.9 | 0.25 |
| polarC18 | 325.216 | 173.8837 | 48.8 | 0.25 |
| polarC18 | 325.216 | 121.4732 | 48.7 | 0.25 |
| polarC18 | 325.216 | 105.2687 | 48.6 | 0.25 |
| polarC18 | 325.216 | 217.0365 | 48.6 | 0.25 |
| polarC18 | 325.216 | 242.8952 | 48.4 | 0.25 |
| polarC18 | 325.216 | 93.8603 | 48.3 | 0.25 |
| polarC18 | 325.216 | 81.2112 | 48.2 | 0.25 |
| polarC18 | 325.216 | 67.9644 | 48.2 | 0.25 |
| polarC18 | 325.216 | 66.2084 | 48 | 0.25 |
| polarC18 | 325.216 | 145.3344 | 47.9 | 0.25 |

| | | | | |
|---|---|---|---|---|
| polarC18 | 325.216 | 94.1265 | 47.9 | 0.25 |
| polarC18 | 325.216 | 93.9987 | 47.9 | 0.25 |
| polarC18 | 325.216 | 129.4359 | 47.8 | 0.25 |
| polarC18 | 325.216 | 54.1422 | 47.8 | 0.25 |
| polarC18 | 325.216 | 59.0037 | 47.8 | 0.25 |
| polarC18 | 325.216 | 89.7539 | 47.7 | 0.24 |
| polarC18 | 325.216 | 109.0754 | 47.6 | 0.24 |
| polarC18 | 325.216 | 281.3249 | 47.6 | 0.24 |
| polarC18 | 325.216 | 311.1228 | 47.5 | 0.24 |
| polarC18 | 325.216 | 202.2255 | 47.4 | 0.24 |
| polarC18 | 325.216 | 93.4256 | 47.4 | 0.24 |
| polarC18 | 325.216 | 69.0703 | 47.4 | 0.24 |
| polarC18 | 325.216 | 135.3003 | 47.4 | 0.24 |
| polarC18 | 325.216 | 108.8724 | 47.3 | 0.24 |
| polarC18 | 325.216 | 51.4916 | 47.1 | 0.24 |
| polarC18 | 325.216 | 115.7408 | 47.1 | 0.24 |
| polarC18 | 325.216 | 222.3437 | 47 | 0.24 |
| polarC18 | 325.216 | 83.5368 | 47 | 0.24 |
| polarC18 | 325.216 | 142.636 | 46.9 | 0.24 |
| polarC18 | 325.216 | 126.1794 | 46.9 | 0.24 |
| polarC18 | 325.216 | 52.4994 | 46.9 | 0.24 |
| polarC18 | 325.216 | 126.4694 | 46.8 | 0.24 |
| polarC18 | 325.216 | 107.9601 | 46.8 | 0.24 |
| polarC18 | 325.216 | 54.0051 | 46.8 | 0.24 |
| polarC18 | 325.216 | 130.8753 | 46.8 | 0.24 |
| polarC18 | 325.216 | 163.7151 | 46.7 | 0.24 |
| polarC18 | 325.216 | 266.6999 | 46.7 | 0.24 |
| polarC18 | 325.216 | 124.5006 | 46.6 | 0.24 |
| polarC18 | 325.216 | 325.3505 | 46.6 | 0.24 |
| polarC18 | 325.216 | 275.2853 | 46.6 | 0.24 |
| polarC18 | 325.216 | 50.435 | 46.5 | 0.24 |
| polarC18 | 325.216 | 52.7421 | 46.5 | 0.24 |
| polarC18 | 325.216 | 351.1249 | 46.5 | 0.24 |
| polarC18 | 325.216 | 111.1968 | 46.4 | 0.24 |
| polarC18 | 325.216 | 88.06 | 46.4 | 0.24 |
| polarC18 | 325.216 | 76.3178 | 46.4 | 0.24 |
| polarC18 | 325.216 | 148.3365 | 46.4 | 0.24 |
| polarC18 | 325.216 | 91.43 | 46.4 | 0.24 |
| polarC18 | 325.216 | 50.6573 | 46.4 | 0.24 |
| polarC18 | 325.216 | 283.6442 | 46.3 | 0.24 |
| polarC18 | 325.216 | 54.1644 | 46.3 | 0.24 |
| polarC18 | 325.216 | 345.7838 | 46.3 | 0.24 |

| | | | | |
|---|---|---|---|---|
| polarC18 | 325.216 | 278.6029 | 46.3 | 0.24 |
| polarC18 | 325.216 | 237.8338 | 46.2 | 0.24 |
| polarC18 | 325.216 | 87.4393 | 46.2 | 0.24 |
| polarC18 | 325.216 | 227.433 | 46.2 | 0.24 |
| polarC18 | 325.216 | 252.0098 | 46.1 | 0.24 |
| polarC18 | 325.216 | 73.0326 | 46 | 0.24 |
| polarC18 | 325.216 | 294.1013 | 46 | 0.24 |
| polarC18 | 325.216 | 149.3672 | 46 | 0.24 |
| polarC18 | 325.216 | 112.6755 | 45.9 | 0.24 |
| polarC18 | 325.216 | 293.0042 | 45.9 | 0.24 |
| polarC18 | 325.216 | 219.9953 | 45.9 | 0.24 |
| polarC18 | 325.216 | 103.0945 | 45.8 | 0.24 |
| polarC18 | 325.216 | 286.6392 | 45.8 | 0.24 |
| polarC18 | 325.216 | 51.5474 | 45.8 | 0.24 |
| polarC18 | 325.216 | 51.5689 | 45.8 | 0.24 |
| polarC18 | 325.216 | 145.8171 | 45.6 | 0.23 |
| polarC18 | 325.216 | 56.9894 | 45.6 | 0.23 |
| polarC18 | 325.216 | 133.4557 | 45.5 | 0.23 |
| polarC18 | 325.216 | 84.1915 | 45.5 | 0.23 |
| polarC18 | 325.216 | 108.9723 | 45.5 | 0.23 |
| polarC18 | 325.216 | 51.2724 | 45.5 | 0.23 |
| polarC18 | 325.216 | 194.291 | 45.5 | 0.23 |
| polarC18 | 325.216 | 70.3398 | 45.5 | 0.23 |
| polarC18 | 325.216 | 119.5072 | 45.4 | 0.23 |
| polarC18 | 325.216 | 105.8886 | 45.4 | 0.23 |
| polarC18 | 325.216 | 151.9252 | 45.3 | 0.23 |
| polarC18 | 325.216 | 233.398 | 45.3 | 0.23 |
| polarC18 | 325.216 | 105.5936 | 45.2 | 0.23 |
| polarC18 | 325.216 | 169.7288 | 45.2 | 0.23 |
| polarC18 | 325.216 | 89.179 | 45.2 | 0.23 |
| polarC18 | 325.216 | 242.0437 | 45.2 | 0.23 |
| polarC18 | 325.216 | 74.3073 | 45.2 | 0.23 |
| polarC18 | 325.216 | 65.8863 | 45.2 | 0.23 |
| polarC18 | 325.216 | 99.0673 | 45.1 | 0.23 |
| polarC18 | 325.216 | 100.7814 | 45.1 | 0.23 |
| polarC18 | 325.216 | 151.9868 | 45.1 | 0.23 |
| polarC18 | 325.216 | 248.7322 | 45.1 | 0.23 |
| polarC18 | 325.216 | 115.8124 | 45 | 0.23 |
| polarC18 | 325.216 | 74.6951 | 44.9 | 0.23 |
| polarC18 | 325.216 | 66.7287 | 44.9 | 0.23 |
| polarC18 | 325.216 | 53.3981 | 44.8 | 0.23 |
| polarC18 | 325.216 | 54.3738 | 44.8 | 0.23 |

| | | | | |
|---|---|---|---|---|
| polarC18 | 325.216 | 252.2808 | 44.7 | 0.23 |
| polarC18 | 325.216 | 134.2816 | 44.7 | 0.23 |
| polarC18 | 325.216 | 108.3221 | 44.6 | 0.23 |
| polarC18 | 325.216 | 115.5122 | 44.6 | 0.23 |
| polarC18 | 325.216 | 91.3599 | 44.5 | 0.23 |
| polarC18 | 325.216 | 201.4525 | 44.5 | 0.23 |
| polarC18 | 325.216 | 187.8676 | 44.4 | 0.23 |
| polarC18 | 325.216 | 130.0735 | 44.4 | 0.23 |
| polarC18 | 325.216 | 315.2585 | 44.3 | 0.23 |
| polarC18 | 325.216 | 60.8521 | 44.3 | 0.23 |
| polarC18 | 325.216 | 117.1116 | 44.3 | 0.23 |
| polarC18 | 325.216 | 51.5708 | 44.3 | 0.23 |
| polarC18 | 325.216 | 61.8765 | 44.2 | 0.23 |
| polarC18 | 325.216 | 139.3828 | 44.2 | 0.23 |
| polarC18 | 325.216 | 291.8466 | 44.2 | 0.23 |
| polarC18 | 325.216 | 120.8179 | 44.2 | 0.23 |
| polarC18 | 325.216 | 83.8185 | 44.2 | 0.23 |
| polarC18 | 325.216 | 103.3983 | 44.1 | 0.23 |
| polarC18 | 325.216 | 89.8645 | 44.1 | 0.23 |
| polarC18 | 325.216 | 76.3743 | 44 | 0.23 |
| polarC18 | 325.216 | 306.7729 | 44 | 0.23 |
| polarC18 | 325.216 | 90.0748 | 43.9 | 0.23 |
| polarC18 | 325.216 | 119.4999 | 43.9 | 0.23 |
| polarC18 | 325.216 | 173.6063 | 43.8 | 0.23 |
| polarC18 | 325.216 | 136.8947 | 43.8 | 0.23 |
| polarC18 | 325.216 | 305.823 | 43.8 | 0.23 |
| polarC18 | 325.216 | 60.5324 | 43.7 | 0.22 |
| polarC18 | 325.216 | 95.8079 | 43.7 | 0.22 |
| polarC18 | 325.216 | 211.8495 | 43.7 | 0.22 |
| polarC18 | 325.216 | 165.6973 | 43.7 | 0.22 |
| polarC18 | 325.216 | 140.5389 | 43.6 | 0.22 |
| polarC18 | 325.216 | 316.1422 | 43.6 | 0.22 |
| polarC18 | 325.216 | 200.3306 | 43.5 | 0.22 |
| polarC18 | 325.216 | 54.5693 | 43.4 | 0.22 |
| polarC18 | 325.216 | 139.5977 | 43.4 | 0.22 |
| polarC18 | 325.216 | 65.7666 | 43.4 | 0.22 |
| polarC18 | 325.216 | 193.3323 | 43.4 | 0.22 |
| polarC18 | 325.216 | 209.2645 | 43.3 | 0.22 |
| polarC18 | 325.216 | 111.4369 | 43.3 | 0.22 |
| polarC18 | 325.216 | 93.3656 | 43.3 | 0.22 |
| polarC18 | 325.216 | 221.1044 | 43.3 | 0.22 |
| polarC18 | 325.216 | 72.9348 | 43.3 | 0.22 |

| | | | | |
|---|---|---|---|---|
| polarC18 | 325.216 | 159.882 | 43.2 | 0.22 |
| polarC18 | 325.216 | 177.0835 | 43.2 | 0.22 |
| polarC18 | 325.216 | 101.6689 | 43.2 | 0.22 |
| polarC18 | 325.216 | 199.8638 | 43.1 | 0.22 |
| polarC18 | 325.216 | 157.1507 | 43 | 0.22 |
| polarC18 | 325.216 | 133.1582 | 43 | 0.22 |
| polarC18 | 325.216 | 51.5603 | 43 | 0.22 |
| polarC18 | 325.216 | 108.5945 | 42.9 | 0.22 |
| polarC18 | 325.216 | 90.1273 | 42.8 | 0.22 |
| polarC18 | 325.216 | 110.676 | 42.7 | 0.22 |
| polarC18 | 325.216 | 322.9278 | 42.7 | 0.22 |
| polarC18 | 325.216 | 69.9611 | 42.6 | 0.22 |
| polarC18 | 325.216 | 93.3329 | 42.6 | 0.22 |
| polarC18 | 325.216 | 307.9652 | 42.6 | 0.22 |
| polarC18 | 325.216 | 72.4114 | 42.5 | 0.22 |
| polarC18 | 325.216 | 219.6645 | 42.5 | 0.22 |
| polarC18 | 325.216 | 55.8603 | 42.4 | 0.22 |
| polarC18 | 325.216 | 59.4814 | 42.4 | 0.22 |
| polarC18 | 325.216 | 72.3813 | 42.3 | 0.22 |
| polarC18 | 325.216 | 87.575 | 42.3 | 0.22 |
| polarC18 | 325.216 | 67.8316 | 42.3 | 0.22 |
| polarC18 | 325.216 | 150.5937 | 42.2 | 0.22 |
| polarC18 | 325.216 | 91.715 | 42 | 0.22 |
| polarC18 | 325.216 | 87.8117 | 41.8 | 0.21 |
| polarC18 | 325.216 | 119.9666 | 41.8 | 0.21 |
| polarC18 | 325.216 | 95.6301 | 41.7 | 0.21 |
| polarC18 | 325.216 | 128.5294 | 41.6 | 0.21 |
| polarC18 | 325.216 | 75.4808 | 41.6 | 0.21 |
| polarC18 | 325.216 | 154.386 | 41.6 | 0.21 |
| polarC18 | 325.216 | 62.1496 | 41.5 | 0.21 |
| polarC18 | 325.216 | 212.0206 | 41.3 | 0.21 |
| polarC18 | 325.216 | 157.7028 | 41.3 | 0.21 |
| polarC18 | 325.216 | 87.9609 | 41.1 | 0.21 |
| polarC18 | 325.216 | 164.0708 | 41.1 | 0.21 |
| polarC18 | 325.216 | 93.4593 | 41.1 | 0.21 |
| polarC18 | 325.216 | 281.9985 | 40.6 | 0.21 |
| polarC18 | 325.216 | 56.6492 | 40.3 | 0.21 |
| polarC18 | 325.216 | 62.4981 | 40.3 | 0.21 |
| polarC18 | 325.216 | 78.5836 | 40 | 0.21 |
| polarC18 | 325.216 | 119.1233 | 39.8 | 0.2 |
| polarC18 | 325.216 | 69.2611 | 39 | 0.2 |
| polarC18 | 325.216 | 60.3831 | 37.7 | 0.19 |

| | | | | |
|---|---|---|---|---|
| polarC18 | 325.216 | 60.1188 | 32.3 | 0.17 |
| polarC18 | 325.216 | 87.0494 | 31.8 | 0.16 |
| polarC18 | 325.216 | 300.9751 | 30.8 | 0.16 |
| polarC18 | 325.216 | 252.0481 | 30.6 | 0.16 |
| polarC18 | 325.216 | 202.9344 | 30.5 | 0.16 |
| polarC18 | 325.216 | 112.2944 | 30.4 | 0.16 |
| polarC18 | 325.216 | 61.4251 | 30.4 | 0.16 |
| polarC18 | 325.216 | 92.2801 | 30.3 | 0.16 |
| polarC18 | 325.216 | 206.8869 | 30.1 | 0.15 |
| polarC18 | 325.216 | 177.1455 | 30 | 0.15 |
| polarC18 | 325.216 | 260.6212 | 29.8 | 0.15 |
| polarC18 | 325.216 | 210.8116 | 29.8 | 0.15 |
| polarC18 | 325.216 | 98.1384 | 29.8 | 0.15 |
| polarC18 | 325.216 | 198.6142 | 29.7 | 0.15 |
| polarC18 | 325.216 | 86.0952 | 29.7 | 0.15 |
| polarC18 | 325.216 | 150.193 | 29.1 | 0.15 |
| polarC18 | 325.216 | 128.8678 | 29 | 0.15 |
| polarC18 | 325.216 | 82.8863 | 28.9 | 0.15 |
| polarC18 | 325.216 | 100.5114 | 28.9 | 0.15 |
| polarC18 | 325.216 | 125.794 | 28.8 | 0.15 |
| polarC18 | 325.216 | 208.984 | 28.7 | 0.15 |
| polarC18 | 325.216 | 318.0542 | 28.6 | 0.15 |
| polarC18 | 325.216 | 76.41 | 28.6 | 0.15 |
| polarC18 | 325.216 | 275.327 | 28.6 | 0.15 |
| polarC18 | 325.216 | 125.1846 | 28.5 | 0.15 |
| polarC18 | 325.216 | 230.7385 | 28.5 | 0.15 |
| polarC18 | 325.216 | 72.7795 | 28.5 | 0.15 |
| polarC18 | 325.216 | 63.1489 | 28.3 | 0.15 |
| polarC18 | 325.216 | 270.7191 | 28.3 | 0.15 |
| polarC18 | 325.216 | 138.3373 | 28.3 | 0.15 |
| polarC18 | 325.216 | 129.2009 | 28.2 | 0.15 |
| polarC18 | 325.216 | 245.8789 | 28.2 | 0.15 |
| polarC18 | 325.216 | 70.381 | 28.2 | 0.14 |
| polarC18 | 325.216 | 220.3098 | 28.2 | 0.14 |
| polarC18 | 325.216 | 66.0601 | 28.1 | 0.14 |
| polarC18 | 325.216 | 90.8954 | 28.1 | 0.14 |
| polarC18 | 325.216 | 67.2404 | 28 | 0.14 |
| polarC18 | 325.216 | 163.4297 | 28 | 0.14 |
| polarC18 | 325.216 | 193.2383 | 27.9 | 0.14 |
| polarC18 | 325.216 | 126.498 | 27.9 | 0.14 |
| polarC18 | 325.216 | 88.0373 | 27.9 | 0.14 |
| polarC18 | 325.216 | 73.4868 | 27.9 | 0.14 |

| | | | | |
|---|---|---|---|---|
| polarC18 | 325.216 | 87.6484 | 27.8 | 0.14 |
| polarC18 | 325.216 | 281.8765 | 27.8 | 0.14 |
| polarC18 | 325.216 | 220.8328 | 27.8 | 0.14 |
| polarC18 | 325.216 | 130.83 | 27.8 | 0.14 |
| polarC18 | 325.216 | 110.0886 | 27.7 | 0.14 |
| polarC18 | 325.216 | 82.7615 | 27.7 | 0.14 |
| polarC18 | 325.216 | 210.6352 | 27.7 | 0.14 |
| polarC18 | 325.216 | 100.1584 | 27.7 | 0.14 |
| polarC18 | 325.216 | 59.0829 | 27.6 | 0.14 |
| polarC18 | 325.216 | 141.4457 | 27.6 | 0.14 |
| polarC18 | 325.216 | 159.0554 | 27.5 | 0.14 |
| polarC18 | 325.216 | 345.6736 | 27.5 | 0.14 |
| polarC18 | 325.216 | 114.1384 | 27.5 | 0.14 |
| polarC18 | 325.216 | 80.2508 | 27.5 | 0.14 |
| polarC18 | 325.216 | 250.1064 | 27.5 | 0.14 |
| polarC18 | 325.216 | 135.4581 | 27.4 | 0.14 |
| polarC18 | 325.216 | 233.6084 | 27.4 | 0.14 |
| polarC18 | 325.216 | 229.6925 | 27.4 | 0.14 |
| polarC18 | 325.216 | 58.0097 | 27.4 | 0.14 |
| polarC18 | 325.216 | 230.0926 | 27.4 | 0.14 |
| polarC18 | 325.216 | 106.4714 | 27.4 | 0.14 |
| polarC18 | 325.216 | 339.2789 | 27.4 | 0.14 |
| polarC18 | 325.216 | 209.3645 | 27.4 | 0.14 |
| polarC18 | 325.216 | 79.4887 | 27.3 | 0.14 |
| polarC18 | 325.216 | 145.0615 | 27.3 | 0.14 |
| polarC18 | 325.216 | 301.6763 | 27.3 | 0.14 |
| polarC18 | 325.216 | 57.5589 | 27.3 | 0.14 |
| polarC18 | 325.216 | 244.4788 | 27.3 | 0.14 |
| polarC18 | 325.216 | 341.4416 | 27.2 | 0.14 |
| polarC18 | 325.216 | 155.7388 | 27.2 | 0.14 |
| polarC18 | 325.216 | 193.5901 | 27.1 | 0.14 |
| polarC18 | 325.216 | 285.2506 | 27.1 | 0.14 |
| polarC18 | 325.216 | 75.5573 | 27.1 | 0.14 |
| polarC18 | 325.216 | 184.7142 | 27.1 | 0.14 |
| polarC18 | 325.216 | 59.1431 | 27.1 | 0.14 |
| polarC18 | 325.216 | 197.4674 | 27 | 0.14 |
| polarC18 | 325.216 | 74.338 | 27 | 0.14 |
| polarC18 | 325.216 | 226.5699 | 27 | 0.14 |
| polarC18 | 325.216 | 165.1221 | 26.9 | 0.14 |
| polarC18 | 325.216 | 63.5006 | 26.9 | 0.14 |
| polarC18 | 325.216 | 159.2376 | 26.9 | 0.14 |
| polarC18 | 325.216 | 63.0199 | 26.9 | 0.14 |

| | | | | |
|---|---|---|---|---|
| polarC18 | 325.216 | 60.7306 | 26.9 | 0.14 |
| polarC18 | 325.216 | 118.4641 | 26.9 | 0.14 |
| polarC18 | 325.216 | 246.5794 | 26.8 | 0.14 |
| polarC18 | 325.216 | 233.2263 | 26.8 | 0.14 |
| polarC18 | 325.216 | 50.8279 | 26.8 | 0.14 |
| polarC18 | 325.216 | 149.895 | 26.8 | 0.14 |
| polarC18 | 325.216 | 174.3185 | 26.8 | 0.14 |
| polarC18 | 325.216 | 52.1041 | 26.8 | 0.14 |
| polarC18 | 325.216 | 69.1453 | 26.8 | 0.14 |
| polarC18 | 325.216 | 293.3746 | 26.8 | 0.14 |
| polarC18 | 325.216 | 105.9751 | 26.8 | 0.14 |
| polarC18 | 325.216 | 51.9899 | 26.7 | 0.14 |
| polarC18 | 325.216 | 59.2113 | 26.7 | 0.14 |
| polarC18 | 325.216 | 85.4017 | 26.7 | 0.14 |
| polarC18 | 325.216 | 174.9611 | 26.7 | 0.14 |
| polarC18 | 325.216 | 109.2899 | 26.6 | 0.14 |
| polarC18 | 325.216 | 143.1923 | 26.6 | 0.14 |
| polarC18 | 325.216 | 181.1374 | 26.6 | 0.14 |
| polarC18 | 325.216 | 73.5058 | 26.6 | 0.14 |
| polarC18 | 325.216 | 159.285 | 26.6 | 0.14 |
| polarC18 | 325.216 | 188.8725 | 26.6 | 0.14 |
| polarC18 | 325.216 | 134.5948 | 26.6 | 0.14 |
| polarC18 | 325.216 | 197.4403 | 26.6 | 0.14 |
| polarC18 | 325.216 | 214.81 | 26.6 | 0.14 |
| polarC18 | 325.216 | 184.7953 | 26.6 | 0.14 |
| polarC18 | 325.216 | 110.6253 | 26.5 | 0.14 |
| polarC18 | 325.216 | 119.2925 | 26.5 | 0.14 |
| polarC18 | 325.216 | 99.5929 | 26.5 | 0.14 |
| polarC18 | 325.216 | 336.2474 | 26.5 | 0.14 |
| polarC18 | 325.216 | 312.9934 | 26.5 | 0.14 |
| polarC18 | 325.216 | 171.879 | 26.5 | 0.14 |
| polarC18 | 325.216 | 113.4443 | 26.5 | 0.14 |
| polarC18 | 325.216 | 92.9075 | 26.5 | 0.14 |
| polarC18 | 325.216 | 58.6764 | 26.5 | 0.14 |
| polarC18 | 325.216 | 139.4607 | 26.5 | 0.14 |
| polarC18 | 325.216 | 190.7513 | 26.4 | 0.14 |
| polarC18 | 325.216 | 71.3108 | 26.4 | 0.14 |
| polarC18 | 325.216 | 53.7699 | 26.4 | 0.14 |
| polarC18 | 325.216 | 120.2887 | 26.4 | 0.14 |
| polarC18 | 325.216 | 101.4978 | 26.4 | 0.14 |
| polarC18 | 325.216 | 227.9327 | 26.3 | 0.14 |
| polarC18 | 325.216 | 194.5926 | 26.3 | 0.14 |

| | | | | |
|---|---|---|---|---|
| polarC18 | 325.216 | 196.3979 | 26.3 | 0.14 |
| polarC18 | 325.216 | 176.2784 | 26.3 | 0.14 |
| polarC18 | 325.216 | 80.9335 | 26.3 | 0.14 |
| polarC18 | 325.216 | 63.6011 | 26.3 | 0.14 |
| polarC18 | 325.216 | 108.2851 | 26.3 | 0.14 |
| polarC18 | 325.216 | 179.4427 | 26.3 | 0.14 |
| polarC18 | 325.216 | 93.5206 | 26.3 | 0.14 |
| polarC18 | 325.216 | 82.0252 | 26.3 | 0.14 |
| polarC18 | 325.216 | 81.0695 | 26.3 | 0.14 |
| polarC18 | 325.216 | 190.0602 | 26.3 | 0.14 |
| polarC18 | 325.216 | 123.8926 | 26.3 | 0.13 |
| polarC18 | 325.216 | 53.1047 | 26.3 | 0.13 |
| polarC18 | 325.216 | 197.6881 | 26.3 | 0.13 |
| polarC18 | 325.216 | 239.655 | 26.2 | 0.13 |
| polarC18 | 325.216 | 70.737 | 26.2 | 0.13 |
| polarC18 | 325.216 | 108.0374 | 26.2 | 0.13 |
| polarC18 | 325.216 | 185.2593 | 26.2 | 0.13 |
| polarC18 | 325.216 | 105.9418 | 26.2 | 0.13 |
| polarC18 | 325.216 | 136.765 | 26.2 | 0.13 |
| polarC18 | 325.216 | 200.8772 | 26.2 | 0.13 |
| polarC18 | 325.216 | 190.1853 | 26.2 | 0.13 |
| polarC18 | 325.216 | 317.5926 | 26.1 | 0.13 |
| polarC18 | 325.216 | 213.3955 | 26.1 | 0.13 |
| polarC18 | 325.216 | 240.8058 | 26.1 | 0.13 |
| polarC18 | 325.216 | 304.6878 | 26.1 | 0.13 |
| polarC18 | 325.216 | 245.4395 | 26.1 | 0.13 |
| polarC18 | 325.216 | 53.4857 | 26 | 0.13 |
| polarC18 | 325.216 | 53.9575 | 26 | 0.13 |
| polarC18 | 325.216 | 93.2932 | 26 | 0.13 |
| polarC18 | 325.216 | 209.8897 | 26 | 0.13 |
| polarC18 | 325.216 | 77.9462 | 26 | 0.13 |
| polarC18 | 325.216 | 126.0321 | 25.9 | 0.13 |
| polarC18 | 325.216 | 173.6313 | 25.9 | 0.13 |
| polarC18 | 325.216 | 59.3116 | 25.9 | 0.13 |
| polarC18 | 325.216 | 93.9554 | 25.9 | 0.13 |
| polarC18 | 325.216 | 124.6506 | 25.9 | 0.13 |
| polarC18 | 325.216 | 288.1506 | 25.9 | 0.13 |
| polarC18 | 325.216 | 74.259 | 25.9 | 0.13 |
| polarC18 | 325.216 | 69.9507 | 25.9 | 0.13 |
| polarC18 | 325.216 | 52.3633 | 25.9 | 0.13 |
| polarC18 | 325.216 | 74.501 | 25.9 | 0.13 |
| polarC18 | 325.216 | 344.3691 | 25.9 | 0.13 |

| | | | | |
|---|---|---|---|---|
| polarC18 | 325.216 | 319.5334 | 25.9 | 0.13 |
| polarC18 | 325.216 | 146.2994 | 25.9 | 0.13 |
| polarC18 | 325.216 | 336.0452 | 25.9 | 0.13 |
| polarC18 | 325.216 | 65.8908 | 25.9 | 0.13 |
| polarC18 | 325.216 | 52.9783 | 25.8 | 0.13 |
| polarC18 | 325.216 | 123.8079 | 25.8 | 0.13 |
| polarC18 | 325.216 | 98.0424 | 25.8 | 0.13 |
| polarC18 | 325.216 | 104.3331 | 25.8 | 0.13 |
| polarC18 | 325.216 | 110.2715 | 25.8 | 0.13 |
| polarC18 | 325.216 | 54.6211 | 25.8 | 0.13 |
| polarC18 | 325.216 | 123.5951 | 25.8 | 0.13 |
| polarC18 | 325.216 | 79.9032 | 25.8 | 0.13 |
| polarC18 | 325.216 | 149.8322 | 25.8 | 0.13 |
| polarC18 | 325.216 | 126.303 | 25.8 | 0.13 |
| polarC18 | 325.216 | 64.2322 | 25.8 | 0.13 |
| polarC18 | 325.216 | 87.0259 | 25.8 | 0.13 |
| polarC18 | 325.216 | 99.9323 | 25.8 | 0.13 |
| polarC18 | 325.216 | 93.7808 | 25.8 | 0.13 |
| polarC18 | 325.216 | 88.4329 | 25.7 | 0.13 |
| polarC18 | 325.216 | 338.4562 | 25.7 | 0.13 |
| polarC18 | 325.216 | 320.4159 | 25.7 | 0.13 |
| polarC18 | 325.216 | 184.3779 | 25.7 | 0.13 |
| polarC18 | 325.216 | 220.1754 | 25.7 | 0.13 |
| polarC18 | 325.216 | 70.7699 | 25.7 | 0.13 |
| polarC18 | 325.216 | 228.4573 | 25.7 | 0.13 |
| polarC18 | 325.216 | 304.314 | 25.7 | 0.13 |
| polarC18 | 325.216 | 144.7435 | 25.7 | 0.13 |
| polarC18 | 325.216 | 162.9967 | 25.7 | 0.13 |
| polarC18 | 325.216 | 193.8461 | 25.7 | 0.13 |
| polarC18 | 325.216 | 215.4548 | 25.7 | 0.13 |
| polarC18 | 325.216 | 104.4869 | 25.7 | 0.13 |
| polarC18 | 325.216 | 53.6447 | 25.6 | 0.13 |
| polarC18 | 325.216 | 74.0517 | 25.6 | 0.13 |
| polarC18 | 325.216 | 256.0612 | 25.6 | 0.13 |
| polarC18 | 325.216 | 112.7325 | 25.6 | 0.13 |
| polarC18 | 325.216 | 281.4194 | 25.6 | 0.13 |
| polarC18 | 325.216 | 63.3957 | 25.6 | 0.13 |
| polarC18 | 325.216 | 207.6393 | 25.6 | 0.13 |
| polarC18 | 325.216 | 187.3152 | 25.6 | 0.13 |
| polarC18 | 325.216 | 297.6796 | 25.6 | 0.13 |
| polarC18 | 325.216 | 115.6087 | 25.6 | 0.13 |
| polarC18 | 325.216 | 147.1181 | 25.6 | 0.13 |

| | | | | |
|---|---|---|---|---|
| polarC18 | 325.216 | 200.158 | 25.6 | 0.13 |
| polarC18 | 325.216 | 252.2299 | 25.5 | 0.13 |
| polarC18 | 325.216 | 241.7616 | 25.5 | 0.13 |
| polarC18 | 325.216 | 222.5491 | 25.5 | 0.13 |
| polarC18 | 325.216 | 61.2133 | 25.5 | 0.13 |
| polarC18 | 325.216 | 147.2849 | 25.5 | 0.13 |
| polarC18 | 325.216 | 54.067 | 25.5 | 0.13 |
| polarC18 | 325.216 | 327.6978 | 25.5 | 0.13 |
| polarC18 | 325.216 | 80.1389 | 25.5 | 0.13 |
| polarC18 | 325.216 | 56.7405 | 25.5 | 0.13 |
| polarC18 | 325.216 | 50.2931 | 25.5 | 0.13 |
| polarC18 | 325.216 | 66.3733 | 25.5 | 0.13 |
| polarC18 | 325.216 | 302.3045 | 25.5 | 0.13 |
| polarC18 | 325.216 | 50.7334 | 25.5 | 0.13 |
| polarC18 | 325.216 | 147.5478 | 25.5 | 0.13 |
| polarC18 | 325.216 | 106.3698 | 25.5 | 0.13 |
| polarC18 | 325.216 | 95.2365 | 25.5 | 0.13 |
| polarC18 | 325.216 | 96.0548 | 25.5 | 0.13 |
| polarC18 | 325.216 | 342.3665 | 25.5 | 0.13 |
| polarC18 | 325.216 | 54.5515 | 25.4 | 0.13 |
| polarC18 | 325.216 | 58.3171 | 25.4 | 0.13 |
| polarC18 | 325.216 | 83.9298 | 25.4 | 0.13 |
| polarC18 | 325.216 | 117.6059 | 25.4 | 0.13 |
| polarC18 | 325.216 | 212.2813 | 25.4 | 0.13 |
| polarC18 | 325.216 | 154.9763 | 25.4 | 0.13 |
| polarC18 | 325.216 | 106.0562 | 25.4 | 0.13 |
| polarC18 | 325.216 | 52.9918 | 25.4 | 0.13 |
| polarC18 | 325.216 | 62.2957 | 25.4 | 0.13 |
| polarC18 | 325.216 | 55.3655 | 25.4 | 0.13 |
| polarC18 | 325.216 | 91.663 | 25.4 | 0.13 |
| polarC18 | 325.216 | 351.3319 | 25.4 | 0.13 |
| polarC18 | 325.216 | 254.4408 | 25.4 | 0.13 |
| polarC18 | 325.216 | 211.7616 | 25.4 | 0.13 |
| polarC18 | 325.216 | 63.9969 | 25.4 | 0.13 |
| polarC18 | 325.216 | 85.2021 | 25.4 | 0.13 |
| polarC18 | 325.216 | 196.5591 | 25.3 | 0.13 |
| polarC18 | 325.216 | 87.4439 | 25.3 | 0.13 |
| polarC18 | 325.216 | 161.4521 | 25.3 | 0.13 |
| polarC18 | 325.216 | 168.3813 | 25.3 | 0.13 |
| polarC18 | 325.216 | 278.9601 | 25.3 | 0.13 |
| polarC18 | 325.216 | 182.3462 | 25.3 | 0.13 |
| polarC18 | 325.216 | 98.3216 | 25.3 | 0.13 |

| | | | | |
|---|---|---|---|---|
| polarC18 | 325.216 | 109.7526 | 25.3 | 0.13 |
| polarC18 | 325.216 | 114.595 | 25.3 | 0.13 |
| polarC18 | 325.216 | 73.1464 | 25.3 | 0.13 |
| polarC18 | 325.216 | 119.0972 | 25.3 | 0.13 |
| polarC18 | 325.216 | 240.6694 | 25.3 | 0.13 |
| polarC18 | 325.216 | 58.6057 | 25.3 | 0.13 |
| polarC18 | 325.216 | 168.7849 | 25.3 | 0.13 |
| polarC18 | 325.216 | 213.3251 | 25.2 | 0.13 |
| polarC18 | 325.216 | 86.7184 | 25.2 | 0.13 |
| polarC18 | 325.216 | 109.6716 | 25.2 | 0.13 |
| polarC18 | 325.216 | 199.1542 | 25.2 | 0.13 |
| polarC18 | 325.216 | 52.8692 | 25.2 | 0.13 |
| polarC18 | 325.216 | 257.4472 | 25.2 | 0.13 |
| polarC18 | 325.216 | 66.9575 | 25.2 | 0.13 |
| polarC18 | 325.216 | 96.6569 | 25.2 | 0.13 |
| polarC18 | 325.216 | 268.161 | 25.2 | 0.13 |
| polarC18 | 325.216 | 107.5093 | 25.2 | 0.13 |
| polarC18 | 325.216 | 214.2793 | 25.2 | 0.13 |
| polarC18 | 325.216 | 149.0803 | 25.2 | 0.13 |
| polarC18 | 325.216 | 284.6486 | 25.2 | 0.13 |
| polarC18 | 325.216 | 193.6397 | 25.2 | 0.13 |
| polarC18 | 325.216 | 117.1959 | 25.2 | 0.13 |
| polarC18 | 325.216 | 97.066 | 25.2 | 0.13 |
| polarC18 | 325.216 | 314.2603 | 25.2 | 0.13 |
| polarC18 | 325.216 | 54.4013 | 25.1 | 0.13 |
| polarC18 | 325.216 | 80.4856 | 25.1 | 0.13 |
| polarC18 | 325.216 | 122.2539 | 25.1 | 0.13 |
| polarC18 | 325.216 | 56.0272 | 25.1 | 0.13 |
| polarC18 | 325.216 | 79.1814 | 25.1 | 0.13 |
| polarC18 | 325.216 | 167.1603 | 25.1 | 0.13 |
| polarC18 | 325.216 | 157.2063 | 25.1 | 0.13 |
| polarC18 | 325.216 | 84.2165 | 25.1 | 0.13 |
| polarC18 | 325.216 | 57.3929 | 25.1 | 0.13 |
| polarC18 | 325.216 | 105.412 | 25.1 | 0.13 |
| polarC18 | 325.216 | 98.1143 | 25.1 | 0.13 |
| polarC18 | 325.216 | 154.2666 | 25.1 | 0.13 |
| polarC18 | 325.216 | 217.7807 | 25.1 | 0.13 |
| polarC18 | 325.216 | 70.322 | 25.1 | 0.13 |
| polarC18 | 325.216 | 54.9161 | 25 | 0.13 |
| polarC18 | 325.216 | 75.2554 | 25 | 0.13 |
| polarC18 | 325.216 | 308.6785 | 25 | 0.13 |
| polarC18 | 325.216 | 223.6854 | 25 | 0.13 |

| | | | | |
|---|---|---|---|---|
| polarC18 | 325.216 | 53.9716 | 25 | 0.13 |
| polarC18 | 325.216 | 311.9224 | 25 | 0.13 |
| polarC18 | 325.216 | 131.1393 | 25 | 0.13 |
| polarC18 | 325.216 | 71.9409 | 25 | 0.13 |
| polarC18 | 325.216 | 97.136 | 25 | 0.13 |
| polarC18 | 325.216 | 228.8456 | 25 | 0.13 |
| polarC18 | 325.216 | 220.5952 | 25 | 0.13 |
| polarC18 | 325.216 | 73.463 | 25 | 0.13 |
| polarC18 | 325.216 | 53.1965 | 25 | 0.13 |
| polarC18 | 325.216 | 89.1512 | 25 | 0.13 |
| polarC18 | 325.216 | 200.6891 | 24.9 | 0.13 |
| polarC18 | 325.216 | 222.3813 | 24.9 | 0.13 |
| polarC18 | 325.216 | 96.5624 | 24.9 | 0.13 |
| polarC18 | 325.216 | 170.696 | 24.9 | 0.13 |
| polarC18 | 325.216 | 66.4109 | 24.9 | 0.13 |
| polarC18 | 325.216 | 72.4596 | 24.9 | 0.13 |
| polarC18 | 325.216 | 125.3993 | 24.9 | 0.13 |
| polarC18 | 325.216 | 86.6961 | 24.9 | 0.13 |
| polarC18 | 325.216 | 113.8576 | 24.9 | 0.13 |
| polarC18 | 325.216 | 100.4751 | 24.9 | 0.13 |
| polarC18 | 325.216 | 147.2537 | 24.9 | 0.13 |
| polarC18 | 325.216 | 264.7398 | 24.9 | 0.13 |
| polarC18 | 325.216 | 70.3312 | 24.9 | 0.13 |
| polarC18 | 325.216 | 93.5512 | 24.9 | 0.13 |
| polarC18 | 325.216 | 230.5797 | 24.8 | 0.13 |
| polarC18 | 325.216 | 161.0305 | 24.8 | 0.13 |
| polarC18 | 325.216 | 93.4092 | 24.8 | 0.13 |
| polarC18 | 325.216 | 119.0337 | 24.8 | 0.13 |
| polarC18 | 325.216 | 117.3367 | 24.8 | 0.13 |
| polarC18 | 325.216 | 134.7588 | 24.8 | 0.13 |
| polarC18 | 325.216 | 201.2231 | 24.8 | 0.13 |
| polarC18 | 325.216 | 164.3452 | 24.8 | 0.13 |
| polarC18 | 325.216 | 286.4731 | 24.8 | 0.13 |
| polarC18 | 325.216 | 72.2551 | 24.8 | 0.13 |
| polarC18 | 325.216 | 91.1709 | 24.8 | 0.13 |
| polarC18 | 325.216 | 99.8536 | 24.8 | 0.13 |
| polarC18 | 325.216 | 207.7703 | 24.8 | 0.13 |
| polarC18 | 325.216 | 57.3681 | 24.8 | 0.13 |
| polarC18 | 325.216 | 102.4054 | 24.8 | 0.13 |
| polarC18 | 325.216 | 82.2705 | 24.8 | 0.13 |
| polarC18 | 325.216 | 56.4657 | 24.8 | 0.13 |
| polarC18 | 325.216 | 248.9317 | 24.8 | 0.13 |

| | | | | |
|---|---|---|---|---|
| polarC18 | 325.216 | 108.8353 | 24.8 | 0.13 |
| polarC18 | 325.216 | 245.107 | 24.8 | 0.13 |
| polarC18 | 325.216 | 223.5418 | 24.8 | 0.13 |
| polarC18 | 325.216 | 56.9039 | 24.8 | 0.13 |
| polarC18 | 325.216 | 275.5311 | 24.8 | 0.13 |
| polarC18 | 325.216 | 337.8934 | 24.7 | 0.13 |
| polarC18 | 325.216 | 186.5822 | 24.7 | 0.13 |
| polarC18 | 325.216 | 133.6605 | 24.7 | 0.13 |
| polarC18 | 325.216 | 56.8135 | 24.7 | 0.13 |
| polarC18 | 325.216 | 72.2577 | 24.7 | 0.13 |
| polarC18 | 325.216 | 58.5135 | 24.7 | 0.13 |
| polarC18 | 325.216 | 223.353 | 24.7 | 0.13 |
| polarC18 | 325.216 | 52.7827 | 24.7 | 0.13 |
| polarC18 | 325.216 | 239.1386 | 24.7 | 0.13 |
| polarC18 | 325.216 | 75.7562 | 24.7 | 0.13 |
| polarC18 | 325.216 | 77.3656 | 24.7 | 0.13 |
| polarC18 | 325.216 | 143.8913 | 24.7 | 0.13 |
| polarC18 | 325.216 | 64.8789 | 24.7 | 0.13 |
| polarC18 | 325.216 | 57.9677 | 24.7 | 0.13 |
| polarC18 | 325.216 | 52.8995 | 24.7 | 0.13 |
| polarC18 | 325.216 | 272.0224 | 24.7 | 0.13 |
| polarC18 | 325.216 | 121.2784 | 24.7 | 0.13 |
| polarC18 | 325.216 | 66.1558 | 24.7 | 0.13 |
| polarC18 | 325.216 | 90.0999 | 24.7 | 0.13 |
| polarC18 | 325.216 | 102.9556 | 24.7 | 0.13 |
| polarC18 | 325.216 | 285.886 | 24.7 | 0.13 |
| polarC18 | 325.216 | 111.9761 | 24.7 | 0.13 |
| polarC18 | 325.216 | 64.2712 | 24.7 | 0.13 |
| polarC18 | 325.216 | 93.3563 | 24.7 | 0.13 |
| polarC18 | 325.216 | 223.292 | 24.6 | 0.13 |
| polarC18 | 325.216 | 120.563 | 24.6 | 0.13 |
| polarC18 | 325.216 | 202.8487 | 24.6 | 0.13 |
| polarC18 | 325.216 | 50.9524 | 24.6 | 0.13 |
| polarC18 | 325.216 | 326.5656 | 24.6 | 0.13 |
| polarC18 | 325.216 | 94.979 | 24.6 | 0.13 |
| polarC18 | 325.216 | 217.7475 | 24.6 | 0.13 |
| polarC18 | 325.216 | 50.3667 | 24.6 | 0.13 |
| polarC18 | 325.216 | 92.8191 | 24.6 | 0.13 |
| polarC18 | 325.216 | 67.1958 | 24.6 | 0.13 |
| polarC18 | 325.216 | 204.1151 | 24.6 | 0.13 |
| polarC18 | 325.216 | 86.2608 | 24.6 | 0.13 |
| polarC18 | 325.216 | 150.0287 | 24.6 | 0.13 |

| | | | | |
|---|---|---|---|---|
| polarC18 | 325.216 | 56.6064 | 24.6 | 0.13 |
| polarC18 | 325.216 | 57.9368 | 24.6 | 0.13 |
| polarC18 | 325.216 | 91.381 | 24.6 | 0.13 |
| polarC18 | 325.216 | 70.9112 | 24.6 | 0.13 |
| polarC18 | 325.216 | 132.8185 | 24.5 | 0.13 |
| polarC18 | 325.216 | 286.2597 | 24.5 | 0.13 |
| polarC18 | 325.216 | 108.5214 | 24.5 | 0.13 |
| polarC18 | 325.216 | 199.5957 | 24.5 | 0.13 |
| polarC18 | 325.216 | 135.7398 | 24.5 | 0.13 |
| polarC18 | 325.216 | 161.1618 | 24.5 | 0.13 |
| polarC18 | 325.216 | 78.2192 | 24.5 | 0.13 |
| polarC18 | 325.216 | 68.6741 | 24.5 | 0.13 |
| polarC18 | 325.216 | 93.0137 | 24.5 | 0.13 |
| polarC18 | 325.216 | 202.6789 | 24.5 | 0.13 |
| polarC18 | 325.216 | 153.3019 | 24.5 | 0.13 |
| polarC18 | 325.216 | 279.1457 | 24.5 | 0.13 |
| polarC18 | 325.216 | 77.5513 | 24.5 | 0.13 |
| polarC18 | 325.216 | 122.3459 | 24.5 | 0.13 |
| polarC18 | 325.216 | 337.3452 | 24.5 | 0.13 |
| polarC18 | 325.216 | 64.3967 | 24.5 | 0.13 |
| polarC18 | 325.216 | 150.9611 | 24.5 | 0.13 |
| polarC18 | 325.216 | 140.216 | 24.5 | 0.13 |
| polarC18 | 325.216 | 220.4124 | 24.5 | 0.13 |
| polarC18 | 325.216 | 174.715 | 24.5 | 0.13 |
| polarC18 | 325.216 | 122.6732 | 24.5 | 0.13 |
| polarC18 | 325.216 | 63.9925 | 24.5 | 0.13 |
| polarC18 | 325.216 | 61.4695 | 24.4 | 0.13 |
| polarC18 | 325.216 | 270.8909 | 24.4 | 0.13 |
| polarC18 | 325.216 | 137.7164 | 24.4 | 0.13 |
| polarC18 | 325.216 | 72.3423 | 24.4 | 0.13 |
| polarC18 | 325.216 | 79.6053 | 24.4 | 0.13 |
| polarC18 | 325.216 | 172.4231 | 24.4 | 0.13 |
| polarC18 | 325.216 | 128.1276 | 24.4 | 0.13 |
| polarC18 | 325.216 | 192.082 | 24.4 | 0.13 |
| polarC18 | 325.216 | 50.7691 | 24.4 | 0.13 |
| polarC18 | 325.216 | 95.6578 | 24.4 | 0.13 |
| polarC18 | 325.216 | 54.3052 | 24.4 | 0.13 |
| polarC18 | 325.216 | 170.3811 | 24.4 | 0.13 |
| polarC18 | 325.216 | 94.5623 | 24.4 | 0.13 |
| polarC18 | 325.216 | 133.8823 | 24.4 | 0.13 |
| polarC18 | 325.216 | 180.9834 | 24.4 | 0.13 |
| polarC18 | 325.216 | 133.3322 | 24.4 | 0.13 |

| | | | | |
|---|---|---|---|---|
| polarC18 | 325.216 | 176.7528 | 24.4 | 0.13 |
| polarC18 | 325.216 | 222.48 | 24.4 | 0.13 |
| polarC18 | 325.216 | 70.3529 | 24.4 | 0.13 |
| polarC18 | 325.216 | 243.5468 | 24.4 | 0.13 |
| polarC18 | 325.216 | 311.5023 | 24.4 | 0.13 |
| polarC18 | 325.216 | 133.9014 | 24.3 | 0.13 |
| polarC18 | 325.216 | 100.7636 | 24.3 | 0.13 |
| polarC18 | 325.216 | 336.7298 | 24.3 | 0.13 |
| polarC18 | 325.216 | 325.9589 | 24.3 | 0.13 |
| polarC18 | 325.216 | 59.2332 | 24.3 | 0.13 |
| polarC18 | 325.216 | 129.209 | 24.3 | 0.13 |
| polarC18 | 325.216 | 151.8124 | 24.3 | 0.13 |
| polarC18 | 325.216 | 96.9978 | 24.3 | 0.13 |
| polarC18 | 325.216 | 87.6904 | 24.3 | 0.13 |
| polarC18 | 325.216 | 130.1137 | 24.3 | 0.12 |
| polarC18 | 325.216 | 94.6142 | 24.3 | 0.12 |
| polarC18 | 325.216 | 278.1895 | 24.3 | 0.12 |
| polarC18 | 325.216 | 92.4128 | 24.3 | 0.12 |
| polarC18 | 325.216 | 53.5904 | 24.3 | 0.12 |
| polarC18 | 325.216 | 146.9975 | 24.3 | 0.12 |
| polarC18 | 325.216 | 98.4287 | 24.3 | 0.12 |
| polarC18 | 325.216 | 141.9393 | 24.3 | 0.12 |
| polarC18 | 325.216 | 94.8543 | 24.3 | 0.12 |
| polarC18 | 325.216 | 238.5323 | 24.3 | 0.12 |
| polarC18 | 325.216 | 57.6323 | 24.3 | 0.12 |
| polarC18 | 325.216 | 110.3803 | 24.2 | 0.12 |
| polarC18 | 325.216 | 238.9087 | 24.2 | 0.12 |
| polarC18 | 325.216 | 113.8315 | 24.2 | 0.12 |
| polarC18 | 325.216 | 276.5129 | 24.2 | 0.12 |
| polarC18 | 325.216 | 233.7949 | 24.2 | 0.12 |
| polarC18 | 325.216 | 336.3105 | 24.2 | 0.12 |
| polarC18 | 325.216 | 50.9474 | 24.2 | 0.12 |
| polarC18 | 325.216 | 113.0223 | 24.2 | 0.12 |
| polarC18 | 325.216 | 151.8292 | 24.2 | 0.12 |
| polarC18 | 325.216 | 197.9594 | 24.2 | 0.12 |
| polarC18 | 325.216 | 65.2727 | 24.2 | 0.12 |
| polarC18 | 325.216 | 72.4818 | 24.2 | 0.12 |
| polarC18 | 325.216 | 111.5993 | 24.2 | 0.12 |
| polarC18 | 325.216 | 95.577 | 24.2 | 0.12 |
| polarC18 | 325.216 | 96.1175 | 24.2 | 0.12 |
| polarC18 | 325.216 | 341.8145 | 24.2 | 0.12 |
| polarC18 | 325.216 | 72.6841 | 24.2 | 0.12 |

| | | | | |
|---|---|---|---|---|
| polarC18 | 325.216 | 146.2155 | 24.2 | 0.12 |
| polarC18 | 325.216 | 194.9964 | 24.2 | 0.12 |
| polarC18 | 325.216 | 213.566 | 24.2 | 0.12 |
| polarC18 | 325.216 | 119.7794 | 24.2 | 0.12 |
| polarC18 | 325.216 | 90.5085 | 24.2 | 0.12 |
| polarC18 | 325.216 | 172.7788 | 24.2 | 0.12 |
| polarC18 | 325.216 | 138.005 | 24.2 | 0.12 |
| polarC18 | 325.216 | 86.1971 | 24.1 | 0.12 |
| polarC18 | 325.216 | 303.5843 | 24.1 | 0.12 |
| polarC18 | 325.216 | 305.2017 | 24.1 | 0.12 |
| polarC18 | 325.216 | 208.8295 | 24.1 | 0.12 |
| polarC18 | 325.216 | 93.393 | 24.1 | 0.12 |
| polarC18 | 325.216 | 54.0689 | 24.1 | 0.12 |
| polarC18 | 325.216 | 342.6049 | 24.1 | 0.12 |
| polarC18 | 325.216 | 142.4581 | 24.1 | 0.12 |
| polarC18 | 325.216 | 183.6432 | 24.1 | 0.12 |
| polarC18 | 325.216 | 89.0144 | 24.1 | 0.12 |
| polarC18 | 325.216 | 158.2317 | 24.1 | 0.12 |
| polarC18 | 325.216 | 61.9524 | 24.1 | 0.12 |
| polarC18 | 325.216 | 55.8454 | 24.1 | 0.12 |
| polarC18 | 325.216 | 241.8884 | 24.1 | 0.12 |
| polarC18 | 325.216 | 242.1874 | 24.1 | 0.12 |
| polarC18 | 325.216 | 53.9513 | 24.1 | 0.12 |
| polarC18 | 325.216 | 59.1519 | 24.1 | 0.12 |
| polarC18 | 325.216 | 186.9916 | 24.1 | 0.12 |
| polarC18 | 325.216 | 274.1073 | 24.1 | 0.12 |
| polarC18 | 325.216 | 60.5452 | 24.1 | 0.12 |
| polarC18 | 325.216 | 98.2135 | 24.1 | 0.12 |
| polarC18 | 325.216 | 80.388 | 24.1 | 0.12 |
| polarC18 | 325.216 | 182.3129 | 24.1 | 0.12 |
| polarC18 | 325.216 | 149.6538 | 24.1 | 0.12 |
| polarC18 | 325.216 | 55.0563 | 24.1 | 0.12 |
| polarC18 | 325.216 | 185.7844 | 24.1 | 0.12 |
| polarC18 | 325.216 | 52.4201 | 24.1 | 0.12 |
| polarC18 | 325.216 | 233.3647 | 24 | 0.12 |
| polarC18 | 325.216 | 200.7637 | 24 | 0.12 |
| polarC18 | 325.216 | 226.9557 | 24 | 0.12 |
| polarC18 | 325.216 | 65.044 | 24 | 0.12 |
| polarC18 | 325.216 | 76.8978 | 24 | 0.12 |
| polarC18 | 325.216 | 344.1514 | 24 | 0.12 |
| polarC18 | 325.216 | 86.8509 | 24 | 0.12 |
| polarC18 | 325.216 | 211.5872 | 24 | 0.12 |

| | | | | |
|---|---|---|---|---|
| polarC18 | 325.216 | 152.9283 | 24 | 0.12 |
| polarC18 | 325.216 | 83.6966 | 24 | 0.12 |
| polarC18 | 325.216 | 196.2684 | 24 | 0.12 |
| polarC18 | 325.216 | 288.8533 | 24 | 0.12 |
| polarC18 | 325.216 | 144.3707 | 24 | 0.12 |
| polarC18 | 325.216 | 121.2906 | 24 | 0.12 |
| polarC18 | 325.216 | 320.6794 | 24 | 0.12 |
| polarC18 | 325.216 | 63.3094 | 24 | 0.12 |
| polarC18 | 325.216 | 125.1258 | 24 | 0.12 |
| polarC18 | 325.216 | 332.5987 | 24 | 0.12 |
| polarC18 | 325.216 | 299.6151 | 24 | 0.12 |
| polarC18 | 325.216 | 53.4883 | 24 | 0.12 |
| polarC18 | 325.216 | 63.9048 | 24 | 0.12 |
| polarC18 | 325.216 | 51.6076 | 24 | 0.12 |
| polarC18 | 325.216 | 101.5938 | 24 | 0.12 |
| polarC18 | 325.216 | 139.5774 | 24 | 0.12 |
| polarC18 | 325.216 | 316.4421 | 24 | 0.12 |
| polarC18 | 325.216 | 72.2447 | 24 | 0.12 |
| polarC18 | 325.216 | 115.7155 | 24 | 0.12 |
| polarC18 | 325.216 | 295.0154 | 24 | 0.12 |
| polarC18 | 325.216 | 96.1452 | 24 | 0.12 |
| polarC18 | 325.216 | 97.923 | 24 | 0.12 |
| polarC18 | 325.216 | 82.0392 | 24 | 0.12 |
| polarC18 | 325.216 | 165.5185 | 24 | 0.12 |
| polarC18 | 325.216 | 56.9835 | 24 | 0.12 |
| polarC18 | 325.216 | 185.2971 | 24 | 0.12 |
| polarC18 | 325.216 | 57.578 | 24 | 0.12 |
| polarC18 | 325.216 | 72.3206 | 24 | 0.12 |
| polarC18 | 325.216 | 81.6237 | 24 | 0.12 |
| polarC18 | 325.216 | 73.289 | 24 | 0.12 |
| polarC18 | 325.216 | 62.862 | 23.9 | 0.12 |
| polarC18 | 325.216 | 94.6777 | 23.9 | 0.12 |
| polarC18 | 325.216 | 98.7275 | 23.9 | 0.12 |
| polarC18 | 325.216 | 102.9038 | 23.9 | 0.12 |
| polarC18 | 325.216 | 332.1126 | 23.9 | 0.12 |
| polarC18 | 325.216 | 57.5204 | 23.9 | 0.12 |
| polarC18 | 325.216 | 64.3127 | 23.9 | 0.12 |
| polarC18 | 325.216 | 208.6684 | 23.9 | 0.12 |
| polarC18 | 325.216 | 129.929 | 23.9 | 0.12 |
| polarC18 | 325.216 | 279.3221 | 23.9 | 0.12 |
| polarC18 | 325.216 | 185.5746 | 23.9 | 0.12 |
| polarC18 | 325.216 | 53.2131 | 23.9 | 0.12 |

| | | | | |
|---|---|---|---|---|
| polarC18 | 325.216 | 108.8517 | 23.9 | 0.12 |
| polarC18 | 325.216 | 72.2831 | 23.9 | 0.12 |
| polarC18 | 325.216 | 84.5265 | 23.9 | 0.12 |
| polarC18 | 325.216 | 100.2688 | 23.9 | 0.12 |
| polarC18 | 325.216 | 94.241 | 23.9 | 0.12 |
| polarC18 | 325.216 | 100.7059 | 23.9 | 0.12 |
| polarC18 | 325.216 | 51.8505 | 23.9 | 0.12 |
| polarC18 | 325.216 | 105.7712 | 23.9 | 0.12 |
| polarC18 | 325.216 | 94.8826 | 23.9 | 0.12 |
| polarC18 | 325.216 | 97.5741 | 23.9 | 0.12 |
| polarC18 | 325.216 | 85.2752 | 23.9 | 0.12 |
| polarC18 | 325.216 | 79.6709 | 23.9 | 0.12 |
| polarC18 | 325.216 | 212.795 | 23.9 | 0.12 |
| polarC18 | 325.216 | 70.0599 | 23.9 | 0.12 |
| polarC18 | 325.216 | 105.3102 | 23.9 | 0.12 |
| polarC18 | 325.216 | 238.2165 | 23.9 | 0.12 |
| polarC18 | 325.216 | 261.2221 | 23.9 | 0.12 |
| polarC18 | 325.216 | 163.8143 | 23.9 | 0.12 |
| polarC18 | 325.216 | 62.575 | 23.9 | 0.12 |
| polarC18 | 325.216 | 56.0569 | 23.9 | 0.12 |
| polarC18 | 325.216 | 53.6175 | 23.9 | 0.12 |
| polarC18 | 325.216 | 97.0491 | 23.9 | 0.12 |
| polarC18 | 325.216 | 58.0526 | 23.9 | 0.12 |
| polarC18 | 325.216 | 51.7457 | 23.9 | 0.12 |
| polarC18 | 325.216 | 85.9158 | 23.8 | 0.12 |
| polarC18 | 325.216 | 274.695 | 23.8 | 0.12 |
| polarC18 | 325.216 | 146.3474 | 23.8 | 0.12 |
| polarC18 | 325.216 | 77.6183 | 23.8 | 0.12 |
| polarC18 | 325.216 | 156.124 | 23.8 | 0.12 |
| polarC18 | 325.216 | 325.5743 | 23.8 | 0.12 |
| polarC18 | 325.216 | 302.9929 | 23.8 | 0.12 |
| polarC18 | 325.216 | 342.8685 | 23.8 | 0.12 |
| polarC18 | 325.216 | 70.9517 | 23.8 | 0.12 |
| polarC18 | 325.216 | 87.4515 | 23.8 | 0.12 |
| polarC18 | 325.216 | 67.0967 | 23.8 | 0.12 |
| polarC18 | 325.216 | 79.1132 | 23.8 | 0.12 |
| polarC18 | 325.216 | 115.0282 | 23.8 | 0.12 |
| polarC18 | 325.216 | 181.4535 | 23.8 | 0.12 |
| polarC18 | 325.216 | 122.8762 | 23.8 | 0.12 |
| polarC18 | 325.216 | 50.6742 | 23.8 | 0.12 |
| polarC18 | 325.216 | 57.0611 | 23.8 | 0.12 |
| polarC18 | 325.216 | 102.8615 | 23.8 | 0.12 |

| | | | | |
|---|---|---|---|---|
| polarC18 | 325.216 | 61.0907 | 23.8 | 0.12 |
| polarC18 | 325.216 | 133.349 | 23.8 | 0.12 |
| polarC18 | 325.216 | 286.3571 | 23.8 | 0.12 |
| polarC18 | 325.216 | 86.9676 | 23.8 | 0.12 |
| polarC18 | 325.216 | 116.8694 | 23.8 | 0.12 |
| polarC18 | 325.216 | 144.8712 | 23.8 | 0.12 |
| polarC18 | 325.216 | 87.2816 | 23.8 | 0.12 |
| polarC18 | 325.216 | 123.2088 | 23.8 | 0.12 |
| polarC18 | 325.216 | 79.9327 | 23.8 | 0.12 |
| polarC18 | 325.216 | 202.4175 | 23.8 | 0.12 |
| polarC18 | 325.216 | 190.6177 | 23.7 | 0.12 |
| polarC18 | 325.216 | 274.3456 | 23.7 | 0.12 |
| polarC18 | 325.216 | 214.8771 | 23.7 | 0.12 |
| polarC18 | 325.216 | 247.3918 | 23.7 | 0.12 |
| polarC18 | 325.216 | 68.2227 | 23.7 | 0.12 |
| polarC18 | 325.216 | 78.3273 | 23.7 | 0.12 |
| polarC18 | 325.216 | 331.7555 | 23.7 | 0.12 |
| polarC18 | 325.216 | 277.8704 | 23.7 | 0.12 |
| polarC18 | 325.216 | 239.5887 | 23.7 | 0.12 |
| polarC18 | 325.216 | 168.707 | 23.7 | 0.12 |
| polarC18 | 325.216 | 76.5561 | 23.7 | 0.12 |
| polarC18 | 325.216 | 267.7386 | 23.7 | 0.12 |
| polarC18 | 325.216 | 72.5835 | 23.7 | 0.12 |
| polarC18 | 325.216 | 62.1956 | 23.7 | 0.12 |
| polarC18 | 325.216 | 50.6426 | 23.7 | 0.12 |
| polarC18 | 325.216 | 89.2179 | 23.7 | 0.12 |
| polarC18 | 325.216 | 120.8516 | 23.7 | 0.12 |
| polarC18 | 325.216 | 60.5639 | 23.7 | 0.12 |
| polarC18 | 325.216 | 59.8729 | 23.7 | 0.12 |
| polarC18 | 325.216 | 116.6953 | 23.7 | 0.12 |
| polarC18 | 325.216 | 148.2273 | 23.7 | 0.12 |
| polarC18 | 325.216 | 113.3475 | 23.7 | 0.12 |
| polarC18 | 325.216 | 58.1134 | 23.7 | 0.12 |
| polarC18 | 325.216 | 89.5034 | 23.7 | 0.12 |
| polarC18 | 325.216 | 54.8022 | 23.7 | 0.12 |
| polarC18 | 325.216 | 121.0437 | 23.7 | 0.12 |
| polarC18 | 325.216 | 217.656 | 23.7 | 0.12 |
| polarC18 | 325.216 | 65.5548 | 23.7 | 0.12 |
| polarC18 | 325.216 | 78.0643 | 23.7 | 0.12 |
| polarC18 | 325.216 | 55.9749 | 23.7 | 0.12 |
| polarC18 | 325.216 | 110.0008 | 23.7 | 0.12 |
| polarC18 | 325.216 | 109.0036 | 23.7 | 0.12 |

| | | | | |
|---|---|---|---|---|
| polarC18 | 325.216 | 52.9824 | 23.7 | 0.12 |
| polarC18 | 325.216 | 56.3478 | 23.7 | 0.12 |
| polarC18 | 325.216 | 125.3856 | 23.7 | 0.12 |
| polarC18 | 325.216 | 55.6551 | 23.6 | 0.12 |
| polarC18 | 325.216 | 107.1773 | 23.6 | 0.12 |
| polarC18 | 325.216 | 242.6715 | 23.6 | 0.12 |
| polarC18 | 325.216 | 52.0974 | 23.6 | 0.12 |
| polarC18 | 325.216 | 74.078 | 23.6 | 0.12 |
| polarC18 | 325.216 | 217.7184 | 23.6 | 0.12 |
| polarC18 | 325.216 | 52.3611 | 23.6 | 0.12 |
| polarC18 | 325.216 | 203.1523 | 23.6 | 0.12 |
| polarC18 | 325.216 | 102.4503 | 23.6 | 0.12 |
| polarC18 | 325.216 | 335.9477 | 23.6 | 0.12 |
| polarC18 | 325.216 | 55.1066 | 23.6 | 0.12 |
| polarC18 | 325.216 | 104.1667 | 23.6 | 0.12 |
| polarC18 | 325.216 | 69.8785 | 23.6 | 0.12 |
| polarC18 | 325.216 | 233.7306 | 23.6 | 0.12 |
| polarC18 | 325.216 | 157.9786 | 23.6 | 0.12 |
| polarC18 | 325.216 | 117.5828 | 23.6 | 0.12 |
| polarC18 | 325.216 | 85.5017 | 23.6 | 0.12 |
| polarC18 | 325.216 | 169.3049 | 23.6 | 0.12 |
| polarC18 | 325.216 | 96.4186 | 23.6 | 0.12 |
| polarC18 | 325.216 | 318.2136 | 23.6 | 0.12 |
| polarC18 | 325.216 | 239.3531 | 23.6 | 0.12 |
| polarC18 | 325.216 | 73.2302 | 23.6 | 0.12 |
| polarC18 | 325.216 | 83.9581 | 23.6 | 0.12 |
| polarC18 | 325.216 | 166.9666 | 23.6 | 0.12 |
| polarC18 | 325.216 | 340.3401 | 23.6 | 0.12 |
| polarC18 | 325.216 | 64.1024 | 23.6 | 0.12 |
| polarC18 | 325.216 | 149.4164 | 23.6 | 0.12 |
| polarC18 | 325.216 | 120.6459 | 23.6 | 0.12 |
| polarC18 | 325.216 | 298.5364 | 23.6 | 0.12 |
| polarC18 | 325.216 | 58.9208 | 23.6 | 0.12 |
| polarC18 | 325.216 | 85.4654 | 23.6 | 0.12 |
| polarC18 | 325.216 | 151.6534 | 23.6 | 0.12 |
| polarC18 | 325.216 | 197.9324 | 23.6 | 0.12 |
| polarC18 | 325.216 | 273.2006 | 23.6 | 0.12 |
| polarC18 | 325.216 | 191.592 | 23.6 | 0.12 |
| polarC18 | 325.216 | 129.0395 | 23.6 | 0.12 |
| polarC18 | 325.216 | 69.0907 | 23.6 | 0.12 |
| polarC18 | 325.216 | 77.5402 | 23.6 | 0.12 |
| polarC18 | 325.216 | 223.1952 | 23.6 | 0.12 |

| | | | | |
|---|---|---|---|---|
| polarC18 | 325.216 | 56.6961 | 23.5 | 0.12 |
| polarC18 | 325.216 | 170.1502 | 23.5 | 0.12 |
| polarC18 | 325.216 | 69.9756 | 23.5 | 0.12 |
| polarC18 | 325.216 | 101.1546 | 23.5 | 0.12 |
| polarC18 | 325.216 | 168.357 | 23.5 | 0.12 |
| polarC18 | 325.216 | 335.2079 | 23.5 | 0.12 |
| polarC18 | 325.216 | 120.7451 | 23.5 | 0.12 |
| polarC18 | 325.216 | 168.8312 | 23.5 | 0.12 |
| polarC18 | 325.216 | 237.5013 | 23.5 | 0.12 |
| polarC18 | 325.216 | 70.8716 | 23.5 | 0.12 |
| polarC18 | 325.216 | 71.4276 | 23.5 | 0.12 |
| polarC18 | 325.216 | 141.6924 | 23.5 | 0.12 |
| polarC18 | 325.216 | 72.3773 | 23.5 | 0.12 |
| polarC18 | 325.216 | 78.6248 | 23.5 | 0.12 |
| polarC18 | 325.216 | 180.7905 | 23.5 | 0.12 |
| polarC18 | 325.216 | 51.564 | 23.5 | 0.12 |
| polarC18 | 325.216 | 61.6861 | 23.5 | 0.12 |
| polarC18 | 325.216 | 104.9903 | 23.5 | 0.12 |
| polarC18 | 325.216 | 340.6334 | 23.5 | 0.12 |
| polarC18 | 325.216 | 85.4391 | 23.5 | 0.12 |
| polarC18 | 325.216 | 79.8217 | 23.5 | 0.12 |
| polarC18 | 325.216 | 51.6395 | 23.5 | 0.12 |
| polarC18 | 325.216 | 136.0836 | 23.5 | 0.12 |
| polarC18 | 325.216 | 76.1595 | 23.5 | 0.12 |
| polarC18 | 325.216 | 214.978 | 23.5 | 0.12 |
| polarC18 | 325.216 | 279.5099 | 23.5 | 0.12 |
| polarC18 | 325.216 | 87.6323 | 23.5 | 0.12 |
| polarC18 | 325.216 | 77.136 | 23.5 | 0.12 |
| polarC18 | 325.216 | 216.4005 | 23.5 | 0.12 |
| polarC18 | 325.216 | 189.1089 | 23.5 | 0.12 |
| polarC18 | 325.216 | 59.821 | 23.5 | 0.12 |
| polarC18 | 325.216 | 247.9319 | 23.5 | 0.12 |
| polarC18 | 325.216 | 165.2335 | 23.4 | 0.12 |
| polarC18 | 325.216 | 206.1052 | 23.4 | 0.12 |
| polarC18 | 325.216 | 104.5274 | 23.4 | 0.12 |
| polarC18 | 325.216 | 139.5137 | 23.4 | 0.12 |
| polarC18 | 325.216 | 92.9335 | 23.4 | 0.12 |
| polarC18 | 325.216 | 172.1966 | 23.4 | 0.12 |
| polarC18 | 325.216 | 135.9216 | 23.4 | 0.12 |
| polarC18 | 325.216 | 152.5113 | 23.4 | 0.12 |
| polarC18 | 325.216 | 157.8424 | 23.4 | 0.12 |
| polarC18 | 325.216 | 53.9548 | 23.4 | 0.12 |

| | | | | |
|---|---|---|---|---|
| polarC18 | 325.216 | 102.3101 | 23.4 | 0.12 |
| polarC18 | 325.216 | 181.3127 | 23.4 | 0.12 |
| polarC18 | 325.216 | 220.6827 | 23.4 | 0.12 |
| polarC18 | 325.216 | 94.6728 | 23.4 | 0.12 |
| polarC18 | 325.216 | 194.924 | 23.4 | 0.12 |
| polarC18 | 325.216 | 184.0766 | 23.4 | 0.12 |
| polarC18 | 325.216 | 115.7804 | 23.4 | 0.12 |
| polarC18 | 325.216 | 68.177 | 23.4 | 0.12 |
| polarC18 | 325.216 | 131.2618 | 23.4 | 0.12 |
| polarC18 | 325.216 | 53.7023 | 23.4 | 0.12 |
| polarC18 | 325.216 | 292.6724 | 23.4 | 0.12 |
| polarC18 | 325.216 | 66.9899 | 23.4 | 0.12 |
| polarC18 | 325.216 | 68.9137 | 23.4 | 0.12 |
| polarC18 | 325.216 | 99.2508 | 23.4 | 0.12 |
| polarC18 | 325.216 | 126.6815 | 23.4 | 0.12 |
| polarC18 | 325.216 | 164.7079 | 23.4 | 0.12 |
| polarC18 | 325.216 | 267.0439 | 23.4 | 0.12 |
| polarC18 | 325.216 | 213.193 | 23.4 | 0.12 |
| polarC18 | 325.216 | 57.6151 | 23.4 | 0.12 |
| polarC18 | 325.216 | 63.673 | 23.4 | 0.12 |
| polarC18 | 325.216 | 105.8353 | 23.4 | 0.12 |
| polarC18 | 325.216 | 169.5843 | 23.4 | 0.12 |
| polarC18 | 325.216 | 194.9548 | 23.4 | 0.12 |
| polarC18 | 325.216 | 205.1287 | 23.4 | 0.12 |
| polarC18 | 325.216 | 130.2452 | 23.4 | 0.12 |
| polarC18 | 325.216 | 53.4017 | 23.4 | 0.12 |
| polarC18 | 325.216 | 86.8631 | 23.4 | 0.12 |
| polarC18 | 325.216 | 76.516 | 23.4 | 0.12 |
| polarC18 | 325.216 | 275.4765 | 23.4 | 0.12 |
| polarC18 | 325.216 | 298.1888 | 23.4 | 0.12 |
| polarC18 | 325.216 | 89.6794 | 23.3 | 0.12 |
| polarC18 | 325.216 | 59.1078 | 23.3 | 0.12 |
| polarC18 | 325.216 | 114.6525 | 23.3 | 0.12 |
| polarC18 | 325.216 | 173.8668 | 23.3 | 0.12 |
| polarC18 | 325.216 | 131.4085 | 23.3 | 0.12 |
| polarC18 | 325.216 | 58.0358 | 23.3 | 0.12 |
| polarC18 | 325.216 | 82.0872 | 23.3 | 0.12 |
| polarC18 | 325.216 | 80.782 | 23.3 | 0.12 |
| polarC18 | 325.216 | 296.2518 | 23.3 | 0.12 |
| polarC18 | 325.216 | 76.0508 | 23.3 | 0.12 |
| polarC18 | 325.216 | 177.3121 | 23.3 | 0.12 |
| polarC18 | 325.216 | 63.006 | 23.3 | 0.12 |

| | | | | |
|---|---|---|---|---|
| polarC18 | 325.216 | 114.3745 | 23.3 | 0.12 |
| polarC18 | 325.216 | 109.2471 | 23.3 | 0.12 |
| polarC18 | 325.216 | 150.4795 | 23.3 | 0.12 |
| polarC18 | 325.216 | 72.4785 | 23.3 | 0.12 |
| polarC18 | 325.216 | 94.4546 | 23.3 | 0.12 |
| polarC18 | 325.216 | 59.4936 | 23.3 | 0.12 |
| polarC18 | 325.216 | 95.8802 | 23.3 | 0.12 |
| polarC18 | 325.216 | 109.0493 | 23.3 | 0.12 |
| polarC18 | 325.216 | 72.8506 | 23.3 | 0.12 |
| polarC18 | 325.216 | 76.0339 | 23.3 | 0.12 |
| polarC18 | 325.216 | 75.3244 | 23.3 | 0.12 |
| polarC18 | 325.216 | 50.3184 | 23.3 | 0.12 |
| polarC18 | 325.216 | 119.1387 | 23.3 | 0.12 |
| polarC18 | 325.216 | 85.0058 | 23.3 | 0.12 |
| polarC18 | 325.216 | 107.9197 | 23.3 | 0.12 |
| polarC18 | 325.216 | 95.2696 | 23.3 | 0.12 |
| polarC18 | 325.216 | 69.459 | 23.3 | 0.12 |
| polarC18 | 325.216 | 62.0993 | 23.3 | 0.12 |
| polarC18 | 325.216 | 74.4419 | 23.3 | 0.12 |
| polarC18 | 325.216 | 190.594 | 23.3 | 0.12 |
| polarC18 | 325.216 | 53.4607 | 23.3 | 0.12 |
| polarC18 | 325.216 | 166.3342 | 23.3 | 0.12 |
| polarC18 | 325.216 | 234.3688 | 23.3 | 0.12 |
| polarC18 | 325.216 | 72.4744 | 23.3 | 0.12 |
| polarC18 | 325.216 | 62.848 | 23.3 | 0.12 |
| polarC18 | 325.216 | 136.6325 | 23.3 | 0.12 |
| polarC18 | 325.216 | 98.9736 | 23.3 | 0.12 |
| polarC18 | 325.216 | 50.3004 | 23.3 | 0.12 |
| polarC18 | 325.216 | 311.185 | 23.3 | 0.12 |
| polarC18 | 325.216 | 60.4608 | 23.3 | 0.12 |
| polarC18 | 325.216 | 142.6159 | 23.3 | 0.12 |
| polarC18 | 325.216 | 80.6015 | 23.3 | 0.12 |
| polarC18 | 325.216 | 80.1673 | 23.3 | 0.12 |
| polarC18 | 325.216 | 66.6691 | 23.3 | 0.12 |
| polarC18 | 325.216 | 55.7999 | 23.3 | 0.12 |
| polarC18 | 325.216 | 93.9101 | 23.3 | 0.12 |
| polarC18 | 325.216 | 126.3143 | 23.2 | 0.12 |
| polarC18 | 325.216 | 54.2562 | 23.2 | 0.12 |
| polarC18 | 325.216 | 284.0519 | 23.2 | 0.12 |
| polarC18 | 325.216 | 168.4002 | 23.2 | 0.12 |
| polarC18 | 325.216 | 208.7206 | 23.2 | 0.12 |
| polarC18 | 325.216 | 80.4691 | 23.2 | 0.12 |

| | | | | |
|---|---|---|---|---|
| polarC18 | 325.216 | 165.5414 | 23.2 | 0.12 |
| polarC18 | 325.216 | 72.0941 | 23.2 | 0.12 |
| polarC18 | 325.216 | 71.589 | 23.2 | 0.12 |
| polarC18 | 325.216 | 209.6488 | 23.2 | 0.12 |
| polarC18 | 325.216 | 176.8552 | 23.2 | 0.12 |
| polarC18 | 325.216 | 220.7305 | 23.2 | 0.12 |
| polarC18 | 325.216 | 95.6708 | 23.2 | 0.12 |
| polarC18 | 325.216 | 51.9254 | 23.2 | 0.12 |
| polarC18 | 325.216 | 240.8945 | 23.2 | 0.12 |
| polarC18 | 325.216 | 56.9144 | 23.2 | 0.12 |
| polarC18 | 325.216 | 89.1118 | 23.2 | 0.12 |
| polarC18 | 325.216 | 133.7792 | 23.2 | 0.12 |
| polarC18 | 325.216 | 51.228 | 23.2 | 0.12 |
| polarC18 | 325.216 | 153.3332 | 23.2 | 0.12 |
| polarC18 | 325.216 | 230.1385 | 23.2 | 0.12 |
| polarC18 | 325.216 | 229.3181 | 23.2 | 0.12 |
| polarC18 | 325.216 | 102.1678 | 23.2 | 0.12 |
| polarC18 | 325.216 | 55.088 | 23.2 | 0.12 |
| polarC18 | 325.216 | 186.4547 | 23.2 | 0.12 |
| polarC18 | 325.216 | 60.1457 | 23.2 | 0.12 |
| polarC18 | 325.216 | 115.4289 | 23.2 | 0.12 |
| polarC18 | 325.216 | 283.0122 | 23.2 | 0.12 |
| polarC18 | 325.216 | 109.6219 | 23.2 | 0.12 |
| polarC18 | 325.216 | 96.2948 | 23.2 | 0.12 |
| polarC18 | 325.216 | 189.1871 | 23.2 | 0.12 |
| polarC18 | 325.216 | 64.6742 | 23.2 | 0.12 |
| polarC18 | 325.216 | 83.835 | 23.2 | 0.12 |
| polarC18 | 325.216 | 280.1362 | 23.2 | 0.12 |
| polarC18 | 325.216 | 98.2199 | 23.2 | 0.12 |
| polarC18 | 325.216 | 59.3024 | 23.2 | 0.12 |
| polarC18 | 325.216 | 106.0466 | 23.2 | 0.12 |
| polarC18 | 325.216 | 156.6315 | 23.2 | 0.12 |
| polarC18 | 325.216 | 78.5659 | 23.2 | 0.12 |
| polarC18 | 325.216 | 102.4599 | 23.2 | 0.12 |
| polarC18 | 325.216 | 116.1816 | 23.2 | 0.12 |
| polarC18 | 325.216 | 58.3965 | 23.2 | 0.12 |
| polarC18 | 325.216 | 78.5106 | 23.2 | 0.12 |
| polarC18 | 325.216 | 65.6744 | 23.2 | 0.12 |
| polarC18 | 325.216 | 264.5601 | 23.2 | 0.12 |
| polarC18 | 325.216 | 56.8323 | 23.2 | 0.12 |
| polarC18 | 325.216 | 68.9017 | 23.2 | 0.12 |
| polarC18 | 325.216 | 65.9099 | 23.2 | 0.12 |

| | | | | |
|---|---|---|---|---|
| polarC18 | 325.216 | 88.973 | 23.2 | 0.12 |
| polarC18 | 325.216 | 62.389 | 23.2 | 0.12 |
| polarC18 | 325.216 | 83.1554 | 23.2 | 0.12 |
| polarC18 | 325.216 | 319.6143 | 23.2 | 0.12 |
| polarC18 | 325.216 | 66.2044 | 23.2 | 0.12 |
| polarC18 | 325.216 | 69.0246 | 23.2 | 0.12 |
| polarC18 | 325.216 | 101.0617 | 23.2 | 0.12 |
| polarC18 | 325.216 | 61.7978 | 23.2 | 0.12 |
| polarC18 | 325.216 | 78.0871 | 23.2 | 0.12 |
| polarC18 | 325.216 | 186.7376 | 23.2 | 0.12 |
| polarC18 | 325.216 | 192.665 | 23.2 | 0.12 |
| polarC18 | 325.216 | 74.3591 | 23.1 | 0.12 |
| polarC18 | 325.216 | 54.7612 | 23.1 | 0.12 |
| polarC18 | 325.216 | 326.369 | 23.1 | 0.12 |
| polarC18 | 325.216 | 58.9518 | 23.1 | 0.12 |
| polarC18 | 325.216 | 188.0242 | 23.1 | 0.12 |
| polarC18 | 325.216 | 80.1201 | 23.1 | 0.12 |
| polarC18 | 325.216 | 87.5685 | 23.1 | 0.12 |
| polarC18 | 325.216 | 240.586 | 23.1 | 0.12 |
| polarC18 | 325.216 | 112.9418 | 23.1 | 0.12 |
| polarC18 | 325.216 | 93.7949 | 23.1 | 0.12 |
| polarC18 | 325.216 | 218.2198 | 23.1 | 0.12 |
| polarC18 | 325.216 | 91.9358 | 23.1 | 0.12 |
| polarC18 | 325.216 | 57.3582 | 23.1 | 0.12 |
| polarC18 | 325.216 | 57.5664 | 23.1 | 0.12 |
| polarC18 | 325.216 | 56.9422 | 23.1 | 0.12 |
| polarC18 | 325.216 | 103.7311 | 23.1 | 0.12 |
| polarC18 | 325.216 | 62.0488 | 23.1 | 0.12 |
| polarC18 | 325.216 | 63.7768 | 23.1 | 0.12 |
| polarC18 | 325.216 | 74.95 | 23.1 | 0.12 |
| polarC18 | 325.216 | 100.837 | 23.1 | 0.12 |
| polarC18 | 325.216 | 119.4719 | 23.1 | 0.12 |
| polarC18 | 325.216 | 175.0755 | 23.1 | 0.12 |
| polarC18 | 325.216 | 177.0003 | 23.1 | 0.12 |
| polarC18 | 325.216 | 210.907 | 23.1 | 0.12 |
| polarC18 | 325.216 | 172.6337 | 23.1 | 0.12 |
| polarC18 | 325.216 | 52.5037 | 23.1 | 0.12 |
| polarC18 | 325.216 | 181.1664 | 23.1 | 0.12 |
| polarC18 | 325.216 | 92.9964 | 23.1 | 0.12 |
| polarC18 | 325.216 | 309.3615 | 23.1 | 0.12 |
| polarC18 | 325.216 | 82.4709 | 23.1 | 0.12 |
| polarC18 | 325.216 | 247.2739 | 23.1 | 0.12 |

| | | | | |
|---|---|---|---|---|
| polarC18 | 325.216 | 64.7761 | 23.1 | 0.12 |
| polarC18 | 325.216 | 77.7486 | 23.1 | 0.12 |
| polarC18 | 325.216 | 76.0617 | 23 | 0.12 |
| polarC18 | 325.216 | 124.0375 | 23 | 0.12 |
| polarC18 | 325.216 | 124.5235 | 23 | 0.12 |
| polarC18 | 325.216 | 211.1106 | 23 | 0.12 |
| polarC18 | 325.216 | 261.6899 | 23 | 0.12 |
| polarC18 | 325.216 | 50.6355 | 23 | 0.12 |
| polarC18 | 325.216 | 112.3051 | 23 | 0.12 |
| polarC18 | 325.216 | 161.9983 | 23 | 0.12 |
| polarC18 | 325.216 | 154.0527 | 23 | 0.12 |
| polarC18 | 325.216 | 58.0705 | 23 | 0.12 |
| polarC18 | 325.216 | 212.6602 | 23 | 0.12 |
| polarC18 | 325.216 | 83.4907 | 23 | 0.12 |
| polarC18 | 325.216 | 77.6986 | 23 | 0.12 |
| polarC18 | 325.216 | 73.4894 | 23 | 0.12 |
| polarC18 | 325.216 | 96.1088 | 23 | 0.12 |
| polarC18 | 325.216 | 92.8511 | 23 | 0.12 |
| polarC18 | 325.216 | 54.3121 | 23 | 0.12 |
| polarC18 | 325.216 | 81.6628 | 23 | 0.12 |
| polarC18 | 325.216 | 90.3585 | 23 | 0.12 |
| polarC18 | 325.216 | 118.3396 | 23 | 0.12 |
| polarC18 | 325.216 | 89.1454 | 23 | 0.12 |
| polarC18 | 325.216 | 262.7086 | 23 | 0.12 |
| polarC18 | 325.216 | 104.2186 | 23 | 0.12 |
| polarC18 | 325.216 | 316.9103 | 23 | 0.12 |
| polarC18 | 325.216 | 69.772 | 23 | 0.12 |
| polarC18 | 325.216 | 112.6355 | 23 | 0.12 |
| polarC18 | 325.216 | 124.7641 | 23 | 0.12 |
| polarC18 | 325.216 | 114.318 | 23 | 0.12 |
| polarC18 | 325.216 | 68.2445 | 23 | 0.12 |
| polarC18 | 325.216 | 94.1322 | 23 | 0.12 |
| polarC18 | 325.216 | 290.9424 | 23 | 0.12 |
| polarC18 | 325.216 | 164.1374 | 23 | 0.12 |
| polarC18 | 325.216 | 136.0285 | 23 | 0.12 |
| polarC18 | 325.216 | 204.3348 | 23 | 0.12 |
| polarC18 | 325.216 | 143.4529 | 23 | 0.12 |
| polarC18 | 325.216 | 116.3541 | 23 | 0.12 |
| polarC18 | 325.216 | 149.5375 | 23 | 0.12 |
| polarC18 | 325.216 | 179.0281 | 23 | 0.12 |
| polarC18 | 325.216 | 99.1644 | 23 | 0.12 |
| polarC18 | 325.216 | 53.4273 | 23 | 0.12 |

| | | | | |
|---|---|---|---|---|
| polarC18 | 325.216 | 173.813 | 23 | 0.12 |
| polarC18 | 325.216 | 61.8092 | 23 | 0.12 |
| polarC18 | 325.216 | 107.3372 | 23 | 0.12 |
| polarC18 | 325.216 | 61.5689 | 23 | 0.12 |
| polarC18 | 325.216 | 122.2028 | 23 | 0.12 |
| polarC18 | 325.216 | 105.0345 | 23 | 0.12 |
| polarC18 | 325.216 | 177.0574 | 23 | 0.12 |
| polarC18 | 325.216 | 205.9347 | 23 | 0.12 |
| polarC18 | 325.216 | 51.1653 | 23 | 0.12 |
| polarC18 | 325.216 | 79.4338 | 23 | 0.12 |
| polarC18 | 325.216 | 72.1956 | 23 | 0.12 |
| polarC18 | 325.216 | 90.4515 | 23 | 0.12 |
| polarC18 | 325.216 | 52.1857 | 23 | 0.12 |
| polarC18 | 325.216 | 171.8992 | 23 | 0.12 |
| polarC18 | 325.216 | 79.189 | 23 | 0.12 |
| polarC18 | 325.216 | 51.574 | 22.9 | 0.12 |
| polarC18 | 325.216 | 289.5056 | 22.9 | 0.12 |
| polarC18 | 325.216 | 51.2264 | 22.9 | 0.12 |
| polarC18 | 325.216 | 186.3848 | 22.9 | 0.12 |
| polarC18 | 325.216 | 211.144 | 22.9 | 0.12 |
| polarC18 | 325.216 | 132.1846 | 22.9 | 0.12 |
| polarC18 | 325.216 | 102.5932 | 22.9 | 0.12 |
| polarC18 | 325.216 | 170.3371 | 22.9 | 0.12 |
| polarC18 | 325.216 | 261.2733 | 22.9 | 0.12 |
| polarC18 | 325.216 | 146.3848 | 22.9 | 0.12 |
| polarC18 | 325.216 | 123.1618 | 22.9 | 0.12 |
| polarC18 | 325.216 | 329.6143 | 22.9 | 0.12 |
| polarC18 | 325.216 | 163.1435 | 22.9 | 0.12 |
| polarC18 | 325.216 | 75.5354 | 22.9 | 0.12 |
| polarC18 | 325.216 | 52.0289 | 22.9 | 0.12 |
| polarC18 | 325.216 | 103.6516 | 22.9 | 0.12 |
| polarC18 | 325.216 | 176.928 | 22.9 | 0.12 |
| polarC18 | 325.216 | 194.7187 | 22.9 | 0.12 |
| polarC18 | 325.216 | 106.3623 | 22.9 | 0.12 |
| polarC18 | 325.216 | 53.5502 | 22.9 | 0.12 |
| polarC18 | 325.216 | 56.5564 | 22.9 | 0.12 |
| polarC18 | 325.216 | 181.8485 | 22.9 | 0.12 |
| polarC18 | 325.216 | 281.2294 | 22.9 | 0.12 |
| polarC18 | 325.216 | 107.9752 | 22.9 | 0.12 |
| polarC18 | 325.216 | 59.0641 | 22.9 | 0.12 |
| polarC18 | 325.216 | 72.5735 | 22.9 | 0.12 |
| polarC18 | 325.216 | 100.2872 | 22.9 | 0.12 |

| | | | | |
|---|---|---|---|---|
| polarC18 | 325.216 | 149.9923 | 22.9 | 0.12 |
| polarC18 | 325.216 | 309.1585 | 22.9 | 0.12 |
| polarC18 | 325.216 | 113.791 | 22.9 | 0.12 |
| polarC18 | 325.216 | 134.6147 | 22.9 | 0.12 |
| polarC18 | 325.216 | 59.0086 | 22.9 | 0.12 |
| polarC18 | 325.216 | 51.6266 | 22.9 | 0.12 |
| polarC18 | 325.216 | 242.2391 | 22.9 | 0.12 |
| polarC18 | 325.216 | 75.4259 | 22.9 | 0.12 |
| polarC18 | 325.216 | 66.7678 | 22.9 | 0.12 |
| polarC18 | 325.216 | 51.503 | 22.9 | 0.12 |
| polarC18 | 325.216 | 66.6166 | 22.9 | 0.12 |
| polarC18 | 325.216 | 57.4362 | 22.9 | 0.12 |
| polarC18 | 325.216 | 52.0838 | 22.9 | 0.12 |
| polarC18 | 325.216 | 136.0434 | 22.9 | 0.12 |
| polarC18 | 325.216 | 292.1871 | 22.9 | 0.12 |
| polarC18 | 325.216 | 184.4542 | 22.9 | 0.12 |
| polarC18 | 325.216 | 218.7559 | 22.9 | 0.12 |
| polarC18 | 325.216 | 74.1467 | 22.9 | 0.12 |
| polarC18 | 325.216 | 103.5959 | 22.9 | 0.12 |
| polarC18 | 325.216 | 171.1165 | 22.9 | 0.12 |
| polarC18 | 325.216 | 271.7867 | 22.9 | 0.12 |
| polarC18 | 325.216 | 234.5879 | 22.9 | 0.12 |
| polarC18 | 325.216 | 127.7785 | 22.8 | 0.12 |
| polarC18 | 325.216 | 96.1616 | 22.8 | 0.12 |
| polarC18 | 325.216 | 52.839 | 22.8 | 0.12 |
| polarC18 | 325.216 | 62.9194 | 22.8 | 0.12 |
| polarC18 | 325.216 | 78.9952 | 22.8 | 0.12 |
| polarC18 | 325.216 | 297.602 | 22.8 | 0.12 |
| polarC18 | 325.216 | 172.6048 | 22.8 | 0.12 |
| polarC18 | 325.216 | 305.3569 | 22.8 | 0.12 |
| polarC18 | 325.216 | 206.9354 | 22.8 | 0.12 |
| polarC18 | 325.216 | 50.7638 | 22.8 | 0.12 |
| polarC18 | 325.216 | 173.4216 | 22.8 | 0.12 |
| polarC18 | 325.216 | 57.178 | 22.8 | 0.12 |
| polarC18 | 325.216 | 67.3902 | 22.8 | 0.12 |
| polarC18 | 325.216 | 89.9651 | 22.8 | 0.12 |
| polarC18 | 325.216 | 154.2123 | 22.8 | 0.12 |
| polarC18 | 325.216 | 65.2162 | 22.8 | 0.12 |
| polarC18 | 325.216 | 100.1731 | 22.8 | 0.12 |
| polarC18 | 325.216 | 268.3981 | 22.8 | 0.12 |
| polarC18 | 325.216 | 52.0527 | 22.8 | 0.12 |
| polarC18 | 325.216 | 96.8102 | 22.8 | 0.12 |

| | | | | |
|---|---|---|---|---|
| polarC18 | 325.216 | 125.3012 | 22.8 | 0.12 |
| polarC18 | 325.216 | 69.0199 | 22.8 | 0.12 |
| polarC18 | 325.216 | 133.9124 | 22.8 | 0.12 |
| polarC18 | 325.216 | 51.054 | 22.8 | 0.12 |
| polarC18 | 325.216 | 86.3639 | 22.8 | 0.12 |
| polarC18 | 325.216 | 54.5854 | 22.8 | 0.12 |
| polarC18 | 325.216 | 224.4245 | 22.8 | 0.12 |
| polarC18 | 325.216 | 68.6502 | 22.8 | 0.12 |
| polarC18 | 325.216 | 82.1973 | 22.8 | 0.12 |
| polarC18 | 325.216 | 101.7739 | 22.8 | 0.12 |
| polarC18 | 325.216 | 104.8899 | 22.8 | 0.12 |
| polarC18 | 325.216 | 93.2187 | 22.8 | 0.12 |
| polarC18 | 325.216 | 170.2646 | 22.8 | 0.12 |
| polarC18 | 325.216 | 112.1745 | 22.8 | 0.12 |
| polarC18 | 325.216 | 335.0378 | 22.8 | 0.12 |
| polarC18 | 325.216 | 109.7756 | 22.8 | 0.12 |
| polarC18 | 325.216 | 98.5146 | 22.8 | 0.12 |
| polarC18 | 325.216 | 69.1673 | 22.8 | 0.12 |
| polarC18 | 325.216 | 52.5531 | 22.8 | 0.12 |
| polarC18 | 325.216 | 96.5119 | 22.8 | 0.12 |
| polarC18 | 325.216 | 57.2432 | 22.8 | 0.12 |
| polarC18 | 325.216 | 84.4441 | 22.8 | 0.12 |
| polarC18 | 325.216 | 63.423 | 22.8 | 0.12 |
| polarC18 | 325.216 | 52.037 | 22.8 | 0.12 |
| polarC18 | 325.216 | 192.8305 | 22.8 | 0.12 |
| polarC18 | 325.216 | 78.2955 | 22.7 | 0.12 |
| polarC18 | 325.216 | 85.0938 | 22.7 | 0.12 |
| polarC18 | 325.216 | 195.6724 | 22.7 | 0.12 |
| polarC18 | 325.216 | 181.6892 | 22.7 | 0.12 |
| polarC18 | 325.216 | 194.1372 | 22.7 | 0.12 |
| polarC18 | 325.216 | 182.442 | 22.7 | 0.12 |
| polarC18 | 325.216 | 227.1071 | 22.7 | 0.12 |
| polarC18 | 325.216 | 75.4598 | 22.7 | 0.12 |
| polarC18 | 325.216 | 64.282 | 22.7 | 0.12 |
| polarC18 | 325.216 | 61.9303 | 22.7 | 0.12 |
| polarC18 | 325.216 | 287.705 | 22.7 | 0.12 |
| polarC18 | 325.216 | 120.4052 | 22.7 | 0.12 |
| polarC18 | 325.216 | 78.6899 | 22.7 | 0.12 |
| polarC18 | 325.216 | 326.4765 | 22.7 | 0.12 |
| polarC18 | 325.216 | 241.1648 | 22.7 | 0.12 |
| polarC18 | 325.216 | 77.7768 | 22.7 | 0.12 |
| polarC18 | 325.216 | 266.2124 | 22.7 | 0.12 |

| | | | | |
|---|---|---|---|---|
| polarC18 | 325.216 | 234.7584 | 22.7 | 0.12 |
| polarC18 | 325.216 | 334.2725 | 22.7 | 0.12 |
| polarC18 | 325.216 | 93.9669 | 22.7 | 0.12 |
| polarC18 | 325.216 | 112.0083 | 22.7 | 0.12 |
| polarC18 | 325.216 | 83.2036 | 22.7 | 0.12 |
| polarC18 | 325.216 | 315.0015 | 22.7 | 0.12 |
| polarC18 | 325.216 | 107.5692 | 22.7 | 0.12 |
| polarC18 | 325.216 | 348.8194 | 22.7 | 0.12 |
| polarC18 | 325.216 | 56.4943 | 22.7 | 0.12 |
| polarC18 | 325.216 | 289.9037 | 22.7 | 0.12 |
| polarC18 | 325.216 | 114.0185 | 22.7 | 0.12 |
| polarC18 | 325.216 | 273.8139 | 22.7 | 0.12 |
| polarC18 | 325.216 | 57.0042 | 22.7 | 0.12 |
| polarC18 | 325.216 | 224.2842 | 22.7 | 0.12 |
| polarC18 | 325.216 | 284.8701 | 22.7 | 0.12 |
| polarC18 | 325.216 | 55.1101 | 22.7 | 0.12 |
| polarC18 | 325.216 | 67.6523 | 22.7 | 0.12 |
| polarC18 | 325.216 | 62.1202 | 22.7 | 0.12 |
| polarC18 | 325.216 | 98.5254 | 22.7 | 0.12 |
| polarC18 | 325.216 | 95.4132 | 22.7 | 0.12 |
| polarC18 | 325.216 | 261.0988 | 22.7 | 0.12 |
| polarC18 | 325.216 | 100.3052 | 22.7 | 0.12 |
| polarC18 | 325.216 | 98.5207 | 22.7 | 0.12 |
| polarC18 | 325.216 | 116.8522 | 22.7 | 0.12 |
| polarC18 | 325.216 | 227.4866 | 22.7 | 0.12 |
| polarC18 | 325.216 | 91.4941 | 22.6 | 0.12 |
| polarC18 | 325.216 | 288.3594 | 22.6 | 0.12 |
| polarC18 | 325.216 | 73.5433 | 22.6 | 0.12 |
| polarC18 | 325.216 | 298.1267 | 22.6 | 0.12 |
| polarC18 | 325.216 | 83.4213 | 22.6 | 0.12 |
| polarC18 | 325.216 | 53.0399 | 22.6 | 0.12 |
| polarC18 | 325.216 | 73.513 | 22.6 | 0.12 |
| polarC18 | 325.216 | 63.8622 | 22.6 | 0.12 |
| polarC18 | 325.216 | 50.408 | 22.6 | 0.12 |
| polarC18 | 325.216 | 243.6279 | 22.6 | 0.12 |
| polarC18 | 325.216 | 74.7093 | 22.6 | 0.12 |
| polarC18 | 325.216 | 70.7243 | 22.6 | 0.12 |
| polarC18 | 325.216 | 51.1605 | 22.6 | 0.12 |
| polarC18 | 325.216 | 289.347 | 22.6 | 0.12 |
| polarC18 | 325.216 | 123.6838 | 22.6 | 0.12 |
| polarC18 | 325.216 | 63.6489 | 22.6 | 0.12 |
| polarC18 | 325.216 | 51.9037 | 22.6 | 0.12 |

| | | | | |
|---|---|---|---|---|
| polarC18 | 325.216 | 199.252 | 22.6 | 0.12 |
| polarC18 | 325.216 | 58.2349 | 22.6 | 0.12 |
| polarC18 | 325.216 | 76.2236 | 22.6 | 0.12 |
| polarC18 | 325.216 | 302.1748 | 22.6 | 0.12 |
| polarC18 | 325.216 | 283.9721 | 22.6 | 0.12 |
| polarC18 | 325.216 | 260.5032 | 22.6 | 0.12 |
| polarC18 | 325.216 | 56.308 | 22.6 | 0.12 |
| polarC18 | 325.216 | 133.0113 | 22.6 | 0.12 |
| polarC18 | 325.216 | 53.8447 | 22.6 | 0.12 |
| polarC18 | 325.216 | 146.2751 | 22.6 | 0.12 |
| polarC18 | 325.216 | 156.228 | 22.6 | 0.12 |
| polarC18 | 325.216 | 71.5161 | 22.6 | 0.12 |
| polarC18 | 325.216 | 179.6135 | 22.6 | 0.12 |
| polarC18 | 325.216 | 78.1271 | 22.6 | 0.12 |
| polarC18 | 325.216 | 199.3359 | 22.6 | 0.12 |
| polarC18 | 325.216 | 163.0629 | 22.6 | 0.12 |
| polarC18 | 325.216 | 58.1815 | 22.6 | 0.12 |
| polarC18 | 325.216 | 50.5773 | 22.6 | 0.12 |
| polarC18 | 325.216 | 135.5984 | 22.6 | 0.12 |
| polarC18 | 325.216 | 157.7931 | 22.6 | 0.12 |
| polarC18 | 325.216 | 70.7427 | 22.6 | 0.12 |
| polarC18 | 325.216 | 147.1905 | 22.6 | 0.12 |
| polarC18 | 325.216 | 53.7499 | 22.6 | 0.12 |
| polarC18 | 325.216 | 93.5039 | 22.6 | 0.12 |
| polarC18 | 325.216 | 182.399 | 22.6 | 0.12 |
| polarC18 | 325.216 | 253.7546 | 22.6 | 0.12 |
| polarC18 | 325.216 | 141.366 | 22.6 | 0.12 |
| polarC18 | 325.216 | 337.5773 | 22.6 | 0.12 |
| polarC18 | 325.216 | 137.0284 | 22.6 | 0.12 |
| polarC18 | 325.216 | 59.4554 | 22.6 | 0.12 |
| polarC18 | 325.216 | 266.8033 | 22.6 | 0.12 |
| polarC18 | 325.216 | 82.5155 | 22.6 | 0.12 |
| polarC18 | 325.216 | 75.9567 | 22.5 | 0.12 |
| polarC18 | 325.216 | 58.7509 | 22.5 | 0.12 |
| polarC18 | 325.216 | 287.2094 | 22.5 | 0.12 |
| polarC18 | 325.216 | 51.2902 | 22.5 | 0.12 |
| polarC18 | 325.216 | 249.3899 | 22.5 | 0.12 |
| polarC18 | 325.216 | 172.5441 | 22.5 | 0.12 |
| polarC18 | 325.216 | 111.0678 | 22.5 | 0.12 |
| polarC18 | 325.216 | 152.7355 | 22.5 | 0.12 |
| polarC18 | 325.216 | 347.4762 | 22.5 | 0.12 |
| polarC18 | 325.216 | 229.7601 | 22.5 | 0.12 |

| | | | | |
|---|---|---|---|---|
| polarC18 | 325.216 | 104.359 | 22.5 | 0.12 |
| polarC18 | 325.216 | 178.584 | 22.5 | 0.12 |
| polarC18 | 325.216 | 155.6334 | 22.5 | 0.12 |
| polarC18 | 325.216 | 121.3463 | 22.5 | 0.12 |
| polarC18 | 325.216 | 53.5683 | 22.5 | 0.12 |
| polarC18 | 325.216 | 212.3461 | 22.5 | 0.12 |
| polarC18 | 325.216 | 79.9886 | 22.5 | 0.12 |
| polarC18 | 325.216 | 201.9586 | 22.5 | 0.12 |
| polarC18 | 325.216 | 137.1014 | 22.5 | 0.12 |
| polarC18 | 325.216 | 156.4835 | 22.5 | 0.12 |
| polarC18 | 325.216 | 116.8446 | 22.5 | 0.12 |
| polarC18 | 325.216 | 137.3418 | 22.5 | 0.12 |
| polarC18 | 325.216 | 240.6387 | 22.5 | 0.12 |
| polarC18 | 325.216 | 325.4578 | 22.5 | 0.12 |
| polarC18 | 325.216 | 273.7694 | 22.5 | 0.12 |
| polarC18 | 325.216 | 139.6709 | 22.5 | 0.12 |
| polarC18 | 325.216 | 326.9705 | 22.5 | 0.12 |
| polarC18 | 325.216 | 55.7277 | 22.5 | 0.12 |
| polarC18 | 325.216 | 175.6046 | 22.5 | 0.12 |
| polarC18 | 325.216 | 267.4258 | 22.5 | 0.12 |
| polarC18 | 325.216 | 64.1424 | 22.5 | 0.12 |
| polarC18 | 325.216 | 80.8129 | 22.5 | 0.12 |
| polarC18 | 325.216 | 93.976 | 22.5 | 0.12 |
| polarC18 | 325.216 | 57.7024 | 22.5 | 0.12 |
| polarC18 | 325.216 | 325.0058 | 22.5 | 0.12 |
| polarC18 | 325.216 | 228.5327 | 22.5 | 0.12 |
| polarC18 | 325.216 | 94.9322 | 22.5 | 0.12 |
| polarC18 | 325.216 | 142.1422 | 22.5 | 0.12 |
| polarC18 | 325.216 | 86.5149 | 22.5 | 0.12 |
| polarC18 | 325.216 | 71.7089 | 22.5 | 0.12 |
| polarC18 | 325.216 | 223.0756 | 22.5 | 0.12 |
| polarC18 | 325.216 | 91.9755 | 22.5 | 0.12 |
| polarC18 | 325.216 | 56.1048 | 22.5 | 0.12 |
| polarC18 | 325.216 | 144.2329 | 22.5 | 0.12 |
| polarC18 | 325.216 | 324.8108 | 22.5 | 0.12 |
| polarC18 | 325.216 | 179.143 | 22.5 | 0.12 |
| polarC18 | 325.216 | 162.4849 | 22.5 | 0.12 |
| polarC18 | 325.216 | 86.9403 | 22.5 | 0.12 |
| polarC18 | 325.216 | 77.0555 | 22.5 | 0.12 |
| polarC18 | 325.216 | 51.5424 | 22.5 | 0.12 |
| polarC18 | 325.216 | 113.6919 | 22.5 | 0.12 |
| polarC18 | 325.216 | 100.4685 | 22.4 | 0.12 |

| | | | | |
|---|---|---|---|---|
| polarC18 | 325.216 | 80.4407 | 22.4 | 0.12 |
| polarC18 | 325.216 | 104.3952 | 22.4 | 0.12 |
| polarC18 | 325.216 | 149.9595 | 22.4 | 0.12 |
| polarC18 | 325.216 | 54.1352 | 22.4 | 0.12 |
| polarC18 | 325.216 | 176.8864 | 22.4 | 0.12 |
| polarC18 | 325.216 | 63.7716 | 22.4 | 0.12 |
| polarC18 | 325.216 | 81.2675 | 22.4 | 0.12 |
| polarC18 | 325.216 | 53.58 | 22.4 | 0.12 |
| polarC18 | 325.216 | 63.8836 | 22.4 | 0.12 |
| polarC18 | 325.216 | 223.0471 | 22.4 | 0.12 |
| polarC18 | 325.216 | 55.6963 | 22.4 | 0.12 |
| polarC18 | 325.216 | 122.7341 | 22.4 | 0.12 |
| polarC18 | 325.216 | 83.0809 | 22.4 | 0.12 |
| polarC18 | 325.216 | 84.5837 | 22.4 | 0.12 |
| polarC18 | 325.216 | 265.2299 | 22.4 | 0.12 |
| polarC18 | 325.216 | 135.6081 | 22.4 | 0.12 |
| polarC18 | 325.216 | 116.1259 | 22.4 | 0.12 |
| polarC18 | 325.216 | 152.607 | 22.4 | 0.12 |
| polarC18 | 325.216 | 71.32 | 22.4 | 0.12 |
| polarC18 | 325.216 | 62.1018 | 22.4 | 0.11 |
| polarC18 | 325.216 | 309.7198 | 22.4 | 0.11 |
| polarC18 | 325.216 | 307.5406 | 22.4 | 0.11 |
| polarC18 | 325.216 | 114.9138 | 22.4 | 0.11 |
| polarC18 | 325.216 | 84.0437 | 22.4 | 0.11 |
| polarC18 | 325.216 | 159.7959 | 22.4 | 0.11 |
| polarC18 | 325.216 | 157.2572 | 22.4 | 0.11 |
| polarC18 | 325.216 | 85.884 | 22.4 | 0.11 |
| polarC18 | 325.216 | 173.6675 | 22.4 | 0.11 |
| polarC18 | 325.216 | 54.1552 | 22.4 | 0.11 |
| polarC18 | 325.216 | 57.4696 | 22.4 | 0.11 |
| polarC18 | 325.216 | 96.5865 | 22.4 | 0.11 |
| polarC18 | 325.216 | 84.7173 | 22.4 | 0.11 |
| polarC18 | 325.216 | 106.2878 | 22.4 | 0.11 |
| polarC18 | 325.216 | 187.1363 | 22.4 | 0.11 |
| polarC18 | 325.216 | 192.2023 | 22.4 | 0.11 |
| polarC18 | 325.216 | 56.1194 | 22.3 | 0.11 |
| polarC18 | 325.216 | 243.7021 | 22.3 | 0.11 |
| polarC18 | 325.216 | 113.6589 | 22.3 | 0.11 |
| polarC18 | 325.216 | 335.4226 | 22.3 | 0.11 |
| polarC18 | 325.216 | 180.4017 | 22.3 | 0.11 |
| polarC18 | 325.216 | 54.0277 | 22.3 | 0.11 |
| polarC18 | 325.216 | 74.6039 | 22.3 | 0.11 |

| | | | | |
|---|---|---|---|---|
| polarC18 | 325.216 | 178.7909 | 22.3 | 0.11 |
| polarC18 | 325.216 | 97.2869 | 22.3 | 0.11 |
| polarC18 | 325.216 | 123.0782 | 22.3 | 0.11 |
| polarC18 | 325.216 | 62.2025 | 22.3 | 0.11 |
| polarC18 | 325.216 | 116.2826 | 22.3 | 0.11 |
| polarC18 | 325.216 | 64.8373 | 22.3 | 0.11 |
| polarC18 | 325.216 | 75.474 | 22.3 | 0.11 |
| polarC18 | 325.216 | 149.2965 | 22.3 | 0.11 |
| polarC18 | 325.216 | 142.1202 | 22.3 | 0.11 |
| polarC18 | 325.216 | 65.0522 | 22.3 | 0.11 |
| polarC18 | 325.216 | 109.8058 | 22.3 | 0.11 |
| polarC18 | 325.216 | 73.9778 | 22.3 | 0.11 |
| polarC18 | 325.216 | 94.9466 | 22.3 | 0.11 |
| polarC18 | 325.216 | 67.8343 | 22.3 | 0.11 |
| polarC18 | 325.216 | 71.5089 | 22.3 | 0.11 |
| polarC18 | 325.216 | 51.5411 | 22.3 | 0.11 |
| polarC18 | 325.216 | 149.7397 | 22.3 | 0.11 |
| polarC18 | 325.216 | 121.5647 | 22.3 | 0.11 |
| polarC18 | 325.216 | 272.1013 | 22.3 | 0.11 |
| polarC18 | 325.216 | 289.6671 | 22.3 | 0.11 |
| polarC18 | 325.216 | 145.5597 | 22.3 | 0.11 |
| polarC18 | 325.216 | 126.1272 | 22.3 | 0.11 |
| polarC18 | 325.216 | 136.7976 | 22.3 | 0.11 |
| polarC18 | 325.216 | 65.1818 | 22.3 | 0.11 |
| polarC18 | 325.216 | 57.4559 | 22.3 | 0.11 |
| polarC18 | 325.216 | 168.979 | 22.3 | 0.11 |
| polarC18 | 325.216 | 56.1311 | 22.3 | 0.11 |
| polarC18 | 325.216 | 121.491 | 22.3 | 0.11 |
| polarC18 | 325.216 | 80.7762 | 22.3 | 0.11 |
| polarC18 | 325.216 | 50.87 | 22.3 | 0.11 |
| polarC18 | 325.216 | 51.1376 | 22.3 | 0.11 |
| polarC18 | 325.216 | 84.6934 | 22.3 | 0.11 |
| polarC18 | 325.216 | 295.6575 | 22.3 | 0.11 |
| polarC18 | 325.216 | 133.8579 | 22.3 | 0.11 |
| polarC18 | 325.216 | 70.1925 | 22.3 | 0.11 |
| polarC18 | 325.216 | 57.549 | 22.3 | 0.11 |
| polarC18 | 325.216 | 80.8323 | 22.3 | 0.11 |
| polarC18 | 325.216 | 144.3261 | 22.3 | 0.11 |
| polarC18 | 325.216 | 63.1943 | 22.3 | 0.11 |
| polarC18 | 325.216 | 181.9122 | 22.3 | 0.11 |
| polarC18 | 325.216 | 170.1983 | 22.3 | 0.11 |
| polarC18 | 325.216 | 235.0253 | 22.3 | 0.11 |

| | | | | |
|---|---|---|---|---|
| polarC18 | 325.216 | 151.7733 | 22.3 | 0.11 |
| polarC18 | 325.216 | 55.2824 | 22.3 | 0.11 |
| polarC18 | 325.216 | 77.4537 | 22.3 | 0.11 |
| polarC18 | 325.216 | 64.3304 | 22.3 | 0.11 |
| polarC18 | 325.216 | 79.0032 | 22.3 | 0.11 |
| polarC18 | 325.216 | 85.2235 | 22.3 | 0.11 |
| polarC18 | 325.216 | 273.0638 | 22.3 | 0.11 |
| polarC18 | 325.216 | 51.8128 | 22.2 | 0.11 |
| polarC18 | 325.216 | 284.9269 | 22.2 | 0.11 |
| polarC18 | 325.216 | 198.3444 | 22.2 | 0.11 |
| polarC18 | 325.216 | 80.2839 | 22.2 | 0.11 |
| polarC18 | 325.216 | 215.6311 | 22.2 | 0.11 |
| polarC18 | 325.216 | 152.854 | 22.2 | 0.11 |
| polarC18 | 325.216 | 51.7931 | 22.2 | 0.11 |
| polarC18 | 325.216 | 103.6945 | 22.2 | 0.11 |
| polarC18 | 325.216 | 297.931 | 22.2 | 0.11 |
| polarC18 | 325.216 | 58.7291 | 22.2 | 0.11 |
| polarC18 | 325.216 | 136.6221 | 22.2 | 0.11 |
| polarC18 | 325.216 | 138.5323 | 22.2 | 0.11 |
| polarC18 | 325.216 | 346.7792 | 22.2 | 0.11 |
| polarC18 | 325.216 | 60.4506 | 22.2 | 0.11 |
| polarC18 | 325.216 | 125.2236 | 22.2 | 0.11 |
| polarC18 | 325.216 | 140.4423 | 22.2 | 0.11 |
| polarC18 | 325.216 | 300.2893 | 22.2 | 0.11 |
| polarC18 | 325.216 | 110.367 | 22.2 | 0.11 |
| polarC18 | 325.216 | 98.9466 | 22.2 | 0.11 |
| polarC18 | 325.216 | 109.0415 | 22.2 | 0.11 |
| polarC18 | 325.216 | 207.0009 | 22.2 | 0.11 |
| polarC18 | 325.216 | 71.9076 | 22.2 | 0.11 |
| polarC18 | 325.216 | 50.852 | 22.2 | 0.11 |
| polarC18 | 325.216 | 71.174 | 22.2 | 0.11 |
| polarC18 | 325.216 | 232.3656 | 22.2 | 0.11 |
| polarC18 | 325.216 | 122.8507 | 22.2 | 0.11 |
| polarC18 | 325.216 | 177.9917 | 22.2 | 0.11 |
| polarC18 | 325.216 | 264.9309 | 22.2 | 0.11 |
| polarC18 | 325.216 | 74.1673 | 22.2 | 0.11 |
| polarC18 | 325.216 | 84.0148 | 22.2 | 0.11 |
| polarC18 | 325.216 | 86.2024 | 22.2 | 0.11 |
| polarC18 | 325.216 | 104.5735 | 22.2 | 0.11 |
| polarC18 | 325.216 | 60.6454 | 22.2 | 0.11 |
| polarC18 | 325.216 | 85.4276 | 22.2 | 0.11 |
| polarC18 | 325.216 | 93.4018 | 22.2 | 0.11 |

| | | | | |
|---|---|---|---|---|
| polarC18 | 325.216 | 300.2023 | 22.2 | 0.11 |
| polarC18 | 325.216 | 164.0077 | 22.2 | 0.11 |
| polarC18 | 325.216 | 230.8891 | 22.2 | 0.11 |
| polarC18 | 325.216 | 182.7586 | 22.2 | 0.11 |
| polarC18 | 325.216 | 134.5246 | 22.2 | 0.11 |
| polarC18 | 325.216 | 150.1366 | 22.2 | 0.11 |
| polarC18 | 325.216 | 55.7238 | 22.2 | 0.11 |
| polarC18 | 325.216 | 59.7313 | 22.2 | 0.11 |
| polarC18 | 325.216 | 106.4285 | 22.2 | 0.11 |
| polarC18 | 325.216 | 76.6073 | 22.2 | 0.11 |
| polarC18 | 325.216 | 155.6552 | 22.2 | 0.11 |
| polarC18 | 325.216 | 57.5891 | 22.2 | 0.11 |
| polarC18 | 325.216 | 71.9714 | 22.2 | 0.11 |
| polarC18 | 325.216 | 308.5931 | 22.2 | 0.11 |
| polarC18 | 325.216 | 142.7671 | 22.2 | 0.11 |
| polarC18 | 325.216 | 91.0846 | 22.1 | 0.11 |
| polarC18 | 325.216 | 156.2812 | 22.1 | 0.11 |
| polarC18 | 325.216 | 219.6189 | 22.1 | 0.11 |
| polarC18 | 325.216 | 81.7738 | 22.1 | 0.11 |
| polarC18 | 325.216 | 246.3309 | 22.1 | 0.11 |
| polarC18 | 325.216 | 169.6012 | 22.1 | 0.11 |
| polarC18 | 325.216 | 236.9408 | 22.1 | 0.11 |
| polarC18 | 325.216 | 100.8013 | 22.1 | 0.11 |
| polarC18 | 325.216 | 180.2079 | 22.1 | 0.11 |
| polarC18 | 325.216 | 121.5055 | 22.1 | 0.11 |
| polarC18 | 325.216 | 85.4587 | 22.1 | 0.11 |
| polarC18 | 325.216 | 245.7923 | 22.1 | 0.11 |
| polarC18 | 325.216 | 232.4253 | 22.1 | 0.11 |
| polarC18 | 325.216 | 229.8602 | 22.1 | 0.11 |
| polarC18 | 325.216 | 95.7384 | 22.1 | 0.11 |
| polarC18 | 325.216 | 60.5522 | 22.1 | 0.11 |
| polarC18 | 325.216 | 69.1495 | 22.1 | 0.11 |
| polarC18 | 325.216 | 75.4386 | 22.1 | 0.11 |
| polarC18 | 325.216 | 56.022 | 22.1 | 0.11 |
| polarC18 | 325.216 | 58.0822 | 22.1 | 0.11 |
| polarC18 | 325.216 | 50.5833 | 22.1 | 0.11 |
| polarC18 | 325.216 | 81.2203 | 22.1 | 0.11 |
| polarC18 | 325.216 | 99.0364 | 22.1 | 0.11 |
| polarC18 | 325.216 | 50.3596 | 22.1 | 0.11 |
| polarC18 | 325.216 | 64.9442 | 22.1 | 0.11 |
| polarC18 | 325.216 | 67.457 | 22.1 | 0.11 |
| polarC18 | 325.216 | 301.1665 | 22.1 | 0.11 |

| | | | | |
|---|---|---|---|---|
| polarC18 | 325.216 | 57.1836 | 22.1 | 0.11 |
| polarC18 | 325.216 | 145.9647 | 22.1 | 0.11 |
| polarC18 | 325.216 | 103.6765 | 22.1 | 0.11 |
| polarC18 | 325.216 | 54.3153 | 22.1 | 0.11 |
| polarC18 | 325.216 | 73.1864 | 22.1 | 0.11 |
| polarC18 | 325.216 | 86.9559 | 22.1 | 0.11 |
| polarC18 | 325.216 | 233.8704 | 22.1 | 0.11 |
| polarC18 | 325.216 | 65.3739 | 22.1 | 0.11 |
| polarC18 | 325.216 | 184.1861 | 22.1 | 0.11 |
| polarC18 | 325.216 | 61.052 | 22.1 | 0.11 |
| polarC18 | 325.216 | 70.5323 | 22.1 | 0.11 |
| polarC18 | 325.216 | 144.8122 | 22.1 | 0.11 |
| polarC18 | 325.216 | 62.743 | 22.1 | 0.11 |
| polarC18 | 325.216 | 212.3212 | 22.1 | 0.11 |
| polarC18 | 325.216 | 88.3968 | 22.1 | 0.11 |
| polarC18 | 325.216 | 237.1106 | 22.1 | 0.11 |
| polarC18 | 325.216 | 65.8801 | 22.1 | 0.11 |
| polarC18 | 325.216 | 144.4588 | 22.1 | 0.11 |
| polarC18 | 325.216 | 118.9569 | 22.1 | 0.11 |
| polarC18 | 325.216 | 52.9733 | 22.1 | 0.11 |
| polarC18 | 325.216 | 131.8987 | 22.1 | 0.11 |
| polarC18 | 325.216 | 185.8816 | 22 | 0.11 |
| polarC18 | 325.216 | 70.86 | 22 | 0.11 |
| polarC18 | 325.216 | 106.034 | 22 | 0.11 |
| polarC18 | 325.216 | 89.8135 | 22 | 0.11 |
| polarC18 | 325.216 | 173.3231 | 22 | 0.11 |
| polarC18 | 325.216 | 95.867 | 22 | 0.11 |
| polarC18 | 325.216 | 75.5283 | 22 | 0.11 |
| polarC18 | 325.216 | 90.4026 | 22 | 0.11 |
| polarC18 | 325.216 | 74.3134 | 22 | 0.11 |
| polarC18 | 325.216 | 59.6703 | 22 | 0.11 |
| polarC18 | 325.216 | 244.6587 | 22 | 0.11 |
| polarC18 | 325.216 | 95.7086 | 22 | 0.11 |
| polarC18 | 325.216 | 56.4866 | 22 | 0.11 |
| polarC18 | 325.216 | 335.5389 | 22 | 0.11 |
| polarC18 | 325.216 | 188.0058 | 22 | 0.11 |
| polarC18 | 325.216 | 71.9516 | 22 | 0.11 |
| polarC18 | 325.216 | 138.2571 | 22 | 0.11 |
| polarC18 | 325.216 | 142.9128 | 22 | 0.11 |
| polarC18 | 325.216 | 53.8252 | 22 | 0.11 |
| polarC18 | 325.216 | 72.4886 | 22 | 0.11 |
| polarC18 | 325.216 | 244.5085 | 22 | 0.11 |

| | | | | |
|---|---|---|---|---|
| polarC18 | 325.216 | 59.7657 | 22 | 0.11 |
| polarC18 | 325.216 | 219.0887 | 22 | 0.11 |
| polarC18 | 325.216 | 187.2583 | 22 | 0.11 |
| polarC18 | 325.216 | 95.1837 | 22 | 0.11 |
| polarC18 | 325.216 | 117.8313 | 22 | 0.11 |
| polarC18 | 325.216 | 121.4602 | 22 | 0.11 |
| polarC18 | 325.216 | 129.4772 | 22 | 0.11 |
| polarC18 | 325.216 | 94.3383 | 22 | 0.11 |
| polarC18 | 325.216 | 158.4705 | 22 | 0.11 |
| polarC18 | 325.216 | 163.2619 | 22 | 0.11 |
| polarC18 | 325.216 | 345.9666 | 22 | 0.11 |
| polarC18 | 325.216 | 72.3015 | 22 | 0.11 |
| polarC18 | 325.216 | 103.0863 | 22 | 0.11 |
| polarC18 | 325.216 | 52.2214 | 22 | 0.11 |
| polarC18 | 325.216 | 92.8376 | 22 | 0.11 |
| polarC18 | 325.216 | 68.6243 | 22 | 0.11 |
| polarC18 | 325.216 | 86.5726 | 22 | 0.11 |
| polarC18 | 325.216 | 131.2016 | 21.9 | 0.11 |
| polarC18 | 325.216 | 57.9142 | 21.9 | 0.11 |
| polarC18 | 325.216 | 67.8224 | 21.9 | 0.11 |
| polarC18 | 325.216 | 92.7806 | 21.9 | 0.11 |
| polarC18 | 325.216 | 61.6077 | 21.9 | 0.11 |
| polarC18 | 325.216 | 59.9797 | 21.9 | 0.11 |
| polarC18 | 325.216 | 332.7826 | 21.9 | 0.11 |
| polarC18 | 325.216 | 282.5969 | 21.9 | 0.11 |
| polarC18 | 325.216 | 294.4352 | 21.9 | 0.11 |
| polarC18 | 325.216 | 138.2118 | 21.9 | 0.11 |
| polarC18 | 325.216 | 52.2907 | 21.9 | 0.11 |
| polarC18 | 325.216 | 175.2956 | 21.9 | 0.11 |
| polarC18 | 325.216 | 113.3771 | 21.9 | 0.11 |
| polarC18 | 325.216 | 197.9117 | 21.9 | 0.11 |
| polarC18 | 325.216 | 83.3232 | 21.9 | 0.11 |
| polarC18 | 325.216 | 52.5557 | 21.9 | 0.11 |
| polarC18 | 325.216 | 134.6573 | 21.9 | 0.11 |
| polarC18 | 325.216 | 81.9216 | 21.9 | 0.11 |
| polarC18 | 325.216 | 121.1491 | 21.9 | 0.11 |
| polarC18 | 325.216 | 295.4632 | 21.9 | 0.11 |
| polarC18 | 325.216 | 55.065 | 21.9 | 0.11 |
| polarC18 | 325.216 | 59.2609 | 21.9 | 0.11 |
| polarC18 | 325.216 | 149.9094 | 21.9 | 0.11 |
| polarC18 | 325.216 | 57.4842 | 21.9 | 0.11 |
| polarC18 | 325.216 | 122.7751 | 21.9 | 0.11 |

| | | | | |
|---|---|---|---|---|
| polarC18 | 325.216 | 121.0177 | 21.9 | 0.11 |
| polarC18 | 325.216 | 216.6796 | 21.9 | 0.11 |
| polarC18 | 325.216 | 123.1998 | 21.9 | 0.11 |
| polarC18 | 325.216 | 54.4638 | 21.9 | 0.11 |
| polarC18 | 325.216 | 50.488 | 21.9 | 0.11 |
| polarC18 | 325.216 | 69.6728 | 21.9 | 0.11 |
| polarC18 | 325.216 | 58.1259 | 21.9 | 0.11 |
| polarC18 | 325.216 | 259.4105 | 21.9 | 0.11 |
| polarC18 | 325.216 | 304.8128 | 21.9 | 0.11 |
| polarC18 | 325.216 | 246.2979 | 21.9 | 0.11 |
| polarC18 | 325.216 | 118.039 | 21.9 | 0.11 |
| polarC18 | 325.216 | 171.4069 | 21.9 | 0.11 |
| polarC18 | 325.216 | 80.0821 | 21.9 | 0.11 |
| polarC18 | 325.216 | 70.1855 | 21.9 | 0.11 |
| polarC18 | 325.216 | 145.4521 | 21.9 | 0.11 |
| polarC18 | 325.216 | 115.2249 | 21.9 | 0.11 |
| polarC18 | 325.216 | 99.9087 | 21.9 | 0.11 |
| polarC18 | 325.216 | 170.0759 | 21.9 | 0.11 |
| polarC18 | 325.216 | 317.1584 | 21.9 | 0.11 |
| polarC18 | 325.216 | 95.9585 | 21.9 | 0.11 |
| polarC18 | 325.216 | 218.3754 | 21.8 | 0.11 |
| polarC18 | 325.216 | 53.2264 | 21.8 | 0.11 |
| polarC18 | 325.216 | 144.9855 | 21.8 | 0.11 |
| polarC18 | 325.216 | 103.4245 | 21.8 | 0.11 |
| polarC18 | 325.216 | 59.84 | 21.8 | 0.11 |
| polarC18 | 325.216 | 57.3614 | 21.8 | 0.11 |
| polarC18 | 325.216 | 146.0922 | 21.8 | 0.11 |
| polarC18 | 325.216 | 147.773 | 21.8 | 0.11 |
| polarC18 | 325.216 | 183.9148 | 21.8 | 0.11 |
| polarC18 | 325.216 | 85.2923 | 21.8 | 0.11 |
| polarC18 | 325.216 | 51.1325 | 21.8 | 0.11 |
| polarC18 | 325.216 | 129.8118 | 21.8 | 0.11 |
| polarC18 | 325.216 | 73.4223 | 21.8 | 0.11 |
| polarC18 | 325.216 | 206.2234 | 21.8 | 0.11 |
| polarC18 | 325.216 | 86.649 | 21.8 | 0.11 |
| polarC18 | 325.216 | 71.8111 | 21.8 | 0.11 |
| polarC18 | 325.216 | 270.953 | 21.8 | 0.11 |
| polarC18 | 325.216 | 79.1509 | 21.8 | 0.11 |
| polarC18 | 325.216 | 59.0066 | 21.8 | 0.11 |
| polarC18 | 325.216 | 158.7514 | 21.8 | 0.11 |
| polarC18 | 325.216 | 63.4904 | 21.8 | 0.11 |
| polarC18 | 325.216 | 302.838 | 21.8 | 0.11 |

| | | | | |
|---|---|---|---|---|
| polarC18 | 325.216 | 109.742 | 21.8 | 0.11 |
| polarC18 | 325.216 | 56.2483 | 21.8 | 0.11 |
| polarC18 | 325.216 | 316.0863 | 21.8 | 0.11 |
| polarC18 | 325.216 | 96.2151 | 21.8 | 0.11 |
| polarC18 | 325.216 | 280.7013 | 21.8 | 0.11 |
| polarC18 | 325.216 | 125.0593 | 21.8 | 0.11 |
| polarC18 | 325.216 | 254.0155 | 21.8 | 0.11 |
| polarC18 | 325.216 | 296.006 | 21.8 | 0.11 |
| polarC18 | 325.216 | 72.3887 | 21.8 | 0.11 |
| polarC18 | 325.216 | 255.1721 | 21.8 | 0.11 |
| polarC18 | 325.216 | 108.9539 | 21.8 | 0.11 |
| polarC18 | 325.216 | 142.5969 | 21.8 | 0.11 |
| polarC18 | 325.216 | 51.7682 | 21.8 | 0.11 |
| polarC18 | 325.216 | 54.9847 | 21.8 | 0.11 |
| polarC18 | 325.216 | 135.8762 | 21.8 | 0.11 |
| polarC18 | 325.216 | 93.4812 | 21.8 | 0.11 |
| polarC18 | 325.216 | 263.6455 | 21.8 | 0.11 |
| polarC18 | 325.216 | 66.5823 | 21.8 | 0.11 |
| polarC18 | 325.216 | 93.261 | 21.8 | 0.11 |
| polarC18 | 325.216 | 157.0869 | 21.8 | 0.11 |
| polarC18 | 325.216 | 316.2273 | 21.8 | 0.11 |
| polarC18 | 325.216 | 273.4378 | 21.8 | 0.11 |
| polarC18 | 325.216 | 112.4419 | 21.8 | 0.11 |
| polarC18 | 325.216 | 97.6377 | 21.8 | 0.11 |
| polarC18 | 325.216 | 103.3279 | 21.8 | 0.11 |
| polarC18 | 325.216 | 90.1516 | 21.8 | 0.11 |
| polarC18 | 325.216 | 61.4725 | 21.8 | 0.11 |
| polarC18 | 325.216 | 233.5785 | 21.8 | 0.11 |
| polarC18 | 325.216 | 80.0879 | 21.8 | 0.11 |
| polarC18 | 325.216 | 111.4581 | 21.8 | 0.11 |
| polarC18 | 325.216 | 76.4498 | 21.8 | 0.11 |
| polarC18 | 325.216 | 138.1772 | 21.8 | 0.11 |
| polarC18 | 325.216 | 101.7217 | 21.8 | 0.11 |
| polarC18 | 325.216 | 51.5364 | 21.7 | 0.11 |
| polarC18 | 325.216 | 88.9017 | 21.7 | 0.11 |
| polarC18 | 325.216 | 57.6268 | 21.7 | 0.11 |
| polarC18 | 325.216 | 68.1148 | 21.7 | 0.11 |
| polarC18 | 325.216 | 62.2178 | 21.7 | 0.11 |
| polarC18 | 325.216 | 58.5887 | 21.7 | 0.11 |
| polarC18 | 325.216 | 78.5866 | 21.7 | 0.11 |
| polarC18 | 325.216 | 204.8717 | 21.7 | 0.11 |
| polarC18 | 325.216 | 109.8737 | 21.7 | 0.11 |

| | | | | |
|---|---|---|---|---|
| polarC18 | 325.216 | 79.5914 | 21.7 | 0.11 |
| polarC18 | 325.216 | 61.5253 | 21.7 | 0.11 |
| polarC18 | 325.216 | 311.719 | 21.7 | 0.11 |
| polarC18 | 325.216 | 132.3163 | 21.7 | 0.11 |
| polarC18 | 325.216 | 163.945 | 21.7 | 0.11 |
| polarC18 | 325.216 | 277.9449 | 21.7 | 0.11 |
| polarC18 | 325.216 | 137.5359 | 21.7 | 0.11 |
| polarC18 | 325.216 | 56.9367 | 21.7 | 0.11 |
| polarC18 | 325.216 | 58.0882 | 21.7 | 0.11 |
| polarC18 | 325.216 | 136.6033 | 21.7 | 0.11 |
| polarC18 | 325.216 | 83.8039 | 21.7 | 0.11 |
| polarC18 | 325.216 | 58.6371 | 21.7 | 0.11 |
| polarC18 | 325.216 | 188.7943 | 21.7 | 0.11 |
| polarC18 | 325.216 | 63.954 | 21.7 | 0.11 |
| polarC18 | 325.216 | 59.6339 | 21.7 | 0.11 |
| polarC18 | 325.216 | 73.6956 | 21.7 | 0.11 |
| polarC18 | 325.216 | 79.0924 | 21.7 | 0.11 |
| polarC18 | 325.216 | 141.0094 | 21.7 | 0.11 |
| polarC18 | 325.216 | 108.1898 | 21.7 | 0.11 |
| polarC18 | 325.216 | 110.8671 | 21.7 | 0.11 |
| polarC18 | 325.216 | 78.1227 | 21.7 | 0.11 |
| polarC18 | 325.216 | 63.6818 | 21.7 | 0.11 |
| polarC18 | 325.216 | 104.3425 | 21.7 | 0.11 |
| polarC18 | 325.216 | 71.033 | 21.7 | 0.11 |
| polarC18 | 325.216 | 300.852 | 21.7 | 0.11 |
| polarC18 | 325.216 | 84.4203 | 21.7 | 0.11 |
| polarC18 | 325.216 | 69.6109 | 21.7 | 0.11 |
| polarC18 | 325.216 | 65.3684 | 21.7 | 0.11 |
| polarC18 | 325.216 | 98.2558 | 21.7 | 0.11 |
| polarC18 | 325.216 | 70.5917 | 21.7 | 0.11 |
| polarC18 | 325.216 | 136.9241 | 21.7 | 0.11 |
| polarC18 | 325.216 | 61.108 | 21.6 | 0.11 |
| polarC18 | 325.216 | 338.7794 | 21.6 | 0.11 |
| polarC18 | 325.216 | 100.3463 | 21.6 | 0.11 |
| polarC18 | 325.216 | 156.5754 | 21.6 | 0.11 |
| polarC18 | 325.216 | 130.3022 | 21.6 | 0.11 |
| polarC18 | 325.216 | 77.9705 | 21.6 | 0.11 |
| polarC18 | 325.216 | 51.1959 | 21.6 | 0.11 |
| polarC18 | 325.216 | 166.9259 | 21.6 | 0.11 |
| polarC18 | 325.216 | 127.0365 | 21.6 | 0.11 |
| polarC18 | 325.216 | 277.3556 | 21.6 | 0.11 |
| polarC18 | 325.216 | 148.6462 | 21.6 | 0.11 |

| | | | | |
|---|---|---|---|---|
| polarC18 | 325.216 | 339.5891 | 21.6 | 0.11 |
| polarC18 | 325.216 | 51.6688 | 21.6 | 0.11 |
| polarC18 | 325.216 | 55.3764 | 21.6 | 0.11 |
| polarC18 | 325.216 | 328.4584 | 21.6 | 0.11 |
| polarC18 | 325.216 | 166.4062 | 21.6 | 0.11 |
| polarC18 | 325.216 | 321.1924 | 21.6 | 0.11 |
| polarC18 | 325.216 | 138.9048 | 21.6 | 0.11 |
| polarC18 | 325.216 | 249.5464 | 21.6 | 0.11 |
| polarC18 | 325.216 | 94.288 | 21.6 | 0.11 |
| polarC18 | 325.216 | 128.2746 | 21.6 | 0.11 |
| polarC18 | 325.216 | 84.8619 | 21.6 | 0.11 |
| polarC18 | 325.216 | 72.0632 | 21.6 | 0.11 |
| polarC18 | 325.216 | 57.8148 | 21.6 | 0.11 |
| polarC18 | 325.216 | 100.4142 | 21.6 | 0.11 |
| polarC18 | 325.216 | 330.1025 | 21.6 | 0.11 |
| polarC18 | 325.216 | 64.494 | 21.6 | 0.11 |
| polarC18 | 325.216 | 278.1329 | 21.6 | 0.11 |
| polarC18 | 325.216 | 261.3377 | 21.6 | 0.11 |
| polarC18 | 325.216 | 55.3101 | 21.6 | 0.11 |
| polarC18 | 325.216 | 159.8159 | 21.6 | 0.11 |
| polarC18 | 325.216 | 237.6073 | 21.6 | 0.11 |
| polarC18 | 325.216 | 180.0071 | 21.6 | 0.11 |
| polarC18 | 325.216 | 197.3709 | 21.6 | 0.11 |
| polarC18 | 325.216 | 160.8812 | 21.6 | 0.11 |
| polarC18 | 325.216 | 105.9147 | 21.6 | 0.11 |
| polarC18 | 325.216 | 129.7395 | 21.6 | 0.11 |
| polarC18 | 325.216 | 113.0819 | 21.6 | 0.11 |
| polarC18 | 325.216 | 53.4471 | 21.6 | 0.11 |
| polarC18 | 325.216 | 63.8719 | 21.6 | 0.11 |
| polarC18 | 325.216 | 73.746 | 21.6 | 0.11 |
| polarC18 | 325.216 | 130.2809 | 21.6 | 0.11 |
| polarC18 | 325.216 | 81.2007 | 21.6 | 0.11 |
| polarC18 | 325.216 | 84.3518 | 21.6 | 0.11 |
| polarC18 | 325.216 | 150.9191 | 21.6 | 0.11 |
| polarC18 | 325.216 | 89.2944 | 21.6 | 0.11 |
| polarC18 | 325.216 | 58.3289 | 21.6 | 0.11 |
| polarC18 | 325.216 | 183.2555 | 21.6 | 0.11 |
| polarC18 | 325.216 | 111.4819 | 21.6 | 0.11 |
| polarC18 | 325.216 | 52.1976 | 21.6 | 0.11 |
| polarC18 | 325.216 | 58.634 | 21.6 | 0.11 |
| polarC18 | 325.216 | 141.2819 | 21.6 | 0.11 |
| polarC18 | 325.216 | 169.5082 | 21.6 | 0.11 |

| | | | | |
|---|---|---|---|---|
| polarC18 | 325.216 | 108.1978 | 21.5 | 0.11 |
| polarC18 | 325.216 | 89.7684 | 21.5 | 0.11 |
| polarC18 | 325.216 | 175.5543 | 21.5 | 0.11 |
| polarC18 | 325.216 | 52.9611 | 21.5 | 0.11 |
| polarC18 | 325.216 | 311.5999 | 21.5 | 0.11 |
| polarC18 | 325.216 | 213.9311 | 21.5 | 0.11 |
| polarC18 | 325.216 | 83.3113 | 21.5 | 0.11 |
| polarC18 | 325.216 | 61.7751 | 21.5 | 0.11 |
| polarC18 | 325.216 | 256.5916 | 21.5 | 0.11 |
| polarC18 | 325.216 | 60.4266 | 21.5 | 0.11 |
| polarC18 | 325.216 | 129.8001 | 21.5 | 0.11 |
| polarC18 | 325.216 | 98.504 | 21.5 | 0.11 |
| polarC18 | 325.216 | 77.8721 | 21.5 | 0.11 |
| polarC18 | 325.216 | 158.5098 | 21.5 | 0.11 |
| polarC18 | 325.216 | 124.9832 | 21.5 | 0.11 |
| polarC18 | 325.216 | 69.6987 | 21.5 | 0.11 |
| polarC18 | 325.216 | 240.7291 | 21.5 | 0.11 |
| polarC18 | 325.216 | 67.7215 | 21.5 | 0.11 |
| polarC18 | 325.216 | 53.6356 | 21.5 | 0.11 |
| polarC18 | 325.216 | 245.511 | 21.5 | 0.11 |
| polarC18 | 325.216 | 124.1937 | 21.5 | 0.11 |
| polarC18 | 325.216 | 95.2001 | 21.5 | 0.11 |
| polarC18 | 325.216 | 135.8338 | 21.5 | 0.11 |
| polarC18 | 325.216 | 206.5255 | 21.5 | 0.11 |
| polarC18 | 325.216 | 55.2444 | 21.5 | 0.11 |
| polarC18 | 325.216 | 144.0241 | 21.5 | 0.11 |
| polarC18 | 325.216 | 87.3407 | 21.5 | 0.11 |
| polarC18 | 325.216 | 78.0475 | 21.5 | 0.11 |
| polarC18 | 325.216 | 51.3248 | 21.5 | 0.11 |
| polarC18 | 325.216 | 162.091 | 21.5 | 0.11 |
| polarC18 | 325.216 | 108.021 | 21.5 | 0.11 |
| polarC18 | 325.216 | 229.4559 | 21.5 | 0.11 |
| polarC18 | 325.216 | 95.0986 | 21.5 | 0.11 |
| polarC18 | 325.216 | 65.1015 | 21.5 | 0.11 |
| polarC18 | 325.216 | 101.1355 | 21.5 | 0.11 |
| polarC18 | 325.216 | 319.2878 | 21.5 | 0.11 |
| polarC18 | 325.216 | 66.7075 | 21.5 | 0.11 |
| polarC18 | 325.216 | 179.1145 | 21.5 | 0.11 |
| polarC18 | 325.216 | 157.24 | 21.5 | 0.11 |
| polarC18 | 325.216 | 259.9996 | 21.5 | 0.11 |
| polarC18 | 325.216 | 89.8379 | 21.5 | 0.11 |
| polarC18 | 325.216 | 148.409 | 21.5 | 0.11 |

| | | | | |
|---|---|---|---|---|
| polarC18 | 325.216 | 183.8003 | 21.5 | 0.11 |
| polarC18 | 325.216 | 193.0368 | 21.5 | 0.11 |
| polarC18 | 325.216 | 197.7948 | 21.5 | 0.11 |
| polarC18 | 325.216 | 85.7921 | 21.5 | 0.11 |
| polarC18 | 325.216 | 211.0524 | 21.5 | 0.11 |
| polarC18 | 325.216 | 83.7266 | 21.5 | 0.11 |
| polarC18 | 325.216 | 228.11 | 21.5 | 0.11 |
| polarC18 | 325.216 | 87.6609 | 21.5 | 0.11 |
| polarC18 | 325.216 | 50.8387 | 21.5 | 0.11 |
| polarC18 | 325.216 | 104.1075 | 21.5 | 0.11 |
| polarC18 | 325.216 | 67.6588 | 21.5 | 0.11 |
| polarC18 | 325.216 | 290.5574 | 21.5 | 0.11 |
| polarC18 | 325.216 | 92.5977 | 21.4 | 0.11 |
| polarC18 | 325.216 | 110.7395 | 21.4 | 0.11 |
| polarC18 | 325.216 | 62.2489 | 21.4 | 0.11 |
| polarC18 | 325.216 | 112.4047 | 21.4 | 0.11 |
| polarC18 | 325.216 | 57.2984 | 21.4 | 0.11 |
| polarC18 | 325.216 | 61.7937 | 21.4 | 0.11 |
| polarC18 | 325.216 | 57.0489 | 21.4 | 0.11 |
| polarC18 | 325.216 | 52.7107 | 21.4 | 0.11 |
| polarC18 | 325.216 | 94.0586 | 21.4 | 0.11 |
| polarC18 | 325.216 | 160.7729 | 21.4 | 0.11 |
| polarC18 | 325.216 | 151.3244 | 21.4 | 0.11 |
| polarC18 | 325.216 | 58.854 | 21.4 | 0.11 |
| polarC18 | 325.216 | 104.0491 | 21.4 | 0.11 |
| polarC18 | 325.216 | 61.7793 | 21.4 | 0.11 |
| polarC18 | 325.216 | 164.6578 | 21.4 | 0.11 |
| polarC18 | 325.216 | 106.6672 | 21.4 | 0.11 |
| polarC18 | 325.216 | 235.3074 | 21.4 | 0.11 |
| polarC18 | 325.216 | 54.3034 | 21.4 | 0.11 |
| polarC18 | 325.216 | 319.8763 | 21.4 | 0.11 |
| polarC18 | 325.216 | 90.9186 | 21.4 | 0.11 |
| polarC18 | 325.216 | 149.5968 | 21.4 | 0.11 |
| polarC18 | 325.216 | 90.135 | 21.4 | 0.11 |
| polarC18 | 325.216 | 267.2496 | 21.4 | 0.11 |
| polarC18 | 325.216 | 85.3094 | 21.4 | 0.11 |
| polarC18 | 325.216 | 250.3327 | 21.4 | 0.11 |
| polarC18 | 325.216 | 70.9677 | 21.4 | 0.11 |
| polarC18 | 325.216 | 50.0778 | 21.4 | 0.11 |
| polarC18 | 325.216 | 70.3693 | 21.4 | 0.11 |
| polarC18 | 325.216 | 73.8563 | 21.4 | 0.11 |
| polarC18 | 325.216 | 51.243 | 21.4 | 0.11 |

| | | | | |
|---|---|---|---|---|
| polarC18 | 325.216 | 67.2848 | 21.4 | 0.11 |
| polarC18 | 325.216 | 63.4026 | 21.4 | 0.11 |
| polarC18 | 325.216 | 51.9585 | 21.4 | 0.11 |
| polarC18 | 325.216 | 64.8682 | 21.4 | 0.11 |
| polarC18 | 325.216 | 58.6123 | 21.4 | 0.11 |
| polarC18 | 325.216 | 87.8525 | 21.4 | 0.11 |
| polarC18 | 325.216 | 116.9537 | 21.4 | 0.11 |
| polarC18 | 325.216 | 87.1373 | 21.4 | 0.11 |
| polarC18 | 325.216 | 89.4009 | 21.4 | 0.11 |
| polarC18 | 325.216 | 52.6159 | 21.4 | 0.11 |
| polarC18 | 325.216 | 63.5957 | 21.4 | 0.11 |
| polarC18 | 325.216 | 69.0481 | 21.4 | 0.11 |
| polarC18 | 325.216 | 66.4473 | 21.4 | 0.11 |
| polarC18 | 325.216 | 53.7873 | 21.4 | 0.11 |
| polarC18 | 325.216 | 60.1542 | 21.4 | 0.11 |
| polarC18 | 325.216 | 152.7112 | 21.4 | 0.11 |
| polarC18 | 325.216 | 181.0327 | 21.3 | 0.11 |
| polarC18 | 325.216 | 92.9194 | 21.3 | 0.11 |
| polarC18 | 325.216 | 73.7208 | 21.3 | 0.11 |
| polarC18 | 325.216 | 95.6523 | 21.3 | 0.11 |
| polarC18 | 325.216 | 88.2353 | 21.3 | 0.11 |
| polarC18 | 325.216 | 243.3539 | 21.3 | 0.11 |
| polarC18 | 325.216 | 55.666 | 21.3 | 0.11 |
| polarC18 | 325.216 | 69.3577 | 21.3 | 0.11 |
| polarC18 | 325.216 | 85.6356 | 21.3 | 0.11 |
| polarC18 | 325.216 | 53.9058 | 21.3 | 0.11 |
| polarC18 | 325.216 | 215.021 | 21.3 | 0.11 |
| polarC18 | 325.216 | 313.384 | 21.3 | 0.11 |
| polarC18 | 325.216 | 190.8403 | 21.3 | 0.11 |
| polarC18 | 325.216 | 137.7293 | 21.3 | 0.11 |
| polarC18 | 325.216 | 271.8914 | 21.3 | 0.11 |
| polarC18 | 325.216 | 65.7489 | 21.3 | 0.11 |
| polarC18 | 325.216 | 183.5548 | 21.3 | 0.11 |
| polarC18 | 325.216 | 113.7564 | 21.3 | 0.11 |
| polarC18 | 325.216 | 239.8258 | 21.3 | 0.11 |
| polarC18 | 325.216 | 54.1791 | 21.3 | 0.11 |
| polarC18 | 325.216 | 60.6416 | 21.3 | 0.11 |
| polarC18 | 325.216 | 64.133 | 21.3 | 0.11 |
| polarC18 | 325.216 | 252.1068 | 21.3 | 0.11 |
| polarC18 | 325.216 | 84.6697 | 21.3 | 0.11 |
| polarC18 | 325.216 | 50.9263 | 21.3 | 0.11 |
| polarC18 | 325.216 | 58.4935 | 21.3 | 0.11 |

| | | | | |
|---|---|---|---|---|
| polarC18 | 325.216 | 86.8462 | 21.3 | 0.11 |
| polarC18 | 325.216 | 135.5835 | 21.3 | 0.11 |
| polarC18 | 325.216 | 83.7221 | 21.3 | 0.11 |
| polarC18 | 325.216 | 95.8951 | 21.3 | 0.11 |
| polarC18 | 325.216 | 219.3695 | 21.3 | 0.11 |
| polarC18 | 325.216 | 66.3981 | 21.3 | 0.11 |
| polarC18 | 325.216 | 131.1704 | 21.3 | 0.11 |
| polarC18 | 325.216 | 161.1381 | 21.3 | 0.11 |
| polarC18 | 325.216 | 88.3099 | 21.3 | 0.11 |
| polarC18 | 325.216 | 51.9065 | 21.3 | 0.11 |
| polarC18 | 325.216 | 56.5137 | 21.3 | 0.11 |
| polarC18 | 325.216 | 67.7149 | 21.3 | 0.11 |
| polarC18 | 325.216 | 290.8251 | 21.3 | 0.11 |
| polarC18 | 325.216 | 191.8799 | 21.3 | 0.11 |
| polarC18 | 325.216 | 220.4429 | 21.3 | 0.11 |
| polarC18 | 325.216 | 227.5648 | 21.3 | 0.11 |
| polarC18 | 325.216 | 180.7546 | 21.3 | 0.11 |
| polarC18 | 325.216 | 244.0267 | 21.3 | 0.11 |
| polarC18 | 325.216 | 55.8108 | 21.3 | 0.11 |
| polarC18 | 325.216 | 100.5007 | 21.3 | 0.11 |
| polarC18 | 325.216 | 312.4132 | 21.3 | 0.11 |
| polarC18 | 325.216 | 70.0459 | 21.3 | 0.11 |
| polarC18 | 325.216 | 174.9243 | 21.3 | 0.11 |
| polarC18 | 325.216 | 281.9308 | 21.3 | 0.11 |
| polarC18 | 325.216 | 68.5766 | 21.3 | 0.11 |
| polarC18 | 325.216 | 163.6156 | 21.3 | 0.11 |
| polarC18 | 325.216 | 96.4091 | 21.3 | 0.11 |
| polarC18 | 325.216 | 280.6505 | 21.2 | 0.11 |
| polarC18 | 325.216 | 336.5741 | 21.2 | 0.11 |
| polarC18 | 325.216 | 74.7025 | 21.2 | 0.11 |
| polarC18 | 325.216 | 162.4282 | 21.2 | 0.11 |
| polarC18 | 325.216 | 115.2536 | 21.2 | 0.11 |
| polarC18 | 325.216 | 148.6004 | 21.2 | 0.11 |
| polarC18 | 325.216 | 145.9947 | 21.2 | 0.11 |
| polarC18 | 325.216 | 87.3967 | 21.2 | 0.11 |
| polarC18 | 325.216 | 57.0142 | 21.2 | 0.11 |
| polarC18 | 325.216 | 152.5634 | 21.2 | 0.11 |
| polarC18 | 325.216 | 193.0794 | 21.2 | 0.11 |
| polarC18 | 325.216 | 232.898 | 21.2 | 0.11 |
| polarC18 | 325.216 | 60.8559 | 21.2 | 0.11 |
| polarC18 | 325.216 | 204.6225 | 21.2 | 0.11 |
| polarC18 | 325.216 | 146.0412 | 21.2 | 0.11 |

| | | | | |
|---|---|---|---|---|
| polarC18 | 325.216 | 79.8603 | 21.2 | 0.11 |
| polarC18 | 325.216 | 55.3817 | 21.2 | 0.11 |
| polarC18 | 325.216 | 54.9826 | 21.2 | 0.11 |
| polarC18 | 325.216 | 77.7024 | 21.2 | 0.11 |
| polarC18 | 325.216 | 127.9935 | 21.2 | 0.11 |
| polarC18 | 325.216 | 73.319 | 21.2 | 0.11 |
| polarC18 | 325.216 | 63.7677 | 21.2 | 0.11 |
| polarC18 | 325.216 | 50.4182 | 21.2 | 0.11 |
| polarC18 | 325.216 | 57.264 | 21.2 | 0.11 |
| polarC18 | 325.216 | 73.9166 | 21.2 | 0.11 |
| polarC18 | 325.216 | 79.5105 | 21.2 | 0.11 |
| polarC18 | 325.216 | 343.0945 | 21.2 | 0.11 |
| polarC18 | 325.216 | 84.8186 | 21.2 | 0.11 |
| polarC18 | 325.216 | 156.6735 | 21.2 | 0.11 |
| polarC18 | 325.216 | 97.6182 | 21.2 | 0.11 |
| polarC18 | 325.216 | 54.4193 | 21.2 | 0.11 |
| polarC18 | 325.216 | 52.4572 | 21.2 | 0.11 |
| polarC18 | 325.216 | 69.512 | 21.2 | 0.11 |
| polarC18 | 325.216 | 63.0657 | 21.2 | 0.11 |
| polarC18 | 325.216 | 69.1026 | 21.2 | 0.11 |
| polarC18 | 325.216 | 264.3052 | 21.1 | 0.11 |
| polarC18 | 325.216 | 65.2101 | 21.1 | 0.11 |
| polarC18 | 325.216 | 95.9344 | 21.1 | 0.11 |
| polarC18 | 325.216 | 77.1423 | 21.1 | 0.11 |
| polarC18 | 325.216 | 236.849 | 21.1 | 0.11 |
| polarC18 | 325.216 | 101.2085 | 21.1 | 0.11 |
| polarC18 | 325.216 | 90.6165 | 21.1 | 0.11 |
| polarC18 | 325.216 | 136.0147 | 21.1 | 0.11 |
| polarC18 | 325.216 | 66.6932 | 21.1 | 0.11 |
| polarC18 | 325.216 | 330.5479 | 21.1 | 0.11 |
| polarC18 | 325.216 | 55.37 | 21.1 | 0.11 |
| polarC18 | 325.216 | 124.9349 | 21.1 | 0.11 |
| polarC18 | 325.216 | 64.2447 | 21.1 | 0.11 |
| polarC18 | 325.216 | 81.6684 | 21.1 | 0.11 |
| polarC18 | 325.216 | 70.9834 | 21.1 | 0.11 |
| polarC18 | 325.216 | 69.3988 | 21.1 | 0.11 |
| polarC18 | 325.216 | 62.966 | 21.1 | 0.11 |
| polarC18 | 325.216 | 139.3979 | 21.1 | 0.11 |
| polarC18 | 325.216 | 159.7632 | 21.1 | 0.11 |
| polarC18 | 325.216 | 66.5757 | 21.1 | 0.11 |
| polarC18 | 325.216 | 52.475 | 21.1 | 0.11 |
| polarC18 | 325.216 | 89.3788 | 21.1 | 0.11 |

| | | | | |
|---|---|---|---|---|
| polarC18 | 325.216 | 55.7188 | 21.1 | 0.11 |
| polarC18 | 325.216 | 123.5745 | 21.1 | 0.11 |
| polarC18 | 325.216 | 95.9956 | 21.1 | 0.11 |
| polarC18 | 325.216 | 271.2757 | 21.1 | 0.11 |
| polarC18 | 325.216 | 257.8636 | 21.1 | 0.11 |
| polarC18 | 325.216 | 77.1789 | 21.1 | 0.11 |
| polarC18 | 325.216 | 241.8497 | 21.1 | 0.11 |
| polarC18 | 325.216 | 136.8456 | 21.1 | 0.11 |
| polarC18 | 325.216 | 68.4622 | 21.1 | 0.11 |
| polarC18 | 325.216 | 102.373 | 21.1 | 0.11 |
| polarC18 | 325.216 | 259.6528 | 21.1 | 0.11 |
| polarC18 | 325.216 | 275.7758 | 21 | 0.11 |
| polarC18 | 325.216 | 307.1144 | 21 | 0.11 |
| polarC18 | 325.216 | 172.229 | 21 | 0.11 |
| polarC18 | 325.216 | 56.5212 | 21 | 0.11 |
| polarC18 | 325.216 | 210.9612 | 21 | 0.11 |
| polarC18 | 325.216 | 69.8667 | 21 | 0.11 |
| polarC18 | 325.216 | 55.5202 | 21 | 0.11 |
| polarC18 | 325.216 | 74.5507 | 21 | 0.11 |
| polarC18 | 325.216 | 153.7906 | 21 | 0.11 |
| polarC18 | 325.216 | 108.3386 | 21 | 0.11 |
| polarC18 | 325.216 | 61.923 | 21 | 0.11 |
| polarC18 | 325.216 | 59.6915 | 21 | 0.11 |
| polarC18 | 325.216 | 100.5867 | 21 | 0.11 |
| polarC18 | 325.216 | 87.7685 | 21 | 0.11 |
| polarC18 | 325.216 | 118.051 | 21 | 0.11 |
| polarC18 | 325.216 | 53.4068 | 21 | 0.11 |
| polarC18 | 325.216 | 108.1123 | 21 | 0.11 |
| polarC18 | 325.216 | 288.5577 | 21 | 0.11 |
| polarC18 | 325.216 | 97.84 | 21 | 0.11 |
| polarC18 | 325.216 | 84.6023 | 21 | 0.11 |
| polarC18 | 325.216 | 64.3087 | 21 | 0.11 |
| polarC18 | 325.216 | 143.8422 | 21 | 0.11 |
| polarC18 | 325.216 | 171.9746 | 21 | 0.11 |
| polarC18 | 325.216 | 73.3076 | 21 | 0.11 |
| polarC18 | 325.216 | 93.495 | 21 | 0.11 |
| polarC18 | 325.216 | 54.5908 | 21 | 0.11 |
| polarC18 | 325.216 | 97.2475 | 21 | 0.11 |
| polarC18 | 325.216 | 229.6064 | 21 | 0.11 |
| polarC18 | 325.216 | 110.9277 | 21 | 0.11 |
| polarC18 | 325.216 | 66.5326 | 21 | 0.11 |
| polarC18 | 325.216 | 221.5466 | 21 | 0.11 |

| | | | | |
|---|---|---|---|---|
| polarC18 | 325.216 | 126.8494 | 21 | 0.11 |
| polarC18 | 325.216 | 66.6656 | 21 | 0.11 |
| polarC18 | 325.216 | 60.2001 | 21 | 0.11 |
| polarC18 | 325.216 | 139.2083 | 21 | 0.11 |
| polarC18 | 325.216 | 50.653 | 21 | 0.11 |
| polarC18 | 325.216 | 57.1395 | 21 | 0.11 |
| polarC18 | 325.216 | 152.813 | 21 | 0.11 |
| polarC18 | 325.216 | 87.4956 | 21 | 0.11 |
| polarC18 | 325.216 | 71.8836 | 21 | 0.11 |
| polarC18 | 325.216 | 95.1233 | 21 | 0.11 |
| polarC18 | 325.216 | 90.4471 | 21 | 0.11 |
| polarC18 | 325.216 | 65.1462 | 20.9 | 0.11 |
| polarC18 | 325.216 | 128.2325 | 20.9 | 0.11 |
| polarC18 | 325.216 | 60.9506 | 20.9 | 0.11 |
| polarC18 | 325.216 | 58.0058 | 20.9 | 0.11 |
| polarC18 | 325.216 | 139.9026 | 20.9 | 0.11 |
| polarC18 | 325.216 | 106.1244 | 20.9 | 0.11 |
| polarC18 | 325.216 | 85.4875 | 20.9 | 0.11 |
| polarC18 | 325.216 | 67.0498 | 20.9 | 0.11 |
| polarC18 | 325.216 | 97.1718 | 20.9 | 0.11 |
| polarC18 | 325.216 | 294.3446 | 20.9 | 0.11 |
| polarC18 | 325.216 | 77.71 | 20.9 | 0.11 |
| polarC18 | 325.216 | 76.7654 | 20.9 | 0.11 |
| polarC18 | 325.216 | 95.0719 | 20.9 | 0.11 |
| polarC18 | 325.216 | 230.6915 | 20.9 | 0.11 |
| polarC18 | 325.216 | 53.6013 | 20.9 | 0.11 |
| polarC18 | 325.216 | 58.4244 | 20.9 | 0.11 |
| polarC18 | 325.216 | 178.9679 | 20.9 | 0.11 |
| polarC18 | 325.216 | 333.7313 | 20.9 | 0.11 |
| polarC18 | 325.216 | 61.578 | 20.9 | 0.11 |
| polarC18 | 325.216 | 125.0924 | 20.9 | 0.11 |
| polarC18 | 325.216 | 91.3049 | 20.9 | 0.11 |
| polarC18 | 325.216 | 69.9959 | 20.9 | 0.11 |
| polarC18 | 325.216 | 258.7565 | 20.9 | 0.11 |
| polarC18 | 325.216 | 149.6206 | 20.9 | 0.11 |
| polarC18 | 325.216 | 126.3992 | 20.9 | 0.11 |
| polarC18 | 325.216 | 50.2431 | 20.9 | 0.11 |
| polarC18 | 325.216 | 56.6111 | 20.9 | 0.11 |
| polarC18 | 325.216 | 57.5048 | 20.9 | 0.11 |
| polarC18 | 325.216 | 107.1225 | 20.9 | 0.11 |
| polarC18 | 325.216 | 94.233 | 20.9 | 0.11 |
| polarC18 | 325.216 | 102.0845 | 20.9 | 0.11 |

| | | | | |
|---|---|---|---|---|
| polarC18 | 325.216 | 59.6787 | 20.9 | 0.11 |
| polarC18 | 325.216 | 66.2134 | 20.9 | 0.11 |
| polarC18 | 325.216 | 128.7766 | 20.9 | 0.11 |
| polarC18 | 325.216 | 160.482 | 20.9 | 0.11 |
| polarC18 | 325.216 | 54.4718 | 20.9 | 0.11 |
| polarC18 | 325.216 | 151.8826 | 20.9 | 0.11 |
| polarC18 | 325.216 | 71.091 | 20.9 | 0.11 |
| polarC18 | 325.216 | 243.4328 | 20.9 | 0.11 |
| polarC18 | 325.216 | 85.3805 | 20.9 | 0.11 |
| polarC18 | 325.216 | 93.9426 | 20.9 | 0.11 |
| polarC18 | 325.216 | 90.8802 | 20.9 | 0.11 |
| polarC18 | 325.216 | 351.3989 | 20.9 | 0.11 |
| polarC18 | 325.216 | 94.4981 | 20.9 | 0.11 |
| polarC18 | 325.216 | 134.9473 | 20.9 | 0.11 |
| polarC18 | 325.216 | 103.0773 | 20.9 | 0.11 |
| polarC18 | 325.216 | 85.8016 | 20.8 | 0.11 |
| polarC18 | 325.216 | 68.0534 | 20.8 | 0.11 |
| polarC18 | 325.216 | 122.7459 | 20.8 | 0.11 |
| polarC18 | 325.216 | 236.6557 | 20.8 | 0.11 |
| polarC18 | 325.216 | 62.0225 | 20.8 | 0.11 |
| polarC18 | 325.216 | 301.1166 | 20.8 | 0.11 |
| polarC18 | 325.216 | 73.267 | 20.8 | 0.11 |
| polarC18 | 325.216 | 122.5687 | 20.8 | 0.11 |
| polarC18 | 325.216 | 237.9727 | 20.8 | 0.11 |
| polarC18 | 325.216 | 75.6364 | 20.8 | 0.11 |
| polarC18 | 325.216 | 300.8026 | 20.8 | 0.11 |
| polarC18 | 325.216 | 55.1495 | 20.8 | 0.11 |
| polarC18 | 325.216 | 123.2779 | 20.8 | 0.11 |
| polarC18 | 325.216 | 317.7251 | 20.8 | 0.11 |
| polarC18 | 325.216 | 112.3748 | 20.8 | 0.11 |
| polarC18 | 325.216 | 76.8019 | 20.8 | 0.11 |
| polarC18 | 325.216 | 77.4982 | 20.8 | 0.11 |
| polarC18 | 325.216 | 53.5122 | 20.8 | 0.11 |
| polarC18 | 325.216 | 284.7643 | 20.8 | 0.11 |
| polarC18 | 325.216 | 178.6845 | 20.8 | 0.11 |
| polarC18 | 325.216 | 57.5853 | 20.8 | 0.11 |
| polarC18 | 325.216 | 57.5167 | 20.8 | 0.11 |
| polarC18 | 325.216 | 74.5443 | 20.8 | 0.11 |
| polarC18 | 325.216 | 64.8719 | 20.8 | 0.11 |
| polarC18 | 325.216 | 81.4977 | 20.8 | 0.11 |
| polarC18 | 325.216 | 65.4387 | 20.8 | 0.11 |
| polarC18 | 325.216 | 62.012 | 20.8 | 0.11 |

| | | | | |
|---|---|---|---|---|
| polarC18 | 325.216 | 108.2233 | 20.8 | 0.11 |
| polarC18 | 325.216 | 147.682 | 20.8 | 0.11 |
| polarC18 | 325.216 | 141.1022 | 20.8 | 0.11 |
| polarC18 | 325.216 | 50.0103 | 20.8 | 0.11 |
| polarC18 | 325.216 | 98.5839 | 20.8 | 0.11 |
| polarC18 | 325.216 | 89.6472 | 20.8 | 0.11 |
| polarC18 | 325.216 | 91.5781 | 20.8 | 0.11 |
| polarC18 | 325.216 | 307.0486 | 20.8 | 0.11 |
| polarC18 | 325.216 | 244.8009 | 20.8 | 0.11 |
| polarC18 | 325.216 | 65.508 | 20.7 | 0.11 |
| polarC18 | 325.216 | 57.2589 | 20.7 | 0.11 |
| polarC18 | 325.216 | 50.7569 | 20.7 | 0.11 |
| polarC18 | 325.216 | 222.0261 | 20.7 | 0.11 |
| polarC18 | 325.216 | 73.6197 | 20.7 | 0.11 |
| polarC18 | 325.216 | 275.1347 | 20.7 | 0.11 |
| polarC18 | 325.216 | 275.5947 | 20.7 | 0.11 |
| polarC18 | 325.216 | 139.0838 | 20.7 | 0.11 |
| polarC18 | 325.216 | 183.7681 | 20.7 | 0.11 |
| polarC18 | 325.216 | 64.1608 | 20.7 | 0.11 |
| polarC18 | 325.216 | 62.799 | 20.7 | 0.11 |
| polarC18 | 325.216 | 88.336 | 20.7 | 0.11 |
| polarC18 | 325.216 | 190.5315 | 20.7 | 0.11 |
| polarC18 | 325.216 | 54.2252 | 20.7 | 0.11 |
| polarC18 | 325.216 | 51.7819 | 20.7 | 0.11 |
| polarC18 | 325.216 | 117.7657 | 20.7 | 0.11 |
| polarC18 | 325.216 | 136.5342 | 20.7 | 0.11 |
| polarC18 | 325.216 | 70.0191 | 20.7 | 0.11 |
| polarC18 | 325.216 | 55.114 | 20.7 | 0.11 |
| polarC18 | 325.216 | 68.9704 | 20.7 | 0.11 |
| polarC18 | 325.216 | 108.0777 | 20.7 | 0.11 |
| polarC18 | 325.216 | 284.2874 | 20.7 | 0.11 |
| polarC18 | 325.216 | 51.0658 | 20.7 | 0.11 |
| polarC18 | 325.216 | 110.2896 | 20.7 | 0.11 |
| polarC18 | 325.216 | 175.2752 | 20.7 | 0.11 |
| polarC18 | 325.216 | 56.9976 | 20.7 | 0.11 |
| polarC18 | 325.216 | 50.9014 | 20.7 | 0.11 |
| polarC18 | 325.216 | 152.227 | 20.7 | 0.11 |
| polarC18 | 325.216 | 74.3564 | 20.7 | 0.11 |
| polarC18 | 325.216 | 204.2916 | 20.7 | 0.11 |
| polarC18 | 325.216 | 81.4095 | 20.7 | 0.11 |
| polarC18 | 325.216 | 95.4673 | 20.7 | 0.11 |
| polarC18 | 325.216 | 56.4062 | 20.7 | 0.11 |

| | | | | |
|---|---|---|---|---|
| polarC18 | 325.216 | 71.133 | 20.7 | 0.11 |
| polarC18 | 325.216 | 120.6605 | 20.7 | 0.11 |
| polarC18 | 325.216 | 82.3046 | 20.7 | 0.11 |
| polarC18 | 325.216 | 91.1491 | 20.7 | 0.11 |
| polarC18 | 325.216 | 106.9483 | 20.7 | 0.11 |
| polarC18 | 325.216 | 77.8599 | 20.7 | 0.11 |
| polarC18 | 325.216 | 225.8368 | 20.7 | 0.11 |
| polarC18 | 325.216 | 52.1651 | 20.6 | 0.11 |
| polarC18 | 325.216 | 107.1541 | 20.6 | 0.11 |
| polarC18 | 325.216 | 73.1918 | 20.6 | 0.11 |
| polarC18 | 325.216 | 64.0568 | 20.6 | 0.11 |
| polarC18 | 325.216 | 347.2207 | 20.6 | 0.11 |
| polarC18 | 325.216 | 76.9338 | 20.6 | 0.11 |
| polarC18 | 325.216 | 253.4428 | 20.6 | 0.11 |
| polarC18 | 325.216 | 56.1371 | 20.6 | 0.11 |
| polarC18 | 325.216 | 81.3027 | 20.6 | 0.11 |
| polarC18 | 325.216 | 55.8658 | 20.6 | 0.11 |
| polarC18 | 325.216 | 118.6957 | 20.6 | 0.11 |
| polarC18 | 325.216 | 50.018 | 20.6 | 0.11 |
| polarC18 | 325.216 | 113.918 | 20.6 | 0.11 |
| polarC18 | 325.216 | 102.0275 | 20.6 | 0.11 |
| polarC18 | 325.216 | 103.6461 | 20.6 | 0.11 |
| polarC18 | 325.216 | 130.504 | 20.6 | 0.11 |
| polarC18 | 325.216 | 149.7702 | 20.6 | 0.11 |
| polarC18 | 325.216 | 107.6085 | 20.6 | 0.11 |
| polarC18 | 325.216 | 129.0309 | 20.6 | 0.11 |
| polarC18 | 325.216 | 66.4403 | 20.6 | 0.11 |
| polarC18 | 325.216 | 75.6498 | 20.6 | 0.11 |
| polarC18 | 325.216 | 88.8675 | 20.6 | 0.11 |
| polarC18 | 325.216 | 73.5682 | 20.6 | 0.11 |
| polarC18 | 325.216 | 63.3365 | 20.6 | 0.11 |
| polarC18 | 325.216 | 184.0401 | 20.6 | 0.11 |
| polarC18 | 325.216 | 245.1602 | 20.6 | 0.11 |
| polarC18 | 325.216 | 50.4608 | 20.6 | 0.11 |
| polarC18 | 325.216 | 148.0537 | 20.6 | 0.11 |
| polarC18 | 325.216 | 249.6267 | 20.6 | 0.11 |
| polarC18 | 325.216 | 77.0085 | 20.6 | 0.11 |
| polarC18 | 325.216 | 190.1277 | 20.6 | 0.11 |
| polarC18 | 325.216 | 84.3983 | 20.6 | 0.11 |
| polarC18 | 325.216 | 59.5038 | 20.6 | 0.11 |
| polarC18 | 325.216 | 81.7268 | 20.6 | 0.11 |
| polarC18 | 325.216 | 52.6566 | 20.6 | 0.11 |

| | | | | |
|---|---|---|---|---|
| polarC18 | 325.216 | 200.0308 | 20.6 | 0.11 |
| polarC18 | 325.216 | 83.9654 | 20.5 | 0.11 |
| polarC18 | 325.216 | 143.761 | 20.5 | 0.11 |
| polarC18 | 325.216 | 52.5495 | 20.5 | 0.11 |
| polarC18 | 325.216 | 54.3246 | 20.5 | 0.11 |
| polarC18 | 325.216 | 53.7236 | 20.5 | 0.11 |
| polarC18 | 325.216 | 161.7711 | 20.5 | 0.11 |
| polarC18 | 325.216 | 56.8739 | 20.5 | 0.11 |
| polarC18 | 325.216 | 70.6755 | 20.5 | 0.11 |
| polarC18 | 325.216 | 225.4006 | 20.5 | 0.11 |
| polarC18 | 325.216 | 113.0907 | 20.5 | 0.11 |
| polarC18 | 325.216 | 87.0426 | 20.5 | 0.11 |
| polarC18 | 325.216 | 67.9692 | 20.5 | 0.11 |
| polarC18 | 325.216 | 129.8475 | 20.5 | 0.11 |
| polarC18 | 325.216 | 62.2626 | 20.5 | 0.11 |
| polarC18 | 325.216 | 67.5455 | 20.5 | 0.11 |
| polarC18 | 325.216 | 201.4205 | 20.5 | 0.11 |
| polarC18 | 325.216 | 149.0098 | 20.5 | 0.11 |
| polarC18 | 325.216 | 63.5671 | 20.5 | 0.11 |
| polarC18 | 325.216 | 105.4332 | 20.5 | 0.11 |
| polarC18 | 325.216 | 150.8477 | 20.5 | 0.11 |
| polarC18 | 325.216 | 50.4584 | 20.5 | 0.11 |
| polarC18 | 325.216 | 85.5443 | 20.5 | 0.11 |
| polarC18 | 325.216 | 156.2951 | 20.5 | 0.11 |
| polarC18 | 325.216 | 135.5132 | 20.5 | 0.11 |
| polarC18 | 325.216 | 80.6945 | 20.5 | 0.11 |
| polarC18 | 325.216 | 110.2225 | 20.5 | 0.11 |
| polarC18 | 325.216 | 97.0579 | 20.5 | 0.11 |
| polarC18 | 325.216 | 50.8491 | 20.5 | 0.11 |
| polarC18 | 325.216 | 162.4526 | 20.5 | 0.11 |
| polarC18 | 325.216 | 298.2768 | 20.5 | 0.11 |
| polarC18 | 325.216 | 276.7625 | 20.5 | 0.11 |
| polarC18 | 325.216 | 93.2406 | 20.5 | 0.11 |
| polarC18 | 325.216 | 106.2953 | 20.5 | 0.11 |
| polarC18 | 325.216 | 152.8398 | 20.5 | 0.11 |
| polarC18 | 325.216 | 63.9657 | 20.5 | 0.11 |
| polarC18 | 325.216 | 95.1912 | 20.5 | 0.11 |
| polarC18 | 325.216 | 112.4565 | 20.5 | 0.11 |
| polarC18 | 325.216 | 87.5579 | 20.5 | 0.11 |
| polarC18 | 325.216 | 332.2144 | 20.5 | 0.11 |
| polarC18 | 325.216 | 139.9866 | 20.5 | 0.11 |
| polarC18 | 325.216 | 75.251 | 20.5 | 0.11 |

| | | | | |
|---|---|---|---|---|
| polarC18 | 325.216 | 86.0011 | 20.4 | 0.11 |
| polarC18 | 325.216 | 56.6581 | 20.4 | 0.11 |
| polarC18 | 325.216 | 89.3055 | 20.4 | 0.11 |
| polarC18 | 325.216 | 60.2698 | 20.4 | 0.11 |
| polarC18 | 325.216 | 55.4413 | 20.4 | 0.11 |
| polarC18 | 325.216 | 50.4471 | 20.4 | 0.11 |
| polarC18 | 325.216 | 58.5375 | 20.4 | 0.1 |
| polarC18 | 325.216 | 228.7194 | 20.4 | 0.1 |
| polarC18 | 325.216 | 94.7746 | 20.4 | 0.1 |
| polarC18 | 325.216 | 111.2622 | 20.4 | 0.1 |
| polarC18 | 325.216 | 55.6999 | 20.4 | 0.1 |
| polarC18 | 325.216 | 198.7436 | 20.4 | 0.1 |
| polarC18 | 325.216 | 62.0936 | 20.4 | 0.1 |
| polarC18 | 325.216 | 50.9796 | 20.4 | 0.1 |
| polarC18 | 325.216 | 76.4275 | 20.4 | 0.1 |
| polarC18 | 325.216 | 85.1338 | 20.4 | 0.1 |
| polarC18 | 325.216 | 131.3419 | 20.4 | 0.1 |
| polarC18 | 325.216 | 85.7014 | 20.4 | 0.1 |
| polarC18 | 325.216 | 79.5989 | 20.4 | 0.1 |
| polarC18 | 325.216 | 119.9871 | 20.4 | 0.1 |
| polarC18 | 325.216 | 95.7917 | 20.4 | 0.1 |
| polarC18 | 325.216 | 58.5713 | 20.4 | 0.1 |
| polarC18 | 325.216 | 68.7371 | 20.4 | 0.1 |
| polarC18 | 325.216 | 162.5847 | 20.4 | 0.1 |
| polarC18 | 325.216 | 146.4893 | 20.4 | 0.1 |
| polarC18 | 325.216 | 71.4914 | 20.4 | 0.1 |
| polarC18 | 325.216 | 122.417 | 20.4 | 0.1 |
| polarC18 | 325.216 | 203.9987 | 20.4 | 0.1 |
| polarC18 | 325.216 | 149.81 | 20.4 | 0.1 |
| polarC18 | 325.216 | 52.087 | 20.4 | 0.1 |
| polarC18 | 325.216 | 134.3264 | 20.4 | 0.1 |
| polarC18 | 325.216 | 249.1201 | 20.4 | 0.1 |
| polarC18 | 325.216 | 142.3667 | 20.4 | 0.1 |
| polarC18 | 325.216 | 72.1337 | 20.4 | 0.1 |
| polarC18 | 325.216 | 115.7273 | 20.4 | 0.1 |
| polarC18 | 325.216 | 65.7327 | 20.4 | 0.1 |
| polarC18 | 325.216 | 237.1541 | 20.3 | 0.1 |
| polarC18 | 325.216 | 111.489 | 20.3 | 0.1 |
| polarC18 | 325.216 | 64.13 | 20.3 | 0.1 |
| polarC18 | 325.216 | 121.115 | 20.3 | 0.1 |
| polarC18 | 325.216 | 233.9655 | 20.3 | 0.1 |
| polarC18 | 325.216 | 56.4778 | 20.3 | 0.1 |

| | | | | |
|---|---|---|---|---|
| polarC18 | 325.216 | 215.1679 | 20.3 | 0.1 |
| polarC18 | 325.216 | 137.5591 | 20.3 | 0.1 |
| polarC18 | 325.216 | 66.8577 | 20.3 | 0.1 |
| polarC18 | 325.216 | 76.0456 | 20.3 | 0.1 |
| polarC18 | 325.216 | 68.9738 | 20.3 | 0.1 |
| polarC18 | 325.216 | 53.821 | 20.3 | 0.1 |
| polarC18 | 325.216 | 66.1215 | 20.3 | 0.1 |
| polarC18 | 325.216 | 60.8296 | 20.3 | 0.1 |
| polarC18 | 325.216 | 148.0238 | 20.3 | 0.1 |
| polarC18 | 325.216 | 52.423 | 20.3 | 0.1 |
| polarC18 | 325.216 | 211.5538 | 20.3 | 0.1 |
| polarC18 | 325.216 | 88.4446 | 20.3 | 0.1 |
| polarC18 | 325.216 | 51.5666 | 20.3 | 0.1 |
| polarC18 | 325.216 | 116.4752 | 20.3 | 0.1 |
| polarC18 | 325.216 | 90.4709 | 20.3 | 0.1 |
| polarC18 | 325.216 | 84.889 | 20.3 | 0.1 |
| polarC18 | 325.216 | 59.8875 | 20.3 | 0.1 |
| polarC18 | 325.216 | 76.6541 | 20.3 | 0.1 |
| polarC18 | 325.216 | 79.3295 | 20.3 | 0.1 |
| polarC18 | 325.216 | 334.1862 | 20.3 | 0.1 |
| polarC18 | 325.216 | 219.7054 | 20.3 | 0.1 |
| polarC18 | 325.216 | 84.8421 | 20.3 | 0.1 |
| polarC18 | 325.216 | 244.0988 | 20.3 | 0.1 |
| polarC18 | 325.216 | 51.9344 | 20.3 | 0.1 |
| polarC18 | 325.216 | 83.7558 | 20.3 | 0.1 |
| polarC18 | 325.216 | 78.338 | 20.3 | 0.1 |
| polarC18 | 325.216 | 62.3948 | 20.3 | 0.1 |
| polarC18 | 325.216 | 63.6627 | 20.2 | 0.1 |
| polarC18 | 325.216 | 72.7665 | 20.2 | 0.1 |
| polarC18 | 325.216 | 199.3061 | 20.2 | 0.1 |
| polarC18 | 325.216 | 134.1266 | 20.2 | 0.1 |
| polarC18 | 325.216 | 52.5778 | 20.2 | 0.1 |
| polarC18 | 325.216 | 52.7701 | 20.2 | 0.1 |
| polarC18 | 325.216 | 61.6409 | 20.2 | 0.1 |
| polarC18 | 325.216 | 126.1148 | 20.2 | 0.1 |
| polarC18 | 325.216 | 305.4587 | 20.2 | 0.1 |
| polarC18 | 325.216 | 78.3071 | 20.2 | 0.1 |
| polarC18 | 325.216 | 59.808 | 20.2 | 0.1 |
| polarC18 | 325.216 | 131.8095 | 20.2 | 0.1 |
| polarC18 | 325.216 | 51.7873 | 20.2 | 0.1 |
| polarC18 | 325.216 | 126.0945 | 20.2 | 0.1 |
| polarC18 | 325.216 | 267.9324 | 20.2 | 0.1 |

| | | | | |
|---|---|---|---|---|
| polarC18 | 325.216 | 52.523 | 20.2 | 0.1 |
| polarC18 | 325.216 | 93.9821 | 20.2 | 0.1 |
| polarC18 | 325.216 | 82.0026 | 20.2 | 0.1 |
| polarC18 | 325.216 | 78.5499 | 20.2 | 0.1 |
| polarC18 | 325.216 | 61.3987 | 20.2 | 0.1 |
| polarC18 | 325.216 | 148.844 | 20.2 | 0.1 |
| polarC18 | 325.216 | 71.8741 | 20.2 | 0.1 |
| polarC18 | 325.216 | 138.7502 | 20.2 | 0.1 |
| polarC18 | 325.216 | 138.4774 | 20.2 | 0.1 |
| polarC18 | 325.216 | 71.3011 | 20.1 | 0.1 |
| polarC18 | 325.216 | 58.7456 | 20.1 | 0.1 |
| polarC18 | 325.216 | 82.5734 | 20.1 | 0.1 |
| polarC18 | 325.216 | 72.0242 | 20.1 | 0.1 |
| polarC18 | 325.216 | 194.0549 | 20.1 | 0.1 |
| polarC18 | 325.216 | 58.0031 | 20.1 | 0.1 |
| polarC18 | 325.216 | 53.7943 | 20.1 | 0.1 |
| polarC18 | 325.216 | 106.9614 | 20.1 | 0.1 |
| polarC18 | 325.216 | 144.9374 | 20.1 | 0.1 |
| polarC18 | 325.216 | 72.0374 | 20.1 | 0.1 |
| polarC18 | 325.216 | 312.3008 | 20.1 | 0.1 |
| polarC18 | 325.216 | 84.4784 | 20.1 | 0.1 |
| polarC18 | 325.216 | 104.567 | 20.1 | 0.1 |
| polarC18 | 325.216 | 308.0396 | 20.1 | 0.1 |
| polarC18 | 325.216 | 63.2537 | 20.1 | 0.1 |
| polarC18 | 325.216 | 185.521 | 20.1 | 0.1 |
| polarC18 | 325.216 | 74.4844 | 20.1 | 0.1 |
| polarC18 | 325.216 | 96.3546 | 20.1 | 0.1 |
| polarC18 | 325.216 | 275.7168 | 20.1 | 0.1 |
| polarC18 | 325.216 | 124.8628 | 20.1 | 0.1 |
| polarC18 | 325.216 | 50.5188 | 20.1 | 0.1 |
| polarC18 | 325.216 | 197.1083 | 20.1 | 0.1 |
| polarC18 | 325.216 | 51.6458 | 20 | 0.1 |
| polarC18 | 325.216 | 84.0707 | 20 | 0.1 |
| polarC18 | 325.216 | 51.1156 | 20 | 0.1 |
| polarC18 | 325.216 | 80.8785 | 20 | 0.1 |
| polarC18 | 325.216 | 101.7993 | 20 | 0.1 |
| polarC18 | 325.216 | 55.4968 | 20 | 0.1 |
| polarC18 | 325.216 | 95.9699 | 20 | 0.1 |
| polarC18 | 325.216 | 228.9557 | 20 | 0.1 |
| polarC18 | 325.216 | 89.4529 | 20 | 0.1 |
| polarC18 | 325.216 | 142.4197 | 20 | 0.1 |
| polarC18 | 325.216 | 56.129 | 20 | 0.1 |

| | | | | |
|---|---|---|---|---|
| polarC18 | 325.216 | 56.2072 | 20 | 0.1 |
| polarC18 | 325.216 | 165.8073 | 19.9 | 0.1 |
| polarC18 | 325.216 | 267.5598 | 19.9 | 0.1 |
| polarC18 | 325.216 | 63.8191 | 19.9 | 0.1 |
| polarC18 | 325.216 | 93.9188 | 19.9 | 0.1 |
| polarC18 | 325.216 | 169.957 | 19.9 | 0.1 |
| polarC18 | 325.216 | 174.7582 | 19.9 | 0.1 |
| polarC18 | 325.216 | 65.6695 | 19.9 | 0.1 |
| polarC18 | 325.216 | 65.1976 | 19.9 | 0.1 |
| polarC18 | 325.216 | 306.9652 | 19.9 | 0.1 |
| polarC18 | 325.216 | 88.0892 | 19.9 | 0.1 |
| polarC18 | 325.216 | 77.7536 | 19.9 | 0.1 |
| polarC18 | 325.216 | 80.3156 | 19.9 | 0.1 |
| polarC18 | 325.216 | 116.2432 | 19.9 | 0.1 |
| polarC18 | 325.216 | 72.7376 | 19.9 | 0.1 |
| polarC18 | 325.216 | 56.6438 | 19.8 | 0.1 |
| polarC18 | 325.216 | 69.5329 | 19.8 | 0.1 |
| polarC18 | 325.216 | 156.8058 | 19.8 | 0.1 |
| polarC18 | 325.216 | 149.1672 | 19.8 | 0.1 |
| polarC18 | 325.216 | 55.8619 | 19.8 | 0.1 |
| polarC18 | 325.216 | 112.6083 | 19.8 | 0.1 |
| polarC18 | 325.216 | 50.9296 | 19.8 | 0.1 |
| polarC18 | 325.216 | 86.5873 | 19.8 | 0.1 |
| polarC18 | 325.216 | 103.9077 | 19.8 | 0.1 |
| polarC18 | 325.216 | 235.823 | 19.8 | 0.1 |
| polarC18 | 325.216 | 155.559 | 19.7 | 0.1 |
| polarC18 | 325.216 | 276.3771 | 19.7 | 0.1 |
| polarC18 | 325.216 | 143.7841 | 19.7 | 0.1 |
| polarC18 | 325.216 | 61.0204 | 19.7 | 0.1 |
| polarC18 | 325.216 | 303.9977 | 19.7 | 0.1 |
| polarC18 | 325.216 | 107.4521 | 19.7 | 0.1 |
| polarC18 | 325.216 | 60.2367 | 19.7 | 0.1 |
| polarC18 | 325.216 | 91.474 | 19.7 | 0.1 |
| polarC18 | 325.216 | 65.3417 | 19.7 | 0.1 |
| polarC18 | 325.216 | 85.2062 | 19.7 | 0.1 |
| polarC18 | 325.216 | 161.075 | 19.6 | 0.1 |
| polarC18 | 325.216 | 56.8363 | 19.6 | 0.1 |
| polarC18 | 325.216 | 76.1022 | 19.6 | 0.1 |
| polarC18 | 325.216 | 57.251 | 19.6 | 0.1 |
| polarC18 | 325.216 | 331.1331 | 19.6 | 0.1 |
| polarC18 | 325.216 | 135.6687 | 19.6 | 0.1 |
| polarC18 | 325.216 | 70.6907 | 19.6 | 0.1 |

| | | | | |
|---|---|---|---|---|
| polarC18 | 325.216 | 160.0462 | 19.6 | 0.1 |
| polarC18 | 325.216 | 314.3259 | 19.6 | 0.1 |
| polarC18 | 325.216 | 65.963 | 19.6 | 0.1 |
| polarC18 | 325.216 | 60.3597 | 19.6 | 0.1 |
| polarC18 | 325.216 | 167.4264 | 19.6 | 0.1 |
| polarC18 | 325.216 | 62.4521 | 19.6 | 0.1 |
| polarC18 | 325.216 | 112.085 | 19.6 | 0.1 |
| polarC18 | 325.216 | 326.8509 | 19.5 | 0.1 |
| polarC18 | 325.216 | 59.708 | 19.5 | 0.1 |
| polarC18 | 325.216 | 185.6088 | 19.5 | 0.1 |
| polarC18 | 325.216 | 71.3293 | 19.5 | 0.1 |
| polarC18 | 325.216 | 128.5435 | 19.5 | 0.1 |
| polarC18 | 325.216 | 86.124 | 19.5 | 0.1 |
| polarC18 | 325.216 | 113.3652 | 19.5 | 0.1 |
| polarC18 | 325.216 | 84.1505 | 19.4 | 0.1 |
| polarC18 | 325.216 | 160.957 | 19.4 | 0.1 |
| polarC18 | 325.216 | 184.3552 | 19.4 | 0.1 |
| polarC18 | 325.216 | 88.1081 | 19.4 | 0.1 |
| polarC18 | 325.216 | 150.659 | 19.4 | 0.1 |
| polarC18 | 325.216 | 50.5125 | 19.4 | 0.1 |
| polarC18 | 325.216 | 148.1794 | 19.4 | 0.1 |
| polarC18 | 325.216 | 70.237 | 19.3 | 0.1 |
| polarC18 | 325.216 | 100.4031 | 19.3 | 0.1 |
| polarC18 | 325.216 | 123.6175 | 19.3 | 0.1 |
| polarC18 | 325.216 | 100.6644 | 19.2 | 0.1 |
| polarC18 | 325.216 | 87.7874 | 19.2 | 0.1 |
| polarC18 | 325.216 | 99.3558 | 19.1 | 0.1 |
| polarC18 | 325.216 | 56.3096 | 19.1 | 0.1 |
| polarC18 | 325.216 | 75.6863 | 19.1 | 0.1 |
| polarC18 | 325.216 | 64.3661 | 19 | 0.1 |
| polarC18 | 325.216 | 65.4795 | 19 | 0.1 |
| polarC18 | 325.216 | 163.3769 | 19 | 0.1 |
| polarC18 | 325.216 | 171.0578 | 18.9 | 0.1 |
| polarC18 | 325.216 | 118.721 | 18.7 | 0.1 |
| polarC18 | 325.216 | 139.0665 | 18.6 | 0.1 |
| polarC18 | 325.216 | 75.6641 | 18.5 | 0.1 |
| polarC18 | 325.216 | 68.8722 | 18.5 | 0.1 |
| polarC18 | 325.216 | 60.9696 | 18.5 | 0.1 |
| polarC18 | 325.216 | 117.8974 | 18.1 | 0.09 |
| polarC18 | 468.308 | 283.1749 | 8882.4 | 100 |
| polarC18 | 468.308 | 89.0596 | 7964.2 | 89.66 |
| polarC18 | 468.308 | 133.0859 | 2947.1 | 33.18 |

| | | | | |
|---|---|---|---|---|
| polarC18 | 468.308 | 184.0732 | 1755.1 | 19.76 |
| polarC18 | 468.308 | 104.107 | 1394.6 | 15.7 |
| polarC18 | 468.308 | 468.3089 | 1263.5 | 14.22 |
| polarC18 | 468.308 | 228.2193 | 1160.8 | 13.07 |
| polarC18 | 468.308 | 451.3631 | 1137.4 | 12.81 |
| polarC18 | 468.308 | 177.112 | 607.3 | 6.84 |
| polarC18 | 468.308 | 86.0965 | 511 | 5.75 |
| polarC18 | 468.308 | 57.0698 | 400.8 | 4.51 |
| polarC18 | 468.308 | 410.1674 | 349.5 | 3.93 |
| polarC18 | 468.308 | 71.0856 | 207.8 | 2.34 |
| polarC18 | 468.308 | 125.0001 | 175.8 | 1.98 |
| polarC18 | 468.308 | 80.9478 | 153.2 | 1.72 |
| polarC18 | 468.308 | 280.6759 | 113.5 | 1.28 |
| polarC18 | 468.308 | 59.0491 | 105.2 | 1.18 |
| polarC18 | 468.308 | 479.8896 | 89.8 | 1.01 |
| polarC18 | 468.308 | 51.5436 | 86.2 | 0.97 |
| polarC18 | 468.308 | 141.7702 | 73.2 | 0.82 |
| polarC18 | 468.308 | 381.6603 | 72.7 | 0.82 |
| polarC18 | 468.308 | 426.806 | 70.9 | 0.8 |
| polarC18 | 468.308 | 97.8147 | 68.9 | 0.78 |
| polarC18 | 468.308 | 290.5752 | 68.8 | 0.77 |
| polarC18 | 468.308 | 78.3357 | 68.4 | 0.77 |
| polarC18 | 468.308 | 489.6871 | 68.3 | 0.77 |
| polarC18 | 468.308 | 307.7163 | 67.5 | 0.76 |
| polarC18 | 468.308 | 213.469 | 67.3 | 0.76 |
| polarC18 | 468.308 | 207.2421 | 67 | 0.75 |
| polarC18 | 468.308 | 134.1093 | 66.9 | 0.75 |
| polarC18 | 468.308 | 173.2786 | 66.6 | 0.75 |
| polarC18 | 468.308 | 137.6037 | 66 | 0.74 |
| polarC18 | 468.308 | 142.861 | 65.9 | 0.74 |
| polarC18 | 468.308 | 145.353 | 65.8 | 0.74 |
| polarC18 | 468.308 | 50.8101 | 65.5 | 0.74 |
| polarC18 | 468.308 | 164.2065 | 65.4 | 0.74 |
| polarC18 | 468.308 | 362.4556 | 65.2 | 0.73 |
| polarC18 | 468.308 | 51.5706 | 64.9 | 0.73 |
| polarC18 | 468.308 | 66.3185 | 64.8 | 0.73 |
| polarC18 | 468.308 | 303.7537 | 64.4 | 0.72 |
| polarC18 | 468.308 | 370.1781 | 63.5 | 0.71 |
| polarC18 | 468.308 | 400.5625 | 63.3 | 0.71 |
| polarC18 | 468.308 | 168.7208 | 63.1 | 0.71 |
| polarC18 | 468.308 | 117.252 | 63 | 0.71 |
| polarC18 | 468.308 | 240.5496 | 62.7 | 0.71 |

| | | | | |
|---|---|---|---|---|
| polarC18 | 468.308 | 71.9454 | 61.3 | 0.69 |
| polarC18 | 468.308 | 84.0175 | 60.8 | 0.68 |
| polarC18 | 468.308 | 135.5312 | 60.3 | 0.68 |
| polarC18 | 468.308 | 203.3307 | 60.3 | 0.68 |
| polarC18 | 468.308 | 121.3603 | 60.1 | 0.68 |
| polarC18 | 468.308 | 113.3086 | 59.1 | 0.67 |
| polarC18 | 468.308 | 475.9898 | 58.3 | 0.66 |
| polarC18 | 468.308 | 93.9914 | 53.5 | 0.6 |
| polarC18 | 468.308 | 364.6965 | 52.5 | 0.59 |
| polarC18 | 468.308 | 169.4961 | 51.6 | 0.58 |
| polarC18 | 468.308 | 160.4326 | 51.4 | 0.58 |
| polarC18 | 468.308 | 223.3113 | 51.2 | 0.58 |
| polarC18 | 468.308 | 496.4617 | 50.9 | 0.57 |
| polarC18 | 468.308 | 144.4312 | 50.8 | 0.57 |
| polarC18 | 468.308 | 243.0068 | 50.8 | 0.57 |
| polarC18 | 468.308 | 93.4151 | 50.8 | 0.57 |
| polarC18 | 468.308 | 190.5077 | 50.8 | 0.57 |
| polarC18 | 468.308 | 78.9471 | 50.7 | 0.57 |
| polarC18 | 468.308 | 85.101 | 50.3 | 0.57 |
| polarC18 | 468.308 | 437.7698 | 50.3 | 0.57 |
| polarC18 | 468.308 | 357.0314 | 50.1 | 0.56 |
| polarC18 | 468.308 | 195.1311 | 50.1 | 0.56 |
| polarC18 | 468.308 | 93.171 | 50 | 0.56 |
| polarC18 | 468.308 | 93.9086 | 49.9 | 0.56 |
| polarC18 | 468.308 | 89.0658 | 49.8 | 0.56 |
| polarC18 | 468.308 | 102.8474 | 49.8 | 0.56 |
| polarC18 | 468.308 | 374.6046 | 49.2 | 0.55 |
| polarC18 | 468.308 | 492.7782 | 49.1 | 0.55 |
| polarC18 | 468.308 | 279.8493 | 49.1 | 0.55 |
| polarC18 | 468.308 | 484.3742 | 49 | 0.55 |
| polarC18 | 468.308 | 160.4057 | 48.9 | 0.55 |
| polarC18 | 468.308 | 227.0504 | 48.6 | 0.55 |
| polarC18 | 468.308 | 152.4978 | 48.5 | 0.55 |
| polarC18 | 468.308 | 168.5314 | 48.5 | 0.55 |
| polarC18 | 468.308 | 233.3055 | 48.5 | 0.55 |
| polarC18 | 468.308 | 390.2555 | 48.4 | 0.55 |
| polarC18 | 468.308 | 378.2283 | 48.1 | 0.54 |
| polarC18 | 468.308 | 326.9976 | 47.8 | 0.54 |
| polarC18 | 468.308 | 118.1115 | 47.7 | 0.54 |
| polarC18 | 468.308 | 216.3199 | 47.6 | 0.54 |
| polarC18 | 468.308 | 227.5939 | 47.6 | 0.54 |
| polarC18 | 468.308 | 301.0819 | 47.6 | 0.54 |

| | | | | |
|---|---|---|---|---|
| polarC18 | 468.308 | 107.6173 | 47.6 | 0.54 |
| polarC18 | 468.308 | 71.3518 | 47.5 | 0.54 |
| polarC18 | 468.308 | 61.6549 | 47.5 | 0.53 |
| polarC18 | 468.308 | 122.8764 | 47.4 | 0.53 |
| polarC18 | 468.308 | 97.5332 | 47.4 | 0.53 |
| polarC18 | 468.308 | 419.7501 | 47.3 | 0.53 |
| polarC18 | 468.308 | 348.395 | 47.3 | 0.53 |
| polarC18 | 468.308 | 421.2486 | 47.3 | 0.53 |
| polarC18 | 468.308 | 109.5005 | 47.2 | 0.53 |
| polarC18 | 468.308 | 67.1171 | 47.1 | 0.53 |
| polarC18 | 468.308 | 486.5939 | 47 | 0.53 |
| polarC18 | 468.308 | 146.3367 | 47 | 0.53 |
| polarC18 | 468.308 | 345.6018 | 47 | 0.53 |
| polarC18 | 468.308 | 213.1178 | 46.9 | 0.53 |
| polarC18 | 468.308 | 422.008 | 46.9 | 0.53 |
| polarC18 | 468.308 | 161.5648 | 46.9 | 0.53 |
| polarC18 | 468.308 | 152.3998 | 46.9 | 0.53 |
| polarC18 | 468.308 | 458.5906 | 46.7 | 0.53 |
| polarC18 | 468.308 | 139.7577 | 46.6 | 0.52 |
| polarC18 | 468.308 | 139.5703 | 46.6 | 0.52 |
| polarC18 | 468.308 | 319.0518 | 46.6 | 0.52 |
| polarC18 | 468.308 | 160.2959 | 46.5 | 0.52 |
| polarC18 | 468.308 | 92.9494 | 46.5 | 0.52 |
| polarC18 | 468.308 | 65.8429 | 46.5 | 0.52 |
| polarC18 | 468.308 | 204.9877 | 46.5 | 0.52 |
| polarC18 | 468.308 | 432.9211 | 46.5 | 0.52 |
| polarC18 | 468.308 | 491.1452 | 46.5 | 0.52 |
| polarC18 | 468.308 | 87.4596 | 46.4 | 0.52 |
| polarC18 | 468.308 | 108.0701 | 46.4 | 0.52 |
| polarC18 | 468.308 | 109.159 | 46.4 | 0.52 |
| polarC18 | 468.308 | 82.3276 | 46.3 | 0.52 |
| polarC18 | 468.308 | 57.7076 | 46.2 | 0.52 |
| polarC18 | 468.308 | 397.7398 | 46.1 | 0.52 |
| polarC18 | 468.308 | 439.4471 | 46.1 | 0.52 |
| polarC18 | 468.308 | 149.7686 | 46.1 | 0.52 |
| polarC18 | 468.308 | 204.8489 | 46 | 0.52 |
| polarC18 | 468.308 | 96.1677 | 46 | 0.52 |
| polarC18 | 468.308 | 142.8781 | 46 | 0.52 |
| polarC18 | 468.308 | 50.1597 | 46 | 0.52 |
| polarC18 | 468.308 | 409.999 | 46 | 0.52 |
| polarC18 | 468.308 | 226.4651 | 46 | 0.52 |
| polarC18 | 468.308 | 463.9489 | 46 | 0.52 |

| | | | | |
|---|---|---|---|---|
| polarC18 | 468.308 | 74.9362 | 45.9 | 0.52 |
| polarC18 | 468.308 | 93.8979 | 45.9 | 0.52 |
| polarC18 | 468.308 | 81.4105 | 45.9 | 0.52 |
| polarC18 | 468.308 | 273.4173 | 45.8 | 0.52 |
| polarC18 | 468.308 | 52.5471 | 45.8 | 0.52 |
| polarC18 | 468.308 | 195.9011 | 45.8 | 0.52 |
| polarC18 | 468.308 | 128.5152 | 45.7 | 0.51 |
| polarC18 | 468.308 | 94.4213 | 45.7 | 0.51 |
| polarC18 | 468.308 | 151.9004 | 45.7 | 0.51 |
| polarC18 | 468.308 | 60.5444 | 45.7 | 0.51 |
| polarC18 | 468.308 | 79.1798 | 45.6 | 0.51 |
| polarC18 | 468.308 | 186.7612 | 45.5 | 0.51 |
| polarC18 | 468.308 | 204.4629 | 45.5 | 0.51 |
| polarC18 | 468.308 | 83.0777 | 45.5 | 0.51 |
| polarC18 | 468.308 | 448.7148 | 45.4 | 0.51 |
| polarC18 | 468.308 | 229.1072 | 45.4 | 0.51 |
| polarC18 | 468.308 | 235.1154 | 45.3 | 0.51 |
| polarC18 | 468.308 | 215.7027 | 45.3 | 0.51 |
| polarC18 | 468.308 | 416.6144 | 45.3 | 0.51 |
| polarC18 | 468.308 | 96.3378 | 45.2 | 0.51 |
| polarC18 | 468.308 | 54.6808 | 45.2 | 0.51 |
| polarC18 | 468.308 | 290.8099 | 45.2 | 0.51 |
| polarC18 | 468.308 | 470.9421 | 45.2 | 0.51 |
| polarC18 | 468.308 | 440.3468 | 45.1 | 0.51 |
| polarC18 | 468.308 | 473.228 | 45.1 | 0.51 |
| polarC18 | 468.308 | 113.2846 | 45.1 | 0.51 |
| polarC18 | 468.308 | 80.7567 | 45.1 | 0.51 |
| polarC18 | 468.308 | 116.4777 | 45.1 | 0.51 |
| polarC18 | 468.308 | 157.5263 | 45 | 0.51 |
| polarC18 | 468.308 | 196.2272 | 45 | 0.51 |
| polarC18 | 468.308 | 205.0551 | 45 | 0.51 |
| polarC18 | 468.308 | 349.4541 | 44.9 | 0.51 |
| polarC18 | 468.308 | 308.6072 | 44.9 | 0.51 |
| polarC18 | 468.308 | 59.9166 | 44.9 | 0.51 |
| polarC18 | 468.308 | 323.9708 | 44.9 | 0.51 |
| polarC18 | 468.308 | 167.5074 | 44.8 | 0.5 |
| polarC18 | 468.308 | 428.4786 | 44.8 | 0.5 |
| polarC18 | 468.308 | 432.4682 | 44.7 | 0.5 |
| polarC18 | 468.308 | 57.3677 | 44.7 | 0.5 |
| polarC18 | 468.308 | 355.5147 | 44.7 | 0.5 |
| polarC18 | 468.308 | 261.4535 | 44.7 | 0.5 |
| polarC18 | 468.308 | 278.2474 | 44.7 | 0.5 |

| | | | | |
|---|---|---|---|---|
| polarC18 | 468.308 | 115.6318 | 44.6 | 0.5 |
| polarC18 | 468.308 | 473.753 | 44.6 | 0.5 |
| polarC18 | 468.308 | 69.4962 | 44.5 | 0.5 |
| polarC18 | 468.308 | 381.1033 | 44.5 | 0.5 |
| polarC18 | 468.308 | 80.8884 | 44.5 | 0.5 |
| polarC18 | 468.308 | 93.9387 | 44.5 | 0.5 |
| polarC18 | 468.308 | 63.6484 | 44.5 | 0.5 |
| polarC18 | 468.308 | 399.0583 | 44.5 | 0.5 |
| polarC18 | 468.308 | 84.8716 | 44.5 | 0.5 |
| polarC18 | 468.308 | 136.9619 | 44.4 | 0.5 |
| polarC18 | 468.308 | 396.9767 | 44.4 | 0.5 |
| polarC18 | 468.308 | 161.0704 | 44.4 | 0.5 |
| polarC18 | 468.308 | 51.6401 | 44.4 | 0.5 |
| polarC18 | 468.308 | 120.9272 | 44.3 | 0.5 |
| polarC18 | 468.308 | 115.4572 | 44.3 | 0.5 |
| polarC18 | 468.308 | 220.3857 | 44.3 | 0.5 |
| polarC18 | 468.308 | 182.4848 | 44.3 | 0.5 |
| polarC18 | 468.308 | 50.4408 | 44.3 | 0.5 |
| polarC18 | 468.308 | 93.956 | 44.3 | 0.5 |
| polarC18 | 468.308 | 162.1101 | 44.2 | 0.5 |
| polarC18 | 468.308 | 467.8168 | 44.2 | 0.5 |
| polarC18 | 468.308 | 254.6707 | 44.1 | 0.5 |
| polarC18 | 468.308 | 276.8522 | 44.1 | 0.5 |
| polarC18 | 468.308 | 53.2815 | 44.1 | 0.5 |
| polarC18 | 468.308 | 403.3184 | 44.1 | 0.5 |
| polarC18 | 468.308 | 118.3548 | 44.1 | 0.5 |
| polarC18 | 468.308 | 233.2323 | 44.1 | 0.5 |
| polarC18 | 468.308 | 136.7606 | 44 | 0.5 |
| polarC18 | 468.308 | 67.8306 | 44 | 0.5 |
| polarC18 | 468.308 | 136.9119 | 44 | 0.5 |
| polarC18 | 468.308 | 148.4666 | 44 | 0.5 |
| polarC18 | 468.308 | 488.4703 | 44 | 0.5 |
| polarC18 | 468.308 | 348.1687 | 44 | 0.5 |
| polarC18 | 468.308 | 122.7938 | 44 | 0.5 |
| polarC18 | 468.308 | 77.1165 | 43.9 | 0.49 |
| polarC18 | 468.308 | 253.4537 | 43.8 | 0.49 |
| polarC18 | 468.308 | 260.9344 | 43.8 | 0.49 |
| polarC18 | 468.308 | 117.9858 | 43.8 | 0.49 |
| polarC18 | 468.308 | 55.9444 | 43.7 | 0.49 |
| polarC18 | 468.308 | 104.4173 | 43.7 | 0.49 |
| polarC18 | 468.308 | 293.6887 | 43.6 | 0.49 |
| polarC18 | 468.308 | 332.028 | 43.6 | 0.49 |

| | | | | |
|---|---|---|---|---|
| polarC18 | 468.308 | 124.2238 | 43.6 | 0.49 |
| polarC18 | 468.308 | 64.8981 | 43.6 | 0.49 |
| polarC18 | 468.308 | 83.7277 | 43.6 | 0.49 |
| polarC18 | 468.308 | 76.5646 | 43.5 | 0.49 |
| polarC18 | 468.308 | 318.6827 | 43.5 | 0.49 |
| polarC18 | 468.308 | 415.9597 | 43.5 | 0.49 |
| polarC18 | 468.308 | 288.5742 | 43.5 | 0.49 |
| polarC18 | 468.308 | 60.9607 | 43.4 | 0.49 |
| polarC18 | 468.308 | 268.4425 | 43.4 | 0.49 |
| polarC18 | 468.308 | 197.2206 | 43.4 | 0.49 |
| polarC18 | 468.308 | 58.9458 | 43.4 | 0.49 |
| polarC18 | 468.308 | 338.5183 | 43.4 | 0.49 |
| polarC18 | 468.308 | 198.2605 | 43.4 | 0.49 |
| polarC18 | 468.308 | 53.7527 | 43.3 | 0.49 |
| polarC18 | 468.308 | 213.3875 | 43.2 | 0.49 |
| polarC18 | 468.308 | 58.6191 | 43.2 | 0.49 |
| polarC18 | 468.308 | 148.6126 | 43.2 | 0.49 |
| polarC18 | 468.308 | 96.3862 | 43.1 | 0.49 |
| polarC18 | 468.308 | 467.3946 | 43.1 | 0.49 |
| polarC18 | 468.308 | 86.2822 | 43.1 | 0.49 |
| polarC18 | 468.308 | 223.3397 | 43.1 | 0.49 |
| polarC18 | 468.308 | 225.1336 | 43.1 | 0.48 |
| polarC18 | 468.308 | 64.7365 | 43 | 0.48 |
| polarC18 | 468.308 | 209.7452 | 43 | 0.48 |
| polarC18 | 468.308 | 262.6724 | 43 | 0.48 |
| polarC18 | 468.308 | 50.2596 | 43 | 0.48 |
| polarC18 | 468.308 | 114.7114 | 42.9 | 0.48 |
| polarC18 | 468.308 | 449.7427 | 42.9 | 0.48 |
| polarC18 | 468.308 | 87.8737 | 42.9 | 0.48 |
| polarC18 | 468.308 | 346.7019 | 42.9 | 0.48 |
| polarC18 | 468.308 | 56.5164 | 42.9 | 0.48 |
| polarC18 | 468.308 | 156.3595 | 42.9 | 0.48 |
| polarC18 | 468.308 | 366.5897 | 42.8 | 0.48 |
| polarC18 | 468.308 | 430.7662 | 42.8 | 0.48 |
| polarC18 | 468.308 | 90.2883 | 42.8 | 0.48 |
| polarC18 | 468.308 | 494.6698 | 42.8 | 0.48 |
| polarC18 | 468.308 | 168.0971 | 42.8 | 0.48 |
| polarC18 | 468.308 | 120.8073 | 42.8 | 0.48 |
| polarC18 | 468.308 | 331.6778 | 42.7 | 0.48 |
| polarC18 | 468.308 | 143.9442 | 42.7 | 0.48 |
| polarC18 | 468.308 | 68.2104 | 42.7 | 0.48 |
| polarC18 | 468.308 | 96.0159 | 42.6 | 0.48 |

| | | | | |
|---|---|---|---|---|
| polarC18 | 468.308 | 442.7062 | 42.6 | 0.48 |
| polarC18 | 468.308 | 335.3625 | 42.6 | 0.48 |
| polarC18 | 468.308 | 101.0207 | 42.6 | 0.48 |
| polarC18 | 468.308 | 67.3241 | 42.6 | 0.48 |
| polarC18 | 468.308 | 66.2095 | 42.6 | 0.48 |
| polarC18 | 468.308 | 128.7449 | 42.6 | 0.48 |
| polarC18 | 468.308 | 156.2191 | 42.6 | 0.48 |
| polarC18 | 468.308 | 97.3583 | 42.5 | 0.48 |
| polarC18 | 468.308 | 55.0013 | 42.5 | 0.48 |
| polarC18 | 468.308 | 224.2979 | 42.5 | 0.48 |
| polarC18 | 468.308 | 183.5507 | 42.4 | 0.48 |
| polarC18 | 468.308 | 109.5218 | 42.4 | 0.48 |
| polarC18 | 468.308 | 211.6283 | 42.4 | 0.48 |
| polarC18 | 468.308 | 94.5985 | 42.4 | 0.48 |
| polarC18 | 468.308 | 106.6911 | 42.3 | 0.48 |
| polarC18 | 468.308 | 198.9791 | 42.3 | 0.48 |
| polarC18 | 468.308 | 137.1084 | 42.3 | 0.48 |
| polarC18 | 468.308 | 57.0067 | 42.3 | 0.48 |
| polarC18 | 468.308 | 99.0865 | 42.2 | 0.48 |
| polarC18 | 468.308 | 444.4602 | 42.2 | 0.48 |
| polarC18 | 468.308 | 163.7459 | 42.2 | 0.48 |
| polarC18 | 468.308 | 61.648 | 42.2 | 0.48 |
| polarC18 | 468.308 | 143.0995 | 42.2 | 0.48 |
| polarC18 | 468.308 | 233.1844 | 42.2 | 0.48 |
| polarC18 | 468.308 | 169.5823 | 42.2 | 0.47 |
| polarC18 | 468.308 | 70.3455 | 42.2 | 0.47 |
| polarC18 | 468.308 | 98.6689 | 42.2 | 0.47 |
| polarC18 | 468.308 | 54.2163 | 42.1 | 0.47 |
| polarC18 | 468.308 | 151.2391 | 42.1 | 0.47 |
| polarC18 | 468.308 | 91.843 | 42.1 | 0.47 |
| polarC18 | 468.308 | 99.1237 | 42.1 | 0.47 |
| polarC18 | 468.308 | 223.9276 | 42 | 0.47 |
| polarC18 | 468.308 | 164.3907 | 42 | 0.47 |
| polarC18 | 468.308 | 191.5789 | 42 | 0.47 |
| polarC18 | 468.308 | 112.6822 | 41.9 | 0.47 |
| polarC18 | 468.308 | 192.9945 | 41.9 | 0.47 |
| polarC18 | 468.308 | 179.9423 | 41.8 | 0.47 |
| polarC18 | 468.308 | 57.7284 | 41.8 | 0.47 |
| polarC18 | 468.308 | 356.2921 | 41.8 | 0.47 |
| polarC18 | 468.308 | 54.5734 | 41.8 | 0.47 |
| polarC18 | 468.308 | 54.5792 | 41.8 | 0.47 |
| polarC18 | 468.308 | 93.4564 | 41.8 | 0.47 |

| | | | | |
|---|---|---|---|---|
| polarC18 | 468.308 | 81.7016 | 41.7 | 0.47 |
| polarC18 | 468.308 | 91.7059 | 41.7 | 0.47 |
| polarC18 | 468.308 | 272.7967 | 41.7 | 0.47 |
| polarC18 | 468.308 | 70.9142 | 41.7 | 0.47 |
| polarC18 | 468.308 | 79.0459 | 41.7 | 0.47 |
| polarC18 | 468.308 | 395.4839 | 41.7 | 0.47 |
| polarC18 | 468.308 | 441.0963 | 41.7 | 0.47 |
| polarC18 | 468.308 | 100.9384 | 41.6 | 0.47 |
| polarC18 | 468.308 | 468.4169 | 41.6 | 0.47 |
| polarC18 | 468.308 | 382.7549 | 41.5 | 0.47 |
| polarC18 | 468.308 | 90.328 | 41.5 | 0.47 |
| polarC18 | 468.308 | 477.9263 | 41.5 | 0.47 |
| polarC18 | 468.308 | 128.7587 | 41.4 | 0.47 |
| polarC18 | 468.308 | 91.0882 | 41.4 | 0.47 |
| polarC18 | 468.308 | 89.0826 | 41.4 | 0.47 |
| polarC18 | 468.308 | 138.3547 | 41.3 | 0.47 |
| polarC18 | 468.308 | 116.4608 | 41.3 | 0.47 |
| polarC18 | 468.308 | 174.6348 | 41.3 | 0.47 |
| polarC18 | 468.308 | 65.408 | 41.3 | 0.46 |
| polarC18 | 468.308 | 345.9657 | 41.3 | 0.46 |
| polarC18 | 468.308 | 467.0703 | 41.3 | 0.46 |
| polarC18 | 468.308 | 52.2029 | 41.3 | 0.46 |
| polarC18 | 468.308 | 89.5699 | 41.2 | 0.46 |
| polarC18 | 468.308 | 105.1005 | 41.2 | 0.46 |
| polarC18 | 468.308 | 66.4319 | 41.2 | 0.46 |
| polarC18 | 468.308 | 133.7771 | 41.2 | 0.46 |
| polarC18 | 468.308 | 160.093 | 41.1 | 0.46 |
| polarC18 | 468.308 | 51.9314 | 41.1 | 0.46 |
| polarC18 | 468.308 | 78.2851 | 41.1 | 0.46 |
| polarC18 | 468.308 | 110.731 | 41.1 | 0.46 |
| polarC18 | 468.308 | 248.7196 | 41 | 0.46 |
| polarC18 | 468.308 | 171.5606 | 41 | 0.46 |
| polarC18 | 468.308 | 95.9252 | 40.9 | 0.46 |
| polarC18 | 468.308 | 104.3912 | 40.8 | 0.46 |
| polarC18 | 468.308 | 62.6087 | 40.7 | 0.46 |
| polarC18 | 468.308 | 376.2418 | 40.7 | 0.46 |
| polarC18 | 468.308 | 79.0128 | 40.7 | 0.46 |
| polarC18 | 468.308 | 80.2151 | 40.7 | 0.46 |
| polarC18 | 468.308 | 129.9944 | 40.7 | 0.46 |
| polarC18 | 468.308 | 97.7369 | 40.7 | 0.46 |
| polarC18 | 468.308 | 100.5533 | 40.7 | 0.46 |
| polarC18 | 468.308 | 358.8615 | 40.6 | 0.46 |

| | | | | |
|---|---|---|---|---|
| polarC18 | 468.308 | 189.7581 | 40.6 | 0.46 |
| polarC18 | 468.308 | 68.8598 | 40.6 | 0.46 |
| polarC18 | 468.308 | 81.1658 | 40.6 | 0.46 |
| polarC18 | 468.308 | 465.2244 | 40.5 | 0.46 |
| polarC18 | 468.308 | 71.0032 | 40.5 | 0.46 |
| polarC18 | 468.308 | 341.9874 | 40.5 | 0.46 |
| polarC18 | 468.308 | 93.8584 | 40.4 | 0.45 |
| polarC18 | 468.308 | 138.1928 | 40.3 | 0.45 |
| polarC18 | 468.308 | 93.2866 | 40.3 | 0.45 |
| polarC18 | 468.308 | 59.3566 | 40.3 | 0.45 |
| polarC18 | 468.308 | 59.9371 | 40.3 | 0.45 |
| polarC18 | 468.308 | 110.5366 | 40.2 | 0.45 |
| polarC18 | 468.308 | 82.6009 | 40.2 | 0.45 |
| polarC18 | 468.308 | 53.7606 | 40.2 | 0.45 |
| polarC18 | 468.308 | 130.5004 | 40 | 0.45 |
| polarC18 | 468.308 | 81.1329 | 40 | 0.45 |
| polarC18 | 468.308 | 194.8051 | 40 | 0.45 |
| polarC18 | 468.308 | 275.5324 | 39.9 | 0.45 |
| polarC18 | 468.308 | 129.1892 | 39.8 | 0.45 |
| polarC18 | 468.308 | 139.4124 | 39.8 | 0.45 |
| polarC18 | 468.308 | 65.6944 | 39.7 | 0.45 |
| polarC18 | 468.308 | 161.8326 | 39.7 | 0.45 |
| polarC18 | 468.308 | 56.9907 | 39.5 | 0.45 |
| polarC18 | 468.308 | 86.3734 | 39.5 | 0.44 |
| polarC18 | 468.308 | 85.2551 | 39.5 | 0.44 |
| polarC18 | 468.308 | 157.2706 | 39.2 | 0.44 |
| polarC18 | 468.308 | 177.8658 | 39.2 | 0.44 |
| polarC18 | 468.308 | 78.8951 | 39.1 | 0.44 |
| polarC18 | 468.308 | 68.2002 | 39.1 | 0.44 |
| polarC18 | 468.308 | 231.4217 | 39.1 | 0.44 |
| polarC18 | 468.308 | 145.3374 | 39 | 0.44 |
| polarC18 | 468.308 | 52.9765 | 38.8 | 0.44 |
| polarC18 | 468.308 | 88.3785 | 38.7 | 0.44 |
| polarC18 | 468.308 | 81.707 | 38.7 | 0.44 |
| polarC18 | 468.308 | 56.6588 | 38.6 | 0.43 |
| polarC18 | 468.308 | 167.0408 | 38.5 | 0.43 |
| polarC18 | 468.308 | 249.9551 | 38.3 | 0.43 |
| polarC18 | 468.308 | 175.7691 | 38 | 0.43 |
| polarC18 | 468.308 | 252.9528 | 37.9 | 0.43 |
| polarC18 | 468.308 | 149.8291 | 37.9 | 0.43 |
| polarC18 | 468.308 | 160.8902 | 37.9 | 0.43 |
| polarC18 | 468.308 | 97.8412 | 37.9 | 0.43 |

| | | | | |
|---|---|---|---|---|
| polarC18 | 468.308 | 67.1943 | 37.5 | 0.42 |
| polarC18 | 468.308 | 61.3479 | 37.4 | 0.42 |
| polarC18 | 468.308 | 56.9774 | 37 | 0.42 |
| polarC18 | 468.308 | 150.0403 | 36 | 0.41 |
| polarC18 | 468.308 | 63.5601 | 35.9 | 0.4 |
| polarC18 | 468.308 | 272.6898 | 35.3 | 0.4 |
| polarC18 | 468.308 | 357.7087 | 32.1 | 0.36 |
| polarC18 | 468.308 | 437.132 | 31.4 | 0.35 |
| polarC18 | 468.308 | 166.2728 | 31.3 | 0.35 |
| polarC18 | 468.308 | 99.5221 | 31.2 | 0.35 |
| polarC18 | 468.308 | 172.2006 | 31 | 0.35 |
| polarC18 | 468.308 | 74.6531 | 30.9 | 0.35 |
| polarC18 | 468.308 | 281.6645 | 30.7 | 0.35 |
| polarC18 | 468.308 | 148.1882 | 30.2 | 0.34 |
| polarC18 | 468.308 | 68.7266 | 30.1 | 0.34 |
| polarC18 | 468.308 | 474.2347 | 29.9 | 0.34 |
| polarC18 | 468.308 | 336.7535 | 29.7 | 0.33 |
| polarC18 | 468.308 | 452.3802 | 29.7 | 0.33 |
| polarC18 | 468.308 | 133.0745 | 29.7 | 0.33 |
| polarC18 | 468.308 | 334.43 | 29.6 | 0.33 |
| polarC18 | 468.308 | 68.4255 | 29.4 | 0.33 |
| polarC18 | 468.308 | 128.0361 | 29 | 0.33 |
| polarC18 | 468.308 | 93.9757 | 29 | 0.33 |
| polarC18 | 468.308 | 51.2626 | 28.9 | 0.33 |
| polarC18 | 468.308 | 170.7456 | 28.9 | 0.33 |
| polarC18 | 468.308 | 200.8448 | 28.8 | 0.32 |
| polarC18 | 468.308 | 275.3973 | 28.8 | 0.32 |
| polarC18 | 468.308 | 280.6201 | 28.6 | 0.32 |
| polarC18 | 468.308 | 240.1567 | 28.6 | 0.32 |
| polarC18 | 468.308 | 490.3394 | 28.6 | 0.32 |
| polarC18 | 468.308 | 469.1344 | 28.6 | 0.32 |
| polarC18 | 468.308 | 63.6343 | 28.5 | 0.32 |
| polarC18 | 468.308 | 332.2953 | 28.4 | 0.32 |
| polarC18 | 468.308 | 159.5161 | 28.4 | 0.32 |
| polarC18 | 468.308 | 162.4399 | 28.3 | 0.32 |
| polarC18 | 468.308 | 316.2664 | 28.2 | 0.32 |
| polarC18 | 468.308 | 190.2462 | 28.2 | 0.32 |
| polarC18 | 468.308 | 390.0577 | 28.2 | 0.32 |
| polarC18 | 468.308 | 145.8882 | 28.2 | 0.32 |
| polarC18 | 468.308 | 396.893 | 28.2 | 0.32 |
| polarC18 | 468.308 | 406.6169 | 28.1 | 0.32 |
| polarC18 | 468.308 | 374.9975 | 28.1 | 0.32 |

| | | | | |
|---|---|---|---|---|
| polarC18 | 468.308 | 96.4892 | 28 | 0.32 |
| polarC18 | 468.308 | 59.4152 | 28 | 0.32 |
| polarC18 | 468.308 | 101.2101 | 28 | 0.32 |
| polarC18 | 468.308 | 87.1137 | 28 | 0.32 |
| polarC18 | 468.308 | 52.4617 | 27.9 | 0.31 |
| polarC18 | 468.308 | 82.7424 | 27.9 | 0.31 |
| polarC18 | 468.308 | 454.1078 | 27.9 | 0.31 |
| polarC18 | 468.308 | 51.8802 | 27.8 | 0.31 |
| polarC18 | 468.308 | 255.7936 | 27.8 | 0.31 |
| polarC18 | 468.308 | 237.7666 | 27.8 | 0.31 |
| polarC18 | 468.308 | 83.3676 | 27.7 | 0.31 |
| polarC18 | 468.308 | 52.7809 | 27.7 | 0.31 |
| polarC18 | 468.308 | 65.3045 | 27.6 | 0.31 |
| polarC18 | 468.308 | 60.0838 | 27.6 | 0.31 |
| polarC18 | 468.308 | 120.9449 | 27.6 | 0.31 |
| polarC18 | 468.308 | 59.316 | 27.5 | 0.31 |
| polarC18 | 468.308 | 216.2687 | 27.5 | 0.31 |
| polarC18 | 468.308 | 327.6221 | 27.4 | 0.31 |
| polarC18 | 468.308 | 325.9548 | 27.3 | 0.31 |
| polarC18 | 468.308 | 104.1874 | 27.3 | 0.31 |
| polarC18 | 468.308 | 297.519 | 27.3 | 0.31 |
| polarC18 | 468.308 | 187.9342 | 27.3 | 0.31 |
| polarC18 | 468.308 | 245.2511 | 27.3 | 0.31 |
| polarC18 | 468.308 | 54.1372 | 27.1 | 0.31 |
| polarC18 | 468.308 | 78.3711 | 27.1 | 0.31 |
| polarC18 | 468.308 | 447.8441 | 27.1 | 0.31 |
| polarC18 | 468.308 | 254.2238 | 27.1 | 0.3 |
| polarC18 | 468.308 | 360.3854 | 27 | 0.3 |
| polarC18 | 468.308 | 97.3276 | 27 | 0.3 |
| polarC18 | 468.308 | 94.0168 | 27 | 0.3 |
| polarC18 | 468.308 | 296.7904 | 26.9 | 0.3 |
| polarC18 | 468.308 | 56.7938 | 26.9 | 0.3 |
| polarC18 | 468.308 | 69.5298 | 26.9 | 0.3 |
| polarC18 | 468.308 | 412.1247 | 26.9 | 0.3 |
| polarC18 | 468.308 | 481.5474 | 26.9 | 0.3 |
| polarC18 | 468.308 | 254.1056 | 26.8 | 0.3 |
| polarC18 | 468.308 | 147.0336 | 26.8 | 0.3 |
| polarC18 | 468.308 | 83.374 | 26.8 | 0.3 |
| polarC18 | 468.308 | 108.2317 | 26.8 | 0.3 |
| polarC18 | 468.308 | 111.5115 | 26.8 | 0.3 |
| polarC18 | 468.308 | 66.8591 | 26.7 | 0.3 |
| polarC18 | 468.308 | 69.1503 | 26.7 | 0.3 |

| | | | | |
|---|---|---|---|---|
| polarC18 | 468.308 | 208.6017 | 26.7 | 0.3 |
| polarC18 | 468.308 | 89.0373 | 26.7 | 0.3 |
| polarC18 | 468.308 | 99.6004 | 26.6 | 0.3 |
| polarC18 | 468.308 | 386.4689 | 26.6 | 0.3 |
| polarC18 | 468.308 | 129.8357 | 26.6 | 0.3 |
| polarC18 | 468.308 | 134.5795 | 26.6 | 0.3 |
| polarC18 | 468.308 | 334.2233 | 26.6 | 0.3 |
| polarC18 | 468.308 | 312.8355 | 26.5 | 0.3 |
| polarC18 | 468.308 | 256.2887 | 26.5 | 0.3 |
| polarC18 | 468.308 | 51.6108 | 26.5 | 0.3 |
| polarC18 | 468.308 | 128.4766 | 26.5 | 0.3 |
| polarC18 | 468.308 | 402.0283 | 26.5 | 0.3 |
| polarC18 | 468.308 | 103.0689 | 26.5 | 0.3 |
| polarC18 | 468.308 | 51.767 | 26.5 | 0.3 |
| polarC18 | 468.308 | 191.2171 | 26.5 | 0.3 |
| polarC18 | 468.308 | 84.5857 | 26.5 | 0.3 |
| polarC18 | 468.308 | 449.5302 | 26.5 | 0.3 |
| polarC18 | 468.308 | 51.4259 | 26.4 | 0.3 |
| polarC18 | 468.308 | 88.0353 | 26.4 | 0.3 |
| polarC18 | 468.308 | 93.6567 | 26.4 | 0.3 |
| polarC18 | 468.308 | 226.0786 | 26.4 | 0.3 |
| polarC18 | 468.308 | 167.1763 | 26.4 | 0.3 |
| polarC18 | 468.308 | 267.7174 | 26.4 | 0.3 |
| polarC18 | 468.308 | 56.8222 | 26.3 | 0.3 |
| polarC18 | 468.308 | 249.3371 | 26.3 | 0.3 |
| polarC18 | 468.308 | 95.0769 | 26.3 | 0.3 |
| polarC18 | 468.308 | 83.9214 | 26.3 | 0.3 |
| polarC18 | 468.308 | 147.2666 | 26.2 | 0.3 |
| polarC18 | 468.308 | 82.5855 | 26.2 | 0.3 |
| polarC18 | 468.308 | 51.0843 | 26.2 | 0.29 |
| polarC18 | 468.308 | 313.9144 | 26.2 | 0.29 |
| polarC18 | 468.308 | 163.9651 | 26.1 | 0.29 |
| polarC18 | 468.308 | 208.6959 | 26.1 | 0.29 |
| polarC18 | 468.308 | 80.5302 | 26.1 | 0.29 |
| polarC18 | 468.308 | 101.2398 | 26.1 | 0.29 |
| polarC18 | 468.308 | 120.3357 | 26.1 | 0.29 |
| polarC18 | 468.308 | 459.5123 | 26.1 | 0.29 |
| polarC18 | 468.308 | 222.8237 | 26 | 0.29 |
| polarC18 | 468.308 | 454.5822 | 26 | 0.29 |
| polarC18 | 468.308 | 57.6073 | 26 | 0.29 |
| polarC18 | 468.308 | 198.021 | 26 | 0.29 |
| polarC18 | 468.308 | 57.0507 | 26 | 0.29 |

| | | | | |
|---|---|---|---|---|
| polarC18 | 468.308 | 129.7401 | 26 | 0.29 |
| polarC18 | 468.308 | 372.6008 | 25.9 | 0.29 |
| polarC18 | 468.308 | 232.381 | 25.9 | 0.29 |
| polarC18 | 468.308 | 195.1775 | 25.9 | 0.29 |
| polarC18 | 468.308 | 87.3801 | 25.9 | 0.29 |
| polarC18 | 468.308 | 449.1236 | 25.9 | 0.29 |
| polarC18 | 468.308 | 330.9913 | 25.8 | 0.29 |
| polarC18 | 468.308 | 254.492 | 25.8 | 0.29 |
| polarC18 | 468.308 | 288.0035 | 25.8 | 0.29 |
| polarC18 | 468.308 | 53.208 | 25.8 | 0.29 |
| polarC18 | 468.308 | 67.2915 | 25.8 | 0.29 |
| polarC18 | 468.308 | 140.2939 | 25.8 | 0.29 |
| polarC18 | 468.308 | 66.8206 | 25.8 | 0.29 |
| polarC18 | 468.308 | 182.229 | 25.8 | 0.29 |
| polarC18 | 468.308 | 130.3314 | 25.8 | 0.29 |
| polarC18 | 468.308 | 142.7252 | 25.8 | 0.29 |
| polarC18 | 468.308 | 289.2348 | 25.8 | 0.29 |
| polarC18 | 468.308 | 172.9749 | 25.8 | 0.29 |
| polarC18 | 468.308 | 339.7253 | 25.8 | 0.29 |
| polarC18 | 468.308 | 218.8041 | 25.8 | 0.29 |
| polarC18 | 468.308 | 463.3775 | 25.8 | 0.29 |
| polarC18 | 468.308 | 290.9359 | 25.7 | 0.29 |
| polarC18 | 468.308 | 66.0704 | 25.7 | 0.29 |
| polarC18 | 468.308 | 286.3451 | 25.7 | 0.29 |
| polarC18 | 468.308 | 314.5164 | 25.7 | 0.29 |
| polarC18 | 468.308 | 239.5408 | 25.7 | 0.29 |
| polarC18 | 468.308 | 99.0404 | 25.7 | 0.29 |
| polarC18 | 468.308 | 398.7277 | 25.7 | 0.29 |
| polarC18 | 468.308 | 144.4119 | 25.7 | 0.29 |
| polarC18 | 468.308 | 74.3147 | 25.7 | 0.29 |
| polarC18 | 468.308 | 67.6127 | 25.7 | 0.29 |
| polarC18 | 468.308 | 128.6034 | 25.7 | 0.29 |
| polarC18 | 468.308 | 61.0432 | 25.6 | 0.29 |
| polarC18 | 468.308 | 50.7172 | 25.6 | 0.29 |
| polarC18 | 468.308 | 141.6292 | 25.6 | 0.29 |
| polarC18 | 468.308 | 368.0899 | 25.6 | 0.29 |
| polarC18 | 468.308 | 113.5201 | 25.6 | 0.29 |
| polarC18 | 468.308 | 75.0491 | 25.6 | 0.29 |
| polarC18 | 468.308 | 85.7484 | 25.6 | 0.29 |
| polarC18 | 468.308 | 111.4829 | 25.5 | 0.29 |
| polarC18 | 468.308 | 470.2924 | 25.5 | 0.29 |
| polarC18 | 468.308 | 356.0909 | 25.5 | 0.29 |

| | | | | |
|---|---|---|---|---|
| polarC18 | 468.308 | 215.3754 | 25.5 | 0.29 |
| polarC18 | 468.308 | 195.838 | 25.5 | 0.29 |
| polarC18 | 468.308 | 398.1149 | 25.5 | 0.29 |
| polarC18 | 468.308 | 169.2972 | 25.5 | 0.29 |
| polarC18 | 468.308 | 172.6938 | 25.5 | 0.29 |
| polarC18 | 468.308 | 62.1333 | 25.5 | 0.29 |
| polarC18 | 468.308 | 196.6855 | 25.5 | 0.29 |
| polarC18 | 468.308 | 125.0107 | 25.5 | 0.29 |
| polarC18 | 468.308 | 329.8387 | 25.5 | 0.29 |
| polarC18 | 468.308 | 123.9768 | 25.5 | 0.29 |
| polarC18 | 468.308 | 444.3063 | 25.5 | 0.29 |
| polarC18 | 468.308 | 67.4427 | 25.4 | 0.29 |
| polarC18 | 468.308 | 93.6073 | 25.4 | 0.29 |
| polarC18 | 468.308 | 94.392 | 25.4 | 0.29 |
| polarC18 | 468.308 | 78.9096 | 25.4 | 0.29 |
| polarC18 | 468.308 | 130.6458 | 25.4 | 0.29 |
| polarC18 | 468.308 | 285.6099 | 25.4 | 0.29 |
| polarC18 | 468.308 | 193.1118 | 25.4 | 0.29 |
| polarC18 | 468.308 | 478.6544 | 25.4 | 0.29 |
| polarC18 | 468.308 | 50.9202 | 25.4 | 0.29 |
| polarC18 | 468.308 | 75.5293 | 25.4 | 0.29 |
| polarC18 | 468.308 | 82.0102 | 25.4 | 0.29 |
| polarC18 | 468.308 | 89.7694 | 25.4 | 0.29 |
| polarC18 | 468.308 | 120.9145 | 25.4 | 0.29 |
| polarC18 | 468.308 | 57.5482 | 25.4 | 0.29 |
| polarC18 | 468.308 | 86.5936 | 25.3 | 0.29 |
| polarC18 | 468.308 | 67.1871 | 25.3 | 0.29 |
| polarC18 | 468.308 | 446.4631 | 25.3 | 0.29 |
| polarC18 | 468.308 | 51.7706 | 25.3 | 0.29 |
| polarC18 | 468.308 | 51.5904 | 25.3 | 0.29 |
| polarC18 | 468.308 | 214.7364 | 25.3 | 0.29 |
| polarC18 | 468.308 | 63.3903 | 25.3 | 0.29 |
| polarC18 | 468.308 | 344.4347 | 25.3 | 0.29 |
| polarC18 | 468.308 | 167.7369 | 25.3 | 0.28 |
| polarC18 | 468.308 | 198.6907 | 25.3 | 0.28 |
| polarC18 | 468.308 | 247.6122 | 25.3 | 0.28 |
| polarC18 | 468.308 | 380.596 | 25.3 | 0.28 |
| polarC18 | 468.308 | 458.2882 | 25.3 | 0.28 |
| polarC18 | 468.308 | 245.6794 | 25.3 | 0.28 |
| polarC18 | 468.308 | 126.3136 | 25.3 | 0.28 |
| polarC18 | 468.308 | 153.3042 | 25.3 | 0.28 |
| polarC18 | 468.308 | 94.2079 | 25.2 | 0.28 |

| | | | | |
|---|---|---|---|---|
| polarC18 | 468.308 | 214.0294 | 25.2 | 0.28 |
| polarC18 | 468.308 | 376.4102 | 25.2 | 0.28 |
| polarC18 | 468.308 | 149.2479 | 25.2 | 0.28 |
| polarC18 | 468.308 | 50.9295 | 25.2 | 0.28 |
| polarC18 | 468.308 | 60.5163 | 25.2 | 0.28 |
| polarC18 | 468.308 | 129.7996 | 25.2 | 0.28 |
| polarC18 | 468.308 | 54.9309 | 25.2 | 0.28 |
| polarC18 | 468.308 | 52.8716 | 25.2 | 0.28 |
| polarC18 | 468.308 | 123.468 | 25.2 | 0.28 |
| polarC18 | 468.308 | 75.6296 | 25.2 | 0.28 |
| polarC18 | 468.308 | 94.4532 | 25.2 | 0.28 |
| polarC18 | 468.308 | 61.3324 | 25.2 | 0.28 |
| polarC18 | 468.308 | 124.934 | 25.2 | 0.28 |
| polarC18 | 468.308 | 93.4025 | 25.1 | 0.28 |
| polarC18 | 468.308 | 215.6729 | 25.1 | 0.28 |
| polarC18 | 468.308 | 56.9091 | 25.1 | 0.28 |
| polarC18 | 468.308 | 111.2778 | 25.1 | 0.28 |
| polarC18 | 468.308 | 56.4491 | 25.1 | 0.28 |
| polarC18 | 468.308 | 360.0273 | 25.1 | 0.28 |
| polarC18 | 468.308 | 124.1259 | 25.1 | 0.28 |
| polarC18 | 468.308 | 94.1699 | 25.1 | 0.28 |
| polarC18 | 468.308 | 473.4122 | 25.1 | 0.28 |
| polarC18 | 468.308 | 497.643 | 25.1 | 0.28 |
| polarC18 | 468.308 | 132.2887 | 25.1 | 0.28 |
| polarC18 | 468.308 | 234.5492 | 25.1 | 0.28 |
| polarC18 | 468.308 | 111.2714 | 25.1 | 0.28 |
| polarC18 | 468.308 | 100.0797 | 25.1 | 0.28 |
| polarC18 | 468.308 | 226.8256 | 25.1 | 0.28 |
| polarC18 | 468.308 | 54.0242 | 25 | 0.28 |
| polarC18 | 468.308 | 90.8329 | 25 | 0.28 |
| polarC18 | 468.308 | 81.674 | 25 | 0.28 |
| polarC18 | 468.308 | 135.4722 | 25 | 0.28 |
| polarC18 | 468.308 | 50.5167 | 25 | 0.28 |
| polarC18 | 468.308 | 229.5645 | 25 | 0.28 |
| polarC18 | 468.308 | 61.4393 | 25 | 0.28 |
| polarC18 | 468.308 | 240.4882 | 25 | 0.28 |
| polarC18 | 468.308 | 261.6263 | 25 | 0.28 |
| polarC18 | 468.308 | 58.0979 | 25 | 0.28 |
| polarC18 | 468.308 | 321.9038 | 25 | 0.28 |
| polarC18 | 468.308 | 485.3089 | 25 | 0.28 |
| polarC18 | 468.308 | 71.6563 | 25 | 0.28 |
| polarC18 | 468.308 | 94.0063 | 25 | 0.28 |

| | | | | |
|---|---|---|---|---|
| polarC18 | 468.308 | 108.5446 | 25 | 0.28 |
| polarC18 | 468.308 | 71.5233 | 25 | 0.28 |
| polarC18 | 468.308 | 270.3849 | 25 | 0.28 |
| polarC18 | 468.308 | 64.5479 | 24.9 | 0.28 |
| polarC18 | 468.308 | 459.8268 | 24.9 | 0.28 |
| polarC18 | 468.308 | 463.1056 | 24.9 | 0.28 |
| polarC18 | 468.308 | 68.7415 | 24.9 | 0.28 |
| polarC18 | 468.308 | 54.1621 | 24.9 | 0.28 |
| polarC18 | 468.308 | 186.6913 | 24.9 | 0.28 |
| polarC18 | 468.308 | 280.0148 | 24.9 | 0.28 |
| polarC18 | 468.308 | 116.1455 | 24.9 | 0.28 |
| polarC18 | 468.308 | 246.5784 | 24.9 | 0.28 |
| polarC18 | 468.308 | 74.3528 | 24.9 | 0.28 |
| polarC18 | 468.308 | 121.8862 | 24.9 | 0.28 |
| polarC18 | 468.308 | 314.0404 | 24.9 | 0.28 |
| polarC18 | 468.308 | 98.9456 | 24.9 | 0.28 |
| polarC18 | 468.308 | 122.8272 | 24.9 | 0.28 |
| polarC18 | 468.308 | 81.752 | 24.9 | 0.28 |
| polarC18 | 468.308 | 52.2687 | 24.8 | 0.28 |
| polarC18 | 468.308 | 62.2059 | 24.8 | 0.28 |
| polarC18 | 468.308 | 128.7735 | 24.8 | 0.28 |
| polarC18 | 468.308 | 120.6782 | 24.8 | 0.28 |
| polarC18 | 468.308 | 57.6496 | 24.8 | 0.28 |
| polarC18 | 468.308 | 55.9506 | 24.8 | 0.28 |
| polarC18 | 468.308 | 90.099 | 24.8 | 0.28 |
| polarC18 | 468.308 | 67.0445 | 24.8 | 0.28 |
| polarC18 | 468.308 | 96.0344 | 24.8 | 0.28 |
| polarC18 | 468.308 | 128.5742 | 24.8 | 0.28 |
| polarC18 | 468.308 | 113.4592 | 24.8 | 0.28 |
| polarC18 | 468.308 | 424.2035 | 24.8 | 0.28 |
| polarC18 | 468.308 | 124.0007 | 24.8 | 0.28 |
| polarC18 | 468.308 | 68.476 | 24.8 | 0.28 |
| polarC18 | 468.308 | 188.8152 | 24.8 | 0.28 |
| polarC18 | 468.308 | 86.8356 | 24.8 | 0.28 |
| polarC18 | 468.308 | 74.3213 | 24.8 | 0.28 |
| polarC18 | 468.308 | 79.1017 | 24.8 | 0.28 |
| polarC18 | 468.308 | 183.3803 | 24.7 | 0.28 |
| polarC18 | 468.308 | 193.0262 | 24.7 | 0.28 |
| polarC18 | 468.308 | 297.01 | 24.7 | 0.28 |
| polarC18 | 468.308 | 191.5187 | 24.7 | 0.28 |
| polarC18 | 468.308 | 171.3893 | 24.7 | 0.28 |
| polarC18 | 468.308 | 130.6656 | 24.7 | 0.28 |

| | | | | |
|---|---|---|---|---|
| polarC18 | 468.308 | 263.1494 | 24.7 | 0.28 |
| polarC18 | 468.308 | 58.983 | 24.7 | 0.28 |
| polarC18 | 468.308 | 177.0306 | 24.7 | 0.28 |
| polarC18 | 468.308 | 75.6902 | 24.7 | 0.28 |
| polarC18 | 468.308 | 111.5714 | 24.7 | 0.28 |
| polarC18 | 468.308 | 183.3465 | 24.7 | 0.28 |
| polarC18 | 468.308 | 98.0928 | 24.7 | 0.28 |
| polarC18 | 468.308 | 86.622 | 24.7 | 0.28 |
| polarC18 | 468.308 | 54.0458 | 24.7 | 0.28 |
| polarC18 | 468.308 | 258.0337 | 24.7 | 0.28 |
| polarC18 | 468.308 | 408.8151 | 24.7 | 0.28 |
| polarC18 | 468.308 | 162.7262 | 24.7 | 0.28 |
| polarC18 | 468.308 | 164.6423 | 24.7 | 0.28 |
| polarC18 | 468.308 | 168.0255 | 24.6 | 0.28 |
| polarC18 | 468.308 | 186.0461 | 24.6 | 0.28 |
| polarC18 | 468.308 | 225.6966 | 24.6 | 0.28 |
| polarC18 | 468.308 | 190.4063 | 24.6 | 0.28 |
| polarC18 | 468.308 | 143.2513 | 24.6 | 0.28 |
| polarC18 | 468.308 | 488.9513 | 24.6 | 0.28 |
| polarC18 | 468.308 | 159.1994 | 24.6 | 0.28 |
| polarC18 | 468.308 | 211.7215 | 24.6 | 0.28 |
| polarC18 | 468.308 | 228.3697 | 24.6 | 0.28 |
| polarC18 | 468.308 | 54.5243 | 24.6 | 0.28 |
| polarC18 | 468.308 | 195.1543 | 24.6 | 0.28 |
| polarC18 | 468.308 | 390.8327 | 24.6 | 0.28 |
| polarC18 | 468.308 | 133.4853 | 24.6 | 0.28 |
| polarC18 | 468.308 | 187.6636 | 24.6 | 0.28 |
| polarC18 | 468.308 | 89.8647 | 24.6 | 0.28 |
| polarC18 | 468.308 | 94.9901 | 24.6 | 0.28 |
| polarC18 | 468.308 | 72.0112 | 24.6 | 0.28 |
| polarC18 | 468.308 | 119.2386 | 24.6 | 0.28 |
| polarC18 | 468.308 | 200.8961 | 24.6 | 0.28 |
| polarC18 | 468.308 | 323.6785 | 24.6 | 0.28 |
| polarC18 | 468.308 | 121.4891 | 24.6 | 0.28 |
| polarC18 | 468.308 | 105.5093 | 24.6 | 0.28 |
| polarC18 | 468.308 | 62.5819 | 24.5 | 0.28 |
| polarC18 | 468.308 | 231.0965 | 24.5 | 0.28 |
| polarC18 | 468.308 | 222.9618 | 24.5 | 0.28 |
| polarC18 | 468.308 | 63.9026 | 24.5 | 0.28 |
| polarC18 | 468.308 | 76.68 | 24.5 | 0.28 |
| polarC18 | 468.308 | 144.4744 | 24.5 | 0.28 |
| polarC18 | 468.308 | 465.7417 | 24.5 | 0.28 |

| | | | | |
|---|---|---|---|---|
| polarC18 | 468.308 | 166.2589 | 24.5 | 0.28 |
| polarC18 | 468.308 | 379.1161 | 24.5 | 0.28 |
| polarC18 | 468.308 | 336.4028 | 24.5 | 0.28 |
| polarC18 | 468.308 | 51.3303 | 24.5 | 0.28 |
| polarC18 | 468.308 | 430.4353 | 24.5 | 0.28 |
| polarC18 | 468.308 | 55.6932 | 24.5 | 0.28 |
| polarC18 | 468.308 | 82.5622 | 24.5 | 0.28 |
| polarC18 | 468.308 | 189.734 | 24.5 | 0.28 |
| polarC18 | 468.308 | 492.3781 | 24.5 | 0.28 |
| polarC18 | 468.308 | 91.3143 | 24.4 | 0.28 |
| polarC18 | 468.308 | 118.5877 | 24.4 | 0.28 |
| polarC18 | 468.308 | 63.957 | 24.4 | 0.28 |
| polarC18 | 468.308 | 100.3075 | 24.4 | 0.28 |
| polarC18 | 468.308 | 83.1629 | 24.4 | 0.28 |
| polarC18 | 468.308 | 176.0878 | 24.4 | 0.28 |
| polarC18 | 468.308 | 433.6606 | 24.4 | 0.28 |
| polarC18 | 468.308 | 97.2258 | 24.4 | 0.28 |
| polarC18 | 468.308 | 68.2991 | 24.4 | 0.27 |
| polarC18 | 468.308 | 225.5376 | 24.4 | 0.27 |
| polarC18 | 468.308 | 74.9664 | 24.4 | 0.27 |
| polarC18 | 468.308 | 449.9126 | 24.4 | 0.27 |
| polarC18 | 468.308 | 447.0723 | 24.4 | 0.27 |
| polarC18 | 468.308 | 126.8212 | 24.4 | 0.27 |
| polarC18 | 468.308 | 173.9231 | 24.4 | 0.27 |
| polarC18 | 468.308 | 253.5927 | 24.4 | 0.27 |
| polarC18 | 468.308 | 55.2312 | 24.4 | 0.27 |
| polarC18 | 468.308 | 137.9359 | 24.4 | 0.27 |
| polarC18 | 468.308 | 262.4255 | 24.4 | 0.27 |
| polarC18 | 468.308 | 99.2224 | 24.4 | 0.27 |
| polarC18 | 468.308 | 76.6902 | 24.4 | 0.27 |
| polarC18 | 468.308 | 194.4217 | 24.4 | 0.27 |
| polarC18 | 468.308 | 182.3867 | 24.4 | 0.27 |
| polarC18 | 468.308 | 213.0527 | 24.3 | 0.27 |
| polarC18 | 468.308 | 51.7786 | 24.3 | 0.27 |
| polarC18 | 468.308 | 314.6859 | 24.3 | 0.27 |
| polarC18 | 468.308 | 101.4138 | 24.3 | 0.27 |
| polarC18 | 468.308 | 459.1485 | 24.3 | 0.27 |
| polarC18 | 468.308 | 163.1164 | 24.3 | 0.27 |
| polarC18 | 468.308 | 84.2152 | 24.3 | 0.27 |
| polarC18 | 468.308 | 50.7947 | 24.3 | 0.27 |
| polarC18 | 468.308 | 235.1502 | 24.3 | 0.27 |
| polarC18 | 468.308 | 58.7509 | 24.3 | 0.27 |

| | | | | |
|---|---|---|---|---|
| polarC18 | 468.308 | 55.0867 | 24.3 | 0.27 |
| polarC18 | 468.308 | 246.0782 | 24.3 | 0.27 |
| polarC18 | 468.308 | 192.2007 | 24.3 | 0.27 |
| polarC18 | 468.308 | 233.8805 | 24.2 | 0.27 |
| polarC18 | 468.308 | 88.422 | 24.2 | 0.27 |
| polarC18 | 468.308 | 106.9308 | 24.2 | 0.27 |
| polarC18 | 468.308 | 68.0465 | 24.2 | 0.27 |
| polarC18 | 468.308 | 56.4793 | 24.2 | 0.27 |
| polarC18 | 468.308 | 206.3735 | 24.2 | 0.27 |
| polarC18 | 468.308 | 106.1127 | 24.2 | 0.27 |
| polarC18 | 468.308 | 69.1805 | 24.2 | 0.27 |
| polarC18 | 468.308 | 247.2223 | 24.2 | 0.27 |
| polarC18 | 468.308 | 156.8609 | 24.2 | 0.27 |
| polarC18 | 468.308 | 66.8947 | 24.2 | 0.27 |
| polarC18 | 468.308 | 298.348 | 24.2 | 0.27 |
| polarC18 | 468.308 | 288.4542 | 24.2 | 0.27 |
| polarC18 | 468.308 | 290.4685 | 24.2 | 0.27 |
| polarC18 | 468.308 | 189.118 | 24.2 | 0.27 |
| polarC18 | 468.308 | 90.0017 | 24.2 | 0.27 |
| polarC18 | 468.308 | 192.4487 | 24.2 | 0.27 |
| polarC18 | 468.308 | 328.0609 | 24.2 | 0.27 |
| polarC18 | 468.308 | 124.7365 | 24.2 | 0.27 |
| polarC18 | 468.308 | 54.6469 | 24.2 | 0.27 |
| polarC18 | 468.308 | 63.0473 | 24.2 | 0.27 |
| polarC18 | 468.308 | 133.6092 | 24.2 | 0.27 |
| polarC18 | 468.308 | 180.4638 | 24.2 | 0.27 |
| polarC18 | 468.308 | 51.0333 | 24.2 | 0.27 |
| polarC18 | 468.308 | 145.1436 | 24.2 | 0.27 |
| polarC18 | 468.308 | 207.3598 | 24.1 | 0.27 |
| polarC18 | 468.308 | 240.381 | 24.1 | 0.27 |
| polarC18 | 468.308 | 268.2522 | 24.1 | 0.27 |
| polarC18 | 468.308 | 53.5194 | 24.1 | 0.27 |
| polarC18 | 468.308 | 359.3844 | 24.1 | 0.27 |
| polarC18 | 468.308 | 52.8911 | 24.1 | 0.27 |
| polarC18 | 468.308 | 401.3636 | 24.1 | 0.27 |
| polarC18 | 468.308 | 67.8875 | 24.1 | 0.27 |
| polarC18 | 468.308 | 242.903 | 24.1 | 0.27 |
| polarC18 | 468.308 | 194.1325 | 24.1 | 0.27 |
| polarC18 | 468.308 | 83.0875 | 24.1 | 0.27 |
| polarC18 | 468.308 | 325.5128 | 24.1 | 0.27 |
| polarC18 | 468.308 | 88.1478 | 24.1 | 0.27 |
| polarC18 | 468.308 | 109.0916 | 24.1 | 0.27 |

| | | | | |
|---|---|---|---|---|
| polarC18 | 468.308 | 125.3882 | 24.1 | 0.27 |
| polarC18 | 468.308 | 252.1968 | 24.1 | 0.27 |
| polarC18 | 468.308 | 53.0062 | 24.1 | 0.27 |
| polarC18 | 468.308 | 179.0836 | 24.1 | 0.27 |
| polarC18 | 468.308 | 145.2145 | 24.1 | 0.27 |
| polarC18 | 468.308 | 410.435 | 24.1 | 0.27 |
| polarC18 | 468.308 | 54.5928 | 24.1 | 0.27 |
| polarC18 | 468.308 | 364.0381 | 24.1 | 0.27 |
| polarC18 | 468.308 | 59.9987 | 24.1 | 0.27 |
| polarC18 | 468.308 | 81.9008 | 24.1 | 0.27 |
| polarC18 | 468.308 | 262.9486 | 24.1 | 0.27 |
| polarC18 | 468.308 | 185.5665 | 24.1 | 0.27 |
| polarC18 | 468.308 | 193.2454 | 24.1 | 0.27 |
| polarC18 | 468.308 | 57.6151 | 24 | 0.27 |
| polarC18 | 468.308 | 56.3444 | 24 | 0.27 |
| polarC18 | 468.308 | 56.164 | 24 | 0.27 |
| polarC18 | 468.308 | 83.1197 | 24 | 0.27 |
| polarC18 | 468.308 | 420.5938 | 24 | 0.27 |
| polarC18 | 468.308 | 88.0254 | 24 | 0.27 |
| polarC18 | 468.308 | 385.0078 | 24 | 0.27 |
| polarC18 | 468.308 | 102.1909 | 24 | 0.27 |
| polarC18 | 468.308 | 298.4241 | 24 | 0.27 |
| polarC18 | 468.308 | 70.1661 | 24 | 0.27 |
| polarC18 | 468.308 | 419.3975 | 24 | 0.27 |
| polarC18 | 468.308 | 105.84 | 24 | 0.27 |
| polarC18 | 468.308 | 188.8344 | 24 | 0.27 |
| polarC18 | 468.308 | 358.4875 | 24 | 0.27 |
| polarC18 | 468.308 | 377.8837 | 24 | 0.27 |
| polarC18 | 468.308 | 317.1143 | 24 | 0.27 |
| polarC18 | 468.308 | 61.0649 | 24 | 0.27 |
| polarC18 | 468.308 | 271.5237 | 24 | 0.27 |
| polarC18 | 468.308 | 60.9135 | 24 | 0.27 |
| polarC18 | 468.308 | 89.2272 | 24 | 0.27 |
| polarC18 | 468.308 | 52.196 | 24 | 0.27 |
| polarC18 | 468.308 | 274.7576 | 24 | 0.27 |
| polarC18 | 468.308 | 209.1827 | 23.9 | 0.27 |
| polarC18 | 468.308 | 129.1648 | 23.9 | 0.27 |
| polarC18 | 468.308 | 235.0336 | 23.9 | 0.27 |
| polarC18 | 468.308 | 157.3524 | 23.9 | 0.27 |
| polarC18 | 468.308 | 67.0987 | 23.9 | 0.27 |
| polarC18 | 468.308 | 113.5087 | 23.9 | 0.27 |
| polarC18 | 468.308 | 74.7753 | 23.9 | 0.27 |

| | | | | |
|---|---|---|---|---|
| polarC18 | 468.308 | 63.1774 | 23.9 | 0.27 |
| polarC18 | 468.308 | 118.9611 | 23.9 | 0.27 |
| polarC18 | 468.308 | 69.3451 | 23.9 | 0.27 |
| polarC18 | 468.308 | 363.2612 | 23.9 | 0.27 |
| polarC18 | 468.308 | 260.0589 | 23.9 | 0.27 |
| polarC18 | 468.308 | 281.9698 | 23.9 | 0.27 |
| polarC18 | 468.308 | 189.4651 | 23.9 | 0.27 |
| polarC18 | 468.308 | 197.8023 | 23.9 | 0.27 |
| polarC18 | 468.308 | 64.8782 | 23.9 | 0.27 |
| polarC18 | 468.308 | 252.1608 | 23.9 | 0.27 |
| polarC18 | 468.308 | 75.0116 | 23.9 | 0.27 |
| polarC18 | 468.308 | 61.7305 | 23.9 | 0.27 |
| polarC18 | 468.308 | 78.2476 | 23.9 | 0.27 |
| polarC18 | 468.308 | 155.8943 | 23.9 | 0.27 |
| polarC18 | 468.308 | 297.2545 | 23.9 | 0.27 |
| polarC18 | 468.308 | 276.541 | 23.9 | 0.27 |
| polarC18 | 468.308 | 70.3955 | 23.9 | 0.27 |
| polarC18 | 468.308 | 123.7892 | 23.9 | 0.27 |
| polarC18 | 468.308 | 57.4548 | 23.9 | 0.27 |
| polarC18 | 468.308 | 58.8334 | 23.9 | 0.27 |
| polarC18 | 468.308 | 63.4825 | 23.9 | 0.27 |
| polarC18 | 468.308 | 58.3615 | 23.9 | 0.27 |
| polarC18 | 468.308 | 132.7275 | 23.9 | 0.27 |
| polarC18 | 468.308 | 117.3676 | 23.9 | 0.27 |
| polarC18 | 468.308 | 68.6797 | 23.8 | 0.27 |
| polarC18 | 468.308 | 59.3805 | 23.8 | 0.27 |
| polarC18 | 468.308 | 154.712 | 23.8 | 0.27 |
| polarC18 | 468.308 | 94.3541 | 23.8 | 0.27 |
| polarC18 | 468.308 | 258.8022 | 23.8 | 0.27 |
| polarC18 | 468.308 | 322.6155 | 23.8 | 0.27 |
| polarC18 | 468.308 | 304.0625 | 23.8 | 0.27 |
| polarC18 | 468.308 | 242.3803 | 23.8 | 0.27 |
| polarC18 | 468.308 | 161.8759 | 23.8 | 0.27 |
| polarC18 | 468.308 | 224.499 | 23.8 | 0.27 |
| polarC18 | 468.308 | 440.5525 | 23.8 | 0.27 |
| polarC18 | 468.308 | 66.792 | 23.8 | 0.27 |
| polarC18 | 468.308 | 66.3005 | 23.8 | 0.27 |
| polarC18 | 468.308 | 71.7315 | 23.8 | 0.27 |
| polarC18 | 468.308 | 111.2958 | 23.8 | 0.27 |
| polarC18 | 468.308 | 105.7287 | 23.8 | 0.27 |
| polarC18 | 468.308 | 50.6647 | 23.8 | 0.27 |
| polarC18 | 468.308 | 74.6229 | 23.8 | 0.27 |

| | | | | |
|---|---|---|---|---|
| polarC18 | 468.308 | 56.2421 | 23.8 | 0.27 |
| polarC18 | 468.308 | 53.1931 | 23.8 | 0.27 |
| polarC18 | 468.308 | 102.6712 | 23.8 | 0.27 |
| polarC18 | 468.308 | 345.2801 | 23.8 | 0.27 |
| polarC18 | 468.308 | 50.2932 | 23.7 | 0.27 |
| polarC18 | 468.308 | 88.1594 | 23.7 | 0.27 |
| polarC18 | 468.308 | 118.6729 | 23.7 | 0.27 |
| polarC18 | 468.308 | 69.4874 | 23.7 | 0.27 |
| polarC18 | 468.308 | 369.4272 | 23.7 | 0.27 |
| polarC18 | 468.308 | 128.488 | 23.7 | 0.27 |
| polarC18 | 468.308 | 97.6021 | 23.7 | 0.27 |
| polarC18 | 468.308 | 146.4601 | 23.7 | 0.27 |
| polarC18 | 468.308 | 98.7527 | 23.7 | 0.27 |
| polarC18 | 468.308 | 361.0441 | 23.7 | 0.27 |
| polarC18 | 468.308 | 277.7049 | 23.7 | 0.27 |
| polarC18 | 468.308 | 92.2926 | 23.7 | 0.27 |
| polarC18 | 468.308 | 77.1314 | 23.7 | 0.27 |
| polarC18 | 468.308 | 56.2489 | 23.7 | 0.27 |
| polarC18 | 468.308 | 178.7254 | 23.7 | 0.27 |
| polarC18 | 468.308 | 347.4785 | 23.7 | 0.27 |
| polarC18 | 468.308 | 90.7275 | 23.7 | 0.27 |
| polarC18 | 468.308 | 395.8272 | 23.7 | 0.27 |
| polarC18 | 468.308 | 117.124 | 23.7 | 0.27 |
| polarC18 | 468.308 | 85.4464 | 23.7 | 0.27 |
| polarC18 | 468.308 | 93.5114 | 23.7 | 0.27 |
| polarC18 | 468.308 | 237.8167 | 23.7 | 0.27 |
| polarC18 | 468.308 | 120.6346 | 23.7 | 0.27 |
| polarC18 | 468.308 | 92.9992 | 23.7 | 0.27 |
| polarC18 | 468.308 | 313.4465 | 23.7 | 0.27 |
| polarC18 | 468.308 | 50.5105 | 23.7 | 0.27 |
| polarC18 | 468.308 | 140.3701 | 23.7 | 0.27 |
| polarC18 | 468.308 | 162.8529 | 23.7 | 0.27 |
| polarC18 | 468.308 | 155.7334 | 23.7 | 0.27 |
| polarC18 | 468.308 | 63.2993 | 23.7 | 0.27 |
| polarC18 | 468.308 | 150.8182 | 23.7 | 0.27 |
| polarC18 | 468.308 | 268.9253 | 23.7 | 0.27 |
| polarC18 | 468.308 | 113.132 | 23.7 | 0.27 |
| polarC18 | 468.308 | 51.5591 | 23.7 | 0.27 |
| polarC18 | 468.308 | 67.977 | 23.7 | 0.27 |
| polarC18 | 468.308 | 50.4648 | 23.7 | 0.27 |
| polarC18 | 468.308 | 82.1223 | 23.6 | 0.27 |
| polarC18 | 468.308 | 62.7561 | 23.6 | 0.27 |

| | | | | |
|---|---|---|---|---|
| polarC18 | 468.308 | 339.9856 | 23.6 | 0.27 |
| polarC18 | 468.308 | 397.4931 | 23.6 | 0.27 |
| polarC18 | 468.308 | 129.2819 | 23.6 | 0.27 |
| polarC18 | 468.308 | 245.4095 | 23.6 | 0.27 |
| polarC18 | 468.308 | 224.2704 | 23.6 | 0.27 |
| polarC18 | 468.308 | 137.163 | 23.6 | 0.27 |
| polarC18 | 468.308 | 51.3323 | 23.6 | 0.27 |
| polarC18 | 468.308 | 184.5422 | 23.6 | 0.27 |
| polarC18 | 468.308 | 186.6634 | 23.6 | 0.27 |
| polarC18 | 468.308 | 56.1719 | 23.6 | 0.27 |
| polarC18 | 468.308 | 431.7456 | 23.6 | 0.27 |
| polarC18 | 468.308 | 80.1411 | 23.6 | 0.27 |
| polarC18 | 468.308 | 62.3996 | 23.6 | 0.27 |
| polarC18 | 468.308 | 125.8536 | 23.6 | 0.27 |
| polarC18 | 468.308 | 56.7733 | 23.6 | 0.27 |
| polarC18 | 468.308 | 91.2655 | 23.6 | 0.27 |
| polarC18 | 468.308 | 162.8785 | 23.6 | 0.27 |
| polarC18 | 468.308 | 54.3258 | 23.6 | 0.27 |
| polarC18 | 468.308 | 105.1601 | 23.6 | 0.27 |
| polarC18 | 468.308 | 143.5003 | 23.6 | 0.27 |
| polarC18 | 468.308 | 208.3475 | 23.6 | 0.27 |
| polarC18 | 468.308 | 77.39 | 23.6 | 0.27 |
| polarC18 | 468.308 | 127.3217 | 23.6 | 0.27 |
| polarC18 | 468.308 | 83.6942 | 23.6 | 0.27 |
| polarC18 | 468.308 | 442.5821 | 23.6 | 0.27 |
| polarC18 | 468.308 | 315.4152 | 23.6 | 0.27 |
| polarC18 | 468.308 | 161.1064 | 23.6 | 0.27 |
| polarC18 | 468.308 | 53.4389 | 23.6 | 0.27 |
| polarC18 | 468.308 | 62.8318 | 23.6 | 0.27 |
| polarC18 | 468.308 | 230.7309 | 23.6 | 0.27 |
| polarC18 | 468.308 | 73.7178 | 23.6 | 0.27 |
| polarC18 | 468.308 | 228.8926 | 23.6 | 0.27 |
| polarC18 | 468.308 | 90.7886 | 23.6 | 0.27 |
| polarC18 | 468.308 | 318.5713 | 23.6 | 0.27 |
| polarC18 | 468.308 | 90.2635 | 23.6 | 0.27 |
| polarC18 | 468.308 | 71.4895 | 23.5 | 0.27 |
| polarC18 | 468.308 | 82.1812 | 23.5 | 0.27 |
| polarC18 | 468.308 | 89.8351 | 23.5 | 0.27 |
| polarC18 | 468.308 | 55.2237 | 23.5 | 0.27 |
| polarC18 | 468.308 | 71.3438 | 23.5 | 0.27 |
| polarC18 | 468.308 | 78.3099 | 23.5 | 0.27 |
| polarC18 | 468.308 | 437.9473 | 23.5 | 0.26 |

| | | | | |
|---|---|---|---|---|
| polarC18 | 468.308 | 58.322 | 23.5 | 0.26 |
| polarC18 | 468.308 | 58.0004 | 23.5 | 0.26 |
| polarC18 | 468.308 | 349.2603 | 23.5 | 0.26 |
| polarC18 | 468.308 | 113.1603 | 23.5 | 0.26 |
| polarC18 | 468.308 | 205.5574 | 23.5 | 0.26 |
| polarC18 | 468.308 | 430.1821 | 23.5 | 0.26 |
| polarC18 | 468.308 | 221.9739 | 23.5 | 0.26 |
| polarC18 | 468.308 | 268.343 | 23.5 | 0.26 |
| polarC18 | 468.308 | 188.3119 | 23.5 | 0.26 |
| polarC18 | 468.308 | 57.0773 | 23.5 | 0.26 |
| polarC18 | 468.308 | 70.7314 | 23.5 | 0.26 |
| polarC18 | 468.308 | 139.6828 | 23.5 | 0.26 |
| polarC18 | 468.308 | 94.4166 | 23.5 | 0.26 |
| polarC18 | 468.308 | 73.0195 | 23.5 | 0.26 |
| polarC18 | 468.308 | 315.276 | 23.5 | 0.26 |
| polarC18 | 468.308 | 109.3791 | 23.5 | 0.26 |
| polarC18 | 468.308 | 331.7594 | 23.5 | 0.26 |
| polarC18 | 468.308 | 170.6534 | 23.5 | 0.26 |
| polarC18 | 468.308 | 346.7937 | 23.5 | 0.26 |
| polarC18 | 468.308 | 81.8875 | 23.5 | 0.26 |
| polarC18 | 468.308 | 76.6685 | 23.5 | 0.26 |
| polarC18 | 468.308 | 146.3499 | 23.5 | 0.26 |
| polarC18 | 468.308 | 420.498 | 23.5 | 0.26 |
| polarC18 | 468.308 | 189.1615 | 23.5 | 0.26 |
| polarC18 | 468.308 | 368.8088 | 23.5 | 0.26 |
| polarC18 | 468.308 | 111.0736 | 23.5 | 0.26 |
| polarC18 | 468.308 | 120.7606 | 23.5 | 0.26 |
| polarC18 | 468.308 | 59.9019 | 23.5 | 0.26 |
| polarC18 | 468.308 | 484.9708 | 23.5 | 0.26 |
| polarC18 | 468.308 | 58.4538 | 23.4 | 0.26 |
| polarC18 | 468.308 | 80.3438 | 23.4 | 0.26 |
| polarC18 | 468.308 | 59.5783 | 23.4 | 0.26 |
| polarC18 | 468.308 | 80.795 | 23.4 | 0.26 |
| polarC18 | 468.308 | 203.4652 | 23.4 | 0.26 |
| polarC18 | 468.308 | 58.2952 | 23.4 | 0.26 |
| polarC18 | 468.308 | 59.1283 | 23.4 | 0.26 |
| polarC18 | 468.308 | 228.9841 | 23.4 | 0.26 |
| polarC18 | 468.308 | 469.7358 | 23.4 | 0.26 |
| polarC18 | 468.308 | 261.039 | 23.4 | 0.26 |
| polarC18 | 468.308 | 59.5621 | 23.4 | 0.26 |
| polarC18 | 468.308 | 200.72 | 23.4 | 0.26 |
| polarC18 | 468.308 | 110.1536 | 23.4 | 0.26 |

| | | | | |
|---|---|---|---|---|
| polarC18 | 468.308 | 106.6219 | 23.4 | 0.26 |
| polarC18 | 468.308 | 291.4931 | 23.4 | 0.26 |
| polarC18 | 468.308 | 376.1551 | 23.4 | 0.26 |
| polarC18 | 468.308 | 306.2491 | 23.4 | 0.26 |
| polarC18 | 468.308 | 395.9559 | 23.4 | 0.26 |
| polarC18 | 468.308 | 70.1754 | 23.4 | 0.26 |
| polarC18 | 468.308 | 425.7462 | 23.4 | 0.26 |
| polarC18 | 468.308 | 155.0119 | 23.4 | 0.26 |
| polarC18 | 468.308 | 184.1906 | 23.4 | 0.26 |
| polarC18 | 468.308 | 303.3168 | 23.4 | 0.26 |
| polarC18 | 468.308 | 104.6486 | 23.4 | 0.26 |
| polarC18 | 468.308 | 280.7591 | 23.4 | 0.26 |
| polarC18 | 468.308 | 153.3896 | 23.4 | 0.26 |
| polarC18 | 468.308 | 251.2301 | 23.4 | 0.26 |
| polarC18 | 468.308 | 205.1955 | 23.4 | 0.26 |
| polarC18 | 468.308 | 51.6522 | 23.4 | 0.26 |
| polarC18 | 468.308 | 391.0264 | 23.4 | 0.26 |
| polarC18 | 468.308 | 174.8758 | 23.4 | 0.26 |
| polarC18 | 468.308 | 105.8728 | 23.4 | 0.26 |
| polarC18 | 468.308 | 247.4359 | 23.4 | 0.26 |
| polarC18 | 468.308 | 66.2428 | 23.4 | 0.26 |
| polarC18 | 468.308 | 295.4316 | 23.4 | 0.26 |
| polarC18 | 468.308 | 312.5884 | 23.4 | 0.26 |
| polarC18 | 468.308 | 58.805 | 23.4 | 0.26 |
| polarC18 | 468.308 | 61.5679 | 23.4 | 0.26 |
| polarC18 | 468.308 | 79.8768 | 23.4 | 0.26 |
| polarC18 | 468.308 | 129.5146 | 23.4 | 0.26 |
| polarC18 | 468.308 | 64.9753 | 23.4 | 0.26 |
| polarC18 | 468.308 | 495.166 | 23.4 | 0.26 |
| polarC18 | 468.308 | 407.1486 | 23.3 | 0.26 |
| polarC18 | 468.308 | 106.3686 | 23.3 | 0.26 |
| polarC18 | 468.308 | 57.7418 | 23.3 | 0.26 |
| polarC18 | 468.308 | 396.2206 | 23.3 | 0.26 |
| polarC18 | 468.308 | 136.2157 | 23.3 | 0.26 |
| polarC18 | 468.308 | 93.9308 | 23.3 | 0.26 |
| polarC18 | 468.308 | 171.0829 | 23.3 | 0.26 |
| polarC18 | 468.308 | 133.3028 | 23.3 | 0.26 |
| polarC18 | 468.308 | 394.8309 | 23.3 | 0.26 |
| polarC18 | 468.308 | 52.2647 | 23.3 | 0.26 |
| polarC18 | 468.308 | 116.6546 | 23.3 | 0.26 |
| polarC18 | 468.308 | 71.3227 | 23.3 | 0.26 |
| polarC18 | 468.308 | 190.9507 | 23.3 | 0.26 |

| | | | | |
|---|---|---|---|---|
| polarC18 | 468.308 | 148.9392 | 23.3 | 0.26 |
| polarC18 | 468.308 | 88.6957 | 23.3 | 0.26 |
| polarC18 | 468.308 | 202.4197 | 23.3 | 0.26 |
| polarC18 | 468.308 | 183.9601 | 23.3 | 0.26 |
| polarC18 | 468.308 | 209.4762 | 23.3 | 0.26 |
| polarC18 | 468.308 | 124.0712 | 23.3 | 0.26 |
| polarC18 | 468.308 | 64.5602 | 23.3 | 0.26 |
| polarC18 | 468.308 | 70.5431 | 23.3 | 0.26 |
| polarC18 | 468.308 | 406.0166 | 23.3 | 0.26 |
| polarC18 | 468.308 | 52.2893 | 23.3 | 0.26 |
| polarC18 | 468.308 | 103.9465 | 23.3 | 0.26 |
| polarC18 | 468.308 | 245.6044 | 23.3 | 0.26 |
| polarC18 | 468.308 | 89.1031 | 23.3 | 0.26 |
| polarC18 | 468.308 | 232.236 | 23.3 | 0.26 |
| polarC18 | 468.308 | 91.5492 | 23.3 | 0.26 |
| polarC18 | 468.308 | 156.0446 | 23.3 | 0.26 |
| polarC18 | 468.308 | 175.8338 | 23.3 | 0.26 |
| polarC18 | 468.308 | 386.8869 | 23.3 | 0.26 |
| polarC18 | 468.308 | 59.0129 | 23.3 | 0.26 |
| polarC18 | 468.308 | 171.6746 | 23.3 | 0.26 |
| polarC18 | 468.308 | 139.8785 | 23.3 | 0.26 |
| polarC18 | 468.308 | 206.7993 | 23.3 | 0.26 |
| polarC18 | 468.308 | 97.2181 | 23.3 | 0.26 |
| polarC18 | 468.308 | 105.8131 | 23.3 | 0.26 |
| polarC18 | 468.308 | 120.6977 | 23.3 | 0.26 |
| polarC18 | 468.308 | 71.9208 | 23.3 | 0.26 |
| polarC18 | 468.308 | 87.6944 | 23.3 | 0.26 |
| polarC18 | 468.308 | 70.8456 | 23.3 | 0.26 |
| polarC18 | 468.308 | 58.4778 | 23.3 | 0.26 |
| polarC18 | 468.308 | 150.069 | 23.3 | 0.26 |
| polarC18 | 468.308 | 95.7128 | 23.3 | 0.26 |
| polarC18 | 468.308 | 400.0942 | 23.3 | 0.26 |
| polarC18 | 468.308 | 486.925 | 23.2 | 0.26 |
| polarC18 | 468.308 | 57.4881 | 23.2 | 0.26 |
| polarC18 | 468.308 | 234.6772 | 23.2 | 0.26 |
| polarC18 | 468.308 | 54.8814 | 23.2 | 0.26 |
| polarC18 | 468.308 | 241.9583 | 23.2 | 0.26 |
| polarC18 | 468.308 | 152.2994 | 23.2 | 0.26 |
| polarC18 | 468.308 | 159.716 | 23.2 | 0.26 |
| polarC18 | 468.308 | 54.2404 | 23.2 | 0.26 |
| polarC18 | 468.308 | 167.8308 | 23.2 | 0.26 |
| polarC18 | 468.308 | 76.9015 | 23.2 | 0.26 |

| | | | | |
|---|---|---|---|---|
| polarC18 | 468.308 | 118.8583 | 23.2 | 0.26 |
| polarC18 | 468.308 | 120.5495 | 23.2 | 0.26 |
| polarC18 | 468.308 | 431.2226 | 23.2 | 0.26 |
| polarC18 | 468.308 | 489.8806 | 23.2 | 0.26 |
| polarC18 | 468.308 | 74.0552 | 23.2 | 0.26 |
| polarC18 | 468.308 | 248.5208 | 23.2 | 0.26 |
| polarC18 | 468.308 | 383.0595 | 23.2 | 0.26 |
| polarC18 | 468.308 | 77.7435 | 23.2 | 0.26 |
| polarC18 | 468.308 | 64.4368 | 23.2 | 0.26 |
| polarC18 | 468.308 | 146.4805 | 23.2 | 0.26 |
| polarC18 | 468.308 | 467.9793 | 23.2 | 0.26 |
| polarC18 | 468.308 | 224.7838 | 23.2 | 0.26 |
| polarC18 | 468.308 | 103.2862 | 23.2 | 0.26 |
| polarC18 | 468.308 | 141.1726 | 23.2 | 0.26 |
| polarC18 | 468.308 | 377.2027 | 23.2 | 0.26 |
| polarC18 | 468.308 | 367.1887 | 23.2 | 0.26 |
| polarC18 | 468.308 | 217.3935 | 23.2 | 0.26 |
| polarC18 | 468.308 | 56.9943 | 23.2 | 0.26 |
| polarC18 | 468.308 | 63.607 | 23.2 | 0.26 |
| polarC18 | 468.308 | 67.8358 | 23.2 | 0.26 |
| polarC18 | 468.308 | 460.5373 | 23.2 | 0.26 |
| polarC18 | 468.308 | 402.9737 | 23.2 | 0.26 |
| polarC18 | 468.308 | 149.8805 | 23.2 | 0.26 |
| polarC18 | 468.308 | 157.2244 | 23.2 | 0.26 |
| polarC18 | 468.308 | 110.1624 | 23.2 | 0.26 |
| polarC18 | 468.308 | 78.578 | 23.2 | 0.26 |
| polarC18 | 468.308 | 128.3218 | 23.2 | 0.26 |
| polarC18 | 468.308 | 154.9523 | 23.2 | 0.26 |
| polarC18 | 468.308 | 54.0503 | 23.2 | 0.26 |
| polarC18 | 468.308 | 234.0382 | 23.2 | 0.26 |
| polarC18 | 468.308 | 359.7538 | 23.2 | 0.26 |
| polarC18 | 468.308 | 61.8413 | 23.2 | 0.26 |
| polarC18 | 468.308 | 70.1583 | 23.2 | 0.26 |
| polarC18 | 468.308 | 372.0948 | 23.2 | 0.26 |
| polarC18 | 468.308 | 104.1329 | 23.1 | 0.26 |
| polarC18 | 468.308 | 245.192 | 23.1 | 0.26 |
| polarC18 | 468.308 | 67.8644 | 23.1 | 0.26 |
| polarC18 | 468.308 | 66.7683 | 23.1 | 0.26 |
| polarC18 | 468.308 | 95.7812 | 23.1 | 0.26 |
| polarC18 | 468.308 | 102.2092 | 23.1 | 0.26 |
| polarC18 | 468.308 | 254.7652 | 23.1 | 0.26 |
| polarC18 | 468.308 | 81.088 | 23.1 | 0.26 |

| | | | | |
|---|---|---|---|---|
| polarC18 | 468.308 | 120.2055 | 23.1 | 0.26 |
| polarC18 | 468.308 | 94.5392 | 23.1 | 0.26 |
| polarC18 | 468.308 | 77.2068 | 23.1 | 0.26 |
| polarC18 | 468.308 | 452.686 | 23.1 | 0.26 |
| polarC18 | 468.308 | 82.1108 | 23.1 | 0.26 |
| polarC18 | 468.308 | 178.2625 | 23.1 | 0.26 |
| polarC18 | 468.308 | 89.897 | 23.1 | 0.26 |
| polarC18 | 468.308 | 249.9001 | 23.1 | 0.26 |
| polarC18 | 468.308 | 281.1014 | 23.1 | 0.26 |
| polarC18 | 468.308 | 210.4161 | 23.1 | 0.26 |
| polarC18 | 468.308 | 156.9592 | 23.1 | 0.26 |
| polarC18 | 468.308 | 117.6238 | 23.1 | 0.26 |
| polarC18 | 468.308 | 274.4443 | 23.1 | 0.26 |
| polarC18 | 468.308 | 179.4783 | 23.1 | 0.26 |
| polarC18 | 468.308 | 155.3877 | 23.1 | 0.26 |
| polarC18 | 468.308 | 356.8515 | 23.1 | 0.26 |
| polarC18 | 468.308 | 88.8351 | 23.1 | 0.26 |
| polarC18 | 468.308 | 292.2867 | 23.1 | 0.26 |
| polarC18 | 468.308 | 174.823 | 23.1 | 0.26 |
| polarC18 | 468.308 | 183.0153 | 23.1 | 0.26 |
| polarC18 | 468.308 | 480.2102 | 23.1 | 0.26 |
| polarC18 | 468.308 | 53.0998 | 23.1 | 0.26 |
| polarC18 | 468.308 | 134.7513 | 23.1 | 0.26 |
| polarC18 | 468.308 | 73.1822 | 23.1 | 0.26 |
| polarC18 | 468.308 | 56.2608 | 23.1 | 0.26 |
| polarC18 | 468.308 | 310.8781 | 23.1 | 0.26 |
| polarC18 | 468.308 | 266.9541 | 23.1 | 0.26 |
| polarC18 | 468.308 | 266.761 | 23.1 | 0.26 |
| polarC18 | 468.308 | 62.2623 | 23.1 | 0.26 |
| polarC18 | 468.308 | 77.2663 | 23 | 0.26 |
| polarC18 | 468.308 | 158.6562 | 23 | 0.26 |
| polarC18 | 468.308 | 79.4565 | 23 | 0.26 |
| polarC18 | 468.308 | 95.8009 | 23 | 0.26 |
| polarC18 | 468.308 | 130.8365 | 23 | 0.26 |
| polarC18 | 468.308 | 244.8749 | 23 | 0.26 |
| polarC18 | 468.308 | 94.2513 | 23 | 0.26 |
| polarC18 | 468.308 | 99.0534 | 23 | 0.26 |
| polarC18 | 468.308 | 119.9822 | 23 | 0.26 |
| polarC18 | 468.308 | 119.7455 | 23 | 0.26 |
| polarC18 | 468.308 | 52.2091 | 23 | 0.26 |
| polarC18 | 468.308 | 133.9314 | 23 | 0.26 |
| polarC18 | 468.308 | 418.218 | 23 | 0.26 |

| | | | | |
|---|---|---|---|---|
| polarC18 | 468.308 | 406.7355 | 23 | 0.26 |
| polarC18 | 468.308 | 68.3934 | 23 | 0.26 |
| polarC18 | 468.308 | 169.3783 | 23 | 0.26 |
| polarC18 | 468.308 | 51.0193 | 23 | 0.26 |
| polarC18 | 468.308 | 124.3898 | 23 | 0.26 |
| polarC18 | 468.308 | 250.0817 | 23 | 0.26 |
| polarC18 | 468.308 | 340.3957 | 23 | 0.26 |
| polarC18 | 468.308 | 67.1421 | 23 | 0.26 |
| polarC18 | 468.308 | 50.5664 | 23 | 0.26 |
| polarC18 | 468.308 | 73.197 | 23 | 0.26 |
| polarC18 | 468.308 | 94.9158 | 23 | 0.26 |
| polarC18 | 468.308 | 457.2205 | 23 | 0.26 |
| polarC18 | 468.308 | 54.5715 | 23 | 0.26 |
| polarC18 | 468.308 | 145.1068 | 23 | 0.26 |
| polarC18 | 468.308 | 238.3028 | 23 | 0.26 |
| polarC18 | 468.308 | 362.5269 | 23 | 0.26 |
| polarC18 | 468.308 | 248.5886 | 23 | 0.26 |
| polarC18 | 468.308 | 291.5381 | 23 | 0.26 |
| polarC18 | 468.308 | 84.6904 | 23 | 0.26 |
| polarC18 | 468.308 | 294.3505 | 23 | 0.26 |
| polarC18 | 468.308 | 135.978 | 23 | 0.26 |
| polarC18 | 468.308 | 61.9649 | 23 | 0.26 |
| polarC18 | 468.308 | 270.3234 | 23 | 0.26 |
| polarC18 | 468.308 | 399.9266 | 23 | 0.26 |
| polarC18 | 468.308 | 258.4731 | 23 | 0.26 |
| polarC18 | 468.308 | 165.029 | 23 | 0.26 |
| polarC18 | 468.308 | 230.5037 | 23 | 0.26 |
| polarC18 | 468.308 | 72.6686 | 23 | 0.26 |
| polarC18 | 468.308 | 82.5925 | 23 | 0.26 |
| polarC18 | 468.308 | 492.1969 | 23 | 0.26 |
| polarC18 | 468.308 | 71.7483 | 23 | 0.26 |
| polarC18 | 468.308 | 295.6203 | 23 | 0.26 |
| polarC18 | 468.308 | 377.601 | 23 | 0.26 |
| polarC18 | 468.308 | 112.9846 | 23 | 0.26 |
| polarC18 | 468.308 | 170.0301 | 22.9 | 0.26 |
| polarC18 | 468.308 | 104.6608 | 22.9 | 0.26 |
| polarC18 | 468.308 | 54.1605 | 22.9 | 0.26 |
| polarC18 | 468.308 | 310.1389 | 22.9 | 0.26 |
| polarC18 | 468.308 | 93.8859 | 22.9 | 0.26 |
| polarC18 | 468.308 | 61.9503 | 22.9 | 0.26 |
| polarC18 | 468.308 | 75.8563 | 22.9 | 0.26 |
| polarC18 | 468.308 | 108.3965 | 22.9 | 0.26 |

| | | | | |
|---|---|---|---|---|
| polarC18 | 468.308 | 238.0447 | 22.9 | 0.26 |
| polarC18 | 468.308 | 209.7746 | 22.9 | 0.26 |
| polarC18 | 468.308 | 198.8806 | 22.9 | 0.26 |
| polarC18 | 468.308 | 64.0379 | 22.9 | 0.26 |
| polarC18 | 468.308 | 78.8321 | 22.9 | 0.26 |
| polarC18 | 468.308 | 83.7838 | 22.9 | 0.26 |
| polarC18 | 468.308 | 477.311 | 22.9 | 0.26 |
| polarC18 | 468.308 | 258.1382 | 22.9 | 0.26 |
| polarC18 | 468.308 | 472.6493 | 22.9 | 0.26 |
| polarC18 | 468.308 | 254.5513 | 22.9 | 0.26 |
| polarC18 | 468.308 | 236.2291 | 22.9 | 0.26 |
| polarC18 | 468.308 | 54.6542 | 22.9 | 0.26 |
| polarC18 | 468.308 | 60.2789 | 22.9 | 0.26 |
| polarC18 | 468.308 | 103.1475 | 22.9 | 0.26 |
| polarC18 | 468.308 | 78.5713 | 22.9 | 0.26 |
| polarC18 | 468.308 | 61.8851 | 22.9 | 0.26 |
| polarC18 | 468.308 | 241.0778 | 22.9 | 0.26 |
| polarC18 | 468.308 | 138.9176 | 22.9 | 0.26 |
| polarC18 | 468.308 | 239.5087 | 22.9 | 0.26 |
| polarC18 | 468.308 | 425.1637 | 22.9 | 0.26 |
| polarC18 | 468.308 | 117.1648 | 22.9 | 0.26 |
| polarC18 | 468.308 | 218.1928 | 22.9 | 0.26 |
| polarC18 | 468.308 | 149.0301 | 22.8 | 0.26 |
| polarC18 | 468.308 | 70.7261 | 22.8 | 0.26 |
| polarC18 | 468.308 | 144.7765 | 22.8 | 0.26 |
| polarC18 | 468.308 | 169.6026 | 22.8 | 0.26 |
| polarC18 | 468.308 | 171.0061 | 22.8 | 0.26 |
| polarC18 | 468.308 | 194.9611 | 22.8 | 0.26 |
| polarC18 | 468.308 | 118.0848 | 22.8 | 0.26 |
| polarC18 | 468.308 | 89.9277 | 22.8 | 0.26 |
| polarC18 | 468.308 | 160.0715 | 22.8 | 0.26 |
| polarC18 | 468.308 | 115.8307 | 22.8 | 0.26 |
| polarC18 | 468.308 | 132.636 | 22.8 | 0.26 |
| polarC18 | 468.308 | 171.5958 | 22.8 | 0.26 |
| polarC18 | 468.308 | 221.8648 | 22.8 | 0.26 |
| polarC18 | 468.308 | 160.1659 | 22.8 | 0.26 |
| polarC18 | 468.308 | 59.3501 | 22.8 | 0.26 |
| polarC18 | 468.308 | 70.5371 | 22.8 | 0.26 |
| polarC18 | 468.308 | 115.944 | 22.8 | 0.26 |
| polarC18 | 468.308 | 106.2053 | 22.8 | 0.26 |
| polarC18 | 468.308 | 165.0853 | 22.8 | 0.26 |
| polarC18 | 468.308 | 304.8767 | 22.8 | 0.26 |

| | | | | |
|---|---|---|---|---|
| polarC18 | 468.308 | 72.3953 | 22.8 | 0.26 |
| polarC18 | 468.308 | 293.8203 | 22.8 | 0.26 |
| polarC18 | 468.308 | 222.5108 | 22.8 | 0.26 |
| polarC18 | 468.308 | 227.2822 | 22.8 | 0.26 |
| polarC18 | 468.308 | 51.4509 | 22.8 | 0.26 |
| polarC18 | 468.308 | 352.2314 | 22.8 | 0.26 |
| polarC18 | 468.308 | 117.7728 | 22.8 | 0.26 |
| polarC18 | 468.308 | 397.2254 | 22.8 | 0.26 |
| polarC18 | 468.308 | 114.0024 | 22.8 | 0.26 |
| polarC18 | 468.308 | 234.3731 | 22.8 | 0.26 |
| polarC18 | 468.308 | 270.0566 | 22.8 | 0.26 |
| polarC18 | 468.308 | 177.5862 | 22.8 | 0.26 |
| polarC18 | 468.308 | 271.7057 | 22.8 | 0.26 |
| polarC18 | 468.308 | 105.0216 | 22.8 | 0.26 |
| polarC18 | 468.308 | 66.6808 | 22.8 | 0.26 |
| polarC18 | 468.308 | 180.7409 | 22.8 | 0.26 |
| polarC18 | 468.308 | 108.6004 | 22.8 | 0.26 |
| polarC18 | 468.308 | 97.4351 | 22.8 | 0.26 |
| polarC18 | 468.308 | 160.6991 | 22.8 | 0.26 |
| polarC18 | 468.308 | 169.2085 | 22.7 | 0.26 |
| polarC18 | 468.308 | 111.7027 | 22.7 | 0.26 |
| polarC18 | 468.308 | 497.3325 | 22.7 | 0.26 |
| polarC18 | 468.308 | 177.9472 | 22.7 | 0.26 |
| polarC18 | 468.308 | 230.8933 | 22.7 | 0.26 |
| polarC18 | 468.308 | 409.5457 | 22.7 | 0.26 |
| polarC18 | 468.308 | 53.1199 | 22.7 | 0.26 |
| polarC18 | 468.308 | 144.9169 | 22.7 | 0.26 |
| polarC18 | 468.308 | 54.3754 | 22.7 | 0.26 |
| polarC18 | 468.308 | 68.6228 | 22.7 | 0.26 |
| polarC18 | 468.308 | 277.0448 | 22.7 | 0.26 |
| polarC18 | 468.308 | 169.7222 | 22.7 | 0.26 |
| polarC18 | 468.308 | 85.7614 | 22.7 | 0.26 |
| polarC18 | 468.308 | 95.197 | 22.7 | 0.26 |
| polarC18 | 468.308 | 76.3006 | 22.7 | 0.26 |
| polarC18 | 468.308 | 75.3127 | 22.7 | 0.26 |
| polarC18 | 468.308 | 234.6334 | 22.7 | 0.26 |
| polarC18 | 468.308 | 66.9129 | 22.7 | 0.26 |
| polarC18 | 468.308 | 185.0454 | 22.7 | 0.26 |
| polarC18 | 468.308 | 118.3751 | 22.7 | 0.26 |
| polarC18 | 468.308 | 123.1982 | 22.7 | 0.26 |
| polarC18 | 468.308 | 88.2204 | 22.7 | 0.26 |
| polarC18 | 468.308 | 75.0186 | 22.7 | 0.26 |

| | | | | |
|---|---|---|---|---|
| polarC18 | 468.308 | 209.9901 | 22.7 | 0.26 |
| polarC18 | 468.308 | 111.1409 | 22.7 | 0.26 |
| polarC18 | 468.308 | 124.3445 | 22.7 | 0.26 |
| polarC18 | 468.308 | 214.6808 | 22.7 | 0.26 |
| polarC18 | 468.308 | 51.9409 | 22.7 | 0.26 |
| polarC18 | 468.308 | 57.1198 | 22.7 | 0.26 |
| polarC18 | 468.308 | 210.352 | 22.7 | 0.26 |
| polarC18 | 468.308 | 67.6333 | 22.7 | 0.26 |
| polarC18 | 468.308 | 197.759 | 22.7 | 0.26 |
| polarC18 | 468.308 | 68.8036 | 22.7 | 0.26 |
| polarC18 | 468.308 | 481.2222 | 22.7 | 0.26 |
| polarC18 | 468.308 | 186.3063 | 22.7 | 0.26 |
| polarC18 | 468.308 | 195.5643 | 22.7 | 0.26 |
| polarC18 | 468.308 | 118.2608 | 22.7 | 0.26 |
| polarC18 | 468.308 | 55.4738 | 22.7 | 0.26 |
| polarC18 | 468.308 | 354.8294 | 22.7 | 0.26 |
| polarC18 | 468.308 | 298.2936 | 22.7 | 0.26 |
| polarC18 | 468.308 | 320.7172 | 22.7 | 0.26 |
| polarC18 | 468.308 | 99.2794 | 22.6 | 0.25 |
| polarC18 | 468.308 | 106.1989 | 22.6 | 0.25 |
| polarC18 | 468.308 | 384.9207 | 22.6 | 0.25 |
| polarC18 | 468.308 | 193.0887 | 22.6 | 0.25 |
| polarC18 | 468.308 | 53.8865 | 22.6 | 0.25 |
| polarC18 | 468.308 | 144.5847 | 22.6 | 0.25 |
| polarC18 | 468.308 | 90.2047 | 22.6 | 0.25 |
| polarC18 | 468.308 | 56.6025 | 22.6 | 0.25 |
| polarC18 | 468.308 | 57.6364 | 22.6 | 0.25 |
| polarC18 | 468.308 | 367.6768 | 22.6 | 0.25 |
| polarC18 | 468.308 | 53.6724 | 22.6 | 0.25 |
| polarC18 | 468.308 | 149.5241 | 22.6 | 0.25 |
| polarC18 | 468.308 | 63.9302 | 22.6 | 0.25 |
| polarC18 | 468.308 | 175.9401 | 22.6 | 0.25 |
| polarC18 | 468.308 | 67.2347 | 22.6 | 0.25 |
| polarC18 | 468.308 | 98.5725 | 22.6 | 0.25 |
| polarC18 | 468.308 | 51.3349 | 22.6 | 0.25 |
| polarC18 | 468.308 | 79.3898 | 22.6 | 0.25 |
| polarC18 | 468.308 | 249.5257 | 22.6 | 0.25 |
| polarC18 | 468.308 | 73.2227 | 22.6 | 0.25 |
| polarC18 | 468.308 | 123.0343 | 22.6 | 0.25 |
| polarC18 | 468.308 | 299.4223 | 22.6 | 0.25 |
| polarC18 | 468.308 | 62.2365 | 22.6 | 0.25 |
| polarC18 | 468.308 | 262.9888 | 22.6 | 0.25 |

| | | | | |
|---|---|---|---|---|
| polarC18 | 468.308 | 88.8997 | 22.6 | 0.25 |
| polarC18 | 468.308 | 346.4076 | 22.6 | 0.25 |
| polarC18 | 468.308 | 280.2345 | 22.6 | 0.25 |
| polarC18 | 468.308 | 343.2941 | 22.6 | 0.25 |
| polarC18 | 468.308 | 229.6403 | 22.6 | 0.25 |
| polarC18 | 468.308 | 54.9046 | 22.6 | 0.25 |
| polarC18 | 468.308 | 59.8085 | 22.6 | 0.25 |
| polarC18 | 468.308 | 83.5802 | 22.6 | 0.25 |
| polarC18 | 468.308 | 160.114 | 22.6 | 0.25 |
| polarC18 | 468.308 | 57.691 | 22.6 | 0.25 |
| polarC18 | 468.308 | 100.8905 | 22.6 | 0.25 |
| polarC18 | 468.308 | 56.3461 | 22.6 | 0.25 |
| polarC18 | 468.308 | 375.9589 | 22.6 | 0.25 |
| polarC18 | 468.308 | 201.6154 | 22.6 | 0.25 |
| polarC18 | 468.308 | 52.1653 | 22.6 | 0.25 |
| polarC18 | 468.308 | 319.8004 | 22.6 | 0.25 |
| polarC18 | 468.308 | 82.4913 | 22.6 | 0.25 |
| polarC18 | 468.308 | 165.7759 | 22.6 | 0.25 |
| polarC18 | 468.308 | 107.869 | 22.6 | 0.25 |
| polarC18 | 468.308 | 56.6407 | 22.6 | 0.25 |
| polarC18 | 468.308 | 59.7729 | 22.5 | 0.25 |
| polarC18 | 468.308 | 94.0795 | 22.5 | 0.25 |
| polarC18 | 468.308 | 154.6412 | 22.5 | 0.25 |
| polarC18 | 468.308 | 158.7141 | 22.5 | 0.25 |
| polarC18 | 468.308 | 403.6934 | 22.5 | 0.25 |
| polarC18 | 468.308 | 327.8958 | 22.5 | 0.25 |
| polarC18 | 468.308 | 201.9671 | 22.5 | 0.25 |
| polarC18 | 468.308 | 148.9797 | 22.5 | 0.25 |
| polarC18 | 468.308 | 92.8665 | 22.5 | 0.25 |
| polarC18 | 468.308 | 378.4919 | 22.5 | 0.25 |
| polarC18 | 468.308 | 55.7656 | 22.5 | 0.25 |
| polarC18 | 468.308 | 97.3512 | 22.5 | 0.25 |
| polarC18 | 468.308 | 307.8206 | 22.5 | 0.25 |
| polarC18 | 468.308 | 122.9109 | 22.5 | 0.25 |
| polarC18 | 468.308 | 77.7958 | 22.5 | 0.25 |
| polarC18 | 468.308 | 120.8929 | 22.5 | 0.25 |
| polarC18 | 468.308 | 79.1948 | 22.5 | 0.25 |
| polarC18 | 468.308 | 54.3894 | 22.5 | 0.25 |
| polarC18 | 468.308 | 74.5715 | 22.5 | 0.25 |
| polarC18 | 468.308 | 117.6716 | 22.5 | 0.25 |
| polarC18 | 468.308 | 118.0239 | 22.5 | 0.25 |
| polarC18 | 468.308 | 118.7249 | 22.5 | 0.25 |

| | | | | |
|---|---|---|---|---|
| polarC18 | 468.308 | 239.8472 | 22.5 | 0.25 |
| polarC18 | 468.308 | 64.4772 | 22.5 | 0.25 |
| polarC18 | 468.308 | 162.739 | 22.5 | 0.25 |
| polarC18 | 468.308 | 53.8589 | 22.5 | 0.25 |
| polarC18 | 468.308 | 55.3008 | 22.5 | 0.25 |
| polarC18 | 468.308 | 57.8533 | 22.5 | 0.25 |
| polarC18 | 468.308 | 82.0969 | 22.5 | 0.25 |
| polarC18 | 468.308 | 96.3027 | 22.5 | 0.25 |
| polarC18 | 468.308 | 381.7386 | 22.5 | 0.25 |
| polarC18 | 468.308 | 385.9619 | 22.5 | 0.25 |
| polarC18 | 468.308 | 261.8141 | 22.5 | 0.25 |
| polarC18 | 468.308 | 248.938 | 22.5 | 0.25 |
| polarC18 | 468.308 | 241.9261 | 22.5 | 0.25 |
| polarC18 | 468.308 | 51.5813 | 22.5 | 0.25 |
| polarC18 | 468.308 | 64.1941 | 22.5 | 0.25 |
| polarC18 | 468.308 | 186.9328 | 22.5 | 0.25 |
| polarC18 | 468.308 | 191.3069 | 22.4 | 0.25 |
| polarC18 | 468.308 | 125.6076 | 22.4 | 0.25 |
| polarC18 | 468.308 | 141.0685 | 22.4 | 0.25 |
| polarC18 | 468.308 | 86.3036 | 22.4 | 0.25 |
| polarC18 | 468.308 | 59.6085 | 22.4 | 0.25 |
| polarC18 | 468.308 | 323.8852 | 22.4 | 0.25 |
| polarC18 | 468.308 | 360.2863 | 22.4 | 0.25 |
| polarC18 | 468.308 | 242.2553 | 22.4 | 0.25 |
| polarC18 | 468.308 | 122.1811 | 22.4 | 0.25 |
| polarC18 | 468.308 | 255.7137 | 22.4 | 0.25 |
| polarC18 | 468.308 | 206.8678 | 22.4 | 0.25 |
| polarC18 | 468.308 | 71.9629 | 22.4 | 0.25 |
| polarC18 | 468.308 | 191.7595 | 22.4 | 0.25 |
| polarC18 | 468.308 | 84.3799 | 22.4 | 0.25 |
| polarC18 | 468.308 | 67.239 | 22.4 | 0.25 |
| polarC18 | 468.308 | 66.2344 | 22.4 | 0.25 |
| polarC18 | 468.308 | 95.4615 | 22.4 | 0.25 |
| polarC18 | 468.308 | 267.9893 | 22.4 | 0.25 |
| polarC18 | 468.308 | 415.6933 | 22.4 | 0.25 |
| polarC18 | 468.308 | 64.0497 | 22.4 | 0.25 |
| polarC18 | 468.308 | 162.1895 | 22.4 | 0.25 |
| polarC18 | 468.308 | 91.9186 | 22.4 | 0.25 |
| polarC18 | 468.308 | 94.4981 | 22.4 | 0.25 |
| polarC18 | 468.308 | 337.4877 | 22.4 | 0.25 |
| polarC18 | 468.308 | 90.8988 | 22.4 | 0.25 |
| polarC18 | 468.308 | 69.7096 | 22.4 | 0.25 |

| | | | | |
|---|---|---|---|---|
| polarC18 | 468.308 | 93.6654 | 22.4 | 0.25 |
| polarC18 | 468.308 | 149.4348 | 22.4 | 0.25 |
| polarC18 | 468.308 | 93.7277 | 22.4 | 0.25 |
| polarC18 | 468.308 | 371.4086 | 22.4 | 0.25 |
| polarC18 | 468.308 | 90.1786 | 22.4 | 0.25 |
| polarC18 | 468.308 | 178.5182 | 22.4 | 0.25 |
| polarC18 | 468.308 | 108.1164 | 22.4 | 0.25 |
| polarC18 | 468.308 | 51.579 | 22.4 | 0.25 |
| polarC18 | 468.308 | 273.7693 | 22.4 | 0.25 |
| polarC18 | 468.308 | 334.5232 | 22.4 | 0.25 |
| polarC18 | 468.308 | 137.4162 | 22.4 | 0.25 |
| polarC18 | 468.308 | 61.0256 | 22.4 | 0.25 |
| polarC18 | 468.308 | 161.3443 | 22.4 | 0.25 |
| polarC18 | 468.308 | 72.5312 | 22.4 | 0.25 |
| polarC18 | 468.308 | 50.7146 | 22.4 | 0.25 |
| polarC18 | 468.308 | 117.7364 | 22.4 | 0.25 |
| polarC18 | 468.308 | 66.0175 | 22.4 | 0.25 |
| polarC18 | 468.308 | 69.9932 | 22.4 | 0.25 |
| polarC18 | 468.308 | 57.2848 | 22.4 | 0.25 |
| polarC18 | 468.308 | 70.3623 | 22.4 | 0.25 |
| polarC18 | 468.308 | 102.0534 | 22.4 | 0.25 |
| polarC18 | 468.308 | 55.9281 | 22.4 | 0.25 |
| polarC18 | 468.308 | 72.3061 | 22.4 | 0.25 |
| polarC18 | 468.308 | 98.4187 | 22.4 | 0.25 |
| polarC18 | 468.308 | 258.2218 | 22.4 | 0.25 |
| polarC18 | 468.308 | 258.3731 | 22.4 | 0.25 |
| polarC18 | 468.308 | 143.7343 | 22.4 | 0.25 |
| polarC18 | 468.308 | 408.495 | 22.4 | 0.25 |
| polarC18 | 468.308 | 88.0726 | 22.3 | 0.25 |
| polarC18 | 468.308 | 57.1595 | 22.3 | 0.25 |
| polarC18 | 468.308 | 307.1788 | 22.3 | 0.25 |
| polarC18 | 468.308 | 129.8678 | 22.3 | 0.25 |
| polarC18 | 468.308 | 61.9798 | 22.3 | 0.25 |
| polarC18 | 468.308 | 136.2451 | 22.3 | 0.25 |
| polarC18 | 468.308 | 472.2643 | 22.3 | 0.25 |
| polarC18 | 468.308 | 54.2827 | 22.3 | 0.25 |
| polarC18 | 468.308 | 186.1842 | 22.3 | 0.25 |
| polarC18 | 468.308 | 178.7838 | 22.3 | 0.25 |
| polarC18 | 468.308 | 120.325 | 22.3 | 0.25 |
| polarC18 | 468.308 | 79.6674 | 22.3 | 0.25 |
| polarC18 | 468.308 | 100.4887 | 22.3 | 0.25 |
| polarC18 | 468.308 | 64.2066 | 22.3 | 0.25 |

| | | | | |
|---|---|---|---|---|
| polarC18 | 468.308 | 452.9322 | 22.3 | 0.25 |
| polarC18 | 468.308 | 108.8937 | 22.3 | 0.25 |
| polarC18 | 468.308 | 74.0484 | 22.3 | 0.25 |
| polarC18 | 468.308 | 224.727 | 22.3 | 0.25 |
| polarC18 | 468.308 | 278.3513 | 22.3 | 0.25 |
| polarC18 | 468.308 | 179.7816 | 22.3 | 0.25 |
| polarC18 | 468.308 | 236.3632 | 22.3 | 0.25 |
| polarC18 | 468.308 | 56.5563 | 22.3 | 0.25 |
| polarC18 | 468.308 | 238.583 | 22.3 | 0.25 |
| polarC18 | 468.308 | 239.7862 | 22.3 | 0.25 |
| polarC18 | 468.308 | 87.2852 | 22.3 | 0.25 |
| polarC18 | 468.308 | 55.4914 | 22.3 | 0.25 |
| polarC18 | 468.308 | 239.8885 | 22.3 | 0.25 |
| polarC18 | 468.308 | 96.5595 | 22.3 | 0.25 |
| polarC18 | 468.308 | 189.0587 | 22.3 | 0.25 |
| polarC18 | 468.308 | 329.4536 | 22.3 | 0.25 |
| polarC18 | 468.308 | 57.4495 | 22.3 | 0.25 |
| polarC18 | 468.308 | 193.5447 | 22.3 | 0.25 |
| polarC18 | 468.308 | 153.8848 | 22.3 | 0.25 |
| polarC18 | 468.308 | 81.9997 | 22.3 | 0.25 |
| polarC18 | 468.308 | 466.2358 | 22.3 | 0.25 |
| polarC18 | 468.308 | 281.3348 | 22.3 | 0.25 |
| polarC18 | 468.308 | 181.138 | 22.3 | 0.25 |
| polarC18 | 468.308 | 73.4206 | 22.3 | 0.25 |
| polarC18 | 468.308 | 77.0213 | 22.3 | 0.25 |
| polarC18 | 468.308 | 387.106 | 22.3 | 0.25 |
| polarC18 | 468.308 | 208.3761 | 22.3 | 0.25 |
| polarC18 | 468.308 | 342.4132 | 22.3 | 0.25 |
| polarC18 | 468.308 | 197.6213 | 22.3 | 0.25 |
| polarC18 | 468.308 | 277.1393 | 22.3 | 0.25 |
| polarC18 | 468.308 | 159.7729 | 22.3 | 0.25 |
| polarC18 | 468.308 | 71.2736 | 22.3 | 0.25 |
| polarC18 | 468.308 | 53.3103 | 22.3 | 0.25 |
| polarC18 | 468.308 | 376.5117 | 22.3 | 0.25 |
| polarC18 | 468.308 | 134.5167 | 22.3 | 0.25 |
| polarC18 | 468.308 | 70.0531 | 22.3 | 0.25 |
| polarC18 | 468.308 | 122.109 | 22.3 | 0.25 |
| polarC18 | 468.308 | 122.5932 | 22.2 | 0.25 |
| polarC18 | 468.308 | 413.9202 | 22.2 | 0.25 |
| polarC18 | 468.308 | 210.4577 | 22.2 | 0.25 |
| polarC18 | 468.308 | 153.0081 | 22.2 | 0.25 |
| polarC18 | 468.308 | 125.5107 | 22.2 | 0.25 |

| | | | | |
|---|---|---|---|---|
| polarC18 | 468.308 | 121.4267 | 22.2 | 0.25 |
| polarC18 | 468.308 | 175.4624 | 22.2 | 0.25 |
| polarC18 | 468.308 | 237.2114 | 22.2 | 0.25 |
| polarC18 | 468.308 | 76.6302 | 22.2 | 0.25 |
| polarC18 | 468.308 | 344.9494 | 22.2 | 0.25 |
| polarC18 | 468.308 | 332.3653 | 22.2 | 0.25 |
| polarC18 | 468.308 | 171.8119 | 22.2 | 0.25 |
| polarC18 | 468.308 | 376.8803 | 22.2 | 0.25 |
| polarC18 | 468.308 | 51.2692 | 22.2 | 0.25 |
| polarC18 | 468.308 | 51.8904 | 22.2 | 0.25 |
| polarC18 | 468.308 | 100.3609 | 22.2 | 0.25 |
| polarC18 | 468.308 | 484.1373 | 22.2 | 0.25 |
| polarC18 | 468.308 | 94.8247 | 22.2 | 0.25 |
| polarC18 | 468.308 | 213.9747 | 22.2 | 0.25 |
| polarC18 | 468.308 | 88.1726 | 22.2 | 0.25 |
| polarC18 | 468.308 | 80.1905 | 22.2 | 0.25 |
| polarC18 | 468.308 | 394.11 | 22.2 | 0.25 |
| polarC18 | 468.308 | 128.6199 | 22.2 | 0.25 |
| polarC18 | 468.308 | 129.4 | 22.2 | 0.25 |
| polarC18 | 468.308 | 155.7667 | 22.2 | 0.25 |
| polarC18 | 468.308 | 126.8388 | 22.2 | 0.25 |
| polarC18 | 468.308 | 55.0783 | 22.2 | 0.25 |
| polarC18 | 468.308 | 112.2416 | 22.2 | 0.25 |
| polarC18 | 468.308 | 141.4233 | 22.2 | 0.25 |
| polarC18 | 468.308 | 309.4014 | 22.2 | 0.25 |
| polarC18 | 468.308 | 159.2869 | 22.2 | 0.25 |
| polarC18 | 468.308 | 142.0591 | 22.2 | 0.25 |
| polarC18 | 468.308 | 141.2496 | 22.2 | 0.25 |
| polarC18 | 468.308 | 325.3319 | 22.2 | 0.25 |
| polarC18 | 468.308 | 443.8477 | 22.2 | 0.25 |
| polarC18 | 468.308 | 74.1451 | 22.2 | 0.25 |
| polarC18 | 468.308 | 132.8883 | 22.2 | 0.25 |
| polarC18 | 468.308 | 326.4968 | 22.2 | 0.25 |
| polarC18 | 468.308 | 61.3675 | 22.2 | 0.25 |
| polarC18 | 468.308 | 113.1037 | 22.2 | 0.25 |
| polarC18 | 468.308 | 61.3052 | 22.2 | 0.25 |
| polarC18 | 468.308 | 95.7364 | 22.2 | 0.25 |
| polarC18 | 468.308 | 250.735 | 22.2 | 0.25 |
| polarC18 | 468.308 | 491.6281 | 22.2 | 0.25 |
| polarC18 | 468.308 | 86.4522 | 22.2 | 0.25 |
| polarC18 | 468.308 | 223.2752 | 22.2 | 0.25 |
| polarC18 | 468.308 | 222.4315 | 22.2 | 0.25 |

| | | | | |
|---|---|---|---|---|
| polarC18 | 468.308 | 86.4143 | 22.2 | 0.25 |
| polarC18 | 468.308 | 109.9436 | 22.2 | 0.25 |
| polarC18 | 468.308 | 179.2353 | 22.2 | 0.25 |
| polarC18 | 468.308 | 133.7603 | 22.2 | 0.25 |
| polarC18 | 468.308 | 287.8579 | 22.2 | 0.25 |
| polarC18 | 468.308 | 77.0086 | 22.2 | 0.25 |
| polarC18 | 468.308 | 62.0961 | 22.2 | 0.25 |
| polarC18 | 468.308 | 172.0745 | 22.2 | 0.25 |
| polarC18 | 468.308 | 78.4465 | 22.2 | 0.25 |
| polarC18 | 468.308 | 94.3079 | 22.2 | 0.25 |
| polarC18 | 468.308 | 72.1183 | 22.2 | 0.25 |
| polarC18 | 468.308 | 275.666 | 22.2 | 0.25 |
| polarC18 | 468.308 | 60.8155 | 22.2 | 0.25 |
| polarC18 | 468.308 | 51.6822 | 22.2 | 0.25 |
| polarC18 | 468.308 | 414.317 | 22.2 | 0.25 |
| polarC18 | 468.308 | 53.7354 | 22.1 | 0.25 |
| polarC18 | 468.308 | 117.9615 | 22.1 | 0.25 |
| polarC18 | 468.308 | 284.2246 | 22.1 | 0.25 |
| polarC18 | 468.308 | 162.2737 | 22.1 | 0.25 |
| polarC18 | 468.308 | 462.8958 | 22.1 | 0.25 |
| polarC18 | 468.308 | 65.3492 | 22.1 | 0.25 |
| polarC18 | 468.308 | 108.9022 | 22.1 | 0.25 |
| polarC18 | 468.308 | 217.2635 | 22.1 | 0.25 |
| polarC18 | 468.308 | 58.8002 | 22.1 | 0.25 |
| polarC18 | 468.308 | 412.8135 | 22.1 | 0.25 |
| polarC18 | 468.308 | 53.4556 | 22.1 | 0.25 |
| polarC18 | 468.308 | 102.6587 | 22.1 | 0.25 |
| polarC18 | 468.308 | 163.0065 | 22.1 | 0.25 |
| polarC18 | 468.308 | 71.5423 | 22.1 | 0.25 |
| polarC18 | 468.308 | 191.8955 | 22.1 | 0.25 |
| polarC18 | 468.308 | 299.9917 | 22.1 | 0.25 |
| polarC18 | 468.308 | 71.9954 | 22.1 | 0.25 |
| polarC18 | 468.308 | 186.2195 | 22.1 | 0.25 |
| polarC18 | 468.308 | 420.8855 | 22.1 | 0.25 |
| polarC18 | 468.308 | 155.3386 | 22.1 | 0.25 |
| polarC18 | 468.308 | 457.7973 | 22.1 | 0.25 |
| polarC18 | 468.308 | 110.7652 | 22.1 | 0.25 |
| polarC18 | 468.308 | 73.7589 | 22.1 | 0.25 |
| polarC18 | 468.308 | 144.0449 | 22.1 | 0.25 |
| polarC18 | 468.308 | 170.822 | 22.1 | 0.25 |
| polarC18 | 468.308 | 314.8668 | 22.1 | 0.25 |
| polarC18 | 468.308 | 369.6593 | 22.1 | 0.25 |

| | | | | |
|---|---|---|---|---|
| polarC18 | 468.308 | 485.8307 | 22.1 | 0.25 |
| polarC18 | 468.308 | 75.3911 | 22.1 | 0.25 |
| polarC18 | 468.308 | 260.2141 | 22.1 | 0.25 |
| polarC18 | 468.308 | 57.2157 | 22.1 | 0.25 |
| polarC18 | 468.308 | 50.6101 | 22.1 | 0.25 |
| polarC18 | 468.308 | 160.4703 | 22.1 | 0.25 |
| polarC18 | 468.308 | 159.1362 | 22.1 | 0.25 |
| polarC18 | 468.308 | 104.8305 | 22.1 | 0.25 |
| polarC18 | 468.308 | 321.3078 | 22.1 | 0.25 |
| polarC18 | 468.308 | 199.1171 | 22.1 | 0.25 |
| polarC18 | 468.308 | 54.2308 | 22.1 | 0.25 |
| polarC18 | 468.308 | 160.7311 | 22.1 | 0.25 |
| polarC18 | 468.308 | 78.8631 | 22.1 | 0.25 |
| polarC18 | 468.308 | 389.9536 | 22.1 | 0.25 |
| polarC18 | 468.308 | 94.1514 | 22.1 | 0.25 |
| polarC18 | 468.308 | 207.3879 | 22.1 | 0.25 |
| polarC18 | 468.308 | 107.092 | 22.1 | 0.25 |
| polarC18 | 468.308 | 98.0434 | 22.1 | 0.25 |
| polarC18 | 468.308 | 73.8217 | 22.1 | 0.25 |
| polarC18 | 468.308 | 57.5241 | 22.1 | 0.25 |
| polarC18 | 468.308 | 464.1787 | 22.1 | 0.25 |
| polarC18 | 468.308 | 476.4591 | 22 | 0.25 |
| polarC18 | 468.308 | 63.5324 | 22 | 0.25 |
| polarC18 | 468.308 | 69.4036 | 22 | 0.25 |
| polarC18 | 468.308 | 68.2379 | 22 | 0.25 |
| polarC18 | 468.308 | 355.7325 | 22 | 0.25 |
| polarC18 | 468.308 | 67.4854 | 22 | 0.25 |
| polarC18 | 468.308 | 193.9965 | 22 | 0.25 |
| polarC18 | 468.308 | 136.4381 | 22 | 0.25 |
| polarC18 | 468.308 | 269.9656 | 22 | 0.25 |
| polarC18 | 468.308 | 298.4998 | 22 | 0.25 |
| polarC18 | 468.308 | 164.2221 | 22 | 0.25 |
| polarC18 | 468.308 | 150.4678 | 22 | 0.25 |
| polarC18 | 468.308 | 382.1945 | 22 | 0.25 |
| polarC18 | 468.308 | 110.9714 | 22 | 0.25 |
| polarC18 | 468.308 | 365.2608 | 22 | 0.25 |
| polarC18 | 468.308 | 123.0149 | 22 | 0.25 |
| polarC18 | 468.308 | 64.2412 | 22 | 0.25 |
| polarC18 | 468.308 | 78.5963 | 22 | 0.25 |
| polarC18 | 468.308 | 80.4799 | 22 | 0.25 |
| polarC18 | 468.308 | 81.0413 | 22 | 0.25 |
| polarC18 | 468.308 | 99.5483 | 22 | 0.25 |

| | | | | |
|---|---|---|---|---|
| polarC18 | 468.308 | 82.4386 | 22 | 0.25 |
| polarC18 | 468.308 | 94.7256 | 22 | 0.25 |
| polarC18 | 468.308 | 93.9151 | 22 | 0.25 |
| polarC18 | 468.308 | 296.2815 | 22 | 0.25 |
| polarC18 | 468.308 | 52.4754 | 22 | 0.25 |
| polarC18 | 468.308 | 164.9911 | 22 | 0.25 |
| polarC18 | 468.308 | 105.8859 | 22 | 0.25 |
| polarC18 | 468.308 | 405.5006 | 22 | 0.25 |
| polarC18 | 468.308 | 421.897 | 22 | 0.25 |
| polarC18 | 468.308 | 85.4327 | 22 | 0.25 |
| polarC18 | 468.308 | 311.8965 | 22 | 0.25 |
| polarC18 | 468.308 | 68.0951 | 22 | 0.25 |
| polarC18 | 468.308 | 100.8212 | 22 | 0.25 |
| polarC18 | 468.308 | 492.0301 | 22 | 0.25 |
| polarC18 | 468.308 | 198.5143 | 22 | 0.25 |
| polarC18 | 468.308 | 74.722 | 22 | 0.25 |
| polarC18 | 468.308 | 220.3387 | 22 | 0.25 |
| polarC18 | 468.308 | 60.2699 | 22 | 0.25 |
| polarC18 | 468.308 | 61.7713 | 22 | 0.25 |
| polarC18 | 468.308 | 366.2629 | 22 | 0.25 |
| polarC18 | 468.308 | 233.5459 | 22 | 0.25 |
| polarC18 | 468.308 | 332.8922 | 22 | 0.25 |
| polarC18 | 468.308 | 176.2324 | 22 | 0.25 |
| polarC18 | 468.308 | 94.1948 | 22 | 0.25 |
| polarC18 | 468.308 | 232.9766 | 22 | 0.25 |
| polarC18 | 468.308 | 138.1187 | 21.9 | 0.25 |
| polarC18 | 468.308 | 312.1454 | 21.9 | 0.25 |
| polarC18 | 468.308 | 201.2519 | 21.9 | 0.25 |
| polarC18 | 468.308 | 101.8717 | 21.9 | 0.25 |
| polarC18 | 468.308 | 271.3133 | 21.9 | 0.25 |
| polarC18 | 468.308 | 125.6858 | 21.9 | 0.25 |
| polarC18 | 468.308 | 130.5914 | 21.9 | 0.25 |
| polarC18 | 468.308 | 300.5633 | 21.9 | 0.25 |
| polarC18 | 468.308 | 163.026 | 21.9 | 0.25 |
| polarC18 | 468.308 | 200.4805 | 21.9 | 0.25 |
| polarC18 | 468.308 | 64.0874 | 21.9 | 0.25 |
| polarC18 | 468.308 | 87.7258 | 21.9 | 0.25 |
| polarC18 | 468.308 | 103.8824 | 21.9 | 0.25 |
| polarC18 | 468.308 | 88.6389 | 21.9 | 0.25 |
| polarC18 | 468.308 | 292.0502 | 21.9 | 0.25 |
| polarC18 | 468.308 | 208.87 | 21.9 | 0.25 |
| polarC18 | 468.308 | 301.8828 | 21.9 | 0.25 |

| | | | | |
|---|---|---|---|---|
| polarC18 | 468.308 | 117.0384 | 21.9 | 0.25 |
| polarC18 | 468.308 | 114.4779 | 21.9 | 0.25 |
| polarC18 | 468.308 | 234.5852 | 21.9 | 0.25 |
| polarC18 | 468.308 | 86.9664 | 21.9 | 0.25 |
| polarC18 | 468.308 | 51.4334 | 21.9 | 0.25 |
| polarC18 | 468.308 | 54.8476 | 21.9 | 0.25 |
| polarC18 | 468.308 | 72.1291 | 21.9 | 0.25 |
| polarC18 | 468.308 | 238.458 | 21.9 | 0.25 |
| polarC18 | 468.308 | 75.3195 | 21.9 | 0.25 |
| polarC18 | 468.308 | 117.6808 | 21.9 | 0.25 |
| polarC18 | 468.308 | 140.9316 | 21.9 | 0.25 |
| polarC18 | 468.308 | 242.0085 | 21.9 | 0.25 |
| polarC18 | 468.308 | 244.6 | 21.9 | 0.25 |
| polarC18 | 468.308 | 100.156 | 21.9 | 0.25 |
| polarC18 | 468.308 | 164.0128 | 21.9 | 0.25 |
| polarC18 | 468.308 | 59.1383 | 21.9 | 0.25 |
| polarC18 | 468.308 | 156.4998 | 21.9 | 0.25 |
| polarC18 | 468.308 | 83.6326 | 21.9 | 0.25 |
| polarC18 | 468.308 | 124.616 | 21.9 | 0.25 |
| polarC18 | 468.308 | 435.7445 | 21.9 | 0.25 |
| polarC18 | 468.308 | 70.2546 | 21.9 | 0.25 |
| polarC18 | 468.308 | 66.0009 | 21.9 | 0.25 |
| polarC18 | 468.308 | 152.7761 | 21.9 | 0.25 |
| polarC18 | 468.308 | 64.8587 | 21.9 | 0.25 |
| polarC18 | 468.308 | 89.2588 | 21.9 | 0.25 |
| polarC18 | 468.308 | 83.8877 | 21.9 | 0.25 |
| polarC18 | 468.308 | 102.4654 | 21.9 | 0.25 |
| polarC18 | 468.308 | 77.6707 | 21.9 | 0.25 |
| polarC18 | 468.308 | 59.7617 | 21.8 | 0.25 |
| polarC18 | 468.308 | 159.8308 | 21.8 | 0.25 |
| polarC18 | 468.308 | 263.2219 | 21.8 | 0.25 |
| polarC18 | 468.308 | 180.3358 | 21.8 | 0.25 |
| polarC18 | 468.308 | 53.9465 | 21.8 | 0.25 |
| polarC18 | 468.308 | 102.502 | 21.8 | 0.25 |
| polarC18 | 468.308 | 146.2791 | 21.8 | 0.25 |
| polarC18 | 468.308 | 189.2207 | 21.8 | 0.25 |
| polarC18 | 468.308 | 75.2887 | 21.8 | 0.25 |
| polarC18 | 468.308 | 111.5937 | 21.8 | 0.25 |
| polarC18 | 468.308 | 135.2646 | 21.8 | 0.25 |
| polarC18 | 468.308 | 56.3689 | 21.8 | 0.25 |
| polarC18 | 468.308 | 60.6904 | 21.8 | 0.25 |
| polarC18 | 468.308 | 68.6423 | 21.8 | 0.25 |

| | | | | |
|---|---|---|---|---|
| polarC18 | 468.308 | 205.5172 | 21.8 | 0.25 |
| polarC18 | 468.308 | 260.9944 | 21.8 | 0.25 |
| polarC18 | 468.308 | 280.1476 | 21.8 | 0.25 |
| polarC18 | 468.308 | 159.561 | 21.8 | 0.25 |
| polarC18 | 468.308 | 206.641 | 21.8 | 0.25 |
| polarC18 | 468.308 | 180.569 | 21.8 | 0.25 |
| polarC18 | 468.308 | 267.2639 | 21.8 | 0.25 |
| polarC18 | 468.308 | 303.9657 | 21.8 | 0.25 |
| polarC18 | 468.308 | 93.387 | 21.8 | 0.25 |
| polarC18 | 468.308 | 57.3486 | 21.8 | 0.25 |
| polarC18 | 468.308 | 302.9243 | 21.8 | 0.25 |
| polarC18 | 468.308 | 386.1646 | 21.8 | 0.25 |
| polarC18 | 468.308 | 154.5579 | 21.8 | 0.25 |
| polarC18 | 468.308 | 61.0211 | 21.8 | 0.25 |
| polarC18 | 468.308 | 204.5128 | 21.8 | 0.25 |
| polarC18 | 468.308 | 56.7528 | 21.8 | 0.25 |
| polarC18 | 468.308 | 54.3909 | 21.8 | 0.25 |
| polarC18 | 468.308 | 93.2381 | 21.8 | 0.25 |
| polarC18 | 468.308 | 98.5145 | 21.8 | 0.25 |
| polarC18 | 468.308 | 136.8892 | 21.8 | 0.25 |
| polarC18 | 468.308 | 51.5483 | 21.8 | 0.25 |
| polarC18 | 468.308 | 88.8465 | 21.8 | 0.25 |
| polarC18 | 468.308 | 178.4929 | 21.8 | 0.25 |
| polarC18 | 468.308 | 97.5254 | 21.8 | 0.25 |
| polarC18 | 468.308 | 52.0125 | 21.8 | 0.25 |
| polarC18 | 468.308 | 66.2774 | 21.8 | 0.25 |
| polarC18 | 468.308 | 148.6604 | 21.8 | 0.25 |
| polarC18 | 468.308 | 125.2204 | 21.8 | 0.25 |
| polarC18 | 468.308 | 58.8831 | 21.8 | 0.25 |
| polarC18 | 468.308 | 341.756 | 21.8 | 0.24 |
| polarC18 | 468.308 | 263.5951 | 21.8 | 0.24 |
| polarC18 | 468.308 | 257.4432 | 21.8 | 0.24 |
| polarC18 | 468.308 | 119.2272 | 21.8 | 0.24 |
| polarC18 | 468.308 | 236.4427 | 21.7 | 0.24 |
| polarC18 | 468.308 | 113.5296 | 21.7 | 0.24 |
| polarC18 | 468.308 | 223.0746 | 21.7 | 0.24 |
| polarC18 | 468.308 | 60.8608 | 21.7 | 0.24 |
| polarC18 | 468.308 | 57.7873 | 21.7 | 0.24 |
| polarC18 | 468.308 | 77.7263 | 21.7 | 0.24 |
| polarC18 | 468.308 | 277.2062 | 21.7 | 0.24 |
| polarC18 | 468.308 | 181.992 | 21.7 | 0.24 |
| polarC18 | 468.308 | 180.0887 | 21.7 | 0.24 |

| | | | | |
|---|---|---|---|---|
| polarC18 | 468.308 | 76.0114 | 21.7 | 0.24 |
| polarC18 | 468.308 | 300.616 | 21.7 | 0.24 |
| polarC18 | 468.308 | 260.4529 | 21.7 | 0.24 |
| polarC18 | 468.308 | 167.9736 | 21.7 | 0.24 |
| polarC18 | 468.308 | 51.4751 | 21.7 | 0.24 |
| polarC18 | 468.308 | 198.3734 | 21.7 | 0.24 |
| polarC18 | 468.308 | 78.1585 | 21.7 | 0.24 |
| polarC18 | 468.308 | 99.2727 | 21.7 | 0.24 |
| polarC18 | 468.308 | 62.8224 | 21.7 | 0.24 |
| polarC18 | 468.308 | 81.4825 | 21.7 | 0.24 |
| polarC18 | 468.308 | 93.9472 | 21.7 | 0.24 |
| polarC18 | 468.308 | 125.0943 | 21.7 | 0.24 |
| polarC18 | 468.308 | 437.6834 | 21.7 | 0.24 |
| polarC18 | 468.308 | 393.9295 | 21.7 | 0.24 |
| polarC18 | 468.308 | 92.2816 | 21.7 | 0.24 |
| polarC18 | 468.308 | 227.6839 | 21.7 | 0.24 |
| polarC18 | 468.308 | 54.1506 | 21.7 | 0.24 |
| polarC18 | 468.308 | 66.3689 | 21.7 | 0.24 |
| polarC18 | 468.308 | 75.8044 | 21.7 | 0.24 |
| polarC18 | 468.308 | 182.8566 | 21.7 | 0.24 |
| polarC18 | 468.308 | 64.7848 | 21.7 | 0.24 |
| polarC18 | 468.308 | 54.2123 | 21.7 | 0.24 |
| polarC18 | 468.308 | 219.6673 | 21.7 | 0.24 |
| polarC18 | 468.308 | 97.7697 | 21.7 | 0.24 |
| polarC18 | 468.308 | 60.2093 | 21.7 | 0.24 |
| polarC18 | 468.308 | 64.2746 | 21.7 | 0.24 |
| polarC18 | 468.308 | 228.4372 | 21.7 | 0.24 |
| polarC18 | 468.308 | 402.1391 | 21.7 | 0.24 |
| polarC18 | 468.308 | 77.9408 | 21.7 | 0.24 |
| polarC18 | 468.308 | 332.8254 | 21.7 | 0.24 |
| polarC18 | 468.308 | 55.8654 | 21.7 | 0.24 |
| polarC18 | 468.308 | 90.7464 | 21.7 | 0.24 |
| polarC18 | 468.308 | 66.0202 | 21.7 | 0.24 |
| polarC18 | 468.308 | 299.1867 | 21.7 | 0.24 |
| polarC18 | 468.308 | 66.2232 | 21.7 | 0.24 |
| polarC18 | 468.308 | 105.6561 | 21.7 | 0.24 |
| polarC18 | 468.308 | 150.5394 | 21.7 | 0.24 |
| polarC18 | 468.308 | 244.0995 | 21.7 | 0.24 |
| polarC18 | 468.308 | 51.0989 | 21.7 | 0.24 |
| polarC18 | 468.308 | 82.9875 | 21.7 | 0.24 |
| polarC18 | 468.308 | 176.1422 | 21.7 | 0.24 |
| polarC18 | 468.308 | 216.1869 | 21.7 | 0.24 |

| | | | | |
|---|---|---|---|---|
| polarC18 | 468.308 | 74.8139 | 21.7 | 0.24 |
| polarC18 | 468.308 | 106.7293 | 21.7 | 0.24 |
| polarC18 | 468.308 | 74.2357 | 21.7 | 0.24 |
| polarC18 | 468.308 | 62.9127 | 21.7 | 0.24 |
| polarC18 | 468.308 | 181.0819 | 21.7 | 0.24 |
| polarC18 | 468.308 | 219.9338 | 21.7 | 0.24 |
| polarC18 | 468.308 | 204.0224 | 21.7 | 0.24 |
| polarC18 | 468.308 | 71.4805 | 21.7 | 0.24 |
| polarC18 | 468.308 | 134.7818 | 21.7 | 0.24 |
| polarC18 | 468.308 | 81.5659 | 21.6 | 0.24 |
| polarC18 | 468.308 | 119.5027 | 21.6 | 0.24 |
| polarC18 | 468.308 | 98.353 | 21.6 | 0.24 |
| polarC18 | 468.308 | 342.9684 | 21.6 | 0.24 |
| polarC18 | 468.308 | 63.7543 | 21.6 | 0.24 |
| polarC18 | 468.308 | 54.5269 | 21.6 | 0.24 |
| polarC18 | 468.308 | 77.6768 | 21.6 | 0.24 |
| polarC18 | 468.308 | 116.8921 | 21.6 | 0.24 |
| polarC18 | 468.308 | 203.8835 | 21.6 | 0.24 |
| polarC18 | 468.308 | 108.4195 | 21.6 | 0.24 |
| polarC18 | 468.308 | 105.7659 | 21.6 | 0.24 |
| polarC18 | 468.308 | 66.9996 | 21.6 | 0.24 |
| polarC18 | 468.308 | 77.4189 | 21.6 | 0.24 |
| polarC18 | 468.308 | 119.0381 | 21.6 | 0.24 |
| polarC18 | 468.308 | 233.6616 | 21.6 | 0.24 |
| polarC18 | 468.308 | 184.2471 | 21.6 | 0.24 |
| polarC18 | 468.308 | 134.9751 | 21.6 | 0.24 |
| polarC18 | 468.308 | 93.84 | 21.6 | 0.24 |
| polarC18 | 468.308 | 56.8424 | 21.6 | 0.24 |
| polarC18 | 468.308 | 188.7102 | 21.6 | 0.24 |
| polarC18 | 468.308 | 182.5863 | 21.6 | 0.24 |
| polarC18 | 468.308 | 53.295 | 21.6 | 0.24 |
| polarC18 | 468.308 | 94.0556 | 21.6 | 0.24 |
| polarC18 | 468.308 | 171.0287 | 21.6 | 0.24 |
| polarC18 | 468.308 | 50.7287 | 21.6 | 0.24 |
| polarC18 | 468.308 | 271.6341 | 21.6 | 0.24 |
| polarC18 | 468.308 | 56.7232 | 21.6 | 0.24 |
| polarC18 | 468.308 | 75.218 | 21.6 | 0.24 |
| polarC18 | 468.308 | 61.3999 | 21.6 | 0.24 |
| polarC18 | 468.308 | 204.2598 | 21.6 | 0.24 |
| polarC18 | 468.308 | 59.2555 | 21.6 | 0.24 |
| polarC18 | 468.308 | 73.234 | 21.6 | 0.24 |
| polarC18 | 468.308 | 68.8797 | 21.6 | 0.24 |

| | | | | |
|---|---|---|---|---|
| polarC18 | 468.308 | 122.0761 | 21.6 | 0.24 |
| polarC18 | 468.308 | 374.0331 | 21.6 | 0.24 |
| polarC18 | 468.308 | 100.5058 | 21.6 | 0.24 |
| polarC18 | 468.308 | 141.2804 | 21.6 | 0.24 |
| polarC18 | 468.308 | 136.3313 | 21.6 | 0.24 |
| polarC18 | 468.308 | 70.0865 | 21.6 | 0.24 |
| polarC18 | 468.308 | 266.8932 | 21.6 | 0.24 |
| polarC18 | 468.308 | 63.0157 | 21.6 | 0.24 |
| polarC18 | 468.308 | 112.5224 | 21.6 | 0.24 |
| polarC18 | 468.308 | 72.8876 | 21.6 | 0.24 |
| polarC18 | 468.308 | 141.27 | 21.6 | 0.24 |
| polarC18 | 468.308 | 68.019 | 21.6 | 0.24 |
| polarC18 | 468.308 | 56.0952 | 21.6 | 0.24 |
| polarC18 | 468.308 | 211.8345 | 21.6 | 0.24 |
| polarC18 | 468.308 | 72.0416 | 21.6 | 0.24 |
| polarC18 | 468.308 | 87.9938 | 21.6 | 0.24 |
| polarC18 | 468.308 | 65.0648 | 21.6 | 0.24 |
| polarC18 | 468.308 | 85.6995 | 21.6 | 0.24 |
| polarC18 | 468.308 | 183.6064 | 21.6 | 0.24 |
| polarC18 | 468.308 | 187.8314 | 21.6 | 0.24 |
| polarC18 | 468.308 | 98.8277 | 21.6 | 0.24 |
| polarC18 | 468.308 | 63.4529 | 21.6 | 0.24 |
| polarC18 | 468.308 | 58.0541 | 21.6 | 0.24 |
| polarC18 | 468.308 | 66.9509 | 21.6 | 0.24 |
| polarC18 | 468.308 | 81.9125 | 21.6 | 0.24 |
| polarC18 | 468.308 | 188.7573 | 21.6 | 0.24 |
| polarC18 | 468.308 | 130.765 | 21.6 | 0.24 |
| polarC18 | 468.308 | 136.5018 | 21.6 | 0.24 |
| polarC18 | 468.308 | 184.8752 | 21.6 | 0.24 |
| polarC18 | 468.308 | 57.2033 | 21.6 | 0.24 |
| polarC18 | 468.308 | 69.137 | 21.6 | 0.24 |
| polarC18 | 468.308 | 112.782 | 21.6 | 0.24 |
| polarC18 | 468.308 | 152.1345 | 21.6 | 0.24 |
| polarC18 | 468.308 | 304.2506 | 21.6 | 0.24 |
| polarC18 | 468.308 | 278.7076 | 21.6 | 0.24 |
| polarC18 | 468.308 | 180.0391 | 21.6 | 0.24 |
| polarC18 | 468.308 | 92.0606 | 21.6 | 0.24 |
| polarC18 | 468.308 | 60.6042 | 21.5 | 0.24 |
| polarC18 | 468.308 | 188.8929 | 21.5 | 0.24 |
| polarC18 | 468.308 | 102.393 | 21.5 | 0.24 |
| polarC18 | 468.308 | 105.7071 | 21.5 | 0.24 |
| polarC18 | 468.308 | 131.6299 | 21.5 | 0.24 |

| | | | | |
|---|---|---|---|---|
| polarC18 | 468.308 | 171.2624 | 21.5 | 0.24 |
| polarC18 | 468.308 | 106.8258 | 21.5 | 0.24 |
| polarC18 | 468.308 | 270.8834 | 21.5 | 0.24 |
| polarC18 | 468.308 | 57.0648 | 21.5 | 0.24 |
| polarC18 | 468.308 | 427.8057 | 21.5 | 0.24 |
| polarC18 | 468.308 | 69.0048 | 21.5 | 0.24 |
| polarC18 | 468.308 | 408.9008 | 21.5 | 0.24 |
| polarC18 | 468.308 | 52.8396 | 21.5 | 0.24 |
| polarC18 | 468.308 | 139.4294 | 21.5 | 0.24 |
| polarC18 | 468.308 | 68.886 | 21.5 | 0.24 |
| polarC18 | 468.308 | 429.4049 | 21.5 | 0.24 |
| polarC18 | 468.308 | 133.1156 | 21.5 | 0.24 |
| polarC18 | 468.308 | 190.0894 | 21.5 | 0.24 |
| polarC18 | 468.308 | 115.9246 | 21.5 | 0.24 |
| polarC18 | 468.308 | 151.5336 | 21.5 | 0.24 |
| polarC18 | 468.308 | 55.6904 | 21.5 | 0.24 |
| polarC18 | 468.308 | 141.6876 | 21.5 | 0.24 |
| polarC18 | 468.308 | 114.9214 | 21.5 | 0.24 |
| polarC18 | 468.308 | 59.6727 | 21.5 | 0.24 |
| polarC18 | 468.308 | 73.6313 | 21.5 | 0.24 |
| polarC18 | 468.308 | 330.18 | 21.5 | 0.24 |
| polarC18 | 468.308 | 284.1555 | 21.5 | 0.24 |
| polarC18 | 468.308 | 88.4345 | 21.5 | 0.24 |
| polarC18 | 468.308 | 68.5107 | 21.5 | 0.24 |
| polarC18 | 468.308 | 200.265 | 21.5 | 0.24 |
| polarC18 | 468.308 | 56.227 | 21.5 | 0.24 |
| polarC18 | 468.308 | 198.3378 | 21.5 | 0.24 |
| polarC18 | 468.308 | 158.0553 | 21.5 | 0.24 |
| polarC18 | 468.308 | 62.5215 | 21.5 | 0.24 |
| polarC18 | 468.308 | 462.4062 | 21.5 | 0.24 |
| polarC18 | 468.308 | 96.638 | 21.5 | 0.24 |
| polarC18 | 468.308 | 67.5321 | 21.5 | 0.24 |
| polarC18 | 468.308 | 89.2177 | 21.5 | 0.24 |
| polarC18 | 468.308 | 187.6843 | 21.5 | 0.24 |
| polarC18 | 468.308 | 96.8975 | 21.5 | 0.24 |
| polarC18 | 468.308 | 107.0609 | 21.5 | 0.24 |
| polarC18 | 468.308 | 467.5075 | 21.5 | 0.24 |
| polarC18 | 468.308 | 50.6348 | 21.5 | 0.24 |
| polarC18 | 468.308 | 151.2045 | 21.5 | 0.24 |
| polarC18 | 468.308 | 218.0506 | 21.5 | 0.24 |
| polarC18 | 468.308 | 112.3277 | 21.5 | 0.24 |
| polarC18 | 468.308 | 341.2253 | 21.5 | 0.24 |

| | | | | |
|---|---|---|---|---|
| polarC18 | 468.308 | 69.7918 | 21.5 | 0.24 |
| polarC18 | 468.308 | 74.9129 | 21.5 | 0.24 |
| polarC18 | 468.308 | 179.6392 | 21.5 | 0.24 |
| polarC18 | 468.308 | 104.2083 | 21.5 | 0.24 |
| polarC18 | 468.308 | 128.811 | 21.5 | 0.24 |
| polarC18 | 468.308 | 214.5225 | 21.4 | 0.24 |
| polarC18 | 468.308 | 58.3056 | 21.4 | 0.24 |
| polarC18 | 468.308 | 65.5039 | 21.4 | 0.24 |
| polarC18 | 468.308 | 56.3476 | 21.4 | 0.24 |
| polarC18 | 468.308 | 110.6353 | 21.4 | 0.24 |
| polarC18 | 468.308 | 75.7932 | 21.4 | 0.24 |
| polarC18 | 468.308 | 103.3631 | 21.4 | 0.24 |
| polarC18 | 468.308 | 328.2026 | 21.4 | 0.24 |
| polarC18 | 468.308 | 240.764 | 21.4 | 0.24 |
| polarC18 | 468.308 | 69.784 | 21.4 | 0.24 |
| polarC18 | 468.308 | 168.616 | 21.4 | 0.24 |
| polarC18 | 468.308 | 70.5227 | 21.4 | 0.24 |
| polarC18 | 468.308 | 51.2722 | 21.4 | 0.24 |
| polarC18 | 468.308 | 223.0194 | 21.4 | 0.24 |
| polarC18 | 468.308 | 363.4925 | 21.4 | 0.24 |
| polarC18 | 468.308 | 208.7934 | 21.4 | 0.24 |
| polarC18 | 468.308 | 86.221 | 21.4 | 0.24 |
| polarC18 | 468.308 | 73.5501 | 21.4 | 0.24 |
| polarC18 | 468.308 | 55.1594 | 21.4 | 0.24 |
| polarC18 | 468.308 | 163.0594 | 21.4 | 0.24 |
| polarC18 | 468.308 | 107.1382 | 21.4 | 0.24 |
| polarC18 | 468.308 | 265.0966 | 21.4 | 0.24 |
| polarC18 | 468.308 | 154.361 | 21.4 | 0.24 |
| polarC18 | 468.308 | 54.3346 | 21.4 | 0.24 |
| polarC18 | 468.308 | 83.7881 | 21.4 | 0.24 |
| polarC18 | 468.308 | 133.0403 | 21.4 | 0.24 |
| polarC18 | 468.308 | 63.3449 | 21.4 | 0.24 |
| polarC18 | 468.308 | 102.8901 | 21.4 | 0.24 |
| polarC18 | 468.308 | 185.1016 | 21.4 | 0.24 |
| polarC18 | 468.308 | 243.6326 | 21.4 | 0.24 |
| polarC18 | 468.308 | 67.5455 | 21.4 | 0.24 |
| polarC18 | 468.308 | 56.4087 | 21.4 | 0.24 |
| polarC18 | 468.308 | 332.6556 | 21.4 | 0.24 |
| polarC18 | 468.308 | 129.4829 | 21.4 | 0.24 |
| polarC18 | 468.308 | 238.3471 | 21.4 | 0.24 |
| polarC18 | 468.308 | 78.4846 | 21.4 | 0.24 |
| polarC18 | 468.308 | 342.1917 | 21.4 | 0.24 |

| | | | | |
|---|---|---|---|---|
| polarC18 | 468.308 | 219.503 | 21.4 | 0.24 |
| polarC18 | 468.308 | 78.1072 | 21.4 | 0.24 |
| polarC18 | 468.308 | 59.0083 | 21.4 | 0.24 |
| polarC18 | 468.308 | 80.6579 | 21.4 | 0.24 |
| polarC18 | 468.308 | 54.0473 | 21.4 | 0.24 |
| polarC18 | 468.308 | 134.1306 | 21.4 | 0.24 |
| polarC18 | 468.308 | 119.175 | 21.4 | 0.24 |
| polarC18 | 468.308 | 322.7256 | 21.4 | 0.24 |
| polarC18 | 468.308 | 81.8797 | 21.4 | 0.24 |
| polarC18 | 468.308 | 177.2813 | 21.4 | 0.24 |
| polarC18 | 468.308 | 103.9585 | 21.4 | 0.24 |
| polarC18 | 468.308 | 120.7494 | 21.4 | 0.24 |
| polarC18 | 468.308 | 70.8234 | 21.4 | 0.24 |
| polarC18 | 468.308 | 174.3208 | 21.4 | 0.24 |
| polarC18 | 468.308 | 104.0606 | 21.4 | 0.24 |
| polarC18 | 468.308 | 493.5073 | 21.4 | 0.24 |
| polarC18 | 468.308 | 53.2689 | 21.4 | 0.24 |
| polarC18 | 468.308 | 129.258 | 21.4 | 0.24 |
| polarC18 | 468.308 | 139.9807 | 21.4 | 0.24 |
| polarC18 | 468.308 | 127.0109 | 21.4 | 0.24 |
| polarC18 | 468.308 | 144.1276 | 21.3 | 0.24 |
| polarC18 | 468.308 | 140.5208 | 21.3 | 0.24 |
| polarC18 | 468.308 | 112.0997 | 21.3 | 0.24 |
| polarC18 | 468.308 | 154.2353 | 21.3 | 0.24 |
| polarC18 | 468.308 | 390.5369 | 21.3 | 0.24 |
| polarC18 | 468.308 | 51.8102 | 21.3 | 0.24 |
| polarC18 | 468.308 | 94.4726 | 21.3 | 0.24 |
| polarC18 | 468.308 | 164.6828 | 21.3 | 0.24 |
| polarC18 | 468.308 | 115.6404 | 21.3 | 0.24 |
| polarC18 | 468.308 | 73.0528 | 21.3 | 0.24 |
| polarC18 | 468.308 | 73.2075 | 21.3 | 0.24 |
| polarC18 | 468.308 | 127.4581 | 21.3 | 0.24 |
| polarC18 | 468.308 | 53.5641 | 21.3 | 0.24 |
| polarC18 | 468.308 | 95.749 | 21.3 | 0.24 |
| polarC18 | 468.308 | 82.8917 | 21.3 | 0.24 |
| polarC18 | 468.308 | 161.6608 | 21.3 | 0.24 |
| polarC18 | 468.308 | 61.5294 | 21.3 | 0.24 |
| polarC18 | 468.308 | 95.833 | 21.3 | 0.24 |
| polarC18 | 468.308 | 149.7346 | 21.3 | 0.24 |
| polarC18 | 468.308 | 55.3794 | 21.3 | 0.24 |
| polarC18 | 468.308 | 96.7552 | 21.3 | 0.24 |
| polarC18 | 468.308 | 431.3527 | 21.3 | 0.24 |

| | | | | |
|---|---|---|---|---|
| polarC18 | 468.308 | 50.417 | 21.3 | 0.24 |
| polarC18 | 468.308 | 167.8534 | 21.3 | 0.24 |
| polarC18 | 468.308 | 89.3338 | 21.3 | 0.24 |
| polarC18 | 468.308 | 52.8278 | 21.3 | 0.24 |
| polarC18 | 468.308 | 62.9109 | 21.3 | 0.24 |
| polarC18 | 468.308 | 229.3102 | 21.3 | 0.24 |
| polarC18 | 468.308 | 118.9291 | 21.3 | 0.24 |
| polarC18 | 468.308 | 178.039 | 21.3 | 0.24 |
| polarC18 | 468.308 | 97.703 | 21.3 | 0.24 |
| polarC18 | 468.308 | 78.4152 | 21.3 | 0.24 |
| polarC18 | 468.308 | 88.0674 | 21.3 | 0.24 |
| polarC18 | 468.308 | 60.3827 | 21.3 | 0.24 |
| polarC18 | 468.308 | 79.9257 | 21.3 | 0.24 |
| polarC18 | 468.308 | 276.2005 | 21.3 | 0.24 |
| polarC18 | 468.308 | 389.8423 | 21.3 | 0.24 |
| polarC18 | 468.308 | 57.9173 | 21.3 | 0.24 |
| polarC18 | 468.308 | 183.5821 | 21.3 | 0.24 |
| polarC18 | 468.308 | 149.2876 | 21.3 | 0.24 |
| polarC18 | 468.308 | 50.0571 | 21.3 | 0.24 |
| polarC18 | 468.308 | 108.735 | 21.3 | 0.24 |
| polarC18 | 468.308 | 59.2712 | 21.3 | 0.24 |
| polarC18 | 468.308 | 103.5369 | 21.3 | 0.24 |
| polarC18 | 468.308 | 333.294 | 21.3 | 0.24 |
| polarC18 | 468.308 | 443.5121 | 21.3 | 0.24 |
| polarC18 | 468.308 | 155.6341 | 21.2 | 0.24 |
| polarC18 | 468.308 | 354.6027 | 21.2 | 0.24 |
| polarC18 | 468.308 | 357.1779 | 21.2 | 0.24 |
| polarC18 | 468.308 | 104.6335 | 21.2 | 0.24 |
| polarC18 | 468.308 | 261.9597 | 21.2 | 0.24 |
| polarC18 | 468.308 | 91.1992 | 21.2 | 0.24 |
| polarC18 | 468.308 | 135.5871 | 21.2 | 0.24 |
| polarC18 | 468.308 | 162.8393 | 21.2 | 0.24 |
| polarC18 | 468.308 | 98.3471 | 21.2 | 0.24 |
| polarC18 | 468.308 | 386.3451 | 21.2 | 0.24 |
| polarC18 | 468.308 | 252.405 | 21.2 | 0.24 |
| polarC18 | 468.308 | 170.7598 | 21.2 | 0.24 |
| polarC18 | 468.308 | 107.6355 | 21.2 | 0.24 |
| polarC18 | 468.308 | 192.3111 | 21.2 | 0.24 |
| polarC18 | 468.308 | 96.4354 | 21.2 | 0.24 |
| polarC18 | 468.308 | 51.2175 | 21.2 | 0.24 |
| polarC18 | 468.308 | 89.8171 | 21.2 | 0.24 |
| polarC18 | 468.308 | 406.1189 | 21.2 | 0.24 |

| | | | | |
|---|---|---|---|---|
| polarC18 | 468.308 | 132.3832 | 21.2 | 0.24 |
| polarC18 | 468.308 | 84.0662 | 21.2 | 0.24 |
| polarC18 | 468.308 | 390.3697 | 21.2 | 0.24 |
| polarC18 | 468.308 | 115.374 | 21.2 | 0.24 |
| polarC18 | 468.308 | 55.6157 | 21.2 | 0.24 |
| polarC18 | 468.308 | 125.3782 | 21.2 | 0.24 |
| polarC18 | 468.308 | 56.3112 | 21.2 | 0.24 |
| polarC18 | 468.308 | 101.8964 | 21.2 | 0.24 |
| polarC18 | 468.308 | 96.6277 | 21.2 | 0.24 |
| polarC18 | 468.308 | 64.2226 | 21.2 | 0.24 |
| polarC18 | 468.308 | 58.9365 | 21.2 | 0.24 |
| polarC18 | 468.308 | 182.9598 | 21.2 | 0.24 |
| polarC18 | 468.308 | 61.9738 | 21.2 | 0.24 |
| polarC18 | 468.308 | 61.989 | 21.2 | 0.24 |
| polarC18 | 468.308 | 417.1862 | 21.2 | 0.24 |
| polarC18 | 468.308 | 58.5189 | 21.2 | 0.24 |
| polarC18 | 468.308 | 65.6173 | 21.2 | 0.24 |
| polarC18 | 468.308 | 81.2552 | 21.2 | 0.24 |
| polarC18 | 468.308 | 349.8372 | 21.2 | 0.24 |
| polarC18 | 468.308 | 75.8975 | 21.2 | 0.24 |
| polarC18 | 468.308 | 54.2852 | 21.2 | 0.24 |
| polarC18 | 468.308 | 61.2194 | 21.2 | 0.24 |
| polarC18 | 468.308 | 134.8677 | 21.2 | 0.24 |
| polarC18 | 468.308 | 174.4038 | 21.2 | 0.24 |
| polarC18 | 468.308 | 170.5599 | 21.2 | 0.24 |
| polarC18 | 468.308 | 201.7469 | 21.2 | 0.24 |
| polarC18 | 468.308 | 94.4839 | 21.2 | 0.24 |
| polarC18 | 468.308 | 155.1474 | 21.2 | 0.24 |
| polarC18 | 468.308 | 389.0652 | 21.2 | 0.24 |
| polarC18 | 468.308 | 69.6703 | 21.2 | 0.24 |
| polarC18 | 468.308 | 472.9302 | 21.1 | 0.24 |
| polarC18 | 468.308 | 66.0473 | 21.1 | 0.24 |
| polarC18 | 468.308 | 70.1072 | 21.1 | 0.24 |
| polarC18 | 468.308 | 72.1772 | 21.1 | 0.24 |
| polarC18 | 468.308 | 407.7724 | 21.1 | 0.24 |
| polarC18 | 468.308 | 51.9704 | 21.1 | 0.24 |
| polarC18 | 468.308 | 60.8233 | 21.1 | 0.24 |
| polarC18 | 468.308 | 51.5164 | 21.1 | 0.24 |
| polarC18 | 468.308 | 111.3239 | 21.1 | 0.24 |
| polarC18 | 468.308 | 50.8429 | 21.1 | 0.24 |
| polarC18 | 468.308 | 59.6838 | 21.1 | 0.24 |
| polarC18 | 468.308 | 50.5001 | 21.1 | 0.24 |

| | | | | |
|---|---|---|---|---|
| polarC18 | 468.308 | 86.1611 | 21.1 | 0.24 |
| polarC18 | 468.308 | 89.8728 | 21.1 | 0.24 |
| polarC18 | 468.308 | 55.3946 | 21.1 | 0.24 |
| polarC18 | 468.308 | 122.4525 | 21.1 | 0.24 |
| polarC18 | 468.308 | 440.033 | 21.1 | 0.24 |
| polarC18 | 468.308 | 117.6628 | 21.1 | 0.24 |
| polarC18 | 468.308 | 100.639 | 21.1 | 0.24 |
| polarC18 | 468.308 | 55.8956 | 21.1 | 0.24 |
| polarC18 | 468.308 | 172.8248 | 21.1 | 0.24 |
| polarC18 | 468.308 | 186.5833 | 21.1 | 0.24 |
| polarC18 | 468.308 | 52.9649 | 21.1 | 0.24 |
| polarC18 | 468.308 | 57.8686 | 21.1 | 0.24 |
| polarC18 | 468.308 | 164.167 | 21.1 | 0.24 |
| polarC18 | 468.308 | 119.4399 | 21.1 | 0.24 |
| polarC18 | 468.308 | 283.3411 | 21.1 | 0.24 |
| polarC18 | 468.308 | 361.7004 | 21.1 | 0.24 |
| polarC18 | 468.308 | 119.0914 | 21.1 | 0.24 |
| polarC18 | 468.308 | 132.8176 | 21.1 | 0.24 |
| polarC18 | 468.308 | 53.344 | 21.1 | 0.24 |
| polarC18 | 468.308 | 57.3859 | 21.1 | 0.24 |
| polarC18 | 468.308 | 130.2538 | 21.1 | 0.24 |
| polarC18 | 468.308 | 276.8085 | 21.1 | 0.24 |
| polarC18 | 468.308 | 268.8676 | 21.1 | 0.24 |
| polarC18 | 468.308 | 51.2449 | 21.1 | 0.24 |
| polarC18 | 468.308 | 57.9844 | 21.1 | 0.24 |
| polarC18 | 468.308 | 172.0177 | 21.1 | 0.24 |
| polarC18 | 468.308 | 79.535 | 21.1 | 0.24 |
| polarC18 | 468.308 | 58.1222 | 21.1 | 0.24 |
| polarC18 | 468.308 | 183.8783 | 21.1 | 0.24 |
| polarC18 | 468.308 | 132.6837 | 21.1 | 0.24 |
| polarC18 | 468.308 | 109.7037 | 21.1 | 0.24 |
| polarC18 | 468.308 | 51.5497 | 21.1 | 0.24 |
| polarC18 | 468.308 | 55.5168 | 21.1 | 0.24 |
| polarC18 | 468.308 | 249.83 | 21.1 | 0.24 |
| polarC18 | 468.308 | 59.0567 | 21.1 | 0.24 |
| polarC18 | 468.308 | 261.2729 | 21.1 | 0.24 |
| polarC18 | 468.308 | 113.3814 | 21.1 | 0.24 |
| polarC18 | 468.308 | 122.5174 | 21.1 | 0.24 |
| polarC18 | 468.308 | 62.351 | 21.1 | 0.24 |
| polarC18 | 468.308 | 199.0871 | 21.1 | 0.24 |
| polarC18 | 468.308 | 91.5258 | 21.1 | 0.24 |
| polarC18 | 468.308 | 306.7923 | 21.1 | 0.24 |

| | | | | |
|---|---|---|---|---|
| polarC18 | 468.308 | 118.9913 | 21.1 | 0.24 |
| polarC18 | 468.308 | 273.5463 | 21.1 | 0.24 |
| polarC18 | 468.308 | 58.8274 | 21.1 | 0.24 |
| polarC18 | 468.308 | 64.6316 | 21.1 | 0.24 |
| polarC18 | 468.308 | 187.4779 | 21.1 | 0.24 |
| polarC18 | 468.308 | 147.1625 | 21.1 | 0.24 |
| polarC18 | 468.308 | 62.7436 | 21.1 | 0.24 |
| polarC18 | 468.308 | 65.2677 | 21.1 | 0.24 |
| polarC18 | 468.308 | 443.3982 | 21.1 | 0.24 |
| polarC18 | 468.308 | 337.6368 | 21.1 | 0.24 |
| polarC18 | 468.308 | 182.3581 | 21 | 0.24 |
| polarC18 | 468.308 | 121.2084 | 21 | 0.24 |
| polarC18 | 468.308 | 140.2473 | 21 | 0.24 |
| polarC18 | 468.308 | 67.1678 | 21 | 0.24 |
| polarC18 | 468.308 | 91.142 | 21 | 0.24 |
| polarC18 | 468.308 | 366.0882 | 21 | 0.24 |
| polarC18 | 468.308 | 68.4962 | 21 | 0.24 |
| polarC18 | 468.308 | 411.6447 | 21 | 0.24 |
| polarC18 | 468.308 | 124.2022 | 21 | 0.24 |
| polarC18 | 468.308 | 478.3507 | 21 | 0.24 |
| polarC18 | 468.308 | 136.7411 | 21 | 0.24 |
| polarC18 | 468.308 | 354.2864 | 21 | 0.24 |
| polarC18 | 468.308 | 281.4704 | 21 | 0.24 |
| polarC18 | 468.308 | 226.2681 | 21 | 0.24 |
| polarC18 | 468.308 | 197.678 | 21 | 0.24 |
| polarC18 | 468.308 | 73.623 | 21 | 0.24 |
| polarC18 | 468.308 | 99.47 | 21 | 0.24 |
| polarC18 | 468.308 | 78.1519 | 21 | 0.24 |
| polarC18 | 468.308 | 67.6474 | 21 | 0.24 |
| polarC18 | 468.308 | 309.5507 | 21 | 0.24 |
| polarC18 | 468.308 | 61.1252 | 21 | 0.24 |
| polarC18 | 468.308 | 310.4293 | 21 | 0.24 |
| polarC18 | 468.308 | 52.6717 | 21 | 0.24 |
| polarC18 | 468.308 | 75.3519 | 21 | 0.24 |
| polarC18 | 468.308 | 270.6387 | 21 | 0.24 |
| polarC18 | 468.308 | 361.8369 | 21 | 0.24 |
| polarC18 | 468.308 | 211.1634 | 21 | 0.24 |
| polarC18 | 468.308 | 141.1381 | 21 | 0.24 |
| polarC18 | 468.308 | 51.5191 | 21 | 0.24 |
| polarC18 | 468.308 | 87.6777 | 21 | 0.24 |
| polarC18 | 468.308 | 70.2877 | 21 | 0.24 |
| polarC18 | 468.308 | 118.4787 | 21 | 0.24 |

| | | | | |
|---|---|---|---|---|
| polarC18 | 468.308 | 93.3064 | 21 | 0.24 |
| polarC18 | 468.308 | 272.7503 | 21 | 0.24 |
| polarC18 | 468.308 | 72.8308 | 21 | 0.24 |
| polarC18 | 468.308 | 56.4597 | 21 | 0.24 |
| polarC18 | 468.308 | 76.8185 | 21 | 0.24 |
| polarC18 | 468.308 | 107.4132 | 21 | 0.24 |
| polarC18 | 468.308 | 153.156 | 21 | 0.24 |
| polarC18 | 468.308 | 144.6861 | 21 | 0.24 |
| polarC18 | 468.308 | 57.4817 | 21 | 0.24 |
| polarC18 | 468.308 | 144.06 | 21 | 0.24 |
| polarC18 | 468.308 | 143.7635 | 21 | 0.24 |
| polarC18 | 468.308 | 130.0252 | 21 | 0.24 |
| polarC18 | 468.308 | 86.4471 | 21 | 0.24 |
| polarC18 | 468.308 | 71.382 | 21 | 0.24 |
| polarC18 | 468.308 | 482.6339 | 21 | 0.24 |
| polarC18 | 468.308 | 82.7332 | 21 | 0.24 |
| polarC18 | 468.308 | 128.7068 | 21 | 0.24 |
| polarC18 | 468.308 | 95.0357 | 21 | 0.24 |
| polarC18 | 468.308 | 61.8206 | 21 | 0.24 |
| polarC18 | 468.308 | 99.6874 | 21 | 0.24 |
| polarC18 | 468.308 | 63.0542 | 21 | 0.24 |
| polarC18 | 468.308 | 62.1034 | 20.9 | 0.24 |
| polarC18 | 468.308 | 87.8389 | 20.9 | 0.24 |
| polarC18 | 468.308 | 96.2839 | 20.9 | 0.24 |
| polarC18 | 468.308 | 50.5971 | 20.9 | 0.24 |
| polarC18 | 468.308 | 125.3445 | 20.9 | 0.24 |
| polarC18 | 468.308 | 133.1695 | 20.9 | 0.24 |
| polarC18 | 468.308 | 411.8361 | 20.9 | 0.24 |
| polarC18 | 468.308 | 61.778 | 20.9 | 0.24 |
| polarC18 | 468.308 | 280.0584 | 20.9 | 0.24 |
| polarC18 | 468.308 | 114.8814 | 20.9 | 0.24 |
| polarC18 | 468.308 | 260.6913 | 20.9 | 0.24 |
| polarC18 | 468.308 | 95.1656 | 20.9 | 0.24 |
| polarC18 | 468.308 | 54.3972 | 20.9 | 0.24 |
| polarC18 | 468.308 | 84.1097 | 20.9 | 0.24 |
| polarC18 | 468.308 | 284.8289 | 20.9 | 0.24 |
| polarC18 | 468.308 | 363.0431 | 20.9 | 0.24 |
| polarC18 | 468.308 | 91.64 | 20.9 | 0.24 |
| polarC18 | 468.308 | 154.0239 | 20.9 | 0.24 |
| polarC18 | 468.308 | 54.0884 | 20.9 | 0.24 |
| polarC18 | 468.308 | 111.6614 | 20.9 | 0.24 |
| polarC18 | 468.308 | 417.44 | 20.9 | 0.24 |

| | | | | |
|---|---|---|---|---|
| polarC18 | 468.308 | 116.6035 | 20.9 | 0.24 |
| polarC18 | 468.308 | 135.8039 | 20.9 | 0.24 |
| polarC18 | 468.308 | 312.3218 | 20.9 | 0.24 |
| polarC18 | 468.308 | 347.313 | 20.9 | 0.24 |
| polarC18 | 468.308 | 146.101 | 20.9 | 0.24 |
| polarC18 | 468.308 | 67.2988 | 20.9 | 0.24 |
| polarC18 | 468.308 | 111.3414 | 20.9 | 0.24 |
| polarC18 | 468.308 | 353.917 | 20.9 | 0.24 |
| polarC18 | 468.308 | 407.3285 | 20.9 | 0.24 |
| polarC18 | 468.308 | 83.5097 | 20.9 | 0.24 |
| polarC18 | 468.308 | 61.812 | 20.9 | 0.24 |
| polarC18 | 468.308 | 73.211 | 20.9 | 0.24 |
| polarC18 | 468.308 | 245.8954 | 20.9 | 0.24 |
| polarC18 | 468.308 | 67.4588 | 20.9 | 0.24 |
| polarC18 | 468.308 | 56.2929 | 20.9 | 0.24 |
| polarC18 | 468.308 | 76.3049 | 20.9 | 0.24 |
| polarC18 | 468.308 | 126.144 | 20.9 | 0.24 |
| polarC18 | 468.308 | 92.7697 | 20.9 | 0.24 |
| polarC18 | 468.308 | 60.3196 | 20.9 | 0.24 |
| polarC18 | 468.308 | 56.5943 | 20.9 | 0.24 |
| polarC18 | 468.308 | 76.9453 | 20.9 | 0.24 |
| polarC18 | 468.308 | 436.4555 | 20.9 | 0.24 |
| polarC18 | 468.308 | 62.2796 | 20.9 | 0.24 |
| polarC18 | 468.308 | 360.6629 | 20.9 | 0.24 |
| polarC18 | 468.308 | 62.2295 | 20.9 | 0.24 |
| polarC18 | 468.308 | 220.4301 | 20.9 | 0.24 |
| polarC18 | 468.308 | 67.5046 | 20.9 | 0.24 |
| polarC18 | 468.308 | 93.1539 | 20.9 | 0.24 |
| polarC18 | 468.308 | 195.8067 | 20.9 | 0.24 |
| polarC18 | 468.308 | 55.5916 | 20.9 | 0.24 |
| polarC18 | 468.308 | 161.461 | 20.9 | 0.24 |
| polarC18 | 468.308 | 197.2548 | 20.9 | 0.23 |
| polarC18 | 468.308 | 72.5398 | 20.9 | 0.23 |
| polarC18 | 468.308 | 89.4487 | 20.9 | 0.23 |
| polarC18 | 468.308 | 193.4083 | 20.9 | 0.23 |
| polarC18 | 468.308 | 89.2701 | 20.9 | 0.23 |
| polarC18 | 468.308 | 81.201 | 20.9 | 0.23 |
| polarC18 | 468.308 | 440.8556 | 20.9 | 0.23 |
| polarC18 | 468.308 | 265.6044 | 20.9 | 0.23 |
| polarC18 | 468.308 | 61.6739 | 20.9 | 0.23 |
| polarC18 | 468.308 | 149.1566 | 20.9 | 0.23 |
| polarC18 | 468.308 | 104.1258 | 20.9 | 0.23 |

| | | | | |
|---|---|---|---|---|
| polarC18 | 468.308 | 82.4537 | 20.9 | 0.23 |
| polarC18 | 468.308 | 69.9228 | 20.9 | 0.23 |
| polarC18 | 468.308 | 225.9949 | 20.9 | 0.23 |
| polarC18 | 468.308 | 205.9609 | 20.8 | 0.23 |
| polarC18 | 468.308 | 306.4811 | 20.8 | 0.23 |
| polarC18 | 468.308 | 100.019 | 20.8 | 0.23 |
| polarC18 | 468.308 | 143.6703 | 20.8 | 0.23 |
| polarC18 | 468.308 | 51.6252 | 20.8 | 0.23 |
| polarC18 | 468.308 | 71.5059 | 20.8 | 0.23 |
| polarC18 | 468.308 | 55.8933 | 20.8 | 0.23 |
| polarC18 | 468.308 | 56.7591 | 20.8 | 0.23 |
| polarC18 | 468.308 | 205.6203 | 20.8 | 0.23 |
| polarC18 | 468.308 | 62.113 | 20.8 | 0.23 |
| polarC18 | 468.308 | 378.0396 | 20.8 | 0.23 |
| polarC18 | 468.308 | 211.7572 | 20.8 | 0.23 |
| polarC18 | 468.308 | 56.1774 | 20.8 | 0.23 |
| polarC18 | 468.308 | 122.1705 | 20.8 | 0.23 |
| polarC18 | 468.308 | 84.7914 | 20.8 | 0.23 |
| polarC18 | 468.308 | 50.2357 | 20.8 | 0.23 |
| polarC18 | 468.308 | 66.199 | 20.8 | 0.23 |
| polarC18 | 468.308 | 70.3793 | 20.8 | 0.23 |
| polarC18 | 468.308 | 66.9278 | 20.8 | 0.23 |
| polarC18 | 468.308 | 63.2828 | 20.8 | 0.23 |
| polarC18 | 468.308 | 166.7933 | 20.8 | 0.23 |
| polarC18 | 468.308 | 61.9147 | 20.8 | 0.23 |
| polarC18 | 468.308 | 53.8029 | 20.8 | 0.23 |
| polarC18 | 468.308 | 53.3941 | 20.8 | 0.23 |
| polarC18 | 468.308 | 294.5944 | 20.8 | 0.23 |
| polarC18 | 468.308 | 69.0531 | 20.8 | 0.23 |
| polarC18 | 468.308 | 66.0629 | 20.8 | 0.23 |
| polarC18 | 468.308 | 438.4096 | 20.8 | 0.23 |
| polarC18 | 468.308 | 142.509 | 20.8 | 0.23 |
| polarC18 | 468.308 | 165.5716 | 20.8 | 0.23 |
| polarC18 | 468.308 | 82.5247 | 20.8 | 0.23 |
| polarC18 | 468.308 | 57.2429 | 20.8 | 0.23 |
| polarC18 | 468.308 | 122.4253 | 20.8 | 0.23 |
| polarC18 | 468.308 | 95.025 | 20.8 | 0.23 |
| polarC18 | 468.308 | 158.3806 | 20.8 | 0.23 |
| polarC18 | 468.308 | 120.2436 | 20.8 | 0.23 |
| polarC18 | 468.308 | 100.6908 | 20.8 | 0.23 |
| polarC18 | 468.308 | 81.2859 | 20.8 | 0.23 |
| polarC18 | 468.308 | 53.5602 | 20.8 | 0.23 |

| | | | | |
|---|---|---|---|---|
| polarC18 | 468.308 | 53.1568 | 20.8 | 0.23 |
| polarC18 | 468.308 | 57.9522 | 20.8 | 0.23 |
| polarC18 | 468.308 | 59.9468 | 20.8 | 0.23 |
| polarC18 | 468.308 | 150.8636 | 20.8 | 0.23 |
| polarC18 | 468.308 | 97.9232 | 20.8 | 0.23 |
| polarC18 | 468.308 | 233.7036 | 20.8 | 0.23 |
| polarC18 | 468.308 | 318.4861 | 20.8 | 0.23 |
| polarC18 | 468.308 | 276.3835 | 20.8 | 0.23 |
| polarC18 | 468.308 | 103.7396 | 20.8 | 0.23 |
| polarC18 | 468.308 | 126.1523 | 20.8 | 0.23 |
| polarC18 | 468.308 | 240.7142 | 20.8 | 0.23 |
| polarC18 | 468.308 | 166.8613 | 20.8 | 0.23 |
| polarC18 | 468.308 | 50.0703 | 20.8 | 0.23 |
| polarC18 | 468.308 | 109.5132 | 20.8 | 0.23 |
| polarC18 | 468.308 | 123.6206 | 20.8 | 0.23 |
| polarC18 | 468.308 | 196.5521 | 20.8 | 0.23 |
| polarC18 | 468.308 | 86.9394 | 20.8 | 0.23 |
| polarC18 | 468.308 | 252.4953 | 20.8 | 0.23 |
| polarC18 | 468.308 | 147.297 | 20.8 | 0.23 |
| polarC18 | 468.308 | 154.8234 | 20.8 | 0.23 |
| polarC18 | 468.308 | 112.4252 | 20.8 | 0.23 |
| polarC18 | 468.308 | 211.521 | 20.8 | 0.23 |
| polarC18 | 468.308 | 92.4082 | 20.8 | 0.23 |
| polarC18 | 468.308 | 157.0011 | 20.8 | 0.23 |
| polarC18 | 468.308 | 135.9385 | 20.7 | 0.23 |
| polarC18 | 468.308 | 50.3659 | 20.7 | 0.23 |
| polarC18 | 468.308 | 121.1633 | 20.7 | 0.23 |
| polarC18 | 468.308 | 330.3323 | 20.7 | 0.23 |
| polarC18 | 468.308 | 436.8163 | 20.7 | 0.23 |
| polarC18 | 468.308 | 87.422 | 20.7 | 0.23 |
| polarC18 | 468.308 | 50.7933 | 20.7 | 0.23 |
| polarC18 | 468.308 | 392.5189 | 20.7 | 0.23 |
| polarC18 | 468.308 | 89.4357 | 20.7 | 0.23 |
| polarC18 | 468.308 | 60.5991 | 20.7 | 0.23 |
| polarC18 | 468.308 | 176.3472 | 20.7 | 0.23 |
| polarC18 | 468.308 | 399.3926 | 20.7 | 0.23 |
| polarC18 | 468.308 | 122.5438 | 20.7 | 0.23 |
| polarC18 | 468.308 | 50.6403 | 20.7 | 0.23 |
| polarC18 | 468.308 | 322.5182 | 20.7 | 0.23 |
| polarC18 | 468.308 | 111.0103 | 20.7 | 0.23 |
| polarC18 | 468.308 | 50.763 | 20.7 | 0.23 |
| polarC18 | 468.308 | 67.3325 | 20.7 | 0.23 |

| | | | | |
|---|---|---|---|---|
| polarC18 | 468.308 | 99.8759 | 20.7 | 0.23 |
| polarC18 | 468.308 | 63.8791 | 20.7 | 0.23 |
| polarC18 | 468.308 | 261.8812 | 20.7 | 0.23 |
| polarC18 | 468.308 | 53.0195 | 20.7 | 0.23 |
| polarC18 | 468.308 | 108.5063 | 20.7 | 0.23 |
| polarC18 | 468.308 | 159.424 | 20.7 | 0.23 |
| polarC18 | 468.308 | 114.4275 | 20.7 | 0.23 |
| polarC18 | 468.308 | 410.6857 | 20.7 | 0.23 |
| polarC18 | 468.308 | 64.993 | 20.7 | 0.23 |
| polarC18 | 468.308 | 61.7107 | 20.7 | 0.23 |
| polarC18 | 468.308 | 56.0609 | 20.7 | 0.23 |
| polarC18 | 468.308 | 342.7313 | 20.7 | 0.23 |
| polarC18 | 468.308 | 64.1551 | 20.7 | 0.23 |
| polarC18 | 468.308 | 146.9177 | 20.7 | 0.23 |
| polarC18 | 468.308 | 58.4024 | 20.7 | 0.23 |
| polarC18 | 468.308 | 64.6647 | 20.7 | 0.23 |
| polarC18 | 468.308 | 102.0423 | 20.7 | 0.23 |
| polarC18 | 468.308 | 222.9354 | 20.7 | 0.23 |
| polarC18 | 468.308 | 246.2561 | 20.7 | 0.23 |
| polarC18 | 468.308 | 169.0634 | 20.7 | 0.23 |
| polarC18 | 468.308 | 83.1302 | 20.7 | 0.23 |
| polarC18 | 468.308 | 89.3382 | 20.7 | 0.23 |
| polarC18 | 468.308 | 91.455 | 20.7 | 0.23 |
| polarC18 | 468.308 | 84.5652 | 20.7 | 0.23 |
| polarC18 | 468.308 | 120.6481 | 20.7 | 0.23 |
| polarC18 | 468.308 | 140.4652 | 20.7 | 0.23 |
| polarC18 | 468.308 | 57.0333 | 20.7 | 0.23 |
| polarC18 | 468.308 | 52.0351 | 20.7 | 0.23 |
| polarC18 | 468.308 | 220.7652 | 20.7 | 0.23 |
| polarC18 | 468.308 | 222.0069 | 20.7 | 0.23 |
| polarC18 | 468.308 | 70.8321 | 20.7 | 0.23 |
| polarC18 | 468.308 | 115.3916 | 20.7 | 0.23 |
| polarC18 | 468.308 | 133.3596 | 20.7 | 0.23 |
| polarC18 | 468.308 | 52.5358 | 20.7 | 0.23 |
| polarC18 | 468.308 | 188.5538 | 20.7 | 0.23 |
| polarC18 | 468.308 | 75.5947 | 20.7 | 0.23 |
| polarC18 | 468.308 | 66.2126 | 20.7 | 0.23 |
| polarC18 | 468.308 | 80.0332 | 20.7 | 0.23 |
| polarC18 | 468.308 | 50.4583 | 20.7 | 0.23 |
| polarC18 | 468.308 | 88.3341 | 20.7 | 0.23 |
| polarC18 | 468.308 | 71.6849 | 20.7 | 0.23 |
| polarC18 | 468.308 | 191.8156 | 20.6 | 0.23 |

| | | | | |
|---|---|---|---|---|
| polarC18 | 468.308 | 50.9453 | 20.6 | 0.23 |
| polarC18 | 468.308 | 58.5371 | 20.6 | 0.23 |
| polarC18 | 468.308 | 144.751 | 20.6 | 0.23 |
| polarC18 | 468.308 | 458.1146 | 20.6 | 0.23 |
| polarC18 | 468.308 | 88.7391 | 20.6 | 0.23 |
| polarC18 | 468.308 | 193.5633 | 20.6 | 0.23 |
| polarC18 | 468.308 | 68.4372 | 20.6 | 0.23 |
| polarC18 | 468.308 | 58.6079 | 20.6 | 0.23 |
| polarC18 | 468.308 | 94.7711 | 20.6 | 0.23 |
| polarC18 | 468.308 | 102.7963 | 20.6 | 0.23 |
| polarC18 | 468.308 | 278.9879 | 20.6 | 0.23 |
| polarC18 | 468.308 | 99.3606 | 20.6 | 0.23 |
| polarC18 | 468.308 | 242.4864 | 20.6 | 0.23 |
| polarC18 | 468.308 | 51.0814 | 20.6 | 0.23 |
| polarC18 | 468.308 | 69.0242 | 20.6 | 0.23 |
| polarC18 | 468.308 | 64.3877 | 20.6 | 0.23 |
| polarC18 | 468.308 | 94.156 | 20.6 | 0.23 |
| polarC18 | 468.308 | 115.1956 | 20.6 | 0.23 |
| polarC18 | 468.308 | 197.0449 | 20.6 | 0.23 |
| polarC18 | 468.308 | 55.8402 | 20.6 | 0.23 |
| polarC18 | 468.308 | 50.2882 | 20.6 | 0.23 |
| polarC18 | 468.308 | 124.532 | 20.6 | 0.23 |
| polarC18 | 468.308 | 63.8768 | 20.6 | 0.23 |
| polarC18 | 468.308 | 196.0077 | 20.6 | 0.23 |
| polarC18 | 468.308 | 110.6276 | 20.6 | 0.23 |
| polarC18 | 468.308 | 317.006 | 20.6 | 0.23 |
| polarC18 | 468.308 | 106.1821 | 20.6 | 0.23 |
| polarC18 | 468.308 | 61.4571 | 20.6 | 0.23 |
| polarC18 | 468.308 | 68.2805 | 20.6 | 0.23 |
| polarC18 | 468.308 | 248.3987 | 20.6 | 0.23 |
| polarC18 | 468.308 | 69.366 | 20.6 | 0.23 |
| polarC18 | 468.308 | 108.6652 | 20.6 | 0.23 |
| polarC18 | 468.308 | 178.5722 | 20.6 | 0.23 |
| polarC18 | 468.308 | 106.9072 | 20.6 | 0.23 |
| polarC18 | 468.308 | 71.1941 | 20.6 | 0.23 |
| polarC18 | 468.308 | 116.7063 | 20.6 | 0.23 |
| polarC18 | 468.308 | 56.965 | 20.6 | 0.23 |
| polarC18 | 468.308 | 80.9912 | 20.6 | 0.23 |
| polarC18 | 468.308 | 76.7737 | 20.6 | 0.23 |
| polarC18 | 468.308 | 91.8848 | 20.6 | 0.23 |
| polarC18 | 468.308 | 85.0292 | 20.6 | 0.23 |
| polarC18 | 468.308 | 96.196 | 20.6 | 0.23 |

| | | | | |
|---|---|---|---|---|
| polarC18 | 468.308 | 279.0602 | 20.5 | 0.23 |
| polarC18 | 468.308 | 114.7234 | 20.5 | 0.23 |
| polarC18 | 468.308 | 105.3272 | 20.5 | 0.23 |
| polarC18 | 468.308 | 250.3715 | 20.5 | 0.23 |
| polarC18 | 468.308 | 148.2138 | 20.5 | 0.23 |
| polarC18 | 468.308 | 78.2679 | 20.5 | 0.23 |
| polarC18 | 468.308 | 105.9121 | 20.5 | 0.23 |
| polarC18 | 468.308 | 59.4569 | 20.5 | 0.23 |
| polarC18 | 468.308 | 50.1733 | 20.5 | 0.23 |
| polarC18 | 468.308 | 73.2602 | 20.5 | 0.23 |
| polarC18 | 468.308 | 192.8828 | 20.5 | 0.23 |
| polarC18 | 468.308 | 174.54 | 20.5 | 0.23 |
| polarC18 | 468.308 | 117.1982 | 20.5 | 0.23 |
| polarC18 | 468.308 | 61.8961 | 20.5 | 0.23 |
| polarC18 | 468.308 | 105.1978 | 20.5 | 0.23 |
| polarC18 | 468.308 | 63.3655 | 20.5 | 0.23 |
| polarC18 | 468.308 | 54.8543 | 20.5 | 0.23 |
| polarC18 | 468.308 | 64.7922 | 20.5 | 0.23 |
| polarC18 | 468.308 | 113.1884 | 20.5 | 0.23 |
| polarC18 | 468.308 | 60.0045 | 20.5 | 0.23 |
| polarC18 | 468.308 | 57.3266 | 20.5 | 0.23 |
| polarC18 | 468.308 | 201.1974 | 20.5 | 0.23 |
| polarC18 | 468.308 | 94.0335 | 20.5 | 0.23 |
| polarC18 | 468.308 | 59.511 | 20.5 | 0.23 |
| polarC18 | 468.308 | 74.8491 | 20.5 | 0.23 |
| polarC18 | 468.308 | 131.9566 | 20.5 | 0.23 |
| polarC18 | 468.308 | 139.086 | 20.5 | 0.23 |
| polarC18 | 468.308 | 247.1275 | 20.5 | 0.23 |
| polarC18 | 468.308 | 108.2068 | 20.5 | 0.23 |
| polarC18 | 468.308 | 91.7983 | 20.5 | 0.23 |
| polarC18 | 468.308 | 57.3802 | 20.5 | 0.23 |
| polarC18 | 468.308 | 116.2704 | 20.5 | 0.23 |
| polarC18 | 468.308 | 82.9356 | 20.5 | 0.23 |
| polarC18 | 468.308 | 402.2924 | 20.5 | 0.23 |
| polarC18 | 468.308 | 450.308 | 20.5 | 0.23 |
| polarC18 | 468.308 | 240.2271 | 20.5 | 0.23 |
| polarC18 | 468.308 | 76.8585 | 20.5 | 0.23 |
| polarC18 | 468.308 | 70.6463 | 20.5 | 0.23 |
| polarC18 | 468.308 | 71.9325 | 20.5 | 0.23 |
| polarC18 | 468.308 | 127.7248 | 20.5 | 0.23 |
| polarC18 | 468.308 | 55.6699 | 20.5 | 0.23 |
| polarC18 | 468.308 | 133.4667 | 20.5 | 0.23 |

| | | | | |
|---|---|---|---|---|
| polarC18 | 468.308 | 95.0141 | 20.5 | 0.23 |
| polarC18 | 468.308 | 468.896 | 20.5 | 0.23 |
| polarC18 | 468.308 | 82.3593 | 20.5 | 0.23 |
| polarC18 | 468.308 | 163.7208 | 20.5 | 0.23 |
| polarC18 | 468.308 | 97.5807 | 20.5 | 0.23 |
| polarC18 | 468.308 | 110.1906 | 20.5 | 0.23 |
| polarC18 | 468.308 | 62.9282 | 20.5 | 0.23 |
| polarC18 | 468.308 | 361.9152 | 20.5 | 0.23 |
| polarC18 | 468.308 | 229.1768 | 20.5 | 0.23 |
| polarC18 | 468.308 | 50.2841 | 20.5 | 0.23 |
| polarC18 | 468.308 | 96.5982 | 20.5 | 0.23 |
| polarC18 | 468.308 | 102.3528 | 20.5 | 0.23 |
| polarC18 | 468.308 | 162.7073 | 20.5 | 0.23 |
| polarC18 | 468.308 | 52.7402 | 20.5 | 0.23 |
| polarC18 | 468.308 | 57.1383 | 20.4 | 0.23 |
| polarC18 | 468.308 | 135.9157 | 20.4 | 0.23 |
| polarC18 | 468.308 | 73.2142 | 20.4 | 0.23 |
| polarC18 | 468.308 | 55.5549 | 20.4 | 0.23 |
| polarC18 | 468.308 | 493.8023 | 20.4 | 0.23 |
| polarC18 | 468.308 | 178.5467 | 20.4 | 0.23 |
| polarC18 | 468.308 | 127.4064 | 20.4 | 0.23 |
| polarC18 | 468.308 | 126.4934 | 20.4 | 0.23 |
| polarC18 | 468.308 | 98.9137 | 20.4 | 0.23 |
| polarC18 | 468.308 | 132.6689 | 20.4 | 0.23 |
| polarC18 | 468.308 | 251.7081 | 20.4 | 0.23 |
| polarC18 | 468.308 | 50.2126 | 20.4 | 0.23 |
| polarC18 | 468.308 | 67.4469 | 20.4 | 0.23 |
| polarC18 | 468.308 | 62.4449 | 20.4 | 0.23 |
| polarC18 | 468.308 | 81.4902 | 20.4 | 0.23 |
| polarC18 | 468.308 | 55.1865 | 20.4 | 0.23 |
| polarC18 | 468.308 | 228.8562 | 20.4 | 0.23 |
| polarC18 | 468.308 | 431.9288 | 20.4 | 0.23 |
| polarC18 | 468.308 | 102.0709 | 20.4 | 0.23 |
| polarC18 | 468.308 | 142.489 | 20.4 | 0.23 |
| polarC18 | 468.308 | 54.207 | 20.4 | 0.23 |
| polarC18 | 468.308 | 175.546 | 20.4 | 0.23 |
| polarC18 | 468.308 | 139.2161 | 20.4 | 0.23 |
| polarC18 | 468.308 | 202.4957 | 20.4 | 0.23 |
| polarC18 | 468.308 | 81.9827 | 20.4 | 0.23 |
| polarC18 | 468.308 | 50.4601 | 20.4 | 0.23 |
| polarC18 | 468.308 | 77.5065 | 20.4 | 0.23 |
| polarC18 | 468.308 | 104.8895 | 20.4 | 0.23 |

| | | | | |
|---|---|---|---|---|
| polarC18 | 468.308 | 77.3475 | 20.4 | 0.23 |
| polarC18 | 468.308 | 95.2702 | 20.4 | 0.23 |
| polarC18 | 468.308 | 56.0891 | 20.4 | 0.23 |
| polarC18 | 468.308 | 76.8059 | 20.4 | 0.23 |
| polarC18 | 468.308 | 63.8033 | 20.4 | 0.23 |
| polarC18 | 468.308 | 67.4043 | 20.4 | 0.23 |
| polarC18 | 468.308 | 129.6523 | 20.4 | 0.23 |
| polarC18 | 468.308 | 205.7689 | 20.4 | 0.23 |
| polarC18 | 468.308 | 217.1901 | 20.4 | 0.23 |
| polarC18 | 468.308 | 313.3372 | 20.4 | 0.23 |
| polarC18 | 468.308 | 96.8435 | 20.4 | 0.23 |
| polarC18 | 468.308 | 80.0202 | 20.4 | 0.23 |
| polarC18 | 468.308 | 94.1883 | 20.4 | 0.23 |
| polarC18 | 468.308 | 197.1388 | 20.4 | 0.23 |
| polarC18 | 468.308 | 108.8695 | 20.4 | 0.23 |
| polarC18 | 468.308 | 54.575 | 20.4 | 0.23 |
| polarC18 | 468.308 | 331.4011 | 20.4 | 0.23 |
| polarC18 | 468.308 | 97.8023 | 20.4 | 0.23 |
| polarC18 | 468.308 | 54.9005 | 20.4 | 0.23 |
| polarC18 | 468.308 | 78.0109 | 20.4 | 0.23 |
| polarC18 | 468.308 | 298.5803 | 20.4 | 0.23 |
| polarC18 | 468.308 | 343.4525 | 20.4 | 0.23 |
| polarC18 | 468.308 | 113.4684 | 20.4 | 0.23 |
| polarC18 | 468.308 | 62.4302 | 20.4 | 0.23 |
| polarC18 | 468.308 | 186.4455 | 20.4 | 0.23 |
| polarC18 | 468.308 | 179.1434 | 20.4 | 0.23 |
| polarC18 | 468.308 | 53.7098 | 20.4 | 0.23 |
| polarC18 | 468.308 | 50.2772 | 20.4 | 0.23 |
| polarC18 | 468.308 | 51.6547 | 20.3 | 0.23 |
| polarC18 | 468.308 | 51.3499 | 20.3 | 0.23 |
| polarC18 | 468.308 | 97.2982 | 20.3 | 0.23 |
| polarC18 | 468.308 | 335.0381 | 20.3 | 0.23 |
| polarC18 | 468.308 | 176.5506 | 20.3 | 0.23 |
| polarC18 | 468.308 | 114.8414 | 20.3 | 0.23 |
| polarC18 | 468.308 | 62.3706 | 20.3 | 0.23 |
| polarC18 | 468.308 | 69.2055 | 20.3 | 0.23 |
| polarC18 | 468.308 | 196.6115 | 20.3 | 0.23 |
| polarC18 | 468.308 | 125.3263 | 20.3 | 0.23 |
| polarC18 | 468.308 | 93.3585 | 20.3 | 0.23 |
| polarC18 | 468.308 | 59.5003 | 20.3 | 0.23 |
| polarC18 | 468.308 | 80.5869 | 20.3 | 0.23 |
| polarC18 | 468.308 | 50.4798 | 20.3 | 0.23 |

| | | | | |
|---|---|---|---|---|
| polarC18 | 468.308 | 64.7702 | 20.3 | 0.23 |
| polarC18 | 468.308 | 159.4749 | 20.3 | 0.23 |
| polarC18 | 468.308 | 70.7607 | 20.3 | 0.23 |
| polarC18 | 468.308 | 70.2086 | 20.3 | 0.23 |
| polarC18 | 468.308 | 58.8691 | 20.3 | 0.23 |
| polarC18 | 468.308 | 162.3957 | 20.3 | 0.23 |
| polarC18 | 468.308 | 157.0584 | 20.3 | 0.23 |
| polarC18 | 468.308 | 71.7891 | 20.3 | 0.23 |
| polarC18 | 468.308 | 81.2201 | 20.3 | 0.23 |
| polarC18 | 468.308 | 69.8543 | 20.3 | 0.23 |
| polarC18 | 468.308 | 56.3234 | 20.3 | 0.23 |
| polarC18 | 468.308 | 94.0466 | 20.3 | 0.23 |
| polarC18 | 468.308 | 75.7439 | 20.3 | 0.23 |
| polarC18 | 468.308 | 58.8442 | 20.3 | 0.23 |
| polarC18 | 468.308 | 67.974 | 20.3 | 0.23 |
| polarC18 | 468.308 | 98.8383 | 20.3 | 0.23 |
| polarC18 | 468.308 | 117.2641 | 20.3 | 0.23 |
| polarC18 | 468.308 | 151.5191 | 20.3 | 0.23 |
| polarC18 | 468.308 | 78.0184 | 20.3 | 0.23 |
| polarC18 | 468.308 | 55.1009 | 20.3 | 0.23 |
| polarC18 | 468.308 | 263.4662 | 20.3 | 0.23 |
| polarC18 | 468.308 | 239.4522 | 20.3 | 0.23 |
| polarC18 | 468.308 | 134.9179 | 20.3 | 0.23 |
| polarC18 | 468.308 | 82.3727 | 20.3 | 0.23 |
| polarC18 | 468.308 | 73.1392 | 20.3 | 0.23 |
| polarC18 | 468.308 | 96.6504 | 20.3 | 0.23 |
| polarC18 | 468.308 | 83.8407 | 20.3 | 0.23 |
| polarC18 | 468.308 | 74.9502 | 20.3 | 0.23 |
| polarC18 | 468.308 | 68.7201 | 20.3 | 0.23 |
| polarC18 | 468.308 | 93.9031 | 20.3 | 0.23 |
| polarC18 | 468.308 | 78.9154 | 20.3 | 0.23 |
| polarC18 | 468.308 | 94.5299 | 20.3 | 0.23 |
| polarC18 | 468.308 | 457.3986 | 20.3 | 0.23 |
| polarC18 | 468.308 | 60.1845 | 20.3 | 0.23 |
| polarC18 | 468.308 | 67.9376 | 20.3 | 0.23 |
| polarC18 | 468.308 | 124.8577 | 20.3 | 0.23 |
| polarC18 | 468.308 | 392.7223 | 20.3 | 0.23 |
| polarC18 | 468.308 | 59.193 | 20.3 | 0.23 |
| polarC18 | 468.308 | 63.2537 | 20.3 | 0.23 |
| polarC18 | 468.308 | 51.0875 | 20.3 | 0.23 |
| polarC18 | 468.308 | 103.4111 | 20.3 | 0.23 |
| polarC18 | 468.308 | 372.5071 | 20.3 | 0.23 |

| | | | | |
|---|---|---|---|---|
| polarC18 | 468.308 | 318.9155 | 20.3 | 0.23 |
| polarC18 | 468.308 | 93.3778 | 20.3 | 0.23 |
| polarC18 | 468.308 | 69.5036 | 20.3 | 0.23 |
| polarC18 | 468.308 | 65.0675 | 20.3 | 0.23 |
| polarC18 | 468.308 | 79.3387 | 20.3 | 0.23 |
| polarC18 | 468.308 | 145.406 | 20.3 | 0.23 |
| polarC18 | 468.308 | 87.198 | 20.3 | 0.23 |
| polarC18 | 468.308 | 134.6619 | 20.2 | 0.23 |
| polarC18 | 468.308 | 141.8475 | 20.2 | 0.23 |
| polarC18 | 468.308 | 305.9405 | 20.2 | 0.23 |
| polarC18 | 468.308 | 136.4516 | 20.2 | 0.23 |
| polarC18 | 468.308 | 104.344 | 20.2 | 0.23 |
| polarC18 | 468.308 | 51.0289 | 20.2 | 0.23 |
| polarC18 | 468.308 | 112.4477 | 20.2 | 0.23 |
| polarC18 | 468.308 | 50.2234 | 20.2 | 0.23 |
| polarC18 | 468.308 | 140.1529 | 20.2 | 0.23 |
| polarC18 | 468.308 | 134.2552 | 20.2 | 0.23 |
| polarC18 | 468.308 | 151.8018 | 20.2 | 0.23 |
| polarC18 | 468.308 | 106.3914 | 20.2 | 0.23 |
| polarC18 | 468.308 | 71.3286 | 20.2 | 0.23 |
| polarC18 | 468.308 | 69.9292 | 20.2 | 0.23 |
| polarC18 | 468.308 | 182.0306 | 20.2 | 0.23 |
| polarC18 | 468.308 | 122.1621 | 20.2 | 0.23 |
| polarC18 | 468.308 | 187.3847 | 20.2 | 0.23 |
| polarC18 | 468.308 | 154.7978 | 20.2 | 0.23 |
| polarC18 | 468.308 | 60.9106 | 20.2 | 0.23 |
| polarC18 | 468.308 | 68.7242 | 20.2 | 0.23 |
| polarC18 | 468.308 | 52.9529 | 20.2 | 0.23 |
| polarC18 | 468.308 | 76.5199 | 20.2 | 0.23 |
| polarC18 | 468.308 | 130.1163 | 20.2 | 0.23 |
| polarC18 | 468.308 | 54.301 | 20.2 | 0.23 |
| polarC18 | 468.308 | 53.0158 | 20.2 | 0.23 |
| polarC18 | 468.308 | 150.112 | 20.2 | 0.23 |
| polarC18 | 468.308 | 60.7763 | 20.2 | 0.23 |
| polarC18 | 468.308 | 332.4831 | 20.2 | 0.23 |
| polarC18 | 468.308 | 58.3471 | 20.2 | 0.23 |
| polarC18 | 468.308 | 164.0707 | 20.2 | 0.23 |
| polarC18 | 468.308 | 74.821 | 20.2 | 0.23 |
| polarC18 | 468.308 | 102.7337 | 20.2 | 0.23 |
| polarC18 | 468.308 | 196.7903 | 20.2 | 0.23 |
| polarC18 | 468.308 | 59.2227 | 20.2 | 0.23 |
| polarC18 | 468.308 | 84.1668 | 20.2 | 0.23 |

| | | | | |
|---|---|---|---|---|
| polarC18 | 468.308 | 317.8764 | 20.2 | 0.23 |
| polarC18 | 468.308 | 121.3794 | 20.2 | 0.23 |
| polarC18 | 468.308 | 165.2933 | 20.2 | 0.23 |
| polarC18 | 468.308 | 67.0824 | 20.2 | 0.23 |
| polarC18 | 468.308 | 68.5528 | 20.2 | 0.23 |
| polarC18 | 468.308 | 55.4278 | 20.2 | 0.23 |
| polarC18 | 468.308 | 64.1499 | 20.2 | 0.23 |
| polarC18 | 468.308 | 86.1546 | 20.2 | 0.23 |
| polarC18 | 468.308 | 93.8792 | 20.2 | 0.23 |
| polarC18 | 468.308 | 93.7497 | 20.2 | 0.23 |
| polarC18 | 468.308 | 80.1948 | 20.2 | 0.23 |
| polarC18 | 468.308 | 56.0362 | 20.2 | 0.23 |
| polarC18 | 468.308 | 157.7027 | 20.2 | 0.23 |
| polarC18 | 468.308 | 93.3011 | 20.2 | 0.23 |
| polarC18 | 468.308 | 144.9835 | 20.2 | 0.23 |
| polarC18 | 468.308 | 90.5413 | 20.2 | 0.23 |
| polarC18 | 468.308 | 102.9456 | 20.2 | 0.23 |
| polarC18 | 468.308 | 410.8637 | 20.2 | 0.23 |
| polarC18 | 468.308 | 409.3585 | 20.2 | 0.23 |
| polarC18 | 468.308 | 150.7275 | 20.1 | 0.23 |
| polarC18 | 468.308 | 144.6382 | 20.1 | 0.23 |
| polarC18 | 468.308 | 91.0678 | 20.1 | 0.23 |
| polarC18 | 468.308 | 73.7337 | 20.1 | 0.23 |
| polarC18 | 468.308 | 100.7263 | 20.1 | 0.23 |
| polarC18 | 468.308 | 65.7073 | 20.1 | 0.23 |
| polarC18 | 468.308 | 70.6657 | 20.1 | 0.23 |
| polarC18 | 468.308 | 324.2548 | 20.1 | 0.23 |
| polarC18 | 468.308 | 85.6681 | 20.1 | 0.23 |
| polarC18 | 468.308 | 304.1125 | 20.1 | 0.23 |
| polarC18 | 468.308 | 118.0112 | 20.1 | 0.23 |
| polarC18 | 468.308 | 164.3033 | 20.1 | 0.23 |
| polarC18 | 468.308 | 239.7009 | 20.1 | 0.23 |
| polarC18 | 468.308 | 170.9828 | 20.1 | 0.23 |
| polarC18 | 468.308 | 85.2818 | 20.1 | 0.23 |
| polarC18 | 468.308 | 425.0166 | 20.1 | 0.23 |
| polarC18 | 468.308 | 80.4426 | 20.1 | 0.23 |
| polarC18 | 468.308 | 50.8165 | 20.1 | 0.23 |
| polarC18 | 468.308 | 71.0684 | 20.1 | 0.23 |
| polarC18 | 468.308 | 95.6792 | 20.1 | 0.23 |
| polarC18 | 468.308 | 61.0788 | 20.1 | 0.23 |
| polarC18 | 468.308 | 91.5778 | 20.1 | 0.23 |
| polarC18 | 468.308 | 235.4292 | 20.1 | 0.23 |

| | | | | |
|---|---|---|---|---|
| polarC18 | 468.308 | 77.1587 | 20.1 | 0.23 |
| polarC18 | 468.308 | 70.7842 | 20.1 | 0.23 |
| polarC18 | 468.308 | 51.0903 | 20.1 | 0.23 |
| polarC18 | 468.308 | 62.4634 | 20.1 | 0.23 |
| polarC18 | 468.308 | 62.4896 | 20.1 | 0.23 |
| polarC18 | 468.308 | 153.9922 | 20.1 | 0.23 |
| polarC18 | 468.308 | 69.8674 | 20.1 | 0.23 |
| polarC18 | 468.308 | 99.5678 | 20.1 | 0.23 |
| polarC18 | 468.308 | 248.7521 | 20.1 | 0.23 |
| polarC18 | 468.308 | 193.1591 | 20.1 | 0.23 |
| polarC18 | 468.308 | 136.0661 | 20.1 | 0.23 |
| polarC18 | 468.308 | 153.7648 | 20.1 | 0.23 |
| polarC18 | 468.308 | 83.326 | 20.1 | 0.23 |
| polarC18 | 468.308 | 83.0493 | 20.1 | 0.23 |
| polarC18 | 468.308 | 53.59 | 20.1 | 0.23 |
| polarC18 | 468.308 | 157.5623 | 20.1 | 0.23 |
| polarC18 | 468.308 | 94.7466 | 20.1 | 0.23 |
| polarC18 | 468.308 | 225.9657 | 20.1 | 0.23 |
| polarC18 | 468.308 | 140.1315 | 20.1 | 0.23 |
| polarC18 | 468.308 | 60.5053 | 20.1 | 0.23 |
| polarC18 | 468.308 | 50.2183 | 20.1 | 0.23 |
| polarC18 | 468.308 | 59.1576 | 20.1 | 0.23 |
| polarC18 | 468.308 | 190.8656 | 20.1 | 0.23 |
| polarC18 | 468.308 | 307.4372 | 20.1 | 0.23 |
| polarC18 | 468.308 | 76.4123 | 20.1 | 0.23 |
| polarC18 | 468.308 | 70.1938 | 20.1 | 0.23 |
| polarC18 | 468.308 | 101.149 | 20.1 | 0.23 |
| polarC18 | 468.308 | 104.7895 | 20.1 | 0.23 |
| polarC18 | 468.308 | 244.7926 | 20.1 | 0.23 |
| polarC18 | 468.308 | 53.1313 | 20.1 | 0.23 |
| polarC18 | 468.308 | 155.5686 | 20 | 0.23 |
| polarC18 | 468.308 | 131.0387 | 20 | 0.23 |
| polarC18 | 468.308 | 445.3337 | 20 | 0.23 |
| polarC18 | 468.308 | 96.727 | 20 | 0.23 |
| polarC18 | 468.308 | 94.0936 | 20 | 0.23 |
| polarC18 | 468.308 | 396.8084 | 20 | 0.23 |
| polarC18 | 468.308 | 53.6003 | 20 | 0.23 |
| polarC18 | 468.308 | 236.9185 | 20 | 0.23 |
| polarC18 | 468.308 | 60.5612 | 20 | 0.23 |
| polarC18 | 468.308 | 181.8743 | 20 | 0.23 |
| polarC18 | 468.308 | 77.7133 | 20 | 0.23 |
| polarC18 | 468.308 | 229.6694 | 20 | 0.23 |

| | | | | |
|---|---|---|---|---|
| polarC18 | 468.308 | 327.532 | 20 | 0.23 |
| polarC18 | 468.308 | 317.4301 | 20 | 0.23 |
| polarC18 | 468.308 | 204.096 | 20 | 0.23 |
| polarC18 | 468.308 | 62.5068 | 20 | 0.23 |
| polarC18 | 468.308 | 56.4677 | 20 | 0.23 |
| polarC18 | 468.308 | 211.329 | 20 | 0.23 |
| polarC18 | 468.308 | 245.1098 | 20 | 0.23 |
| polarC18 | 468.308 | 50.0528 | 20 | 0.23 |
| polarC18 | 468.308 | 207.0725 | 20 | 0.23 |
| polarC18 | 468.308 | 84.4773 | 20 | 0.23 |
| polarC18 | 468.308 | 136.295 | 20 | 0.23 |
| polarC18 | 468.308 | 103.4993 | 20 | 0.23 |
| polarC18 | 468.308 | 74.6055 | 20 | 0.23 |
| polarC18 | 468.308 | 125.3063 | 20 | 0.22 |
| polarC18 | 468.308 | 182.2111 | 20 | 0.22 |
| polarC18 | 468.308 | 53.6247 | 20 | 0.22 |
| polarC18 | 468.308 | 121.869 | 20 | 0.22 |
| polarC18 | 468.308 | 148.0041 | 20 | 0.22 |
| polarC18 | 468.308 | 177.5431 | 20 | 0.22 |
| polarC18 | 468.308 | 94.3012 | 20 | 0.22 |
| polarC18 | 468.308 | 158.6317 | 20 | 0.22 |
| polarC18 | 468.308 | 405.8903 | 20 | 0.22 |
| polarC18 | 468.308 | 78.3487 | 20 | 0.22 |
| polarC18 | 468.308 | 90.8025 | 20 | 0.22 |
| polarC18 | 468.308 | 75.4056 | 20 | 0.22 |
| polarC18 | 468.308 | 115.907 | 20 | 0.22 |
| polarC18 | 468.308 | 109.9634 | 20 | 0.22 |
| polarC18 | 468.308 | 54.4811 | 20 | 0.22 |
| polarC18 | 468.308 | 53.2361 | 20 | 0.22 |
| polarC18 | 468.308 | 85.6156 | 19.9 | 0.22 |
| polarC18 | 468.308 | 65.7039 | 19.9 | 0.22 |
| polarC18 | 468.308 | 88.1409 | 19.9 | 0.22 |
| polarC18 | 468.308 | 149.4601 | 19.9 | 0.22 |
| polarC18 | 468.308 | 196.4428 | 19.9 | 0.22 |
| polarC18 | 468.308 | 69.3175 | 19.9 | 0.22 |
| polarC18 | 468.308 | 70.3183 | 19.9 | 0.22 |
| polarC18 | 468.308 | 102.5502 | 19.9 | 0.22 |
| polarC18 | 468.308 | 69.9497 | 19.9 | 0.22 |
| polarC18 | 468.308 | 58.1816 | 19.9 | 0.22 |
| polarC18 | 468.308 | 59.7802 | 19.9 | 0.22 |
| polarC18 | 468.308 | 275.5868 | 19.9 | 0.22 |
| polarC18 | 468.308 | 80.8427 | 19.9 | 0.22 |

| | | | | |
|---|---|---|---|---|
| polarC18 | 468.308 | 85.9784 | 19.9 | 0.22 |
| polarC18 | 468.308 | 70.222 | 19.9 | 0.22 |
| polarC18 | 468.308 | 422.2505 | 19.9 | 0.22 |
| polarC18 | 468.308 | 62.4765 | 19.9 | 0.22 |
| polarC18 | 468.308 | 63.3807 | 19.9 | 0.22 |
| polarC18 | 468.308 | 84.7052 | 19.9 | 0.22 |
| polarC18 | 468.308 | 279.3845 | 19.9 | 0.22 |
| polarC18 | 468.308 | 62.9955 | 19.9 | 0.22 |
| polarC18 | 468.308 | 122.2628 | 19.9 | 0.22 |
| polarC18 | 468.308 | 83.7626 | 19.9 | 0.22 |
| polarC18 | 468.308 | 88.0585 | 19.9 | 0.22 |
| polarC18 | 468.308 | 226.7879 | 19.9 | 0.22 |
| polarC18 | 468.308 | 154.1642 | 19.9 | 0.22 |
| polarC18 | 468.308 | 216.653 | 19.9 | 0.22 |
| polarC18 | 468.308 | 64.2313 | 19.9 | 0.22 |
| polarC18 | 468.308 | 58.5213 | 19.9 | 0.22 |
| polarC18 | 468.308 | 51.8626 | 19.9 | 0.22 |
| polarC18 | 468.308 | 363.3841 | 19.9 | 0.22 |
| polarC18 | 468.308 | 50.5993 | 19.9 | 0.22 |
| polarC18 | 468.308 | 69.7736 | 19.9 | 0.22 |
| polarC18 | 468.308 | 120.4619 | 19.8 | 0.22 |
| polarC18 | 468.308 | 173.7757 | 19.8 | 0.22 |
| polarC18 | 468.308 | 73.4608 | 19.8 | 0.22 |
| polarC18 | 468.308 | 253.9705 | 19.8 | 0.22 |
| polarC18 | 468.308 | 352.6288 | 19.8 | 0.22 |
| polarC18 | 468.308 | 122.0115 | 19.8 | 0.22 |
| polarC18 | 468.308 | 91.7243 | 19.8 | 0.22 |
| polarC18 | 468.308 | 60.1301 | 19.8 | 0.22 |
| polarC18 | 468.308 | 60.6316 | 19.8 | 0.22 |
| polarC18 | 468.308 | 73.4495 | 19.8 | 0.22 |
| polarC18 | 468.308 | 130.8953 | 19.8 | 0.22 |
| polarC18 | 468.308 | 58.1156 | 19.8 | 0.22 |
| polarC18 | 468.308 | 244.8382 | 19.8 | 0.22 |
| polarC18 | 468.308 | 59.314 | 19.8 | 0.22 |
| polarC18 | 468.308 | 55.5668 | 19.8 | 0.22 |
| polarC18 | 468.308 | 188.1598 | 19.8 | 0.22 |
| polarC18 | 468.308 | 101.2783 | 19.8 | 0.22 |
| polarC18 | 468.308 | 370.4878 | 19.8 | 0.22 |
| polarC18 | 468.308 | 93.7004 | 19.8 | 0.22 |
| polarC18 | 468.308 | 51.5036 | 19.8 | 0.22 |
| polarC18 | 468.308 | 138.2141 | 19.8 | 0.22 |
| polarC18 | 468.308 | 68.6689 | 19.8 | 0.22 |

| | | | | |
|---|---|---|---|---|
| polarC18 | 468.308 | 72.2766 | 19.8 | 0.22 |
| polarC18 | 468.308 | 69.2898 | 19.8 | 0.22 |
| polarC18 | 468.308 | 68.6831 | 19.8 | 0.22 |
| polarC18 | 468.308 | 67.9148 | 19.8 | 0.22 |
| polarC18 | 468.308 | 198.7838 | 19.8 | 0.22 |
| polarC18 | 468.308 | 179.0125 | 19.8 | 0.22 |
| polarC18 | 468.308 | 85.0365 | 19.8 | 0.22 |
| polarC18 | 468.308 | 138.493 | 19.8 | 0.22 |
| polarC18 | 468.308 | 53.8707 | 19.8 | 0.22 |
| polarC18 | 468.308 | 59.3964 | 19.8 | 0.22 |
| polarC18 | 468.308 | 85.1147 | 19.8 | 0.22 |
| polarC18 | 468.308 | 94.6362 | 19.8 | 0.22 |
| polarC18 | 468.308 | 99.9566 | 19.8 | 0.22 |
| polarC18 | 468.308 | 86.1708 | 19.8 | 0.22 |
| polarC18 | 468.308 | 65.718 | 19.7 | 0.22 |
| polarC18 | 468.308 | 282.552 | 19.7 | 0.22 |
| polarC18 | 468.308 | 55.6011 | 19.7 | 0.22 |
| polarC18 | 468.308 | 52.6974 | 19.7 | 0.22 |
| polarC18 | 468.308 | 56.3756 | 19.7 | 0.22 |
| polarC18 | 468.308 | 114.7607 | 19.7 | 0.22 |
| polarC18 | 468.308 | 169.442 | 19.7 | 0.22 |
| polarC18 | 468.308 | 122.4997 | 19.7 | 0.22 |
| polarC18 | 468.308 | 60.7221 | 19.7 | 0.22 |
| polarC18 | 468.308 | 59.3014 | 19.7 | 0.22 |
| polarC18 | 468.308 | 101.6599 | 19.7 | 0.22 |
| polarC18 | 468.308 | 178.8333 | 19.7 | 0.22 |
| polarC18 | 468.308 | 120.449 | 19.7 | 0.22 |
| polarC18 | 468.308 | 58.743 | 19.7 | 0.22 |
| polarC18 | 468.308 | 57.1395 | 19.7 | 0.22 |
| polarC18 | 468.308 | 63.1228 | 19.7 | 0.22 |
| polarC18 | 468.308 | 219.1438 | 19.7 | 0.22 |
| polarC18 | 468.308 | 322.7864 | 19.7 | 0.22 |
| polarC18 | 468.308 | 196.7639 | 19.7 | 0.22 |
| polarC18 | 468.308 | 61.3145 | 19.7 | 0.22 |
| polarC18 | 468.308 | 110.3463 | 19.7 | 0.22 |
| polarC18 | 468.308 | 116.8608 | 19.7 | 0.22 |
| polarC18 | 468.308 | 51.007 | 19.7 | 0.22 |
| polarC18 | 468.308 | 90.9282 | 19.7 | 0.22 |
| polarC18 | 468.308 | 107.8326 | 19.7 | 0.22 |
| polarC18 | 468.308 | 109.1442 | 19.7 | 0.22 |
| polarC18 | 468.308 | 106.0156 | 19.7 | 0.22 |
| polarC18 | 468.308 | 115.5676 | 19.7 | 0.22 |

| | | | | |
|---|---|---|---|---|
| polarC18 | 468.308 | 76.0319 | 19.7 | 0.22 |
| polarC18 | 468.308 | 107.6661 | 19.6 | 0.22 |
| polarC18 | 468.308 | 151.9883 | 19.6 | 0.22 |
| polarC18 | 468.308 | 433.7669 | 19.6 | 0.22 |
| polarC18 | 468.308 | 65.4496 | 19.6 | 0.22 |
| polarC18 | 468.308 | 356.191 | 19.6 | 0.22 |
| polarC18 | 468.308 | 54.1304 | 19.6 | 0.22 |
| polarC18 | 468.308 | 56.1497 | 19.6 | 0.22 |
| polarC18 | 468.308 | 85.8569 | 19.6 | 0.22 |
| polarC18 | 468.308 | 66.6175 | 19.6 | 0.22 |
| polarC18 | 468.308 | 69.1618 | 19.6 | 0.22 |
| polarC18 | 468.308 | 67.8611 | 19.6 | 0.22 |
| polarC18 | 468.308 | 55.715 | 19.6 | 0.22 |
| polarC18 | 468.308 | 72.9472 | 19.6 | 0.22 |
| polarC18 | 468.308 | 385.5531 | 19.6 | 0.22 |
| polarC18 | 468.308 | 64.0956 | 19.6 | 0.22 |
| polarC18 | 468.308 | 160.2682 | 19.6 | 0.22 |
| polarC18 | 468.308 | 60.109 | 19.6 | 0.22 |
| polarC18 | 468.308 | 129.947 | 19.6 | 0.22 |
| polarC18 | 468.308 | 289.3478 | 19.6 | 0.22 |
| polarC18 | 468.308 | 50.9978 | 19.6 | 0.22 |
| polarC18 | 468.308 | 108.3654 | 19.6 | 0.22 |
| polarC18 | 468.308 | 174.4787 | 19.6 | 0.22 |
| polarC18 | 468.308 | 192.4237 | 19.6 | 0.22 |
| polarC18 | 468.308 | 96.9226 | 19.6 | 0.22 |
| polarC18 | 468.308 | 55.9085 | 19.6 | 0.22 |
| polarC18 | 468.308 | 52.1104 | 19.6 | 0.22 |
| polarC18 | 468.308 | 271.0214 | 19.6 | 0.22 |
| polarC18 | 468.308 | 76.3779 | 19.6 | 0.22 |
| polarC18 | 468.308 | 87.5401 | 19.6 | 0.22 |
| polarC18 | 468.308 | 217.4203 | 19.6 | 0.22 |
| polarC18 | 468.308 | 428.6805 | 19.5 | 0.22 |
| polarC18 | 468.308 | 308.4101 | 19.5 | 0.22 |
| polarC18 | 468.308 | 116.0899 | 19.5 | 0.22 |
| polarC18 | 468.308 | 56.6475 | 19.5 | 0.22 |
| polarC18 | 468.308 | 96.2762 | 19.5 | 0.22 |
| polarC18 | 468.308 | 56.7034 | 19.5 | 0.22 |
| polarC18 | 468.308 | 52.8185 | 19.5 | 0.22 |
| polarC18 | 468.308 | 64.8118 | 19.5 | 0.22 |
| polarC18 | 468.308 | 144.994 | 19.5 | 0.22 |
| polarC18 | 468.308 | 53.265 | 19.5 | 0.22 |
| polarC18 | 468.308 | 112.1451 | 19.5 | 0.22 |

| | | | | |
|---|---|---|---|---|
| polarC18 | 468.308 | 102.9017 | 19.5 | 0.22 |
| polarC18 | 468.308 | 58.7642 | 19.5 | 0.22 |
| polarC18 | 468.308 | 142.2714 | 19.5 | 0.22 |
| polarC18 | 468.308 | 88.6826 | 19.5 | 0.22 |
| polarC18 | 468.308 | 77.0768 | 19.5 | 0.22 |
| polarC18 | 468.308 | 59.308 | 19.5 | 0.22 |
| polarC18 | 468.308 | 66.5927 | 19.5 | 0.22 |
| polarC18 | 468.308 | 85.6013 | 19.5 | 0.22 |
| polarC18 | 468.308 | 66.1674 | 19.5 | 0.22 |
| polarC18 | 468.308 | 77.9061 | 19.5 | 0.22 |
| polarC18 | 468.308 | 92.1095 | 19.5 | 0.22 |
| polarC18 | 468.308 | 50.6602 | 19.5 | 0.22 |
| polarC18 | 468.308 | 83.4019 | 19.5 | 0.22 |
| polarC18 | 468.308 | 175.3863 | 19.5 | 0.22 |
| polarC18 | 468.308 | 80.5098 | 19.5 | 0.22 |
| polarC18 | 468.308 | 55.0657 | 19.4 | 0.22 |
| polarC18 | 468.308 | 55.1775 | 19.4 | 0.22 |
| polarC18 | 468.308 | 221.6305 | 19.4 | 0.22 |
| polarC18 | 468.308 | 95.4492 | 19.4 | 0.22 |
| polarC18 | 468.308 | 72.6047 | 19.4 | 0.22 |
| polarC18 | 468.308 | 113.2957 | 19.4 | 0.22 |
| polarC18 | 468.308 | 74.8368 | 19.4 | 0.22 |
| polarC18 | 468.308 | 58.543 | 19.4 | 0.22 |
| polarC18 | 468.308 | 51.5523 | 19.4 | 0.22 |
| polarC18 | 468.308 | 62.9487 | 19.4 | 0.22 |
| polarC18 | 468.308 | 96.9959 | 19.4 | 0.22 |
| polarC18 | 468.308 | 52.8643 | 19.4 | 0.22 |
| polarC18 | 468.308 | 252.8692 | 19.4 | 0.22 |
| polarC18 | 468.308 | 67.6215 | 19.4 | 0.22 |
| polarC18 | 468.308 | 63.0742 | 19.4 | 0.22 |
| polarC18 | 468.308 | 169.1375 | 19.4 | 0.22 |
| polarC18 | 468.308 | 78.2926 | 19.4 | 0.22 |
| polarC18 | 468.308 | 59.024 | 19.4 | 0.22 |
| polarC18 | 468.308 | 117.7086 | 19.4 | 0.22 |
| polarC18 | 468.308 | 93.0786 | 19.4 | 0.22 |
| polarC18 | 468.308 | 70.0014 | 19.4 | 0.22 |
| polarC18 | 468.308 | 56.8651 | 19.4 | 0.22 |
| polarC18 | 468.308 | 58.5483 | 19.3 | 0.22 |
| polarC18 | 468.308 | 148.1196 | 19.3 | 0.22 |
| polarC18 | 468.308 | 87.8942 | 19.3 | 0.22 |
| polarC18 | 468.308 | 57.2338 | 19.3 | 0.22 |
| polarC18 | 468.308 | 65.435 | 19.3 | 0.22 |

| | | | | |
|---|---|---|---|---|
| polarC18 | 468.308 | 228.0926 | 19.3 | 0.22 |
| polarC18 | 468.308 | 63.4759 | 19.3 | 0.22 |
| polarC18 | 468.308 | 198.7106 | 19.3 | 0.22 |
| polarC18 | 468.308 | 78.4897 | 19.3 | 0.22 |
| polarC18 | 468.308 | 95.9346 | 19.3 | 0.22 |
| polarC18 | 468.308 | 329.7496 | 19.3 | 0.22 |
| polarC18 | 468.308 | 70.7577 | 19.3 | 0.22 |
| polarC18 | 468.308 | 199.3604 | 19.3 | 0.22 |
| polarC18 | 468.308 | 55.9608 | 19.3 | 0.22 |
| polarC18 | 468.308 | 145.3794 | 19.3 | 0.22 |
| polarC18 | 468.308 | 53.8952 | 19.3 | 0.22 |
| polarC18 | 468.308 | 407.0213 | 19.2 | 0.22 |
| polarC18 | 468.308 | 150.5144 | 19.2 | 0.22 |
| polarC18 | 468.308 | 60.6819 | 19.2 | 0.22 |
| polarC18 | 468.308 | 218.4332 | 19.2 | 0.22 |
| polarC18 | 468.308 | 200.7603 | 19.2 | 0.22 |
| polarC18 | 468.308 | 53.9835 | 19.2 | 0.22 |
| polarC18 | 468.308 | 79.216 | 19.2 | 0.22 |
| polarC18 | 468.308 | 110.3174 | 19.2 | 0.22 |
| polarC18 | 468.308 | 55.1918 | 19.2 | 0.22 |
| polarC18 | 468.308 | 51.1363 | 19.2 | 0.22 |
| polarC18 | 468.308 | 87.0098 | 19.2 | 0.22 |
| polarC18 | 468.308 | 87.5519 | 19.2 | 0.22 |
| polarC18 | 468.308 | 159.6512 | 19.2 | 0.22 |
| polarC18 | 468.308 | 88.4717 | 19.1 | 0.22 |
| polarC18 | 468.308 | 236.146 | 19.1 | 0.22 |
| polarC18 | 468.308 | 98.0259 | 19.1 | 0.22 |
| polarC18 | 468.308 | 78.3006 | 19.1 | 0.22 |
| polarC18 | 468.308 | 74.7698 | 19.1 | 0.22 |
| polarC18 | 468.308 | 112.0337 | 19.1 | 0.22 |
| polarC18 | 468.308 | 85.8711 | 19.1 | 0.22 |
| polarC18 | 468.308 | 110.4605 | 19.1 | 0.21 |
| polarC18 | 468.308 | 156.7113 | 19.1 | 0.21 |
| polarC18 | 468.308 | 137.3517 | 19.1 | 0.21 |
| polarC18 | 468.308 | 188.4427 | 19.1 | 0.21 |
| polarC18 | 468.308 | 63.9945 | 19.1 | 0.21 |
| polarC18 | 468.308 | 61.4708 | 19.1 | 0.21 |
| polarC18 | 468.308 | 169.0372 | 19.1 | 0.21 |
| polarC18 | 468.308 | 249.5892 | 19.1 | 0.21 |
| polarC18 | 468.308 | 57.0441 | 19.1 | 0.21 |
| polarC18 | 468.308 | 50.3208 | 19.1 | 0.21 |
| polarC18 | 468.308 | 55.6963 | 19 | 0.21 |

| | | | | |
|---|---|---|---|---|
| polarC18 | 468.308 | 152.0579 | 19 | 0.21 |
| polarC18 | 468.308 | 63.6238 | 19 | 0.21 |
| polarC18 | 468.308 | 328.5597 | 19 | 0.21 |
| polarC18 | 468.308 | 58.5308 | 19 | 0.21 |
| polarC18 | 468.308 | 152.5809 | 19 | 0.21 |
| polarC18 | 468.308 | 50.4962 | 19 | 0.21 |
| polarC18 | 468.308 | 110.529 | 19 | 0.21 |
| polarC18 | 468.308 | 56.5304 | 19 | 0.21 |
| polarC18 | 468.308 | 50.3492 | 19 | 0.21 |
| polarC18 | 468.308 | 67.5268 | 19 | 0.21 |
| polarC18 | 468.308 | 162.173 | 19 | 0.21 |
| polarC18 | 468.308 | 168.4149 | 19 | 0.21 |
| polarC18 | 468.308 | 67.8994 | 19 | 0.21 |
| polarC18 | 468.308 | 268.511 | 19 | 0.21 |
| polarC18 | 468.308 | 69.7946 | 19 | 0.21 |
| polarC18 | 468.308 | 65.1802 | 19 | 0.21 |
| polarC18 | 468.308 | 232.6887 | 18.9 | 0.21 |
| polarC18 | 468.308 | 76.2155 | 18.9 | 0.21 |
| polarC18 | 468.308 | 243.9615 | 18.9 | 0.21 |
| polarC18 | 468.308 | 269.2886 | 18.9 | 0.21 |
| polarC18 | 468.308 | 56.4147 | 18.9 | 0.21 |
| polarC18 | 468.308 | 83.1801 | 18.9 | 0.21 |
| polarC18 | 468.308 | 151.1067 | 18.9 | 0.21 |
| polarC18 | 468.308 | 61.1792 | 18.9 | 0.21 |
| polarC18 | 468.308 | 119.4901 | 18.9 | 0.21 |
| polarC18 | 468.308 | 112.8828 | 18.9 | 0.21 |
| polarC18 | 468.308 | 73.9553 | 18.9 | 0.21 |
| polarC18 | 468.308 | 88.8112 | 18.9 | 0.21 |
| polarC18 | 468.308 | 60.5713 | 18.9 | 0.21 |
| polarC18 | 468.308 | 66.2055 | 18.9 | 0.21 |
| polarC18 | 468.308 | 97.113 | 18.9 | 0.21 |
| polarC18 | 468.308 | 149.6714 | 18.9 | 0.21 |
| polarC18 | 468.308 | 206.2054 | 18.9 | 0.21 |
| polarC18 | 468.308 | 360.89 | 18.9 | 0.21 |
| polarC18 | 468.308 | 62.0295 | 18.9 | 0.21 |
| polarC18 | 468.308 | 188.2229 | 18.9 | 0.21 |
| polarC18 | 468.308 | 249.3853 | 18.9 | 0.21 |
| polarC18 | 468.308 | 74.8444 | 18.9 | 0.21 |
| polarC18 | 468.308 | 154.6034 | 18.8 | 0.21 |
| polarC18 | 468.308 | 84.9839 | 18.8 | 0.21 |
| polarC18 | 468.308 | 128.9626 | 18.8 | 0.21 |
| polarC18 | 468.308 | 117.1526 | 18.8 | 0.21 |

| | | | | |
|---|---|---|---|---|
| polarC18 | 468.308 | 127.7472 | 18.8 | 0.21 |
| polarC18 | 468.308 | 119.5211 | 18.8 | 0.21 |
| polarC18 | 468.308 | 62.433 | 18.8 | 0.21 |
| polarC18 | 468.308 | 56.3624 | 18.8 | 0.21 |
| polarC18 | 468.308 | 94.4883 | 18.7 | 0.21 |
| polarC18 | 468.308 | 127.141 | 18.7 | 0.21 |
| polarC18 | 468.308 | 209.4467 | 18.7 | 0.21 |
| polarC18 | 468.308 | 59.5263 | 18.7 | 0.21 |
| polarC18 | 468.308 | 58.389 | 18.7 | 0.21 |
| polarC18 | 468.308 | 145.7024 | 18.7 | 0.21 |
| polarC18 | 468.308 | 88.4544 | 18.7 | 0.21 |
| polarC18 | 468.308 | 89.9349 | 18.7 | 0.21 |
| polarC18 | 468.308 | 51.24 | 18.7 | 0.21 |
| polarC18 | 468.308 | 69.321 | 18.6 | 0.21 |
| polarC18 | 468.308 | 127.4379 | 18.6 | 0.21 |
| polarC18 | 468.308 | 343.7583 | 18.6 | 0.21 |
| polarC18 | 468.308 | 168.2458 | 18.6 | 0.21 |
| polarC18 | 468.308 | 104.6416 | 18.6 | 0.21 |
| polarC18 | 468.308 | 94.4632 | 18.6 | 0.21 |
| polarC18 | 468.308 | 53.982 | 18.6 | 0.21 |
| polarC18 | 468.308 | 109.5629 | 18.5 | 0.21 |
| polarC18 | 468.308 | 161.91 | 18.5 | 0.21 |
| polarC18 | 468.308 | 59.353 | 18.5 | 0.21 |
| polarC18 | 468.308 | 94.2957 | 18.5 | 0.21 |
| polarC18 | 468.308 | 119.1028 | 18.5 | 0.21 |
| polarC18 | 468.308 | 61.175 | 18.5 | 0.21 |
| polarC18 | 468.308 | 63.1657 | 18.5 | 0.21 |
| polarC18 | 468.308 | 83.9677 | 18.5 | 0.21 |
| polarC18 | 468.308 | 85.5073 | 18.5 | 0.21 |
| polarC18 | 468.308 | 60.1889 | 18.4 | 0.21 |
| polarC18 | 468.308 | 175.2532 | 18.4 | 0.21 |
| polarC18 | 468.308 | 67.2664 | 18.4 | 0.21 |
| polarC18 | 468.308 | 329.3841 | 18.4 | 0.21 |
| polarC18 | 468.308 | 54.785 | 18.4 | 0.21 |
| polarC18 | 468.308 | 296.5465 | 18.4 | 0.21 |
| polarC18 | 468.308 | 181.3098 | 18.4 | 0.21 |
| polarC18 | 468.308 | 57.5455 | 18.3 | 0.21 |
| polarC18 | 468.308 | 99.9369 | 18.3 | 0.21 |
| polarC18 | 468.308 | 272.96 | 18.3 | 0.21 |
| polarC18 | 468.308 | 406.4125 | 18.3 | 0.21 |
| polarC18 | 468.308 | 136.5764 | 18.2 | 0.21 |
| polarC18 | 468.308 | 136.5249 | 18.2 | 0.21 |

| | | | | |
|---|---|---|---|---|
| polarC18 | 468.308 | 90.9822 | 18.2 | 0.21 |
| polarC18 | 468.308 | 168.1176 | 18.2 | 0.2 |
| polarC18 | 468.308 | 206.269 | 18.2 | 0.2 |
| polarC18 | 468.308 | 89.349 | 18.2 | 0.2 |
| polarC18 | 468.308 | 84.4824 | 18.2 | 0.2 |
| polarC18 | 468.308 | 55.1803 | 18.2 | 0.2 |
| polarC18 | 468.308 | 460.3854 | 18.1 | 0.2 |
| polarC18 | 468.308 | 164.5413 | 18 | 0.2 |
| polarC18 | 468.308 | 337.4156 | 18 | 0.2 |
| polarC18 | 468.308 | 92.2062 | 18 | 0.2 |
| polarC18 | 468.308 | 58.9136 | 18 | 0.2 |
| polarC18 | 468.308 | 83.8464 | 18 | 0.2 |
| polarC18 | 468.308 | 64.4258 | 18 | 0.2 |
| polarC18 | 468.308 | 191.6125 | 18 | 0.2 |
| polarC18 | 468.308 | 77.5894 | 18 | 0.2 |
| polarC18 | 468.308 | 274.0865 | 18 | 0.2 |
| polarC18 | 468.308 | 53.2515 | 18 | 0.2 |
| polarC18 | 468.308 | 108.2007 | 17.9 | 0.2 |
| polarC18 | 468.308 | 117.1399 | 17.9 | 0.2 |
| polarC18 | 468.308 | 55.6469 | 17.9 | 0.2 |
| polarC18 | 468.308 | 113.1452 | 17.9 | 0.2 |
| polarC18 | 468.308 | 66.383 | 17.9 | 0.2 |
| polarC18 | 468.308 | 134.905 | 17.8 | 0.2 |
| polarC18 | 468.308 | 166.1353 | 17.8 | 0.2 |
| polarC18 | 468.308 | 216.8734 | 17.8 | 0.2 |
| polarC18 | 468.308 | 57.94 | 17.8 | 0.2 |
| polarC18 | 468.308 | 57.6777 | 17.8 | 0.2 |
| polarC18 | 468.308 | 84.1374 | 17.8 | 0.2 |
| polarC18 | 468.308 | 52.4529 | 17.8 | 0.2 |
| polarC18 | 468.308 | 471.9194 | 17.7 | 0.2 |
| polarC18 | 468.308 | 196.37 | 17.7 | 0.2 |
| polarC18 | 468.308 | 136.1504 | 17.7 | 0.2 |
| polarC18 | 468.308 | 230.1451 | 17.7 | 0.2 |
| polarC18 | 468.308 | 237.34 | 17.7 | 0.2 |
| polarC18 | 468.308 | 92.6004 | 17.7 | 0.2 |
| polarC18 | 468.308 | 83.6289 | 17.7 | 0.2 |
| polarC18 | 468.308 | 56.1519 | 17.6 | 0.2 |
| polarC18 | 468.308 | 153.6239 | 17.6 | 0.2 |
| polarC18 | 468.308 | 214.8917 | 17.6 | 0.2 |
| polarC18 | 468.308 | 102.488 | 17.6 | 0.2 |
| polarC18 | 468.308 | 57.5692 | 17.6 | 0.2 |
| polarC18 | 468.308 | 60.124 | 17.6 | 0.2 |

| | | | | |
|---|---|---|---|---|
| polarC18 | 468.308 | 224.5604 | 17.6 | 0.2 |
| polarC18 | 468.308 | 191.3603 | 17.5 | 0.2 |
| polarC18 | 468.308 | 111.0639 | 17.5 | 0.2 |
| polarC18 | 468.308 | 442.4814 | 17.5 | 0.2 |
| polarC18 | 468.308 | 70.9318 | 17.5 | 0.2 |
| polarC18 | 468.308 | 260.6562 | 17.4 | 0.2 |
| polarC18 | 468.308 | 137.1243 | 17.4 | 0.2 |
| polarC18 | 468.308 | 177.8236 | 17.4 | 0.2 |
| polarC18 | 468.308 | 121.5774 | 17.4 | 0.2 |
| polarC18 | 468.308 | 118.1558 | 17.3 | 0.19 |
| polarC18 | 468.308 | 128.4556 | 17.3 | 0.19 |
| polarC18 | 468.308 | 118.0563 | 17.3 | 0.19 |
| polarC18 | 468.308 | 116.6111 | 17.3 | 0.19 |
| polarC18 | 468.308 | 352.0415 | 17.3 | 0.19 |
| polarC18 | 468.308 | 172.6255 | 17.3 | 0.19 |
| polarC18 | 468.308 | 100.2366 | 17.2 | 0.19 |
| polarC18 | 468.308 | 152.9232 | 17.2 | 0.19 |
| polarC18 | 468.308 | 78.9535 | 17.2 | 0.19 |
| polarC18 | 468.308 | 92.746 | 17.2 | 0.19 |
| polarC18 | 468.308 | 60.7901 | 17.2 | 0.19 |
| polarC18 | 468.308 | 299.28 | 17.2 | 0.19 |
| polarC18 | 468.308 | 146.6537 | 17.2 | 0.19 |
| polarC18 | 468.308 | 293.1859 | 17.1 | 0.19 |
| polarC18 | 468.308 | 257.1944 | 17.1 | 0.19 |
| polarC18 | 468.308 | 450.4451 | 17.1 | 0.19 |
| polarC18 | 468.308 | 61.7752 | 17.1 | 0.19 |
| polarC18 | 468.308 | 54.4938 | 17 | 0.19 |
| polarC18 | 468.308 | 86.3584 | 17 | 0.19 |
| polarC18 | 468.308 | 244.9299 | 17 | 0.19 |
| polarC18 | 468.308 | 61.2326 | 16.9 | 0.19 |
| polarC18 | 468.308 | 156.2711 | 16.9 | 0.19 |
| polarC18 | 468.308 | 80.3069 | 16.9 | 0.19 |
| polarC18 | 468.308 | 142.6242 | 16.9 | 0.19 |
| polarC18 | 468.308 | 52.8763 | 16.9 | 0.19 |
| polarC18 | 468.308 | 55.5593 | 16.9 | 0.19 |
| polarC18 | 468.308 | 93.2599 | 16.9 | 0.19 |
| polarC18 | 468.308 | 137.1931 | 16.8 | 0.19 |
| polarC18 | 468.308 | 164.425 | 16.8 | 0.19 |
| polarC18 | 468.308 | 69.5905 | 16.8 | 0.19 |
| polarC18 | 468.308 | 147.1767 | 16.8 | 0.19 |
| polarC18 | 468.308 | 191.0051 | 16.8 | 0.19 |
| polarC18 | 468.308 | 56.1855 | 16.8 | 0.19 |

| | | | | |
|---|---|---|---|---|
| polarC18 | 468.308 | 81.403 | 16.7 | 0.19 |
| polarC18 | 468.308 | 52.5418 | 16.7 | 0.19 |
| polarC18 | 468.308 | 99.5618 | 16.7 | 0.19 |
| polarC18 | 468.308 | 262.5915 | 16.7 | 0.19 |
| polarC18 | 468.308 | 110.691 | 16.7 | 0.19 |
| polarC18 | 468.308 | 263.6401 | 16.6 | 0.19 |
| polarC18 | 468.308 | 80.124 | 16.6 | 0.19 |
| polarC18 | 468.308 | 76.8286 | 16.6 | 0.19 |
| polarC18 | 468.308 | 55.2402 | 16.6 | 0.19 |
| polarC18 | 468.308 | 192.5557 | 16.6 | 0.19 |
| polarC18 | 468.308 | 55.7433 | 16.5 | 0.19 |
| polarC18 | 468.308 | 108.7853 | 16.5 | 0.19 |
| polarC18 | 468.308 | 66.2924 | 16.5 | 0.19 |
| polarC18 | 468.308 | 56.0815 | 16.5 | 0.19 |
| polarC18 | 468.308 | 93.8117 | 16.5 | 0.19 |
| polarC18 | 468.308 | 369.0709 | 16.5 | 0.19 |
| polarC18 | 468.308 | 55.1421 | 16.5 | 0.19 |
| polarC18 | 468.308 | 92.5557 | 16.5 | 0.19 |
| polarC18 | 468.308 | 110.8067 | 16.4 | 0.18 |
| polarC18 | 468.308 | 79.9937 | 16.4 | 0.18 |
| polarC18 | 468.308 | 102.653 | 16.4 | 0.18 |
| polarC18 | 468.308 | 245.9357 | 16.4 | 0.18 |
| polarC18 | 468.308 | 82.9466 | 16.4 | 0.18 |
| polarC18 | 468.308 | 64.9051 | 16.3 | 0.18 |
| polarC18 | 468.308 | 153.8175 | 16.3 | 0.18 |
| polarC18 | 468.308 | 294.8492 | 16.3 | 0.18 |
| polarC18 | 468.308 | 358.3325 | 16.3 | 0.18 |
| polarC18 | 468.308 | 71.9494 | 16.3 | 0.18 |
| polarC18 | 468.308 | 56.9926 | 16.3 | 0.18 |
| polarC18 | 468.308 | 126.4812 | 16.2 | 0.18 |
| polarC18 | 468.308 | 142.0694 | 16.2 | 0.18 |
| polarC18 | 468.308 | 272.5008 | 16.2 | 0.18 |
| polarC18 | 468.308 | 154.4723 | 16.2 | 0.18 |
| polarC18 | 468.308 | 175.5876 | 16.2 | 0.18 |
| polarC18 | 468.308 | 55.072 | 16.2 | 0.18 |
| polarC18 | 468.308 | 331.2056 | 16.2 | 0.18 |
| polarC18 | 468.308 | 140.6272 | 16.1 | 0.18 |
| polarC18 | 468.308 | 455.173 | 16.1 | 0.18 |
| polarC18 | 468.308 | 293.3057 | 16.1 | 0.18 |
| polarC18 | 468.308 | 300.7784 | 16.1 | 0.18 |
| polarC18 | 468.308 | 229.0404 | 16.1 | 0.18 |
| polarC18 | 468.308 | 138.7718 | 16 | 0.18 |

| | | | | |
|---|---|---|---|---|
| polarC18 | 468.308 | 91.0617 | 16 | 0.18 |
| polarC18 | 468.308 | 109.3007 | 16 | 0.18 |
| polarC18 | 468.308 | 242.7858 | 15.9 | 0.18 |
| polarC18 | 468.308 | 112.9348 | 15.9 | 0.18 |
| polarC18 | 468.308 | 131.1616 | 15.9 | 0.18 |
| polarC18 | 468.308 | 54.9489 | 15.9 | 0.18 |
| polarC18 | 468.308 | 260.3536 | 15.9 | 0.18 |
| polarC18 | 468.308 | 126.3466 | 15.8 | 0.18 |
| polarC18 | 468.308 | 227.4586 | 15.8 | 0.18 |
| polarC18 | 468.308 | 52.1276 | 15.8 | 0.18 |
| polarC18 | 468.308 | 145.2967 | 15.8 | 0.18 |
| polarC18 | 468.308 | 128.2462 | 15.7 | 0.18 |
| polarC18 | 468.308 | 309.1707 | 15.7 | 0.18 |
| polarC18 | 468.308 | 85.1192 | 15.6 | 0.18 |
| polarC18 | 468.308 | 51.7029 | 15.5 | 0.17 |
| polarC18 | 468.308 | 118.516 | 15.4 | 0.17 |
| polarC18 | 468.308 | 74.1075 | 15.4 | 0.17 |
| polarC18 | 468.308 | 90.21 | 15.3 | 0.17 |
| polarC18 | 468.308 | 101.6733 | 15.2 | 0.17 |
| polarC18 | 468.308 | 63.2013 | 15.2 | 0.17 |
| polarC18 | 468.308 | 174.1555 | 15.2 | 0.17 |
| polarC18 | 468.308 | 109.5931 | 15.1 | 0.17 |
| polarC18 | 468.308 | 200.4291 | 15 | 0.17 |
| polarC18 | 468.308 | 50.89 | 14.9 | 0.17 |
| polarC18 | 468.308 | 282.6079 | 14.9 | 0.17 |
| polarC18 | 468.308 | 428.2324 | 14.9 | 0.17 |
| polarC18 | 468.308 | 131.1289 | 14.6 | 0.16 |
| polarC18 | 468.308 | 77.44 | 14.6 | 0.16 |
| polarC18 | 468.308 | 65.2335 | 14.3 | 0.16 |
| polarC18 | 468.308 | 54.6155 | 13.4 | 0.15 |
| polarC18 | 758.568 | 228.2196 | 9316.7 | 100 |
| polarC18 | 758.568 | 184.0733 | 4171.5 | 44.77 |
| polarC18 | 758.568 | 80.9476 | 325.6 | 3.49 |
| polarC18 | 758.568 | 283.2633 | 307 | 3.3 |
| polarC18 | 758.568 | 758.5828 | 106 | 1.14 |
| polarC18 | 758.568 | 86.0962 | 102.8 | 1.1 |
| polarC18 | 758.568 | 685.9013 | 70.8 | 0.76 |
| polarC18 | 758.568 | 546.6157 | 68.7 | 0.74 |
| polarC18 | 758.568 | 749.1876 | 68.2 | 0.73 |
| polarC18 | 758.568 | 603.0737 | 66.3 | 0.71 |
| polarC18 | 758.568 | 523.6202 | 64.9 | 0.7 |
| polarC18 | 758.568 | 93.9383 | 64.8 | 0.7 |

| | | | | |
|---|---|---|---|---|
| polarC18 | 758.568 | 227.9026 | 63.8 | 0.68 |
| polarC18 | 758.568 | 94.7056 | 62.5 | 0.67 |
| polarC18 | 758.568 | 658.2052 | 58.5 | 0.63 |
| polarC18 | 758.568 | 338.9183 | 57.5 | 0.62 |
| polarC18 | 758.568 | 571.056 | 57.4 | 0.62 |
| polarC18 | 758.568 | 506.791 | 55.2 | 0.59 |
| polarC18 | 758.568 | 431.8391 | 54.5 | 0.58 |
| polarC18 | 758.568 | 707.8763 | 54.5 | 0.58 |
| polarC18 | 758.568 | 274.667 | 54.3 | 0.58 |
| polarC18 | 758.568 | 95.0604 | 54 | 0.58 |
| polarC18 | 758.568 | 245.86 | 53.8 | 0.58 |
| polarC18 | 758.568 | 572.4643 | 53.4 | 0.57 |
| polarC18 | 758.568 | 93.4144 | 52.4 | 0.56 |
| polarC18 | 758.568 | 380.5815 | 52 | 0.56 |
| polarC18 | 758.568 | 93.3703 | 51.9 | 0.56 |
| polarC18 | 758.568 | 780.1095 | 51.5 | 0.55 |
| polarC18 | 758.568 | 592.5334 | 51.3 | 0.55 |
| polarC18 | 758.568 | 436.6915 | 50.9 | 0.55 |
| polarC18 | 758.568 | 247.6387 | 50.9 | 0.55 |
| polarC18 | 758.568 | 785.5495 | 50.9 | 0.55 |
| polarC18 | 758.568 | 519.7933 | 50.6 | 0.54 |
| polarC18 | 758.568 | 311.679 | 50.6 | 0.54 |
| polarC18 | 758.568 | 359.7979 | 50.5 | 0.54 |
| polarC18 | 758.568 | 582.477 | 50 | 0.54 |
| polarC18 | 758.568 | 198.19 | 50 | 0.54 |
| polarC18 | 758.568 | 490.8261 | 49.9 | 0.54 |
| polarC18 | 758.568 | 158.6494 | 49.9 | 0.54 |
| polarC18 | 758.568 | 668.8039 | 49.6 | 0.53 |
| polarC18 | 758.568 | 305.3617 | 49.6 | 0.53 |
| polarC18 | 758.568 | 548.0305 | 49.2 | 0.53 |
| polarC18 | 758.568 | 772.8191 | 48.8 | 0.52 |
| polarC18 | 758.568 | 722.1379 | 48.7 | 0.52 |
| polarC18 | 758.568 | 671.1683 | 48.7 | 0.52 |
| polarC18 | 758.568 | 158.256 | 48.5 | 0.52 |
| polarC18 | 758.568 | 229.2558 | 48.4 | 0.52 |
| polarC18 | 758.568 | 380.6846 | 48.4 | 0.52 |
| polarC18 | 758.568 | 324.5238 | 48.3 | 0.52 |
| polarC18 | 758.568 | 447.4049 | 48.1 | 0.52 |
| polarC18 | 758.568 | 393.7745 | 48.1 | 0.52 |
| polarC18 | 758.568 | 463.8046 | 48 | 0.52 |
| polarC18 | 758.568 | 339.1254 | 48 | 0.51 |
| polarC18 | 758.568 | 398.8716 | 48 | 0.51 |

| | | | | |
|---|---|---|---|---|
| polarC18 | 758.568 | 407.5179 | 47.9 | 0.51 |
| polarC18 | 758.568 | 87.5337 | 47.8 | 0.51 |
| polarC18 | 758.568 | 554.3678 | 47.8 | 0.51 |
| polarC18 | 758.568 | 214.8357 | 47.7 | 0.51 |
| polarC18 | 758.568 | 452.3963 | 47.6 | 0.51 |
| polarC18 | 758.568 | 100.7908 | 47.4 | 0.51 |
| polarC18 | 758.568 | 93.4522 | 47.4 | 0.51 |
| polarC18 | 758.568 | 641.9314 | 47.4 | 0.51 |
| polarC18 | 758.568 | 375.8646 | 47.3 | 0.51 |
| polarC18 | 758.568 | 143.3616 | 47.3 | 0.51 |
| polarC18 | 758.568 | 308.7648 | 47.2 | 0.51 |
| polarC18 | 758.568 | 570.314 | 46.9 | 0.5 |
| polarC18 | 758.568 | 319.0327 | 46.6 | 0.5 |
| polarC18 | 758.568 | 131.4076 | 46.6 | 0.5 |
| polarC18 | 758.568 | 93.9247 | 46.5 | 0.5 |
| polarC18 | 758.568 | 93.386 | 46.3 | 0.5 |
| polarC18 | 758.568 | 193.6683 | 46.3 | 0.5 |
| polarC18 | 758.568 | 528.6418 | 46.2 | 0.5 |
| polarC18 | 758.568 | 269.4034 | 46.1 | 0.49 |
| polarC18 | 758.568 | 125.8838 | 46 | 0.49 |
| polarC18 | 758.568 | 85.5656 | 46 | 0.49 |
| polarC18 | 758.568 | 269.8897 | 46 | 0.49 |
| polarC18 | 758.568 | 413.7778 | 45.6 | 0.49 |
| polarC18 | 758.568 | 653.1388 | 45.4 | 0.49 |
| polarC18 | 758.568 | 478.74 | 45.3 | 0.49 |
| polarC18 | 758.568 | 239.5223 | 45.2 | 0.49 |
| polarC18 | 758.568 | 189.5261 | 45.1 | 0.48 |
| polarC18 | 758.568 | 117.9866 | 44.9 | 0.48 |
| polarC18 | 758.568 | 654.3525 | 44.8 | 0.48 |
| polarC18 | 758.568 | 597.7595 | 44.6 | 0.48 |
| polarC18 | 758.568 | 547.588 | 44.4 | 0.48 |
| polarC18 | 758.568 | 314.6454 | 44.2 | 0.47 |
| polarC18 | 758.568 | 227.5494 | 44.1 | 0.47 |
| polarC18 | 758.568 | 262.5202 | 43.5 | 0.47 |
| polarC18 | 758.568 | 119.2767 | 43.5 | 0.47 |
| polarC18 | 758.568 | 261.9866 | 43.3 | 0.47 |
| polarC18 | 758.568 | 267.5997 | 43.3 | 0.46 |
| polarC18 | 758.568 | 420.6748 | 43.1 | 0.46 |
| polarC18 | 758.568 | 211.0142 | 42.8 | 0.46 |
| polarC18 | 758.568 | 152.3277 | 42.7 | 0.46 |
| polarC18 | 758.568 | 93.4585 | 42.6 | 0.46 |
| polarC18 | 758.568 | 525.6935 | 42.5 | 0.46 |

| | | | | |
|---|---|---|---|---|
| polarC18 | 758.568 | 616.334 | 42.4 | 0.46 |
| polarC18 | 758.568 | 387.5555 | 42.2 | 0.45 |
| polarC18 | 758.568 | 258.0292 | 41.9 | 0.45 |
| polarC18 | 758.568 | 199.4868 | 41.3 | 0.44 |
| polarC18 | 758.568 | 287.2945 | 41.2 | 0.44 |
| polarC18 | 758.568 | 189.6294 | 41.1 | 0.44 |
| polarC18 | 758.568 | 205.9785 | 41 | 0.44 |
| polarC18 | 758.568 | 748.4072 | 40.9 | 0.44 |
| polarC18 | 758.568 | 344.9695 | 39.7 | 0.43 |
| polarC18 | 758.568 | 125.5066 | 39.7 | 0.43 |
| polarC18 | 758.568 | 506.1869 | 39.4 | 0.42 |
| polarC18 | 758.568 | 494.4079 | 39.3 | 0.42 |
| polarC18 | 758.568 | 775.9101 | 39.3 | 0.42 |
| polarC18 | 758.568 | 168.8479 | 38.9 | 0.42 |
| polarC18 | 758.568 | 93.7011 | 38.8 | 0.42 |
| polarC18 | 758.568 | 302.128 | 38.7 | 0.42 |
| polarC18 | 758.568 | 551.6885 | 38.4 | 0.41 |
| polarC18 | 758.568 | 662.208 | 38.4 | 0.41 |
| polarC18 | 758.568 | 742.5981 | 38.2 | 0.41 |
| polarC18 | 758.568 | 134.794 | 38 | 0.41 |
| polarC18 | 758.568 | 692.4539 | 37.5 | 0.4 |
| polarC18 | 758.568 | 663.2755 | 37.5 | 0.4 |
| polarC18 | 758.568 | 686.7601 | 37.5 | 0.4 |
| polarC18 | 758.568 | 522.1764 | 37.4 | 0.4 |
| polarC18 | 758.568 | 478.0647 | 37.4 | 0.4 |
| polarC18 | 758.568 | 222.749 | 37.1 | 0.4 |
| polarC18 | 758.568 | 442.2067 | 37 | 0.4 |
| polarC18 | 758.568 | 98.572 | 37 | 0.4 |
| polarC18 | 758.568 | 332.952 | 36.7 | 0.39 |
| polarC18 | 758.568 | 690.5519 | 36.6 | 0.39 |
| polarC18 | 758.568 | 219.4436 | 36.6 | 0.39 |
| polarC18 | 758.568 | 128.0896 | 36.4 | 0.39 |
| polarC18 | 758.568 | 462.7189 | 36.4 | 0.39 |
| polarC18 | 758.568 | 593.8166 | 36.2 | 0.39 |
| polarC18 | 758.568 | 111.014 | 36.1 | 0.39 |
| polarC18 | 758.568 | 723.7404 | 36.1 | 0.39 |
| polarC18 | 758.568 | 93.4277 | 36.1 | 0.39 |
| polarC18 | 758.568 | 581.7805 | 36.1 | 0.39 |
| polarC18 | 758.568 | 640.0057 | 36.1 | 0.39 |
| polarC18 | 758.568 | 563.8545 | 36 | 0.39 |
| polarC18 | 758.568 | 727.9621 | 36 | 0.39 |
| polarC18 | 758.568 | 732.1804 | 35.8 | 0.38 |

| | | | | |
|---|---|---|---|---|
| polarC18 | 758.568 | 443.7851 | 35.8 | 0.38 |
| polarC18 | 758.568 | 130.1371 | 35.8 | 0.38 |
| polarC18 | 758.568 | 138.4702 | 35.8 | 0.38 |
| polarC18 | 758.568 | 369.1139 | 35.7 | 0.38 |
| polarC18 | 758.568 | 229.3053 | 35.7 | 0.38 |
| polarC18 | 758.568 | 455.1856 | 35.7 | 0.38 |
| polarC18 | 758.568 | 290.3991 | 35.5 | 0.38 |
| polarC18 | 758.568 | 469.1597 | 35.5 | 0.38 |
| polarC18 | 758.568 | 287.5642 | 35.5 | 0.38 |
| polarC18 | 758.568 | 122.4688 | 35.4 | 0.38 |
| polarC18 | 758.568 | 90.5538 | 35.4 | 0.38 |
| polarC18 | 758.568 | 121.9551 | 35.4 | 0.38 |
| polarC18 | 758.568 | 146.4522 | 35.3 | 0.38 |
| polarC18 | 758.568 | 762.4174 | 35.3 | 0.38 |
| polarC18 | 758.568 | 234.8077 | 35.3 | 0.38 |
| polarC18 | 758.568 | 104.4192 | 35.3 | 0.38 |
| polarC18 | 758.568 | 93.9446 | 35.2 | 0.38 |
| polarC18 | 758.568 | 154.5259 | 35.2 | 0.38 |
| polarC18 | 758.568 | 466.9383 | 35.1 | 0.38 |
| polarC18 | 758.568 | 138.0922 | 35.1 | 0.38 |
| polarC18 | 758.568 | 512.6006 | 35.1 | 0.38 |
| polarC18 | 758.568 | 377.5099 | 35.1 | 0.38 |
| polarC18 | 758.568 | 280.9907 | 35 | 0.38 |
| polarC18 | 758.568 | 334.5865 | 35 | 0.38 |
| polarC18 | 758.568 | 449.4788 | 35 | 0.38 |
| polarC18 | 758.568 | 84.7894 | 35 | 0.38 |
| polarC18 | 758.568 | 271.5934 | 35 | 0.38 |
| polarC18 | 758.568 | 653.9694 | 35 | 0.38 |
| polarC18 | 758.568 | 228.9636 | 35 | 0.38 |
| polarC18 | 758.568 | 746.3232 | 35 | 0.38 |
| polarC18 | 758.568 | 317.9961 | 35 | 0.38 |
| polarC18 | 758.568 | 417.145 | 34.9 | 0.37 |
| polarC18 | 758.568 | 159.2579 | 34.9 | 0.37 |
| polarC18 | 758.568 | 103.4872 | 34.9 | 0.37 |
| polarC18 | 758.568 | 186.2218 | 34.9 | 0.37 |
| polarC18 | 758.568 | 139.8485 | 34.8 | 0.37 |
| polarC18 | 758.568 | 93.3526 | 34.8 | 0.37 |
| polarC18 | 758.568 | 99.7526 | 34.8 | 0.37 |
| polarC18 | 758.568 | 168.5527 | 34.8 | 0.37 |
| polarC18 | 758.568 | 367.0843 | 34.8 | 0.37 |
| polarC18 | 758.568 | 532.8755 | 34.7 | 0.37 |
| polarC18 | 758.568 | 627.9096 | 34.7 | 0.37 |

| | | | | |
|---|---|---|---|---|
| polarC18 | 758.568 | 790.5912 | 34.7 | 0.37 |
| polarC18 | 758.568 | 442.8712 | 34.6 | 0.37 |
| polarC18 | 758.568 | 98.2701 | 34.6 | 0.37 |
| polarC18 | 758.568 | 95.0058 | 34.6 | 0.37 |
| polarC18 | 758.568 | 153.1723 | 34.6 | 0.37 |
| polarC18 | 758.568 | 445.5588 | 34.6 | 0.37 |
| polarC18 | 758.568 | 97.5781 | 34.6 | 0.37 |
| polarC18 | 758.568 | 300.656 | 34.6 | 0.37 |
| polarC18 | 758.568 | 407.1206 | 34.5 | 0.37 |
| polarC18 | 758.568 | 738.1826 | 34.5 | 0.37 |
| polarC18 | 758.568 | 129.9919 | 34.5 | 0.37 |
| polarC18 | 758.568 | 195.4475 | 34.5 | 0.37 |
| polarC18 | 758.568 | 358.8507 | 34.5 | 0.37 |
| polarC18 | 758.568 | 430.5591 | 34.5 | 0.37 |
| polarC18 | 758.568 | 83.8863 | 34.4 | 0.37 |
| polarC18 | 758.568 | 639.6126 | 34.4 | 0.37 |
| polarC18 | 758.568 | 548.4518 | 34.3 | 0.37 |
| polarC18 | 758.568 | 358.3887 | 34.3 | 0.37 |
| polarC18 | 758.568 | 396.7701 | 34.3 | 0.37 |
| polarC18 | 758.568 | 404.3492 | 34.3 | 0.37 |
| polarC18 | 758.568 | 553.7496 | 34.2 | 0.37 |
| polarC18 | 758.568 | 517.4261 | 34.2 | 0.37 |
| polarC18 | 758.568 | 207.4381 | 34.2 | 0.37 |
| polarC18 | 758.568 | 764.5138 | 34.2 | 0.37 |
| polarC18 | 758.568 | 724.4214 | 34.1 | 0.37 |
| polarC18 | 758.568 | 491.9996 | 34.1 | 0.37 |
| polarC18 | 758.568 | 399.1041 | 34.1 | 0.37 |
| polarC18 | 758.568 | 139.1883 | 34.1 | 0.37 |
| polarC18 | 758.568 | 136.348 | 34.1 | 0.37 |
| polarC18 | 758.568 | 379.997 | 34.1 | 0.37 |
| polarC18 | 758.568 | 712.9356 | 34.1 | 0.37 |
| polarC18 | 758.568 | 739.9495 | 34.1 | 0.37 |
| polarC18 | 758.568 | 539.651 | 34.1 | 0.37 |
| polarC18 | 758.568 | 623.5792 | 34.1 | 0.37 |
| polarC18 | 758.568 | 726.9003 | 34.1 | 0.37 |
| polarC18 | 758.568 | 617.1781 | 34 | 0.37 |
| polarC18 | 758.568 | 93.3785 | 34 | 0.36 |
| polarC18 | 758.568 | 772.361 | 34 | 0.36 |
| polarC18 | 758.568 | 114.9334 | 34 | 0.36 |
| polarC18 | 758.568 | 165.2001 | 33.9 | 0.36 |
| polarC18 | 758.568 | 95.0008 | 33.9 | 0.36 |
| polarC18 | 758.568 | 407.937 | 33.9 | 0.36 |

| | | | | |
|---|---|---|---|---|
| polarC18 | 758.568 | 325.7238 | 33.9 | 0.36 |
| polarC18 | 758.568 | 657.6373 | 33.9 | 0.36 |
| polarC18 | 758.568 | 163.4093 | 33.9 | 0.36 |
| polarC18 | 758.568 | 268.7472 | 33.9 | 0.36 |
| polarC18 | 758.568 | 110.2097 | 33.9 | 0.36 |
| polarC18 | 758.568 | 81.0826 | 33.8 | 0.36 |
| polarC18 | 758.568 | 341.8485 | 33.7 | 0.36 |
| polarC18 | 758.568 | 189.2466 | 33.6 | 0.36 |
| polarC18 | 758.568 | 110.2471 | 33.6 | 0.36 |
| polarC18 | 758.568 | 282.6335 | 33.6 | 0.36 |
| polarC18 | 758.568 | 79.7164 | 33.6 | 0.36 |
| polarC18 | 758.568 | 504.3513 | 33.6 | 0.36 |
| polarC18 | 758.568 | 384.8176 | 33.5 | 0.36 |
| polarC18 | 758.568 | 248.7251 | 33.5 | 0.36 |
| polarC18 | 758.568 | 334.9328 | 33.5 | 0.36 |
| polarC18 | 758.568 | 397.0063 | 33.5 | 0.36 |
| polarC18 | 758.568 | 188.3444 | 33.5 | 0.36 |
| polarC18 | 758.568 | 159.3675 | 33.5 | 0.36 |
| polarC18 | 758.568 | 294.0754 | 33.5 | 0.36 |
| polarC18 | 758.568 | 604.1656 | 33.5 | 0.36 |
| polarC18 | 758.568 | 260.7783 | 33.5 | 0.36 |
| polarC18 | 758.568 | 100.9588 | 33.5 | 0.36 |
| polarC18 | 758.568 | 763.5898 | 33.5 | 0.36 |
| polarC18 | 758.568 | 628.7153 | 33.4 | 0.36 |
| polarC18 | 758.568 | 777.9099 | 33.4 | 0.36 |
| polarC18 | 758.568 | 383.163 | 33.4 | 0.36 |
| polarC18 | 758.568 | 192.4688 | 33.4 | 0.36 |
| polarC18 | 758.568 | 117.2029 | 33.4 | 0.36 |
| polarC18 | 758.568 | 615.4261 | 33.4 | 0.36 |
| polarC18 | 758.568 | 703.5682 | 33.4 | 0.36 |
| polarC18 | 758.568 | 537.6917 | 33.4 | 0.36 |
| polarC18 | 758.568 | 740.7366 | 33.3 | 0.36 |
| polarC18 | 758.568 | 514.2135 | 33.3 | 0.36 |
| polarC18 | 758.568 | 602.04 | 33.3 | 0.36 |
| polarC18 | 758.568 | 394.302 | 33.3 | 0.36 |
| polarC18 | 758.568 | 578.9018 | 33.3 | 0.36 |
| polarC18 | 758.568 | 109.8561 | 33.2 | 0.36 |
| polarC18 | 758.568 | 309.9158 | 33.2 | 0.36 |
| polarC18 | 758.568 | 94.0327 | 33.2 | 0.36 |
| polarC18 | 758.568 | 180.3673 | 33.2 | 0.36 |
| polarC18 | 758.568 | 707.5275 | 33.1 | 0.36 |
| polarC18 | 758.568 | 100.5134 | 33.1 | 0.36 |

| | | | | |
|---|---|---|---|---|
| polarC18 | 758.568 | 403.8265 | 33.1 | 0.36 |
| polarC18 | 758.568 | 145.9906 | 33.1 | 0.36 |
| polarC18 | 758.568 | 401.8022 | 33.1 | 0.35 |
| polarC18 | 758.568 | 699.8247 | 33.1 | 0.35 |
| polarC18 | 758.568 | 265.6851 | 33 | 0.35 |
| polarC18 | 758.568 | 263.9776 | 33 | 0.35 |
| polarC18 | 758.568 | 562.8438 | 33 | 0.35 |
| polarC18 | 758.568 | 608.5333 | 33 | 0.35 |
| polarC18 | 758.568 | 121.7928 | 33 | 0.35 |
| polarC18 | 758.568 | 316.5715 | 33 | 0.35 |
| polarC18 | 758.568 | 151.626 | 33 | 0.35 |
| polarC18 | 758.568 | 97.5677 | 32.9 | 0.35 |
| polarC18 | 758.568 | 100.9761 | 32.9 | 0.35 |
| polarC18 | 758.568 | 594.2816 | 32.9 | 0.35 |
| polarC18 | 758.568 | 569.5732 | 32.9 | 0.35 |
| polarC18 | 758.568 | 270.4519 | 32.9 | 0.35 |
| polarC18 | 758.568 | 90.3988 | 32.9 | 0.35 |
| polarC18 | 758.568 | 85.6984 | 32.9 | 0.35 |
| polarC18 | 758.568 | 613.331 | 32.9 | 0.35 |
| polarC18 | 758.568 | 780.6965 | 32.9 | 0.35 |
| polarC18 | 758.568 | 125.7994 | 32.9 | 0.35 |
| polarC18 | 758.568 | 196.5059 | 32.9 | 0.35 |
| polarC18 | 758.568 | 93.7552 | 32.9 | 0.35 |
| polarC18 | 758.568 | 210.5599 | 32.8 | 0.35 |
| polarC18 | 758.568 | 294.9973 | 32.8 | 0.35 |
| polarC18 | 758.568 | 494.0072 | 32.8 | 0.35 |
| polarC18 | 758.568 | 180.4654 | 32.8 | 0.35 |
| polarC18 | 758.568 | 204.3889 | 32.8 | 0.35 |
| polarC18 | 758.568 | 94.3822 | 32.8 | 0.35 |
| polarC18 | 758.568 | 277.4173 | 32.8 | 0.35 |
| polarC18 | 758.568 | 250.2504 | 32.8 | 0.35 |
| polarC18 | 758.568 | 413.609 | 32.8 | 0.35 |
| polarC18 | 758.568 | 394.5324 | 32.8 | 0.35 |
| polarC18 | 758.568 | 223.7453 | 32.8 | 0.35 |
| polarC18 | 758.568 | 779.7459 | 32.8 | 0.35 |
| polarC18 | 758.568 | 792.5705 | 32.7 | 0.35 |
| polarC18 | 758.568 | 616.6008 | 32.7 | 0.35 |
| polarC18 | 758.568 | 191.9056 | 32.7 | 0.35 |
| polarC18 | 758.568 | 367.4212 | 32.7 | 0.35 |
| polarC18 | 758.568 | 671.9351 | 32.7 | 0.35 |
| polarC18 | 758.568 | 262.9411 | 32.7 | 0.35 |
| polarC18 | 758.568 | 668.5292 | 32.7 | 0.35 |

| | | | | |
|---|---|---|---|---|
| polarC18 | 758.568 | 223.5365 | 32.6 | 0.35 |
| polarC18 | 758.568 | 734.6414 | 32.6 | 0.35 |
| polarC18 | 758.568 | 262.0211 | 32.6 | 0.35 |
| polarC18 | 758.568 | 83.432 | 32.6 | 0.35 |
| polarC18 | 758.568 | 115.8952 | 32.6 | 0.35 |
| polarC18 | 758.568 | 456.7033 | 32.6 | 0.35 |
| polarC18 | 758.568 | 223.0664 | 32.6 | 0.35 |
| polarC18 | 758.568 | 644.6122 | 32.6 | 0.35 |
| polarC18 | 758.568 | 148.6638 | 32.6 | 0.35 |
| polarC18 | 758.568 | 290.6449 | 32.6 | 0.35 |
| polarC18 | 758.568 | 443.3531 | 32.6 | 0.35 |
| polarC18 | 758.568 | 421.7093 | 32.6 | 0.35 |
| polarC18 | 758.568 | 537.9645 | 32.6 | 0.35 |
| polarC18 | 758.568 | 133.2781 | 32.5 | 0.35 |
| polarC18 | 758.568 | 393.0114 | 32.5 | 0.35 |
| polarC18 | 758.568 | 81.3089 | 32.5 | 0.35 |
| polarC18 | 758.568 | 687.8834 | 32.5 | 0.35 |
| polarC18 | 758.568 | 233.3133 | 32.5 | 0.35 |
| polarC18 | 758.568 | 598.2519 | 32.5 | 0.35 |
| polarC18 | 758.568 | 98.3631 | 32.5 | 0.35 |
| polarC18 | 758.568 | 93.5901 | 32.5 | 0.35 |
| polarC18 | 758.568 | 110.6505 | 32.5 | 0.35 |
| polarC18 | 758.568 | 302.7272 | 32.5 | 0.35 |
| polarC18 | 758.568 | 369.8182 | 32.5 | 0.35 |
| polarC18 | 758.568 | 452.668 | 32.4 | 0.35 |
| polarC18 | 758.568 | 174.7338 | 32.4 | 0.35 |
| polarC18 | 758.568 | 414.3767 | 32.4 | 0.35 |
| polarC18 | 758.568 | 528.2806 | 32.4 | 0.35 |
| polarC18 | 758.568 | 93.3993 | 32.4 | 0.35 |
| polarC18 | 758.568 | 757.8485 | 32.4 | 0.35 |
| polarC18 | 758.568 | 86.7305 | 32.3 | 0.35 |
| polarC18 | 758.568 | 85.0751 | 32.3 | 0.35 |
| polarC18 | 758.568 | 537.0958 | 32.3 | 0.35 |
| polarC18 | 758.568 | 94.291 | 32.3 | 0.35 |
| polarC18 | 758.568 | 174.7891 | 32.3 | 0.35 |
| polarC18 | 758.568 | 181.6977 | 32.3 | 0.35 |
| polarC18 | 758.568 | 149.2804 | 32.3 | 0.35 |
| polarC18 | 758.568 | 781.2257 | 32.3 | 0.35 |
| polarC18 | 758.568 | 187.8287 | 32.2 | 0.35 |
| polarC18 | 758.568 | 174.2048 | 32.2 | 0.35 |
| polarC18 | 758.568 | 136.5728 | 32.2 | 0.35 |
| polarC18 | 758.568 | 648.1195 | 32.2 | 0.35 |

| | | | | |
|---|---|---|---|---|
| polarC18 | 758.568 | 792.1048 | 32.2 | 0.35 |
| polarC18 | 758.568 | 81.7516 | 32.2 | 0.35 |
| polarC18 | 758.568 | 229.4863 | 32.2 | 0.35 |
| polarC18 | 758.568 | 109.5571 | 32.2 | 0.35 |
| polarC18 | 758.568 | 114.1373 | 32.2 | 0.35 |
| polarC18 | 758.568 | 324.9828 | 32.2 | 0.35 |
| polarC18 | 758.568 | 148.1 | 32.2 | 0.35 |
| polarC18 | 758.568 | 355.9929 | 32.2 | 0.35 |
| polarC18 | 758.568 | 407.724 | 32.2 | 0.35 |
| polarC18 | 758.568 | 225.0606 | 32.1 | 0.35 |
| polarC18 | 758.568 | 348.8825 | 32.1 | 0.34 |
| polarC18 | 758.568 | 585.9821 | 32.1 | 0.34 |
| polarC18 | 758.568 | 659.4742 | 32.1 | 0.34 |
| polarC18 | 758.568 | 93.9645 | 32.1 | 0.34 |
| polarC18 | 758.568 | 226.8631 | 32.1 | 0.34 |
| polarC18 | 758.568 | 299.4558 | 32.1 | 0.34 |
| polarC18 | 758.568 | 312.454 | 32.1 | 0.34 |
| polarC18 | 758.568 | 720.8606 | 32.1 | 0.34 |
| polarC18 | 758.568 | 345.5343 | 32.1 | 0.34 |
| polarC18 | 758.568 | 97.2317 | 32 | 0.34 |
| polarC18 | 758.568 | 177.4849 | 32 | 0.34 |
| polarC18 | 758.568 | 290.0515 | 32 | 0.34 |
| polarC18 | 758.568 | 457.1325 | 32 | 0.34 |
| polarC18 | 758.568 | 328.5214 | 32 | 0.34 |
| polarC18 | 758.568 | 562.6259 | 32 | 0.34 |
| polarC18 | 758.568 | 145.7727 | 31.9 | 0.34 |
| polarC18 | 758.568 | 400.412 | 31.9 | 0.34 |
| polarC18 | 758.568 | 171.6142 | 31.9 | 0.34 |
| polarC18 | 758.568 | 156.6826 | 31.9 | 0.34 |
| polarC18 | 758.568 | 571.9882 | 31.9 | 0.34 |
| polarC18 | 758.568 | 547.207 | 31.9 | 0.34 |
| polarC18 | 758.568 | 155.0771 | 31.9 | 0.34 |
| polarC18 | 758.568 | 544.952 | 31.9 | 0.34 |
| polarC18 | 758.568 | 158.1699 | 31.9 | 0.34 |
| polarC18 | 758.568 | 116.7759 | 31.8 | 0.34 |
| polarC18 | 758.568 | 188.7096 | 31.8 | 0.34 |
| polarC18 | 758.568 | 102.1094 | 31.8 | 0.34 |
| polarC18 | 758.568 | 199.6775 | 31.8 | 0.34 |
| polarC18 | 758.568 | 255.9706 | 31.7 | 0.34 |
| polarC18 | 758.568 | 456.4896 | 31.7 | 0.34 |
| polarC18 | 758.568 | 196.4465 | 31.7 | 0.34 |
| polarC18 | 758.568 | 239.1548 | 31.7 | 0.34 |

| | | | | |
|---|---|---|---|---|
| polarC18 | 758.568 | 132.5963 | 31.7 | 0.34 |
| polarC18 | 758.568 | 748.1441 | 31.7 | 0.34 |
| polarC18 | 758.568 | 452.9364 | 31.7 | 0.34 |
| polarC18 | 758.568 | 131.3083 | 31.7 | 0.34 |
| polarC18 | 758.568 | 562.0761 | 31.7 | 0.34 |
| polarC18 | 758.568 | 292.8553 | 31.7 | 0.34 |
| polarC18 | 758.568 | 111.6591 | 31.7 | 0.34 |
| polarC18 | 758.568 | 673.1863 | 31.7 | 0.34 |
| polarC18 | 758.568 | 462.5187 | 31.6 | 0.34 |
| polarC18 | 758.568 | 558.635 | 31.6 | 0.34 |
| polarC18 | 758.568 | 380.1002 | 31.6 | 0.34 |
| polarC18 | 758.568 | 257.6799 | 31.6 | 0.34 |
| polarC18 | 758.568 | 584.0684 | 31.6 | 0.34 |
| polarC18 | 758.568 | 482.1384 | 31.6 | 0.34 |
| polarC18 | 758.568 | 280.1658 | 31.6 | 0.34 |
| polarC18 | 758.568 | 368.5799 | 31.6 | 0.34 |
| polarC18 | 758.568 | 364.2084 | 31.6 | 0.34 |
| polarC18 | 758.568 | 471.5998 | 31.6 | 0.34 |
| polarC18 | 758.568 | 137.2389 | 31.5 | 0.34 |
| polarC18 | 758.568 | 93.9192 | 31.5 | 0.34 |
| polarC18 | 758.568 | 109.9571 | 31.5 | 0.34 |
| polarC18 | 758.568 | 105.3999 | 31.5 | 0.34 |
| polarC18 | 758.568 | 169.5297 | 31.5 | 0.34 |
| polarC18 | 758.568 | 88.2819 | 31.5 | 0.34 |
| polarC18 | 758.568 | 501.149 | 31.5 | 0.34 |
| polarC18 | 758.568 | 307.3125 | 31.5 | 0.34 |
| polarC18 | 758.568 | 155.2538 | 31.5 | 0.34 |
| polarC18 | 758.568 | 108.3236 | 31.5 | 0.34 |
| polarC18 | 758.568 | 404.7584 | 31.5 | 0.34 |
| polarC18 | 758.568 | 109.6955 | 31.4 | 0.34 |
| polarC18 | 758.568 | 79.9236 | 31.4 | 0.34 |
| polarC18 | 758.568 | 99.8686 | 31.4 | 0.34 |
| polarC18 | 758.568 | 118.5213 | 31.4 | 0.34 |
| polarC18 | 758.568 | 163.4764 | 31.4 | 0.34 |
| polarC18 | 758.568 | 118.0148 | 31.4 | 0.34 |
| polarC18 | 758.568 | 263.2984 | 31.4 | 0.34 |
| polarC18 | 758.568 | 494.6165 | 31.4 | 0.34 |
| polarC18 | 758.568 | 104.5486 | 31.4 | 0.34 |
| polarC18 | 758.568 | 342.2711 | 31.4 | 0.34 |
| polarC18 | 758.568 | 339.3367 | 31.4 | 0.34 |
| polarC18 | 758.568 | 114.0823 | 31.3 | 0.34 |
| polarC18 | 758.568 | 341.7318 | 31.3 | 0.34 |

| | | | | |
|---|---|---|---|---|
| polarC18 | 758.568 | 423.9513 | 31.3 | 0.34 |
| polarC18 | 758.568 | 435.6414 | 31.3 | 0.34 |
| polarC18 | 758.568 | 692.9607 | 31.3 | 0.34 |
| polarC18 | 758.568 | 752.6704 | 31.3 | 0.34 |
| polarC18 | 758.568 | 646.2062 | 31.3 | 0.34 |
| polarC18 | 758.568 | 575.2594 | 31.3 | 0.34 |
| polarC18 | 758.568 | 93.2421 | 31.3 | 0.34 |
| polarC18 | 758.568 | 356.4217 | 31.3 | 0.34 |
| polarC18 | 758.568 | 95.4555 | 31.3 | 0.34 |
| polarC18 | 758.568 | 481.8675 | 31.3 | 0.34 |
| polarC18 | 758.568 | 186.9165 | 31.3 | 0.34 |
| polarC18 | 758.568 | 227.1876 | 31.3 | 0.34 |
| polarC18 | 758.568 | 310.9984 | 31.3 | 0.34 |
| polarC18 | 758.568 | 359.8533 | 31.3 | 0.34 |
| polarC18 | 758.568 | 91.4251 | 31.3 | 0.34 |
| polarC18 | 758.568 | 123.3489 | 31.3 | 0.34 |
| polarC18 | 758.568 | 173.9792 | 31.3 | 0.34 |
| polarC18 | 758.568 | 290.3383 | 31.2 | 0.34 |
| polarC18 | 758.568 | 470.0814 | 31.2 | 0.34 |
| polarC18 | 758.568 | 407.3211 | 31.2 | 0.34 |
| polarC18 | 758.568 | 279.8703 | 31.2 | 0.34 |
| polarC18 | 758.568 | 206.7728 | 31.2 | 0.33 |
| polarC18 | 758.568 | 198.4384 | 31.2 | 0.33 |
| polarC18 | 758.568 | 90.9808 | 31.2 | 0.33 |
| polarC18 | 758.568 | 206.3253 | 31.2 | 0.33 |
| polarC18 | 758.568 | 179.0202 | 31.2 | 0.33 |
| polarC18 | 758.568 | 197.5316 | 31.1 | 0.33 |
| polarC18 | 758.568 | 107.3845 | 31.1 | 0.33 |
| polarC18 | 758.568 | 324.3395 | 31.1 | 0.33 |
| polarC18 | 758.568 | 171.6417 | 31.1 | 0.33 |
| polarC18 | 758.568 | 683.1754 | 31.1 | 0.33 |
| polarC18 | 758.568 | 253.4659 | 31.1 | 0.33 |
| polarC18 | 758.568 | 118.9532 | 31.1 | 0.33 |
| polarC18 | 758.568 | 365.0384 | 31.1 | 0.33 |
| polarC18 | 758.568 | 423.3487 | 31.1 | 0.33 |
| polarC18 | 758.568 | 92.4325 | 31 | 0.33 |
| polarC18 | 758.568 | 359.0484 | 31 | 0.33 |
| polarC18 | 758.568 | 454.6865 | 31 | 0.33 |
| polarC18 | 758.568 | 700.3984 | 30.9 | 0.33 |
| polarC18 | 758.568 | 235.0869 | 30.9 | 0.33 |
| polarC18 | 758.568 | 747.879 | 30.9 | 0.33 |
| polarC18 | 758.568 | 126.0981 | 30.9 | 0.33 |

| | | | | |
|---|---|---|---|---|
| polarC18 | 758.568 | 195.8879 | 30.9 | 0.33 |
| polarC18 | 758.568 | 433.9607 | 30.9 | 0.33 |
| polarC18 | 758.568 | 112.898 | 30.9 | 0.33 |
| polarC18 | 758.568 | 260.0175 | 30.9 | 0.33 |
| polarC18 | 758.568 | 139.2203 | 30.9 | 0.33 |
| polarC18 | 758.568 | 102.6233 | 30.9 | 0.33 |
| polarC18 | 758.568 | 313.3568 | 30.9 | 0.33 |
| polarC18 | 758.568 | 606.7385 | 30.8 | 0.33 |
| polarC18 | 758.568 | 157.1642 | 30.8 | 0.33 |
| polarC18 | 758.568 | 261.3435 | 30.8 | 0.33 |
| polarC18 | 758.568 | 91.636 | 30.8 | 0.33 |
| polarC18 | 758.568 | 113.4161 | 30.8 | 0.33 |
| polarC18 | 758.568 | 93.9814 | 30.8 | 0.33 |
| polarC18 | 758.568 | 276.2289 | 30.8 | 0.33 |
| polarC18 | 758.568 | 235.3685 | 30.8 | 0.33 |
| polarC18 | 758.568 | 97.421 | 30.8 | 0.33 |
| polarC18 | 758.568 | 187.9578 | 30.8 | 0.33 |
| polarC18 | 758.568 | 274.1265 | 30.8 | 0.33 |
| polarC18 | 758.568 | 116.0203 | 30.8 | 0.33 |
| polarC18 | 758.568 | 89.8561 | 30.8 | 0.33 |
| polarC18 | 758.568 | 717.8957 | 30.8 | 0.33 |
| polarC18 | 758.568 | 600.9663 | 30.7 | 0.33 |
| polarC18 | 758.568 | 108.4919 | 30.7 | 0.33 |
| polarC18 | 758.568 | 87.3583 | 30.7 | 0.33 |
| polarC18 | 758.568 | 174.9012 | 30.7 | 0.33 |
| polarC18 | 758.568 | 84.4083 | 30.7 | 0.33 |
| polarC18 | 758.568 | 372.969 | 30.7 | 0.33 |
| polarC18 | 758.568 | 116.8509 | 30.7 | 0.33 |
| polarC18 | 758.568 | 609.7682 | 30.7 | 0.33 |
| polarC18 | 758.568 | 174.3535 | 30.7 | 0.33 |
| polarC18 | 758.568 | 280.5232 | 30.6 | 0.33 |
| polarC18 | 758.568 | 94.5108 | 30.6 | 0.33 |
| polarC18 | 758.568 | 125.8994 | 30.6 | 0.33 |
| polarC18 | 758.568 | 330.9275 | 30.6 | 0.33 |
| polarC18 | 758.568 | 585.3892 | 30.6 | 0.33 |
| polarC18 | 758.568 | 122.1206 | 30.6 | 0.33 |
| polarC18 | 758.568 | 132.3341 | 30.6 | 0.33 |
| polarC18 | 758.568 | 310.5643 | 30.6 | 0.33 |
| polarC18 | 758.568 | 211.2748 | 30.6 | 0.33 |
| polarC18 | 758.568 | 367.7742 | 30.5 | 0.33 |
| polarC18 | 758.568 | 178.4731 | 30.5 | 0.33 |
| polarC18 | 758.568 | 104.1476 | 30.5 | 0.33 |

| | | | | |
|---|---|---|---|---|
| polarC18 | 758.568 | 124.1039 | 30.5 | 0.33 |
| polarC18 | 758.568 | 162.1384 | 30.4 | 0.33 |
| polarC18 | 758.568 | 248.6463 | 30.4 | 0.33 |
| polarC18 | 758.568 | 87.1546 | 30.4 | 0.33 |
| polarC18 | 758.568 | 348.0287 | 30.4 | 0.33 |
| polarC18 | 758.568 | 147.3708 | 30.3 | 0.33 |
| polarC18 | 758.568 | 176.1789 | 30.3 | 0.33 |
| polarC18 | 758.568 | 547.0593 | 30.3 | 0.33 |
| polarC18 | 758.568 | 315.8644 | 30.3 | 0.33 |
| polarC18 | 758.568 | 245.9693 | 30.3 | 0.33 |
| polarC18 | 758.568 | 209.7357 | 30.3 | 0.33 |
| polarC18 | 758.568 | 422.129 | 30.3 | 0.33 |
| polarC18 | 758.568 | 100.3949 | 30.3 | 0.33 |
| polarC18 | 758.568 | 435.7941 | 30.3 | 0.33 |
| polarC18 | 758.568 | 139.7903 | 30.3 | 0.33 |
| polarC18 | 758.568 | 126.113 | 30.3 | 0.33 |
| polarC18 | 758.568 | 218.1371 | 30.3 | 0.32 |
| polarC18 | 758.568 | 105.6901 | 30.2 | 0.32 |
| polarC18 | 758.568 | 438.7132 | 30.2 | 0.32 |
| polarC18 | 758.568 | 743.5072 | 30.2 | 0.32 |
| polarC18 | 758.568 | 116.1313 | 30.2 | 0.32 |
| polarC18 | 758.568 | 91.0813 | 30.2 | 0.32 |
| polarC18 | 758.568 | 232.6719 | 30.2 | 0.32 |
| polarC18 | 758.568 | 209.379 | 30.2 | 0.32 |
| polarC18 | 758.568 | 385.2193 | 30.2 | 0.32 |
| polarC18 | 758.568 | 226.521 | 30.1 | 0.32 |
| polarC18 | 758.568 | 479.6604 | 30.1 | 0.32 |
| polarC18 | 758.568 | 93.4222 | 30.1 | 0.32 |
| polarC18 | 758.568 | 228.4215 | 30.1 | 0.32 |
| polarC18 | 758.568 | 81.9924 | 30.1 | 0.32 |
| polarC18 | 758.568 | 134.9206 | 30.1 | 0.32 |
| polarC18 | 758.568 | 179.2883 | 30.1 | 0.32 |
| polarC18 | 758.568 | 786.9216 | 30.1 | 0.32 |
| polarC18 | 758.568 | 228.8853 | 30.1 | 0.32 |
| polarC18 | 758.568 | 272.5478 | 30.1 | 0.32 |
| polarC18 | 758.568 | 93.4745 | 30.1 | 0.32 |
| polarC18 | 758.568 | 645.134 | 30 | 0.32 |
| polarC18 | 758.568 | 507.8274 | 30 | 0.32 |
| polarC18 | 758.568 | 216.8735 | 30 | 0.32 |
| polarC18 | 758.568 | 152.4778 | 30 | 0.32 |
| polarC18 | 758.568 | 229.7696 | 30 | 0.32 |
| polarC18 | 758.568 | 238.7044 | 30 | 0.32 |

| | | | | |
|---|---|---|---|---|
| polarC18 | 758.568 | 100.0386 | 30 | 0.32 |
| polarC18 | 758.568 | 128.149 | 30 | 0.32 |
| polarC18 | 758.568 | 144.6174 | 30 | 0.32 |
| polarC18 | 758.568 | 131.0559 | 29.9 | 0.32 |
| polarC18 | 758.568 | 189.414 | 29.9 | 0.32 |
| polarC18 | 758.568 | 176.5034 | 29.9 | 0.32 |
| polarC18 | 758.568 | 268.6875 | 29.9 | 0.32 |
| polarC18 | 758.568 | 93.4688 | 29.9 | 0.32 |
| polarC18 | 758.568 | 711.0777 | 29.9 | 0.32 |
| polarC18 | 758.568 | 150.243 | 29.9 | 0.32 |
| polarC18 | 758.568 | 134.9904 | 29.9 | 0.32 |
| polarC18 | 758.568 | 588.3075 | 29.9 | 0.32 |
| polarC18 | 758.568 | 142.6643 | 29.9 | 0.32 |
| polarC18 | 758.568 | 93.8812 | 29.8 | 0.32 |
| polarC18 | 758.568 | 207.8977 | 29.8 | 0.32 |
| polarC18 | 758.568 | 156.8911 | 29.8 | 0.32 |
| polarC18 | 758.568 | 652.0257 | 29.8 | 0.32 |
| polarC18 | 758.568 | 140.4459 | 29.7 | 0.32 |
| polarC18 | 758.568 | 116.6192 | 29.7 | 0.32 |
| polarC18 | 758.568 | 311.1647 | 29.7 | 0.32 |
| polarC18 | 758.568 | 297.9914 | 29.7 | 0.32 |
| polarC18 | 758.568 | 381.7497 | 29.6 | 0.32 |
| polarC18 | 758.568 | 84.7474 | 29.6 | 0.32 |
| polarC18 | 758.568 | 673.9249 | 29.6 | 0.32 |
| polarC18 | 758.568 | 366.0798 | 29.6 | 0.32 |
| polarC18 | 758.568 | 121.2681 | 29.6 | 0.32 |
| polarC18 | 758.568 | 137.3096 | 29.6 | 0.32 |
| polarC18 | 758.568 | 176.2381 | 29.6 | 0.32 |
| polarC18 | 758.568 | 401.3882 | 29.6 | 0.32 |
| polarC18 | 758.568 | 81.4779 | 29.6 | 0.32 |
| polarC18 | 758.568 | 93.4445 | 29.6 | 0.32 |
| polarC18 | 758.568 | 298.1816 | 29.6 | 0.32 |
| polarC18 | 758.568 | 569.8754 | 29.6 | 0.32 |
| polarC18 | 758.568 | 158.0191 | 29.5 | 0.32 |
| polarC18 | 758.568 | 93.9306 | 29.5 | 0.32 |
| polarC18 | 758.568 | 91.3045 | 29.5 | 0.32 |
| polarC18 | 758.568 | 136.8117 | 29.5 | 0.32 |
| polarC18 | 758.568 | 92.2876 | 29.5 | 0.32 |
| polarC18 | 758.568 | 352.1293 | 29.4 | 0.32 |
| polarC18 | 758.568 | 214.8721 | 29.4 | 0.32 |
| polarC18 | 758.568 | 255.4946 | 29.4 | 0.32 |
| polarC18 | 758.568 | 142.7752 | 29.4 | 0.32 |

| | | | | |
|---|---|---|---|---|
| polarC18 | 758.568 | 220.3723 | 29.4 | 0.32 |
| polarC18 | 758.568 | 549.5766 | 29.3 | 0.31 |
| polarC18 | 758.568 | 79.6377 | 29.3 | 0.31 |
| polarC18 | 758.568 | 105.9525 | 29.3 | 0.31 |
| polarC18 | 758.568 | 122.756 | 29.3 | 0.31 |
| polarC18 | 758.568 | 368.6894 | 29.3 | 0.31 |
| polarC18 | 758.568 | 236.5224 | 29.3 | 0.31 |
| polarC18 | 758.568 | 273.6802 | 29.3 | 0.31 |
| polarC18 | 758.568 | 81.2957 | 29.3 | 0.31 |
| polarC18 | 758.568 | 99.709 | 29.3 | 0.31 |
| polarC18 | 758.568 | 331.4672 | 29.2 | 0.31 |
| polarC18 | 758.568 | 486.1061 | 29.2 | 0.31 |
| polarC18 | 758.568 | 680.8493 | 29.2 | 0.31 |
| polarC18 | 758.568 | 203.4067 | 29.2 | 0.31 |
| polarC18 | 758.568 | 606.1878 | 29.2 | 0.31 |
| polarC18 | 758.568 | 706.8303 | 29.2 | 0.31 |
| polarC18 | 758.568 | 240.348 | 29.1 | 0.31 |
| polarC18 | 758.568 | 182.009 | 29.1 | 0.31 |
| polarC18 | 758.568 | 173.0052 | 29.1 | 0.31 |
| polarC18 | 758.568 | 82.1131 | 29.1 | 0.31 |
| polarC18 | 758.568 | 429.0609 | 29.1 | 0.31 |
| polarC18 | 758.568 | 183.3913 | 29 | 0.31 |
| polarC18 | 758.568 | 303.0898 | 29 | 0.31 |
| polarC18 | 758.568 | 136.4847 | 29 | 0.31 |
| polarC18 | 758.568 | 755.4148 | 29 | 0.31 |
| polarC18 | 758.568 | 209.8401 | 29 | 0.31 |
| polarC18 | 758.568 | 105.026 | 29 | 0.31 |
| polarC18 | 758.568 | 175.3219 | 29 | 0.31 |
| polarC18 | 758.568 | 125.2635 | 29 | 0.31 |
| polarC18 | 758.568 | 121.7825 | 28.9 | 0.31 |
| polarC18 | 758.568 | 569.2958 | 28.9 | 0.31 |
| polarC18 | 758.568 | 179.4363 | 28.9 | 0.31 |
| polarC18 | 758.568 | 144.2631 | 28.9 | 0.31 |
| polarC18 | 758.568 | 104.2445 | 28.9 | 0.31 |
| polarC18 | 758.568 | 261.1863 | 28.9 | 0.31 |
| polarC18 | 758.568 | 125.8259 | 28.8 | 0.31 |
| polarC18 | 758.568 | 100.4902 | 28.8 | 0.31 |
| polarC18 | 758.568 | 90.1796 | 28.8 | 0.31 |
| polarC18 | 758.568 | 187.5284 | 28.7 | 0.31 |
| polarC18 | 758.568 | 110.6858 | 28.7 | 0.31 |
| polarC18 | 758.568 | 634.957 | 28.7 | 0.31 |
| polarC18 | 758.568 | 123.2774 | 28.7 | 0.31 |

| | | | | |
|---|---|---|---|---|
| polarC18 | 758.568 | 137.7344 | 28.7 | 0.31 |
| polarC18 | 758.568 | 737.8631 | 28.7 | 0.31 |
| polarC18 | 758.568 | 552.5989 | 28.7 | 0.31 |
| polarC18 | 758.568 | 566.5476 | 28.7 | 0.31 |
| polarC18 | 758.568 | 139.1736 | 28.7 | 0.31 |
| polarC18 | 758.568 | 96.6038 | 28.6 | 0.31 |
| polarC18 | 758.568 | 168.971 | 28.6 | 0.31 |
| polarC18 | 758.568 | 157.5796 | 28.6 | 0.31 |
| polarC18 | 758.568 | 160.0186 | 28.6 | 0.31 |
| polarC18 | 758.568 | 114.2141 | 28.6 | 0.31 |
| polarC18 | 758.568 | 89.8964 | 28.5 | 0.31 |
| polarC18 | 758.568 | 418.9903 | 28.5 | 0.31 |
| polarC18 | 758.568 | 90.2212 | 28.5 | 0.31 |
| polarC18 | 758.568 | 216.28 | 28.5 | 0.31 |
| polarC18 | 758.568 | 738.9446 | 28.5 | 0.31 |
| polarC18 | 758.568 | 432.2214 | 28.5 | 0.31 |
| polarC18 | 758.568 | 99.0901 | 28.5 | 0.31 |
| polarC18 | 758.568 | 192.5421 | 28.4 | 0.31 |
| polarC18 | 758.568 | 156.3294 | 28.4 | 0.3 |
| polarC18 | 758.568 | 206.3851 | 28.4 | 0.3 |
| polarC18 | 758.568 | 178.432 | 28.4 | 0.3 |
| polarC18 | 758.568 | 172.9626 | 28.4 | 0.3 |
| polarC18 | 758.568 | 190.0567 | 28.3 | 0.3 |
| polarC18 | 758.568 | 117.5959 | 28.3 | 0.3 |
| polarC18 | 758.568 | 447.2133 | 28.2 | 0.3 |
| polarC18 | 758.568 | 501.5552 | 28.1 | 0.3 |
| polarC18 | 758.568 | 82.1697 | 28 | 0.3 |
| polarC18 | 758.568 | 79.8492 | 28 | 0.3 |
| polarC18 | 758.568 | 304.0016 | 28 | 0.3 |
| polarC18 | 758.568 | 153.6346 | 28 | 0.3 |
| polarC18 | 758.568 | 91.783 | 27.8 | 0.3 |
| polarC18 | 758.568 | 434.1165 | 27.8 | 0.3 |
| polarC18 | 758.568 | 254.2403 | 27.8 | 0.3 |
| polarC18 | 758.568 | 175.6719 | 27.8 | 0.3 |
| polarC18 | 758.568 | 488.9827 | 27.8 | 0.3 |
| polarC18 | 758.568 | 325.1521 | 27.7 | 0.3 |
| polarC18 | 758.568 | 168.246 | 27.7 | 0.3 |
| polarC18 | 758.568 | 192.7636 | 27.6 | 0.3 |
| polarC18 | 758.568 | 289.684 | 27.6 | 0.3 |
| polarC18 | 758.568 | 209.3103 | 27.6 | 0.3 |
| polarC18 | 758.568 | 301.6291 | 27.5 | 0.3 |
| polarC18 | 758.568 | 219.3062 | 27.5 | 0.3 |

| | | | | |
|---|---|---|---|---|
| polarC18 | 758.568 | 194.5328 | 27.5 | 0.29 |
| polarC18 | 758.568 | 201.0284 | 27.5 | 0.29 |
| polarC18 | 758.568 | 196.7933 | 27.4 | 0.29 |
| polarC18 | 758.568 | 368.7808 | 27.4 | 0.29 |
| polarC18 | 758.568 | 166.5863 | 27.4 | 0.29 |
| polarC18 | 758.568 | 81.271 | 27.4 | 0.29 |
| polarC18 | 758.568 | 674.8447 | 27.3 | 0.29 |
| polarC18 | 758.568 | 177.3909 | 27.3 | 0.29 |
| polarC18 | 758.568 | 93.5477 | 27.3 | 0.29 |
| polarC18 | 758.568 | 121.855 | 27.3 | 0.29 |
| polarC18 | 758.568 | 317.1262 | 27.3 | 0.29 |
| polarC18 | 758.568 | 293.316 | 27.2 | 0.29 |
| polarC18 | 758.568 | 103.7125 | 27.2 | 0.29 |
| polarC18 | 758.568 | 84.5225 | 27.2 | 0.29 |
| polarC18 | 758.568 | 181.4287 | 27.1 | 0.29 |
| polarC18 | 758.568 | 443.1698 | 27.1 | 0.29 |
| polarC18 | 758.568 | 384.6351 | 27 | 0.29 |
| polarC18 | 758.568 | 88.204 | 27 | 0.29 |
| polarC18 | 758.568 | 173.8705 | 27 | 0.29 |
| polarC18 | 758.568 | 94.0133 | 27 | 0.29 |
| polarC18 | 758.568 | 205.8761 | 26.9 | 0.29 |
| polarC18 | 758.568 | 107.1312 | 26.9 | 0.29 |
| polarC18 | 758.568 | 622.8901 | 26.8 | 0.29 |
| polarC18 | 758.568 | 746.5902 | 26.8 | 0.29 |
| polarC18 | 758.568 | 125.8612 | 26.8 | 0.29 |
| polarC18 | 758.568 | 134.6454 | 26.7 | 0.29 |
| polarC18 | 758.568 | 326.6097 | 26.7 | 0.29 |
| polarC18 | 758.568 | 117.2185 | 26.7 | 0.29 |
| polarC18 | 758.568 | 86.2383 | 26.6 | 0.29 |
| polarC18 | 758.568 | 90.1575 | 26.6 | 0.29 |
| polarC18 | 758.568 | 171.132 | 26.6 | 0.29 |
| polarC18 | 758.568 | 708.1892 | 26.6 | 0.29 |
| polarC18 | 758.568 | 430.3363 | 26.5 | 0.28 |
| polarC18 | 758.568 | 129.4351 | 26.5 | 0.28 |
| polarC18 | 758.568 | 385.9273 | 26.4 | 0.28 |
| polarC18 | 758.568 | 89.3918 | 26.4 | 0.28 |
| polarC18 | 758.568 | 91.2497 | 26.4 | 0.28 |
| polarC18 | 758.568 | 90.6236 | 26.4 | 0.28 |
| polarC18 | 758.568 | 258.6185 | 26.3 | 0.28 |
| polarC18 | 758.568 | 270.135 | 26.3 | 0.28 |
| polarC18 | 758.568 | 195.6202 | 26.2 | 0.28 |
| polarC18 | 758.568 | 79.5983 | 26.2 | 0.28 |

| | | | | |
|---|---|---|---|---|
| polarC18 | 758.568 | 721.4362 | 25.8 | 0.28 |
| polarC18 | 758.568 | 212.6148 | 25.8 | 0.28 |
| polarC18 | 758.568 | 545.669 | 25.7 | 0.28 |
| polarC18 | 758.568 | 211.7874 | 25.7 | 0.28 |
| polarC18 | 758.568 | 193.7525 | 25.6 | 0.28 |
| polarC18 | 758.568 | 116.6529 | 25.6 | 0.27 |
| polarC18 | 758.568 | 186.4366 | 25.3 | 0.27 |
| polarC18 | 758.568 | 223.268 | 25.3 | 0.27 |
| polarC18 | 758.568 | 171.6736 | 25.3 | 0.27 |
| polarC18 | 758.568 | 120.7999 | 25.3 | 0.27 |
| polarC18 | 758.568 | 92.7421 | 25.3 | 0.27 |
| polarC18 | 758.568 | 268.471 | 25.3 | 0.27 |
| polarC18 | 758.568 | 498.3634 | 25.1 | 0.27 |
| polarC18 | 758.568 | 357.0838 | 25 | 0.27 |
| polarC18 | 758.568 | 158.7121 | 24.7 | 0.26 |
| polarC18 | 758.568 | 154.7845 | 24.6 | 0.26 |
| polarC18 | 758.568 | 148.3458 | 24.2 | 0.26 |
| polarC18 | 758.568 | 94.1406 | 24.2 | 0.26 |
| polarC18 | 758.568 | 296.3155 | 23.9 | 0.26 |
| polarC18 | 758.568 | 121.5701 | 23.9 | 0.26 |
| polarC18 | 758.568 | 209.0339 | 23 | 0.25 |
| polarC18 | 758.568 | 415.9072 | 22.2 | 0.24 |
| polarC18 | 758.568 | 113.1372 | 21.8 | 0.23 |
| polarC18 | 758.568 | 465.3558 | 21.6 | 0.23 |
| polarC18 | 758.568 | 367.1633 | 21.5 | 0.23 |
| polarC18 | 758.568 | 84.0282 | 21.3 | 0.23 |
| polarC18 | 758.568 | 681.6482 | 21.1 | 0.23 |
| polarC18 | 758.568 | 771.5707 | 21.1 | 0.23 |
| polarC18 | 758.568 | 343.7894 | 21.1 | 0.23 |
| polarC18 | 758.568 | 412.6109 | 21 | 0.23 |
| polarC18 | 758.568 | 468.8224 | 20.9 | 0.22 |
| polarC18 | 758.568 | 258.9432 | 20.9 | 0.22 |
| polarC18 | 758.568 | 647.6249 | 20.8 | 0.22 |
| polarC18 | 758.568 | 157.9933 | 20.7 | 0.22 |
| polarC18 | 758.568 | 323.8842 | 20.7 | 0.22 |
| polarC18 | 758.568 | 184.4439 | 20.7 | 0.22 |
| polarC18 | 758.568 | 93.4631 | 20.7 | 0.22 |
| polarC18 | 758.568 | 126.3834 | 20.7 | 0.22 |
| polarC18 | 758.568 | 106.9646 | 20.7 | 0.22 |
| polarC18 | 758.568 | 86.5904 | 20.6 | 0.22 |
| polarC18 | 758.568 | 93.899 | 20.6 | 0.22 |
| polarC18 | 758.568 | 353.5486 | 20.6 | 0.22 |

| | | | | |
|---|---|---|---|---|
| polarC18 | 758.568 | 545.5001 | 20.5 | 0.22 |
| polarC18 | 758.568 | 280.8355 | 20.5 | 0.22 |
| polarC18 | 758.568 | 106.5075 | 20.5 | 0.22 |
| polarC18 | 758.568 | 142.7106 | 20.5 | 0.22 |
| polarC18 | 758.568 | 782.3963 | 20.5 | 0.22 |
| polarC18 | 758.568 | 93.686 | 20.4 | 0.22 |
| polarC18 | 758.568 | 666.0823 | 20.3 | 0.22 |
| polarC18 | 758.568 | 456.9084 | 20.3 | 0.22 |
| polarC18 | 758.568 | 510.9522 | 20.3 | 0.22 |
| polarC18 | 758.568 | 235.7506 | 20.3 | 0.22 |
| polarC18 | 758.568 | 202.9224 | 20.3 | 0.22 |
| polarC18 | 758.568 | 96.1827 | 20.3 | 0.22 |
| polarC18 | 758.568 | 118.728 | 20.3 | 0.22 |
| polarC18 | 758.568 | 470.8615 | 20.2 | 0.22 |
| polarC18 | 758.568 | 83.2974 | 20.2 | 0.22 |
| polarC18 | 758.568 | 135.4774 | 20.2 | 0.22 |
| polarC18 | 758.568 | 592.7523 | 20.2 | 0.22 |
| polarC18 | 758.568 | 333.547 | 20.2 | 0.22 |
| polarC18 | 758.568 | 102.1365 | 20.1 | 0.22 |
| polarC18 | 758.568 | 638.9346 | 20.1 | 0.22 |
| polarC18 | 758.568 | 243.2843 | 20.1 | 0.22 |
| polarC18 | 758.568 | 257.7477 | 20 | 0.21 |
| polarC18 | 758.568 | 326.9029 | 20 | 0.21 |
| polarC18 | 758.568 | 507.3285 | 19.9 | 0.21 |
| polarC18 | 758.568 | 116.3214 | 19.9 | 0.21 |
| polarC18 | 758.568 | 221.7281 | 19.9 | 0.21 |
| polarC18 | 758.568 | 759.9887 | 19.9 | 0.21 |
| polarC18 | 758.568 | 586.2961 | 19.9 | 0.21 |
| polarC18 | 758.568 | 173.7878 | 19.8 | 0.21 |
| polarC18 | 758.568 | 112.4995 | 19.8 | 0.21 |
| polarC18 | 758.568 | 754.1876 | 19.8 | 0.21 |
| polarC18 | 758.568 | 443.0338 | 19.8 | 0.21 |
| polarC18 | 758.568 | 564.9983 | 19.8 | 0.21 |
| polarC18 | 758.568 | 171.4131 | 19.8 | 0.21 |
| polarC18 | 758.568 | 217.1141 | 19.8 | 0.21 |
| polarC18 | 758.568 | 92.2454 | 19.8 | 0.21 |
| polarC18 | 758.568 | 187.3981 | 19.7 | 0.21 |
| polarC18 | 758.568 | 106.7168 | 19.7 | 0.21 |
| polarC18 | 758.568 | 529.7488 | 19.7 | 0.21 |
| polarC18 | 758.568 | 546.2333 | 19.7 | 0.21 |
| polarC18 | 758.568 | 243.5547 | 19.7 | 0.21 |
| polarC18 | 758.568 | 431.3796 | 19.7 | 0.21 |

| | | | | |
|---|---|---|---|---|
| polarC18 | 758.568 | 689.9322 | 19.7 | 0.21 |
| polarC18 | 758.568 | 363.3965 | 19.7 | 0.21 |
| polarC18 | 758.568 | 136.9327 | 19.7 | 0.21 |
| polarC18 | 758.568 | 169.0205 | 19.7 | 0.21 |
| polarC18 | 758.568 | 345.87 | 19.6 | 0.21 |
| polarC18 | 758.568 | 782.1089 | 19.6 | 0.21 |
| polarC18 | 758.568 | 427.8143 | 19.6 | 0.21 |
| polarC18 | 758.568 | 146.0492 | 19.6 | 0.21 |
| polarC18 | 758.568 | 697.8943 | 19.6 | 0.21 |
| polarC18 | 758.568 | 770.605 | 19.6 | 0.21 |
| polarC18 | 758.568 | 150.9421 | 19.5 | 0.21 |
| polarC18 | 758.568 | 741.0549 | 19.5 | 0.21 |
| polarC18 | 758.568 | 568.1618 | 19.5 | 0.21 |
| polarC18 | 758.568 | 94.0201 | 19.5 | 0.21 |
| polarC18 | 758.568 | 94.3067 | 19.5 | 0.21 |
| polarC18 | 758.568 | 404.8617 | 19.5 | 0.21 |
| polarC18 | 758.568 | 271.0197 | 19.5 | 0.21 |
| polarC18 | 758.568 | 774.6835 | 19.5 | 0.21 |
| polarC18 | 758.568 | 253.9966 | 19.4 | 0.21 |
| polarC18 | 758.568 | 174.7103 | 19.4 | 0.21 |
| polarC18 | 758.568 | 435.9536 | 19.4 | 0.21 |
| polarC18 | 758.568 | 185.7154 | 19.4 | 0.21 |
| polarC18 | 758.568 | 172.7684 | 19.4 | 0.21 |
| polarC18 | 758.568 | 119.9007 | 19.4 | 0.21 |
| polarC18 | 758.568 | 210.7465 | 19.4 | 0.21 |
| polarC18 | 758.568 | 129.2932 | 19.4 | 0.21 |
| polarC18 | 758.568 | 487.0315 | 19.3 | 0.21 |
| polarC18 | 758.568 | 409.0102 | 19.3 | 0.21 |
| polarC18 | 758.568 | 147.8952 | 19.3 | 0.21 |
| polarC18 | 758.568 | 251.0031 | 19.3 | 0.21 |
| polarC18 | 758.568 | 217.7197 | 19.3 | 0.21 |
| polarC18 | 758.568 | 400.9369 | 19.3 | 0.21 |
| polarC18 | 758.568 | 127.8815 | 19.3 | 0.21 |
| polarC18 | 758.568 | 306.7353 | 19.3 | 0.21 |
| polarC18 | 758.568 | 662.4434 | 19.3 | 0.21 |
| polarC18 | 758.568 | 290.1539 | 19.3 | 0.21 |
| polarC18 | 758.568 | 737.2006 | 19.3 | 0.21 |
| polarC18 | 758.568 | 80.8089 | 19.3 | 0.21 |
| polarC18 | 758.568 | 123.4298 | 19.3 | 0.21 |
| polarC18 | 758.568 | 110.6753 | 19.3 | 0.21 |
| polarC18 | 758.568 | 283.3555 | 19.2 | 0.21 |
| polarC18 | 758.568 | 620.4891 | 19.2 | 0.21 |

| | | | | |
|---|---|---|---|---|
| polarC18 | 758.568 | 101.6085 | 19.2 | 0.21 |
| polarC18 | 758.568 | 127.1808 | 19.2 | 0.21 |
| polarC18 | 758.568 | 263.6476 | 19.2 | 0.21 |
| polarC18 | 758.568 | 108.3969 | 19.2 | 0.21 |
| polarC18 | 758.568 | 102.3584 | 19.2 | 0.21 |
| polarC18 | 758.568 | 640.599 | 19.2 | 0.21 |
| polarC18 | 758.568 | 488.7227 | 19.2 | 0.21 |
| polarC18 | 758.568 | 637.2679 | 19.1 | 0.21 |
| polarC18 | 758.568 | 100.8191 | 19.1 | 0.21 |
| polarC18 | 758.568 | 505.7814 | 19.1 | 0.21 |
| polarC18 | 758.568 | 501.3109 | 19.1 | 0.21 |
| polarC18 | 758.568 | 313.4755 | 19.1 | 0.21 |
| polarC18 | 758.568 | 106.859 | 19.1 | 0.21 |
| polarC18 | 758.568 | 606.9551 | 19.1 | 0.21 |
| polarC18 | 758.568 | 106.6583 | 19.1 | 0.21 |
| polarC18 | 758.568 | 509.2198 | 19.1 | 0.2 |
| polarC18 | 758.568 | 184.1871 | 19.1 | 0.2 |
| polarC18 | 758.568 | 494.5038 | 19.1 | 0.2 |
| polarC18 | 758.568 | 565.3222 | 19.1 | 0.2 |
| polarC18 | 758.568 | 95.5385 | 19.1 | 0.2 |
| polarC18 | 758.568 | 330.3364 | 19 | 0.2 |
| polarC18 | 758.568 | 139.0751 | 19 | 0.2 |
| polarC18 | 758.568 | 383.9101 | 19 | 0.2 |
| polarC18 | 758.568 | 382.9674 | 19 | 0.2 |
| polarC18 | 758.568 | 104.397 | 19 | 0.2 |
| polarC18 | 758.568 | 144.7428 | 19 | 0.2 |
| polarC18 | 758.568 | 225.5867 | 19 | 0.2 |
| polarC18 | 758.568 | 104.4427 | 19 | 0.2 |
| polarC18 | 758.568 | 265.0622 | 19 | 0.2 |
| polarC18 | 758.568 | 226.5589 | 19 | 0.2 |
| polarC18 | 758.568 | 675.8099 | 19 | 0.2 |
| polarC18 | 758.568 | 129.1835 | 18.9 | 0.2 |
| polarC18 | 758.568 | 114.3033 | 18.9 | 0.2 |
| polarC18 | 758.568 | 163.9646 | 18.9 | 0.2 |
| polarC18 | 758.568 | 517.8408 | 18.9 | 0.2 |
| polarC18 | 758.568 | 768.8212 | 18.9 | 0.2 |
| polarC18 | 758.568 | 163.5663 | 18.9 | 0.2 |
| polarC18 | 758.568 | 106.0619 | 18.9 | 0.2 |
| polarC18 | 758.568 | 264.0293 | 18.9 | 0.2 |
| polarC18 | 758.568 | 580.925 | 18.9 | 0.2 |
| polarC18 | 758.568 | 206.0292 | 18.9 | 0.2 |
| polarC18 | 758.568 | 83.5079 | 18.9 | 0.2 |

| | | | | |
|---|---|---|---|---|
| polarC18 | 758.568 | 270.7456 | 18.9 | 0.2 |
| polarC18 | 758.568 | 127.766 | 18.8 | 0.2 |
| polarC18 | 758.568 | 446.3443 | 18.8 | 0.2 |
| polarC18 | 758.568 | 110.885 | 18.8 | 0.2 |
| polarC18 | 758.568 | 712.5464 | 18.8 | 0.2 |
| polarC18 | 758.568 | 158.6269 | 18.8 | 0.2 |
| polarC18 | 758.568 | 265.2163 | 18.8 | 0.2 |
| polarC18 | 758.568 | 207.0143 | 18.8 | 0.2 |
| polarC18 | 758.568 | 188.6673 | 18.8 | 0.2 |
| polarC18 | 758.568 | 607.4593 | 18.8 | 0.2 |
| polarC18 | 758.568 | 580.69 | 18.8 | 0.2 |
| polarC18 | 758.568 | 121.284 | 18.8 | 0.2 |
| polarC18 | 758.568 | 574.939 | 18.8 | 0.2 |
| polarC18 | 758.568 | 520.5898 | 18.8 | 0.2 |
| polarC18 | 758.568 | 274.5555 | 18.8 | 0.2 |
| polarC18 | 758.568 | 190.6938 | 18.8 | 0.2 |
| polarC18 | 758.568 | 352.8483 | 18.8 | 0.2 |
| polarC18 | 758.568 | 445.6648 | 18.8 | 0.2 |
| polarC18 | 758.568 | 143.7143 | 18.7 | 0.2 |
| polarC18 | 758.568 | 773.4917 | 18.7 | 0.2 |
| polarC18 | 758.568 | 103.5976 | 18.7 | 0.2 |
| polarC18 | 758.568 | 85.0384 | 18.7 | 0.2 |
| polarC18 | 758.568 | 138.8905 | 18.7 | 0.2 |
| polarC18 | 758.568 | 110.4293 | 18.7 | 0.2 |
| polarC18 | 758.568 | 133.8535 | 18.7 | 0.2 |
| polarC18 | 758.568 | 276.7692 | 18.7 | 0.2 |
| polarC18 | 758.568 | 762.1191 | 18.7 | 0.2 |
| polarC18 | 758.568 | 377.7543 | 18.7 | 0.2 |
| polarC18 | 758.568 | 201.2209 | 18.6 | 0.2 |
| polarC18 | 758.568 | 515.663 | 18.6 | 0.2 |
| polarC18 | 758.568 | 161.5088 | 18.6 | 0.2 |
| polarC18 | 758.568 | 449.0629 | 18.6 | 0.2 |
| polarC18 | 758.568 | 107.156 | 18.6 | 0.2 |
| polarC18 | 758.568 | 385.5256 | 18.6 | 0.2 |
| polarC18 | 758.568 | 250.1041 | 18.6 | 0.2 |
| polarC18 | 758.568 | 237.7785 | 18.6 | 0.2 |
| polarC18 | 758.568 | 111.7413 | 18.6 | 0.2 |
| polarC18 | 758.568 | 82.526 | 18.6 | 0.2 |
| polarC18 | 758.568 | 388.787 | 18.6 | 0.2 |
| polarC18 | 758.568 | 300.1398 | 18.6 | 0.2 |
| polarC18 | 758.568 | 541.2696 | 18.6 | 0.2 |
| polarC18 | 758.568 | 129.1477 | 18.6 | 0.2 |

| | | | | |
|---|---|---|---|---|
| polarC18 | 758.568 | 91.5956 | 18.5 | 0.2 |
| polarC18 | 758.568 | 139.538 | 18.5 | 0.2 |
| polarC18 | 758.568 | 113.7796 | 18.5 | 0.2 |
| polarC18 | 758.568 | 414.5151 | 18.5 | 0.2 |
| polarC18 | 758.568 | 684.2885 | 18.5 | 0.2 |
| polarC18 | 758.568 | 209.477 | 18.5 | 0.2 |
| polarC18 | 758.568 | 119.4349 | 18.5 | 0.2 |
| polarC18 | 758.568 | 126.718 | 18.5 | 0.2 |
| polarC18 | 758.568 | 290.1208 | 18.5 | 0.2 |
| polarC18 | 758.568 | 337.6046 | 18.5 | 0.2 |
| polarC18 | 758.568 | 104.7457 | 18.5 | 0.2 |
| polarC18 | 758.568 | 246.7982 | 18.5 | 0.2 |
| polarC18 | 758.568 | 87.8464 | 18.5 | 0.2 |
| polarC18 | 758.568 | 488.3003 | 18.5 | 0.2 |
| polarC18 | 758.568 | 486.3936 | 18.5 | 0.2 |
| polarC18 | 758.568 | 130.4656 | 18.4 | 0.2 |
| polarC18 | 758.568 | 165.3329 | 18.4 | 0.2 |
| polarC18 | 758.568 | 226.7398 | 18.4 | 0.2 |
| polarC18 | 758.568 | 158.6104 | 18.4 | 0.2 |
| polarC18 | 758.568 | 596.8832 | 18.4 | 0.2 |
| polarC18 | 758.568 | 730.6577 | 18.4 | 0.2 |
| polarC18 | 758.568 | 657.8761 | 18.4 | 0.2 |
| polarC18 | 758.568 | 438.296 | 18.4 | 0.2 |
| polarC18 | 758.568 | 108.2226 | 18.4 | 0.2 |
| polarC18 | 758.568 | 128.3689 | 18.4 | 0.2 |
| polarC18 | 758.568 | 153.0085 | 18.4 | 0.2 |
| polarC18 | 758.568 | 340.8951 | 18.4 | 0.2 |
| polarC18 | 758.568 | 187.1239 | 18.4 | 0.2 |
| polarC18 | 758.568 | 282.1106 | 18.4 | 0.2 |
| polarC18 | 758.568 | 157.0663 | 18.4 | 0.2 |
| polarC18 | 758.568 | 137.2607 | 18.4 | 0.2 |
| polarC18 | 758.568 | 223.0058 | 18.4 | 0.2 |
| polarC18 | 758.568 | 125.497 | 18.4 | 0.2 |
| polarC18 | 758.568 | 370.9913 | 18.3 | 0.2 |
| polarC18 | 758.568 | 275.3757 | 18.3 | 0.2 |
| polarC18 | 758.568 | 385.3277 | 18.3 | 0.2 |
| polarC18 | 758.568 | 131.4345 | 18.3 | 0.2 |
| polarC18 | 758.568 | 154.5069 | 18.3 | 0.2 |
| polarC18 | 758.568 | 230.4861 | 18.3 | 0.2 |
| polarC18 | 758.568 | 103.6251 | 18.3 | 0.2 |
| polarC18 | 758.568 | 252.4656 | 18.3 | 0.2 |
| polarC18 | 758.568 | 93.0884 | 18.3 | 0.2 |

| | | | | |
|---|---|---|---|---|
| polarC18 | 758.568 | 90.6448 | 18.3 | 0.2 |
| polarC18 | 758.568 | 421.5224 | 18.3 | 0.2 |
| polarC18 | 758.568 | 282.2571 | 18.3 | 0.2 |
| polarC18 | 758.568 | 141.4279 | 18.3 | 0.2 |
| polarC18 | 758.568 | 261.8926 | 18.3 | 0.2 |
| polarC18 | 758.568 | 95.0756 | 18.3 | 0.2 |
| polarC18 | 758.568 | 107.4091 | 18.3 | 0.2 |
| polarC18 | 758.568 | 112.4363 | 18.3 | 0.2 |
| polarC18 | 758.568 | 89.5417 | 18.2 | 0.2 |
| polarC18 | 758.568 | 89.0235 | 18.2 | 0.2 |
| polarC18 | 758.568 | 117.6369 | 18.2 | 0.2 |
| polarC18 | 758.568 | 113.5875 | 18.2 | 0.2 |
| polarC18 | 758.568 | 682.2628 | 18.2 | 0.2 |
| polarC18 | 758.568 | 96.2271 | 18.2 | 0.2 |
| polarC18 | 758.568 | 140.3399 | 18.2 | 0.2 |
| polarC18 | 758.568 | 551.5516 | 18.2 | 0.2 |
| polarC18 | 758.568 | 301.986 | 18.2 | 0.2 |
| polarC18 | 758.568 | 207.1811 | 18.2 | 0.2 |
| polarC18 | 758.568 | 287.6743 | 18.2 | 0.2 |
| polarC18 | 758.568 | 162.2798 | 18.2 | 0.2 |
| polarC18 | 758.568 | 104.3804 | 18.2 | 0.2 |
| polarC18 | 758.568 | 345.3932 | 18.2 | 0.2 |
| polarC18 | 758.568 | 444.2228 | 18.2 | 0.2 |
| polarC18 | 758.568 | 262.4036 | 18.2 | 0.2 |
| polarC18 | 758.568 | 550.9045 | 18.2 | 0.2 |
| polarC18 | 758.568 | 103.1167 | 18.2 | 0.2 |
| polarC18 | 758.568 | 189.3395 | 18.2 | 0.2 |
| polarC18 | 758.568 | 110.6278 | 18.2 | 0.19 |
| polarC18 | 758.568 | 129.2856 | 18.2 | 0.19 |
| polarC18 | 758.568 | 104.8468 | 18.1 | 0.19 |
| polarC18 | 758.568 | 143.9801 | 18.1 | 0.19 |
| polarC18 | 758.568 | 130.3765 | 18.1 | 0.19 |
| polarC18 | 758.568 | 97.2134 | 18.1 | 0.19 |
| polarC18 | 758.568 | 147.24 | 18.1 | 0.19 |
| polarC18 | 758.568 | 589.4304 | 18.1 | 0.19 |
| polarC18 | 758.568 | 787.6293 | 18.1 | 0.19 |
| polarC18 | 758.568 | 158.1269 | 18.1 | 0.19 |
| polarC18 | 758.568 | 183.786 | 18.1 | 0.19 |
| polarC18 | 758.568 | 229.641 | 18.1 | 0.19 |
| polarC18 | 758.568 | 604.8594 | 18.1 | 0.19 |
| polarC18 | 758.568 | 345.6603 | 18.1 | 0.19 |
| polarC18 | 758.568 | 639.4299 | 18.1 | 0.19 |

| | | | | |
|---|---|---|---|---|
| polarC18 | 758.568 | 154.9591 | 18.1 | 0.19 |
| polarC18 | 758.568 | 208.9744 | 18.1 | 0.19 |
| polarC18 | 758.568 | 137.2178 | 18.1 | 0.19 |
| polarC18 | 758.568 | 159.2683 | 18.1 | 0.19 |
| polarC18 | 758.568 | 117.0579 | 18.1 | 0.19 |
| polarC18 | 758.568 | 98.497 | 18.1 | 0.19 |
| polarC18 | 758.568 | 118.8495 | 18.1 | 0.19 |
| polarC18 | 758.568 | 122.2243 | 18.1 | 0.19 |
| polarC18 | 758.568 | 560.7272 | 18.1 | 0.19 |
| polarC18 | 758.568 | 172.5136 | 18.1 | 0.19 |
| polarC18 | 758.568 | 702.4642 | 18.1 | 0.19 |
| polarC18 | 758.568 | 577.7904 | 18.1 | 0.19 |
| polarC18 | 758.568 | 79.8187 | 18 | 0.19 |
| polarC18 | 758.568 | 476.7024 | 18 | 0.19 |
| polarC18 | 758.568 | 294.7278 | 18 | 0.19 |
| polarC18 | 758.568 | 90.1929 | 18 | 0.19 |
| polarC18 | 758.568 | 229.1614 | 18 | 0.19 |
| polarC18 | 758.568 | 404.9719 | 18 | 0.19 |
| polarC18 | 758.568 | 406.9966 | 18 | 0.19 |
| polarC18 | 758.568 | 173.1003 | 18 | 0.19 |
| polarC18 | 758.568 | 115.1116 | 18 | 0.19 |
| polarC18 | 758.568 | 329.0844 | 18 | 0.19 |
| polarC18 | 758.568 | 136.707 | 18 | 0.19 |
| polarC18 | 758.568 | 93.9849 | 18 | 0.19 |
| polarC18 | 758.568 | 80.1986 | 18 | 0.19 |
| polarC18 | 758.568 | 197.1436 | 18 | 0.19 |
| polarC18 | 758.568 | 129.6868 | 18 | 0.19 |
| polarC18 | 758.568 | 105.7809 | 18 | 0.19 |
| polarC18 | 758.568 | 281.1842 | 18 | 0.19 |
| polarC18 | 758.568 | 355.4993 | 18 | 0.19 |
| polarC18 | 758.568 | 123.301 | 18 | 0.19 |
| polarC18 | 758.568 | 107.6094 | 17.9 | 0.19 |
| polarC18 | 758.568 | 264.8459 | 17.9 | 0.19 |
| polarC18 | 758.568 | 201.0961 | 17.9 | 0.19 |
| polarC18 | 758.568 | 102.6664 | 17.9 | 0.19 |
| polarC18 | 758.568 | 788.1239 | 17.9 | 0.19 |
| polarC18 | 758.568 | 84.286 | 17.9 | 0.19 |
| polarC18 | 758.568 | 286.6053 | 17.9 | 0.19 |
| polarC18 | 758.568 | 89.6957 | 17.9 | 0.19 |
| polarC18 | 758.568 | 119.7918 | 17.9 | 0.19 |
| polarC18 | 758.568 | 430.034 | 17.9 | 0.19 |
| polarC18 | 758.568 | 337.7226 | 17.9 | 0.19 |

| | | | | |
|---|---|---|---|---|
| polarC18 | 758.568 | 706.4899 | 17.9 | 0.19 |
| polarC18 | 758.568 | 156.2805 | 17.9 | 0.19 |
| polarC18 | 758.568 | 510.8444 | 17.9 | 0.19 |
| polarC18 | 758.568 | 125.2063 | 17.9 | 0.19 |
| polarC18 | 758.568 | 134.8783 | 17.9 | 0.19 |
| polarC18 | 758.568 | 346.9574 | 17.9 | 0.19 |
| polarC18 | 758.568 | 402.1677 | 17.9 | 0.19 |
| polarC18 | 758.568 | 97.886 | 17.9 | 0.19 |
| polarC18 | 758.568 | 275.4548 | 17.9 | 0.19 |
| polarC18 | 758.568 | 591.8104 | 17.9 | 0.19 |
| polarC18 | 758.568 | 153.1943 | 17.9 | 0.19 |
| polarC18 | 758.568 | 467.5083 | 17.9 | 0.19 |
| polarC18 | 758.568 | 247.9421 | 17.9 | 0.19 |
| polarC18 | 758.568 | 674.6337 | 17.9 | 0.19 |
| polarC18 | 758.568 | 100.6589 | 17.9 | 0.19 |
| polarC18 | 758.568 | 281.6547 | 17.9 | 0.19 |
| polarC18 | 758.568 | 111.7777 | 17.9 | 0.19 |
| polarC18 | 758.568 | 693.3245 | 17.9 | 0.19 |
| polarC18 | 758.568 | 759.6169 | 17.9 | 0.19 |
| polarC18 | 758.568 | 570.7897 | 17.8 | 0.19 |
| polarC18 | 758.568 | 203.2973 | 17.8 | 0.19 |
| polarC18 | 758.568 | 207.5468 | 17.8 | 0.19 |
| polarC18 | 758.568 | 304.528 | 17.8 | 0.19 |
| polarC18 | 758.568 | 91.9609 | 17.8 | 0.19 |
| polarC18 | 758.568 | 143.6354 | 17.8 | 0.19 |
| polarC18 | 758.568 | 527.2343 | 17.8 | 0.19 |
| polarC18 | 758.568 | 218.8635 | 17.8 | 0.19 |
| polarC18 | 758.568 | 87.4054 | 17.8 | 0.19 |
| polarC18 | 758.568 | 229.1046 | 17.8 | 0.19 |
| polarC18 | 758.568 | 268.6387 | 17.8 | 0.19 |
| polarC18 | 758.568 | 98.0914 | 17.8 | 0.19 |
| polarC18 | 758.568 | 277.2785 | 17.8 | 0.19 |
| polarC18 | 758.568 | 317.3287 | 17.8 | 0.19 |
| polarC18 | 758.568 | 559.0583 | 17.8 | 0.19 |
| polarC18 | 758.568 | 744.0687 | 17.8 | 0.19 |
| polarC18 | 758.568 | 744.8548 | 17.8 | 0.19 |
| polarC18 | 758.568 | 774.516 | 17.8 | 0.19 |
| polarC18 | 758.568 | 94.9203 | 17.8 | 0.19 |
| polarC18 | 758.568 | 319.5832 | 17.8 | 0.19 |
| polarC18 | 758.568 | 500.4275 | 17.8 | 0.19 |
| polarC18 | 758.568 | 229.6658 | 17.8 | 0.19 |
| polarC18 | 758.568 | 643.0098 | 17.8 | 0.19 |

| | | | | |
|---|---|---|---|---|
| polarC18 | 758.568 | 154.475 | 17.8 | 0.19 |
| polarC18 | 758.568 | 215.4484 | 17.8 | 0.19 |
| polarC18 | 758.568 | 618.752 | 17.8 | 0.19 |
| polarC18 | 758.568 | 109.7717 | 17.8 | 0.19 |
| polarC18 | 758.568 | 248.3616 | 17.8 | 0.19 |
| polarC18 | 758.568 | 261.2818 | 17.8 | 0.19 |
| polarC18 | 758.568 | 214.142 | 17.7 | 0.19 |
| polarC18 | 758.568 | 454.0067 | 17.7 | 0.19 |
| polarC18 | 758.568 | 337.8036 | 17.7 | 0.19 |
| polarC18 | 758.568 | 233.1718 | 17.7 | 0.19 |
| polarC18 | 758.568 | 722.7641 | 17.7 | 0.19 |
| polarC18 | 758.568 | 300.0338 | 17.7 | 0.19 |
| polarC18 | 758.568 | 211.0983 | 17.7 | 0.19 |
| polarC18 | 758.568 | 93.7174 | 17.7 | 0.19 |
| polarC18 | 758.568 | 103.7392 | 17.7 | 0.19 |
| polarC18 | 758.568 | 160.3977 | 17.7 | 0.19 |
| polarC18 | 758.568 | 457.319 | 17.7 | 0.19 |
| polarC18 | 758.568 | 130.6188 | 17.7 | 0.19 |
| polarC18 | 758.568 | 89.7607 | 17.7 | 0.19 |
| polarC18 | 758.568 | 476.3091 | 17.7 | 0.19 |
| polarC18 | 758.568 | 572.5879 | 17.7 | 0.19 |
| polarC18 | 758.568 | 395.6931 | 17.7 | 0.19 |
| polarC18 | 758.568 | 93.3227 | 17.7 | 0.19 |
| polarC18 | 758.568 | 429.7623 | 17.7 | 0.19 |
| polarC18 | 758.568 | 105.0941 | 17.7 | 0.19 |
| polarC18 | 758.568 | 83.0189 | 17.7 | 0.19 |
| polarC18 | 758.568 | 139.2021 | 17.7 | 0.19 |
| polarC18 | 758.568 | 307.6765 | 17.7 | 0.19 |
| polarC18 | 758.568 | 268.252 | 17.7 | 0.19 |
| polarC18 | 758.568 | 358.0944 | 17.7 | 0.19 |
| polarC18 | 758.568 | 94.5553 | 17.7 | 0.19 |
| polarC18 | 758.568 | 120.9099 | 17.7 | 0.19 |
| polarC18 | 758.568 | 339.7427 | 17.7 | 0.19 |
| polarC18 | 758.568 | 277.1715 | 17.7 | 0.19 |
| polarC18 | 758.568 | 275.993 | 17.7 | 0.19 |
| polarC18 | 758.568 | 378.3032 | 17.6 | 0.19 |
| polarC18 | 758.568 | 375.5608 | 17.6 | 0.19 |
| polarC18 | 758.568 | 440.7132 | 17.6 | 0.19 |
| polarC18 | 758.568 | 403.7179 | 17.6 | 0.19 |
| polarC18 | 758.568 | 114.1585 | 17.6 | 0.19 |
| polarC18 | 758.568 | 522.5705 | 17.6 | 0.19 |
| polarC18 | 758.568 | 289.8811 | 17.6 | 0.19 |

| | | | | |
|---|---|---|---|---|
| polarC18 | 758.568 | 85.3657 | 17.6 | 0.19 |
| polarC18 | 758.568 | 518.9308 | 17.6 | 0.19 |
| polarC18 | 758.568 | 238.5372 | 17.6 | 0.19 |
| polarC18 | 758.568 | 184.428 | 17.6 | 0.19 |
| polarC18 | 758.568 | 734.2395 | 17.6 | 0.19 |
| polarC18 | 758.568 | 417.433 | 17.6 | 0.19 |
| polarC18 | 758.568 | 420.7898 | 17.6 | 0.19 |
| polarC18 | 758.568 | 125.019 | 17.6 | 0.19 |
| polarC18 | 758.568 | 193.3757 | 17.6 | 0.19 |
| polarC18 | 758.568 | 185.5773 | 17.6 | 0.19 |
| polarC18 | 758.568 | 87.6262 | 17.6 | 0.19 |
| polarC18 | 758.568 | 184.496 | 17.6 | 0.19 |
| polarC18 | 758.568 | 87.0335 | 17.6 | 0.19 |
| polarC18 | 758.568 | 205.2789 | 17.6 | 0.19 |
| polarC18 | 758.568 | 147.949 | 17.6 | 0.19 |
| polarC18 | 758.568 | 243.3729 | 17.6 | 0.19 |
| polarC18 | 758.568 | 112.9235 | 17.6 | 0.19 |
| polarC18 | 758.568 | 103.8313 | 17.6 | 0.19 |
| polarC18 | 758.568 | 749.9736 | 17.6 | 0.19 |
| polarC18 | 758.568 | 709.993 | 17.6 | 0.19 |
| polarC18 | 758.568 | 129.9413 | 17.6 | 0.19 |
| polarC18 | 758.568 | 98.968 | 17.6 | 0.19 |
| polarC18 | 758.568 | 167.1757 | 17.6 | 0.19 |
| polarC18 | 758.568 | 524.9812 | 17.6 | 0.19 |
| polarC18 | 758.568 | 190.0056 | 17.6 | 0.19 |
| polarC18 | 758.568 | 264.1835 | 17.6 | 0.19 |
| polarC18 | 758.568 | 179.7936 | 17.6 | 0.19 |
| polarC18 | 758.568 | 341.5117 | 17.6 | 0.19 |
| polarC18 | 758.568 | 95.932 | 17.6 | 0.19 |
| polarC18 | 758.568 | 304.2113 | 17.6 | 0.19 |
| polarC18 | 758.568 | 93.7277 | 17.6 | 0.19 |
| polarC18 | 758.568 | 139.7116 | 17.6 | 0.19 |
| polarC18 | 758.568 | 174.6174 | 17.6 | 0.19 |
| polarC18 | 758.568 | 198.56 | 17.6 | 0.19 |
| polarC18 | 758.568 | 311.8446 | 17.5 | 0.19 |
| polarC18 | 758.568 | 125.2534 | 17.5 | 0.19 |
| polarC18 | 758.568 | 266.648 | 17.5 | 0.19 |
| polarC18 | 758.568 | 100.0582 | 17.5 | 0.19 |
| polarC18 | 758.568 | 111.1908 | 17.5 | 0.19 |
| polarC18 | 758.568 | 198.9854 | 17.5 | 0.19 |
| polarC18 | 758.568 | 332.7788 | 17.5 | 0.19 |
| polarC18 | 758.568 | 331.0442 | 17.5 | 0.19 |

| | | | | |
|---|---|---|---|---|
| polarC18 | 758.568 | 354.0448 | 17.5 | 0.19 |
| polarC18 | 758.568 | 113.8386 | 17.5 | 0.19 |
| polarC18 | 758.568 | 269.6553 | 17.5 | 0.19 |
| polarC18 | 758.568 | 109.6845 | 17.5 | 0.19 |
| polarC18 | 758.568 | 187.452 | 17.5 | 0.19 |
| polarC18 | 758.568 | 194.2101 | 17.5 | 0.19 |
| polarC18 | 758.568 | 411.949 | 17.5 | 0.19 |
| polarC18 | 758.568 | 334.179 | 17.5 | 0.19 |
| polarC18 | 758.568 | 570.0958 | 17.5 | 0.19 |
| polarC18 | 758.568 | 230.343 | 17.5 | 0.19 |
| polarC18 | 758.568 | 603.5773 | 17.5 | 0.19 |
| polarC18 | 758.568 | 176.5685 | 17.5 | 0.19 |
| polarC18 | 758.568 | 191.0273 | 17.5 | 0.19 |
| polarC18 | 758.568 | 293.588 | 17.5 | 0.19 |
| polarC18 | 758.568 | 268.0894 | 17.5 | 0.19 |
| polarC18 | 758.568 | 87.0418 | 17.5 | 0.19 |
| polarC18 | 758.568 | 475.9446 | 17.5 | 0.19 |
| polarC18 | 758.568 | 130.6387 | 17.5 | 0.19 |
| polarC18 | 758.568 | 763.2436 | 17.5 | 0.19 |
| polarC18 | 758.568 | 130.0732 | 17.5 | 0.19 |
| polarC18 | 758.568 | 129.7827 | 17.5 | 0.19 |
| polarC18 | 758.568 | 155.3887 | 17.5 | 0.19 |
| polarC18 | 758.568 | 220.8185 | 17.5 | 0.19 |
| polarC18 | 758.568 | 82.1631 | 17.5 | 0.19 |
| polarC18 | 758.568 | 323.975 | 17.5 | 0.19 |
| polarC18 | 758.568 | 107.7366 | 17.5 | 0.19 |
| polarC18 | 758.568 | 221.5003 | 17.5 | 0.19 |
| polarC18 | 758.568 | 99.9983 | 17.5 | 0.19 |
| polarC18 | 758.568 | 383.4756 | 17.5 | 0.19 |
| polarC18 | 758.568 | 173.3686 | 17.5 | 0.19 |
| polarC18 | 758.568 | 117.0814 | 17.5 | 0.19 |
| polarC18 | 758.568 | 144.3806 | 17.5 | 0.19 |
| polarC18 | 758.568 | 205.7359 | 17.5 | 0.19 |
| polarC18 | 758.568 | 86.6111 | 17.5 | 0.19 |
| polarC18 | 758.568 | 343.1602 | 17.5 | 0.19 |
| polarC18 | 758.568 | 398.9414 | 17.5 | 0.19 |
| polarC18 | 758.568 | 171.8976 | 17.5 | 0.19 |
| polarC18 | 758.568 | 290.7986 | 17.5 | 0.19 |
| polarC18 | 758.568 | 708.4603 | 17.5 | 0.19 |
| polarC18 | 758.568 | 249.0483 | 17.5 | 0.19 |
| polarC18 | 758.568 | 174.4277 | 17.4 | 0.19 |
| polarC18 | 758.568 | 368.0533 | 17.4 | 0.19 |

| | | | | |
|---|---|---|---|---|
| polarC18 | 758.568 | 143.1941 | 17.4 | 0.19 |
| polarC18 | 758.568 | 727.3499 | 17.4 | 0.19 |
| polarC18 | 758.568 | 199.8829 | 17.4 | 0.19 |
| polarC18 | 758.568 | 463.0892 | 17.4 | 0.19 |
| polarC18 | 758.568 | 134.0092 | 17.4 | 0.19 |
| polarC18 | 758.568 | 95.0155 | 17.4 | 0.19 |
| polarC18 | 758.568 | 101.8903 | 17.4 | 0.19 |
| polarC18 | 758.568 | 121.9302 | 17.4 | 0.19 |
| polarC18 | 758.568 | 100.7308 | 17.4 | 0.19 |
| polarC18 | 758.568 | 340.0963 | 17.4 | 0.19 |
| polarC18 | 758.568 | 667.3208 | 17.4 | 0.19 |
| polarC18 | 758.568 | 375.3645 | 17.4 | 0.19 |
| polarC18 | 758.568 | 199.0375 | 17.4 | 0.19 |
| polarC18 | 758.568 | 513.6851 | 17.4 | 0.19 |
| polarC18 | 758.568 | 152.4651 | 17.4 | 0.19 |
| polarC18 | 758.568 | 287.9217 | 17.4 | 0.19 |
| polarC18 | 758.568 | 698.8494 | 17.4 | 0.19 |
| polarC18 | 758.568 | 321.6542 | 17.4 | 0.19 |
| polarC18 | 758.568 | 153.2229 | 17.4 | 0.19 |
| polarC18 | 758.568 | 638.5596 | 17.4 | 0.19 |
| polarC18 | 758.568 | 617.9875 | 17.4 | 0.19 |
| polarC18 | 758.568 | 342.9221 | 17.4 | 0.19 |
| polarC18 | 758.568 | 103.5747 | 17.4 | 0.19 |
| polarC18 | 758.568 | 745.3603 | 17.4 | 0.19 |
| polarC18 | 758.568 | 93.555 | 17.4 | 0.19 |
| polarC18 | 758.568 | 503.8463 | 17.4 | 0.19 |
| polarC18 | 758.568 | 625.8473 | 17.4 | 0.19 |
| polarC18 | 758.568 | 559.2994 | 17.4 | 0.19 |
| polarC18 | 758.568 | 137.5606 | 17.4 | 0.19 |
| polarC18 | 758.568 | 141.8501 | 17.4 | 0.19 |
| polarC18 | 758.568 | 90.3456 | 17.4 | 0.19 |
| polarC18 | 758.568 | 242.3158 | 17.4 | 0.19 |
| polarC18 | 758.568 | 508.5279 | 17.4 | 0.19 |
| polarC18 | 758.568 | 486.887 | 17.4 | 0.19 |
| polarC18 | 758.568 | 401.1347 | 17.4 | 0.19 |
| polarC18 | 758.568 | 529.1574 | 17.4 | 0.19 |
| polarC18 | 758.568 | 607.2969 | 17.4 | 0.19 |
| polarC18 | 758.568 | 96.6894 | 17.4 | 0.19 |
| polarC18 | 758.568 | 663.8499 | 17.4 | 0.19 |
| polarC18 | 758.568 | 172.0831 | 17.4 | 0.19 |
| polarC18 | 758.568 | 200.7528 | 17.4 | 0.19 |
| polarC18 | 758.568 | 81.4628 | 17.4 | 0.19 |

| | | | | |
|---|---|---|---|---|
| polarC18 | 758.568 | 329.4777 | 17.4 | 0.19 |
| polarC18 | 758.568 | 87.8794 | 17.3 | 0.19 |
| polarC18 | 758.568 | 105.3319 | 17.3 | 0.19 |
| polarC18 | 758.568 | 109.3971 | 17.3 | 0.19 |
| polarC18 | 758.568 | 80.6049 | 17.3 | 0.19 |
| polarC18 | 758.568 | 373.3003 | 17.3 | 0.19 |
| polarC18 | 758.568 | 95.1686 | 17.3 | 0.19 |
| polarC18 | 758.568 | 201.9367 | 17.3 | 0.19 |
| polarC18 | 758.568 | 305.7296 | 17.3 | 0.19 |
| polarC18 | 758.568 | 136.7549 | 17.3 | 0.19 |
| polarC18 | 758.568 | 404.6167 | 17.3 | 0.19 |
| polarC18 | 758.568 | 239.9558 | 17.3 | 0.19 |
| polarC18 | 758.568 | 85.3896 | 17.3 | 0.19 |
| polarC18 | 758.568 | 92.4879 | 17.3 | 0.19 |
| polarC18 | 758.568 | 108.5706 | 17.3 | 0.19 |
| polarC18 | 758.568 | 149.9648 | 17.3 | 0.19 |
| polarC18 | 758.568 | 267.5009 | 17.3 | 0.19 |
| polarC18 | 758.568 | 83.7489 | 17.3 | 0.19 |
| polarC18 | 758.568 | 122.0446 | 17.3 | 0.19 |
| polarC18 | 758.568 | 693.845 | 17.3 | 0.19 |
| polarC18 | 758.568 | 83.992 | 17.3 | 0.19 |
| polarC18 | 758.568 | 148.116 | 17.3 | 0.19 |
| polarC18 | 758.568 | 500.5747 | 17.3 | 0.19 |
| polarC18 | 758.568 | 532.4947 | 17.3 | 0.19 |
| polarC18 | 758.568 | 81.4971 | 17.3 | 0.19 |
| polarC18 | 758.568 | 699.3387 | 17.3 | 0.19 |
| polarC18 | 758.568 | 353.8746 | 17.3 | 0.19 |
| polarC18 | 758.568 | 95.0192 | 17.3 | 0.19 |
| polarC18 | 758.568 | 188.315 | 17.3 | 0.19 |
| polarC18 | 758.568 | 114.1278 | 17.3 | 0.19 |
| polarC18 | 758.568 | 322.808 | 17.3 | 0.19 |
| polarC18 | 758.568 | 507.1833 | 17.3 | 0.19 |
| polarC18 | 758.568 | 208.8514 | 17.3 | 0.19 |
| polarC18 | 758.568 | 127.7016 | 17.3 | 0.19 |
| polarC18 | 758.568 | 144.5866 | 17.3 | 0.19 |
| polarC18 | 758.568 | 119.3757 | 17.3 | 0.19 |
| polarC18 | 758.568 | 136.1934 | 17.3 | 0.19 |
| polarC18 | 758.568 | 256.8172 | 17.3 | 0.19 |
| polarC18 | 758.568 | 636.6482 | 17.2 | 0.19 |
| polarC18 | 758.568 | 81.2155 | 17.2 | 0.19 |
| polarC18 | 758.568 | 275.9549 | 17.2 | 0.19 |
| polarC18 | 758.568 | 89.8795 | 17.2 | 0.19 |

| | | | | |
|---|---|---|---|---|
| polarC18 | 758.568 | 143.3768 | 17.2 | 0.19 |
| polarC18 | 758.568 | 215.6439 | 17.2 | 0.19 |
| polarC18 | 758.568 | 330.542 | 17.2 | 0.19 |
| polarC18 | 758.568 | 221.6524 | 17.2 | 0.19 |
| polarC18 | 758.568 | 83.562 | 17.2 | 0.19 |
| polarC18 | 758.568 | 196.2342 | 17.2 | 0.19 |
| polarC18 | 758.568 | 717.1204 | 17.2 | 0.18 |
| polarC18 | 758.568 | 108.1022 | 17.2 | 0.18 |
| polarC18 | 758.568 | 652.8816 | 17.2 | 0.18 |
| polarC18 | 758.568 | 334.0771 | 17.2 | 0.18 |
| polarC18 | 758.568 | 87.4511 | 17.2 | 0.18 |
| polarC18 | 758.568 | 730.928 | 17.2 | 0.18 |
| polarC18 | 758.568 | 146.0304 | 17.2 | 0.18 |
| polarC18 | 758.568 | 152.6809 | 17.2 | 0.18 |
| polarC18 | 758.568 | 112.2141 | 17.2 | 0.18 |
| polarC18 | 758.568 | 372.8379 | 17.2 | 0.18 |
| polarC18 | 758.568 | 136.0274 | 17.2 | 0.18 |
| polarC18 | 758.568 | 266.8529 | 17.2 | 0.18 |
| polarC18 | 758.568 | 83.6846 | 17.2 | 0.18 |
| polarC18 | 758.568 | 316.8603 | 17.2 | 0.18 |
| polarC18 | 758.568 | 313.9763 | 17.2 | 0.18 |
| polarC18 | 758.568 | 327.8082 | 17.2 | 0.18 |
| polarC18 | 758.568 | 120.4979 | 17.2 | 0.18 |
| polarC18 | 758.568 | 248.141 | 17.2 | 0.18 |
| polarC18 | 758.568 | 87.4317 | 17.2 | 0.18 |
| polarC18 | 758.568 | 725.728 | 17.2 | 0.18 |
| polarC18 | 758.568 | 135.8114 | 17.2 | 0.18 |
| polarC18 | 758.568 | 388.4434 | 17.2 | 0.18 |
| polarC18 | 758.568 | 543.4165 | 17.2 | 0.18 |
| polarC18 | 758.568 | 322.4914 | 17.2 | 0.18 |
| polarC18 | 758.568 | 86.3922 | 17.2 | 0.18 |
| polarC18 | 758.568 | 339.4155 | 17.2 | 0.18 |
| polarC18 | 758.568 | 273.1199 | 17.2 | 0.18 |
| polarC18 | 758.568 | 138.0164 | 17.2 | 0.18 |
| polarC18 | 758.568 | 282.0777 | 17.2 | 0.18 |
| polarC18 | 758.568 | 180.9677 | 17.2 | 0.18 |
| polarC18 | 758.568 | 533.2825 | 17.2 | 0.18 |
| polarC18 | 758.568 | 79.8821 | 17.2 | 0.18 |
| polarC18 | 758.568 | 629.4055 | 17.2 | 0.18 |
| polarC18 | 758.568 | 195.1722 | 17.2 | 0.18 |
| polarC18 | 758.568 | 731.5939 | 17.1 | 0.18 |
| polarC18 | 758.568 | 97.3516 | 17.1 | 0.18 |

| | | | | |
|---|---|---|---|---|
| polarC18 | 758.568 | 151.1053 | 17.1 | 0.18 |
| polarC18 | 758.568 | 257.7151 | 17.1 | 0.18 |
| polarC18 | 758.568 | 426.2963 | 17.1 | 0.18 |
| polarC18 | 758.568 | 167.1899 | 17.1 | 0.18 |
| polarC18 | 758.568 | 382.0075 | 17.1 | 0.18 |
| polarC18 | 758.568 | 250.1303 | 17.1 | 0.18 |
| polarC18 | 758.568 | 79.8043 | 17.1 | 0.18 |
| polarC18 | 758.568 | 721.8277 | 17.1 | 0.18 |
| polarC18 | 758.568 | 553.0522 | 17.1 | 0.18 |
| polarC18 | 758.568 | 327.1541 | 17.1 | 0.18 |
| polarC18 | 758.568 | 163.1006 | 17.1 | 0.18 |
| polarC18 | 758.568 | 103.9314 | 17.1 | 0.18 |
| polarC18 | 758.568 | 145.8654 | 17.1 | 0.18 |
| polarC18 | 758.568 | 104.0247 | 17.1 | 0.18 |
| polarC18 | 758.568 | 121.3892 | 17.1 | 0.18 |
| polarC18 | 758.568 | 95.0835 | 17.1 | 0.18 |
| polarC18 | 758.568 | 199.4662 | 17.1 | 0.18 |
| polarC18 | 758.568 | 86.1927 | 17.1 | 0.18 |
| polarC18 | 758.568 | 106.181 | 17.1 | 0.18 |
| polarC18 | 758.568 | 371.6214 | 17.1 | 0.18 |
| polarC18 | 758.568 | 230.7103 | 17.1 | 0.18 |
| polarC18 | 758.568 | 714.4579 | 17.1 | 0.18 |
| polarC18 | 758.568 | 273.4292 | 17.1 | 0.18 |
| polarC18 | 758.568 | 88.6596 | 17.1 | 0.18 |
| polarC18 | 758.568 | 394.4414 | 17.1 | 0.18 |
| polarC18 | 758.568 | 541.5112 | 17.1 | 0.18 |
| polarC18 | 758.568 | 773.9875 | 17.1 | 0.18 |
| polarC18 | 758.568 | 177.6699 | 17.1 | 0.18 |
| polarC18 | 758.568 | 197.7806 | 17.1 | 0.18 |
| polarC18 | 758.568 | 282.575 | 17.1 | 0.18 |
| polarC18 | 758.568 | 276.3064 | 17.1 | 0.18 |
| polarC18 | 758.568 | 310.0628 | 17.1 | 0.18 |
| polarC18 | 758.568 | 110.2013 | 17.1 | 0.18 |
| polarC18 | 758.568 | 765.7136 | 17.1 | 0.18 |
| polarC18 | 758.568 | 166.9416 | 17.1 | 0.18 |
| polarC18 | 758.568 | 109.5682 | 17.1 | 0.18 |
| polarC18 | 758.568 | 207.4083 | 17.1 | 0.18 |
| polarC18 | 758.568 | 344.0364 | 17.1 | 0.18 |
| polarC18 | 758.568 | 173.5116 | 17.1 | 0.18 |
| polarC18 | 758.568 | 340.0381 | 17.1 | 0.18 |
| polarC18 | 758.568 | 467.9834 | 17.1 | 0.18 |
| polarC18 | 758.568 | 183.8311 | 17.1 | 0.18 |

| | | | | |
|---|---|---|---|---|
| polarC18 | 758.568 | 146.695 | 17.1 | 0.18 |
| polarC18 | 758.568 | 736.2866 | 17.1 | 0.18 |
| polarC18 | 758.568 | 158.8732 | 17.1 | 0.18 |
| polarC18 | 758.568 | 330.6766 | 17.1 | 0.18 |
| polarC18 | 758.568 | 542.1097 | 17.1 | 0.18 |
| polarC18 | 758.568 | 88.2958 | 17.1 | 0.18 |
| polarC18 | 758.568 | 503.2429 | 17.1 | 0.18 |
| polarC18 | 758.568 | 173.0799 | 17.1 | 0.18 |
| polarC18 | 758.568 | 86.2182 | 17.1 | 0.18 |
| polarC18 | 758.568 | 307.9211 | 17.1 | 0.18 |
| polarC18 | 758.568 | 419.3973 | 17.1 | 0.18 |
| polarC18 | 758.568 | 239.3926 | 17.1 | 0.18 |
| polarC18 | 758.568 | 230.4195 | 17 | 0.18 |
| polarC18 | 758.568 | 598.5119 | 17 | 0.18 |
| polarC18 | 758.568 | 148.8374 | 17 | 0.18 |
| polarC18 | 758.568 | 177.5907 | 17 | 0.18 |
| polarC18 | 758.568 | 479.1919 | 17 | 0.18 |
| polarC18 | 758.568 | 83.8262 | 17 | 0.18 |
| polarC18 | 758.568 | 375.2651 | 17 | 0.18 |
| polarC18 | 758.568 | 105.7854 | 17 | 0.18 |
| polarC18 | 758.568 | 93.4856 | 17 | 0.18 |
| polarC18 | 758.568 | 674.1382 | 17 | 0.18 |
| polarC18 | 758.568 | 151.251 | 17 | 0.18 |
| polarC18 | 758.568 | 319.1723 | 17 | 0.18 |
| polarC18 | 758.568 | 563.2303 | 17 | 0.18 |
| polarC18 | 758.568 | 258.2904 | 17 | 0.18 |
| polarC18 | 758.568 | 83.0577 | 17 | 0.18 |
| polarC18 | 758.568 | 423.4828 | 17 | 0.18 |
| polarC18 | 758.568 | 83.2054 | 17 | 0.18 |
| polarC18 | 758.568 | 112.9485 | 17 | 0.18 |
| polarC18 | 758.568 | 550.1952 | 17 | 0.18 |
| polarC18 | 758.568 | 694.9625 | 17 | 0.18 |
| polarC18 | 758.568 | 102.2094 | 17 | 0.18 |
| polarC18 | 758.568 | 79.4834 | 17 | 0.18 |
| polarC18 | 758.568 | 96.307 | 17 | 0.18 |
| polarC18 | 758.568 | 106.6834 | 17 | 0.18 |
| polarC18 | 758.568 | 100.0993 | 17 | 0.18 |
| polarC18 | 758.568 | 91.5599 | 17 | 0.18 |
| polarC18 | 758.568 | 165.4364 | 17 | 0.18 |
| polarC18 | 758.568 | 83.7636 | 17 | 0.18 |
| polarC18 | 758.568 | 104.0444 | 17 | 0.18 |
| polarC18 | 758.568 | 650.2105 | 17 | 0.18 |

| | | | | |
|---|---|---|---|---|
| polarC18 | 758.568 | 231.1118 | 17 | 0.18 |
| polarC18 | 758.568 | 187.2572 | 17 | 0.18 |
| polarC18 | 758.568 | 91.182 | 17 | 0.18 |
| polarC18 | 758.568 | 138.8188 | 17 | 0.18 |
| polarC18 | 758.568 | 173.7005 | 17 | 0.18 |
| polarC18 | 758.568 | 648.5642 | 17 | 0.18 |
| polarC18 | 758.568 | 94.9145 | 17 | 0.18 |
| polarC18 | 758.568 | 619.6038 | 17 | 0.18 |
| polarC18 | 758.568 | 703.1038 | 17 | 0.18 |
| polarC18 | 758.568 | 102.0608 | 17 | 0.18 |
| polarC18 | 758.568 | 84.9514 | 17 | 0.18 |
| polarC18 | 758.568 | 278.2001 | 17 | 0.18 |
| polarC18 | 758.568 | 101.0262 | 17 | 0.18 |
| polarC18 | 758.568 | 591.6637 | 17 | 0.18 |
| polarC18 | 758.568 | 613.1049 | 17 | 0.18 |
| polarC18 | 758.568 | 280.4803 | 17 | 0.18 |
| polarC18 | 758.568 | 93.642 | 17 | 0.18 |
| polarC18 | 758.568 | 695.9827 | 17 | 0.18 |
| polarC18 | 758.568 | 191.9685 | 17 | 0.18 |
| polarC18 | 758.568 | 534.2426 | 17 | 0.18 |
| polarC18 | 758.568 | 83.5668 | 17 | 0.18 |
| polarC18 | 758.568 | 253.1131 | 17 | 0.18 |
| polarC18 | 758.568 | 514.9058 | 17 | 0.18 |
| polarC18 | 758.568 | 191.6168 | 17 | 0.18 |
| polarC18 | 758.568 | 99.9251 | 16.9 | 0.18 |
| polarC18 | 758.568 | 388.9401 | 16.9 | 0.18 |
| polarC18 | 758.568 | 164.1092 | 16.9 | 0.18 |
| polarC18 | 758.568 | 312.6479 | 16.9 | 0.18 |
| polarC18 | 758.568 | 345.9408 | 16.9 | 0.18 |
| polarC18 | 758.568 | 133.4722 | 16.9 | 0.18 |
| polarC18 | 758.568 | 122.1961 | 16.9 | 0.18 |
| polarC18 | 758.568 | 350.6415 | 16.9 | 0.18 |
| polarC18 | 758.568 | 198.0089 | 16.9 | 0.18 |
| polarC18 | 758.568 | 144.9786 | 16.9 | 0.18 |
| polarC18 | 758.568 | 96.6531 | 16.9 | 0.18 |
| polarC18 | 758.568 | 213.3615 | 16.9 | 0.18 |
| polarC18 | 758.568 | 227.599 | 16.9 | 0.18 |
| polarC18 | 758.568 | 131.7168 | 16.9 | 0.18 |
| polarC18 | 758.568 | 149.8385 | 16.9 | 0.18 |
| polarC18 | 758.568 | 111.3733 | 16.9 | 0.18 |
| polarC18 | 758.568 | 103.7021 | 16.9 | 0.18 |
| polarC18 | 758.568 | 191.5734 | 16.9 | 0.18 |

| | | | | |
|---|---|---|---|---|
| polarC18 | 758.568 | 96.3765 | 16.9 | 0.18 |
| polarC18 | 758.568 | 497.3742 | 16.9 | 0.18 |
| polarC18 | 758.568 | 447.6434 | 16.9 | 0.18 |
| polarC18 | 758.568 | 198.1328 | 16.9 | 0.18 |
| polarC18 | 758.568 | 217.578 | 16.9 | 0.18 |
| polarC18 | 758.568 | 87.0547 | 16.9 | 0.18 |
| polarC18 | 758.568 | 365.4494 | 16.9 | 0.18 |
| polarC18 | 758.568 | 460.3684 | 16.9 | 0.18 |
| polarC18 | 758.568 | 108.1594 | 16.9 | 0.18 |
| polarC18 | 758.568 | 105.6539 | 16.9 | 0.18 |
| polarC18 | 758.568 | 89.7065 | 16.9 | 0.18 |
| polarC18 | 758.568 | 80.0981 | 16.9 | 0.18 |
| polarC18 | 758.568 | 133.767 | 16.9 | 0.18 |
| polarC18 | 758.568 | 80.2607 | 16.9 | 0.18 |
| polarC18 | 758.568 | 115.2819 | 16.9 | 0.18 |
| polarC18 | 758.568 | 294.6127 | 16.9 | 0.18 |
| polarC18 | 758.568 | 283.7633 | 16.9 | 0.18 |
| polarC18 | 758.568 | 539.8664 | 16.9 | 0.18 |
| polarC18 | 758.568 | 82.5008 | 16.9 | 0.18 |
| polarC18 | 758.568 | 93.1398 | 16.9 | 0.18 |
| polarC18 | 758.568 | 108.6399 | 16.9 | 0.18 |
| polarC18 | 758.568 | 205.8956 | 16.9 | 0.18 |
| polarC18 | 758.568 | 93.7943 | 16.9 | 0.18 |
| polarC18 | 758.568 | 81.9287 | 16.9 | 0.18 |
| polarC18 | 758.568 | 83.8031 | 16.9 | 0.18 |
| polarC18 | 758.568 | 315.8022 | 16.9 | 0.18 |
| polarC18 | 758.568 | 220.0344 | 16.9 | 0.18 |
| polarC18 | 758.568 | 313.6249 | 16.9 | 0.18 |
| polarC18 | 758.568 | 480.3097 | 16.9 | 0.18 |
| polarC18 | 758.568 | 775.2417 | 16.9 | 0.18 |
| polarC18 | 758.568 | 456.1911 | 16.9 | 0.18 |
| polarC18 | 758.568 | 328.4391 | 16.9 | 0.18 |
| polarC18 | 758.568 | 82.9107 | 16.9 | 0.18 |
| polarC18 | 758.568 | 266.4236 | 16.9 | 0.18 |
| polarC18 | 758.568 | 98.6927 | 16.9 | 0.18 |
| polarC18 | 758.568 | 90.6684 | 16.9 | 0.18 |
| polarC18 | 758.568 | 202.3602 | 16.9 | 0.18 |
| polarC18 | 758.568 | 518.8033 | 16.9 | 0.18 |
| polarC18 | 758.568 | 112.2232 | 16.9 | 0.18 |
| polarC18 | 758.568 | 230.0071 | 16.9 | 0.18 |
| polarC18 | 758.568 | 335.1939 | 16.9 | 0.18 |
| polarC18 | 758.568 | 146.134 | 16.9 | 0.18 |

| | | | | |
|---|---|---|---|---|
| polarC18 | 758.568 | 197.6467 | 16.9 | 0.18 |
| polarC18 | 758.568 | 233.6865 | 16.9 | 0.18 |
| polarC18 | 758.568 | 94.3117 | 16.9 | 0.18 |
| polarC18 | 758.568 | 757.128 | 16.8 | 0.18 |
| polarC18 | 758.568 | 361.2537 | 16.8 | 0.18 |
| polarC18 | 758.568 | 715.7186 | 16.8 | 0.18 |
| polarC18 | 758.568 | 237.6783 | 16.8 | 0.18 |
| polarC18 | 758.568 | 364.4766 | 16.8 | 0.18 |
| polarC18 | 758.568 | 111.9743 | 16.8 | 0.18 |
| polarC18 | 758.568 | 93.9061 | 16.8 | 0.18 |
| polarC18 | 758.568 | 129.9047 | 16.8 | 0.18 |
| polarC18 | 758.568 | 597.1021 | 16.8 | 0.18 |
| polarC18 | 758.568 | 233.958 | 16.8 | 0.18 |
| polarC18 | 758.568 | 454.3232 | 16.8 | 0.18 |
| polarC18 | 758.568 | 89.4742 | 16.8 | 0.18 |
| polarC18 | 758.568 | 112.3707 | 16.8 | 0.18 |
| polarC18 | 758.568 | 491.8673 | 16.8 | 0.18 |
| polarC18 | 758.568 | 527.6467 | 16.8 | 0.18 |
| polarC18 | 758.568 | 128.9346 | 16.8 | 0.18 |
| polarC18 | 758.568 | 87.0586 | 16.8 | 0.18 |
| polarC18 | 758.568 | 87.3439 | 16.8 | 0.18 |
| polarC18 | 758.568 | 84.7813 | 16.8 | 0.18 |
| polarC18 | 758.568 | 122.8296 | 16.8 | 0.18 |
| polarC18 | 758.568 | 437.1249 | 16.8 | 0.18 |
| polarC18 | 758.568 | 195.1425 | 16.8 | 0.18 |
| polarC18 | 758.568 | 283.6963 | 16.8 | 0.18 |
| polarC18 | 758.568 | 625.5013 | 16.8 | 0.18 |
| polarC18 | 758.568 | 456.6227 | 16.8 | 0.18 |
| polarC18 | 758.568 | 226.0388 | 16.8 | 0.18 |
| polarC18 | 758.568 | 231.8658 | 16.8 | 0.18 |
| polarC18 | 758.568 | 154.766 | 16.8 | 0.18 |
| polarC18 | 758.568 | 85.1247 | 16.8 | 0.18 |
| polarC18 | 758.568 | 400.2797 | 16.8 | 0.18 |
| polarC18 | 758.568 | 210.3046 | 16.8 | 0.18 |
| polarC18 | 758.568 | 226.7627 | 16.8 | 0.18 |
| polarC18 | 758.568 | 564.5897 | 16.8 | 0.18 |
| polarC18 | 758.568 | 215.2814 | 16.8 | 0.18 |
| polarC18 | 758.568 | 494.2836 | 16.8 | 0.18 |
| polarC18 | 758.568 | 210.8103 | 16.8 | 0.18 |
| polarC18 | 758.568 | 434.893 | 16.8 | 0.18 |
| polarC18 | 758.568 | 150.2801 | 16.8 | 0.18 |
| polarC18 | 758.568 | 615.7186 | 16.8 | 0.18 |

| | | | | |
|---|---|---|---|---|
| polarC18 | 758.568 | 168.1039 | 16.8 | 0.18 |
| polarC18 | 758.568 | 624.8665 | 16.8 | 0.18 |
| polarC18 | 758.568 | 264.9348 | 16.8 | 0.18 |
| polarC18 | 758.568 | 193.6398 | 16.8 | 0.18 |
| polarC18 | 758.568 | 645.639 | 16.8 | 0.18 |
| polarC18 | 758.568 | 540.1763 | 16.8 | 0.18 |
| polarC18 | 758.568 | 673.6717 | 16.8 | 0.18 |
| polarC18 | 758.568 | 156.4203 | 16.8 | 0.18 |
| polarC18 | 758.568 | 544.6158 | 16.8 | 0.18 |
| polarC18 | 758.568 | 443.6449 | 16.8 | 0.18 |
| polarC18 | 758.568 | 238.135 | 16.7 | 0.18 |
| polarC18 | 758.568 | 90.7845 | 16.7 | 0.18 |
| polarC18 | 758.568 | 87.2361 | 16.7 | 0.18 |
| polarC18 | 758.568 | 419.2776 | 16.7 | 0.18 |
| polarC18 | 758.568 | 509.028 | 16.7 | 0.18 |
| polarC18 | 758.568 | 94.7311 | 16.7 | 0.18 |
| polarC18 | 758.568 | 604.4677 | 16.7 | 0.18 |
| polarC18 | 758.568 | 462.9664 | 16.7 | 0.18 |
| polarC18 | 758.568 | 148.2166 | 16.7 | 0.18 |
| polarC18 | 758.568 | 301.0864 | 16.7 | 0.18 |
| polarC18 | 758.568 | 274.2167 | 16.7 | 0.18 |
| polarC18 | 758.568 | 132.0098 | 16.7 | 0.18 |
| polarC18 | 758.568 | 286.5435 | 16.7 | 0.18 |
| polarC18 | 758.568 | 278.1295 | 16.7 | 0.18 |
| polarC18 | 758.568 | 90.0653 | 16.7 | 0.18 |
| polarC18 | 758.568 | 98.4331 | 16.7 | 0.18 |
| polarC18 | 758.568 | 102.2825 | 16.7 | 0.18 |
| polarC18 | 758.568 | 130.1786 | 16.7 | 0.18 |
| polarC18 | 758.568 | 227.9483 | 16.7 | 0.18 |
| polarC18 | 758.568 | 241.2342 | 16.7 | 0.18 |
| polarC18 | 758.568 | 260.0595 | 16.7 | 0.18 |
| polarC18 | 758.568 | 114.9272 | 16.7 | 0.18 |
| polarC18 | 758.568 | 113.7707 | 16.7 | 0.18 |
| polarC18 | 758.568 | 120.135 | 16.7 | 0.18 |
| polarC18 | 758.568 | 255.4157 | 16.7 | 0.18 |
| polarC18 | 758.568 | 83.3628 | 16.7 | 0.18 |
| polarC18 | 758.568 | 279.1398 | 16.7 | 0.18 |
| polarC18 | 758.568 | 479.8881 | 16.7 | 0.18 |
| polarC18 | 758.568 | 506.9742 | 16.7 | 0.18 |
| polarC18 | 758.568 | 495.723 | 16.7 | 0.18 |
| polarC18 | 758.568 | 88.0056 | 16.7 | 0.18 |
| polarC18 | 758.568 | 101.4972 | 16.7 | 0.18 |

| | | | | |
|---|---|---|---|---|
| polarC18 | 758.568 | 197.8253 | 16.7 | 0.18 |
| polarC18 | 758.568 | 200.6363 | 16.7 | 0.18 |
| polarC18 | 758.568 | 293.072 | 16.7 | 0.18 |
| polarC18 | 758.568 | 381.0126 | 16.7 | 0.18 |
| polarC18 | 758.568 | 234.6401 | 16.7 | 0.18 |
| polarC18 | 758.568 | 90.9061 | 16.7 | 0.18 |
| polarC18 | 758.568 | 150.7469 | 16.7 | 0.18 |
| polarC18 | 758.568 | 235.7926 | 16.7 | 0.18 |
| polarC18 | 758.568 | 85.1297 | 16.7 | 0.18 |
| polarC18 | 758.568 | 190.5177 | 16.7 | 0.18 |
| polarC18 | 758.568 | 490.6508 | 16.7 | 0.18 |
| polarC18 | 758.568 | 165.8826 | 16.7 | 0.18 |
| polarC18 | 758.568 | 82.1576 | 16.7 | 0.18 |
| polarC18 | 758.568 | 106.531 | 16.7 | 0.18 |
| polarC18 | 758.568 | 152.5107 | 16.7 | 0.18 |
| polarC18 | 758.568 | 108.9573 | 16.6 | 0.18 |
| polarC18 | 758.568 | 109.2567 | 16.6 | 0.18 |
| polarC18 | 758.568 | 220.1315 | 16.6 | 0.18 |
| polarC18 | 758.568 | 124.7854 | 16.6 | 0.18 |
| polarC18 | 758.568 | 345.7318 | 16.6 | 0.18 |
| polarC18 | 758.568 | 83.7101 | 16.6 | 0.18 |
| polarC18 | 758.568 | 217.4502 | 16.6 | 0.18 |
| polarC18 | 758.568 | 782.9918 | 16.6 | 0.18 |
| polarC18 | 758.568 | 490.9361 | 16.6 | 0.18 |
| polarC18 | 758.568 | 116.5895 | 16.6 | 0.18 |
| polarC18 | 758.568 | 337.6745 | 16.6 | 0.18 |
| polarC18 | 758.568 | 237.4368 | 16.6 | 0.18 |
| polarC18 | 758.568 | 730.3303 | 16.6 | 0.18 |
| polarC18 | 758.568 | 83.5743 | 16.6 | 0.18 |
| polarC18 | 758.568 | 292.2795 | 16.6 | 0.18 |
| polarC18 | 758.568 | 86.0392 | 16.6 | 0.18 |
| polarC18 | 758.568 | 226.1407 | 16.6 | 0.18 |
| polarC18 | 758.568 | 578.384 | 16.6 | 0.18 |
| polarC18 | 758.568 | 626.9479 | 16.6 | 0.18 |
| polarC18 | 758.568 | 93.344 | 16.6 | 0.18 |
| polarC18 | 758.568 | 271.6396 | 16.6 | 0.18 |
| polarC18 | 758.568 | 282.04 | 16.6 | 0.18 |
| polarC18 | 758.568 | 136.5529 | 16.6 | 0.18 |
| polarC18 | 758.568 | 146.3299 | 16.6 | 0.18 |
| polarC18 | 758.568 | 137.3717 | 16.6 | 0.18 |
| polarC18 | 758.568 | 144.5992 | 16.6 | 0.18 |
| polarC18 | 758.568 | 91.6013 | 16.6 | 0.18 |

| | | | | |
|---|---|---|---|---|
| polarC18 | 758.568 | 154.332 | 16.6 | 0.18 |
| polarC18 | 758.568 | 291.6699 | 16.6 | 0.18 |
| polarC18 | 758.568 | 172.8949 | 16.6 | 0.18 |
| polarC18 | 758.568 | 146.7413 | 16.6 | 0.18 |
| polarC18 | 758.568 | 159.1103 | 16.6 | 0.18 |
| polarC18 | 758.568 | 112.7151 | 16.6 | 0.18 |
| polarC18 | 758.568 | 181.1916 | 16.6 | 0.18 |
| polarC18 | 758.568 | 81.7048 | 16.6 | 0.18 |
| polarC18 | 758.568 | 328.6807 | 16.6 | 0.18 |
| polarC18 | 758.568 | 699.1277 | 16.6 | 0.18 |
| polarC18 | 758.568 | 82.0094 | 16.6 | 0.18 |
| polarC18 | 758.568 | 405.071 | 16.6 | 0.18 |
| polarC18 | 758.568 | 120.2395 | 16.6 | 0.18 |
| polarC18 | 758.568 | 123.3654 | 16.6 | 0.18 |
| polarC18 | 758.568 | 128.844 | 16.6 | 0.18 |
| polarC18 | 758.568 | 87.3876 | 16.6 | 0.18 |
| polarC18 | 758.568 | 137.0278 | 16.6 | 0.18 |
| polarC18 | 758.568 | 150.1091 | 16.6 | 0.18 |
| polarC18 | 758.568 | 120.0347 | 16.6 | 0.18 |
| polarC18 | 758.568 | 197.7546 | 16.6 | 0.18 |
| polarC18 | 758.568 | 327.5693 | 16.6 | 0.18 |
| polarC18 | 758.568 | 97.919 | 16.6 | 0.18 |
| polarC18 | 758.568 | 371.1664 | 16.6 | 0.18 |
| polarC18 | 758.568 | 183.6902 | 16.6 | 0.18 |
| polarC18 | 758.568 | 498.2094 | 16.6 | 0.18 |
| polarC18 | 758.568 | 184.6252 | 16.6 | 0.18 |
| polarC18 | 758.568 | 207.4767 | 16.6 | 0.18 |
| polarC18 | 758.568 | 84.097 | 16.6 | 0.18 |
| polarC18 | 758.568 | 129.197 | 16.6 | 0.18 |
| polarC18 | 758.568 | 214.1668 | 16.6 | 0.18 |
| polarC18 | 758.568 | 630.106 | 16.6 | 0.18 |
| polarC18 | 758.568 | 80.1717 | 16.6 | 0.18 |
| polarC18 | 758.568 | 115.9696 | 16.6 | 0.18 |
| polarC18 | 758.568 | 741.9062 | 16.6 | 0.18 |
| polarC18 | 758.568 | 408.6965 | 16.6 | 0.18 |
| polarC18 | 758.568 | 154.8381 | 16.6 | 0.18 |
| polarC18 | 758.568 | 184.2275 | 16.6 | 0.18 |
| polarC18 | 758.568 | 476.9739 | 16.6 | 0.18 |
| polarC18 | 758.568 | 363.86 | 16.6 | 0.18 |
| polarC18 | 758.568 | 98.2099 | 16.6 | 0.18 |
| polarC18 | 758.568 | 485.8697 | 16.6 | 0.18 |
| polarC18 | 758.568 | 220.4177 | 16.6 | 0.18 |

| | | | | |
|---|---|---|---|---|
| polarC18 | 758.568 | 142.26 | 16.5 | 0.18 |
| polarC18 | 758.568 | 173.1918 | 16.5 | 0.18 |
| polarC18 | 758.568 | 129.2315 | 16.5 | 0.18 |
| polarC18 | 758.568 | 323.2719 | 16.5 | 0.18 |
| polarC18 | 758.568 | 101.0784 | 16.5 | 0.18 |
| polarC18 | 758.568 | 82.4677 | 16.5 | 0.18 |
| polarC18 | 758.568 | 138.1364 | 16.5 | 0.18 |
| polarC18 | 758.568 | 576.4243 | 16.5 | 0.18 |
| polarC18 | 758.568 | 107.3652 | 16.5 | 0.18 |
| polarC18 | 758.568 | 369.2793 | 16.5 | 0.18 |
| polarC18 | 758.568 | 399.7409 | 16.5 | 0.18 |
| polarC18 | 758.568 | 163.269 | 16.5 | 0.18 |
| polarC18 | 758.568 | 684.9195 | 16.5 | 0.18 |
| polarC18 | 758.568 | 247.8986 | 16.5 | 0.18 |
| polarC18 | 758.568 | 201.1297 | 16.5 | 0.18 |
| polarC18 | 758.568 | 131.1161 | 16.5 | 0.18 |
| polarC18 | 758.568 | 88.5677 | 16.5 | 0.18 |
| polarC18 | 758.568 | 325.4414 | 16.5 | 0.18 |
| polarC18 | 758.568 | 397.8956 | 16.5 | 0.18 |
| polarC18 | 758.568 | 84.8634 | 16.5 | 0.18 |
| polarC18 | 758.568 | 361.7051 | 16.5 | 0.18 |
| polarC18 | 758.568 | 132.2663 | 16.5 | 0.18 |
| polarC18 | 758.568 | 86.3698 | 16.5 | 0.18 |
| polarC18 | 758.568 | 125.119 | 16.5 | 0.18 |
| polarC18 | 758.568 | 80.4222 | 16.5 | 0.18 |
| polarC18 | 758.568 | 93.5148 | 16.5 | 0.18 |
| polarC18 | 758.568 | 538.0924 | 16.5 | 0.18 |
| polarC18 | 758.568 | 87.3142 | 16.5 | 0.18 |
| polarC18 | 758.568 | 273.7762 | 16.5 | 0.18 |
| polarC18 | 758.568 | 188.1222 | 16.5 | 0.18 |
| polarC18 | 758.568 | 236.2776 | 16.5 | 0.18 |
| polarC18 | 758.568 | 575.1078 | 16.5 | 0.18 |
| polarC18 | 758.568 | 162.9862 | 16.5 | 0.18 |
| polarC18 | 758.568 | 80.6841 | 16.5 | 0.18 |
| polarC18 | 758.568 | 229.8702 | 16.5 | 0.18 |
| polarC18 | 758.568 | 130.0525 | 16.5 | 0.18 |
| polarC18 | 758.568 | 660.6063 | 16.5 | 0.18 |
| polarC18 | 758.568 | 93.8016 | 16.5 | 0.18 |
| polarC18 | 758.568 | 111.0814 | 16.5 | 0.18 |
| polarC18 | 758.568 | 288.6997 | 16.5 | 0.18 |
| polarC18 | 758.568 | 306.0778 | 16.5 | 0.18 |
| polarC18 | 758.568 | 428.5591 | 16.5 | 0.18 |

| | | | | |
|---|---|---|---|---|
| polarC18 | 758.568 | 143.7798 | 16.5 | 0.18 |
| polarC18 | 758.568 | 224.6105 | 16.5 | 0.18 |
| polarC18 | 758.568 | 161.0295 | 16.5 | 0.18 |
| polarC18 | 758.568 | 88.6286 | 16.5 | 0.18 |
| polarC18 | 758.568 | 171.0123 | 16.5 | 0.18 |
| polarC18 | 758.568 | 390.8053 | 16.5 | 0.18 |
| polarC18 | 758.568 | 339.5308 | 16.5 | 0.18 |
| polarC18 | 758.568 | 96.9879 | 16.5 | 0.18 |
| polarC18 | 758.568 | 379.9254 | 16.5 | 0.18 |
| polarC18 | 758.568 | 133.5655 | 16.5 | 0.18 |
| polarC18 | 758.568 | 115.0252 | 16.5 | 0.18 |
| polarC18 | 758.568 | 202.548 | 16.5 | 0.18 |
| polarC18 | 758.568 | 256.5579 | 16.5 | 0.18 |
| polarC18 | 758.568 | 134.976 | 16.5 | 0.18 |
| polarC18 | 758.568 | 91.2248 | 16.5 | 0.18 |
| polarC18 | 758.568 | 322.665 | 16.5 | 0.18 |
| polarC18 | 758.568 | 232.4977 | 16.5 | 0.18 |
| polarC18 | 758.568 | 328.9197 | 16.5 | 0.18 |
| polarC18 | 758.568 | 214.5151 | 16.5 | 0.18 |
| polarC18 | 758.568 | 88.6892 | 16.5 | 0.18 |
| polarC18 | 758.568 | 672.9537 | 16.4 | 0.18 |
| polarC18 | 758.568 | 127.0058 | 16.4 | 0.18 |
| polarC18 | 758.568 | 417.2971 | 16.4 | 0.18 |
| polarC18 | 758.568 | 207.9954 | 16.4 | 0.18 |
| polarC18 | 758.568 | 190.1005 | 16.4 | 0.18 |
| polarC18 | 758.568 | 111.7833 | 16.4 | 0.18 |
| polarC18 | 758.568 | 90.3058 | 16.4 | 0.18 |
| polarC18 | 758.568 | 119.8408 | 16.4 | 0.18 |
| polarC18 | 758.568 | 112.8848 | 16.4 | 0.18 |
| polarC18 | 758.568 | 378.122 | 16.4 | 0.18 |
| polarC18 | 758.568 | 136.9558 | 16.4 | 0.18 |
| polarC18 | 758.568 | 690.742 | 16.4 | 0.18 |
| polarC18 | 758.568 | 269.1624 | 16.4 | 0.18 |
| polarC18 | 758.568 | 99.878 | 16.4 | 0.18 |
| polarC18 | 758.568 | 104.5588 | 16.4 | 0.18 |
| polarC18 | 758.568 | 297.2403 | 16.4 | 0.18 |
| polarC18 | 758.568 | 85.3171 | 16.4 | 0.18 |
| polarC18 | 758.568 | 94.6969 | 16.4 | 0.18 |
| polarC18 | 758.568 | 124.921 | 16.4 | 0.18 |
| polarC18 | 758.568 | 136.6492 | 16.4 | 0.18 |
| polarC18 | 758.568 | 206.6643 | 16.4 | 0.18 |
| polarC18 | 758.568 | 147.2123 | 16.4 | 0.18 |

| | | | | |
|---|---|---|---|---|
| polarC18 | 758.568 | 199.6552 | 16.4 | 0.18 |
| polarC18 | 758.568 | 306.6562 | 16.4 | 0.18 |
| polarC18 | 758.568 | 221.9595 | 16.4 | 0.18 |
| polarC18 | 758.568 | 143.5868 | 16.4 | 0.18 |
| polarC18 | 758.568 | 725.1391 | 16.4 | 0.18 |
| polarC18 | 758.568 | 130.1283 | 16.4 | 0.18 |
| polarC18 | 758.568 | 406.159 | 16.4 | 0.18 |
| polarC18 | 758.568 | 401.9918 | 16.4 | 0.18 |
| polarC18 | 758.568 | 92.1566 | 16.4 | 0.18 |
| polarC18 | 758.568 | 151.4388 | 16.4 | 0.18 |
| polarC18 | 758.568 | 153.5157 | 16.4 | 0.18 |
| polarC18 | 758.568 | 123.1075 | 16.4 | 0.18 |
| polarC18 | 758.568 | 334.4797 | 16.4 | 0.18 |
| polarC18 | 758.568 | 102.555 | 16.4 | 0.18 |
| polarC18 | 758.568 | 151.7632 | 16.4 | 0.18 |
| polarC18 | 758.568 | 208.0637 | 16.4 | 0.18 |
| polarC18 | 758.568 | 215.7843 | 16.4 | 0.18 |
| polarC18 | 758.568 | 160.3371 | 16.4 | 0.18 |
| polarC18 | 758.568 | 209.9572 | 16.4 | 0.18 |
| polarC18 | 758.568 | 91.5252 | 16.4 | 0.18 |
| polarC18 | 758.568 | 672.5037 | 16.4 | 0.18 |
| polarC18 | 758.568 | 93.9692 | 16.4 | 0.18 |
| polarC18 | 758.568 | 86.632 | 16.4 | 0.18 |
| polarC18 | 758.568 | 209.2744 | 16.4 | 0.18 |
| polarC18 | 758.568 | 80.5799 | 16.4 | 0.18 |
| polarC18 | 758.568 | 225.687 | 16.4 | 0.18 |
| polarC18 | 758.568 | 102.3166 | 16.4 | 0.18 |
| polarC18 | 758.568 | 81.4123 | 16.4 | 0.18 |
| polarC18 | 758.568 | 197.2114 | 16.4 | 0.18 |
| polarC18 | 758.568 | 120.257 | 16.4 | 0.18 |
| polarC18 | 758.568 | 118.9949 | 16.4 | 0.18 |
| polarC18 | 758.568 | 420.2799 | 16.4 | 0.18 |
| polarC18 | 758.568 | 361.9857 | 16.4 | 0.18 |
| polarC18 | 758.568 | 344.3708 | 16.4 | 0.18 |
| polarC18 | 758.568 | 552.3137 | 16.4 | 0.18 |
| polarC18 | 758.568 | 106.4113 | 16.4 | 0.18 |
| polarC18 | 758.568 | 179.308 | 16.4 | 0.18 |
| polarC18 | 758.568 | 470.2408 | 16.4 | 0.18 |
| polarC18 | 758.568 | 111.726 | 16.4 | 0.18 |
| polarC18 | 758.568 | 159.5399 | 16.4 | 0.18 |
| polarC18 | 758.568 | 100.004 | 16.4 | 0.18 |
| polarC18 | 758.568 | 104.3045 | 16.4 | 0.18 |

| | | | | |
|---|---|---|---|---|
| polarC18 | 758.568 | 380.8889 | 16.4 | 0.18 |
| polarC18 | 758.568 | 387.1744 | 16.4 | 0.18 |
| polarC18 | 758.568 | 463.2584 | 16.4 | 0.18 |
| polarC18 | 758.568 | 697.1523 | 16.4 | 0.18 |
| polarC18 | 758.568 | 373.1479 | 16.4 | 0.18 |
| polarC18 | 758.568 | 226.1917 | 16.3 | 0.18 |
| polarC18 | 758.568 | 99.3258 | 16.3 | 0.18 |
| polarC18 | 758.568 | 511.1294 | 16.3 | 0.18 |
| polarC18 | 758.568 | 145.4779 | 16.3 | 0.18 |
| polarC18 | 758.568 | 85.4948 | 16.3 | 0.18 |
| polarC18 | 758.568 | 232.9285 | 16.3 | 0.18 |
| polarC18 | 758.568 | 98.597 | 16.3 | 0.18 |
| polarC18 | 758.568 | 633.7086 | 16.3 | 0.18 |
| polarC18 | 758.568 | 244.5159 | 16.3 | 0.18 |
| polarC18 | 758.568 | 331.2868 | 16.3 | 0.18 |
| polarC18 | 758.568 | 164.9044 | 16.3 | 0.18 |
| polarC18 | 758.568 | 240.6521 | 16.3 | 0.18 |
| polarC18 | 758.568 | 588.6008 | 16.3 | 0.18 |
| polarC18 | 758.568 | 80.1242 | 16.3 | 0.18 |
| polarC18 | 758.568 | 599.6228 | 16.3 | 0.18 |
| polarC18 | 758.568 | 115.0079 | 16.3 | 0.18 |
| polarC18 | 758.568 | 95.8526 | 16.3 | 0.18 |
| polarC18 | 758.568 | 161.5462 | 16.3 | 0.18 |
| polarC18 | 758.568 | 206.1286 | 16.3 | 0.18 |
| polarC18 | 758.568 | 137.8278 | 16.3 | 0.18 |
| polarC18 | 758.568 | 305.4421 | 16.3 | 0.18 |
| polarC18 | 758.568 | 166.8104 | 16.3 | 0.18 |
| polarC18 | 758.568 | 178.3343 | 16.3 | 0.18 |
| polarC18 | 758.568 | 98.0448 | 16.3 | 0.18 |
| polarC18 | 758.568 | 165.6928 | 16.3 | 0.18 |
| polarC18 | 758.568 | 230.0498 | 16.3 | 0.18 |
| polarC18 | 758.568 | 539.4567 | 16.3 | 0.18 |
| polarC18 | 758.568 | 190.6775 | 16.3 | 0.18 |
| polarC18 | 758.568 | 305.5931 | 16.3 | 0.18 |
| polarC18 | 758.568 | 95.944 | 16.3 | 0.18 |
| polarC18 | 758.568 | 204.4671 | 16.3 | 0.18 |
| polarC18 | 758.568 | 279.1882 | 16.3 | 0.18 |
| polarC18 | 758.568 | 396.4758 | 16.3 | 0.18 |
| polarC18 | 758.568 | 135.4391 | 16.3 | 0.18 |
| polarC18 | 758.568 | 109.538 | 16.3 | 0.17 |
| polarC18 | 758.568 | 580.3633 | 16.3 | 0.17 |
| polarC18 | 758.568 | 160.6787 | 16.3 | 0.17 |

| | | | | |
|---|---|---|---|---|
| polarC18 | 758.568 | 421.3416 | 16.3 | 0.17 |
| polarC18 | 758.568 | 189.7155 | 16.3 | 0.17 |
| polarC18 | 758.568 | 256.9528 | 16.3 | 0.17 |
| polarC18 | 758.568 | 130.3464 | 16.3 | 0.17 |
| polarC18 | 758.568 | 102.1815 | 16.3 | 0.17 |
| polarC18 | 758.568 | 234.9348 | 16.3 | 0.17 |
| polarC18 | 758.568 | 105.452 | 16.3 | 0.17 |
| polarC18 | 758.568 | 262.8275 | 16.3 | 0.17 |
| polarC18 | 758.568 | 168.9996 | 16.3 | 0.17 |
| polarC18 | 758.568 | 136.4199 | 16.3 | 0.17 |
| polarC18 | 758.568 | 143.6946 | 16.3 | 0.17 |
| polarC18 | 758.568 | 333.7448 | 16.3 | 0.17 |
| polarC18 | 758.568 | 212.9174 | 16.3 | 0.17 |
| polarC18 | 758.568 | 295.7016 | 16.3 | 0.17 |
| polarC18 | 758.568 | 588.845 | 16.3 | 0.17 |
| polarC18 | 758.568 | 81.7713 | 16.3 | 0.17 |
| polarC18 | 758.568 | 510.2098 | 16.3 | 0.17 |
| polarC18 | 758.568 | 276.7066 | 16.2 | 0.17 |
| polarC18 | 758.568 | 705.5109 | 16.2 | 0.17 |
| polarC18 | 758.568 | 88.4545 | 16.2 | 0.17 |
| polarC18 | 758.568 | 91.6516 | 16.2 | 0.17 |
| polarC18 | 758.568 | 219.8702 | 16.2 | 0.17 |
| polarC18 | 758.568 | 90.1509 | 16.2 | 0.17 |
| polarC18 | 758.568 | 168.5666 | 16.2 | 0.17 |
| polarC18 | 758.568 | 168.4331 | 16.2 | 0.17 |
| polarC18 | 758.568 | 92.6924 | 16.2 | 0.17 |
| polarC18 | 758.568 | 101.4812 | 16.2 | 0.17 |
| polarC18 | 758.568 | 349.1164 | 16.2 | 0.17 |
| polarC18 | 758.568 | 166.6418 | 16.2 | 0.17 |
| polarC18 | 758.568 | 398.0823 | 16.2 | 0.17 |
| polarC18 | 758.568 | 120.2507 | 16.2 | 0.17 |
| polarC18 | 758.568 | 201.366 | 16.2 | 0.17 |
| polarC18 | 758.568 | 119.6866 | 16.2 | 0.17 |
| polarC18 | 758.568 | 113.6424 | 16.2 | 0.17 |
| polarC18 | 758.568 | 91.8206 | 16.2 | 0.17 |
| polarC18 | 758.568 | 500.949 | 16.2 | 0.17 |
| polarC18 | 758.568 | 723.3238 | 16.2 | 0.17 |
| polarC18 | 758.568 | 130.5874 | 16.2 | 0.17 |
| polarC18 | 758.568 | 96.4857 | 16.2 | 0.17 |
| polarC18 | 758.568 | 477.6465 | 16.2 | 0.17 |
| polarC18 | 758.568 | 229.5477 | 16.2 | 0.17 |
| polarC18 | 758.568 | 482.5154 | 16.2 | 0.17 |

| | | | | |
|---|---|---|---|---|
| polarC18 | 758.568 | 140.2395 | 16.2 | 0.17 |
| polarC18 | 758.568 | 716.2006 | 16.2 | 0.17 |
| polarC18 | 758.568 | 179.6447 | 16.2 | 0.17 |
| polarC18 | 758.568 | 93.6352 | 16.2 | 0.17 |
| polarC18 | 758.568 | 115.6714 | 16.2 | 0.17 |
| polarC18 | 758.568 | 272.2058 | 16.2 | 0.17 |
| polarC18 | 758.568 | 278.5459 | 16.2 | 0.17 |
| polarC18 | 758.568 | 257.0697 | 16.2 | 0.17 |
| polarC18 | 758.568 | 625.2034 | 16.2 | 0.17 |
| polarC18 | 758.568 | 104.6494 | 16.2 | 0.17 |
| polarC18 | 758.568 | 228.8239 | 16.2 | 0.17 |
| polarC18 | 758.568 | 144.7698 | 16.2 | 0.17 |
| polarC18 | 758.568 | 151.1978 | 16.2 | 0.17 |
| polarC18 | 758.568 | 118.7546 | 16.2 | 0.17 |
| polarC18 | 758.568 | 612.0649 | 16.2 | 0.17 |
| polarC18 | 758.568 | 90.5605 | 16.2 | 0.17 |
| polarC18 | 758.568 | 425.1439 | 16.2 | 0.17 |
| polarC18 | 758.568 | 273.9221 | 16.2 | 0.17 |
| polarC18 | 758.568 | 394.6559 | 16.2 | 0.17 |
| polarC18 | 758.568 | 88.7741 | 16.2 | 0.17 |
| polarC18 | 758.568 | 186.4915 | 16.2 | 0.17 |
| polarC18 | 758.568 | 81.5693 | 16.2 | 0.17 |
| polarC18 | 758.568 | 90.2058 | 16.2 | 0.17 |
| polarC18 | 758.568 | 95.3468 | 16.2 | 0.17 |
| polarC18 | 758.568 | 86.8395 | 16.2 | 0.17 |
| polarC18 | 758.568 | 370.5579 | 16.2 | 0.17 |
| polarC18 | 758.568 | 169.7802 | 16.2 | 0.17 |
| polarC18 | 758.568 | 109.6687 | 16.2 | 0.17 |
| polarC18 | 758.568 | 296.9433 | 16.2 | 0.17 |
| polarC18 | 758.568 | 114.7115 | 16.2 | 0.17 |
| polarC18 | 758.568 | 265.8221 | 16.2 | 0.17 |
| polarC18 | 758.568 | 202.0085 | 16.2 | 0.17 |
| polarC18 | 758.568 | 86.1998 | 16.2 | 0.17 |
| polarC18 | 758.568 | 255.5423 | 16.2 | 0.17 |
| polarC18 | 758.568 | 225.8781 | 16.2 | 0.17 |
| polarC18 | 758.568 | 506.4563 | 16.2 | 0.17 |
| polarC18 | 758.568 | 784.4114 | 16.2 | 0.17 |
| polarC18 | 758.568 | 219.0775 | 16.2 | 0.17 |
| polarC18 | 758.568 | 83.3779 | 16.2 | 0.17 |
| polarC18 | 758.568 | 122.1113 | 16.2 | 0.17 |
| polarC18 | 758.568 | 319.6586 | 16.2 | 0.17 |
| polarC18 | 758.568 | 622.5983 | 16.2 | 0.17 |

| | | | | |
|---|---|---|---|---|
| polarC18 | 758.568 | 164.0982 | 16.2 | 0.17 |
| polarC18 | 758.568 | 102.6412 | 16.2 | 0.17 |
| polarC18 | 758.568 | 83.2524 | 16.2 | 0.17 |
| polarC18 | 758.568 | 182.2289 | 16.1 | 0.17 |
| polarC18 | 758.568 | 380.3806 | 16.1 | 0.17 |
| polarC18 | 758.568 | 119.2029 | 16.1 | 0.17 |
| polarC18 | 758.568 | 159.9757 | 16.1 | 0.17 |
| polarC18 | 758.568 | 234.3168 | 16.1 | 0.17 |
| polarC18 | 758.568 | 104.6065 | 16.1 | 0.17 |
| polarC18 | 758.568 | 132.3903 | 16.1 | 0.17 |
| polarC18 | 758.568 | 84.2959 | 16.1 | 0.17 |
| polarC18 | 758.568 | 211.928 | 16.1 | 0.17 |
| polarC18 | 758.568 | 385.1118 | 16.1 | 0.17 |
| polarC18 | 758.568 | 293.6662 | 16.1 | 0.17 |
| polarC18 | 758.568 | 125.93 | 16.1 | 0.17 |
| polarC18 | 758.568 | 123.2146 | 16.1 | 0.17 |
| polarC18 | 758.568 | 260.0866 | 16.1 | 0.17 |
| polarC18 | 758.568 | 130.277 | 16.1 | 0.17 |
| polarC18 | 758.568 | 133.0714 | 16.1 | 0.17 |
| polarC18 | 758.568 | 116.0503 | 16.1 | 0.17 |
| polarC18 | 758.568 | 93.8934 | 16.1 | 0.17 |
| polarC18 | 758.568 | 93.7363 | 16.1 | 0.17 |
| polarC18 | 758.568 | 181.3513 | 16.1 | 0.17 |
| polarC18 | 758.568 | 440.3783 | 16.1 | 0.17 |
| polarC18 | 758.568 | 438.4762 | 16.1 | 0.17 |
| polarC18 | 758.568 | 286.3879 | 16.1 | 0.17 |
| polarC18 | 758.568 | 139.6533 | 16.1 | 0.17 |
| polarC18 | 758.568 | 113.7457 | 16.1 | 0.17 |
| polarC18 | 758.568 | 104.3888 | 16.1 | 0.17 |
| polarC18 | 758.568 | 215.2166 | 16.1 | 0.17 |
| polarC18 | 758.568 | 745.785 | 16.1 | 0.17 |
| polarC18 | 758.568 | 186.7621 | 16.1 | 0.17 |
| polarC18 | 758.568 | 222.3693 | 16.1 | 0.17 |
| polarC18 | 758.568 | 100.0898 | 16.1 | 0.17 |
| polarC18 | 758.568 | 208.4312 | 16.1 | 0.17 |
| polarC18 | 758.568 | 576.0682 | 16.1 | 0.17 |
| polarC18 | 758.568 | 162.021 | 16.1 | 0.17 |
| polarC18 | 758.568 | 624.399 | 16.1 | 0.17 |
| polarC18 | 758.568 | 623.844 | 16.1 | 0.17 |
| polarC18 | 758.568 | 612.5767 | 16.1 | 0.17 |
| polarC18 | 758.568 | 391.1408 | 16.1 | 0.17 |
| polarC18 | 758.568 | 87.5372 | 16.1 | 0.17 |

| | | | | |
|---|---|---|---|---|
| polarC18 | 758.568 | 231.6108 | 16.1 | 0.17 |
| polarC18 | 758.568 | 287.9932 | 16.1 | 0.17 |
| polarC18 | 758.568 | 754.5876 | 16.1 | 0.17 |
| polarC18 | 758.568 | 199.8517 | 16.1 | 0.17 |
| polarC18 | 758.568 | 285.4532 | 16.1 | 0.17 |
| polarC18 | 758.568 | 79.6836 | 16.1 | 0.17 |
| polarC18 | 758.568 | 213.2073 | 16.1 | 0.17 |
| polarC18 | 758.568 | 208.5642 | 16.1 | 0.17 |
| polarC18 | 758.568 | 108.0736 | 16.1 | 0.17 |
| polarC18 | 758.568 | 100.434 | 16.1 | 0.17 |
| polarC18 | 758.568 | 448.1104 | 16.1 | 0.17 |
| polarC18 | 758.568 | 165.8563 | 16.1 | 0.17 |
| polarC18 | 758.568 | 151.0897 | 16.1 | 0.17 |
| polarC18 | 758.568 | 101.8224 | 16.1 | 0.17 |
| polarC18 | 758.568 | 181.9105 | 16.1 | 0.17 |
| polarC18 | 758.568 | 185.1336 | 16.1 | 0.17 |
| polarC18 | 758.568 | 458.5983 | 16.1 | 0.17 |
| polarC18 | 758.568 | 89.6439 | 16.1 | 0.17 |
| polarC18 | 758.568 | 167.6086 | 16.1 | 0.17 |
| polarC18 | 758.568 | 517.0339 | 16.1 | 0.17 |
| polarC18 | 758.568 | 92.7922 | 16.1 | 0.17 |
| polarC18 | 758.568 | 142.3114 | 16.1 | 0.17 |
| polarC18 | 758.568 | 266.0426 | 16.1 | 0.17 |
| polarC18 | 758.568 | 81.1259 | 16.1 | 0.17 |
| polarC18 | 758.568 | 274.3847 | 16.1 | 0.17 |
| polarC18 | 758.568 | 322.1377 | 16.1 | 0.17 |
| polarC18 | 758.568 | 134.0183 | 16.1 | 0.17 |
| polarC18 | 758.568 | 138.5168 | 16.1 | 0.17 |
| polarC18 | 758.568 | 211.6236 | 16.1 | 0.17 |
| polarC18 | 758.568 | 305.9407 | 16.1 | 0.17 |
| polarC18 | 758.568 | 119.0976 | 16.1 | 0.17 |
| polarC18 | 758.568 | 583.4132 | 16.1 | 0.17 |
| polarC18 | 758.568 | 139.8594 | 16.1 | 0.17 |
| polarC18 | 758.568 | 562.2874 | 16.1 | 0.17 |
| polarC18 | 758.568 | 80.468 | 16.1 | 0.17 |
| polarC18 | 758.568 | 164.1736 | 16.1 | 0.17 |
| polarC18 | 758.568 | 384.2042 | 16.1 | 0.17 |
| polarC18 | 758.568 | 194.4841 | 16 | 0.17 |
| polarC18 | 758.568 | 256.0744 | 16 | 0.17 |
| polarC18 | 758.568 | 629.6114 | 16 | 0.17 |
| polarC18 | 758.568 | 149.9263 | 16 | 0.17 |
| polarC18 | 758.568 | 237.9022 | 16 | 0.17 |

| | | | | |
|---|---|---|---|---|
| polarC18 | 758.568 | 88.6223 | 16 | 0.17 |
| polarC18 | 758.568 | 184.4037 | 16 | 0.17 |
| polarC18 | 758.568 | 729.5812 | 16 | 0.17 |
| polarC18 | 758.568 | 304.3554 | 16 | 0.17 |
| polarC18 | 758.568 | 157.0017 | 16 | 0.17 |
| polarC18 | 758.568 | 537.2352 | 16 | 0.17 |
| polarC18 | 758.568 | 101.0619 | 16 | 0.17 |
| polarC18 | 758.568 | 615.0569 | 16 | 0.17 |
| polarC18 | 758.568 | 602.8405 | 16 | 0.17 |
| polarC18 | 758.568 | 108.1722 | 16 | 0.17 |
| polarC18 | 758.568 | 101.7511 | 16 | 0.17 |
| polarC18 | 758.568 | 543.636 | 16 | 0.17 |
| polarC18 | 758.568 | 115.9738 | 16 | 0.17 |
| polarC18 | 758.568 | 132.7604 | 16 | 0.17 |
| polarC18 | 758.568 | 259.4567 | 16 | 0.17 |
| polarC18 | 758.568 | 144.2972 | 16 | 0.17 |
| polarC18 | 758.568 | 88.4699 | 16 | 0.17 |
| polarC18 | 758.568 | 322.0015 | 16 | 0.17 |
| polarC18 | 758.568 | 181.1521 | 16 | 0.17 |
| polarC18 | 758.568 | 90.4019 | 16 | 0.17 |
| polarC18 | 758.568 | 381.4258 | 16 | 0.17 |
| polarC18 | 758.568 | 218.6605 | 16 | 0.17 |
| polarC18 | 758.568 | 95.8568 | 16 | 0.17 |
| polarC18 | 758.568 | 162.9692 | 16 | 0.17 |
| polarC18 | 758.568 | 102.039 | 16 | 0.17 |
| polarC18 | 758.568 | 87.1704 | 16 | 0.17 |
| polarC18 | 758.568 | 770.9493 | 16 | 0.17 |
| polarC18 | 758.568 | 90.2365 | 16 | 0.17 |
| polarC18 | 758.568 | 285.7832 | 16 | 0.17 |
| polarC18 | 758.568 | 283.6269 | 16 | 0.17 |
| polarC18 | 758.568 | 218.3792 | 16 | 0.17 |
| polarC18 | 758.568 | 413.0042 | 16 | 0.17 |
| polarC18 | 758.568 | 212.048 | 16 | 0.17 |
| polarC18 | 758.568 | 115.4867 | 16 | 0.17 |
| polarC18 | 758.568 | 150.8368 | 16 | 0.17 |
| polarC18 | 758.568 | 329.6646 | 16 | 0.17 |
| polarC18 | 758.568 | 80.2872 | 16 | 0.17 |
| polarC18 | 758.568 | 243.0807 | 16 | 0.17 |
| polarC18 | 758.568 | 118.6523 | 16 | 0.17 |
| polarC18 | 758.568 | 493.1762 | 16 | 0.17 |
| polarC18 | 758.568 | 223.3703 | 16 | 0.17 |
| polarC18 | 758.568 | 329.7573 | 16 | 0.17 |

| | | | | |
|---|---|---|---|---|
| polarC18 | 758.568 | 127.0927 | 16 | 0.17 |
| polarC18 | 758.568 | 110.5306 | 16 | 0.17 |
| polarC18 | 758.568 | 162.5742 | 16 | 0.17 |
| polarC18 | 758.568 | 140.8197 | 16 | 0.17 |
| polarC18 | 758.568 | 84.1707 | 16 | 0.17 |
| polarC18 | 758.568 | 263.8628 | 16 | 0.17 |
| polarC18 | 758.568 | 634.2458 | 16 | 0.17 |
| polarC18 | 758.568 | 238.941 | 16 | 0.17 |
| polarC18 | 758.568 | 127.1405 | 16 | 0.17 |
| polarC18 | 758.568 | 599.0571 | 16 | 0.17 |
| polarC18 | 758.568 | 154.1798 | 16 | 0.17 |
| polarC18 | 758.568 | 382.6142 | 16 | 0.17 |
| polarC18 | 758.568 | 433.7323 | 16 | 0.17 |
| polarC18 | 758.568 | 83.4029 | 16 | 0.17 |
| polarC18 | 758.568 | 139.3147 | 16 | 0.17 |
| polarC18 | 758.568 | 344.5542 | 16 | 0.17 |
| polarC18 | 758.568 | 470.9976 | 16 | 0.17 |
| polarC18 | 758.568 | 378.9267 | 16 | 0.17 |
| polarC18 | 758.568 | 140.9175 | 16 | 0.17 |
| polarC18 | 758.568 | 143.6249 | 16 | 0.17 |
| polarC18 | 758.568 | 134.1994 | 16 | 0.17 |
| polarC18 | 758.568 | 98.1298 | 16 | 0.17 |
| polarC18 | 758.568 | 271.142 | 16 | 0.17 |
| polarC18 | 758.568 | 409.2087 | 16 | 0.17 |
| polarC18 | 758.568 | 129.0869 | 16 | 0.17 |
| polarC18 | 758.568 | 88.3018 | 16 | 0.17 |
| polarC18 | 758.568 | 109.8832 | 16 | 0.17 |
| polarC18 | 758.568 | 134.2438 | 16 | 0.17 |
| polarC18 | 758.568 | 197.5878 | 16 | 0.17 |
| polarC18 | 758.568 | 113.219 | 16 | 0.17 |
| polarC18 | 758.568 | 296.4715 | 16 | 0.17 |
| polarC18 | 758.568 | 97.0731 | 16 | 0.17 |
| polarC18 | 758.568 | 171.7529 | 16 | 0.17 |
| polarC18 | 758.568 | 473.5273 | 16 | 0.17 |
| polarC18 | 758.568 | 681.2936 | 15.9 | 0.17 |
| polarC18 | 758.568 | 107.1382 | 15.9 | 0.17 |
| polarC18 | 758.568 | 133.3643 | 15.9 | 0.17 |
| polarC18 | 758.568 | 685.452 | 15.9 | 0.17 |
| polarC18 | 758.568 | 203.5437 | 15.9 | 0.17 |
| polarC18 | 758.568 | 118.679 | 15.9 | 0.17 |
| polarC18 | 758.568 | 338.3855 | 15.9 | 0.17 |
| polarC18 | 758.568 | 139.0362 | 15.9 | 0.17 |

| | | | | |
|---|---|---|---|---|
| polarC18 | 758.568 | 683.4772 | 15.9 | 0.17 |
| polarC18 | 758.568 | 640.9343 | 15.9 | 0.17 |
| polarC18 | 758.568 | 320.3853 | 15.9 | 0.17 |
| polarC18 | 758.568 | 725.3549 | 15.9 | 0.17 |
| polarC18 | 758.568 | 81.4467 | 15.9 | 0.17 |
| polarC18 | 758.568 | 113.0187 | 15.9 | 0.17 |
| polarC18 | 758.568 | 164.0602 | 15.9 | 0.17 |
| polarC18 | 758.568 | 525.1158 | 15.9 | 0.17 |
| polarC18 | 758.568 | 356.2301 | 15.9 | 0.17 |
| polarC18 | 758.568 | 189.0681 | 15.9 | 0.17 |
| polarC18 | 758.568 | 82.9532 | 15.9 | 0.17 |
| polarC18 | 758.568 | 761.554 | 15.9 | 0.17 |
| polarC18 | 758.568 | 117.8683 | 15.9 | 0.17 |
| polarC18 | 758.568 | 138.351 | 15.9 | 0.17 |
| polarC18 | 758.568 | 369.7099 | 15.9 | 0.17 |
| polarC18 | 758.568 | 446.532 | 15.9 | 0.17 |
| polarC18 | 758.568 | 217.3936 | 15.9 | 0.17 |
| polarC18 | 758.568 | 429.609 | 15.9 | 0.17 |
| polarC18 | 758.568 | 172.9282 | 15.9 | 0.17 |
| polarC18 | 758.568 | 210.103 | 15.9 | 0.17 |
| polarC18 | 758.568 | 104.8216 | 15.9 | 0.17 |
| polarC18 | 758.568 | 97.461 | 15.9 | 0.17 |
| polarC18 | 758.568 | 548.3041 | 15.9 | 0.17 |
| polarC18 | 758.568 | 101.7053 | 15.9 | 0.17 |
| polarC18 | 758.568 | 234.4641 | 15.9 | 0.17 |
| polarC18 | 758.568 | 86.6853 | 15.9 | 0.17 |
| polarC18 | 758.568 | 446.7875 | 15.9 | 0.17 |
| polarC18 | 758.568 | 156.518 | 15.9 | 0.17 |
| polarC18 | 758.568 | 236.4305 | 15.9 | 0.17 |
| polarC18 | 758.568 | 410.6854 | 15.9 | 0.17 |
| polarC18 | 758.568 | 387.3861 | 15.9 | 0.17 |
| polarC18 | 758.568 | 666.3258 | 15.9 | 0.17 |
| polarC18 | 758.568 | 759.1656 | 15.9 | 0.17 |
| polarC18 | 758.568 | 116.8182 | 15.9 | 0.17 |
| polarC18 | 758.568 | 145.6719 | 15.9 | 0.17 |
| polarC18 | 758.568 | 204.2263 | 15.9 | 0.17 |
| polarC18 | 758.568 | 201.7169 | 15.9 | 0.17 |
| polarC18 | 758.568 | 258.0544 | 15.9 | 0.17 |
| polarC18 | 758.568 | 89.1103 | 15.9 | 0.17 |
| polarC18 | 758.568 | 108.6697 | 15.9 | 0.17 |
| polarC18 | 758.568 | 226.6403 | 15.9 | 0.17 |
| polarC18 | 758.568 | 222.2025 | 15.9 | 0.17 |

| | | | | |
|---|---|---|---|---|
| polarC18 | 758.568 | 384.5207 | 15.9 | 0.17 |
| polarC18 | 758.568 | 190.6062 | 15.9 | 0.17 |
| polarC18 | 758.568 | 175.098 | 15.9 | 0.17 |
| polarC18 | 758.568 | 390.3241 | 15.9 | 0.17 |
| polarC18 | 758.568 | 257.4859 | 15.9 | 0.17 |
| polarC18 | 758.568 | 153.5631 | 15.9 | 0.17 |
| polarC18 | 758.568 | 361.3801 | 15.9 | 0.17 |
| polarC18 | 758.568 | 750.8445 | 15.9 | 0.17 |
| polarC18 | 758.568 | 195.7077 | 15.9 | 0.17 |
| polarC18 | 758.568 | 236.5561 | 15.9 | 0.17 |
| polarC18 | 758.568 | 242.0535 | 15.9 | 0.17 |
| polarC18 | 758.568 | 112.6841 | 15.9 | 0.17 |
| polarC18 | 758.568 | 354.1103 | 15.9 | 0.17 |
| polarC18 | 758.568 | 777.1826 | 15.9 | 0.17 |
| polarC18 | 758.568 | 209.6104 | 15.9 | 0.17 |
| polarC18 | 758.568 | 92.1763 | 15.9 | 0.17 |
| polarC18 | 758.568 | 113.3219 | 15.9 | 0.17 |
| polarC18 | 758.568 | 88.8553 | 15.9 | 0.17 |
| polarC18 | 758.568 | 486.7089 | 15.9 | 0.17 |
| polarC18 | 758.568 | 91.8108 | 15.9 | 0.17 |
| polarC18 | 758.568 | 107.6547 | 15.9 | 0.17 |
| polarC18 | 758.568 | 99.7405 | 15.9 | 0.17 |
| polarC18 | 758.568 | 413.1846 | 15.9 | 0.17 |
| polarC18 | 758.568 | 90.5218 | 15.9 | 0.17 |
| polarC18 | 758.568 | 224.7269 | 15.9 | 0.17 |
| polarC18 | 758.568 | 88.7688 | 15.9 | 0.17 |
| polarC18 | 758.568 | 91.6845 | 15.9 | 0.17 |
| polarC18 | 758.568 | 691.8998 | 15.9 | 0.17 |
| polarC18 | 758.568 | 133.3288 | 15.9 | 0.17 |
| polarC18 | 758.568 | 270.634 | 15.9 | 0.17 |
| polarC18 | 758.568 | 372.303 | 15.9 | 0.17 |
| polarC18 | 758.568 | 161.8489 | 15.8 | 0.17 |
| polarC18 | 758.568 | 685.2437 | 15.8 | 0.17 |
| polarC18 | 758.568 | 484.4864 | 15.8 | 0.17 |
| polarC18 | 758.568 | 372.7676 | 15.8 | 0.17 |
| polarC18 | 758.568 | 319.8697 | 15.8 | 0.17 |
| polarC18 | 758.568 | 90.0742 | 15.8 | 0.17 |
| polarC18 | 758.568 | 129.4684 | 15.8 | 0.17 |
| polarC18 | 758.568 | 102.0959 | 15.8 | 0.17 |
| polarC18 | 758.568 | 126.5704 | 15.8 | 0.17 |
| polarC18 | 758.568 | 276.3491 | 15.8 | 0.17 |
| polarC18 | 758.568 | 133.514 | 15.8 | 0.17 |

| | | | | |
|---|---|---|---|---|
| polarC18 | 758.568 | 83.4675 | 15.8 | 0.17 |
| polarC18 | 758.568 | 389.4473 | 15.8 | 0.17 |
| polarC18 | 758.568 | 152.3628 | 15.8 | 0.17 |
| polarC18 | 758.568 | 102.8491 | 15.8 | 0.17 |
| polarC18 | 758.568 | 192.1002 | 15.8 | 0.17 |
| polarC18 | 758.568 | 108.2616 | 15.8 | 0.17 |
| polarC18 | 758.568 | 153.4998 | 15.8 | 0.17 |
| polarC18 | 758.568 | 447.7369 | 15.8 | 0.17 |
| polarC18 | 758.568 | 166.545 | 15.8 | 0.17 |
| polarC18 | 758.568 | 201.3209 | 15.8 | 0.17 |
| polarC18 | 758.568 | 135.0417 | 15.8 | 0.17 |
| polarC18 | 758.568 | 226.2153 | 15.8 | 0.17 |
| polarC18 | 758.568 | 94.1804 | 15.8 | 0.17 |
| polarC18 | 758.568 | 79.9699 | 15.8 | 0.17 |
| polarC18 | 758.568 | 235.6194 | 15.8 | 0.17 |
| polarC18 | 758.568 | 332.4102 | 15.8 | 0.17 |
| polarC18 | 758.568 | 400.6215 | 15.8 | 0.17 |
| polarC18 | 758.568 | 246.4199 | 15.8 | 0.17 |
| polarC18 | 758.568 | 116.442 | 15.8 | 0.17 |
| polarC18 | 758.568 | 278.3387 | 15.8 | 0.17 |
| polarC18 | 758.568 | 120.1275 | 15.8 | 0.17 |
| polarC18 | 758.568 | 141.6979 | 15.8 | 0.17 |
| polarC18 | 758.568 | 84.6683 | 15.8 | 0.17 |
| polarC18 | 758.568 | 426.1441 | 15.8 | 0.17 |
| polarC18 | 758.568 | 277.1327 | 15.8 | 0.17 |
| polarC18 | 758.568 | 95.9198 | 15.8 | 0.17 |
| polarC18 | 758.568 | 152.4906 | 15.8 | 0.17 |
| polarC18 | 758.568 | 252.5824 | 15.8 | 0.17 |
| polarC18 | 758.568 | 177.2617 | 15.8 | 0.17 |
| polarC18 | 758.568 | 232.5918 | 15.8 | 0.17 |
| polarC18 | 758.568 | 104.9977 | 15.8 | 0.17 |
| polarC18 | 758.568 | 167.3974 | 15.8 | 0.17 |
| polarC18 | 758.568 | 418.386 | 15.8 | 0.17 |
| polarC18 | 758.568 | 194.4057 | 15.8 | 0.17 |
| polarC18 | 758.568 | 789.6841 | 15.8 | 0.17 |
| polarC18 | 758.568 | 446.1914 | 15.8 | 0.17 |
| polarC18 | 758.568 | 82.0893 | 15.8 | 0.17 |
| polarC18 | 758.568 | 147.0983 | 15.8 | 0.17 |
| polarC18 | 758.568 | 84.8888 | 15.8 | 0.17 |
| polarC18 | 758.568 | 283.1721 | 15.8 | 0.17 |
| polarC18 | 758.568 | 180.6201 | 15.8 | 0.17 |
| polarC18 | 758.568 | 89.0188 | 15.8 | 0.17 |

| | | | | |
|---|---|---|---|---|
| polarC18 | 758.568 | 96.2468 | 15.8 | 0.17 |
| polarC18 | 758.568 | 93.2316 | 15.8 | 0.17 |
| polarC18 | 758.568 | 203.7644 | 15.8 | 0.17 |
| polarC18 | 758.568 | 174.2497 | 15.8 | 0.17 |
| polarC18 | 758.568 | 101.6831 | 15.8 | 0.17 |
| polarC18 | 758.568 | 231.2685 | 15.8 | 0.17 |
| polarC18 | 758.568 | 285.5642 | 15.8 | 0.17 |
| polarC18 | 758.568 | 362.8681 | 15.8 | 0.17 |
| polarC18 | 758.568 | 135.6104 | 15.8 | 0.17 |
| polarC18 | 758.568 | 182.5415 | 15.8 | 0.17 |
| polarC18 | 758.568 | 84.2548 | 15.8 | 0.17 |
| polarC18 | 758.568 | 124.7693 | 15.8 | 0.17 |
| polarC18 | 758.568 | 332.1571 | 15.8 | 0.17 |
| polarC18 | 758.568 | 393.0891 | 15.8 | 0.17 |
| polarC18 | 758.568 | 283.9069 | 15.8 | 0.17 |
| polarC18 | 758.568 | 88.8281 | 15.8 | 0.17 |
| polarC18 | 758.568 | 377.3898 | 15.8 | 0.17 |
| polarC18 | 758.568 | 97.9056 | 15.8 | 0.17 |
| polarC18 | 758.568 | 108.0307 | 15.8 | 0.17 |
| polarC18 | 758.568 | 136.0696 | 15.8 | 0.17 |
| polarC18 | 758.568 | 83.1047 | 15.8 | 0.17 |
| polarC18 | 758.568 | 114.757 | 15.8 | 0.17 |
| polarC18 | 758.568 | 267.362 | 15.8 | 0.17 |
| polarC18 | 758.568 | 80.7285 | 15.8 | 0.17 |
| polarC18 | 758.568 | 452.7905 | 15.8 | 0.17 |
| polarC18 | 758.568 | 357.2729 | 15.8 | 0.17 |
| polarC18 | 758.568 | 136.0356 | 15.8 | 0.17 |
| polarC18 | 758.568 | 262.2492 | 15.8 | 0.17 |
| polarC18 | 758.568 | 285.1759 | 15.8 | 0.17 |
| polarC18 | 758.568 | 291.4979 | 15.8 | 0.17 |
| polarC18 | 758.568 | 149.2562 | 15.7 | 0.17 |
| polarC18 | 758.568 | 742.3031 | 15.7 | 0.17 |
| polarC18 | 758.568 | 82.3377 | 15.7 | 0.17 |
| polarC18 | 758.568 | 204.135 | 15.7 | 0.17 |
| polarC18 | 758.568 | 237.2425 | 15.7 | 0.17 |
| polarC18 | 758.568 | 456.2859 | 15.7 | 0.17 |
| polarC18 | 758.568 | 637.5571 | 15.7 | 0.17 |
| polarC18 | 758.568 | 363.5169 | 15.7 | 0.17 |
| polarC18 | 758.568 | 108.4817 | 15.7 | 0.17 |
| polarC18 | 758.568 | 336.0115 | 15.7 | 0.17 |
| polarC18 | 758.568 | 288.5503 | 15.7 | 0.17 |
| polarC18 | 758.568 | 106.7336 | 15.7 | 0.17 |

| | | | | |
|---|---|---|---|---|
| polarC18 | 758.568 | 144.4327 | 15.7 | 0.17 |
| polarC18 | 758.568 | 238.741 | 15.7 | 0.17 |
| polarC18 | 758.568 | 219.8071 | 15.7 | 0.17 |
| polarC18 | 758.568 | 214.5509 | 15.7 | 0.17 |
| polarC18 | 758.568 | 267.1076 | 15.7 | 0.17 |
| polarC18 | 758.568 | 362.0976 | 15.7 | 0.17 |
| polarC18 | 758.568 | 353.1669 | 15.7 | 0.17 |
| polarC18 | 758.568 | 114.8356 | 15.7 | 0.17 |
| polarC18 | 758.568 | 236.2512 | 15.7 | 0.17 |
| polarC18 | 758.568 | 154.6271 | 15.7 | 0.17 |
| polarC18 | 758.568 | 138.9747 | 15.7 | 0.17 |
| polarC18 | 758.568 | 178.4008 | 15.7 | 0.17 |
| polarC18 | 758.568 | 139.6826 | 15.7 | 0.17 |
| polarC18 | 758.568 | 142.6955 | 15.7 | 0.17 |
| polarC18 | 758.568 | 97.3927 | 15.7 | 0.17 |
| polarC18 | 758.568 | 141.653 | 15.7 | 0.17 |
| polarC18 | 758.568 | 163.7205 | 15.7 | 0.17 |
| polarC18 | 758.568 | 122.801 | 15.7 | 0.17 |
| polarC18 | 758.568 | 338.2921 | 15.7 | 0.17 |
| polarC18 | 758.568 | 93.9746 | 15.7 | 0.17 |
| polarC18 | 758.568 | 126.8868 | 15.7 | 0.17 |
| polarC18 | 758.568 | 640.4103 | 15.7 | 0.17 |
| polarC18 | 758.568 | 130.9807 | 15.7 | 0.17 |
| polarC18 | 758.568 | 483.4755 | 15.7 | 0.17 |
| polarC18 | 758.568 | 331.3659 | 15.7 | 0.17 |
| polarC18 | 758.568 | 94.7366 | 15.7 | 0.17 |
| polarC18 | 758.568 | 115.1509 | 15.7 | 0.17 |
| polarC18 | 758.568 | 108.8206 | 15.7 | 0.17 |
| polarC18 | 758.568 | 127.2239 | 15.7 | 0.17 |
| polarC18 | 758.568 | 149.0189 | 15.7 | 0.17 |
| polarC18 | 758.568 | 502.915 | 15.7 | 0.17 |
| polarC18 | 758.568 | 304.3139 | 15.7 | 0.17 |
| polarC18 | 758.568 | 94.2109 | 15.7 | 0.17 |
| polarC18 | 758.568 | 426.7082 | 15.7 | 0.17 |
| polarC18 | 758.568 | 244.1044 | 15.7 | 0.17 |
| polarC18 | 758.568 | 220.6346 | 15.7 | 0.17 |
| polarC18 | 758.568 | 83.8322 | 15.7 | 0.17 |
| polarC18 | 758.568 | 223.6489 | 15.7 | 0.17 |
| polarC18 | 758.568 | 98.8809 | 15.7 | 0.17 |
| polarC18 | 758.568 | 196.3069 | 15.7 | 0.17 |
| polarC18 | 758.568 | 569.0499 | 15.7 | 0.17 |
| polarC18 | 758.568 | 150.5032 | 15.7 | 0.17 |

| | | | | |
|---|---|---|---|---|
| polarC18 | 758.568 | 176.0926 | 15.7 | 0.17 |
| polarC18 | 758.568 | 84.3488 | 15.7 | 0.17 |
| polarC18 | 758.568 | 103.9233 | 15.7 | 0.17 |
| polarC18 | 758.568 | 199.7461 | 15.7 | 0.17 |
| polarC18 | 758.568 | 185.548 | 15.7 | 0.17 |
| polarC18 | 758.568 | 80.7641 | 15.7 | 0.17 |
| polarC18 | 758.568 | 166.2179 | 15.7 | 0.17 |
| polarC18 | 758.568 | 484.1122 | 15.7 | 0.17 |
| polarC18 | 758.568 | 98.5169 | 15.7 | 0.17 |
| polarC18 | 758.568 | 679.337 | 15.7 | 0.17 |
| polarC18 | 758.568 | 150.0085 | 15.7 | 0.17 |
| polarC18 | 758.568 | 381.596 | 15.7 | 0.17 |
| polarC18 | 758.568 | 145.1428 | 15.7 | 0.17 |
| polarC18 | 758.568 | 271.0503 | 15.7 | 0.17 |
| polarC18 | 758.568 | 580.2363 | 15.7 | 0.17 |
| polarC18 | 758.568 | 124.4301 | 15.7 | 0.17 |
| polarC18 | 758.568 | 93.1529 | 15.7 | 0.17 |
| polarC18 | 758.568 | 165.0606 | 15.7 | 0.17 |
| polarC18 | 758.568 | 110.7777 | 15.7 | 0.17 |
| polarC18 | 758.568 | 138.7034 | 15.7 | 0.17 |
| polarC18 | 758.568 | 225.7293 | 15.7 | 0.17 |
| polarC18 | 758.568 | 115.783 | 15.7 | 0.17 |
| polarC18 | 758.568 | 104.7506 | 15.7 | 0.17 |
| polarC18 | 758.568 | 581.2689 | 15.7 | 0.17 |
| polarC18 | 758.568 | 701.1968 | 15.7 | 0.17 |
| polarC18 | 758.568 | 159.6433 | 15.7 | 0.17 |
| polarC18 | 758.568 | 408.4059 | 15.7 | 0.17 |
| polarC18 | 758.568 | 164.9215 | 15.7 | 0.17 |
| polarC18 | 758.568 | 202.6119 | 15.7 | 0.17 |
| polarC18 | 758.568 | 132.2114 | 15.7 | 0.17 |
| polarC18 | 758.568 | 85.0048 | 15.7 | 0.17 |
| polarC18 | 758.568 | 244.0404 | 15.7 | 0.17 |
| polarC18 | 758.568 | 289.0481 | 15.7 | 0.17 |
| polarC18 | 758.568 | 322.7467 | 15.7 | 0.17 |
| polarC18 | 758.568 | 386.5353 | 15.7 | 0.17 |
| polarC18 | 758.568 | 614.5608 | 15.6 | 0.17 |
| polarC18 | 758.568 | 498.0368 | 15.6 | 0.17 |
| polarC18 | 758.568 | 224.6996 | 15.6 | 0.17 |
| polarC18 | 758.568 | 90.9103 | 15.6 | 0.17 |
| polarC18 | 758.568 | 181.7589 | 15.6 | 0.17 |
| polarC18 | 758.568 | 488.5178 | 15.6 | 0.17 |
| polarC18 | 758.568 | 136.989 | 15.6 | 0.17 |

| | | | | |
|---|---|---|---|---|
| polarC18 | 758.568 | 290.7058 | 15.6 | 0.17 |
| polarC18 | 758.568 | 122.3215 | 15.6 | 0.17 |
| polarC18 | 758.568 | 80.7599 | 15.6 | 0.17 |
| polarC18 | 758.568 | 557.0977 | 15.6 | 0.17 |
| polarC18 | 758.568 | 460.1706 | 15.6 | 0.17 |
| polarC18 | 758.568 | 192.7154 | 15.6 | 0.17 |
| polarC18 | 758.568 | 95.3511 | 15.6 | 0.17 |
| polarC18 | 758.568 | 183.6732 | 15.6 | 0.17 |
| polarC18 | 758.568 | 283.4243 | 15.6 | 0.17 |
| polarC18 | 758.568 | 155.6526 | 15.6 | 0.17 |
| polarC18 | 758.568 | 90.4667 | 15.6 | 0.17 |
| polarC18 | 758.568 | 151.804 | 15.6 | 0.17 |
| polarC18 | 758.568 | 621.9994 | 15.6 | 0.17 |
| polarC18 | 758.568 | 405.3638 | 15.6 | 0.17 |
| polarC18 | 758.568 | 192.624 | 15.6 | 0.17 |
| polarC18 | 758.568 | 125.2854 | 15.6 | 0.17 |
| polarC18 | 758.568 | 97.3835 | 15.6 | 0.17 |
| polarC18 | 758.568 | 135.618 | 15.6 | 0.17 |
| polarC18 | 758.568 | 193.9483 | 15.6 | 0.17 |
| polarC18 | 758.568 | 88.7067 | 15.6 | 0.17 |
| polarC18 | 758.568 | 133.7269 | 15.6 | 0.17 |
| polarC18 | 758.568 | 266.364 | 15.6 | 0.17 |
| polarC18 | 758.568 | 79.457 | 15.6 | 0.17 |
| polarC18 | 758.568 | 232.1632 | 15.6 | 0.17 |
| polarC18 | 758.568 | 281.357 | 15.6 | 0.17 |
| polarC18 | 758.568 | 225.9902 | 15.6 | 0.17 |
| polarC18 | 758.568 | 281.0354 | 15.6 | 0.17 |
| polarC18 | 758.568 | 135.7863 | 15.6 | 0.17 |
| polarC18 | 758.568 | 118.5856 | 15.6 | 0.17 |
| polarC18 | 758.568 | 135.8232 | 15.6 | 0.17 |
| polarC18 | 758.568 | 659.6854 | 15.6 | 0.17 |
| polarC18 | 758.568 | 243.0134 | 15.6 | 0.17 |
| polarC18 | 758.568 | 94.4187 | 15.6 | 0.17 |
| polarC18 | 758.568 | 170.2412 | 15.6 | 0.17 |
| polarC18 | 758.568 | 177.779 | 15.6 | 0.17 |
| polarC18 | 758.568 | 185.4568 | 15.6 | 0.17 |
| polarC18 | 758.568 | 340.6513 | 15.6 | 0.17 |
| polarC18 | 758.568 | 90.0171 | 15.6 | 0.17 |
| polarC18 | 758.568 | 147.1132 | 15.6 | 0.17 |
| polarC18 | 758.568 | 316.701 | 15.6 | 0.17 |
| polarC18 | 758.568 | 261.4562 | 15.6 | 0.17 |
| polarC18 | 758.568 | 789.285 | 15.6 | 0.17 |

| | | | | |
|---|---|---|---|---|
| polarC18 | 758.568 | 85.5727 | 15.6 | 0.17 |
| polarC18 | 758.568 | 148.0339 | 15.6 | 0.17 |
| polarC18 | 758.568 | 291.1947 | 15.6 | 0.17 |
| polarC18 | 758.568 | 97.8817 | 15.6 | 0.17 |
| polarC18 | 758.568 | 113.2812 | 15.6 | 0.17 |
| polarC18 | 758.568 | 122.5168 | 15.6 | 0.17 |
| polarC18 | 758.568 | 122.5732 | 15.6 | 0.17 |
| polarC18 | 758.568 | 365.1292 | 15.6 | 0.17 |
| polarC18 | 758.568 | 279.3904 | 15.6 | 0.17 |
| polarC18 | 758.568 | 214.2307 | 15.6 | 0.17 |
| polarC18 | 758.568 | 232.0382 | 15.6 | 0.17 |
| polarC18 | 758.568 | 146.7621 | 15.6 | 0.17 |
| polarC18 | 758.568 | 395.4657 | 15.6 | 0.17 |
| polarC18 | 758.568 | 136.4295 | 15.6 | 0.17 |
| polarC18 | 758.568 | 107.9378 | 15.6 | 0.17 |
| polarC18 | 758.568 | 185.0944 | 15.6 | 0.17 |
| polarC18 | 758.568 | 347.1744 | 15.6 | 0.17 |
| polarC18 | 758.568 | 111.6381 | 15.6 | 0.17 |
| polarC18 | 758.568 | 168.8274 | 15.6 | 0.17 |
| polarC18 | 758.568 | 79.8355 | 15.6 | 0.17 |
| polarC18 | 758.568 | 487.6357 | 15.6 | 0.17 |
| polarC18 | 758.568 | 124.7571 | 15.6 | 0.17 |
| polarC18 | 758.568 | 677.2809 | 15.6 | 0.17 |
| polarC18 | 758.568 | 406.5203 | 15.6 | 0.17 |
| polarC18 | 758.568 | 99.1775 | 15.6 | 0.17 |
| polarC18 | 758.568 | 141.0602 | 15.6 | 0.17 |
| polarC18 | 758.568 | 284.6499 | 15.6 | 0.17 |
| polarC18 | 758.568 | 104.6859 | 15.6 | 0.17 |
| polarC18 | 758.568 | 148.8899 | 15.6 | 0.17 |
| polarC18 | 758.568 | 79.6922 | 15.6 | 0.17 |
| polarC18 | 758.568 | 126.7648 | 15.6 | 0.17 |
| polarC18 | 758.568 | 81.7865 | 15.6 | 0.17 |
| polarC18 | 758.568 | 155.6359 | 15.6 | 0.17 |
| polarC18 | 758.568 | 153.9983 | 15.6 | 0.17 |
| polarC18 | 758.568 | 208.9639 | 15.6 | 0.17 |
| polarC18 | 758.568 | 114.5301 | 15.6 | 0.17 |
| polarC18 | 758.568 | 139.2462 | 15.6 | 0.17 |
| polarC18 | 758.568 | 158.1583 | 15.6 | 0.17 |
| polarC18 | 758.568 | 79.5284 | 15.6 | 0.17 |
| polarC18 | 758.568 | 85.1962 | 15.6 | 0.17 |
| polarC18 | 758.568 | 194.7324 | 15.6 | 0.17 |
| polarC18 | 758.568 | 128.1571 | 15.6 | 0.17 |

| | | | | |
|---|---|---|---|---|
| polarC18 | 758.568 | 181.6475 | 15.6 | 0.17 |
| polarC18 | 758.568 | 94.2386 | 15.6 | 0.17 |
| polarC18 | 758.568 | 218.8902 | 15.6 | 0.17 |
| polarC18 | 758.568 | 157.4245 | 15.6 | 0.17 |
| polarC18 | 758.568 | 107.7303 | 15.5 | 0.17 |
| polarC18 | 758.568 | 135.5001 | 15.5 | 0.17 |
| polarC18 | 758.568 | 154.226 | 15.5 | 0.17 |
| polarC18 | 758.568 | 92.5677 | 15.5 | 0.17 |
| polarC18 | 758.568 | 405.7234 | 15.5 | 0.17 |
| polarC18 | 758.568 | 200.2316 | 15.5 | 0.17 |
| polarC18 | 758.568 | 238.45 | 15.5 | 0.17 |
| polarC18 | 758.568 | 539.9949 | 15.5 | 0.17 |
| polarC18 | 758.568 | 262.7752 | 15.5 | 0.17 |
| polarC18 | 758.568 | 114.6044 | 15.5 | 0.17 |
| polarC18 | 758.568 | 161.3227 | 15.5 | 0.17 |
| polarC18 | 758.568 | 715.0134 | 15.5 | 0.17 |
| polarC18 | 758.568 | 138.4583 | 15.5 | 0.17 |
| polarC18 | 758.568 | 756.2155 | 15.5 | 0.17 |
| polarC18 | 758.568 | 101.7747 | 15.5 | 0.17 |
| polarC18 | 758.568 | 295.0748 | 15.5 | 0.17 |
| polarC18 | 758.568 | 98.0681 | 15.5 | 0.17 |
| polarC18 | 758.568 | 123.4004 | 15.5 | 0.17 |
| polarC18 | 758.568 | 193.3315 | 15.5 | 0.17 |
| polarC18 | 758.568 | 154.7517 | 15.5 | 0.17 |
| polarC18 | 758.568 | 124.2134 | 15.5 | 0.17 |
| polarC18 | 758.568 | 92.3521 | 15.5 | 0.17 |
| polarC18 | 758.568 | 231.9801 | 15.5 | 0.17 |
| polarC18 | 758.568 | 227.5775 | 15.5 | 0.17 |
| polarC18 | 758.568 | 218.0732 | 15.5 | 0.17 |
| polarC18 | 758.568 | 207.6022 | 15.5 | 0.17 |
| polarC18 | 758.568 | 387.9741 | 15.5 | 0.17 |
| polarC18 | 758.568 | 95.9785 | 15.5 | 0.17 |
| polarC18 | 758.568 | 577.0314 | 15.5 | 0.17 |
| polarC18 | 758.568 | 416.9806 | 15.5 | 0.17 |
| polarC18 | 758.568 | 138.3059 | 15.5 | 0.17 |
| polarC18 | 758.568 | 123.0798 | 15.5 | 0.17 |
| polarC18 | 758.568 | 82.479 | 15.5 | 0.17 |
| polarC18 | 758.568 | 246.1885 | 15.5 | 0.17 |
| polarC18 | 758.568 | 266.2763 | 15.5 | 0.17 |
| polarC18 | 758.568 | 179.7701 | 15.5 | 0.17 |
| polarC18 | 758.568 | 153.3952 | 15.5 | 0.17 |
| polarC18 | 758.568 | 190.9427 | 15.5 | 0.17 |

| | | | | |
|---|---|---|---|---|
| polarC18 | 758.568 | 308.2746 | 15.5 | 0.17 |
| polarC18 | 758.568 | 452.1713 | 15.5 | 0.17 |
| polarC18 | 758.568 | 98.538 | 15.5 | 0.17 |
| polarC18 | 758.568 | 788.4615 | 15.5 | 0.17 |
| polarC18 | 758.568 | 433.3048 | 15.5 | 0.17 |
| polarC18 | 758.568 | 234.9767 | 15.5 | 0.17 |
| polarC18 | 758.568 | 82.4959 | 15.5 | 0.17 |
| polarC18 | 758.568 | 146.9015 | 15.5 | 0.17 |
| polarC18 | 758.568 | 87.0771 | 15.5 | 0.17 |
| polarC18 | 758.568 | 147.5553 | 15.5 | 0.17 |
| polarC18 | 758.568 | 104.9428 | 15.5 | 0.17 |
| polarC18 | 758.568 | 133.291 | 15.5 | 0.17 |
| polarC18 | 758.568 | 93.9523 | 15.5 | 0.17 |
| polarC18 | 758.568 | 146.5485 | 15.5 | 0.17 |
| polarC18 | 758.568 | 437.7053 | 15.5 | 0.17 |
| polarC18 | 758.568 | 152.5317 | 15.5 | 0.17 |
| polarC18 | 758.568 | 134.37 | 15.5 | 0.17 |
| polarC18 | 758.568 | 115.2086 | 15.5 | 0.17 |
| polarC18 | 758.568 | 523.9293 | 15.5 | 0.17 |
| polarC18 | 758.568 | 194.1237 | 15.5 | 0.17 |
| polarC18 | 758.568 | 127.257 | 15.5 | 0.17 |
| polarC18 | 758.568 | 262.1105 | 15.5 | 0.17 |
| polarC18 | 758.568 | 528.4791 | 15.5 | 0.17 |
| polarC18 | 758.568 | 102.6803 | 15.5 | 0.17 |
| polarC18 | 758.568 | 109.0938 | 15.5 | 0.17 |
| polarC18 | 758.568 | 199.7106 | 15.5 | 0.17 |
| polarC18 | 758.568 | 88.962 | 15.5 | 0.17 |
| polarC18 | 758.568 | 114.2919 | 15.5 | 0.17 |
| polarC18 | 758.568 | 84.5895 | 15.5 | 0.17 |
| polarC18 | 758.568 | 83.6 | 15.5 | 0.17 |
| polarC18 | 758.568 | 81.8708 | 15.5 | 0.17 |
| polarC18 | 758.568 | 435.4943 | 15.5 | 0.17 |
| polarC18 | 758.568 | 677.5358 | 15.5 | 0.17 |
| polarC18 | 758.568 | 147.7701 | 15.5 | 0.17 |
| polarC18 | 758.568 | 109.9077 | 15.5 | 0.17 |
| polarC18 | 758.568 | 224.8011 | 15.5 | 0.17 |
| polarC18 | 758.568 | 319.8015 | 15.4 | 0.17 |
| polarC18 | 758.568 | 166.4755 | 15.4 | 0.17 |
| polarC18 | 758.568 | 121.7501 | 15.4 | 0.17 |
| polarC18 | 758.568 | 81.5791 | 15.4 | 0.17 |
| polarC18 | 758.568 | 86.1586 | 15.4 | 0.17 |
| polarC18 | 758.568 | 131.8858 | 15.4 | 0.17 |

| | | | | |
|---|---|---|---|---|
| polarC18 | 758.568 | 83.9809 | 15.4 | 0.17 |
| polarC18 | 758.568 | 675.5898 | 15.4 | 0.17 |
| polarC18 | 758.568 | 94.9508 | 15.4 | 0.17 |
| polarC18 | 758.568 | 103.9014 | 15.4 | 0.17 |
| polarC18 | 758.568 | 217.8442 | 15.4 | 0.17 |
| polarC18 | 758.568 | 120.514 | 15.4 | 0.17 |
| polarC18 | 758.568 | 86.4126 | 15.4 | 0.17 |
| polarC18 | 758.568 | 145.9385 | 15.4 | 0.17 |
| polarC18 | 758.568 | 111.1168 | 15.4 | 0.17 |
| polarC18 | 758.568 | 702.9082 | 15.4 | 0.17 |
| polarC18 | 758.568 | 507.6507 | 15.4 | 0.17 |
| polarC18 | 758.568 | 84.3771 | 15.4 | 0.17 |
| polarC18 | 758.568 | 129.5001 | 15.4 | 0.17 |
| polarC18 | 758.568 | 100.6497 | 15.4 | 0.17 |
| polarC18 | 758.568 | 91.1076 | 15.4 | 0.17 |
| polarC18 | 758.568 | 256.4583 | 15.4 | 0.17 |
| polarC18 | 758.568 | 105.0153 | 15.4 | 0.17 |
| polarC18 | 758.568 | 248.7757 | 15.4 | 0.17 |
| polarC18 | 758.568 | 214.9716 | 15.4 | 0.17 |
| polarC18 | 758.568 | 126.5394 | 15.4 | 0.17 |
| polarC18 | 758.568 | 113.3084 | 15.4 | 0.17 |
| polarC18 | 758.568 | 140.1709 | 15.4 | 0.17 |
| polarC18 | 758.568 | 583.8667 | 15.4 | 0.17 |
| polarC18 | 758.568 | 93.7875 | 15.4 | 0.17 |
| polarC18 | 758.568 | 193.2847 | 15.4 | 0.17 |
| polarC18 | 758.568 | 428.3597 | 15.4 | 0.17 |
| polarC18 | 758.568 | 606.3443 | 15.4 | 0.17 |
| polarC18 | 758.568 | 214.0699 | 15.4 | 0.17 |
| polarC18 | 758.568 | 94.0446 | 15.4 | 0.17 |
| polarC18 | 758.568 | 128.4313 | 15.4 | 0.17 |
| polarC18 | 758.568 | 309.5357 | 15.4 | 0.17 |
| polarC18 | 758.568 | 79.9778 | 15.4 | 0.17 |
| polarC18 | 758.568 | 276.526 | 15.4 | 0.17 |
| polarC18 | 758.568 | 84.3721 | 15.4 | 0.17 |
| polarC18 | 758.568 | 84.8767 | 15.4 | 0.17 |
| polarC18 | 758.568 | 123.4838 | 15.4 | 0.17 |
| polarC18 | 758.568 | 192.5938 | 15.4 | 0.17 |
| polarC18 | 758.568 | 442.0932 | 15.4 | 0.17 |
| polarC18 | 758.568 | 79.7234 | 15.4 | 0.17 |
| polarC18 | 758.568 | 187.7802 | 15.4 | 0.17 |
| polarC18 | 758.568 | 90.7377 | 15.4 | 0.17 |
| polarC18 | 758.568 | 106.6209 | 15.4 | 0.17 |

| | | | | |
|---|---|---|---|---|
| polarC18 | 758.568 | 97.3212 | 15.4 | 0.17 |
| polarC18 | 758.568 | 91.1378 | 15.4 | 0.17 |
| polarC18 | 758.568 | 135.8906 | 15.4 | 0.17 |
| polarC18 | 758.568 | 392.3384 | 15.4 | 0.17 |
| polarC18 | 758.568 | 95.1732 | 15.4 | 0.17 |
| polarC18 | 758.568 | 232.473 | 15.4 | 0.17 |
| polarC18 | 758.568 | 589.1366 | 15.4 | 0.17 |
| polarC18 | 758.568 | 116.691 | 15.4 | 0.17 |
| polarC18 | 758.568 | 495.3165 | 15.4 | 0.17 |
| polarC18 | 758.568 | 145.8278 | 15.4 | 0.17 |
| polarC18 | 758.568 | 204.2659 | 15.4 | 0.17 |
| polarC18 | 758.568 | 272.2483 | 15.4 | 0.17 |
| polarC18 | 758.568 | 85.6823 | 15.4 | 0.17 |
| polarC18 | 758.568 | 80.9764 | 15.4 | 0.17 |
| polarC18 | 758.568 | 345.0787 | 15.4 | 0.17 |
| polarC18 | 758.568 | 264.5335 | 15.4 | 0.17 |
| polarC18 | 758.568 | 192.7954 | 15.4 | 0.17 |
| polarC18 | 758.568 | 123.672 | 15.4 | 0.17 |
| polarC18 | 758.568 | 232.0011 | 15.4 | 0.17 |
| polarC18 | 758.568 | 81.984 | 15.4 | 0.17 |
| polarC18 | 758.568 | 127.8049 | 15.4 | 0.17 |
| polarC18 | 758.568 | 93.3267 | 15.4 | 0.17 |
| polarC18 | 758.568 | 704.3694 | 15.4 | 0.17 |
| polarC18 | 758.568 | 123.882 | 15.4 | 0.16 |
| polarC18 | 758.568 | 390.5934 | 15.4 | 0.16 |
| polarC18 | 758.568 | 167.7158 | 15.4 | 0.16 |
| polarC18 | 758.568 | 144.5709 | 15.4 | 0.16 |
| polarC18 | 758.568 | 148.8617 | 15.4 | 0.16 |
| polarC18 | 758.568 | 167.0526 | 15.4 | 0.16 |
| polarC18 | 758.568 | 259.4176 | 15.4 | 0.16 |
| polarC18 | 758.568 | 275.3264 | 15.4 | 0.16 |
| polarC18 | 758.568 | 189.3774 | 15.4 | 0.16 |
| polarC18 | 758.568 | 116.2356 | 15.4 | 0.16 |
| polarC18 | 758.568 | 112.1663 | 15.4 | 0.16 |
| polarC18 | 758.568 | 107.8243 | 15.4 | 0.16 |
| polarC18 | 758.568 | 535.3331 | 15.4 | 0.16 |
| polarC18 | 758.568 | 147.4364 | 15.4 | 0.16 |
| polarC18 | 758.568 | 92.3756 | 15.4 | 0.16 |
| polarC18 | 758.568 | 89.2488 | 15.4 | 0.16 |
| polarC18 | 758.568 | 107.1223 | 15.4 | 0.16 |
| polarC18 | 758.568 | 168.7001 | 15.3 | 0.16 |
| polarC18 | 758.568 | 98.1178 | 15.3 | 0.16 |

| | | | | |
|---|---|---|---|---|
| polarC18 | 758.568 | 84.9698 | 15.3 | 0.16 |
| polarC18 | 758.568 | 98.0098 | 15.3 | 0.16 |
| polarC18 | 758.568 | 109.2402 | 15.3 | 0.16 |
| polarC18 | 758.568 | 312.1691 | 15.3 | 0.16 |
| polarC18 | 758.568 | 221.2926 | 15.3 | 0.16 |
| polarC18 | 758.568 | 84.3036 | 15.3 | 0.16 |
| polarC18 | 758.568 | 439.1035 | 15.3 | 0.16 |
| polarC18 | 758.568 | 361.6478 | 15.3 | 0.16 |
| polarC18 | 758.568 | 190.7842 | 15.3 | 0.16 |
| polarC18 | 758.568 | 112.246 | 15.3 | 0.16 |
| polarC18 | 758.568 | 109.3144 | 15.3 | 0.16 |
| polarC18 | 758.568 | 143.844 | 15.3 | 0.16 |
| polarC18 | 758.568 | 81.1735 | 15.3 | 0.16 |
| polarC18 | 758.568 | 97.5881 | 15.3 | 0.16 |
| polarC18 | 758.568 | 107.749 | 15.3 | 0.16 |
| polarC18 | 758.568 | 106.4648 | 15.3 | 0.16 |
| polarC18 | 758.568 | 452.5265 | 15.3 | 0.16 |
| polarC18 | 758.568 | 666.497 | 15.3 | 0.16 |
| polarC18 | 758.568 | 558.2849 | 15.3 | 0.16 |
| polarC18 | 758.568 | 543.1262 | 15.3 | 0.16 |
| polarC18 | 758.568 | 125.9601 | 15.3 | 0.16 |
| polarC18 | 758.568 | 451.2596 | 15.3 | 0.16 |
| polarC18 | 758.568 | 136.0833 | 15.3 | 0.16 |
| polarC18 | 758.568 | 366.5984 | 15.3 | 0.16 |
| polarC18 | 758.568 | 147.7107 | 15.3 | 0.16 |
| polarC18 | 758.568 | 473.6862 | 15.3 | 0.16 |
| polarC18 | 758.568 | 93.3076 | 15.3 | 0.16 |
| polarC18 | 758.568 | 138.8366 | 15.3 | 0.16 |
| polarC18 | 758.568 | 156.8312 | 15.3 | 0.16 |
| polarC18 | 758.568 | 175.2165 | 15.3 | 0.16 |
| polarC18 | 758.568 | 113.0507 | 15.3 | 0.16 |
| polarC18 | 758.568 | 111.4991 | 15.3 | 0.16 |
| polarC18 | 758.568 | 96.0079 | 15.3 | 0.16 |
| polarC18 | 758.568 | 129.3198 | 15.3 | 0.16 |
| polarC18 | 758.568 | 397.379 | 15.3 | 0.16 |
| polarC18 | 758.568 | 360.9254 | 15.3 | 0.16 |
| polarC18 | 758.568 | 120.3208 | 15.3 | 0.16 |
| polarC18 | 758.568 | 160.8767 | 15.3 | 0.16 |
| polarC18 | 758.568 | 169.1614 | 15.3 | 0.16 |
| polarC18 | 758.568 | 110.6013 | 15.3 | 0.16 |
| polarC18 | 758.568 | 198.5147 | 15.3 | 0.16 |
| polarC18 | 758.568 | 105.9579 | 15.3 | 0.16 |

| | | | | |
|---|---|---|---|---|
| polarC18 | 758.568 | 110.7104 | 15.3 | 0.16 |
| polarC18 | 758.568 | 136.445 | 15.3 | 0.16 |
| polarC18 | 758.568 | 609.3514 | 15.3 | 0.16 |
| polarC18 | 758.568 | 152.3962 | 15.3 | 0.16 |
| polarC18 | 758.568 | 91.8879 | 15.3 | 0.16 |
| polarC18 | 758.568 | 408.3295 | 15.3 | 0.16 |
| polarC18 | 758.568 | 418.0865 | 15.3 | 0.16 |
| polarC18 | 758.568 | 650.6285 | 15.3 | 0.16 |
| polarC18 | 758.568 | 118.9072 | 15.3 | 0.16 |
| polarC18 | 758.568 | 82.6382 | 15.3 | 0.16 |
| polarC18 | 758.568 | 357.9027 | 15.3 | 0.16 |
| polarC18 | 758.568 | 85.7746 | 15.3 | 0.16 |
| polarC18 | 758.568 | 186.0947 | 15.3 | 0.16 |
| polarC18 | 758.568 | 310.9217 | 15.3 | 0.16 |
| polarC18 | 758.568 | 121.1988 | 15.3 | 0.16 |
| polarC18 | 758.568 | 318.6021 | 15.3 | 0.16 |
| polarC18 | 758.568 | 95.9879 | 15.3 | 0.16 |
| polarC18 | 758.568 | 272.6461 | 15.3 | 0.16 |
| polarC18 | 758.568 | 89.944 | 15.3 | 0.16 |
| polarC18 | 758.568 | 584.262 | 15.3 | 0.16 |
| polarC18 | 758.568 | 260.7219 | 15.3 | 0.16 |
| polarC18 | 758.568 | 94.6758 | 15.3 | 0.16 |
| polarC18 | 758.568 | 427.0006 | 15.3 | 0.16 |
| polarC18 | 758.568 | 230.583 | 15.3 | 0.16 |
| polarC18 | 758.568 | 127.272 | 15.3 | 0.16 |
| polarC18 | 758.568 | 266.7311 | 15.3 | 0.16 |
| polarC18 | 758.568 | 88.907 | 15.3 | 0.16 |
| polarC18 | 758.568 | 347.067 | 15.3 | 0.16 |
| polarC18 | 758.568 | 82.2327 | 15.3 | 0.16 |
| polarC18 | 758.568 | 96.257 | 15.3 | 0.16 |
| polarC18 | 758.568 | 106.0934 | 15.3 | 0.16 |
| polarC18 | 758.568 | 145.7411 | 15.3 | 0.16 |
| polarC18 | 758.568 | 128.2031 | 15.2 | 0.16 |
| polarC18 | 758.568 | 148.552 | 15.2 | 0.16 |
| polarC18 | 758.568 | 190.4273 | 15.2 | 0.16 |
| polarC18 | 758.568 | 131.6675 | 15.2 | 0.16 |
| polarC18 | 758.568 | 99.6931 | 15.2 | 0.16 |
| polarC18 | 758.568 | 192.3348 | 15.2 | 0.16 |
| polarC18 | 758.568 | 130.4109 | 15.2 | 0.16 |
| polarC18 | 758.568 | 122.7264 | 15.2 | 0.16 |
| polarC18 | 758.568 | 211.5877 | 15.2 | 0.16 |
| polarC18 | 758.568 | 152.9325 | 15.2 | 0.16 |

| | | | | |
|---|---|---|---|---|
| polarC18 | 758.568 | 156.7068 | 15.2 | 0.16 |
| polarC18 | 758.568 | 258.7907 | 15.2 | 0.16 |
| polarC18 | 758.568 | 347.7497 | 15.2 | 0.16 |
| polarC18 | 758.568 | 103.2472 | 15.2 | 0.16 |
| polarC18 | 758.568 | 184.1182 | 15.2 | 0.16 |
| polarC18 | 758.568 | 292.2212 | 15.2 | 0.16 |
| polarC18 | 758.568 | 282.3469 | 15.2 | 0.16 |
| polarC18 | 758.568 | 86.0921 | 15.2 | 0.16 |
| polarC18 | 758.568 | 348.1853 | 15.2 | 0.16 |
| polarC18 | 758.568 | 347.9123 | 15.2 | 0.16 |
| polarC18 | 758.568 | 89.4993 | 15.2 | 0.16 |
| polarC18 | 758.568 | 366.4467 | 15.2 | 0.16 |
| polarC18 | 758.568 | 172.102 | 15.2 | 0.16 |
| polarC18 | 758.568 | 83.2322 | 15.2 | 0.16 |
| polarC18 | 758.568 | 352.0064 | 15.2 | 0.16 |
| polarC18 | 758.568 | 260.9685 | 15.2 | 0.16 |
| polarC18 | 758.568 | 179.1571 | 15.2 | 0.16 |
| polarC18 | 758.568 | 215.1952 | 15.2 | 0.16 |
| polarC18 | 758.568 | 330.7437 | 15.2 | 0.16 |
| polarC18 | 758.568 | 163.6406 | 15.2 | 0.16 |
| polarC18 | 758.568 | 418.7343 | 15.2 | 0.16 |
| polarC18 | 758.568 | 119.3992 | 15.2 | 0.16 |
| polarC18 | 758.568 | 89.5126 | 15.2 | 0.16 |
| polarC18 | 758.568 | 163.7684 | 15.2 | 0.16 |
| polarC18 | 758.568 | 95.8266 | 15.2 | 0.16 |
| polarC18 | 758.568 | 205.3876 | 15.2 | 0.16 |
| polarC18 | 758.568 | 115.4209 | 15.2 | 0.16 |
| polarC18 | 758.568 | 103.1361 | 15.2 | 0.16 |
| polarC18 | 758.568 | 419.8759 | 15.2 | 0.16 |
| polarC18 | 758.568 | 97.9525 | 15.2 | 0.16 |
| polarC18 | 758.568 | 150.9251 | 15.2 | 0.16 |
| polarC18 | 758.568 | 90.6283 | 15.2 | 0.16 |
| polarC18 | 758.568 | 367.2896 | 15.2 | 0.16 |
| polarC18 | 758.568 | 214.5826 | 15.2 | 0.16 |
| polarC18 | 758.568 | 764.1661 | 15.2 | 0.16 |
| polarC18 | 758.568 | 91.6083 | 15.2 | 0.16 |
| polarC18 | 758.568 | 573.093 | 15.2 | 0.16 |
| polarC18 | 758.568 | 312.7871 | 15.2 | 0.16 |
| polarC18 | 758.568 | 166.3055 | 15.2 | 0.16 |
| polarC18 | 758.568 | 789.9269 | 15.2 | 0.16 |
| polarC18 | 758.568 | 237.5814 | 15.2 | 0.16 |
| polarC18 | 758.568 | 90.0405 | 15.2 | 0.16 |

| | | | | |
|---|---|---|---|---|
| polarC18 | 758.568 | 138.6423 | 15.2 | 0.16 |
| polarC18 | 758.568 | 342.868 | 15.2 | 0.16 |
| polarC18 | 758.568 | 101.6169 | 15.2 | 0.16 |
| polarC18 | 758.568 | 157.7769 | 15.2 | 0.16 |
| polarC18 | 758.568 | 190.5028 | 15.2 | 0.16 |
| polarC18 | 758.568 | 316.37 | 15.2 | 0.16 |
| polarC18 | 758.568 | 110.0865 | 15.2 | 0.16 |
| polarC18 | 758.568 | 221.5243 | 15.2 | 0.16 |
| polarC18 | 758.568 | 216.891 | 15.2 | 0.16 |
| polarC18 | 758.568 | 244.173 | 15.2 | 0.16 |
| polarC18 | 758.568 | 263.7493 | 15.2 | 0.16 |
| polarC18 | 758.568 | 139.8132 | 15.2 | 0.16 |
| polarC18 | 758.568 | 123.0387 | 15.2 | 0.16 |
| polarC18 | 758.568 | 87.162 | 15.2 | 0.16 |
| polarC18 | 758.568 | 258.5162 | 15.2 | 0.16 |
| polarC18 | 758.568 | 191.5173 | 15.2 | 0.16 |
| polarC18 | 758.568 | 391.5767 | 15.2 | 0.16 |
| polarC18 | 758.568 | 109.4141 | 15.2 | 0.16 |
| polarC18 | 758.568 | 85.0931 | 15.2 | 0.16 |
| polarC18 | 758.568 | 155.1638 | 15.2 | 0.16 |
| polarC18 | 758.568 | 120.9827 | 15.2 | 0.16 |
| polarC18 | 758.568 | 157.7217 | 15.2 | 0.16 |
| polarC18 | 758.568 | 100.9208 | 15.2 | 0.16 |
| polarC18 | 758.568 | 107.8583 | 15.2 | 0.16 |
| polarC18 | 758.568 | 161.0794 | 15.2 | 0.16 |
| polarC18 | 758.568 | 771.2422 | 15.2 | 0.16 |
| polarC18 | 758.568 | 753.1111 | 15.2 | 0.16 |
| polarC18 | 758.568 | 134.1658 | 15.2 | 0.16 |
| polarC18 | 758.568 | 81.6905 | 15.2 | 0.16 |
| polarC18 | 758.568 | 453.7681 | 15.2 | 0.16 |
| polarC18 | 758.568 | 155.1522 | 15.2 | 0.16 |
| polarC18 | 758.568 | 85.8444 | 15.2 | 0.16 |
| polarC18 | 758.568 | 83.7799 | 15.2 | 0.16 |
| polarC18 | 758.568 | 195.3857 | 15.2 | 0.16 |
| polarC18 | 758.568 | 459.024 | 15.2 | 0.16 |
| polarC18 | 758.568 | 156.7603 | 15.2 | 0.16 |
| polarC18 | 758.568 | 131.4792 | 15.2 | 0.16 |
| polarC18 | 758.568 | 97.7597 | 15.2 | 0.16 |
| polarC18 | 758.568 | 416.7569 | 15.1 | 0.16 |
| polarC18 | 758.568 | 405.1443 | 15.1 | 0.16 |
| polarC18 | 758.568 | 205.0093 | 15.1 | 0.16 |
| polarC18 | 758.568 | 170.716 | 15.1 | 0.16 |

| | | | | |
|---|---|---|---|---|
| polarC18 | 758.568 | 151.2913 | 15.1 | 0.16 |
| polarC18 | 758.568 | 101.7558 | 15.1 | 0.16 |
| polarC18 | 758.568 | 758.2371 | 15.1 | 0.16 |
| polarC18 | 758.568 | 111.0704 | 15.1 | 0.16 |
| polarC18 | 758.568 | 163.4575 | 15.1 | 0.16 |
| polarC18 | 758.568 | 371.3964 | 15.1 | 0.16 |
| polarC18 | 758.568 | 82.2983 | 15.1 | 0.16 |
| polarC18 | 758.568 | 116.3829 | 15.1 | 0.16 |
| polarC18 | 758.568 | 113.1022 | 15.1 | 0.16 |
| polarC18 | 758.568 | 192.886 | 15.1 | 0.16 |
| polarC18 | 758.568 | 122.5632 | 15.1 | 0.16 |
| polarC18 | 758.568 | 134.8438 | 15.1 | 0.16 |
| polarC18 | 758.568 | 101.7857 | 15.1 | 0.16 |
| polarC18 | 758.568 | 249.6223 | 15.1 | 0.16 |
| polarC18 | 758.568 | 112.2904 | 15.1 | 0.16 |
| polarC18 | 758.568 | 308.0008 | 15.1 | 0.16 |
| polarC18 | 758.568 | 80.6283 | 15.1 | 0.16 |
| polarC18 | 758.568 | 162.5356 | 15.1 | 0.16 |
| polarC18 | 758.568 | 580.5438 | 15.1 | 0.16 |
| polarC18 | 758.568 | 222.4075 | 15.1 | 0.16 |
| polarC18 | 758.568 | 213.6749 | 15.1 | 0.16 |
| polarC18 | 758.568 | 143.3295 | 15.1 | 0.16 |
| polarC18 | 758.568 | 329.8647 | 15.1 | 0.16 |
| polarC18 | 758.568 | 93.9993 | 15.1 | 0.16 |
| polarC18 | 758.568 | 101.4501 | 15.1 | 0.16 |
| polarC18 | 758.568 | 83.4951 | 15.1 | 0.16 |
| polarC18 | 758.568 | 206.801 | 15.1 | 0.16 |
| polarC18 | 758.568 | 277.5945 | 15.1 | 0.16 |
| polarC18 | 758.568 | 308.1434 | 15.1 | 0.16 |
| polarC18 | 758.568 | 163.5042 | 15.1 | 0.16 |
| polarC18 | 758.568 | 294.3035 | 15.1 | 0.16 |
| polarC18 | 758.568 | 403.4116 | 15.1 | 0.16 |
| polarC18 | 758.568 | 442.6602 | 15.1 | 0.16 |
| polarC18 | 758.568 | 297.0984 | 15.1 | 0.16 |
| polarC18 | 758.568 | 98.3526 | 15.1 | 0.16 |
| polarC18 | 758.568 | 128.2622 | 15.1 | 0.16 |
| polarC18 | 758.568 | 700.6075 | 15.1 | 0.16 |
| polarC18 | 758.568 | 122.0988 | 15.1 | 0.16 |
| polarC18 | 758.568 | 396.1728 | 15.1 | 0.16 |
| polarC18 | 758.568 | 266.469 | 15.1 | 0.16 |
| polarC18 | 758.568 | 135.8704 | 15.1 | 0.16 |
| polarC18 | 758.568 | 151.3809 | 15.1 | 0.16 |

| | | | | |
|---|---|---|---|---|
| polarC18 | 758.568 | 96.1405 | 15.1 | 0.16 |
| polarC18 | 758.568 | 100.881 | 15.1 | 0.16 |
| polarC18 | 758.568 | 161.1292 | 15.1 | 0.16 |
| polarC18 | 758.568 | 313.7209 | 15.1 | 0.16 |
| polarC18 | 758.568 | 326.4106 | 15.1 | 0.16 |
| polarC18 | 758.568 | 601.4636 | 15.1 | 0.16 |
| polarC18 | 758.568 | 95.9618 | 15.1 | 0.16 |
| polarC18 | 758.568 | 173.0487 | 15.1 | 0.16 |
| polarC18 | 758.568 | 572.8482 | 15.1 | 0.16 |
| polarC18 | 758.568 | 124.8524 | 15.1 | 0.16 |
| polarC18 | 758.568 | 144.9881 | 15.1 | 0.16 |
| polarC18 | 758.568 | 214.4499 | 15.1 | 0.16 |
| polarC18 | 758.568 | 93.6581 | 15.1 | 0.16 |
| polarC18 | 758.568 | 120.3511 | 15.1 | 0.16 |
| polarC18 | 758.568 | 117.2564 | 15.1 | 0.16 |
| polarC18 | 758.568 | 481.0101 | 15.1 | 0.16 |
| polarC18 | 758.568 | 103.5574 | 15.1 | 0.16 |
| polarC18 | 758.568 | 250.9483 | 15.1 | 0.16 |
| polarC18 | 758.568 | 272.3339 | 15.1 | 0.16 |
| polarC18 | 758.568 | 177.7933 | 15.1 | 0.16 |
| polarC18 | 758.568 | 149.4058 | 15.1 | 0.16 |
| polarC18 | 758.568 | 321.186 | 15.1 | 0.16 |
| polarC18 | 758.568 | 100.1766 | 15.1 | 0.16 |
| polarC18 | 758.568 | 87.9833 | 15.1 | 0.16 |
| polarC18 | 758.568 | 93.6937 | 15.1 | 0.16 |
| polarC18 | 758.568 | 380.9242 | 15.1 | 0.16 |
| polarC18 | 758.568 | 177.406 | 15.1 | 0.16 |
| polarC18 | 758.568 | 119.581 | 15.1 | 0.16 |
| polarC18 | 758.568 | 81.9117 | 15.1 | 0.16 |
| polarC18 | 758.568 | 100.2112 | 15.1 | 0.16 |
| polarC18 | 758.568 | 232.3535 | 15.1 | 0.16 |
| polarC18 | 758.568 | 177.6354 | 15.1 | 0.16 |
| polarC18 | 758.568 | 84.1121 | 15.1 | 0.16 |
| polarC18 | 758.568 | 105.4151 | 15.1 | 0.16 |
| polarC18 | 758.568 | 497.2267 | 15.1 | 0.16 |
| polarC18 | 758.568 | 93.3348 | 15 | 0.16 |
| polarC18 | 758.568 | 81.4692 | 15 | 0.16 |
| polarC18 | 758.568 | 84.4877 | 15 | 0.16 |
| polarC18 | 758.568 | 181.6153 | 15 | 0.16 |
| polarC18 | 758.568 | 356.1592 | 15 | 0.16 |
| polarC18 | 758.568 | 87.8843 | 15 | 0.16 |
| polarC18 | 758.568 | 290.6 | 15 | 0.16 |

| | | | | |
|---|---|---|---|---|
| polarC18 | 758.568 | 121.6883 | 15 | 0.16 |
| polarC18 | 758.568 | 99.4403 | 15 | 0.16 |
| polarC18 | 758.568 | 460.0391 | 15 | 0.16 |
| polarC18 | 758.568 | 231.7219 | 15 | 0.16 |
| polarC18 | 758.568 | 469.8718 | 15 | 0.16 |
| polarC18 | 758.568 | 224.7542 | 15 | 0.16 |
| polarC18 | 758.568 | 243.8485 | 15 | 0.16 |
| polarC18 | 758.568 | 96.7924 | 15 | 0.16 |
| polarC18 | 758.568 | 132.8858 | 15 | 0.16 |
| polarC18 | 758.568 | 694.308 | 15 | 0.16 |
| polarC18 | 758.568 | 274.3204 | 15 | 0.16 |
| polarC18 | 758.568 | 111.8901 | 15 | 0.16 |
| polarC18 | 758.568 | 417.6259 | 15 | 0.16 |
| polarC18 | 758.568 | 101.3816 | 15 | 0.16 |
| polarC18 | 758.568 | 85.2801 | 15 | 0.16 |
| polarC18 | 758.568 | 189.8764 | 15 | 0.16 |
| polarC18 | 758.568 | 302.4014 | 15 | 0.16 |
| polarC18 | 758.568 | 96.0453 | 15 | 0.16 |
| polarC18 | 758.568 | 497.0401 | 15 | 0.16 |
| polarC18 | 758.568 | 253.0486 | 15 | 0.16 |
| polarC18 | 758.568 | 183.2808 | 15 | 0.16 |
| polarC18 | 758.568 | 150.5768 | 15 | 0.16 |
| polarC18 | 758.568 | 199.0537 | 15 | 0.16 |
| polarC18 | 758.568 | 384.0949 | 15 | 0.16 |
| polarC18 | 758.568 | 111.6096 | 15 | 0.16 |
| polarC18 | 758.568 | 122.5095 | 15 | 0.16 |
| polarC18 | 758.568 | 530.9853 | 15 | 0.16 |
| polarC18 | 758.568 | 134.3245 | 15 | 0.16 |
| polarC18 | 758.568 | 193.5305 | 15 | 0.16 |
| polarC18 | 758.568 | 213.3841 | 15 | 0.16 |
| polarC18 | 758.568 | 106.1246 | 15 | 0.16 |
| polarC18 | 758.568 | 136.1403 | 15 | 0.16 |
| polarC18 | 758.568 | 92.1054 | 15 | 0.16 |
| polarC18 | 758.568 | 84.0558 | 15 | 0.16 |
| polarC18 | 758.568 | 370.2578 | 15 | 0.16 |
| polarC18 | 758.568 | 499.1862 | 15 | 0.16 |
| polarC18 | 758.568 | 227.6849 | 15 | 0.16 |
| polarC18 | 758.568 | 264.7706 | 15 | 0.16 |
| polarC18 | 758.568 | 255.9022 | 15 | 0.16 |
| polarC18 | 758.568 | 104.9633 | 15 | 0.16 |
| polarC18 | 758.568 | 270.2971 | 15 | 0.16 |
| polarC18 | 758.568 | 132.6476 | 15 | 0.16 |

| | | | | |
|---|---|---|---|---|
| polarC18 | 758.568 | 95.7365 | 15 | 0.16 |
| polarC18 | 758.568 | 163.7939 | 15 | 0.16 |
| polarC18 | 758.568 | 129.3678 | 15 | 0.16 |
| polarC18 | 758.568 | 318.3727 | 15 | 0.16 |
| polarC18 | 758.568 | 170.5597 | 15 | 0.16 |
| polarC18 | 758.568 | 101.9955 | 15 | 0.16 |
| polarC18 | 758.568 | 194.3187 | 15 | 0.16 |
| polarC18 | 758.568 | 450.5225 | 15 | 0.16 |
| polarC18 | 758.568 | 475.0379 | 15 | 0.16 |
| polarC18 | 758.568 | 87.9046 | 15 | 0.16 |
| polarC18 | 758.568 | 313.1261 | 15 | 0.16 |
| polarC18 | 758.568 | 151.7274 | 15 | 0.16 |
| polarC18 | 758.568 | 86.6268 | 15 | 0.16 |
| polarC18 | 758.568 | 711.5798 | 15 | 0.16 |
| polarC18 | 758.568 | 109.492 | 15 | 0.16 |
| polarC18 | 758.568 | 134.7147 | 15 | 0.16 |
| polarC18 | 758.568 | 136.5032 | 15 | 0.16 |
| polarC18 | 758.568 | 115.8005 | 15 | 0.16 |
| polarC18 | 758.568 | 85.1759 | 15 | 0.16 |
| polarC18 | 758.568 | 84.7145 | 15 | 0.16 |
| polarC18 | 758.568 | 165.5084 | 15 | 0.16 |
| polarC18 | 758.568 | 98.5286 | 15 | 0.16 |
| polarC18 | 758.568 | 458.0819 | 15 | 0.16 |
| polarC18 | 758.568 | 92.9675 | 15 | 0.16 |
| polarC18 | 758.568 | 262.4472 | 15 | 0.16 |
| polarC18 | 758.568 | 291.1417 | 15 | 0.16 |
| polarC18 | 758.568 | 159.1202 | 15 | 0.16 |
| polarC18 | 758.568 | 132.4183 | 15 | 0.16 |
| polarC18 | 758.568 | 281.851 | 15 | 0.16 |
| polarC18 | 758.568 | 95.5061 | 15 | 0.16 |
| polarC18 | 758.568 | 130.956 | 15 | 0.16 |
| polarC18 | 758.568 | 349.5373 | 15 | 0.16 |
| polarC18 | 758.568 | 636.8405 | 14.9 | 0.16 |
| polarC18 | 758.568 | 83.2467 | 14.9 | 0.16 |
| polarC18 | 758.568 | 100.0538 | 14.9 | 0.16 |
| polarC18 | 758.568 | 117.4604 | 14.9 | 0.16 |
| polarC18 | 758.568 | 140.7773 | 14.9 | 0.16 |
| polarC18 | 758.568 | 116.8437 | 14.9 | 0.16 |
| polarC18 | 758.568 | 134.4776 | 14.9 | 0.16 |
| polarC18 | 758.568 | 80.4265 | 14.9 | 0.16 |
| polarC18 | 758.568 | 94.8811 | 14.9 | 0.16 |
| polarC18 | 758.568 | 440.202 | 14.9 | 0.16 |

| | | | | |
|---|---|---|---|---|
| polarC18 | 758.568 | 231.3617 | 14.9 | 0.16 |
| polarC18 | 758.568 | 120.0722 | 14.9 | 0.16 |
| polarC18 | 758.568 | 122.2173 | 14.9 | 0.16 |
| polarC18 | 758.568 | 235.8824 | 14.9 | 0.16 |
| polarC18 | 758.568 | 122.7333 | 14.9 | 0.16 |
| polarC18 | 758.568 | 102.9204 | 14.9 | 0.16 |
| polarC18 | 758.568 | 189.6596 | 14.9 | 0.16 |
| polarC18 | 758.568 | 473.4134 | 14.9 | 0.16 |
| polarC18 | 758.568 | 90.1043 | 14.9 | 0.16 |
| polarC18 | 758.568 | 101.972 | 14.9 | 0.16 |
| polarC18 | 758.568 | 194.6642 | 14.9 | 0.16 |
| polarC18 | 758.568 | 318.6586 | 14.9 | 0.16 |
| polarC18 | 758.568 | 263.5221 | 14.9 | 0.16 |
| polarC18 | 758.568 | 432.4783 | 14.9 | 0.16 |
| polarC18 | 758.568 | 136.1668 | 14.9 | 0.16 |
| polarC18 | 758.568 | 125.002 | 14.9 | 0.16 |
| polarC18 | 758.568 | 206.5725 | 14.9 | 0.16 |
| polarC18 | 758.568 | 215.4143 | 14.9 | 0.16 |
| polarC18 | 758.568 | 176.6547 | 14.9 | 0.16 |
| polarC18 | 758.568 | 129.1716 | 14.9 | 0.16 |
| polarC18 | 758.568 | 139.4093 | 14.9 | 0.16 |
| polarC18 | 758.568 | 227.2439 | 14.9 | 0.16 |
| polarC18 | 758.568 | 85.9296 | 14.9 | 0.16 |
| polarC18 | 758.568 | 93.8737 | 14.9 | 0.16 |
| polarC18 | 758.568 | 101.0976 | 14.9 | 0.16 |
| polarC18 | 758.568 | 283.317 | 14.9 | 0.16 |
| polarC18 | 758.568 | 483.1425 | 14.9 | 0.16 |
| polarC18 | 758.568 | 99.667 | 14.9 | 0.16 |
| polarC18 | 758.568 | 116.3544 | 14.9 | 0.16 |
| polarC18 | 758.568 | 93.7114 | 14.9 | 0.16 |
| polarC18 | 758.568 | 83.8406 | 14.9 | 0.16 |
| polarC18 | 758.568 | 96.1496 | 14.9 | 0.16 |
| polarC18 | 758.568 | 439.7482 | 14.9 | 0.16 |
| polarC18 | 758.568 | 534.9133 | 14.9 | 0.16 |
| polarC18 | 758.568 | 94.1218 | 14.9 | 0.16 |
| polarC18 | 758.568 | 148.6324 | 14.9 | 0.16 |
| polarC18 | 758.568 | 331.2347 | 14.9 | 0.16 |
| polarC18 | 758.568 | 168.9875 | 14.9 | 0.16 |
| polarC18 | 758.568 | 210.4802 | 14.9 | 0.16 |
| polarC18 | 758.568 | 397.1814 | 14.9 | 0.16 |
| polarC18 | 758.568 | 89.2419 | 14.9 | 0.16 |
| polarC18 | 758.568 | 146.2322 | 14.9 | 0.16 |

| | | | | |
|---|---|---|---|---|
| polarC18 | 758.568 | 175.6916 | 14.9 | 0.16 |
| polarC18 | 758.568 | 177.3642 | 14.9 | 0.16 |
| polarC18 | 758.568 | 90.6034 | 14.9 | 0.16 |
| polarC18 | 758.568 | 94.1604 | 14.9 | 0.16 |
| polarC18 | 758.568 | 113.9747 | 14.9 | 0.16 |
| polarC18 | 758.568 | 333.3806 | 14.9 | 0.16 |
| polarC18 | 758.568 | 420.4113 | 14.9 | 0.16 |
| polarC18 | 758.568 | 162.488 | 14.9 | 0.16 |
| polarC18 | 758.568 | 272.9558 | 14.9 | 0.16 |
| polarC18 | 758.568 | 362.4296 | 14.9 | 0.16 |
| polarC18 | 758.568 | 733.1101 | 14.9 | 0.16 |
| polarC18 | 758.568 | 129.3902 | 14.9 | 0.16 |
| polarC18 | 758.568 | 89.9636 | 14.9 | 0.16 |
| polarC18 | 758.568 | 286.9182 | 14.9 | 0.16 |
| polarC18 | 758.568 | 144.4697 | 14.9 | 0.16 |
| polarC18 | 758.568 | 402.3622 | 14.9 | 0.16 |
| polarC18 | 758.568 | 80.5189 | 14.9 | 0.16 |
| polarC18 | 758.568 | 82.4179 | 14.9 | 0.16 |
| polarC18 | 758.568 | 93.0991 | 14.9 | 0.16 |
| polarC18 | 758.568 | 309.1203 | 14.9 | 0.16 |
| polarC18 | 758.568 | 86.2142 | 14.9 | 0.16 |
| polarC18 | 758.568 | 166.7131 | 14.9 | 0.16 |
| polarC18 | 758.568 | 150.8191 | 14.9 | 0.16 |
| polarC18 | 758.568 | 357.5919 | 14.9 | 0.16 |
| polarC18 | 758.568 | 170.3534 | 14.9 | 0.16 |
| polarC18 | 758.568 | 85.3858 | 14.9 | 0.16 |
| polarC18 | 758.568 | 208.1789 | 14.9 | 0.16 |
| polarC18 | 758.568 | 118.0061 | 14.8 | 0.16 |
| polarC18 | 758.568 | 121.0177 | 14.8 | 0.16 |
| polarC18 | 758.568 | 93.6154 | 14.8 | 0.16 |
| polarC18 | 758.568 | 83.2615 | 14.8 | 0.16 |
| polarC18 | 758.568 | 88.6735 | 14.8 | 0.16 |
| polarC18 | 758.568 | 81.6855 | 14.8 | 0.16 |
| polarC18 | 758.568 | 97.4792 | 14.8 | 0.16 |
| polarC18 | 758.568 | 220.8707 | 14.8 | 0.16 |
| polarC18 | 758.568 | 247.4204 | 14.8 | 0.16 |
| polarC18 | 758.568 | 644.3217 | 14.8 | 0.16 |
| polarC18 | 758.568 | 565.9174 | 14.8 | 0.16 |
| polarC18 | 758.568 | 84.7184 | 14.8 | 0.16 |
| polarC18 | 758.568 | 246.0627 | 14.8 | 0.16 |
| polarC18 | 758.568 | 194.9119 | 14.8 | 0.16 |
| polarC18 | 758.568 | 147.0488 | 14.8 | 0.16 |

| | | | | |
|---|---|---|---|---|
| polarC18 | 758.568 | 97.7037 | 14.8 | 0.16 |
| polarC18 | 758.568 | 179.0716 | 14.8 | 0.16 |
| polarC18 | 758.568 | 165.9617 | 14.8 | 0.16 |
| polarC18 | 758.568 | 146.9205 | 14.8 | 0.16 |
| polarC18 | 758.568 | 94.9298 | 14.8 | 0.16 |
| polarC18 | 758.568 | 82.9819 | 14.8 | 0.16 |
| polarC18 | 758.568 | 594.9128 | 14.8 | 0.16 |
| polarC18 | 758.568 | 202.9632 | 14.8 | 0.16 |
| polarC18 | 758.568 | 93.5198 | 14.8 | 0.16 |
| polarC18 | 758.568 | 99.2133 | 14.8 | 0.16 |
| polarC18 | 758.568 | 441.7338 | 14.8 | 0.16 |
| polarC18 | 758.568 | 400.0767 | 14.8 | 0.16 |
| polarC18 | 758.568 | 80.8769 | 14.8 | 0.16 |
| polarC18 | 758.568 | 761.2531 | 14.8 | 0.16 |
| polarC18 | 758.568 | 85.4999 | 14.8 | 0.16 |
| polarC18 | 758.568 | 99.104 | 14.8 | 0.16 |
| polarC18 | 758.568 | 247.448 | 14.8 | 0.16 |
| polarC18 | 758.568 | 94.004 | 14.8 | 0.16 |
| polarC18 | 758.568 | 120.2176 | 14.8 | 0.16 |
| polarC18 | 758.568 | 199.5493 | 14.8 | 0.16 |
| polarC18 | 758.568 | 120.6559 | 14.8 | 0.16 |
| polarC18 | 758.568 | 112.0695 | 14.8 | 0.16 |
| polarC18 | 758.568 | 128.189 | 14.8 | 0.16 |
| polarC18 | 758.568 | 291.5818 | 14.8 | 0.16 |
| polarC18 | 758.568 | 88.2606 | 14.8 | 0.16 |
| polarC18 | 758.568 | 93.495 | 14.8 | 0.16 |
| polarC18 | 758.568 | 87.8703 | 14.8 | 0.16 |
| polarC18 | 758.568 | 183.3108 | 14.8 | 0.16 |
| polarC18 | 758.568 | 80.5148 | 14.8 | 0.16 |
| polarC18 | 758.568 | 127.4622 | 14.8 | 0.16 |
| polarC18 | 758.568 | 98.83 | 14.8 | 0.16 |
| polarC18 | 758.568 | 336.1583 | 14.8 | 0.16 |
| polarC18 | 758.568 | 107.265 | 14.8 | 0.16 |
| polarC18 | 758.568 | 82.148 | 14.8 | 0.16 |
| polarC18 | 758.568 | 166.8375 | 14.8 | 0.16 |
| polarC18 | 758.568 | 205.8576 | 14.8 | 0.16 |
| polarC18 | 758.568 | 106.4716 | 14.8 | 0.16 |
| polarC18 | 758.568 | 212.1951 | 14.8 | 0.16 |
| polarC18 | 758.568 | 85.646 | 14.8 | 0.16 |
| polarC18 | 758.568 | 81.5761 | 14.8 | 0.16 |
| polarC18 | 758.568 | 96.4818 | 14.8 | 0.16 |
| polarC18 | 758.568 | 366.6976 | 14.8 | 0.16 |

| | | | | |
|---|---|---|---|---|
| polarC18 | 758.568 | 91.4436 | 14.8 | 0.16 |
| polarC18 | 758.568 | 85.8138 | 14.8 | 0.16 |
| polarC18 | 758.568 | 127.0415 | 14.8 | 0.16 |
| polarC18 | 758.568 | 242.2467 | 14.8 | 0.16 |
| polarC18 | 758.568 | 84.3694 | 14.8 | 0.16 |
| polarC18 | 758.568 | 140.8852 | 14.8 | 0.16 |
| polarC18 | 758.568 | 183.415 | 14.8 | 0.16 |
| polarC18 | 758.568 | 89.1347 | 14.8 | 0.16 |
| polarC18 | 758.568 | 85.0704 | 14.8 | 0.16 |
| polarC18 | 758.568 | 209.6939 | 14.8 | 0.16 |
| polarC18 | 758.568 | 88.2534 | 14.8 | 0.16 |
| polarC18 | 758.568 | 159.3059 | 14.8 | 0.16 |
| polarC18 | 758.568 | 201.4102 | 14.8 | 0.16 |
| polarC18 | 758.568 | 197.6146 | 14.7 | 0.16 |
| polarC18 | 758.568 | 103.1814 | 14.7 | 0.16 |
| polarC18 | 758.568 | 108.0095 | 14.7 | 0.16 |
| polarC18 | 758.568 | 142.7523 | 14.7 | 0.16 |
| polarC18 | 758.568 | 282.1805 | 14.7 | 0.16 |
| polarC18 | 758.568 | 82.4554 | 14.7 | 0.16 |
| polarC18 | 758.568 | 107.5923 | 14.7 | 0.16 |
| polarC18 | 758.568 | 210.2482 | 14.7 | 0.16 |
| polarC18 | 758.568 | 106.9015 | 14.7 | 0.16 |
| polarC18 | 758.568 | 83.9053 | 14.7 | 0.16 |
| polarC18 | 758.568 | 118.1284 | 14.7 | 0.16 |
| polarC18 | 758.568 | 248.2767 | 14.7 | 0.16 |
| polarC18 | 758.568 | 195.317 | 14.7 | 0.16 |
| polarC18 | 758.568 | 217.2133 | 14.7 | 0.16 |
| polarC18 | 758.568 | 92.5098 | 14.7 | 0.16 |
| polarC18 | 758.568 | 115.9817 | 14.7 | 0.16 |
| polarC18 | 758.568 | 219.1757 | 14.7 | 0.16 |
| polarC18 | 758.568 | 320.661 | 14.7 | 0.16 |
| polarC18 | 758.568 | 81.6373 | 14.7 | 0.16 |
| polarC18 | 758.568 | 286.0501 | 14.7 | 0.16 |
| polarC18 | 758.568 | 225.1253 | 14.7 | 0.16 |
| polarC18 | 758.568 | 105.0642 | 14.7 | 0.16 |
| polarC18 | 758.568 | 187.1736 | 14.7 | 0.16 |
| polarC18 | 758.568 | 264.2318 | 14.7 | 0.16 |
| polarC18 | 758.568 | 167.1301 | 14.7 | 0.16 |
| polarC18 | 758.568 | 99.4239 | 14.7 | 0.16 |
| polarC18 | 758.568 | 92.9753 | 14.7 | 0.16 |
| polarC18 | 758.568 | 92.1512 | 14.7 | 0.16 |
| polarC18 | 758.568 | 194.1816 | 14.7 | 0.16 |

| | | | | |
|---|---|---|---|---|
| polarC18 | 758.568 | 231.4954 | 14.7 | 0.16 |
| polarC18 | 758.568 | 256.2717 | 14.7 | 0.16 |
| polarC18 | 758.568 | 399.9673 | 14.7 | 0.16 |
| polarC18 | 758.568 | 101.0911 | 14.7 | 0.16 |
| polarC18 | 758.568 | 223.9738 | 14.7 | 0.16 |
| polarC18 | 758.568 | 111.0359 | 14.7 | 0.16 |
| polarC18 | 758.568 | 88.8704 | 14.7 | 0.16 |
| polarC18 | 758.568 | 151.0683 | 14.7 | 0.16 |
| polarC18 | 758.568 | 128.7742 | 14.7 | 0.16 |
| polarC18 | 758.568 | 249.5218 | 14.7 | 0.16 |
| polarC18 | 758.568 | 318.8633 | 14.7 | 0.16 |
| polarC18 | 758.568 | 118.6025 | 14.7 | 0.16 |
| polarC18 | 758.568 | 288.8464 | 14.7 | 0.16 |
| polarC18 | 758.568 | 90.2522 | 14.7 | 0.16 |
| polarC18 | 758.568 | 174.5117 | 14.7 | 0.16 |
| polarC18 | 758.568 | 96.0225 | 14.7 | 0.16 |
| polarC18 | 758.568 | 132.0202 | 14.7 | 0.16 |
| polarC18 | 758.568 | 141.2639 | 14.7 | 0.16 |
| polarC18 | 758.568 | 501.8244 | 14.7 | 0.16 |
| polarC18 | 758.568 | 113.8902 | 14.7 | 0.16 |
| polarC18 | 758.568 | 114.257 | 14.7 | 0.16 |
| polarC18 | 758.568 | 258.193 | 14.7 | 0.16 |
| polarC18 | 758.568 | 352.2518 | 14.7 | 0.16 |
| polarC18 | 758.568 | 139.7417 | 14.7 | 0.16 |
| polarC18 | 758.568 | 154.8886 | 14.7 | 0.16 |
| polarC18 | 758.568 | 310.1134 | 14.7 | 0.16 |
| polarC18 | 758.568 | 87.821 | 14.7 | 0.16 |
| polarC18 | 758.568 | 258.486 | 14.7 | 0.16 |
| polarC18 | 758.568 | 357.4959 | 14.7 | 0.16 |
| polarC18 | 758.568 | 136.7819 | 14.7 | 0.16 |
| polarC18 | 758.568 | 118.6604 | 14.7 | 0.16 |
| polarC18 | 758.568 | 152.4434 | 14.7 | 0.16 |
| polarC18 | 758.568 | 288.4395 | 14.7 | 0.16 |
| polarC18 | 758.568 | 370.4572 | 14.7 | 0.16 |
| polarC18 | 758.568 | 568.5302 | 14.7 | 0.16 |
| polarC18 | 758.568 | 106.0091 | 14.7 | 0.16 |
| polarC18 | 758.568 | 93.8326 | 14.7 | 0.16 |
| polarC18 | 758.568 | 341.013 | 14.7 | 0.16 |
| polarC18 | 758.568 | 182.7731 | 14.7 | 0.16 |
| polarC18 | 758.568 | 313.0191 | 14.6 | 0.16 |
| polarC18 | 758.568 | 173.9484 | 14.6 | 0.16 |
| polarC18 | 758.568 | 269.8375 | 14.6 | 0.16 |

| | | | | |
|---|---|---|---|---|
| polarC18 | 758.568 | 218.3515 | 14.6 | 0.16 |
| polarC18 | 758.568 | 153.2124 | 14.6 | 0.16 |
| polarC18 | 758.568 | 158.7473 | 14.6 | 0.16 |
| polarC18 | 758.568 | 99.9135 | 14.6 | 0.16 |
| polarC18 | 758.568 | 108.0546 | 14.6 | 0.16 |
| polarC18 | 758.568 | 139.7697 | 14.6 | 0.16 |
| polarC18 | 758.568 | 449.3777 | 14.6 | 0.16 |
| polarC18 | 758.568 | 109.5092 | 14.6 | 0.16 |
| polarC18 | 758.568 | 114.5009 | 14.6 | 0.16 |
| polarC18 | 758.568 | 90.539 | 14.6 | 0.16 |
| polarC18 | 758.568 | 186.0101 | 14.6 | 0.16 |
| polarC18 | 758.568 | 369.3583 | 14.6 | 0.16 |
| polarC18 | 758.568 | 418.8552 | 14.6 | 0.16 |
| polarC18 | 758.568 | 126.9001 | 14.6 | 0.16 |
| polarC18 | 758.568 | 137.5801 | 14.6 | 0.16 |
| polarC18 | 758.568 | 129.4258 | 14.6 | 0.16 |
| polarC18 | 758.568 | 102.3748 | 14.6 | 0.16 |
| polarC18 | 758.568 | 474.3215 | 14.6 | 0.16 |
| polarC18 | 758.568 | 184.9003 | 14.6 | 0.16 |
| polarC18 | 758.568 | 231.9552 | 14.6 | 0.16 |
| polarC18 | 758.568 | 264.6107 | 14.6 | 0.16 |
| polarC18 | 758.568 | 335.4495 | 14.6 | 0.16 |
| polarC18 | 758.568 | 91.4077 | 14.6 | 0.16 |
| polarC18 | 758.568 | 103.1993 | 14.6 | 0.16 |
| polarC18 | 758.568 | 567.3521 | 14.6 | 0.16 |
| polarC18 | 758.568 | 154.7259 | 14.6 | 0.16 |
| polarC18 | 758.568 | 268.808 | 14.6 | 0.16 |
| polarC18 | 758.568 | 126.0329 | 14.6 | 0.16 |
| polarC18 | 758.568 | 83.9255 | 14.6 | 0.16 |
| polarC18 | 758.568 | 269.3225 | 14.6 | 0.16 |
| polarC18 | 758.568 | 100.0769 | 14.6 | 0.16 |
| polarC18 | 758.568 | 175.8369 | 14.6 | 0.16 |
| polarC18 | 758.568 | 302.9878 | 14.6 | 0.16 |
| polarC18 | 758.568 | 116.9277 | 14.6 | 0.16 |
| polarC18 | 758.568 | 102.5842 | 14.6 | 0.16 |
| polarC18 | 758.568 | 235.2494 | 14.6 | 0.16 |
| polarC18 | 758.568 | 124.8307 | 14.6 | 0.16 |
| polarC18 | 758.568 | 150.2532 | 14.6 | 0.16 |
| polarC18 | 758.568 | 100.685 | 14.6 | 0.16 |
| polarC18 | 758.568 | 224.5129 | 14.6 | 0.16 |
| polarC18 | 758.568 | 179.3329 | 14.6 | 0.16 |
| polarC18 | 758.568 | 432.0777 | 14.6 | 0.16 |

| | | | | |
|---|---|---|---|---|
| polarC18 | 758.568 | 280.0879 | 14.6 | 0.16 |
| polarC18 | 758.568 | 290.2542 | 14.6 | 0.16 |
| polarC18 | 758.568 | 217.9804 | 14.6 | 0.16 |
| polarC18 | 758.568 | 359.5055 | 14.6 | 0.16 |
| polarC18 | 758.568 | 99.0156 | 14.6 | 0.16 |
| polarC18 | 758.568 | 81.9614 | 14.6 | 0.16 |
| polarC18 | 758.568 | 83.6117 | 14.6 | 0.16 |
| polarC18 | 758.568 | 266.7933 | 14.6 | 0.16 |
| polarC18 | 758.568 | 154.3072 | 14.6 | 0.16 |
| polarC18 | 758.568 | 294.2093 | 14.6 | 0.16 |
| polarC18 | 758.568 | 259.8642 | 14.6 | 0.16 |
| polarC18 | 758.568 | 174.8503 | 14.6 | 0.16 |
| polarC18 | 758.568 | 89.0045 | 14.6 | 0.16 |
| polarC18 | 758.568 | 136.8272 | 14.6 | 0.16 |
| polarC18 | 758.568 | 89.6214 | 14.6 | 0.16 |
| polarC18 | 758.568 | 100.2544 | 14.6 | 0.16 |
| polarC18 | 758.568 | 197.5148 | 14.6 | 0.16 |
| polarC18 | 758.568 | 592.0511 | 14.6 | 0.16 |
| polarC18 | 758.568 | 164.7436 | 14.6 | 0.16 |
| polarC18 | 758.568 | 220.9103 | 14.6 | 0.16 |
| polarC18 | 758.568 | 243.1554 | 14.6 | 0.16 |
| polarC18 | 758.568 | 180.673 | 14.6 | 0.16 |
| polarC18 | 758.568 | 151.71 | 14.6 | 0.16 |
| polarC18 | 758.568 | 164.9957 | 14.6 | 0.16 |
| polarC18 | 758.568 | 119.9412 | 14.6 | 0.16 |
| polarC18 | 758.568 | 133.1215 | 14.6 | 0.16 |
| polarC18 | 758.568 | 126.3446 | 14.6 | 0.16 |
| polarC18 | 758.568 | 129.5297 | 14.5 | 0.16 |
| polarC18 | 758.568 | 240.1331 | 14.5 | 0.16 |
| polarC18 | 758.568 | 83.6768 | 14.5 | 0.16 |
| polarC18 | 758.568 | 211.1478 | 14.5 | 0.16 |
| polarC18 | 758.568 | 408.6339 | 14.5 | 0.16 |
| polarC18 | 758.568 | 134.9004 | 14.5 | 0.16 |
| polarC18 | 758.568 | 126.5132 | 14.5 | 0.16 |
| polarC18 | 758.568 | 92.9786 | 14.5 | 0.16 |
| polarC18 | 758.568 | 101.1598 | 14.5 | 0.16 |
| polarC18 | 758.568 | 90.1353 | 14.5 | 0.16 |
| polarC18 | 758.568 | 128.5821 | 14.5 | 0.16 |
| polarC18 | 758.568 | 317.2208 | 14.5 | 0.16 |
| polarC18 | 758.568 | 361.0402 | 14.5 | 0.16 |
| polarC18 | 758.568 | 123.0075 | 14.5 | 0.16 |
| polarC18 | 758.568 | 95.7962 | 14.5 | 0.16 |

| | | | | |
|---|---|---|---|---|
| polarC18 | 758.568 | 110.3503 | 14.5 | 0.16 |
| polarC18 | 758.568 | 88.3678 | 14.5 | 0.16 |
| polarC18 | 758.568 | 83.7566 | 14.5 | 0.16 |
| polarC18 | 758.568 | 94.074 | 14.5 | 0.16 |
| polarC18 | 758.568 | 156.4973 | 14.5 | 0.16 |
| polarC18 | 758.568 | 213.4729 | 14.5 | 0.16 |
| polarC18 | 758.568 | 149.858 | 14.5 | 0.16 |
| polarC18 | 758.568 | 110.8533 | 14.5 | 0.16 |
| polarC18 | 758.568 | 218.9416 | 14.5 | 0.16 |
| polarC18 | 758.568 | 91.6477 | 14.5 | 0.16 |
| polarC18 | 758.568 | 227.3834 | 14.5 | 0.16 |
| polarC18 | 758.568 | 84.4243 | 14.5 | 0.16 |
| polarC18 | 758.568 | 126.418 | 14.5 | 0.16 |
| polarC18 | 758.568 | 409.9957 | 14.5 | 0.16 |
| polarC18 | 758.568 | 87.5134 | 14.5 | 0.16 |
| polarC18 | 758.568 | 212.2799 | 14.5 | 0.16 |
| polarC18 | 758.568 | 143.476 | 14.5 | 0.16 |
| polarC18 | 758.568 | 111.7014 | 14.5 | 0.16 |
| polarC18 | 758.568 | 132.1311 | 14.5 | 0.16 |
| polarC18 | 758.568 | 130.7583 | 14.5 | 0.16 |
| polarC18 | 758.568 | 91.364 | 14.5 | 0.16 |
| polarC18 | 758.568 | 190.2161 | 14.5 | 0.16 |
| polarC18 | 758.568 | 176.6767 | 14.5 | 0.16 |
| polarC18 | 758.568 | 93.4354 | 14.5 | 0.16 |
| polarC18 | 758.568 | 141.9879 | 14.5 | 0.16 |
| polarC18 | 758.568 | 123.5024 | 14.5 | 0.16 |
| polarC18 | 758.568 | 167.8457 | 14.5 | 0.16 |
| polarC18 | 758.568 | 136.9664 | 14.5 | 0.16 |
| polarC18 | 758.568 | 203.7392 | 14.5 | 0.16 |
| polarC18 | 758.568 | 146.8477 | 14.5 | 0.16 |
| polarC18 | 758.568 | 783.2773 | 14.5 | 0.16 |
| polarC18 | 758.568 | 134.9474 | 14.5 | 0.16 |
| polarC18 | 758.568 | 129.0979 | 14.5 | 0.16 |
| polarC18 | 758.568 | 232.7413 | 14.5 | 0.16 |
| polarC18 | 758.568 | 93.767 | 14.5 | 0.16 |
| polarC18 | 758.568 | 82.69 | 14.5 | 0.16 |
| polarC18 | 758.568 | 121.7337 | 14.5 | 0.16 |
| polarC18 | 758.568 | 274.1556 | 14.5 | 0.16 |
| polarC18 | 758.568 | 262.8716 | 14.5 | 0.16 |
| polarC18 | 758.568 | 261.5473 | 14.5 | 0.16 |
| polarC18 | 758.568 | 83.9412 | 14.5 | 0.16 |
| polarC18 | 758.568 | 101.1964 | 14.5 | 0.16 |

| | | | | |
|---|---|---|---|---|
| polarC18 | 758.568 | 90.273 | 14.5 | 0.16 |
| polarC18 | 758.568 | 106.9494 | 14.5 | 0.16 |
| polarC18 | 758.568 | 86.324 | 14.5 | 0.16 |
| polarC18 | 758.568 | 88.6819 | 14.5 | 0.16 |
| polarC18 | 758.568 | 105.0717 | 14.5 | 0.16 |
| polarC18 | 758.568 | 91.3136 | 14.5 | 0.16 |
| polarC18 | 758.568 | 246.5909 | 14.5 | 0.16 |
| polarC18 | 758.568 | 413.3954 | 14.5 | 0.16 |
| polarC18 | 758.568 | 496.5873 | 14.5 | 0.16 |
| polarC18 | 758.568 | 188.2718 | 14.5 | 0.16 |
| polarC18 | 758.568 | 203.3278 | 14.5 | 0.16 |
| polarC18 | 758.568 | 665.7444 | 14.4 | 0.16 |
| polarC18 | 758.568 | 110.505 | 14.4 | 0.16 |
| polarC18 | 758.568 | 101.0171 | 14.4 | 0.16 |
| polarC18 | 758.568 | 148.4654 | 14.4 | 0.15 |
| polarC18 | 758.568 | 195.5931 | 14.4 | 0.15 |
| polarC18 | 758.568 | 89.756 | 14.4 | 0.15 |
| polarC18 | 758.568 | 205.2273 | 14.4 | 0.15 |
| polarC18 | 758.568 | 99.5379 | 14.4 | 0.15 |
| polarC18 | 758.568 | 304.2708 | 14.4 | 0.15 |
| polarC18 | 758.568 | 85.0593 | 14.4 | 0.15 |
| polarC18 | 758.568 | 83.2742 | 14.4 | 0.15 |
| polarC18 | 758.568 | 97.8475 | 14.4 | 0.15 |
| polarC18 | 758.568 | 249.1189 | 14.4 | 0.15 |
| polarC18 | 758.568 | 88.5906 | 14.4 | 0.15 |
| polarC18 | 758.568 | 89.789 | 14.4 | 0.15 |
| polarC18 | 758.568 | 219.3662 | 14.4 | 0.15 |
| polarC18 | 758.568 | 93.4408 | 14.4 | 0.15 |
| polarC18 | 758.568 | 151.4703 | 14.4 | 0.15 |
| polarC18 | 758.568 | 276.9008 | 14.4 | 0.15 |
| polarC18 | 758.568 | 145.2249 | 14.4 | 0.15 |
| polarC18 | 758.568 | 337.2335 | 14.4 | 0.15 |
| polarC18 | 758.568 | 618.6086 | 14.4 | 0.15 |
| polarC18 | 758.568 | 535.9537 | 14.4 | 0.15 |
| polarC18 | 758.568 | 528.8455 | 14.4 | 0.15 |
| polarC18 | 758.568 | 81.2245 | 14.4 | 0.15 |
| polarC18 | 758.568 | 82.7012 | 14.4 | 0.15 |
| polarC18 | 758.568 | 224.4197 | 14.4 | 0.15 |
| polarC18 | 758.568 | 206.9069 | 14.4 | 0.15 |
| polarC18 | 758.568 | 303.2394 | 14.4 | 0.15 |
| polarC18 | 758.568 | 86.4027 | 14.4 | 0.15 |
| polarC18 | 758.568 | 360.3186 | 14.4 | 0.15 |

| | | | | |
|---|---|---|---|---|
| polarC18 | 758.568 | 127.248 | 14.4 | 0.15 |
| polarC18 | 758.568 | 129.2453 | 14.4 | 0.15 |
| polarC18 | 758.568 | 95.7208 | 14.4 | 0.15 |
| polarC18 | 758.568 | 143.2966 | 14.4 | 0.15 |
| polarC18 | 758.568 | 89.2609 | 14.4 | 0.15 |
| polarC18 | 758.568 | 91.2991 | 14.4 | 0.15 |
| polarC18 | 758.568 | 102.2679 | 14.4 | 0.15 |
| polarC18 | 758.568 | 390.741 | 14.4 | 0.15 |
| polarC18 | 758.568 | 165.2733 | 14.4 | 0.15 |
| polarC18 | 758.568 | 108.935 | 14.4 | 0.15 |
| polarC18 | 758.568 | 132.1848 | 14.4 | 0.15 |
| polarC18 | 758.568 | 174.6465 | 14.4 | 0.15 |
| polarC18 | 758.568 | 180.0898 | 14.4 | 0.15 |
| polarC18 | 758.568 | 101.8769 | 14.4 | 0.15 |
| polarC18 | 758.568 | 517.145 | 14.4 | 0.15 |
| polarC18 | 758.568 | 94.6462 | 14.4 | 0.15 |
| polarC18 | 758.568 | 79.9385 | 14.4 | 0.15 |
| polarC18 | 758.568 | 135.9806 | 14.4 | 0.15 |
| polarC18 | 758.568 | 141.5664 | 14.4 | 0.15 |
| polarC18 | 758.568 | 87.5685 | 14.4 | 0.15 |
| polarC18 | 758.568 | 97.935 | 14.4 | 0.15 |
| polarC18 | 758.568 | 80.7422 | 14.4 | 0.15 |
| polarC18 | 758.568 | 199.9021 | 14.4 | 0.15 |
| polarC18 | 758.568 | 123.3886 | 14.4 | 0.15 |
| polarC18 | 758.568 | 244.8151 | 14.4 | 0.15 |
| polarC18 | 758.568 | 279.0829 | 14.4 | 0.15 |
| polarC18 | 758.568 | 129.1557 | 14.4 | 0.15 |
| polarC18 | 758.568 | 160.6439 | 14.4 | 0.15 |
| polarC18 | 758.568 | 129.879 | 14.3 | 0.15 |
| polarC18 | 758.568 | 257.8085 | 14.3 | 0.15 |
| polarC18 | 758.568 | 195.6781 | 14.3 | 0.15 |
| polarC18 | 758.568 | 236.0845 | 14.3 | 0.15 |
| polarC18 | 758.568 | 187.3387 | 14.3 | 0.15 |
| polarC18 | 758.568 | 177.0721 | 14.3 | 0.15 |
| polarC18 | 758.568 | 81.6408 | 14.3 | 0.15 |
| polarC18 | 758.568 | 343.0945 | 14.3 | 0.15 |
| polarC18 | 758.568 | 100.5228 | 14.3 | 0.15 |
| polarC18 | 758.568 | 84.308 | 14.3 | 0.15 |
| polarC18 | 758.568 | 135.5551 | 14.3 | 0.15 |
| polarC18 | 758.568 | 217.9376 | 14.3 | 0.15 |
| polarC18 | 758.568 | 82.2079 | 14.3 | 0.15 |
| polarC18 | 758.568 | 190.6513 | 14.3 | 0.15 |

| | | | | |
|---|---|---|---|---|
| polarC18 | 758.568 | 97.2716 | 14.3 | 0.15 |
| polarC18 | 758.568 | 155.5911 | 14.3 | 0.15 |
| polarC18 | 758.568 | 82.8947 | 14.3 | 0.15 |
| polarC18 | 758.568 | 262.0703 | 14.3 | 0.15 |
| polarC18 | 758.568 | 219.9598 | 14.3 | 0.15 |
| polarC18 | 758.568 | 295.9723 | 14.3 | 0.15 |
| polarC18 | 758.568 | 163.1608 | 14.3 | 0.15 |
| polarC18 | 758.568 | 188.0242 | 14.3 | 0.15 |
| polarC18 | 758.568 | 404.1138 | 14.3 | 0.15 |
| polarC18 | 758.568 | 216.6919 | 14.3 | 0.15 |
| polarC18 | 758.568 | 166.8251 | 14.3 | 0.15 |
| polarC18 | 758.568 | 131.1977 | 14.3 | 0.15 |
| polarC18 | 758.568 | 141.6331 | 14.3 | 0.15 |
| polarC18 | 758.568 | 110.7464 | 14.3 | 0.15 |
| polarC18 | 758.568 | 124.0235 | 14.3 | 0.15 |
| polarC18 | 758.568 | 89.9319 | 14.3 | 0.15 |
| polarC18 | 758.568 | 84.8036 | 14.3 | 0.15 |
| polarC18 | 758.568 | 102.2915 | 14.3 | 0.15 |
| polarC18 | 758.568 | 90.79 | 14.3 | 0.15 |
| polarC18 | 758.568 | 118.8981 | 14.3 | 0.15 |
| polarC18 | 758.568 | 90.2782 | 14.3 | 0.15 |
| polarC18 | 758.568 | 89.6622 | 14.3 | 0.15 |
| polarC18 | 758.568 | 610.0295 | 14.3 | 0.15 |
| polarC18 | 758.568 | 169.8181 | 14.3 | 0.15 |
| polarC18 | 758.568 | 350.6873 | 14.3 | 0.15 |
| polarC18 | 758.568 | 180.8131 | 14.3 | 0.15 |
| polarC18 | 758.568 | 114.781 | 14.3 | 0.15 |
| polarC18 | 758.568 | 134.8532 | 14.3 | 0.15 |
| polarC18 | 758.568 | 266.2037 | 14.3 | 0.15 |
| polarC18 | 758.568 | 552.1396 | 14.3 | 0.15 |
| polarC18 | 758.568 | 173.5689 | 14.3 | 0.15 |
| polarC18 | 758.568 | 221.1236 | 14.3 | 0.15 |
| polarC18 | 758.568 | 88.748 | 14.3 | 0.15 |
| polarC18 | 758.568 | 129.3394 | 14.3 | 0.15 |
| polarC18 | 758.568 | 142.3807 | 14.3 | 0.15 |
| polarC18 | 758.568 | 318.4803 | 14.2 | 0.15 |
| polarC18 | 758.568 | 243.7343 | 14.2 | 0.15 |
| polarC18 | 758.568 | 127.5516 | 14.2 | 0.15 |
| polarC18 | 758.568 | 136.471 | 14.2 | 0.15 |
| polarC18 | 758.568 | 85.2979 | 14.2 | 0.15 |
| polarC18 | 758.568 | 171.332 | 14.2 | 0.15 |
| polarC18 | 758.568 | 251.3255 | 14.2 | 0.15 |

| | | | | |
|---|---|---|---|---|
| polarC18 | 758.568 | 130.7405 | 14.2 | 0.15 |
| polarC18 | 758.568 | 123.657 | 14.2 | 0.15 |
| polarC18 | 758.568 | 161.6829 | 14.2 | 0.15 |
| polarC18 | 758.568 | 117.4689 | 14.2 | 0.15 |
| polarC18 | 758.568 | 101.2329 | 14.2 | 0.15 |
| polarC18 | 758.568 | 94.6705 | 14.2 | 0.15 |
| polarC18 | 758.568 | 194.8498 | 14.2 | 0.15 |
| polarC18 | 758.568 | 84.3274 | 14.2 | 0.15 |
| polarC18 | 758.568 | 104.6782 | 14.2 | 0.15 |
| polarC18 | 758.568 | 317.2694 | 14.2 | 0.15 |
| polarC18 | 758.568 | 131.136 | 14.2 | 0.15 |
| polarC18 | 758.568 | 177.7272 | 14.2 | 0.15 |
| polarC18 | 758.568 | 180.9211 | 14.2 | 0.15 |
| polarC18 | 758.568 | 380.4435 | 14.2 | 0.15 |
| polarC18 | 758.568 | 143.9185 | 14.2 | 0.15 |
| polarC18 | 758.568 | 161.6359 | 14.2 | 0.15 |
| polarC18 | 758.568 | 109.3871 | 14.2 | 0.15 |
| polarC18 | 758.568 | 83.8896 | 14.2 | 0.15 |
| polarC18 | 758.568 | 90.0006 | 14.2 | 0.15 |
| polarC18 | 758.568 | 83.9551 | 14.2 | 0.15 |
| polarC18 | 758.568 | 109.7448 | 14.2 | 0.15 |
| polarC18 | 758.568 | 596.4286 | 14.2 | 0.15 |
| polarC18 | 758.568 | 94.5986 | 14.2 | 0.15 |
| polarC18 | 758.568 | 371.8312 | 14.2 | 0.15 |
| polarC18 | 758.568 | 261.7313 | 14.2 | 0.15 |
| polarC18 | 758.568 | 138.5875 | 14.2 | 0.15 |
| polarC18 | 758.568 | 147.9085 | 14.2 | 0.15 |
| polarC18 | 758.568 | 115.1644 | 14.2 | 0.15 |
| polarC18 | 758.568 | 146.5152 | 14.2 | 0.15 |
| polarC18 | 758.568 | 131.3871 | 14.2 | 0.15 |
| polarC18 | 758.568 | 246.6788 | 14.2 | 0.15 |
| polarC18 | 758.568 | 197.3893 | 14.1 | 0.15 |
| polarC18 | 758.568 | 156.1618 | 14.1 | 0.15 |
| polarC18 | 758.568 | 90.408 | 14.1 | 0.15 |
| polarC18 | 758.568 | 118.1165 | 14.1 | 0.15 |
| polarC18 | 758.568 | 125.8416 | 14.1 | 0.15 |
| polarC18 | 758.568 | 526.5921 | 14.1 | 0.15 |
| polarC18 | 758.568 | 102.1725 | 14.1 | 0.15 |
| polarC18 | 758.568 | 239.8389 | 14.1 | 0.15 |
| polarC18 | 758.568 | 81.7117 | 14.1 | 0.15 |
| polarC18 | 758.568 | 124.5232 | 14.1 | 0.15 |
| polarC18 | 758.568 | 113.5164 | 14.1 | 0.15 |

| | | | | |
|---|---|---|---|---|
| polarC18 | 758.568 | 214.5333 | 14.1 | 0.15 |
| polarC18 | 758.568 | 133.5781 | 14.1 | 0.15 |
| polarC18 | 758.568 | 129.2758 | 14.1 | 0.15 |
| polarC18 | 758.568 | 121.2902 | 14.1 | 0.15 |
| polarC18 | 758.568 | 112.9882 | 14.1 | 0.15 |
| polarC18 | 758.568 | 188.0889 | 14.1 | 0.15 |
| polarC18 | 758.568 | 143.9306 | 14.1 | 0.15 |
| polarC18 | 758.568 | 94.4549 | 14.1 | 0.15 |
| polarC18 | 758.568 | 92.4538 | 14.1 | 0.15 |
| polarC18 | 758.568 | 95.2392 | 14.1 | 0.15 |
| polarC18 | 758.568 | 90.3707 | 14.1 | 0.15 |
| polarC18 | 758.568 | 86.4713 | 14.1 | 0.15 |
| polarC18 | 758.568 | 477.144 | 14.1 | 0.15 |
| polarC18 | 758.568 | 273.4789 | 14.1 | 0.15 |
| polarC18 | 758.568 | 83.5932 | 14.1 | 0.15 |
| polarC18 | 758.568 | 147.5732 | 14.1 | 0.15 |
| polarC18 | 758.568 | 184.828 | 14.1 | 0.15 |
| polarC18 | 758.568 | 93.4095 | 14.1 | 0.15 |
| polarC18 | 758.568 | 179.6745 | 14.1 | 0.15 |
| polarC18 | 758.568 | 535.1094 | 14.1 | 0.15 |
| polarC18 | 758.568 | 91.8437 | 14.1 | 0.15 |
| polarC18 | 758.568 | 304.6594 | 14.1 | 0.15 |
| polarC18 | 758.568 | 110.5959 | 14.1 | 0.15 |
| polarC18 | 758.568 | 152.2903 | 14.1 | 0.15 |
| polarC18 | 758.568 | 412.0801 | 14.1 | 0.15 |
| polarC18 | 758.568 | 202.2016 | 14 | 0.15 |
| polarC18 | 758.568 | 129.0643 | 14 | 0.15 |
| polarC18 | 758.568 | 133.6043 | 14 | 0.15 |
| polarC18 | 758.568 | 84.5156 | 14 | 0.15 |
| polarC18 | 758.568 | 121.0302 | 14 | 0.15 |
| polarC18 | 758.568 | 155.4217 | 14 | 0.15 |
| polarC18 | 758.568 | 217.3476 | 14 | 0.15 |
| polarC18 | 758.568 | 156.5607 | 14 | 0.15 |
| polarC18 | 758.568 | 90.1968 | 14 | 0.15 |
| polarC18 | 758.568 | 159.7018 | 14 | 0.15 |
| polarC18 | 758.568 | 100.6935 | 14 | 0.15 |
| polarC18 | 758.568 | 253.3654 | 14 | 0.15 |
| polarC18 | 758.568 | 93.0657 | 14 | 0.15 |
| polarC18 | 758.568 | 103.6576 | 14 | 0.15 |
| polarC18 | 758.568 | 286.091 | 14 | 0.15 |
| polarC18 | 758.568 | 178.3595 | 14 | 0.15 |
| polarC18 | 758.568 | 81.6304 | 14 | 0.15 |

| | | | | |
|---|---|---|---|---|
| polarC18 | 758.568 | 406.877 | 14 | 0.15 |
| polarC18 | 758.568 | 161.4584 | 14 | 0.15 |
| polarC18 | 758.568 | 186.3403 | 14 | 0.15 |
| polarC18 | 758.568 | 224.1104 | 14 | 0.15 |
| polarC18 | 758.568 | 447.8404 | 14 | 0.15 |
| polarC18 | 758.568 | 385.6361 | 14 | 0.15 |
| polarC18 | 758.568 | 226.6617 | 14 | 0.15 |
| polarC18 | 758.568 | 408.7856 | 14 | 0.15 |
| polarC18 | 758.568 | 100.8527 | 14 | 0.15 |
| polarC18 | 758.568 | 84.857 | 14 | 0.15 |
| polarC18 | 758.568 | 124.0787 | 14 | 0.15 |
| polarC18 | 758.568 | 91.5045 | 14 | 0.15 |
| polarC18 | 758.568 | 91.0355 | 14 | 0.15 |
| polarC18 | 758.568 | 570.5369 | 14 | 0.15 |
| polarC18 | 758.568 | 379.5299 | 14 | 0.15 |
| polarC18 | 758.568 | 93.6287 | 14 | 0.15 |
| polarC18 | 758.568 | 84.4365 | 14 | 0.15 |
| polarC18 | 758.568 | 320.7982 | 14 | 0.15 |
| polarC18 | 758.568 | 104.8394 | 14 | 0.15 |
| polarC18 | 758.568 | 243.4043 | 14 | 0.15 |
| polarC18 | 758.568 | 129.6666 | 14 | 0.15 |
| polarC18 | 758.568 | 112.2797 | 14 | 0.15 |
| polarC18 | 758.568 | 204.8266 | 13.9 | 0.15 |
| polarC18 | 758.568 | 119.6184 | 13.9 | 0.15 |
| polarC18 | 758.568 | 115.2742 | 13.9 | 0.15 |
| polarC18 | 758.568 | 571.5544 | 13.9 | 0.15 |
| polarC18 | 758.568 | 178.9599 | 13.9 | 0.15 |
| polarC18 | 758.568 | 525.4064 | 13.9 | 0.15 |
| polarC18 | 758.568 | 81.2027 | 13.9 | 0.15 |
| polarC18 | 758.568 | 120.7464 | 13.9 | 0.15 |
| polarC18 | 758.568 | 166.1903 | 13.9 | 0.15 |
| polarC18 | 758.568 | 128.6297 | 13.9 | 0.15 |
| polarC18 | 758.568 | 126.251 | 13.9 | 0.15 |
| polarC18 | 758.568 | 95.209 | 13.9 | 0.15 |
| polarC18 | 758.568 | 278.69 | 13.9 | 0.15 |
| polarC18 | 758.568 | 163.6548 | 13.9 | 0.15 |
| polarC18 | 758.568 | 649.7507 | 13.9 | 0.15 |
| polarC18 | 758.568 | 81.7907 | 13.9 | 0.15 |
| polarC18 | 758.568 | 92.2838 | 13.9 | 0.15 |
| polarC18 | 758.568 | 148.6166 | 13.9 | 0.15 |
| polarC18 | 758.568 | 87.1216 | 13.9 | 0.15 |
| polarC18 | 758.568 | 91.8923 | 13.9 | 0.15 |

| | | | | |
|---|---|---|---|---|
| polarC18 | 758.568 | 203.0434 | 13.9 | 0.15 |
| polarC18 | 758.568 | 153.4566 | 13.9 | 0.15 |
| polarC18 | 758.568 | 505.6577 | 13.9 | 0.15 |
| polarC18 | 758.568 | 114.8505 | 13.9 | 0.15 |
| polarC18 | 758.568 | 128.3371 | 13.9 | 0.15 |
| polarC18 | 758.568 | 273.0588 | 13.9 | 0.15 |
| polarC18 | 758.568 | 135.3372 | 13.9 | 0.15 |
| polarC18 | 758.568 | 123.7312 | 13.9 | 0.15 |
| polarC18 | 758.568 | 91.8339 | 13.9 | 0.15 |
| polarC18 | 758.568 | 117.4887 | 13.9 | 0.15 |
| polarC18 | 758.568 | 206.0947 | 13.9 | 0.15 |
| polarC18 | 758.568 | 105.0068 | 13.9 | 0.15 |
| polarC18 | 758.568 | 252.9042 | 13.9 | 0.15 |
| polarC18 | 758.568 | 106.4494 | 13.9 | 0.15 |
| polarC18 | 758.568 | 86.3392 | 13.9 | 0.15 |
| polarC18 | 758.568 | 369.9306 | 13.9 | 0.15 |
| polarC18 | 758.568 | 88.6537 | 13.9 | 0.15 |
| polarC18 | 758.568 | 140.7576 | 13.8 | 0.15 |
| polarC18 | 758.568 | 85.5425 | 13.8 | 0.15 |
| polarC18 | 758.568 | 95.0495 | 13.8 | 0.15 |
| polarC18 | 758.568 | 97.2243 | 13.8 | 0.15 |
| polarC18 | 758.568 | 448.7718 | 13.8 | 0.15 |
| polarC18 | 758.568 | 123.6357 | 13.8 | 0.15 |
| polarC18 | 758.568 | 182.2481 | 13.8 | 0.15 |
| polarC18 | 758.568 | 567.1292 | 13.8 | 0.15 |
| polarC18 | 758.568 | 149.8254 | 13.8 | 0.15 |
| polarC18 | 758.568 | 110.2639 | 13.8 | 0.15 |
| polarC18 | 758.568 | 88.5417 | 13.8 | 0.15 |
| polarC18 | 758.568 | 239.5657 | 13.8 | 0.15 |
| polarC18 | 758.568 | 211.9791 | 13.8 | 0.15 |
| polarC18 | 758.568 | 184.5529 | 13.8 | 0.15 |
| polarC18 | 758.568 | 207.7258 | 13.8 | 0.15 |
| polarC18 | 758.568 | 223.4029 | 13.8 | 0.15 |
| polarC18 | 758.568 | 121.9208 | 13.8 | 0.15 |
| polarC18 | 758.568 | 166.7312 | 13.8 | 0.15 |
| polarC18 | 758.568 | 86.4472 | 13.8 | 0.15 |
| polarC18 | 758.568 | 499.0111 | 13.8 | 0.15 |
| polarC18 | 758.568 | 88.8364 | 13.8 | 0.15 |
| polarC18 | 758.568 | 185.9857 | 13.8 | 0.15 |
| polarC18 | 758.568 | 87.8255 | 13.8 | 0.15 |
| polarC18 | 758.568 | 86.6006 | 13.8 | 0.15 |
| polarC18 | 758.568 | 95.5018 | 13.8 | 0.15 |

| | | | | |
|---|---|---|---|---|
| polarC18 | 758.568 | 183.5088 | 13.8 | 0.15 |
| polarC18 | 758.568 | 184.4727 | 13.7 | 0.15 |
| polarC18 | 758.568 | 145.837 | 13.7 | 0.15 |
| polarC18 | 758.568 | 172.727 | 13.7 | 0.15 |
| polarC18 | 758.568 | 152.6543 | 13.7 | 0.15 |
| polarC18 | 758.568 | 145.0575 | 13.7 | 0.15 |
| polarC18 | 758.568 | 119.7165 | 13.7 | 0.15 |
| polarC18 | 758.568 | 91.1606 | 13.7 | 0.15 |
| polarC18 | 758.568 | 80.4394 | 13.7 | 0.15 |
| polarC18 | 758.568 | 766.3043 | 13.7 | 0.15 |
| polarC18 | 758.568 | 100.1212 | 13.7 | 0.15 |
| polarC18 | 758.568 | 111.6525 | 13.7 | 0.15 |
| polarC18 | 758.568 | 493.5909 | 13.7 | 0.15 |
| polarC18 | 758.568 | 111.4006 | 13.7 | 0.15 |
| polarC18 | 758.568 | 169.9938 | 13.7 | 0.15 |
| polarC18 | 758.568 | 120.1974 | 13.7 | 0.15 |
| polarC18 | 758.568 | 203.8416 | 13.7 | 0.15 |
| polarC18 | 758.568 | 102.6026 | 13.7 | 0.15 |
| polarC18 | 758.568 | 107.6728 | 13.7 | 0.15 |
| polarC18 | 758.568 | 141.2011 | 13.7 | 0.15 |
| polarC18 | 758.568 | 107.9223 | 13.7 | 0.15 |
| polarC18 | 758.568 | 136.6952 | 13.7 | 0.15 |
| polarC18 | 758.568 | 112.7464 | 13.7 | 0.15 |
| polarC18 | 758.568 | 185.067 | 13.7 | 0.15 |
| polarC18 | 758.568 | 135.9161 | 13.7 | 0.15 |
| polarC18 | 758.568 | 84.8307 | 13.7 | 0.15 |
| polarC18 | 758.568 | 207.6419 | 13.7 | 0.15 |
| polarC18 | 758.568 | 93.1268 | 13.7 | 0.15 |
| polarC18 | 758.568 | 113.66 | 13.7 | 0.15 |
| polarC18 | 758.568 | 128.5956 | 13.7 | 0.15 |
| polarC18 | 758.568 | 549.8418 | 13.7 | 0.15 |
| polarC18 | 758.568 | 541.8533 | 13.7 | 0.15 |
| polarC18 | 758.568 | 118.2937 | 13.7 | 0.15 |
| polarC18 | 758.568 | 314.2861 | 13.7 | 0.15 |
| polarC18 | 758.568 | 175.0263 | 13.7 | 0.15 |
| polarC18 | 758.568 | 120.6377 | 13.7 | 0.15 |
| polarC18 | 758.568 | 114.4469 | 13.7 | 0.15 |
| polarC18 | 758.568 | 110.9793 | 13.7 | 0.15 |
| polarC18 | 758.568 | 189.2649 | 13.6 | 0.15 |
| polarC18 | 758.568 | 136.9755 | 13.6 | 0.15 |
| polarC18 | 758.568 | 254.8284 | 13.6 | 0.15 |
| polarC18 | 758.568 | 169.7482 | 13.6 | 0.15 |

| | | | | |
|---|---|---|---|---|
| polarC18 | 758.568 | 152.781 | 13.6 | 0.15 |
| polarC18 | 758.568 | 426.8532 | 13.6 | 0.15 |
| polarC18 | 758.568 | 247.4855 | 13.6 | 0.15 |
| polarC18 | 758.568 | 767.6375 | 13.6 | 0.15 |
| polarC18 | 758.568 | 230.6079 | 13.6 | 0.15 |
| polarC18 | 758.568 | 141.6692 | 13.6 | 0.15 |
| polarC18 | 758.568 | 92.187 | 13.6 | 0.15 |
| polarC18 | 758.568 | 484.2045 | 13.6 | 0.15 |
| polarC18 | 758.568 | 123.7525 | 13.6 | 0.15 |
| polarC18 | 758.568 | 221.3236 | 13.6 | 0.15 |
| polarC18 | 758.568 | 86.1833 | 13.6 | 0.15 |
| polarC18 | 758.568 | 184.1636 | 13.6 | 0.15 |
| polarC18 | 758.568 | 238.4218 | 13.6 | 0.15 |
| polarC18 | 758.568 | 321.9408 | 13.6 | 0.15 |
| polarC18 | 758.568 | 243.232 | 13.6 | 0.15 |
| polarC18 | 758.568 | 175.8587 | 13.6 | 0.15 |
| polarC18 | 758.568 | 116.4007 | 13.6 | 0.15 |
| polarC18 | 758.568 | 176.8718 | 13.6 | 0.15 |
| polarC18 | 758.568 | 379.3778 | 13.6 | 0.15 |
| polarC18 | 758.568 | 348.6941 | 13.6 | 0.15 |
| polarC18 | 758.568 | 86.1532 | 13.6 | 0.15 |
| polarC18 | 758.568 | 172.6204 | 13.5 | 0.15 |
| polarC18 | 758.568 | 158.9419 | 13.5 | 0.15 |
| polarC18 | 758.568 | 190.5749 | 13.5 | 0.15 |
| polarC18 | 758.568 | 82.9791 | 13.5 | 0.15 |
| polarC18 | 758.568 | 92.9302 | 13.5 | 0.15 |
| polarC18 | 758.568 | 128.4071 | 13.5 | 0.15 |
| polarC18 | 758.568 | 210.2258 | 13.5 | 0.15 |
| polarC18 | 758.568 | 193.8076 | 13.5 | 0.14 |
| polarC18 | 758.568 | 97.1897 | 13.5 | 0.14 |
| polarC18 | 758.568 | 209.2055 | 13.5 | 0.14 |
| polarC18 | 758.568 | 353.4292 | 13.5 | 0.14 |
| polarC18 | 758.568 | 344.8105 | 13.5 | 0.14 |
| polarC18 | 758.568 | 254.4929 | 13.5 | 0.14 |
| polarC18 | 758.568 | 373.6957 | 13.5 | 0.14 |
| polarC18 | 758.568 | 793.6683 | 13.5 | 0.14 |
| polarC18 | 758.568 | 439.993 | 13.5 | 0.14 |
| polarC18 | 758.568 | 95.4681 | 13.4 | 0.14 |
| polarC18 | 758.568 | 139.6193 | 13.4 | 0.14 |
| polarC18 | 758.568 | 242.8633 | 13.4 | 0.14 |
| polarC18 | 758.568 | 183.2377 | 13.4 | 0.14 |
| polarC18 | 758.568 | 80.9045 | 13.4 | 0.14 |

| | | | | |
|---|---|---|---|---|
| polarC18 | 758.568 | 120.2845 | 13.4 | 0.14 |
| polarC18 | 758.568 | 557.7137 | 13.4 | 0.14 |
| polarC18 | 758.568 | 243.8158 | 13.4 | 0.14 |
| polarC18 | 758.568 | 111.1608 | 13.4 | 0.14 |
| polarC18 | 758.568 | 536.7117 | 13.4 | 0.14 |
| polarC18 | 758.568 | 155.9534 | 13.4 | 0.14 |
| polarC18 | 758.568 | 93.8525 | 13.4 | 0.14 |
| polarC18 | 758.568 | 207.9545 | 13.4 | 0.14 |
| polarC18 | 758.568 | 182.0697 | 13.4 | 0.14 |
| polarC18 | 758.568 | 144.8576 | 13.4 | 0.14 |
| polarC18 | 758.568 | 281.9551 | 13.4 | 0.14 |
| polarC18 | 758.568 | 106.769 | 13.4 | 0.14 |
| polarC18 | 758.568 | 102.4955 | 13.4 | 0.14 |
| polarC18 | 758.568 | 765.347 | 13.4 | 0.14 |
| polarC18 | 758.568 | 159.5568 | 13.4 | 0.14 |
| polarC18 | 758.568 | 165.4581 | 13.4 | 0.14 |
| polarC18 | 758.568 | 268.0291 | 13.4 | 0.14 |
| polarC18 | 758.568 | 559.8107 | 13.4 | 0.14 |
| polarC18 | 758.568 | 557.5296 | 13.3 | 0.14 |
| polarC18 | 758.568 | 700.8837 | 13.3 | 0.14 |
| polarC18 | 758.568 | 275.5359 | 13.3 | 0.14 |
| polarC18 | 758.568 | 328.1086 | 13.3 | 0.14 |
| polarC18 | 758.568 | 287.4604 | 13.3 | 0.14 |
| polarC18 | 758.568 | 132.0392 | 13.3 | 0.14 |
| polarC18 | 758.568 | 101.8519 | 13.3 | 0.14 |
| polarC18 | 758.568 | 321.5039 | 13.3 | 0.14 |
| polarC18 | 758.568 | 105.7554 | 13.3 | 0.14 |
| polarC18 | 758.568 | 525.2636 | 13.3 | 0.14 |
| polarC18 | 758.568 | 84.0168 | 13.3 | 0.14 |
| polarC18 | 758.568 | 92.1906 | 13.3 | 0.14 |
| polarC18 | 758.568 | 106.5394 | 13.3 | 0.14 |
| polarC18 | 758.568 | 102.4656 | 13.3 | 0.14 |
| polarC18 | 758.568 | 174.1023 | 13.3 | 0.14 |
| polarC18 | 758.568 | 212.8855 | 13.3 | 0.14 |
| polarC18 | 758.568 | 85.4459 | 13.3 | 0.14 |
| polarC18 | 758.568 | 248.5488 | 13.3 | 0.14 |
| polarC18 | 758.568 | 150.3284 | 13.2 | 0.14 |
| polarC18 | 758.568 | 95.0282 | 13.2 | 0.14 |
| polarC18 | 758.568 | 228.3547 | 13.2 | 0.14 |
| polarC18 | 758.568 | 177.8844 | 13.2 | 0.14 |
| polarC18 | 758.568 | 475.5299 | 13.2 | 0.14 |
| polarC18 | 758.568 | 129.6064 | 13.2 | 0.14 |

| | | | | |
|---|---|---|---|---|
| polarC18 | 758.568 | 101.5442 | 13.2 | 0.14 |
| polarC18 | 758.568 | 87.5841 | 13.2 | 0.14 |
| polarC18 | 758.568 | 420.5717 | 13.2 | 0.14 |
| polarC18 | 758.568 | 148.7567 | 13.2 | 0.14 |
| polarC18 | 758.568 | 82.1007 | 13.2 | 0.14 |
| polarC18 | 758.568 | 156.0515 | 13.2 | 0.14 |
| polarC18 | 758.568 | 368.9663 | 13.2 | 0.14 |
| polarC18 | 758.568 | 82.6501 | 13.2 | 0.14 |
| polarC18 | 758.568 | 92.1806 | 13.2 | 0.14 |
| polarC18 | 758.568 | 98.6965 | 13.1 | 0.14 |
| polarC18 | 758.568 | 128.6361 | 13.1 | 0.14 |
| polarC18 | 758.568 | 112.1895 | 13.1 | 0.14 |
| polarC18 | 758.568 | 373.5388 | 13.1 | 0.14 |
| polarC18 | 758.568 | 118.7145 | 13.1 | 0.14 |
| polarC18 | 758.568 | 530.4362 | 13.1 | 0.14 |
| polarC18 | 758.568 | 142.045 | 13.1 | 0.14 |
| polarC18 | 758.568 | 439.6037 | 13.1 | 0.14 |
| polarC18 | 758.568 | 171.0978 | 13.1 | 0.14 |
| polarC18 | 758.568 | 381.1228 | 13.1 | 0.14 |
| polarC18 | 758.568 | 155.2093 | 13.1 | 0.14 |
| polarC18 | 758.568 | 227.3296 | 13.1 | 0.14 |
| polarC18 | 758.568 | 88.4126 | 13.1 | 0.14 |
| polarC18 | 758.568 | 374.1302 | 13.1 | 0.14 |
| polarC18 | 758.568 | 279.818 | 13.1 | 0.14 |
| polarC18 | 758.568 | 309.7712 | 13.1 | 0.14 |
| polarC18 | 758.568 | 107.8639 | 13.1 | 0.14 |
| polarC18 | 758.568 | 445.2138 | 13.1 | 0.14 |
| polarC18 | 758.568 | 89.4042 | 13 | 0.14 |
| polarC18 | 758.568 | 226.3943 | 13 | 0.14 |
| polarC18 | 758.568 | 99.2876 | 13 | 0.14 |
| polarC18 | 758.568 | 117.4763 | 13 | 0.14 |
| polarC18 | 758.568 | 130.3357 | 13 | 0.14 |
| polarC18 | 758.568 | 92.1945 | 13 | 0.14 |
| polarC18 | 758.568 | 135.9349 | 13 | 0.14 |
| polarC18 | 758.568 | 151.5862 | 13 | 0.14 |
| polarC18 | 758.568 | 341.6284 | 13 | 0.14 |
| polarC18 | 758.568 | 164.2475 | 13 | 0.14 |
| polarC18 | 758.568 | 200.5175 | 13 | 0.14 |
| polarC18 | 758.568 | 88.7955 | 13 | 0.14 |
| polarC18 | 758.568 | 146.4938 | 13 | 0.14 |
| polarC18 | 758.568 | 109.8197 | 13 | 0.14 |
| polarC18 | 758.568 | 129.6584 | 13 | 0.14 |

| | | | | |
|---|---|---|---|---|
| polarC18 | 758.568 | 157.0766 | 13 | 0.14 |
| polarC18 | 758.568 | 93.392 | 13 | 0.14 |
| polarC18 | 758.568 | 150.7059 | 13 | 0.14 |
| polarC18 | 758.568 | 688.585 | 13 | 0.14 |
| polarC18 | 758.568 | 261.7661 | 13 | 0.14 |
| polarC18 | 758.568 | 151.1634 | 13 | 0.14 |
| polarC18 | 758.568 | 83.6321 | 13 | 0.14 |
| polarC18 | 758.568 | 462.1192 | 13 | 0.14 |
| polarC18 | 758.568 | 551.9995 | 13 | 0.14 |
| polarC18 | 758.568 | 187.038 | 13 | 0.14 |
| polarC18 | 758.568 | 602.239 | 12.9 | 0.14 |
| polarC18 | 758.568 | 108.7623 | 12.9 | 0.14 |
| polarC18 | 758.568 | 79.8 | 12.9 | 0.14 |
| polarC18 | 758.568 | 279.5816 | 12.9 | 0.14 |
| polarC18 | 758.568 | 224.9094 | 12.9 | 0.14 |
| polarC18 | 758.568 | 497.4819 | 12.9 | 0.14 |
| polarC18 | 758.568 | 365.6206 | 12.9 | 0.14 |
| polarC18 | 758.568 | 151.0799 | 12.9 | 0.14 |
| polarC18 | 758.568 | 112.4019 | 12.9 | 0.14 |
| polarC18 | 758.568 | 231.7945 | 12.9 | 0.14 |
| polarC18 | 758.568 | 233.6159 | 12.9 | 0.14 |
| polarC18 | 758.568 | 173.3144 | 12.9 | 0.14 |
| polarC18 | 758.568 | 566.9152 | 12.9 | 0.14 |
| polarC18 | 758.568 | 594.0587 | 12.9 | 0.14 |
| polarC18 | 758.568 | 180.1479 | 12.9 | 0.14 |
| polarC18 | 758.568 | 222.3099 | 12.9 | 0.14 |
| polarC18 | 758.568 | 133.6157 | 12.9 | 0.14 |
| polarC18 | 758.568 | 141.8882 | 12.9 | 0.14 |
| polarC18 | 758.568 | 148.7195 | 12.9 | 0.14 |
| polarC18 | 758.568 | 120.6297 | 12.9 | 0.14 |
| polarC18 | 758.568 | 222.6745 | 12.9 | 0.14 |
| polarC18 | 758.568 | 86.8154 | 12.9 | 0.14 |
| polarC18 | 758.568 | 185.8712 | 12.8 | 0.14 |
| polarC18 | 758.568 | 83.813 | 12.8 | 0.14 |
| polarC18 | 758.568 | 103.4549 | 12.8 | 0.14 |
| polarC18 | 758.568 | 213.2739 | 12.8 | 0.14 |
| polarC18 | 758.568 | 228.9306 | 12.8 | 0.14 |
| polarC18 | 758.568 | 736.6785 | 12.8 | 0.14 |
| polarC18 | 758.568 | 749.6639 | 12.8 | 0.14 |
| polarC18 | 758.568 | 91.4707 | 12.8 | 0.14 |
| polarC18 | 758.568 | 138.2865 | 12.8 | 0.14 |
| polarC18 | 758.568 | 100.6757 | 12.8 | 0.14 |

| | | | | |
|---|---|---|---|---|
| polarC18 | 758.568 | 105.2321 | 12.8 | 0.14 |
| polarC18 | 758.568 | 160.7532 | 12.8 | 0.14 |
| polarC18 | 758.568 | 96.2807 | 12.8 | 0.14 |
| polarC18 | 758.568 | 390.9187 | 12.8 | 0.14 |
| polarC18 | 758.568 | 159.0737 | 12.8 | 0.14 |
| polarC18 | 758.568 | 142.7259 | 12.8 | 0.14 |
| polarC18 | 758.568 | 508.3501 | 12.8 | 0.14 |
| polarC18 | 758.568 | 316.8994 | 12.8 | 0.14 |
| polarC18 | 758.568 | 221.6024 | 12.8 | 0.14 |
| polarC18 | 758.568 | 215.8221 | 12.8 | 0.14 |
| polarC18 | 758.568 | 489.4626 | 12.7 | 0.14 |
| polarC18 | 758.568 | 125.464 | 12.7 | 0.14 |
| polarC18 | 758.568 | 180.0066 | 12.7 | 0.14 |
| polarC18 | 758.568 | 655.6404 | 12.7 | 0.14 |
| polarC18 | 758.568 | 186.3089 | 12.7 | 0.14 |
| polarC18 | 758.568 | 109.4296 | 12.7 | 0.14 |
| polarC18 | 758.568 | 412.7337 | 12.7 | 0.14 |
| polarC18 | 758.568 | 289.6025 | 12.7 | 0.14 |
| polarC18 | 758.568 | 331.6545 | 12.7 | 0.14 |
| polarC18 | 758.568 | 348.7809 | 12.7 | 0.14 |
| polarC18 | 758.568 | 93.0352 | 12.7 | 0.14 |
| polarC18 | 758.568 | 98.5425 | 12.7 | 0.14 |
| polarC18 | 758.568 | 98.6445 | 12.7 | 0.14 |
| polarC18 | 758.568 | 171.1671 | 12.7 | 0.14 |
| polarC18 | 758.568 | 102.7477 | 12.7 | 0.14 |
| polarC18 | 758.568 | 103.9638 | 12.7 | 0.14 |
| polarC18 | 758.568 | 357.3669 | 12.7 | 0.14 |
| polarC18 | 758.568 | 127.4926 | 12.7 | 0.14 |
| polarC18 | 758.568 | 158.3111 | 12.7 | 0.14 |
| polarC18 | 758.568 | 164.0813 | 12.7 | 0.14 |
| polarC18 | 758.568 | 484.9058 | 12.7 | 0.14 |
| polarC18 | 758.568 | 271.4134 | 12.7 | 0.14 |
| polarC18 | 758.568 | 471.8481 | 12.7 | 0.14 |
| polarC18 | 758.568 | 89.8317 | 12.6 | 0.14 |
| polarC18 | 758.568 | 139.3698 | 12.6 | 0.14 |
| polarC18 | 758.568 | 111.3819 | 12.6 | 0.14 |
| polarC18 | 758.568 | 222.5908 | 12.6 | 0.14 |
| polarC18 | 758.568 | 379.0462 | 12.6 | 0.14 |
| polarC18 | 758.568 | 175.8891 | 12.6 | 0.14 |
| polarC18 | 758.568 | 113.025 | 12.6 | 0.14 |
| polarC18 | 758.568 | 294.8053 | 12.6 | 0.14 |
| polarC18 | 758.568 | 675.1672 | 12.6 | 0.14 |

| | | | | |
|---|---|---|---|---|
| polarC18 | 758.568 | 276.5703 | 12.6 | 0.14 |
| polarC18 | 758.568 | 285.6679 | 12.6 | 0.14 |
| polarC18 | 758.568 | 169.5059 | 12.6 | 0.14 |
| polarC18 | 758.568 | 200.6705 | 12.6 | 0.14 |
| polarC18 | 758.568 | 85.6306 | 12.6 | 0.14 |
| polarC18 | 758.568 | 359.1033 | 12.6 | 0.13 |
| polarC18 | 758.568 | 125.687 | 12.6 | 0.13 |
| polarC18 | 758.568 | 661.4561 | 12.6 | 0.13 |
| polarC18 | 758.568 | 250.3133 | 12.6 | 0.13 |
| polarC18 | 758.568 | 283.0491 | 12.6 | 0.13 |
| polarC18 | 758.568 | 375.1785 | 12.6 | 0.13 |
| polarC18 | 758.568 | 289.5118 | 12.6 | 0.13 |
| polarC18 | 758.568 | 348.2418 | 12.6 | 0.13 |
| polarC18 | 758.568 | 121.2353 | 12.6 | 0.13 |
| polarC18 | 758.568 | 79.6971 | 12.6 | 0.13 |
| polarC18 | 758.568 | 142.9424 | 12.6 | 0.13 |
| polarC18 | 758.568 | 100.4167 | 12.6 | 0.13 |
| polarC18 | 758.568 | 318.8077 | 12.6 | 0.13 |
| polarC18 | 758.568 | 392.199 | 12.6 | 0.13 |
| polarC18 | 758.568 | 388.515 | 12.5 | 0.13 |
| polarC18 | 758.568 | 107.4703 | 12.5 | 0.13 |
| polarC18 | 758.568 | 217.5976 | 12.5 | 0.13 |
| polarC18 | 758.568 | 573.8054 | 12.5 | 0.13 |
| polarC18 | 758.568 | 186.0622 | 12.5 | 0.13 |
| polarC18 | 758.568 | 198.6898 | 12.5 | 0.13 |
| polarC18 | 758.568 | 209.0126 | 12.5 | 0.13 |
| polarC18 | 758.568 | 134.5282 | 12.5 | 0.13 |
| polarC18 | 758.568 | 678.1868 | 12.5 | 0.13 |
| polarC18 | 758.568 | 109.5154 | 12.5 | 0.13 |
| polarC18 | 758.568 | 98.3136 | 12.5 | 0.13 |
| polarC18 | 758.568 | 100.3429 | 12.5 | 0.13 |
| polarC18 | 758.568 | 151.227 | 12.5 | 0.13 |
| polarC18 | 758.568 | 307.1065 | 12.5 | 0.13 |
| polarC18 | 758.568 | 81.0032 | 12.5 | 0.13 |
| polarC18 | 758.568 | 126.9859 | 12.5 | 0.13 |
| polarC18 | 758.568 | 129.773 | 12.5 | 0.13 |
| polarC18 | 758.568 | 417.8428 | 12.5 | 0.13 |
| polarC18 | 758.568 | 138.7623 | 12.5 | 0.13 |
| polarC18 | 758.568 | 611.3007 | 12.4 | 0.13 |
| polarC18 | 758.568 | 119.674 | 12.4 | 0.13 |
| polarC18 | 758.568 | 364.9207 | 12.4 | 0.13 |
| polarC18 | 758.568 | 366.9352 | 12.4 | 0.13 |

| | | | | |
|---|---|---|---|---|
| polarC18 | 758.568 | 174.0151 | 12.4 | 0.13 |
| polarC18 | 758.568 | 500.3307 | 12.4 | 0.13 |
| polarC18 | 758.568 | 363.7972 | 12.4 | 0.13 |
| polarC18 | 758.568 | 259.1376 | 12.4 | 0.13 |
| polarC18 | 758.568 | 139.4411 | 12.4 | 0.13 |
| polarC18 | 758.568 | 303.7364 | 12.4 | 0.13 |
| polarC18 | 758.568 | 308.5477 | 12.4 | 0.13 |
| polarC18 | 758.568 | 622.3163 | 12.4 | 0.13 |
| polarC18 | 758.568 | 200.4319 | 12.4 | 0.13 |
| polarC18 | 758.568 | 132.3063 | 12.4 | 0.13 |
| polarC18 | 758.568 | 149.0275 | 12.4 | 0.13 |
| polarC18 | 758.568 | 148.7366 | 12.4 | 0.13 |
| polarC18 | 758.568 | 110.7871 | 12.4 | 0.13 |
| polarC18 | 758.568 | 277.6291 | 12.4 | 0.13 |
| polarC18 | 758.568 | 340.7406 | 12.4 | 0.13 |
| polarC18 | 758.568 | 278.876 | 12.3 | 0.13 |
| polarC18 | 758.568 | 300.7851 | 12.3 | 0.13 |
| polarC18 | 758.568 | 502.7575 | 12.3 | 0.13 |
| polarC18 | 758.568 | 601.6758 | 12.3 | 0.13 |
| polarC18 | 758.568 | 180.5199 | 12.3 | 0.13 |
| polarC18 | 758.568 | 91.6551 | 12.3 | 0.13 |
| polarC18 | 758.568 | 87.5901 | 12.3 | 0.13 |
| polarC18 | 758.568 | 163.3089 | 12.3 | 0.13 |
| polarC18 | 758.568 | 567.4857 | 12.3 | 0.13 |
| polarC18 | 758.568 | 496.2151 | 12.3 | 0.13 |
| polarC18 | 758.568 | 422.4605 | 12.3 | 0.13 |
| polarC18 | 758.568 | 647.437 | 12.3 | 0.13 |
| polarC18 | 758.568 | 555.5529 | 12.3 | 0.13 |
| polarC18 | 758.568 | 301.7513 | 12.3 | 0.13 |
| polarC18 | 758.568 | 188.4228 | 12.3 | 0.13 |
| polarC18 | 758.568 | 80.5032 | 12.3 | 0.13 |
| polarC18 | 758.568 | 313.783 | 12.3 | 0.13 |
| polarC18 | 758.568 | 95.3668 | 12.3 | 0.13 |
| polarC18 | 758.568 | 561.8183 | 12.3 | 0.13 |
| polarC18 | 758.568 | 94.756 | 12.3 | 0.13 |
| polarC18 | 758.568 | 158.9997 | 12.3 | 0.13 |
| polarC18 | 758.568 | 189.7827 | 12.3 | 0.13 |
| polarC18 | 758.568 | 128.2233 | 12.3 | 0.13 |
| polarC18 | 758.568 | 127.3808 | 12.3 | 0.13 |
| polarC18 | 758.568 | 95.6108 | 12.3 | 0.13 |
| polarC18 | 758.568 | 95.6284 | 12.3 | 0.13 |
| polarC18 | 758.568 | 103.7843 | 12.3 | 0.13 |

| | | | | |
|---|---|---|---|---|
| polarC18 | 758.568 | 85.7442 | 12.3 | 0.13 |
| polarC18 | 758.568 | 87.9109 | 12.3 | 0.13 |
| polarC18 | 758.568 | 567.7263 | 12.3 | 0.13 |
| polarC18 | 758.568 | 164.1915 | 12.3 | 0.13 |
| polarC18 | 758.568 | 165.6051 | 12.3 | 0.13 |
| polarC18 | 758.568 | 110.2894 | 12.2 | 0.13 |
| polarC18 | 758.568 | 427.6284 | 12.2 | 0.13 |
| polarC18 | 758.568 | 80.1404 | 12.2 | 0.13 |
| polarC18 | 758.568 | 140.7253 | 12.2 | 0.13 |
| polarC18 | 758.568 | 180.3034 | 12.2 | 0.13 |
| polarC18 | 758.568 | 248.2349 | 12.2 | 0.13 |
| polarC18 | 758.568 | 700.0969 | 12.2 | 0.13 |
| polarC18 | 758.568 | 137.2018 | 12.2 | 0.13 |
| polarC18 | 758.568 | 111.3922 | 12.2 | 0.13 |
| polarC18 | 758.568 | 109.7832 | 12.2 | 0.13 |
| polarC18 | 758.568 | 119.0449 | 12.2 | 0.13 |
| polarC18 | 758.568 | 187.4894 | 12.2 | 0.13 |
| polarC18 | 758.568 | 245.512 | 12.2 | 0.13 |
| polarC18 | 758.568 | 80.2205 | 12.2 | 0.13 |
| polarC18 | 758.568 | 118.3256 | 12.2 | 0.13 |
| polarC18 | 758.568 | 240.483 | 12.2 | 0.13 |
| polarC18 | 758.568 | 311.5283 | 12.2 | 0.13 |
| polarC18 | 758.568 | 136.2167 | 12.2 | 0.13 |
| polarC18 | 758.568 | 89.0117 | 12.2 | 0.13 |
| polarC18 | 758.568 | 94.4012 | 12.2 | 0.13 |
| polarC18 | 758.568 | 218.5533 | 12.2 | 0.13 |
| polarC18 | 758.568 | 84.6157 | 12.2 | 0.13 |
| polarC18 | 758.568 | 292.6206 | 12.2 | 0.13 |
| polarC18 | 758.568 | 224.954 | 12.2 | 0.13 |
| polarC18 | 758.568 | 312.8396 | 12.1 | 0.13 |
| polarC18 | 758.568 | 97.6532 | 12.1 | 0.13 |
| polarC18 | 758.568 | 340.5924 | 12.1 | 0.13 |
| polarC18 | 758.568 | 98.4486 | 12.1 | 0.13 |
| polarC18 | 758.568 | 237.6296 | 12.1 | 0.13 |
| polarC18 | 758.568 | 156.2158 | 12.1 | 0.13 |
| polarC18 | 758.568 | 149.5645 | 12.1 | 0.13 |
| polarC18 | 758.568 | 89.8521 | 12.1 | 0.13 |
| polarC18 | 758.568 | 466.5389 | 12.1 | 0.13 |
| polarC18 | 758.568 | 270.4834 | 12.1 | 0.13 |
| polarC18 | 758.568 | 276.9732 | 12.1 | 0.13 |
| polarC18 | 758.568 | 341.4539 | 12.1 | 0.13 |
| polarC18 | 758.568 | 133.4817 | 12.1 | 0.13 |

| | | | | |
|---|---|---|---|---|
| polarC18 | 758.568 | 170.4737 | 12.1 | 0.13 |
| polarC18 | 758.568 | 561.2138 | 12.1 | 0.13 |
| polarC18 | 758.568 | 81.0551 | 12.1 | 0.13 |
| polarC18 | 758.568 | 154.493 | 12.1 | 0.13 |
| polarC18 | 758.568 | 122.2566 | 12.1 | 0.13 |
| polarC18 | 758.568 | 106.3198 | 12.1 | 0.13 |
| polarC18 | 758.568 | 82.0357 | 12.1 | 0.13 |
| polarC18 | 758.568 | 330.0543 | 12.1 | 0.13 |
| polarC18 | 758.568 | 109.2948 | 12.1 | 0.13 |
| polarC18 | 758.568 | 82.0741 | 12.1 | 0.13 |
| polarC18 | 758.568 | 276.9356 | 12.1 | 0.13 |
| polarC18 | 758.568 | 109.9656 | 12.1 | 0.13 |
| polarC18 | 758.568 | 121.8332 | 12.1 | 0.13 |
| polarC18 | 758.568 | 249.2203 | 12.1 | 0.13 |
| polarC18 | 758.568 | 540.788 | 12 | 0.13 |
| polarC18 | 758.568 | 231.1607 | 12 | 0.13 |
| polarC18 | 758.568 | 121.8822 | 12 | 0.13 |
| polarC18 | 758.568 | 131.4434 | 12 | 0.13 |
| polarC18 | 758.568 | 194.4413 | 12 | 0.13 |
| polarC18 | 758.568 | 103.0987 | 12 | 0.13 |
| polarC18 | 758.568 | 90.8129 | 12 | 0.13 |
| polarC18 | 758.568 | 109.9796 | 12 | 0.13 |
| polarC18 | 758.568 | 92.6832 | 12 | 0.13 |
| polarC18 | 758.568 | 175.9172 | 12 | 0.13 |
| polarC18 | 758.568 | 335.5321 | 12 | 0.13 |
| polarC18 | 758.568 | 153.1574 | 12 | 0.13 |
| polarC18 | 758.568 | 403.2708 | 12 | 0.13 |
| polarC18 | 758.568 | 114.0914 | 11.9 | 0.13 |
| polarC18 | 758.568 | 261.5826 | 11.9 | 0.13 |
| polarC18 | 758.568 | 85.7812 | 11.9 | 0.13 |
| polarC18 | 758.568 | 712.3097 | 11.9 | 0.13 |
| polarC18 | 758.568 | 165.8455 | 11.9 | 0.13 |
| polarC18 | 758.568 | 112.6441 | 11.9 | 0.13 |
| polarC18 | 758.568 | 464.0197 | 11.9 | 0.13 |
| polarC18 | 758.568 | 326.7354 | 11.9 | 0.13 |
| polarC18 | 758.568 | 192.5255 | 11.9 | 0.13 |
| polarC18 | 758.568 | 155.7181 | 11.9 | 0.13 |
| polarC18 | 758.568 | 98.1817 | 11.9 | 0.13 |
| polarC18 | 758.568 | 98.2603 | 11.9 | 0.13 |
| polarC18 | 758.568 | 443.5552 | 11.9 | 0.13 |
| polarC18 | 758.568 | 133.8794 | 11.9 | 0.13 |
| polarC18 | 758.568 | 274.4225 | 11.9 | 0.13 |

| | | | | |
|---|---|---|---|---|
| polarC18 | 758.568 | 392.7796 | 11.8 | 0.13 |
| polarC18 | 758.568 | 145.5445 | 11.8 | 0.13 |
| polarC18 | 758.568 | 295.6038 | 11.8 | 0.13 |
| polarC18 | 758.568 | 96.0557 | 11.8 | 0.13 |
| polarC18 | 758.568 | 93.0783 | 11.8 | 0.13 |
| polarC18 | 758.568 | 478.4635 | 11.8 | 0.13 |
| polarC18 | 758.568 | 90.4939 | 11.8 | 0.13 |
| polarC18 | 758.568 | 354.777 | 11.8 | 0.13 |
| polarC18 | 758.568 | 463.517 | 11.8 | 0.13 |
| polarC18 | 758.568 | 194.9893 | 11.8 | 0.13 |
| polarC18 | 758.568 | 496.6962 | 11.8 | 0.13 |
| polarC18 | 758.568 | 101.2744 | 11.7 | 0.13 |
| polarC18 | 758.568 | 129.8905 | 11.7 | 0.13 |
| polarC18 | 758.568 | 107.9508 | 11.7 | 0.13 |
| polarC18 | 758.568 | 86.8744 | 11.7 | 0.13 |
| polarC18 | 758.568 | 304.9024 | 11.7 | 0.13 |
| polarC18 | 758.568 | 199.1097 | 11.7 | 0.13 |
| polarC18 | 758.568 | 126.8772 | 11.7 | 0.13 |
| polarC18 | 758.568 | 467.3642 | 11.7 | 0.13 |
| polarC18 | 758.568 | 87.0116 | 11.7 | 0.13 |
| polarC18 | 758.568 | 106.7389 | 11.7 | 0.13 |
| polarC18 | 758.568 | 530.8663 | 11.7 | 0.13 |
| polarC18 | 758.568 | 160.2802 | 11.7 | 0.13 |
| polarC18 | 758.568 | 141.018 | 11.7 | 0.13 |
| polarC18 | 758.568 | 133.1416 | 11.7 | 0.13 |
| polarC18 | 758.568 | 167.8821 | 11.7 | 0.13 |
| polarC18 | 758.568 | 216.9639 | 11.7 | 0.13 |
| polarC18 | 758.568 | 226.4691 | 11.7 | 0.13 |
| polarC18 | 758.568 | 193.7859 | 11.7 | 0.13 |
| polarC18 | 758.568 | 266.1548 | 11.7 | 0.13 |
| polarC18 | 758.568 | 645.9874 | 11.7 | 0.13 |
| polarC18 | 758.568 | 299.1389 | 11.7 | 0.13 |
| polarC18 | 758.568 | 122.4468 | 11.6 | 0.13 |
| polarC18 | 758.568 | 541.6885 | 11.6 | 0.12 |
| polarC18 | 758.568 | 422.3279 | 11.6 | 0.12 |
| polarC18 | 758.568 | 277.7492 | 11.6 | 0.12 |
| polarC18 | 758.568 | 101.7163 | 11.6 | 0.12 |
| polarC18 | 758.568 | 131.7915 | 11.6 | 0.12 |
| polarC18 | 758.568 | 106.675 | 11.6 | 0.12 |
| polarC18 | 758.568 | 146.1436 | 11.6 | 0.12 |
| polarC18 | 758.568 | 90.5132 | 11.6 | 0.12 |
| polarC18 | 758.568 | 176.9953 | 11.6 | 0.12 |

| | | | | |
|---|---|---|---|---|
| polarC18 | 758.568 | 154.0076 | 11.6 | 0.12 |
| polarC18 | 758.568 | 119.0259 | 11.6 | 0.12 |
| polarC18 | 758.568 | 126.731 | 11.5 | 0.12 |
| polarC18 | 758.568 | 767.9294 | 11.5 | 0.12 |
| polarC18 | 758.568 | 245.2756 | 11.5 | 0.12 |
| polarC18 | 758.568 | 135.6882 | 11.5 | 0.12 |
| polarC18 | 758.568 | 207.5175 | 11.5 | 0.12 |
| polarC18 | 758.568 | 108.4493 | 11.5 | 0.12 |
| polarC18 | 758.568 | 144.1203 | 11.5 | 0.12 |
| polarC18 | 758.568 | 86.9987 | 11.5 | 0.12 |
| polarC18 | 758.568 | 142.9283 | 11.5 | 0.12 |
| polarC18 | 758.568 | 585.6508 | 11.5 | 0.12 |
| polarC18 | 758.568 | 90.1679 | 11.5 | 0.12 |
| polarC18 | 758.568 | 79.5658 | 11.5 | 0.12 |
| polarC18 | 758.568 | 85.9608 | 11.5 | 0.12 |
| polarC18 | 758.568 | 100.4116 | 11.5 | 0.12 |
| polarC18 | 758.568 | 133.6671 | 11.5 | 0.12 |
| polarC18 | 758.568 | 293.9531 | 11.4 | 0.12 |
| polarC18 | 758.568 | 270.8192 | 11.4 | 0.12 |
| polarC18 | 758.568 | 216.987 | 11.4 | 0.12 |
| polarC18 | 758.568 | 190.4677 | 11.4 | 0.12 |
| polarC18 | 758.568 | 117.3231 | 11.4 | 0.12 |
| polarC18 | 758.568 | 85.351 | 11.4 | 0.12 |
| polarC18 | 758.568 | 196.8586 | 11.4 | 0.12 |
| polarC18 | 758.568 | 252.4197 | 11.4 | 0.12 |
| polarC18 | 758.568 | 101.9324 | 11.4 | 0.12 |
| polarC18 | 758.568 | 324.4387 | 11.4 | 0.12 |
| polarC18 | 758.568 | 210.007 | 11.4 | 0.12 |
| polarC18 | 758.568 | 208.6372 | 11.4 | 0.12 |
| polarC18 | 758.568 | 101.6872 | 11.4 | 0.12 |
| polarC18 | 758.568 | 293.1547 | 11.4 | 0.12 |
| polarC18 | 758.568 | 630.2596 | 11.4 | 0.12 |
| polarC18 | 758.568 | 126.6745 | 11.4 | 0.12 |
| polarC18 | 758.568 | 90.6164 | 11.4 | 0.12 |
| polarC18 | 758.568 | 127.4203 | 11.3 | 0.12 |
| polarC18 | 758.568 | 80.9083 | 11.3 | 0.12 |
| polarC18 | 758.568 | 89.1941 | 11.3 | 0.12 |
| polarC18 | 758.568 | 120.0283 | 11.3 | 0.12 |
| polarC18 | 758.568 | 247.7564 | 11.3 | 0.12 |
| polarC18 | 758.568 | 148.5084 | 11.3 | 0.12 |
| polarC18 | 758.568 | 91.3961 | 11.3 | 0.12 |
| polarC18 | 758.568 | 242.9369 | 11.3 | 0.12 |

| | | | | |
|---|---|---|---|---|
| polarC18 | 758.568 | 253.229 | 11.3 | 0.12 |
| polarC18 | 758.568 | 197.8612 | 11.3 | 0.12 |
| polarC18 | 758.568 | 152.3759 | 11.3 | 0.12 |
| polarC18 | 758.568 | 86.623 | 11.2 | 0.12 |
| polarC18 | 758.568 | 169.5474 | 11.2 | 0.12 |
| polarC18 | 758.568 | 99.5149 | 11.2 | 0.12 |
| polarC18 | 758.568 | 89.6328 | 11.2 | 0.12 |
| polarC18 | 758.568 | 244.4442 | 11.2 | 0.12 |
| polarC18 | 758.568 | 111.0003 | 11.2 | 0.12 |
| polarC18 | 758.568 | 101.7621 | 11.2 | 0.12 |
| polarC18 | 758.568 | 141.7162 | 11.2 | 0.12 |
| polarC18 | 758.568 | 173.8027 | 11.2 | 0.12 |
| polarC18 | 758.568 | 167.2213 | 11.2 | 0.12 |
| polarC18 | 758.568 | 98.1656 | 11.2 | 0.12 |
| polarC18 | 758.568 | 739.6096 | 11.2 | 0.12 |
| polarC18 | 758.568 | 265.1741 | 11.1 | 0.12 |
| polarC18 | 758.568 | 422.6175 | 11.1 | 0.12 |
| polarC18 | 758.568 | 116.0562 | 11.1 | 0.12 |
| polarC18 | 758.568 | 156.0327 | 11.1 | 0.12 |
| polarC18 | 758.568 | 243.6107 | 11.1 | 0.12 |
| polarC18 | 758.568 | 411.4041 | 11.1 | 0.12 |
| polarC18 | 758.568 | 190.8689 | 11.1 | 0.12 |
| polarC18 | 758.568 | 87.6503 | 11.1 | 0.12 |
| polarC18 | 758.568 | 134.8043 | 11.1 | 0.12 |
| polarC18 | 758.568 | 107.3398 | 11.1 | 0.12 |
| polarC18 | 758.568 | 196.3568 | 11.1 | 0.12 |
| polarC18 | 758.568 | 157.4864 | 11.1 | 0.12 |
| polarC18 | 758.568 | 115.2442 | 11.1 | 0.12 |
| polarC18 | 758.568 | 189.5745 | 11.1 | 0.12 |
| polarC18 | 758.568 | 131.0045 | 11.1 | 0.12 |
| polarC18 | 758.568 | 146.7091 | 11 | 0.12 |
| polarC18 | 758.568 | 144.7576 | 11 | 0.12 |
| polarC18 | 758.568 | 96.68 | 11 | 0.12 |
| polarC18 | 758.568 | 186.7754 | 11 | 0.12 |
| polarC18 | 758.568 | 388.3788 | 11 | 0.12 |
| polarC18 | 758.568 | 191.7012 | 11 | 0.12 |
| polarC18 | 758.568 | 157.6716 | 11 | 0.12 |
| polarC18 | 758.568 | 206.5442 | 11 | 0.12 |
| polarC18 | 758.568 | 197.2651 | 11 | 0.12 |
| polarC18 | 758.568 | 119.183 | 11 | 0.12 |
| polarC18 | 758.568 | 212.1542 | 10.9 | 0.12 |
| polarC18 | 758.568 | 191.3831 | 10.9 | 0.12 |

| | | | | |
|---|---|---|---|---|
| polarC18 | 758.568 | 146.3389 | 10.9 | 0.12 |
| polarC18 | 758.568 | 79.4286 | 10.9 | 0.12 |
| polarC18 | 758.568 | 373.2252 | 10.9 | 0.12 |
| polarC18 | 758.568 | 193.4304 | 10.9 | 0.12 |
| polarC18 | 758.568 | 223.3273 | 10.8 | 0.12 |
| polarC18 | 758.568 | 121.6243 | 10.8 | 0.12 |
| polarC18 | 758.568 | 275.2058 | 10.8 | 0.12 |
| polarC18 | 758.568 | 91.5918 | 10.8 | 0.12 |
| polarC18 | 758.568 | 86.5638 | 10.7 | 0.12 |
| polarC18 | 758.568 | 101.564 | 10.7 | 0.11 |
| polarC18 | 758.568 | 289.2108 | 10.7 | 0.11 |
| polarC18 | 758.568 | 97.5462 | 10.6 | 0.11 |
| polarC18 | 758.568 | 203.353 | 10.6 | 0.11 |
| polarC18 | 758.568 | 111.903 | 10.6 | 0.11 |
| polarC18 | 758.568 | 142.0942 | 10.6 | 0.11 |
| polarC18 | 758.568 | 86.5739 | 10.5 | 0.11 |
| polarC18 | 758.568 | 79.4381 | 10.5 | 0.11 |
| polarC18 | 758.568 | 85.2631 | 10.4 | 0.11 |
| polarC18 | 758.568 | 156.6523 | 10.4 | 0.11 |
| polarC18 | 758.568 | 151.4117 | 10.4 | 0.11 |
| polarC18 | 758.568 | 84.6857 | 10.4 | 0.11 |
| polarC18 | 758.568 | 85.5263 | 10.3 | 0.11 |
| polarC18 | 758.568 | 262.3521 | 10.1 | 0.11 |
| polarC18 | 758.568 | 177.9668 | 10.1 | 0.11 |
| polarC18 | 780.55 | 780.6341 | 13643.8 | 100 |
| polarC18 | 780.55 | 104.107 | 7234.7 | 53.03 |
| polarC18 | 780.55 | 228.2181 | 5271.3 | 38.64 |
| polarC18 | 780.55 | 118.1227 | 414.2 | 3.04 |
| polarC18 | 780.55 | 804.2326 | 127.5 | 0.93 |
| polarC18 | 780.55 | 93.4482 | 123 | 0.9 |
| polarC18 | 780.55 | 93.375 | 90.1 | 0.66 |
| polarC18 | 780.55 | 706.1065 | 89 | 0.65 |
| polarC18 | 780.55 | 431.313 | 86.6 | 0.63 |
| polarC18 | 780.55 | 93.4872 | 83.4 | 0.61 |
| polarC18 | 780.55 | 123.8198 | 78.7 | 0.58 |
| polarC18 | 780.55 | 224.0009 | 78.3 | 0.57 |
| polarC18 | 780.55 | 756.7769 | 74.1 | 0.54 |
| polarC18 | 780.55 | 746.929 | 72.2 | 0.53 |
| polarC18 | 780.55 | 529.7969 | 70.9 | 0.52 |
| polarC18 | 780.55 | 776.6425 | 70.4 | 0.52 |
| polarC18 | 780.55 | 682.3742 | 69 | 0.51 |
| polarC18 | 780.55 | 155.3067 | 68.6 | 0.5 |

| | | | | |
|---|---|---|---|---|
| polarC18 | 780.55 | 693.4119 | 67.6 | 0.5 |
| polarC18 | 780.55 | 615.1097 | 67.4 | 0.49 |
| polarC18 | 780.55 | 93.417 | 66.9 | 0.49 |
| polarC18 | 780.55 | 452.4729 | 66.7 | 0.49 |
| polarC18 | 780.55 | 621.2481 | 66.6 | 0.49 |
| polarC18 | 780.55 | 93.4655 | 66.5 | 0.49 |
| polarC18 | 780.55 | 305.4098 | 65.9 | 0.48 |
| polarC18 | 780.55 | 640.6816 | 65.6 | 0.48 |
| polarC18 | 780.55 | 129.4316 | 65.3 | 0.48 |
| polarC18 | 780.55 | 728.3072 | 64.5 | 0.47 |
| polarC18 | 780.55 | 655.0634 | 64.3 | 0.47 |
| polarC18 | 780.55 | 93.4761 | 63.3 | 0.46 |
| polarC18 | 780.55 | 93.4117 | 63.2 | 0.46 |
| polarC18 | 780.55 | 310.0429 | 61.7 | 0.45 |
| polarC18 | 780.55 | 362.5343 | 60.8 | 0.45 |
| polarC18 | 780.55 | 429.235 | 59.8 | 0.44 |
| polarC18 | 780.55 | 93.4258 | 58.6 | 0.43 |
| polarC18 | 780.55 | 548.6627 | 57.5 | 0.42 |
| polarC18 | 780.55 | 146.5604 | 57 | 0.42 |
| polarC18 | 780.55 | 612.8576 | 56.1 | 0.41 |
| polarC18 | 780.55 | 83.499 | 56 | 0.41 |
| polarC18 | 780.55 | 575.948 | 55.1 | 0.4 |
| polarC18 | 780.55 | 753.4405 | 54.5 | 0.4 |
| polarC18 | 780.55 | 639.3461 | 54 | 0.4 |
| polarC18 | 780.55 | 144.8592 | 53.5 | 0.39 |
| polarC18 | 780.55 | 657.8488 | 53.3 | 0.39 |
| polarC18 | 780.55 | 376.7043 | 52.9 | 0.39 |
| polarC18 | 780.55 | 541.1616 | 52.9 | 0.39 |
| polarC18 | 780.55 | 93.4942 | 52.3 | 0.38 |
| polarC18 | 780.55 | 155.0442 | 52 | 0.38 |
| polarC18 | 780.55 | 93.3694 | 52 | 0.38 |
| polarC18 | 780.55 | 509.5244 | 51.8 | 0.38 |
| polarC18 | 780.55 | 371.2593 | 51.5 | 0.38 |
| polarC18 | 780.55 | 459.3477 | 51.3 | 0.38 |
| polarC18 | 780.55 | 82.9367 | 51.2 | 0.38 |
| polarC18 | 780.55 | 750.0883 | 51.1 | 0.37 |
| polarC18 | 780.55 | 314.7694 | 51 | 0.37 |
| polarC18 | 780.55 | 235.0873 | 51 | 0.37 |
| polarC18 | 780.55 | 703.6084 | 50.6 | 0.37 |
| polarC18 | 780.55 | 620.8285 | 50.6 | 0.37 |
| polarC18 | 780.55 | 536.7085 | 50.5 | 0.37 |
| polarC18 | 780.55 | 380.1146 | 50.5 | 0.37 |

| | | | | |
|---|---|---|---|---|
| polarC18 | 780.55 | 793.1915 | 50.4 | 0.37 |
| polarC18 | 780.55 | 433.5466 | 50.3 | 0.37 |
| polarC18 | 780.55 | 93.406 | 50.2 | 0.37 |
| polarC18 | 780.55 | 648.5409 | 50 | 0.37 |
| polarC18 | 780.55 | 106.7718 | 49.9 | 0.37 |
| polarC18 | 780.55 | 702.1067 | 49.8 | 0.37 |
| polarC18 | 780.55 | 615.8997 | 49.6 | 0.36 |
| polarC18 | 780.55 | 419.9974 | 49.5 | 0.36 |
| polarC18 | 780.55 | 164.8922 | 49.1 | 0.36 |
| polarC18 | 780.55 | 117.839 | 49.1 | 0.36 |
| polarC18 | 780.55 | 438.6648 | 49.1 | 0.36 |
| polarC18 | 780.55 | 662.8457 | 49.1 | 0.36 |
| polarC18 | 780.55 | 711.1254 | 49 | 0.36 |
| polarC18 | 780.55 | 747.546 | 49 | 0.36 |
| polarC18 | 780.55 | 499.6269 | 48.9 | 0.36 |
| polarC18 | 780.55 | 125.0648 | 48.8 | 0.36 |
| polarC18 | 780.55 | 284.2648 | 48.7 | 0.36 |
| polarC18 | 780.55 | 173.411 | 48.6 | 0.36 |
| polarC18 | 780.55 | 757.93 | 48.6 | 0.36 |
| polarC18 | 780.55 | 425.5224 | 48.6 | 0.36 |
| polarC18 | 780.55 | 560.0326 | 48.5 | 0.36 |
| polarC18 | 780.55 | 412.1071 | 48.5 | 0.36 |
| polarC18 | 780.55 | 347.4113 | 48.3 | 0.35 |
| polarC18 | 780.55 | 103.0844 | 48.2 | 0.35 |
| polarC18 | 780.55 | 206.9639 | 48 | 0.35 |
| polarC18 | 780.55 | 159.3167 | 47.9 | 0.35 |
| polarC18 | 780.55 | 685.4844 | 47.9 | 0.35 |
| polarC18 | 780.55 | 665.6073 | 47.6 | 0.35 |
| polarC18 | 780.55 | 162.9256 | 47.6 | 0.35 |
| polarC18 | 780.55 | 302.1421 | 47.6 | 0.35 |
| polarC18 | 780.55 | 148.1864 | 47.5 | 0.35 |
| polarC18 | 780.55 | 646.9221 | 47.5 | 0.35 |
| polarC18 | 780.55 | 299.0071 | 47.4 | 0.35 |
| polarC18 | 780.55 | 743.5215 | 47.4 | 0.35 |
| polarC18 | 780.55 | 83.38 | 47.2 | 0.35 |
| polarC18 | 780.55 | 788.9455 | 47 | 0.34 |
| polarC18 | 780.55 | 117.4383 | 46.9 | 0.34 |
| polarC18 | 780.55 | 124.6093 | 46.5 | 0.34 |
| polarC18 | 780.55 | 307.7048 | 46.5 | 0.34 |
| polarC18 | 780.55 | 312.6486 | 46.5 | 0.34 |
| polarC18 | 780.55 | 244.8888 | 46.4 | 0.34 |
| polarC18 | 780.55 | 318.6692 | 46.4 | 0.34 |

| | | | | |
|---|---|---|---|---|
| polarC18 | 780.55 | 245.3103 | 46.3 | 0.34 |
| polarC18 | 780.55 | 722.3897 | 46.2 | 0.34 |
| polarC18 | 780.55 | 562.2027 | 46.2 | 0.34 |
| polarC18 | 780.55 | 744.2305 | 45.9 | 0.34 |
| polarC18 | 780.55 | 438.0358 | 45.7 | 0.33 |
| polarC18 | 780.55 | 402.0419 | 45.6 | 0.33 |
| polarC18 | 780.55 | 109.0686 | 45.6 | 0.33 |
| polarC18 | 780.55 | 330.2626 | 45.4 | 0.33 |
| polarC18 | 780.55 | 114.3759 | 45.4 | 0.33 |
| polarC18 | 780.55 | 215.3793 | 45.4 | 0.33 |
| polarC18 | 780.55 | 291.2336 | 45.3 | 0.33 |
| polarC18 | 780.55 | 479.1915 | 45.3 | 0.33 |
| polarC18 | 780.55 | 227.2222 | 45.1 | 0.33 |
| polarC18 | 780.55 | 323.5331 | 44.8 | 0.33 |
| polarC18 | 780.55 | 727.1617 | 44.7 | 0.33 |
| polarC18 | 780.55 | 287.7473 | 44.7 | 0.33 |
| polarC18 | 780.55 | 100.5264 | 44.5 | 0.33 |
| polarC18 | 780.55 | 119.0742 | 44.4 | 0.33 |
| polarC18 | 780.55 | 526.2536 | 44 | 0.32 |
| polarC18 | 780.55 | 558.8754 | 43.6 | 0.32 |
| polarC18 | 780.55 | 200.888 | 43.5 | 0.32 |
| polarC18 | 780.55 | 121.5935 | 43.5 | 0.32 |
| polarC18 | 780.55 | 154.0903 | 43.4 | 0.32 |
| polarC18 | 780.55 | 93.4619 | 42.8 | 0.31 |
| polarC18 | 780.55 | 330.1615 | 42.7 | 0.31 |
| polarC18 | 780.55 | 258.6898 | 42.3 | 0.31 |
| polarC18 | 780.55 | 331.7786 | 41.8 | 0.31 |
| polarC18 | 780.55 | 369.6362 | 41.1 | 0.3 |
| polarC18 | 780.55 | 391.5498 | 40.9 | 0.3 |
| polarC18 | 780.55 | 497.3432 | 40.7 | 0.3 |
| polarC18 | 780.55 | 163.5244 | 40 | 0.29 |
| polarC18 | 780.55 | 533.8166 | 39.6 | 0.29 |
| polarC18 | 780.55 | 458.5164 | 39.5 | 0.29 |
| polarC18 | 780.55 | 604.3778 | 39.4 | 0.29 |
| polarC18 | 780.55 | 755.8295 | 39.2 | 0.29 |
| polarC18 | 780.55 | 806.341 | 38.9 | 0.29 |
| polarC18 | 780.55 | 209.4608 | 38.3 | 0.28 |
| polarC18 | 780.55 | 514.1865 | 38.1 | 0.28 |
| polarC18 | 780.55 | 247.1565 | 37.9 | 0.28 |
| polarC18 | 780.55 | 593.0628 | 37.7 | 0.28 |
| polarC18 | 780.55 | 510.5752 | 37.7 | 0.28 |
| polarC18 | 780.55 | 813.1807 | 37.6 | 0.28 |

| | | | | |
|---|---|---|---|---|
| polarC18 | 780.55 | 486.0371 | 37.5 | 0.27 |
| polarC18 | 780.55 | 517.1501 | 37.5 | 0.27 |
| polarC18 | 780.55 | 742.4298 | 37.5 | 0.27 |
| polarC18 | 780.55 | 226.5912 | 37.4 | 0.27 |
| polarC18 | 780.55 | 201.2375 | 37.3 | 0.27 |
| polarC18 | 780.55 | 461.7275 | 37.2 | 0.27 |
| polarC18 | 780.55 | 714.1704 | 37.2 | 0.27 |
| polarC18 | 780.55 | 198.0876 | 37 | 0.27 |
| polarC18 | 780.55 | 301.3105 | 37 | 0.27 |
| polarC18 | 780.55 | 463.4945 | 37 | 0.27 |
| polarC18 | 780.55 | 566.3791 | 36.8 | 0.27 |
| polarC18 | 780.55 | 153.0078 | 36.7 | 0.27 |
| polarC18 | 780.55 | 421.9397 | 36.4 | 0.27 |
| polarC18 | 780.55 | 590.8738 | 36.4 | 0.27 |
| polarC18 | 780.55 | 478.7968 | 36.4 | 0.27 |
| polarC18 | 780.55 | 569.6123 | 36.4 | 0.27 |
| polarC18 | 780.55 | 696.8378 | 36.4 | 0.27 |
| polarC18 | 780.55 | 353.2677 | 36.3 | 0.27 |
| polarC18 | 780.55 | 192.3113 | 36.3 | 0.27 |
| polarC18 | 780.55 | 678.8715 | 36.2 | 0.27 |
| polarC18 | 780.55 | 670.0712 | 36.2 | 0.27 |
| polarC18 | 780.55 | 765.1266 | 36.2 | 0.27 |
| polarC18 | 780.55 | 290.0516 | 36.2 | 0.26 |
| polarC18 | 780.55 | 799.7972 | 36.2 | 0.26 |
| polarC18 | 780.55 | 660.722 | 36.1 | 0.26 |
| polarC18 | 780.55 | 602.9453 | 36 | 0.26 |
| polarC18 | 780.55 | 483.8961 | 35.9 | 0.26 |
| polarC18 | 780.55 | 376.8654 | 35.8 | 0.26 |
| polarC18 | 780.55 | 543.2792 | 35.7 | 0.26 |
| polarC18 | 780.55 | 86.0963 | 35.7 | 0.26 |
| polarC18 | 780.55 | 185.9085 | 35.7 | 0.26 |
| polarC18 | 780.55 | 304.9941 | 35.6 | 0.26 |
| polarC18 | 780.55 | 711.9692 | 35.6 | 0.26 |
| polarC18 | 780.55 | 337.2325 | 35.5 | 0.26 |
| polarC18 | 780.55 | 406.7419 | 35.5 | 0.26 |
| polarC18 | 780.55 | 84.8715 | 35.5 | 0.26 |
| polarC18 | 780.55 | 126.659 | 35.5 | 0.26 |
| polarC18 | 780.55 | 662.1725 | 35.5 | 0.26 |
| polarC18 | 780.55 | 620.0299 | 35.5 | 0.26 |
| polarC18 | 780.55 | 646.462 | 35.4 | 0.26 |
| polarC18 | 780.55 | 498.1072 | 35.4 | 0.26 |
| polarC18 | 780.55 | 737.7399 | 35.4 | 0.26 |

| | | | | |
|---|---|---|---|---|
| polarC18 | 780.55 | 266.163 | 35.3 | 0.26 |
| polarC18 | 780.55 | 168.045 | 35.3 | 0.26 |
| polarC18 | 780.55 | 209.1909 | 35.3 | 0.26 |
| polarC18 | 780.55 | 590.3494 | 35.2 | 0.26 |
| polarC18 | 780.55 | 379.0745 | 35.2 | 0.26 |
| polarC18 | 780.55 | 712.9786 | 35.2 | 0.26 |
| polarC18 | 780.55 | 223.5807 | 35.1 | 0.26 |
| polarC18 | 780.55 | 816.1432 | 35.1 | 0.26 |
| polarC18 | 780.55 | 332.5985 | 35.1 | 0.26 |
| polarC18 | 780.55 | 424.94 | 35.1 | 0.26 |
| polarC18 | 780.55 | 343.6438 | 35.1 | 0.26 |
| polarC18 | 780.55 | 310.197 | 35.1 | 0.26 |
| polarC18 | 780.55 | 543.5793 | 35 | 0.26 |
| polarC18 | 780.55 | 784.3579 | 35 | 0.26 |
| polarC18 | 780.55 | 686.856 | 35 | 0.26 |
| polarC18 | 780.55 | 723.6023 | 35 | 0.26 |
| polarC18 | 780.55 | 245.0562 | 35 | 0.26 |
| polarC18 | 780.55 | 123.4236 | 34.9 | 0.26 |
| polarC18 | 780.55 | 730.6275 | 34.9 | 0.26 |
| polarC18 | 780.55 | 125.4975 | 34.9 | 0.26 |
| polarC18 | 780.55 | 409.9103 | 34.9 | 0.26 |
| polarC18 | 780.55 | 252.4935 | 34.9 | 0.26 |
| polarC18 | 780.55 | 239.4347 | 34.8 | 0.26 |
| polarC18 | 780.55 | 210.0489 | 34.8 | 0.26 |
| polarC18 | 780.55 | 93.3631 | 34.8 | 0.26 |
| polarC18 | 780.55 | 395.2146 | 34.8 | 0.26 |
| polarC18 | 780.55 | 322.5716 | 34.8 | 0.25 |
| polarC18 | 780.55 | 601.8891 | 34.8 | 0.25 |
| polarC18 | 780.55 | 90.759 | 34.8 | 0.25 |
| polarC18 | 780.55 | 225.817 | 34.8 | 0.25 |
| polarC18 | 780.55 | 555.4537 | 34.8 | 0.25 |
| polarC18 | 780.55 | 225.205 | 34.7 | 0.25 |
| polarC18 | 780.55 | 131.1614 | 34.7 | 0.25 |
| polarC18 | 780.55 | 529.9681 | 34.6 | 0.25 |
| polarC18 | 780.55 | 193.8655 | 34.6 | 0.25 |
| polarC18 | 780.55 | 238.1856 | 34.6 | 0.25 |
| polarC18 | 780.55 | 540.9978 | 34.6 | 0.25 |
| polarC18 | 780.55 | 676.1848 | 34.6 | 0.25 |
| polarC18 | 780.55 | 508.4423 | 34.6 | 0.25 |
| polarC18 | 780.55 | 440.6735 | 34.6 | 0.25 |
| polarC18 | 780.55 | 472.7128 | 34.6 | 0.25 |
| polarC18 | 780.55 | 318.577 | 34.6 | 0.25 |

| | | | | |
|---|---|---|---|---|
| polarC18 | 780.55 | 108.4426 | 34.6 | 0.25 |
| polarC18 | 780.55 | 396.3293 | 34.5 | 0.25 |
| polarC18 | 780.55 | 650.7221 | 34.5 | 0.25 |
| polarC18 | 780.55 | 525.1371 | 34.5 | 0.25 |
| polarC18 | 780.55 | 394.3608 | 34.5 | 0.25 |
| polarC18 | 780.55 | 96.472 | 34.5 | 0.25 |
| polarC18 | 780.55 | 168.1489 | 34.5 | 0.25 |
| polarC18 | 780.55 | 454.8158 | 34.4 | 0.25 |
| polarC18 | 780.55 | 557.7726 | 34.4 | 0.25 |
| polarC18 | 780.55 | 176.0127 | 34.4 | 0.25 |
| polarC18 | 780.55 | 157.1973 | 34.4 | 0.25 |
| polarC18 | 780.55 | 674.5795 | 34.4 | 0.25 |
| polarC18 | 780.55 | 96.8661 | 34.3 | 0.25 |
| polarC18 | 780.55 | 468.5428 | 34.3 | 0.25 |
| polarC18 | 780.55 | 164.6656 | 34.3 | 0.25 |
| polarC18 | 780.55 | 130.0426 | 34.3 | 0.25 |
| polarC18 | 780.55 | 237.8145 | 34.3 | 0.25 |
| polarC18 | 780.55 | 138.7167 | 34.2 | 0.25 |
| polarC18 | 780.55 | 649.1445 | 34.2 | 0.25 |
| polarC18 | 780.55 | 673.7811 | 34.1 | 0.25 |
| polarC18 | 780.55 | 471.3538 | 34.1 | 0.25 |
| polarC18 | 780.55 | 236.7028 | 34.1 | 0.25 |
| polarC18 | 780.55 | 391.0472 | 34.1 | 0.25 |
| polarC18 | 780.55 | 93.4214 | 34 | 0.25 |
| polarC18 | 780.55 | 298.3164 | 34 | 0.25 |
| polarC18 | 780.55 | 547.2788 | 34 | 0.25 |
| polarC18 | 780.55 | 132.2166 | 34 | 0.25 |
| polarC18 | 780.55 | 251.2455 | 34 | 0.25 |
| polarC18 | 780.55 | 239.6796 | 34 | 0.25 |
| polarC18 | 780.55 | 264.0237 | 34 | 0.25 |
| polarC18 | 780.55 | 805.912 | 34 | 0.25 |
| polarC18 | 780.55 | 591.9515 | 34 | 0.25 |
| polarC18 | 780.55 | 468.1548 | 34 | 0.25 |
| polarC18 | 780.55 | 146.5445 | 34 | 0.25 |
| polarC18 | 780.55 | 484.0124 | 34 | 0.25 |
| polarC18 | 780.55 | 284.1506 | 33.9 | 0.25 |
| polarC18 | 780.55 | 82.5266 | 33.9 | 0.25 |
| polarC18 | 780.55 | 292.1143 | 33.9 | 0.25 |
| polarC18 | 780.55 | 586.2854 | 33.9 | 0.25 |
| polarC18 | 780.55 | 576.3618 | 33.9 | 0.25 |
| polarC18 | 780.55 | 611.4757 | 33.9 | 0.25 |
| polarC18 | 780.55 | 741.8258 | 33.9 | 0.25 |

| | | | | |
|---|---|---|---|---|
| polarC18 | 780.55 | 778.0748 | 33.9 | 0.25 |
| polarC18 | 780.55 | 717.5897 | 33.8 | 0.25 |
| polarC18 | 780.55 | 340.5092 | 33.8 | 0.25 |
| polarC18 | 780.55 | 633.9189 | 33.8 | 0.25 |
| polarC18 | 780.55 | 235.9151 | 33.8 | 0.25 |
| polarC18 | 780.55 | 736.2573 | 33.8 | 0.25 |
| polarC18 | 780.55 | 416.4971 | 33.8 | 0.25 |
| polarC18 | 780.55 | 545.0497 | 33.8 | 0.25 |
| polarC18 | 780.55 | 99.5861 | 33.8 | 0.25 |
| polarC18 | 780.55 | 239.2868 | 33.7 | 0.25 |
| polarC18 | 780.55 | 485.7644 | 33.7 | 0.25 |
| polarC18 | 780.55 | 121.6243 | 33.7 | 0.25 |
| polarC18 | 780.55 | 346.7073 | 33.7 | 0.25 |
| polarC18 | 780.55 | 482.187 | 33.7 | 0.25 |
| polarC18 | 780.55 | 492.8802 | 33.7 | 0.25 |
| polarC18 | 780.55 | 81.6823 | 33.7 | 0.25 |
| polarC18 | 780.55 | 254.4211 | 33.6 | 0.25 |
| polarC18 | 780.55 | 160.6997 | 33.6 | 0.25 |
| polarC18 | 780.55 | 91.9341 | 33.6 | 0.25 |
| polarC18 | 780.55 | 678.3446 | 33.6 | 0.25 |
| polarC18 | 780.55 | 272.0095 | 33.6 | 0.25 |
| polarC18 | 780.55 | 93.2717 | 33.6 | 0.25 |
| polarC18 | 780.55 | 187.3071 | 33.6 | 0.25 |
| polarC18 | 780.55 | 105.4286 | 33.5 | 0.25 |
| polarC18 | 780.55 | 118.5056 | 33.5 | 0.25 |
| polarC18 | 780.55 | 87.7927 | 33.5 | 0.25 |
| polarC18 | 780.55 | 200.2902 | 33.5 | 0.25 |
| polarC18 | 780.55 | 760.5858 | 33.5 | 0.25 |
| polarC18 | 780.55 | 648.1565 | 33.4 | 0.25 |
| polarC18 | 780.55 | 444.6361 | 33.4 | 0.25 |
| polarC18 | 780.55 | 200.0732 | 33.4 | 0.24 |
| polarC18 | 780.55 | 165.577 | 33.4 | 0.24 |
| polarC18 | 780.55 | 163.581 | 33.4 | 0.24 |
| polarC18 | 780.55 | 140.0101 | 33.4 | 0.24 |
| polarC18 | 780.55 | 529.3583 | 33.3 | 0.24 |
| polarC18 | 780.55 | 267.667 | 33.3 | 0.24 |
| polarC18 | 780.55 | 383.6654 | 33.3 | 0.24 |
| polarC18 | 780.55 | 567.8061 | 33.3 | 0.24 |
| polarC18 | 780.55 | 785.2666 | 33.3 | 0.24 |
| polarC18 | 780.55 | 567.46 | 33.3 | 0.24 |
| polarC18 | 780.55 | 222.3784 | 33.3 | 0.24 |
| polarC18 | 780.55 | 280.7437 | 33.2 | 0.24 |

| | | | | |
|---|---|---|---|---|
| polarC18 | 780.55 | 217.6795 | 33.2 | 0.24 |
| polarC18 | 780.55 | 275.9332 | 33.2 | 0.24 |
| polarC18 | 780.55 | 118.0583 | 33.2 | 0.24 |
| polarC18 | 780.55 | 666.4517 | 33.2 | 0.24 |
| polarC18 | 780.55 | 103.9959 | 33.1 | 0.24 |
| polarC18 | 780.55 | 334.3994 | 33.1 | 0.24 |
| polarC18 | 780.55 | 90.0933 | 33.1 | 0.24 |
| polarC18 | 780.55 | 164.2835 | 33.1 | 0.24 |
| polarC18 | 780.55 | 429.7661 | 33.1 | 0.24 |
| polarC18 | 780.55 | 96.9214 | 33.1 | 0.24 |
| polarC18 | 780.55 | 234.0618 | 33.1 | 0.24 |
| polarC18 | 780.55 | 635.2264 | 33.1 | 0.24 |
| polarC18 | 780.55 | 109.7418 | 33.1 | 0.24 |
| polarC18 | 780.55 | 405.3701 | 33.1 | 0.24 |
| polarC18 | 780.55 | 491.9274 | 33 | 0.24 |
| polarC18 | 780.55 | 158.5144 | 33 | 0.24 |
| polarC18 | 780.55 | 582.7387 | 33 | 0.24 |
| polarC18 | 780.55 | 324.458 | 33 | 0.24 |
| polarC18 | 780.55 | 263.4321 | 33 | 0.24 |
| polarC18 | 780.55 | 609.2932 | 33 | 0.24 |
| polarC18 | 780.55 | 134.5073 | 32.9 | 0.24 |
| polarC18 | 780.55 | 107.4888 | 32.9 | 0.24 |
| polarC18 | 780.55 | 129.2068 | 32.9 | 0.24 |
| polarC18 | 780.55 | 466.0646 | 32.9 | 0.24 |
| polarC18 | 780.55 | 553.1442 | 32.9 | 0.24 |
| polarC18 | 780.55 | 150.7097 | 32.9 | 0.24 |
| polarC18 | 780.55 | 462.0894 | 32.8 | 0.24 |
| polarC18 | 780.55 | 400.0308 | 32.8 | 0.24 |
| polarC18 | 780.55 | 106.269 | 32.8 | 0.24 |
| polarC18 | 780.55 | 87.9126 | 32.8 | 0.24 |
| polarC18 | 780.55 | 196.5172 | 32.8 | 0.24 |
| polarC18 | 780.55 | 159.0919 | 32.8 | 0.24 |
| polarC18 | 780.55 | 140.5051 | 32.8 | 0.24 |
| polarC18 | 780.55 | 87.347 | 32.8 | 0.24 |
| polarC18 | 780.55 | 526.4448 | 32.8 | 0.24 |
| polarC18 | 780.55 | 330.7944 | 32.7 | 0.24 |
| polarC18 | 780.55 | 253.345 | 32.7 | 0.24 |
| polarC18 | 780.55 | 253.5884 | 32.7 | 0.24 |
| polarC18 | 780.55 | 408.5253 | 32.7 | 0.24 |
| polarC18 | 780.55 | 198.8731 | 32.7 | 0.24 |
| polarC18 | 780.55 | 84.0533 | 32.7 | 0.24 |
| polarC18 | 780.55 | 145.7517 | 32.7 | 0.24 |

| | | | | |
|---|---|---|---|---|
| polarC18 | 780.55 | 559.8241 | 32.7 | 0.24 |
| polarC18 | 780.55 | 83.8826 | 32.7 | 0.24 |
| polarC18 | 780.55 | 815.3351 | 32.6 | 0.24 |
| polarC18 | 780.55 | 208.3936 | 32.6 | 0.24 |
| polarC18 | 780.55 | 501.4101 | 32.6 | 0.24 |
| polarC18 | 780.55 | 108.2256 | 32.6 | 0.24 |
| polarC18 | 780.55 | 787.3893 | 32.6 | 0.24 |
| polarC18 | 780.55 | 603.8768 | 32.6 | 0.24 |
| polarC18 | 780.55 | 399.7888 | 32.6 | 0.24 |
| polarC18 | 780.55 | 176.835 | 32.6 | 0.24 |
| polarC18 | 780.55 | 440.5052 | 32.6 | 0.24 |
| polarC18 | 780.55 | 132.2849 | 32.6 | 0.24 |
| polarC18 | 780.55 | 187.2873 | 32.5 | 0.24 |
| polarC18 | 780.55 | 93.3855 | 32.5 | 0.24 |
| polarC18 | 780.55 | 735.7621 | 32.5 | 0.24 |
| polarC18 | 780.55 | 85.1663 | 32.5 | 0.24 |
| polarC18 | 780.55 | 172.7853 | 32.5 | 0.24 |
| polarC18 | 780.55 | 106.894 | 32.4 | 0.24 |
| polarC18 | 780.55 | 197.1054 | 32.4 | 0.24 |
| polarC18 | 780.55 | 97.0494 | 32.4 | 0.24 |
| polarC18 | 780.55 | 85.6619 | 32.3 | 0.24 |
| polarC18 | 780.55 | 184.189 | 32.3 | 0.24 |
| polarC18 | 780.55 | 190.654 | 32.3 | 0.24 |
| polarC18 | 780.55 | 116.3696 | 32.3 | 0.24 |
| polarC18 | 780.55 | 95.5894 | 32.3 | 0.24 |
| polarC18 | 780.55 | 415.3295 | 32.3 | 0.24 |
| polarC18 | 780.55 | 169.1313 | 32.3 | 0.24 |
| polarC18 | 780.55 | 271.5279 | 32.3 | 0.24 |
| polarC18 | 780.55 | 330.7296 | 32.3 | 0.24 |
| polarC18 | 780.55 | 205.9984 | 32.3 | 0.24 |
| polarC18 | 780.55 | 126.4364 | 32.3 | 0.24 |
| polarC18 | 780.55 | 286.5186 | 32.3 | 0.24 |
| polarC18 | 780.55 | 141.6653 | 32.2 | 0.24 |
| polarC18 | 780.55 | 138.1125 | 32.2 | 0.24 |
| polarC18 | 780.55 | 394.6788 | 32.2 | 0.24 |
| polarC18 | 780.55 | 449.0945 | 32.2 | 0.24 |
| polarC18 | 780.55 | 252.4091 | 32.2 | 0.24 |
| polarC18 | 780.55 | 127.6275 | 32.1 | 0.24 |
| polarC18 | 780.55 | 432.8893 | 32.1 | 0.24 |
| polarC18 | 780.55 | 241.1009 | 32.1 | 0.24 |
| polarC18 | 780.55 | 464.9893 | 32.1 | 0.24 |
| polarC18 | 780.55 | 118.2351 | 32.1 | 0.24 |

| | | | | |
|---|---|---|---|---|
| polarC18 | 780.55 | 481.441 | 32.1 | 0.24 |
| polarC18 | 780.55 | 169.2276 | 32.1 | 0.24 |
| polarC18 | 780.55 | 460.9097 | 32.1 | 0.24 |
| polarC18 | 780.55 | 214.288 | 32.1 | 0.24 |
| polarC18 | 780.55 | 780.9955 | 32.1 | 0.24 |
| polarC18 | 780.55 | 797.3693 | 32.1 | 0.24 |
| polarC18 | 780.55 | 570.6957 | 32.1 | 0.24 |
| polarC18 | 780.55 | 221.761 | 32.1 | 0.24 |
| polarC18 | 780.55 | 348.199 | 32.1 | 0.24 |
| polarC18 | 780.55 | 81.9246 | 32.1 | 0.24 |
| polarC18 | 780.55 | 309.8246 | 32.1 | 0.24 |
| polarC18 | 780.55 | 113.2948 | 32.1 | 0.24 |
| polarC18 | 780.55 | 506.2425 | 32 | 0.23 |
| polarC18 | 780.55 | 262.1883 | 32 | 0.23 |
| polarC18 | 780.55 | 263.9149 | 32 | 0.23 |
| polarC18 | 780.55 | 103.3374 | 32 | 0.23 |
| polarC18 | 780.55 | 96.3607 | 32 | 0.23 |
| polarC18 | 780.55 | 814.2163 | 32 | 0.23 |
| polarC18 | 780.55 | 221.2838 | 32 | 0.23 |
| polarC18 | 780.55 | 273.3056 | 32 | 0.23 |
| polarC18 | 780.55 | 242.3539 | 32 | 0.23 |
| polarC18 | 780.55 | 103.0127 | 31.9 | 0.23 |
| polarC18 | 780.55 | 134.5683 | 31.9 | 0.23 |
| polarC18 | 780.55 | 259.9931 | 31.9 | 0.23 |
| polarC18 | 780.55 | 94.2285 | 31.9 | 0.23 |
| polarC18 | 780.55 | 114.5375 | 31.9 | 0.23 |
| polarC18 | 780.55 | 682.8138 | 31.9 | 0.23 |
| polarC18 | 780.55 | 148.3488 | 31.9 | 0.23 |
| polarC18 | 780.55 | 173.742 | 31.9 | 0.23 |
| polarC18 | 780.55 | 459.9986 | 31.9 | 0.23 |
| polarC18 | 780.55 | 798.719 | 31.9 | 0.23 |
| polarC18 | 780.55 | 230.2241 | 31.8 | 0.23 |
| polarC18 | 780.55 | 138.8152 | 31.8 | 0.23 |
| polarC18 | 780.55 | 86.3735 | 31.8 | 0.23 |
| polarC18 | 780.55 | 216.36 | 31.8 | 0.23 |
| polarC18 | 780.55 | 120.1143 | 31.8 | 0.23 |
| polarC18 | 780.55 | 161.8451 | 31.8 | 0.23 |
| polarC18 | 780.55 | 752.469 | 31.8 | 0.23 |
| polarC18 | 780.55 | 595.6623 | 31.7 | 0.23 |
| polarC18 | 780.55 | 99.8815 | 31.7 | 0.23 |
| polarC18 | 780.55 | 595.8948 | 31.7 | 0.23 |
| polarC18 | 780.55 | 456.3987 | 31.7 | 0.23 |

| | | | | |
|---|---|---|---|---|
| polarC18 | 780.55 | 239.9076 | 31.7 | 0.23 |
| polarC18 | 780.55 | 564.2592 | 31.7 | 0.23 |
| polarC18 | 780.55 | 418.5147 | 31.7 | 0.23 |
| polarC18 | 780.55 | 669.1342 | 31.7 | 0.23 |
| polarC18 | 780.55 | 494.113 | 31.7 | 0.23 |
| polarC18 | 780.55 | 140.351 | 31.7 | 0.23 |
| polarC18 | 780.55 | 175.995 | 31.7 | 0.23 |
| polarC18 | 780.55 | 93.3895 | 31.7 | 0.23 |
| polarC18 | 780.55 | 159.0352 | 31.7 | 0.23 |
| polarC18 | 780.55 | 359.2493 | 31.7 | 0.23 |
| polarC18 | 780.55 | 81.8522 | 31.7 | 0.23 |
| polarC18 | 780.55 | 367.1844 | 31.6 | 0.23 |
| polarC18 | 780.55 | 127.2343 | 31.6 | 0.23 |
| polarC18 | 780.55 | 109.0549 | 31.6 | 0.23 |
| polarC18 | 780.55 | 197.7288 | 31.6 | 0.23 |
| polarC18 | 780.55 | 562.9711 | 31.6 | 0.23 |
| polarC18 | 780.55 | 345.1856 | 31.6 | 0.23 |
| polarC18 | 780.55 | 128.1372 | 31.6 | 0.23 |
| polarC18 | 780.55 | 166.4991 | 31.6 | 0.23 |
| polarC18 | 780.55 | 130.6698 | 31.6 | 0.23 |
| polarC18 | 780.55 | 296.9747 | 31.6 | 0.23 |
| polarC18 | 780.55 | 244.0501 | 31.5 | 0.23 |
| polarC18 | 780.55 | 517.8285 | 31.5 | 0.23 |
| polarC18 | 780.55 | 507.6128 | 31.4 | 0.23 |
| polarC18 | 780.55 | 88.3209 | 31.4 | 0.23 |
| polarC18 | 780.55 | 658.3154 | 31.4 | 0.23 |
| polarC18 | 780.55 | 519.5084 | 31.4 | 0.23 |
| polarC18 | 780.55 | 240.5336 | 31.4 | 0.23 |
| polarC18 | 780.55 | 209.8496 | 31.3 | 0.23 |
| polarC18 | 780.55 | 359.0174 | 31.3 | 0.23 |
| polarC18 | 780.55 | 136.2761 | 31.3 | 0.23 |
| polarC18 | 780.55 | 633.3067 | 31.3 | 0.23 |
| polarC18 | 780.55 | 336.0382 | 31.3 | 0.23 |
| polarC18 | 780.55 | 280.2218 | 31.3 | 0.23 |
| polarC18 | 780.55 | 388.3594 | 31.3 | 0.23 |
| polarC18 | 780.55 | 198.1928 | 31.2 | 0.23 |
| polarC18 | 780.55 | 284.3176 | 31.2 | 0.23 |
| polarC18 | 780.55 | 408.9225 | 31.2 | 0.23 |
| polarC18 | 780.55 | 121.1941 | 31.2 | 0.23 |
| polarC18 | 780.55 | 322.3704 | 31.2 | 0.23 |
| polarC18 | 780.55 | 596.6166 | 31.2 | 0.23 |
| polarC18 | 780.55 | 134.464 | 31.2 | 0.23 |

| | | | | |
|---|---|---|---|---|
| polarC18 | 780.55 | 355.423 | 31.2 | 0.23 |
| polarC18 | 780.55 | 140.0389 | 31.2 | 0.23 |
| polarC18 | 780.55 | 429.8533 | 31.2 | 0.23 |
| polarC18 | 780.55 | 304.4142 | 31.1 | 0.23 |
| polarC18 | 780.55 | 119.7086 | 31.1 | 0.23 |
| polarC18 | 780.55 | 123.2777 | 31.1 | 0.23 |
| polarC18 | 780.55 | 201.3232 | 31.1 | 0.23 |
| polarC18 | 780.55 | 99.9162 | 31.1 | 0.23 |
| polarC18 | 780.55 | 184.5343 | 31.1 | 0.23 |
| polarC18 | 780.55 | 114.7578 | 31.1 | 0.23 |
| polarC18 | 780.55 | 92.0704 | 31 | 0.23 |
| polarC18 | 780.55 | 113.8114 | 31 | 0.23 |
| polarC18 | 780.55 | 263.6361 | 31 | 0.23 |
| polarC18 | 780.55 | 366.7637 | 31 | 0.23 |
| polarC18 | 780.55 | 492.06 | 31 | 0.23 |
| polarC18 | 780.55 | 252.9688 | 31 | 0.23 |
| polarC18 | 780.55 | 560.2096 | 30.9 | 0.23 |
| polarC18 | 780.55 | 303.8675 | 30.9 | 0.23 |
| polarC18 | 780.55 | 315.0591 | 30.9 | 0.23 |
| polarC18 | 780.55 | 299.206 | 30.9 | 0.23 |
| polarC18 | 780.55 | 379.4862 | 30.9 | 0.23 |
| polarC18 | 780.55 | 532.3912 | 30.9 | 0.23 |
| polarC18 | 780.55 | 239.176 | 30.9 | 0.23 |
| polarC18 | 780.55 | 96.7702 | 30.9 | 0.23 |
| polarC18 | 780.55 | 225.8563 | 30.9 | 0.23 |
| polarC18 | 780.55 | 156.6877 | 30.9 | 0.23 |
| polarC18 | 780.55 | 188.2303 | 30.8 | 0.23 |
| polarC18 | 780.55 | 309.6163 | 30.8 | 0.23 |
| polarC18 | 780.55 | 88.9757 | 30.8 | 0.23 |
| polarC18 | 780.55 | 625.2811 | 30.8 | 0.23 |
| polarC18 | 780.55 | 229.9273 | 30.8 | 0.23 |
| polarC18 | 780.55 | 622.1746 | 30.8 | 0.23 |
| polarC18 | 780.55 | 711.5138 | 30.8 | 0.23 |
| polarC18 | 780.55 | 436.22 | 30.8 | 0.23 |
| polarC18 | 780.55 | 161.7927 | 30.8 | 0.23 |
| polarC18 | 780.55 | 282.8874 | 30.8 | 0.23 |
| polarC18 | 780.55 | 430.7374 | 30.8 | 0.23 |
| polarC18 | 780.55 | 126.6728 | 30.8 | 0.23 |
| polarC18 | 780.55 | 748.7546 | 30.7 | 0.23 |
| polarC18 | 780.55 | 552.7451 | 30.7 | 0.23 |
| polarC18 | 780.55 | 143.8316 | 30.7 | 0.23 |
| polarC18 | 780.55 | 132.7859 | 30.7 | 0.23 |

| | | | | |
|---|---|---|---|---|
| polarC18 | 780.55 | 586.9851 | 30.7 | 0.22 |
| polarC18 | 780.55 | 692.0298 | 30.7 | 0.22 |
| polarC18 | 780.55 | 310.6363 | 30.7 | 0.22 |
| polarC18 | 780.55 | 568.0807 | 30.7 | 0.22 |
| polarC18 | 780.55 | 213.1402 | 30.7 | 0.22 |
| polarC18 | 780.55 | 92.5256 | 30.7 | 0.22 |
| polarC18 | 780.55 | 171.9962 | 30.6 | 0.22 |
| polarC18 | 780.55 | 281.3515 | 30.6 | 0.22 |
| polarC18 | 780.55 | 235.9802 | 30.6 | 0.22 |
| polarC18 | 780.55 | 226.5172 | 30.6 | 0.22 |
| polarC18 | 780.55 | 493.956 | 30.6 | 0.22 |
| polarC18 | 780.55 | 84.2109 | 30.5 | 0.22 |
| polarC18 | 780.55 | 139.5675 | 30.5 | 0.22 |
| polarC18 | 780.55 | 699.4967 | 30.5 | 0.22 |
| polarC18 | 780.55 | 244.2487 | 30.5 | 0.22 |
| polarC18 | 780.55 | 164.4477 | 30.5 | 0.22 |
| polarC18 | 780.55 | 314.3053 | 30.5 | 0.22 |
| polarC18 | 780.55 | 265.4701 | 30.5 | 0.22 |
| polarC18 | 780.55 | 239.3802 | 30.5 | 0.22 |
| polarC18 | 780.55 | 162.728 | 30.5 | 0.22 |
| polarC18 | 780.55 | 314.0961 | 30.4 | 0.22 |
| polarC18 | 780.55 | 387.2259 | 30.4 | 0.22 |
| polarC18 | 780.55 | 197.4179 | 30.4 | 0.22 |
| polarC18 | 780.55 | 259.5241 | 30.4 | 0.22 |
| polarC18 | 780.55 | 258.4192 | 30.4 | 0.22 |
| polarC18 | 780.55 | 267.1496 | 30.3 | 0.22 |
| polarC18 | 780.55 | 126.973 | 30.3 | 0.22 |
| polarC18 | 780.55 | 100.9577 | 30.3 | 0.22 |
| polarC18 | 780.55 | 135.682 | 30.3 | 0.22 |
| polarC18 | 780.55 | 417.9027 | 30.3 | 0.22 |
| polarC18 | 780.55 | 459.1181 | 30.3 | 0.22 |
| polarC18 | 780.55 | 143.6369 | 30.3 | 0.22 |
| polarC18 | 780.55 | 222.0167 | 30.3 | 0.22 |
| polarC18 | 780.55 | 250.8287 | 30.3 | 0.22 |
| polarC18 | 780.55 | 82.664 | 30.2 | 0.22 |
| polarC18 | 780.55 | 89.0611 | 30.2 | 0.22 |
| polarC18 | 780.55 | 290.4863 | 30.2 | 0.22 |
| polarC18 | 780.55 | 179.6104 | 30.2 | 0.22 |
| polarC18 | 780.55 | 489.1741 | 30.2 | 0.22 |
| polarC18 | 780.55 | 302.6251 | 30.2 | 0.22 |
| polarC18 | 780.55 | 85.2538 | 30.2 | 0.22 |
| polarC18 | 780.55 | 244.653 | 30.1 | 0.22 |

| | | | | |
|---|---|---|---|---|
| polarC18 | 780.55 | 120.0227 | 30.1 | 0.22 |
| polarC18 | 780.55 | 401.805 | 30.1 | 0.22 |
| polarC18 | 780.55 | 144.8828 | 30.1 | 0.22 |
| polarC18 | 780.55 | 518.3517 | 30 | 0.22 |
| polarC18 | 780.55 | 132.2565 | 30 | 0.22 |
| polarC18 | 780.55 | 674.8617 | 29.9 | 0.22 |
| polarC18 | 780.55 | 155.6554 | 29.9 | 0.22 |
| polarC18 | 780.55 | 194.9891 | 29.9 | 0.22 |
| polarC18 | 780.55 | 651.9121 | 29.9 | 0.22 |
| polarC18 | 780.55 | 297.357 | 29.9 | 0.22 |
| polarC18 | 780.55 | 112.9684 | 29.8 | 0.22 |
| polarC18 | 780.55 | 453.1009 | 29.8 | 0.22 |
| polarC18 | 780.55 | 133.8661 | 29.8 | 0.22 |
| polarC18 | 780.55 | 359.143 | 29.8 | 0.22 |
| polarC18 | 780.55 | 261.7019 | 29.8 | 0.22 |
| polarC18 | 780.55 | 123.717 | 29.8 | 0.22 |
| polarC18 | 780.55 | 93.5807 | 29.8 | 0.22 |
| polarC18 | 780.55 | 641.9366 | 29.8 | 0.22 |
| polarC18 | 780.55 | 140.5235 | 29.8 | 0.22 |
| polarC18 | 780.55 | 373.24 | 29.8 | 0.22 |
| polarC18 | 780.55 | 310.2545 | 29.8 | 0.22 |
| polarC18 | 780.55 | 236.3161 | 29.7 | 0.22 |
| polarC18 | 780.55 | 233.4769 | 29.7 | 0.22 |
| polarC18 | 780.55 | 112.8075 | 29.7 | 0.22 |
| polarC18 | 780.55 | 370.6667 | 29.7 | 0.22 |
| polarC18 | 780.55 | 339.7197 | 29.6 | 0.22 |
| polarC18 | 780.55 | 198.1093 | 29.6 | 0.22 |
| polarC18 | 780.55 | 314.1973 | 29.6 | 0.22 |
| polarC18 | 780.55 | 91.6092 | 29.6 | 0.22 |
| polarC18 | 780.55 | 295.9029 | 29.6 | 0.22 |
| polarC18 | 780.55 | 258.7837 | 29.5 | 0.22 |
| polarC18 | 780.55 | 100.2746 | 29.5 | 0.22 |
| polarC18 | 780.55 | 113.8624 | 29.5 | 0.22 |
| polarC18 | 780.55 | 106.2637 | 29.4 | 0.22 |
| polarC18 | 780.55 | 274.6391 | 29.4 | 0.22 |
| polarC18 | 780.55 | 190.2526 | 29.4 | 0.22 |
| polarC18 | 780.55 | 108.349 | 29.4 | 0.22 |
| polarC18 | 780.55 | 371.4742 | 29.4 | 0.22 |
| polarC18 | 780.55 | 92.4548 | 29.4 | 0.22 |
| polarC18 | 780.55 | 136.3483 | 29.4 | 0.22 |
| polarC18 | 780.55 | 86.4095 | 29.4 | 0.22 |
| polarC18 | 780.55 | 237.2572 | 29.3 | 0.21 |

| | | | | |
|---|---|---|---|---|
| polarC18 | 780.55 | 647.1972 | 29.3 | 0.21 |
| polarC18 | 780.55 | 505.9652 | 29.3 | 0.21 |
| polarC18 | 780.55 | 224.507 | 29.3 | 0.21 |
| polarC18 | 780.55 | 113.0077 | 29.2 | 0.21 |
| polarC18 | 780.55 | 91.7077 | 29.2 | 0.21 |
| polarC18 | 780.55 | 123.5522 | 29.2 | 0.21 |
| polarC18 | 780.55 | 91.0861 | 29.2 | 0.21 |
| polarC18 | 780.55 | 393.1695 | 29.2 | 0.21 |
| polarC18 | 780.55 | 157.1313 | 29.2 | 0.21 |
| polarC18 | 780.55 | 217.2882 | 29.2 | 0.21 |
| polarC18 | 780.55 | 173.8099 | 29.1 | 0.21 |
| polarC18 | 780.55 | 86.324 | 29.1 | 0.21 |
| polarC18 | 780.55 | 211.8475 | 29.1 | 0.21 |
| polarC18 | 780.55 | 442.2311 | 29.1 | 0.21 |
| polarC18 | 780.55 | 249.1557 | 29.1 | 0.21 |
| polarC18 | 780.55 | 182.0439 | 29.1 | 0.21 |
| polarC18 | 780.55 | 376.966 | 29.1 | 0.21 |
| polarC18 | 780.55 | 100.0897 | 29.1 | 0.21 |
| polarC18 | 780.55 | 288.9112 | 29.1 | 0.21 |
| polarC18 | 780.55 | 440.1947 | 29.1 | 0.21 |
| polarC18 | 780.55 | 111.1094 | 29.1 | 0.21 |
| polarC18 | 780.55 | 618.4646 | 29 | 0.21 |
| polarC18 | 780.55 | 296.136 | 29 | 0.21 |
| polarC18 | 780.55 | 143.5478 | 29 | 0.21 |
| polarC18 | 780.55 | 93.501 | 29 | 0.21 |
| polarC18 | 780.55 | 138.8562 | 29 | 0.21 |
| polarC18 | 780.55 | 156.3669 | 29 | 0.21 |
| polarC18 | 780.55 | 91.1502 | 28.9 | 0.21 |
| polarC18 | 780.55 | 795.9772 | 28.9 | 0.21 |
| polarC18 | 780.55 | 342.9799 | 28.9 | 0.21 |
| polarC18 | 780.55 | 155.7949 | 28.9 | 0.21 |
| polarC18 | 780.55 | 214.6236 | 28.9 | 0.21 |
| polarC18 | 780.55 | 292.354 | 28.9 | 0.21 |
| polarC18 | 780.55 | 183.5555 | 28.8 | 0.21 |
| polarC18 | 780.55 | 581.2003 | 28.8 | 0.21 |
| polarC18 | 780.55 | 157.9615 | 28.8 | 0.21 |
| polarC18 | 780.55 | 462.3122 | 28.8 | 0.21 |
| polarC18 | 780.55 | 112.3906 | 28.8 | 0.21 |
| polarC18 | 780.55 | 109.4987 | 28.7 | 0.21 |
| polarC18 | 780.55 | 236.436 | 28.6 | 0.21 |
| polarC18 | 780.55 | 284.8283 | 28.6 | 0.21 |
| polarC18 | 780.55 | 390.5138 | 28.6 | 0.21 |

| | | | | |
|---|---|---|---|---|
| polarC18 | 780.55 | 424.5527 | 28.6 | 0.21 |
| polarC18 | 780.55 | 97.8269 | 28.6 | 0.21 |
| polarC18 | 780.55 | 372.2205 | 28.6 | 0.21 |
| polarC18 | 780.55 | 570.2197 | 28.5 | 0.21 |
| polarC18 | 780.55 | 263.2134 | 28.5 | 0.21 |
| polarC18 | 780.55 | 156.3906 | 28.5 | 0.21 |
| polarC18 | 780.55 | 177.6848 | 28.5 | 0.21 |
| polarC18 | 780.55 | 138.1982 | 28.4 | 0.21 |
| polarC18 | 780.55 | 216.8538 | 28.4 | 0.21 |
| polarC18 | 780.55 | 208.0845 | 28.4 | 0.21 |
| polarC18 | 780.55 | 421.8009 | 28.4 | 0.21 |
| polarC18 | 780.55 | 206.1663 | 28.4 | 0.21 |
| polarC18 | 780.55 | 189.1177 | 28.3 | 0.21 |
| polarC18 | 780.55 | 127.0686 | 28.3 | 0.21 |
| polarC18 | 780.55 | 214.8641 | 28.3 | 0.21 |
| polarC18 | 780.55 | 110.7552 | 28.3 | 0.21 |
| polarC18 | 780.55 | 92.5612 | 28.3 | 0.21 |
| polarC18 | 780.55 | 171.7315 | 28.3 | 0.21 |
| polarC18 | 780.55 | 119.9265 | 28.3 | 0.21 |
| polarC18 | 780.55 | 132.2327 | 28.3 | 0.21 |
| polarC18 | 780.55 | 204.0316 | 28.2 | 0.21 |
| polarC18 | 780.55 | 300.9175 | 28.2 | 0.21 |
| polarC18 | 780.55 | 211.2772 | 28.2 | 0.21 |
| polarC18 | 780.55 | 91.3538 | 28.2 | 0.21 |
| polarC18 | 780.55 | 111.149 | 28.2 | 0.21 |
| polarC18 | 780.55 | 247.2071 | 28.1 | 0.21 |
| polarC18 | 780.55 | 275.1788 | 28.1 | 0.21 |
| polarC18 | 780.55 | 123.6712 | 28.1 | 0.21 |
| polarC18 | 780.55 | 583.7255 | 28.1 | 0.21 |
| polarC18 | 780.55 | 731.6445 | 28 | 0.21 |
| polarC18 | 780.55 | 173.0538 | 28 | 0.21 |
| polarC18 | 780.55 | 151.647 | 27.9 | 0.2 |
| polarC18 | 780.55 | 151.7555 | 27.8 | 0.2 |
| polarC18 | 780.55 | 323.5885 | 27.8 | 0.2 |
| polarC18 | 780.55 | 296.3835 | 27.8 | 0.2 |
| polarC18 | 780.55 | 217.9746 | 27.7 | 0.2 |
| polarC18 | 780.55 | 142.7832 | 27.7 | 0.2 |
| polarC18 | 780.55 | 235.4854 | 27.7 | 0.2 |
| polarC18 | 780.55 | 519.6437 | 27.7 | 0.2 |
| polarC18 | 780.55 | 350.2071 | 27.7 | 0.2 |
| polarC18 | 780.55 | 161.3355 | 27.6 | 0.2 |
| polarC18 | 780.55 | 158.3652 | 27.6 | 0.2 |

| | | | | |
|---|---|---|---|---|
| polarC18 | 780.55 | 326.3979 | 27.6 | 0.2 |
| polarC18 | 780.55 | 195.5885 | 27.5 | 0.2 |
| polarC18 | 780.55 | 90.256 | 27.5 | 0.2 |
| polarC18 | 780.55 | 90.0267 | 27.5 | 0.2 |
| polarC18 | 780.55 | 411.8255 | 27.5 | 0.2 |
| polarC18 | 780.55 | 186.0844 | 27.4 | 0.2 |
| polarC18 | 780.55 | 234.7369 | 27.4 | 0.2 |
| polarC18 | 780.55 | 283.6934 | 27.4 | 0.2 |
| polarC18 | 780.55 | 579.4627 | 27.3 | 0.2 |
| polarC18 | 780.55 | 90.7369 | 27.3 | 0.2 |
| polarC18 | 780.55 | 118.2425 | 27.1 | 0.2 |
| polarC18 | 780.55 | 357.5611 | 27.1 | 0.2 |
| polarC18 | 780.55 | 138.9917 | 27.1 | 0.2 |
| polarC18 | 780.55 | 493.6588 | 27 | 0.2 |
| polarC18 | 780.55 | 152.5162 | 27 | 0.2 |
| polarC18 | 780.55 | 91.2746 | 26.9 | 0.2 |
| polarC18 | 780.55 | 547.8781 | 26.9 | 0.2 |
| polarC18 | 780.55 | 219.4863 | 26.9 | 0.2 |
| polarC18 | 780.55 | 348.1058 | 26.8 | 0.2 |
| polarC18 | 780.55 | 126.5293 | 26.8 | 0.2 |
| polarC18 | 780.55 | 87.4779 | 26.7 | 0.2 |
| polarC18 | 780.55 | 262.399 | 26.6 | 0.2 |
| polarC18 | 780.55 | 288.031 | 26.5 | 0.19 |
| polarC18 | 780.55 | 216.5089 | 26.5 | 0.19 |
| polarC18 | 780.55 | 124.748 | 26.4 | 0.19 |
| polarC18 | 780.55 | 281.0032 | 26.4 | 0.19 |
| polarC18 | 780.55 | 178.0526 | 26.4 | 0.19 |
| polarC18 | 780.55 | 82.3696 | 26.4 | 0.19 |
| polarC18 | 780.55 | 83.1382 | 26.2 | 0.19 |
| polarC18 | 780.55 | 93.3438 | 26.1 | 0.19 |
| polarC18 | 780.55 | 313.3733 | 26.1 | 0.19 |
| polarC18 | 780.55 | 474.1714 | 26 | 0.19 |
| polarC18 | 780.55 | 680.9312 | 25.9 | 0.19 |
| polarC18 | 780.55 | 213.2175 | 25.9 | 0.19 |
| polarC18 | 780.55 | 111.9362 | 25.4 | 0.19 |
| polarC18 | 780.55 | 98.7649 | 25.3 | 0.19 |
| polarC18 | 780.55 | 88.5607 | 24.9 | 0.18 |
| polarC18 | 780.55 | 788.2111 | 24.5 | 0.18 |
| polarC18 | 780.55 | 183.7337 | 24.4 | 0.18 |
| polarC18 | 780.55 | 609.8277 | 23.7 | 0.17 |
| polarC18 | 780.55 | 93.436 | 22.8 | 0.17 |
| polarC18 | 780.55 | 697.6208 | 22.2 | 0.16 |

| | | | | |
|---|---|---|---|---|
| polarC18 | 780.55 | 445.4754 | 22 | 0.16 |
| polarC18 | 780.55 | 90.7097 | 21.9 | 0.16 |
| polarC18 | 780.55 | 602.6624 | 21.9 | 0.16 |
| polarC18 | 780.55 | 699.1606 | 21.9 | 0.16 |
| polarC18 | 780.55 | 359.6655 | 21.8 | 0.16 |
| polarC18 | 780.55 | 93.7773 | 21.8 | 0.16 |
| polarC18 | 780.55 | 131.6086 | 21.8 | 0.16 |
| polarC18 | 780.55 | 787.0409 | 21.8 | 0.16 |
| polarC18 | 780.55 | 657.3026 | 21.6 | 0.16 |
| polarC18 | 780.55 | 93.3519 | 21.6 | 0.16 |
| polarC18 | 780.55 | 717.2565 | 21.6 | 0.16 |
| polarC18 | 780.55 | 158.3372 | 21.6 | 0.16 |
| polarC18 | 780.55 | 83.7063 | 21.4 | 0.16 |
| polarC18 | 780.55 | 420.335 | 21.3 | 0.16 |
| polarC18 | 780.55 | 174.6201 | 21.2 | 0.16 |
| polarC18 | 780.55 | 313.1971 | 21.2 | 0.16 |
| polarC18 | 780.55 | 368.4199 | 21.2 | 0.16 |
| polarC18 | 780.55 | 439.4226 | 21 | 0.15 |
| polarC18 | 780.55 | 159.4752 | 20.9 | 0.15 |
| polarC18 | 780.55 | 175.9758 | 20.9 | 0.15 |
| polarC18 | 780.55 | 262.6431 | 20.9 | 0.15 |
| polarC18 | 780.55 | 614.7579 | 20.8 | 0.15 |
| polarC18 | 780.55 | 347.1987 | 20.8 | 0.15 |
| polarC18 | 780.55 | 132.3777 | 20.7 | 0.15 |
| polarC18 | 780.55 | 160.5645 | 20.7 | 0.15 |
| polarC18 | 780.55 | 385.2245 | 20.6 | 0.15 |
| polarC18 | 780.55 | 152.0071 | 20.6 | 0.15 |
| polarC18 | 780.55 | 305.8606 | 20.6 | 0.15 |
| polarC18 | 780.55 | 103.9132 | 20.6 | 0.15 |
| polarC18 | 780.55 | 349.9056 | 20.6 | 0.15 |
| polarC18 | 780.55 | 98.4535 | 20.5 | 0.15 |
| polarC18 | 780.55 | 268.1411 | 20.5 | 0.15 |
| polarC18 | 780.55 | 182.5897 | 20.4 | 0.15 |
| polarC18 | 780.55 | 794.4471 | 20.4 | 0.15 |
| polarC18 | 780.55 | 142.0906 | 20.3 | 0.15 |
| polarC18 | 780.55 | 110.0635 | 20.3 | 0.15 |
| polarC18 | 780.55 | 293.917 | 20.3 | 0.15 |
| polarC18 | 780.55 | 263.2737 | 20.2 | 0.15 |
| polarC18 | 780.55 | 191.2393 | 20.2 | 0.15 |
| polarC18 | 780.55 | 107.8949 | 20.2 | 0.15 |
| polarC18 | 780.55 | 532.5585 | 20.2 | 0.15 |
| polarC18 | 780.55 | 446.3102 | 20.2 | 0.15 |

| | | | | |
|---|---|---|---|---|
| polarC18 | 780.55 | 238.8347 | 20.2 | 0.15 |
| polarC18 | 780.55 | 694.3621 | 20.2 | 0.15 |
| polarC18 | 780.55 | 155.8264 | 20.2 | 0.15 |
| polarC18 | 780.55 | 89.396 | 20.1 | 0.15 |
| polarC18 | 780.55 | 606.85 | 20.1 | 0.15 |
| polarC18 | 780.55 | 628.1871 | 20.1 | 0.15 |
| polarC18 | 780.55 | 211.7772 | 20.1 | 0.15 |
| polarC18 | 780.55 | 83.8062 | 20.1 | 0.15 |
| polarC18 | 780.55 | 171.6143 | 20.1 | 0.15 |
| polarC18 | 780.55 | 526.9846 | 20.1 | 0.15 |
| polarC18 | 780.55 | 346.6279 | 20 | 0.15 |
| polarC18 | 780.55 | 125.5079 | 20 | 0.15 |
| polarC18 | 780.55 | 404.7574 | 20 | 0.15 |
| polarC18 | 780.55 | 749.2847 | 19.9 | 0.15 |
| polarC18 | 780.55 | 525.3806 | 19.9 | 0.15 |
| polarC18 | 780.55 | 231.8393 | 19.9 | 0.15 |
| polarC18 | 780.55 | 121.5326 | 19.9 | 0.15 |
| polarC18 | 780.55 | 508.8489 | 19.9 | 0.15 |
| polarC18 | 780.55 | 205.9483 | 19.9 | 0.15 |
| polarC18 | 780.55 | 133.4432 | 19.8 | 0.15 |
| polarC18 | 780.55 | 534.2809 | 19.8 | 0.14 |
| polarC18 | 780.55 | 516.949 | 19.7 | 0.14 |
| polarC18 | 780.55 | 608.0456 | 19.7 | 0.14 |
| polarC18 | 780.55 | 720.4507 | 19.7 | 0.14 |
| polarC18 | 780.55 | 104.0737 | 19.7 | 0.14 |
| polarC18 | 780.55 | 100.3345 | 19.7 | 0.14 |
| polarC18 | 780.55 | 108.7618 | 19.6 | 0.14 |
| polarC18 | 780.55 | 172.953 | 19.6 | 0.14 |
| polarC18 | 780.55 | 176.4435 | 19.6 | 0.14 |
| polarC18 | 780.55 | 217.3251 | 19.6 | 0.14 |
| polarC18 | 780.55 | 258.0133 | 19.6 | 0.14 |
| polarC18 | 780.55 | 143.5785 | 19.6 | 0.14 |
| polarC18 | 780.55 | 764.2575 | 19.6 | 0.14 |
| polarC18 | 780.55 | 83.6294 | 19.5 | 0.14 |
| polarC18 | 780.55 | 139.2572 | 19.5 | 0.14 |
| polarC18 | 780.55 | 209.1185 | 19.5 | 0.14 |
| polarC18 | 780.55 | 215.0902 | 19.5 | 0.14 |
| polarC18 | 780.55 | 183.7873 | 19.5 | 0.14 |
| polarC18 | 780.55 | 606.1968 | 19.5 | 0.14 |
| polarC18 | 780.55 | 674.0777 | 19.5 | 0.14 |
| polarC18 | 780.55 | 450.588 | 19.5 | 0.14 |
| polarC18 | 780.55 | 204.9352 | 19.4 | 0.14 |

| | | | | |
|---|---|---|---|---|
| polarC18 | 780.55 | 89.3396 | 19.4 | 0.14 |
| polarC18 | 780.55 | 85.7618 | 19.4 | 0.14 |
| polarC18 | 780.55 | 122.8444 | 19.4 | 0.14 |
| polarC18 | 780.55 | 587.7771 | 19.4 | 0.14 |
| polarC18 | 780.55 | 656.8661 | 19.4 | 0.14 |
| polarC18 | 780.55 | 403.5323 | 19.4 | 0.14 |
| polarC18 | 780.55 | 477.981 | 19.4 | 0.14 |
| polarC18 | 780.55 | 613.9094 | 19.4 | 0.14 |
| polarC18 | 780.55 | 221.3344 | 19.3 | 0.14 |
| polarC18 | 780.55 | 538.2393 | 19.3 | 0.14 |
| polarC18 | 780.55 | 585.15 | 19.3 | 0.14 |
| polarC18 | 780.55 | 371.7773 | 19.3 | 0.14 |
| polarC18 | 780.55 | 97.6618 | 19.3 | 0.14 |
| polarC18 | 780.55 | 87.8569 | 19.3 | 0.14 |
| polarC18 | 780.55 | 207.4837 | 19.3 | 0.14 |
| polarC18 | 780.55 | 117.2962 | 19.3 | 0.14 |
| polarC18 | 780.55 | 208.8185 | 19.3 | 0.14 |
| polarC18 | 780.55 | 370.353 | 19.3 | 0.14 |
| polarC18 | 780.55 | 221.1972 | 19.3 | 0.14 |
| polarC18 | 780.55 | 200.0008 | 19.3 | 0.14 |
| polarC18 | 780.55 | 89.0423 | 19.3 | 0.14 |
| polarC18 | 780.55 | 93.6369 | 19.3 | 0.14 |
| polarC18 | 780.55 | 283.8285 | 19.3 | 0.14 |
| polarC18 | 780.55 | 118.4333 | 19.2 | 0.14 |
| polarC18 | 780.55 | 313.8297 | 19.2 | 0.14 |
| polarC18 | 780.55 | 216.4535 | 19.2 | 0.14 |
| polarC18 | 780.55 | 154.1656 | 19.2 | 0.14 |
| polarC18 | 780.55 | 103.7272 | 19.2 | 0.14 |
| polarC18 | 780.55 | 104.82 | 19.2 | 0.14 |
| polarC18 | 780.55 | 426.2618 | 19.2 | 0.14 |
| polarC18 | 780.55 | 310.9397 | 19.2 | 0.14 |
| polarC18 | 780.55 | 160.2106 | 19.2 | 0.14 |
| polarC18 | 780.55 | 145.3937 | 19.2 | 0.14 |
| polarC18 | 780.55 | 423.3299 | 19.2 | 0.14 |
| polarC18 | 780.55 | 518.4963 | 19.1 | 0.14 |
| polarC18 | 780.55 | 272.9286 | 19.1 | 0.14 |
| polarC18 | 780.55 | 416.0953 | 19.1 | 0.14 |
| polarC18 | 780.55 | 324.9832 | 19.1 | 0.14 |
| polarC18 | 780.55 | 107.1714 | 19.1 | 0.14 |
| polarC18 | 780.55 | 311.0839 | 19.1 | 0.14 |
| polarC18 | 780.55 | 409.1029 | 19.1 | 0.14 |
| polarC18 | 780.55 | 698.9557 | 19.1 | 0.14 |

| | | | | |
|---|---|---|---|---|
| polarC18 | 780.55 | 433.337 | 19.1 | 0.14 |
| polarC18 | 780.55 | 203.2614 | 19.1 | 0.14 |
| polarC18 | 780.55 | 93.6109 | 19.1 | 0.14 |
| polarC18 | 780.55 | 523.1078 | 19.1 | 0.14 |
| polarC18 | 780.55 | 763.2321 | 19 | 0.14 |
| polarC18 | 780.55 | 540.3949 | 19 | 0.14 |
| polarC18 | 780.55 | 510.2842 | 19 | 0.14 |
| polarC18 | 780.55 | 233.5483 | 19 | 0.14 |
| polarC18 | 780.55 | 126.5381 | 19 | 0.14 |
| polarC18 | 780.55 | 673.5333 | 19 | 0.14 |
| polarC18 | 780.55 | 339.0753 | 19 | 0.14 |
| polarC18 | 780.55 | 300.4089 | 19 | 0.14 |
| polarC18 | 780.55 | 162.1953 | 19 | 0.14 |
| polarC18 | 780.55 | 109.6328 | 19 | 0.14 |
| polarC18 | 780.55 | 679.4779 | 19 | 0.14 |
| polarC18 | 780.55 | 114.2951 | 19 | 0.14 |
| polarC18 | 780.55 | 110.0512 | 19 | 0.14 |
| polarC18 | 780.55 | 141.9499 | 19 | 0.14 |
| polarC18 | 780.55 | 455.2015 | 19 | 0.14 |
| polarC18 | 780.55 | 277.8745 | 19 | 0.14 |
| polarC18 | 780.55 | 92.6679 | 18.9 | 0.14 |
| polarC18 | 780.55 | 684.5983 | 18.9 | 0.14 |
| polarC18 | 780.55 | 84.8789 | 18.9 | 0.14 |
| polarC18 | 780.55 | 359.5294 | 18.9 | 0.14 |
| polarC18 | 780.55 | 271.0963 | 18.9 | 0.14 |
| polarC18 | 780.55 | 128.6758 | 18.9 | 0.14 |
| polarC18 | 780.55 | 203.5649 | 18.9 | 0.14 |
| polarC18 | 780.55 | 109.6246 | 18.9 | 0.14 |
| polarC18 | 780.55 | 179.0191 | 18.9 | 0.14 |
| polarC18 | 780.55 | 92.977 | 18.9 | 0.14 |
| polarC18 | 780.55 | 97.5042 | 18.9 | 0.14 |
| polarC18 | 780.55 | 130.3285 | 18.9 | 0.14 |
| polarC18 | 780.55 | 305.6243 | 18.9 | 0.14 |
| polarC18 | 780.55 | 115.5755 | 18.9 | 0.14 |
| polarC18 | 780.55 | 96.0344 | 18.9 | 0.14 |
| polarC18 | 780.55 | 662.4789 | 18.8 | 0.14 |
| polarC18 | 780.55 | 162.5901 | 18.8 | 0.14 |
| polarC18 | 780.55 | 697.8199 | 18.8 | 0.14 |
| polarC18 | 780.55 | 114.0807 | 18.8 | 0.14 |
| polarC18 | 780.55 | 178.3116 | 18.8 | 0.14 |
| polarC18 | 780.55 | 149.8313 | 18.8 | 0.14 |
| polarC18 | 780.55 | 298.4108 | 18.8 | 0.14 |

| | | | | |
|---|---|---|---|---|
| polarC18 | 780.55 | 486.4678 | 18.8 | 0.14 |
| polarC18 | 780.55 | 676.848 | 18.8 | 0.14 |
| polarC18 | 780.55 | 107.7225 | 18.8 | 0.14 |
| polarC18 | 780.55 | 141.3361 | 18.8 | 0.14 |
| polarC18 | 780.55 | 257.9247 | 18.8 | 0.14 |
| polarC18 | 780.55 | 670.58 | 18.8 | 0.14 |
| polarC18 | 780.55 | 104.5496 | 18.8 | 0.14 |
| polarC18 | 780.55 | 89.7061 | 18.8 | 0.14 |
| polarC18 | 780.55 | 241.7823 | 18.8 | 0.14 |
| polarC18 | 780.55 | 216.1815 | 18.8 | 0.14 |
| polarC18 | 780.55 | 599.9337 | 18.7 | 0.14 |
| polarC18 | 780.55 | 96.4657 | 18.7 | 0.14 |
| polarC18 | 780.55 | 506.9518 | 18.7 | 0.14 |
| polarC18 | 780.55 | 749.6773 | 18.7 | 0.14 |
| polarC18 | 780.55 | 161.7487 | 18.7 | 0.14 |
| polarC18 | 780.55 | 623.9009 | 18.7 | 0.14 |
| polarC18 | 780.55 | 113.6186 | 18.7 | 0.14 |
| polarC18 | 780.55 | 201.4609 | 18.7 | 0.14 |
| polarC18 | 780.55 | 122.8997 | 18.7 | 0.14 |
| polarC18 | 780.55 | 487.6586 | 18.7 | 0.14 |
| polarC18 | 780.55 | 365.2023 | 18.7 | 0.14 |
| polarC18 | 780.55 | 421.5054 | 18.7 | 0.14 |
| polarC18 | 780.55 | 491.483 | 18.7 | 0.14 |
| polarC18 | 780.55 | 142.8744 | 18.7 | 0.14 |
| polarC18 | 780.55 | 151.4686 | 18.7 | 0.14 |
| polarC18 | 780.55 | 110.4763 | 18.7 | 0.14 |
| polarC18 | 780.55 | 123.8824 | 18.7 | 0.14 |
| polarC18 | 780.55 | 274.8039 | 18.6 | 0.14 |
| polarC18 | 780.55 | 234.4355 | 18.6 | 0.14 |
| polarC18 | 780.55 | 147.5202 | 18.6 | 0.14 |
| polarC18 | 780.55 | 86.1427 | 18.6 | 0.14 |
| polarC18 | 780.55 | 130.3214 | 18.6 | 0.14 |
| polarC18 | 780.55 | 85.2953 | 18.6 | 0.14 |
| polarC18 | 780.55 | 778.4258 | 18.6 | 0.14 |
| polarC18 | 780.55 | 453.8992 | 18.6 | 0.14 |
| polarC18 | 780.55 | 427.4113 | 18.6 | 0.14 |
| polarC18 | 780.55 | 150.2756 | 18.6 | 0.14 |
| polarC18 | 780.55 | 88.2887 | 18.6 | 0.14 |
| polarC18 | 780.55 | 94.1896 | 18.6 | 0.14 |
| polarC18 | 780.55 | 247.5222 | 18.6 | 0.14 |
| polarC18 | 780.55 | 107.8885 | 18.5 | 0.14 |
| polarC18 | 780.55 | 200.2614 | 18.5 | 0.14 |

| | | | | |
|---|---|---|---|---|
| polarC18 | 780.55 | 123.2593 | 18.5 | 0.14 |
| polarC18 | 780.55 | 496.5591 | 18.5 | 0.14 |
| polarC18 | 780.55 | 118.4491 | 18.5 | 0.14 |
| polarC18 | 780.55 | 506.3733 | 18.5 | 0.14 |
| polarC18 | 780.55 | 93.7868 | 18.5 | 0.14 |
| polarC18 | 780.55 | 383.2086 | 18.5 | 0.14 |
| polarC18 | 780.55 | 426.0486 | 18.5 | 0.14 |
| polarC18 | 780.55 | 152.273 | 18.5 | 0.14 |
| polarC18 | 780.55 | 359.9579 | 18.5 | 0.14 |
| polarC18 | 780.55 | 516.3781 | 18.5 | 0.14 |
| polarC18 | 780.55 | 181.0245 | 18.5 | 0.14 |
| polarC18 | 780.55 | 84.7068 | 18.4 | 0.14 |
| polarC18 | 780.55 | 162.2931 | 18.4 | 0.14 |
| polarC18 | 780.55 | 103.7316 | 18.4 | 0.14 |
| polarC18 | 780.55 | 416.4148 | 18.4 | 0.13 |
| polarC18 | 780.55 | 308.9766 | 18.4 | 0.13 |
| polarC18 | 780.55 | 94.3796 | 18.4 | 0.13 |
| polarC18 | 780.55 | 739.6901 | 18.4 | 0.13 |
| polarC18 | 780.55 | 125.71 | 18.4 | 0.13 |
| polarC18 | 780.55 | 385.5891 | 18.4 | 0.13 |
| polarC18 | 780.55 | 165.3496 | 18.4 | 0.13 |
| polarC18 | 780.55 | 374.5308 | 18.4 | 0.13 |
| polarC18 | 780.55 | 88.0282 | 18.4 | 0.13 |
| polarC18 | 780.55 | 727.7264 | 18.4 | 0.13 |
| polarC18 | 780.55 | 96.7242 | 18.4 | 0.13 |
| polarC18 | 780.55 | 89.6778 | 18.4 | 0.13 |
| polarC18 | 780.55 | 353.1498 | 18.4 | 0.13 |
| polarC18 | 780.55 | 175.5565 | 18.4 | 0.13 |
| polarC18 | 780.55 | 84.6397 | 18.4 | 0.13 |
| polarC18 | 780.55 | 346.1884 | 18.3 | 0.13 |
| polarC18 | 780.55 | 297.7303 | 18.3 | 0.13 |
| polarC18 | 780.55 | 351.5821 | 18.3 | 0.13 |
| polarC18 | 780.55 | 329.0854 | 18.3 | 0.13 |
| polarC18 | 780.55 | 186.3926 | 18.3 | 0.13 |
| polarC18 | 780.55 | 485.2431 | 18.3 | 0.13 |
| polarC18 | 780.55 | 623.4019 | 18.3 | 0.13 |
| polarC18 | 780.55 | 83.5998 | 18.3 | 0.13 |
| polarC18 | 780.55 | 86.315 | 18.3 | 0.13 |
| polarC18 | 780.55 | 467.851 | 18.3 | 0.13 |
| polarC18 | 780.55 | 110.012 | 18.3 | 0.13 |
| polarC18 | 780.55 | 114.3872 | 18.3 | 0.13 |
| polarC18 | 780.55 | 617.8271 | 18.3 | 0.13 |

| | | | | |
|---|---|---|---|---|
| polarC18 | 780.55 | 645.4927 | 18.3 | 0.13 |
| polarC18 | 780.55 | 252.2062 | 18.3 | 0.13 |
| polarC18 | 780.55 | 174.6406 | 18.3 | 0.13 |
| polarC18 | 780.55 | 145.1539 | 18.3 | 0.13 |
| polarC18 | 780.55 | 686.4603 | 18.3 | 0.13 |
| polarC18 | 780.55 | 251.6193 | 18.3 | 0.13 |
| polarC18 | 780.55 | 320.335 | 18.3 | 0.13 |
| polarC18 | 780.55 | 434.2332 | 18.3 | 0.13 |
| polarC18 | 780.55 | 144.8706 | 18.2 | 0.13 |
| polarC18 | 780.55 | 477.7129 | 18.2 | 0.13 |
| polarC18 | 780.55 | 141.9652 | 18.2 | 0.13 |
| polarC18 | 780.55 | 537.9164 | 18.2 | 0.13 |
| polarC18 | 780.55 | 99.1292 | 18.2 | 0.13 |
| polarC18 | 780.55 | 122.818 | 18.2 | 0.13 |
| polarC18 | 780.55 | 366.2675 | 18.2 | 0.13 |
| polarC18 | 780.55 | 86.7475 | 18.2 | 0.13 |
| polarC18 | 780.55 | 465.9141 | 18.2 | 0.13 |
| polarC18 | 780.55 | 474.4859 | 18.2 | 0.13 |
| polarC18 | 780.55 | 133.3754 | 18.2 | 0.13 |
| polarC18 | 780.55 | 533.1838 | 18.2 | 0.13 |
| polarC18 | 780.55 | 223.6055 | 18.2 | 0.13 |
| polarC18 | 780.55 | 395.7652 | 18.2 | 0.13 |
| polarC18 | 780.55 | 179.3676 | 18.2 | 0.13 |
| polarC18 | 780.55 | 200.5429 | 18.2 | 0.13 |
| polarC18 | 780.55 | 88.5485 | 18.2 | 0.13 |
| polarC18 | 780.55 | 107.7475 | 18.2 | 0.13 |
| polarC18 | 780.55 | 766.5386 | 18.2 | 0.13 |
| polarC18 | 780.55 | 122.3749 | 18.2 | 0.13 |
| polarC18 | 780.55 | 327.9666 | 18.2 | 0.13 |
| polarC18 | 780.55 | 284.623 | 18.2 | 0.13 |
| polarC18 | 780.55 | 246.022 | 18.2 | 0.13 |
| polarC18 | 780.55 | 688.7581 | 18.1 | 0.13 |
| polarC18 | 780.55 | 340.0984 | 18.1 | 0.13 |
| polarC18 | 780.55 | 206.8502 | 18.1 | 0.13 |
| polarC18 | 780.55 | 228.4104 | 18.1 | 0.13 |
| polarC18 | 780.55 | 94.4453 | 18.1 | 0.13 |
| polarC18 | 780.55 | 186.4831 | 18.1 | 0.13 |
| polarC18 | 780.55 | 394.2285 | 18.1 | 0.13 |
| polarC18 | 780.55 | 149.1652 | 18.1 | 0.13 |
| polarC18 | 780.55 | 399.2304 | 18.1 | 0.13 |
| polarC18 | 780.55 | 201.1723 | 18.1 | 0.13 |
| polarC18 | 780.55 | 88.599 | 18.1 | 0.13 |

| | | | | |
|---|---|---|---|---|
| polarC18 | 780.55 | 759.3779 | 18.1 | 0.13 |
| polarC18 | 780.55 | 542.4419 | 18.1 | 0.13 |
| polarC18 | 780.55 | 111.0431 | 18.1 | 0.13 |
| polarC18 | 780.55 | 116.9302 | 18.1 | 0.13 |
| polarC18 | 780.55 | 108.1477 | 18.1 | 0.13 |
| polarC18 | 780.55 | 192.8911 | 18.1 | 0.13 |
| polarC18 | 780.55 | 234.5124 | 18.1 | 0.13 |
| polarC18 | 780.55 | 659.9413 | 18.1 | 0.13 |
| polarC18 | 780.55 | 578.6972 | 18.1 | 0.13 |
| polarC18 | 780.55 | 583.2266 | 18.1 | 0.13 |
| polarC18 | 780.55 | 275.292 | 18.1 | 0.13 |
| polarC18 | 780.55 | 260.2205 | 18.1 | 0.13 |
| polarC18 | 780.55 | 656.0362 | 18.1 | 0.13 |
| polarC18 | 780.55 | 127.3053 | 18.1 | 0.13 |
| polarC18 | 780.55 | 430.1944 | 18.1 | 0.13 |
| polarC18 | 780.55 | 808.645 | 18.1 | 0.13 |
| polarC18 | 780.55 | 660.1193 | 18.1 | 0.13 |
| polarC18 | 780.55 | 349.3788 | 18.1 | 0.13 |
| polarC18 | 780.55 | 97.9977 | 18.1 | 0.13 |
| polarC18 | 780.55 | 96.4781 | 18.1 | 0.13 |
| polarC18 | 780.55 | 271.3192 | 18.1 | 0.13 |
| polarC18 | 780.55 | 389.114 | 18 | 0.13 |
| polarC18 | 780.55 | 129.915 | 18 | 0.13 |
| polarC18 | 780.55 | 82.0857 | 18 | 0.13 |
| polarC18 | 780.55 | 791.0413 | 18 | 0.13 |
| polarC18 | 780.55 | 231.0611 | 18 | 0.13 |
| polarC18 | 780.55 | 132.3232 | 18 | 0.13 |
| polarC18 | 780.55 | 87.6893 | 18 | 0.13 |
| polarC18 | 780.55 | 106.4094 | 18 | 0.13 |
| polarC18 | 780.55 | 288.1973 | 18 | 0.13 |
| polarC18 | 780.55 | 367.0482 | 18 | 0.13 |
| polarC18 | 780.55 | 107.3351 | 18 | 0.13 |
| polarC18 | 780.55 | 127.8331 | 18 | 0.13 |
| polarC18 | 780.55 | 256.6865 | 18 | 0.13 |
| polarC18 | 780.55 | 134.5222 | 18 | 0.13 |
| polarC18 | 780.55 | 223.9433 | 18 | 0.13 |
| polarC18 | 780.55 | 113.3453 | 18 | 0.13 |
| polarC18 | 780.55 | 667.9128 | 18 | 0.13 |
| polarC18 | 780.55 | 121.2703 | 18 | 0.13 |
| polarC18 | 780.55 | 84.7037 | 18 | 0.13 |
| polarC18 | 780.55 | 447.2619 | 18 | 0.13 |
| polarC18 | 780.55 | 125.9187 | 18 | 0.13 |

| | | | | |
|---|---|---|---|---|
| polarC18 | 780.55 | 123.7698 | 18 | 0.13 |
| polarC18 | 780.55 | 124.4729 | 18 | 0.13 |
| polarC18 | 780.55 | 144.331 | 18 | 0.13 |
| polarC18 | 780.55 | 177.5902 | 18 | 0.13 |
| polarC18 | 780.55 | 743.0566 | 18 | 0.13 |
| polarC18 | 780.55 | 163.5615 | 18 | 0.13 |
| polarC18 | 780.55 | 167.4774 | 18 | 0.13 |
| polarC18 | 780.55 | 630.3312 | 18 | 0.13 |
| polarC18 | 780.55 | 779.5759 | 18 | 0.13 |
| polarC18 | 780.55 | 165.212 | 18 | 0.13 |
| polarC18 | 780.55 | 140.8867 | 18 | 0.13 |
| polarC18 | 780.55 | 436.8356 | 18 | 0.13 |
| polarC18 | 780.55 | 342.3777 | 18 | 0.13 |
| polarC18 | 780.55 | 202.4345 | 17.9 | 0.13 |
| polarC18 | 780.55 | 100.6046 | 17.9 | 0.13 |
| polarC18 | 780.55 | 569.8442 | 17.9 | 0.13 |
| polarC18 | 780.55 | 91.2011 | 17.9 | 0.13 |
| polarC18 | 780.55 | 120.294 | 17.9 | 0.13 |
| polarC18 | 780.55 | 106.3876 | 17.9 | 0.13 |
| polarC18 | 780.55 | 167.9305 | 17.9 | 0.13 |
| polarC18 | 780.55 | 511.2027 | 17.9 | 0.13 |
| polarC18 | 780.55 | 792.3279 | 17.9 | 0.13 |
| polarC18 | 780.55 | 192.7133 | 17.9 | 0.13 |
| polarC18 | 780.55 | 130.3841 | 17.9 | 0.13 |
| polarC18 | 780.55 | 109.9411 | 17.9 | 0.13 |
| polarC18 | 780.55 | 126.5187 | 17.9 | 0.13 |
| polarC18 | 780.55 | 516.6403 | 17.9 | 0.13 |
| polarC18 | 780.55 | 377.6399 | 17.9 | 0.13 |
| polarC18 | 780.55 | 95.508 | 17.9 | 0.13 |
| polarC18 | 780.55 | 380.2228 | 17.9 | 0.13 |
| polarC18 | 780.55 | 677.8881 | 17.9 | 0.13 |
| polarC18 | 780.55 | 292.2462 | 17.9 | 0.13 |
| polarC18 | 780.55 | 354.9938 | 17.9 | 0.13 |
| polarC18 | 780.55 | 280.0181 | 17.9 | 0.13 |
| polarC18 | 780.55 | 85.6544 | 17.9 | 0.13 |
| polarC18 | 780.55 | 371.1897 | 17.9 | 0.13 |
| polarC18 | 780.55 | 478.149 | 17.9 | 0.13 |
| polarC18 | 780.55 | 533.5838 | 17.9 | 0.13 |
| polarC18 | 780.55 | 173.011 | 17.9 | 0.13 |
| polarC18 | 780.55 | 108.0415 | 17.9 | 0.13 |
| polarC18 | 780.55 | 732.9974 | 17.9 | 0.13 |
| polarC18 | 780.55 | 338.8432 | 17.9 | 0.13 |

| | | | | |
|---|---|---|---|---|
| polarC18 | 780.55 | 438.528 | 17.9 | 0.13 |
| polarC18 | 780.55 | 114.9072 | 17.9 | 0.13 |
| polarC18 | 780.55 | 252.543 | 17.9 | 0.13 |
| polarC18 | 780.55 | 137.1199 | 17.8 | 0.13 |
| polarC18 | 780.55 | 375.2031 | 17.8 | 0.13 |
| polarC18 | 780.55 | 155.7182 | 17.8 | 0.13 |
| polarC18 | 780.55 | 580.054 | 17.8 | 0.13 |
| polarC18 | 780.55 | 99.6705 | 17.8 | 0.13 |
| polarC18 | 780.55 | 612.5027 | 17.8 | 0.13 |
| polarC18 | 780.55 | 475.5145 | 17.8 | 0.13 |
| polarC18 | 780.55 | 426.9501 | 17.8 | 0.13 |
| polarC18 | 780.55 | 91.09 | 17.8 | 0.13 |
| polarC18 | 780.55 | 437.4205 | 17.8 | 0.13 |
| polarC18 | 780.55 | 571.2679 | 17.8 | 0.13 |
| polarC18 | 780.55 | 389.5779 | 17.8 | 0.13 |
| polarC18 | 780.55 | 478.5845 | 17.8 | 0.13 |
| polarC18 | 780.55 | 90.3217 | 17.8 | 0.13 |
| polarC18 | 780.55 | 166.555 | 17.8 | 0.13 |
| polarC18 | 780.55 | 130.8765 | 17.8 | 0.13 |
| polarC18 | 780.55 | 586.6256 | 17.8 | 0.13 |
| polarC18 | 780.55 | 761.2307 | 17.8 | 0.13 |
| polarC18 | 780.55 | 388.7827 | 17.8 | 0.13 |
| polarC18 | 780.55 | 256.8186 | 17.8 | 0.13 |
| polarC18 | 780.55 | 133.6661 | 17.8 | 0.13 |
| polarC18 | 780.55 | 273.7661 | 17.8 | 0.13 |
| polarC18 | 780.55 | 328.9935 | 17.8 | 0.13 |
| polarC18 | 780.55 | 276.3486 | 17.8 | 0.13 |
| polarC18 | 780.55 | 204.1629 | 17.8 | 0.13 |
| polarC18 | 780.55 | 97.4579 | 17.8 | 0.13 |
| polarC18 | 780.55 | 448.6243 | 17.8 | 0.13 |
| polarC18 | 780.55 | 171.0727 | 17.7 | 0.13 |
| polarC18 | 780.55 | 144.6755 | 17.7 | 0.13 |
| polarC18 | 780.55 | 265.2722 | 17.7 | 0.13 |
| polarC18 | 780.55 | 185.0405 | 17.7 | 0.13 |
| polarC18 | 780.55 | 183.7711 | 17.7 | 0.13 |
| polarC18 | 780.55 | 164.3995 | 17.7 | 0.13 |
| polarC18 | 780.55 | 808.3275 | 17.7 | 0.13 |
| polarC18 | 780.55 | 156.5515 | 17.7 | 0.13 |
| polarC18 | 780.55 | 116.554 | 17.7 | 0.13 |
| polarC18 | 780.55 | 311.7678 | 17.7 | 0.13 |
| polarC18 | 780.55 | 756.212 | 17.7 | 0.13 |
| polarC18 | 780.55 | 601.0919 | 17.7 | 0.13 |

| | | | | |
|---|---|---|---|---|
| polarC18 | 780.55 | 214.1217 | 17.7 | 0.13 |
| polarC18 | 780.55 | 715.4059 | 17.7 | 0.13 |
| polarC18 | 780.55 | 522.3417 | 17.7 | 0.13 |
| polarC18 | 780.55 | 245.159 | 17.7 | 0.13 |
| polarC18 | 780.55 | 126.1444 | 17.7 | 0.13 |
| polarC18 | 780.55 | 467.6942 | 17.7 | 0.13 |
| polarC18 | 780.55 | 221.0997 | 17.7 | 0.13 |
| polarC18 | 780.55 | 88.2009 | 17.7 | 0.13 |
| polarC18 | 780.55 | 104.4181 | 17.7 | 0.13 |
| polarC18 | 780.55 | 338.4009 | 17.7 | 0.13 |
| polarC18 | 780.55 | 146.0348 | 17.7 | 0.13 |
| polarC18 | 780.55 | 165.0564 | 17.7 | 0.13 |
| polarC18 | 780.55 | 209.3783 | 17.7 | 0.13 |
| polarC18 | 780.55 | 806.7352 | 17.7 | 0.13 |
| polarC18 | 780.55 | 153.6308 | 17.7 | 0.13 |
| polarC18 | 780.55 | 585.8847 | 17.7 | 0.13 |
| polarC18 | 780.55 | 738.3553 | 17.7 | 0.13 |
| polarC18 | 780.55 | 105.0158 | 17.7 | 0.13 |
| polarC18 | 780.55 | 176.7767 | 17.7 | 0.13 |
| polarC18 | 780.55 | 209.5217 | 17.7 | 0.13 |
| polarC18 | 780.55 | 642.5451 | 17.7 | 0.13 |
| polarC18 | 780.55 | 265.3338 | 17.7 | 0.13 |
| polarC18 | 780.55 | 157.2629 | 17.7 | 0.13 |
| polarC18 | 780.55 | 146.8426 | 17.7 | 0.13 |
| polarC18 | 780.55 | 370.5045 | 17.7 | 0.13 |
| polarC18 | 780.55 | 188.2559 | 17.7 | 0.13 |
| polarC18 | 780.55 | 92.968 | 17.7 | 0.13 |
| polarC18 | 780.55 | 374.3582 | 17.7 | 0.13 |
| polarC18 | 780.55 | 214.0309 | 17.7 | 0.13 |
| polarC18 | 780.55 | 169.0923 | 17.7 | 0.13 |
| polarC18 | 780.55 | 339.1258 | 17.7 | 0.13 |
| polarC18 | 780.55 | 451.938 | 17.7 | 0.13 |
| polarC18 | 780.55 | 312.0522 | 17.7 | 0.13 |
| polarC18 | 780.55 | 238.2492 | 17.7 | 0.13 |
| polarC18 | 780.55 | 252.7257 | 17.7 | 0.13 |
| polarC18 | 780.55 | 160.4292 | 17.7 | 0.13 |
| polarC18 | 780.55 | 159.5124 | 17.7 | 0.13 |
| polarC18 | 780.55 | 219.8668 | 17.6 | 0.13 |
| polarC18 | 780.55 | 522.0192 | 17.6 | 0.13 |
| polarC18 | 780.55 | 87.4548 | 17.6 | 0.13 |
| polarC18 | 780.55 | 120.0386 | 17.6 | 0.13 |
| polarC18 | 780.55 | 431.9569 | 17.6 | 0.13 |

| | | | | |
|---|---|---|---|---|
| polarC18 | 780.55 | 197.5866 | 17.6 | 0.13 |
| polarC18 | 780.55 | 152.56 | 17.6 | 0.13 |
| polarC18 | 780.55 | 191.5879 | 17.6 | 0.13 |
| polarC18 | 780.55 | 290.9662 | 17.6 | 0.13 |
| polarC18 | 780.55 | 92.5965 | 17.6 | 0.13 |
| polarC18 | 780.55 | 132.7227 | 17.6 | 0.13 |
| polarC18 | 780.55 | 111.325 | 17.6 | 0.13 |
| polarC18 | 780.55 | 628.5511 | 17.6 | 0.13 |
| polarC18 | 780.55 | 252.8447 | 17.6 | 0.13 |
| polarC18 | 780.55 | 209.3348 | 17.6 | 0.13 |
| polarC18 | 780.55 | 377.7673 | 17.6 | 0.13 |
| polarC18 | 780.55 | 311.6643 | 17.6 | 0.13 |
| polarC18 | 780.55 | 437.534 | 17.6 | 0.13 |
| polarC18 | 780.55 | 140.8158 | 17.6 | 0.13 |
| polarC18 | 780.55 | 192.3813 | 17.6 | 0.13 |
| polarC18 | 780.55 | 768.6109 | 17.6 | 0.13 |
| polarC18 | 780.55 | 91.0283 | 17.6 | 0.13 |
| polarC18 | 780.55 | 93.5917 | 17.6 | 0.13 |
| polarC18 | 780.55 | 89.0512 | 17.6 | 0.13 |
| polarC18 | 780.55 | 136.8699 | 17.6 | 0.13 |
| polarC18 | 780.55 | 120.665 | 17.6 | 0.13 |
| polarC18 | 780.55 | 122.2993 | 17.6 | 0.13 |
| polarC18 | 780.55 | 115.2719 | 17.6 | 0.13 |
| polarC18 | 780.55 | 776.0692 | 17.6 | 0.13 |
| polarC18 | 780.55 | 405.6598 | 17.6 | 0.13 |
| polarC18 | 780.55 | 293.322 | 17.6 | 0.13 |
| polarC18 | 780.55 | 101.9975 | 17.6 | 0.13 |
| polarC18 | 780.55 | 237.9938 | 17.6 | 0.13 |
| polarC18 | 780.55 | 605.0222 | 17.5 | 0.13 |
| polarC18 | 780.55 | 92.0007 | 17.5 | 0.13 |
| polarC18 | 780.55 | 213.3926 | 17.5 | 0.13 |
| polarC18 | 780.55 | 364.1302 | 17.5 | 0.13 |
| polarC18 | 780.55 | 369.9346 | 17.5 | 0.13 |
| polarC18 | 780.55 | 417.8219 | 17.5 | 0.13 |
| polarC18 | 780.55 | 132.1674 | 17.5 | 0.13 |
| polarC18 | 780.55 | 342.3264 | 17.5 | 0.13 |
| polarC18 | 780.55 | 196.5788 | 17.5 | 0.13 |
| polarC18 | 780.55 | 143.0754 | 17.5 | 0.13 |
| polarC18 | 780.55 | 649.5455 | 17.5 | 0.13 |
| polarC18 | 780.55 | 243.7365 | 17.5 | 0.13 |
| polarC18 | 780.55 | 179.7442 | 17.5 | 0.13 |
| polarC18 | 780.55 | 376.2765 | 17.5 | 0.13 |

| | | | | |
|---|---|---|---|---|
| polarC18 | 780.55 | 525.2674 | 17.5 | 0.13 |
| polarC18 | 780.55 | 87.6677 | 17.5 | 0.13 |
| polarC18 | 780.55 | 324.534 | 17.5 | 0.13 |
| polarC18 | 780.55 | 596.9757 | 17.5 | 0.13 |
| polarC18 | 780.55 | 85.8481 | 17.5 | 0.13 |
| polarC18 | 780.55 | 543.7507 | 17.5 | 0.13 |
| polarC18 | 780.55 | 443.0193 | 17.5 | 0.13 |
| polarC18 | 780.55 | 560.7054 | 17.5 | 0.13 |
| polarC18 | 780.55 | 206.9204 | 17.5 | 0.13 |
| polarC18 | 780.55 | 104.0295 | 17.5 | 0.13 |
| polarC18 | 780.55 | 582.0799 | 17.5 | 0.13 |
| polarC18 | 780.55 | 632.6317 | 17.5 | 0.13 |
| polarC18 | 780.55 | 653.3373 | 17.5 | 0.13 |
| polarC18 | 780.55 | 199.4119 | 17.5 | 0.13 |
| polarC18 | 780.55 | 774.1813 | 17.5 | 0.13 |
| polarC18 | 780.55 | 507.7812 | 17.5 | 0.13 |
| polarC18 | 780.55 | 239.9754 | 17.4 | 0.13 |
| polarC18 | 780.55 | 182.3228 | 17.4 | 0.13 |
| polarC18 | 780.55 | 498.2672 | 17.4 | 0.13 |
| polarC18 | 780.55 | 634.6547 | 17.4 | 0.13 |
| polarC18 | 780.55 | 369.35 | 17.4 | 0.13 |
| polarC18 | 780.55 | 190.6703 | 17.4 | 0.13 |
| polarC18 | 780.55 | 575.3846 | 17.4 | 0.13 |
| polarC18 | 780.55 | 87.0758 | 17.4 | 0.13 |
| polarC18 | 780.55 | 199.2549 | 17.4 | 0.13 |
| polarC18 | 780.55 | 240.8148 | 17.4 | 0.13 |
| polarC18 | 780.55 | 305.5498 | 17.4 | 0.13 |
| polarC18 | 780.55 | 148.4292 | 17.4 | 0.13 |
| polarC18 | 780.55 | 111.293 | 17.4 | 0.13 |
| polarC18 | 780.55 | 175.5414 | 17.4 | 0.13 |
| polarC18 | 780.55 | 171.8122 | 17.4 | 0.13 |
| polarC18 | 780.55 | 133.1052 | 17.4 | 0.13 |
| polarC18 | 780.55 | 714.8413 | 17.4 | 0.13 |
| polarC18 | 780.55 | 581.7944 | 17.4 | 0.13 |
| polarC18 | 780.55 | 345.119 | 17.4 | 0.13 |
| polarC18 | 780.55 | 90.8812 | 17.4 | 0.13 |
| polarC18 | 780.55 | 205.9232 | 17.4 | 0.13 |
| polarC18 | 780.55 | 127.7478 | 17.4 | 0.13 |
| polarC18 | 780.55 | 210.8712 | 17.4 | 0.13 |
| polarC18 | 780.55 | 90.634 | 17.4 | 0.13 |
| polarC18 | 780.55 | 137.8439 | 17.4 | 0.13 |
| polarC18 | 780.55 | 87.7251 | 17.4 | 0.13 |

| | | | | |
|---|---|---|---|---|
| polarC18 | 780.55 | 593.5596 | 17.4 | 0.13 |
| polarC18 | 780.55 | 131.2497 | 17.4 | 0.13 |
| polarC18 | 780.55 | 165.0843 | 17.4 | 0.13 |
| polarC18 | 780.55 | 234.1838 | 17.4 | 0.13 |
| polarC18 | 780.55 | 444.432 | 17.4 | 0.13 |
| polarC18 | 780.55 | 160.2572 | 17.4 | 0.13 |
| polarC18 | 780.55 | 252.104 | 17.4 | 0.13 |
| polarC18 | 780.55 | 130.3519 | 17.4 | 0.13 |
| polarC18 | 780.55 | 172.0081 | 17.4 | 0.13 |
| polarC18 | 780.55 | 574.3201 | 17.4 | 0.13 |
| polarC18 | 780.55 | 743.96 | 17.4 | 0.13 |
| polarC18 | 780.55 | 95.3053 | 17.4 | 0.13 |
| polarC18 | 780.55 | 642.2093 | 17.4 | 0.13 |
| polarC18 | 780.55 | 115.5595 | 17.4 | 0.13 |
| polarC18 | 780.55 | 296.3458 | 17.4 | 0.13 |
| polarC18 | 780.55 | 586.819 | 17.4 | 0.13 |
| polarC18 | 780.55 | 340.8916 | 17.4 | 0.13 |
| polarC18 | 780.55 | 778.6991 | 17.4 | 0.13 |
| polarC18 | 780.55 | 158.4224 | 17.3 | 0.13 |
| polarC18 | 780.55 | 661.6775 | 17.3 | 0.13 |
| polarC18 | 780.55 | 337.4144 | 17.3 | 0.13 |
| polarC18 | 780.55 | 331.5157 | 17.3 | 0.13 |
| polarC18 | 780.55 | 105.1104 | 17.3 | 0.13 |
| polarC18 | 780.55 | 99.5644 | 17.3 | 0.13 |
| polarC18 | 780.55 | 378.4224 | 17.3 | 0.13 |
| polarC18 | 780.55 | 137.3291 | 17.3 | 0.13 |
| polarC18 | 780.55 | 88.5784 | 17.3 | 0.13 |
| polarC18 | 780.55 | 138.1875 | 17.3 | 0.13 |
| polarC18 | 780.55 | 425.0501 | 17.3 | 0.13 |
| polarC18 | 780.55 | 804.7794 | 17.3 | 0.13 |
| polarC18 | 780.55 | 84.4133 | 17.3 | 0.13 |
| polarC18 | 780.55 | 417.4853 | 17.3 | 0.13 |
| polarC18 | 780.55 | 328.1749 | 17.3 | 0.13 |
| polarC18 | 780.55 | 94.5584 | 17.3 | 0.13 |
| polarC18 | 780.55 | 174.5305 | 17.3 | 0.13 |
| polarC18 | 780.55 | 90.0803 | 17.3 | 0.13 |
| polarC18 | 780.55 | 218.5245 | 17.3 | 0.13 |
| polarC18 | 780.55 | 290.1284 | 17.3 | 0.13 |
| polarC18 | 780.55 | 275.2532 | 17.3 | 0.13 |
| polarC18 | 780.55 | 474.2948 | 17.3 | 0.13 |
| polarC18 | 780.55 | 688.9545 | 17.3 | 0.13 |
| polarC18 | 780.55 | 217.1105 | 17.3 | 0.13 |

| | | | | |
|---|---|---|---|---|
| polarC18 | 780.55 | 135.3559 | 17.3 | 0.13 |
| polarC18 | 780.55 | 698.1186 | 17.3 | 0.13 |
| polarC18 | 780.55 | 236.2345 | 17.3 | 0.13 |
| polarC18 | 780.55 | 123.0682 | 17.3 | 0.13 |
| polarC18 | 780.55 | 236.9957 | 17.3 | 0.13 |
| polarC18 | 780.55 | 296.5449 | 17.3 | 0.13 |
| polarC18 | 780.55 | 162.5146 | 17.3 | 0.13 |
| polarC18 | 780.55 | 123.7053 | 17.3 | 0.13 |
| polarC18 | 780.55 | 101.3415 | 17.3 | 0.13 |
| polarC18 | 780.55 | 487.1446 | 17.3 | 0.13 |
| polarC18 | 780.55 | 390.1884 | 17.3 | 0.13 |
| polarC18 | 780.55 | 580.3082 | 17.3 | 0.13 |
| polarC18 | 780.55 | 229.1694 | 17.3 | 0.13 |
| polarC18 | 780.55 | 104.951 | 17.3 | 0.13 |
| polarC18 | 780.55 | 337.3005 | 17.3 | 0.13 |
| polarC18 | 780.55 | 179.692 | 17.3 | 0.13 |
| polarC18 | 780.55 | 121.9435 | 17.3 | 0.13 |
| polarC18 | 780.55 | 114.5647 | 17.3 | 0.13 |
| polarC18 | 780.55 | 253.1441 | 17.3 | 0.13 |
| polarC18 | 780.55 | 111.4399 | 17.3 | 0.13 |
| polarC18 | 780.55 | 104.3333 | 17.3 | 0.13 |
| polarC18 | 780.55 | 437.2457 | 17.3 | 0.13 |
| polarC18 | 780.55 | 248.3099 | 17.3 | 0.13 |
| polarC18 | 780.55 | 170.3429 | 17.3 | 0.13 |
| polarC18 | 780.55 | 733.3806 | 17.2 | 0.13 |
| polarC18 | 780.55 | 214.3675 | 17.2 | 0.13 |
| polarC18 | 780.55 | 95.1958 | 17.2 | 0.13 |
| polarC18 | 780.55 | 186.8071 | 17.2 | 0.13 |
| polarC18 | 780.55 | 90.7243 | 17.2 | 0.13 |
| polarC18 | 780.55 | 84.5361 | 17.2 | 0.13 |
| polarC18 | 780.55 | 272.4083 | 17.2 | 0.13 |
| polarC18 | 780.55 | 145.9857 | 17.2 | 0.13 |
| polarC18 | 780.55 | 388.9057 | 17.2 | 0.13 |
| polarC18 | 780.55 | 157.2417 | 17.2 | 0.13 |
| polarC18 | 780.55 | 121.6424 | 17.2 | 0.13 |
| polarC18 | 780.55 | 769.2133 | 17.2 | 0.13 |
| polarC18 | 780.55 | 138.5694 | 17.2 | 0.13 |
| polarC18 | 780.55 | 321.8846 | 17.2 | 0.13 |
| polarC18 | 780.55 | 212.5821 | 17.2 | 0.13 |
| polarC18 | 780.55 | 171.8925 | 17.2 | 0.13 |
| polarC18 | 780.55 | 89.039 | 17.2 | 0.13 |
| polarC18 | 780.55 | 155.6737 | 17.2 | 0.13 |

| | | | | |
|---|---|---|---|---|
| polarC18 | 780.55 | 351.6713 | 17.2 | 0.13 |
| polarC18 | 780.55 | 124.5407 | 17.2 | 0.13 |
| polarC18 | 780.55 | 282.0308 | 17.2 | 0.13 |
| polarC18 | 780.55 | 193.1971 | 17.2 | 0.13 |
| polarC18 | 780.55 | 224.2342 | 17.2 | 0.13 |
| polarC18 | 780.55 | 139.9494 | 17.2 | 0.13 |
| polarC18 | 780.55 | 92.5028 | 17.2 | 0.13 |
| polarC18 | 780.55 | 317.3978 | 17.2 | 0.13 |
| polarC18 | 780.55 | 648.8979 | 17.2 | 0.13 |
| polarC18 | 780.55 | 318.4305 | 17.2 | 0.13 |
| polarC18 | 780.55 | 190.1805 | 17.2 | 0.13 |
| polarC18 | 780.55 | 197.8772 | 17.2 | 0.13 |
| polarC18 | 780.55 | 695.3229 | 17.2 | 0.13 |
| polarC18 | 780.55 | 701.0844 | 17.2 | 0.13 |
| polarC18 | 780.55 | 204.9943 | 17.2 | 0.13 |
| polarC18 | 780.55 | 544.1898 | 17.2 | 0.13 |
| polarC18 | 780.55 | 92.9032 | 17.2 | 0.13 |
| polarC18 | 780.55 | 121.0116 | 17.2 | 0.13 |
| polarC18 | 780.55 | 101.5485 | 17.2 | 0.13 |
| polarC18 | 780.55 | 622.7837 | 17.2 | 0.13 |
| polarC18 | 780.55 | 204.7287 | 17.2 | 0.13 |
| polarC18 | 780.55 | 192.8055 | 17.2 | 0.13 |
| polarC18 | 780.55 | 484.3425 | 17.2 | 0.13 |
| polarC18 | 780.55 | 461.1765 | 17.2 | 0.13 |
| polarC18 | 780.55 | 106.9533 | 17.2 | 0.13 |
| polarC18 | 780.55 | 122.5526 | 17.2 | 0.13 |
| polarC18 | 780.55 | 190.1464 | 17.2 | 0.13 |
| polarC18 | 780.55 | 638.6996 | 17.2 | 0.13 |
| polarC18 | 780.55 | 89.7467 | 17.2 | 0.13 |
| polarC18 | 780.55 | 446.9643 | 17.2 | 0.13 |
| polarC18 | 780.55 | 315.9695 | 17.2 | 0.13 |
| polarC18 | 780.55 | 83.677 | 17.2 | 0.13 |
| polarC18 | 780.55 | 273.1462 | 17.1 | 0.13 |
| polarC18 | 780.55 | 160.4901 | 17.1 | 0.13 |
| polarC18 | 780.55 | 565.9594 | 17.1 | 0.13 |
| polarC18 | 780.55 | 161.8685 | 17.1 | 0.13 |
| polarC18 | 780.55 | 771.6782 | 17.1 | 0.13 |
| polarC18 | 780.55 | 204.4199 | 17.1 | 0.13 |
| polarC18 | 780.55 | 560.9982 | 17.1 | 0.13 |
| polarC18 | 780.55 | 99.2281 | 17.1 | 0.13 |
| polarC18 | 780.55 | 141.6349 | 17.1 | 0.13 |
| polarC18 | 780.55 | 617.3318 | 17.1 | 0.13 |

| | | | | |
|---|---|---|---|---|
| polarC18 | 780.55 | 152.8528 | 17.1 | 0.13 |
| polarC18 | 780.55 | 250.9637 | 17.1 | 0.13 |
| polarC18 | 780.55 | 427.0661 | 17.1 | 0.13 |
| polarC18 | 780.55 | 283.3539 | 17.1 | 0.13 |
| polarC18 | 780.55 | 710.7561 | 17.1 | 0.13 |
| polarC18 | 780.55 | 684.9345 | 17.1 | 0.13 |
| polarC18 | 780.55 | 302.0724 | 17.1 | 0.13 |
| polarC18 | 780.55 | 194.5652 | 17.1 | 0.13 |
| polarC18 | 780.55 | 203.6514 | 17.1 | 0.13 |
| polarC18 | 780.55 | 204.5298 | 17.1 | 0.13 |
| polarC18 | 780.55 | 154.2731 | 17.1 | 0.13 |
| polarC18 | 780.55 | 121.3242 | 17.1 | 0.13 |
| polarC18 | 780.55 | 109.6693 | 17.1 | 0.13 |
| polarC18 | 780.55 | 137.6364 | 17.1 | 0.13 |
| polarC18 | 780.55 | 572.4344 | 17.1 | 0.13 |
| polarC18 | 780.55 | 82.164 | 17.1 | 0.13 |
| polarC18 | 780.55 | 164.3358 | 17.1 | 0.13 |
| polarC18 | 780.55 | 111.4085 | 17.1 | 0.13 |
| polarC18 | 780.55 | 85.9744 | 17.1 | 0.13 |
| polarC18 | 780.55 | 291.674 | 17.1 | 0.13 |
| polarC18 | 780.55 | 683.5405 | 17.1 | 0.13 |
| polarC18 | 780.55 | 138.6979 | 17.1 | 0.13 |
| polarC18 | 780.55 | 96.5616 | 17.1 | 0.13 |
| polarC18 | 780.55 | 382.2125 | 17.1 | 0.13 |
| polarC18 | 780.55 | 106.4706 | 17.1 | 0.13 |
| polarC18 | 780.55 | 468.3268 | 17.1 | 0.13 |
| polarC18 | 780.55 | 139.0706 | 17.1 | 0.13 |
| polarC18 | 780.55 | 95.1061 | 17.1 | 0.13 |
| polarC18 | 780.55 | 248.9334 | 17.1 | 0.13 |
| polarC18 | 780.55 | 390.0515 | 17.1 | 0.13 |
| polarC18 | 780.55 | 93.3809 | 17.1 | 0.13 |
| polarC18 | 780.55 | 379.5617 | 17.1 | 0.13 |
| polarC18 | 780.55 | 162.6251 | 17.1 | 0.13 |
| polarC18 | 780.55 | 123.3246 | 17.1 | 0.13 |
| polarC18 | 780.55 | 417.9915 | 17.1 | 0.13 |
| polarC18 | 780.55 | 180.7378 | 17.1 | 0.13 |
| polarC18 | 780.55 | 718.6872 | 17.1 | 0.13 |
| polarC18 | 780.55 | 249.0605 | 17.1 | 0.13 |
| polarC18 | 780.55 | 777.63 | 17.1 | 0.12 |
| polarC18 | 780.55 | 603.4351 | 17.1 | 0.12 |
| polarC18 | 780.55 | 308.8347 | 17 | 0.12 |
| polarC18 | 780.55 | 532.1587 | 17 | 0.12 |

| | | | | |
|---|---|---|---|---|
| polarC18 | 780.55 | 135.6684 | 17 | 0.12 |
| polarC18 | 780.55 | 111.5002 | 17 | 0.12 |
| polarC18 | 780.55 | 592.843 | 17 | 0.12 |
| polarC18 | 780.55 | 156.9685 | 17 | 0.12 |
| polarC18 | 780.55 | 176.0328 | 17 | 0.12 |
| polarC18 | 780.55 | 142.9066 | 17 | 0.12 |
| polarC18 | 780.55 | 228.1433 | 17 | 0.12 |
| polarC18 | 780.55 | 342.8311 | 17 | 0.12 |
| polarC18 | 780.55 | 538.4968 | 17 | 0.12 |
| polarC18 | 780.55 | 468.8134 | 17 | 0.12 |
| polarC18 | 780.55 | 98.6073 | 17 | 0.12 |
| polarC18 | 780.55 | 674.2651 | 17 | 0.12 |
| polarC18 | 780.55 | 232.679 | 17 | 0.12 |
| polarC18 | 780.55 | 328.3561 | 17 | 0.12 |
| polarC18 | 780.55 | 651.3393 | 17 | 0.12 |
| polarC18 | 780.55 | 397.9198 | 17 | 0.12 |
| polarC18 | 780.55 | 465.6961 | 17 | 0.12 |
| polarC18 | 780.55 | 121.0712 | 17 | 0.12 |
| polarC18 | 780.55 | 447.9788 | 17 | 0.12 |
| polarC18 | 780.55 | 248.2792 | 17 | 0.12 |
| polarC18 | 780.55 | 114.894 | 17 | 0.12 |
| polarC18 | 780.55 | 203.85 | 17 | 0.12 |
| polarC18 | 780.55 | 312.5962 | 17 | 0.12 |
| polarC18 | 780.55 | 220.1611 | 17 | 0.12 |
| polarC18 | 780.55 | 584.713 | 17 | 0.12 |
| polarC18 | 780.55 | 393.7639 | 17 | 0.12 |
| polarC18 | 780.55 | 322.7169 | 17 | 0.12 |
| polarC18 | 780.55 | 131.073 | 17 | 0.12 |
| polarC18 | 780.55 | 476.3481 | 17 | 0.12 |
| polarC18 | 780.55 | 198.4809 | 17 | 0.12 |
| polarC18 | 780.55 | 552.1233 | 17 | 0.12 |
| polarC18 | 780.55 | 483.0579 | 17 | 0.12 |
| polarC18 | 780.55 | 196.0245 | 17 | 0.12 |
| polarC18 | 780.55 | 339.2917 | 17 | 0.12 |
| polarC18 | 780.55 | 349.8199 | 17 | 0.12 |
| polarC18 | 780.55 | 254.9892 | 17 | 0.12 |
| polarC18 | 780.55 | 499.3398 | 17 | 0.12 |
| polarC18 | 780.55 | 130.1845 | 17 | 0.12 |
| polarC18 | 780.55 | 271.7461 | 17 | 0.12 |
| polarC18 | 780.55 | 103.4336 | 17 | 0.12 |
| polarC18 | 780.55 | 145.9595 | 17 | 0.12 |
| polarC18 | 780.55 | 210.9827 | 17 | 0.12 |

| | | | | |
|---|---|---|---|---|
| polarC18 | 780.55 | 483.4349 | 17 | 0.12 |
| polarC18 | 780.55 | 372.1309 | 17 | 0.12 |
| polarC18 | 780.55 | 708.1568 | 17 | 0.12 |
| polarC18 | 780.55 | 84.8386 | 17 | 0.12 |
| polarC18 | 780.55 | 366.0654 | 17 | 0.12 |
| polarC18 | 780.55 | 93.6054 | 17 | 0.12 |
| polarC18 | 780.55 | 251.8144 | 17 | 0.12 |
| polarC18 | 780.55 | 137.3704 | 16.9 | 0.12 |
| polarC18 | 780.55 | 172.4209 | 16.9 | 0.12 |
| polarC18 | 780.55 | 177.8415 | 16.9 | 0.12 |
| polarC18 | 780.55 | 91.3653 | 16.9 | 0.12 |
| polarC18 | 780.55 | 110.8193 | 16.9 | 0.12 |
| polarC18 | 780.55 | 113.2174 | 16.9 | 0.12 |
| polarC18 | 780.55 | 352.2499 | 16.9 | 0.12 |
| polarC18 | 780.55 | 178.4822 | 16.9 | 0.12 |
| polarC18 | 780.55 | 215.0346 | 16.9 | 0.12 |
| polarC18 | 780.55 | 120.6441 | 16.9 | 0.12 |
| polarC18 | 780.55 | 328.4229 | 16.9 | 0.12 |
| polarC18 | 780.55 | 231.8884 | 16.9 | 0.12 |
| polarC18 | 780.55 | 274.1894 | 16.9 | 0.12 |
| polarC18 | 780.55 | 87.9584 | 16.9 | 0.12 |
| polarC18 | 780.55 | 398.2274 | 16.9 | 0.12 |
| polarC18 | 780.55 | 308.085 | 16.9 | 0.12 |
| polarC18 | 780.55 | 120.8304 | 16.9 | 0.12 |
| polarC18 | 780.55 | 726.0642 | 16.9 | 0.12 |
| polarC18 | 780.55 | 142.5529 | 16.9 | 0.12 |
| polarC18 | 780.55 | 293.6695 | 16.9 | 0.12 |
| polarC18 | 780.55 | 299.981 | 16.9 | 0.12 |
| polarC18 | 780.55 | 104.0617 | 16.9 | 0.12 |
| polarC18 | 780.55 | 165.6469 | 16.9 | 0.12 |
| polarC18 | 780.55 | 150.1246 | 16.9 | 0.12 |
| polarC18 | 780.55 | 116.0114 | 16.9 | 0.12 |
| polarC18 | 780.55 | 616.9029 | 16.9 | 0.12 |
| polarC18 | 780.55 | 137.1546 | 16.9 | 0.12 |
| polarC18 | 780.55 | 202.6933 | 16.9 | 0.12 |
| polarC18 | 780.55 | 396.1434 | 16.9 | 0.12 |
| polarC18 | 780.55 | 150.6273 | 16.9 | 0.12 |
| polarC18 | 780.55 | 130.5114 | 16.9 | 0.12 |
| polarC18 | 780.55 | 184.3518 | 16.9 | 0.12 |
| polarC18 | 780.55 | 152.3671 | 16.9 | 0.12 |
| polarC18 | 780.55 | 482.4564 | 16.9 | 0.12 |
| polarC18 | 780.55 | 142.0574 | 16.9 | 0.12 |

| | | | | |
|---|---|---|---|---|
| polarC18 | 780.55 | 212.4278 | 16.9 | 0.12 |
| polarC18 | 780.55 | 795.2567 | 16.9 | 0.12 |
| polarC18 | 780.55 | 479.4962 | 16.9 | 0.12 |
| polarC18 | 780.55 | 345.648 | 16.9 | 0.12 |
| polarC18 | 780.55 | 195.109 | 16.9 | 0.12 |
| polarC18 | 780.55 | 113.0802 | 16.9 | 0.12 |
| polarC18 | 780.55 | 364.8387 | 16.9 | 0.12 |
| polarC18 | 780.55 | 90.8458 | 16.9 | 0.12 |
| polarC18 | 780.55 | 170.7092 | 16.9 | 0.12 |
| polarC18 | 780.55 | 483.6082 | 16.9 | 0.12 |
| polarC18 | 780.55 | 624.3159 | 16.9 | 0.12 |
| polarC18 | 780.55 | 88.2179 | 16.9 | 0.12 |
| polarC18 | 780.55 | 217.4671 | 16.9 | 0.12 |
| polarC18 | 780.55 | 242.2621 | 16.9 | 0.12 |
| polarC18 | 780.55 | 175.2695 | 16.9 | 0.12 |
| polarC18 | 780.55 | 109.3294 | 16.8 | 0.12 |
| polarC18 | 780.55 | 116.8538 | 16.8 | 0.12 |
| polarC18 | 780.55 | 572.0141 | 16.8 | 0.12 |
| polarC18 | 780.55 | 193.0607 | 16.8 | 0.12 |
| polarC18 | 780.55 | 259.0503 | 16.8 | 0.12 |
| polarC18 | 780.55 | 93.5162 | 16.8 | 0.12 |
| polarC18 | 780.55 | 131.622 | 16.8 | 0.12 |
| polarC18 | 780.55 | 384.4317 | 16.8 | 0.12 |
| polarC18 | 780.55 | 219.3736 | 16.8 | 0.12 |
| polarC18 | 780.55 | 92.7801 | 16.8 | 0.12 |
| polarC18 | 780.55 | 123.2893 | 16.8 | 0.12 |
| polarC18 | 780.55 | 414.478 | 16.8 | 0.12 |
| polarC18 | 780.55 | 619.1499 | 16.8 | 0.12 |
| polarC18 | 780.55 | 235.5772 | 16.8 | 0.12 |
| polarC18 | 780.55 | 669.7213 | 16.8 | 0.12 |
| polarC18 | 780.55 | 389.9565 | 16.8 | 0.12 |
| polarC18 | 780.55 | 223.3828 | 16.8 | 0.12 |
| polarC18 | 780.55 | 544.4567 | 16.8 | 0.12 |
| polarC18 | 780.55 | 96.0137 | 16.8 | 0.12 |
| polarC18 | 780.55 | 89.9607 | 16.8 | 0.12 |
| polarC18 | 780.55 | 169.6427 | 16.8 | 0.12 |
| polarC18 | 780.55 | 365.9937 | 16.8 | 0.12 |
| polarC18 | 780.55 | 93.1407 | 16.8 | 0.12 |
| polarC18 | 780.55 | 654.7271 | 16.8 | 0.12 |
| polarC18 | 780.55 | 157.4005 | 16.8 | 0.12 |
| polarC18 | 780.55 | 83.3235 | 16.8 | 0.12 |
| polarC18 | 780.55 | 93.9942 | 16.8 | 0.12 |

| | | | | |
|---|---|---|---|---|
| polarC18 | 780.55 | 296.3089 | 16.8 | 0.12 |
| polarC18 | 780.55 | 97.324 | 16.8 | 0.12 |
| polarC18 | 780.55 | 510.4326 | 16.8 | 0.12 |
| polarC18 | 780.55 | 219.5424 | 16.8 | 0.12 |
| polarC18 | 780.55 | 101.7125 | 16.8 | 0.12 |
| polarC18 | 780.55 | 114.0455 | 16.8 | 0.12 |
| polarC18 | 780.55 | 745.7354 | 16.8 | 0.12 |
| polarC18 | 780.55 | 121.2318 | 16.8 | 0.12 |
| polarC18 | 780.55 | 88.1073 | 16.8 | 0.12 |
| polarC18 | 780.55 | 102.954 | 16.8 | 0.12 |
| polarC18 | 780.55 | 389.7116 | 16.8 | 0.12 |
| polarC18 | 780.55 | 95.2037 | 16.8 | 0.12 |
| polarC18 | 780.55 | 239.2171 | 16.8 | 0.12 |
| polarC18 | 780.55 | 127.5247 | 16.8 | 0.12 |
| polarC18 | 780.55 | 121.8884 | 16.8 | 0.12 |
| polarC18 | 780.55 | 87.7355 | 16.8 | 0.12 |
| polarC18 | 780.55 | 220.1004 | 16.8 | 0.12 |
| polarC18 | 780.55 | 687.158 | 16.8 | 0.12 |
| polarC18 | 780.55 | 122.869 | 16.8 | 0.12 |
| polarC18 | 780.55 | 138.0361 | 16.8 | 0.12 |
| polarC18 | 780.55 | 89.6439 | 16.8 | 0.12 |
| polarC18 | 780.55 | 164.8374 | 16.8 | 0.12 |
| polarC18 | 780.55 | 307.7692 | 16.8 | 0.12 |
| polarC18 | 780.55 | 420.631 | 16.8 | 0.12 |
| polarC18 | 780.55 | 155.0242 | 16.8 | 0.12 |
| polarC18 | 780.55 | 189.0085 | 16.8 | 0.12 |
| polarC18 | 780.55 | 355.7793 | 16.8 | 0.12 |
| polarC18 | 780.55 | 498.4017 | 16.8 | 0.12 |
| polarC18 | 780.55 | 83.2413 | 16.8 | 0.12 |
| polarC18 | 780.55 | 675.2502 | 16.8 | 0.12 |
| polarC18 | 780.55 | 217.5187 | 16.8 | 0.12 |
| polarC18 | 780.55 | 94.6175 | 16.7 | 0.12 |
| polarC18 | 780.55 | 297.8454 | 16.7 | 0.12 |
| polarC18 | 780.55 | 202.6063 | 16.7 | 0.12 |
| polarC18 | 780.55 | 96.937 | 16.7 | 0.12 |
| polarC18 | 780.55 | 115.4468 | 16.7 | 0.12 |
| polarC18 | 780.55 | 194.6566 | 16.7 | 0.12 |
| polarC18 | 780.55 | 470.1302 | 16.7 | 0.12 |
| polarC18 | 780.55 | 91.6593 | 16.7 | 0.12 |
| polarC18 | 780.55 | 441.5258 | 16.7 | 0.12 |
| polarC18 | 780.55 | 103.5472 | 16.7 | 0.12 |
| polarC18 | 780.55 | 251.2099 | 16.7 | 0.12 |

| | | | | |
|---|---|---|---|---|
| polarC18 | 780.55 | 179.631 | 16.7 | 0.12 |
| polarC18 | 780.55 | 162.4856 | 16.7 | 0.12 |
| polarC18 | 780.55 | 754.6345 | 16.7 | 0.12 |
| polarC18 | 780.55 | 242.2062 | 16.7 | 0.12 |
| polarC18 | 780.55 | 278.7972 | 16.7 | 0.12 |
| polarC18 | 780.55 | 255.1216 | 16.7 | 0.12 |
| polarC18 | 780.55 | 110.7323 | 16.7 | 0.12 |
| polarC18 | 780.55 | 133.6118 | 16.7 | 0.12 |
| polarC18 | 780.55 | 131.7962 | 16.7 | 0.12 |
| polarC18 | 780.55 | 147.6183 | 16.7 | 0.12 |
| polarC18 | 780.55 | 158.7175 | 16.7 | 0.12 |
| polarC18 | 780.55 | 394.7641 | 16.7 | 0.12 |
| polarC18 | 780.55 | 188.8788 | 16.7 | 0.12 |
| polarC18 | 780.55 | 200.4797 | 16.7 | 0.12 |
| polarC18 | 780.55 | 222.896 | 16.7 | 0.12 |
| polarC18 | 780.55 | 250.6337 | 16.7 | 0.12 |
| polarC18 | 780.55 | 95.0256 | 16.7 | 0.12 |
| polarC18 | 780.55 | 144.5542 | 16.7 | 0.12 |
| polarC18 | 780.55 | 234.227 | 16.7 | 0.12 |
| polarC18 | 780.55 | 188.5433 | 16.7 | 0.12 |
| polarC18 | 780.55 | 110.1906 | 16.7 | 0.12 |
| polarC18 | 780.55 | 129.744 | 16.7 | 0.12 |
| polarC18 | 780.55 | 131.9164 | 16.7 | 0.12 |
| polarC18 | 780.55 | 152.4038 | 16.7 | 0.12 |
| polarC18 | 780.55 | 663.38 | 16.7 | 0.12 |
| polarC18 | 780.55 | 135.2116 | 16.7 | 0.12 |
| polarC18 | 780.55 | 285.17 | 16.7 | 0.12 |
| polarC18 | 780.55 | 114.1327 | 16.7 | 0.12 |
| polarC18 | 780.55 | 206.1462 | 16.7 | 0.12 |
| polarC18 | 780.55 | 218.8594 | 16.7 | 0.12 |
| polarC18 | 780.55 | 156.6011 | 16.7 | 0.12 |
| polarC18 | 780.55 | 555.2624 | 16.7 | 0.12 |
| polarC18 | 780.55 | 414.2435 | 16.7 | 0.12 |
| polarC18 | 780.55 | 142.0266 | 16.7 | 0.12 |
| polarC18 | 780.55 | 284.869 | 16.7 | 0.12 |
| polarC18 | 780.55 | 338.2939 | 16.7 | 0.12 |
| polarC18 | 780.55 | 204.844 | 16.7 | 0.12 |
| polarC18 | 780.55 | 333.9521 | 16.7 | 0.12 |
| polarC18 | 780.55 | 265.366 | 16.7 | 0.12 |
| polarC18 | 780.55 | 473.5399 | 16.7 | 0.12 |
| polarC18 | 780.55 | 88.8471 | 16.7 | 0.12 |
| polarC18 | 780.55 | 113.2233 | 16.7 | 0.12 |

| | | | | |
|---|---|---|---|---|
| polarC18 | 780.55 | 94.2757 | 16.7 | 0.12 |
| polarC18 | 780.55 | 702.5716 | 16.7 | 0.12 |
| polarC18 | 780.55 | 243.2104 | 16.7 | 0.12 |
| polarC18 | 780.55 | 181.2762 | 16.6 | 0.12 |
| polarC18 | 780.55 | 85.341 | 16.6 | 0.12 |
| polarC18 | 780.55 | 86.0852 | 16.6 | 0.12 |
| polarC18 | 780.55 | 356.4177 | 16.6 | 0.12 |
| polarC18 | 780.55 | 268.2108 | 16.6 | 0.12 |
| polarC18 | 780.55 | 133.9925 | 16.6 | 0.12 |
| polarC18 | 780.55 | 245.0941 | 16.6 | 0.12 |
| polarC18 | 780.55 | 123.1882 | 16.6 | 0.12 |
| polarC18 | 780.55 | 90.038 | 16.6 | 0.12 |
| polarC18 | 780.55 | 97.5096 | 16.6 | 0.12 |
| polarC18 | 780.55 | 162.0687 | 16.6 | 0.12 |
| polarC18 | 780.55 | 97.7788 | 16.6 | 0.12 |
| polarC18 | 780.55 | 149.8499 | 16.6 | 0.12 |
| polarC18 | 780.55 | 154.3304 | 16.6 | 0.12 |
| polarC18 | 780.55 | 86.1707 | 16.6 | 0.12 |
| polarC18 | 780.55 | 107.1195 | 16.6 | 0.12 |
| polarC18 | 780.55 | 152.2385 | 16.6 | 0.12 |
| polarC18 | 780.55 | 752.1078 | 16.6 | 0.12 |
| polarC18 | 780.55 | 803.9379 | 16.6 | 0.12 |
| polarC18 | 780.55 | 300.5188 | 16.6 | 0.12 |
| polarC18 | 780.55 | 196.7749 | 16.6 | 0.12 |
| polarC18 | 780.55 | 527.837 | 16.6 | 0.12 |
| polarC18 | 780.55 | 174.5116 | 16.6 | 0.12 |
| polarC18 | 780.55 | 126.8171 | 16.6 | 0.12 |
| polarC18 | 780.55 | 134.2802 | 16.6 | 0.12 |
| polarC18 | 780.55 | 102.8233 | 16.6 | 0.12 |
| polarC18 | 780.55 | 226.6968 | 16.6 | 0.12 |
| polarC18 | 780.55 | 321.3257 | 16.6 | 0.12 |
| polarC18 | 780.55 | 114.1984 | 16.6 | 0.12 |
| polarC18 | 780.55 | 406.4689 | 16.6 | 0.12 |
| polarC18 | 780.55 | 161.8277 | 16.6 | 0.12 |
| polarC18 | 780.55 | 206.6402 | 16.6 | 0.12 |
| polarC18 | 780.55 | 183.755 | 16.6 | 0.12 |
| polarC18 | 780.55 | 400.9619 | 16.6 | 0.12 |
| polarC18 | 780.55 | 546.273 | 16.6 | 0.12 |
| polarC18 | 780.55 | 82.3887 | 16.6 | 0.12 |
| polarC18 | 780.55 | 116.5124 | 16.6 | 0.12 |
| polarC18 | 780.55 | 135.4163 | 16.6 | 0.12 |
| polarC18 | 780.55 | 138.6354 | 16.6 | 0.12 |

| | | | | |
|---|---|---|---|---|
| polarC18 | 780.55 | 135.2294 | 16.6 | 0.12 |
| polarC18 | 780.55 | 526.8076 | 16.6 | 0.12 |
| polarC18 | 780.55 | 84.473 | 16.6 | 0.12 |
| polarC18 | 780.55 | 194.0621 | 16.6 | 0.12 |
| polarC18 | 780.55 | 192.4234 | 16.6 | 0.12 |
| polarC18 | 780.55 | 127.7084 | 16.6 | 0.12 |
| polarC18 | 780.55 | 176.5872 | 16.6 | 0.12 |
| polarC18 | 780.55 | 415.4796 | 16.6 | 0.12 |
| polarC18 | 780.55 | 252.0496 | 16.6 | 0.12 |
| polarC18 | 780.55 | 338.1674 | 16.6 | 0.12 |
| polarC18 | 780.55 | 422.4223 | 16.6 | 0.12 |
| polarC18 | 780.55 | 151.8316 | 16.6 | 0.12 |
| polarC18 | 780.55 | 416.9059 | 16.6 | 0.12 |
| polarC18 | 780.55 | 250.2947 | 16.6 | 0.12 |
| polarC18 | 780.55 | 652.4275 | 16.6 | 0.12 |
| polarC18 | 780.55 | 270.2697 | 16.6 | 0.12 |
| polarC18 | 780.55 | 174.7064 | 16.6 | 0.12 |
| polarC18 | 780.55 | 124.2956 | 16.6 | 0.12 |
| polarC18 | 780.55 | 99.7011 | 16.6 | 0.12 |
| polarC18 | 780.55 | 113.2518 | 16.6 | 0.12 |
| polarC18 | 780.55 | 163.8293 | 16.6 | 0.12 |
| polarC18 | 780.55 | 437.0151 | 16.6 | 0.12 |
| polarC18 | 780.55 | 164.9148 | 16.6 | 0.12 |
| polarC18 | 780.55 | 163.353 | 16.6 | 0.12 |
| polarC18 | 780.55 | 92.7516 | 16.6 | 0.12 |
| polarC18 | 780.55 | 160.3046 | 16.6 | 0.12 |
| polarC18 | 780.55 | 183.8998 | 16.6 | 0.12 |
| polarC18 | 780.55 | 259.1813 | 16.6 | 0.12 |
| polarC18 | 780.55 | 214.155 | 16.6 | 0.12 |
| polarC18 | 780.55 | 154.7862 | 16.5 | 0.12 |
| polarC18 | 780.55 | 366.6154 | 16.5 | 0.12 |
| polarC18 | 780.55 | 809.4171 | 16.5 | 0.12 |
| polarC18 | 780.55 | 190.3232 | 16.5 | 0.12 |
| polarC18 | 780.55 | 151.304 | 16.5 | 0.12 |
| polarC18 | 780.55 | 163.0534 | 16.5 | 0.12 |
| polarC18 | 780.55 | 215.5404 | 16.5 | 0.12 |
| polarC18 | 780.55 | 292.492 | 16.5 | 0.12 |
| polarC18 | 780.55 | 103.0622 | 16.5 | 0.12 |
| polarC18 | 780.55 | 161.4292 | 16.5 | 0.12 |
| polarC18 | 780.55 | 424.2102 | 16.5 | 0.12 |
| polarC18 | 780.55 | 472.4921 | 16.5 | 0.12 |
| polarC18 | 780.55 | 122.2688 | 16.5 | 0.12 |

| | | | | |
|---|---|---|---|---|
| polarC18 | 780.55 | 238.7973 | 16.5 | 0.12 |
| polarC18 | 780.55 | 280.4143 | 16.5 | 0.12 |
| polarC18 | 780.55 | 210.7363 | 16.5 | 0.12 |
| polarC18 | 780.55 | 99.9088 | 16.5 | 0.12 |
| polarC18 | 780.55 | 564.676 | 16.5 | 0.12 |
| polarC18 | 780.55 | 356.2725 | 16.5 | 0.12 |
| polarC18 | 780.55 | 130.8552 | 16.5 | 0.12 |
| polarC18 | 780.55 | 375.7547 | 16.5 | 0.12 |
| polarC18 | 780.55 | 333.4622 | 16.5 | 0.12 |
| polarC18 | 780.55 | 107.8012 | 16.5 | 0.12 |
| polarC18 | 780.55 | 620.5907 | 16.5 | 0.12 |
| polarC18 | 780.55 | 438.8279 | 16.5 | 0.12 |
| polarC18 | 780.55 | 273.0039 | 16.5 | 0.12 |
| polarC18 | 780.55 | 83.8971 | 16.5 | 0.12 |
| polarC18 | 780.55 | 559.4733 | 16.5 | 0.12 |
| polarC18 | 780.55 | 106.1802 | 16.5 | 0.12 |
| polarC18 | 780.55 | 89.9482 | 16.5 | 0.12 |
| polarC18 | 780.55 | 703.8722 | 16.5 | 0.12 |
| polarC18 | 780.55 | 334.7788 | 16.5 | 0.12 |
| polarC18 | 780.55 | 177.6238 | 16.5 | 0.12 |
| polarC18 | 780.55 | 105.8791 | 16.5 | 0.12 |
| polarC18 | 780.55 | 263.0205 | 16.5 | 0.12 |
| polarC18 | 780.55 | 422.7402 | 16.5 | 0.12 |
| polarC18 | 780.55 | 100.6754 | 16.5 | 0.12 |
| polarC18 | 780.55 | 302.8935 | 16.5 | 0.12 |
| polarC18 | 780.55 | 97.7986 | 16.5 | 0.12 |
| polarC18 | 780.55 | 101.2994 | 16.5 | 0.12 |
| polarC18 | 780.55 | 268.7076 | 16.5 | 0.12 |
| polarC18 | 780.55 | 393.2823 | 16.5 | 0.12 |
| polarC18 | 780.55 | 469.5589 | 16.5 | 0.12 |
| polarC18 | 780.55 | 117.0354 | 16.5 | 0.12 |
| polarC18 | 780.55 | 152.3465 | 16.5 | 0.12 |
| polarC18 | 780.55 | 122.7146 | 16.5 | 0.12 |
| polarC18 | 780.55 | 103.2204 | 16.5 | 0.12 |
| polarC18 | 780.55 | 128.0118 | 16.5 | 0.12 |
| polarC18 | 780.55 | 141.3873 | 16.5 | 0.12 |
| polarC18 | 780.55 | 106.3371 | 16.5 | 0.12 |
| polarC18 | 780.55 | 187.4411 | 16.5 | 0.12 |
| polarC18 | 780.55 | 85.1925 | 16.5 | 0.12 |
| polarC18 | 780.55 | 307.503 | 16.5 | 0.12 |
| polarC18 | 780.55 | 802.9017 | 16.5 | 0.12 |
| polarC18 | 780.55 | 166.3744 | 16.5 | 0.12 |

| | | | | |
|---|---|---|---|---|
| polarC18 | 780.55 | 361.3945 | 16.5 | 0.12 |
| polarC18 | 780.55 | 106.3145 | 16.5 | 0.12 |
| polarC18 | 780.55 | 140.5545 | 16.5 | 0.12 |
| polarC18 | 780.55 | 87.036 | 16.5 | 0.12 |
| polarC18 | 780.55 | 339.8237 | 16.5 | 0.12 |
| polarC18 | 780.55 | 270.7755 | 16.5 | 0.12 |
| polarC18 | 780.55 | 209.3997 | 16.5 | 0.12 |
| polarC18 | 780.55 | 332.4911 | 16.5 | 0.12 |
| polarC18 | 780.55 | 105.4789 | 16.5 | 0.12 |
| polarC18 | 780.55 | 222.484 | 16.5 | 0.12 |
| polarC18 | 780.55 | 114.3545 | 16.5 | 0.12 |
| polarC18 | 780.55 | 173.3175 | 16.5 | 0.12 |
| polarC18 | 780.55 | 300.2957 | 16.5 | 0.12 |
| polarC18 | 780.55 | 110.844 | 16.4 | 0.12 |
| polarC18 | 780.55 | 398.0061 | 16.4 | 0.12 |
| polarC18 | 780.55 | 101.966 | 16.4 | 0.12 |
| polarC18 | 780.55 | 249.5763 | 16.4 | 0.12 |
| polarC18 | 780.55 | 580.6622 | 16.4 | 0.12 |
| polarC18 | 780.55 | 110.8871 | 16.4 | 0.12 |
| polarC18 | 780.55 | 103.4285 | 16.4 | 0.12 |
| polarC18 | 780.55 | 515.9358 | 16.4 | 0.12 |
| polarC18 | 780.55 | 285.8687 | 16.4 | 0.12 |
| polarC18 | 780.55 | 535.51 | 16.4 | 0.12 |
| polarC18 | 780.55 | 95.7352 | 16.4 | 0.12 |
| polarC18 | 780.55 | 119.8228 | 16.4 | 0.12 |
| polarC18 | 780.55 | 159.754 | 16.4 | 0.12 |
| polarC18 | 780.55 | 85.0572 | 16.4 | 0.12 |
| polarC18 | 780.55 | 203.237 | 16.4 | 0.12 |
| polarC18 | 780.55 | 373.5126 | 16.4 | 0.12 |
| polarC18 | 780.55 | 90.9753 | 16.4 | 0.12 |
| polarC18 | 780.55 | 135.8664 | 16.4 | 0.12 |
| polarC18 | 780.55 | 95.2895 | 16.4 | 0.12 |
| polarC18 | 780.55 | 144.1786 | 16.4 | 0.12 |
| polarC18 | 780.55 | 306.3064 | 16.4 | 0.12 |
| polarC18 | 780.55 | 709.7626 | 16.4 | 0.12 |
| polarC18 | 780.55 | 157.6839 | 16.4 | 0.12 |
| polarC18 | 780.55 | 86.5328 | 16.4 | 0.12 |
| polarC18 | 780.55 | 292.7142 | 16.4 | 0.12 |
| polarC18 | 780.55 | 718.0701 | 16.4 | 0.12 |
| polarC18 | 780.55 | 158.169 | 16.4 | 0.12 |
| polarC18 | 780.55 | 102.0461 | 16.4 | 0.12 |
| polarC18 | 780.55 | 356.1365 | 16.4 | 0.12 |

| | | | | |
|---|---|---|---|---|
| polarC18 | 780.55 | 323.0826 | 16.4 | 0.12 |
| polarC18 | 780.55 | 402.9058 | 16.4 | 0.12 |
| polarC18 | 780.55 | 225.5055 | 16.4 | 0.12 |
| polarC18 | 780.55 | 163.8962 | 16.4 | 0.12 |
| polarC18 | 780.55 | 153.5936 | 16.4 | 0.12 |
| polarC18 | 780.55 | 463.0375 | 16.4 | 0.12 |
| polarC18 | 780.55 | 179.2407 | 16.4 | 0.12 |
| polarC18 | 780.55 | 219.8991 | 16.4 | 0.12 |
| polarC18 | 780.55 | 417.1231 | 16.4 | 0.12 |
| polarC18 | 780.55 | 117.0609 | 16.4 | 0.12 |
| polarC18 | 780.55 | 149.1914 | 16.4 | 0.12 |
| polarC18 | 780.55 | 207.7842 | 16.4 | 0.12 |
| polarC18 | 780.55 | 117.2074 | 16.4 | 0.12 |
| polarC18 | 780.55 | 112.5669 | 16.4 | 0.12 |
| polarC18 | 780.55 | 129.1645 | 16.4 | 0.12 |
| polarC18 | 780.55 | 146.7352 | 16.4 | 0.12 |
| polarC18 | 780.55 | 188.8482 | 16.4 | 0.12 |
| polarC18 | 780.55 | 195.901 | 16.4 | 0.12 |
| polarC18 | 780.55 | 117.5535 | 16.4 | 0.12 |
| polarC18 | 780.55 | 552.3795 | 16.4 | 0.12 |
| polarC18 | 780.55 | 333.7276 | 16.4 | 0.12 |
| polarC18 | 780.55 | 643.2015 | 16.4 | 0.12 |
| polarC18 | 780.55 | 81.8138 | 16.4 | 0.12 |
| polarC18 | 780.55 | 85.694 | 16.4 | 0.12 |
| polarC18 | 780.55 | 547.6887 | 16.4 | 0.12 |
| polarC18 | 780.55 | 785.7327 | 16.4 | 0.12 |
| polarC18 | 780.55 | 222.0478 | 16.4 | 0.12 |
| polarC18 | 780.55 | 95.6562 | 16.4 | 0.12 |
| polarC18 | 780.55 | 85.1204 | 16.4 | 0.12 |
| polarC18 | 780.55 | 146.5002 | 16.4 | 0.12 |
| polarC18 | 780.55 | 360.1869 | 16.4 | 0.12 |
| polarC18 | 780.55 | 165.3046 | 16.4 | 0.12 |
| polarC18 | 780.55 | 174.0982 | 16.4 | 0.12 |
| polarC18 | 780.55 | 260.2964 | 16.4 | 0.12 |
| polarC18 | 780.55 | 212.7348 | 16.3 | 0.12 |
| polarC18 | 780.55 | 220.725 | 16.3 | 0.12 |
| polarC18 | 780.55 | 654.2253 | 16.3 | 0.12 |
| polarC18 | 780.55 | 462.2144 | 16.3 | 0.12 |
| polarC18 | 780.55 | 464.141 | 16.3 | 0.12 |
| polarC18 | 780.55 | 285.3658 | 16.3 | 0.12 |
| polarC18 | 780.55 | 229.0576 | 16.3 | 0.12 |
| polarC18 | 780.55 | 88.7446 | 16.3 | 0.12 |

| | | | | |
|---|---|---|---|---|
| polarC18 | 780.55 | 126.8273 | 16.3 | 0.12 |
| polarC18 | 780.55 | 294.4509 | 16.3 | 0.12 |
| polarC18 | 780.55 | 314.5719 | 16.3 | 0.12 |
| polarC18 | 780.55 | 204.3955 | 16.3 | 0.12 |
| polarC18 | 780.55 | 121.4804 | 16.3 | 0.12 |
| polarC18 | 780.55 | 123.8588 | 16.3 | 0.12 |
| polarC18 | 780.55 | 138.2721 | 16.3 | 0.12 |
| polarC18 | 780.55 | 177.0671 | 16.3 | 0.12 |
| polarC18 | 780.55 | 126.4092 | 16.3 | 0.12 |
| polarC18 | 780.55 | 168.8288 | 16.3 | 0.12 |
| polarC18 | 780.55 | 121.3301 | 16.3 | 0.12 |
| polarC18 | 780.55 | 169.1942 | 16.3 | 0.12 |
| polarC18 | 780.55 | 146.9524 | 16.3 | 0.12 |
| polarC18 | 780.55 | 279.5916 | 16.3 | 0.12 |
| polarC18 | 780.55 | 107.6983 | 16.3 | 0.12 |
| polarC18 | 780.55 | 92.8259 | 16.3 | 0.12 |
| polarC18 | 780.55 | 151.2439 | 16.3 | 0.12 |
| polarC18 | 780.55 | 159.9524 | 16.3 | 0.12 |
| polarC18 | 780.55 | 188.813 | 16.3 | 0.12 |
| polarC18 | 780.55 | 133.7883 | 16.3 | 0.12 |
| polarC18 | 780.55 | 205.3155 | 16.3 | 0.12 |
| polarC18 | 780.55 | 327.0477 | 16.3 | 0.12 |
| polarC18 | 780.55 | 136.4162 | 16.3 | 0.12 |
| polarC18 | 780.55 | 561.7089 | 16.3 | 0.12 |
| polarC18 | 780.55 | 126.1647 | 16.3 | 0.12 |
| polarC18 | 780.55 | 167.777 | 16.3 | 0.12 |
| polarC18 | 780.55 | 224.9214 | 16.3 | 0.12 |
| polarC18 | 780.55 | 95.7195 | 16.3 | 0.12 |
| polarC18 | 780.55 | 171.7557 | 16.3 | 0.12 |
| polarC18 | 780.55 | 112.7202 | 16.3 | 0.12 |
| polarC18 | 780.55 | 302.3818 | 16.3 | 0.12 |
| polarC18 | 780.55 | 180.1231 | 16.3 | 0.12 |
| polarC18 | 780.55 | 195.7173 | 16.3 | 0.12 |
| polarC18 | 780.55 | 191.7399 | 16.3 | 0.12 |
| polarC18 | 780.55 | 154.4016 | 16.3 | 0.12 |
| polarC18 | 780.55 | 164.5787 | 16.3 | 0.12 |
| polarC18 | 780.55 | 106.5522 | 16.3 | 0.12 |
| polarC18 | 780.55 | 744.4654 | 16.3 | 0.12 |
| polarC18 | 780.55 | 266.8099 | 16.3 | 0.12 |
| polarC18 | 780.55 | 741.3556 | 16.3 | 0.12 |
| polarC18 | 780.55 | 304.2579 | 16.3 | 0.12 |
| polarC18 | 780.55 | 90.2312 | 16.3 | 0.12 |

| | | | | |
|---|---|---|---|---|
| polarC18 | 780.55 | 173.3821 | 16.3 | 0.12 |
| polarC18 | 780.55 | 111.1278 | 16.3 | 0.12 |
| polarC18 | 780.55 | 109.0094 | 16.3 | 0.12 |
| polarC18 | 780.55 | 114.8774 | 16.3 | 0.12 |
| polarC18 | 780.55 | 81.9724 | 16.3 | 0.12 |
| polarC18 | 780.55 | 102.4672 | 16.3 | 0.12 |
| polarC18 | 780.55 | 113.1598 | 16.3 | 0.12 |
| polarC18 | 780.55 | 138.1593 | 16.3 | 0.12 |
| polarC18 | 780.55 | 205.1712 | 16.3 | 0.12 |
| polarC18 | 780.55 | 153.5141 | 16.3 | 0.12 |
| polarC18 | 780.55 | 501.1906 | 16.3 | 0.12 |
| polarC18 | 780.55 | 106.5987 | 16.3 | 0.12 |
| polarC18 | 780.55 | 795.5518 | 16.3 | 0.12 |
| polarC18 | 780.55 | 333.6559 | 16.3 | 0.12 |
| polarC18 | 780.55 | 373.1535 | 16.3 | 0.12 |
| polarC18 | 780.55 | 309.1315 | 16.3 | 0.12 |
| polarC18 | 780.55 | 188.1413 | 16.3 | 0.12 |
| polarC18 | 780.55 | 347.4859 | 16.3 | 0.12 |
| polarC18 | 780.55 | 223.067 | 16.3 | 0.12 |
| polarC18 | 780.55 | 116.8938 | 16.3 | 0.12 |
| polarC18 | 780.55 | 287.303 | 16.3 | 0.12 |
| polarC18 | 780.55 | 93.4309 | 16.3 | 0.12 |
| polarC18 | 780.55 | 115.4885 | 16.3 | 0.12 |
| polarC18 | 780.55 | 301.1333 | 16.3 | 0.12 |
| polarC18 | 780.55 | 359.0737 | 16.2 | 0.12 |
| polarC18 | 780.55 | 91.9897 | 16.2 | 0.12 |
| polarC18 | 780.55 | 93.6922 | 16.2 | 0.12 |
| polarC18 | 780.55 | 150.9263 | 16.2 | 0.12 |
| polarC18 | 780.55 | 399.7029 | 16.2 | 0.12 |
| polarC18 | 780.55 | 176.9029 | 16.2 | 0.12 |
| polarC18 | 780.55 | 123.6337 | 16.2 | 0.12 |
| polarC18 | 780.55 | 137.932 | 16.2 | 0.12 |
| polarC18 | 780.55 | 730.2576 | 16.2 | 0.12 |
| polarC18 | 780.55 | 157.175 | 16.2 | 0.12 |
| polarC18 | 780.55 | 282.1701 | 16.2 | 0.12 |
| polarC18 | 780.55 | 174.9998 | 16.2 | 0.12 |
| polarC18 | 780.55 | 95.031 | 16.2 | 0.12 |
| polarC18 | 780.55 | 127.3124 | 16.2 | 0.12 |
| polarC18 | 780.55 | 191.517 | 16.2 | 0.12 |
| polarC18 | 780.55 | 802.269 | 16.2 | 0.12 |
| polarC18 | 780.55 | 127.2785 | 16.2 | 0.12 |
| polarC18 | 780.55 | 485.5222 | 16.2 | 0.12 |

| | | | | |
|---|---|---|---|---|
| polarC18 | 780.55 | 198.2948 | 16.2 | 0.12 |
| polarC18 | 780.55 | 478.4337 | 16.2 | 0.12 |
| polarC18 | 780.55 | 423.9189 | 16.2 | 0.12 |
| polarC18 | 780.55 | 177.4226 | 16.2 | 0.12 |
| polarC18 | 780.55 | 105.2919 | 16.2 | 0.12 |
| polarC18 | 780.55 | 228.5856 | 16.2 | 0.12 |
| polarC18 | 780.55 | 262.5543 | 16.2 | 0.12 |
| polarC18 | 780.55 | 95.2412 | 16.2 | 0.12 |
| polarC18 | 780.55 | 93.3041 | 16.2 | 0.12 |
| polarC18 | 780.55 | 81.8181 | 16.2 | 0.12 |
| polarC18 | 780.55 | 220.2744 | 16.2 | 0.12 |
| polarC18 | 780.55 | 226.7693 | 16.2 | 0.12 |
| polarC18 | 780.55 | 242.0259 | 16.2 | 0.12 |
| polarC18 | 780.55 | 402.2729 | 16.2 | 0.12 |
| polarC18 | 780.55 | 299.2481 | 16.2 | 0.12 |
| polarC18 | 780.55 | 456.2935 | 16.2 | 0.12 |
| polarC18 | 780.55 | 109.5798 | 16.2 | 0.12 |
| polarC18 | 780.55 | 124.1841 | 16.2 | 0.12 |
| polarC18 | 780.55 | 116.2917 | 16.2 | 0.12 |
| polarC18 | 780.55 | 178.956 | 16.2 | 0.12 |
| polarC18 | 780.55 | 392.208 | 16.2 | 0.12 |
| polarC18 | 780.55 | 138.4133 | 16.2 | 0.12 |
| polarC18 | 780.55 | 145.0173 | 16.2 | 0.12 |
| polarC18 | 780.55 | 455.0912 | 16.2 | 0.12 |
| polarC18 | 780.55 | 97.7912 | 16.2 | 0.12 |
| polarC18 | 780.55 | 527.7101 | 16.2 | 0.12 |
| polarC18 | 780.55 | 512.3051 | 16.2 | 0.12 |
| polarC18 | 780.55 | 287.7867 | 16.2 | 0.12 |
| polarC18 | 780.55 | 462.572 | 16.2 | 0.12 |
| polarC18 | 780.55 | 129.2629 | 16.2 | 0.12 |
| polarC18 | 780.55 | 171.9378 | 16.2 | 0.12 |
| polarC18 | 780.55 | 91.4259 | 16.2 | 0.12 |
| polarC18 | 780.55 | 208.1927 | 16.2 | 0.12 |
| polarC18 | 780.55 | 81.968 | 16.2 | 0.12 |
| polarC18 | 780.55 | 213.9162 | 16.2 | 0.12 |
| polarC18 | 780.55 | 580.9991 | 16.2 | 0.12 |
| polarC18 | 780.55 | 254.2937 | 16.2 | 0.12 |
| polarC18 | 780.55 | 142.7203 | 16.2 | 0.12 |
| polarC18 | 780.55 | 663.6111 | 16.2 | 0.12 |
| polarC18 | 780.55 | 213.6454 | 16.2 | 0.12 |
| polarC18 | 780.55 | 472.0542 | 16.2 | 0.12 |
| polarC18 | 780.55 | 124.3973 | 16.2 | 0.12 |

| | | | | |
|---|---|---|---|---|
| polarC18 | 780.55 | 128.3961 | 16.2 | 0.12 |
| polarC18 | 780.55 | 163.2992 | 16.2 | 0.12 |
| polarC18 | 780.55 | 193.925 | 16.2 | 0.12 |
| polarC18 | 780.55 | 400.4381 | 16.2 | 0.12 |
| polarC18 | 780.55 | 108.2144 | 16.2 | 0.12 |
| polarC18 | 780.55 | 358.5475 | 16.2 | 0.12 |
| polarC18 | 780.55 | 112.2534 | 16.2 | 0.12 |
| polarC18 | 780.55 | 107.8207 | 16.2 | 0.12 |
| polarC18 | 780.55 | 149.9137 | 16.2 | 0.12 |
| polarC18 | 780.55 | 395.9077 | 16.2 | 0.12 |
| polarC18 | 780.55 | 192.1138 | 16.2 | 0.12 |
| polarC18 | 780.55 | 151.0528 | 16.2 | 0.12 |
| polarC18 | 780.55 | 166.9783 | 16.2 | 0.12 |
| polarC18 | 780.55 | 606.5773 | 16.2 | 0.12 |
| polarC18 | 780.55 | 488.7493 | 16.2 | 0.12 |
| polarC18 | 780.55 | 96.0906 | 16.2 | 0.12 |
| polarC18 | 780.55 | 198.653 | 16.2 | 0.12 |
| polarC18 | 780.55 | 571.6699 | 16.2 | 0.12 |
| polarC18 | 780.55 | 618.8514 | 16.2 | 0.12 |
| polarC18 | 780.55 | 354.7855 | 16.2 | 0.12 |
| polarC18 | 780.55 | 119.9904 | 16.2 | 0.12 |
| polarC18 | 780.55 | 129.5295 | 16.2 | 0.12 |
| polarC18 | 780.55 | 246.6508 | 16.1 | 0.12 |
| polarC18 | 780.55 | 326.087 | 16.1 | 0.12 |
| polarC18 | 780.55 | 136.462 | 16.1 | 0.12 |
| polarC18 | 780.55 | 575.1613 | 16.1 | 0.12 |
| polarC18 | 780.55 | 199.1843 | 16.1 | 0.12 |
| polarC18 | 780.55 | 340.6561 | 16.1 | 0.12 |
| polarC18 | 780.55 | 127.4147 | 16.1 | 0.12 |
| polarC18 | 780.55 | 223.5352 | 16.1 | 0.12 |
| polarC18 | 780.55 | 382.9798 | 16.1 | 0.12 |
| polarC18 | 780.55 | 338.5016 | 16.1 | 0.12 |
| polarC18 | 780.55 | 480.1125 | 16.1 | 0.12 |
| polarC18 | 780.55 | 88.3975 | 16.1 | 0.12 |
| polarC18 | 780.55 | 458.3751 | 16.1 | 0.12 |
| polarC18 | 780.55 | 228.4606 | 16.1 | 0.12 |
| polarC18 | 780.55 | 347.747 | 16.1 | 0.12 |
| polarC18 | 780.55 | 113.693 | 16.1 | 0.12 |
| polarC18 | 780.55 | 94.8788 | 16.1 | 0.12 |
| polarC18 | 780.55 | 365.7247 | 16.1 | 0.12 |
| polarC18 | 780.55 | 550.9209 | 16.1 | 0.12 |
| polarC18 | 780.55 | 82.6218 | 16.1 | 0.12 |

| | | | | |
|---|---|---|---|---|
| polarC18 | 780.55 | 241.814 | 16.1 | 0.12 |
| polarC18 | 780.55 | 156.8047 | 16.1 | 0.12 |
| polarC18 | 780.55 | 91.8838 | 16.1 | 0.12 |
| polarC18 | 780.55 | 236.134 | 16.1 | 0.12 |
| polarC18 | 780.55 | 86.4157 | 16.1 | 0.12 |
| polarC18 | 780.55 | 89.9261 | 16.1 | 0.12 |
| polarC18 | 780.55 | 401.2411 | 16.1 | 0.12 |
| polarC18 | 780.55 | 724.7669 | 16.1 | 0.12 |
| polarC18 | 780.55 | 283.4875 | 16.1 | 0.12 |
| polarC18 | 780.55 | 245.2352 | 16.1 | 0.12 |
| polarC18 | 780.55 | 103.3047 | 16.1 | 0.12 |
| polarC18 | 780.55 | 102.1698 | 16.1 | 0.12 |
| polarC18 | 780.55 | 91.5135 | 16.1 | 0.12 |
| polarC18 | 780.55 | 177.9731 | 16.1 | 0.12 |
| polarC18 | 780.55 | 466.3358 | 16.1 | 0.12 |
| polarC18 | 780.55 | 364.2734 | 16.1 | 0.12 |
| polarC18 | 780.55 | 165.371 | 16.1 | 0.12 |
| polarC18 | 780.55 | 204.0607 | 16.1 | 0.12 |
| polarC18 | 780.55 | 102.9775 | 16.1 | 0.12 |
| polarC18 | 780.55 | 204.3522 | 16.1 | 0.12 |
| polarC18 | 780.55 | 122.4511 | 16.1 | 0.12 |
| polarC18 | 780.55 | 117.2314 | 16.1 | 0.12 |
| polarC18 | 780.55 | 760.1125 | 16.1 | 0.12 |
| polarC18 | 780.55 | 174.3213 | 16.1 | 0.12 |
| polarC18 | 780.55 | 170.9746 | 16.1 | 0.12 |
| polarC18 | 780.55 | 253.8902 | 16.1 | 0.12 |
| polarC18 | 780.55 | 164.6014 | 16.1 | 0.12 |
| polarC18 | 780.55 | 506.0748 | 16.1 | 0.12 |
| polarC18 | 780.55 | 502.4025 | 16.1 | 0.12 |
| polarC18 | 780.55 | 85.9581 | 16.1 | 0.12 |
| polarC18 | 780.55 | 391.7885 | 16.1 | 0.12 |
| polarC18 | 780.55 | 355.1378 | 16.1 | 0.12 |
| polarC18 | 780.55 | 245.5938 | 16.1 | 0.12 |
| polarC18 | 780.55 | 111.5277 | 16.1 | 0.12 |
| polarC18 | 780.55 | 273.5587 | 16.1 | 0.12 |
| polarC18 | 780.55 | 201.4368 | 16.1 | 0.12 |
| polarC18 | 780.55 | 191.7972 | 16.1 | 0.12 |
| polarC18 | 780.55 | 740.6204 | 16.1 | 0.12 |
| polarC18 | 780.55 | 178.9964 | 16.1 | 0.12 |
| polarC18 | 780.55 | 88.6986 | 16.1 | 0.12 |
| polarC18 | 780.55 | 355.054 | 16.1 | 0.12 |
| polarC18 | 780.55 | 251.5889 | 16.1 | 0.12 |

| | | | | |
|---|---|---|---|---|
| polarC18 | 780.55 | 135.1916 | 16.1 | 0.12 |
| polarC18 | 780.55 | 86.7506 | 16.1 | 0.12 |
| polarC18 | 780.55 | 111.6759 | 16.1 | 0.12 |
| polarC18 | 780.55 | 208.2529 | 16.1 | 0.12 |
| polarC18 | 780.55 | 196.9528 | 16.1 | 0.12 |
| polarC18 | 780.55 | 257.1986 | 16.1 | 0.12 |
| polarC18 | 780.55 | 229.2977 | 16.1 | 0.12 |
| polarC18 | 780.55 | 426.4698 | 16.1 | 0.12 |
| polarC18 | 780.55 | 128.9257 | 16.1 | 0.12 |
| polarC18 | 780.55 | 110.6422 | 16.1 | 0.12 |
| polarC18 | 780.55 | 101.7678 | 16.1 | 0.12 |
| polarC18 | 780.55 | 263.1629 | 16.1 | 0.12 |
| polarC18 | 780.55 | 224.2554 | 16.1 | 0.12 |
| polarC18 | 780.55 | 209.9673 | 16.1 | 0.12 |
| polarC18 | 780.55 | 441.4434 | 16.1 | 0.12 |
| polarC18 | 780.55 | 100.5177 | 16.1 | 0.12 |
| polarC18 | 780.55 | 167.2643 | 16.1 | 0.12 |
| polarC18 | 780.55 | 128.4102 | 16 | 0.12 |
| polarC18 | 780.55 | 261.5266 | 16 | 0.12 |
| polarC18 | 780.55 | 221.9506 | 16 | 0.12 |
| polarC18 | 780.55 | 166.1727 | 16 | 0.12 |
| polarC18 | 780.55 | 209.6364 | 16 | 0.12 |
| polarC18 | 780.55 | 152.6514 | 16 | 0.12 |
| polarC18 | 780.55 | 230.088 | 16 | 0.12 |
| polarC18 | 780.55 | 115.2504 | 16 | 0.12 |
| polarC18 | 780.55 | 307.9162 | 16 | 0.12 |
| polarC18 | 780.55 | 107.5736 | 16 | 0.12 |
| polarC18 | 780.55 | 337.0402 | 16 | 0.12 |
| polarC18 | 780.55 | 582.5303 | 16 | 0.12 |
| polarC18 | 780.55 | 258.5946 | 16 | 0.12 |
| polarC18 | 780.55 | 279.0588 | 16 | 0.12 |
| polarC18 | 780.55 | 98.5159 | 16 | 0.12 |
| polarC18 | 780.55 | 105.1017 | 16 | 0.12 |
| polarC18 | 780.55 | 106.5176 | 16 | 0.12 |
| polarC18 | 780.55 | 127.0176 | 16 | 0.12 |
| polarC18 | 780.55 | 252.5879 | 16 | 0.12 |
| polarC18 | 780.55 | 134.4291 | 16 | 0.12 |
| polarC18 | 780.55 | 692.9719 | 16 | 0.12 |
| polarC18 | 780.55 | 197.6629 | 16 | 0.12 |
| polarC18 | 780.55 | 82.9448 | 16 | 0.12 |
| polarC18 | 780.55 | 99.4949 | 16 | 0.12 |
| polarC18 | 780.55 | 799.3035 | 16 | 0.12 |

| | | | | |
|---|---|---|---|---|
| polarC18 | 780.55 | 816.5645 | 16 | 0.12 |
| polarC18 | 780.55 | 116.5474 | 16 | 0.12 |
| polarC18 | 780.55 | 218.4513 | 16 | 0.12 |
| polarC18 | 780.55 | 158.1242 | 16 | 0.12 |
| polarC18 | 780.55 | 295.1916 | 16 | 0.12 |
| polarC18 | 780.55 | 335.3712 | 16 | 0.12 |
| polarC18 | 780.55 | 339.1944 | 16 | 0.12 |
| polarC18 | 780.55 | 98.491 | 16 | 0.12 |
| polarC18 | 780.55 | 255.4422 | 16 | 0.12 |
| polarC18 | 780.55 | 445.7052 | 16 | 0.12 |
| polarC18 | 780.55 | 154.6093 | 16 | 0.12 |
| polarC18 | 780.55 | 162.8218 | 16 | 0.12 |
| polarC18 | 780.55 | 145.2709 | 16 | 0.12 |
| polarC18 | 780.55 | 515.3834 | 16 | 0.12 |
| polarC18 | 780.55 | 214.7599 | 16 | 0.12 |
| polarC18 | 780.55 | 126.3685 | 16 | 0.12 |
| polarC18 | 780.55 | 118.934 | 16 | 0.12 |
| polarC18 | 780.55 | 578.4868 | 16 | 0.12 |
| polarC18 | 780.55 | 93.3302 | 16 | 0.12 |
| polarC18 | 780.55 | 606.3597 | 16 | 0.12 |
| polarC18 | 780.55 | 166.9897 | 16 | 0.12 |
| polarC18 | 780.55 | 102.7691 | 16 | 0.12 |
| polarC18 | 780.55 | 96.9641 | 16 | 0.12 |
| polarC18 | 780.55 | 151.1975 | 16 | 0.12 |
| polarC18 | 780.55 | 140.5348 | 16 | 0.12 |
| polarC18 | 780.55 | 172.6732 | 16 | 0.12 |
| polarC18 | 780.55 | 316.1676 | 16 | 0.12 |
| polarC18 | 780.55 | 123.748 | 16 | 0.12 |
| polarC18 | 780.55 | 767.3683 | 16 | 0.12 |
| polarC18 | 780.55 | 86.6349 | 16 | 0.12 |
| polarC18 | 780.55 | 129.8141 | 16 | 0.12 |
| polarC18 | 780.55 | 260.267 | 16 | 0.12 |
| polarC18 | 780.55 | 555.6185 | 16 | 0.12 |
| polarC18 | 780.55 | 234.3557 | 16 | 0.12 |
| polarC18 | 780.55 | 708.5848 | 16 | 0.12 |
| polarC18 | 780.55 | 90.2653 | 16 | 0.12 |
| polarC18 | 780.55 | 428.0644 | 16 | 0.12 |
| polarC18 | 780.55 | 311.5153 | 16 | 0.12 |
| polarC18 | 780.55 | 180.1465 | 16 | 0.12 |
| polarC18 | 780.55 | 299.8414 | 16 | 0.12 |
| polarC18 | 780.55 | 381.9429 | 16 | 0.12 |
| polarC18 | 780.55 | 101.6575 | 16 | 0.12 |

| | | | | |
|---|---|---|---|---|
| polarC18 | 780.55 | 363.5292 | 16 | 0.12 |
| polarC18 | 780.55 | 95.8279 | 16 | 0.12 |
| polarC18 | 780.55 | 376.3716 | 16 | 0.12 |
| polarC18 | 780.55 | 100.7696 | 16 | 0.12 |
| polarC18 | 780.55 | 610.8135 | 16 | 0.12 |
| polarC18 | 780.55 | 261.8508 | 16 | 0.12 |
| polarC18 | 780.55 | 151.1734 | 16 | 0.12 |
| polarC18 | 780.55 | 649.8538 | 16 | 0.12 |
| polarC18 | 780.55 | 130.5591 | 16 | 0.12 |
| polarC18 | 780.55 | 173.5063 | 16 | 0.12 |
| polarC18 | 780.55 | 91.1765 | 16 | 0.12 |
| polarC18 | 780.55 | 170.25 | 16 | 0.12 |
| polarC18 | 780.55 | 86.2601 | 16 | 0.12 |
| polarC18 | 780.55 | 90.7317 | 16 | 0.12 |
| polarC18 | 780.55 | 247.6413 | 16 | 0.12 |
| polarC18 | 780.55 | 110.2629 | 16 | 0.12 |
| polarC18 | 780.55 | 144.2003 | 16 | 0.12 |
| polarC18 | 780.55 | 119.8481 | 15.9 | 0.12 |
| polarC18 | 780.55 | 276.6261 | 15.9 | 0.12 |
| polarC18 | 780.55 | 315.3391 | 15.9 | 0.12 |
| polarC18 | 780.55 | 144.4117 | 15.9 | 0.12 |
| polarC18 | 780.55 | 624.9558 | 15.9 | 0.12 |
| polarC18 | 780.55 | 395.6281 | 15.9 | 0.12 |
| polarC18 | 780.55 | 118.7441 | 15.9 | 0.12 |
| polarC18 | 780.55 | 148.381 | 15.9 | 0.12 |
| polarC18 | 780.55 | 489.8925 | 15.9 | 0.12 |
| polarC18 | 780.55 | 93.4721 | 15.9 | 0.12 |
| polarC18 | 780.55 | 517.5417 | 15.9 | 0.12 |
| polarC18 | 780.55 | 287.9715 | 15.9 | 0.12 |
| polarC18 | 780.55 | 538.3563 | 15.9 | 0.12 |
| polarC18 | 780.55 | 160.4413 | 15.9 | 0.12 |
| polarC18 | 780.55 | 424.0738 | 15.9 | 0.12 |
| polarC18 | 780.55 | 217.8748 | 15.9 | 0.12 |
| polarC18 | 780.55 | 88.6294 | 15.9 | 0.12 |
| polarC18 | 780.55 | 259.4164 | 15.9 | 0.12 |
| polarC18 | 780.55 | 86.1739 | 15.9 | 0.12 |
| polarC18 | 780.55 | 114.259 | 15.9 | 0.12 |
| polarC18 | 780.55 | 638.3853 | 15.9 | 0.12 |
| polarC18 | 780.55 | 123.4905 | 15.9 | 0.12 |
| polarC18 | 780.55 | 412.5603 | 15.9 | 0.12 |
| polarC18 | 780.55 | 98.1099 | 15.9 | 0.12 |
| polarC18 | 780.55 | 93.6557 | 15.9 | 0.12 |

| | | | | |
|---|---|---|---|---|
| polarC18 | 780.55 | 464.8501 | 15.9 | 0.12 |
| polarC18 | 780.55 | 103.3743 | 15.9 | 0.12 |
| polarC18 | 780.55 | 170.0186 | 15.9 | 0.12 |
| polarC18 | 780.55 | 298.6726 | 15.9 | 0.12 |
| polarC18 | 780.55 | 705.6423 | 15.9 | 0.12 |
| polarC18 | 780.55 | 125.3376 | 15.9 | 0.12 |
| polarC18 | 780.55 | 226.3553 | 15.9 | 0.12 |
| polarC18 | 780.55 | 278.0657 | 15.9 | 0.12 |
| polarC18 | 780.55 | 157.0528 | 15.9 | 0.12 |
| polarC18 | 780.55 | 88.7559 | 15.9 | 0.12 |
| polarC18 | 780.55 | 311.4692 | 15.9 | 0.12 |
| polarC18 | 780.55 | 266.0423 | 15.9 | 0.12 |
| polarC18 | 780.55 | 237.1129 | 15.9 | 0.12 |
| polarC18 | 780.55 | 184.6075 | 15.9 | 0.12 |
| polarC18 | 780.55 | 143.8685 | 15.9 | 0.12 |
| polarC18 | 780.55 | 103.5717 | 15.9 | 0.12 |
| polarC18 | 780.55 | 316.0128 | 15.9 | 0.12 |
| polarC18 | 780.55 | 240.8782 | 15.9 | 0.12 |
| polarC18 | 780.55 | 313.0305 | 15.9 | 0.12 |
| polarC18 | 780.55 | 418.9063 | 15.9 | 0.12 |
| polarC18 | 780.55 | 100.9721 | 15.9 | 0.12 |
| polarC18 | 780.55 | 434.9158 | 15.9 | 0.12 |
| polarC18 | 780.55 | 257.3384 | 15.9 | 0.12 |
| polarC18 | 780.55 | 96.7509 | 15.9 | 0.12 |
| polarC18 | 780.55 | 101.8596 | 15.9 | 0.12 |
| polarC18 | 780.55 | 712.334 | 15.9 | 0.12 |
| polarC18 | 780.55 | 97.3043 | 15.9 | 0.12 |
| polarC18 | 780.55 | 200.972 | 15.9 | 0.12 |
| polarC18 | 780.55 | 301.6042 | 15.9 | 0.12 |
| polarC18 | 780.55 | 337.853 | 15.9 | 0.12 |
| polarC18 | 780.55 | 194.5159 | 15.9 | 0.12 |
| polarC18 | 780.55 | 723.8555 | 15.9 | 0.12 |
| polarC18 | 780.55 | 215.2739 | 15.9 | 0.12 |
| polarC18 | 780.55 | 82.7247 | 15.9 | 0.12 |
| polarC18 | 780.55 | 370.9774 | 15.9 | 0.12 |
| polarC18 | 780.55 | 630.5067 | 15.9 | 0.12 |
| polarC18 | 780.55 | 101.6341 | 15.9 | 0.12 |
| polarC18 | 780.55 | 384.6665 | 15.9 | 0.12 |
| polarC18 | 780.55 | 165.4667 | 15.9 | 0.12 |
| polarC18 | 780.55 | 125.1633 | 15.9 | 0.12 |
| polarC18 | 780.55 | 177.2131 | 15.9 | 0.12 |
| polarC18 | 780.55 | 172.7046 | 15.9 | 0.12 |

| | | | | |
|---|---|---|---|---|
| polarC18 | 780.55 | 710.0124 | 15.9 | 0.12 |
| polarC18 | 780.55 | 419.7748 | 15.9 | 0.12 |
| polarC18 | 780.55 | 163.4144 | 15.9 | 0.12 |
| polarC18 | 780.55 | 272.7681 | 15.9 | 0.12 |
| polarC18 | 780.55 | 632.3752 | 15.9 | 0.12 |
| polarC18 | 780.55 | 81.6729 | 15.9 | 0.12 |
| polarC18 | 780.55 | 144.507 | 15.9 | 0.12 |
| polarC18 | 780.55 | 151.3204 | 15.9 | 0.12 |
| polarC18 | 780.55 | 177.468 | 15.9 | 0.12 |
| polarC18 | 780.55 | 715.9034 | 15.9 | 0.12 |
| polarC18 | 780.55 | 87.8665 | 15.9 | 0.12 |
| polarC18 | 780.55 | 303.6718 | 15.9 | 0.12 |
| polarC18 | 780.55 | 88.5849 | 15.9 | 0.12 |
| polarC18 | 780.55 | 99.5409 | 15.9 | 0.12 |
| polarC18 | 780.55 | 166.7865 | 15.9 | 0.12 |
| polarC18 | 780.55 | 154.7165 | 15.9 | 0.12 |
| polarC18 | 780.55 | 396.8587 | 15.8 | 0.12 |
| polarC18 | 780.55 | 109.2784 | 15.8 | 0.12 |
| polarC18 | 780.55 | 396.2336 | 15.8 | 0.12 |
| polarC18 | 780.55 | 181.5593 | 15.8 | 0.12 |
| polarC18 | 780.55 | 194.9023 | 15.8 | 0.12 |
| polarC18 | 780.55 | 156.7109 | 15.8 | 0.12 |
| polarC18 | 780.55 | 145.3105 | 15.8 | 0.12 |
| polarC18 | 780.55 | 85.3077 | 15.8 | 0.12 |
| polarC18 | 780.55 | 392.4523 | 15.8 | 0.12 |
| polarC18 | 780.55 | 174.3373 | 15.8 | 0.12 |
| polarC18 | 780.55 | 271.4479 | 15.8 | 0.12 |
| polarC18 | 780.55 | 282.1302 | 15.8 | 0.12 |
| polarC18 | 780.55 | 119.9699 | 15.8 | 0.12 |
| polarC18 | 780.55 | 138.3502 | 15.8 | 0.12 |
| polarC18 | 780.55 | 83.7635 | 15.8 | 0.12 |
| polarC18 | 780.55 | 397.833 | 15.8 | 0.12 |
| polarC18 | 780.55 | 451.1558 | 15.8 | 0.12 |
| polarC18 | 780.55 | 103.971 | 15.8 | 0.12 |
| polarC18 | 780.55 | 82.484 | 15.8 | 0.12 |
| polarC18 | 780.55 | 134.9808 | 15.8 | 0.12 |
| polarC18 | 780.55 | 489.7928 | 15.8 | 0.12 |
| polarC18 | 780.55 | 293.8724 | 15.8 | 0.12 |
| polarC18 | 780.55 | 310.106 | 15.8 | 0.12 |
| polarC18 | 780.55 | 421.2266 | 15.8 | 0.12 |
| polarC18 | 780.55 | 245.4013 | 15.8 | 0.12 |
| polarC18 | 780.55 | 469.0166 | 15.8 | 0.12 |

| | | | | |
|---|---|---|---|---|
| polarC18 | 780.55 | 317.7129 | 15.8 | 0.12 |
| polarC18 | 780.55 | 276.0127 | 15.8 | 0.12 |
| polarC18 | 780.55 | 179.828 | 15.8 | 0.12 |
| polarC18 | 780.55 | 688.5453 | 15.8 | 0.12 |
| polarC18 | 780.55 | 210.5279 | 15.8 | 0.12 |
| polarC18 | 780.55 | 222.9716 | 15.8 | 0.12 |
| polarC18 | 780.55 | 156.5188 | 15.8 | 0.12 |
| polarC18 | 780.55 | 90.8077 | 15.8 | 0.12 |
| polarC18 | 780.55 | 101.3982 | 15.8 | 0.12 |
| polarC18 | 780.55 | 366.4192 | 15.8 | 0.12 |
| polarC18 | 780.55 | 140.6594 | 15.8 | 0.12 |
| polarC18 | 780.55 | 188.4861 | 15.8 | 0.12 |
| polarC18 | 780.55 | 614.2578 | 15.8 | 0.12 |
| polarC18 | 780.55 | 105.5234 | 15.8 | 0.12 |
| polarC18 | 780.55 | 196.4326 | 15.8 | 0.12 |
| polarC18 | 780.55 | 707.9085 | 15.8 | 0.12 |
| polarC18 | 780.55 | 457.408 | 15.8 | 0.12 |
| polarC18 | 780.55 | 243.6064 | 15.8 | 0.12 |
| polarC18 | 780.55 | 90.2805 | 15.8 | 0.12 |
| polarC18 | 780.55 | 245.8535 | 15.8 | 0.12 |
| polarC18 | 780.55 | 95.7432 | 15.8 | 0.12 |
| polarC18 | 780.55 | 93.0686 | 15.8 | 0.12 |
| polarC18 | 780.55 | 92.9363 | 15.8 | 0.12 |
| polarC18 | 780.55 | 264.6449 | 15.8 | 0.12 |
| polarC18 | 780.55 | 86.2248 | 15.8 | 0.12 |
| polarC18 | 780.55 | 244.1547 | 15.8 | 0.12 |
| polarC18 | 780.55 | 85.0406 | 15.8 | 0.12 |
| polarC18 | 780.55 | 138.9032 | 15.8 | 0.12 |
| polarC18 | 780.55 | 201.0671 | 15.8 | 0.12 |
| polarC18 | 780.55 | 122.809 | 15.8 | 0.12 |
| polarC18 | 780.55 | 239.3229 | 15.8 | 0.12 |
| polarC18 | 780.55 | 87.3964 | 15.8 | 0.12 |
| polarC18 | 780.55 | 241.5965 | 15.8 | 0.12 |
| polarC18 | 780.55 | 403.7662 | 15.8 | 0.12 |
| polarC18 | 780.55 | 229.2054 | 15.8 | 0.12 |
| polarC18 | 780.55 | 234.1514 | 15.8 | 0.12 |
| polarC18 | 780.55 | 129.931 | 15.8 | 0.12 |
| polarC18 | 780.55 | 121.5388 | 15.8 | 0.12 |
| polarC18 | 780.55 | 82.6007 | 15.8 | 0.12 |
| polarC18 | 780.55 | 183.8786 | 15.8 | 0.12 |
| polarC18 | 780.55 | 389.3719 | 15.8 | 0.12 |
| polarC18 | 780.55 | 148.9118 | 15.8 | 0.12 |

| | | | | |
|---|---|---|---|---|
| polarC18 | 780.55 | 468.6668 | 15.8 | 0.12 |
| polarC18 | 780.55 | 343.788 | 15.8 | 0.12 |
| polarC18 | 780.55 | 125.1275 | 15.8 | 0.12 |
| polarC18 | 780.55 | 381.031 | 15.8 | 0.12 |
| polarC18 | 780.55 | 120.2101 | 15.8 | 0.12 |
| polarC18 | 780.55 | 111.9599 | 15.8 | 0.12 |
| polarC18 | 780.55 | 123.5665 | 15.8 | 0.12 |
| polarC18 | 780.55 | 236.9494 | 15.8 | 0.12 |
| polarC18 | 780.55 | 86.3074 | 15.8 | 0.12 |
| polarC18 | 780.55 | 88.7412 | 15.8 | 0.12 |
| polarC18 | 780.55 | 237.3956 | 15.8 | 0.12 |
| polarC18 | 780.55 | 245.9824 | 15.8 | 0.12 |
| polarC18 | 780.55 | 302.5251 | 15.8 | 0.12 |
| polarC18 | 780.55 | 315.2897 | 15.7 | 0.12 |
| polarC18 | 780.55 | 83.424 | 15.7 | 0.12 |
| polarC18 | 780.55 | 130.8156 | 15.7 | 0.12 |
| polarC18 | 780.55 | 429.0821 | 15.7 | 0.12 |
| polarC18 | 780.55 | 121.61 | 15.7 | 0.12 |
| polarC18 | 780.55 | 153.8408 | 15.7 | 0.12 |
| polarC18 | 780.55 | 680.1827 | 15.7 | 0.12 |
| polarC18 | 780.55 | 168.9908 | 15.7 | 0.12 |
| polarC18 | 780.55 | 296.4901 | 15.7 | 0.12 |
| polarC18 | 780.55 | 555.0518 | 15.7 | 0.12 |
| polarC18 | 780.55 | 126.2238 | 15.7 | 0.12 |
| polarC18 | 780.55 | 415.8087 | 15.7 | 0.12 |
| polarC18 | 780.55 | 370.2645 | 15.7 | 0.12 |
| polarC18 | 780.55 | 171.5925 | 15.7 | 0.12 |
| polarC18 | 780.55 | 118.6928 | 15.7 | 0.12 |
| polarC18 | 780.55 | 184.2296 | 15.7 | 0.12 |
| polarC18 | 780.55 | 183.0289 | 15.7 | 0.12 |
| polarC18 | 780.55 | 202.0495 | 15.7 | 0.12 |
| polarC18 | 780.55 | 134.3821 | 15.7 | 0.12 |
| polarC18 | 780.55 | 126.6861 | 15.7 | 0.12 |
| polarC18 | 780.55 | 213.3056 | 15.7 | 0.12 |
| polarC18 | 780.55 | 96.2252 | 15.7 | 0.12 |
| polarC18 | 780.55 | 91.2394 | 15.7 | 0.12 |
| polarC18 | 780.55 | 167.8697 | 15.7 | 0.12 |
| polarC18 | 780.55 | 88.5126 | 15.7 | 0.12 |
| polarC18 | 780.55 | 133.8047 | 15.7 | 0.12 |
| polarC18 | 780.55 | 230.9789 | 15.7 | 0.12 |
| polarC18 | 780.55 | 189.0306 | 15.7 | 0.12 |
| polarC18 | 780.55 | 100.9053 | 15.7 | 0.12 |

| | | | | |
|---|---|---|---|---|
| polarC18 | 780.55 | 83.9826 | 15.7 | 0.12 |
| polarC18 | 780.55 | 395.0025 | 15.7 | 0.12 |
| polarC18 | 780.55 | 172.9946 | 15.7 | 0.12 |
| polarC18 | 780.55 | 194.1303 | 15.7 | 0.12 |
| polarC18 | 780.55 | 94.2967 | 15.7 | 0.12 |
| polarC18 | 780.55 | 82.1999 | 15.7 | 0.12 |
| polarC18 | 780.55 | 350.0159 | 15.7 | 0.12 |
| polarC18 | 780.55 | 671.4554 | 15.7 | 0.12 |
| polarC18 | 780.55 | 144.136 | 15.7 | 0.12 |
| polarC18 | 780.55 | 152.7269 | 15.7 | 0.12 |
| polarC18 | 780.55 | 367.8398 | 15.7 | 0.12 |
| polarC18 | 780.55 | 519.2107 | 15.7 | 0.12 |
| polarC18 | 780.55 | 83.6596 | 15.7 | 0.12 |
| polarC18 | 780.55 | 262.2643 | 15.7 | 0.12 |
| polarC18 | 780.55 | 178.38 | 15.7 | 0.12 |
| polarC18 | 780.55 | 102.4137 | 15.7 | 0.12 |
| polarC18 | 780.55 | 626.6099 | 15.7 | 0.12 |
| polarC18 | 780.55 | 548.1324 | 15.7 | 0.12 |
| polarC18 | 780.55 | 107.1513 | 15.7 | 0.12 |
| polarC18 | 780.55 | 491.1453 | 15.7 | 0.12 |
| polarC18 | 780.55 | 627.9999 | 15.7 | 0.11 |
| polarC18 | 780.55 | 108.3396 | 15.7 | 0.11 |
| polarC18 | 780.55 | 142.324 | 15.7 | 0.11 |
| polarC18 | 780.55 | 363.4511 | 15.7 | 0.11 |
| polarC18 | 780.55 | 105.5804 | 15.7 | 0.11 |
| polarC18 | 780.55 | 300.0642 | 15.7 | 0.11 |
| polarC18 | 780.55 | 651.0439 | 15.7 | 0.11 |
| polarC18 | 780.55 | 407.4664 | 15.7 | 0.11 |
| polarC18 | 780.55 | 176.4569 | 15.7 | 0.11 |
| polarC18 | 780.55 | 312.9376 | 15.7 | 0.11 |
| polarC18 | 780.55 | 137.8302 | 15.7 | 0.11 |
| polarC18 | 780.55 | 720.109 | 15.7 | 0.11 |
| polarC18 | 780.55 | 110.4038 | 15.7 | 0.11 |
| polarC18 | 780.55 | 141.9053 | 15.7 | 0.11 |
| polarC18 | 780.55 | 134.9606 | 15.7 | 0.11 |
| polarC18 | 780.55 | 179.3143 | 15.7 | 0.11 |
| polarC18 | 780.55 | 538.7931 | 15.7 | 0.11 |
| polarC18 | 780.55 | 120.513 | 15.7 | 0.11 |
| polarC18 | 780.55 | 167.5505 | 15.7 | 0.11 |
| polarC18 | 780.55 | 212.2919 | 15.7 | 0.11 |
| polarC18 | 780.55 | 455.6305 | 15.7 | 0.11 |
| polarC18 | 780.55 | 92.6598 | 15.7 | 0.11 |

| | | | | |
|---|---|---|---|---|
| polarC18 | 780.55 | 206.2419 | 15.7 | 0.11 |
| polarC18 | 780.55 | 178.2944 | 15.7 | 0.11 |
| polarC18 | 780.55 | 99.0021 | 15.7 | 0.11 |
| polarC18 | 780.55 | 117.238 | 15.7 | 0.11 |
| polarC18 | 780.55 | 188.4262 | 15.7 | 0.11 |
| polarC18 | 780.55 | 123.5135 | 15.7 | 0.11 |
| polarC18 | 780.55 | 142.3005 | 15.7 | 0.11 |
| polarC18 | 780.55 | 592.2203 | 15.7 | 0.11 |
| polarC18 | 780.55 | 201.2931 | 15.7 | 0.11 |
| polarC18 | 780.55 | 98.6238 | 15.7 | 0.11 |
| polarC18 | 780.55 | 108.9362 | 15.7 | 0.11 |
| polarC18 | 780.55 | 117.575 | 15.6 | 0.11 |
| polarC18 | 780.55 | 292.5838 | 15.6 | 0.11 |
| polarC18 | 780.55 | 103.876 | 15.6 | 0.11 |
| polarC18 | 780.55 | 133.9704 | 15.6 | 0.11 |
| polarC18 | 780.55 | 178.5468 | 15.6 | 0.11 |
| polarC18 | 780.55 | 254.0349 | 15.6 | 0.11 |
| polarC18 | 780.55 | 120.2262 | 15.6 | 0.11 |
| polarC18 | 780.55 | 94.5853 | 15.6 | 0.11 |
| polarC18 | 780.55 | 219.0703 | 15.6 | 0.11 |
| polarC18 | 780.55 | 109.0019 | 15.6 | 0.11 |
| polarC18 | 780.55 | 248.5976 | 15.6 | 0.11 |
| polarC18 | 780.55 | 82.1433 | 15.6 | 0.11 |
| polarC18 | 780.55 | 168.3932 | 15.6 | 0.11 |
| polarC18 | 780.55 | 123.6405 | 15.6 | 0.11 |
| polarC18 | 780.55 | 91.0678 | 15.6 | 0.11 |
| polarC18 | 780.55 | 132.4 | 15.6 | 0.11 |
| polarC18 | 780.55 | 89.9381 | 15.6 | 0.11 |
| polarC18 | 780.55 | 165.0185 | 15.6 | 0.11 |
| polarC18 | 780.55 | 568.3817 | 15.6 | 0.11 |
| polarC18 | 780.55 | 163.6357 | 15.6 | 0.11 |
| polarC18 | 780.55 | 101.4072 | 15.6 | 0.11 |
| polarC18 | 780.55 | 719.2311 | 15.6 | 0.11 |
| polarC18 | 780.55 | 96.4535 | 15.6 | 0.11 |
| polarC18 | 780.55 | 104.8554 | 15.6 | 0.11 |
| polarC18 | 780.55 | 199.7772 | 15.6 | 0.11 |
| polarC18 | 780.55 | 83.9199 | 15.6 | 0.11 |
| polarC18 | 780.55 | 97.043 | 15.6 | 0.11 |
| polarC18 | 780.55 | 317.3224 | 15.6 | 0.11 |
| polarC18 | 780.55 | 111.608 | 15.6 | 0.11 |
| polarC18 | 780.55 | 141.6 | 15.6 | 0.11 |
| polarC18 | 780.55 | 178.619 | 15.6 | 0.11 |

| | | | | |
|---|---|---|---|---|
| polarC18 | 780.55 | 574.71 | 15.6 | 0.11 |
| polarC18 | 780.55 | 397.6244 | 15.6 | 0.11 |
| polarC18 | 780.55 | 110.953 | 15.6 | 0.11 |
| polarC18 | 780.55 | 98.2664 | 15.6 | 0.11 |
| polarC18 | 780.55 | 335.2807 | 15.6 | 0.11 |
| polarC18 | 780.55 | 82.3139 | 15.6 | 0.11 |
| polarC18 | 780.55 | 173.5912 | 15.6 | 0.11 |
| polarC18 | 780.55 | 174.4698 | 15.6 | 0.11 |
| polarC18 | 780.55 | 244.3295 | 15.6 | 0.11 |
| polarC18 | 780.55 | 298.1955 | 15.6 | 0.11 |
| polarC18 | 780.55 | 807.537 | 15.6 | 0.11 |
| polarC18 | 780.55 | 316.4283 | 15.6 | 0.11 |
| polarC18 | 780.55 | 106.7848 | 15.6 | 0.11 |
| polarC18 | 780.55 | 90.3728 | 15.6 | 0.11 |
| polarC18 | 780.55 | 173.9931 | 15.6 | 0.11 |
| polarC18 | 780.55 | 92.2393 | 15.6 | 0.11 |
| polarC18 | 780.55 | 233.1648 | 15.6 | 0.11 |
| polarC18 | 780.55 | 553.6747 | 15.6 | 0.11 |
| polarC18 | 780.55 | 193.6632 | 15.6 | 0.11 |
| polarC18 | 780.55 | 311.9773 | 15.6 | 0.11 |
| polarC18 | 780.55 | 107.5427 | 15.6 | 0.11 |
| polarC18 | 780.55 | 93.5382 | 15.6 | 0.11 |
| polarC18 | 780.55 | 276.547 | 15.6 | 0.11 |
| polarC18 | 780.55 | 281.6823 | 15.6 | 0.11 |
| polarC18 | 780.55 | 148.9617 | 15.6 | 0.11 |
| polarC18 | 780.55 | 163.2791 | 15.6 | 0.11 |
| polarC18 | 780.55 | 91.6453 | 15.6 | 0.11 |
| polarC18 | 780.55 | 101.4288 | 15.6 | 0.11 |
| polarC18 | 780.55 | 369.5297 | 15.6 | 0.11 |
| polarC18 | 780.55 | 174.4095 | 15.6 | 0.11 |
| polarC18 | 780.55 | 248.8177 | 15.6 | 0.11 |
| polarC18 | 780.55 | 152.0824 | 15.6 | 0.11 |
| polarC18 | 780.55 | 301.0732 | 15.6 | 0.11 |
| polarC18 | 780.55 | 523.6593 | 15.6 | 0.11 |
| polarC18 | 780.55 | 353.7498 | 15.6 | 0.11 |
| polarC18 | 780.55 | 105.3379 | 15.6 | 0.11 |
| polarC18 | 780.55 | 277.1187 | 15.6 | 0.11 |
| polarC18 | 780.55 | 127.8586 | 15.6 | 0.11 |
| polarC18 | 780.55 | 358.7652 | 15.6 | 0.11 |
| polarC18 | 780.55 | 98.062 | 15.6 | 0.11 |
| polarC18 | 780.55 | 326.2418 | 15.6 | 0.11 |
| polarC18 | 780.55 | 173.2154 | 15.6 | 0.11 |

| | | | | |
|---|---|---|---|---|
| polarC18 | 780.55 | 120.3331 | 15.6 | 0.11 |
| polarC18 | 780.55 | 298.5553 | 15.6 | 0.11 |
| polarC18 | 780.55 | 201.043 | 15.6 | 0.11 |
| polarC18 | 780.55 | 118.6062 | 15.6 | 0.11 |
| polarC18 | 780.55 | 185.5086 | 15.6 | 0.11 |
| polarC18 | 780.55 | 159.0809 | 15.6 | 0.11 |
| polarC18 | 780.55 | 383.1202 | 15.6 | 0.11 |
| polarC18 | 780.55 | 656.6436 | 15.6 | 0.11 |
| polarC18 | 780.55 | 235.3986 | 15.6 | 0.11 |
| polarC18 | 780.55 | 518.7319 | 15.6 | 0.11 |
| polarC18 | 780.55 | 409.6167 | 15.6 | 0.11 |
| polarC18 | 780.55 | 148.8263 | 15.6 | 0.11 |
| polarC18 | 780.55 | 384.9626 | 15.6 | 0.11 |
| polarC18 | 780.55 | 93.3087 | 15.6 | 0.11 |
| polarC18 | 780.55 | 182.3438 | 15.5 | 0.11 |
| polarC18 | 780.55 | 120.9186 | 15.5 | 0.11 |
| polarC18 | 780.55 | 88.3728 | 15.5 | 0.11 |
| polarC18 | 780.55 | 218.6664 | 15.5 | 0.11 |
| polarC18 | 780.55 | 96.6973 | 15.5 | 0.11 |
| polarC18 | 780.55 | 427.724 | 15.5 | 0.11 |
| polarC18 | 780.55 | 121.7644 | 15.5 | 0.11 |
| polarC18 | 780.55 | 99.349 | 15.5 | 0.11 |
| polarC18 | 780.55 | 93.287 | 15.5 | 0.11 |
| polarC18 | 780.55 | 183.2168 | 15.5 | 0.11 |
| polarC18 | 780.55 | 126.943 | 15.5 | 0.11 |
| polarC18 | 780.55 | 90.2171 | 15.5 | 0.11 |
| polarC18 | 780.55 | 192.6228 | 15.5 | 0.11 |
| polarC18 | 780.55 | 303.0435 | 15.5 | 0.11 |
| polarC18 | 780.55 | 368.8314 | 15.5 | 0.11 |
| polarC18 | 780.55 | 92.7268 | 15.5 | 0.11 |
| polarC18 | 780.55 | 253.0695 | 15.5 | 0.11 |
| polarC18 | 780.55 | 191.3611 | 15.5 | 0.11 |
| polarC18 | 780.55 | 204.1388 | 15.5 | 0.11 |
| polarC18 | 780.55 | 644.202 | 15.5 | 0.11 |
| polarC18 | 780.55 | 216.1418 | 15.5 | 0.11 |
| polarC18 | 780.55 | 329.7558 | 15.5 | 0.11 |
| polarC18 | 780.55 | 187.2095 | 15.5 | 0.11 |
| polarC18 | 780.55 | 184.0643 | 15.5 | 0.11 |
| polarC18 | 780.55 | 120.5878 | 15.5 | 0.11 |
| polarC18 | 780.55 | 102.6201 | 15.5 | 0.11 |
| polarC18 | 780.55 | 251.177 | 15.5 | 0.11 |
| polarC18 | 780.55 | 94.3321 | 15.5 | 0.11 |

| | | | | |
|---|---|---|---|---|
| polarC18 | 780.55 | 159.8118 | 15.5 | 0.11 |
| polarC18 | 780.55 | 84.5611 | 15.5 | 0.11 |
| polarC18 | 780.55 | 88.0395 | 15.5 | 0.11 |
| polarC18 | 780.55 | 129.1505 | 15.5 | 0.11 |
| polarC18 | 780.55 | 104.3476 | 15.5 | 0.11 |
| polarC18 | 780.55 | 137.4448 | 15.5 | 0.11 |
| polarC18 | 780.55 | 104.0987 | 15.5 | 0.11 |
| polarC18 | 780.55 | 135.1415 | 15.5 | 0.11 |
| polarC18 | 780.55 | 140.0619 | 15.5 | 0.11 |
| polarC18 | 780.55 | 100.9838 | 15.5 | 0.11 |
| polarC18 | 780.55 | 129.5978 | 15.5 | 0.11 |
| polarC18 | 780.55 | 238.6745 | 15.5 | 0.11 |
| polarC18 | 780.55 | 130.2107 | 15.5 | 0.11 |
| polarC18 | 780.55 | 167.1347 | 15.5 | 0.11 |
| polarC18 | 780.55 | 164.7057 | 15.5 | 0.11 |
| polarC18 | 780.55 | 466.8569 | 15.5 | 0.11 |
| polarC18 | 780.55 | 405.0602 | 15.5 | 0.11 |
| polarC18 | 780.55 | 366.1927 | 15.5 | 0.11 |
| polarC18 | 780.55 | 514.3892 | 15.5 | 0.11 |
| polarC18 | 780.55 | 190.7423 | 15.5 | 0.11 |
| polarC18 | 780.55 | 265.4134 | 15.5 | 0.11 |
| polarC18 | 780.55 | 352.8787 | 15.5 | 0.11 |
| polarC18 | 780.55 | 161.0427 | 15.5 | 0.11 |
| polarC18 | 780.55 | 273.8181 | 15.5 | 0.11 |
| polarC18 | 780.55 | 544.6696 | 15.5 | 0.11 |
| polarC18 | 780.55 | 147.8359 | 15.5 | 0.11 |
| polarC18 | 780.55 | 144.2759 | 15.5 | 0.11 |
| polarC18 | 780.55 | 99.7778 | 15.5 | 0.11 |
| polarC18 | 780.55 | 415.1071 | 15.5 | 0.11 |
| polarC18 | 780.55 | 88.5735 | 15.5 | 0.11 |
| polarC18 | 780.55 | 99.9599 | 15.5 | 0.11 |
| polarC18 | 780.55 | 224.877 | 15.5 | 0.11 |
| polarC18 | 780.55 | 198.7064 | 15.5 | 0.11 |
| polarC18 | 780.55 | 349.468 | 15.5 | 0.11 |
| polarC18 | 780.55 | 286.7544 | 15.5 | 0.11 |
| polarC18 | 780.55 | 169.1106 | 15.5 | 0.11 |
| polarC18 | 780.55 | 240.4677 | 15.5 | 0.11 |
| polarC18 | 780.55 | 234.3785 | 15.5 | 0.11 |
| polarC18 | 780.55 | 391.4162 | 15.5 | 0.11 |
| polarC18 | 780.55 | 159.404 | 15.5 | 0.11 |
| polarC18 | 780.55 | 161.927 | 15.5 | 0.11 |
| polarC18 | 780.55 | 116.3216 | 15.5 | 0.11 |

| | | | | |
|---|---|---|---|---|
| polarC18 | 780.55 | 117.6928 | 15.5 | 0.11 |
| polarC18 | 780.55 | 104.6541 | 15.5 | 0.11 |
| polarC18 | 780.55 | 100.2019 | 15.5 | 0.11 |
| polarC18 | 780.55 | 151.3986 | 15.5 | 0.11 |
| polarC18 | 780.55 | 135.735 | 15.5 | 0.11 |
| polarC18 | 780.55 | 117.7589 | 15.5 | 0.11 |
| polarC18 | 780.55 | 147.538 | 15.5 | 0.11 |
| polarC18 | 780.55 | 211.7471 | 15.5 | 0.11 |
| polarC18 | 780.55 | 81.704 | 15.5 | 0.11 |
| polarC18 | 780.55 | 94.0894 | 15.5 | 0.11 |
| polarC18 | 780.55 | 427.9884 | 15.5 | 0.11 |
| polarC18 | 780.55 | 411.9358 | 15.5 | 0.11 |
| polarC18 | 780.55 | 175.2928 | 15.5 | 0.11 |
| polarC18 | 780.55 | 415.583 | 15.5 | 0.11 |
| polarC18 | 780.55 | 152.1928 | 15.5 | 0.11 |
| polarC18 | 780.55 | 290.5459 | 15.4 | 0.11 |
| polarC18 | 780.55 | 364.4829 | 15.4 | 0.11 |
| polarC18 | 780.55 | 499.8603 | 15.4 | 0.11 |
| polarC18 | 780.55 | 474.7142 | 15.4 | 0.11 |
| polarC18 | 780.55 | 179.4962 | 15.4 | 0.11 |
| polarC18 | 780.55 | 108.1993 | 15.4 | 0.11 |
| polarC18 | 780.55 | 94.0753 | 15.4 | 0.11 |
| polarC18 | 780.55 | 134.6618 | 15.4 | 0.11 |
| polarC18 | 780.55 | 85.5683 | 15.4 | 0.11 |
| polarC18 | 780.55 | 189.6754 | 15.4 | 0.11 |
| polarC18 | 780.55 | 107.9495 | 15.4 | 0.11 |
| polarC18 | 780.55 | 86.2843 | 15.4 | 0.11 |
| polarC18 | 780.55 | 102.6675 | 15.4 | 0.11 |
| polarC18 | 780.55 | 173.7835 | 15.4 | 0.11 |
| polarC18 | 780.55 | 129.3127 | 15.4 | 0.11 |
| polarC18 | 780.55 | 458.8553 | 15.4 | 0.11 |
| polarC18 | 780.55 | 189.1711 | 15.4 | 0.11 |
| polarC18 | 780.55 | 145.7971 | 15.4 | 0.11 |
| polarC18 | 780.55 | 358.8703 | 15.4 | 0.11 |
| polarC18 | 780.55 | 293.9756 | 15.4 | 0.11 |
| polarC18 | 780.55 | 266.0097 | 15.4 | 0.11 |
| polarC18 | 780.55 | 83.2474 | 15.4 | 0.11 |
| polarC18 | 780.55 | 124.4516 | 15.4 | 0.11 |
| polarC18 | 780.55 | 411.626 | 15.4 | 0.11 |
| polarC18 | 780.55 | 131.5643 | 15.4 | 0.11 |
| polarC18 | 780.55 | 99.7161 | 15.4 | 0.11 |
| polarC18 | 780.55 | 119.9565 | 15.4 | 0.11 |

| | | | | |
|---|---|---|---|---|
| polarC18 | 780.55 | 88.0059 | 15.4 | 0.11 |
| polarC18 | 780.55 | 304.8123 | 15.4 | 0.11 |
| polarC18 | 780.55 | 93.7831 | 15.4 | 0.11 |
| polarC18 | 780.55 | 509.3164 | 15.4 | 0.11 |
| polarC18 | 780.55 | 231.2808 | 15.4 | 0.11 |
| polarC18 | 780.55 | 82.2669 | 15.4 | 0.11 |
| polarC18 | 780.55 | 197.231 | 15.4 | 0.11 |
| polarC18 | 780.55 | 181.7887 | 15.4 | 0.11 |
| polarC18 | 780.55 | 140.8994 | 15.4 | 0.11 |
| polarC18 | 780.55 | 330.5054 | 15.4 | 0.11 |
| polarC18 | 780.55 | 105.2625 | 15.4 | 0.11 |
| polarC18 | 780.55 | 264.8896 | 15.4 | 0.11 |
| polarC18 | 780.55 | 83.5941 | 15.4 | 0.11 |
| polarC18 | 780.55 | 128.4837 | 15.4 | 0.11 |
| polarC18 | 780.55 | 298.747 | 15.4 | 0.11 |
| polarC18 | 780.55 | 475.8084 | 15.4 | 0.11 |
| polarC18 | 780.55 | 148.4823 | 15.4 | 0.11 |
| polarC18 | 780.55 | 401.1651 | 15.4 | 0.11 |
| polarC18 | 780.55 | 227.2935 | 15.4 | 0.11 |
| polarC18 | 780.55 | 358.0633 | 15.4 | 0.11 |
| polarC18 | 780.55 | 133.1917 | 15.4 | 0.11 |
| polarC18 | 780.55 | 193.8443 | 15.4 | 0.11 |
| polarC18 | 780.55 | 450.2379 | 15.4 | 0.11 |
| polarC18 | 780.55 | 495.6605 | 15.4 | 0.11 |
| polarC18 | 780.55 | 102.7388 | 15.4 | 0.11 |
| polarC18 | 780.55 | 135.6155 | 15.4 | 0.11 |
| polarC18 | 780.55 | 164.2125 | 15.4 | 0.11 |
| polarC18 | 780.55 | 119.2165 | 15.4 | 0.11 |
| polarC18 | 780.55 | 199.8071 | 15.4 | 0.11 |
| polarC18 | 780.55 | 164.6315 | 15.4 | 0.11 |
| polarC18 | 780.55 | 167.1542 | 15.4 | 0.11 |
| polarC18 | 780.55 | 116.224 | 15.4 | 0.11 |
| polarC18 | 780.55 | 117.4039 | 15.4 | 0.11 |
| polarC18 | 780.55 | 383.4192 | 15.4 | 0.11 |
| polarC18 | 780.55 | 94.9833 | 15.4 | 0.11 |
| polarC18 | 780.55 | 362.6941 | 15.4 | 0.11 |
| polarC18 | 780.55 | 225.7332 | 15.4 | 0.11 |
| polarC18 | 780.55 | 655.3151 | 15.4 | 0.11 |
| polarC18 | 780.55 | 232.9161 | 15.4 | 0.11 |
| polarC18 | 780.55 | 600.7402 | 15.4 | 0.11 |
| polarC18 | 780.55 | 99.2692 | 15.4 | 0.11 |
| polarC18 | 780.55 | 322.9137 | 15.4 | 0.11 |

| | | | | |
|---|---|---|---|---|
| polarC18 | 780.55 | 82.282 | 15.4 | 0.11 |
| polarC18 | 780.55 | 120.3051 | 15.4 | 0.11 |
| polarC18 | 780.55 | 217.721 | 15.4 | 0.11 |
| polarC18 | 780.55 | 102.76 | 15.4 | 0.11 |
| polarC18 | 780.55 | 310.8827 | 15.4 | 0.11 |
| polarC18 | 780.55 | 267.2809 | 15.4 | 0.11 |
| polarC18 | 780.55 | 187.7175 | 15.4 | 0.11 |
| polarC18 | 780.55 | 95.0455 | 15.4 | 0.11 |
| polarC18 | 780.55 | 554.9269 | 15.4 | 0.11 |
| polarC18 | 780.55 | 200.6859 | 15.4 | 0.11 |
| polarC18 | 780.55 | 86.3913 | 15.4 | 0.11 |
| polarC18 | 780.55 | 116.4058 | 15.4 | 0.11 |
| polarC18 | 780.55 | 500.8522 | 15.4 | 0.11 |
| polarC18 | 780.55 | 89.403 | 15.4 | 0.11 |
| polarC18 | 780.55 | 156.9214 | 15.4 | 0.11 |
| polarC18 | 780.55 | 115.7634 | 15.4 | 0.11 |
| polarC18 | 780.55 | 141.7512 | 15.4 | 0.11 |
| polarC18 | 780.55 | 97.7273 | 15.4 | 0.11 |
| polarC18 | 780.55 | 186.8679 | 15.4 | 0.11 |
| polarC18 | 780.55 | 231.0378 | 15.4 | 0.11 |
| polarC18 | 780.55 | 85.2705 | 15.4 | 0.11 |
| polarC18 | 780.55 | 135.4071 | 15.4 | 0.11 |
| polarC18 | 780.55 | 159.2696 | 15.4 | 0.11 |
| polarC18 | 780.55 | 87.0458 | 15.4 | 0.11 |
| polarC18 | 780.55 | 84.1085 | 15.4 | 0.11 |
| polarC18 | 780.55 | 134.8796 | 15.4 | 0.11 |
| polarC18 | 780.55 | 435.7427 | 15.4 | 0.11 |
| polarC18 | 780.55 | 231.5471 | 15.3 | 0.11 |
| polarC18 | 780.55 | 159.6591 | 15.3 | 0.11 |
| polarC18 | 780.55 | 206.6753 | 15.3 | 0.11 |
| polarC18 | 780.55 | 331.0546 | 15.3 | 0.11 |
| polarC18 | 780.55 | 284.0232 | 15.3 | 0.11 |
| polarC18 | 780.55 | 139.4571 | 15.3 | 0.11 |
| polarC18 | 780.55 | 118.3138 | 15.3 | 0.11 |
| polarC18 | 780.55 | 351.7322 | 15.3 | 0.11 |
| polarC18 | 780.55 | 162.222 | 15.3 | 0.11 |
| polarC18 | 780.55 | 231.947 | 15.3 | 0.11 |
| polarC18 | 780.55 | 360.6307 | 15.3 | 0.11 |
| polarC18 | 780.55 | 95.1667 | 15.3 | 0.11 |
| polarC18 | 780.55 | 282.4606 | 15.3 | 0.11 |
| polarC18 | 780.55 | 83.6657 | 15.3 | 0.11 |
| polarC18 | 780.55 | 164.3614 | 15.3 | 0.11 |

| | | | | |
|---|---|---|---|---|
| polarC18 | 780.55 | 149.5063 | 15.3 | 0.11 |
| polarC18 | 780.55 | 181.862 | 15.3 | 0.11 |
| polarC18 | 780.55 | 132.0278 | 15.3 | 0.11 |
| polarC18 | 780.55 | 141.4344 | 15.3 | 0.11 |
| polarC18 | 780.55 | 382.3754 | 15.3 | 0.11 |
| polarC18 | 780.55 | 154.4819 | 15.3 | 0.11 |
| polarC18 | 780.55 | 124.7954 | 15.3 | 0.11 |
| polarC18 | 780.55 | 333.2639 | 15.3 | 0.11 |
| polarC18 | 780.55 | 307.819 | 15.3 | 0.11 |
| polarC18 | 780.55 | 147.0794 | 15.3 | 0.11 |
| polarC18 | 780.55 | 225.3775 | 15.3 | 0.11 |
| polarC18 | 780.55 | 94.4363 | 15.3 | 0.11 |
| polarC18 | 780.55 | 154.0565 | 15.3 | 0.11 |
| polarC18 | 780.55 | 166.8163 | 15.3 | 0.11 |
| polarC18 | 780.55 | 91.6931 | 15.3 | 0.11 |
| polarC18 | 780.55 | 130.359 | 15.3 | 0.11 |
| polarC18 | 780.55 | 122.8281 | 15.3 | 0.11 |
| polarC18 | 780.55 | 128.5278 | 15.3 | 0.11 |
| polarC18 | 780.55 | 116.0897 | 15.3 | 0.11 |
| polarC18 | 780.55 | 307.5477 | 15.3 | 0.11 |
| polarC18 | 780.55 | 119.5543 | 15.3 | 0.11 |
| polarC18 | 780.55 | 302.2541 | 15.3 | 0.11 |
| polarC18 | 780.55 | 82.4449 | 15.3 | 0.11 |
| polarC18 | 780.55 | 227.6508 | 15.3 | 0.11 |
| polarC18 | 780.55 | 419.4569 | 15.3 | 0.11 |
| polarC18 | 780.55 | 151.6068 | 15.3 | 0.11 |
| polarC18 | 780.55 | 188.1608 | 15.3 | 0.11 |
| polarC18 | 780.55 | 94.7917 | 15.3 | 0.11 |
| polarC18 | 780.55 | 95.8978 | 15.3 | 0.11 |
| polarC18 | 780.55 | 200.0478 | 15.3 | 0.11 |
| polarC18 | 780.55 | 269.3591 | 15.3 | 0.11 |
| polarC18 | 780.55 | 86.694 | 15.3 | 0.11 |
| polarC18 | 780.55 | 256.0586 | 15.3 | 0.11 |
| polarC18 | 780.55 | 273.3475 | 15.3 | 0.11 |
| polarC18 | 780.55 | 161.5225 | 15.3 | 0.11 |
| polarC18 | 780.55 | 337.4653 | 15.3 | 0.11 |
| polarC18 | 780.55 | 461.4061 | 15.3 | 0.11 |
| polarC18 | 780.55 | 130.6057 | 15.3 | 0.11 |
| polarC18 | 780.55 | 159.5595 | 15.3 | 0.11 |
| polarC18 | 780.55 | 109.1452 | 15.3 | 0.11 |
| polarC18 | 780.55 | 230.3896 | 15.3 | 0.11 |
| polarC18 | 780.55 | 101.6797 | 15.3 | 0.11 |

| | | | | |
|---|---|---|---|---|
| polarC18 | 780.55 | 188.3068 | 15.3 | 0.11 |
| polarC18 | 780.55 | 231.6297 | 15.3 | 0.11 |
| polarC18 | 780.55 | 723.3481 | 15.3 | 0.11 |
| polarC18 | 780.55 | 731.0248 | 15.3 | 0.11 |
| polarC18 | 780.55 | 92.3095 | 15.3 | 0.11 |
| polarC18 | 780.55 | 151.1615 | 15.3 | 0.11 |
| polarC18 | 780.55 | 408.7254 | 15.3 | 0.11 |
| polarC18 | 780.55 | 122.9534 | 15.3 | 0.11 |
| polarC18 | 780.55 | 145.6824 | 15.3 | 0.11 |
| polarC18 | 780.55 | 135.4985 | 15.3 | 0.11 |
| polarC18 | 780.55 | 519.0718 | 15.3 | 0.11 |
| polarC18 | 780.55 | 121.6837 | 15.3 | 0.11 |
| polarC18 | 780.55 | 108.5851 | 15.3 | 0.11 |
| polarC18 | 780.55 | 140.321 | 15.3 | 0.11 |
| polarC18 | 780.55 | 360.0456 | 15.3 | 0.11 |
| polarC18 | 780.55 | 107.8502 | 15.3 | 0.11 |
| polarC18 | 780.55 | 204.3337 | 15.3 | 0.11 |
| polarC18 | 780.55 | 186.9241 | 15.3 | 0.11 |
| polarC18 | 780.55 | 312.5328 | 15.2 | 0.11 |
| polarC18 | 780.55 | 494.9955 | 15.2 | 0.11 |
| polarC18 | 780.55 | 84.3255 | 15.2 | 0.11 |
| polarC18 | 780.55 | 174.3883 | 15.2 | 0.11 |
| polarC18 | 780.55 | 159.3052 | 15.2 | 0.11 |
| polarC18 | 780.55 | 219.1253 | 15.2 | 0.11 |
| polarC18 | 780.55 | 84.2522 | 15.2 | 0.11 |
| polarC18 | 780.55 | 309.4235 | 15.2 | 0.11 |
| polarC18 | 780.55 | 434.7859 | 15.2 | 0.11 |
| polarC18 | 780.55 | 89.7143 | 15.2 | 0.11 |
| polarC18 | 780.55 | 347.8451 | 15.2 | 0.11 |
| polarC18 | 780.55 | 121.2065 | 15.2 | 0.11 |
| polarC18 | 780.55 | 150.8186 | 15.2 | 0.11 |
| polarC18 | 780.55 | 384.7933 | 15.2 | 0.11 |
| polarC18 | 780.55 | 119.3914 | 15.2 | 0.11 |
| polarC18 | 780.55 | 105.7445 | 15.2 | 0.11 |
| polarC18 | 780.55 | 147.6861 | 15.2 | 0.11 |
| polarC18 | 780.55 | 159.1747 | 15.2 | 0.11 |
| polarC18 | 780.55 | 119.3117 | 15.2 | 0.11 |
| polarC18 | 780.55 | 226.441 | 15.2 | 0.11 |
| polarC18 | 780.55 | 165.8899 | 15.2 | 0.11 |
| polarC18 | 780.55 | 223.1302 | 15.2 | 0.11 |
| polarC18 | 780.55 | 197.4816 | 15.2 | 0.11 |
| polarC18 | 780.55 | 256.1084 | 15.2 | 0.11 |

| | | | | |
|---|---|---|---|---|
| polarC18 | 780.55 | 194.1493 | 15.2 | 0.11 |
| polarC18 | 780.55 | 258.9563 | 15.2 | 0.11 |
| polarC18 | 780.55 | 320.5645 | 15.2 | 0.11 |
| polarC18 | 780.55 | 381.7491 | 15.2 | 0.11 |
| polarC18 | 780.55 | 411.1859 | 15.2 | 0.11 |
| polarC18 | 780.55 | 261.0301 | 15.2 | 0.11 |
| polarC18 | 780.55 | 152.3802 | 15.2 | 0.11 |
| polarC18 | 780.55 | 87.1355 | 15.2 | 0.11 |
| polarC18 | 780.55 | 725.0744 | 15.2 | 0.11 |
| polarC18 | 780.55 | 138.8404 | 15.2 | 0.11 |
| polarC18 | 780.55 | 211.5462 | 15.2 | 0.11 |
| polarC18 | 780.55 | 93.5962 | 15.2 | 0.11 |
| polarC18 | 780.55 | 178.6869 | 15.2 | 0.11 |
| polarC18 | 780.55 | 133.0774 | 15.2 | 0.11 |
| polarC18 | 780.55 | 233.3259 | 15.2 | 0.11 |
| polarC18 | 780.55 | 104.6734 | 15.2 | 0.11 |
| polarC18 | 780.55 | 97.0726 | 15.2 | 0.11 |
| polarC18 | 780.55 | 94.1038 | 15.2 | 0.11 |
| polarC18 | 780.55 | 84.9811 | 15.2 | 0.11 |
| polarC18 | 780.55 | 302.01 | 15.2 | 0.11 |
| polarC18 | 780.55 | 184.6673 | 15.2 | 0.11 |
| polarC18 | 780.55 | 293.4186 | 15.2 | 0.11 |
| polarC18 | 780.55 | 113.3764 | 15.2 | 0.11 |
| polarC18 | 780.55 | 199.2352 | 15.2 | 0.11 |
| polarC18 | 780.55 | 443.3424 | 15.2 | 0.11 |
| polarC18 | 780.55 | 139.6316 | 15.2 | 0.11 |
| polarC18 | 780.55 | 85.3026 | 15.2 | 0.11 |
| polarC18 | 780.55 | 106.8287 | 15.2 | 0.11 |
| polarC18 | 780.55 | 134.8232 | 15.2 | 0.11 |
| polarC18 | 780.55 | 102.4222 | 15.2 | 0.11 |
| polarC18 | 780.55 | 377.899 | 15.2 | 0.11 |
| polarC18 | 780.55 | 156.8218 | 15.2 | 0.11 |
| polarC18 | 780.55 | 295.4094 | 15.2 | 0.11 |
| polarC18 | 780.55 | 728.8892 | 15.2 | 0.11 |
| polarC18 | 780.55 | 128.5419 | 15.2 | 0.11 |
| polarC18 | 780.55 | 154.5111 | 15.2 | 0.11 |
| polarC18 | 780.55 | 88.4386 | 15.2 | 0.11 |
| polarC18 | 780.55 | 544.8983 | 15.2 | 0.11 |
| polarC18 | 780.55 | 197.1307 | 15.2 | 0.11 |
| polarC18 | 780.55 | 151.1387 | 15.2 | 0.11 |
| polarC18 | 780.55 | 238.2764 | 15.1 | 0.11 |
| polarC18 | 780.55 | 88.9067 | 15.1 | 0.11 |

| | | | | |
|---|---|---|---|---|
| polarC18 | 780.55 | 323.932 | 15.1 | 0.11 |
| polarC18 | 780.55 | 549.9073 | 15.1 | 0.11 |
| polarC18 | 780.55 | 144.5804 | 15.1 | 0.11 |
| polarC18 | 780.55 | 455.9088 | 15.1 | 0.11 |
| polarC18 | 780.55 | 146.4874 | 15.1 | 0.11 |
| polarC18 | 780.55 | 120.8772 | 15.1 | 0.11 |
| polarC18 | 780.55 | 105.3462 | 15.1 | 0.11 |
| polarC18 | 780.55 | 86.3545 | 15.1 | 0.11 |
| polarC18 | 780.55 | 231.2138 | 15.1 | 0.11 |
| polarC18 | 780.55 | 106.2266 | 15.1 | 0.11 |
| polarC18 | 780.55 | 86.943 | 15.1 | 0.11 |
| polarC18 | 780.55 | 395.0695 | 15.1 | 0.11 |
| polarC18 | 780.55 | 380.4686 | 15.1 | 0.11 |
| polarC18 | 780.55 | 148.2442 | 15.1 | 0.11 |
| polarC18 | 780.55 | 151.0767 | 15.1 | 0.11 |
| polarC18 | 780.55 | 160.8913 | 15.1 | 0.11 |
| polarC18 | 780.55 | 256.7401 | 15.1 | 0.11 |
| polarC18 | 780.55 | 342.454 | 15.1 | 0.11 |
| polarC18 | 780.55 | 204.5623 | 15.1 | 0.11 |
| polarC18 | 780.55 | 86.3342 | 15.1 | 0.11 |
| polarC18 | 780.55 | 143.4312 | 15.1 | 0.11 |
| polarC18 | 780.55 | 93.3974 | 15.1 | 0.11 |
| polarC18 | 780.55 | 99.4118 | 15.1 | 0.11 |
| polarC18 | 780.55 | 267.6265 | 15.1 | 0.11 |
| polarC18 | 780.55 | 256.0008 | 15.1 | 0.11 |
| polarC18 | 780.55 | 116.4885 | 15.1 | 0.11 |
| polarC18 | 780.55 | 85.7907 | 15.1 | 0.11 |
| polarC18 | 780.55 | 119.6013 | 15.1 | 0.11 |
| polarC18 | 780.55 | 107.7574 | 15.1 | 0.11 |
| polarC18 | 780.55 | 473.0052 | 15.1 | 0.11 |
| polarC18 | 780.55 | 264.7808 | 15.1 | 0.11 |
| polarC18 | 780.55 | 150.9696 | 15.1 | 0.11 |
| polarC18 | 780.55 | 106.5664 | 15.1 | 0.11 |
| polarC18 | 780.55 | 94.6947 | 15.1 | 0.11 |
| polarC18 | 780.55 | 215.0089 | 15.1 | 0.11 |
| polarC18 | 780.55 | 89.2409 | 15.1 | 0.11 |
| polarC18 | 780.55 | 111.9755 | 15.1 | 0.11 |
| polarC18 | 780.55 | 322.4201 | 15.1 | 0.11 |
| polarC18 | 780.55 | 203.9532 | 15.1 | 0.11 |
| polarC18 | 780.55 | 194.0044 | 15.1 | 0.11 |
| polarC18 | 780.55 | 465.5966 | 15.1 | 0.11 |
| polarC18 | 780.55 | 345.4005 | 15.1 | 0.11 |

| | | | | |
|---|---|---|---|---|
| polarC18 | 780.55 | 86.7187 | 15.1 | 0.11 |
| polarC18 | 780.55 | 130.5429 | 15.1 | 0.11 |
| polarC18 | 780.55 | 125.3901 | 15.1 | 0.11 |
| polarC18 | 780.55 | 124.888 | 15.1 | 0.11 |
| polarC18 | 780.55 | 345.5099 | 15.1 | 0.11 |
| polarC18 | 780.55 | 162.8372 | 15.1 | 0.11 |
| polarC18 | 780.55 | 365.4735 | 15.1 | 0.11 |
| polarC18 | 780.55 | 99.5119 | 15.1 | 0.11 |
| polarC18 | 780.55 | 122.8783 | 15.1 | 0.11 |
| polarC18 | 780.55 | 93.3563 | 15.1 | 0.11 |
| polarC18 | 780.55 | 733.9243 | 15.1 | 0.11 |
| polarC18 | 780.55 | 226.0883 | 15.1 | 0.11 |
| polarC18 | 780.55 | 88.3081 | 15.1 | 0.11 |
| polarC18 | 780.55 | 235.4504 | 15.1 | 0.11 |
| polarC18 | 780.55 | 146.8157 | 15.1 | 0.11 |
| polarC18 | 780.55 | 198.4335 | 15.1 | 0.11 |
| polarC18 | 780.55 | 413.4744 | 15.1 | 0.11 |
| polarC18 | 780.55 | 90.3956 | 15.1 | 0.11 |
| polarC18 | 780.55 | 124.0709 | 15.1 | 0.11 |
| polarC18 | 780.55 | 187.2439 | 15.1 | 0.11 |
| polarC18 | 780.55 | 184.7704 | 15.1 | 0.11 |
| polarC18 | 780.55 | 278.4344 | 15.1 | 0.11 |
| polarC18 | 780.55 | 122.1721 | 15.1 | 0.11 |
| polarC18 | 780.55 | 271.5652 | 15.1 | 0.11 |
| polarC18 | 780.55 | 192.8449 | 15.1 | 0.11 |
| polarC18 | 780.55 | 160.5927 | 15.1 | 0.11 |
| polarC18 | 780.55 | 271.6689 | 15.1 | 0.11 |
| polarC18 | 780.55 | 123.9898 | 15.1 | 0.11 |
| polarC18 | 780.55 | 297.8878 | 15.1 | 0.11 |
| polarC18 | 780.55 | 188.4111 | 15.1 | 0.11 |
| polarC18 | 780.55 | 130.0678 | 15.1 | 0.11 |
| polarC18 | 780.55 | 85.7739 | 15.1 | 0.11 |
| polarC18 | 780.55 | 134.5416 | 15.1 | 0.11 |
| polarC18 | 780.55 | 95.4742 | 15.1 | 0.11 |
| polarC18 | 780.55 | 207.6172 | 15.1 | 0.11 |
| polarC18 | 780.55 | 179.0427 | 15.1 | 0.11 |
| polarC18 | 780.55 | 110.7217 | 15.1 | 0.11 |
| polarC18 | 780.55 | 200.917 | 15 | 0.11 |
| polarC18 | 780.55 | 86.0522 | 15 | 0.11 |
| polarC18 | 780.55 | 308.3701 | 15 | 0.11 |
| polarC18 | 780.55 | 291.9282 | 15 | 0.11 |
| polarC18 | 780.55 | 86.5931 | 15 | 0.11 |

| | | | | |
|---|---|---|---|---|
| polarC18 | 780.55 | 87.7162 | 15 | 0.11 |
| polarC18 | 780.55 | 85.3501 | 15 | 0.11 |
| polarC18 | 780.55 | 95.5679 | 15 | 0.11 |
| polarC18 | 780.55 | 140.4793 | 15 | 0.11 |
| polarC18 | 780.55 | 116.3035 | 15 | 0.11 |
| polarC18 | 780.55 | 182.4611 | 15 | 0.11 |
| polarC18 | 780.55 | 539.4691 | 15 | 0.11 |
| polarC18 | 780.55 | 187.9129 | 15 | 0.11 |
| polarC18 | 780.55 | 289.8914 | 15 | 0.11 |
| polarC18 | 780.55 | 213.7758 | 15 | 0.11 |
| polarC18 | 780.55 | 102.7989 | 15 | 0.11 |
| polarC18 | 780.55 | 143.1242 | 15 | 0.11 |
| polarC18 | 780.55 | 256.9146 | 15 | 0.11 |
| polarC18 | 780.55 | 211.3847 | 15 | 0.11 |
| polarC18 | 780.55 | 85.9075 | 15 | 0.11 |
| polarC18 | 780.55 | 305.1196 | 15 | 0.11 |
| polarC18 | 780.55 | 153.7762 | 15 | 0.11 |
| polarC18 | 780.55 | 110.0976 | 15 | 0.11 |
| polarC18 | 780.55 | 215.6276 | 15 | 0.11 |
| polarC18 | 780.55 | 349.2082 | 15 | 0.11 |
| polarC18 | 780.55 | 511.6245 | 15 | 0.11 |
| polarC18 | 780.55 | 93.0999 | 15 | 0.11 |
| polarC18 | 780.55 | 189.481 | 15 | 0.11 |
| polarC18 | 780.55 | 84.3889 | 15 | 0.11 |
| polarC18 | 780.55 | 93.2826 | 15 | 0.11 |
| polarC18 | 780.55 | 84.0641 | 15 | 0.11 |
| polarC18 | 780.55 | 96.4349 | 15 | 0.11 |
| polarC18 | 780.55 | 736.9144 | 15 | 0.11 |
| polarC18 | 780.55 | 89.1458 | 15 | 0.11 |
| polarC18 | 780.55 | 158.0363 | 15 | 0.11 |
| polarC18 | 780.55 | 120.7705 | 15 | 0.11 |
| polarC18 | 780.55 | 97.7548 | 15 | 0.11 |
| polarC18 | 780.55 | 206.0783 | 15 | 0.11 |
| polarC18 | 780.55 | 228.8932 | 15 | 0.11 |
| polarC18 | 780.55 | 118.6851 | 15 | 0.11 |
| polarC18 | 780.55 | 86.7655 | 15 | 0.11 |
| polarC18 | 780.55 | 82.8378 | 15 | 0.11 |
| polarC18 | 780.55 | 135.1089 | 15 | 0.11 |
| polarC18 | 780.55 | 92.3177 | 15 | 0.11 |
| polarC18 | 780.55 | 199.0131 | 15 | 0.11 |
| polarC18 | 780.55 | 148.5196 | 15 | 0.11 |
| polarC18 | 780.55 | 112.7321 | 15 | 0.11 |

| | | | | |
|---|---|---|---|---|
| polarC18 | 780.55 | 419.1402 | 15 | 0.11 |
| polarC18 | 780.55 | 110.38 | 15 | 0.11 |
| polarC18 | 780.55 | 328.111 | 15 | 0.11 |
| polarC18 | 780.55 | 323.4375 | 15 | 0.11 |
| polarC18 | 780.55 | 605.5663 | 15 | 0.11 |
| polarC18 | 780.55 | 117.1022 | 15 | 0.11 |
| polarC18 | 780.55 | 389.8588 | 15 | 0.11 |
| polarC18 | 780.55 | 85.4209 | 15 | 0.11 |
| polarC18 | 780.55 | 621.9003 | 15 | 0.11 |
| polarC18 | 780.55 | 85.1282 | 15 | 0.11 |
| polarC18 | 780.55 | 193.6348 | 15 | 0.11 |
| polarC18 | 780.55 | 175.192 | 15 | 0.11 |
| polarC18 | 780.55 | 157.0202 | 15 | 0.11 |
| polarC18 | 780.55 | 109.2486 | 15 | 0.11 |
| polarC18 | 780.55 | 287.4831 | 15 | 0.11 |
| polarC18 | 780.55 | 147.589 | 15 | 0.11 |
| polarC18 | 780.55 | 201.795 | 15 | 0.11 |
| polarC18 | 780.55 | 165.4463 | 15 | 0.11 |
| polarC18 | 780.55 | 89.7856 | 15 | 0.11 |
| polarC18 | 780.55 | 117.3722 | 15 | 0.11 |
| polarC18 | 780.55 | 111.7073 | 15 | 0.11 |
| polarC18 | 780.55 | 207.7176 | 15 | 0.11 |
| polarC18 | 780.55 | 81.8653 | 15 | 0.11 |
| polarC18 | 780.55 | 184.2679 | 15 | 0.11 |
| polarC18 | 780.55 | 268.1015 | 15 | 0.11 |
| polarC18 | 780.55 | 89.3504 | 15 | 0.11 |
| polarC18 | 780.55 | 168.3081 | 15 | 0.11 |
| polarC18 | 780.55 | 133.4059 | 15 | 0.11 |
| polarC18 | 780.55 | 224.1143 | 15 | 0.11 |
| polarC18 | 780.55 | 113.4943 | 14.9 | 0.11 |
| polarC18 | 780.55 | 82.0044 | 14.9 | 0.11 |
| polarC18 | 780.55 | 186.2503 | 14.9 | 0.11 |
| polarC18 | 780.55 | 185.2471 | 14.9 | 0.11 |
| polarC18 | 780.55 | 127.6729 | 14.9 | 0.11 |
| polarC18 | 780.55 | 384.8733 | 14.9 | 0.11 |
| polarC18 | 780.55 | 107.8405 | 14.9 | 0.11 |
| polarC18 | 780.55 | 178.5119 | 14.9 | 0.11 |
| polarC18 | 780.55 | 388.9748 | 14.9 | 0.11 |
| polarC18 | 780.55 | 114.2806 | 14.9 | 0.11 |
| polarC18 | 780.55 | 153.3016 | 14.9 | 0.11 |
| polarC18 | 780.55 | 292.0076 | 14.9 | 0.11 |
| polarC18 | 780.55 | 641.6541 | 14.9 | 0.11 |

| | | | | |
|---|---|---|---|---|
| polarC18 | 780.55 | 241.5642 | 14.9 | 0.11 |
| polarC18 | 780.55 | 307.0368 | 14.9 | 0.11 |
| polarC18 | 780.55 | 110.5222 | 14.9 | 0.11 |
| polarC18 | 780.55 | 132.858 | 14.9 | 0.11 |
| polarC18 | 780.55 | 83.9378 | 14.9 | 0.11 |
| polarC18 | 780.55 | 83.0192 | 14.9 | 0.11 |
| polarC18 | 780.55 | 288.5238 | 14.9 | 0.11 |
| polarC18 | 780.55 | 109.165 | 14.9 | 0.11 |
| polarC18 | 780.55 | 167.5701 | 14.9 | 0.11 |
| polarC18 | 780.55 | 96.6148 | 14.9 | 0.11 |
| polarC18 | 780.55 | 158.985 | 14.9 | 0.11 |
| polarC18 | 780.55 | 162.0536 | 14.9 | 0.11 |
| polarC18 | 780.55 | 127.8843 | 14.9 | 0.11 |
| polarC18 | 780.55 | 86.3006 | 14.9 | 0.11 |
| polarC18 | 780.55 | 102.1402 | 14.9 | 0.11 |
| polarC18 | 780.55 | 211.4327 | 14.9 | 0.11 |
| polarC18 | 780.55 | 140.5947 | 14.9 | 0.11 |
| polarC18 | 780.55 | 118.287 | 14.9 | 0.11 |
| polarC18 | 780.55 | 551.8937 | 14.9 | 0.11 |
| polarC18 | 780.55 | 247.4702 | 14.9 | 0.11 |
| polarC18 | 780.55 | 287.8298 | 14.9 | 0.11 |
| polarC18 | 780.55 | 96.8345 | 14.9 | 0.11 |
| polarC18 | 780.55 | 457.1275 | 14.9 | 0.11 |
| polarC18 | 780.55 | 498.6675 | 14.9 | 0.11 |
| polarC18 | 780.55 | 93.5547 | 14.9 | 0.11 |
| polarC18 | 780.55 | 163.7635 | 14.9 | 0.11 |
| polarC18 | 780.55 | 163.8059 | 14.9 | 0.11 |
| polarC18 | 780.55 | 111.203 | 14.9 | 0.11 |
| polarC18 | 780.55 | 114.6755 | 14.9 | 0.11 |
| polarC18 | 780.55 | 343.9974 | 14.9 | 0.11 |
| polarC18 | 780.55 | 86.8897 | 14.9 | 0.11 |
| polarC18 | 780.55 | 264.5681 | 14.9 | 0.11 |
| polarC18 | 780.55 | 143.9622 | 14.9 | 0.11 |
| polarC18 | 780.55 | 416.7365 | 14.9 | 0.11 |
| polarC18 | 780.55 | 189.5815 | 14.9 | 0.11 |
| polarC18 | 780.55 | 86.2497 | 14.9 | 0.11 |
| polarC18 | 780.55 | 86.6589 | 14.9 | 0.11 |
| polarC18 | 780.55 | 325.8733 | 14.9 | 0.11 |
| polarC18 | 780.55 | 126.0822 | 14.9 | 0.11 |
| polarC18 | 780.55 | 283.7744 | 14.9 | 0.11 |
| polarC18 | 780.55 | 170.4902 | 14.9 | 0.11 |
| polarC18 | 780.55 | 100.4231 | 14.9 | 0.11 |

| | | | | |
|---|---|---|---|---|
| polarC18 | 780.55 | 307.307 | 14.9 | 0.11 |
| polarC18 | 780.55 | 122.5163 | 14.9 | 0.11 |
| polarC18 | 780.55 | 164.7495 | 14.9 | 0.11 |
| polarC18 | 780.55 | 136.5169 | 14.9 | 0.11 |
| polarC18 | 780.55 | 271.2045 | 14.8 | 0.11 |
| polarC18 | 780.55 | 97.0107 | 14.8 | 0.11 |
| polarC18 | 780.55 | 89.5544 | 14.8 | 0.11 |
| polarC18 | 780.55 | 431.0871 | 14.8 | 0.11 |
| polarC18 | 780.55 | 101.898 | 14.8 | 0.11 |
| polarC18 | 780.55 | 138.7322 | 14.8 | 0.11 |
| polarC18 | 780.55 | 83.6208 | 14.8 | 0.11 |
| polarC18 | 780.55 | 107.0718 | 14.8 | 0.11 |
| polarC18 | 780.55 | 365.9387 | 14.8 | 0.11 |
| polarC18 | 780.55 | 249.8908 | 14.8 | 0.11 |
| polarC18 | 780.55 | 172.1353 | 14.8 | 0.11 |
| polarC18 | 780.55 | 142.1977 | 14.8 | 0.11 |
| polarC18 | 780.55 | 231.155 | 14.8 | 0.11 |
| polarC18 | 780.55 | 173.4798 | 14.8 | 0.11 |
| polarC18 | 780.55 | 116.0732 | 14.8 | 0.11 |
| polarC18 | 780.55 | 115.9458 | 14.8 | 0.11 |
| polarC18 | 780.55 | 346.5142 | 14.8 | 0.11 |
| polarC18 | 780.55 | 82.688 | 14.8 | 0.11 |
| polarC18 | 780.55 | 85.9918 | 14.8 | 0.11 |
| polarC18 | 780.55 | 121.3845 | 14.8 | 0.11 |
| polarC18 | 780.55 | 150.2951 | 14.8 | 0.11 |
| polarC18 | 780.55 | 172.0613 | 14.8 | 0.11 |
| polarC18 | 780.55 | 199.8922 | 14.8 | 0.11 |
| polarC18 | 780.55 | 770.1937 | 14.8 | 0.11 |
| polarC18 | 780.55 | 122.2839 | 14.8 | 0.11 |
| polarC18 | 780.55 | 167.6305 | 14.8 | 0.11 |
| polarC18 | 780.55 | 118.0791 | 14.8 | 0.11 |
| polarC18 | 780.55 | 82.292 | 14.8 | 0.11 |
| polarC18 | 780.55 | 224.5698 | 14.8 | 0.11 |
| polarC18 | 780.55 | 86.1842 | 14.8 | 0.11 |
| polarC18 | 780.55 | 320.6408 | 14.8 | 0.11 |
| polarC18 | 780.55 | 186.602 | 14.8 | 0.11 |
| polarC18 | 780.55 | 124.343 | 14.8 | 0.11 |
| polarC18 | 780.55 | 324.192 | 14.8 | 0.11 |
| polarC18 | 780.55 | 105.8937 | 14.8 | 0.11 |
| polarC18 | 780.55 | 229.6438 | 14.8 | 0.11 |
| polarC18 | 780.55 | 116.1771 | 14.8 | 0.11 |
| polarC18 | 780.55 | 205.3519 | 14.8 | 0.11 |

| | | | | |
|---|---|---|---|---|
| polarC18 | 780.55 | 155.4448 | 14.8 | 0.11 |
| polarC18 | 780.55 | 105.6238 | 14.8 | 0.11 |
| polarC18 | 780.55 | 204.8755 | 14.8 | 0.11 |
| polarC18 | 780.55 | 122.0782 | 14.8 | 0.11 |
| polarC18 | 780.55 | 226.9286 | 14.8 | 0.11 |
| polarC18 | 780.55 | 151.2912 | 14.8 | 0.11 |
| polarC18 | 780.55 | 155.2231 | 14.8 | 0.11 |
| polarC18 | 780.55 | 300.5591 | 14.8 | 0.11 |
| polarC18 | 780.55 | 146.1894 | 14.8 | 0.11 |
| polarC18 | 780.55 | 117.1965 | 14.8 | 0.11 |
| polarC18 | 780.55 | 196.2852 | 14.8 | 0.11 |
| polarC18 | 780.55 | 156.5353 | 14.8 | 0.11 |
| polarC18 | 780.55 | 111.4781 | 14.8 | 0.11 |
| polarC18 | 780.55 | 161.3921 | 14.8 | 0.11 |
| polarC18 | 780.55 | 284.0815 | 14.8 | 0.11 |
| polarC18 | 780.55 | 100.1153 | 14.8 | 0.11 |
| polarC18 | 780.55 | 385.8914 | 14.8 | 0.11 |
| polarC18 | 780.55 | 120.1641 | 14.8 | 0.11 |
| polarC18 | 780.55 | 166.2161 | 14.8 | 0.11 |
| polarC18 | 780.55 | 200.5718 | 14.8 | 0.11 |
| polarC18 | 780.55 | 247.7194 | 14.7 | 0.11 |
| polarC18 | 780.55 | 115.9894 | 14.7 | 0.11 |
| polarC18 | 780.55 | 355.7161 | 14.7 | 0.11 |
| polarC18 | 780.55 | 399.6039 | 14.7 | 0.11 |
| polarC18 | 780.55 | 107.2731 | 14.7 | 0.11 |
| polarC18 | 780.55 | 87.9676 | 14.7 | 0.11 |
| polarC18 | 780.55 | 153.1171 | 14.7 | 0.11 |
| polarC18 | 780.55 | 107.9831 | 14.7 | 0.11 |
| polarC18 | 780.55 | 144.3404 | 14.7 | 0.11 |
| polarC18 | 780.55 | 90.1835 | 14.7 | 0.11 |
| polarC18 | 780.55 | 136.8968 | 14.7 | 0.11 |
| polarC18 | 780.55 | 135.5224 | 14.7 | 0.11 |
| polarC18 | 780.55 | 172.1731 | 14.7 | 0.11 |
| polarC18 | 780.55 | 268.9036 | 14.7 | 0.11 |
| polarC18 | 780.55 | 102.3912 | 14.7 | 0.11 |
| polarC18 | 780.55 | 208.6846 | 14.7 | 0.11 |
| polarC18 | 780.55 | 102.8654 | 14.7 | 0.11 |
| polarC18 | 780.55 | 119.1284 | 14.7 | 0.11 |
| polarC18 | 780.55 | 410.5925 | 14.7 | 0.11 |
| polarC18 | 780.55 | 114.5997 | 14.7 | 0.11 |
| polarC18 | 780.55 | 83.7671 | 14.7 | 0.11 |
| polarC18 | 780.55 | 112.61 | 14.7 | 0.11 |

| | | | | |
|---|---|---|---|---|
| polarC18 | 780.55 | 165.6824 | 14.7 | 0.11 |
| polarC18 | 780.55 | 101.3197 | 14.7 | 0.11 |
| polarC18 | 780.55 | 100.0309 | 14.7 | 0.11 |
| polarC18 | 780.55 | 202.4082 | 14.7 | 0.11 |
| polarC18 | 780.55 | 188.9121 | 14.7 | 0.11 |
| polarC18 | 780.55 | 215.9969 | 14.7 | 0.11 |
| polarC18 | 780.55 | 147.2675 | 14.7 | 0.11 |
| polarC18 | 780.55 | 85.1241 | 14.7 | 0.11 |
| polarC18 | 780.55 | 551.4139 | 14.7 | 0.11 |
| polarC18 | 780.55 | 143.6765 | 14.7 | 0.11 |
| polarC18 | 780.55 | 100.6426 | 14.7 | 0.11 |
| polarC18 | 780.55 | 118.8169 | 14.7 | 0.11 |
| polarC18 | 780.55 | 89.5451 | 14.7 | 0.11 |
| polarC18 | 780.55 | 175.2057 | 14.7 | 0.11 |
| polarC18 | 780.55 | 135.8163 | 14.7 | 0.11 |
| polarC18 | 780.55 | 264.3831 | 14.7 | 0.11 |
| polarC18 | 780.55 | 81.9116 | 14.7 | 0.11 |
| polarC18 | 780.55 | 163.3366 | 14.7 | 0.11 |
| polarC18 | 780.55 | 151.4317 | 14.7 | 0.11 |
| polarC18 | 780.55 | 106.3295 | 14.7 | 0.11 |
| polarC18 | 780.55 | 219.8471 | 14.7 | 0.11 |
| polarC18 | 780.55 | 209.3116 | 14.7 | 0.11 |
| polarC18 | 780.55 | 110.3924 | 14.7 | 0.11 |
| polarC18 | 780.55 | 156.2509 | 14.7 | 0.11 |
| polarC18 | 780.55 | 296.7778 | 14.7 | 0.11 |
| polarC18 | 780.55 | 106.8552 | 14.7 | 0.11 |
| polarC18 | 780.55 | 517.9493 | 14.7 | 0.11 |
| polarC18 | 780.55 | 120.9623 | 14.7 | 0.11 |
| polarC18 | 780.55 | 141.7122 | 14.7 | 0.11 |
| polarC18 | 780.55 | 97.75 | 14.7 | 0.11 |
| polarC18 | 780.55 | 151.6671 | 14.7 | 0.11 |
| polarC18 | 780.55 | 140.9077 | 14.7 | 0.11 |
| polarC18 | 780.55 | 129.4171 | 14.7 | 0.11 |
| polarC18 | 780.55 | 252.1404 | 14.7 | 0.11 |
| polarC18 | 780.55 | 210.3996 | 14.7 | 0.11 |
| polarC18 | 780.55 | 377.1689 | 14.7 | 0.11 |
| polarC18 | 780.55 | 101.8828 | 14.7 | 0.11 |
| polarC18 | 780.55 | 285.2319 | 14.7 | 0.11 |
| polarC18 | 780.55 | 263.3863 | 14.7 | 0.11 |
| polarC18 | 780.55 | 102.1068 | 14.7 | 0.11 |
| polarC18 | 780.55 | 106.0326 | 14.7 | 0.11 |
| polarC18 | 780.55 | 98.4334 | 14.6 | 0.11 |

| | | | | |
|---|---|---|---|---|
| polarC18 | 780.55 | 208.2996 | 14.6 | 0.11 |
| polarC18 | 780.55 | 347.6635 | 14.6 | 0.11 |
| polarC18 | 780.55 | 183.1854 | 14.6 | 0.11 |
| polarC18 | 780.55 | 185.7212 | 14.6 | 0.11 |
| polarC18 | 780.55 | 139.0464 | 14.6 | 0.11 |
| polarC18 | 780.55 | 179.8595 | 14.6 | 0.11 |
| polarC18 | 780.55 | 230.8946 | 14.6 | 0.11 |
| polarC18 | 780.55 | 132.9562 | 14.6 | 0.11 |
| polarC18 | 780.55 | 223.8006 | 14.6 | 0.11 |
| polarC18 | 780.55 | 393.0601 | 14.6 | 0.11 |
| polarC18 | 780.55 | 116.6804 | 14.6 | 0.11 |
| polarC18 | 780.55 | 206.7366 | 14.6 | 0.11 |
| polarC18 | 780.55 | 116.999 | 14.6 | 0.11 |
| polarC18 | 780.55 | 281.1145 | 14.6 | 0.11 |
| polarC18 | 780.55 | 115.1154 | 14.6 | 0.11 |
| polarC18 | 780.55 | 353.9076 | 14.6 | 0.11 |
| polarC18 | 780.55 | 165.549 | 14.6 | 0.11 |
| polarC18 | 780.55 | 127.4445 | 14.6 | 0.11 |
| polarC18 | 780.55 | 111.0934 | 14.6 | 0.11 |
| polarC18 | 780.55 | 141.6858 | 14.6 | 0.11 |
| polarC18 | 780.55 | 83.9284 | 14.6 | 0.11 |
| polarC18 | 780.55 | 269.7578 | 14.6 | 0.11 |
| polarC18 | 780.55 | 414.3766 | 14.6 | 0.11 |
| polarC18 | 780.55 | 179.9555 | 14.6 | 0.11 |
| polarC18 | 780.55 | 110.985 | 14.6 | 0.11 |
| polarC18 | 780.55 | 140.4467 | 14.6 | 0.11 |
| polarC18 | 780.55 | 129.381 | 14.6 | 0.11 |
| polarC18 | 780.55 | 131.1397 | 14.6 | 0.11 |
| polarC18 | 780.55 | 86.7849 | 14.6 | 0.11 |
| polarC18 | 780.55 | 222.4637 | 14.6 | 0.11 |
| polarC18 | 780.55 | 85.044 | 14.6 | 0.11 |
| polarC18 | 780.55 | 389.2901 | 14.6 | 0.11 |
| polarC18 | 780.55 | 117.244 | 14.6 | 0.11 |
| polarC18 | 780.55 | 221.5434 | 14.6 | 0.11 |
| polarC18 | 780.55 | 87.6787 | 14.6 | 0.11 |
| polarC18 | 780.55 | 92.0238 | 14.6 | 0.11 |
| polarC18 | 780.55 | 549.2734 | 14.6 | 0.11 |
| polarC18 | 780.55 | 104.3246 | 14.6 | 0.11 |
| polarC18 | 780.55 | 193.492 | 14.6 | 0.11 |
| polarC18 | 780.55 | 186.5443 | 14.6 | 0.11 |
| polarC18 | 780.55 | 153.3586 | 14.6 | 0.11 |
| polarC18 | 780.55 | 331.9459 | 14.6 | 0.11 |

| | | | | |
|---|---|---|---|---|
| polarC18 | 780.55 | 153.5503 | 14.6 | 0.11 |
| polarC18 | 780.55 | 112.339 | 14.6 | 0.11 |
| polarC18 | 780.55 | 510.111 | 14.6 | 0.11 |
| polarC18 | 780.55 | 136.5607 | 14.6 | 0.11 |
| polarC18 | 780.55 | 110.2101 | 14.6 | 0.11 |
| polarC18 | 780.55 | 132.5626 | 14.6 | 0.11 |
| polarC18 | 780.55 | 134.7635 | 14.6 | 0.11 |
| polarC18 | 780.55 | 224.046 | 14.6 | 0.11 |
| polarC18 | 780.55 | 151.775 | 14.6 | 0.11 |
| polarC18 | 780.55 | 218.0536 | 14.6 | 0.11 |
| polarC18 | 780.55 | 149.0138 | 14.6 | 0.11 |
| polarC18 | 780.55 | 136.1005 | 14.6 | 0.11 |
| polarC18 | 780.55 | 196.3605 | 14.6 | 0.11 |
| polarC18 | 780.55 | 176.3077 | 14.6 | 0.11 |
| polarC18 | 780.55 | 208.7352 | 14.6 | 0.11 |
| polarC18 | 780.55 | 84.6983 | 14.6 | 0.11 |
| polarC18 | 780.55 | 140.9342 | 14.6 | 0.11 |
| polarC18 | 780.55 | 123.0568 | 14.6 | 0.11 |
| polarC18 | 780.55 | 183.995 | 14.6 | 0.11 |
| polarC18 | 780.55 | 358.4294 | 14.6 | 0.11 |
| polarC18 | 780.55 | 259.5973 | 14.6 | 0.11 |
| polarC18 | 780.55 | 97.1703 | 14.5 | 0.11 |
| polarC18 | 780.55 | 540.0369 | 14.5 | 0.11 |
| polarC18 | 780.55 | 133.9014 | 14.5 | 0.11 |
| polarC18 | 780.55 | 143.4593 | 14.5 | 0.11 |
| polarC18 | 780.55 | 91.6728 | 14.5 | 0.11 |
| polarC18 | 780.55 | 508.1846 | 14.5 | 0.11 |
| polarC18 | 780.55 | 130.4448 | 14.5 | 0.11 |
| polarC18 | 780.55 | 170.0311 | 14.5 | 0.11 |
| polarC18 | 780.55 | 491.3824 | 14.5 | 0.11 |
| polarC18 | 780.55 | 224.5298 | 14.5 | 0.11 |
| polarC18 | 780.55 | 227.4043 | 14.5 | 0.11 |
| polarC18 | 780.55 | 293.0584 | 14.5 | 0.11 |
| polarC18 | 780.55 | 333.4017 | 14.5 | 0.11 |
| polarC18 | 780.55 | 82.4707 | 14.5 | 0.11 |
| polarC18 | 780.55 | 109.712 | 14.5 | 0.11 |
| polarC18 | 780.55 | 129.0734 | 14.5 | 0.11 |
| polarC18 | 780.55 | 89.1781 | 14.5 | 0.11 |
| polarC18 | 780.55 | 225.7095 | 14.5 | 0.11 |
| polarC18 | 780.55 | 345.5729 | 14.5 | 0.11 |
| polarC18 | 780.55 | 92.8709 | 14.5 | 0.11 |
| polarC18 | 780.55 | 91.3303 | 14.5 | 0.11 |

| | | | | |
|---|---|---|---|---|
| polarC18 | 780.55 | 366.8903 | 14.5 | 0.11 |
| polarC18 | 780.55 | 85.4626 | 14.5 | 0.11 |
| polarC18 | 780.55 | 107.6833 | 14.5 | 0.11 |
| polarC18 | 780.55 | 280.0861 | 14.5 | 0.11 |
| polarC18 | 780.55 | 118.917 | 14.5 | 0.11 |
| polarC18 | 780.55 | 110.7033 | 14.5 | 0.11 |
| polarC18 | 780.55 | 348.5602 | 14.5 | 0.11 |
| polarC18 | 780.55 | 138.603 | 14.5 | 0.11 |
| polarC18 | 780.55 | 99.6905 | 14.5 | 0.11 |
| polarC18 | 780.55 | 156.334 | 14.5 | 0.11 |
| polarC18 | 780.55 | 173.8659 | 14.5 | 0.11 |
| polarC18 | 780.55 | 117.502 | 14.5 | 0.11 |
| polarC18 | 780.55 | 139.9741 | 14.5 | 0.11 |
| polarC18 | 780.55 | 409.7081 | 14.5 | 0.11 |
| polarC18 | 780.55 | 761.7405 | 14.5 | 0.11 |
| polarC18 | 780.55 | 217.6558 | 14.5 | 0.11 |
| polarC18 | 780.55 | 274.9106 | 14.5 | 0.11 |
| polarC18 | 780.55 | 120.4456 | 14.5 | 0.11 |
| polarC18 | 780.55 | 490.648 | 14.5 | 0.11 |
| polarC18 | 780.55 | 173.0827 | 14.5 | 0.11 |
| polarC18 | 780.55 | 208.2137 | 14.5 | 0.11 |
| polarC18 | 780.55 | 121.314 | 14.5 | 0.11 |
| polarC18 | 780.55 | 84.8205 | 14.5 | 0.11 |
| polarC18 | 780.55 | 186.706 | 14.5 | 0.11 |
| polarC18 | 780.55 | 186.7858 | 14.5 | 0.11 |
| polarC18 | 780.55 | 286.0134 | 14.5 | 0.11 |
| polarC18 | 780.55 | 136.4862 | 14.5 | 0.11 |
| polarC18 | 780.55 | 120.7644 | 14.5 | 0.11 |
| polarC18 | 780.55 | 116.3455 | 14.4 | 0.11 |
| polarC18 | 780.55 | 110.4591 | 14.4 | 0.11 |
| polarC18 | 780.55 | 118.1894 | 14.4 | 0.11 |
| polarC18 | 780.55 | 517.4219 | 14.4 | 0.11 |
| polarC18 | 780.55 | 155.4337 | 14.4 | 0.11 |
| polarC18 | 780.55 | 90.3338 | 14.4 | 0.11 |
| polarC18 | 780.55 | 292.307 | 14.4 | 0.11 |
| polarC18 | 780.55 | 633.6786 | 14.4 | 0.11 |
| polarC18 | 780.55 | 157.0696 | 14.4 | 0.11 |
| polarC18 | 780.55 | 103.6245 | 14.4 | 0.11 |
| polarC18 | 780.55 | 141.85 | 14.4 | 0.11 |
| polarC18 | 780.55 | 384.2842 | 14.4 | 0.11 |
| polarC18 | 780.55 | 181.0817 | 14.4 | 0.11 |
| polarC18 | 780.55 | 94.3945 | 14.4 | 0.11 |

| | | | | |
|---|---|---|---|---|
| polarC18 | 780.55 | 83.1347 | 14.4 | 0.11 |
| polarC18 | 780.55 | 170.7873 | 14.4 | 0.11 |
| polarC18 | 780.55 | 104.0501 | 14.4 | 0.11 |
| polarC18 | 780.55 | 149.6839 | 14.4 | 0.11 |
| polarC18 | 780.55 | 87.1579 | 14.4 | 0.11 |
| polarC18 | 780.55 | 82.0739 | 14.4 | 0.11 |
| polarC18 | 780.55 | 454.279 | 14.4 | 0.11 |
| polarC18 | 780.55 | 162.6947 | 14.4 | 0.11 |
| polarC18 | 780.55 | 151.6164 | 14.4 | 0.11 |
| polarC18 | 780.55 | 189.4308 | 14.4 | 0.11 |
| polarC18 | 780.55 | 104.3053 | 14.4 | 0.11 |
| polarC18 | 780.55 | 134.9127 | 14.4 | 0.11 |
| polarC18 | 780.55 | 361.561 | 14.4 | 0.11 |
| polarC18 | 780.55 | 91.8164 | 14.4 | 0.11 |
| polarC18 | 780.55 | 128.0758 | 14.4 | 0.11 |
| polarC18 | 780.55 | 85.7384 | 14.4 | 0.11 |
| polarC18 | 780.55 | 260.0706 | 14.4 | 0.11 |
| polarC18 | 780.55 | 211.3068 | 14.4 | 0.11 |
| polarC18 | 780.55 | 110.16 | 14.4 | 0.11 |
| polarC18 | 780.55 | 123.3045 | 14.4 | 0.11 |
| polarC18 | 780.55 | 156.1618 | 14.4 | 0.11 |
| polarC18 | 780.55 | 416.2557 | 14.4 | 0.11 |
| polarC18 | 780.55 | 625.519 | 14.4 | 0.11 |
| polarC18 | 780.55 | 224.4653 | 14.4 | 0.11 |
| polarC18 | 780.55 | 181.3656 | 14.4 | 0.11 |
| polarC18 | 780.55 | 508.9733 | 14.4 | 0.11 |
| polarC18 | 780.55 | 252.3564 | 14.4 | 0.11 |
| polarC18 | 780.55 | 205.8899 | 14.4 | 0.11 |
| polarC18 | 780.55 | 345.8966 | 14.4 | 0.11 |
| polarC18 | 780.55 | 452.9904 | 14.4 | 0.11 |
| polarC18 | 780.55 | 94.4584 | 14.4 | 0.11 |
| polarC18 | 780.55 | 589.308 | 14.4 | 0.11 |
| polarC18 | 780.55 | 123.6618 | 14.4 | 0.11 |
| polarC18 | 780.55 | 158.2102 | 14.4 | 0.11 |
| polarC18 | 780.55 | 99.3603 | 14.4 | 0.11 |
| polarC18 | 780.55 | 217.9421 | 14.4 | 0.11 |
| polarC18 | 780.55 | 91.4109 | 14.4 | 0.11 |
| polarC18 | 780.55 | 163.2388 | 14.4 | 0.11 |
| polarC18 | 780.55 | 98.3212 | 14.4 | 0.11 |
| polarC18 | 780.55 | 110.1405 | 14.3 | 0.11 |
| polarC18 | 780.55 | 179.1238 | 14.3 | 0.11 |
| polarC18 | 780.55 | 107.3582 | 14.3 | 0.11 |

| | | | | |
|---|---|---|---|---|
| polarC18 | 780.55 | 86.8274 | 14.3 | 0.11 |
| polarC18 | 780.55 | 186.4296 | 14.3 | 0.11 |
| polarC18 | 780.55 | 89.4823 | 14.3 | 0.11 |
| polarC18 | 780.55 | 295.1539 | 14.3 | 0.11 |
| polarC18 | 780.55 | 112.0639 | 14.3 | 0.11 |
| polarC18 | 780.55 | 165.1251 | 14.3 | 0.11 |
| polarC18 | 780.55 | 124.3783 | 14.3 | 0.11 |
| polarC18 | 780.55 | 217.0674 | 14.3 | 0.1 |
| polarC18 | 780.55 | 182.3933 | 14.3 | 0.1 |
| polarC18 | 780.55 | 153.3128 | 14.3 | 0.1 |
| polarC18 | 780.55 | 175.1527 | 14.3 | 0.1 |
| polarC18 | 780.55 | 83.8406 | 14.3 | 0.1 |
| polarC18 | 780.55 | 103.7692 | 14.3 | 0.1 |
| polarC18 | 780.55 | 91.6327 | 14.3 | 0.1 |
| polarC18 | 780.55 | 92.5568 | 14.3 | 0.1 |
| polarC18 | 780.55 | 106.6515 | 14.3 | 0.1 |
| polarC18 | 780.55 | 141.4066 | 14.3 | 0.1 |
| polarC18 | 780.55 | 114.4907 | 14.3 | 0.1 |
| polarC18 | 780.55 | 122.3574 | 14.3 | 0.1 |
| polarC18 | 780.55 | 98.0743 | 14.3 | 0.1 |
| polarC18 | 780.55 | 266.3562 | 14.3 | 0.1 |
| polarC18 | 780.55 | 96.6071 | 14.3 | 0.1 |
| polarC18 | 780.55 | 227.918 | 14.3 | 0.1 |
| polarC18 | 780.55 | 116.2069 | 14.3 | 0.1 |
| polarC18 | 780.55 | 126.9159 | 14.3 | 0.1 |
| polarC18 | 780.55 | 170.6878 | 14.3 | 0.1 |
| polarC18 | 780.55 | 133.4204 | 14.3 | 0.1 |
| polarC18 | 780.55 | 354.0783 | 14.3 | 0.1 |
| polarC18 | 780.55 | 469.8109 | 14.3 | 0.1 |
| polarC18 | 780.55 | 191.1005 | 14.3 | 0.1 |
| polarC18 | 780.55 | 243.3366 | 14.3 | 0.1 |
| polarC18 | 780.55 | 124.7722 | 14.3 | 0.1 |
| polarC18 | 780.55 | 151.9613 | 14.3 | 0.1 |
| polarC18 | 780.55 | 172.4088 | 14.3 | 0.1 |
| polarC18 | 780.55 | 180.2224 | 14.3 | 0.1 |
| polarC18 | 780.55 | 103.1578 | 14.3 | 0.1 |
| polarC18 | 780.55 | 317.8463 | 14.3 | 0.1 |
| polarC18 | 780.55 | 150.2316 | 14.2 | 0.1 |
| polarC18 | 780.55 | 220.9419 | 14.2 | 0.1 |
| polarC18 | 780.55 | 353.8376 | 14.2 | 0.1 |
| polarC18 | 780.55 | 192.8217 | 14.2 | 0.1 |
| polarC18 | 780.55 | 310.4634 | 14.2 | 0.1 |

| | | | | |
|---|---|---|---|---|
| polarC18 | 780.55 | 512.8785 | 14.2 | 0.1 |
| polarC18 | 780.55 | 125.642 | 14.2 | 0.1 |
| polarC18 | 780.55 | 101.1681 | 14.2 | 0.1 |
| polarC18 | 780.55 | 126.9899 | 14.2 | 0.1 |
| polarC18 | 780.55 | 378.104 | 14.2 | 0.1 |
| polarC18 | 780.55 | 523.8031 | 14.2 | 0.1 |
| polarC18 | 780.55 | 125.8819 | 14.2 | 0.1 |
| polarC18 | 780.55 | 143.2635 | 14.2 | 0.1 |
| polarC18 | 780.55 | 236.6345 | 14.2 | 0.1 |
| polarC18 | 780.55 | 90.8369 | 14.2 | 0.1 |
| polarC18 | 780.55 | 147.0895 | 14.2 | 0.1 |
| polarC18 | 780.55 | 85.8595 | 14.2 | 0.1 |
| polarC18 | 780.55 | 122.0568 | 14.2 | 0.1 |
| polarC18 | 780.55 | 102.9356 | 14.2 | 0.1 |
| polarC18 | 780.55 | 211.6172 | 14.2 | 0.1 |
| polarC18 | 780.55 | 186.3545 | 14.2 | 0.1 |
| polarC18 | 780.55 | 101.0642 | 14.2 | 0.1 |
| polarC18 | 780.55 | 305.9862 | 14.2 | 0.1 |
| polarC18 | 780.55 | 199.6994 | 14.2 | 0.1 |
| polarC18 | 780.55 | 193.3336 | 14.2 | 0.1 |
| polarC18 | 780.55 | 539.1845 | 14.2 | 0.1 |
| polarC18 | 780.55 | 149.8031 | 14.2 | 0.1 |
| polarC18 | 780.55 | 124.2626 | 14.2 | 0.1 |
| polarC18 | 780.55 | 116.1158 | 14.2 | 0.1 |
| polarC18 | 780.55 | 148.3657 | 14.2 | 0.1 |
| polarC18 | 780.55 | 104.1919 | 14.2 | 0.1 |
| polarC18 | 780.55 | 95.1755 | 14.2 | 0.1 |
| polarC18 | 780.55 | 81.8442 | 14.1 | 0.1 |
| polarC18 | 780.55 | 82.5444 | 14.1 | 0.1 |
| polarC18 | 780.55 | 232.3282 | 14.1 | 0.1 |
| polarC18 | 780.55 | 125.0153 | 14.1 | 0.1 |
| polarC18 | 780.55 | 84.5966 | 14.1 | 0.1 |
| polarC18 | 780.55 | 88.7331 | 14.1 | 0.1 |
| polarC18 | 780.55 | 272.4803 | 14.1 | 0.1 |
| polarC18 | 780.55 | 510.9456 | 14.1 | 0.1 |
| polarC18 | 780.55 | 518.8439 | 14.1 | 0.1 |
| polarC18 | 780.55 | 199.6104 | 14.1 | 0.1 |
| polarC18 | 780.55 | 86.6142 | 14.1 | 0.1 |
| polarC18 | 780.55 | 95.6443 | 14.1 | 0.1 |
| polarC18 | 780.55 | 90.8327 | 14.1 | 0.1 |
| polarC18 | 780.55 | 109.857 | 14.1 | 0.1 |
| polarC18 | 780.55 | 86.6997 | 14.1 | 0.1 |

| | | | | |
|---|---|---|---|---|
| polarC18 | 780.55 | 419.2845 | 14.1 | 0.1 |
| polarC18 | 780.55 | 137.6231 | 14.1 | 0.1 |
| polarC18 | 780.55 | 515.8165 | 14.1 | 0.1 |
| polarC18 | 780.55 | 117.8129 | 14.1 | 0.1 |
| polarC18 | 780.55 | 255.8195 | 14.1 | 0.1 |
| polarC18 | 780.55 | 275.6458 | 14.1 | 0.1 |
| polarC18 | 780.55 | 168.4507 | 14.1 | 0.1 |
| polarC18 | 780.55 | 144.7429 | 14.1 | 0.1 |
| polarC18 | 780.55 | 85.0979 | 14.1 | 0.1 |
| polarC18 | 780.55 | 195.2572 | 14.1 | 0.1 |
| polarC18 | 780.55 | 190.1998 | 14.1 | 0.1 |
| polarC18 | 780.55 | 501.8607 | 14.1 | 0.1 |
| polarC18 | 780.55 | 217.5902 | 14.1 | 0.1 |
| polarC18 | 780.55 | 286.6602 | 14.1 | 0.1 |
| polarC18 | 780.55 | 82.0097 | 14.1 | 0.1 |
| polarC18 | 780.55 | 283.5284 | 14.1 | 0.1 |
| polarC18 | 780.55 | 88.27 | 14.1 | 0.1 |
| polarC18 | 780.55 | 160.3958 | 14.1 | 0.1 |
| polarC18 | 780.55 | 322.0023 | 14.1 | 0.1 |
| polarC18 | 780.55 | 175.3284 | 14.1 | 0.1 |
| polarC18 | 780.55 | 117.7654 | 14.1 | 0.1 |
| polarC18 | 780.55 | 230.1875 | 14.1 | 0.1 |
| polarC18 | 780.55 | 376.1964 | 14.1 | 0.1 |
| polarC18 | 780.55 | 110.7425 | 14.1 | 0.1 |
| polarC18 | 780.55 | 139.1255 | 14.1 | 0.1 |
| polarC18 | 780.55 | 94.4927 | 14 | 0.1 |
| polarC18 | 780.55 | 138.3138 | 14 | 0.1 |
| polarC18 | 780.55 | 90.9489 | 14 | 0.1 |
| polarC18 | 780.55 | 134.33 | 14 | 0.1 |
| polarC18 | 780.55 | 810.5251 | 14 | 0.1 |
| polarC18 | 780.55 | 141.2716 | 14 | 0.1 |
| polarC18 | 780.55 | 191.8452 | 14 | 0.1 |
| polarC18 | 780.55 | 164.9525 | 14 | 0.1 |
| polarC18 | 780.55 | 81.9963 | 14 | 0.1 |
| polarC18 | 780.55 | 172.8988 | 14 | 0.1 |
| polarC18 | 780.55 | 700.2328 | 14 | 0.1 |
| polarC18 | 780.55 | 132.2464 | 14 | 0.1 |
| polarC18 | 780.55 | 218.2977 | 14 | 0.1 |
| polarC18 | 780.55 | 88.8825 | 14 | 0.1 |
| polarC18 | 780.55 | 164.9666 | 14 | 0.1 |
| polarC18 | 780.55 | 153.9756 | 14 | 0.1 |
| polarC18 | 780.55 | 161.7718 | 14 | 0.1 |

| | | | | |
|---|---|---|---|---|
| polarC18 | 780.55 | 116.6987 | 14 | 0.1 |
| polarC18 | 780.55 | 98.0689 | 14 | 0.1 |
| polarC18 | 780.55 | 188.7254 | 14 | 0.1 |
| polarC18 | 780.55 | 207.1477 | 14 | 0.1 |
| polarC18 | 780.55 | 122.2237 | 14 | 0.1 |
| polarC18 | 780.55 | 214.3484 | 14 | 0.1 |
| polarC18 | 780.55 | 258.6263 | 14 | 0.1 |
| polarC18 | 780.55 | 114.0068 | 14 | 0.1 |
| polarC18 | 780.55 | 703.3105 | 14 | 0.1 |
| polarC18 | 780.55 | 111.8557 | 13.9 | 0.1 |
| polarC18 | 780.55 | 269.4858 | 13.9 | 0.1 |
| polarC18 | 780.55 | 144.7076 | 13.9 | 0.1 |
| polarC18 | 780.55 | 104.4726 | 13.9 | 0.1 |
| polarC18 | 780.55 | 82.1927 | 13.9 | 0.1 |
| polarC18 | 780.55 | 224.8535 | 13.9 | 0.1 |
| polarC18 | 780.55 | 121.4891 | 13.9 | 0.1 |
| polarC18 | 780.55 | 99.0413 | 13.9 | 0.1 |
| polarC18 | 780.55 | 100.4142 | 13.9 | 0.1 |
| polarC18 | 780.55 | 729.9703 | 13.9 | 0.1 |
| polarC18 | 780.55 | 102.5453 | 13.9 | 0.1 |
| polarC18 | 780.55 | 85.4656 | 13.9 | 0.1 |
| polarC18 | 780.55 | 277.8242 | 13.9 | 0.1 |
| polarC18 | 780.55 | 107.5976 | 13.9 | 0.1 |
| polarC18 | 780.55 | 85.6369 | 13.9 | 0.1 |
| polarC18 | 780.55 | 369.0484 | 13.9 | 0.1 |
| polarC18 | 780.55 | 207.9045 | 13.9 | 0.1 |
| polarC18 | 780.55 | 127.2525 | 13.9 | 0.1 |
| polarC18 | 780.55 | 173.6488 | 13.9 | 0.1 |
| polarC18 | 780.55 | 184.6411 | 13.9 | 0.1 |
| polarC18 | 780.55 | 417.3527 | 13.9 | 0.1 |
| polarC18 | 780.55 | 790.3814 | 13.9 | 0.1 |
| polarC18 | 780.55 | 141.1166 | 13.9 | 0.1 |
| polarC18 | 780.55 | 585.545 | 13.9 | 0.1 |
| polarC18 | 780.55 | 129.1843 | 13.9 | 0.1 |
| polarC18 | 780.55 | 223.0137 | 13.9 | 0.1 |
| polarC18 | 780.55 | 223.84 | 13.9 | 0.1 |
| polarC18 | 780.55 | 513.909 | 13.9 | 0.1 |
| polarC18 | 780.55 | 208.9843 | 13.8 | 0.1 |
| polarC18 | 780.55 | 111.3798 | 13.8 | 0.1 |
| polarC18 | 780.55 | 128.5775 | 13.8 | 0.1 |
| polarC18 | 780.55 | 155.8104 | 13.8 | 0.1 |
| polarC18 | 780.55 | 121.0641 | 13.8 | 0.1 |

| | | | | |
|---|---|---|---|---|
| polarC18 | 780.55 | 95.9663 | 13.8 | 0.1 |
| polarC18 | 780.55 | 140.1882 | 13.8 | 0.1 |
| polarC18 | 780.55 | 113.2872 | 13.8 | 0.1 |
| polarC18 | 780.55 | 134.6091 | 13.8 | 0.1 |
| polarC18 | 780.55 | 447.8827 | 13.8 | 0.1 |
| polarC18 | 780.55 | 208.7962 | 13.8 | 0.1 |
| polarC18 | 780.55 | 88.0694 | 13.8 | 0.1 |
| polarC18 | 780.55 | 108.7765 | 13.8 | 0.1 |
| polarC18 | 780.55 | 87.903 | 13.8 | 0.1 |
| polarC18 | 780.55 | 240.9581 | 13.8 | 0.1 |
| polarC18 | 780.55 | 207.398 | 13.8 | 0.1 |
| polarC18 | 780.55 | 149.9534 | 13.8 | 0.1 |
| polarC18 | 780.55 | 178.0851 | 13.8 | 0.1 |
| polarC18 | 780.55 | 172.1128 | 13.8 | 0.1 |
| polarC18 | 780.55 | 286.4268 | 13.8 | 0.1 |
| polarC18 | 780.55 | 407.1437 | 13.8 | 0.1 |
| polarC18 | 780.55 | 499.1859 | 13.8 | 0.1 |
| polarC18 | 780.55 | 81.8587 | 13.8 | 0.1 |
| polarC18 | 780.55 | 167.3225 | 13.8 | 0.1 |
| polarC18 | 780.55 | 497.0955 | 13.7 | 0.1 |
| polarC18 | 780.55 | 131.39 | 13.7 | 0.1 |
| polarC18 | 780.55 | 322.2796 | 13.7 | 0.1 |
| polarC18 | 780.55 | 131.2355 | 13.7 | 0.1 |
| polarC18 | 780.55 | 219.1485 | 13.7 | 0.1 |
| polarC18 | 780.55 | 750.9537 | 13.7 | 0.1 |
| polarC18 | 780.55 | 116.733 | 13.7 | 0.1 |
| polarC18 | 780.55 | 144.3833 | 13.7 | 0.1 |
| polarC18 | 780.55 | 83.4695 | 13.7 | 0.1 |
| polarC18 | 780.55 | 87.2953 | 13.7 | 0.1 |
| polarC18 | 780.55 | 337.6815 | 13.7 | 0.1 |
| polarC18 | 780.55 | 92.1837 | 13.7 | 0.1 |
| polarC18 | 780.55 | 107.125 | 13.7 | 0.1 |
| polarC18 | 780.55 | 101.7498 | 13.7 | 0.1 |
| polarC18 | 780.55 | 128.3459 | 13.7 | 0.1 |
| polarC18 | 780.55 | 152.7584 | 13.7 | 0.1 |
| polarC18 | 780.55 | 100.8952 | 13.7 | 0.1 |
| polarC18 | 780.55 | 118.2763 | 13.7 | 0.1 |
| polarC18 | 780.55 | 215.5809 | 13.7 | 0.1 |
| polarC18 | 780.55 | 186.2176 | 13.6 | 0.1 |
| polarC18 | 780.55 | 91.5929 | 13.6 | 0.1 |
| polarC18 | 780.55 | 219.4055 | 13.6 | 0.1 |
| polarC18 | 780.55 | 86.1009 | 13.6 | 0.1 |

| | | | | |
|---|---|---|---|---|
| polarC18 | 780.55 | 102.9497 | 13.6 | 0.1 |
| polarC18 | 780.55 | 199.0435 | 13.6 | 0.1 |
| polarC18 | 780.55 | 131.4868 | 13.6 | 0.1 |
| polarC18 | 780.55 | 290.2133 | 13.6 | 0.1 |
| polarC18 | 780.55 | 198.5319 | 13.6 | 0.1 |
| polarC18 | 780.55 | 111.2415 | 13.6 | 0.1 |
| polarC18 | 780.55 | 297.6198 | 13.6 | 0.1 |
| polarC18 | 780.55 | 171.1698 | 13.6 | 0.1 |
| polarC18 | 780.55 | 772.8374 | 13.6 | 0.1 |
| polarC18 | 780.55 | 168.1673 | 13.6 | 0.1 |
| polarC18 | 780.55 | 160.0223 | 13.6 | 0.1 |
| polarC18 | 780.55 | 101.4711 | 13.6 | 0.1 |
| polarC18 | 780.55 | 90.7447 | 13.6 | 0.1 |
| polarC18 | 780.55 | 84.7599 | 13.6 | 0.1 |
| polarC18 | 780.55 | 98.6678 | 13.6 | 0.1 |
| polarC18 | 780.55 | 85.111 | 13.6 | 0.1 |
| polarC18 | 780.55 | 146.6073 | 13.6 | 0.1 |
| polarC18 | 780.55 | 246.5307 | 13.6 | 0.1 |
| polarC18 | 780.55 | 82.7513 | 13.6 | 0.1 |
| polarC18 | 780.55 | 218.0918 | 13.6 | 0.1 |
| polarC18 | 780.55 | 417.7492 | 13.6 | 0.1 |
| polarC18 | 780.55 | 213.9991 | 13.6 | 0.1 |
| polarC18 | 780.55 | 97.1861 | 13.5 | 0.1 |
| polarC18 | 780.55 | 416.0096 | 13.5 | 0.1 |
| polarC18 | 780.55 | 122.4415 | 13.5 | 0.1 |
| polarC18 | 780.55 | 116.6095 | 13.5 | 0.1 |
| polarC18 | 780.55 | 86.9822 | 13.5 | 0.1 |
| polarC18 | 780.55 | 261.3836 | 13.5 | 0.1 |
| polarC18 | 780.55 | 91.683 | 13.5 | 0.1 |
| polarC18 | 780.55 | 120.3203 | 13.5 | 0.1 |
| polarC18 | 780.55 | 229.5639 | 13.5 | 0.1 |
| polarC18 | 780.55 | 255.5624 | 13.5 | 0.1 |
| polarC18 | 780.55 | 154.1105 | 13.5 | 0.1 |
| polarC18 | 780.55 | 246.399 | 13.5 | 0.1 |
| polarC18 | 780.55 | 432.3474 | 13.5 | 0.1 |
| polarC18 | 780.55 | 729.7122 | 13.5 | 0.1 |
| polarC18 | 780.55 | 120.6155 | 13.5 | 0.1 |
| polarC18 | 780.55 | 104.1759 | 13.5 | 0.1 |
| polarC18 | 780.55 | 550.5565 | 13.5 | 0.1 |
| polarC18 | 780.55 | 84.1022 | 13.5 | 0.1 |
| polarC18 | 780.55 | 545.1757 | 13.5 | 0.1 |
| polarC18 | 780.55 | 253.1814 | 13.4 | 0.1 |

| | | | | |
|---|---|---|---|---|
| polarC18 | 780.55 | 88.6757 | 13.4 | 0.1 |
| polarC18 | 780.55 | 105.2818 | 13.4 | 0.1 |
| polarC18 | 780.55 | 603.627 | 13.4 | 0.1 |
| polarC18 | 780.55 | 171.3854 | 13.4 | 0.1 |
| polarC18 | 780.55 | 109.2693 | 13.4 | 0.1 |
| polarC18 | 780.55 | 92.0883 | 13.4 | 0.1 |
| polarC18 | 780.55 | 525.9365 | 13.4 | 0.1 |
| polarC18 | 780.55 | 97.424 | 13.4 | 0.1 |
| polarC18 | 780.55 | 270.1607 | 13.4 | 0.1 |
| polarC18 | 780.55 | 146.4746 | 13.4 | 0.1 |
| polarC18 | 780.55 | 658.0623 | 13.4 | 0.1 |
| polarC18 | 780.55 | 692.2527 | 13.4 | 0.1 |
| polarC18 | 780.55 | 217.1444 | 13.4 | 0.1 |
| polarC18 | 780.55 | 410.3307 | 13.4 | 0.1 |
| polarC18 | 780.55 | 278.5778 | 13.4 | 0.1 |
| polarC18 | 780.55 | 221.4453 | 13.4 | 0.1 |
| polarC18 | 780.55 | 203.2181 | 13.4 | 0.1 |
| polarC18 | 780.55 | 500.4391 | 13.4 | 0.1 |
| polarC18 | 780.55 | 413.3931 | 13.3 | 0.1 |
| polarC18 | 780.55 | 189.0456 | 13.3 | 0.1 |
| polarC18 | 780.55 | 562.3553 | 13.3 | 0.1 |
| polarC18 | 780.55 | 675.8178 | 13.3 | 0.1 |
| polarC18 | 780.55 | 125.691 | 13.3 | 0.1 |
| polarC18 | 780.55 | 105.1831 | 13.3 | 0.1 |
| polarC18 | 780.55 | 432.1654 | 13.3 | 0.1 |
| polarC18 | 780.55 | 140.6873 | 13.3 | 0.1 |
| polarC18 | 780.55 | 142.6744 | 13.3 | 0.1 |
| polarC18 | 780.55 | 99.1007 | 13.3 | 0.1 |
| polarC18 | 780.55 | 497.6817 | 13.3 | 0.1 |
| polarC18 | 780.55 | 393.9557 | 13.3 | 0.1 |
| polarC18 | 780.55 | 141.1423 | 13.3 | 0.1 |
| polarC18 | 780.55 | 179.4786 | 13.3 | 0.1 |
| polarC18 | 780.55 | 164.5082 | 13.2 | 0.1 |
| polarC18 | 780.55 | 120.9878 | 13.2 | 0.1 |
| polarC18 | 780.55 | 683.0871 | 13.2 | 0.1 |
| polarC18 | 780.55 | 203.3066 | 13.2 | 0.1 |
| polarC18 | 780.55 | 147.7659 | 13.2 | 0.1 |
| polarC18 | 780.55 | 119.5234 | 13.2 | 0.1 |
| polarC18 | 780.55 | 117.4863 | 13.2 | 0.1 |
| polarC18 | 780.55 | 231.6924 | 13.2 | 0.1 |
| polarC18 | 780.55 | 198.6188 | 13.2 | 0.1 |
| polarC18 | 780.55 | 121.1485 | 13.2 | 0.1 |

| | | | | |
|---|---|---|---|---|
| polarC18 | 780.55 | 436.345 | 13.1 | 0.1 |
| polarC18 | 780.55 | 470.7625 | 13.1 | 0.1 |
| polarC18 | 780.55 | 145.3536 | 13.1 | 0.1 |
| polarC18 | 780.55 | 191.4024 | 13.1 | 0.1 |
| polarC18 | 780.55 | 324.243 | 13.1 | 0.1 |
| polarC18 | 780.55 | 251.5558 | 13.1 | 0.1 |
| polarC18 | 780.55 | 125.3071 | 13.1 | 0.1 |
| polarC18 | 780.55 | 103.7947 | 13.1 | 0.1 |
| polarC18 | 780.55 | 291.9704 | 13.1 | 0.1 |
| polarC18 | 780.55 | 250.1563 | 13.1 | 0.1 |
| polarC18 | 780.55 | 569.2443 | 13.1 | 0.1 |
| polarC18 | 780.55 | 282.6537 | 13.1 | 0.1 |
| polarC18 | 780.55 | 192.2776 | 13.1 | 0.1 |
| polarC18 | 780.55 | 392.5823 | 13.1 | 0.1 |
| polarC18 | 780.55 | 292.877 | 13.1 | 0.1 |
| polarC18 | 780.55 | 171.8291 | 13.1 | 0.1 |
| polarC18 | 780.55 | 270.8991 | 13.1 | 0.1 |
| polarC18 | 780.55 | 143.2472 | 13 | 0.1 |
| polarC18 | 780.55 | 259.4778 | 13 | 0.1 |
| polarC18 | 780.55 | 293.2383 | 13 | 0.1 |
| polarC18 | 780.55 | 196.9029 | 13 | 0.1 |
| polarC18 | 780.55 | 628.9669 | 13 | 0.1 |
| polarC18 | 780.55 | 281.2106 | 13 | 0.1 |
| polarC18 | 780.55 | 538.0629 | 13 | 0.1 |
| polarC18 | 780.55 | 93.9255 | 13 | 0.1 |
| polarC18 | 780.55 | 203.1291 | 13 | 0.1 |
| polarC18 | 780.55 | 93.0872 | 13 | 0.1 |
| polarC18 | 780.55 | 155.3466 | 13 | 0.1 |
| polarC18 | 780.55 | 240.6857 | 13 | 0.09 |
| polarC18 | 780.55 | 95.5422 | 13 | 0.09 |
| polarC18 | 780.55 | 82.9877 | 13 | 0.09 |
| polarC18 | 780.55 | 165.0389 | 12.9 | 0.09 |
| polarC18 | 780.55 | 459.6977 | 12.9 | 0.09 |
| polarC18 | 780.55 | 191.9092 | 12.9 | 0.09 |
| polarC18 | 780.55 | 185.8552 | 12.9 | 0.09 |
| polarC18 | 780.55 | 552.5667 | 12.9 | 0.09 |
| polarC18 | 780.55 | 212.2166 | 12.9 | 0.09 |
| polarC18 | 780.55 | 710.258 | 12.9 | 0.09 |
| polarC18 | 780.55 | 138.2883 | 12.9 | 0.09 |
| polarC18 | 780.55 | 94.327 | 12.9 | 0.09 |
| polarC18 | 780.55 | 150.6963 | 12.9 | 0.09 |
| polarC18 | 780.55 | 175.1285 | 12.9 | 0.09 |

| | | | | |
|---|---|---|---|---|
| polarC18 | 780.55 | 171.3596 | 12.9 | 0.09 |
| polarC18 | 780.55 | 93.5599 | 12.9 | 0.09 |
| polarC18 | 780.55 | 170.6177 | 12.9 | 0.09 |
| polarC18 | 780.55 | 153.3473 | 12.9 | 0.09 |
| polarC18 | 780.55 | 145.1243 | 12.9 | 0.09 |
| polarC18 | 780.55 | 101.4763 | 12.9 | 0.09 |
| polarC18 | 780.55 | 83.5178 | 12.9 | 0.09 |
| polarC18 | 780.55 | 106.706 | 12.9 | 0.09 |
| polarC18 | 780.55 | 107.5886 | 12.9 | 0.09 |
| polarC18 | 780.55 | 300.7271 | 12.8 | 0.09 |
| polarC18 | 780.55 | 95.6165 | 12.8 | 0.09 |
| polarC18 | 780.55 | 748.2351 | 12.8 | 0.09 |
| polarC18 | 780.55 | 303.4961 | 12.8 | 0.09 |
| polarC18 | 780.55 | 216.721 | 12.8 | 0.09 |
| polarC18 | 780.55 | 183.6485 | 12.8 | 0.09 |
| polarC18 | 780.55 | 146.6552 | 12.8 | 0.09 |
| polarC18 | 780.55 | 84.9362 | 12.8 | 0.09 |
| polarC18 | 780.55 | 105.3117 | 12.8 | 0.09 |
| polarC18 | 780.55 | 306.5979 | 12.8 | 0.09 |
| polarC18 | 780.55 | 98.8161 | 12.8 | 0.09 |
| polarC18 | 780.55 | 425.2356 | 12.8 | 0.09 |
| polarC18 | 780.55 | 85.215 | 12.8 | 0.09 |
| polarC18 | 780.55 | 119.5767 | 12.8 | 0.09 |
| polarC18 | 780.55 | 90.0229 | 12.8 | 0.09 |
| polarC18 | 780.55 | 217.6127 | 12.8 | 0.09 |
| polarC18 | 780.55 | 162.6673 | 12.8 | 0.09 |
| polarC18 | 780.55 | 96.3895 | 12.7 | 0.09 |
| polarC18 | 780.55 | 179.2711 | 12.7 | 0.09 |
| polarC18 | 780.55 | 129.6984 | 12.7 | 0.09 |
| polarC18 | 780.55 | 272.1573 | 12.7 | 0.09 |
| polarC18 | 780.55 | 96.5966 | 12.7 | 0.09 |
| polarC18 | 780.55 | 282.4935 | 12.7 | 0.09 |
| polarC18 | 780.55 | 86.4423 | 12.7 | 0.09 |
| polarC18 | 780.55 | 305.9467 | 12.7 | 0.09 |
| polarC18 | 780.55 | 419.8541 | 12.7 | 0.09 |
| polarC18 | 780.55 | 721.09 | 12.7 | 0.09 |
| polarC18 | 780.55 | 93.8914 | 12.7 | 0.09 |
| polarC18 | 780.55 | 147.8047 | 12.6 | 0.09 |
| polarC18 | 780.55 | 105.2106 | 12.6 | 0.09 |
| polarC18 | 780.55 | 91.4175 | 12.6 | 0.09 |
| polarC18 | 780.55 | 212.6823 | 12.6 | 0.09 |
| polarC18 | 780.55 | 186.0613 | 12.6 | 0.09 |

| | | | | |
|---|---|---|---|---|
| polarC18 | 780.55 | 171.3217 | 12.6 | 0.09 |
| polarC18 | 780.55 | 424.7869 | 12.6 | 0.09 |
| polarC18 | 780.55 | 189.2766 | 12.6 | 0.09 |
| polarC18 | 780.55 | 138.0213 | 12.6 | 0.09 |
| polarC18 | 780.55 | 187.0429 | 12.6 | 0.09 |
| polarC18 | 780.55 | 87.3635 | 12.6 | 0.09 |
| polarC18 | 780.55 | 223.1638 | 12.6 | 0.09 |
| polarC18 | 780.55 | 90.5264 | 12.6 | 0.09 |
| polarC18 | 780.55 | 312.2279 | 12.6 | 0.09 |
| polarC18 | 780.55 | 89.4412 | 12.6 | 0.09 |
| polarC18 | 780.55 | 193.9488 | 12.6 | 0.09 |
| polarC18 | 780.55 | 137.5687 | 12.6 | 0.09 |
| polarC18 | 780.55 | 83.4927 | 12.6 | 0.09 |
| polarC18 | 780.55 | 230.4271 | 12.6 | 0.09 |
| polarC18 | 780.55 | 402.8102 | 12.6 | 0.09 |
| polarC18 | 780.55 | 267.7633 | 12.5 | 0.09 |
| polarC18 | 780.55 | 239.0873 | 12.5 | 0.09 |
| polarC18 | 780.55 | 441.3082 | 12.5 | 0.09 |
| polarC18 | 780.55 | 520.8901 | 12.5 | 0.09 |
| polarC18 | 780.55 | 186.8895 | 12.5 | 0.09 |
| polarC18 | 780.55 | 102.6584 | 12.5 | 0.09 |
| polarC18 | 780.55 | 88.134 | 12.5 | 0.09 |
| polarC18 | 780.55 | 152.0372 | 12.5 | 0.09 |
| polarC18 | 780.55 | 102.4383 | 12.5 | 0.09 |
| polarC18 | 780.55 | 145.623 | 12.5 | 0.09 |
| polarC18 | 780.55 | 88.9317 | 12.5 | 0.09 |
| polarC18 | 780.55 | 132.4218 | 12.5 | 0.09 |
| polarC18 | 780.55 | 260.7975 | 12.5 | 0.09 |
| polarC18 | 780.55 | 659.6199 | 12.5 | 0.09 |
| polarC18 | 780.55 | 141.0676 | 12.5 | 0.09 |
| polarC18 | 780.55 | 83.5603 | 12.5 | 0.09 |
| polarC18 | 780.55 | 99.8376 | 12.5 | 0.09 |
| polarC18 | 780.55 | 94.0258 | 12.5 | 0.09 |
| polarC18 | 780.55 | 114.2431 | 12.5 | 0.09 |
| polarC18 | 780.55 | 201.2736 | 12.5 | 0.09 |
| polarC18 | 780.55 | 466.7195 | 12.5 | 0.09 |
| polarC18 | 780.55 | 154.0374 | 12.5 | 0.09 |
| polarC18 | 780.55 | 368.0775 | 12.5 | 0.09 |
| polarC18 | 780.55 | 188.6289 | 12.5 | 0.09 |
| polarC18 | 780.55 | 384.1862 | 12.5 | 0.09 |
| polarC18 | 780.55 | 136.1497 | 12.5 | 0.09 |
| polarC18 | 780.55 | 274.0682 | 12.5 | 0.09 |

| | | | | |
|---|---|---|---|---|
| polarC18 | 780.55 | 120.8201 | 12.5 | 0.09 |
| polarC18 | 780.55 | 262.0764 | 12.4 | 0.09 |
| polarC18 | 780.55 | 194.1882 | 12.4 | 0.09 |
| polarC18 | 780.55 | 125.0248 | 12.4 | 0.09 |
| polarC18 | 780.55 | 611.6288 | 12.4 | 0.09 |
| polarC18 | 780.55 | 382.5449 | 12.4 | 0.09 |
| polarC18 | 780.55 | 136.6834 | 12.4 | 0.09 |
| polarC18 | 780.55 | 579.9144 | 12.4 | 0.09 |
| polarC18 | 780.55 | 93.1711 | 12.4 | 0.09 |
| polarC18 | 780.55 | 88.3865 | 12.4 | 0.09 |
| polarC18 | 780.55 | 681.3979 | 12.4 | 0.09 |
| polarC18 | 780.55 | 92.7744 | 12.4 | 0.09 |
| polarC18 | 780.55 | 267.0464 | 12.4 | 0.09 |
| polarC18 | 780.55 | 85.4977 | 12.4 | 0.09 |
| polarC18 | 780.55 | 128.8094 | 12.4 | 0.09 |
| polarC18 | 780.55 | 136.7607 | 12.4 | 0.09 |
| polarC18 | 780.55 | 226.4071 | 12.4 | 0.09 |
| polarC18 | 780.55 | 122.7539 | 12.4 | 0.09 |
| polarC18 | 780.55 | 162.1655 | 12.3 | 0.09 |
| polarC18 | 780.55 | 98.9587 | 12.3 | 0.09 |
| polarC18 | 780.55 | 203.3606 | 12.3 | 0.09 |
| polarC18 | 780.55 | 206.3304 | 12.3 | 0.09 |
| polarC18 | 780.55 | 84.5865 | 12.3 | 0.09 |
| polarC18 | 780.55 | 206.6083 | 12.3 | 0.09 |
| polarC18 | 780.55 | 168.409 | 12.3 | 0.09 |
| polarC18 | 780.55 | 94.7001 | 12.3 | 0.09 |
| polarC18 | 780.55 | 146.2905 | 12.3 | 0.09 |
| polarC18 | 780.55 | 279.8011 | 12.3 | 0.09 |
| polarC18 | 780.55 | 107.4731 | 12.3 | 0.09 |
| polarC18 | 780.55 | 253.7729 | 12.3 | 0.09 |
| polarC18 | 780.55 | 185.6747 | 12.3 | 0.09 |
| polarC18 | 780.55 | 385.3551 | 12.3 | 0.09 |
| polarC18 | 780.55 | 283.4539 | 12.3 | 0.09 |
| polarC18 | 780.55 | 100.4366 | 12.3 | 0.09 |
| polarC18 | 780.55 | 85.0934 | 12.3 | 0.09 |
| polarC18 | 780.55 | 184.889 | 12.2 | 0.09 |
| polarC18 | 780.55 | 120.1453 | 12.2 | 0.09 |
| polarC18 | 780.55 | 210.6565 | 12.2 | 0.09 |
| polarC18 | 780.55 | 293.5838 | 12.2 | 0.09 |
| polarC18 | 780.55 | 270.0463 | 12.2 | 0.09 |
| polarC18 | 780.55 | 262.3449 | 12.2 | 0.09 |
| polarC18 | 780.55 | 600.8965 | 12.2 | 0.09 |

| | | | | |
|---|---|---|---|---|
| polarC18 | 780.55 | 276.2055 | 12.2 | 0.09 |
| polarC18 | 780.55 | 208.1599 | 12.2 | 0.09 |
| polarC18 | 780.55 | 122.8576 | 12.2 | 0.09 |
| polarC18 | 780.55 | 95.2696 | 12.2 | 0.09 |
| polarC18 | 780.55 | 138.5874 | 12.2 | 0.09 |
| polarC18 | 780.55 | 128.3736 | 12.2 | 0.09 |
| polarC18 | 780.55 | 144.9117 | 12.2 | 0.09 |
| polarC18 | 780.55 | 314.412 | 12.2 | 0.09 |
| polarC18 | 780.55 | 95.6336 | 12.2 | 0.09 |
| polarC18 | 780.55 | 175.7865 | 12.1 | 0.09 |
| polarC18 | 780.55 | 452.2385 | 12.1 | 0.09 |
| polarC18 | 780.55 | 145.6733 | 12.1 | 0.09 |
| polarC18 | 780.55 | 257.9689 | 12.1 | 0.09 |
| polarC18 | 780.55 | 121.9987 | 12.1 | 0.09 |
| polarC18 | 780.55 | 266.3164 | 12.1 | 0.09 |
| polarC18 | 780.55 | 645.0868 | 12.1 | 0.09 |
| polarC18 | 780.55 | 568.8184 | 12.1 | 0.09 |
| polarC18 | 780.55 | 203.6109 | 12.1 | 0.09 |
| polarC18 | 780.55 | 126.7307 | 12.1 | 0.09 |
| polarC18 | 780.55 | 113.1946 | 12.1 | 0.09 |
| polarC18 | 780.55 | 298.4563 | 12.1 | 0.09 |
| polarC18 | 780.55 | 260.7103 | 12.1 | 0.09 |
| polarC18 | 780.55 | 190.0172 | 12.1 | 0.09 |
| polarC18 | 780.55 | 104.0783 | 12 | 0.09 |
| polarC18 | 780.55 | 495.2803 | 12 | 0.09 |
| polarC18 | 780.55 | 92.8141 | 12 | 0.09 |
| polarC18 | 780.55 | 112.4822 | 12 | 0.09 |
| polarC18 | 780.55 | 156.8879 | 12 | 0.09 |
| polarC18 | 780.55 | 217.7606 | 12 | 0.09 |
| polarC18 | 780.55 | 115.395 | 12 | 0.09 |
| polarC18 | 780.55 | 280.9492 | 12 | 0.09 |
| polarC18 | 780.55 | 124.4251 | 12 | 0.09 |
| polarC18 | 780.55 | 628.3821 | 12 | 0.09 |
| polarC18 | 780.55 | 168.4678 | 12 | 0.09 |
| polarC18 | 780.55 | 112.4465 | 12 | 0.09 |
| polarC18 | 780.55 | 214.3895 | 12 | 0.09 |
| polarC18 | 780.55 | 286.3022 | 12 | 0.09 |
| polarC18 | 780.55 | 102.1959 | 12 | 0.09 |
| polarC18 | 780.55 | 99.695 | 12 | 0.09 |
| polarC18 | 780.55 | 221.06 | 12 | 0.09 |
| polarC18 | 780.55 | 85.1799 | 12 | 0.09 |
| polarC18 | 780.55 | 140.1118 | 12 | 0.09 |

| | | | | |
|---|---|---|---|---|
| polarC18 | 780.55 | 735.3438 | 12 | 0.09 |
| polarC18 | 780.55 | 601.6035 | 12 | 0.09 |
| polarC18 | 780.55 | 201.2149 | 11.9 | 0.09 |
| polarC18 | 780.55 | 332.2272 | 11.9 | 0.09 |
| polarC18 | 780.55 | 155.4615 | 11.9 | 0.09 |
| polarC18 | 780.55 | 118.5672 | 11.9 | 0.09 |
| polarC18 | 780.55 | 89.1821 | 11.9 | 0.09 |
| polarC18 | 780.55 | 154.8414 | 11.9 | 0.09 |
| polarC18 | 780.55 | 378.6601 | 11.9 | 0.09 |
| polarC18 | 780.55 | 83.286 | 11.9 | 0.09 |
| polarC18 | 780.55 | 327.553 | 11.9 | 0.09 |
| polarC18 | 780.55 | 132.9415 | 11.9 | 0.09 |
| polarC18 | 780.55 | 114.8639 | 11.9 | 0.09 |
| polarC18 | 780.55 | 106.7518 | 11.9 | 0.09 |
| polarC18 | 780.55 | 85.2913 | 11.9 | 0.09 |
| polarC18 | 780.55 | 301.4253 | 11.9 | 0.09 |
| polarC18 | 780.55 | 88.6509 | 11.9 | 0.09 |
| polarC18 | 780.55 | 142.466 | 11.9 | 0.09 |
| polarC18 | 780.55 | 215.6803 | 11.9 | 0.09 |
| polarC18 | 780.55 | 336.9073 | 11.8 | 0.09 |
| polarC18 | 780.55 | 121.1169 | 11.8 | 0.09 |
| polarC18 | 780.55 | 247.9776 | 11.8 | 0.09 |
| polarC18 | 780.55 | 103.6778 | 11.8 | 0.09 |
| polarC18 | 780.55 | 113.7317 | 11.8 | 0.09 |
| polarC18 | 780.55 | 615.5135 | 11.8 | 0.09 |
| polarC18 | 780.55 | 566.6142 | 11.8 | 0.09 |
| polarC18 | 780.55 | 83.7594 | 11.8 | 0.09 |
| polarC18 | 780.55 | 135.5376 | 11.8 | 0.09 |
| polarC18 | 780.55 | 186.1185 | 11.8 | 0.09 |
| polarC18 | 780.55 | 94.513 | 11.8 | 0.09 |
| polarC18 | 780.55 | 792.6166 | 11.8 | 0.09 |
| polarC18 | 780.55 | 225.1131 | 11.8 | 0.09 |
| polarC18 | 780.55 | 112.9766 | 11.7 | 0.09 |
| polarC18 | 780.55 | 442.5175 | 11.7 | 0.09 |
| polarC18 | 780.55 | 81.7551 | 11.7 | 0.09 |
| polarC18 | 780.55 | 269.4286 | 11.7 | 0.09 |
| polarC18 | 780.55 | 174.944 | 11.7 | 0.09 |
| polarC18 | 780.55 | 108.516 | 11.7 | 0.09 |
| polarC18 | 780.55 | 84.1186 | 11.7 | 0.09 |
| polarC18 | 780.55 | 170.2997 | 11.7 | 0.09 |
| polarC18 | 780.55 | 209.737 | 11.7 | 0.09 |
| polarC18 | 780.55 | 420.5078 | 11.7 | 0.09 |

| | | | | |
|---|---|---|---|---|
| polarC18 | 780.55 | 213.942 | 11.7 | 0.09 |
| polarC18 | 780.55 | 109.8169 | 11.7 | 0.09 |
| polarC18 | 780.55 | 92.803 | 11.7 | 0.09 |
| polarC18 | 780.55 | 88.8266 | 11.7 | 0.09 |
| polarC18 | 780.55 | 96.9753 | 11.6 | 0.09 |
| polarC18 | 780.55 | 154.9786 | 11.6 | 0.09 |
| polarC18 | 780.55 | 103.2886 | 11.6 | 0.09 |
| polarC18 | 780.55 | 260.9585 | 11.6 | 0.09 |
| polarC18 | 780.55 | 352.692 | 11.6 | 0.09 |
| polarC18 | 780.55 | 548.5162 | 11.6 | 0.09 |
| polarC18 | 780.55 | 85.4161 | 11.6 | 0.09 |
| polarC18 | 780.55 | 255.3039 | 11.6 | 0.08 |
| polarC18 | 780.55 | 100.6573 | 11.6 | 0.08 |
| polarC18 | 780.55 | 163.1985 | 11.6 | 0.08 |
| polarC18 | 780.55 | 229.7019 | 11.6 | 0.08 |
| polarC18 | 780.55 | 645.2806 | 11.6 | 0.08 |
| polarC18 | 780.55 | 148.4411 | 11.6 | 0.08 |
| polarC18 | 780.55 | 138.4699 | 11.6 | 0.08 |
| polarC18 | 780.55 | 256.5278 | 11.6 | 0.08 |
| polarC18 | 780.55 | 425.3244 | 11.6 | 0.08 |
| polarC18 | 780.55 | 356.5056 | 11.6 | 0.08 |
| polarC18 | 780.55 | 91.3195 | 11.6 | 0.08 |
| polarC18 | 780.55 | 89.629 | 11.6 | 0.08 |
| polarC18 | 780.55 | 283.5863 | 11.5 | 0.08 |
| polarC18 | 780.55 | 272.3026 | 11.5 | 0.08 |
| polarC18 | 780.55 | 129.8069 | 11.5 | 0.08 |
| polarC18 | 780.55 | 257.8841 | 11.5 | 0.08 |
| polarC18 | 780.55 | 210.0921 | 11.5 | 0.08 |
| polarC18 | 780.55 | 87.4223 | 11.5 | 0.08 |
| polarC18 | 780.55 | 146.3321 | 11.5 | 0.08 |
| polarC18 | 780.55 | 200.1542 | 11.5 | 0.08 |
| polarC18 | 780.55 | 523.4284 | 11.5 | 0.08 |
| polarC18 | 780.55 | 97.2453 | 11.5 | 0.08 |
| polarC18 | 780.55 | 188.0587 | 11.5 | 0.08 |
| polarC18 | 780.55 | 133.5892 | 11.5 | 0.08 |
| polarC18 | 780.55 | 605.305 | 11.4 | 0.08 |
| polarC18 | 780.55 | 90.2912 | 11.4 | 0.08 |
| polarC18 | 780.55 | 201.0087 | 11.4 | 0.08 |
| polarC18 | 780.55 | 199.2855 | 11.4 | 0.08 |
| polarC18 | 780.55 | 82.9941 | 11.4 | 0.08 |
| polarC18 | 780.55 | 82.2278 | 11.4 | 0.08 |
| polarC18 | 780.55 | 598.1819 | 11.4 | 0.08 |

| | | | | |
|---|---|---|---|---|
| polarC18 | 780.55 | 157.9811 | 11.4 | 0.08 |
| polarC18 | 780.55 | 93.9766 | 11.4 | 0.08 |
| polarC18 | 780.55 | 116.0327 | 11.4 | 0.08 |
| polarC18 | 780.55 | 220.1242 | 11.4 | 0.08 |
| polarC18 | 780.55 | 90.518 | 11.4 | 0.08 |
| polarC18 | 780.55 | 266.8575 | 11.4 | 0.08 |
| polarC18 | 780.55 | 376.4653 | 11.3 | 0.08 |
| polarC18 | 780.55 | 96.2299 | 11.3 | 0.08 |
| polarC18 | 780.55 | 294.0204 | 11.3 | 0.08 |
| polarC18 | 780.55 | 197.9018 | 11.3 | 0.08 |
| polarC18 | 780.55 | 109.4337 | 11.3 | 0.08 |
| polarC18 | 780.55 | 443.636 | 11.2 | 0.08 |
| polarC18 | 780.55 | 175.3426 | 11.2 | 0.08 |
| polarC18 | 780.55 | 284.5349 | 11.2 | 0.08 |
| polarC18 | 780.55 | 98.9301 | 11.2 | 0.08 |
| polarC18 | 780.55 | 85 | 11.2 | 0.08 |
| polarC18 | 780.55 | 95.3096 | 11.2 | 0.08 |
| polarC18 | 780.55 | 294.9696 | 11.1 | 0.08 |
| polarC18 | 780.55 | 146.6281 | 11.1 | 0.08 |
| polarC18 | 780.55 | 152.6225 | 11.1 | 0.08 |
| polarC18 | 780.55 | 84.4076 | 11.1 | 0.08 |
| polarC18 | 780.55 | 244.6959 | 11.1 | 0.08 |
| polarC18 | 780.55 | 99.833 | 11 | 0.08 |
| polarC18 | 780.55 | 155.5338 | 11 | 0.08 |
| polarC18 | 780.55 | 189.087 | 11 | 0.08 |
| polarC18 | 780.55 | 119.5937 | 11 | 0.08 |
| polarC18 | 780.55 | 486.8364 | 11 | 0.08 |
| polarC18 | 780.55 | 82.9829 | 11 | 0.08 |
| polarC18 | 780.55 | 93.7723 | 11 | 0.08 |
| polarC18 | 780.55 | 83.2231 | 11 | 0.08 |
| polarC18 | 780.55 | 136.8082 | 11 | 0.08 |
| polarC18 | 780.55 | 100.3258 | 11 | 0.08 |
| polarC18 | 780.55 | 113.6354 | 10.9 | 0.08 |
| polarC18 | 780.55 | 87.73 | 10.9 | 0.08 |
| polarC18 | 780.55 | 143.5584 | 10.9 | 0.08 |
| polarC18 | 780.55 | 95.946 | 10.9 | 0.08 |
| polarC18 | 780.55 | 89.335 | 10.9 | 0.08 |
| polarC18 | 780.55 | 204.7489 | 10.9 | 0.08 |
| polarC18 | 780.55 | 99.6664 | 10.9 | 0.08 |
| polarC18 | 780.55 | 265.2097 | 10.9 | 0.08 |
| polarC18 | 780.55 | 197.1714 | 10.9 | 0.08 |
| polarC18 | 780.55 | 132.4366 | 10.9 | 0.08 |

| | | | | |
|---|---|---|---|---|
| polarC18 | 780.55 | 82.4877 | 10.8 | 0.08 |
| polarC18 | 780.55 | 341.9153 | 10.8 | 0.08 |
| polarC18 | 780.55 | 369.2902 | 10.8 | 0.08 |
| polarC18 | 780.55 | 119.0273 | 10.8 | 0.08 |
| polarC18 | 780.55 | 84.8009 | 10.7 | 0.08 |
| polarC18 | 780.55 | 95.5857 | 10.7 | 0.08 |
| polarC18 | 780.55 | 237.0628 | 10.7 | 0.08 |
| polarC18 | 780.55 | 83.5742 | 10.6 | 0.08 |
| polarC18 | 780.55 | 160.3658 | 10.6 | 0.08 |
| polarC18 | 780.55 | 109.898 | 10.6 | 0.08 |
| polarC18 | 780.55 | 164.3792 | 10.5 | 0.08 |
| polarC18 | 780.55 | 100.4865 | 10.5 | 0.08 |
| polarC18 | 780.55 | 319.507 | 10.5 | 0.08 |
| polarC18 | 780.55 | 86.1786 | 10.5 | 0.08 |
| polarC18 | 780.55 | 149.5371 | 10.5 | 0.08 |
| polarC18 | 780.55 | 491.2645 | 10.5 | 0.08 |
| polarC18 | 780.55 | 133.4655 | 10.4 | 0.08 |
| polarC18 | 780.55 | 86.8556 | 10.4 | 0.08 |
| polarC18 | 780.55 | 102.3688 | 10.3 | 0.08 |
| polarC18 | 780.55 | 116.3288 | 10.2 | 0.07 |
